# Characteristics that modify the effect of small-quantity lipid-based nutrient supplementation on child growth: an individual participant data meta-analysis of randomized controlled trials

**DOI:** 10.1101/2021.02.05.21251105

**Authors:** Kathryn G. Dewey, K. Ryan Wessells, Charles D. Arnold, Elizabeth L. Prado, Souheila Abbeddou, Seth Adu-Afarwuah, Hasmot Ali, Benjamin F. Arnold, Per Ashorn, Ulla Ashorn, Sania Ashraf, Elodie Becquey, Jaden Bendabenda, Kenneth H. Brown, Parul Christian, John M. Colford, Sherlie J. L. Dulience, Lia C.H. Fernald, Emanuela Galasso, Lotta Hallamaa, Sonja Y. Hess, Jean H. Humphrey, Lieven Huybregts, Lora L. Iannotti, Kaniz Jannat, Anna Lartey, Agnes Le Port, Jef L. Leroy, Stephen P. Luby, Kenneth Maleta, Susana L. Matias, Mduduzi NN Mbuya, Malay K. Mridha, Minyanga Nkhoma, Clair Null, Rina R. Paul, Harriet Okronipa, Jean-Bosco Ouédraogo, Amy J. Pickering, Andrew J. Prendergast, Marie Ruel, Saijuddin Shaikh, Ann M. Weber, Patricia Wolff, Amanda Zongrone, Christine P. Stewart

## Abstract

**Background:** Meta-analyses have demonstrated that small-quantity lipid-based nutrient supplements (SQ-LNS) reduce stunting and wasting prevalence among infants and young children. Identification of subgroups who benefit most from SQ-LNS may facilitate program design.

**Objective:** Our objective was to identify study-level and individual-level modifiers of the effect of SQ-LNS on child growth outcomes.

**Methods:** We conducted a two-stage meta-analysis of individual participant data from 14 randomized controlled trials of SQ-LNS provided to children 6 to 24 months of age in low- and middle-income countries (n=37,066). We generated study-specific and subgroup estimates of SQ-LNS vs. control and pooled the estimates using fixed-effects models, with random-effects models as sensitivity analyses. We used random effects meta-regression to examine study-level effect modifiers. Heterogeneity was assessed using I^2^ and Tau^2^ statistics. Sensitivity analyses were conducted to examine whether results differed depending on inclusion criteria for arms within trials and types of comparisons.

**Results:** SQ-LNS provision decreased stunting (length-for-age z-score < −2) by 12% (relative reduction), wasting (weight-for-length (WLZ) z-score < −2) by 14%, low mid-upper arm circumference (MUAC < 125 mm or MUACZ < −2) by 18%, acute malnutrition (WLZ < −2 or MUAC < 125 mm) by 14%, underweight (weight-for-age z-score < −2) by 13%, and small head size (head-circumference z-score < −2) by 9%. Effects of SQ-LNS on growth outcomes generally did not differ by study-level characteristics including region, stunting burden, malaria prevalence, sanitation, water quality, duration of supplementation, frequency of contact or average reported compliance with SQ-LNS. Effects of SQ-LNS on stunting, wasting, low MUAC and small head size were greater among girls than among boys; effects on stunting, underweight and low MUAC were greater among later-born (vs. first-born) children; and effects on wasting and acute malnutrition were greater among children in households with improved (vs. unimproved) sanitation. Results were similar across sensitivity analyses.

**Conclusions:** The positive impact of SQ-LNS on growth is apparent across a wide variety of study-level contexts. Policy-makers and program planners should consider including SQ-LNS in the mix of interventions to prevent both stunting and wasting. This study was registered at www.crd.york.ac.uk/PROSPERO as CRD42019146592.

## Introduction

Undernutrition, including stunting, wasting and micronutrient deficiencies, is prevalent among infants and young children in low- and middle-income countries and is associated with increased morbidity and mortality and delayed psychomotor and neurocognitive development (1). Among children under 5 years of age globally, 21.3% (144 million) were stunted and 6.9% (47 million) were wasted in 2019 (2). The etiology of stunting and wasting is complex and multi-factorial (3–8), which may explain the limited effectiveness of interventions that focus solely on improving nutrition in improving these outcomes (5, 9). Nonetheless, low quality diets that lack adequate amounts of key nutrients during the complementary feeding period from 6 to 24 months of age are recognized as a critical contributory factor (3). Increased dietary diversity, with foods from all of the key food groups, and selection of nutrient-rich complementary foods within each of those food groups, should be universally promoted (10, 11). However, even under the best of circumstances it is difficult to meet all nutrient needs during this age interval (12), and for low-income populations the cost of certain nutrient-rich foods is often prohibitive (13, 14). For this reason, various types of fortified products designed to fill nutrient gaps have been evaluated, including fortified blended foods and products used for home fortification such as micronutrient powders (MNP) and small-quantity lipid-based nutrient supplements (SQ-LNS) (15).

SQ-LNS were developed to provide multiple micronutrients embedded in a small amount of food that also provides energy, protein, and essential fatty acids (16). This combination of macro- and micro-nutrients addresses multiple potential nutritional deficiencies, including gaps in the key nutrients required for growth. Because SQ-LNS typically provide only ∼100-120 kcal/d (∼ 4 teaspoons) and can be mixed with other foods, they are considered a type of home fortification product (17), although they can also be consumed alone. Unlike medium-quantity (MQ) and large-quantity LNS, which are generally aimed at treatment of moderate and severe acute malnutrition (16), SQ-LNS were designed for the prevention of undernutrition, including stunting.

In a recent meta-analysis of LNS given during the period of complementary feeding (18), most of the included trials (13 out of 17) provided SQ-LNS in at least one arm; the other 4 trials provided MQ-LNS only. LNS significantly reduced the prevalence of moderate stunting (by 7%, relative reduction), severe stunting (by 15%), moderate wasting (by 17%) and moderate underweight (by 15%) compared to no intervention. Exploratory subgroup analysis suggested that MQ-LNS did not have a greater impact than SQ-LNS on these outcomes. The meta-analysis also suggested that LNS was more effective than fortified blended foods or MNP at improving child anthropometric outcomes. Although the meta-analysis included some analyses disaggregated by study characteristics (such as SQ-vs. MQ-LNS, duration of supplementation, and age at follow-up), analyses stratified by individual-level characteristics were not conducted.

Differences in study design and context and the characteristics of individuals may modify the effect of SQ-LNS on child growth and other outcomes. Identification of subgroups of infants and young children who experience greater benefits from SQ-LNS, or are more likely to respond to the intervention, may be useful in informing the development of public health programs and policies (9). To examine effect modification, we conducted an individual participant data (IPD) meta-analysis of randomized controlled trials of SQ-LNS provided to infants and young children 6 to 24 months of age. For this paper, our objectives were to generate pooled estimates of the effect of SQ-LNS on each growth outcome and identify study-level and individual-level modifiers of the effect of SQ-LNS on these outcomes. Two companion papers report results for other outcome domains, anemia and micronutrient status (Wessells et al.) and development (Prado et al.).

## Methods

The protocol for this IPD meta-analysis was registered as PROSPERO CRD42019146592 (https://www.crd.york.ac.uk/prospero) (19). The statistical analysis plan was posted to Open Science Framework (https://osf.io/ymsfu) prior to analysis (20), and the results are reported according to PRISMA-IPD guidelines (21). The analyses were approved by the institutional review board of the University of California Davis (1463609-1). All individual trial protocols were approved by the relevant institutional ethics committees.

### Inclusion and exclusion criteria for this IPD meta-analysis

We included randomized controlled trials of SQ-LNS provided to children age 6-24 months that met the following study-level inclusion criteria: 1) the trial was conducted in a low- or middle-income country (22); 2) SQ-LNS (< ∼125 kcal/d) was provided to the intervention group for at least 3 months between 6 and 24 months of age; 3) at least one trial group did not receive SQ-LNS or other type of child supplementation; 4) the trial reported at least one outcome of interest; and 5) the trial used an individual or cluster randomized design in which the same participants were measured at baseline (prior to child supplementation) and again after completion of the intervention (longitudinal follow-up), or different participants were measured at baseline and post-intervention (repeated cross-sectional data collection). Trials were excluded if: 1) only children with severe or moderate malnutrition were eligible to participate (i.e., LNS was used for treatment, not prevention of malnutrition); 2) the trial was conducted in a hospitalized population or among children with a pre-existing disease; or 3) SQ-LNS provision was combined with additional supplemental food or nutrients within a single arm (e.g. SQ-LNS + food rations vs. control), and there was no appropriate comparison group that would allow isolation of the SQ-LNS effect (e.g., food rations alone).

Trials in which there were multiple relevant SQ-LNS interventions (e.g., varying dosages or formulations of SQ-LNS in different arms), or that combined provision of SQ-LNS with other non-nutritional interventions (i.e., water, sanitation and hygiene (WASH)) were eligible for inclusion. In such trials, all arms that provided SQ-LNS were combined into one group, and all non-LNS arms were combined into a single comparator group for each trial (herein labeled “control”), excluding intervention arms that received non-LNS child supplementation (e.g., MNP, fortified-blended food). We also conducted a sensitivity analysis restricting the comparison to specified contrasts of intervention arms within multiple intervention trials (see below).

At the individual participant level, we included children if their age at baseline would have allowed them to receive at least 3 months of intervention (supplementation or control group components) between 6 and 24 months of age.

### Search methods and identification of studies

First, we identified studies cited in a recent systematic review and meta-analysis of child LNS (18). We then used the same search terms used by that systematic review to search 16 international and 9 regional databases (see **Supplemental Methods**) for additional studies published from July 1, 2018 until May 1, 2019. One author (KRW) reviewed the titles and abstracts of all studies included in the previous systematic review, as well as the additional studies identified by our database searches, to select all potentially relevant studies for full text review. These were screened based on the inclusion and exclusion criteria. In September 2019, KRW searched for additional publications from studies that met inclusion and exclusion criteria to determine if results for outcomes of interest had been published subsequent to the search.

### Data collection

We invited all principal investigators of eligible trials to participate in this IPD meta-analysis. We provided a data dictionary listing definitions of variables requested for pooled analysis. Those variables were provided to the IPD analyst (CDA) in de-identified individual participant datasets. The IPD analyst communicated with investigators to request any missing variables or other clarifications, as needed.

### IPD integrity

We conducted a complete-case intention to treat analysis (23). We checked data for completeness by evaluating whether the study sample sizes in our pooled dataset were the same as in study protocols and publications. We calculated length-for-age z-score (LAZ), weight-for-length z-score (WLZ), weight-for-age z-score (WAZ), mid-upper arm circumference z-score (MUACZ), and head circumference-for-age z-score (HCZ) using the 2006 WHO child growth standards and checked the values for acceptable standard deviations and to be within published WHO acceptable ranges (24). Biologically implausible values were flagged, as recommended by WHO, in the following way: LAZ <-6 or >6; WAZ <-6 or >5; WLZ<-5 or >5; HCZ<-5 or >5, MUACZ <-5 or >5. These were inspected for errors and either winsorized (25) or removed from analysis on an outcome by outcome basis. Such cleaning was necessary for less than 0.5% of participants, with a consistently low rate of implausibility across outcomes and studies. We also checked summary statistics, such as means and standard deviations, in our dataset against published values for each trial.

### Assessment of risk of bias in each study and quality of evidence across studies

Two independent reviewers (KRW and CDA) assessed risk of bias in each trial against the following criteria: random sequence generation and allocation concealment (selection bias), blinding of participants and personnel (performance bias), blinding of outcome assessment (detection bias), incomplete outcome data (attrition bias), selective reporting (reporting bias), and other sources of bias (26). Any discrepancies were resolved by discussion or consultation with the core working group, as needed. The same reviewers also assessed the quality of evidence for anthropometric outcomes across all trials based on the five GRADE criteria: risk of bias, inconsistency of effect, imprecision, indirectness, and publication bias (27).

### Specification of outcomes and effect measures

We pre-specified all anthropometric outcomes in the statistical analysis plan (20). Continuous outcomes included LAZ, WLZ, WAZ, MUACZ, and HCAZ. Binary outcomes were stunting (LAZ < −2 SD), wasting (WLZ < −2 SD), underweight (WAZ < −2 SD), small head size (HCAZ <-2 SD), low MUAC (MUACZ < −2 SD or MUAC < 125 mm); and acute malnutrition (WLZ < −2 SD or MUAC < 125 mm). For estimation of main effects, the primary outcomes were LAZ, WLZ, stunting and wasting.

The principal measure of effect for continuous outcomes was the mean difference between intervention and comparison groups at endline, defined as the principal post-intervention time point as reported for trials with infrequent child assessment or at an age closest to the end of the supplementation period for trials with monthly child assessment. The principal measure of effect for binary outcomes was the prevalence ratio (relative difference in proportions between groups) at endline. We also estimated prevalence differences (in absolute percentage points) but considered them as secondary assessments of binary outcomes because such estimates are less consistent than prevalence ratios (26).

The treatment and comparisons of interest were provision of children with SQ-LNS (< ∼125 kcal/d, with or without co-interventions), compared to provision of no intervention or an intervention without any type of LNS or other child supplement. Other types of interventions have been delivered with or without LNS, such as WASH interventions and child morbidity monitoring and treatment. In several trials, child LNS has been delivered to children whose mothers received maternal LNS during pregnancy and lactation. Given that maternal supplementation may have an additive effect when LNS is provided to both mothers and their children, we originally planned to include trial arms that provided both maternal and child LNS in a sensitivity analysis only (i.e., the all-trials analysis). However, to maximize study inclusion and participant sample size, we decided post-hoc that if the main effects did not differ between the child-LNS-only analysis and the all-trials analysis (including maternal plus child LNS arms) by more than 20% for continuous outcomes or by 0.05 for prevalence ratios, the results of the all-trials analyses would be presented as the principal findings. Three additional pre-specified sensitivity analyses were also conducted, as described below.

### Synthesis methods and exploration of variation in effects

We conducted 3 types of analyses to separately investigate: 1) full sample main effects of the intervention, 2) effect modification by study-level characteristics, and 3) effect modification by individual-level characteristics. We used a two-stage approach for all 3 sets of analyses. This approach is preferred when incorporating cluster-randomized trials, as it allows intra-cluster correlations to be study specific (28). In the first stage, we generated intervention effect estimates within each individual study according to its study design. For longitudinal study designs we controlled for baseline anthropometric status (for each outcome) when estimating the intervention effect to gain efficiency. To deal with outcome dependence in cluster-randomized trials, we used robust standard errors with randomization clusters as the independent unit. In the second stage, we pooled the first stage estimates using inverse-variance weighted fixed effects. We also conducted sensitivity analyses in which we pooled estimates using inverse variance weighted random effects (29, 30). If there were fewer than 3 comparisons to include in a pooled estimate then the pooled estimate was not generated (e.g., if fewer than 3 comparisons were represented within a study-level effect modification category).

1. Full sample main effects of the intervention: We first estimated the intervention effect for each study. We then pooled the first stage estimates to generate a pooled point estimate, 95% confidence interval, and corresponding p-value.
2. Effect modification by study-level characteristics: We identified potential study-level effect modifiers prior to receipt of data, and categorized individual studies based on the distribution of effect modifier values across all studies (Box 1). Study-level characteristics included variables reflecting context as well as aspects of study design. We used random effects meta-regression to test the association between each effect modifier and the intervention. In the first stage of analysis, we estimated the parameter corresponding to the intervention effect as described above. In the second stage, we used a bivariate random effects meta-regression to test the association between study intervention effect and study-level characteristics.
3. Effect modification by individual-level characteristics: We identified potential individual-level characteristics based on a comprehensive review of effect modifiers considered by individual trials (either listed a priori in statistical analysis plans or as published) and selected based on biological plausibility (Box 1). Individual-level effect modifiers included maternal, child and household characteristics. We estimated the parameter corresponding to the interaction term of the effect modifier and the intervention for each study (31), as follows. For categorical effect modifiers, we first recoded them to create binary variables if needed, and then determined the interaction between the intervention and the binary effect modifier. All continuous effect modifiers were transformed into binary variables for the analysis by modeling the relationship within each study using splines and then pooling the first stage estimates to generate a pooled, fitted line. We defined programmatically useful dichotomous cutoffs based on the pooled fitted spline results and relevant context. We then generated pooled intervention effect estimates within each category to determine how the intervention effect in one subgroup differed from the intervention effect in the specified reference subgroup.

Heterogeneity of effect estimates was assessed using I^2^ and Tau^2^ statistics, within strata when relevant (32). We used a p-value of < 0.05 for main effects and a p-diff or p-for-interaction <0.10 for effect modification by study-level or individual-level characteristics, respectively. Because growth outcomes are inter-related and the effect modification analyses are inherently exploratory, we did not adjust for multiple hypothesis testing (33).

#### Box 1.

##### Potential effect modifiers

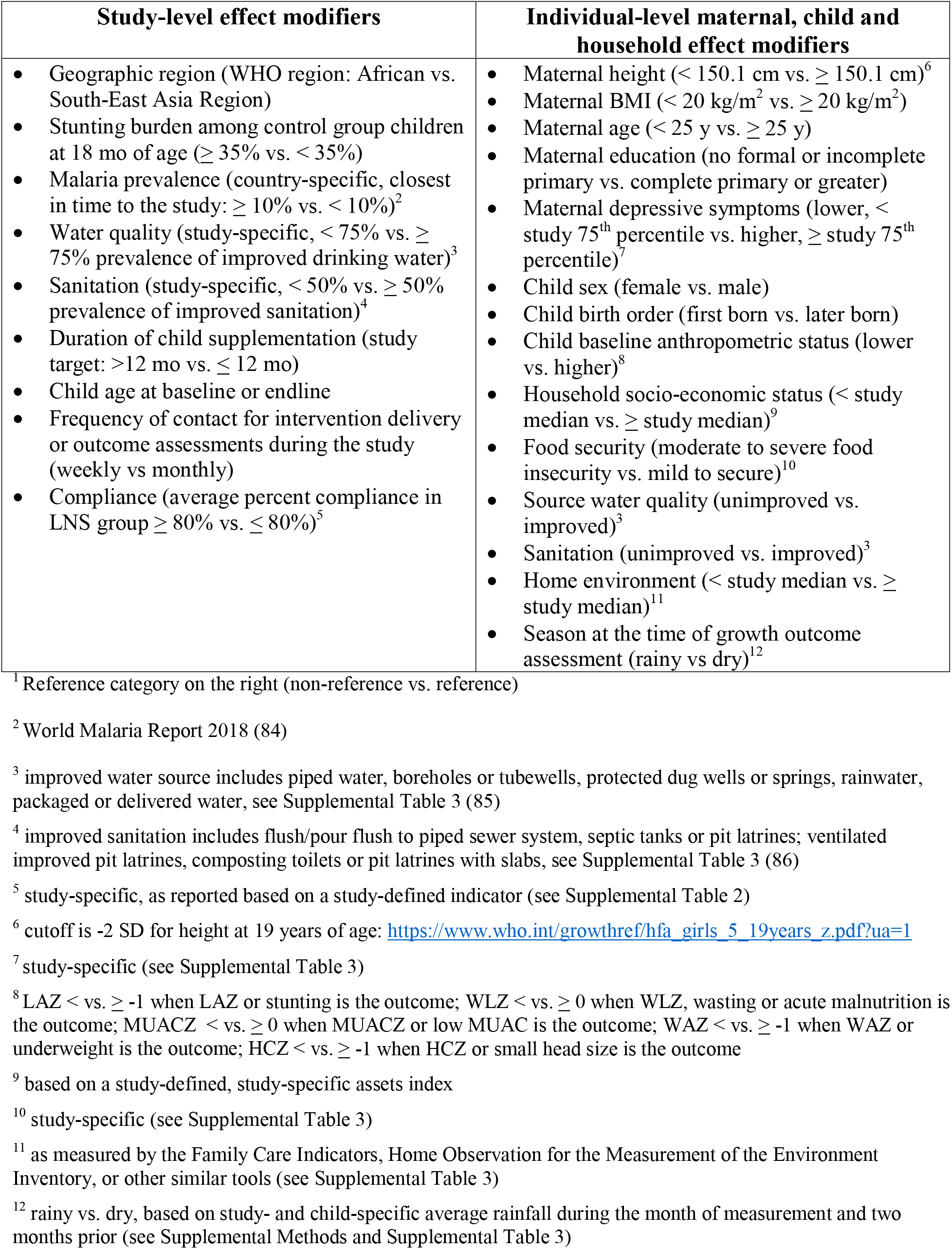

To aid in interpretation of individual-level effect modification, we evaluated the results for binary outcomes to identify what we will call the “cutoff effect”. The distribution of the continuous outcome relative to the cutoff for the corresponding binary outcome (e.g., distribution of LAZ around the −2 SD stunting cutoff) in the 2 effect modifier subgroups can influence the prevalence ratio and prevalence difference. When the mean in each of the 2 subgroups falls in a different location relative to the cutoff, the proportion of children close to the cutoff may be different between subgroups. This can lead to a greater reduction in the adverse binary outcome within one subgroup than within the other even if the shift in the mean value due to SQ-LNS is the same in both subgroups. To examine this, we simulated what would happen if we shifted the distribution of the non-reference effect modification subgroup to align with the reference subgroup (see Box 1), while maintaining the observed intervention effect mean difference in the continuous outcome within each subgroup. Based on ad-hoc, pragmatic criteria, if the p-for-interaction shifted from < 0.1 to > 0.2, we concluded that the cutoff effect explained the apparent effect modification; if it shifted from < 0.1 to > 0.1 but < 0.2, we concluded that the cutoff effect partially contributed to the apparent effect modification.

### Additional sensitivity analyses

Most trials have utilized similar SQ-LNS distribution mechanisms (e.g., weekly or monthly rations provided by study staff, community health workers, or other health extension agents), accompanied by messages to reinforce recommended infant and young child feeding (IYCF) practices. In addition, most trials have used similar formulations of SQ-LNS, specifically peanut and milk-based products providing approximately 1 RDA of most micronutrients (16). However, variations in trial design (e.g., integration of SQ-LNS supplementation with WASH interventions or enhanced morbidity monitoring and treatment; use of passive vs. active control arms) might influence the effect size estimates. In addition, some trials used several different formulations of SQ-LNS. We therefore conducted several pre-specified sensitivity analyses:

1. Separate comparisons within multi-component intervention trials, such that the SQ-LNS vs. no SQ-LNS comparisons were conducted separately between pairs of arms with the same non-nutrition components (e.g. SQ-LNS+WASH vs. WASH; SQ-LNS vs. Control). IYCF behavior change communication was not considered an additional component.
2. Exclusion of passive control arms, i.e., control group participants received no intervention and had no contact with project staff between baseline and endline.
3. Exclusion of intervention arms with SQ-LNS formulations that did not include both milk and peanut.

## Results

### Literature search and trial characteristics

We identified 15 trials that met our inclusion criteria, 14 of which provided individual participant data and are included in this analysis (**Figure 1**, **Table 1**) (34–48). Investigators for one trial were unable to participate (49). One trial was designed *a priori* to present results separately for HIV-exposed and HIV-unexposed children; therefore we present it herein as two separate comparisons in all analyses (47, 48). Similarly, the two PROMIS trials in Burkina Faso and Mali each included an independent longitudinal cohort and repeated (at baseline and endline) cross-sectional samples, so the longitudinal and cross-sectional results are presented as separate comparisons for each trial (38, 46).

**Figure 1:**
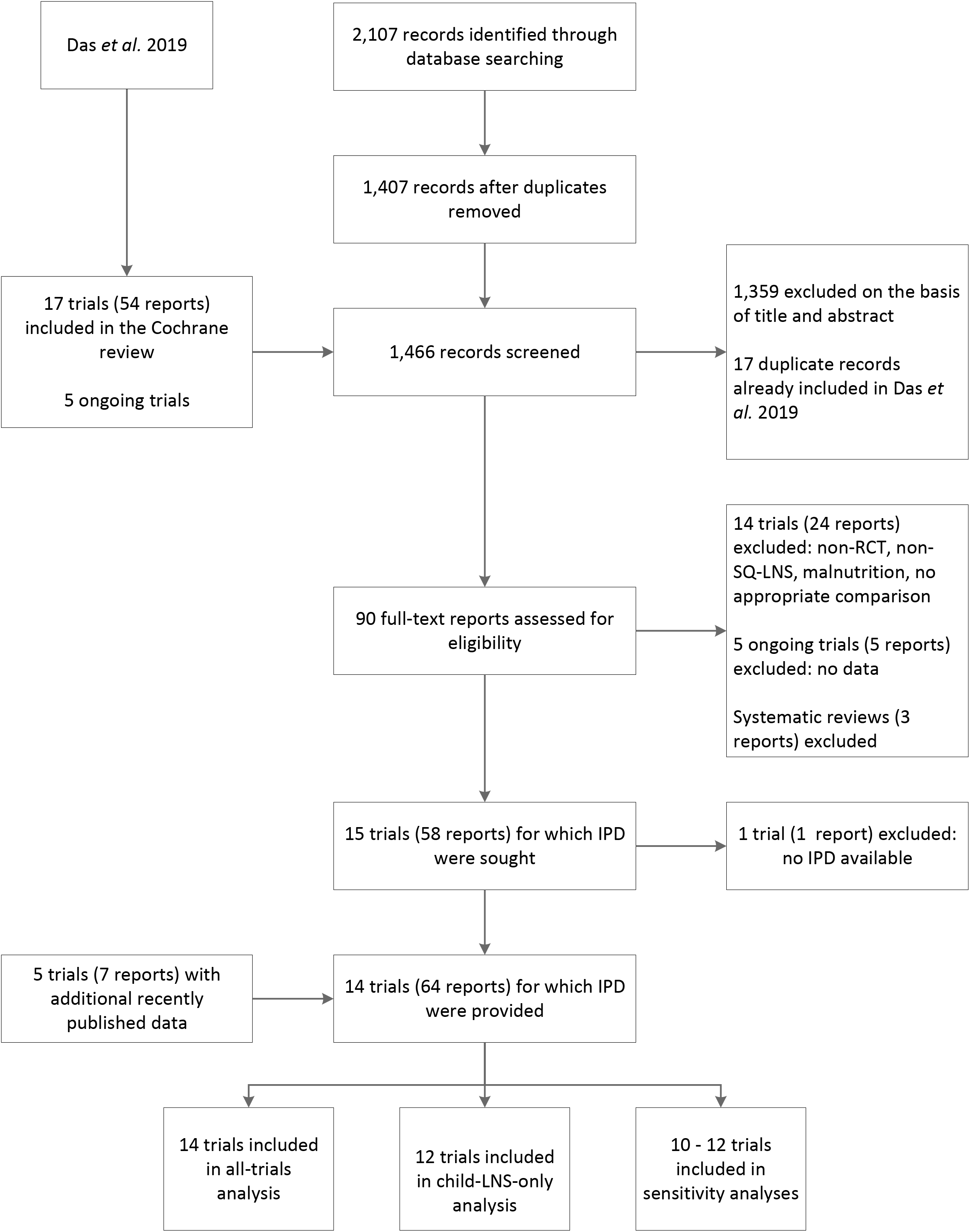
Study flow diagram.

**Table 1.** Characteristics of trials included in the individual participant data analysis and the analytic contrasts in which they were included.

| Country, years of study, study name, N, trial design, (author date) | Intervention Groups | Child SQ-LNS Supplementation |  | IYCF messages<sup>1</sup> | Analysis Contrasts |  |  |  |  |
| --- | --- | --- | --- | --- | --- | --- | --- | --- | --- |
|  |  | Age at start | Duration |  | All-trials analysis | Child-LNS only analysis | Separation of multi-component arms | Passive control arms excluded | Non-milk, non-peanut SQ-LNS arms excluded |
| Bangladesh, 2012-2014, Jivita-4, N=4218, cluster RCT, longitudinal follow-up (Christian 2015) (34) | Plumpy'Doz<sup>2</sup> + IYCF counseling | 6 mo | 12 mo | Expanded | LNS | LNS | LNS | LNS | LNS |
|  | Chickpea based LNS + IYCF counseling | 6 mo | 12 mo | Expanded | LNS | LNS | LNS | LNS | ---- |
|  | Rice-lentil LNS + IYCF counseling | 6 mo | 12 mo | Expanded | LNS | LNS | LNS | LNS | ---- |
|  | Wheat-soy blend (++) + IYCF counseling | 6 mo | 12 mo | Expanded | ---- | ---- | ---- | ---- | ---- |
|  | IYCF counseling only (Control) |  |  | Expanded | control | control | control | control | control |
| Bangladesh, 2011-2015, RDNS, N=2478, cluster RCT; longitudinal follow-up (Dewey 2017) (35) | LNS-LNS: maternal SQ-LNS in pregnancy + 6 mo postpartum, child SQ-LNS 6-24 mo | 6 mo | 18 mo | Minimal | LNS | ---- | ---- | ---- | ---- |
|  | IFA-LNS: maternal IFA in pregnancy + 3 mo postpartum, child SQ-LNS 6-24 mo | 6 mo | 18 mo | Minimal | LNS | LNS | LNS | LNS | LNS |
|  | IFA-MNP: maternal IFA in pregnancy + 3 mo postpartum, child MNP 6-24 mo |  |  | Minimal | ---- | ---- | ---- | ---- | ---- |
|  | IFA-Control: maternal IFA in pregnancy + 3 mo postpartum, no child supplementation |  |  | Minimal | control | control | control | control | control |
| Bangladesh, 2012-2015, WASH-Benefits, N=4633, cluster RCT, cross-sectional surveys (Luby 2018) (36) | Nutrition: SQ-LNS + IYCF counseling | 6 mo | 18 mo | Expanded | LNS | LNS | LNS | LNS | LNS |
|  | Water: family received chlorine for drinking water |  |  |  | control | control | ---- | control | control |
|  | Sanitation: family received upgraded latrine, sani-scoop and child potty |  |  |  | control | control | ---- | control | control |
|  | Handwashing: family received handwashing stations with soap |  |  |  | control | control | ---- | control | control |
|  | WASH: family received all water, sanitation and hygiene interventions |  |  |  | control | control | control-WSH | control | control |
|  | WASH + nutrition: all water, sanitation, hygiene and nutrition interventions | 6 mo | 18 mo | Expanded | LNS | LNS | LNS-WSH | LNS | LNS |
|  | Passive control (no intervention) |  |  |  | control | control | control | ---- | control |
| Burkina Faso, 2010-2012, iLINS-ZINC, N=2626, cluster RCT, longitudinal follow-up (Hess 2015) (37) | LNS-Zn0: SQ-LNS containing 0 mg/d Zn and placebo tablet<sup>3</sup> | 9 mo | 9 mo | Minimal | LNS | LNS | LNS | ---- | LNS |
|  | LNS-Zn5: SQ-LNS containing 5 mg/d Zn and placebo tablet | 9 mo | 9 mo | Minimal | LNS | LNS | LNS | ---- | LNS |
|  | LNS-Zn10: SQ-LNS containing 10 mg/d Zn and placebo tablet | 9 mo | 9 mo | Minimal | LNS | LNS | LNS | ---- | LNS |
|  | LNS-TabZn5: SQ-LNS containing 0 mg/d Zn and Zn tablet containing 5 mg/d Zn | 9 mo | 9 mo | Minimal | LNS | LNS | LNS | ---- | LNS |
|  | Passive control (no intervention) |  |  |  | control | control | control | ---- | control |
| Burkina Faso,<br>2015-2017,<br>PROMIS, N=2651,<br>cluster RCT, longitudinal<br>follow-up and cross-sectional<br>surveys<sup>a</sup> (Becquey 2019) (38) | SQ-LNS + IYCF counseling | 6 mo | 12 mo | Expanded | LNS | LNS | LNS | LNS | LNS |
|  | Active control (standard of care) |  |  |  | control | control | control | control | control |
| Ghana,<br>2004-2005,<br>N=194,<br>RCT, longitudinal follow-up<br>(Adu-Afarwuah 2007) (39) | SQ-LNS | 6 mo | 6 mo | Minimal | LNS | LNS | LNS | ---- | LNS |
|  | MNP |  |  | Minimal | ---- | ---- | ---- | ---- | ---- |
|  | Nutritabs (MMN) |  |  | Minimal | ---- | ---- | ---- | ---- | ---- |
|  | Passive control (no intervention) |  |  | Minimal | control | control | control | ---- | control |
| Ghana,<br>2009-2014,<br>iLINS-DYAD-G, N=1040,<br>RCT; longitudinal follow-up<br>(Adu-Afarwuah 2016) (40) | LNS: maternal SQ-LNS in pregnancy + 6 mo<br>postpartum, child SQ-LNS 6-18 mo | 6 mo | 12 mo | Minimal | LNS | ---- | ---- | ---- | ---- |
|  | MMN: maternal MMN in pregnancy + 6 mo<br>postpartum, no child supplementation |  |  | Minimal | control | ---- | ---- | ---- | ---- |
|  | IFA: maternal IFA in pregnancy and placebo for 6<br>mo postpartum, no child supplementation |  |  | Minimal | control | ---- | ---- | ---- | ---- |
| Haiti,<br>2011-2012,<br>N=300,<br>RCT; longitudinal follow-up<br>(Iannotti 2014) (41) | SQ-LNS for 6 mo | 6-11 mo | 6 mo<sup>5</sup> | Minimal | LNS | LNS | LNS | LNS | LNS |
|  | Active control (standard of care) |  |  | Minimal | control | control | control | control | control |
| Kenya,<br>2012-2016,<br>WASH-Benefits, N=6649,<br>cluster RCT, cross-sectional<br>surveys (Null 2018) (42) | Nutrition: SQ-LNS + IYCF counseling | 6 mo | 18 mo | Expanded | LNS | LNS | LNS | LNS | LNS |
|  | Water: family received chlorine for drinking water |  |  |  | control | control | ---- | control | control |
|  | Sanitation: family received upgraded latrine, sani-scoop and child potty |  |  |  | control | control | ---- | control | control |
|  | Handwashing: family received handwashing<br>stations with soap |  |  |  | control | control | ---- | control | control |
|  | WASH: family received all water, sanitation and<br>hygiene interventions |  |  |  | control | control | control-WSH | control | control |
|  | WASH + nutrition: all water, sanitation, hygiene<br>and nutrition interventions | 6 mo | 18 mo | Expanded | LNS | LNS | LNS-WSH | LNS | LNS |
|  | Passive control (no intervention) |  |  |  | control | control | control | ---- | control |
|  | Active control (visits to measure MUAC) |  |  |  | control | control | control | control | control |
| Madagascar,<br>2014-2016,<br>MAHAY, N=3390,<br>cluster RCT, longitudinal<br>follow-up (Galasso 2019) (43) | T4: early child stimulation + IYCF counseling |  |  | Expanded | ---- | ---- | ---- | ---- | ---- |
|  | T3: maternal SQ-LNS in pregnancy + 6 mo<br>postpartum, child SQ-LNS 6-18 mo + IYCF<br>counseling | 6-11 mo | 6-12 mo | Expanded | LNS | ---- | ---- | ---- | ---- |
|  | T2: child SQ-LNS 6-18 mo + IYCF counseling | 6-11 mo | 6-12 mo | Expanded | LNS | LNS | LNS | LNS | LNS |
|  | T1: IYCF counseling |  |  | Expanded | control | control | control | control | control |
|  | T0: Control (standard of care) |  |  |  | control | control | control | control | control |
| Malawi,<br>2011-2014,<br>iLINS-DYAD-M, N=664,<br>RCT, longitudinal follow-up<br>(Ashorn 2015) (44) | LNS: maternal SQ-LNS in pregnancy + 6 mo postpartum, child SQ-LNS 6-18 mo | 6 mo | 12 mo | Minimal | LNS | ----- | ----- | ----- | ----- |
|  | MMN: maternal MMN in pregnancy + 6 mo postpartum, no child supplementation |  |  | Minimal | control |  | ----- | ----- | ----- |
|  | IFA: maternal IFA in pregnancy and placebo for 6 mo postpartum, no child supplementation |  |  | Minimal | control |  | ----- | ----- | ----- |
| Malawi,<br>2009-2012,<br>iLINS-DOSE, N=943,<br>RCT, longitudinal follow-up<br>(Maleta 2015<sup>6</sup>) (45) | SQ-LNS containing milk (10 g/d) | 6 mo | 12 mo | Minimal | LNS | LNS | LNS | LNS | LNS |
|  | SQ-LNS containing milk (20 g/d) | 6 mo | 12 mo | Minimal | LNS | LNS | LNS | LNS | LNS |
|  | SQ-LNS without milk (20 g/d) | 6 mo | 12 mo | Minimal | LNS | LNS | LNS | LNS | ----- |
|  | MQ-LNS containing milk (40 g/d) |  |  | Minimal | ----- | ----- | ----- | ----- | ----- |
|  | MQ-LNS without milk (40 g/d) |  |  | Minimal | ----- | ----- | ----- | ----- | ----- |
|  | Active control |  |  | Minimal | control | control | control | control | control |
| Mali,<br>2015-2017,<br>PROMIS, N=2937,<br>cluster RCT, longitudinal<br>follow-up and cross-sectional<br>surveys<sup>4</sup> (Huybregts 2019) (46) | SQ-LNS + IYCF counseling | 6 mo | 18 mo | Expanded | LNS | LNS | LNS | LNS | LNS |
|  | Active control (standard of care) + IYCF counseling |  |  |  | control | control | control | control | control |
| Zimbabwe,<br>2013-2017,<br>SHINE<sup>7</sup>, N=4343,<br>cluster RCT, longitudinal<br>follow-up (Humphrey 2019;<br>Prendergast 2019) (47, 48) | IYCF: child SQ-LNS + IYCF counseling | 6 mo | 12 mo | Expanded | LNS | LNS | LNS | LNS | LNS |
|  | WASH : family received ventilated improved pit latrine, handwashing stations, soap, chlorine, child play space |  |  |  | control | control | control-WSH | control | control |
|  | WASH and IYCF: child SQ-LNS + IYCF counseling, family received ventilated improved pit latrine, handwashing stations, soap, chlorine, child play space | 6 mo | 12 mo | Expanded | LNS | LNS | LNS-WSH | LNS | LNS |
|  | Active control (standard of care) |  |  |  | control | control | control | control | control |

IFA, iron-folic acid; IYCF, infant and young child feeding; LNS, lipid-based nutrient supplement; MMN, multiple micronutrients; MNP, multiple micronutrient powder; MQ-LNS, medium-quantity lipid-based nutrient supplements; ORS, oral rehydration solution; SQ-LNS, small-quantity lipid-based nutrient supplements; WASH, water sanitation and hygiene; WSB, wheat soy blend; RCT, randomized controlled trial.
<sup>1</sup> Minimal IYCF messages defined as providing minimal counseling on IYCF other than reinforcing the normal IYCF messages already promoted in that setting. Expanded IYCF messages defined as providing expanded counseling on IYCF that went beyond the usual messaging.
<sup>2</sup> All supplements were isocaloric, children 6-12 mo received 125 kcal/d, children 12-18 mo received 250 kcal/d
<sup>3</sup> All children in the four intervention groups received ORS for diarrhea and treatment for malaria
<sup>4</sup> Cross-sectional and longitudinal cohorts within this trial are considered as separate comparisons in all analyses and the presentation of results
<sup>5</sup> Trial also included a 3 mo duration intervention arm which is excluded from these analyses as there is no comparable control arm available
<sup>6</sup> Trial is cited as Kumwenda 2014 in Das et al. (Cochrane Database of Systematic Reviews 2019)
<sup>7</sup> Trial was designed *a priori* to present results separately for HIV exposed and un-exposed children; thus considered as two comparisons in all analyses and the presentation of results

The 14 trials in these analyses were conducted in Sub-Saharan Africa (10 trials in 7 countries), Bangladesh (3 trials), and Haiti (1 trial), and included a total of 37,066 infants and young children with anthropometric data. The majority of trials began child supplementation with SQ-LNS at 6 months of age and the intended duration ranged from 6 to 18 months of supplementation; four trials included intervention arms that also provided SQ-LNS to mothers during pregnancy and the first 6 months postpartum (35, 40, 43, 44). All trials provided a peanut- and milk-based SQ-LNS in at least one of the arms (Table 1 and **Supplemental Table 1**). Generally, this provided approximately 120 kcal/d and ∼1 RDA of most micronutrients (19 micronutrients in 3 trials, 22 micronutrients in 11 trials); in one trial the ration was ∼120 kcal/d between 6 and 12 months of age and ∼250 kcal/d between 12 and 24 months of age (34). Two trials included additional arms with different formulations or doses of SQ-LNS (34, 45). Six trials were conducted within existing community-based or clinic-based programs (35, 38, 41, 43, 46–48); in the other trials, all activities were conducted by research teams. Seven trials provided minimal social and behavior change communication (SBCC) on IYCF other than reinforcing the normal IYCF messages already promoted in that setting (35, 37, 39–41, 44, 45), and 7 trials provided expanded SBCC on IYCF that went beyond the usual messaging, either in just the SQ-LNS intervention arms (36, 38, 42, 47–48) or in all arms including the non-SQ-LNS control arm (34, 43, 46). Three trials included arms with WASH interventions (36, 42, 47, 48). Most trials included an active control arm (i.e., similar contact frequency as for intervention arms) but 3 included only a passive control arm (36, 37, 39).

Descriptive information on the potential study-level and individual-level effect modifiers (defined in Box 1), by trial, is presented in **Supplemental Tables 2** and **3**, respectively. At the study level, 8 of the 14 study sites had a high burden of stunting (≥ 35% in the control group at 18 mo). Country-level malaria prevalence ranged from <1% in Bangladesh and Haiti to 59% in Burkina Faso. Study-specific prevalence of improved water quality ranged from 27% to 100%, and prevalence of improved sanitation ranged from 0% to 97%. Frequency of contact during the study was weekly in 7 trials and monthly in 7 trials. Average estimated reported compliance with SQ-LNS consumption was categorized as high (≥80%) in 7 trials and ranged between 37% and 77% in the other trials. The following maternal characteristics varied widely across trials: short stature (< 150.1 cm) ranged from <2% in Burkina Faso (37) to >45% in Bangladesh (35, 36); body mass index (BMI) < 20 kg/m^2^ ranged from 9% in Ghana (39) to 55% in Bangladesh (35); age < 25 y ranged from 24% among HIV-positive women in Zimbabwe (48) to 73% in Bangladesh (35); completion of primary education ranged from 3.8% in Burkina Faso (37) to 96.3% in Zimbabwe (47); and reported moderate to severe household food insecurity ranged from 10.3% in Kenya (42) to 73.5% in Malawi (45).

Growth outcomes in the control groups at endline showed that the burden of child malnutrition varied across studies (**Supplemental Table 4).** Prevalence of stunting ranged from 7.3% in the first Ghana trial (39) to 58.5% in Madagascar (43), and prevalence of wasting ranged from <2% (in the Haiti, and WASH-Benefits Kenya trials) (41, 42) to 16.4% in Bangladesh (34–36). The range in prevalence for the other binary outcomes was 1.5-18.3% for low MUAC, 3.2-21.1% for acute malnutrition, 4.7-39.2% for underweight, and 4.3-42.9% for small head size. High WLZ (> 1) was uncommon, with a prevalence > 10% in only 5 trials.

In general, we considered the trials to have a low risk of bias, with the exception of lack of blinding of participants owing to the nature of the intervention (**Supplemental Table 5** and **Supplemental Figure 1**).

### Main effects of SQ-LNS on growth outcomes

Results from the child-LNS-only and all-trials analyses were similar for all outcomes (**Supplemental Figures 2A, B and C**). Therefore, results from the all-trials analyses, inclusive of maternal + child LNS trials/arms, are presented below, and in **Table 2**. For LAZ, WLZ, WAZ, stunting and wasting, all 14 trials (17 comparisons) were represented. Some trials did not measure MUAC or head circumference, so the number of trials (comparisons) was 11 (14) for MUACZ, low MUAC and acute malnutrition and 10 (11) for HCZ and small head size. SQ-LNS had a significant positive effect on all growth outcomes, both continuous and binary. Among the continuous outcomes, the mean difference between intervention groups was largest for LAZ (+0.14) and smallest for WLZ (+0.08). SQ-LNS reduced the prevalence of adverse growth outcomes by 12% (5 percentage points) for stunting, 14% for wasting and acute malnutrition (1 percentage point for each), 18% for low MUAC (1 percentage point), 13% for underweight (3 percentage points), and 9% for small head size (1 percentage point). We rated the quality of the evidence for all outcomes as high based on the GRADE criteria listed above in Methods: at least 10 randomized controlled trials were available for all outcomes, risk of bias was low, heterogeneity was generally low to moderate (Table 2), precision was rated as high because all but 2 trials had sample sizes > 600, all trials were directly aimed at evaluating SQ-LNS, and funnel plots revealed no indication of publication bias (27).

**Table 2.** Main effects of SQ-LNS on growth outcomes.

| **Continuous outcomes** | **Number of participants (comparisons)** | **Mean difference: LNS - Control (95% CI)** | **P value** | **Heterogeneity I<sup>2</sup> (p-for-heterogeneity)<sup>1</sup>** | **Grade** |
| --- | --- | --- | --- | --- | --- |
| Length-for-age Z score (LAZ)<sup>2</sup> | 36795 (17) | 0.14 (0.11, 0.16) | <0.001 | 0.65 (<0.001) | High |
| Weight-for-length Z score (WLZ)<sup>2</sup> | 36608 (17) | 0.08 (0.06, 0.10) | <0.001 | 0.51 (0.008) | High |
| MUAC-for-age Z score (MUACZ) | 31774 (14) | 0.09 (0.06, 0.11) | <0.001 | 0.62 (0.001) | High |
| Weight-for-age Z score (WAZ) | 36787 (17) | 0.13 (0.11, 0.15) | <0.001 | 0.66 (<0.001) | High |
| Head-circumference-for-age Z-score (HCZ) | 27650 (11) | 0.09 (0.06, 0.11) | <0.001 | 0.46 (0.045) | High |
| **Binary outcomes** | **Number of participants (comparisons)** | **Prevalence ratio or difference: LNS vs Control (95% CI)** | **P value** | **Heterogeneity I<sup>2</sup> (p-for-heterogeneity)<sup>1</sup>** | **Grade** |
| Stunting (LAZ < -2 SD)<sup>2</sup> | 36795 (17) | 0.88 (0.85, 0.91) | <0.001 | 0.49 (0.013) | High |
|  |  | -5.0 (-4.1, -5.9) | <0.001 | 0.54 (0.005) | High |
| Wasting (WLZ < -2 SD)<sup>2</sup> | 36311 (16) | 0.86 (0.80, 0.93) | <0.001 | 0.00 (0.872) | High |
|  |  | -0.6 (-0.1, -1.0) | 0.010 | 0.12 (0.321) | High |
| Low MUAC (MUACZ < -2 SD or MUAC < 125 mm) | 31774 (14) | 0.82 (0.75, 0.89) | <0.001 | 0.16 (0.281) | High |
|  |  | -0.9 (-0.5, -1.4) | <0.001 | 0.50 (0.018) | High |
| Acute malnutrition (WLZ < -2 or MUAC < 125 mm) | 31440 (14) | 0.86 (0.80, 0.93) | <0.001 | 0.00 (0.554) | High |
|  |  | -1.1 (-0.5, -1.6) | <0.001 | 0.21 (0.228) | High |
| Underweight (WAZ < -2 SD) | 36787 (17) | 0.87 (0.83, 0.91) | <0.001 | 0.42 (0.031) | High |
|  |  | -3.1 (-2.3, -3.8) | <0.001 | 0.60 (0.001) | High |
| Small head size (HCZ < -2 SD) | 27456 (10) | 0.91 (0.86, 0.95) | <0.001 | 0.00 (0.816) | High |
|  |  | -1.2 (-0.4, -1.9) | 0.002 | 0.23 (0.230) | High |

<sup>1</sup>I<sup>2</sup> describes the percentage of variability in effect estimates that may be due to heterogeneity rather than chance. Roughly, 0.3 to 0.6 may be considered moderate heterogeneity. P-value from chi-squared test for heterogeneity. P < 0.05 indicates statistically significant evidence of heterogeneity of intervention effects beyond chance.
<sup>2</sup>Primary outcomes

Forest plots for stunting and wasting prevalence ratios are shown in **Figures 2** and **3**. Forest plots for all other outcomes are presented as supplemental figures (**Supplemental Figures 3A to 3Q**). Figure 2 illustrates that there was moderate heterogeneity (I^2^ = 0.49) in the effect on stunting prevalence across trials. For wasting, the I^2^ value was 0 (Figure 3), but this is attributable to relatively wide confidence intervals for all of the point estimates rather than low variability in the prevalence ratios. For all outcomes, fixed-effects and random-effects models generated nearly identical estimates.

**Figure 2:**
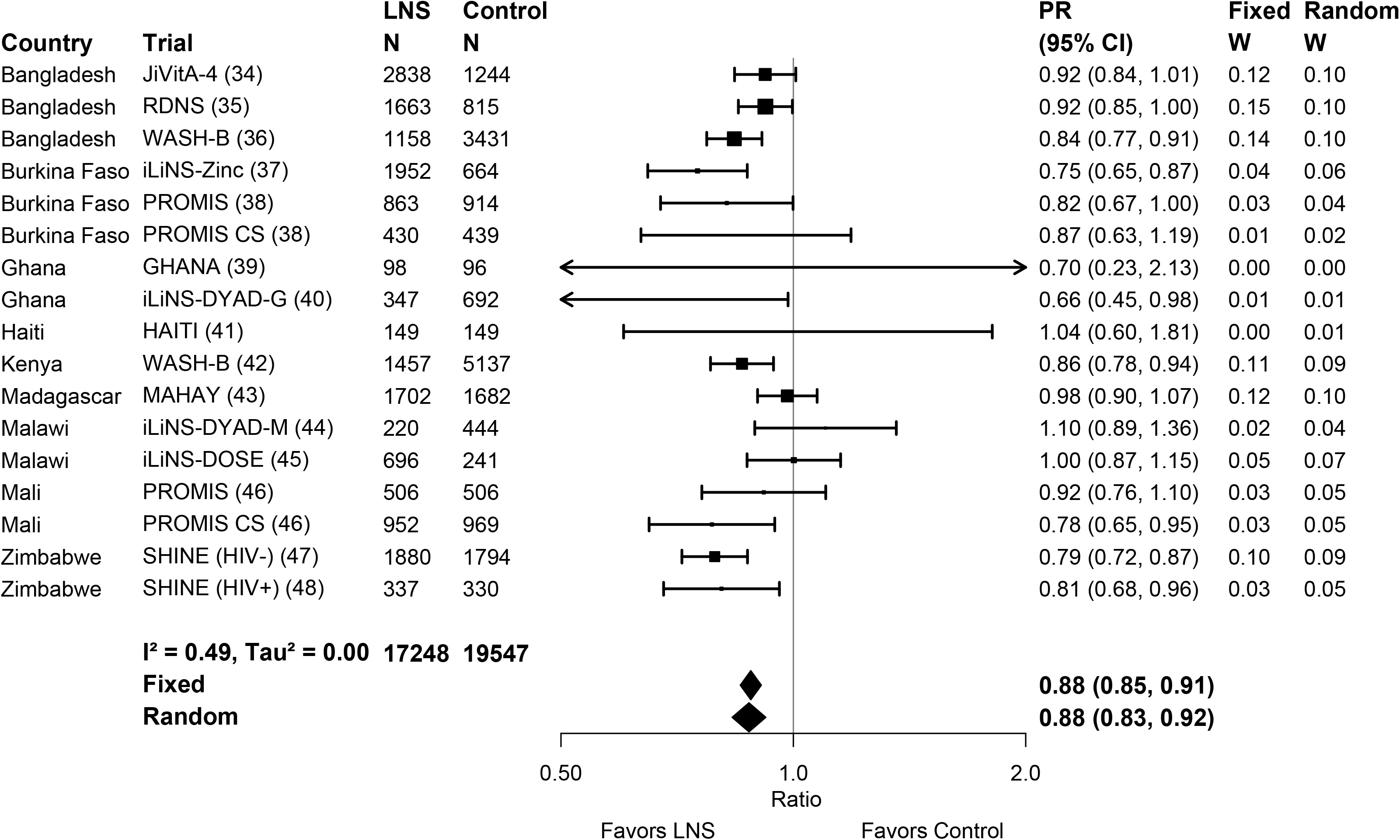
Main effect on stunting. Forest plot of effect of SQ-LNS on stunting prevalence. PR, prevalence ratio.

**Figure 3:**
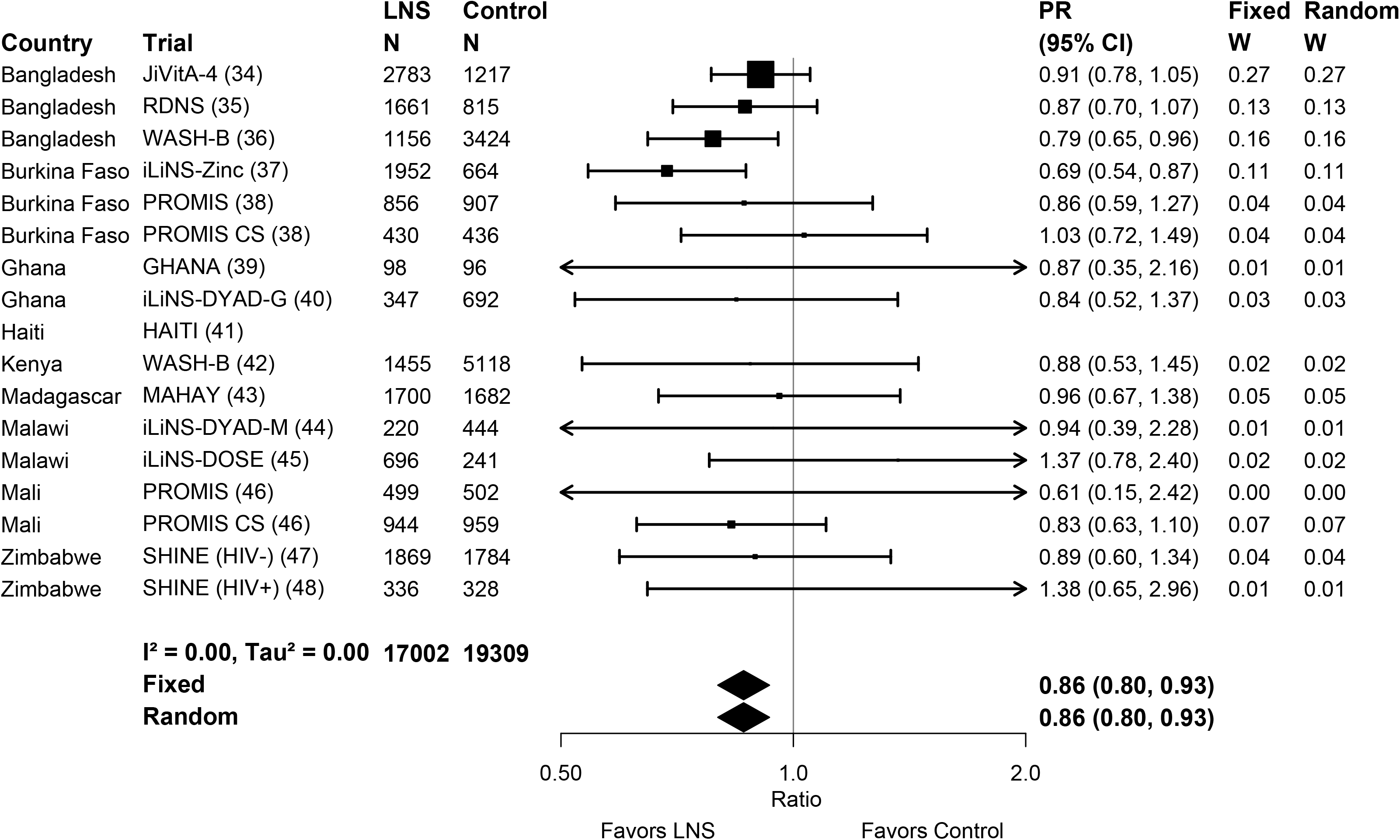
Main effect on wasting. Forest plot of effect of SQ-LNS on wasting prevalence. PR, prevalence ratio.

Results were similar in the sensitivity analyses (**Supplemental Figures 2A, B and C**) that included a) separation of comparisons within multi-component intervention trials, such that the SQ-LNS vs. no SQ-LNS comparisons were conducted separately between pairs of arms with the same non-nutrition components, b) exclusion of passive control arms, or c) exclusion of intervention arms with SQ-LNS formulations that were not milk- and peanut-based. For example, the prevalence ratios for stunting were all 0.87-0.88 and those for wasting were 0.85-0.89.

### Effect modification by study-level characteristics

Forest plots for all outcomes stratified by study-level effect modifiers are presented in **Supplemental Figures 4A to 4Q**. For some outcomes, we were unable to generate pooled estimates for effect modification by certain potential study-level effect modifiers because fewer than 3 comparisons were categorized into one of the study-level effect modification categories (e.g., acute malnutrition and low MUAC by region). Effect modification results were consistent across all sensitivity analyses (data not shown; available upon request); the results presented below refer to the all-trials analysis.

#### LAZ and stunting

None of the study-level characteristics significantly modified the effect of SQ-LNS on mean LAZ (Supplemental Figure 4A) or stunting prevalence, whether expressed as a prevalence ratio (Supplemental Figure 4B) or as a prevalence difference (Supplemental Figure 4C). The upper bound of the 95% confidence interval for the prevalence ratios was ≤ 1 in all categories (**Figure 4**), indicating that there were significant reductions in stunting prevalence among children receiving SQ-LNS regardless of region, stunting burden, malaria prevalence, water quality, sanitation, duration of supplementation, frequency of contact, or average reported compliance.

**Figure 4:**
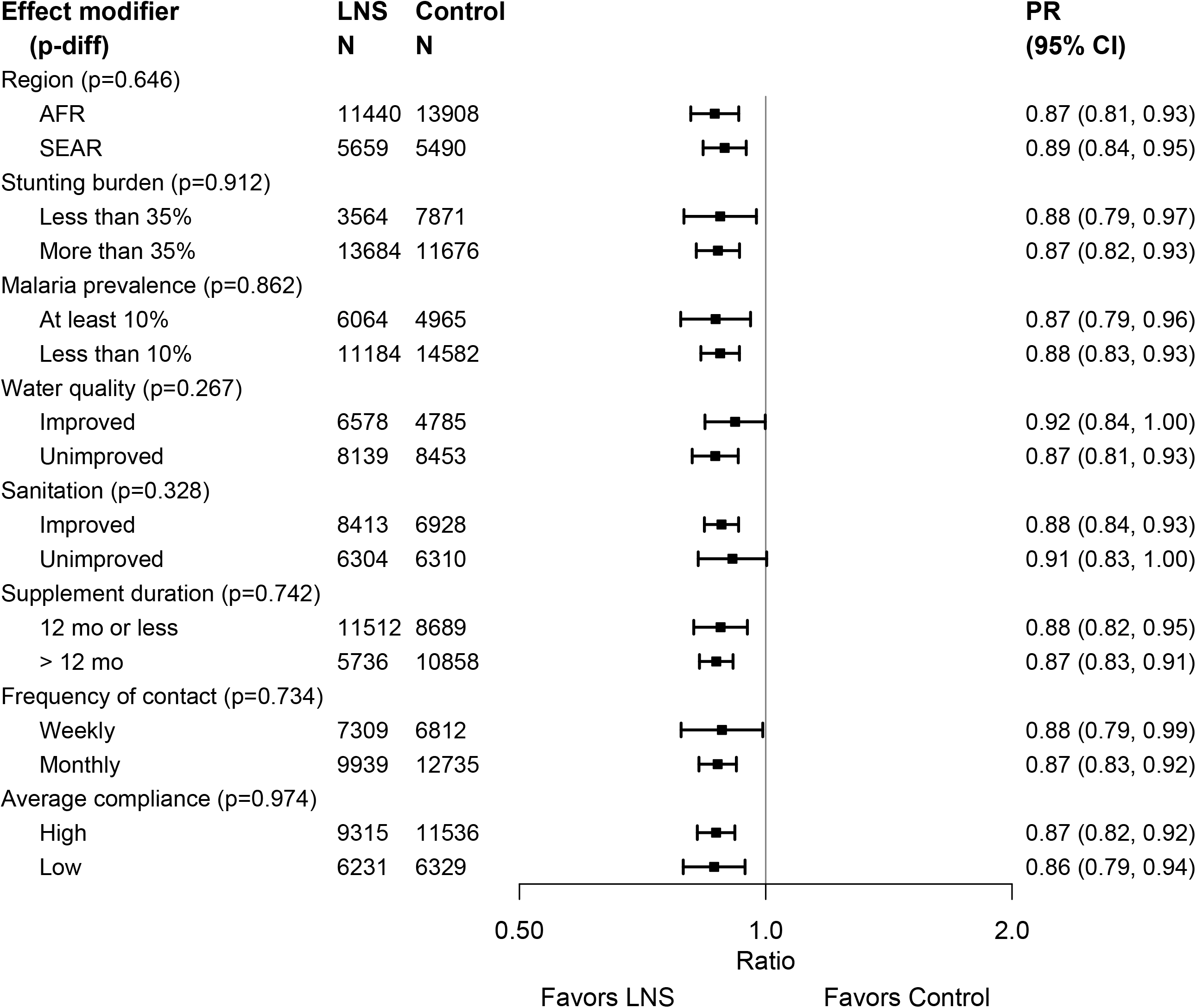
Pooled effect of SQ-LNS on stunting Study-level effect modifiers. Pooled effect of SQ-LNS on stunting stratified by study-level characteristics. P-diff, p-value for the difference in effects of SQ-LNS between the two levels of the effect modifier; PR, prevalence ratio.

#### WLZ, MUACZ, wasting, low MUAC and acute malnutrition

None of the study-level characteristics significantly modified the effect of SQ-LNS on mean WLZ or MUACZ, or the prevalence ratios for wasting, low MUAC or acute malnutrition (**Supplemental Figures 4D to 4K, Figure 5** and **Supplemental Figure 5)**. **Figure 5** shows that for 5 of the 8 study-level characteristics (region, malaria prevalence, water quality, sanitation, and duration of supplementation), the upper bound of the 95% confidence interval for wasting was ≤ 1 in both categories. A few of the study-level characteristics modified the effect of SQ-LNS on the prevalence of wasting, low MUAC or acute malnutrition when these outcomes were examined as prevalence differences (but not prevalence ratios). The percentage point reduction in wasting associated with SQ-LNS was greater in Bangladesh than in the other sites (**Supplemental Figure 4F.1**), and among sites with weekly (vs. monthly) contact (**Supplemental Figure 4F.7**). The percentage point reduction in low MUAC associated with SQ-LNS was greater among sites with average reported compliance ≥ 80% (vs. < 80%) (**Supplemental Figure 4I.8**).

**Figure 5:**
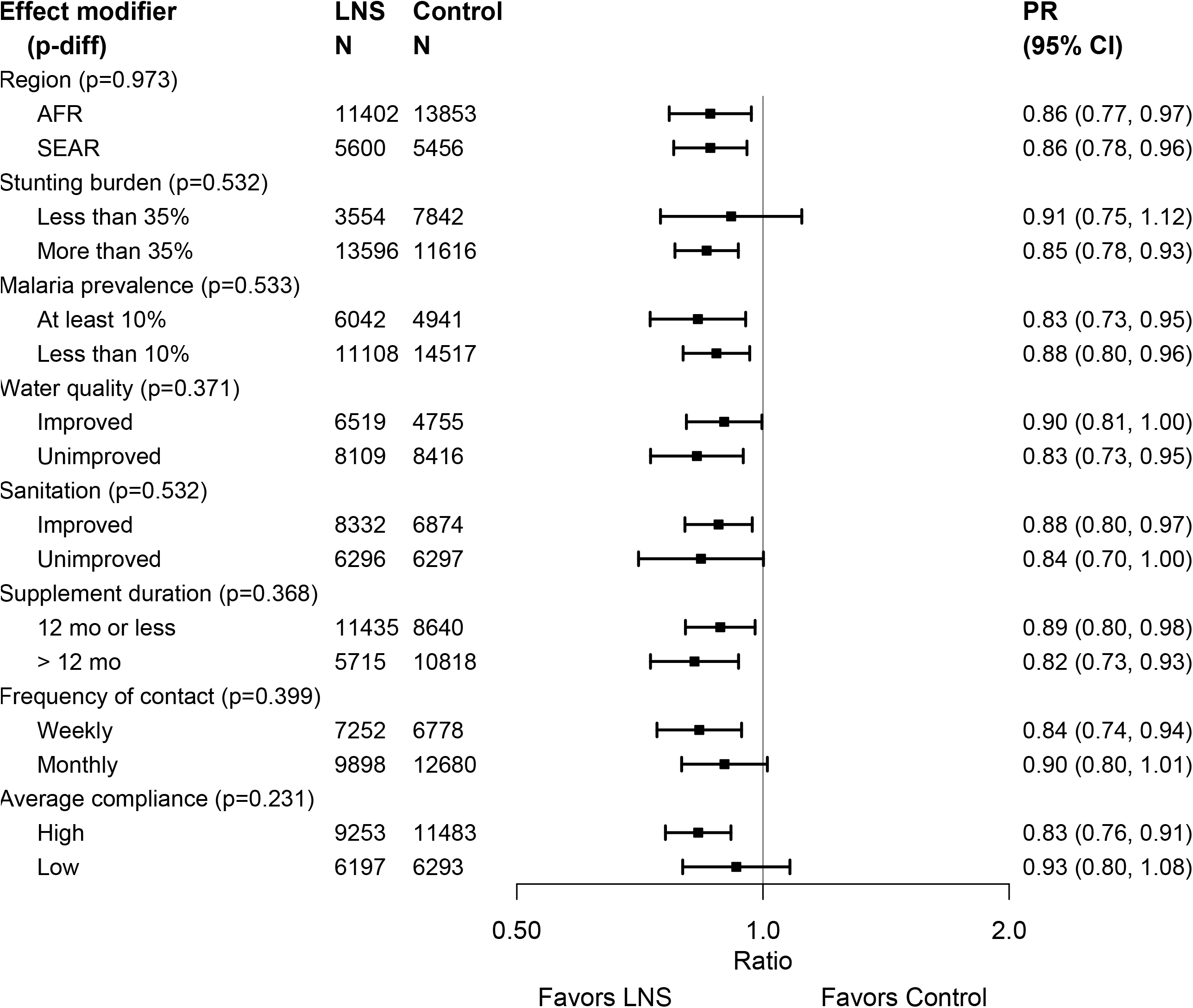
Pooled effect of SQ-LNS on wasting Study-level effect modifiers. Pooled effect of SQ-LNS on wasting stratified by study-level effect characteristics. P-diff, p-value for the difference in effects of SQ-LNS between the two levels of the effect modifier; PR, prevalence ratio.

#### WAZ and underweight

None of the study-level characteristics significantly modified the effect of SQ-LNS on mean WAZ or the prevalence of underweight when examined as a prevalence ratio (**Supplemental Figures 4L to 4M, Supplemental Figure 5)**. However, the percentage point reduction in underweight (prevalence difference) associated with SQ-LNS was greater among sites with weekly (vs. monthly) contact (**Supplemental Figure 4N.7**), and among sites with average reported compliance ≥ 80% (vs. < 80%) (**Supplemental Figure 4N.8**).

#### HCZ and small head size

Only one of the study-level characteristics significantly modified the effect of SQ-LNS on HCZ: the increase in mean HCZ associated with SQ-LNS was greater in sites with a high stunting burden (≥ 35%) than in sites with a stunting burden < 35% (**Supplemental Figure 4O.2**). None of the study-level characteristics significantly modified the effect of SQ-LNS on the prevalence ratio for small head size (**Supplemental Figure 5),** but 2 were effect modifiers for the prevalence difference: the percentage point reduction in small head size associated with SQ-LNS was greater in sites with a malaria prevalence ≥ 10% than in sites with a lower malaria prevalence (**Supplemental Figure 4Q.3**), and in sites with weekly (vs. monthly) contact (**Supplemental Figure 4Q.7**).

### Effect modification by individual-level maternal, child and household characteristics

Forest plots for all outcomes stratified by potential individual-level effect modifiers are presented in **Supplemental Figures 6 and 7**. Effect modification results were consistent across all sensitivity analyses (data not shown, available upon request), so the results presented below refer to the all-trials analysis. Results were generally similar in fixed-effects and random-effects models, although confidence intervals were wider for the latter as expected. Results presented are for the fixed-effects models; when the effect size of effect modification differed substantially in the random-effects model, this is mentioned below.

#### LAZ and stunting

Only one of the individual-level characteristics modified the effect of SQ-LNS on LAZ: the increase in mean LAZ associated with SQ-LNS was greater among children whose mothers scored lower for depressive symptoms than among those whose mothers scored in the top quartile, although the effect was significant in both subgroups (**Supplemental Figure 6A.5**). Several individual-level characteristics modified the effect of SQ-LNS on stunting prevalence. **Figure 6** shows the prevalence ratios for stunting stratified by maternal and child characteristics, and **Figure 7** shows the prevalence ratios stratified by household-level characteristics. There were significant effects in all subgroups (except among children of mothers in the top quartile for depressive symptoms). However, there was a greater effect of SQ-LNS on stunting among children of mothers who were taller, had higher BMI, had more education, and reported fewer depressive symptoms. There was also a greater effect of SQ-LNS on stunting among female (vs. male) children and children of higher birth order (hereafter termed “later-born”) compared to first-born children, although the latter difference was attenuated in the random-effects model. Baseline LAZ modified the effect of SQ-LNS on stunting, but in opposite directions depending on whether the outcome was examined as the prevalence ratio or the prevalence difference. For the former, the effect of SQ-LNS was greater among children whose LAZ was above −1 at baseline (27% reduction) (vs. those with lower LAZ at baseline, 9% reduction), but for the latter there was a greater percentage point reduction in stunting associated with SQ-LNS among children with lower baseline LAZ (6 percentage points) than among those with higher baseline LAZ (3 percentage points, **Supplemental Figure 6C.8**). None of the household-level characteristics modified the effect of SQ-LNS on LAZ or stunting prevalence; **Figure 7** illustrates significant reductions in stunting prevalence among children receiving SQ-LNS regardless of socio-economic status, food security, water quality, sanitation, home environment, or season at endline.

**Figure 6:**
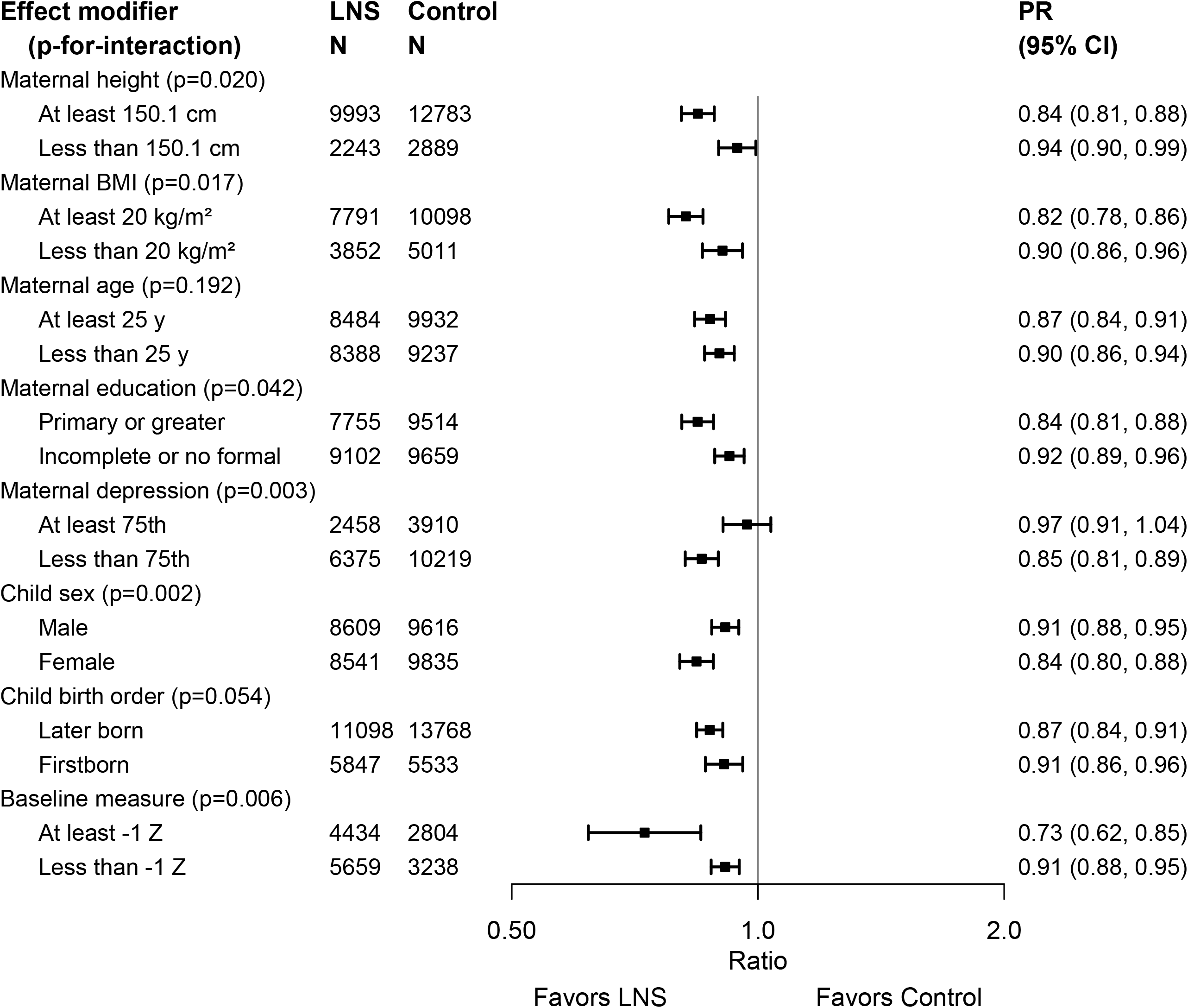
Pooled effect of SQ-LNS on stunting Maternal and child effect modifiers. Pooled effect of SQ-LNS on stunting stratified by individual-level maternal and child characteristics. P-for-interaction, p-value for the interaction indicating the difference in effects of SQ-LNS between the two levels of the effect modifier; PR, prevalence ratio.

**Figure 7:**
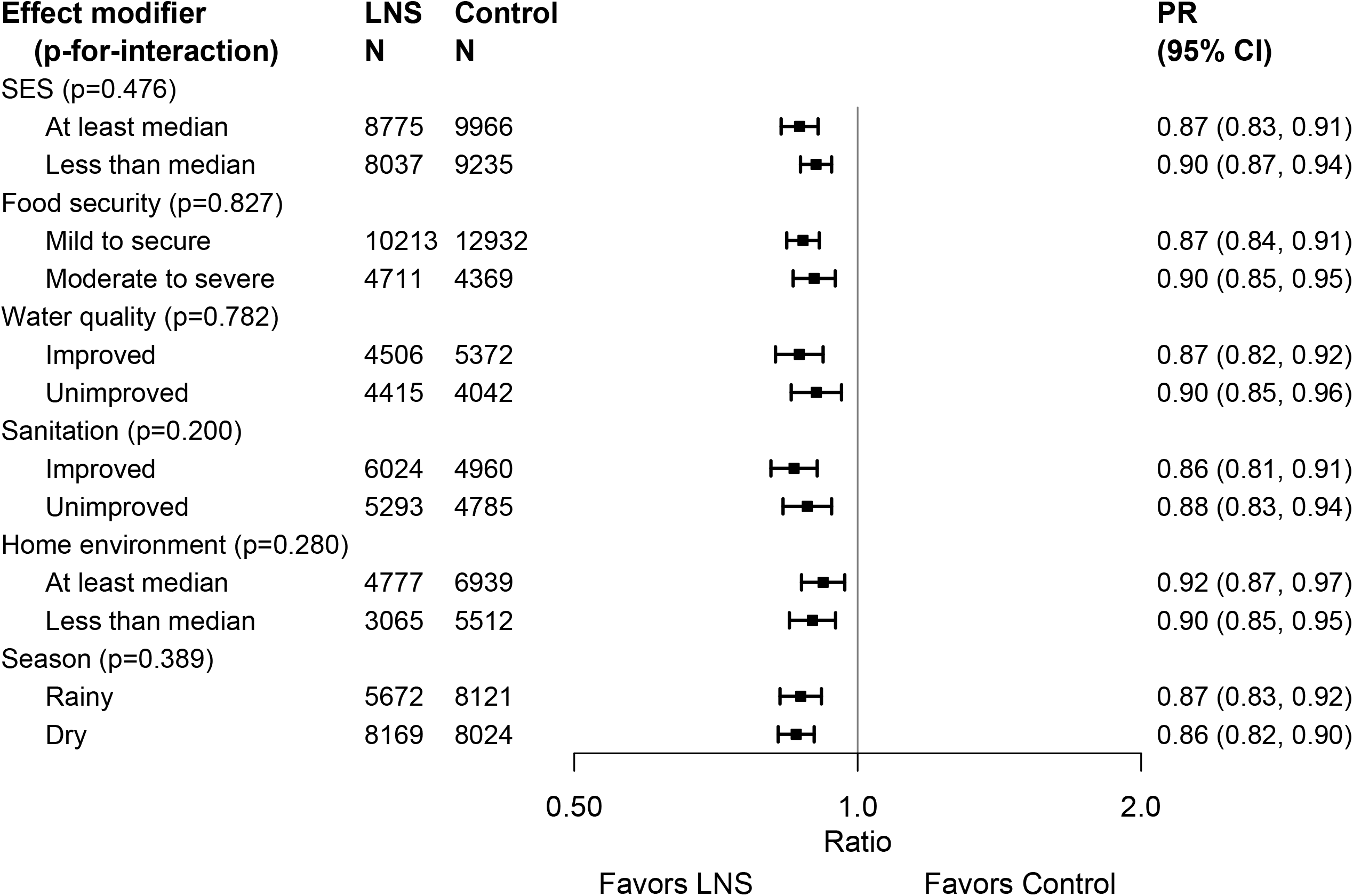
Pooled effect of SQ-LNS on stunting Household-level effect modifiers. Pooled effect of SQ-LNS on stunting stratified by individual-level household characteristics. P-for-interaction, p-value for the interaction indicating the difference in effects of SQ-LNS between the two levels of the effect modifier; PR, prevalence ratio.

#### WLZ, MUACZ, wasting, low MUAC and acute malnutrition

Several individual-level characteristics modified the effects of SQ-LNS on mean WLZ or MUACZ, or prevalence of wasting, low MUAC or acute malnutrition. Prevalence ratios for wasting, stratified by maternal and child characteristics and by household-level characteristics, are shown in **Figures 8 and 9**, respectively; all other outcomes are shown in **Supplemental Figures 6 and 7.** There was a greater effect of SQ-LNS on prevalence of wasting and low MUAC among female (vs. male) children. For mean WLZ and MUACZ, as well as the prevalence of wasting and acute malnutrition, the effects of SQ-LNS were greater among children in households with improved sanitation. For wasting the effects of SQ-LNS were also greater among children in households with improved water quality and children whose endline measurement occurred in the dry season. For mean MUACZ and the prevalence of low MUAC, the effects of SQ-LNS were greater among later-born (vs. first-born) children; for mean WLZ and MUACZ there was a greater effect among children in households with home environment scores below the study median (vs. above), although these differences were attenuated in the random-effects models; and for mean MUACZ there was a greater effect among children of taller (vs. shorter) mothers. Lastly, there was a greater percentage point reduction in wasting and acute malnutrition associated with SQ-LNS among children with lower baseline WLZ than among those with higher baseline WLZ, and a greater percentage point reduction in acute malnutrition among children of households with moderate to severe food insecurity (vs. mild or none).

**Figure 8:**
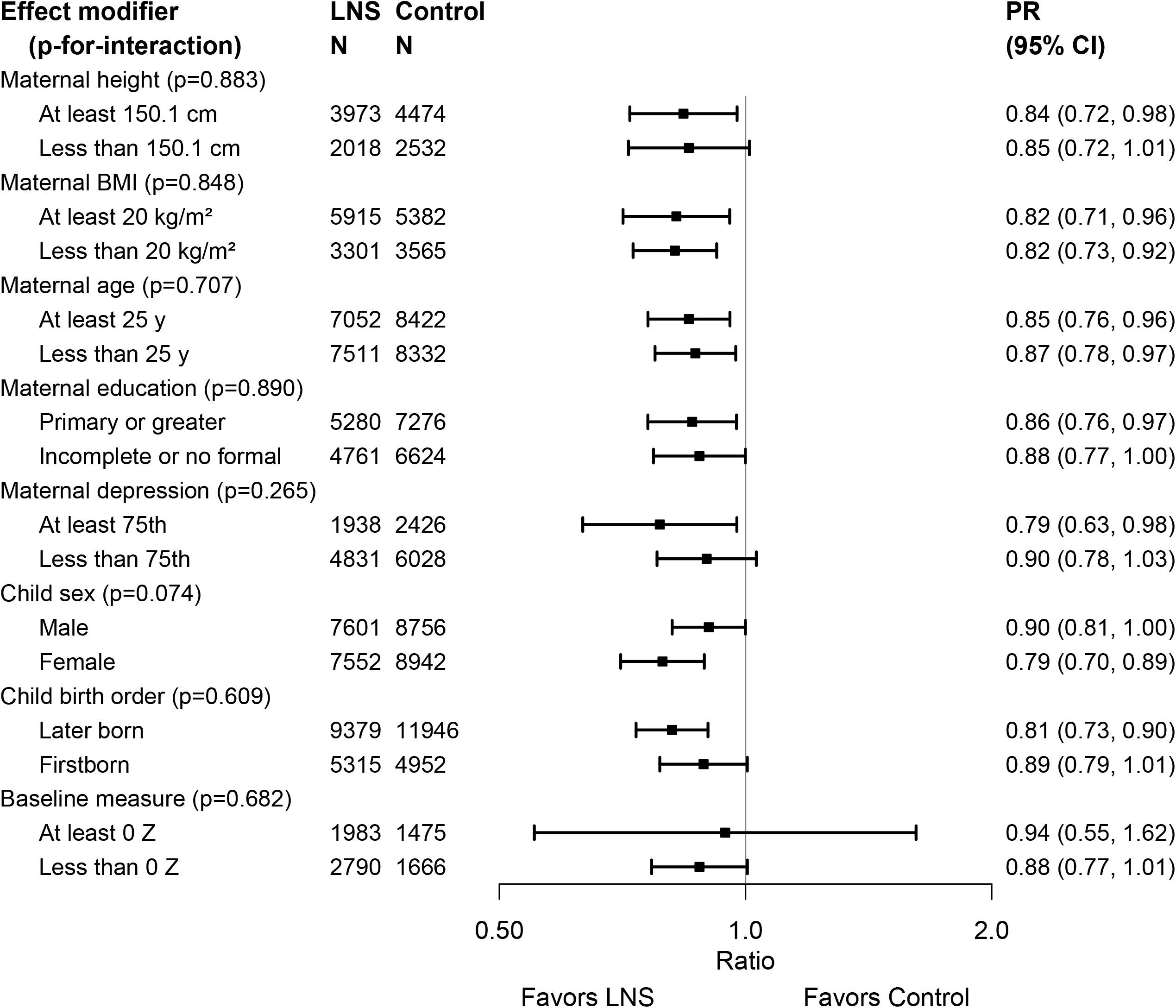
Pooled effect of SQ-LNS on wasting Maternal and child effect modifiers. Pooled effect of SQ-LNS on wasting stratified by individual-level maternal and child characteristics. P-for-interaction, p-value for the interaction indicating the difference in effects of SQ-LNS between the two levels of the effect modifier; PR, prevalence ratio.

**Figure 9:**
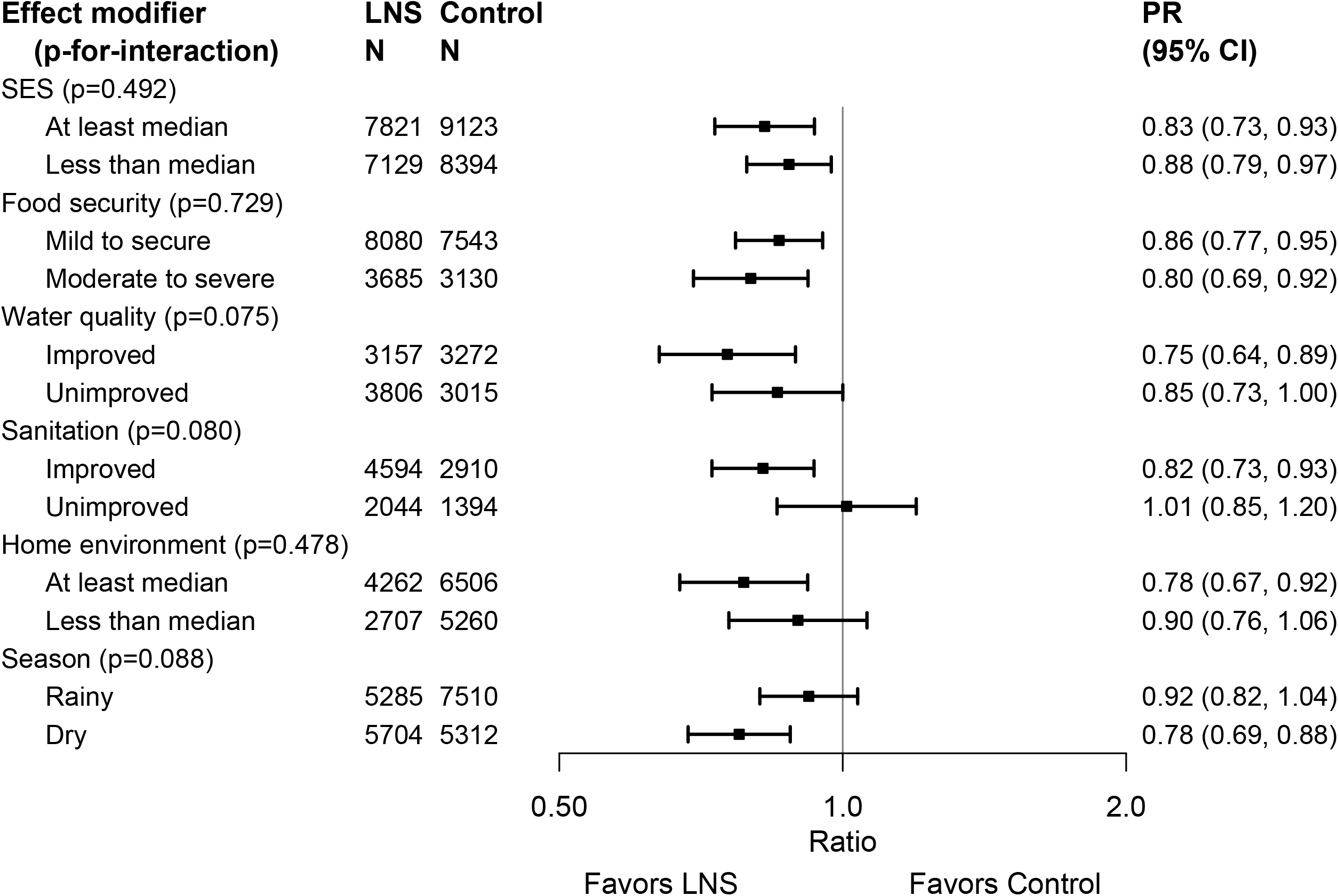
Pooled effect of SQ-LNS on wasting Household-level effect modifiers. Pooled effect of SQ-LNS on wasting stratified by individual-level household characteristics. P-for-interaction, p-value for the interaction indicating the difference in effects of SQ-LNS between the two levels of the effect modifier; PR, prevalence ratio.

#### WAZ and underweight

There was a greater effect of SQ-LNS on mean WAZ and prevalence of underweight among later-born (vs. first-born) children. In addition, effects of SQ-LNS on mean WAZ were greater among children with baseline WAZ < −1 (vs. > −1), and among children in households with improved sanitation (vs. unimproved) or a home environment score below the median. Effects of SQ-LNS on prevalence of underweight (prevalence ratio) were greater among children whose endline measurement occurred in the dry (vs. rainy) season. Lastly, there was a greater percentage point reduction in underweight associated with SQ-LNS among children with baseline WAZ < −1 (vs. > −1) and among children of households with moderate to severe food insecurity (vs. mild or none) (**Supplemental Figures 6 and 7)**.

#### HCZ and small head size

None of the individual-level characteristics modified the effect of SQ-LNS on mean HCZ. However, effects of SQ-LNS on the prevalence of small head size were greater among female (vs. male) children, those whose mothers scored in the top quartile for depressive symptoms (vs. lower), and those whose mothers had more education (vs. less). In addition, there was a greater percentage point reduction in small head size associated with SQ-LNS among children with baseline HCZ < −1 (vs. > −1) (**Supplemental Figures 6 and 7**).

#### Overview of individual-level effect modification

Table 3 shows that some characteristics (e.g., child sex, birth order, household sanitation and home environment) modified the effect of SQ-LNS on several different growth outcomes while others (e.g., maternal height, BMI, education, age; household SES, food security and water quality) exhibited effect modification for only 1 or 2 outcomes or none at all. The table also indicates that effect modification was more likely to be observed for binary outcomes than for continuous outcomes. When there is no significant effect modification for a continuous outcome (e.g., LAZ) but there is for the prevalence ratio or prevalence difference for the corresponding binary outcome (e.g. stunting), the results could be due to the “cutoff effect”, as described in Methods. The results of the simulations to identify cutoff effects are indicated in Table 3. They showed that for stunting prevalence, the cutoff effect explained the apparent effect modification by maternal stature and BMI, and some (but not all) of the apparent effect modification by maternal education. The cutoff effect also appeared to contribute, at least partially, to the apparent effect modification by baseline anthropometric status for each of the corresponding binary outcomes, and to apparent effect modification by household food security for the prevalence difference in acute malnutrition (but not underweight). For maternal depressive symptoms, however, there was significant effect modification for LAZ and both the prevalence ratio and prevalence difference for stunting, so this was not due to the cutoff effect. In addition, the cutoff effect did not explain effect modification by child sex, birth order, household water quality, sanitation, home environment or season.

**Table 3.**
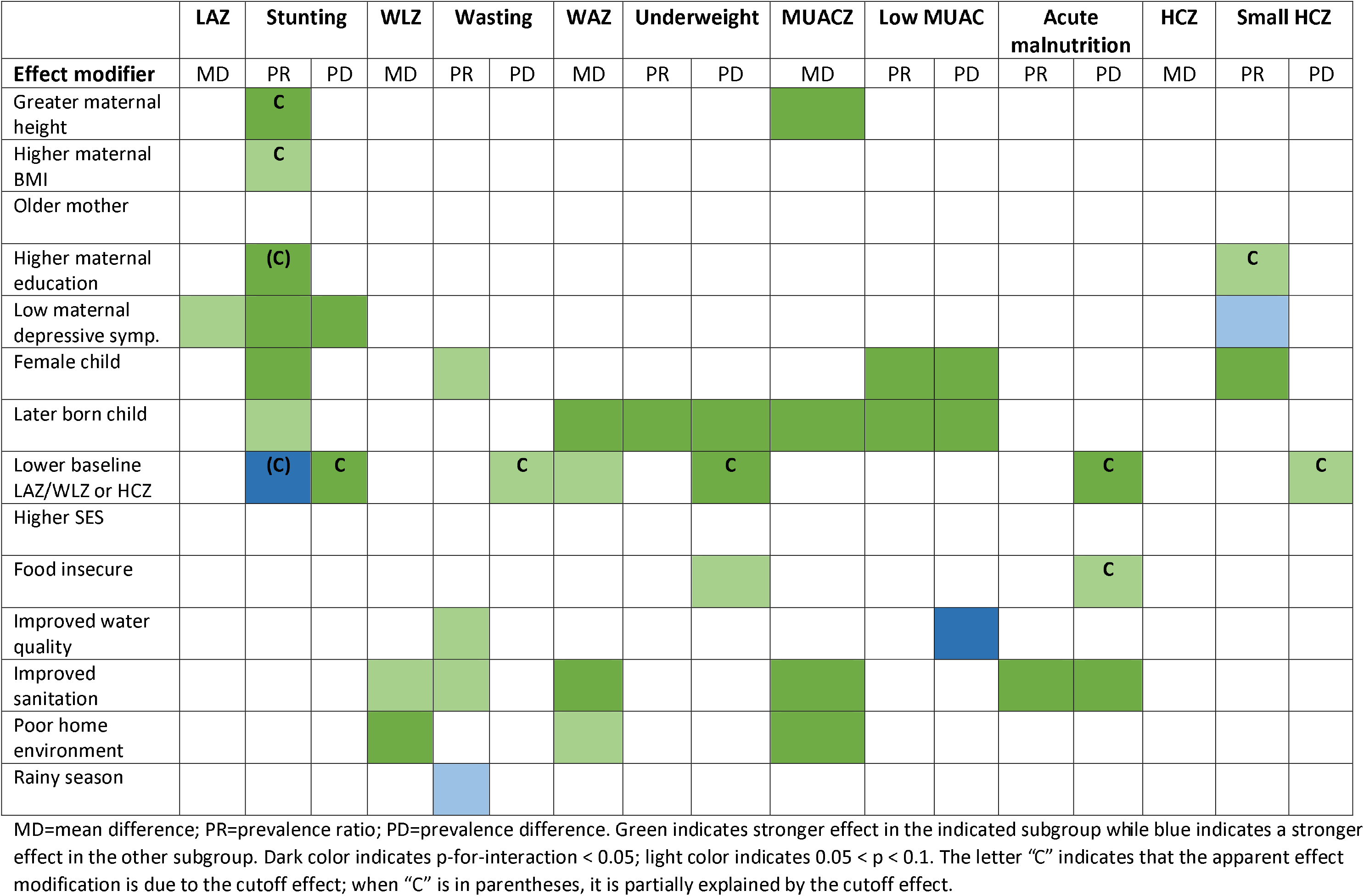
Overview of individual-level effect modification.

## Discussion

In this IPD analysis of 14 randomized controlled trials in 9 countries, with a total sample size of > 37,000 children, the relative reductions in adverse growth outcomes following provision of SQ-LNS to infants and young children 6 to 24 months of age were 12% for stunting, 14% for wasting and acute malnutrition, 18% for low MUAC, 13% for underweight, and 9% for small head size. The beneficial effects of SQ-LNS on stunting were evident regardless of region, stunting burden, malaria prevalence, sanitation, water quality, duration of supplementation, frequency of contact or average reported compliance with SQ-LNS. For wasting (and other adverse growth outcomes), there was also little evidence of effect modification by study-level characteristics, although within some subgroups of trials the effect estimates did not reach statistical significance (such as sites with a lower stunting burden and trials with lower average reported compliance). Several of the individual-level characteristics appeared to modify the effects of SQ-LNS on growth outcomes. For example, the effects of SQ-LNS on stunting, wasting, low MUAC and small head size were larger among girls than among boys; the effects on stunting, underweight and low MUAC were greater among later-born children than among first-born children; the effects on stunting were greater among children of mothers with higher (vs. lower) education levels as well as mothers with fewer depressive symptoms (vs. top quartile); and the effects on wasting and acute malnutrition were greater among children in households with improved sanitation than among those in households with unimproved sanitation.

### Main effects

In comparison to the meta-analysis by Das et al. (18), which included 13,372 children in 9 trials for the comparison of growth outcomes between LNS (SQ- or MQ-LNS) during the period of complementary feeding vs. no intervention, this IPD analysis includes nearly three times as many participants even though we restricted the analysis to trials that provided SQ-LNS. For all except 1 of the 14 trials in this IPD analysis, results were published in the 6-year period between 2014 and 2019. This rapid expansion in the evidence base permitted us to generate new estimates of impact on growth outcomes specific to SQ-LNS. These new estimates for wasting and underweight were similar to those of Das et al., but the new estimate for stunting (12% reduction) was larger than previously reported (7% reduction). Moreover, we report estimates for acute malnutrition, low MUAC and small head size, which were not included in the previous meta-analysis. Overall, the reductions in prevalence of adverse growth outcomes were modest, which is in line with previous reports for nutrition interventions (50). For example, the pooled prevalence difference for stunting was 5 percentage points. However, there was substantial heterogeneity in the effect of SQ-LNS on stunting, with some trials indicating no impact and others reporting reductions of 10-11 percentage points (37, 48). We did not find strong evidence that this heterogeneity in impact on stunting was explained by study-level characteristics, as discussed below.

The sensitivity analyses demonstrated similar results for all outcomes regardless of inclusion/exclusion of arms with maternal plus child SQ-LNS, trials with passive control arms, or arms with non-standard SQ-LNS formulations, as well as when analyses of multi-component intervention trials were structured to more specifically isolate the effects of SQ-LNS. This consistency across sensitivity analyses indicates that the findings are robust.

Because the impetus for this IPD analysis was to explore effect modification, we did not report results for rare outcomes (such as severe acute malnutrition) or outcomes that were evaluated in fewer than 3 trials. An example of the latter is the incidence rate of wasting or of moderate-to-severe acute malnutrition, which warrants further exploration because incidence rate is a more complete assessment of the wasting burden than prevalence of low WLZ at a single time point. For example, cross-sectional results from the trial in Mali (46) indicated an endline prevalence of 15% for acute malnutrition in the control group, but longitudinal results indicated that more than half of control group children experienced at least one episode of acute malnutrition during the 18-month follow-up period. In that trial, the SQ-LNS intervention had no effect on the prevalence of acute malnutrition in the cross-sectional sample but reduced the incidence rate of acute malnutrition in the longitudinal sample by 29%.

The objectives of this IPD analysis were focused on outcomes related to undernutrition, but there is considerable interest in whether interventions using energy-dense products such as SQ-LNS might increase the risk of child overweight. In the trials included in this analysis, the prevalence of child overweight at endline (WLZ > 2) was 0-4.6%, too low to calculate a pooled estimate for the prevalence ratio without excluding more than half of the trials. The prevalence of WLZ > 1 was 0.6-18%, and in only 5 of the 14 trials was the prevalence > 10% even though the prevalence in a normal healthy population should be ∼16%.

### Study-level effect modification

Study site characteristics did not appear to modify the impact of SQ-LNS on linear growth or stunting prevalence, suggesting that those effects are not restricted to certain regions, relevant only to populations with a high burden of stunting, or limited by malaria prevalence, study-level water quality or study-level sanitation. Study-level effect modification was also not significant for most of the other growth outcomes, but in some cases the differences in prevalence ratios were sizable (e.g., > 0.05) even though the p-diff for interaction was not significant, presumably due to limited statistical power for these types of analyses. For example, the relative reduction in wasting with provision of SQ-LNS was 17% in sites with unimproved water quality vs. 10% in sites with improved water quality, with similar findings for low MUAC for both water quality and sanitation. In addition, the relative reduction in wasting was 15% in sites with a higher stunting burden vs. 9% in sites with a lower stunting burden.

Aspects of study design also did not appear to modify the effects of SQ-LNS on linear growth or stunting, even though there was wide variation in setting, duration of supplementation, frequency of contact and average compliance with SQ-LNS. For other growth outcomes, effect modification by study design characteristics was also generally not significant, but again there were some differences that are notable. For example, trials with average reported compliance with consumption of SQ-LNS ≥ 80% tended to have greater relative reductions following provision of SQ-LNS (compared to trials with lower compliance) in prevalences of wasting (17% vs. 7%), low MUAC (23% vs. 14%), acute malnutrition (19% vs. 10%) and underweight (18% vs. 12%), and there was a significantly greater percentage point reduction in low MUAC and underweight in the former than in the latter. Average study compliance with SQ-LNS consumption ranged from 37% to 100%, but it should be noted that methods for assessing compliance varied widely across trials. Estimated compliance rates from self-reported and disappearance data may be overestimates when compared to observed intakes (51). In this IPD, effect modification by compliance was investigated only at the study-level, as compliance at the individual-level cannot be calculated for control group children who receive no supplements.

Thus, further research is needed to investigate potential dose-response relationships. In a recent evaluation of a program in the Democratic Republic of Congo in which SQ-LNS was distributed to infants 6-12 months of age (52), dose-response was examined among children in the intervention zone who were 8-13 months of age at endline, who were expected to have sufficient program exposure to expect a biological impact. Within that subsample, and adjusting for multiple potential confounding variables, children who had received at least 3 monthly distributions of SQ-LNS (n=80) had significantly greater LAZ (+0.4 (0.02, 0.78)) and a lower prevalence of stunting (−16.7 (−32.1, −1.2) percentage points) compared to those who did not receive SQ-LNS (n=89), whereas those who received 1-2 monthly distributions of SQ-LNS (n=136) did not differ from those who did not receive SQ-LNS (LAZ +0.08 (−0.24, 0.41); stunting prevalence difference −9.3 (−22.6, 3.9)).

Trials in this IPD analysis in which SQ-LNS was provided for > 12 months (vs. ≤ 12 months) tended to demonstrate greater relative reductions in wasting (18% vs. 11%), although the p-diff for interaction was not significant. The meta-analysis by Das et al. (18) suggested a somewhat greater impact on stunting when the duration of LNS supplementation was greater than 12 months (vs. ≤ 12 months), but we did not observe this. We also did not find significant effect modification by frequency of contact when growth results were expressed as mean differences or prevalence ratios, but trials with weekly (vs. monthly) contact tended to demonstrate greater relative reductions in wasting (16% vs. 10%) and low MUAC (23% vs. 14%) following SQ-LNS provision, and the percentage point reductions in wasting, underweight and small head size were significantly greater in the former than the latter.

We were unable to examine potential effect modification by child age at baseline because there was insufficient heterogeneity in this aspect of study design: most of the trials in this IPD analysis began supplementation at 6 months of age. However, in the Madagascar trial, enrollment occurred across a wide age range and the investigators reported significant effects on LAZ and stunting only among children who started SQ-LNS at 6 months (43).

Six of the 14 trials in this IPD analysis were conducted within existing community-based or clinic-based programs (35, 38, 41, 43, 46–48), so the findings reflect impact across the spectrum from efficacy trials to effectiveness studies in a real-world context. There were no clear dividing lines between “efficacy” and “effectiveness” trials among the 14 trials because some of the program-based trials had very high reported compliance rates and strong study designs. For that reason, we did not consider this dichotomy as a formal study-level effect modifier. However, there were no obvious differences in effect sizes between the program-based studies and the trials in which all activities were conducted by the research teams. Similarly, there were no clear differences in results between trials that included enhanced SBCC for IYCF (34, 36, 38, 42, 43, 46–48) and those that simply reinforced the normal IYCF messages already promoted in that setting (35, 37, 39–41, 44, 45).

### Individual-level effect modification

For individual-level effect modifiers, it is important to distinguish the potential to *benefit* from the potential to *respond* (9). The former is more likely when the child is more vulnerable, for example when the child is already undernourished at baseline or lives in a household with food insecurity or inadequate resources to provide adequate care. However, some children who exhibit signs of undernutrition (e.g., stunting) may actually be *less* likely to respond to a nutritional intervention because of other constraints on growth due to infection or inflammation, inadequate care, or other factors. Interpretation of effect modification also needs to take into account the pattern of results across continuous and binary outcomes. Significant effect modification for a continuous outcome (e.g., LAZ) is usually a clear-cut indication that the growth response to the intervention differs between subgroups. However, as explained above, effect modification for binary outcomes (e.g. stunting) could be due to the “cutoff effect”, depending on the distribution of the continuous outcome within each of the subgroups in relation to the cutoff. Interpretation of prevalence ratios is further complicated by the fact that the relative reduction in an adverse outcome will be greater in subgroups with a lower control group prevalence of the outcome (the denominator for the prevalence ratio) than in those with a higher control group prevalence. We therefore attempt to take into account all of these considerations in the following discussion of individual-level effect modifiers.

A consistent finding was that there was a greater effect of SQ-LNS on growth outcomes in girls than in boys. Among girls, SQ-LNS reduced stunting by 16%, wasting by 21%, low MUAC by 27% and small head size by 15%, whereas the reductions among boys were 9%, 10%, 7% and 4%, respectively. The cutoff effect did not explain these differences, nor were they explained by lower control group prevalences in girls than in boys. Although the p-for-interaction for child sex and intervention group was not significant for the continuous outcomes (LAZ, WLZ, MUACZ and HCZ), mean differences were somewhat greater in girls than in boys, particularly when length, MUAC and head circumference were examined in absolute units (cm) rather than as z-scores. We thus conclude that there is a real difference in response to SQ-LNS between boys and girls. Girls generally had better growth status than boys: in the control groups across the 14 trials at endline, prevalence of stunting was lower among girls in 13 trials and prevalence of wasting was lower among girls in 12, compared to boys. Thus, it is unlikely that girls had greater potential to benefit. Rather, the difference probably reflects a greater potential to respond to nutritional supplementation among girls. Other studies suggest that males are more vulnerable than are girls to adverse conditions in early life, which may be driven by biological factors (53, 54) that could also constrain responses to nutrition interventions. In a recent meta-analysis, antenatal multiple micronutrient supplementation significantly reduced neonatal mortality in girls but not in boys (55), which is consistent with this hypothesis.

Among later-born children, SQ-LNS reduced stunting by 13%, underweight by 17% and low MUAC by 23%, whereas the reductions among first-born children were 9%, 6% and 5%, respectively. These results were not explained by the cutoff effect. Moreover, significant effect modification was also observed for two of the continuous outcomes: WAZ and MUACZ. Later-born children have at least one older sibling who may compete for caregiving and family resources, making them potentially more vulnerable to undernutrition and thus more likely to benefit from nutritional supplementation. Although later-born children tend to be larger at birth than first-born children (56), prevalences of stunting and underweight at endline in the control groups for this IPD were greater among later-born children than first-born children in 8 of the 14 trials. This suggests that there may be greater potential to benefit among later-born children in some settings.

We observed greater effects of SQ-LNS on stunting among children of taller (vs. shorter) mothers (16% vs. 6% relative reduction). This could be due to greater constraints on a linear growth response among children of shorter mothers due to genetic, intra-uterine or environmental factors (57). Similarly, there were greater effects of SQ-LNS on stunting among children of mothers with higher (vs. lower) BMI (18% vs. 11% relative reduction), which could be related to a higher risk of fetal growth restriction and persistent postnatal constraints on growth among children of thinner mothers (8). However, for both of these examples, there was no significant effect modification for the continuous outcome, the mean difference in LAZ, and the cutoff effect was the most likely explanation for the findings. The cutoff effect also appeared to explain the greater effects of SQ-LNS on prevalence of small head size among children of women with higher levels of education (vs. less), but it did not fully explain effect modification by maternal education for stunting. Although the effects of SQ-LNS on LAZ did not differ between children of women with higher vs. lower educational level, the relative reduction in stunting was 16% vs. 8%, respectively. This may reflect a greater potential to respond to nutrient supplementation among children of mothers who are better educated; those mothers may have greater autonomy and agency, and be better able to adhere to advice regarding recommended frequency and dosage of supplementation.

Information on maternal depressive symptoms was not collected in all of the trials in this IPD analysis, but among those with such data, there was evidence of effect modification for 2 different growth outcomes in opposite directions. The effects of SQ-LNS on LAZ and stunting (both prevalence ratio and prevalence difference) were greater among children of mothers with lower (vs. higher) scores for depressive symptoms whereas the effects of SQ-LNS on the prevalence of small head size (but not mean HCZ) were greater among children of mothers in the top quartile for depressive symptoms (compared to those with lower scores). The former was not explained by the cutoff effect, and may reflect greater potential to respond to a nutritional supplement among children of less-depressed mothers (who may have more capability to effectively use nutritional supplements in ways that improve the health of the child). Among children in the control groups in this IPD, those whose mothers had lower scores for depressive symptoms had slightly higher mean LAZ and HCZ than those whose mothers were in the top quartile for depressive symptoms (a difference of ∼ −0.10 Z), suggesting that the former subgroup was less vulnerable, which is inconsistent with a greater potential to benefit. However, in the SHINE trial in Zimbabwe (Tome et al. in submission), the effect of the intervention on LAZ was greater among children of mothers who scored ≥ 12 on the Edinburgh Postnatal Depression Scale (a cutoff validated against clinically diagnosed major depression in Zimbabwean women (58)), such that the difference in mean LAZ between children of depressed vs. non-depressed mothers observed in the control group was not evident in the intervention group. This may reflect amelioration by SQ-LNS of the adverse influence of maternal depression on growth. A similar phenomenon might underlie the IPD analysis findings for small head size, but those results could also be spurious. Differences in the way maternal depression affects child caregiving across cultures or in the methods used to assess and define depression across studies may contribute to variation in results. Additional research to understand these relationships would be useful.

We observed greater effects of SQ-LNS on the percentage point reduction in acute malnutrition and underweight among children in households with moderate to severe food insecurity, compared to those in households with less food insecurity, though the results for acute malnutrition may be due to the cutoff effect. SQ-LNS may have been more important for helping to fill gaps in energy and micronutrient intakes for children in food insecure households than for children in households with greater food security. The former may have had greater potential to benefit, given that the prevalences of acute malnutrition and underweight in this IPD analysis were higher among control group children in food insecure households (by 4 and 8 percentage points, respectively) than among those in households with greater food security. Similarly, there were greater effects of SQ-LNS on WLZ, WAZ and MUACZ among children in households with home environment scores below the study median (vs. above). Among children in the control groups in this analysis, mean WLZ, WAZ and MUACZ were lower (by ∼0.11-0.18 Z) among those with lower (vs. higher) home environment scores in 7 of the 8 trials that included home environment information, suggesting greater potential to benefit.

By contrast, greater effects of SQ-LNS on WLZ, MUACZ, wasting and acute malnutrition were seen among children in households with improved (vs. unimproved) sanitation, and for wasting the effects were also greater among children in households with improved (vs. unimproved) water quality. These findings likely reflect a greater potential to respond. Better sanitation and water quality may reduce the constraints on growth that are linked to clinical and subclinical gastrointestinal disorders and inflammation (4, 5). However, these findings differ somewhat from what was seen in the 3 trials that included household level WASH interventions, which demonstrated that there was no added benefit of providing WASH interventions together with SQ-LNS as compared to providing SQ-LNS alone on wasting (36, 42, 47, 48). In this IPD analysis, greater effects of SQ-LNS on wasting and underweight were also seen when outcome measurements occurred during the dry (vs. rainy) season, which is consistent with the findings for household sanitation given that bacterial pathogens that cause diarrhea may be less prevalent during the dry season (59). There may also be more time for child care in farming households during the dry season. However, household sanitation and season did not significantly modify the effects of SQ-LNS on *stunting*, which is consistent with the results of the 3 trials mentioned above. It is likely that large improvements in sanitation and water quality at the community level, not just at the individual household level, are needed before a positive synergy between WASH and nutrition interventions is observed with respect to reductions in stunting (60–62).

The effect of SQ-LNS on risk of stunting (prevalence ratio) was greater among children whose baseline LAZ was higher (> −1) than among those with baseline LAZ < −1 (27% vs 9% reduction). However, the opposite was true when examining the prevalence difference in stunting: the percentage point reduction associated with SQ-LNS among children with higher baseline LAZ was less than the reduction among those with lower baseline LAZ (3 vs 6 percentage points). This is because the overall prevalence of stunting at endline was lower among children with a higher baseline LAZ, and thus the prevalence ratio calculation yielded a greater relative reduction in stunting than was the case for children with a lower baseline LAZ.

### Strengths and limitations

Strengths of these analyses include the large sample size, the substantial number of high-quality randomized controlled trials available, and the high participation rate among investigators invited to contribute data. In addition, the 14 study sites were highly diverse in terms of geographic location, stunting burden, malaria prevalence, water quality, sanitation and several aspects of study design, which provided heterogeneity for exploration of study-level potential effect modifiers. In general, the results were similar regardless of whether fixed-effects or random-effects models were used. The consistency in the results of the sensitivity analyses also strengthens the conclusions.

These analyses have a few limitations. Bangladesh was the only country represented in the Southeast Asia Region, and Haiti was the only country represented in Latin America and the Caribbean. Thus, more data from countries outside of Africa are needed. Data were unavailable from some of the trials for certain outcomes (MUACZ for 3 trials and HCZ for 4 trials) and for several individual-level potential effect modifiers (particularly maternal depressive symptoms and home environment), so fewer than 14 trials were represented in those analyses. In addition, fewer trials were represented in the analyses for outcomes with relatively low prevalences (e.g., wasting and acute malnutrition) because effect estimates could not be generated, especially when the number of trials was further restricted by low proportions of children within one of the effect modifier subgroups in some trials. Overall, statistical power for study-level effect modification was constrained by the limited number of trials, so there may be meaningful differences in effect estimates between categories of trials even if the p-diff for the association between the effect modifier and effect size was not significant. On the other hand, the individual-level effect modification analyses involved multiple effect modifiers and numerous outcomes, so several of the significant p-for-interaction values are likely due to chance. As stated in Methods, we did not adjust for multiple hypothesis testing because the effect modification analyses are inherently exploratory. Although we made every effort to standardize definitions and cutoffs for potential effect modifiers, there was variation in the methods used in the field to collect information on certain characteristics, such as household food insecurity and socioeconomic status. Lastly, caution is needed when interpreting the effect modification results because many of the potential effect modifiers are inter-related and also may be confounded by unmeasured variables. This is particularly important for the study-level characteristics, as there is substantial overlap in terms of which trials fall into each category for certain potential effect modifiers (e.g., the categorization of water quality and sanitation as “improved” or “unimproved” was the same within a trial for all but 3 of the trials). Thus, attribution of the relative potential to benefit or respond to SQ-LNS to a particular characteristic may not be warranted.

### Programmatic implications

These results suggest that policy-makers and program planners should consider including SQ-LNS in the mix of interventions to prevent both stunting and wasting. The overall effects on stunting (12% reduction) and wasting (14% reduction) may seem modest, but they are more substantial and more consistent than has been observed for other nutrition interventions for children under 2 years of age. Educational interventions to improve complementary feeding have shown positive effects on feeding practices, however there is insufficient evidence to demonstrate an impact of education alone on growth outcomes (63). Micronutrient supplementation, MNPs, and food fortification are effective for reducing anemia, but in a recent comparison of five different types of interventions for children under five, growth was improved only among children provided with LNS (50). Similarly, fortified cereals and milks for young children have shown little to no effect on growth outcomes, particularly linear growth (64–66). Provision of animal-source foods such as eggs is a promising strategy, but the evidence is too limited to draw conclusions regarding an impact on growth (67), and cost issues need to be considered. The cost of SQ-LNS is estimated at $0.07-0.14 per day (not including distribution costs), depending on scale and location of production (68, 69). A full discussion of cost issues is beyond the scope of this paper, but information on costs and willingness-to-pay is available elsewhere (68–71).

It is reassuring that no adverse effects of SQ-LNS on breast milk intake or infant feeding practices have been observed in any of the trials that reported on these outcomes, and in some settings there have been positive effects on feeding frequency and consumption of animal-source foods (72–76). However, SQ-LNS is not a stand-alone intervention and should always be accompanied by messaging to reinforce recommended IYCF practices in addition to appropriate use of the product. No adverse effects of SQ-LNS on child fatness or high WLZ or BMI have been observed, either at the end of the intervention period (77) or in longer-term follow-up studies (78, 79). One of the Bangladesh trials included in this IPD analysis showed that the LNS groups had greater increases in fat-free mass than in fat mass (77), which is consistent with improved linear growth and no increase in the risk of excess adiposity.

As demonstrated by the effect modification results herein, effects of SQ-LNS on stunting and wasting appear to be greater in girls (vs. boys) and later-born (vs. first-born) children. However, significant effects were also seen in boys and in first-born children, so we do not suggest targeting SQ-LNS to these subgroups, which would in any case be ethically and logistically challenging. However, the greater impact of SQ-LNS on growth of girls could be viewed as a positive finding with regard to the potential to reduce intergenerational stunting, if the increased height in early life persists later in life. Follow-up studies of 2 of the trials included in these analyses (35, 40) examined whether height differed between intervention groups at preschool age (3-6 years). In Bangladesh, the difference in height-for-age z-score between SQ-LNS and comparison groups was significant among female preschoolers (+0.09), though not among males; in households with moderate to severe food insecurity at baseline, stunting prevalence at 3-6 years of age was lower in the SQ-LNS group, by ∼6 percentage points in the total sample and ∼9 percentage points among females (79). In Ghana, height at 4-6 years of age did not differ significantly between intervention groups in the total sample, but there was a difference of +1.1 (95% CI: 0.2, 2.1) cm in the SQ-LNS group (vs. comparison groups) among children of women who were not overweight at baseline (78). Additional follow-up of other cohorts, and at older ages, is needed to determine if there is long-term persistence of growth differences among children who received SQ-LNS in early life.

The effect modification results regarding potential to *benefit* generally did not provide a strong rationale for targeting SQ-LNS only to the neediest, as growth benefits were generally similar regardless of study-level characteristics or household-level socio-economic status. However, some of the results suggested that a greater impact of SQ-LNS may be obtained by combining supplementation with interventions that address factors related to potential to *respond.* A growth response to nutritional supplementation may be constrained by infection, fetal growth restriction, or sub-optimal caregiving (3). Hence, integrated programs that combine SQ-LNS with interventions to prevent and control pre- and postnatal infection and inflammation, optimize fetal growth via improved maternal nutrition and other strategies, and support care for women and children (including maternal mental health promotion and education regarding optimal IYCF practices) should be further evaluated (15). Reducing constraints on a linear growth response may facilitate a larger reduction in stunting than observed in the pooled results of these analyses. Integrated programs that encompass an even broader set of interventions within food systems should also be a high priority. For example, the CHANGE project in Burkina Faso was designed to tackle multiple factors affecting child undernutrition by providing, in a staged process, a) inputs for home gardening and poultry production, with a focus on gender equity, b) education and training on agriculture, health, hygiene, and nutrition, c) WASH interventions, and d) SQ-LNS for children 6-24 months of age. Earlier phases of the program (without SQ-LNS) resulted in improvements in some outcomes, such as anemia, but stunting prevalence was reduced only in the group receiving all components, including SQ-LNS (by 7.7 percentage points) (80).

To our knowledge, SQ-LNS is the only nutrition intervention that has been documented to have positive effects not only on child growth, but also on iron deficiency and anemia (Wessells et al., this supplement), child development (Prado et al., this supplement) and child mortality (81). SQ-LNS can fill key nutrient gaps and reduce the cost of a nutritionally adequate diet (15), thereby potentially mitigating the adverse impact of rising food insecurity on vulnerable children (82). Although SQ-LNS is not a substitute for a diverse diet that includes healthy foods from each of the major food groups, it can play a protective role when access to certain foods (e.g. animal-source foods) is limited due to cost or other circumstances. A critical next step that we plan to undertake is a set of formal cost-effectiveness and cost-benefit analyses that take into account the multiple outcomes that may be influenced by SQ-LNS, similar to the recent cost-benefit analyses of MNPs (83). Also needed are additional rigorously designed evaluations of large-scale programs that include SQ-LNS, particularly in low and middle-income countries that are considering scaling-up this intervention.

## Supporting information

PRISMA Checklist

## Data Availability

Data described in the manuscript, code book, and analytic code will not be made available because they are compiled from 14 different trials, and access is under the control of the investigators of each of those trials.

## Acknowledgments

We thank all of the co-investigators, collaborators, study teams, participants and local communities involved in the trials included in these analyses. These trials benefitted from the contributions of many partner organizations, including: icddr,b (JiVitA-4, Rang-Din Nutrition Study and WASH Benefits trial in Bangladesh); the World Food Program (JiVitA-4 trial in Bangladesh); the Health District of Dandé and the relevant local health-care authorities (iLiNS-ZINC trial in Burkina Faso); AfricSanté and Helen Keller International (PROMIS trials in Burkina Faso and Mali); Ministry of Public Health and Population (Haiti trial); Innovations for Poverty Action and the Kenya Medical Research Institute (WASH-Benefits trial in Kenya); Unité Programme National de Nutrition Communautaire, Government of Madagascar, and World Bank Health and Nutrition and Population Global Practice (MAHAY trial in Madagascar); the Ministry of Health and Child Care in Harare, Chirumanzu and Shurugwi districts, and Midlands Province (SHINE trial in Zimbabwe); the International Lipid-based Nutrient Supplements Project Steering Committee (iLiNS Project trials); and Nutriset (for development of SQ-LNS). We thank Emily Smith for advice on IPD analysis methods.

The authors’ responsibilities were as follows—KGD: drafted the manuscript with input from KRW, CDA, ELP, CPS and other coauthors; KRW, CDA, KGD, ELP and CPS: wrote the statistical analysis plan; BFA, PA, EB, LH and JHH: reviewed, contributed to, and approved the statistical analysis plan; KRW and CDA: compiled the data; CDA: conducted the data analysis; and all authors: read, contributed to, and approved the final manuscript.

Supported by Bill & Melinda Gates Foundation grant OPP49817 (to KGD). BA received travel support (airfare and hotel) covered by the Bill & Melinda Gates Foundation to attend meetings in Seattle during the period of this IPD analysis project; PC was an employee of the Bill & Melinda Gates Foundation when this project was conceived until December 2019. All other authors report no conflicts of interest.

## Supplemental Tables

Supplemental Table 1: Amount of SQ-LNS provided (g/day) and nutrient value (per daily ration)

Supplemental Table 2: Descriptive information on potential study-level effect modifiers, by trial

Supplemental Table 3: Descriptive information on potential individual-level effect modifiers, by trial

Supplemental Table 4: Growth outcomes at endline among control groups, by trial

Supplemental Table 5: Risk of bias assessment in each trial

## Supplemental Figures

Supplemental Figure 1: Summary risk of bias as a percentage of all included studies

Supplemental Figure 2: Sensitivity analyses of main effects

Supplemental Figure 3: Forest plots for all main effects

Supplemental Figure 4: Forest plots for all outcomes stratified by study-level effect modifiers

Supplemental Figure 5: Pooled effect of SQ-LNS on low MUAC, acute malnutrition, underweight and small head size, stratified by study-level and individual-level effect characteristics

Supplemental Figure 6: Forest plots for all outcomes stratified by individual-level maternal and child effect modifiers

Supplemental Figure 7: Forest plots for all outcomes stratified by individual-level household effect modifiers

