## Supplemental Tables 1-5 for "Characteristics that modify the effect of small-quantity lipid-based nutrient supplementation on child growth: an individual participant data meta-analysis of randomized controlled trials"

**Supplemental Table 1. Amount of LNS provided (g/day) and nutrient value (per daily ration)**

| | Nutributter[1] | iLiNS Project formulation[2,3] | Revised iLiNS Project formulation[4] | Plumpy'Doz (6-11 mo)[5] | Plumpy'Doz (12-18 mo)[5] | Rice-lentil LNS (6-11 mo)[5] | Rice-lentil LNS (12-18 mo)[5] | Chickpea LNS (6-11 mo)[5] | Chickpea LNS (12-18 mo)[5] |
|---|---|---|---|---|---|---|---|---|---|
| Ration (g/day) | 20 | 20 | 20 | 23.2 | 46.4 | 25.7 | 51.4 | 23.6 | 47.2 |
| Total energy (kcal) | 108 | 118 | 118 | 123.4 | 246.8 | 133.9 | 267.8 | 128.6 | 257.2 |
| Protein (g) | 2.56 | 2.6 | 2.6 | 2.9 | 5.9 | 2.8 | 5.7 | 3.5 | 7.1 |
| Fat (g) | 7.08 | 9.6 | 9.6 | 7.9 | 15.8 | 6.9 | 13.9 | 6.6 | 13.2 |
| Linoleic acid (g) | 1.29 | 4.46 | 4.46 | | | | | | |
| α-Linolenic acid (g) | 0.29 | 0.58 | 0.58 | | | | | | |
| Vitamin A (µg RE) | 400 | 400 | 400 | 200 | 400 | 117.7 | 235.4 | 118.5 | 236.9 |
| Vitamin C (mg) | 30 | 30 | 30 | | | | | | |
| Vitamin B1 (mg) | 0.3 | 0.3 | 0.5 | 0.3 | 0.5 | 0.3 | 0.6 | 0.3 | 0.6 |
| Vitamin B2 (mg) | 0.4 | 0.4 | 0.5 | 0.3 | 0.5 | 0.2 | 0.5 | 0.2 | 0.5 |
| Niacin (mg) | 4 | 4 | 6 | 2.8 | 5.6 | 2.6 | 5.1 | 2.4 | 4.7 |
| Folic acid (µg) | 80 | 80 | 150 | 80 | 160.1 | 100 | 199.9 | 118.2 | 236.5 |
| Pantothenic acid (mg) | 1.8 | 1.8 | 2 | 1 | 2 | 1.1 | 2.2 | 1 | 2 |
| Vitamin B6 (mg) | 0.3 | 0.3 | 0.5 | 0.3 | 0.5 | 0.3 | 0.6 | 0.3 | 0.6 |
| Vitamin B12 (µg) | 0.5 | 0.5 | 0.9 | 0.4 | 0.8 | 0.5 | 0.9 | 0.5 | 0.9 |
| Vitamin D (IU) 1 IU = 0.025 ug | 0 | 200 | 200 | | | 226.2 | 452.3 | 226.6 | 453.1 |
| Vitamin E (mg) | 0 | 6 | 6 | 3 | 6 | 4.9 | 9.8 | 4.7 | 9.4 |
| Vitamin K (µg) | 0 | 30 | 30 | | | 11.3 | 22.6 | 11.3 | 22.7 |
| Iron (mg) | 9 | 6 | 9 | 4.5 | 9 | 3.3 | 6.7 | 3.5 | 7.1 |
| Zinc (mg) | 4 | 8 | 8 | 2 | 4 | 2.4 | 4.8 | 2.5 | 5.1 |
| Copper (mg) | 0.2 | 0.34 | 0.34 | 0.2 | 0.3 | 0.2 | 0.4 | 0.3 | 0.5 |
| Calcium (mg) | 100 | 280 | 280 | 193.5 | 387 | 208.2 | 416.3 | 219.5 | 439 |
| Phosphorus (mg) | 82 | 190 | 190 | 137.6 | 275.2 | 61.7 | 123.4 | 72.9 | 145.8 |
| Potassium (mg) | 152 | 200 | 200 | 155 | 310 | 206.6 | 413.3 | 220.7 | 441.3 |
| Magnesium (mg) | 16 | 40 | 40 | 29.9 | 59.9 | 41.6 | 83.3 | 47.7 | 95.3 |
| Selenium (µg) | 10 | 20 | 20 | 8.6 | 17.2 | 7.5 | 14.9 | 7.6 | 15.1 |
| Iodine (µg) | 90 | 90 | 90 | 27.6 | 55.2 | 33.7 | 67.3 | 33.7 | 67.5 |
| Manganese (mg) | 0.08 | 1.2 | 1.2 | 0.1 | 0.1 | 0.5 | 0.9 | 0.4 | 0.9 |

[1]Provided by Adu-Afarwuah 2007 (39), Iannotti 2014 (41)

[2]Provided by Hess 2015 (37), Becquey 2019 (38), Adu-Afarwuah 2016 (40), Galasso 2019 (43), Ashorn 2015 (44), Maleta 2015 (45), Huybregts 2019 (46), Humphrey 2019 (47), Prendergast 2019 (48). iLiNS (International Lipid-based Nutrient Supplements) Project formulation described in Arimond et al. 2013 (16).

[3]Hess 2015 (37) provided 0-10 mg zinc/d in the SQ-LNS product, plus a 5 mg/d zinc supplement in one intervention arm. Maleta 2015 provided 10-40 g/d of LNS, with and without milk powder, varying by intervention arm (the 40 g arms were not included in this IPD analysis); the micronutrient composition of the LNS was identical across intervention arms, but there were differences in total kcal, protein, fat, linoleic acid and α-linolenic acid.

[4]Provided by Dewey 2017 (35), Luby 2018 (36), Null 2018 (42).

[5]Provided by Christian 2015 (34). Product and quantities differed by intervention arm and age.

**Supplemental Table 2: Descriptive information on potential study-level effect modifiers, by trial**

| Country | First author, year | Region | Stunting prevalence at 18 mo (control) (%) | Malaria Prevalence (%) | Water quality (% improved) | Sanitation (% improved) | Duration of supplementation | Intensity of visitation | Average SQ-LNS compliance (%) | Compliance definition (in the SQ-LNS group) |
|---|---|---|---|---|---|---|---|---|---|---|
| Bangladesh | Christian, 2015 (34) | SEAR | 44.2[b] | 0.2[a] | 100.0[a] | 77.0[a] | ≤ 12 mo | Weekly | 93.0[a] | % of total intended SQ-LNS consumed (quantity * day) |
| Bangladesh | Dewey, 2017 (35) | SEAR | 35.2[b] | 0.2[a] | 100.0[a] | 71.1[a] | > 12 mo | Monthly | 97.4[a] | % reporting "high adherence" (≥ 4 days/week) |
| Bangladesh | Luby, 2018 (36) | SEAR | 43.4[b] | 0.1[a] | 88.5[a] | 94.7[a] | > 12 mo | Weekly | 93.0[a] | Number of sachets consumed in 14 days prior to annual survey/14 |
| Burkina Faso | Hess, 2015 (37) | AFR | 39.4[b] | 59.1[b] | 26.7[b] | 2.3[b] | ≤ 12 mo | Weekly | 96.8[a] | % of days SQ-LNS reported consumed |
| Burkina Faso | Becquey, 2019 (38)[1] | AFR | 30.5[a] | 42.9[b] | 50.6[b] | 53.4[a] | ≤ 12 mo | Monthly | 37.0[b] | caregiver reported receiving SQ-LNS in the previous month |
| Ghana | Adu Afarwuah,2007 (39) | AFR | 7.3[a] | 37.4[b] | 91.9[a] | 91.5[a] | ≤ 12 mo | Weekly | 88.2[a] | % of days SQ-LNS reported consumed |
| Ghana | Adu Afarwuah, 2016 (40) | AFR | 13.0[a] | 36.8[b] | 98.4[a] | 97.3[a] | ≤ 12 mo | Weekly | 73.5[b] | % of days SQ-LNS reported consumed |
| Haiti | Iannotti, 2014 (41) | AMR | 13.4[a] | 0.9[a] | 98.7[a] | 94.0[a] | ≤ 12 mo | Monthly | 97.0[a] | Reported consuming all of the monthly supply of the SQ-LNS during the supplementation period |
| Kenya | Null, 2018 (42) | AFR | 32.2[a] | 8.5[a] | 68.0[b] | 15.8[b] | > 12 mo | Monthly | 115.0[a] | Number of sachets consumed in 14 days prior to annual survey/14 |
| Madagascar | Galasso, 2019 (43) | AFR | 63.0[b] | 5.5[a] | 26.9[b] | 0.0[b] | ≤ 12 mo | Monthly | - | Data unavailable. |
| Malawi | Ashorn, 2015 (44) | AFR | 34.7[a] | 26.8[b] | 91.7[a] | 9.2[b] | ≤ 12 mo | Weekly | 77.1[b] | % of days SQ-LNS reported consumed |
| Malawi | Maleta, 2015 (45) | AFR | 46.5[b] | 30.3[b] | 91.6[a] | 2.9[b] | ≤ 12 mo | Weekly | 71.6[b] | % of days SQ-LNS reported consumed (considering missed delivery visits) |
| Mali | Huybregts, 2019 (46)[1] | AFR | 36.4[b] | 39.1[b] | 50.8[a] | 75.3[a] | > 12 mo | Monthly | 47.0[b] | caregiver reported receiving SQ-LNS in the previous month |
| Zimbabwe | Humphrey, 2019 (47); Prendergast, 2019 (48)[2] | AFR | 37.2[b] | 9.0[a] | 63.6[b] | 34.0[b] | ≤ 12 mo | Monthly | 73.5[b] | received > 11 (80% of expected) deliveries * consumed SQ-LNS in past 24 h (at 12 month visit) |

[1]Study-level effect modifier categorization based on longitudinal cohort

[2]Study-level effect modifier categorization was the same for both HIV exposed and HIV un-exposed children

Superscripts a and b designate the two categories, as described in Notes below.

**Abbreviations**: SQ-LNS, small-quantity lipid-based nutrient supplements

**Notes:**

Geograhic region based on WHO regions

Stunting was defined as length-for-age Z score < -2 SD. Stunting prevalence was based on study-specific data at 18 months (when available) in the control arms. Stunting was assessed at 12 months for Adu-Afarwuah 2007 (39) and Iannotti 2014 (41), and at ~25 months of age for Null 2018 (42). These 3 studies were then categorized based on expected changes in stunting prevalence with age. Trials were categorized as (a) low/moderate burden when stunting was < 35% and (b) high burden when stunting was ≥35% .

Malaria prevalence: Data extracted from Annex, Data table F: Population at risk and reestimated malaria cases and deaths, 2010-2017 (wmr2018-annex-table-f.xls); Point estimate (presumed and confirmed malaria cases), divided by population at risk, per 100 persons. Trials were categorized as (a) low burden when malaria was < 10% and (b) high burden when malaria was ≥10%.

Water quality: Data based on sample prevalences of improved source water quality. Study-level water quality was considered improved if the main source of drinking water for ≥ 75% of participants was improved; study-level water quality was considered unimproved if the main source of drinking water for < 75% of participants was improved. "Improved water sources are those that have the potential to deliver safe water by nature of their design and construction, and include: piped water, boreholes or tubewells, protected dug wells, protected springs, rainwater and packaged or delivered water". Unimproved water sources include: water from an unprotected dug well or unprotected spring, or surface water (e.g., river, dam, lake, pond, stream, canal or irrigation canal). (washdata.org/monitoring/drinking-water).

Sanitation: Data based on sample prevalences of improved sanitation. Study-level sanitation was considered improved if sanitation services for ≥ 50% of participants were improved; study-level sanitation was considered unimproved if sanitation services for < 50% of participants were improved. "Improved sanitation facilities are those designed to hygienically separate excreta from human contact, and include: flush/pour flush to piped sewer system, septic tanks or pit latrines; ventilated improved pit latrines, composting toilets or pit latrines with slabs". Unimproved sanitation services include: use of pit latrines without a slab or platform, hanging latrines or bucket latrines, or open defecation. (https://washdata.org/monitoring/sanitation)

Compliance: Data extracted from publication; study-specific definitions of compliance are noted in the table. Trials were categorized as (a) high compliance when complinace was ≥ 80% or (b) low compliance when compliance was < 80% compliance.

**Supplemental Table 3: Descriptive information on potential individual-level effect modifiers, by trial**

| Country | First author, year | Maternal height < 150.1 cm (%) | Maternal BMI < 20 $kg/m^2$ (%) | Maternal age < 25 y (%) | Maternal education, completed primary (%) | Maternal depression, below 75th (%) | Child sex, male (%) | Child birth order, first born (%) | Child baseline LAZ < - 1 (%) | Child baseline WLZ < 0 (%) | Child baseline MUACZ < 0 (%) | Child baseline WAZ < -1 (%) | Child baseline HCZ < -1 (%) | Moderate to severe food insecurity (%) | SES index, below median (%) | Improved source water quality (%) | Improved sanitation access (%) | Home environment, below median (%) | Season at endline, dry (%) |
|---|---|---|---|---|---|---|---|---|---|---|---|---|---|---|---|---|---|---|---|
| Bangladesh | Christian, 2015 (34) | | | 56.8 | 63.5 | | 50.2 | 79.0* | 62.9 | 65.2 | 76.2 | 57.2 | 50.8 | 29.6[a] | 49.4 | 100.0 | 77.2 | | 53.7 |
| Bangladesh | Dewey, 2017 (35) | 45.7 | 55.5 | 72.8 | 74.0 | 65.8[a] | 50.2 | 40.2 | 60.3 | 63.5 | 56.7 | 52.6 | 67.8 | 37.4[b] | 49.9 | 100.0 | 70.8 | 28.2[b] | 44.3 |
| Bangladesh | Luby, 2018 (36) | 45.5 | 54.2 | 56.2 | 71.2 | 75.1[d] | 49.7 | 34.1 | | | | | | 22.1[b] | 49.8 | 89.0 | 94.7 | 48.8[b] | 55.8 |
| Burkina Faso | Hess, 2015 (37) | 1.9 | 38.7 | 41.2 | 3.8 | | 50.7 | 22.1 | 56.8 | 82.7 | 81.8 | 63.7 | 72.1 | 48.9[b] | 41.4 | 26.6 | 2.3 | 39.9[a] | 71.5 |
| Burkina Faso | Becquey, 2019 (cross-sectional) (38) | 2.3 | 40.5 | 54.7 | 8.4 | | 52.5 | 16.5 | | | | | | | 50.5 | 63.1 | 39.3 | | 47.6 |
| Burkina Faso | Becquey, 2019 (longitudinal) (38) | 2.8 | 25.2 | 43.2 | 7.0 | 71.1[b] | 51.9 | 17.2 | 36.3 | 56.2 | | 37.1 | | 40.8[b] | 49.9 | 50.6 | 53.5 | | 49.6 |
| Ghana | Adu Afarwuah, 2007 (39) | 4.3 | 9.2 | 29.0 | 88.3 | | 52.1 | 40.0 | | | | | | | 46.3 | 91.9 | 91.5 | | 57.7 |
| Ghana | Adu Afarwuah, 2016 (40) | 5.1 | 15.2 | 38.8 | 78.5 | 70.9[a] | 48.0 | 33.1 | 38.9 | 51.7 | 54.5 | 32.8 | 40.6 | 31.0[b] | 50.6 | 98.5 | 97.2 | 37.1[a] | 58.8 |
| Haiti | Iannotti, 2014 (41) | | | 30.9 | 85.3 | | 43.7 | 37.0* | 29.8 | 49.0 | | 25.2 | | | 49.3 | 99.0 | 94.2 | | |
| Kenya | Null, 2018 (42) | 4.0 | 21.6 | 44.2 | 47.6 | 74.0[e] | 48.2 | 21.4 | | | | | | 10.3[b] | 43.1 | 67.9 | 15.8 | 43.2[b] | 22.7 |
| Madagascar | Galasso, 2019 (43) | 36.2 | | 45.4 | 23.1 | 72.2[f] | 49.1 | 26.4 | | | | | | 28.5[b] | 49.7 | 26.8 | 0.0 | 46.2[c] | 99.9 |
| Malawi | Ashorn, 2015 (44) | 13.3 | 40.3 | 50.0 | 15.8 | 74.2[a] | 47.3 | 20.5 | 58.4 | 35.3 | 45.4 | 33.1 | 23.9 | 70.7[b] | 46.5 | 91.5 | 9.4 | 34.8[a] | 71.1 |
| Malawi | Maleta, 2015 (45) | 16.7 | 26.1 | 45.1 | 23.9 | | 50.6 | 23.7 | 63.9 | 42.2 | 42.5 | 40.4 | 44.9 | 73.5[b] | 49.1 | 91.6 | 2.9 | 41.3[a] | 58.9 |
| Mali | Huybregts, 2019 (cross-sectional) (46) | 3.1 | 27.4 | 47.5 | 10.6 | | 52.2 | 14.4 | | | | | | | 49.7 | 59.7 | 74.9 | | |
| Mali | Huybregts, 2019 (longitudinal) (46) | 2.8 | 25.3 | 30.3 | 7.7 | 74.7[a] | 52.3 | 12.1 | 44.4 | 68.0 | 66.3 | 47.8 | | 33.4[b] | 49.6 | 50.8 | 75.3 | | |
| Zimbabwe | Humphrey, 2019 (HIV-unexposed) (47) | 3.8 | 15.5 | 48.9 | 96.3 | 70.0[c] | 50.1 | 27.9* | 46.3 | 41.8 | 48.4 | 29.6 | 19.5 | 18.7[c] | 49.8 | 63.7 | 34.1 | | 92.7 |
| Zimbabwe | Prendergast, 2019 (HIV-exposed) (48) | 4.6 | 18.5 | 24.1 | 93.8 | 70.4[c] | 49.6 | 20.0* | 58.5 | 47.0 | 56.3 | 41.4 | 27.2 | 25.4[c] | 49.8 | 60.8 | 30.1 | | 93.7 |

**Abbreviations**: BMI, body-mass index; LAZ, length-for-age z-score; WLZ, weight-for-length z-score; MUACZ, mid-upper arm circumference z-score; WAZ, weight-for-age z-score; HCZ, head circumference z-score; SES, socio-economic status.

**Notes:**

Maternal depression scales used: a) Edinburgh Postnatal Depression Scale (EPDS) at 6 mo postpartum; b) EPDS at 2 mo postpartum; c) EPDS during pregnancy at enrollment; d) Center for Epidemiological Studies Depression (CESD) Scale at 12 mo postpartum; e) Patient Health Questionnaire (modified) at 24 mo postpartum; f) CESD Scale during pregnancy at

Baseline anthropometry is measured at enrollment into the study or start of supplementation if supplementation did not begin at enrollment.

Food security scales used: a) Food Access Survey Tool; b) Household Food Insecurity Access Scale (HFAIS); c) Coping Strategy Index

Water and sanitation references - as for study-level effect modifiers

Home environment assessed by the Family Care Indictors tool at: a) 18 mo of age; b) 24 mo of age; c) endline survey

Season is defined at time of outcome assessment as a dichotomous "Rainy" vs "Dry" category based on child-specific average rainfall during the month of measurement and 2 months prior.

*Data on birth order were not available for all children. Consequently, first-born vs later-born status was estimated based on the number of children under 5 years old in the household.

**Supplemental Table 4: Growth outcomes at endline among control groups, by trial**

| Country | First author, year | LAZ | Stunted (%) | WLZ | Wasting (%) | MUACZ | Low MUAC (%) | Acute malnutrition (%) | WAZ | Underweight (%) | HCZ | Small head size (%) | WLZ > 1 (%)[1] |
|---|---|---|---|---|---|---|---|---|---|---|---|---|---|
| Bangladesh | Christian, 2015 (34) | -1.9 ± 1.0 | 44.2 | -1.2 ± 0.9 | 16.4 | -1.1 ± 0.8 | 15.2 | 21.1 | -1.8 ± 1.0 | 39.2 | -1.4 ± 0.9 | 24.0 | 0.6 |
| Bangladesh | Dewey, 2017 (35) | -1.8 ± 0.9 | 42.0 | -1.1 ± 0.9 | 15.1 | -0.7 ± 0.8 | 5.2 | 15.3 | -1.7 ± 0.9 | 37.8 | -1.9 ± 0.9 | 42.9 | 0.9 |
| Bangladesh | Luby, 2018 (36) | -1.8 ± 1.0 | 41.9 | -0.9 ± 1.0 | 11.2 | | | | -1.6 ± 1.0 | 32.4 | -1.6 ± 0.9 | 32.8 | 2.6 |
| Burkina Faso | Hess, 2015 (37) | -1.8 ± 1.2 | 39.4 | -0.9 ± 1.0 | 13.5 | -1.1 ± 1.0 | 18.3 | 19.2 | -1.5 ± 1.1 | 30.9 | -1.8 ± 0.9 | 41.0 | 2.7 |
| Burkina Faso | Becquey, 2019 (38) | -1.4 ± 1.0 | 28.5 | -0.8 ± 1.0 | 10.1 | -0.8 ± 0.9 | 10.2 | 14.2 | -1.2 ± 1.0 | 21.2 | | | 2.8 |
| Ghana | Adu Afarwuah, 2007 (39) | -0.4 ± 1.1 | 7.3 | -0.6 ± 1.2 | 9.4 | | | | -0.7 ± 1.1 | 11.5 | -0.6 ± 0.9 | 6.3 | 6.3 |
| Ghana | Adu Afarwuah, 2016 (40) | -0.9 ± 1.0 | 13.0 | -0.6 ± 1.0 | 7.5 | -0.5 ± 0.9 | 3.8 | 8.1 | -0.9 ± 1.0 | 13.0 | -1.1 ± 0.9 | 16.0 | 4.5 |
| Haiti | Iannotti, 2014 (41) | -0.7 ± 1.2 | 13.4 | 0.0 ± 1.0 | 1.3 | | | | -0.4 ± 1.1 | 4.7 | | | 17.4 |
| Kenya | Null, 2018 (42) | -1.6 ± 1.1 | 32.2 | 0.1 ± 0.9 | 1.5 | -0.5 ± 1.0 | 5.5 | 3.2 | -0.8 ± 1.0 | 10.0 | -0.3 ± 1.0 | 4.3 | 16.2 |
| Madagascar | Galasso, 2019 (43) | -2.2 ± 1.2 | 58.5 | -0.4 ± 1.0 | 5.8 | -0.7 ± 1.0 | 9.7 | 9.4 | -1.4 ± 1.0 | 26.0 | | | 7.1 |
| Malawi | Ashorn, 2015 (44) | -1.6 ± 1.1 | 34.7 | -0.1 ± 1.0 | 3.4 | 0.1 ± 1.0 | 3.4 | 4.5 | -0.9 ± 1.1 | 11.7 | -0.8 ± 1.0 | 10.8 | 12.4 |
| Malawi | Maleta, 2015 (45) | -1.9 ± 1.2 | 46.5 | -0.2 ± 1.1 | 5.4 | 0.0 ± 1.0 | 2.1 | 5.8 | -1.1 ± 1.1 | 19.9 | -1.1 ± 1.0 | 21.8 | 10.8 |
| Mali | Huybregts, 2019 (46) | -1.6 ± 1.1 | 34.6 | -0.6 ± 1.0 | 7.3 | -0.8 ± 0.9 | 9.2 | 9.6 | -1.3 ± 1.0 | 20.4 | | | 4.1 |
| Zimbabwe | Humphrey, 2019 (47) | -1.6 ± 1.1 | 34.8 | 0.0 ± 1.1 | 2.6 | 0.0 ± 0.9 | 1.5 | 3.4 | -0.8 ± 1.0 | 10.6 | -0.3 ± 1.1 | 5.8 | 16.5 |
| Zimbabwe | Prendergast, 2019 (48) | -2.0 ± 1.2 | 50.0 | 0.0 ± 1.2 | 3.7 | -0.2 ± 0.9 | 1.5 | 4.6 | -0.9 ± 1.1 | 17.1 | -0.6 ± 1.2 | 10.0 | 18.0 |

**Abbreviations**: LAZ, length-for-age z-score; WLZ, weight-for-length z-score; MUACZ, mid-upper arm circumference z-score; WAZ, weight-for-age z-score; HCZ, head circumference z-score.

**Notes:**

Values are mean ± SD or prevalence

[1]Not an outcome, made available for context.

### Supplemental Table 5: Risk of bias assessment in each trial

| Country | Author | Random sequence generation | Allocation concealment | Blinding participants | Outcome assessment[1] | Incomplete outcome | Selective reporting | Other |
|---|---|---|---|---|---|---|---|---|
| Bangladesh | Christian 2015 (34) | low | low | high | high | low | low | low |
| Bangladesh | Dewey 2017 (35) | low | low | high | low | low | low | low |
| Bangladesh | Luby 2018 (36) | low | low | high | high | low | low | low |
| Burkina Faso | Hess 2015 (37) | low | low | high | high | low | low | low |
| Burkina Faso | Becquey 2019 (38) | low | low | high | high | low | low | low |
| Ghana | Adu Afarwuah 2007 (39) | low | low | high | low | low | low | low |
| Ghana | Adu Afarwuah 2016 (40) | low | low | high | low | low | low | low |
| Haiti | Iannotti 2014 (41) | unclear | low | high | high | low | low | low |
| Kenya | Null 2018 (42) | low | low | high | high | low | low | low |
| Madagascar | Galasso 2019 (43) | low | low | high | high | low | low | low |
| Malawi | Ashorn 2015 (44) | low | low | high | low | low | low | low |
| Malawi | Maleta 2015 (45) | low | low | high | low | low | low | low |
| Mali | Huybregts 2019 (46) | low | low | high | high | low | low | low |
| Zimbabwe | Humphrey 2019 (47) | low | low | high | high | low | low | low |
| Zimbabwe | Prendergast 2019 (48) | low | low | high | high | low | low | low |

1. Due to the nature of the intervention, blinding of participants was not possible. We considered anthropometric outcome assessment to be at low risk of bias only when it was clearly specified that data collectors who performed the anthropometric measurements were not aware of group allocation, and it would be unlikely that they could easily become aware of group allocation (i.e. observation of interventions materials in study communities, non-intervention passive control arms, etc.)

| Adu-Afarwuah 2007 (39) | | |
|---|---|---|
| **Bias** | **Authors' judgement** | **Support for judgement** |
| Random sequence generation (selection bias) | Low risk | **Quote:** "we randomly selected ~75% of the total number of eligible infants to enter the intervention trial. This was done on a weekly basis, when infants were 5 mo of age, by entering the identification numbers of the eligible infants in a dataset, and using an SAS data step (ranuni [1] le 0.75) to select those for the intervention…the NI infants were randomly selected from the pool of initially eligible infants"<br>**Comment:** adequately done |
| Allocation concealment (selection bias) | Low risk | **Quote:** "the infants were randomly assigned (with the use of opaque envelopes with group designation) to receive SP, NT, or NB until 12 mo of age"<br>**Comment:** adequately done |
| Blinding of participants and personnel (performance bias) | High risk | **Quote**: "NT was provided to the mothers in plastic bags, and the NB (20 g/d) was provided in foil packs with screw caps"<br>**Comment:** not adequate |
| Blinding of outcome assessment (detection bias) | Low risk | **Personal communication with investigator (SAA):** "All anthropometric measurements were carried out by dedicated anthropometrists separate from the field workers who delivered the supplements in the homes, and the anthropometrists had no knowledge of the group assignments. At both 6 and 12 months of age, all children were brought to the laboratory where the anthropometric measurements were conducted; in this case, it was neither possible for the anthropometrists to determine group assignments within the intervention groups, nor was it likely that the anthropometrists could remember or know which children belonged to the intervention versus non-intervention groups."<br>**Comment:** adequately done |
| Incomplete outcome data (attrition bias) | Low risk | **Attrition**: SP group = 98/105; NT group = 102/105; NB group = 98/103; Control group = 96/97<br>**Comment:** Minimal attrition and reasons given for loss to follow-up |
| Selective reporting (reporting bias) | Low risk | **Comment**: The trial was registered at clinicaltrials.gov (NCT00379158); outcomes described in the methods section reported in the results section |
| Other bias | Low risk | **Comment**: no other potential sources of bias reported |
| **Funding**<br>Supported by the Nestlé Foundation with additional support from USAID's MGL Research Program through ILSI. | | |

| **Adu-Afarwuah 2016 (40)** | | |
|---|---|---|
| **Bias** | **Authors' judgement** | **Support for judgement** |
| Random sequence generation (selection bias) | Low risk | **Quote**: "The study statistician at University of California, Davis developed group allocations with the use of a computer generated (SAS version 9.3; SAS Institute) randomization scheme in blocks of 9"<br>**Comment:** adequately done |
| Allocation concealment (selection bias) | Low risk | **Quote**: "At each enrollment, the study nurse offered sealed, opaque envelopes bearing group allocations, 9 envelopes at a time, and the woman picked one to reveal the allocation"<br>**Comment:** adequately done |
| Blinding of participants and personnel (performance bias) | High risk | **Quote:** "It was not possible to blind study workers and participants to the capsules (IFA and MMN supplements) compared with the LNS supplements because of their different appearances" |
| Blinding of outcome assessment (detection bias) | Low risk | **Quote**: "It was not possible to blind study workers and participants to the capsules (IFA and MMN supplements) compared with the LNS supplements because of their apparent differences, but laboratory staff, anthropometrists, and data analysts had no knowledge of group assignment until all preliminary analyses had been completed"<br>**Comment:** adequately done |
| Incomplete outcome data (attrition bias) | Low risk | **Attrition:** IFA group = 393/408; MMN group = 401/411; LNS group = 391/409<br>**Comment:** Minimal attrition and reasons given for loss to follow-up |
| Selective reporting (reporting bias) | Low risk | **Comment**: The trial was registered at clinicaltrials.gov (NCT00970866); SAP available online; outcomes described in the methods section reported in the results section |
| Other bias | Low risk | **Comment**: no other potential sources of bias reported |
| **Funding**<br>Funded by a grant to the University of California, Davis, from the Bill & Melinda Gates Foundation. | | |

| Ashorn 2015 (44) | | |
|---|---|---|
| **Bias** | **Authors' judgement** | **Support for judgement** |
| Random sequence generation (selection bias) | Low risk | **Quote:** "Researcher not involved with the trial created individual randomisation slips (in blocks of 9)"<br>**Comment**: Additional details on randomization provided in Ashorn *et al.* AJCN 2015. |
| Allocation concealment (selection bias) | Low risk | **Quote**: "…packed them in sealed, numbered, opaque randomization envelopes that were stored in numerical order…Eligible pregnant women were requested to choose 1 of the top 6 envelopes in the stack, and the contents of the envelope indicated her participant number and group allocation""<br>**Comment:** adequately done |
| Blinding of participants and personnel (performance bias) | High risk | **Quote:** "The IFA and MMN interventions were provided with double-masked procedures…For the LNS group, we used single-masked procedures; that is field workers who delivered the supplements knew which mothers were receiving LNS, and the participants were advised not to disclose information about their supplements to anyone other than an iLiNS team member"<br>**Comment:** not done |
| Blinding of outcome assessment (detection bias) | Low risk | **Quote:** "The data collectors who performed the anthropometric measurements or assessed other outcomes were not aware of group allocation. Researchers responsible for the data cleaning remained blind to the trial code, until the database was considered fully cleaned"<br>**Comment**: adequately done |
| Incomplete outcome data (attrition bias) | Low risk | **Attrition:** IFA group = 220/223; MMN group = 222/233; LNS group = 214/222<br>**Comment:** Minimal attrition and reasons given for loss to follow-up |
| Selective reporting (reporting bias) | Low risk | **Comment**: The trial was registered at clinicaltrials.gov ( NCT01239693 ); SAP available online; outcomes described in the methods section reported in the results section |
| Other bias | Low risk | **Comment:** no other potential sources of bias reported |

**Funding**

Supported in part by a grant to the University of California, Davis, from the Bill & Melinda Gates Foundation, with additional funding from the Office of Health, Infectious Diseases, and Nutrition, Bureau for Global Health, US Agency for International Development (USAID) under terms of cooperative agreement AID-OAAA-1200005, through the Food and Nutrition Technical Assistance III Project (FANTA), managed by FHI 360. For data management and statistical analysis, the team received additional support from the Academy of Finland grant 252075 and the Medical Research Fund of Tampere University Hospital grant 9M004. YBC was supported by the Singapore Ministry of Health's National Medical Council under its Clinician Scientist Award.

| Becquey 2019 (38) | | |
|---|---|---|
| **Bias** | **Authors' judgement** | **Support for judgement** |
| Random sequence generation (selection bias) | Low risk | **Quote:** "simple(i.e. non-stratified) random allocation was used"; "randomization took place at a community event…with local health authorities"; "32 identical pieces of paper with either 'control' or 'intervention' written on them were mixed in a bag for randomization"<br>"In each health center catchment area, a census was conducted 1 month prior to the cross-sectional baseline and endline surveys to identify all pregnant women and eligible children. A random sample of children was drawn from the census list."<br>**Comment**: adequately done |
| Allocation concealment (selection bias) | Low risk | **Quote:** "32 identical pieces of paper with either 'control' or 'intervention' written on them were mixed in a bag for randomization"<br>**Comment**: adequately done |
| Blinding of participants and personnel (performance bias) | High risk | **Quote:** "non-masked, community-based, trial"<br>**Comment**: blinding of participants who received no intervention was not possible |
| Blinding of outcome assessment (detection bias) | High risk | **Quote:** "A two-arm, cluster-randomized, non-blinded, effectiveness trial"<br>"All anthropometric measurements and checking of bilateral pitting edema were systematically performed by a trained and standardized anthropometrist with the help of a trained assistant."<br>**Comment:** Non-blinded trial |
| Incomplete outcome data (attrition bias) | Low risk | **Attrition:** 21.2% loss to follow-up (7.2% exited study one month early due to misunderstanding by study team, remained of attrition (14% of study sample) primarily due to moving, child death.<br>**Quote**: "the proportion of enrolled children lost to follow-up was balanced across study groups"<br>"We conducted multiple imputation of missing longitudinal outcome data using a 2-fold fully conditional specification algorithm, which imputes missing values under the missing at random assumption, respecting the temporal ordering of observations".<br>**Comment:** reasons provided for loss to follow-up; missing outcome data balanced in numbers across intervention groups; multiple imputation procedures used |
| Selective reporting (reporting bias) | Low risk | **Comment**: trial registered as NCT02245152 at ClinicalTrials.gov, published protocol (Huybregts BMC Public Health 2017); outcomes described in the methods section reported in the results section; data made available to IPD investigators. |
| Other bias | Low risk | **Comment**: no other potential sources of bias reported |
| **Funding**<br>The PROMIS studies were funded by Global Affairs Canada (GAC) (https://www.international.gc.ca/); grant 52308/5252/0200 and CGIAR Agriculture for Nutrition and Health (A4NH) program (http://a4nh.cgiar.org/) led by the International Food Policy Research Institute (IFPRI), with no role by either funder in study design, data collection and analysis, decision to publish, or preparation of the manuscript. | | |

| **Christian 2015 (34)** | | |
|---|---|---|
| **Bias** | **Authors' judgement** | **Support for judgement** |
| Random sequence generation (selection bias) | Low risk | **Quote:** "A random-number seed was selected by a statistician not involved in the study, using a random number generator, and a random number between 0 and 1 drawn from a uniform distribution was assigned to each sector"<br>**Comment:** adequately done |
| Allocation concealment (selection bias) | Low risk | **Quote: "**Cluster-randomization of the 596 predefined communities in JiVitA, called 'sectors', was done by blocks of 19 (total 32 blocks, last block had 7 sectors). A random-number seed was selected by a statistician not involved in the study, using a random number generator, and a random number between 0 and 1 drawn from a uniform distribution was assigned to each sector. Additionally, a block number was assigned to each sector in groups of 19. For blocks 1–31, the first five sectors by sort order were assigned to treatment group 1, the next five to treatment group 2, and so on. For block 32, the two larger controls were assigned two sectors and the intervention groups 1 sector each.<br>**Comment**: central randomization of a cluster-randomized trial |
| Blinding of participants and personnel (performance bias) | High risk | **Quote:** "Our trial was unblinded"<br>**Comment:** not done |
| Blinding of outcome assessment (detection bias) | High risk | **Quote:** "Our trial was unblinded"; "Anthropometry was done at this time using standard methods…Repeat infant anthropometry assessments were conducted during home visits every 3 months after the baseline at 9, 12, 15 and 18 months of age."<br>**Comment**: not done; cluster-randomized trial |
| Incomplete outcome data (attrition bias) | Low risk | **Attrition:** control group = 1312/1591; Plumpy'Doz group = 1395/1599; rice lentil group = 785/901; chickpea group = 786/920; WSB++ group = 789/928<br>**Comments**: reasons given for loss to follow-up; missing outcome data balanced in numbers across intervention groups |
| Selective reporting (reporting bias) | Low risk | **Comment:** trial registered as NCT01562379 at ClinicalTrials.gov, outcomes described in the methodology section reported in the results section |
| Other bias | Low risk | **Comment:** no other potential sources of bias reported |
| **Funding**<br>The JiVitA-4 study was funded by the US Department of Agriculture, NIFA under the FANEP [Award no. 2010-38418-21732]. In kind support in the form of micronutrient premix for the local food supplements was provided by DSM, Basel, Switzerland and Plumpy'doz was provided by Nutriset (Malaunay, France). | | |

| Dewey 2017 (35) | | |
|---|---|---|
| **Bias** | **Authors' judgement** | **Support for judgement** |
| Random sequence generation (selection bias) | Low risk | **Quote:** "For the randomization, the study statistician at UCD first stratified all 64 clusters in the 11 unions by subdistrict and union and then randomly assigned each cluster to 1 of the 4 arms (each containing 16 clusters)"<br>**Comment:** adequately done |
| Allocation concealment (selection bias) | Low risk | **Quote:** "For the randomization, the study statistician at UCD first stratified all 64 clusters in the 11 unions by subdistrict and union and then randomly assigned each cluster to 1 of the 4 arms (each containing 16 clusters)"<br>**Comment**: central randomization of a cluster-randomized trial |
| Blinding of participants and personnel (performance bias) | High risk | **Comment**: participant blinding not possible due to the nature of the intervention (LNS, MNP, Control) |
| Blinding of outcome assessment (detection bias) | Low risk | **Quote:** "The trial was a researcher-blind, longitudinal, cluster randomized effectiveness trial"; "SDU team members conducting anthropometric measurements were not aware of group assignment"<br>**Comment:** adequately done |
| Incomplete outcome data (attrition bias) | Low risk | **Attrition**: LNS-LNS = 884/1047; IFA-LNS = 785/930; IFA-MNP = 895/1052; IFA-Control = 816/982<br>**Comment:** reasons given for loss to follow-up; missing outcome data balanced in numbers across intervention groups |
| Selective reporting (reporting bias) | Low risk | **Comment:** The trial was registered at ClinicalTrials.gov ( NCT01715038 ); outcomes described in the methods section reported in the results section |
| Other bias | Low risk | **Comment:** no other potential sources of bias reported |

**Funding**
Supported by the Office of Health, Infectious Diseases, and Nutrition, Bureau for Global Health, US Agency for International Development (USAID) under the terms of cooperative agreement AID-OAA-A-12-00005, through the Food and Nutrition Technical Assistance III Project (FANTA), managed by FHI 360. Our research intervention was incorporated into the community heath and development program of LAMB, which was supported by Plan-Bangladesh in 6 of the 11 study unions. Nutriset S.A.S. prepared the lipid-based nutrient supplements for this trial, and Hudson Pharmaceuticals Ltd. prepared the iron and folic acid tablets.

| **Hess 2015 (37)** | | |
|---|---|---|
| **Bias** | **Authors' judgement** | **Support for judgement** |
| Random sequence generation (selection bias) | Low risk | **Quote:** "computer-generated an assignment within strata to participate in the intervention cohort ... The same statistician, who was blinded to the intervention, generated a random allocation sequence at the level of the concession for the enrollment of eligible infants in the intervention cohort"<br>**Comment:** adequately done |
| Allocation concealment (selection bias) | Low risk | **Quote: "**The same statistician, who was blinded to the intervention, generated a random allocation sequence at the level of the concession for the enrollment of eligible infants in the intervention cohort"<br>**Comment:** central randomization of a cluster-randomized trial |
| Blinding of participants and personnel (performance bias) | High risk | **Quote:** "The trial was partially masked, as all participants, field staff and researchers remained blinded to the four intervention groups until data analyses were completed, but were aware which communities were assigned to intervention cohort and non-intervention cohort"<br>**Comment:** intervention and non-intervention cohorts non-blinded |
| Blinding of outcome assessment (detection bias) | High risk | **Quote:** "The trial was partially masked, as all participants, field staff and researchers remained blinded to the four intervention groups until data analyses were completed, but were aware which communities were assigned to intervention cohort and non-intervention cohort."<br>**Comment:** IC and NIC non-blinded |
| Incomplete outcome data (attrition bias) | Low risk | **Attrition:** LNS-Zn0 group = 489/602; LNS-Zn5 group = 499/613; LNS-Zn10 group = 491/603; LNS-TabZn5 group = 481/617; NIC group = 666/785<br>**Comment:** reasons for loss to follow-up mentioned |
| Selective reporting (reporting bias) | Low risk | **Comment**: Protocol attached as a supplement in the study paper; registered at ClinicalTrials.gov as NCT 00944281; outcomes described in the methods section reported in the results section |
| Other bias | Low risk | **Comment:** no other potential sources of bias reported |

**Funding**
The project was funded by a grant from the Bill & Melinda Gates Foundation to the University of California, Davis. The funder had no role in study design, data collection and analysis, decision to publish, or preparation of the manuscript.

| **Humphrey 2019 and Prendergast 2019 (47, 48)** | | |
|---|---|---|
| **Bias** | **Authors' judgement** | **Support for judgement** |
| Random sequence generation (selection bias) | Low risk | **Quote:** "clusters were allocated (1:1:1:1) to one of four treatment groups"; "the study's statistician used a constrained randomization technique to identify 500 allocation schemes…From these, 10 allocations were randomly selected. The final allocation was selected at a public randomization event attended by elected representatives"<br>**Comments:** Additional details available in Supplementary Materials (Appendix) and at https://osf.io/w93hy and in (SHINE Trial Team, Clin Infect Dis, 2015). |
| Allocation concealment (selection bias) | Low risk | **Quote:** "the study's statistician used a constrained randomization technique to identify 500 allocation schemes…From these, 10 allocations were randomly selected. The final allocation was selected at a public randomization event attended by elected representatives"<br>**Comments:** Additional details available at https://osf.io/w93hy and in (SHINE Trial Team, Clin Infect Dis, 2015). |
| Blinding of participants and personnel (performance bias) | High risk | **Quote**: "masking of participants and fieldworkers was not possible because of the obvious visual differences between interventions"<br>**Comment:** not done |
| Blinding of outcome assessment (detection bias) | High risk | **Quote**: "masking of participants and fieldworkers was not possible because of the obvious visual differences between interventions, but investigators were blinded to treatment groups until the final analysis of each pre-specified outcome."<br>**Comment:** not done |
| Incomplete outcome data (attrition bias) | Low risk | **Comment:** attrition similar across all seven arms with reasons given |
| Selective reporting (reporting bias) | Low risk | **Comment:** trial registered as NCT01824940 at ClinicalTrials.gov, published protocol (SHINE Trial Team, Clin Infect Dis 2015), research and statistical analysis plan available at https://osf.io/w93hy ; outcomes described in the methods section reported in the results section |
| Other bias | Low risk | **Comment**: no other potential sources of bias reported |
| **Funding**<br>The SHINE trial is funded by the Bill & Melinda Gates Foundation (OPP1021542 to Johns Hopkins Bloomberg School of Public Health and OPP1143707 to Zvitambo Institute for Maternal and Child Health Research), the UK Department for International Development, the Wellcome Trust (093768/Z/10/Z and 108065/Z/15/Z), the Swiss Agency for Development and Cooperation (8106727), UNICEF (PCA-2017-0002), and the US National Institutes of Health (R01 HD060338/HD/NICHD). | | |

| **Huybregts 2019 (46)** | | |
|---|---|---|
| **Bias** | **Authors' judgement** | **Support for judgement** |
| Random sequence generation (selection bias) | Low risk | **Quote**: "we applied stratified random allocation of the HC catchment areas to control and intervention study groups"; "we first stratified the [health centers] by hierarchical clustering"; "random allocation to control or intervention groups was conducted within each stratum during a community ceremony…forty-eight identical pieces of paper with either 'control' or 'intervention' were mixed in a bag…each [health center] director drew one piece of paper"<br>**Comment:** adequately done |
| Allocation concealment (selection bias) | Low risk | "random allocation to control or intervention groups was conducted within each stratum during a community ceremony…forty-eight identical pieces of paper with either 'control' or 'intervention' were mixed in a bag…each [health center] director drew one piece of paper"<br>**Comment**: adequately done |
| Blinding of participants and personnel (performance bias) | High risk | **Quote**: "non-masked, community-based, trial"<br>**Comment**: blinding of participants who received no intervention was not possible |
| Blinding of outcome assessment (detection bias) | High risk | **Quote:** "We used a two-arm, cluster-randomized, non-blinded effectiveness trial"<br>**Comment**: not done, cluster randomized trial at level of HC |
| Incomplete outcome data (attrition bias) | Low risk | **Attrition:** LNS group = 494/577; Control group = 488/577<br>**Quote**: "A total of 150 (13%) children were lost to follow-up with similar attrition patterns for the intervention and control group"; "we conducted multiple imputations of missing longitudinal continuous and binary outcome data using a 2-fold fully conditional specification (FCS) algorithm".<br>**Comment**: reasons provided for loss to follow-up; missing outcome data balanced in numbers across intervention groups; multiple imputation procedures used |
| Selective reporting (reporting bias) | Low risk | **Comment**: trial registered as NCT02323815 at ClinicalTrials.gov, published protocol (Huybregts BMC Public Health 2017); outcomes described in the methods section reported in the results section; data made available to IPD investigators. |
| Other bias | Low risk | **Comment**: no other potential sources of bias reported |
| **Funding**<br>The PROMIS studies were funded by: Global Affairs Canada (GAC) (https://www.international.gc.ca/); grant 52308/5252/0200, and CGIAR Agriculture for Nutrition and Health (A4NH) program (http://a4nh.cgiar.org/) led by the International Food Policy Research Institute (IFPRI), with no role by either funder in study design, data collection and analysis, decision to publish, or preparation of the manuscript. | | |

| **Iannotti 2014 (41)** | | |
|---|---|---|
| **Bias** | **Authors' judgement** | **Support for judgement** |
| Random sequence generation (selection bias) | Unclear risk | **Quote:** "Random assignment was carried out through an allocation-concealment mechanism whereby sealed paper forms that masked group assignments were drawn from a container by mothers by using a simple random assignment ratio of 1:1:1 for group assignments"<br>**Comment:** randomization procedure not specified |
| Allocation concealment (selection bias) | Low risk | **Quote:** "Random assignment was carried out through an allocation-concealment mechanism whereby sealed paper forms that masked group assignments were drawn from a container by mothers by using a simple random assignment ratio of 1:1:1 for group assignments"<br>**Comment:** adequately done |
| Blinding of participants and personnel (performance bias) | High risk | **Comment**: participant blinding not possible due to the nature of the intervention (LNS, Control) |
| Blinding of outcome assessment (detection bias) | High risk | **Quote:** "The study team was comprised of one study coordinator and 3 enumerators who participated in a 1-wk training session at the beginning of the trial and another refresher training midway through covering the protocol of anthropometric measures, survey administration, and ethics."<br>**Comment:** same enumerators conducting anthropometric assessments as providing supplements |
| Incomplete outcome data (attrition bias) | Low risk | **Attrition:** control group = 156/191; 6-month LNS group = 159/202<br>**Comment:** reasons for loss-to-follow-up mentioned |
| Selective reporting (reporting bias) | Low risk | **Comment**: Trial registered at ClinicalTrials.gov (NCT01552512); outcomes described in the methodology section reported in the results section |
| Other bias | Low risk | **Comment**: no other potential sources of bias reported |
| **Funding**<br>Supported by the Bill & Melinda Gates Foundation to FHI 360 through the Alive & Thrive Small Grants Program managed by University of California Davis; The Inter-American Development Bank; The World Bank; and The United Nations World Food Program. | | |

| **Luby 2018 (36)** | | |
|---|---|---|
| **Bias** | **Authors' judgement** | **Support for judgement** |
| Random sequence generation (selection bias) | Low risk | **Quote**: Clusters were randomly allocated to treatment using a random number generator by a coinvestigator at University of California, Berkeley (BFA). Each of the eight geographically adjacent clusters was block randomized to the double-sized control arm or one of the six interventions…Geographical matching ensured that arms were balanced across locations and time of measurement."<br>**Comment**: adequately done |
| Allocation concealment (selection bias) | Low risk | **Quote** "Clusters were randomly allocated to treatment using a random number generator by a coinvestigator at University of California, Berkeley (BFA)."<br>**Comment**: central randomization of a cluster-randomized trial |
| Blinding of participants and personnel (performance bias) | High risk | **Comment:** Not done due to the nature of the intervention |
| Blinding of outcome assessment (detection bias) | High risk | **Quote: "**Interventions included distinct visible components so neither participants nor data collectors were masked to intervention assignment, although the data collection and intervention teams were different individuals".<br>"Outcome and adherence was assessed by a team of university graduates who were not involved in the delivery or promotion of interventions."<br>**Comment**: passive control arm; blinding not possible due to the nature of the intervention |
| Incomplete outcome data (attrition bias) | Low risk | **Comment:** attrition similar across all seven arms with reasons given |
| Selective reporting (reporting bias) | Low risk | **Comment** The trial was registered at clinicaltrials.gov (NCT 01590095); SAP and trial protocol available, and published (Arnold BMJ Open 2013); outcomes described in the methods section reported in the results section |
| Other bias | Low risk | **Comment:** no other potential sources of bias reported |
| **Funding**<br>Supported by a global development grant (OPPGD759) from the Bill & Melinda Gates Foundation to the University of California, Berkeley, CA, USA. | | |

| **Maleta 2015 (45)** | | |
|---|---|---|
| **Bias** | **Authors' judgement** | **Support for judgement** |
| Random sequence generation (selection bias) | Low risk | **Quote:** "We used block randomization and a set of opaque envelopes to assign participants to the intervention groups. The randomization list and envelopes were prepared by a study statistician not involved in trial implementation"<br>**Comment:** adequately done |
| Allocation concealment (selection bias) | Low risk | **Quote:** "We used block randomization and a set of opaque envelopes to assign participants to the intervention groups."<br>**Comment:** adequately done |
| Blinding of participants and personnel (performance bias) | High risk | **Comment**: participant blinding not possible due to the nature of the intervention (LNS, Control) |
| Blinding of outcome assessment (detection bias) | Low risk | **Quote:** "…and the code was not disclosed to the researchers or to those assessing the outcomes until all data had been entered and verified in a database." "For the LNS group, we used single-masked procedures (i.e., fieldworkers who delivered the supplements knew which children were receiving LNSs, but those who performed the anthropometric measurements or assessed other outcomes were not aware of group allocation)."<br>**Comment:** adequately done |
| Incomplete outcome data (attrition bias) | Low risk | **Attrition:** 40g/day milk-free LNS group =239/324; 40g/day milk LNS group = 242/322; 20g/day milk-free LNS group = 247/323; 20g/day milk LNS group = 236/322;10g/day milk LNS group = 221/321; control group = 242/320<br>**Comment**: similar levels of attrition across groups, reasons for dropout provided |
| Selective reporting (reporting bias) | Low risk | **Comment**: The trial was registered at clinicaltrials.gov ( NCT00945698); SAP and trial protocol available online; outcomes described in the methods section reported in the results section |
| Other bias | Low risk | **Comment:** no other potential sources of bias reported |
| **Funding**<br>Funded by a grant to the University of California, Davis, from the Bill & Melinda Gates Foundation. | | |

| **Null 2018 (42)** | | |
|---|---|---|
| **Bias** | **Authors' judgement** | **Support for judgement** |
| Random sequence generation (selection bias) | Low risk | **Quote:** "Clusters were randomly allocated to treatment at the University of California, Berkeley using a random number generator with reproducible seed"<br>**Comment:** adequately done |
| Allocation concealment (selection bias) | Low risk | **Quote:** "Clusters were randomly allocated to treatment at the University of California, Berkeley using a random number generator with reproducible seed"<br>**Comment**: central randomization of a cluster-randomized trial |
| Blinding of participants and personnel (performance bias) | High risk | **Quote:** "Masking participants was not possible"<br>**Comment:** blinding of participants was not possible due to the nature of the intervention |
| Blinding of outcome assessment (detection bias) | High risk | **Quote**: "The health promoters and staff who delivered the interventions were not involved in data collection, but the data collection team could have inferred treatment status if they saw intervention materials in study communities."<br>**Comment:** blinding not possible due to the nature of the intervention |
| Incomplete outcome data (attrition bias) | Low risk | **Comment:** attrition similar across all seven arms with reasons given |
| Selective reporting (reporting bias) | Low risk | **Comment: T**rial registered as<br>NCT01704105 at ClincialTrials.gov. SAP and trial protocol available online, and published (Arnold BMJ Open 2013); outcomes described in the methods section reported in the results section |
| Other bias | Low risk | **Comment:** no other potential sources of bias reported |
| **Funding**<br>Supported in part by Global Development grant OPPGD759 from the Bill & Melinda Gates Foundation to the University of California, Berkeley, CA, USA, and grant AID-OAA-F-13-00040 from United States Agency for International Development (USAID) to Innovations for Poverty Action. This manuscript was made possible by the generous support of the American people through the USAID. The contents are the responsibility of the authors and do not necessarily reflect the views of USAID or the US Government. | | |

| **Galasso 2019 (43)** | | |
|---|---|---|
| **Bias** | **Authors' judgement** | **Support for judgement** |
| Random sequence generation (selection bias) | Low risk | **Quote: "**a random generator was used to block-randomise five sites per intervention group per region"… "An up-to-date registry of government-programme eligible women and children… was used as a sampling frame to select households eligible for enrolment in the trial. 30 households were randomly sampled per site, stratified by children's age at baseline"<br>**Comment:** adequately done |
| Allocation concealment (selection bias) | Low risk | **Comment:** central randomization of a cluster-randomized trial |
| Blinding of participants and personnel (performance bias) | High risk | **Quote:** "Due to the nature of the interventions, masking of participants and community health workers was not possible."<br>**Comment:** not done |
| Blinding of outcome assessment (detection bias) | High risk | **Quote:** "Due to the nature of the interventions, masking of participants and community health workers was not possible. Data analysts were not blinded to intervention group assignment due to differences in survey information"<br>**Comment:** not done |
| Incomplete outcome data (attrition bias) | Low risk | **Quote:** "Mothers or children who died before the final assessment were not replaced. Children who had permanently moved outside the programme site catchment area before final assessment were replaced with a randomly drawn child from the site within the same age range. Children and their households who returned to the site between the baseline and final assessment were re-interviewed."<br>**Comment**: similar levels of attrition across groups, reasons for dropout provided |
| Selective reporting (reporting bias) | Low risk | **Quote:** "This trial has been registered with the ISRCTN registry, number ISRCTN14393738."<br>**Comment:** published protocol (Fernald BMC Public Health 2016); outcomes described in the methods section reported in the results section |
| Other bias | Low risk | **Comment:** no other potential sources of bias reported |
| **Funding**<br>Funded by the Eunice Kennedy Shriver National Institutes of Child Health and Human Development, Strategic Impact Evaluation Fund, Early Learning Partnership Program, World Bank Innovation Grant, Grand Challenges Canada, World Bank Research Committee, Japan Nutrition Trust Fund, Power of Nutrition Trust Fund. | | |
