## Supplemental Figures 1-7 for "Characteristics that modify the effect of small-quantity lipid-based nutrient supplementation on child growth: an individual participant data meta-analysis of randomized controlled trials"

Supplemental Figure 1: Summary risk of bias as a percentage of all included studies for the effects of SQ-LNS on growth outcomes

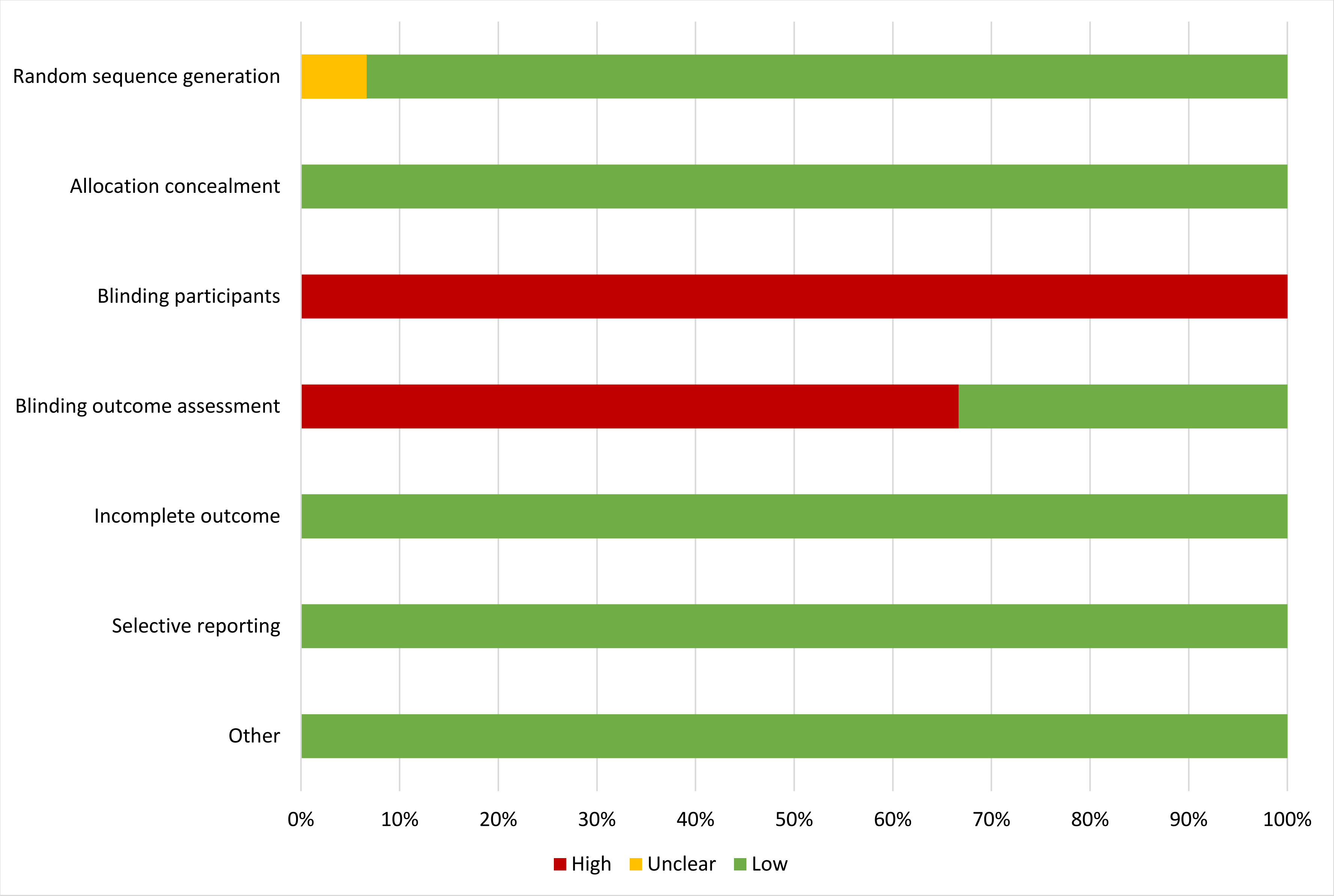

### Supplemental figure 2: Sensitivity analyses of main effects of SQ-LNS on growth outcomes

#### Contents

**Supplemental figure 2A: Mean differences for continuous outcomes** **2**

**Supplemental figure 2B: Prevalence ratios for dichotomous outcomes** **3**

**Supplemental figure 2C: Prevalence differences for dichotomous outcomes** **4**

These figures show the pooled estimates of intervention effects by different pooling methods and different sensitivity analyses. For continuous outcomes, the intervention effect is measured by the difference in mean of the LNS group minus control. For dichotomous outcomes analyzed via prevalence ratios, the effect estimate is the prevalence in the LNS group divided by the prevalence in the control group. For dichotomous outcomes analyzed via prevalence differences, the effect estimate is the prevalence in the LNS group minus the prevalence in the control group. The labels on the left y-axis indicate which outcome is assessed. The different columns correspond to sensitivity analyses in which intervention group categorization differs. All-trial analysis includes all trials; Child-LNS-only excludes trial arms that provided both maternal and child LNS; Multi-component analysis separates comparisons within trials that included multi-component interventions, so that the SQ-LNS vs. no SQ-LNS comparisons were conducted separately between pairs of arms that included the same non-nutrition components (e.g. SQ-LNS+WASH vs. WASH; SQ-LNS vs. Control); Passive arms excluded analysis excludes passive control arms; Milk-peanut LNS only analysis excludes arms with SQ-LNS formulations that were not milk and peanut based. LAZ, length-for-age z-score; WLZ, weight-for-length z-score; WAZ, weight-for-age z-score; MUACZ, midupper arm circumference z-score; HCZ, head circumference-for-age z-score.

#### Supplemental figure 2A: Mean differences for continuous outcomes

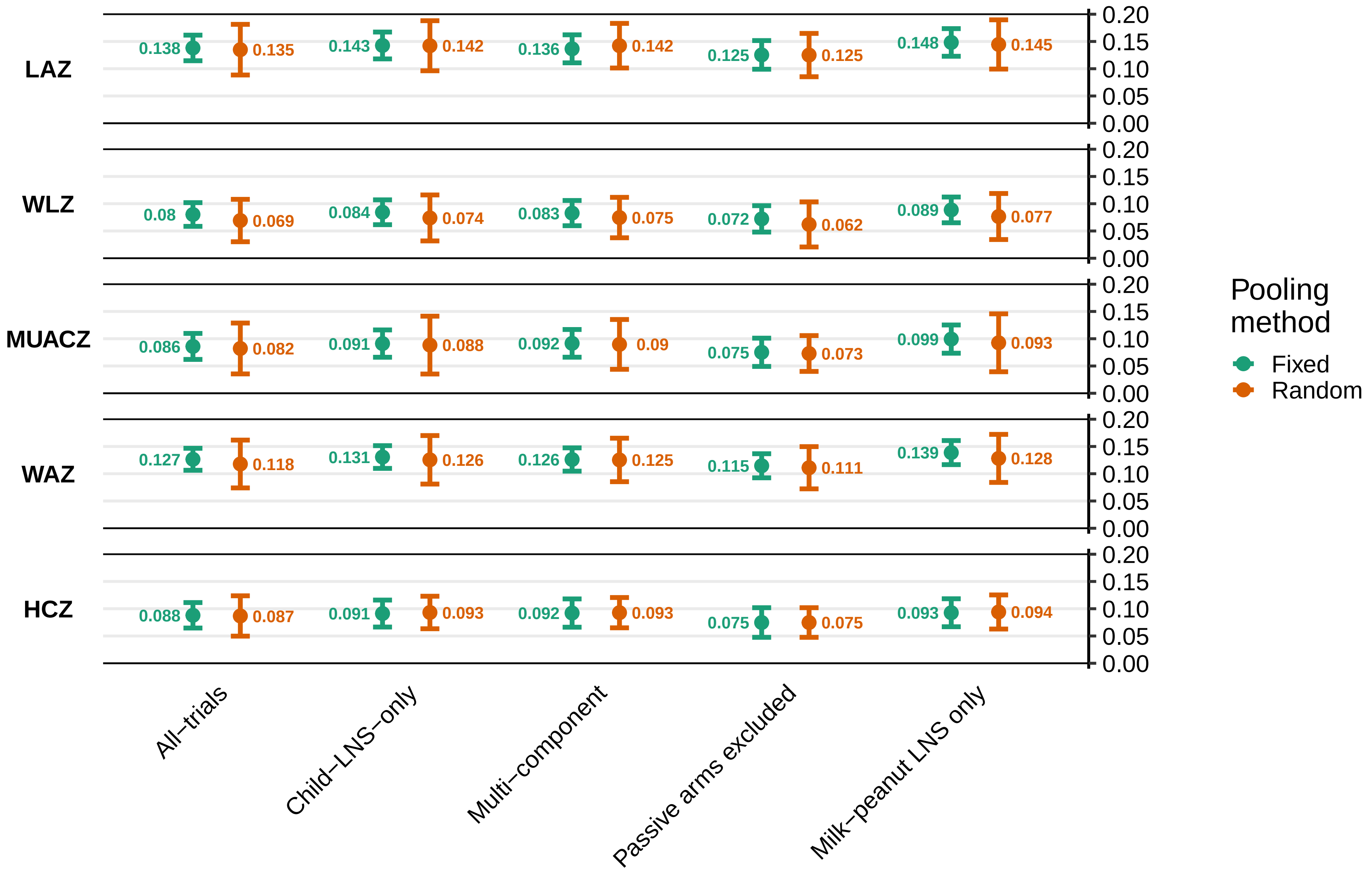

#### Supplemental figure 2B: Prevalence ratios for dichotomous outcomes

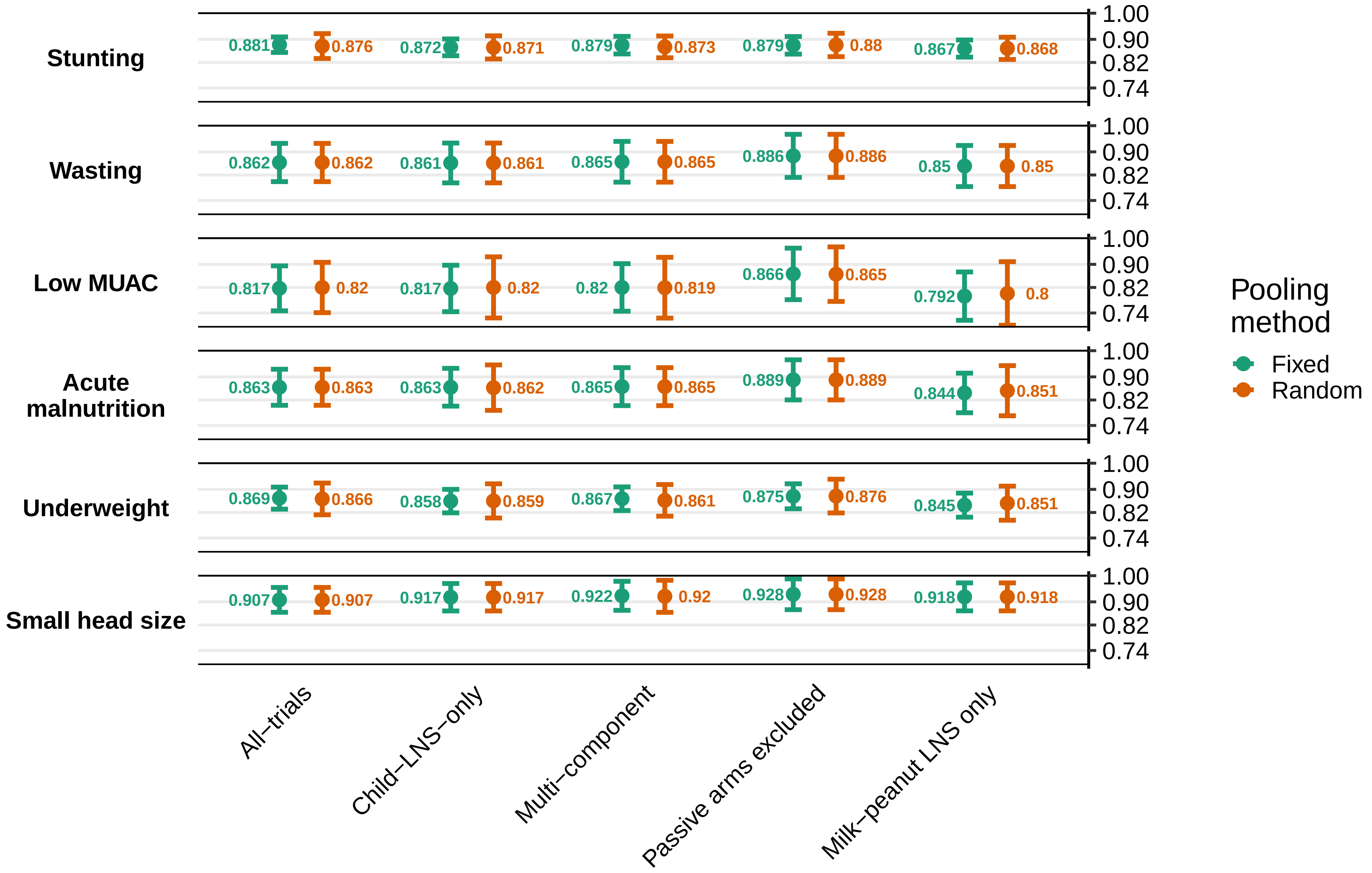

#### Supplemental figure 2C: Prevalence differences for dichotomous outcomes

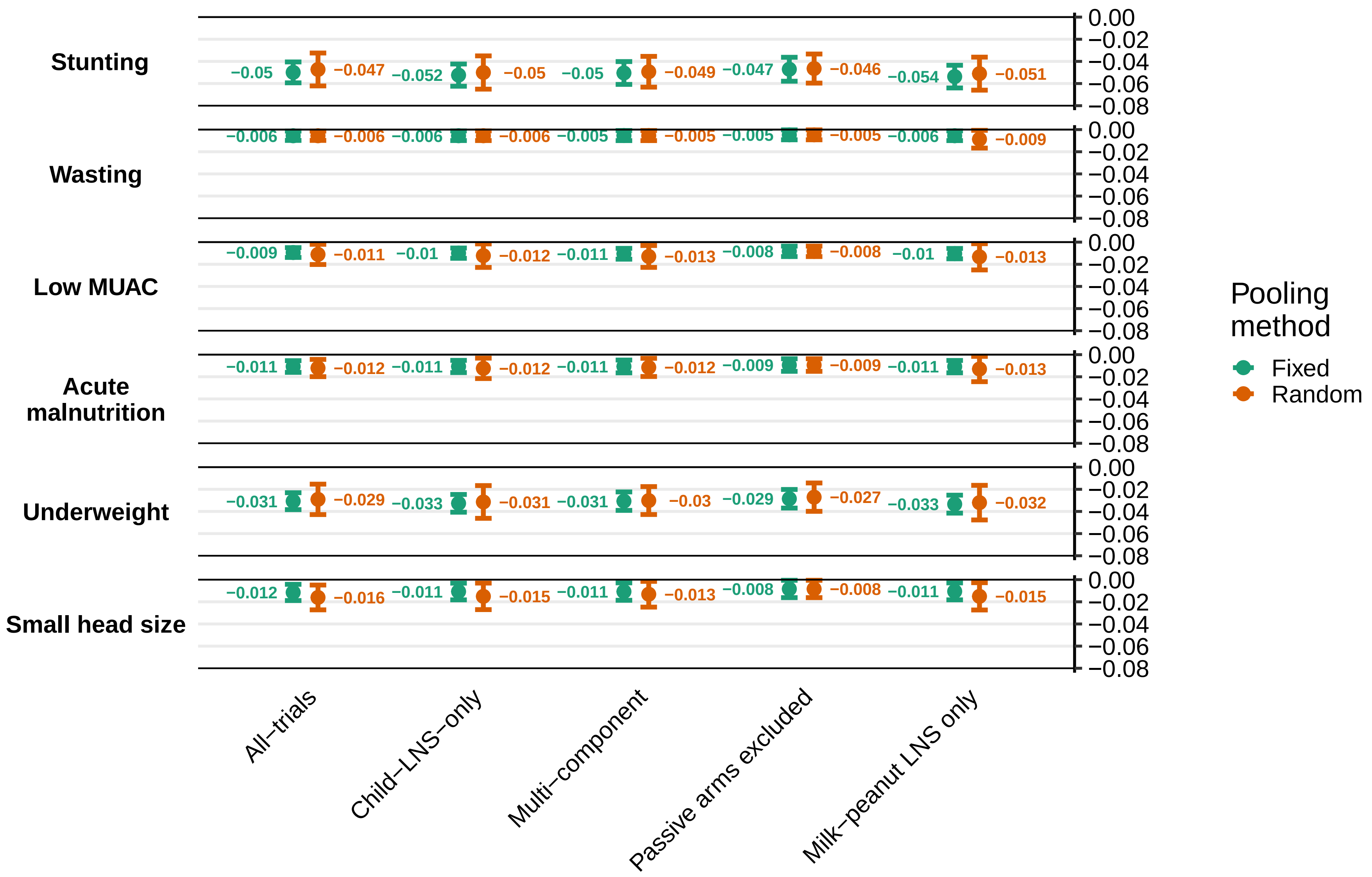

### Supplemental figure 3: Forest plots for all main effects of SQ-LNS on growth outcomes

#### Contents

**Supplemental figure 3A: Mean difference in LAZ** **2**

**Supplemental figure 3B: Stunting prevalence ratio** **3**

**Supplemental figure 3C: Stunting prevalence difference** **4**

**Supplemental figure 3D: Mean difference in WLZ** **5**

**Supplemental figure 3E: Wasting prevalence ratio** **6**

**Supplemental figure 3F: Wasting prevalence difference** **7**

**Supplemental figure 3G: Mean difference in MUACZ** **8**

**Supplemental figure 3H: Low MUAC prevalence ratio** **9**

**Supplemental figure 3I: Low MUAC prevalence difference** **10**

**Supplemental figure 3J: Acute malnutrition prevalence ratio** **11**

**Supplemental figure 3K: Acute malnutrition prevalence difference** **12**

**Supplemental figure 3L: Mean difference in WAZ** **13**

**Supplemental figure 3M: Underweight prevalence ratio** **14**

**Supplemental figure 3N: Underweight prevalence difference** **15**

**Supplemental figure 3O: Mean difference in HCZ** **16**

**Supplemental figure 3P: Small head size prevalence ratio** **17**

**Supplemental figure 3Q: Small head size prevalence difference** **18**

These figures are forest plots showing the study-level estimates of intervention effect with the pooled estimate in the bottom summary rows. For continuous outcomes the intervention effect is measured by the difference in mean of the LNS group minus control. For dichotomous outcomes analyzed via prevalence ratios the effect estimate is the prevalence in the LNS group divided by the prevalence in the control group. For dichotomous outcomes analyzed via prevalence differences the effect estimate is the prevalence in the LNS group minus the prevalence in the control group. The labels on the left y-axis correspond to trial level information. The values on the right indicate the study level effect estimate, confidence interval, and weighting for deriving the pooled estimate. LAZ, length-for-age z-score; WLZ, weight-for-length z-score; WAZ, weight-for-age z-score; MUACZ, mid-upper arm circumference z-score; HCZ, head circumference-for-age z-score.

### Supplemental figure 3A: Mean difference in LAZ

| Country | Trial | LNS N | Control N | MD (95% CI) | Fixed W | Random W |
|---|---|---|---|---|---|---|
| Bangladesh | JiVitA−4 (34) | 2838 | 1244 | 0.09 (0.03, 0.14) | 0.21 | 0.09 |
| Bangladesh | RDNS (35) | 1663 | 815 | 0.08 (−0.01, 0.16) | 0.08 | 0.08 |
| Bangladesh | WASH−B (36) | 1158 | 3431 | 0.21 (0.14, 0.28) | 0.12 | 0.08 |
| Burkina Faso | iLiNS−Zinc (37) | 1952 | 664 | 0.31 (0.22, 0.39) | 0.08 | 0.07 |
| Burkina Faso | PROMIS (38) | 863 | 914 | 0.15 (0.02, 0.27) | 0.03 | 0.06 |
| Burkina Faso | PROMIS CS (38) | 430 | 439 | 0.04 (−0.14, 0.22) | 0.02 | 0.04 |
| Ghana | GHANA (39) | 98 | 96 | 0.28 (−0.02, 0.58) | 0.01 | 0.02 |
| Ghana | iLiNS−DYADG (40) | 347 | 692 | 0.20 (0.07, 0.33) | 0.03 | 0.06 |
| Haiti | HAITI (41) | 149 | 149 | −0.01 (−0.14, 0.13) | 0.03 | 0.05 |
| Kenya | WASH−B (42) | 1457 | 5137 | 0.15 (0.09, 0.21) | 0.15 | 0.08 |
| Madagascar | MAHAY (43) | 1702 | 1682 | 0.03 (−0.11, 0.17) | 0.03 | 0.05 |
| Malawi | iLiNS−DYADM (44) | 220 | 444 | −0.08 (−0.25, 0.10) | 0.02 | 0.04 |
| Malawi | iLiNS−DOSE (45) | 696 | 241 | 0.06 (−0.04, 0.17) | 0.05 | 0.07 |
| Mali | PROMIS (46) | 506 | 506 | 0.16 (0.01, 0.31) | 0.02 | 0.05 |
| Mali | PROMIS CS (46) | 952 | 969 | 0.22 (0.05, 0.39) | 0.02 | 0.04 |
| Zimbabwe | SHINE (HIV−) (47) | 1880 | 1794 | 0.16 (0.08, 0.23) | 0.09 | 0.08 |
| Zimbabwe | SHINE (HIV+) (48) | 337 | 330 | 0.25 (0.08, 0.43) | 0.02 | 0.04 |
| | **I² = 0.65, Tau² = 0.01** | **17248** | **19547** | | | |
| | **Fixed** | | | **0.14 (0.11, 0.16)** | | |
| | **Random** | | | **0.13 (0.09, 0.18)** | | |

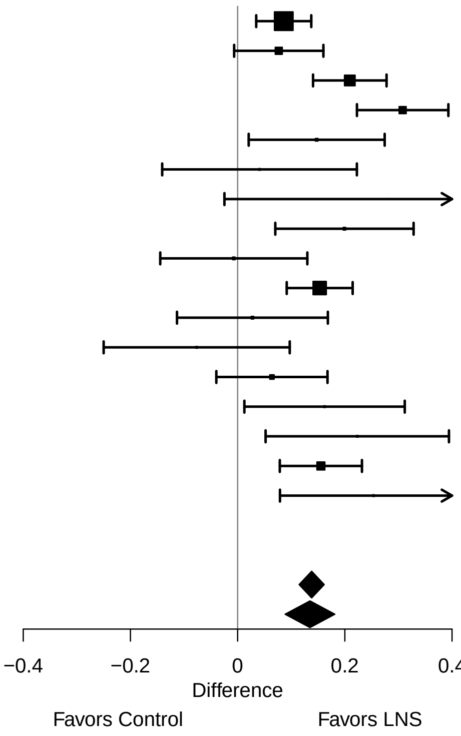

### Supplemental figure 3B: Stunting prevalence ratio

| Country | Trial | LNS N | Control N | PR (95% CI) | Fixed W | Random W |
|---|---|---|---|---|---|---|
| Bangladesh | JiVitA−4 (34) | 2838 | 1244 | 0.92 (0.84, 1.01) | 0.12 | 0.10 |
| Bangladesh | RDNS (35) | 1663 | 815 | 0.92 (0.85, 1.00) | 0.15 | 0.10 |
| Bangladesh | WASH−B (36) | 1158 | 3431 | 0.84 (0.77, 0.91) | 0.14 | 0.10 |
| Burkina Faso | iLiNS−Zinc (37) | 1952 | 664 | 0.75 (0.65, 0.87) | 0.04 | 0.06 |
| Burkina Faso | PROMIS (38) | 863 | 914 | 0.82 (0.67, 1.00) | 0.03 | 0.04 |
| Burkina Faso | PROMIS CS (38) | 430 | 439 | 0.87 (0.63, 1.19) | 0.01 | 0.02 |
| Ghana | GHANA (39) | 98 | 96 | 0.70 (0.23, 2.13) | 0.00 | 0.00 |
| Ghana | iLiNS−DYADG (40) | 347 | 692 | 0.66 (0.45, 0.98) | 0.01 | 0.01 |
| Haiti | HAITI (41) | 149 | 149 | 1.04 (0.60, 1.81) | 0.00 | 0.01 |
| Kenya | WASH−B (42) | 1457 | 5137 | 0.86 (0.78, 0.94) | 0.11 | 0.09 |
| Madagascar | MAHAY (43) | 1702 | 1682 | 0.98 (0.90, 1.07) | 0.12 | 0.10 |
| Malawi | iLiNS−DYADM (44) | 220 | 444 | 1.10 (0.89, 1.36) | 0.02 | 0.04 |
| Malawi | iLiNS−DOSE (45) | 696 | 241 | 1.00 (0.87, 1.15) | 0.05 | 0.07 |
| Mali | PROMIS (46) | 506 | 506 | 0.92 (0.76, 1.10) | 0.03 | 0.05 |
| Mali | PROMIS CS (46) | 952 | 969 | 0.78 (0.65, 0.95) | 0.03 | 0.05 |
| Zimbabwe | SHINE (HIV−) (47) | 1880 | 1794 | 0.79 (0.72, 0.87) | 0.10 | 0.09 |
| Zimbabwe | SHINE (HIV+) (48) | 337 | 330 | 0.81 (0.68, 0.96) | 0.03 | 0.05 |
| | **$I^2$ = 0.49, $Tau^2$ = 0.00** | **17248** | **19547** | | | |
| | **Fixed** | | | **0.88 (0.85, 0.91)** | | |
| | **Random** | | | **0.88 (0.83, 0.92)** | | |

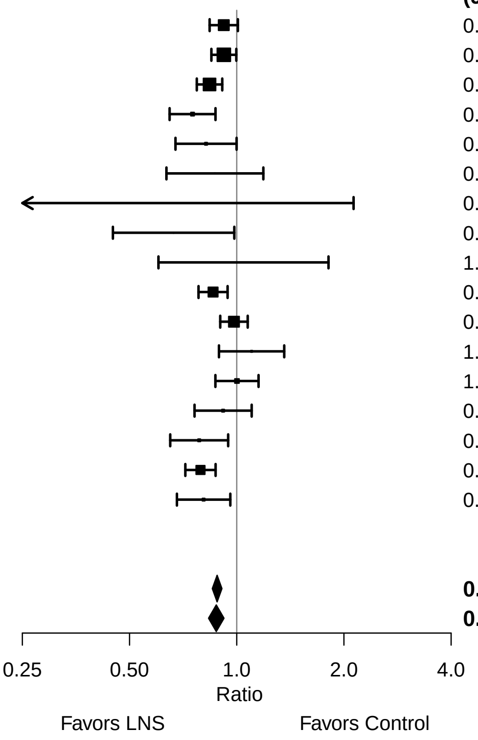

#### Supplemental figure 3C: Stunting prevalence difference

| Country | Trial | LNS N | Control N | PD (95% CI) | Fixed W | Random W |
|---|---|---|---|---|---|---|
| Bangladesh | JiVitA−4 (34) | 2838 | 1244 | −0.04 (−0.06, −0.01) | 0.13 | 0.09 |
| Bangladesh | RDNS (35) | 1663 | 815 | −0.03 (−0.07, 0.01) | 0.05 | 0.06 |
| Bangladesh | WASH−B (36) | 1158 | 3431 | −0.07 (−0.10, −0.04) | 0.09 | 0.08 |
| Burkina Faso | iLiNS−Zinc (37) | 1952 | 664 | −0.11 (−0.14, −0.08) | 0.09 | 0.08 |
| Burkina Faso | PROMIS (38) | 863 | 914 | −0.05 (−0.10, −0.01) | 0.05 | 0.06 |
| Burkina Faso | PROMIS CS (38) | 430 | 439 | −0.03 (−0.09, 0.02) | 0.03 | 0.04 |
| Ghana | GHANA (39) | 98 | 96 | −0.02 (−0.09, 0.05) | 0.02 | 0.03 |
| Ghana | iLiNS−DYADG (40) | 347 | 692 | −0.04 (−0.08, 0.00) | 0.06 | 0.07 |
| Haiti | HAITI (41) | 149 | 149 | −0.02 (−0.08, 0.04) | 0.02 | 0.04 |
| Kenya | WASH−B (42) | 1457 | 5137 | −0.05 (−0.07, −0.02) | 0.13 | 0.09 |
| Madagascar | MAHAY (43) | 1702 | 1682 | −0.01 (−0.04, 0.02) | 0.08 | 0.07 |
| Malawi | iLiNS−DYADM (44) | 220 | 444 | 0.03 (−0.04, 0.11) | 0.01 | 0.03 |
| Malawi | iLiNS−DOSE (45) | 696 | 241 | −0.02 (−0.07, 0.04) | 0.03 | 0.04 |
| Mali | PROMIS (46) | 506 | 506 | −0.04 (−0.09, 0.01) | 0.04 | 0.05 |
| Mali | PROMIS CS (46) | 952 | 969 | −0.07 (−0.11, −0.03) | 0.05 | 0.06 |
| Zimbabwe | SHINE (HIV−) (47) | 1880 | 1794 | −0.07 (−0.10, −0.04) | 0.10 | 0.08 |
| Zimbabwe | SHINE (HIV+) (48) | 337 | 330 | −0.10 (−0.17, −0.02) | 0.02 | 0.03 |
| | **I² = 0.54, Tau² = 0.00** | **17248** | **19547** | | | |
| | **Fixed** | | | **−0.05 (−0.06, −0.04)** | | |
| | **Random** | | | **−0.05 (−0.06, −0.03)** | | |

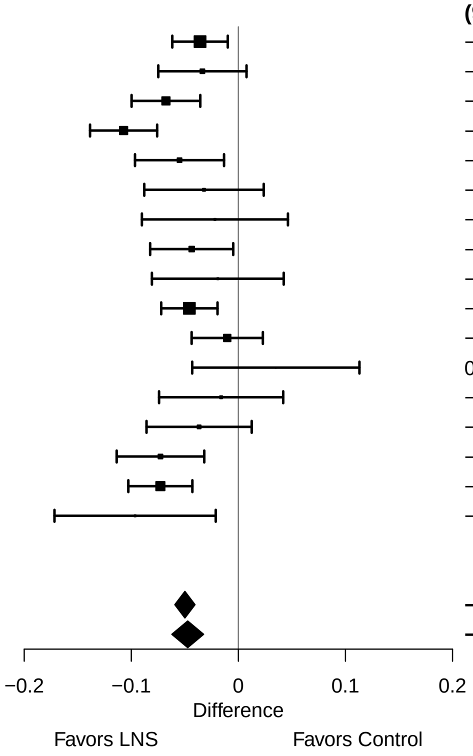

### Supplemental figure 3D: Mean difference in WLZ

| Country | Trial | LNS N | Control N | MD (95% CI) | Fixed W | Random W |
|---|---|---|---|---|---|---|
| Bangladesh | JiVitA−4 (34) | 2783 | 1217 | 0.08 (0.03, 0.13) | 0.22 | 0.10 |
| Bangladesh | RDNS (35) | 1661 | 815 | 0.07 (0.00, 0.15) | 0.08 | 0.08 |
| Bangladesh | WASH−B (36) | 1156 | 3424 | 0.12 (0.05, 0.19) | 0.10 | 0.08 |
| Burkina Faso | iLiNS−Zinc (37) | 1952 | 664 | 0.18 (0.11, 0.26) | 0.09 | 0.08 |
| Burkina Faso | PROMIS (38) | 856 | 907 | 0.10 (−0.03, 0.22) | 0.03 | 0.05 |
| Burkina Faso | PROMIS CS (38) | 430 | 436 | −0.11 (−0.24, 0.02) | 0.03 | 0.05 |
| Ghana | GHANA (39) | 98 | 96 | 0.21 (−0.13, 0.54) | 0.00 | 0.01 |
| Ghana | iLiNS−DYADG (40) | 347 | 692 | 0.09 (−0.04, 0.22) | 0.03 | 0.05 |
| Haiti | HAITI (41) | 148 | 149 | 0.09 (−0.07, 0.24) | 0.02 | 0.04 |
| Kenya | WASH−B (42) | 1455 | 5118 | 0.06 (0.00, 0.13) | 0.13 | 0.09 |
| Madagascar | MAHAY (43) | 1700 | 1682 | 0.00 (−0.11, 0.11) | 0.04 | 0.06 |
| Malawi | iLiNS−DYADM (44) | 220 | 444 | −0.05 (−0.21, 0.11) | 0.02 | 0.04 |
| Malawi | iLiNS−DOSE (45) | 696 | 241 | 0.01 (−0.12, 0.14) | 0.03 | 0.05 |
| Mali | PROMIS (46) | 499 | 502 | 0.10 (−0.02, 0.23) | 0.03 | 0.05 |
| Mali | PROMIS CS (46) | 944 | 959 | 0.16 (0.06, 0.26) | 0.05 | 0.06 |
| Zimbabwe | SHINE (HIV−) (47) | 1869 | 1784 | 0.08 (0.00, 0.15) | 0.09 | 0.08 |
| Zimbabwe | SHINE (HIV+) (48) | 336 | 328 | −0.14 (−0.32, 0.04) | 0.01 | 0.03 |
| | **$I^2 = 0.51$, $Tau^2 = 0.00$** | **17150** | **19458** | | | |
| | **Fixed** | | | **0.08 (0.06, 0.10)** | | |
| | **Random** | | | **0.07 (0.03, 0.11)** | | |

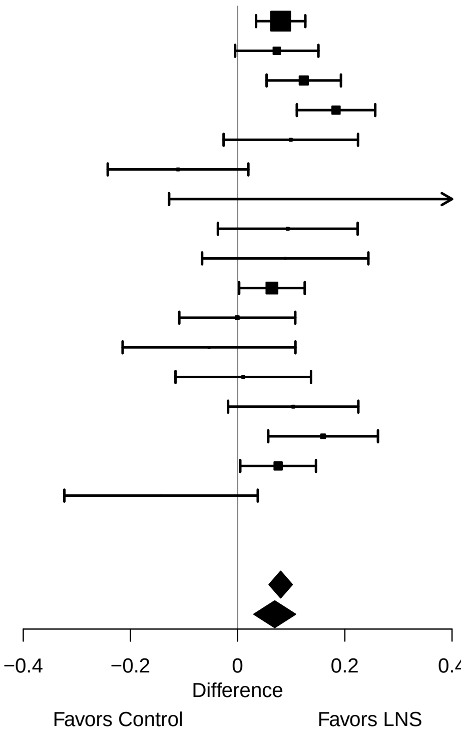

### Supplemental figure 3E: Wasting prevalence ratio

| Country | Trial | LNS N | Control N | PR (95% CI) | Fixed W | Random W |
|---|---|---|---|---|---|---|
| Bangladesh | JiVitA−4 (34) | 2783 | 1217 | 0.91 (0.78, 1.05) | 0.27 | 0.27 |
| Bangladesh | RDNS (35) | 1661 | 815 | 0.87 (0.70, 1.07) | 0.13 | 0.13 |
| Bangladesh | WASH−B (36) | 1156 | 3424 | 0.79 (0.65, 0.96) | 0.16 | 0.16 |
| Burkina Faso | iLiNS−Zinc (37) | 1952 | 664 | 0.69 (0.54, 0.87) | 0.11 | 0.11 |
| Burkina Faso | PROMIS (38) | 856 | 907 | 0.86 (0.59, 1.27) | 0.04 | 0.04 |
| Burkina Faso | PROMIS CS (38) | 430 | 436 | 1.03 (0.72, 1.49) | 0.04 | 0.04 |
| Ghana | GHANA (39) | 98 | 96 | 0.87 (0.35, 2.16) | 0.01 | 0.01 |
| Ghana | iLiNS−DYADG (40) | 347 | 692 | 0.84 (0.52, 1.37) | 0.03 | 0.03 |
| Haiti | HAITI (41) | | | | | |
| Kenya | WASH−B (42) | 1455 | 5118 | 0.88 (0.53, 1.45) | 0.02 | 0.02 |
| Madagascar | MAHAY (43) | 1700 | 1682 | 0.96 (0.67, 1.38) | 0.05 | 0.05 |
| Malawi | iLiNS−DYADM (44) | 220 | 444 | 0.94 (0.39, 2.28) | 0.01 | 0.01 |
| Malawi | iLiNS−DOSE (45) | 696 | 241 | 1.37 (0.78, 2.40) | 0.02 | 0.02 |
| Mali | PROMIS (46) | 499 | 502 | 0.61 (0.15, 2.42) | 0.00 | 0.00 |
| Mali | PROMIS CS (46) | 944 | 959 | 0.83 (0.63, 1.10) | 0.07 | 0.07 |
| Zimbabwe | SHINE (HIV−) (47) | 1869 | 1784 | 0.89 (0.60, 1.34) | 0.04 | 0.04 |
| Zimbabwe | SHINE (HIV+) (48) | 336 | 328 | 1.38 (0.65, 2.96) | 0.01 | 0.01 |
| | **I² = 0.00, Tau² = 0.00** | **17002** | **19309** | | | |
| | **Fixed** | | | **0.86 (0.80, 0.93)** | | |
| | **Random** | | | **0.86 (0.80, 0.93)** | | |

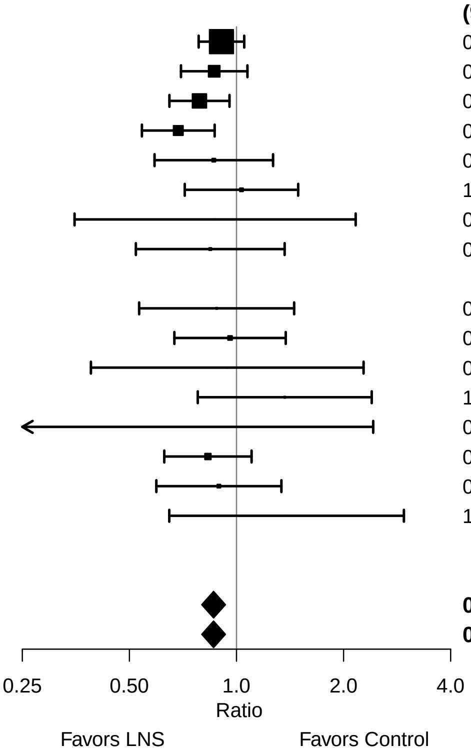

### Supplemental figure 3F: Wasting prevalence difference

| Country | Trial | LNS N | Control N | PD (95% CI) | Fixed W | Random W |
|---|---|---|---|---|---|---|
| Bangladesh | JiVitA−4 (34) | 2783 | 1217 | −0.01 (−0.04, 0.01) | 0.04 | 0.04 |
| Bangladesh | RDNS (35) | 1661 | 815 | −0.02 (−0.05, 0.01) | 0.02 | 0.02 |
| Bangladesh | WASH−B (36) | 1156 | 3424 | −0.02 (−0.04, 0.00) | 0.05 | 0.05 |
| Burkina Faso | iLiNS−Zinc (37) | 1952 | 664 | −0.04 (−0.07, −0.01) | 0.03 | 0.03 |
| Burkina Faso | PROMIS (38) | 856 | 907 | −0.01 (−0.04, 0.01) | 0.03 | 0.03 |
| Burkina Faso | PROMIS CS (38) | 430 | 436 | 0.00 (−0.04, 0.05) | 0.01 | 0.01 |
| Ghana | GHANA (39) | 98 | 96 | −0.01 (−0.09, 0.07) | 0.00 | 0.00 |
| Ghana | iLiNS−DYADG (40) | 347 | 692 | −0.01 (−0.04, 0.02) | 0.02 | 0.02 |
| Haiti | HAITI (41) | | | | | |
| Kenya | WASH−B (42) | 1455 | 5118 | 0.00 (−0.01, 0.00) | 0.39 | 0.39 |
| Madagascar | MAHAY (43) | 1700 | 1682 | 0.00 (−0.02, 0.01) | 0.07 | 0.07 |
| Malawi | iLiNS−DYADM (44) | 220 | 444 | 0.00 (−0.03, 0.03) | 0.02 | 0.02 |
| Malawi | iLiNS−DOSE (45) | 696 | 241 | 0.01 (−0.03, 0.04) | 0.02 | 0.02 |
| Mali | PROMIS (46) | 499 | 502 | −0.01 (−0.02, 0.01) | 0.09 | 0.09 |
| Mali | PROMIS CS (46) | 944 | 959 | −0.02 (−0.04, 0.01) | 0.03 | 0.03 |
| Zimbabwe | SHINE (HIV−) (47) | 1869 | 1784 | 0.00 (−0.01, 0.01) | 0.18 | 0.18 |
| Zimbabwe | SHINE (HIV+) (48) | 336 | 328 | 0.01 (−0.02, 0.05) | 0.02 | 0.02 |
| | **I² = 0.12, Tau² = 0.00** | **17002** | **19309** | | | |
| | **Fixed** | | | **−0.01 (−0.01, 0.00)** | | |
| | **Random** | | | **−0.01 (−0.01, 0.00)** | | |

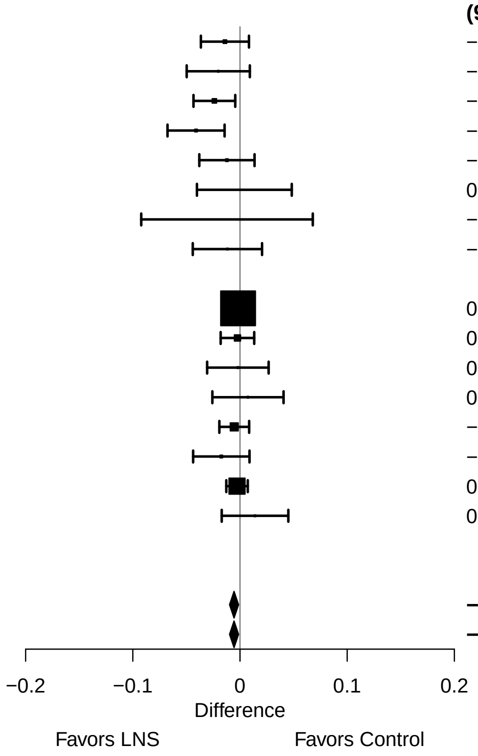

### Supplemental figure 3G: Mean difference in MUACZ

| Country | Trial | LNS N | Control N | MD (95% CI) | Fixed W | Random W |
|---|---|---|---|---|---|---|
| Bangladesh | JiVitA−4 (34) | 2897 | 1271 | 0.08 (0.03, 0.13) | 0.25 | 0.10 |
| Bangladesh | RDNS (35) | 1663 | 815 | 0.04 (−0.04, 0.12) | 0.09 | 0.09 |
| Bangladesh | WASH−B (36) | | | | | |
| Burkina Faso | iLiNS−Zinc (37) | 1959 | 666 | 0.30 (0.20, 0.39) | 0.06 | 0.08 |
| Burkina Faso | PROMIS (38) | 854 | 912 | 0.09 (−0.03, 0.21) | 0.04 | 0.06 |
| Burkina Faso | PROMIS CS (38) | 424 | 436 | −0.06 (−0.22, 0.10) | 0.02 | 0.05 |
| Ghana | GHANA (39) | | | | | |
| Ghana | iLiNS−DYADG (40) | 347 | 693 | 0.09 (−0.03, 0.20) | 0.04 | 0.07 |
| Haiti | HAITI (41) | | | | | |
| Kenya | WASH−B (42) | 1461 | 5185 | 0.12 (0.05, 0.18) | 0.13 | 0.09 |
| Madagascar | MAHAY (43) | 1680 | 1658 | −0.03 (−0.16, 0.09) | 0.04 | 0.06 |
| Malawi | iLiNS−DYADM (44) | 220 | 444 | −0.04 (−0.20, 0.12) | 0.02 | 0.05 |
| Malawi | iLiNS−DOSE (45) | 696 | 240 | 0.06 (−0.05, 0.18) | 0.05 | 0.07 |
| Mali | PROMIS (46) | 507 | 505 | 0.19 (0.05, 0.32) | 0.03 | 0.06 |
| Mali | PROMIS CS (46) | 954 | 970 | 0.13 (0.01, 0.25) | 0.04 | 0.07 |
| Zimbabwe | SHINE (HIV−) (47) | 1872 | 1780 | 0.06 (0.00, 0.12) | 0.15 | 0.10 |
| Zimbabwe | SHINE (HIV+) (48) | 337 | 328 | 0.04 (−0.10, 0.17) | 0.03 | 0.06 |
| | **$I^2$ = 0.62, $Tau^2$ = 0.00** | **15871** | **15903** | | | |
| | **Fixed** | | | **0.09 (0.06, 0.11)** | | |
| | **Random** | | | **0.08 (0.04, 0.13)** | | |

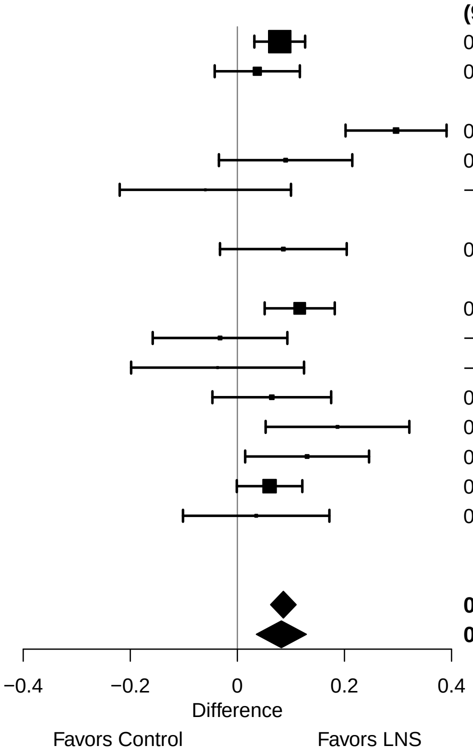

### Supplemental figure 3H: Low MUAC prevalence ratio

| Country | Trial | LNS N | Control N | PR (95% CI) | Fixed W | Random W |
|---|---|---|---|---|---|---|
| Bangladesh | JiVitA−4 (34) | 2897 | 1271 | 0.85 (0.70, 1.03) | 0.22 | 0.19 |
| Bangladesh | RDNS (35) | 1663 | 815 | 1.02 (0.69, 1.49) | 0.06 | 0.06 |
| Bangladesh | WASH−B (36) | | | | | |
| Burkina Faso | iLiNS−Zinc (37) | 1959 | 666 | 0.64 (0.51, 0.79) | 0.18 | 0.16 |
| Burkina Faso | PROMIS (38) | 854 | 912 | 0.94 (0.63, 1.40) | 0.05 | 0.06 |
| Burkina Faso | PROMIS CS (38) | 424 | 436 | 0.92 (0.66, 1.28) | 0.08 | 0.08 |
| Ghana | GHANA (39) | | | | | |
| Ghana | iLiNS−DYADG (40) | 347 | 693 | 0.77 (0.37, 1.57) | 0.02 | 0.02 |
| Haiti | HAITI (41) | | | | | |
| Kenya | WASH−B (42) | 1461 | 5185 | 0.71 (0.54, 0.94) | 0.11 | 0.11 |
| Madagascar | MAHAY (43) | 1680 | 1658 | 0.97 (0.76, 1.26) | 0.13 | 0.13 |
| Malawi | iLiNS−DYADM (44) | 220 | 444 | 0.94 (0.39, 2.28) | 0.01 | 0.01 |
| Malawi | iLiNS−DOSE (45) | 696 | 240 | 1.35 (0.54, 3.36) | 0.01 | 0.01 |
| Mali | PROMIS (46) | 507 | 505 | 0.58 (0.31, 1.07) | 0.02 | 0.03 |
| Mali | PROMIS CS (46) | 954 | 970 | 0.90 (0.68, 1.19) | 0.10 | 0.11 |
| Zimbabwe | SHINE (HIV−) (47) | 1872 | 1780 | 0.51 (0.27, 0.97) | 0.02 | 0.02 |
| Zimbabwe | SHINE (HIV+) (48) | 337 | 328 | 0.97 (0.29, 3.28) | 0.01 | 0.01 |
| | **I² = 0.16, Tau² = 0.00** | **15871** | **15903** | | | |
| | **Fixed** | | | **0.82 (0.75, 0.89)** | | |
| | **Random** | | | **0.82 (0.74, 0.91)** | | |

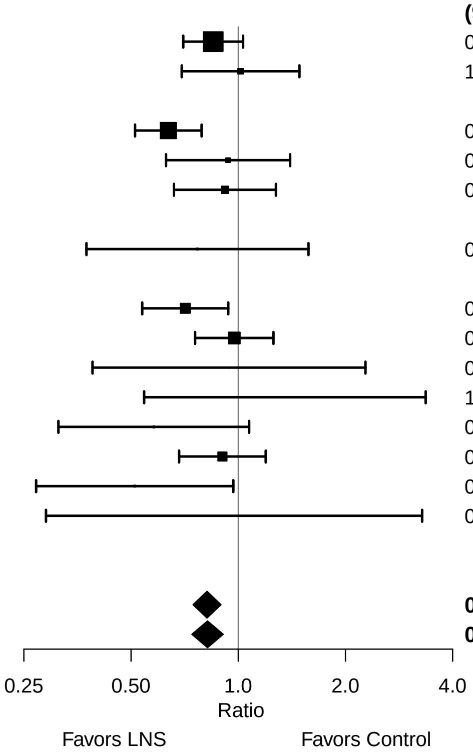

### Supplemental figure 3I: Low MUAC prevalence difference

| Country | Trial | LNS N | Control N | PD (95% CI) | Fixed W | Random W |
|---|---|---|---|---|---|---|
| Bangladesh | JiVitA−4 (34) | 2897 | 1271 | −0.02 (−0.04, 0.00) | 0.05 | 0.08 |
| Bangladesh | RDNS (35) | 1663 | 815 | 0.00 (−0.02, 0.02) | 0.06 | 0.08 |
| Bangladesh | WASH−B (36) | | | | | |
| Burkina Faso | iLiNS−Zinc (37) | 1959 | 666 | −0.07 (−0.10, −0.04) | 0.02 | 0.05 |
| Burkina Faso | PROMIS (38) | 854 | 912 | −0.01 (−0.03, 0.02) | 0.03 | 0.06 |
| Burkina Faso | PROMIS CS (38) | 424 | 436 | −0.01 (−0.06, 0.03) | 0.01 | 0.03 |
| Ghana | GHANA (39) | | | | | |
| Ghana | iLiNS−DYADG (40) | 347 | 693 | −0.01 (−0.03, 0.01) | 0.04 | 0.07 |
| Haiti | HAITI (41) | | | | | |
| Kenya | WASH−B (42) | 1461 | 5185 | −0.02 (−0.03, 0.00) | 0.14 | 0.10 |
| Madagascar | MAHAY (43) | 1680 | 1658 | 0.00 (−0.02, 0.02) | 0.05 | 0.08 |
| Malawi | iLiNS−DYADM (44) | 220 | 444 | 0.00 (−0.03, 0.03) | 0.02 | 0.06 |
| Malawi | iLiNS−DOSE (45) | 696 | 240 | 0.01 (−0.01, 0.03) | 0.04 | 0.07 |
| Mali | PROMIS (46) | 507 | 505 | −0.02 (−0.05, 0.00) | 0.03 | 0.06 |
| Mali | PROMIS CS (46) | 954 | 970 | −0.01 (−0.04, 0.02) | 0.03 | 0.06 |
| Zimbabwe | SHINE (HIV−) (47) | 1872 | 1780 | −0.01 (−0.01, 0.00) | 0.42 | 0.11 |
| Zimbabwe | SHINE (HIV+) (48) | 337 | 328 | 0.00 (−0.02, 0.02) | 0.06 | 0.08 |
| | **I² = 0.50, Tau² = 0.00** | **15871** | **15903** | | | |
| | **Fixed** | | | **−0.01 (−0.01, 0.00)** | | |
| | **Random** | | | **−0.01 (−0.02, 0.00)** | | |

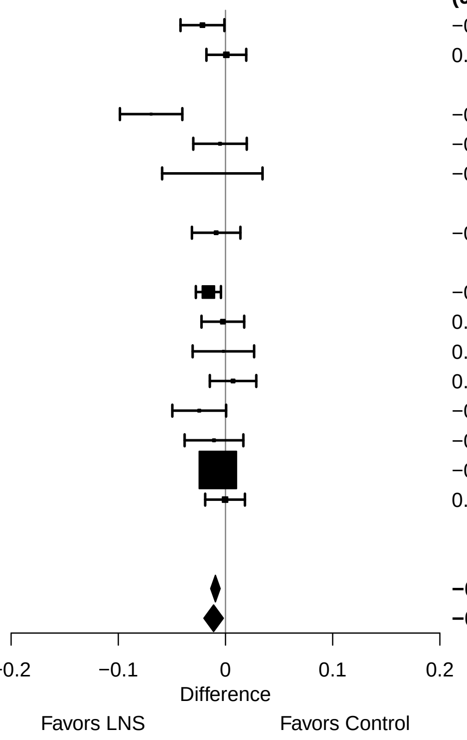

### Supplemental figure 3J: Acute malnutrition prevalence ratio

| Country | Trial | LNS N | Control N | PR (95% CI) | Fixed W | Random W |
|---|---|---|---|---|---|---|
| Bangladesh | JiVitA−4 (34) | 2783 | 1216 | 0.89 (0.78, 1.01) | 0.33 | 0.33 |
| Bangladesh | RDNS (35) | 1661 | 815 | 0.86 (0.70, 1.05) | 0.13 | 0.13 |
| Bangladesh | WASH−B (36) | | | | | |
| Burkina Faso | iLiNS−Zinc (37) | 1951 | 664 | 0.67 (0.54, 0.85) | 0.10 | 0.10 |
| Burkina Faso | PROMIS (38) | 846 | 901 | 0.83 (0.60, 1.14) | 0.05 | 0.05 |
| Burkina Faso | PROMIS CS (38) | 424 | 434 | 0.95 (0.73, 1.23) | 0.08 | 0.08 |
| Ghana | GHANA (39) | | | | | |
| Ghana | iLiNS−DYADG (40) | 347 | 692 | 0.85 (0.54, 1.35) | 0.02 | 0.02 |
| Haiti | HAITI (41) | | | | | |
| Kenya | WASH−B (42) | 1454 | 5117 | 0.71 (0.49, 1.02) | 0.04 | 0.04 |
| Madagascar | MAHAY (43) | 1675 | 1655 | 0.98 (0.75, 1.29) | 0.07 | 0.07 |
| Malawi | iLiNS−DYADM (44) | 220 | 444 | 0.81 (0.36, 1.80) | 0.01 | 0.01 |
| Malawi | iLiNS−DOSE (45) | 690 | 240 | 1.42 (0.83, 2.43) | 0.02 | 0.02 |
| Mali | PROMIS (46) | 499 | 502 | 0.59 (0.23, 1.47) | 0.01 | 0.01 |
| Mali | PROMIS CS (46) | 944 | 959 | 0.89 (0.70, 1.13) | 0.09 | 0.09 |
| Zimbabwe | SHINE (HIV−) (47) | 1867 | 1777 | 0.79 (0.55, 1.14) | 0.04 | 0.04 |
| Zimbabwe | SHINE (HIV+) (48) | 336 | 327 | 1.23 (0.61, 2.48) | 0.01 | 0.01 |
| | **I² = 0.00, Tau² = 0.00** | **15697** | **15743** | | | |
| | **Fixed** | | | **0.86 (0.80, 0.93)** | | |
| | **Random** | | | **0.86 (0.80, 0.93)** | | |

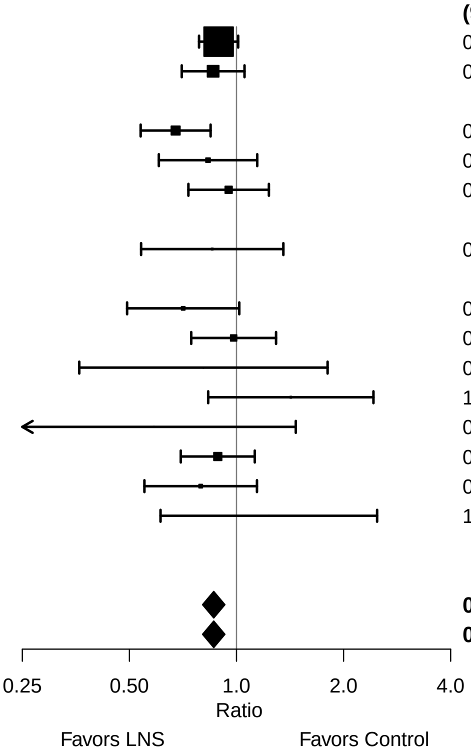

### Supplemental figure 3K: Acute malnutrition prevalence difference

| Country | Trial | LNS N | Control N | PD (95% CI) | Fixed W | Random W |
|---|---|---|---|---|---|---|
| Bangladesh | JiVitA−4 (34) | 2783 | 1216 | −0.02 (−0.05, 0.00) | 0.05 | 0.07 |
| Bangladesh | RDNS (35) | 1661 | 815 | −0.02 (−0.05, 0.01) | 0.03 | 0.05 |
| Bangladesh | WASH−B (36) | | | | | |
| Burkina Faso | iLiNS−Zinc (37) | 1951 | 664 | −0.06 (−0.09, −0.03) | 0.03 | 0.05 |
| Burkina Faso | PROMIS (38) | 846 | 901 | −0.02 (−0.05, 0.01) | 0.03 | 0.06 |
| Burkina Faso | PROMIS CS (38) | 424 | 434 | −0.01 (−0.06, 0.04) | 0.01 | 0.02 |
| Ghana | GHANA (39) | | | | | |
| Ghana | iLiNS−DYADG (40) | 347 | 692 | −0.01 (−0.05, 0.02) | 0.02 | 0.04 |
| Haiti | HAITI (41) | | | | | |
| Kenya | WASH−B (42) | 1454 | 5117 | −0.01 (−0.02, 0.00) | 0.34 | 0.17 |
| Madagascar | MAHAY (43) | 1675 | 1655 | 0.00 (−0.02, 0.02) | 0.07 | 0.09 |
| Malawi | iLiNS−DYADM (44) | 220 | 444 | −0.01 (−0.04, 0.02) | 0.03 | 0.05 |
| Malawi | iLiNS−DOSE (45) | 690 | 240 | 0.01 (−0.03, 0.04) | 0.02 | 0.04 |
| Mali | PROMIS (46) | 499 | 502 | −0.01 (−0.03, 0.01) | 0.08 | 0.10 |
| Mali | PROMIS CS (46) | 944 | 959 | −0.01 (−0.04, 0.01) | 0.03 | 0.05 |
| Zimbabwe | SHINE (HIV−) (47) | 1867 | 1777 | −0.01 (−0.02, 0.00) | 0.23 | 0.16 |
| Zimbabwe | SHINE (HIV+) (48) | 336 | 327 | 0.01 (−0.02, 0.04) | 0.02 | 0.04 |
| | **I² = 0.21, Tau² = 0.00** | **15697** | **15743** | | | |
| | **Fixed** | | | **−0.01 (−0.02, −0.01)** | | |
| | **Random** | | | **−0.01 (−0.02, 0.00)** | | |

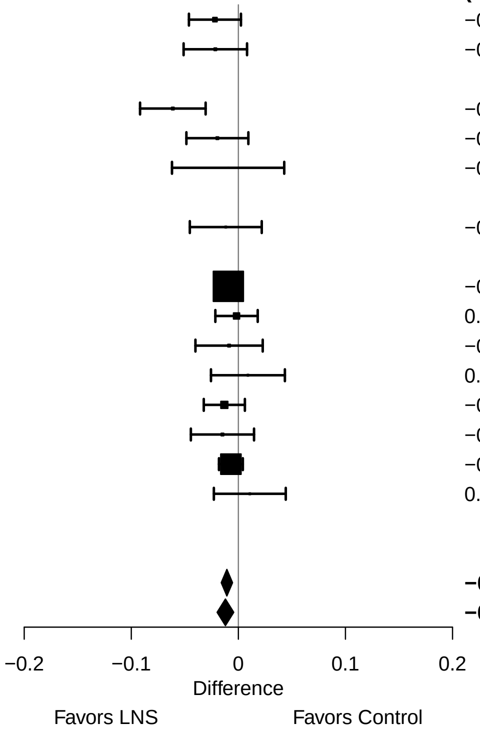

### Supplemental figure 3L: Mean difference in WAZ

| Country | Trial | LNS N | Control N | MD (95% CI) | Fixed W | Random W |
|---|---|---|---|---|---|---|
| Bangladesh | JiVitA−4 (34) | 2814 | 1232 | 0.10 (0.07, 0.14) | 0.33 | 0.09 |
| Bangladesh | RDNS (35) | 1663 | 815 | 0.09 (0.01, 0.18) | 0.05 | 0.07 |
| Bangladesh | WASH−B (36) | 1163 | 3461 | 0.20 (0.13, 0.26) | 0.09 | 0.08 |
| Burkina Faso | iLiNS−Zinc (37) | 1960 | 666 | 0.28 (0.21, 0.35) | 0.08 | 0.08 |
| Burkina Faso | PROMIS (38) | 857 | 911 | 0.15 (0.03, 0.27) | 0.03 | 0.06 |
| Burkina Faso | PROMIS CS (38) | 430 | 436 | −0.05 (−0.19, 0.09) | 0.02 | 0.05 |
| Ghana | GHANA (39) | 98 | 96 | 0.28 (−0.05, 0.61) | 0.00 | 0.01 |
| Ghana | iLiNS−DYADG (40) | 347 | 693 | 0.16 (0.03, 0.29) | 0.02 | 0.05 |
| Haiti | HAITI (41) | 148 | 150 | 0.06 (−0.06, 0.19) | 0.03 | 0.05 |
| Kenya | WASH−B (42) | 1461 | 5165 | 0.13 (0.07, 0.20) | 0.10 | 0.08 |
| Madagascar | MAHAY (43) | 1702 | 1685 | 0.01 (−0.10, 0.13) | 0.03 | 0.06 |
| Malawi | iLiNS−DYADM (44) | 220 | 444 | −0.07 (−0.23, 0.10) | 0.01 | 0.04 |
| Malawi | iLiNS−DOSE (45) | 702 | 241 | 0.05 (−0.05, 0.16) | 0.04 | 0.06 |
| Mali | PROMIS (46) | 500 | 502 | 0.16 (0.05, 0.28) | 0.03 | 0.06 |
| Mali | PROMIS CS (46) | 946 | 960 | 0.23 (0.11, 0.36) | 0.03 | 0.05 |
| Zimbabwe | SHINE (HIV−) (47) | 1869 | 1786 | 0.13 (0.07, 0.20) | 0.09 | 0.08 |
| Zimbabwe | SHINE (HIV+) (48) | 336 | 328 | 0.01 (−0.16, 0.18) | 0.01 | 0.04 |
| | **I² = 0.66, Tau² = 0.01** | **17216** | **19571** | | | |
| | **Fixed** | | | **0.13 (0.11, 0.15)** | | |
| | **Random** | | | **0.12 (0.07, 0.16)** | | |

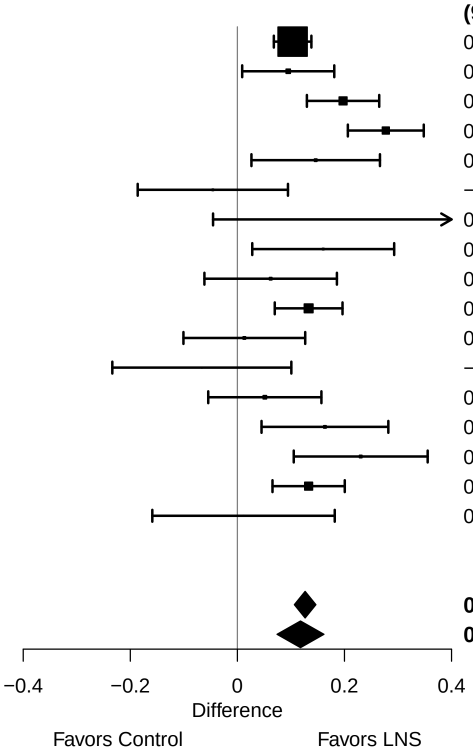

### Supplemental figure 3M: Underweight prevalence ratio

| Country | Trial | LNS N | Control N | PR (95% CI) | Fixed W | Random W |
|---|---|---|---|---|---|---|
| Bangladesh | JiVitA−4 (34) | 2814 | 1232 | 0.91 (0.83, 1.01) | 0.21 | 0.13 |
| Bangladesh | RDNS (35) | 1663 | 815 | 0.92 (0.83, 1.02) | 0.18 | 0.12 |
| Bangladesh | WASH−B (36) | 1163 | 3461 | 0.79 (0.70, 0.89) | 0.14 | 0.11 |
| Burkina Faso | iLiNS−Zinc (37) | 1960 | 666 | 0.70 (0.59, 0.82) | 0.08 | 0.09 |
| Burkina Faso | PROMIS (38) | 857 | 911 | 0.86 (0.71, 1.05) | 0.05 | 0.07 |
| Burkina Faso | PROMIS CS (38) | 430 | 436 | 0.99 (0.77, 1.26) | 0.03 | 0.05 |
| Ghana | GHANA (39) | 98 | 96 | 0.71 (0.30, 1.69) | 0.00 | 0.01 |
| Ghana | iLiNS−DYADG (40) | 347 | 693 | 0.82 (0.57, 1.18) | 0.02 | 0.03 |
| Haiti | HAITI (41) | 148 | 150 | 0.71 (0.28, 1.78) | 0.00 | 0.00 |
| Kenya | WASH−B (42) | 1461 | 5165 | 0.76 (0.62, 0.93) | 0.05 | 0.06 |
| Madagascar | MAHAY (43) | 1702 | 1685 | 1.04 (0.89, 1.21) | 0.08 | 0.09 |
| Malawi | iLiNS−DYADM (44) | 220 | 444 | 1.13 (0.74, 1.72) | 0.01 | 0.02 |
| Malawi | iLiNS−DOSE (45) | 702 | 241 | 1.01 (0.78, 1.31) | 0.03 | 0.05 |
| Mali | PROMIS (46) | 500 | 502 | 0.85 (0.62, 1.18) | 0.02 | 0.03 |
| Mali | PROMIS CS (46) | 946 | 960 | 0.88 (0.71, 1.10) | 0.04 | 0.06 |
| Zimbabwe | SHINE (HIV−) (47) | 1869 | 1786 | 0.71 (0.57, 0.87) | 0.04 | 0.06 |
| Zimbabwe | SHINE (HIV+) (48) | 336 | 328 | 1.01 (0.73, 1.40) | 0.02 | 0.03 |
| | **I² = 0.42, Tau² = 0.01** | **17216** | **19571** | | | |
| | **Fixed** | | | **0.87 (0.83, 0.91)** | | |
| | **Random** | | | **0.87 (0.81, 0.92)** | | |

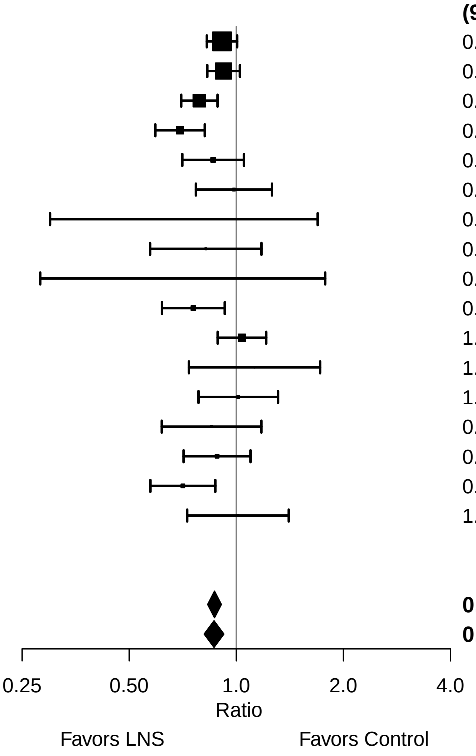

### Supplemental figure 3N: Underweight prevalence difference

| Country | Trial | LNS N | Control N | PD (95% CI) | Fixed W | Random W |
|---|---|---|---|---|---|---|
| Bangladesh | JiVitA−4 (34) | 2814 | 1232 | −0.04 (−0.07, −0.02) | 0.09 | 0.08 |
| Bangladesh | RDNS (35) | 1663 | 815 | −0.03 (−0.07, 0.01) | 0.04 | 0.06 |
| Bangladesh | WASH−B (36) | 1163 | 3461 | −0.07 (−0.10, −0.04) | 0.07 | 0.07 |
| Burkina Faso | iLiNS−Zinc (37) | 1960 | 666 | −0.10 (−0.13, −0.07) | 0.06 | 0.07 |
| Burkina Faso | PROMIS (38) | 857 | 911 | −0.03 (−0.06, 0.01) | 0.05 | 0.06 |
| Burkina Faso | PROMIS CS (38) | 430 | 436 | 0.00 (−0.06, 0.05) | 0.02 | 0.04 |
| Ghana | GHANA (39) | 98 | 96 | −0.03 (−0.12, 0.05) | 0.01 | 0.02 |
| Ghana | iLiNS−DYADG (40) | 347 | 693 | −0.02 (−0.06, 0.02) | 0.03 | 0.05 |
| Haiti | HAITI (41) | 148 | 150 | 0.00 (−0.05, 0.04) | 0.03 | 0.05 |
| Kenya | WASH−B (42) | 1461 | 5165 | −0.02 (−0.04, −0.01) | 0.23 | 0.09 |
| Madagascar | MAHAY (43) | 1702 | 1685 | 0.01 (−0.02, 0.04) | 0.07 | 0.07 |
| Malawi | iLiNS−DYADM (44) | 220 | 444 | 0.01 (−0.04, 0.07) | 0.02 | 0.04 |
| Malawi | iLiNS−DOSE (45) | 702 | 241 | −0.02 (−0.07, 0.03) | 0.02 | 0.05 |
| Mali | PROMIS (46) | 500 | 502 | −0.04 (−0.08, 0.00) | 0.04 | 0.06 |
| Mali | PROMIS CS (46) | 946 | 960 | −0.03 (−0.06, 0.01) | 0.04 | 0.06 |
| Zimbabwe | SHINE (HIV−) (47) | 1869 | 1786 | −0.03 (−0.05, −0.01) | 0.17 | 0.09 |
| Zimbabwe | SHINE (HIV+) (48) | 336 | 328 | 0.00 (−0.06, 0.06) | 0.02 | 0.04 |
| | **I² = 0.60, Tau² = 0.00** | **17216** | **19571** | | | |
| | **Fixed** | | | **−0.03 (−0.04, −0.02)** | | |
| | **Random** | | | **−0.03 (−0.04, −0.02)** | | |

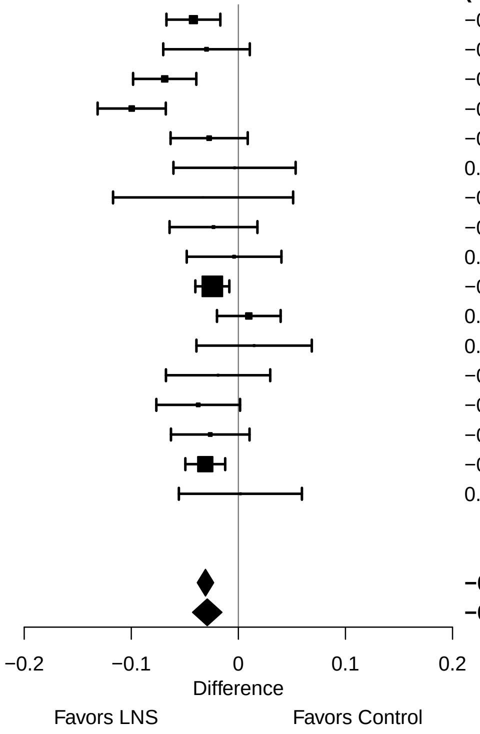

### Supplemental figure 3O: Mean difference in HCZ

| Country | Trial | LNS N | Control N | MD (95% CI) | Fixed W | Random W |
|---|---|---|---|---|---|---|
| Bangladesh | JiVitA−4 (34) | 2885 | 1267 | 0.06 (0.01, 0.11) | 0.23 | 0.15 |
| Bangladesh | RDNS (35) | 1663 | 815 | 0.10 (0.03, 0.17) | 0.11 | 0.12 |
| Bangladesh | WASH−B (36) | 1163 | 3463 | 0.12 (0.06, 0.19) | 0.13 | 0.13 |
| Burkina Faso | iLiNS−Zinc (37) | 1946 | 664 | 0.16 (0.10, 0.23) | 0.14 | 0.13 |
| Burkina Faso | PROMIS (38) | | | | | |
| Burkina Faso | PROMIS CS (38) | | | | | |
| Ghana | GHANA (39) | 98 | 96 | 0.20 (−0.06, 0.46) | 0.01 | 0.02 |
| Ghana | iLiNS−DYADG (40) | 347 | 692 | 0.07 (−0.05, 0.18) | 0.04 | 0.07 |
| Haiti | HAITI (41) | | | | | |
| Kenya | WASH−B (42) | 1461 | 5176 | 0.04 (−0.02, 0.10) | 0.15 | 0.13 |
| Madagascar | MAHAY (43) | | | | | |
| Malawi | iLiNS−DYADM (44) | 220 | 444 | −0.13 (−0.28, 0.03) | 0.02 | 0.04 |
| Malawi | iLiNS−DOSE (45) | 692 | 238 | 0.11 (−0.02, 0.23) | 0.04 | 0.06 |
| Mali | PROMIS (46) | | | | | |
| Mali | PROMIS CS (46) | | | | | |
| Zimbabwe | SHINE (HIV−) (47) | 1871 | 1783 | 0.09 (0.02, 0.16) | 0.11 | 0.12 |
| Zimbabwe | SHINE (HIV+) (48) | 337 | 329 | 0.10 (−0.07, 0.28) | 0.02 | 0.04 |
| | **I² = 0.46, Tau² = 0.00** | **12683** | **14967** | | | |
| | **Fixed** | | | **0.09 (0.06, 0.11)** | | |
| | **Random** | | | **0.09 (0.05, 0.12)** | | |

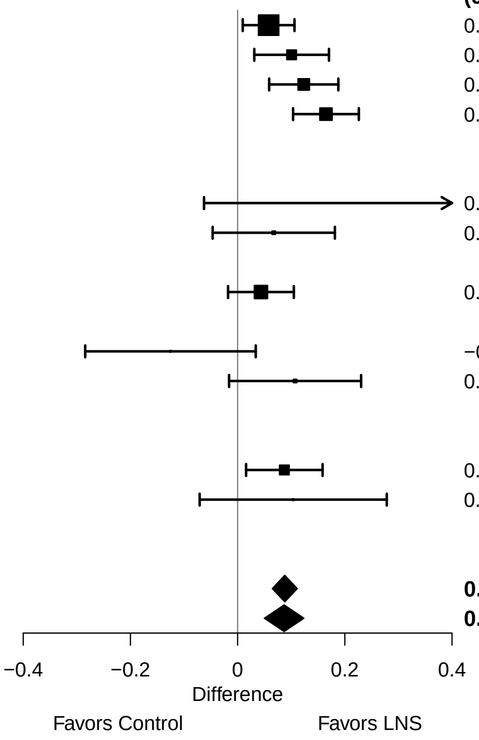

### Supplemental figure 3P: Small head size prevalence ratio

| Country | Trial | LNS N | Control N | PR (95% CI) | Fixed W | Random W |
|---|---|---|---|---|---|---|
| Bangladesh | JiVitA−4 (34) | 2885 | 1267 | 1.03 (0.90, 1.18) | 0.13 | 0.13 |
| Bangladesh | RDNS (35) | 1663 | 815 | 0.90 (0.82, 0.98) | 0.33 | 0.33 |
| Bangladesh | WASH−B (36) | 1163 | 3463 | 0.90 (0.82, 1.00) | 0.24 | 0.24 |
| Burkina Faso | iLiNS−Zinc (37) | 1946 | 664 | 0.89 (0.78, 1.01) | 0.14 | 0.14 |
| Burkina Faso | PROMIS (38) | | | | | |
| Burkina Faso | PROMIS CS (38) | | | | | |
| Ghana | GHANA (39) | | | | | |
| Ghana | iLiNS−DYADG (40) | 347 | 692 | 0.88 (0.65, 1.20) | 0.03 | 0.03 |
| Haiti | HAITI (41) | | | | | |
| Kenya | WASH−B (42) | 1461 | 5176 | 0.92 (0.67, 1.25) | 0.03 | 0.03 |
| Madagascar | MAHAY (43) | | | | | |
| Malawi | iLiNS−DYADM (44) | 220 | 444 | 0.88 (0.54, 1.44) | 0.01 | 0.01 |
| Malawi | iLiNS−DOSE (45) | 692 | 238 | 0.79 (0.60, 1.03) | 0.04 | 0.04 |
| Mali | PROMIS (46) | | | | | |
| Mali | PROMIS CS (46) | | | | | |
| Zimbabwe | SHINE (HIV−) (47) | 1871 | 1783 | 0.81 (0.63, 1.05) | 0.04 | 0.04 |
| Zimbabwe | SHINE (HIV+) (48) | 337 | 329 | 0.98 (0.61, 1.57) | 0.01 | 0.01 |
| | **$I^2$ = 0.00, $Tau^2$ = 0.00** | **12585** | **14871** | | | |
| | **Fixed** | | | **0.91 (0.86, 0.95)** | | |
| | **Random** | | | **0.91 (0.86, 0.95)** | | |

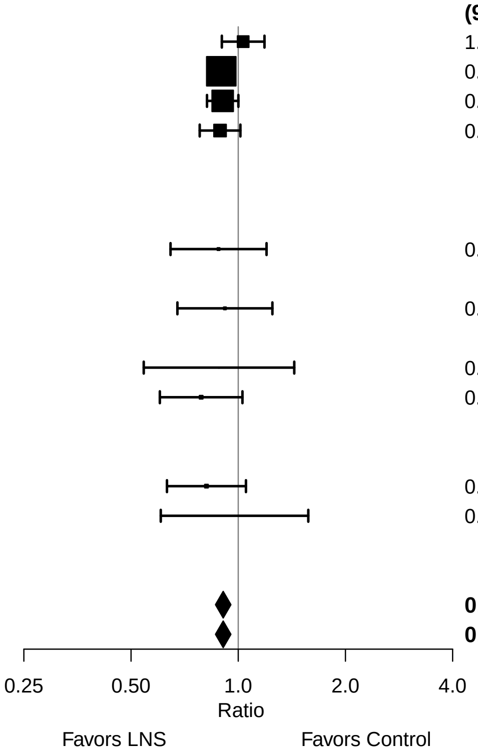

### Supplemental figure 3Q: Small head size prevalence difference

| Country | Trial | LNS N | Control N | PD (95% CI) | Fixed W | Random W |
|---|---|---|---|---|---|---|
| Bangladesh | JiVitA−4 (34) | 2885 | 1267 | 0.00 (−0.03, 0.02) | 0.10 | 0.14 |
| Bangladesh | RDNS (35) | 1663 | 815 | −0.04 (−0.09, 0.00) | 0.03 | 0.06 |
| Bangladesh | WASH−B (36) | 1163 | 3463 | −0.03 (−0.06, 0.00) | 0.06 | 0.09 |
| Burkina Faso | iLiNS−Zinc (37) | 1946 | 664 | −0.04 (−0.08, −0.01) | 0.05 | 0.09 |
| Burkina Faso | PROMIS (38) | | | | | |
| Burkina Faso | PROMIS CS (38) | | | | | |
| Ghana | GHANA (39) | | | | | |
| Ghana | iLiNS−DYADG (40) | 347 | 692 | −0.02 (−0.06, 0.03) | 0.03 | 0.05 |
| Haiti | HAITI (41) | | | | | |
| Kenya | WASH−B (42) | 1461 | 5176 | 0.00 (−0.02, 0.01) | 0.41 | 0.24 |
| Madagascar | MAHAY (43) | | | | | |
| Malawi | iLiNS−DYADM (44) | 220 | 444 | −0.01 (−0.06, 0.04) | 0.02 | 0.05 |
| Malawi | iLiNS−DOSE (45) | 692 | 238 | −0.04 (−0.10, 0.01) | 0.02 | 0.04 |
| Mali | PROMIS (46) | | | | | |
| Mali | PROMIS CS (46) | | | | | |
| Zimbabwe | SHINE (HIV−) (47) | 1871 | 1783 | −0.01 (−0.03, 0.00) | 0.26 | 0.21 |
| Zimbabwe | SHINE (HIV+) (48) | 337 | 329 | 0.00 (−0.05, 0.04) | 0.03 | 0.05 |
| | **I² = 0.23, Tau² = 0.00** | **12585** | **14871** | | | |
| | **Fixed** | | | **−0.01 (−0.02, 0.00)** | | |
| | **Random** | | | **−0.02 (−0.03, 0.00)** | | |

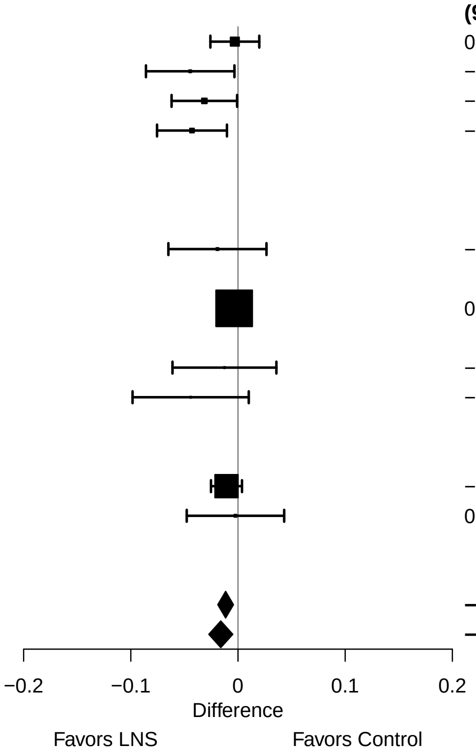

### Supplemental figure 4: Forest plots for effects of SQ-LNS on growth outcomes stratified by study-level effect modifiers

These figures are forest plots showing the study-level effect modification of intervention effects. Each figure shows the study-level estimates along with the corresponding pooled estimate grouped by study-level effect modifier category. For dichotomous outcomes analyzed via prevalence ratios, the effect estimate is the prevalence in the LNS group divided by the prevalence in the control group. For dichotomous outcomes analyzed via prevalence differences, the effect estimate is the prevalence in the LNS group minus the prevalence in the control group. The labels on the left y-axis correspond to trial level information. The values on the right indicate the study level effect estimate, confidence interval, and weighting for deriving the pooled estimates. LAZ, length-for-age z-score; WLZ, weight-for-length z-score; WAZ, weight-for-age z-score; MUACZ, mid-upper arm circumference z-score; HCZ, head circumference-for-age z-score.

### Supplemental figure 4A: Mean difference in LAZ

#### 4A1: Stratified by Geographic region

**Geographic region**
**(p−diff = 0.636)**

| Country | Trial | N | N | MD (95% CI) | W |
|---|---|---|---|---|---|
| **Geographic region − SEAR** | | | | | |
| Bangladesh | JiVitA−4 (34) | 2838 | 1244 | 0.09 (0.03, 0.14) | 0.36 |
| Bangladesh | RDNS (35) | 1663 | 815 | 0.08 (−0.01, 0.16) | 0.30 |
| Bangladesh | WASH−B (36) | 1158 | 3431 | 0.21 (0.14, 0.28) | 0.33 |
| **I² = 0.78, Tau² = 0.00** | | **5659** | **5490** | **0.12 (0.04, 0.21)** | |
| **Geographic region − AFR** | | | | | |
| Burkina Faso | iLiNS−Zinc (37) | 1952 | 664 | 0.31 (0.22, 0.39) | 0.11 |
| Burkina Faso | PROMIS (38) | 863 | 914 | 0.15 (0.02, 0.27) | 0.08 |
| Burkina Faso | PROMIS CS (38) | 430 | 439 | 0.04 (−0.14, 0.22) | 0.05 |
| Ghana | GHANA (39) | 98 | 96 | 0.28 (−0.02, 0.58) | 0.03 |
| Ghana | iLiNS−DYADG (40) | 347 | 692 | 0.20 (0.07, 0.33) | 0.08 |
| Kenya | WASH−B (42) | 1457 | 5137 | 0.15 (0.09, 0.21) | 0.12 |
| Madagascar | MAHAY (43) | 1702 | 1682 | 0.03 (−0.11, 0.17) | 0.07 |
| Malawi | iLiNS−DYADM (44) | 220 | 444 | −0.08 (−0.25, 0.10) | 0.06 |
| Malawi | iLiNS−DOSE (45) | 696 | 241 | 0.06 (−0.04, 0.17) | 0.10 |
| Mali | PROMIS (46) | 506 | 506 | 0.16 (0.01, 0.31) | 0.07 |
| Mali | PROMIS CS (46) | 952 | 969 | 0.22 (0.05, 0.39) | 0.06 |
| Zimbabwe | SHINE (HIV−) (47) | 1880 | 1794 | 0.16 (0.08, 0.23) | 0.11 |
| Zimbabwe | SHINE (HIV+) (48) | 337 | 330 | 0.25 (0.08, 0.43) | 0.06 |
| **I² = 0.60, Tau² = 0.01** | | **11440** | **13908** | **0.15 (0.09, 0.21)** | |

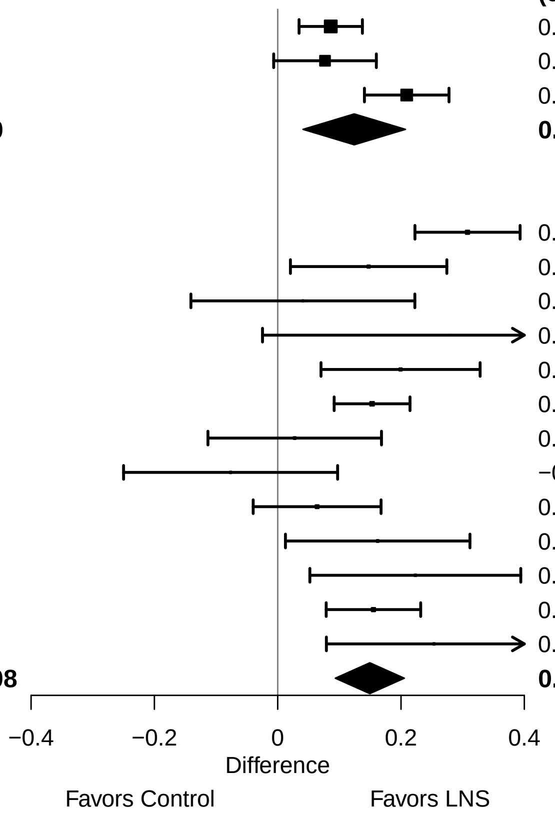

#### Supplemental figure 4A: Mean difference in LAZ

#### 4A2: Stratified by Stunting burden

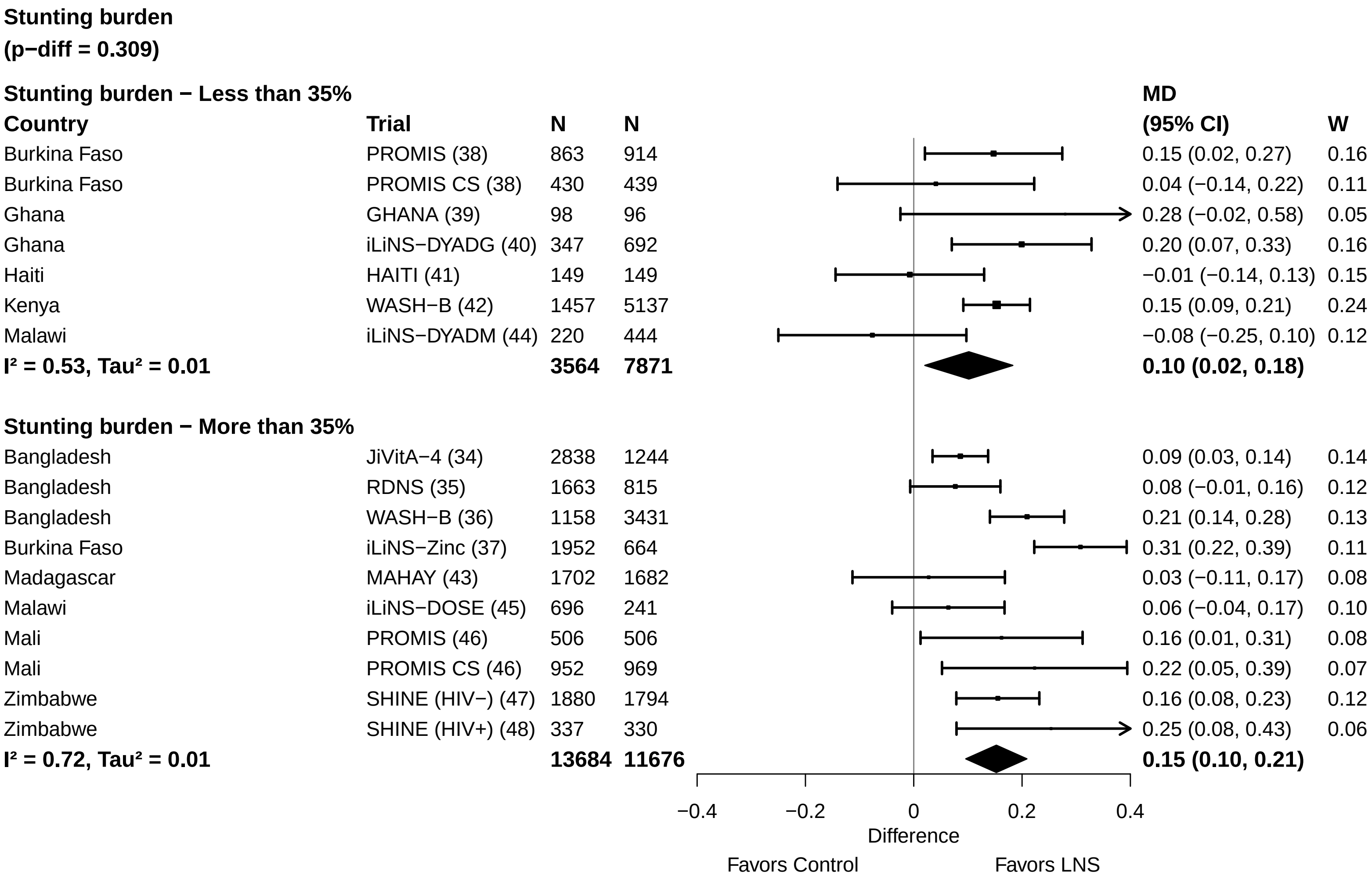

#### Supplemental figure 4A: Mean difference in LAZ

#### 4A3: Stratified by Malaria prevalence

**Malaria prevalence**
**(p−diff = 0.508)**

| Country | Trial | N | N | MD (95% CI) | W |
|---|---|---|---|---|---|
| **Malaria prevalence − Less than 10%** | | | | | |
| Bangladesh | JiVitA−4 (34) | 2838 | 1244 | 0.09 (0.03, 0.14) | 0.16 |
| Bangladesh | RDNS (35) | 1663 | 815 | 0.08 (−0.01, 0.16) | 0.14 |
| Bangladesh | WASH−B (36) | 1158 | 3431 | 0.21 (0.14, 0.28) | 0.15 |
| Haiti | HAITI (41) | 149 | 149 | −0.01 (−0.14, 0.13) | 0.10 |
| Kenya | WASH−B (42) | 1457 | 5137 | 0.15 (0.09, 0.21) | 0.15 |
| Madagascar | MAHAY (43) | 1702 | 1682 | 0.03 (−0.11, 0.17) | 0.09 |
| Zimbabwe | SHINE (HIV−) (47) | 1880 | 1794 | 0.16 (0.08, 0.23) | 0.14 |
| Zimbabwe | SHINE (HIV+) (48) | 337 | 330 | 0.25 (0.08, 0.43) | 0.07 |
| **I² = 0.62, Tau² = 0.00** | | **11184** | **14582** | **0.12 (0.07, 0.18)** | |
| **Malaria prevalence − At least 10%** | | | | | |
| Burkina Faso | iLiNS−Zinc (37) | 1952 | 664 | 0.31 (0.22, 0.39) | 0.17 |
| Burkina Faso | PROMIS (38) | 863 | 914 | 0.15 (0.02, 0.27) | 0.13 |
| Burkina Faso | PROMIS CS (38) | 430 | 439 | 0.04 (−0.14, 0.22) | 0.09 |
| Ghana | GHANA (39) | 98 | 96 | 0.28 (−0.02, 0.58) | 0.04 |
| Ghana | iLiNS−DYADG (40) | 347 | 692 | 0.20 (0.07, 0.33) | 0.13 |
| Malawi | iLiNS−DYADM (44) | 220 | 444 | −0.08 (−0.25, 0.10) | 0.09 |
| Malawi | iLiNS−DOSE (45) | 696 | 241 | 0.06 (−0.04, 0.17) | 0.15 |
| Mali | PROMIS (46) | 506 | 506 | 0.16 (0.01, 0.31) | 0.11 |
| Mali | PROMIS CS (46) | 952 | 969 | 0.22 (0.05, 0.39) | 0.10 |
| **I² = 0.68, Tau² = 0.01** | | **6064** | **4965** | **0.15 (0.07, 0.23)** | |

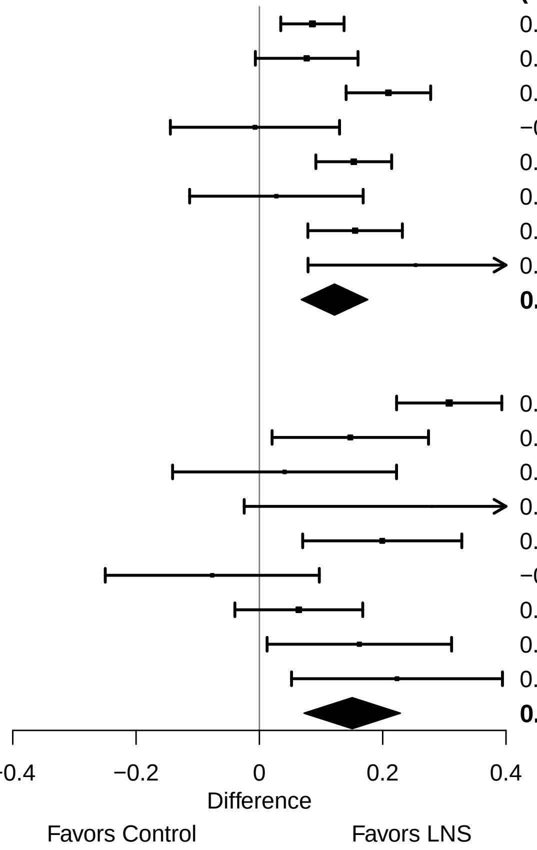

#### Supplemental figure 4A: Mean difference in LAZ

#### 4A4: Stratified by Source water quality

**Source water quality**
**(p−diff = 0.313)**

| Country | Trial | N | N | MD (95% CI) | W |
|---|---|---|---|---|---|
| **Source water quality − Improved** | | | | | |
| Bangladesh | JiVitA−4 (34) | 2838 | 1244 | 0.09 (0.03, 0.14) | 0.19 |
| Bangladesh | RDNS (35) | 1663 | 815 | 0.08 (−0.01, 0.16) | 0.16 |
| Bangladesh | WASH−B (36) | 567 | 1104 | 0.26 (0.16, 0.36) | 0.14 |
| Ghana | GHANA (39) | 98 | 96 | 0.28 (−0.02, 0.58) | 0.04 |
| Ghana | iLiNS−DYADG (40) | 347 | 692 | 0.20 (0.07, 0.33) | 0.12 |
| Haiti | HAITI (41) | 149 | 149 | −0.01 (−0.14, 0.13) | 0.11 |
| Malawi | iLiNS−DYADM (44) | 220 | 444 | −0.08 (−0.25, 0.10) | 0.09 |
| Malawi | iLiNS−DOSE (45) | 696 | 241 | 0.06 (−0.04, 0.17) | 0.14 |
| **I² = 0.65, Tau² = 0.01** | | **6578** | **4785** | **0.10 (0.02, 0.18)** | |
| **Source water quality − Unimproved** | | | | | |
| Burkina Faso | iLiNS−Zinc (37) | 1952 | 664 | 0.31 (0.22, 0.39) | 0.16 |
| Burkina Faso | PROMIS (38) | 863 | 914 | 0.15 (0.02, 0.27) | 0.12 |
| Burkina Faso | PROMIS CS (38) | 430 | 439 | 0.04 (−0.14, 0.22) | 0.09 |
| Kenya | WASH−B (42) | 695 | 2253 | 0.11 (0.00, 0.21) | 0.14 |
| Madagascar | MAHAY (43) | 1702 | 1682 | 0.03 (−0.11, 0.17) | 0.11 |
| Mali | PROMIS (46) | 506 | 506 | 0.16 (0.01, 0.31) | 0.11 |
| Mali | PROMIS CS (46) | 952 | 969 | 0.22 (0.05, 0.39) | 0.09 |
| Zimbabwe | SHINE (HIV−) (47) | 892 | 880 | 0.11 (0.00, 0.21) | 0.14 |
| Zimbabwe | SHINE (HIV+) (48) | 147 | 146 | 0.27 (−0.01, 0.55) | 0.05 |
| **I² = 0.59, Tau² = 0.00** | | **8139** | **8453** | **0.15 (0.09, 0.22)** | |

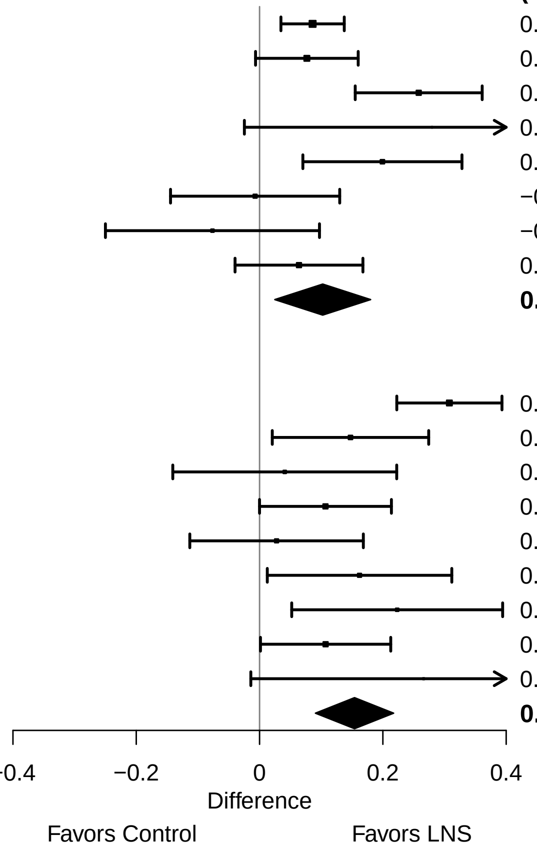

#### Supplemental figure 4A: Mean difference in LAZ

### 4A5: Stratified by Sanitation

**Sanitation**
**(p−diff = 0.694)**

| **Sanitation − Improved** | | | | **MD** | |
|---|---|---|---|---|---|
| **Country** | **Trial** | **N** | **N** | **(95% CI)** | **W** |
| Bangladesh | JiVitA−4 (34) | 2838 | 1244 | 0.09 (0.03, 0.14) | 0.16 |
| Bangladesh | RDNS (35) | 1663 | 815 | 0.08 (−0.01, 0.16) | 0.14 |
| Bangladesh | WASH−B (36) | 567 | 1104 | 0.26 (0.16, 0.36) | 0.12 |
| Burkina Faso | PROMIS (38) | 863 | 914 | 0.15 (0.02, 0.27) | 0.10 |
| Burkina Faso | PROMIS CS (38) | 430 | 439 | 0.04 (−0.14, 0.22) | 0.07 |
| Ghana | GHANA (39) | 98 | 96 | 0.28 (−0.02, 0.58) | 0.04 |
| Ghana | iLiNS−DYADG (40) | 347 | 692 | 0.20 (0.07, 0.33) | 0.10 |
| Haiti | HAITI (41) | 149 | 149 | −0.01 (−0.14, 0.13) | 0.10 |
| Mali | PROMIS (46) | 506 | 506 | 0.16 (0.01, 0.31) | 0.09 |
| Mali | PROMIS CS (46) | 952 | 969 | 0.22 (0.05, 0.39) | 0.08 |
| **I² = 0.50, Tau² = 0.00** | | **8413** | **6928** | **0.13 (0.08, 0.19)** | |
| **Sanitation − Unimproved** | | | | | |
| Burkina Faso | iLiNS−Zinc (37) | 1952 | 664 | 0.31 (0.22, 0.39) | 0.19 |
| Kenya | WASH−B (42) | 695 | 2253 | 0.11 (0.00, 0.21) | 0.17 |
| Madagascar | MAHAY (43) | 1702 | 1682 | 0.03 (−0.11, 0.17) | 0.14 |
| Malawi | iLiNS−DYADM (44) | 220 | 444 | −0.08 (−0.25, 0.10) | 0.11 |
| Malawi | iLiNS−DOSE (45) | 696 | 241 | 0.06 (−0.04, 0.17) | 0.17 |
| Zimbabwe | SHINE (HIV−) (47) | 892 | 880 | 0.11 (0.00, 0.21) | 0.17 |
| Zimbabwe | SHINE (HIV+) (48) | 147 | 146 | 0.27 (−0.01, 0.55) | 0.06 |
| **I² = 0.78, Tau² = 0.01** | | **6304** | **6310** | **0.11 (0.02, 0.21)** | |

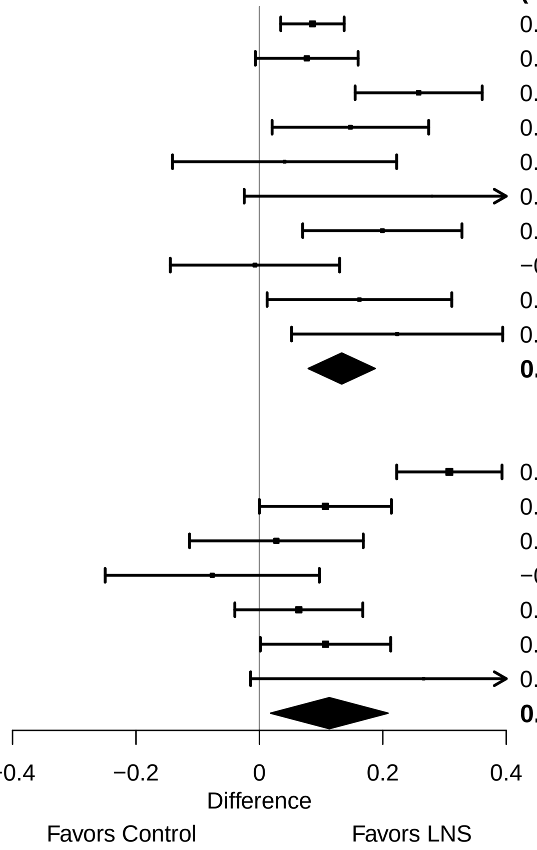

#### Supplemental figure 4A: Mean difference in LAZ

### 4A6: Stratified by Supplement duration

**Supplement duration**
**(p−diff = 0.461)**

| Country | Trial | N | N | MD (95% CI) | W |
|---|---|---|---|---|---|
| **Supplement duration − 12m or less** | | | | | |
| Bangladesh | JiVitA−4 (34) | 2838 | 1244 | 0.09 (0.03, 0.14) | 0.13 |
| Burkina Faso | iLiNS−Zinc (37) | 1952 | 664 | 0.31 (0.22, 0.39) | 0.11 |
| Burkina Faso | PROMIS (38) | 863 | 914 | 0.15 (0.02, 0.27) | 0.09 |
| Burkina Faso | PROMIS CS (38) | 430 | 439 | 0.04 (−0.14, 0.22) | 0.06 |
| Ghana | GHANA (39) | 98 | 96 | 0.28 (−0.02, 0.58) | 0.03 |
| Ghana | iLiNS−DYADG (40) | 347 | 692 | 0.20 (0.07, 0.33) | 0.08 |
| Haiti | HAITI (41) | 149 | 149 | −0.01 (−0.14, 0.13) | 0.08 |
| Madagascar | MAHAY (43) | 1702 | 1682 | 0.03 (−0.11, 0.17) | 0.08 |
| Malawi | iLiNS−DYADM (44) | 220 | 444 | −0.08 (−0.25, 0.10) | 0.06 |
| Malawi | iLiNS−DOSE (45) | 696 | 241 | 0.06 (−0.04, 0.17) | 0.10 |
| Zimbabwe | SHINE (HIV−) (47) | 1880 | 1794 | 0.16 (0.08, 0.23) | 0.12 |
| Zimbabwe | SHINE (HIV+) (48) | 337 | 330 | 0.25 (0.08, 0.43) | 0.06 |
| **$I^2$ = 0.71, $Tau^2$ = 0.01** | | **11512** | **8689** | **0.12 (0.06, 0.19)** | |
| **Supplement duration − > 12m** | | | | | |
| Bangladesh | RDNS (35) | 1663 | 815 | 0.08 (−0.01, 0.16) | 0.23 |
| Bangladesh | WASH−B (36) | 1158 | 3431 | 0.21 (0.14, 0.28) | 0.25 |
| Kenya | WASH−B (42) | 1457 | 5137 | 0.15 (0.09, 0.21) | 0.25 |
| Mali | PROMIS (46) | 506 | 506 | 0.16 (0.01, 0.31) | 0.15 |
| Mali | PROMIS CS (46) | 952 | 969 | 0.22 (0.05, 0.39) | 0.13 |
| **$I^2$ = 0.37, $Tau^2$ = 0.00** | | **5736** | **10858** | **0.16 (0.11, 0.21)** | |

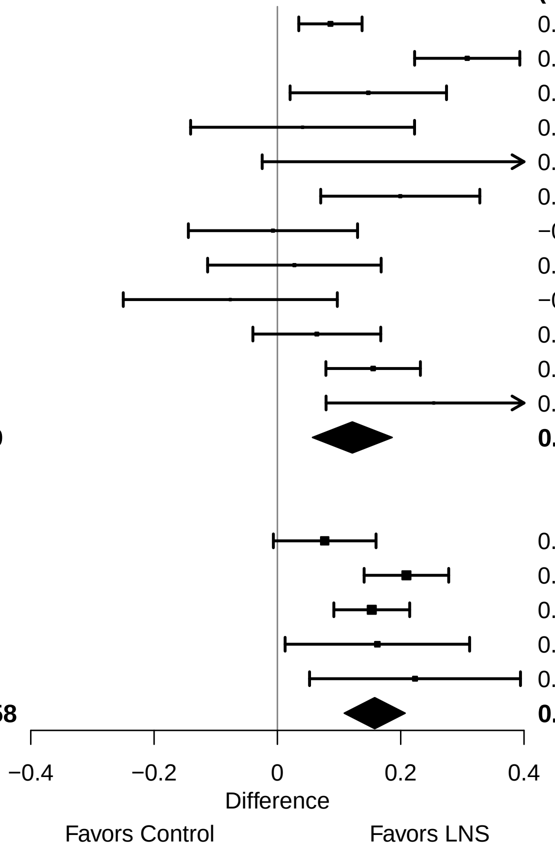

#### Supplemental figure 4A: Mean difference in LAZ

#### 4A7: Stratified by Frequency of contact

**Frequency of contact**
**(p−diff = 0.527)**

**Frequency of contact − Monthly**

| Country | Trial | N | N | MD (95% CI) | W |
|---|---|---|---|---|---|
| Bangladesh | RDNS (35) | 1663 | 815 | 0.08 (−0.01, 0.16) | 0.13 |
| Burkina Faso | PROMIS (38) | 863 | 914 | 0.15 (0.02, 0.27) | 0.10 |
| Burkina Faso | PROMIS CS (38) | 430 | 439 | 0.04 (−0.14, 0.22) | 0.07 |
| Haiti | HAITI (41) | 149 | 149 | −0.01 (−0.14, 0.13) | 0.09 |
| Kenya | WASH−B (42) | 1457 | 5137 | 0.15 (0.09, 0.21) | 0.15 |
| Madagascar | MAHAY (43) | 1702 | 1682 | 0.03 (−0.11, 0.17) | 0.09 |
| Mali | PROMIS (46) | 506 | 506 | 0.16 (0.01, 0.31) | 0.09 |
| Mali | PROMIS CS (46) | 952 | 969 | 0.22 (0.05, 0.39) | 0.07 |
| Zimbabwe | SHINE (HIV−) (47) | 1880 | 1794 | 0.16 (0.08, 0.23) | 0.14 |
| Zimbabwe | SHINE (HIV+) (48) | 337 | 330 | 0.25 (0.08, 0.43) | 0.07 |
| **$I^2$ = 0.29, $Tau^2$ = 0.00** | | **9939** | **12735** | **0.12 (0.08, 0.17)** | |

**Frequency of contact − Weekly**

| Country | Trial | N | N | MD (95% CI) | W |
|---|---|---|---|---|---|
| Bangladesh | JiVitA−4 (34) | 2838 | 1244 | 0.09 (0.03, 0.14) | 0.21 |
| Bangladesh | WASH−B (36) | 1158 | 3431 | 0.21 (0.14, 0.28) | 0.19 |
| Burkina Faso | iLiNS−Zinc (37) | 1952 | 664 | 0.31 (0.22, 0.39) | 0.17 |
| Ghana | GHANA (39) | 98 | 96 | 0.28 (−0.02, 0.58) | 0.04 |
| Ghana | iLiNS−DYADG (40) | 347 | 692 | 0.20 (0.07, 0.33) | 0.13 |
| Malawi | iLiNS−DYADM (44) | 220 | 444 | −0.08 (−0.25, 0.10) | 0.10 |
| Malawi | iLiNS−DOSE (45) | 696 | 241 | 0.06 (−0.04, 0.17) | 0.16 |
| **$I^2$ = 0.81, $Tau^2$ = 0.01** | | **7309** | **6812** | **0.15 (0.06, 0.25)** | |

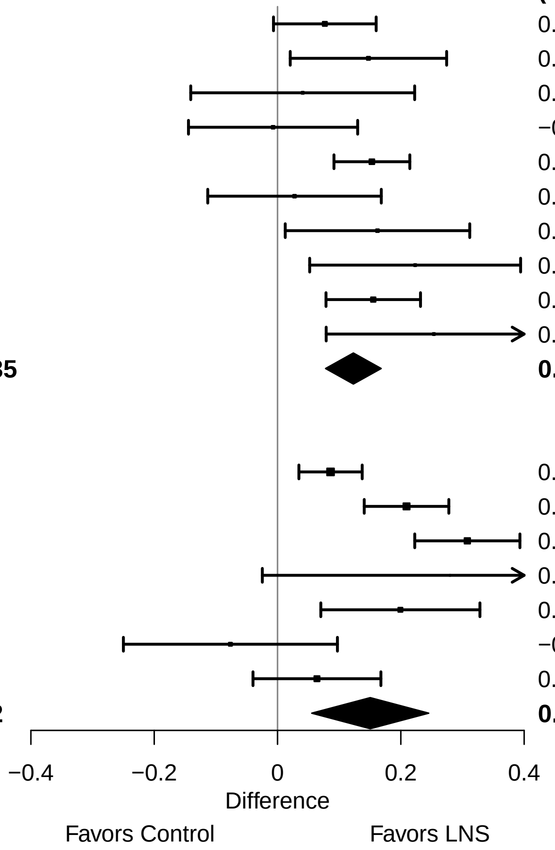

#### Supplemental figure 4A: Mean difference in LAZ

### 4A8: Stratified by Average SQ-LNS compliance

**Average SQ−LNS compliance**
**(p−diff = 0.724)**

**Average SQ−LNS compliance − Low**

| Country | Trial | N | N | MD (95% CI) | W |
|---|---|---|---|---|---|
| Burkina Faso | PROMIS (38) | 863 | 914 | 0.15 (0.02, 0.27) | 0.12 |
| Burkina Faso | PROMIS CS (38) | 430 | 439 | 0.04 (−0.14, 0.22) | 0.08 |
| Ghana | iLiNS−DYADG (40) | 347 | 692 | 0.20 (0.07, 0.33) | 0.12 |
| Malawi | iLiNS−DYADM (44) | 220 | 444 | −0.08 (−0.25, 0.10) | 0.09 |
| Malawi | iLiNS−DOSE (45) | 696 | 241 | 0.06 (−0.04, 0.17) | 0.14 |
| Mali | PROMIS (46) | 506 | 506 | 0.16 (0.01, 0.31) | 0.10 |
| Mali | PROMIS CS (46) | 952 | 969 | 0.22 (0.05, 0.39) | 0.09 |
| Zimbabwe | SHINE (HIV−) (47) | 1880 | 1794 | 0.16 (0.08, 0.23) | 0.17 |
| Zimbabwe | SHINE (HIV+) (48) | 337 | 330 | 0.25 (0.08, 0.43) | 0.09 |
| **I² = 0.37, Tau² = 0.00** | | **6231** | **6329** | **0.13 (0.07, 0.19)** | |

**Average SQ−LNS compliance − High**

| Country | Trial | N | N | MD (95% CI) | W |
|---|---|---|---|---|---|
| Bangladesh | JiVitA−4 (34) | 2838 | 1244 | 0.09 (0.03, 0.14) | 0.19 |
| Bangladesh | RDNS (35) | 1663 | 815 | 0.08 (−0.01, 0.16) | 0.16 |
| Bangladesh | WASH−B (36) | 1158 | 3431 | 0.21 (0.14, 0.28) | 0.17 |
| Burkina Faso | iLiNS−Zinc (37) | 1952 | 664 | 0.31 (0.22, 0.39) | 0.16 |
| Ghana | GHANA (39) | 98 | 96 | 0.28 (−0.02, 0.58) | 0.04 |
| Haiti | HAITI (41) | 149 | 149 | −0.01 (−0.14, 0.13) | 0.11 |
| Kenya | WASH−B (42) | 1457 | 5137 | 0.15 (0.09, 0.21) | 0.18 |
| **I² = 0.80, Tau² = 0.01** | | **9315** | **11536** | **0.15 (0.07, 0.23)** | |

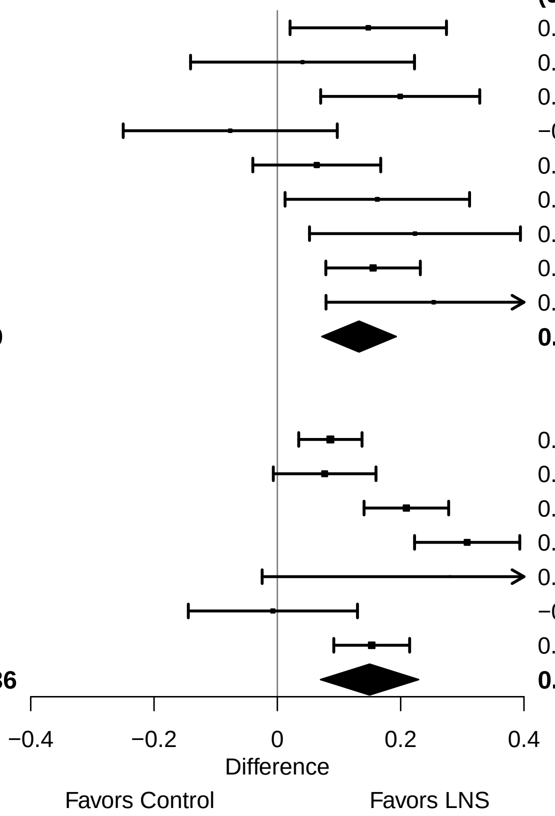

### Supplemental figure 4B: Stunting prevalence ratio

#### 4B1: Stratified by Geographic region

**Geographic region**
**(p−diff = 0.646)**

| Country | Trial | N | N | PR (95% CI) | W |
|---|---|---|---|---|---|
| **Geographic region − SEAR** | | | | | |
| Bangladesh | JiVitA−4 (34) | 2838 | 1244 | 0.92 (0.84, 1.01) | 0.32 |
| Bangladesh | RDNS (35) | 1663 | 815 | 0.92 (0.85, 1.00) | 0.34 |
| Bangladesh | WASH−B (36) | 1158 | 3431 | 0.84 (0.77, 0.91) | 0.34 |
| **I² = 0.36, Tau² = 0.00** | | **5659** | **5490** | **0.89 (0.84, 0.95)** | |
| **Geographic region − AFR** | | | | | |
| Burkina Faso | iLiNS−Zinc (37) | 1952 | 664 | 0.75 (0.65, 0.87) | 0.09 |
| Burkina Faso | PROMIS (38) | 863 | 914 | 0.82 (0.67, 1.00) | 0.07 |
| Burkina Faso | PROMIS CS (38) | 430 | 439 | 0.87 (0.63, 1.19) | 0.03 |
| Ghana | GHANA (39) | 98 | 96 | 0.70 (0.23, 2.13) | 0.00 |
| Ghana | iLiNS−DYADG (40) | 347 | 692 | 0.66 (0.45, 0.98) | 0.02 |
| Kenya | WASH−B (42) | 1457 | 5137 | 0.86 (0.78, 0.94) | 0.14 |
| Madagascar | MAHAY (43) | 1702 | 1682 | 0.98 (0.90, 1.07) | 0.14 |
| Malawi | iLiNS−DYADM (44) | 220 | 444 | 1.10 (0.89, 1.36) | 0.06 |
| Malawi | iLiNS−DOSE (45) | 696 | 241 | 1.00 (0.87, 1.15) | 0.10 |
| Mali | PROMIS (46) | 506 | 506 | 0.92 (0.76, 1.10) | 0.07 |
| Mali | PROMIS CS (46) | 952 | 969 | 0.78 (0.65, 0.95) | 0.07 |
| Zimbabwe | SHINE (HIV−) (47) | 1880 | 1794 | 0.79 (0.72, 0.87) | 0.13 |
| Zimbabwe | SHINE (HIV+) (48) | 337 | 330 | 0.81 (0.68, 0.96) | 0.08 |
| **I² = 0.57, Tau² = 0.01** | | **11440** | **13908** | **0.87 (0.81, 0.93)** | |

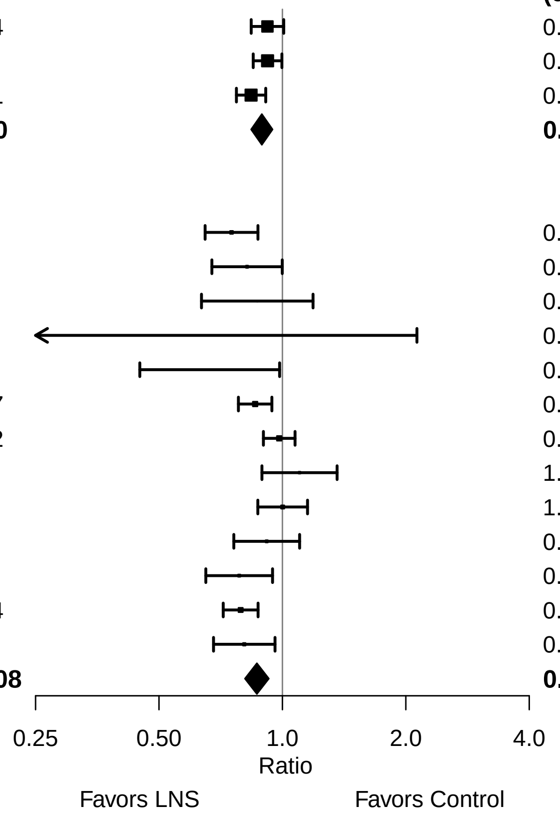

##### Supplemental figure 4B: Stunting prevalence ratio

#### 4B2: Stratified by Stunting burden

**Stunting burden**
**(p−diff = 0.912)**

| Country | Trial | N | N | PR (95% CI) | W |
|---|---|---|---|---|---|
| **Stunting burden − Less than 35%** | | | | | |
| Burkina Faso | PROMIS (38) | 863 | 914 | 0.82 (0.67, 1.00) | 0.20 |
| Burkina Faso | PROMIS CS (38) | 430 | 439 | 0.87 (0.63, 1.19) | 0.10 |
| Ghana | GHANA (39) | 98 | 96 | 0.70 (0.23, 2.13) | 0.01 |
| Ghana | iLiNS−DYADG (40) | 347 | 692 | 0.66 (0.45, 0.98) | 0.06 |
| Haiti | HAITI (41) | 149 | 149 | 1.04 (0.60, 1.81) | 0.03 |
| Kenya | WASH−B (42) | 1457 | 5137 | 0.86 (0.78, 0.94) | 0.42 |
| Malawi | iLiNS−DYADM (44) | 220 | 444 | 1.10 (0.89, 1.36) | 0.18 |
| **I² = 0.21, Tau² = 0.00** | | **3564** | **7871** | **0.88 (0.79, 0.97)** | |
| **Stunting burden − More than 35%** | | | | | |
| Bangladesh | JiVitA−4 (34) | 2838 | 1244 | 0.92 (0.84, 1.01) | 0.12 |
| Bangladesh | RDNS (35) | 1663 | 815 | 0.92 (0.85, 1.00) | 0.13 |
| Bangladesh | WASH−B (36) | 1158 | 3431 | 0.84 (0.77, 0.91) | 0.13 |
| Burkina Faso | iLiNS−Zinc (37) | 1952 | 664 | 0.75 (0.65, 0.87) | 0.08 |
| Madagascar | MAHAY (43) | 1702 | 1682 | 0.98 (0.90, 1.07) | 0.13 |
| Malawi | iLiNS−DOSE (45) | 696 | 241 | 1.00 (0.87, 1.15) | 0.09 |
| Mali | PROMIS (46) | 506 | 506 | 0.92 (0.76, 1.10) | 0.06 |
| Mali | PROMIS CS (46) | 952 | 969 | 0.78 (0.65, 0.95) | 0.06 |
| Zimbabwe | SHINE (HIV−) (47) | 1880 | 1794 | 0.79 (0.72, 0.87) | 0.12 |
| Zimbabwe | SHINE (HIV+) (48) | 337 | 330 | 0.81 (0.68, 0.96) | 0.07 |
| **I² = 0.63, Tau² = 0.01** | | **13684** | **11676** | **0.87 (0.82, 0.93)** | |

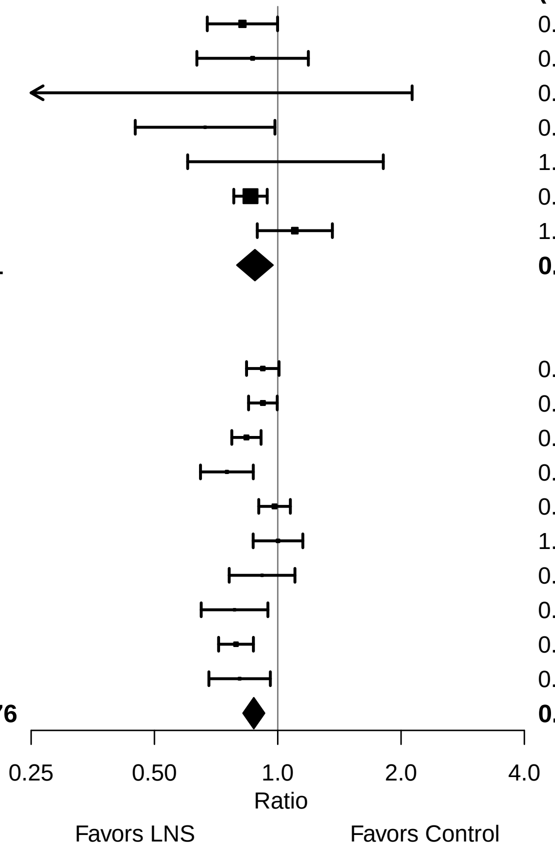

#### Supplemental figure 4B: Stunting prevalence ratio

### 4B3: Stratified by Malaria prevalence

**Malaria prevalence**
**(p−diff = 0.862)**

| Country | Trial | N | N | PR (95% CI) | W |
|---|---|---|---|---|---|
| **Malaria prevalence − Less than 10%** | | | | | |
| Bangladesh | JiVitA−4 (34) | 2838 | 1244 | 0.92 (0.84, 1.01) | 0.15 |
| Bangladesh | RDNS (35) | 1663 | 815 | 0.92 (0.85, 1.00) | 0.16 |
| Bangladesh | WASH−B (36) | 1158 | 3431 | 0.84 (0.77, 0.91) | 0.16 |
| Haiti | HAITI (41) | 149 | 149 | 1.04 (0.60, 1.81) | 0.01 |
| Kenya | WASH−B (42) | 1457 | 5137 | 0.86 (0.78, 0.94) | 0.15 |
| Madagascar | MAHAY (43) | 1702 | 1682 | 0.98 (0.90, 1.07) | 0.15 |
| Zimbabwe | SHINE (HIV−) (47) | 1880 | 1794 | 0.79 (0.72, 0.87) | 0.14 |
| Zimbabwe | SHINE (HIV+) (48) | 337 | 330 | 0.81 (0.68, 0.96) | 0.08 |
| **I² = 0.55, Tau² = 0.00** | | **11184** | **14582** | **0.88 (0.83, 0.93)** | |
| **Malaria prevalence − At least 10%** | | | | | |
| Burkina Faso | iLiNS−Zinc (37) | 1952 | 664 | 0.75 (0.65, 0.87) | 0.18 |
| Burkina Faso | PROMIS (38) | 863 | 914 | 0.82 (0.67, 1.00) | 0.13 |
| Burkina Faso | PROMIS CS (38) | 430 | 439 | 0.87 (0.63, 1.19) | 0.06 |
| Ghana | GHANA (39) | 98 | 96 | 0.70 (0.23, 2.13) | 0.01 |
| Ghana | iLiNS−DYADG (40) | 347 | 692 | 0.66 (0.45, 0.98) | 0.04 |
| Malawi | iLiNS−DYADM (44) | 220 | 444 | 1.10 (0.89, 1.36) | 0.12 |
| Malawi | iLiNS−DOSE (45) | 696 | 241 | 1.00 (0.87, 1.15) | 0.19 |
| Mali | PROMIS (46) | 506 | 506 | 0.92 (0.76, 1.10) | 0.14 |
| Mali | PROMIS CS (46) | 952 | 969 | 0.78 (0.65, 0.95) | 0.14 |
| **I² = 0.51, Tau² = 0.01** | | **6064** | **4965** | **0.87 (0.79, 0.96)** | |

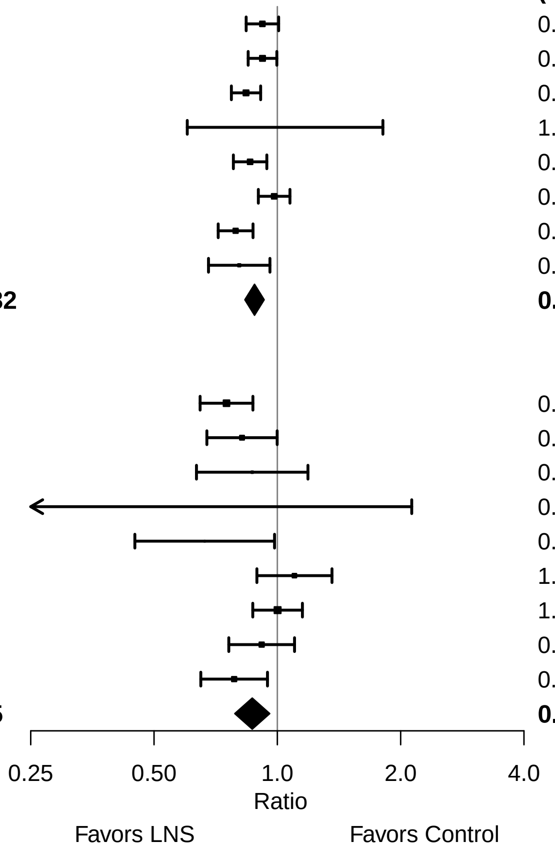

**Supplemental figure 4B: Stunting prevalence ratio**

#### 4B4: Stratified by Source water quality

**Source water quality**
**(p−diff = 0.267)**

| Country | Trial | N | N | PR (95% CI) | W |
|---|---|---|---|---|---|
| **Source water quality − Improved** | | | | | |
| Bangladesh | JiVitA−4 (34) | 2838 | 1244 | 0.92 (0.84, 1.01) | 0.24 |
| Bangladesh | RDNS (35) | 1663 | 815 | 0.92 (0.85, 1.00) | 0.27 |
| Bangladesh | WASH−B (36) | 567 | 1104 | 0.80 (0.70, 0.92) | 0.17 |
| Ghana | GHANA (39) | 98 | 96 | 0.70 (0.23, 2.13) | 0.00 |
| Ghana | iLiNS−DYADG (40) | 347 | 692 | 0.66 (0.45, 0.98) | 0.03 |
| Haiti | HAITI (41) | 149 | 149 | 1.04 (0.60, 1.81) | 0.02 |
| Malawi | iLiNS−DYADM (44) | 220 | 444 | 1.10 (0.89, 1.36) | 0.10 |
| Malawi | iLiNS−DOSE (45) | 696 | 241 | 1.00 (0.87, 1.15) | 0.17 |
| **I² = 0.38, Tau² = 0.01** | | **6578** | **4785** | **0.92 (0.84, 1.00)** | |
| **Source water quality − Unimproved** | | | | | |
| Burkina Faso | iLiNS−Zinc (37) | 1952 | 664 | 0.75 (0.65, 0.87) | 0.13 |
| Burkina Faso | PROMIS (38) | 863 | 914 | 0.82 (0.67, 1.00) | 0.09 |
| Burkina Faso | PROMIS CS (38) | 430 | 439 | 0.87 (0.63, 1.19) | 0.04 |
| Kenya | WASH−B (42) | 695 | 2253 | 0.92 (0.79, 1.08) | 0.12 |
| Madagascar | MAHAY (43) | 1702 | 1682 | 0.98 (0.90, 1.07) | 0.21 |
| Mali | PROMIS (46) | 506 | 506 | 0.92 (0.76, 1.10) | 0.10 |
| Mali | PROMIS CS (46) | 952 | 969 | 0.78 (0.65, 0.95) | 0.10 |
| Zimbabwe | SHINE (HIV−) (47) | 892 | 880 | 0.85 (0.74, 0.97) | 0.15 |
| Zimbabwe | SHINE (HIV+) (48) | 147 | 146 | 0.79 (0.61, 1.04) | 0.06 |
| **I² = 0.41, Tau² = 0.00** | | **8139** | **8453** | **0.87 (0.81, 0.93)** | |

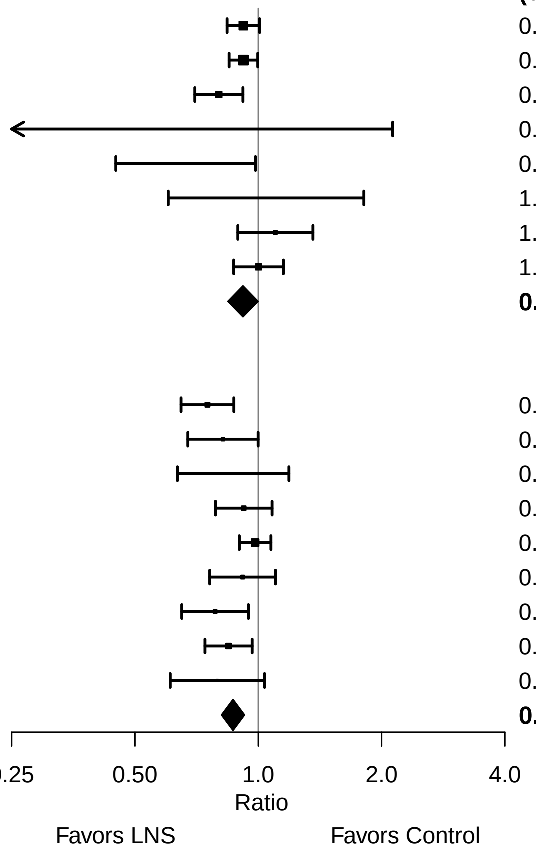

#### Supplemental figure 4B: Stunting prevalence ratio

##### 4B5: Stratified by Sanitation

**Sanitation**
**(p−diff = 0.328)**

| Country | Trial | N | N | PR (95% CI) | W |
|---|---|---|---|---|---|
| **Sanitation − Improved** | | | | | |
| Bangladesh | JiVitA−4 (34) | 2838 | 1244 | 0.92 (0.84, 1.01) | 0.22 |
| Bangladesh | RDNS (35) | 1663 | 815 | 0.92 (0.85, 1.00) | 0.24 |
| Bangladesh | WASH−B (36) | 567 | 1104 | 0.80 (0.70, 0.92) | 0.15 |
| Burkina Faso | PROMIS (38) | 863 | 914 | 0.82 (0.67, 1.00) | 0.09 |
| Burkina Faso | PROMIS CS (38) | 430 | 439 | 0.87 (0.63, 1.19) | 0.04 |
| Ghana | GHANA (39) | 98 | 96 | 0.70 (0.23, 2.13) | 0.00 |
| Ghana | iLiNS−DYADG (40) | 347 | 692 | 0.66 (0.45, 0.98) | 0.03 |
| Haiti | HAITI (41) | 149 | 149 | 1.04 (0.60, 1.81) | 0.02 |
| Mali | PROMIS (46) | 506 | 506 | 0.92 (0.76, 1.10) | 0.10 |
| Mali | PROMIS CS (46) | 952 | 969 | 0.78 (0.65, 0.95) | 0.10 |
| **I² = 0.00, Tau² = 0.00** | | **8413** | **6928** | **0.88 (0.84, 0.93)** | |
| **Sanitation − Unimproved** | | | | | |
| Burkina Faso | iLiNS−Zinc (37) | 1952 | 664 | 0.75 (0.65, 0.87) | 0.15 |
| Kenya | WASH−B (42) | 695 | 2253 | 0.92 (0.79, 1.08) | 0.13 |
| Madagascar | MAHAY (43) | 1702 | 1682 | 0.98 (0.90, 1.07) | 0.24 |
| Malawi | iLiNS−DYADM (44) | 220 | 444 | 1.10 (0.89, 1.36) | 0.09 |
| Malawi | iLiNS−DOSE (45) | 696 | 241 | 1.00 (0.87, 1.15) | 0.16 |
| Zimbabwe | SHINE (HIV−) (47) | 892 | 880 | 0.85 (0.74, 0.97) | 0.17 |
| Zimbabwe | SHINE (HIV+) (48) | 147 | 146 | 0.79 (0.61, 1.04) | 0.06 |
| **I² = 0.63, Tau² = 0.01** | | **6304** | **6310** | **0.91 (0.83, 1.00)** | |

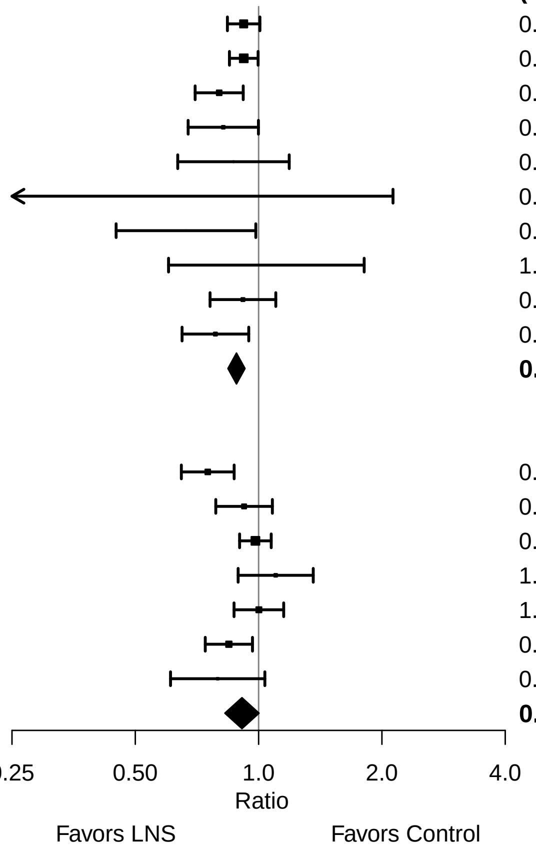

#### Supplemental figure 4B: Stunting prevalence ratio

### 4B6: Stratified by Supplement duration

**Supplement duration**
**(p−diff = 0.742)**

| Country | Trial | N | N | PR (95% CI) | W |
|---|---|---|---|---|---|
| **Supplement duration − 12m or less** | | | | | |
| Bangladesh | JiVitA−4 (34) | 2838 | 1244 | 0.92 (0.84, 1.01) | 0.16 |
| Burkina Faso | iLiNS−Zinc (37) | 1952 | 664 | 0.75 (0.65, 0.87) | 0.11 |
| Burkina Faso | PROMIS (38) | 863 | 914 | 0.82 (0.67, 1.00) | 0.07 |
| Burkina Faso | PROMIS CS (38) | 430 | 439 | 0.87 (0.63, 1.19) | 0.04 |
| Ghana | GHANA (39) | 98 | 96 | 0.70 (0.23, 2.13) | 0.00 |
| Ghana | iLiNS−DYADG (40) | 347 | 692 | 0.66 (0.45, 0.98) | 0.02 |
| Haiti | HAITI (41) | 149 | 149 | 1.04 (0.60, 1.81) | 0.01 |
| Madagascar | MAHAY (43) | 1702 | 1682 | 0.98 (0.90, 1.07) | 0.16 |
| Malawi | iLiNS−DYADM (44) | 220 | 444 | 1.10 (0.89, 1.36) | 0.07 |
| Malawi | iLiNS−DOSE (45) | 696 | 241 | 1.00 (0.87, 1.15) | 0.11 |
| Zimbabwe | SHINE (HIV−) (47) | 1880 | 1794 | 0.79 (0.72, 0.87) | 0.15 |
| Zimbabwe | SHINE (HIV+) (48) | 337 | 330 | 0.81 (0.68, 0.96) | 0.09 |
| **I² = 0.59, Tau² = 0.01** | | **11512** | **8689** | **0.88 (0.82, 0.95)** | |
| **Supplement duration − > 12m** | | | | | |
| Bangladesh | RDNS (35) | 1663 | 815 | 0.92 (0.85, 1.00) | 0.26 |
| Bangladesh | WASH−B (36) | 1158 | 3431 | 0.84 (0.77, 0.91) | 0.26 |
| Kenya | WASH−B (42) | 1457 | 5137 | 0.86 (0.78, 0.94) | 0.24 |
| Mali | PROMIS (46) | 506 | 506 | 0.92 (0.76, 1.10) | 0.12 |
| Mali | PROMIS CS (46) | 952 | 969 | 0.78 (0.65, 0.95) | 0.12 |
| **I² = 0.04, Tau² = 0.00** | | **5736** | **10858** | **0.87 (0.83, 0.91)** | |

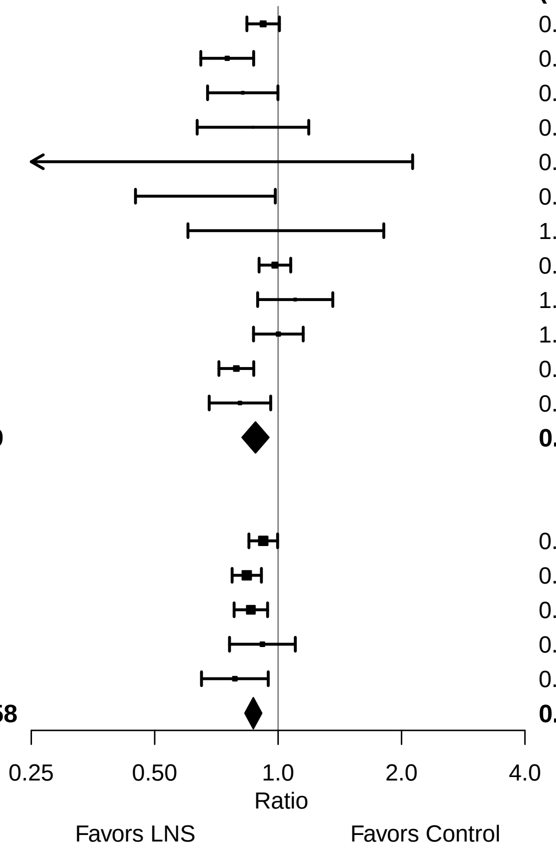

#### Supplemental figure 4B: Stunting prevalence ratio

### 4B7: Stratified by Frequency of contact

**Frequency of contact**
**(p−diff = 0.734)**

**Frequency of contact − Monthly**

| Country | Trial | N | N | PR (95% CI) | W |
|---|---|---|---|---|---|
| Bangladesh | RDNS (35) | 1663 | 815 | 0.92 (0.85, 1.00) | 0.17 |
| Burkina Faso | PROMIS (38) | 863 | 914 | 0.82 (0.67, 1.00) | 0.07 |
| Burkina Faso | PROMIS CS (38) | 430 | 439 | 0.87 (0.63, 1.19) | 0.04 |
| Haiti | HAITI (41) | 149 | 149 | 1.04 (0.60, 1.81) | 0.01 |
| Kenya | WASH−B (42) | 1457 | 5137 | 0.86 (0.78, 0.94) | 0.15 |
| Madagascar | MAHAY (43) | 1702 | 1682 | 0.98 (0.90, 1.07) | 0.16 |
| Mali | PROMIS (46) | 506 | 506 | 0.92 (0.76, 1.10) | 0.08 |
| Mali | PROMIS CS (46) | 952 | 969 | 0.78 (0.65, 0.95) | 0.08 |
| Zimbabwe | SHINE (HIV−) (47) | 1880 | 1794 | 0.79 (0.72, 0.87) | 0.15 |
| Zimbabwe | SHINE (HIV+) (48) | 337 | 330 | 0.81 (0.68, 0.96) | 0.09 |
| **I² = 0.41, Tau² = 0.00** | | **9939** | **12735** | **0.87 (0.83, 0.92)** | |

**Frequency of contact − Weekly**

| Country | Trial | N | N | PR (95% CI) | W |
|---|---|---|---|---|---|
| Bangladesh | JiVitA−4 (34) | 2838 | 1244 | 0.92 (0.84, 1.01) | 0.25 |
| Bangladesh | WASH−B (36) | 1158 | 3431 | 0.84 (0.77, 0.91) | 0.27 |
| Burkina Faso | iLiNS−Zinc (37) | 1952 | 664 | 0.75 (0.65, 0.87) | 0.16 |
| Ghana | GHANA (39) | 98 | 96 | 0.70 (0.23, 2.13) | 0.01 |
| Ghana | iLiNS−DYADG (40) | 347 | 692 | 0.66 (0.45, 0.98) | 0.04 |
| Malawi | iLiNS−DYADM (44) | 220 | 444 | 1.10 (0.89, 1.36) | 0.10 |
| Malawi | iLiNS−DOSE (45) | 696 | 241 | 1.00 (0.87, 1.15) | 0.17 |
| **I² = 0.63, Tau² = 0.01** | | **7309** | **6812** | **0.88 (0.79, 0.99)** | |

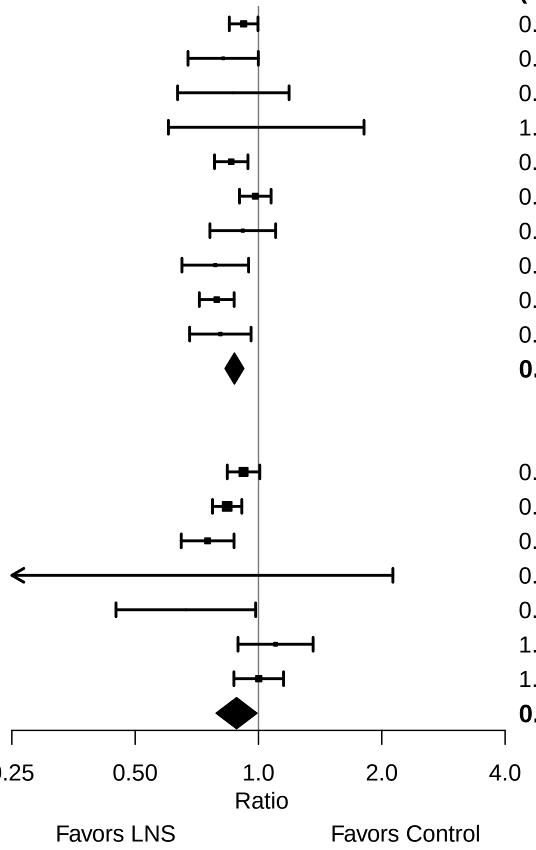

**Supplemental figure 4B: Stunting prevalence ratio**

#### 4B8: Stratified by Average SQ-LNS compliance

**Average SQ−LNS compliance**
**(p−diff = 0.974)**

**Average SQ−LNS compliance − Low**

| Country | Trial | N | N | PR (95% CI) | W |
|---|---|---|---|---|---|
| Burkina Faso | PROMIS (38) | 863 | 914 | 0.82 (0.67, 1.00) | 0.10 |
| Burkina Faso | PROMIS CS (38) | 430 | 439 | 0.87 (0.63, 1.19) | 0.05 |
| Ghana | iLiNS−DYADG (40) | 347 | 692 | 0.66 (0.45, 0.98) | 0.03 |
| Malawi | iLiNS−DYADM (44) | 220 | 444 | 1.10 (0.89, 1.36) | 0.09 |
| Malawi | iLiNS−DOSE (45) | 696 | 241 | 1.00 (0.87, 1.15) | 0.16 |
| Mali | PROMIS (46) | 506 | 506 | 0.92 (0.76, 1.10) | 0.11 |
| Mali | PROMIS CS (46) | 952 | 969 | 0.78 (0.65, 0.95) | 0.11 |
| Zimbabwe | SHINE (HIV−) (47) | 1880 | 1794 | 0.79 (0.72, 0.87) | 0.23 |
| Zimbabwe | SHINE (HIV+) (48) | 337 | 330 | 0.81 (0.68, 0.96) | 0.12 |
| **I² = 0.51, Tau² = 0.01** | | **6231** | **6329** | **0.86 (0.79, 0.94)** | |

**Average SQ−LNS compliance − High**

| Country | Trial | N | N | PR (95% CI) | W |
|---|---|---|---|---|---|
| Bangladesh | JiVitA−4 (34) | 2838 | 1244 | 0.92 (0.84, 1.01) | 0.21 |
| Bangladesh | RDNS (35) | 1663 | 815 | 0.92 (0.85, 1.00) | 0.22 |
| Bangladesh | WASH−B (36) | 1158 | 3431 | 0.84 (0.77, 0.91) | 0.22 |
| Burkina Faso | iLiNS−Zinc (37) | 1952 | 664 | 0.75 (0.65, 0.87) | 0.13 |
| Ghana | GHANA (39) | 98 | 96 | 0.70 (0.23, 2.13) | 0.00 |
| Haiti | HAITI (41) | 149 | 149 | 1.04 (0.60, 1.81) | 0.01 |
| Kenya | WASH−B (42) | 1457 | 5137 | 0.86 (0.78, 0.94) | 0.20 |
| **I² = 0.29, Tau² = 0.00** | | **9315** | **11536** | **0.87 (0.82, 0.92)** | |

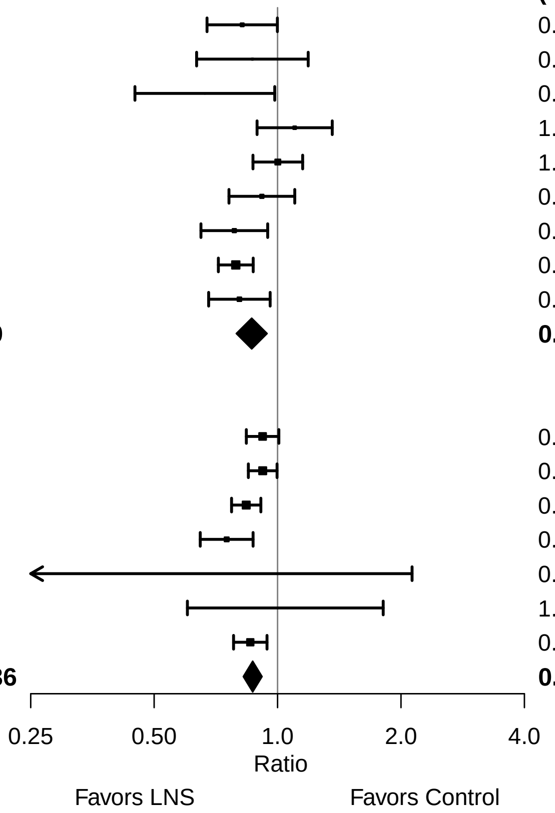

### Supplemental figure 4C: Stunting prevalence difference

#### 4C1: Stratified by Geographic region

**Geographic region**
**(p−diff = 0.874)**

| Country | Trial | N | N | PD (95% CI) | W |
|---|---|---|---|---|---|
| **Geographic region − SEAR** | | | | | |
| Bangladesh | JiVitA−4 (34) | 2838 | 1244 | −0.04 (−0.06, −0.01) | 0.38 |
| Bangladesh | RDNS (35) | 1663 | 815 | −0.03 (−0.07, 0.01) | 0.28 |
| Bangladesh | WASH−B (36) | 1158 | 3431 | −0.07 (−0.10, −0.04) | 0.34 |
| **I² = 0.26, Tau² = 0.00** | | **5659** | **5490** | **−0.05 (−0.07, −0.03)** | |
| **Geographic region − AFR** | | | | | |
| Burkina Faso | iLiNS−Zinc (37) | 1952 | 664 | −0.11 (−0.14, −0.08) | 0.10 |
| Burkina Faso | PROMIS (38) | 863 | 914 | −0.05 (−0.10, −0.01) | 0.08 |
| Burkina Faso | PROMIS CS (38) | 430 | 439 | −0.03 (−0.09, 0.02) | 0.06 |
| Ghana | GHANA (39) | 98 | 96 | −0.02 (−0.09, 0.05) | 0.05 |
| Ghana | iLiNS−DYADG (40) | 347 | 692 | −0.04 (−0.08, 0.00) | 0.09 |
| Kenya | WASH−B (42) | 1457 | 5137 | −0.05 (−0.07, −0.02) | 0.12 |
| Madagascar | MAHAY (43) | 1702 | 1682 | −0.01 (−0.04, 0.02) | 0.10 |
| Malawi | iLiNS−DYADM (44) | 220 | 444 | 0.03 (−0.04, 0.11) | 0.04 |
| Malawi | iLiNS−DOSE (45) | 696 | 241 | −0.02 (−0.07, 0.04) | 0.06 |
| Mali | PROMIS (46) | 506 | 506 | −0.04 (−0.09, 0.01) | 0.07 |
| Mali | PROMIS CS (46) | 952 | 969 | −0.07 (−0.11, −0.03) | 0.08 |
| Zimbabwe | SHINE (HIV−) (47) | 1880 | 1794 | −0.07 (−0.10, −0.04) | 0.11 |
| Zimbabwe | SHINE (HIV+) (48) | 337 | 330 | −0.10 (−0.17, −0.02) | 0.04 |
| **I² = 0.60, Tau² = 0.00** | | **11440** | **13908** | **−0.05 (−0.07, −0.03)** | |

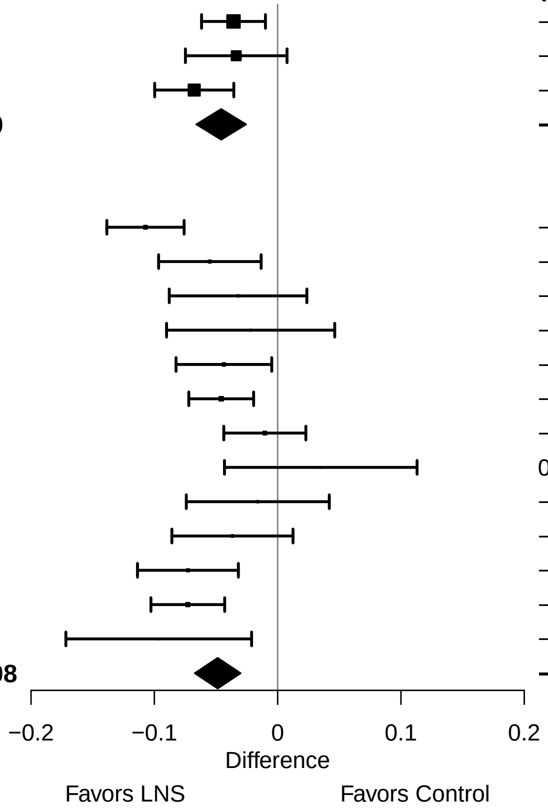

**Supplemental figure 4C: Stunting prevalence difference**

#### 4C2: Stratified by Stunting burden

**Stunting burden**
**(p−diff = 0.181)**

| Country | Trial | N | N | PD (95% CI) | W |
|---|---|---|---|---|---|
| **Stunting burden − Less than 35%** | | | | | |
| Burkina Faso | PROMIS (38) | 863 | 914 | −0.05 (−0.10, −0.01) | 0.17 |
| Burkina Faso | PROMIS CS (38) | 430 | 439 | −0.03 (−0.09, 0.02) | 0.12 |
| Ghana | GHANA (39) | 98 | 96 | −0.02 (−0.09, 0.05) | 0.09 |
| Ghana | iLiNS−DYADG (40) | 347 | 692 | −0.04 (−0.08, 0.00) | 0.18 |
| Haiti | HAITI (41) | 149 | 149 | −0.02 (−0.08, 0.04) | 0.11 |
| Kenya | WASH−B (42) | 1457 | 5137 | −0.05 (−0.07, −0.02) | 0.24 |
| Malawi | iLiNS−DYADM (44) | 220 | 444 | 0.03 (−0.04, 0.11) | 0.08 |
| **I² = 0.00, Tau² = 0.00** | | **3564** | **7871** | **−0.04 (−0.06, −0.02)** | |
| **Stunting burden − More than 35%** | | | | | |
| Bangladesh | JiVitA−4 (34) | 2838 | 1244 | −0.04 (−0.06, −0.01) | 0.14 |
| Bangladesh | RDNS (35) | 1663 | 815 | −0.03 (−0.07, 0.01) | 0.10 |
| Bangladesh | WASH−B (36) | 1158 | 3431 | −0.07 (−0.10, −0.04) | 0.12 |
| Burkina Faso | iLiNS−Zinc (37) | 1952 | 664 | −0.11 (−0.14, −0.08) | 0.12 |
| Madagascar | MAHAY (43) | 1702 | 1682 | −0.01 (−0.04, 0.02) | 0.12 |
| Malawi | iLiNS−DOSE (45) | 696 | 241 | −0.02 (−0.07, 0.04) | 0.07 |
| Mali | PROMIS (46) | 506 | 506 | −0.04 (−0.09, 0.01) | 0.08 |
| Mali | PROMIS CS (46) | 952 | 969 | −0.07 (−0.11, −0.03) | 0.10 |
| Zimbabwe | SHINE (HIV−) (47) | 1880 | 1794 | −0.07 (−0.10, −0.04) | 0.12 |
| Zimbabwe | SHINE (HIV+) (48) | 337 | 330 | −0.10 (−0.17, −0.02) | 0.05 |
| **I² = 0.66, Tau² = 0.00** | | **13684** | **11676** | **−0.05 (−0.07, −0.03)** | |

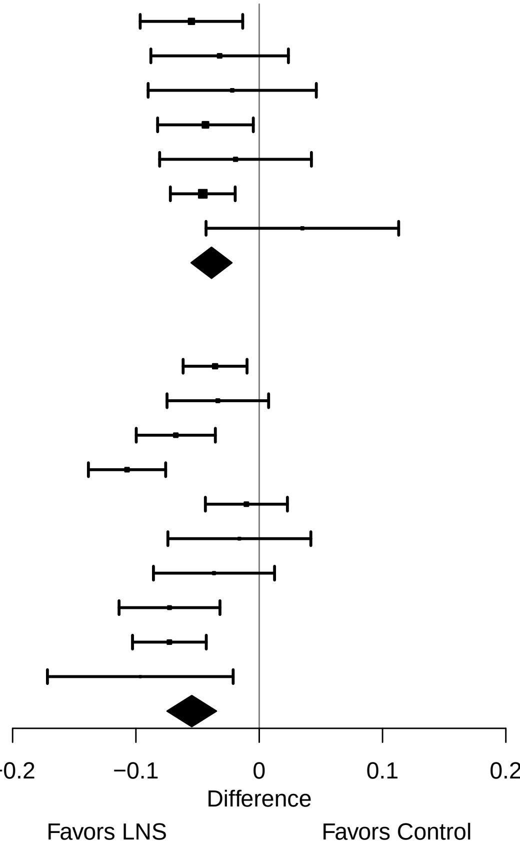

#### Supplemental figure 4C: Stunting prevalence difference

### 4C3: Stratified by Malaria prevalence

**Malaria prevalence**
**(p−diff = 0.848)**

| Country | Trial | N | N | PD (95% CI) | W |
|---|---|---|---|---|---|
| **Malaria prevalence − Less than 10%** | | | | | |
| Bangladesh | JiVitA−4 (34) | 2838 | 1244 | −0.04 (−0.06, −0.01) | 0.16 |
| Bangladesh | RDNS (35) | 1663 | 815 | −0.03 (−0.07, 0.01) | 0.12 |
| Bangladesh | WASH−B (36) | 1158 | 3431 | −0.07 (−0.10, −0.04) | 0.14 |
| Haiti | HAITI (41) | 149 | 149 | −0.02 (−0.08, 0.04) | 0.07 |
| Kenya | WASH−B (42) | 1457 | 5137 | −0.05 (−0.07, −0.02) | 0.16 |
| Madagascar | MAHAY (43) | 1702 | 1682 | −0.01 (−0.04, 0.02) | 0.14 |
| Zimbabwe | SHINE (HIV−) (47) | 1880 | 1794 | −0.07 (−0.10, −0.04) | 0.15 |
| Zimbabwe | SHINE (HIV+) (48) | 337 | 330 | −0.10 (−0.17, −0.02) | 0.05 |
| **I² = 0.45, Tau² = 0.00** | | **11184** | **14582** | **−0.05 (−0.06, −0.03)** | |
| **Malaria prevalence − At least 10%** | | | | | |
| Burkina Faso | iLiNS−Zinc (37) | 1952 | 664 | −0.11 (−0.14, −0.08) | 0.17 |
| Burkina Faso | PROMIS (38) | 863 | 914 | −0.05 (−0.10, −0.01) | 0.13 |
| Burkina Faso | PROMIS CS (38) | 430 | 439 | −0.03 (−0.09, 0.02) | 0.10 |
| Ghana | GHANA (39) | 98 | 96 | −0.02 (−0.09, 0.05) | 0.07 |
| Ghana | iLiNS−DYADG (40) | 347 | 692 | −0.04 (−0.08, 0.00) | 0.14 |
| Malawi | iLiNS−DYADM (44) | 220 | 444 | 0.03 (−0.04, 0.11) | 0.06 |
| Malawi | iLiNS−DOSE (45) | 696 | 241 | −0.02 (−0.07, 0.04) | 0.09 |
| Mali | PROMIS (46) | 506 | 506 | −0.04 (−0.09, 0.01) | 0.11 |
| Mali | PROMIS CS (46) | 952 | 969 | −0.07 (−0.11, −0.03) | 0.13 |
| **I² = 0.61, Tau² = 0.00** | | **6064** | **4965** | **−0.05 (−0.07, −0.02)** | |

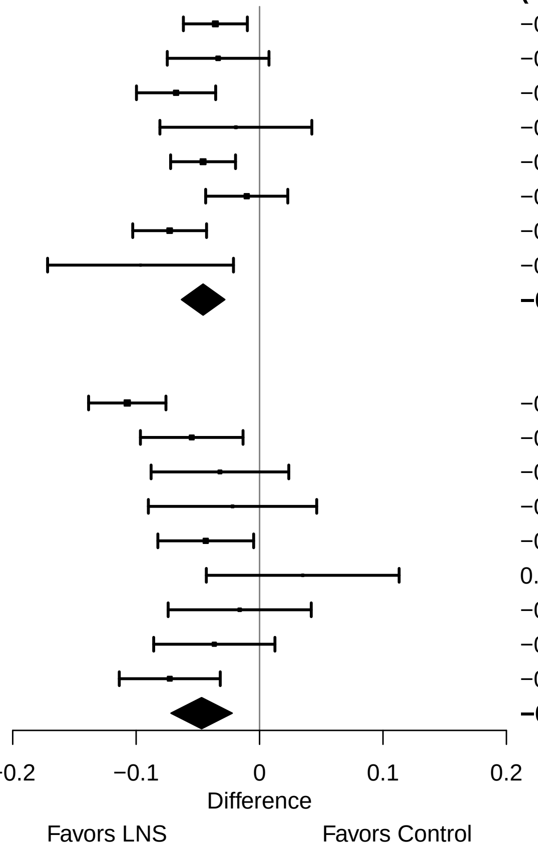

#### Supplemental figure 4C: Stunting prevalence difference

### 4C4: Stratified by Source water quality

**Source water quality**
**(p−diff = 0.234)**

**Source water quality − Improved**

| Country | Trial | N | N | PD (95% CI) | W |
|---|---|---|---|---|---|
| Bangladesh | JiVitA−4 (34) | 2838 | 1244 | −0.04 (−0.06, −0.01) | 0.22 |
| Bangladesh | RDNS (35) | 1663 | 815 | −0.03 (−0.07, 0.01) | 0.15 |
| Bangladesh | WASH−B (36) | 567 | 1104 | −0.08 (−0.13, −0.03) | 0.13 |
| Ghana | GHANA (39) | 98 | 96 | −0.02 (−0.09, 0.05) | 0.08 |
| Ghana | iLiNS−DYADG (40) | 347 | 692 | −0.04 (−0.08, 0.00) | 0.16 |
| Haiti | HAITI (41) | 149 | 149 | −0.02 (−0.08, 0.04) | 0.09 |
| Malawi | iLiNS−DYADM (44) | 220 | 444 | 0.03 (−0.04, 0.11) | 0.07 |
| Malawi | iLiNS−DOSE (45) | 696 | 241 | −0.02 (−0.07, 0.04) | 0.10 |
| **I² = 0.08, Tau² = 0.00** | | **6578** | **4785** | **−0.03 (−0.05, −0.02)** | |

**Source water quality − Unimproved**

| Country | Trial | N | N | PD (95% CI) | W |
|---|---|---|---|---|---|
| Burkina Faso | iLiNS−Zinc (37) | 1952 | 664 | −0.11 (−0.14, −0.08) | 0.15 |
| Burkina Faso | PROMIS (38) | 863 | 914 | −0.05 (−0.10, −0.01) | 0.12 |
| Burkina Faso | PROMIS CS (38) | 430 | 439 | −0.03 (−0.09, 0.02) | 0.09 |
| Kenya | WASH−B (42) | 695 | 2253 | −0.02 (−0.06, 0.01) | 0.13 |
| Madagascar | MAHAY (43) | 1702 | 1682 | −0.01 (−0.04, 0.02) | 0.15 |
| Mali | PROMIS (46) | 506 | 506 | −0.04 (−0.09, 0.01) | 0.10 |
| Mali | PROMIS CS (46) | 952 | 969 | −0.07 (−0.11, −0.03) | 0.12 |
| Zimbabwe | SHINE (HIV−) (47) | 892 | 880 | −0.05 (−0.09, −0.01) | 0.12 |
| Zimbabwe | SHINE (HIV+) (48) | 147 | 146 | −0.11 (−0.22, 0.01) | 0.03 |
| **I² = 0.65, Tau² = 0.00** | | **8139** | **8453** | **−0.05 (−0.07, −0.03)** | |

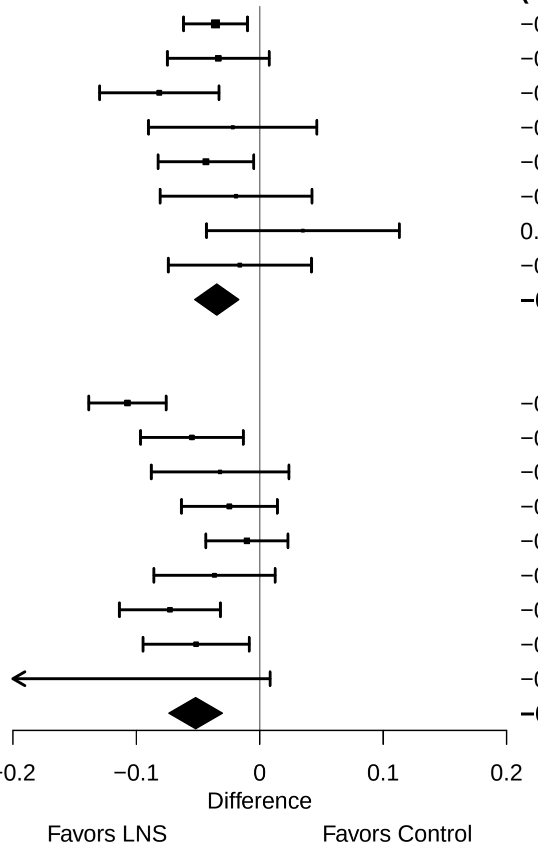

##### Supplemental figure 4C: Stunting prevalence difference

#### 4C5: Stratified by Sanitation

**Sanitation**
**(p−diff = 0.839)**

| Country | Trial | N | N | PD (95% CI) | W |
|---|---|---|---|---|---|
| **Sanitation − Improved** | | | | | |
| Bangladesh | JiVitA−4 (34) | 2838 | 1244 | −0.04 (−0.06, −0.01) | 0.16 |
| Bangladesh | RDNS (35) | 1663 | 815 | −0.03 (−0.07, 0.01) | 0.11 |
| Bangladesh | WASH−B (36) | 567 | 1104 | −0.08 (−0.13, −0.03) | 0.09 |
| Burkina Faso | PROMIS (38) | 863 | 914 | −0.05 (−0.10, −0.01) | 0.11 |
| Burkina Faso | PROMIS CS (38) | 430 | 439 | −0.03 (−0.09, 0.02) | 0.08 |
| Ghana | GHANA (39) | 98 | 96 | −0.02 (−0.09, 0.05) | 0.06 |
| Ghana | iLiNS−DYADG (40) | 347 | 692 | −0.04 (−0.08, 0.00) | 0.12 |
| Haiti | HAITI (41) | 149 | 149 | −0.02 (−0.08, 0.04) | 0.07 |
| Mali | PROMIS (46) | 506 | 506 | −0.04 (−0.09, 0.01) | 0.09 |
| Mali | PROMIS CS (46) | 952 | 969 | −0.07 (−0.11, −0.03) | 0.11 |
| **I² = 0.00, Tau² = 0.00** | | **8413** | **6928** | **−0.04 (−0.06, −0.03)** | |
| **Sanitation − Unimproved** | | | | | |
| Burkina Faso | iLiNS−Zinc (37) | 1952 | 664 | −0.11 (−0.14, −0.08) | 0.22 |
| Kenya | WASH−B (42) | 695 | 2253 | −0.02 (−0.06, 0.01) | 0.18 |
| Madagascar | MAHAY (43) | 1702 | 1682 | −0.01 (−0.04, 0.02) | 0.21 |
| Malawi | iLiNS−DYADM (44) | 220 | 444 | 0.03 (−0.04, 0.11) | 0.08 |
| Malawi | iLiNS−DOSE (45) | 696 | 241 | −0.02 (−0.07, 0.04) | 0.12 |
| Zimbabwe | SHINE (HIV−) (47) | 892 | 880 | −0.05 (−0.09, −0.01) | 0.16 |
| Zimbabwe | SHINE (HIV+) (48) | 147 | 146 | −0.11 (−0.22, 0.01) | 0.04 |
| **I² = 0.77, Tau² = 0.00** | | **6304** | **6310** | **−0.04 (−0.08, 0.00)** | |

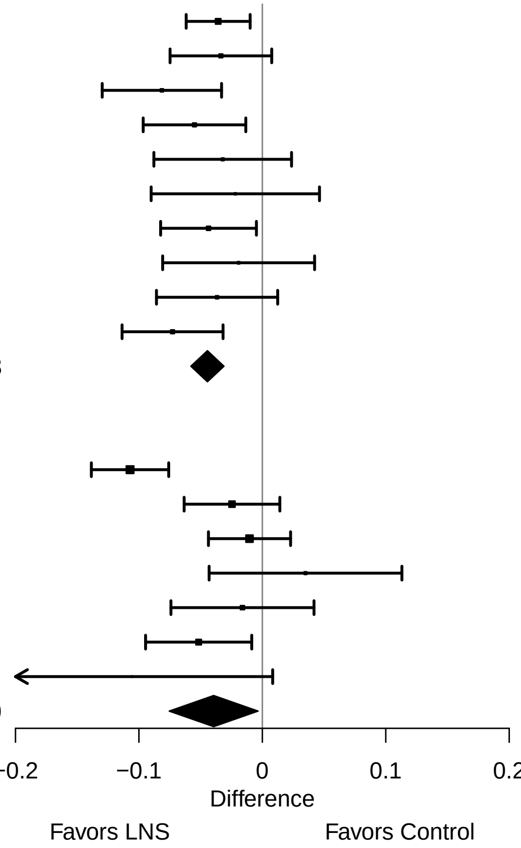

#### Supplemental figure 4C: Stunting prevalence difference

### 4C6: Stratified by Supplement duration

**Supplement duration**
**(p−diff = 0.654)**

| Country | Trial | N | N | PD (95% CI) | W |
|---|---|---|---|---|---|
| **Supplement duration − 12m or less** | | | | | |
| Bangladesh | JiVitA−4 (34) | 2838 | 1244 | −0.04 (−0.06, −0.01) | 0.13 |
| Burkina Faso | iLiNS−Zinc (37) | 1952 | 664 | −0.11 (−0.14, −0.08) | 0.12 |
| Burkina Faso | PROMIS (38) | 863 | 914 | −0.05 (−0.10, −0.01) | 0.09 |
| Burkina Faso | PROMIS CS (38) | 430 | 439 | −0.03 (−0.09, 0.02) | 0.07 |
| Ghana | GHANA (39) | 98 | 96 | −0.02 (−0.09, 0.05) | 0.05 |
| Ghana | iLiNS−DYADG (40) | 347 | 692 | −0.04 (−0.08, 0.00) | 0.10 |
| Haiti | HAITI (41) | 149 | 149 | −0.02 (−0.08, 0.04) | 0.06 |
| Madagascar | MAHAY (43) | 1702 | 1682 | −0.01 (−0.04, 0.02) | 0.11 |
| Malawi | iLiNS−DYADM (44) | 220 | 444 | 0.03 (−0.04, 0.11) | 0.04 |
| Malawi | iLiNS−DOSE (45) | 696 | 241 | −0.02 (−0.07, 0.04) | 0.06 |
| Zimbabwe | SHINE (HIV−) (47) | 1880 | 1794 | −0.07 (−0.10, −0.04) | 0.12 |
| Zimbabwe | SHINE (HIV+) (48) | 337 | 330 | −0.10 (−0.17, −0.02) | 0.04 |
| **I² = 0.65, Tau² = 0.00** | | **11512** | **8689** | **−0.04 (−0.06, −0.02)** | |
| **Supplement duration − > 12m** | | | | | |
| Bangladesh | RDNS (35) | 1663 | 815 | −0.03 (−0.07, 0.01) | 0.18 |
| Bangladesh | WASH−B (36) | 1158 | 3431 | −0.07 (−0.10, −0.04) | 0.23 |
| Kenya | WASH−B (42) | 1457 | 5137 | −0.05 (−0.07, −0.02) | 0.25 |
| Mali | PROMIS (46) | 506 | 506 | −0.04 (−0.09, 0.01) | 0.15 |
| Mali | PROMIS CS (46) | 952 | 969 | −0.07 (−0.11, −0.03) | 0.18 |
| **I² = 0.00, Tau² = 0.00** | | **5736** | **10858** | **−0.05 (−0.07, −0.04)** | |

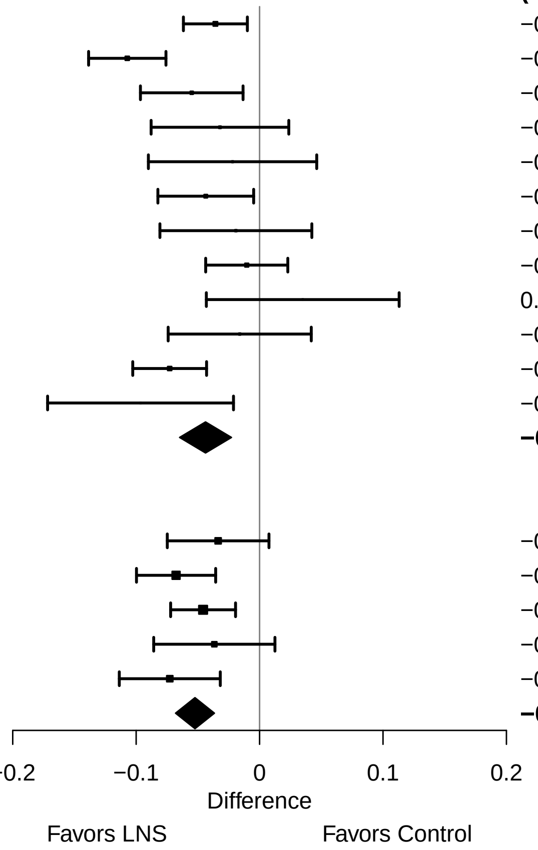

#### Supplemental figure 4C: Stunting prevalence difference

### 4C7: Stratified by Frequency of contact

**Frequency of contact**
**(p−diff = 0.910)**

**Frequency of contact − Monthly**

| Country | Trial | N | N | PD (95% CI) | W |
|---|---|---|---|---|---|
| Bangladesh | RDNS (35) | 1663 | 815 | −0.03 (−0.07, 0.01) | 0.10 |
| Burkina Faso | PROMIS (38) | 863 | 914 | −0.05 (−0.10, −0.01) | 0.10 |
| Burkina Faso | PROMIS CS (38) | 430 | 439 | −0.03 (−0.09, 0.02) | 0.08 |
| Haiti | HAITI (41) | 149 | 149 | −0.02 (−0.08, 0.04) | 0.07 |
| Kenya | WASH−B (42) | 1457 | 5137 | −0.05 (−0.07, −0.02) | 0.15 |
| Madagascar | MAHAY (43) | 1702 | 1682 | −0.01 (−0.04, 0.02) | 0.13 |
| Mali | PROMIS (46) | 506 | 506 | −0.04 (−0.09, 0.01) | 0.09 |
| Mali | PROMIS CS (46) | 952 | 969 | −0.07 (−0.11, −0.03) | 0.11 |
| Zimbabwe | SHINE (HIV−) (47) | 1880 | 1794 | −0.07 (−0.10, −0.04) | 0.14 |
| Zimbabwe | SHINE (HIV+) (48) | 337 | 330 | −0.10 (−0.17, −0.02) | 0.05 |
| **$I^2$ = 0.28, $Tau^2$ = 0.00** | | **9939** | **12735** | **−0.05 (−0.06, −0.03)** | |

**Frequency of contact − Weekly**

| Country | Trial | N | N | PD (95% CI) | W |
|---|---|---|---|---|---|
| Bangladesh | JiVitA−4 (34) | 2838 | 1244 | −0.04 (−0.06, −0.01) | 0.21 |
| Bangladesh | WASH−B (36) | 1158 | 3431 | −0.07 (−0.10, −0.04) | 0.19 |
| Burkina Faso | iLiNS−Zinc (37) | 1952 | 664 | −0.11 (−0.14, −0.08) | 0.19 |
| Ghana | GHANA (39) | 98 | 96 | −0.02 (−0.09, 0.05) | 0.08 |
| Ghana | iLiNS−DYADG (40) | 347 | 692 | −0.04 (−0.08, 0.00) | 0.16 |
| Malawi | iLiNS−DYADM (44) | 220 | 444 | 0.03 (−0.04, 0.11) | 0.07 |
| Malawi | iLiNS−DOSE (45) | 696 | 241 | −0.02 (−0.07, 0.04) | 0.10 |
| **$I^2$ = 0.72, $Tau^2$ = 0.00** | | **7309** | **6812** | **−0.04 (−0.08, −0.01)** | |

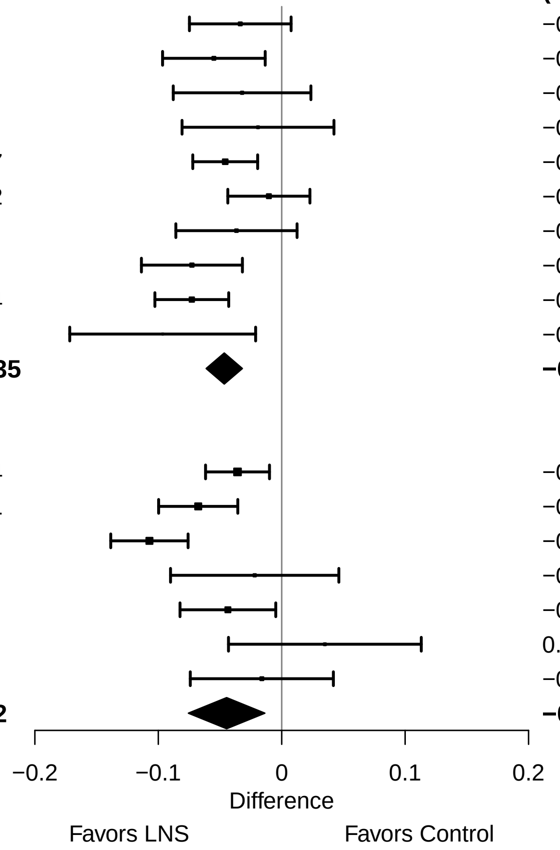

#### Supplemental figure 4C: Stunting prevalence difference

### 4C8: Stratified by Average SQ-LNS compliance

**Average SQ−LNS compliance**
**(p−diff = 0.803)**

| Country | Trial | N | N | PD (95% CI) | W |
|---|---|---|---|---|---|
| **Average SQ−LNS compliance − Low** | | | | | |
| Burkina Faso | PROMIS (38) | 863 | 914 | −0.05 (−0.10, −0.01) | 0.13 |
| Burkina Faso | PROMIS CS (38) | 430 | 439 | −0.03 (−0.09, 0.02) | 0.09 |
| Ghana | iLiNS−DYADG (40) | 347 | 692 | −0.04 (−0.08, 0.00) | 0.14 |
| Malawi | iLiNS−DYADM (44) | 220 | 444 | 0.03 (−0.04, 0.11) | 0.06 |
| Malawi | iLiNS−DOSE (45) | 696 | 241 | −0.02 (−0.07, 0.04) | 0.09 |
| Mali | PROMIS (46) | 506 | 506 | −0.04 (−0.09, 0.01) | 0.11 |
| Mali | PROMIS CS (46) | 952 | 969 | −0.07 (−0.11, −0.03) | 0.14 |
| Zimbabwe | SHINE (HIV−) (47) | 1880 | 1794 | −0.07 (−0.10, −0.04) | 0.18 |
| Zimbabwe | SHINE (HIV+) (48) | 337 | 330 | −0.10 (−0.17, −0.02) | 0.06 |
| **$I^2$ = 0.30, $Tau^2$ = 0.00** | | **6231** | **6329** | **−0.05 (−0.07, −0.03)** | |
| **Average SQ−LNS compliance − High** | | | | | |
| Bangladesh | JiVitA−4 (34) | 2838 | 1244 | −0.04 (−0.06, −0.01) | 0.19 |
| Bangladesh | RDNS (35) | 1663 | 815 | −0.03 (−0.07, 0.01) | 0.13 |
| Bangladesh | WASH−B (36) | 1158 | 3431 | −0.07 (−0.10, −0.04) | 0.17 |
| Burkina Faso | iLiNS−Zinc (37) | 1952 | 664 | −0.11 (−0.14, −0.08) | 0.17 |
| Ghana | GHANA (39) | 98 | 96 | −0.02 (−0.09, 0.05) | 0.07 |
| Haiti | HAITI (41) | 149 | 149 | −0.02 (−0.08, 0.04) | 0.08 |
| Kenya | WASH−B (42) | 1457 | 5137 | −0.05 (−0.07, −0.02) | 0.19 |
| **$I^2$ = 0.65, $Tau^2$ = 0.00** | | **9315** | **11536** | **−0.05 (−0.08, −0.03)** | |

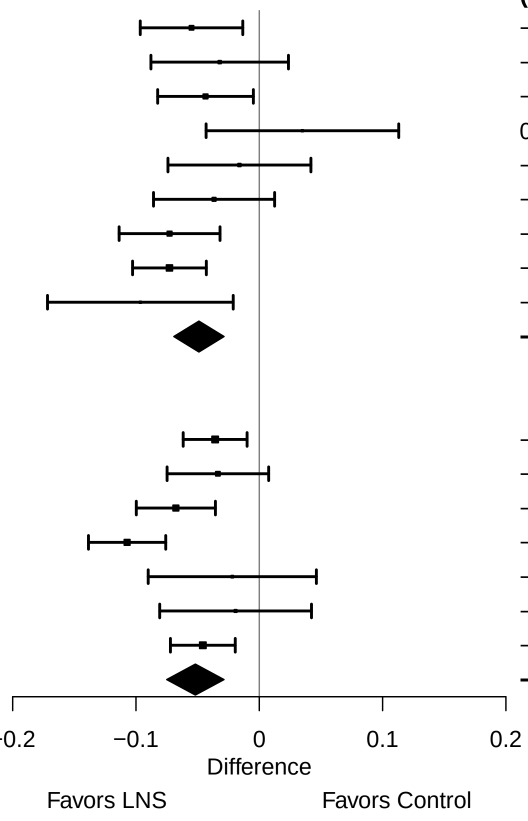

### Supplemental figure 4D: Mean difference in WLZ

#### 4D1: Stratified by Geographic region

**Geographic region**
**(p−diff = 0.482)**

| Country | Trial | N | N | MD (95% CI) | W |
|---|---|---|---|---|---|
| **Geographic region − SEAR** | | | | | |
| Bangladesh | JiVitA−4 (34) | 2783 | 1217 | 0.08 (0.03, 0.13) | 0.37 |
| Bangladesh | RDNS (35) | 1661 | 815 | 0.07 (0.00, 0.15) | 0.30 |
| Bangladesh | WASH−B (36) | 1156 | 3424 | 0.12 (0.05, 0.19) | 0.32 |
| **I² = 0.00, Tau² = 0.00** | | **5600** | **5456** | **0.09 (0.06, 0.12)** | |
| **Geographic region − AFR** | | | | | |
| Burkina Faso | iLiNS−Zinc (37) | 1952 | 664 | 0.18 (0.11, 0.26) | 0.11 |
| Burkina Faso | PROMIS (38) | 856 | 907 | 0.10 (−0.03, 0.22) | 0.07 |
| Burkina Faso | PROMIS CS (38) | 430 | 436 | −0.11 (−0.24, 0.02) | 0.07 |
| Ghana | GHANA (39) | 98 | 96 | 0.21 (−0.13, 0.54) | 0.02 |
| Ghana | iLiNS−DYADG (40) | 347 | 692 | 0.09 (−0.04, 0.22) | 0.07 |
| Kenya | WASH−B (42) | 1455 | 5118 | 0.06 (0.00, 0.13) | 0.12 |
| Madagascar | MAHAY (43) | 1700 | 1682 | 0.00 (−0.11, 0.11) | 0.08 |
| Malawi | iLiNS−DYADM (44) | 220 | 444 | −0.05 (−0.21, 0.11) | 0.06 |
| Malawi | iLiNS−DOSE (45) | 696 | 241 | 0.01 (−0.12, 0.14) | 0.07 |
| Mali | PROMIS (46) | 499 | 502 | 0.10 (−0.02, 0.23) | 0.08 |
| Mali | PROMIS CS (46) | 944 | 959 | 0.16 (0.06, 0.26) | 0.09 |
| Zimbabwe | SHINE (HIV−) (47) | 1869 | 1784 | 0.08 (0.00, 0.15) | 0.11 |
| Zimbabwe | SHINE (HIV+) (48) | 336 | 328 | −0.14 (−0.32, 0.04) | 0.05 |
| **I² = 0.61, Tau² = 0.01** | | **11402** | **13853** | **0.06 (0.00, 0.11)** | |

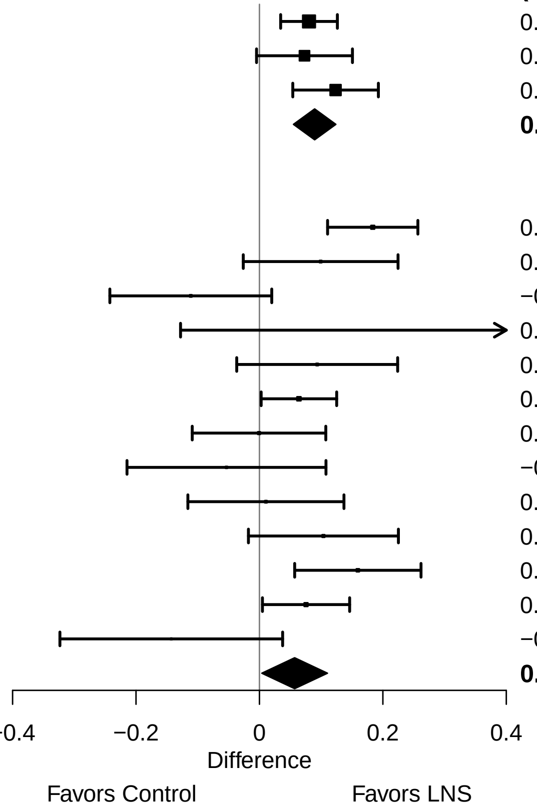

#### Supplemental figure 4D: Mean difference in WLZ

### 4D2: Stratified by Stunting burden

**Stunting burden**
**(p−diff = 0.264)**

| Country | Trial | N | N | MD (95% CI) | W |
|---|---|---|---|---|---|
| **Stunting burden − Less than 35%** | | | | | |
| Burkina Faso | PROMIS (38) | 856 | 907 | 0.10 (−0.03, 0.22) | 0.16 |
| Burkina Faso | PROMIS CS (38) | 430 | 436 | −0.11 (−0.24, 0.02) | 0.15 |
| Ghana | GHANA (39) | 98 | 96 | 0.21 (−0.13, 0.54) | 0.04 |
| Ghana | iLiNS−DYADG (40) | 347 | 692 | 0.09 (−0.04, 0.22) | 0.15 |
| Haiti | HAITI (41) | 148 | 149 | 0.09 (−0.07, 0.24) | 0.12 |
| Kenya | WASH−B (42) | 1455 | 5118 | 0.06 (0.00, 0.13) | 0.27 |
| Malawi | iLiNS−DYADM (44) | 220 | 444 | −0.05 (−0.21, 0.11) | 0.12 |
| **I² = 0.38, Tau² = 0.00** | | **3554** | **7842** | **0.04 (−0.02, 0.11)** | |
| **Stunting burden − More than 35%** | | | | | |
| Bangladesh | JiVitA−4 (34) | 2783 | 1217 | 0.08 (0.03, 0.13) | 0.14 |
| Bangladesh | RDNS (35) | 1661 | 815 | 0.07 (0.00, 0.15) | 0.11 |
| Bangladesh | WASH−B (36) | 1156 | 3424 | 0.12 (0.05, 0.19) | 0.12 |
| Burkina Faso | iLiNS−Zinc (37) | 1952 | 664 | 0.18 (0.11, 0.26) | 0.12 |
| Madagascar | MAHAY (43) | 1700 | 1682 | 0.00 (−0.11, 0.11) | 0.09 |
| Malawi | iLiNS−DOSE (45) | 696 | 241 | 0.01 (−0.12, 0.14) | 0.08 |
| Mali | PROMIS (46) | 499 | 502 | 0.10 (−0.02, 0.23) | 0.08 |
| Mali | PROMIS CS (46) | 944 | 959 | 0.16 (0.06, 0.26) | 0.09 |
| Zimbabwe | SHINE (HIV−) (47) | 1869 | 1784 | 0.08 (0.00, 0.15) | 0.12 |
| Zimbabwe | SHINE (HIV+) (48) | 336 | 328 | −0.14 (−0.32, 0.04) | 0.05 |
| **I² = 0.55, Tau² = 0.00** | | **13596** | **11616** | **0.08 (0.03, 0.13)** | |

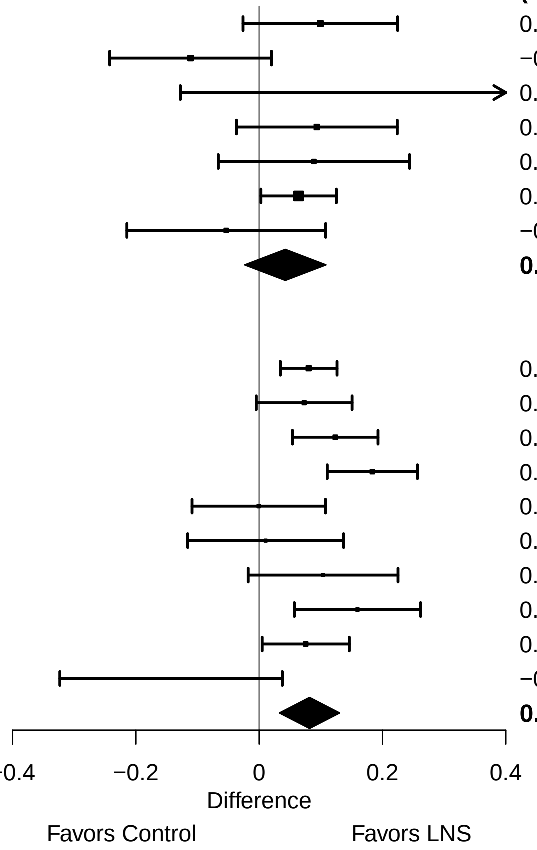

#### Supplemental figure 4D: Mean difference in WLZ

### 4D3: Stratified by Malaria prevalence

**Malaria prevalence**
**(p−diff = 0.629)**

| Country | Trial | N | N | MD (95% CI) | W |
|---|---|---|---|---|---|
| **Malaria prevalence − Less than 10%** | | | | | |
| Bangladesh | JiVitA−4 (34) | 2783 | 1217 | 0.08 (0.03, 0.13) | 0.17 |
| Bangladesh | RDNS (35) | 1661 | 815 | 0.07 (0.00, 0.15) | 0.14 |
| Bangladesh | WASH−B (36) | 1156 | 3424 | 0.12 (0.05, 0.19) | 0.15 |
| Haiti | HAITI (41) | 148 | 149 | 0.09 (−0.07, 0.24) | 0.07 |
| Kenya | WASH−B (42) | 1455 | 5118 | 0.06 (0.00, 0.13) | 0.16 |
| Madagascar | MAHAY (43) | 1700 | 1682 | 0.00 (−0.11, 0.11) | 0.11 |
| Zimbabwe | SHINE (HIV−) (47) | 1869 | 1784 | 0.08 (0.00, 0.15) | 0.15 |
| Zimbabwe | SHINE (HIV+) (48) | 336 | 328 | −0.14 (−0.32, 0.04) | 0.06 |
| **I² = 0.27, Tau² = 0.00** | | **11108** | **14517** | **0.07 (0.03, 0.11)** | |
| **Malaria prevalence − At least 10%** | | | | | |
| Burkina Faso | iLiNS−Zinc (37) | 1952 | 664 | 0.18 (0.11, 0.26) | 0.18 |
| Burkina Faso | PROMIS (38) | 856 | 907 | 0.10 (−0.03, 0.22) | 0.12 |
| Burkina Faso | PROMIS CS (38) | 430 | 436 | −0.11 (−0.24, 0.02) | 0.11 |
| Ghana | GHANA (39) | 98 | 96 | 0.21 (−0.13, 0.54) | 0.03 |
| Ghana | iLiNS−DYADG (40) | 347 | 692 | 0.09 (−0.04, 0.22) | 0.11 |
| Malawi | iLiNS−DYADM (44) | 220 | 444 | −0.05 (−0.21, 0.11) | 0.09 |
| Malawi | iLiNS−DOSE (45) | 696 | 241 | 0.01 (−0.12, 0.14) | 0.11 |
| Mali | PROMIS (46) | 499 | 502 | 0.10 (−0.02, 0.23) | 0.12 |
| Mali | PROMIS CS (46) | 944 | 959 | 0.16 (0.06, 0.26) | 0.14 |
| **I² = 0.64, Tau² = 0.01** | | **6042** | **4941** | **0.08 (0.01, 0.14)** | |

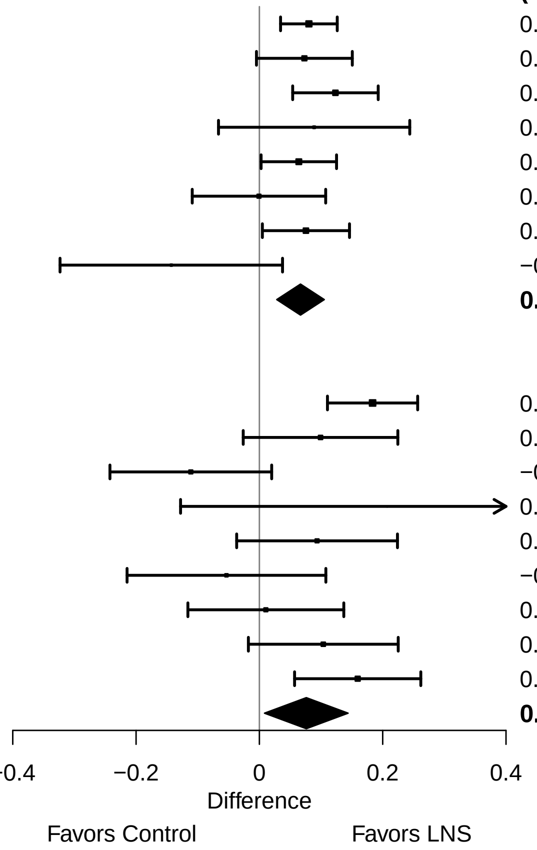

#### Supplemental figure 4D: Mean difference in WLZ

### 4D4: Stratified by Source water quality

**Source water quality**
**(p−diff = 0.707)**

| Country | Trial | N | N | MD (95% CI) | W |
|---|---|---|---|---|---|
| **Source water quality − Improved** | | | | | |
| Bangladesh | JiVitA−4 (34) | 2783 | 1217 | 0.08 (0.03, 0.13) | 0.23 |
| Bangladesh | RDNS (35) | 1661 | 815 | 0.07 (0.00, 0.15) | 0.18 |
| Bangladesh | WASH−B (36) | 566 | 1101 | 0.16 (0.05, 0.27) | 0.14 |
| Ghana | GHANA (39) | 98 | 96 | 0.21 (−0.13, 0.54) | 0.03 |
| Ghana | iLiNS−DYADG (40) | 347 | 692 | 0.09 (−0.04, 0.22) | 0.12 |
| Haiti | HAITI (41) | 148 | 149 | 0.09 (−0.07, 0.24) | 0.09 |
| Malawi | iLiNS−DYADM (44) | 220 | 444 | −0.05 (−0.21, 0.11) | 0.09 |
| Malawi | iLiNS−DOSE (45) | 696 | 241 | 0.01 (−0.12, 0.14) | 0.12 |
| **I² = 0.00, Tau² = 0.00** | | **6519** | **4755** | **0.08 (0.05, 0.11)** | |
| **Source water quality − Unimproved** | | | | | |
| Burkina Faso | iLiNS−Zinc (37) | 1952 | 664 | 0.18 (0.11, 0.26) | 0.16 |
| Burkina Faso | PROMIS (38) | 856 | 907 | 0.10 (−0.03, 0.22) | 0.10 |
| Burkina Faso | PROMIS CS (38) | 430 | 436 | −0.11 (−0.24, 0.02) | 0.10 |
| Kenya | WASH−B (42) | 694 | 2244 | 0.03 (−0.04, 0.11) | 0.15 |
| Madagascar | MAHAY (43) | 1700 | 1682 | 0.00 (−0.11, 0.11) | 0.12 |
| Mali | PROMIS (46) | 499 | 502 | 0.10 (−0.02, 0.23) | 0.10 |
| Mali | PROMIS CS (46) | 944 | 959 | 0.16 (0.06, 0.26) | 0.12 |
| Zimbabwe | SHINE (HIV−) (47) | 887 | 876 | 0.02 (−0.09, 0.13) | 0.12 |
| Zimbabwe | SHINE (HIV+) (48) | 147 | 146 | −0.15 (−0.40, 0.10) | 0.04 |
| **I² = 0.69, Tau² = 0.01** | | **8109** | **8416** | **0.05 (−0.01, 0.12)** | |

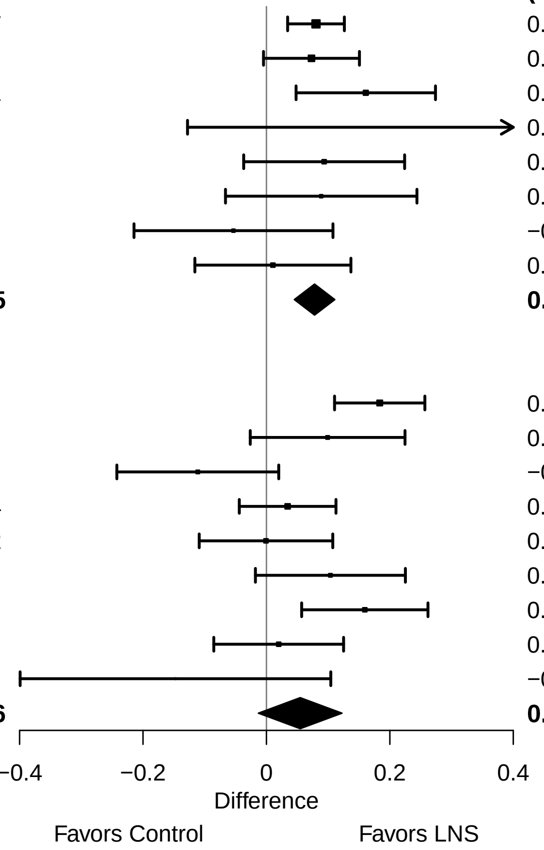

#### Supplemental figure 4D: Mean difference in WLZ

### 4D5: Stratified by Sanitation

**Sanitation**
**(p−diff = 0.211)**

| **Sanitation − Improved**<br>**Country** | **Trial** | **N** | **N** | **MD (95% CI)** | **W** |
|---|---|---|---|---|---|
| Bangladesh | JiVitA−4 (34) | 2783 | 1217 | 0.08 (0.03, 0.13) | 0.18 |
| Bangladesh | RDNS (35) | 1661 | 815 | 0.07 (0.00, 0.15) | 0.14 |
| Bangladesh | WASH−B (36) | 566 | 1101 | 0.16 (0.05, 0.27) | 0.10 |
| Burkina Faso | PROMIS (38) | 856 | 907 | 0.10 (−0.03, 0.22) | 0.09 |
| Burkina Faso | PROMIS CS (38) | 430 | 436 | −0.11 (−0.24, 0.02) | 0.09 |
| Ghana | GHANA (39) | 98 | 96 | 0.21 (−0.13, 0.54) | 0.02 |
| Ghana | iLiNS−DYADG (40) | 347 | 692 | 0.09 (−0.04, 0.22) | 0.09 |
| Haiti | HAITI (41) | 148 | 149 | 0.09 (−0.07, 0.24) | 0.07 |
| Mali | PROMIS (46) | 499 | 502 | 0.10 (−0.02, 0.23) | 0.10 |
| Mali | PROMIS CS (46) | 944 | 959 | 0.16 (0.06, 0.26) | 0.11 |
| **I² = 0.32, Tau² = 0.00** | | **8332** | **6874** | **0.09 (0.04, 0.13)** | |
| **Sanitation − Unimproved** | | | | | |
| Burkina Faso | iLiNS−Zinc (37) | 1952 | 664 | 0.18 (0.11, 0.26) | 0.21 |
| Kenya | WASH−B (42) | 694 | 2244 | 0.03 (−0.04, 0.11) | 0.20 |
| Madagascar | MAHAY (43) | 1700 | 1682 | 0.00 (−0.11, 0.11) | 0.15 |
| Malawi | iLiNS−DYADM (44) | 220 | 444 | −0.05 (−0.21, 0.11) | 0.10 |
| Malawi | iLiNS−DOSE (45) | 696 | 241 | 0.01 (−0.12, 0.14) | 0.13 |
| Zimbabwe | SHINE (HIV−) (47) | 887 | 876 | 0.02 (−0.09, 0.13) | 0.16 |
| Zimbabwe | SHINE (HIV+) (48) | 147 | 146 | −0.15 (−0.40, 0.10) | 0.05 |
| **I² = 0.67, Tau² = 0.00** | | **6296** | **6297** | **0.03 (−0.04, 0.10)** | |

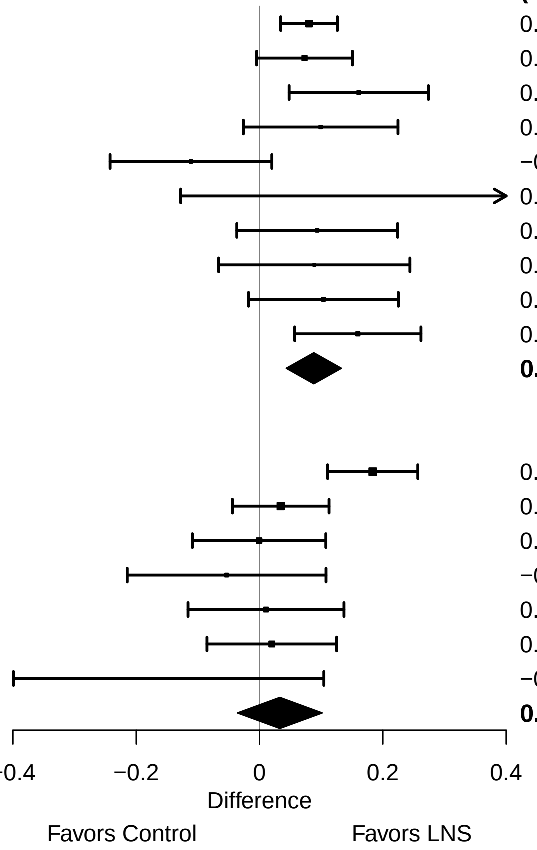

##### Supplemental figure 4D: Mean difference in WLZ

#### 4D6: Stratified by Supplement duration

**Supplement duration**
**(p−diff = 0.207)**

| Country | Trial | N | N | MD (95% CI) | W |
|---|---|---|---|---|---|
| **Supplement duration − 12m or less** | | | | | |
| Bangladesh | JiVitA−4 (34) | 2783 | 1217 | 0.08 (0.03, 0.13) | 0.15 |
| Burkina Faso | iLiNS−Zinc (37) | 1952 | 664 | 0.18 (0.11, 0.26) | 0.12 |
| Burkina Faso | PROMIS (38) | 856 | 907 | 0.10 (−0.03, 0.22) | 0.08 |
| Burkina Faso | PROMIS CS (38) | 430 | 436 | −0.11 (−0.24, 0.02) | 0.08 |
| Ghana | GHANA (39) | 98 | 96 | 0.21 (−0.13, 0.54) | 0.02 |
| Ghana | iLiNS−DYADG (40) | 347 | 692 | 0.09 (−0.04, 0.22) | 0.08 |
| Haiti | HAITI (41) | 148 | 149 | 0.09 (−0.07, 0.24) | 0.06 |
| Madagascar | MAHAY (43) | 1700 | 1682 | 0.00 (−0.11, 0.11) | 0.09 |
| Malawi | iLiNS−DYADM (44) | 220 | 444 | −0.05 (−0.21, 0.11) | 0.06 |
| Malawi | iLiNS−DOSE (45) | 696 | 241 | 0.01 (−0.12, 0.14) | 0.08 |
| Zimbabwe | SHINE (HIV−) (47) | 1869 | 1784 | 0.08 (0.00, 0.15) | 0.13 |
| Zimbabwe | SHINE (HIV+) (48) | 336 | 328 | −0.14 (−0.32, 0.04) | 0.05 |
| **I² = 0.61, Tau² = 0.01** | | **11435** | **8640** | **0.05 (−0.01, 0.10)** | |
| **Supplement duration − > 12m** | | | | | |
| Bangladesh | RDNS (35) | 1661 | 815 | 0.07 (0.00, 0.15) | 0.21 |
| Bangladesh | WASH−B (36) | 1156 | 3424 | 0.12 (0.05, 0.19) | 0.23 |
| Kenya | WASH−B (42) | 1455 | 5118 | 0.06 (0.00, 0.13) | 0.24 |
| Mali | PROMIS (46) | 499 | 502 | 0.10 (−0.02, 0.23) | 0.15 |
| Mali | PROMIS CS (46) | 944 | 959 | 0.16 (0.06, 0.26) | 0.17 |
| **I² = 0.00, Tau² = 0.00** | | **5715** | **10818** | **0.10 (0.06, 0.13)** | |

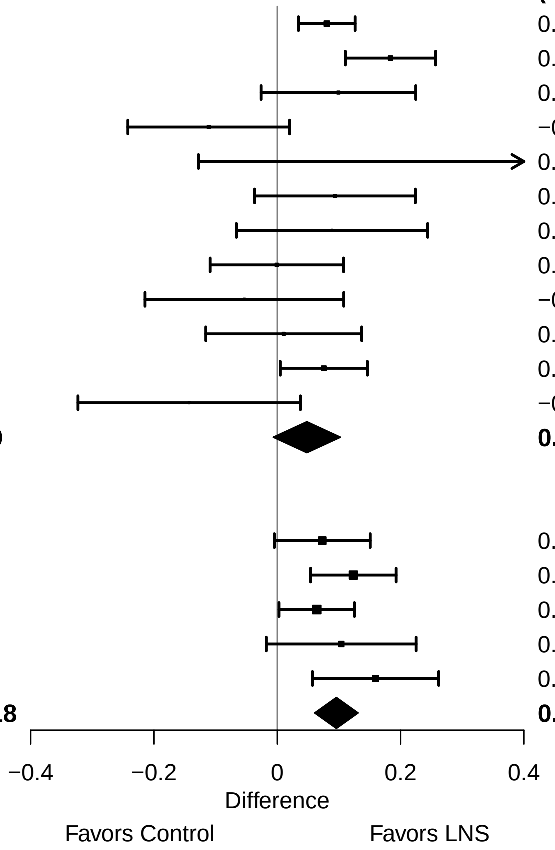

#### Supplemental figure 4D: Mean difference in WLZ

### 4D7: Stratified by Frequency of contact

**Frequency of contact**
**(p−diff = 0.251)**

| Country | Trial | N | N | MD (95% CI) | W |
|---|---|---|---|---|---|
| **Frequency of contact − Monthly** | | | | | |
| Bangladesh | RDNS (35) | 1661 | 815 | 0.07 (0.00, 0.15) | 0.13 |
| Burkina Faso | PROMIS (38) | 856 | 907 | 0.10 (−0.03, 0.22) | 0.09 |
| Burkina Faso | PROMIS CS (38) | 430 | 436 | −0.11 (−0.24, 0.02) | 0.08 |
| Haiti | HAITI (41) | 148 | 149 | 0.09 (−0.07, 0.24) | 0.07 |
| Kenya | WASH−B (42) | 1455 | 5118 | 0.06 (0.00, 0.13) | 0.15 |
| Madagascar | MAHAY (43) | 1700 | 1682 | 0.00 (−0.11, 0.11) | 0.10 |
| Mali | PROMIS (46) | 499 | 502 | 0.10 (−0.02, 0.23) | 0.09 |
| Mali | PROMIS CS (46) | 944 | 959 | 0.16 (0.06, 0.26) | 0.11 |
| Zimbabwe | SHINE (HIV−) (47) | 1869 | 1784 | 0.08 (0.00, 0.15) | 0.14 |
| Zimbabwe | SHINE (HIV+) (48) | 336 | 328 | −0.14 (−0.32, 0.04) | 0.06 |
| **$I^2$ = 0.49, $Tau^2$ = 0.00** | | **9898** | **12680** | **0.05 (0.00, 0.11)** | |
| **Frequency of contact − Weekly** | | | | | |
| Bangladesh | JiVitA−4 (34) | 2783 | 1217 | 0.08 (0.03, 0.13) | 0.23 |
| Bangladesh | WASH−B (36) | 1156 | 3424 | 0.12 (0.05, 0.19) | 0.20 |
| Burkina Faso | iLiNS−Zinc (37) | 1952 | 664 | 0.18 (0.11, 0.26) | 0.20 |
| Ghana | GHANA (39) | 98 | 96 | 0.21 (−0.13, 0.54) | 0.03 |
| Ghana | iLiNS−DYADG (40) | 347 | 692 | 0.09 (−0.04, 0.22) | 0.12 |
| Malawi | iLiNS−DYADM (44) | 220 | 444 | −0.05 (−0.21, 0.11) | 0.09 |
| Malawi | iLiNS−DOSE (45) | 696 | 241 | 0.01 (−0.12, 0.14) | 0.13 |
| **$I^2$ = 0.50, $Tau^2$ = 0.00** | | **7252** | **6778** | **0.09 (0.04, 0.15)** | |

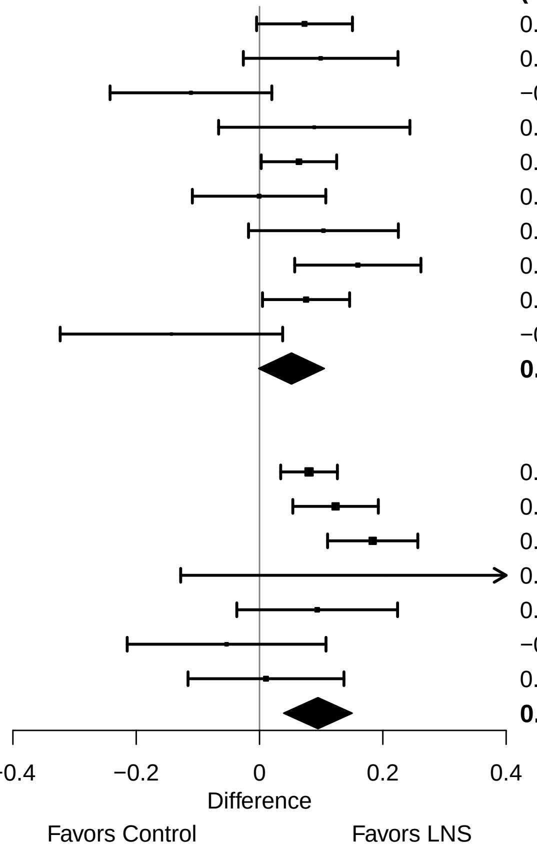

**Supplemental figure 4D: Mean difference in WLZ**

#### 4D8: Stratified by Average SQ-LNS compliance

**Average SQ−LNS compliance**
**(p−diff = 0.101)**

**Average SQ−LNS compliance − Low**

| Country | Trial | N | N | MD (95% CI) | W |
|---|---|---|---|---|---|
| Burkina Faso | PROMIS (38) | 856 | 907 | 0.10 (−0.03, 0.22) | 0.11 |
| Burkina Faso | PROMIS CS (38) | 430 | 436 | −0.11 (−0.24, 0.02) | 0.10 |
| Ghana | iLiNS−DYADG (40) | 347 | 692 | 0.09 (−0.04, 0.22) | 0.11 |
| Malawi | iLiNS−DYADM (44) | 220 | 444 | −0.05 (−0.21, 0.11) | 0.08 |
| Malawi | iLiNS−DOSE (45) | 696 | 241 | 0.01 (−0.12, 0.14) | 0.11 |
| Mali | PROMIS (46) | 499 | 502 | 0.10 (−0.02, 0.23) | 0.11 |
| Mali | PROMIS CS (46) | 944 | 959 | 0.16 (0.06, 0.26) | 0.13 |
| Zimbabwe | SHINE (HIV−) (47) | 1869 | 1784 | 0.08 (0.00, 0.15) | 0.17 |
| Zimbabwe | SHINE (HIV+) (48) | 336 | 328 | −0.14 (−0.32, 0.04) | 0.07 |
| **$I^2$ = 0.57, $Tau^2$ = 0.01** | | **6197** | **6293** | **0.04 (−0.03, 0.10)** | |

**Average SQ−LNS compliance − High**

| Country | Trial | N | N | MD (95% CI) | W |
|---|---|---|---|---|---|
| Bangladesh | JiVitA−4 (34) | 2783 | 1217 | 0.08 (0.03, 0.13) | 0.20 |
| Bangladesh | RDNS (35) | 1661 | 815 | 0.07 (0.00, 0.15) | 0.16 |
| Bangladesh | WASH−B (36) | 1156 | 3424 | 0.12 (0.05, 0.19) | 0.17 |
| Burkina Faso | iLiNS−Zinc (37) | 1952 | 664 | 0.18 (0.11, 0.26) | 0.17 |
| Ghana | GHANA (39) | 98 | 96 | 0.21 (−0.13, 0.54) | 0.03 |
| Haiti | HAITI (41) | 148 | 149 | 0.09 (−0.07, 0.24) | 0.09 |
| Kenya | WASH−B (42) | 1455 | 5118 | 0.06 (0.00, 0.13) | 0.18 |
| **$I^2$ = 0.28, $Tau^2$ = 0.00** | | **9253** | **11483** | **0.10 (0.07, 0.13)** | |

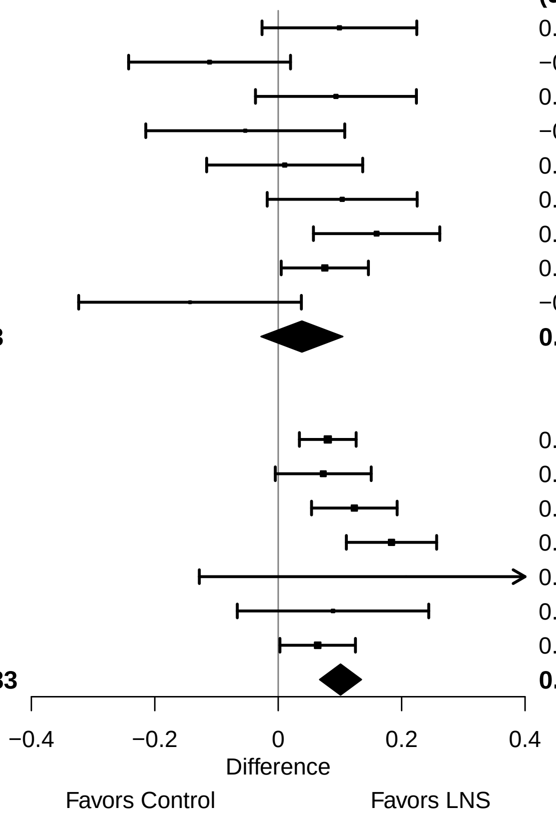

### Supplemental figure 4E: Wasting prevalence ratio

#### 4E1: Stratified by Geographic region

**Geographic region**
**(p−diff = 0.973)**

| Country | Trial | N | N | PR (95% CI) | W |
|---|---|---|---|---|---|
| **Geographic region − SEAR** | | | | | |
| Bangladesh | JiVitA−4 (34) | 2783 | 1217 | 0.91 (0.78, 1.05) | 0.49 |
| Bangladesh | RDNS (35) | 1661 | 815 | 0.87 (0.70, 1.07) | 0.23 |
| Bangladesh | WASH−B (36) | 1156 | 3424 | 0.79 (0.65, 0.96) | 0.28 |
| **I² = 0.00, Tau² = 0.00** | | **5600** | **5456** | **0.86 (0.78, 0.96)** | |
| **Geographic region − AFR** | | | | | |
| Burkina Faso | iLiNS−Zinc (37) | 1952 | 664 | 0.69 (0.54, 0.87) | 0.24 |
| Burkina Faso | PROMIS (38) | 856 | 907 | 0.86 (0.59, 1.27) | 0.09 |
| Burkina Faso | PROMIS CS (38) | 430 | 436 | 1.03 (0.72, 1.49) | 0.10 |
| Ghana | GHANA (39) | 98 | 96 | 0.87 (0.35, 2.16) | 0.02 |
| Ghana | iLiNS−DYADG (40) | 347 | 692 | 0.84 (0.52, 1.37) | 0.06 |
| Kenya | WASH−B (42) | 1455 | 5118 | 0.88 (0.53, 1.45) | 0.05 |
| Madagascar | MAHAY (43) | 1700 | 1682 | 0.96 (0.67, 1.38) | 0.10 |
| Malawi | iLiNS−DYADM (44) | 220 | 444 | 0.94 (0.39, 2.28) | 0.02 |
| Malawi | iLiNS−DOSE (45) | 696 | 241 | 1.37 (0.78, 2.40) | 0.04 |
| Mali | PROMIS (46) | 499 | 502 | 0.61 (0.15, 2.42) | 0.01 |
| Mali | PROMIS CS (46) | 944 | 959 | 0.83 (0.63, 1.10) | 0.17 |
| Zimbabwe | SHINE (HIV−) (47) | 1869 | 1784 | 0.89 (0.60, 1.34) | 0.08 |
| Zimbabwe | SHINE (HIV+) (48) | 336 | 328 | 1.38 (0.65, 2.96) | 0.02 |
| **I² = 0.00, Tau² = 0.00** | | **11402** | **13853** | **0.86 (0.77, 0.97)** | |

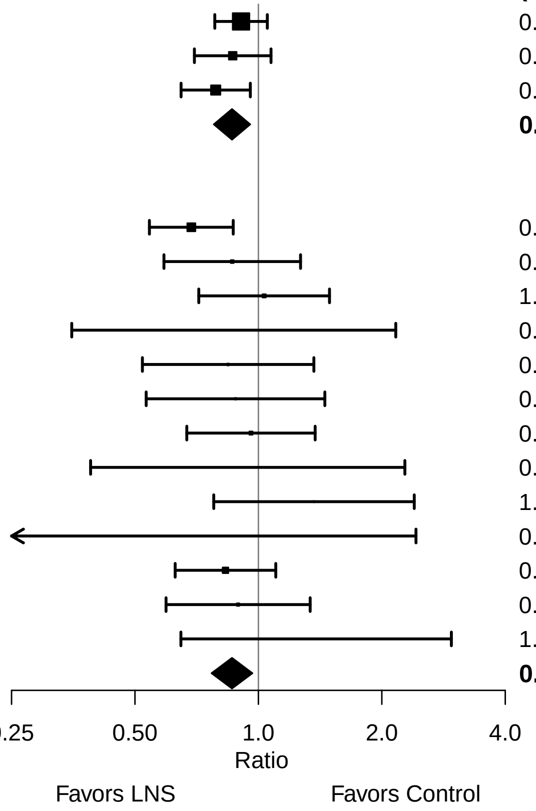

**Supplemental figure 4E: Wasting prevalence ratio**

#### 4E2: Stratified by Stunting burden

**Stunting burden**
**(p−diff = 0.532)**

| Country | Trial | N | N | PR (95% CI) | W |
|---|---|---|---|---|---|
| **Stunting burden − Less than 35%** | | | | | |
| Burkina Faso | PROMIS (38) | 856 | 907 | 0.86 (0.59, 1.27) | 0.27 |
| Burkina Faso | PROMIS CS (38) | 430 | 436 | 1.03 (0.72, 1.49) | 0.29 |
| Ghana | GHANA (39) | 98 | 96 | 0.87 (0.35, 2.16) | 0.05 |
| Ghana | iLiNS−DYADG (40) | 347 | 692 | 0.84 (0.52, 1.37) | 0.17 |
| Haiti | HAITI (41) | 148 | 149 | 0.94 (0.13, 6.52) | 0.01 |
| Kenya | WASH−B (42) | 1455 | 5118 | 0.88 (0.53, 1.45) | 0.16 |
| Malawi | iLiNS−DYADM (44) | 220 | 444 | 0.94 (0.39, 2.28) | 0.05 |
| **I² = 0.00, Tau² = 0.00** | | **3554** | **7842** | **0.91 (0.75, 1.12)** | |
| **Stunting burden − More than 35%** | | | | | |
| Bangladesh | JiVitA−4 (34) | 2783 | 1217 | 0.91 (0.78, 1.05) | 0.32 |
| Bangladesh | RDNS (35) | 1661 | 815 | 0.87 (0.70, 1.07) | 0.15 |
| Bangladesh | WASH−B (36) | 1156 | 3424 | 0.79 (0.65, 0.96) | 0.18 |
| Burkina Faso | iLiNS−Zinc (37) | 1952 | 664 | 0.69 (0.54, 0.87) | 0.13 |
| Madagascar | MAHAY (43) | 1700 | 1682 | 0.96 (0.67, 1.38) | 0.05 |
| Malawi | iLiNS−DOSE (45) | 696 | 241 | 1.37 (0.78, 2.40) | 0.02 |
| Mali | PROMIS (46) | 499 | 502 | 0.61 (0.15, 2.42) | 0.00 |
| Mali | PROMIS CS (46) | 944 | 959 | 0.83 (0.63, 1.10) | 0.09 |
| Zimbabwe | SHINE (HIV−) (47) | 1869 | 1784 | 0.89 (0.60, 1.34) | 0.04 |
| Zimbabwe | SHINE (HIV+) (48) | 336 | 328 | 1.38 (0.65, 2.96) | 0.01 |
| **I² = 0.06, Tau² = 0.00** | | **13596** | **11616** | **0.85 (0.78, 0.93)** | |

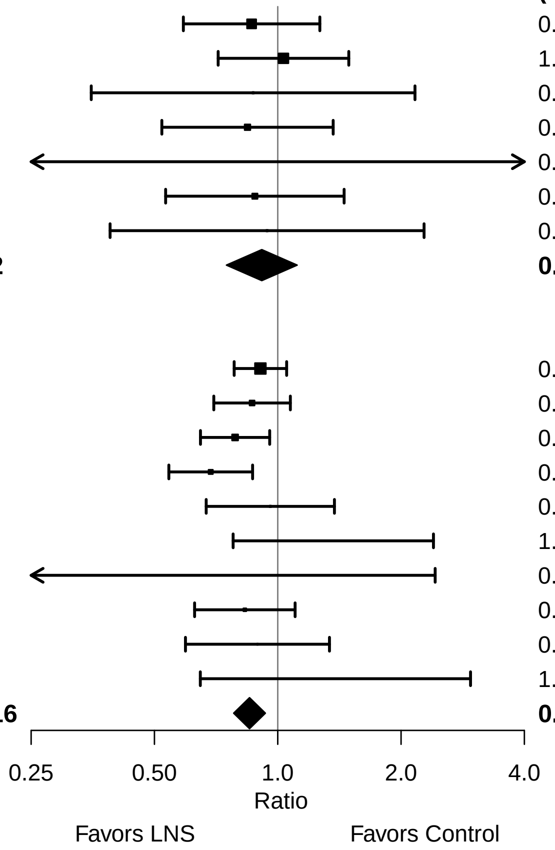

#### Supplemental figure 4E: Wasting prevalence ratio

### 4E3: Stratified by Malaria prevalence

**Malaria prevalence**
**(p−diff = 0.533)**

| Country | Trial | N | N | PR (95% CI) | W |
|---|---|---|---|---|---|
| **Malaria prevalence − Less than 10%** | | | | | |
| Bangladesh | JiVitA−4 (34) | 2783 | 1217 | 0.91 (0.78, 1.05) | 0.40 |
| Bangladesh | RDNS (35) | 1661 | 815 | 0.87 (0.70, 1.07) | 0.19 |
| Bangladesh | WASH−B (36) | 1156 | 3424 | 0.79 (0.65, 0.96) | 0.23 |
| Haiti | HAITI (41) | 148 | 149 | 0.94 (0.13, 6.52) | 0.00 |
| Kenya | WASH−B (42) | 1455 | 5118 | 0.88 (0.53, 1.45) | 0.03 |
| Madagascar | MAHAY (43) | 1700 | 1682 | 0.96 (0.67, 1.38) | 0.07 |
| Zimbabwe | SHINE (HIV−) (47) | 1869 | 1784 | 0.89 (0.60, 1.34) | 0.05 |
| Zimbabwe | SHINE (HIV+) (48) | 336 | 328 | 1.38 (0.65, 2.96) | 0.02 |
| **I² = 0.00, Tau² = 0.00** | | **11108** | **14517** | **0.88 (0.80, 0.96)** | |
| **Malaria prevalence − At least 10%** | | | | | |
| Burkina Faso | iLiNS−Zinc (37) | 1952 | 664 | 0.69 (0.54, 0.87) | 0.33 |
| Burkina Faso | PROMIS (38) | 856 | 907 | 0.86 (0.59, 1.27) | 0.12 |
| Burkina Faso | PROMIS CS (38) | 430 | 436 | 1.03 (0.72, 1.49) | 0.13 |
| Ghana | GHANA (39) | 98 | 96 | 0.87 (0.35, 2.16) | 0.02 |
| Ghana | iLiNS−DYADG (40) | 347 | 692 | 0.84 (0.52, 1.37) | 0.08 |
| Malawi | iLiNS−DYADM (44) | 220 | 444 | 0.94 (0.39, 2.28) | 0.02 |
| Malawi | iLiNS−DOSE (45) | 696 | 241 | 1.37 (0.78, 2.40) | 0.06 |
| Mali | PROMIS (46) | 499 | 502 | 0.61 (0.15, 2.42) | 0.01 |
| Mali | PROMIS CS (46) | 944 | 959 | 0.83 (0.63, 1.10) | 0.23 |
| **I² = 0.00, Tau² = 0.00** | | **6042** | **4941** | **0.83 (0.73, 0.95)** | |

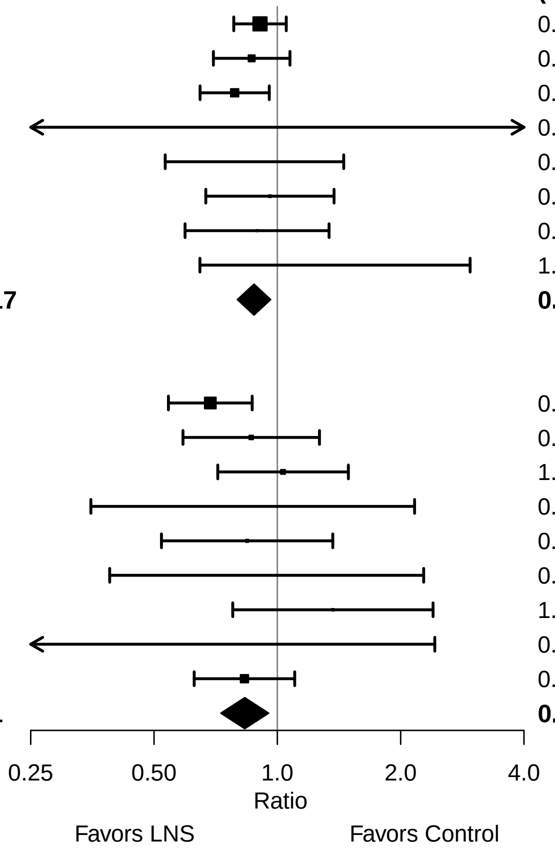

#### Supplemental figure 4E: Wasting prevalence ratio

##### 4E4: Stratified by Source water quality

**Source water quality**
**(p−diff = 0.371)**

| Country | Trial | N | N | PR (95% CI) | W |
|---|---|---|---|---|---|
| **Source water quality − Improved** | | | | | |
| Bangladesh | JiVitA−4 (34) | 2783 | 1217 | 0.91 (0.78, 1.05) | 0.52 |
| Bangladesh | RDNS (35) | 1661 | 815 | 0.87 (0.70, 1.07) | 0.24 |
| Bangladesh | WASH−B (36) | 566 | 1101 | 0.82 (0.61, 1.12) | 0.12 |
| Ghana | GHANA (39) | 98 | 96 | 0.87 (0.35, 2.16) | 0.01 |
| Ghana | iLiNS−DYADG (40) | 347 | 692 | 0.84 (0.52, 1.37) | 0.05 |
| Haiti | HAITI (41) | 148 | 149 | 0.94 (0.13, 6.52) | 0.00 |
| Malawi | iLiNS−DYADM (44) | 220 | 444 | 0.94 (0.39, 2.28) | 0.01 |
| Malawi | iLiNS−DOSE (45) | 696 | 241 | 1.37 (0.78, 2.40) | 0.04 |
| **I² = 0.00, Tau² = 0.00** | | **6519** | **4755** | **0.90 (0.81, 1.00)** | |
| **Source water quality − Unimproved** | | | | | |
| Burkina Faso | iLiNS−Zinc (37) | 1952 | 664 | 0.69 (0.54, 0.87) | 0.31 |
| Burkina Faso | PROMIS (38) | 856 | 907 | 0.86 (0.59, 1.27) | 0.12 |
| Burkina Faso | PROMIS CS (38) | 430 | 436 | 1.03 (0.72, 1.49) | 0.13 |
| Kenya | WASH−B (42) | 694 | 2244 | 0.83 (0.37, 1.90) | 0.03 |
| Madagascar | MAHAY (43) | 1700 | 1682 | 0.96 (0.67, 1.38) | 0.13 |
| Mali | PROMIS (46) | 499 | 502 | 0.61 (0.15, 2.42) | 0.01 |
| Mali | PROMIS CS (46) | 944 | 959 | 0.83 (0.63, 1.10) | 0.21 |
| Zimbabwe | SHINE (HIV−) (47) | 887 | 876 | 0.95 (0.56, 1.61) | 0.06 |
| Zimbabwe | SHINE (HIV+) (48) | 147 | 146 | 0.99 (0.28, 3.54) | 0.01 |
| **I² = 0.00, Tau² = 0.00** | | **8109** | **8416** | **0.83 (0.73, 0.95)** | |

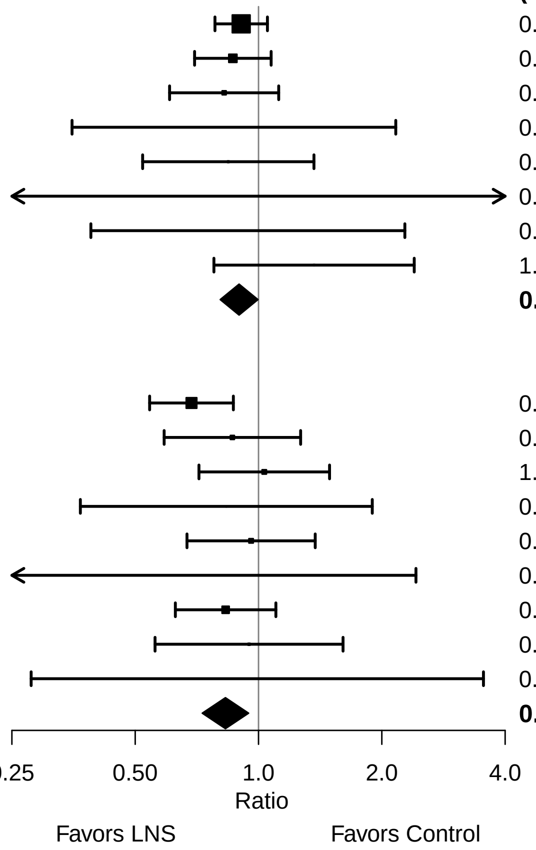

#### Supplemental figure 4E: Wasting prevalence ratio

### 4E5: Stratified by Sanitation

**Sanitation**
**(p−diff = 0.532)**

| Country | Trial | N | N | PR (95% CI) | W |
|---|---|---|---|---|---|
| **Sanitation − Improved** | | | | | |
| Bangladesh | JiVitA−4 (34) | 2783 | 1217 | 0.91 (0.78, 1.05) | 0.41 |
| Bangladesh | RDNS (35) | 1661 | 815 | 0.87 (0.70, 1.07) | 0.19 |
| Bangladesh | WASH−B (36) | 566 | 1101 | 0.82 (0.61, 1.12) | 0.10 |
| Burkina Faso | PROMIS (38) | 856 | 907 | 0.86 (0.59, 1.27) | 0.06 |
| Burkina Faso | PROMIS CS (38) | 430 | 436 | 1.03 (0.72, 1.49) | 0.07 |
| Ghana | GHANA (39) | 98 | 96 | 0.87 (0.35, 2.16) | 0.01 |
| Ghana | iLiNS−DYADG (40) | 347 | 692 | 0.84 (0.52, 1.37) | 0.04 |
| Haiti | HAITI (41) | 148 | 149 | 0.94 (0.13, 6.52) | 0.00 |
| Mali | PROMIS (46) | 499 | 502 | 0.61 (0.15, 2.42) | 0.00 |
| Mali | PROMIS CS (46) | 944 | 959 | 0.83 (0.63, 1.10) | 0.11 |
| **I² = 0.00, Tau² = 0.00** | | **8332** | **6874** | **0.88 (0.80, 0.97)** | |
| **Sanitation − Unimproved** | | | | | |
| Burkina Faso | iLiNS−Zinc (37) | 1952 | 664 | 0.69 (0.54, 0.87) | 0.50 |
| Kenya | WASH−B (42) | 694 | 2244 | 0.83 (0.37, 1.90) | 0.04 |
| Madagascar | MAHAY (43) | 1700 | 1682 | 0.96 (0.67, 1.38) | 0.21 |
| Malawi | iLiNS−DYADM (44) | 220 | 444 | 0.94 (0.39, 2.28) | 0.04 |
| Malawi | iLiNS−DOSE (45) | 696 | 241 | 1.37 (0.78, 2.40) | 0.09 |
| Zimbabwe | SHINE (HIV−) (47) | 887 | 876 | 0.95 (0.56, 1.61) | 0.10 |
| Zimbabwe | SHINE (HIV+) (48) | 147 | 146 | 0.99 (0.28, 3.54) | 0.02 |
| **I² = 0.08, Tau² = 0.00** | | **6296** | **6297** | **0.84 (0.70, 1.00)** | |

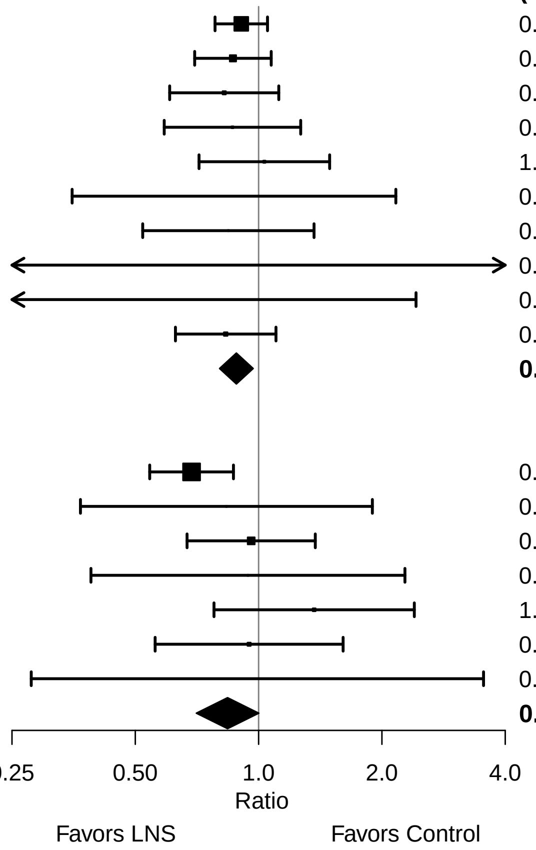

#### Supplemental figure 4E: Wasting prevalence ratio

#### 4E6: Stratified by Supplement duration

**Supplement duration**
**(p−diff = 0.368)**

**Supplement duration − 12m or less**

| Country | Trial | N | N | PR (95% CI) | W |
|---|---|---|---|---|---|
| Bangladesh | JiVitA−4 (34) | 2783 | 1217 | 0.91 (0.78, 1.05) | 0.44 |
| Burkina Faso | iLiNS−Zinc (37) | 1952 | 664 | 0.69 (0.54, 0.87) | 0.17 |
| Burkina Faso | PROMIS (38) | 856 | 907 | 0.86 (0.59, 1.27) | 0.07 |
| Burkina Faso | PROMIS CS (38) | 430 | 436 | 1.03 (0.72, 1.49) | 0.07 |
| Ghana | GHANA (39) | 98 | 96 | 0.87 (0.35, 2.16) | 0.01 |
| Ghana | iLiNS−DYADG (40) | 347 | 692 | 0.84 (0.52, 1.37) | 0.04 |
| Haiti | HAITI (41) | 148 | 149 | 0.94 (0.13, 6.52) | 0.00 |
| Madagascar | MAHAY (43) | 1700 | 1682 | 0.96 (0.67, 1.38) | 0.07 |
| Malawi | iLiNS−DYADM (44) | 220 | 444 | 0.94 (0.39, 2.28) | 0.01 |
| Malawi | iLiNS−DOSE (45) | 696 | 241 | 1.37 (0.78, 2.40) | 0.03 |
| Zimbabwe | SHINE (HIV−) (47) | 1869 | 1784 | 0.89 (0.60, 1.34) | 0.06 |
| Zimbabwe | SHINE (HIV+) (48) | 336 | 328 | 1.38 (0.65, 2.96) | 0.02 |
| **I² = 0.00, Tau² = 0.00** | | **11435** | **8640** | **0.89 (0.80, 0.98)** | |

**Supplement duration − > 12m**

| Country | Trial | N | N | PR (95% CI) | W |
|---|---|---|---|---|---|
| Bangladesh | RDNS (35) | 1661 | 815 | 0.87 (0.70, 1.07) | 0.33 |
| Bangladesh | WASH−B (36) | 1156 | 3424 | 0.79 (0.65, 0.96) | 0.41 |
| Kenya | WASH−B (42) | 1455 | 5118 | 0.88 (0.53, 1.45) | 0.06 |
| Mali | PROMIS (46) | 499 | 502 | 0.61 (0.15, 2.42) | 0.01 |
| Mali | PROMIS CS (46) | 944 | 959 | 0.83 (0.63, 1.10) | 0.19 |
| **I² = 0.00, Tau² = 0.00** | | **5715** | **10818** | **0.82 (0.73, 0.93)** | |

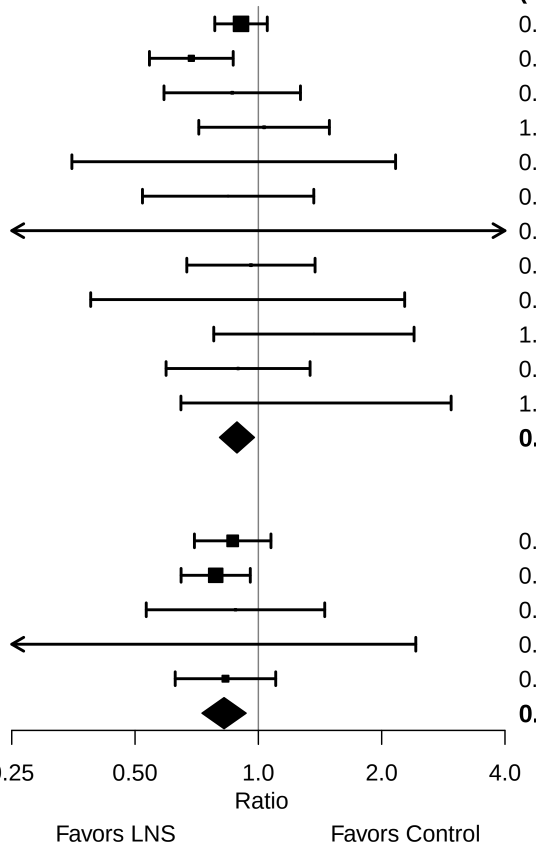

**Supplemental figure 4E: Wasting prevalence ratio**

#### 4E7: Stratified by Frequency of contact

**Frequency of contact**
**(p−diff = 0.399)**

| Country | Trial | N | N | PR (95% CI) | W |
|---|---|---|---|---|---|
| **Frequency of contact − Monthly** | | | | | |
| Bangladesh | RDNS (35) | 1661 | 815 | 0.87 (0.70, 1.07) | 0.31 |
| Burkina Faso | PROMIS (38) | 856 | 907 | 0.86 (0.59, 1.27) | 0.10 |
| Burkina Faso | PROMIS CS (38) | 430 | 436 | 1.03 (0.72, 1.49) | 0.11 |
| Haiti | HAITI (41) | 148 | 149 | 0.94 (0.13, 6.52) | 0.00 |
| Kenya | WASH−B (42) | 1455 | 5118 | 0.88 (0.53, 1.45) | 0.06 |
| Madagascar | MAHAY (43) | 1700 | 1682 | 0.96 (0.67, 1.38) | 0.11 |
| Mali | PROMIS (46) | 499 | 502 | 0.61 (0.15, 2.42) | 0.01 |
| Mali | PROMIS CS (46) | 944 | 959 | 0.83 (0.63, 1.10) | 0.18 |
| Zimbabwe | SHINE (HIV−) (47) | 1869 | 1784 | 0.89 (0.60, 1.34) | 0.09 |
| Zimbabwe | SHINE (HIV+) (48) | 336 | 328 | 1.38 (0.65, 2.96) | 0.03 |
| **I² = 0.00, Tau² = 0.00** | | **9898** | **12680** | **0.90 (0.80, 1.01)** | |
| **Frequency of contact − Weekly** | | | | | |
| Bangladesh | JiVitA−4 (34) | 2783 | 1217 | 0.91 (0.78, 1.05) | 0.46 |
| Bangladesh | WASH−B (36) | 1156 | 3424 | 0.79 (0.65, 0.96) | 0.26 |
| Burkina Faso | iLiNS−Zinc (37) | 1952 | 664 | 0.69 (0.54, 0.87) | 0.18 |
| Ghana | GHANA (39) | 98 | 96 | 0.87 (0.35, 2.16) | 0.01 |
| Ghana | iLiNS−DYADG (40) | 347 | 692 | 0.84 (0.52, 1.37) | 0.04 |
| Malawi | iLiNS−DYADM (44) | 220 | 444 | 0.94 (0.39, 2.28) | 0.01 |
| Malawi | iLiNS−DOSE (45) | 696 | 241 | 1.37 (0.78, 2.40) | 0.03 |
| **I² = 0.17, Tau² = 0.00** | | **7252** | **6778** | **0.84 (0.74, 0.94)** | |

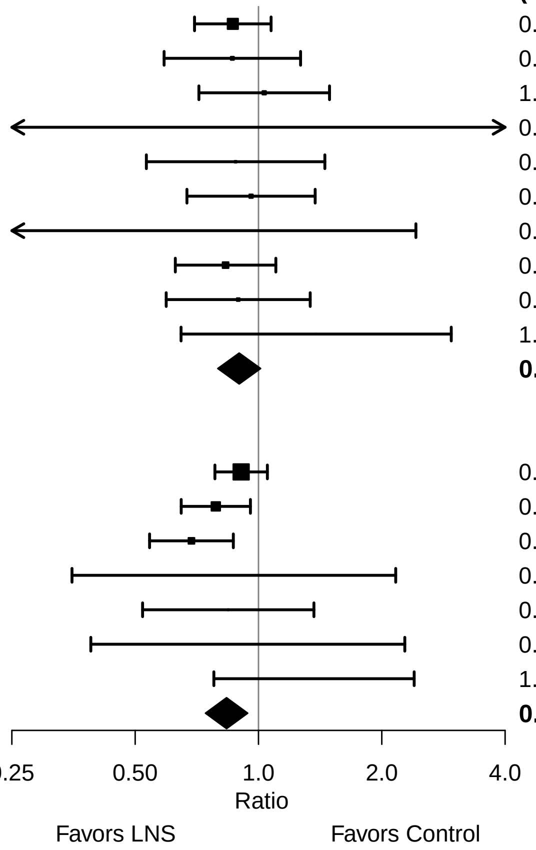

#### Supplemental figure 4E: Wasting prevalence ratio

### 4E8: Stratified by Average SQ-LNS compliance

**Average SQ−LNS compliance**
**(p−diff = 0.231)**

**Average SQ−LNS compliance − Low**

| **Country** | **Trial** | **N** | **N** | **PR (95% CI)** | **W** |
|---|---|---|---|---|---|
| Burkina Faso | PROMIS (38) | 856 | 907 | 0.86 (0.59, 1.27) | 0.15 |
| Burkina Faso | PROMIS CS (38) | 430 | 436 | 1.03 (0.72, 1.49) | 0.17 |
| Ghana | iLiNS−DYADG (40) | 347 | 692 | 0.84 (0.52, 1.37) | 0.10 |
| Malawi | iLiNS−DYADM (44) | 220 | 444 | 0.94 (0.39, 2.28) | 0.03 |
| Malawi | iLiNS−DOSE (45) | 696 | 241 | 1.37 (0.78, 2.40) | 0.07 |
| Mali | PROMIS (46) | 499 | 502 | 0.61 (0.15, 2.42) | 0.01 |
| Mali | PROMIS CS (46) | 944 | 959 | 0.83 (0.63, 1.10) | 0.29 |
| Zimbabwe | SHINE (HIV−) (47) | 1869 | 1784 | 0.89 (0.60, 1.34) | 0.14 |
| Zimbabwe | SHINE (HIV+) (48) | 336 | 328 | 1.38 (0.65, 2.96) | 0.04 |
| **I² = 0.00, Tau² = 0.00** | | **6197** | **6293** | **0.93 (0.80, 1.08)** | |

**Average SQ−LNS compliance − High**

| **Country** | **Trial** | **N** | **N** | **PR (95% CI)** | **W** |
|---|---|---|---|---|---|
| Bangladesh | JiVitA−4 (34) | 2783 | 1217 | 0.91 (0.78, 1.05) | 0.39 |
| Bangladesh | RDNS (35) | 1661 | 815 | 0.87 (0.70, 1.07) | 0.18 |
| Bangladesh | WASH−B (36) | 1156 | 3424 | 0.79 (0.65, 0.96) | 0.22 |
| Burkina Faso | iLiNS−Zinc (37) | 1952 | 664 | 0.69 (0.54, 0.87) | 0.15 |
| Ghana | GHANA (39) | 98 | 96 | 0.87 (0.35, 2.16) | 0.01 |
| Haiti | HAITI (41) | 148 | 149 | 0.94 (0.13, 6.52) | 0.00 |
| Kenya | WASH−B (42) | 1455 | 5118 | 0.88 (0.53, 1.45) | 0.03 |
| **I² = 0.00, Tau² = 0.00** | | **9253** | **11483** | **0.83 (0.76, 0.91)** | |

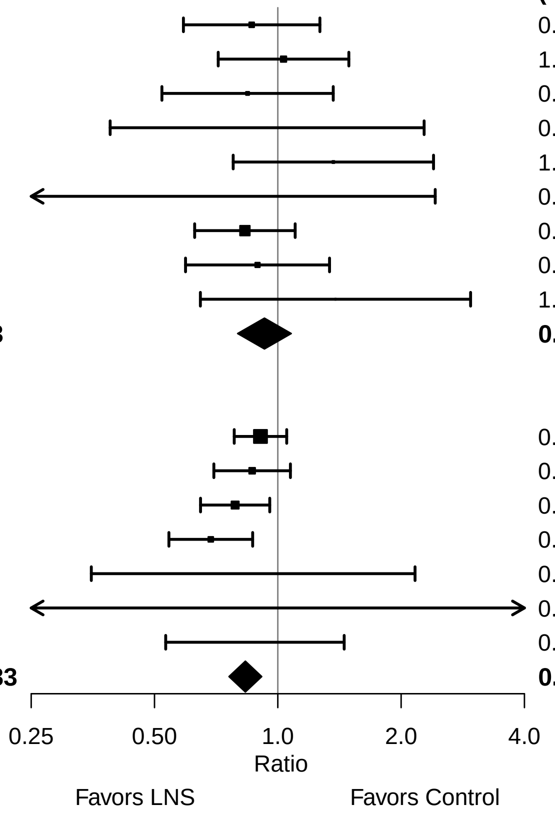

### Supplemental figure 4F: Wasting prevalence difference

#### 4F1: Stratified by Geographic region

**Geographic region**
**(p−diff = 0.025)**

| Country | Trial | N | N | PD (95% CI) | W |
|---|---|---|---|---|---|
| **Geographic region − SEAR** | | | | | |
| Bangladesh | JiVitA−4 (34) | 2783 | 1217 | −0.01 (−0.04, 0.01) | 0.34 |
| Bangladesh | RDNS (35) | 1661 | 815 | −0.02 (−0.05, 0.01) | 0.20 |
| Bangladesh | WASH−B (36) | 1156 | 3424 | −0.02 (−0.04, 0.00) | 0.46 |
| **I² = 0.00, Tau² = 0.00** | | **5600** | **5456** | **−0.02 (−0.03, −0.01)** | |
| **Geographic region − AFR** | | | | | |
| Burkina Faso | iLiNS−Zinc (37) | 1952 | 664 | −0.04 (−0.07, −0.01) | 0.03 |
| Burkina Faso | PROMIS (38) | 856 | 907 | −0.01 (−0.04, 0.01) | 0.03 |
| Burkina Faso | PROMIS CS (38) | 430 | 436 | 0.00 (−0.04, 0.05) | 0.01 |
| Ghana | GHANA (39) | 98 | 96 | −0.01 (−0.09, 0.07) | 0.00 |
| Ghana | iLiNS−DYADG (40) | 347 | 692 | −0.01 (−0.04, 0.02) | 0.02 |
| Kenya | WASH−B (42) | 1455 | 5118 | 0.00 (−0.01, 0.00) | 0.44 |
| Madagascar | MAHAY (43) | 1700 | 1682 | 0.00 (−0.02, 0.01) | 0.08 |
| Malawi | iLiNS−DYADM (44) | 220 | 444 | 0.00 (−0.03, 0.03) | 0.02 |
| Malawi | iLiNS−DOSE (45) | 696 | 241 | 0.01 (−0.03, 0.04) | 0.02 |
| Mali | PROMIS (46) | 499 | 502 | −0.01 (−0.02, 0.01) | 0.10 |
| Mali | PROMIS CS (46) | 944 | 959 | −0.02 (−0.04, 0.01) | 0.03 |
| Zimbabwe | SHINE (HIV−) (47) | 1869 | 1784 | 0.00 (−0.01, 0.01) | 0.20 |
| Zimbabwe | SHINE (HIV+) (48) | 336 | 328 | 0.01 (−0.02, 0.05) | 0.02 |
| **I² = 0.00, Tau² = 0.00** | | **11402** | **13853** | **0.00 (−0.01, 0.00)** | |

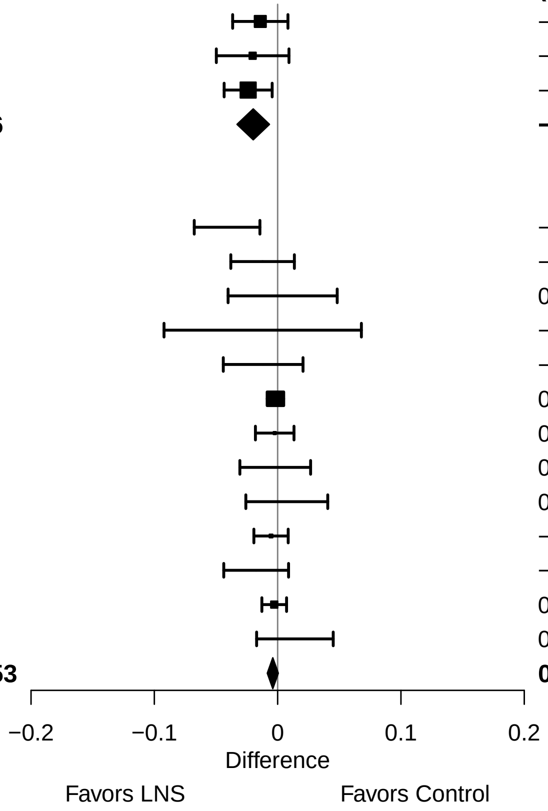

#### Supplemental figure 4F: Wasting prevalence difference

### 4F2: Stratified by Stunting burden

**Stunting burden**
**(p−diff = 0.203)**

| Country | Trial | N | N | PD (95% CI) | W |
|---|---|---|---|---|---|
| **Stunting burden − Less than 35%** | | | | | |
| Burkina Faso | PROMIS (38) | 856 | 907 | −0.01 (−0.04, 0.01) | 0.05 |
| Burkina Faso | PROMIS CS (38) | 430 | 436 | 0.00 (−0.04, 0.05) | 0.02 |
| Ghana | GHANA (39) | 98 | 96 | −0.01 (−0.09, 0.07) | 0.01 |
| Ghana | iLiNS−DYADG (40) | 347 | 692 | −0.01 (−0.04, 0.02) | 0.03 |
| Haiti | HAITI (41) | 148 | 149 | 0.00 (−0.03, 0.03) | 0.05 |
| Kenya | WASH−B (42) | 1455 | 5118 | 0.00 (−0.01, 0.00) | 0.79 |
| Malawi | iLiNS−DYADM (44) | 220 | 444 | 0.00 (−0.03, 0.03) | 0.04 |
| **I² = 0.00, Tau² = 0.00** | | **3554** | **7842** | **0.00 (−0.01, 0.00)** | |
| **Stunting burden − More than 35%** | | | | | |
| Bangladesh | JiVitA−4 (34) | 2783 | 1217 | −0.01 (−0.04, 0.01) | 0.07 |
| Bangladesh | RDNS (35) | 1661 | 815 | −0.02 (−0.05, 0.01) | 0.04 |
| Bangladesh | WASH−B (36) | 1156 | 3424 | −0.02 (−0.04, 0.00) | 0.09 |
| Burkina Faso | iLiNS−Zinc (37) | 1952 | 664 | −0.04 (−0.07, −0.01) | 0.05 |
| Madagascar | MAHAY (43) | 1700 | 1682 | 0.00 (−0.02, 0.01) | 0.14 |
| Malawi | iLiNS−DOSE (45) | 696 | 241 | 0.01 (−0.03, 0.04) | 0.03 |
| Mali | PROMIS (46) | 499 | 502 | −0.01 (−0.02, 0.01) | 0.17 |
| Mali | PROMIS CS (46) | 944 | 959 | −0.02 (−0.04, 0.01) | 0.05 |
| Zimbabwe | SHINE (HIV−) (47) | 1869 | 1784 | 0.00 (−0.01, 0.01) | 0.33 |
| Zimbabwe | SHINE (HIV+) (48) | 336 | 328 | 0.01 (−0.02, 0.05) | 0.03 |
| **I² = 0.37, Tau² = 0.00** | | **13596** | **11616** | **−0.01 (−0.02, 0.00)** | |

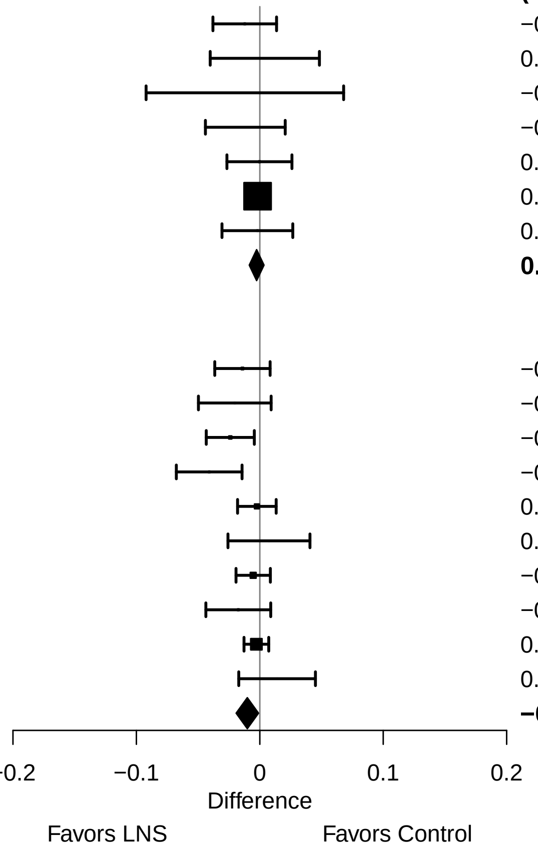

#### Supplemental figure 4F: Wasting prevalence difference

### 4F3: Stratified by Malaria prevalence

**Malaria prevalence**
**(p−diff = 0.216)**

| Country | Trial | N | N | PD (95% CI) | W |
|---|---|---|---|---|---|
| **Malaria prevalence − Less than 10%** | | | | | |
| Bangladesh | JiVitA−4 (34) | 2783 | 1217 | −0.01 (−0.04, 0.01) | 0.04 |
| Bangladesh | RDNS (35) | 1661 | 815 | −0.02 (−0.05, 0.01) | 0.03 |
| Bangladesh | WASH−B (36) | 1156 | 3424 | −0.02 (−0.04, 0.00) | 0.06 |
| Haiti | HAITI (41) | 148 | 149 | 0.00 (−0.03, 0.03) | 0.03 |
| Kenya | WASH−B (42) | 1455 | 5118 | 0.00 (−0.01, 0.00) | 0.50 |
| Madagascar | MAHAY (43) | 1700 | 1682 | 0.00 (−0.02, 0.01) | 0.09 |
| Zimbabwe | SHINE (HIV−) (47) | 1869 | 1784 | 0.00 (−0.01, 0.01) | 0.22 |
| Zimbabwe | SHINE (HIV+) (48) | 336 | 328 | 0.01 (−0.02, 0.05) | 0.02 |
| **I² = 0.11, Tau² = 0.00** | | **11108** | **14517** | **0.00 (−0.01, 0.00)** | |
| **Malaria prevalence − At least 10%** | | | | | |
| Burkina Faso | iLiNS−Zinc (37) | 1952 | 664 | −0.04 (−0.07, −0.01) | 0.11 |
| Burkina Faso | PROMIS (38) | 856 | 907 | −0.01 (−0.04, 0.01) | 0.11 |
| Burkina Faso | PROMIS CS (38) | 430 | 436 | 0.00 (−0.04, 0.05) | 0.04 |
| Ghana | GHANA (39) | 98 | 96 | −0.01 (−0.09, 0.07) | 0.01 |
| Ghana | iLiNS−DYADG (40) | 347 | 692 | −0.01 (−0.04, 0.02) | 0.07 |
| Malawi | iLiNS−DYADM (44) | 220 | 444 | 0.00 (−0.03, 0.03) | 0.09 |
| Malawi | iLiNS−DOSE (45) | 696 | 241 | 0.01 (−0.03, 0.04) | 0.07 |
| Mali | PROMIS (46) | 499 | 502 | −0.01 (−0.02, 0.01) | 0.39 |
| Mali | PROMIS CS (46) | 944 | 959 | −0.02 (−0.04, 0.01) | 0.11 |
| **I² = 0.00, Tau² = 0.00** | | **6042** | **4941** | **−0.01 (−0.02, 0.00)** | |

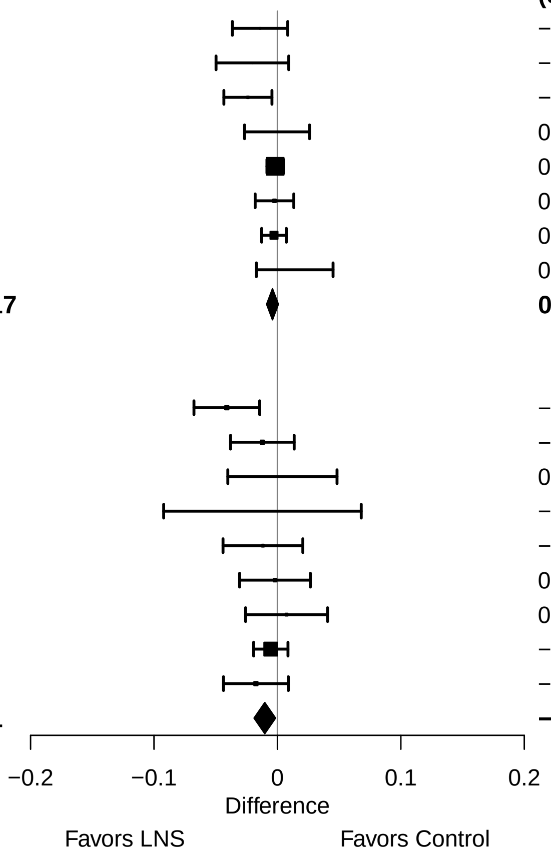

#### Supplemental figure 4F: Wasting prevalence difference

### 4F4: Stratified by Source water quality

**Source water quality**
**(p−diff = 0.560)**

| Country | Trial | N | N | PD (95% CI) | W |
|---|---|---|---|---|---|
| **Source water quality − Improved** | | | | | |
| Bangladesh | JiVitA−4 (34) | 2783 | 1217 | −0.01 (−0.04, 0.01) | 0.22 |
| Bangladesh | RDNS (35) | 1661 | 815 | −0.02 (−0.05, 0.01) | 0.13 |
| Bangladesh | WASH−B (36) | 566 | 1101 | −0.02 (−0.05, 0.01) | 0.13 |
| Ghana | GHANA (39) | 98 | 96 | −0.01 (−0.09, 0.07) | 0.02 |
| Ghana | iLiNS−DYADG (40) | 347 | 692 | −0.01 (−0.04, 0.02) | 0.11 |
| Haiti | HAITI (41) | 148 | 149 | 0.00 (−0.03, 0.03) | 0.16 |
| Malawi | iLiNS−DYADM (44) | 220 | 444 | 0.00 (−0.03, 0.03) | 0.14 |
| Malawi | iLiNS−DOSE (45) | 696 | 241 | 0.01 (−0.03, 0.04) | 0.10 |
| **I² = 0.00, Tau² = 0.00** | | **6519** | **4755** | **−0.01 (−0.02, 0.00)** | |
| **Source water quality − Unimproved** | | | | | |
| Burkina Faso | iLiNS−Zinc (37) | 1952 | 664 | −0.04 (−0.07, −0.01) | 0.05 |
| Burkina Faso | PROMIS (38) | 856 | 907 | −0.01 (−0.04, 0.01) | 0.05 |
| Burkina Faso | PROMIS CS (38) | 430 | 436 | 0.00 (−0.04, 0.05) | 0.02 |
| Kenya | WASH−B (42) | 694 | 2244 | 0.00 (−0.01, 0.01) | 0.38 |
| Madagascar | MAHAY (43) | 1700 | 1682 | 0.00 (−0.02, 0.01) | 0.13 |
| Mali | PROMIS (46) | 499 | 502 | −0.01 (−0.02, 0.01) | 0.17 |
| Mali | PROMIS CS (46) | 944 | 959 | −0.02 (−0.04, 0.01) | 0.05 |
| Zimbabwe | SHINE (HIV−) (47) | 887 | 876 | 0.00 (−0.02, 0.01) | 0.14 |
| Zimbabwe | SHINE (HIV+) (48) | 147 | 146 | 0.00 (−0.05, 0.05) | 0.02 |
| **I² = 0.11, Tau² = 0.00** | | **8109** | **8416** | **−0.01 (−0.01, 0.00)** | |

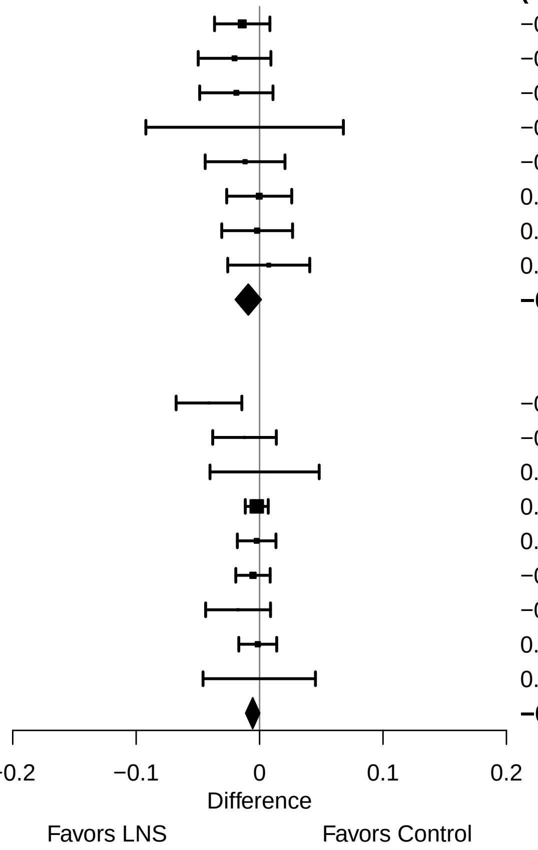

**Supplemental figure 4F: Wasting prevalence difference**

#### 4F5: Stratified by Sanitation

**Sanitation**
**(p−diff = 0.251)**

| Country | Trial | N | N | PD (95% CI) | W |
|---|---|---|---|---|---|
| **Sanitation − Improved** | | | | | |
| Bangladesh | JiVitA−4 (34) | 2783 | 1217 | −0.01 (−0.04, 0.01) | 0.13 |
| Bangladesh | RDNS (35) | 1661 | 815 | −0.02 (−0.05, 0.01) | 0.07 |
| Bangladesh | WASH−B (36) | 566 | 1101 | −0.02 (−0.05, 0.01) | 0.07 |
| Burkina Faso | PROMIS (38) | 856 | 907 | −0.01 (−0.04, 0.01) | 0.10 |
| Burkina Faso | PROMIS CS (38) | 430 | 436 | 0.00 (−0.04, 0.05) | 0.03 |
| Ghana | GHANA (39) | 98 | 96 | −0.01 (−0.09, 0.07) | 0.01 |
| Ghana | iLiNS−DYADG (40) | 347 | 692 | −0.01 (−0.04, 0.02) | 0.06 |
| Haiti | HAITI (41) | 148 | 149 | 0.00 (−0.03, 0.03) | 0.09 |
| Mali | PROMIS (46) | 499 | 502 | −0.01 (−0.02, 0.01) | 0.33 |
| Mali | PROMIS CS (46) | 944 | 959 | −0.02 (−0.04, 0.01) | 0.09 |
| **I² = 0.00, Tau² = 0.00** | | **8332** | **6874** | **−0.01 (−0.02, 0.00)** | |
| **Sanitation − Unimproved** | | | | | |
| Burkina Faso | iLiNS−Zinc (37) | 1952 | 664 | −0.04 (−0.07, −0.01) | 0.06 |
| Kenya | WASH−B (42) | 694 | 2244 | 0.00 (−0.01, 0.01) | 0.48 |
| Madagascar | MAHAY (43) | 1700 | 1682 | 0.00 (−0.02, 0.01) | 0.17 |
| Malawi | iLiNS−DYADM (44) | 220 | 444 | 0.00 (−0.03, 0.03) | 0.05 |
| Malawi | iLiNS−DOSE (45) | 696 | 241 | 0.01 (−0.03, 0.04) | 0.04 |
| Zimbabwe | SHINE (HIV−) (47) | 887 | 876 | 0.00 (−0.02, 0.01) | 0.18 |
| Zimbabwe | SHINE (HIV+) (48) | 147 | 146 | 0.00 (−0.05, 0.05) | 0.02 |
| **I² = 0.27, Tau² = 0.00** | | **6296** | **6297** | **−0.01 (−0.02, 0.01)** | |

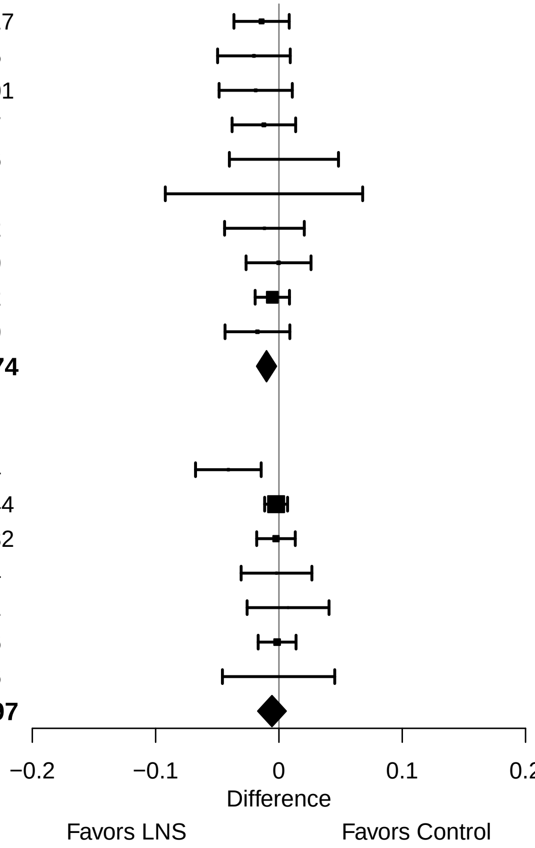

#### Supplemental figure 4F: Wasting prevalence difference

### 4F6: Stratified by Supplement duration

**Supplement duration**
**(p−diff = 0.696)**

| Country | Trial | N | N | PD (95% CI) | W |
|---|---|---|---|---|---|
| **Supplement duration − 12m or less** | | | | | |
| Bangladesh | JiVitA−4 (34) | 2783 | 1217 | −0.01 (−0.04, 0.01) | 0.08 |
| Burkina Faso | iLiNS−Zinc (37) | 1952 | 664 | −0.04 (−0.07, −0.01) | 0.06 |
| Burkina Faso | PROMIS (38) | 856 | 907 | −0.01 (−0.04, 0.01) | 0.06 |
| Burkina Faso | PROMIS CS (38) | 430 | 436 | 0.00 (−0.04, 0.05) | 0.02 |
| Ghana | GHANA (39) | 98 | 96 | −0.01 (−0.09, 0.07) | 0.01 |
| Ghana | iLiNS−DYADG (40) | 347 | 692 | −0.01 (−0.04, 0.02) | 0.04 |
| Haiti | HAITI (41) | 148 | 149 | 0.00 (−0.03, 0.03) | 0.06 |
| Madagascar | MAHAY (43) | 1700 | 1682 | 0.00 (−0.02, 0.01) | 0.16 |
| Malawi | iLiNS−DYADM (44) | 220 | 444 | 0.00 (−0.03, 0.03) | 0.05 |
| Malawi | iLiNS−DOSE (45) | 696 | 241 | 0.01 (−0.03, 0.04) | 0.04 |
| Zimbabwe | SHINE (HIV−) (47) | 1869 | 1784 | 0.00 (−0.01, 0.01) | 0.40 |
| Zimbabwe | SHINE (HIV+) (48) | 336 | 328 | 0.01 (−0.02, 0.05) | 0.04 |
| **$I^2$ = 0.00, $Tau^2$ = 0.00** | | **11435** | **8640** | **−0.01 (−0.01, 0.00)** | |
| **Supplement duration − > 12m** | | | | | |
| Bangladesh | RDNS (35) | 1661 | 815 | −0.02 (−0.05, 0.01) | 0.04 |
| Bangladesh | WASH−B (36) | 1156 | 3424 | −0.02 (−0.04, 0.00) | 0.08 |
| Kenya | WASH−B (42) | 1455 | 5118 | 0.00 (−0.01, 0.00) | 0.68 |
| Mali | PROMIS (46) | 499 | 502 | −0.01 (−0.02, 0.01) | 0.16 |
| Mali | PROMIS CS (46) | 944 | 959 | −0.02 (−0.04, 0.01) | 0.04 |
| **$I^2$ = 0.37, $Tau^2$ = 0.00** | | **5715** | **10818** | **−0.01 (−0.02, 0.00)** | |

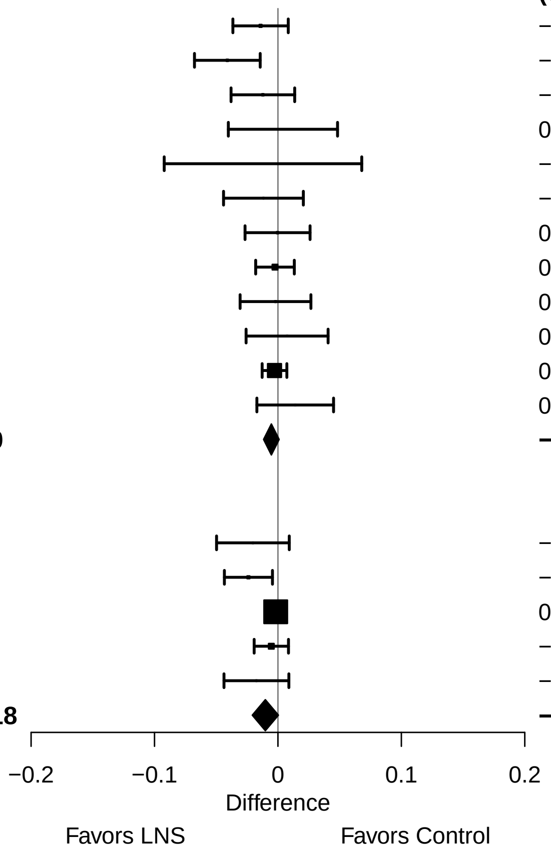

#### Supplemental figure 4F: Wasting prevalence difference

### 4F7: Stratified by Frequency of contact

**Frequency of contact**
**(p−diff = 0.017)**

| Country | Trial | N | N | PD (95% CI) | W |
|---|---|---|---|---|---|
| **Frequency of contact − Monthly** | | | | | |
| Bangladesh | RDNS (35) | 1661 | 815 | −0.02 (−0.05, 0.01) | 0.02 |
| Burkina Faso | PROMIS (38) | 856 | 907 | −0.01 (−0.04, 0.01) | 0.03 |
| Burkina Faso | PROMIS CS (38) | 430 | 436 | 0.00 (−0.04, 0.05) | 0.01 |
| Haiti | HAITI (41) | 148 | 149 | 0.00 (−0.03, 0.03) | 0.03 |
| Kenya | WASH−B (42) | 1455 | 5118 | 0.00 (−0.01, 0.00) | 0.46 |
| Madagascar | MAHAY (43) | 1700 | 1682 | 0.00 (−0.02, 0.01) | 0.08 |
| Mali | PROMIS (46) | 499 | 502 | −0.01 (−0.02, 0.01) | 0.11 |
| Mali | PROMIS CS (46) | 944 | 959 | −0.02 (−0.04, 0.01) | 0.03 |
| Zimbabwe | SHINE (HIV−) (47) | 1869 | 1784 | 0.00 (−0.01, 0.01) | 0.21 |
| Zimbabwe | SHINE (HIV+) (48) | 336 | 328 | 0.01 (−0.02, 0.05) | 0.02 |
| **I² = 0.00, Tau² = 0.00** | | **9898** | **12680** | **0.00 (−0.01, 0.00)** | |
| **Frequency of contact − Weekly** | | | | | |
| Bangladesh | JiVitA−4 (34) | 2783 | 1217 | −0.01 (−0.04, 0.01) | 0.21 |
| Bangladesh | WASH−B (36) | 1156 | 3424 | −0.02 (−0.04, 0.00) | 0.28 |
| Burkina Faso | iLiNS−Zinc (37) | 1952 | 664 | −0.04 (−0.07, −0.01) | 0.15 |
| Ghana | GHANA (39) | 98 | 96 | −0.01 (−0.09, 0.07) | 0.02 |
| Ghana | iLiNS−DYADG (40) | 347 | 692 | −0.01 (−0.04, 0.02) | 0.10 |
| Malawi | iLiNS−DYADM (44) | 220 | 444 | 0.00 (−0.03, 0.03) | 0.13 |
| Malawi | iLiNS−DOSE (45) | 696 | 241 | 0.01 (−0.03, 0.04) | 0.10 |
| **I² = 0.13, Tau² = 0.00** | | **7252** | **6778** | **−0.02 (−0.03, −0.01)** | |

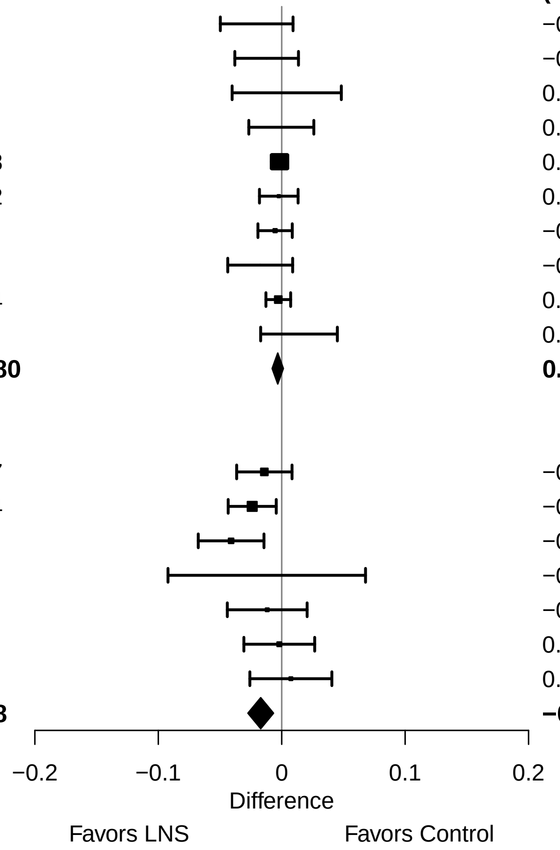

#### Supplemental figure 4F: Wasting prevalence difference

##### 4F8: Stratified by Average SQ-LNS compliance

**Average SQ−LNS compliance**
**(p−diff = 0.204)**

| Country | Trial | N | N | PD (95% CI) | W |
|---|---|---|---|---|---|
| **Average SQ−LNS compliance − Low** | | | | | |
| Burkina Faso | PROMIS (38) | 856 | 907 | −0.01 (−0.04, 0.01) | 0.07 |
| Burkina Faso | PROMIS CS (38) | 430 | 436 | 0.00 (−0.04, 0.05) | 0.02 |
| Ghana | iLiNS−DYADG (40) | 347 | 692 | −0.01 (−0.04, 0.02) | 0.04 |
| Malawi | iLiNS−DYADM (44) | 220 | 444 | 0.00 (−0.03, 0.03) | 0.05 |
| Malawi | iLiNS−DOSE (45) | 696 | 241 | 0.01 (−0.03, 0.04) | 0.04 |
| Mali | PROMIS (46) | 499 | 502 | −0.01 (−0.02, 0.01) | 0.23 |
| Mali | PROMIS CS (46) | 944 | 959 | −0.02 (−0.04, 0.01) | 0.06 |
| Zimbabwe | SHINE (HIV−) (47) | 1869 | 1784 | 0.00 (−0.01, 0.01) | 0.44 |
| Zimbabwe | SHINE (HIV+) (48) | 336 | 328 | 0.01 (−0.02, 0.05) | 0.05 |
| **I² = 0.00, Tau² = 0.00** | | **6197** | **6293** | **0.00 (−0.01, 0.00)** | |
| **Average SQ−LNS compliance − High** | | | | | |
| Bangladesh | JiVitA−4 (34) | 2783 | 1217 | −0.01 (−0.04, 0.01) | 0.06 |
| Bangladesh | RDNS (35) | 1661 | 815 | −0.02 (−0.05, 0.01) | 0.04 |
| Bangladesh | WASH−B (36) | 1156 | 3424 | −0.02 (−0.04, 0.00) | 0.09 |
| Burkina Faso | iLiNS−Zinc (37) | 1952 | 664 | −0.04 (−0.07, −0.01) | 0.05 |
| Ghana | GHANA (39) | 98 | 96 | −0.01 (−0.09, 0.07) | 0.01 |
| Haiti | HAITI (41) | 148 | 149 | 0.00 (−0.03, 0.03) | 0.05 |
| Kenya | WASH−B (42) | 1455 | 5118 | 0.00 (−0.01, 0.00) | 0.72 |
| **I² = 0.54, Tau² = 0.00** | | **9253** | **11483** | **−0.01 (−0.03, 0.00)** | |

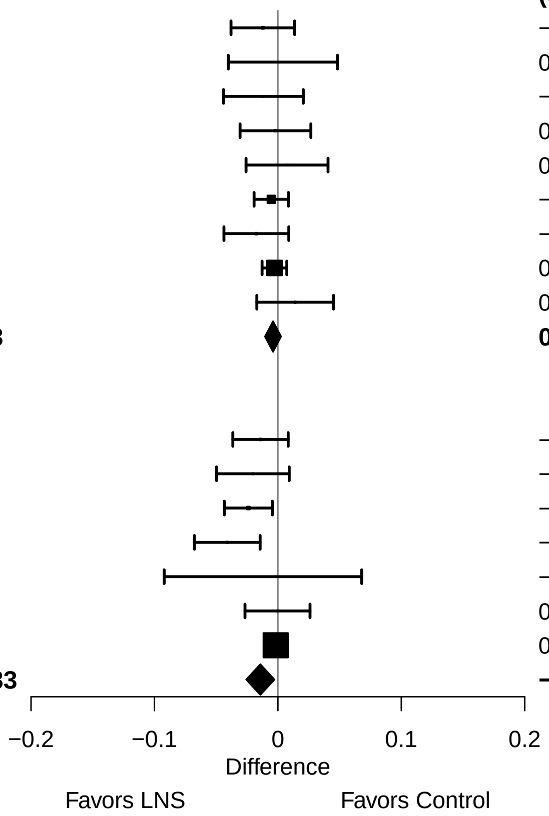

### Supplemental figure 4G: Mean difference in MUACZ

#### 4G1: Stratified by Geographic region (insufficient comparisons)

#### Supplemental figure 4G: Mean difference in MUACZ

#### 4G2: Stratified by Stunting burden

**Stunting burden**
**(p−diff = 0.433)**

| Country | Trial | N | N | MD (95% CI) | W |
|---|---|---|---|---|---|
| **Stunting burden − Less than 35%** | | | | | |
| Burkina Faso | PROMIS (38) | 854 | 912 | 0.09 (−0.03, 0.21) | 0.20 |
| Burkina Faso | PROMIS CS (38) | 424 | 436 | −0.06 (−0.22, 0.10) | 0.15 |
| Ghana | GHANA (39) | | | | |
| Ghana | iLiNS−DYADG (40) | 347 | 693 | 0.09 (−0.03, 0.20) | 0.21 |
| Haiti | HAITI (41) | | | | |
| Kenya | WASH−B (42) | 1461 | 5185 | 0.12 (0.05, 0.18) | 0.29 |
| Malawi | iLiNS−DYADM (44) | 220 | 444 | −0.04 (−0.20, 0.12) | 0.15 |
| **I² = 0.35, Tau² = 0.00** | | **3306** | **7670** | **0.06 (0.00, 0.13)** | |
| **Stunting burden − More than 35%** | | | | | |
| Bangladesh | JiVitA−4 (34) | 2897 | 1271 | 0.08 (0.03, 0.13) | 0.15 |
| Bangladesh | RDNS (35) | 1663 | 815 | 0.04 (−0.04, 0.12) | 0.13 |
| Bangladesh | WASH−B (36) | | | | |
| Burkina Faso | iLiNS−Zinc (37) | 1959 | 666 | 0.30 (0.20, 0.39) | 0.12 |
| Madagascar | MAHAY (43) | 1680 | 1658 | −0.03 (−0.16, 0.09) | 0.09 |
| Malawi | iLiNS−DOSE (45) | 696 | 240 | 0.06 (−0.05, 0.18) | 0.10 |
| Mali | PROMIS (46) | 507 | 505 | 0.19 (0.05, 0.32) | 0.09 |
| Mali | PROMIS CS (46) | 954 | 970 | 0.13 (0.01, 0.25) | 0.10 |
| Zimbabwe | SHINE (HIV−) (47) | 1872 | 1780 | 0.06 (0.00, 0.12) | 0.14 |
| Zimbabwe | SHINE (HIV+) (48) | 337 | 328 | 0.04 (−0.10, 0.17) | 0.09 |
| **I² = 0.72, Tau² = 0.01** | | **12565** | **8233** | **0.10 (0.03, 0.16)** | |

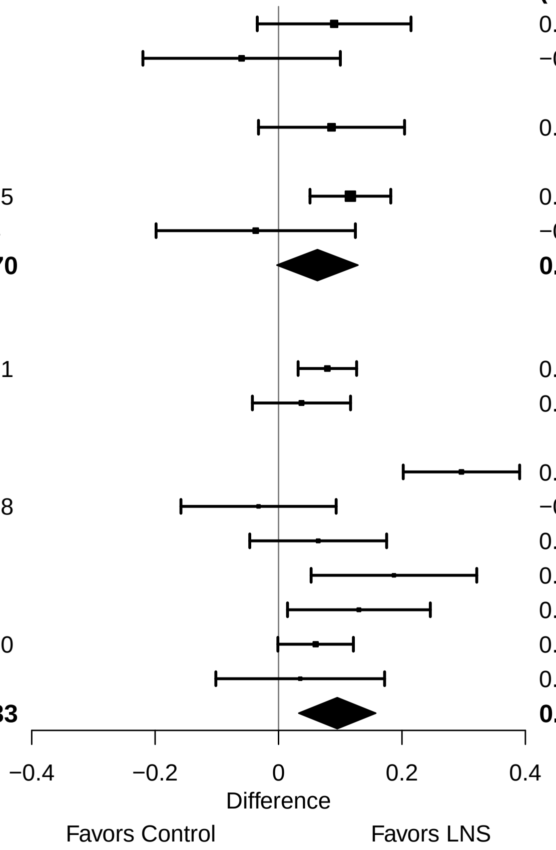

#### Supplemental figure 4G: Mean difference in MUACZ

### 4G3: Stratified by Malaria prevalence

**Malaria prevalence**
**(p−diff = 0.248)**

| Country | Trial | N | N | MD (95% CI) | W |
|---|---|---|---|---|---|
| **Malaria prevalence − Less than 10%** | | | | | |
| Bangladesh | JiVitA−4 (34) | 2897 | 1271 | 0.08 (0.03, 0.13) | 0.21 |
| Bangladesh | RDNS (35) | 1663 | 815 | 0.04 (−0.04, 0.12) | 0.17 |
| Bangladesh | WASH−B (36) | | | | |
| Haiti | HAITI (41) | | | | |
| Kenya | WASH−B (42) | 1461 | 5185 | 0.12 (0.05, 0.18) | 0.19 |
| Madagascar | MAHAY (43) | 1680 | 1658 | −0.03 (−0.16, 0.09) | 0.13 |
| Zimbabwe | SHINE (HIV−) (47) | 1872 | 1780 | 0.06 (0.00, 0.12) | 0.19 |
| Zimbabwe | SHINE (HIV+) (48) | 337 | 328 | 0.04 (−0.10, 0.17) | 0.12 |
| **I² = 0.11, Tau² = 0.00** | | **9910** | **11037** | **0.07 (0.03, 0.10)** | |
| **Malaria prevalence − At least 10%** | | | | | |
| Burkina Faso | iLiNS−Zinc (37) | 1959 | 666 | 0.30 (0.20, 0.39) | 0.16 |
| Burkina Faso | PROMIS (38) | 854 | 912 | 0.09 (−0.03, 0.21) | 0.13 |
| Burkina Faso | PROMIS CS (38) | 424 | 436 | −0.06 (−0.22, 0.10) | 0.10 |
| Ghana | GHANA (39) | | | | |
| Ghana | iLiNS−DYADG (40) | 347 | 693 | 0.09 (−0.03, 0.20) | 0.13 |
| Malawi | iLiNS−DYADM (44) | 220 | 444 | −0.04 (−0.20, 0.12) | 0.10 |
| Malawi | iLiNS−DOSE (45) | 696 | 240 | 0.06 (−0.05, 0.18) | 0.14 |
| Mali | PROMIS (46) | 507 | 505 | 0.19 (0.05, 0.32) | 0.12 |
| Mali | PROMIS CS (46) | 954 | 970 | 0.13 (0.01, 0.25) | 0.13 |
| **I² = 0.71, Tau² = 0.01** | | **5961** | **4866** | **0.10 (0.02, 0.18)** | |

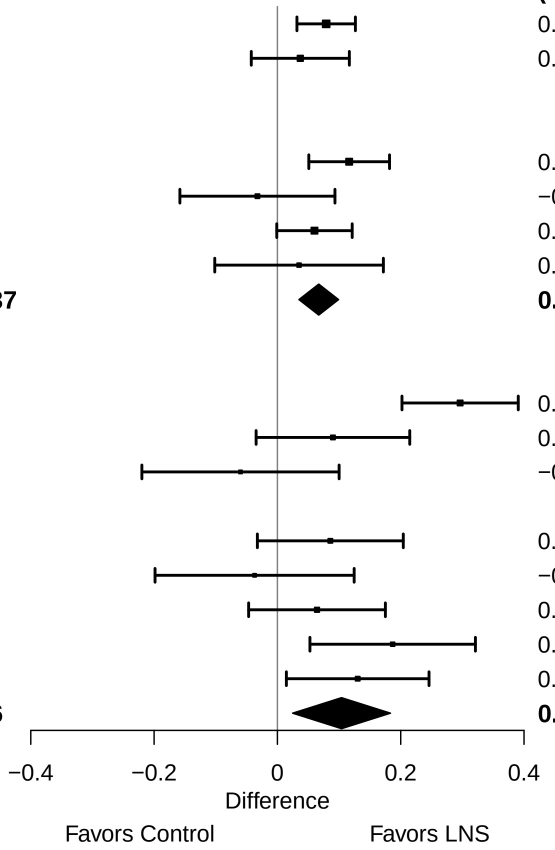

##### Supplemental figure 4G: Mean difference in MUACZ

#### 4G4: Stratified by Source water quality

**Source water quality**
**(p−diff = 0.438)**

**Source water quality − Improved**

| **Country** | **Trial** | **N** | **N** | **MD (95% CI)** | **W** |
|---|---|---|---|---|---|
| Bangladesh | JiVitA−4 (34) | 2897 | 1271 | 0.08 (0.03, 0.13) | 0.27 |
| Bangladesh | RDNS (35) | 1663 | 815 | 0.04 (−0.04, 0.12) | 0.23 |
| Bangladesh | WASH−B (36) | | | | |
| Ghana | GHANA (39) | | | | |
| Ghana | iLiNS−DYADG (40) | 347 | 693 | 0.09 (−0.03, 0.20) | 0.18 |
| Haiti | HAITI (41) | | | | |
| Malawi | iLiNS−DYADM (44) | 220 | 444 | −0.04 (−0.20, 0.12) | 0.13 |
| Malawi | iLiNS−DOSE (45) | 696 | 240 | 0.06 (−0.05, 0.18) | 0.19 |
| **I² = 0.00, Tau² = 0.00** | | **5823** | **3463** | **0.06 (0.03, 0.10)** | |

**Source water quality − Unimproved**

| **Country** | **Trial** | **N** | **N** | **MD (95% CI)** | **W** |
|---|---|---|---|---|---|
| Burkina Faso | iLiNS−Zinc (37) | 1959 | 666 | 0.30 (0.20, 0.39) | 0.14 |
| Burkina Faso | PROMIS (38) | 854 | 912 | 0.09 (−0.03, 0.21) | 0.11 |
| Burkina Faso | PROMIS CS (38) | 424 | 436 | −0.06 (−0.22, 0.10) | 0.09 |
| Kenya | WASH−B (42) | 697 | 2270 | 0.10 (0.01, 0.19) | 0.14 |
| Madagascar | MAHAY (43) | 1680 | 1658 | −0.03 (−0.16, 0.09) | 0.11 |
| Mali | PROMIS (46) | 507 | 505 | 0.19 (0.05, 0.32) | 0.10 |
| Mali | PROMIS CS (46) | 954 | 970 | 0.13 (0.01, 0.25) | 0.12 |
| Zimbabwe | SHINE (HIV−) (47) | 889 | 871 | 0.02 (−0.07, 0.11) | 0.14 |
| Zimbabwe | SHINE (HIV+) (48) | 147 | 146 | 0.00 (−0.23, 0.24) | 0.05 |
| **I² = 0.74, Tau² = 0.01** | | **8111** | **8434** | **0.09 (0.02, 0.17)** | |

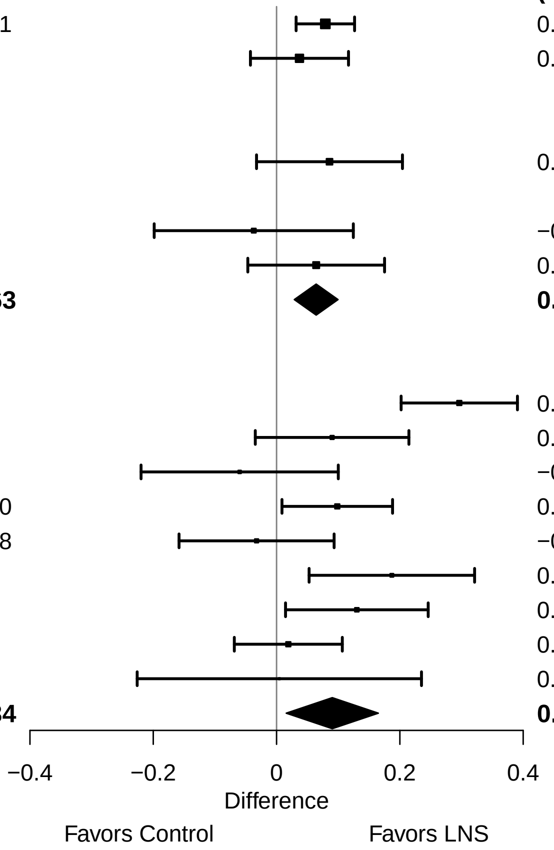

#### Supplemental figure 4G: Mean difference in MUACZ

### 4G5: Stratified by Sanitation

**Sanitation**
**(p−diff = 0.887)**

| **Sanitation − Improved** | | | | **MD** | |
|---|---|---|---|---|---|
| **Country** | **Trial** | **N** | **N** | **(95% CI)** | **W** |
| Bangladesh | JiVitA−4 (34) | 2897 | 1271 | 0.08 (0.03, 0.13) | 0.21 |
| Bangladesh | RDNS (35) | 1663 | 815 | 0.04 (−0.04, 0.12) | 0.17 |
| Bangladesh | WASH−B (36) | | | | |
| Burkina Faso | PROMIS (38) | 854 | 912 | 0.09 (−0.03, 0.21) | 0.13 |
| Burkina Faso | PROMIS CS (38) | 424 | 436 | −0.06 (−0.22, 0.10) | 0.10 |
| Ghana | GHANA (39) | | | | |
| Ghana | iLiNS−DYADG (40) | 347 | 693 | 0.09 (−0.03, 0.20) | 0.13 |
| Haiti | HAITI (41) | | | | |
| Mali | PROMIS (46) | 507 | 505 | 0.19 (0.05, 0.32) | 0.12 |
| Mali | PROMIS CS (46) | 954 | 970 | 0.13 (0.01, 0.25) | 0.14 |
| **I² = 0.17, Tau² = 0.00** | | **7646** | **5602** | **0.08 (0.04, 0.12)** | |
| **Sanitation − Unimproved** | | | | | |
| Burkina Faso | iLiNS−Zinc (37) | 1959 | 666 | 0.30 (0.20, 0.39) | 0.17 |
| Kenya | WASH−B (42) | 697 | 2270 | 0.10 (0.01, 0.19) | 0.18 |
| Madagascar | MAHAY (43) | 1680 | 1658 | −0.03 (−0.16, 0.09) | 0.14 |
| Malawi | iLiNS−DYADM (44) | 220 | 444 | −0.04 (−0.20, 0.12) | 0.11 |
| Malawi | iLiNS−DOSE (45) | 696 | 240 | 0.06 (−0.05, 0.18) | 0.15 |
| Zimbabwe | SHINE (HIV−) (47) | 889 | 871 | 0.02 (−0.07, 0.11) | 0.18 |
| Zimbabwe | SHINE (HIV+) (48) | 147 | 146 | 0.00 (−0.23, 0.24) | 0.07 |
| **I² = 0.78, Tau² = 0.01** | | **6288** | **6295** | **0.07 (−0.02, 0.16)** | |

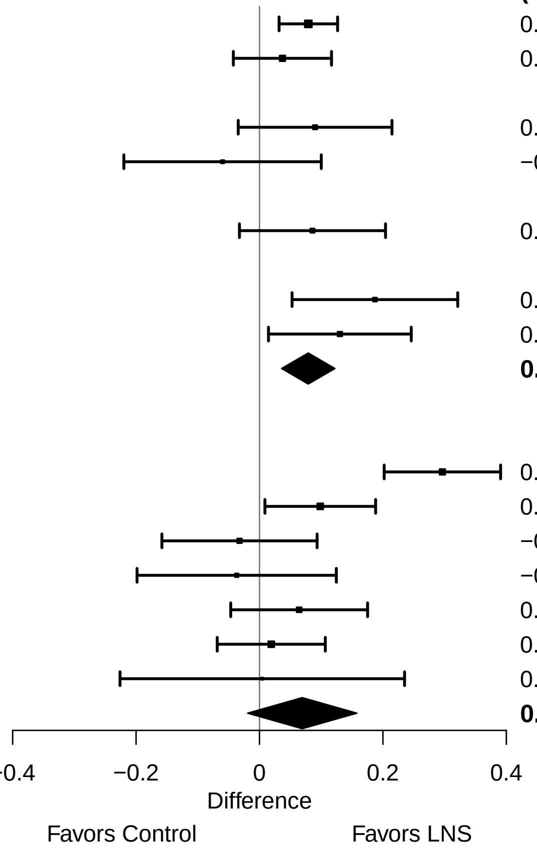

#### Supplemental figure 4G: Mean difference in MUACZ

### 4G6: Stratified by Supplement duration

**Supplement duration**
**(p−diff = 0.427)**

| Country | Trial | N | N | MD (95% CI) | W |
|---|---|---|---|---|---|
| **Supplement duration − 12m or less** | | | | | |
| Bangladesh | JiVitA−4 (34) | 2897 | 1271 | 0.08 (0.03, 0.13) | 0.15 |
| Burkina Faso | iLiNS−Zinc (37) | 1959 | 666 | 0.30 (0.20, 0.39) | 0.11 |
| Burkina Faso | PROMIS (38) | 854 | 912 | 0.09 (−0.03, 0.21) | 0.09 |
| Burkina Faso | PROMIS CS (38) | 424 | 436 | −0.06 (−0.22, 0.10) | 0.07 |
| Ghana | GHANA (39) | | | | |
| Ghana | iLiNS−DYADG (40) | 347 | 693 | 0.09 (−0.03, 0.20) | 0.10 |
| Haiti | HAITI (41) | | | | |
| Madagascar | MAHAY (43) | 1680 | 1658 | −0.03 (−0.16, 0.09) | 0.09 |
| Malawi | iLiNS−DYADM (44) | 220 | 444 | −0.04 (−0.20, 0.12) | 0.07 |
| Malawi | iLiNS−DOSE (45) | 696 | 240 | 0.06 (−0.05, 0.18) | 0.10 |
| Zimbabwe | SHINE (HIV−) (47) | 1872 | 1780 | 0.06 (0.00, 0.12) | 0.14 |
| Zimbabwe | SHINE (HIV+) (48) | 337 | 328 | 0.04 (−0.10, 0.17) | 0.08 |
| **I² = 0.69, Tau² = 0.01** | | **11286** | **8428** | **0.07 (0.01, 0.13)** | |
| **Supplement duration − > 12m** | | | | | |
| Bangladesh | RDNS (35) | 1663 | 815 | 0.04 (−0.04, 0.12) | 0.28 |
| Bangladesh | WASH−B (36) | | | | |
| Kenya | WASH−B (42) | 1461 | 5185 | 0.12 (0.05, 0.18) | 0.31 |
| Mali | PROMIS (46) | 507 | 505 | 0.19 (0.05, 0.32) | 0.19 |
| Mali | PROMIS CS (46) | 954 | 970 | 0.13 (0.01, 0.25) | 0.22 |
| **I² = 0.33, Tau² = 0.00** | | **4585** | **7475** | **0.11 (0.05, 0.16)** | |

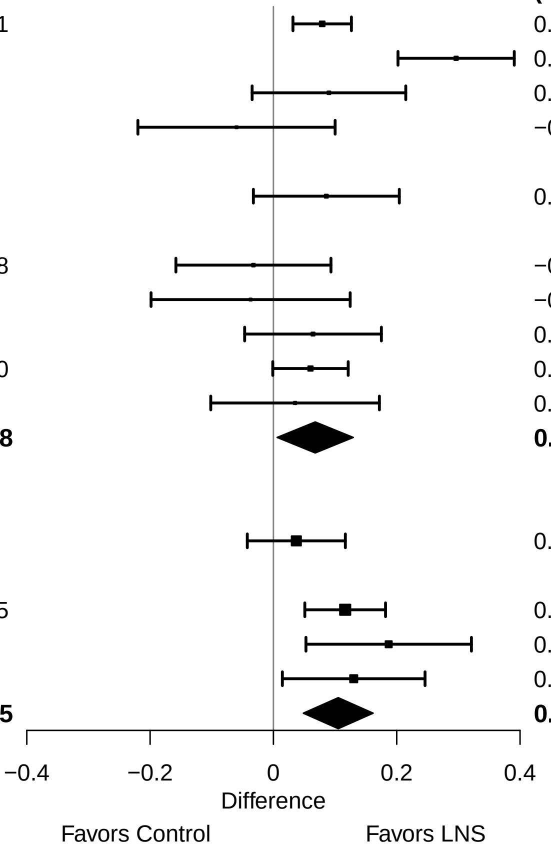

#### Supplemental figure 4G: Mean difference in MUACZ

#### 4G7: Stratified by Frequency of contact

**Frequency of contact**
**(p−diff = 0.388)**

| Country | Trial | N | N | MD (95% CI) | W |
|---|---|---|---|---|---|
| **Frequency of contact − Monthly** | | | | | |
| Bangladesh | RDNS (35) | 1663 | 815 | 0.04 (−0.04, 0.12) | 0.14 |
| Burkina Faso | PROMIS (38) | 854 | 912 | 0.09 (−0.03, 0.21) | 0.10 |
| Burkina Faso | PROMIS CS (38) | 424 | 436 | −0.06 (−0.22, 0.10) | 0.08 |
| Haiti | HAITI (41) | | | | |
| Kenya | WASH−B (42) | 1461 | 5185 | 0.12 (0.05, 0.18) | 0.15 |
| Madagascar | MAHAY (43) | 1680 | 1658 | −0.03 (−0.16, 0.09) | 0.10 |
| Mali | PROMIS (46) | 507 | 505 | 0.19 (0.05, 0.32) | 0.09 |
| Mali | PROMIS CS (46) | 954 | 970 | 0.13 (0.01, 0.25) | 0.11 |
| Zimbabwe | SHINE (HIV−) (47) | 1872 | 1780 | 0.06 (0.00, 0.12) | 0.15 |
| Zimbabwe | SHINE (HIV+) (48) | 337 | 328 | 0.04 (−0.10, 0.17) | 0.09 |
| **I² = 0.34, Tau² = 0.00** | | **9752** | **12589** | **0.07 (0.03, 0.11)** | |
| **Frequency of contact − Weekly** | | | | | |
| Bangladesh | JiVitA−4 (34) | 2897 | 1271 | 0.08 (0.03, 0.13) | 0.28 |
| Bangladesh | WASH−B (36) | | | | |
| Burkina Faso | iLiNS−Zinc (37) | 1959 | 666 | 0.30 (0.20, 0.39) | 0.21 |
| Ghana | GHANA (39) | | | | |
| Ghana | iLiNS−DYADG (40) | 347 | 693 | 0.09 (−0.03, 0.20) | 0.18 |
| Malawi | iLiNS−DYADM (44) | 220 | 444 | −0.04 (−0.20, 0.12) | 0.13 |
| Malawi | iLiNS−DOSE (45) | 696 | 240 | 0.06 (−0.05, 0.18) | 0.19 |
| **I² = 0.81, Tau² = 0.01** | | **6119** | **3314** | **0.10 (0.00, 0.21)** | |

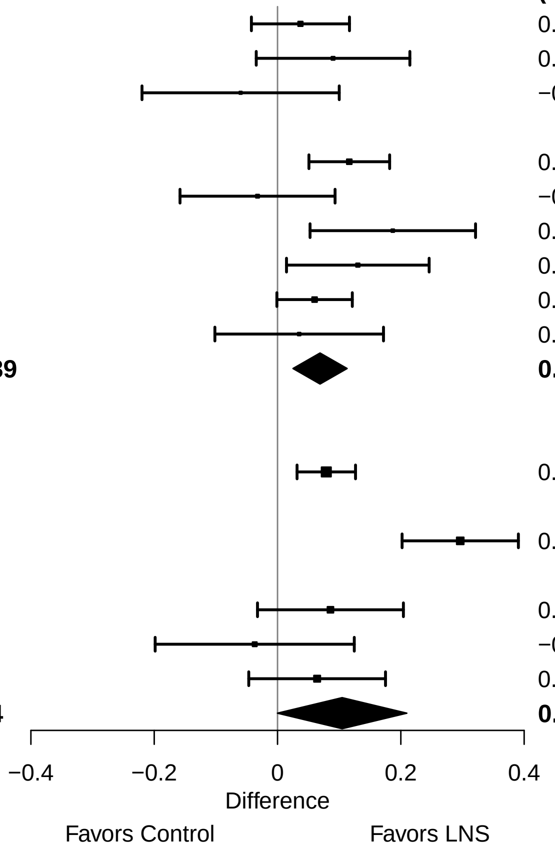

#### Supplemental figure 4G: Mean difference in MUACZ

### 4G8: Stratified by Average SQ-LNS compliance

**Average SQ−LNS compliance**
**(p−diff = 0.228)**

**Average SQ−LNS compliance − Low**

| Country | Trial | N | N | MD (95% CI) | W |
|---|---|---|---|---|---|
| Burkina Faso | PROMIS (38) | 854 | 912 | 0.09 (−0.03, 0.21) | 0.11 |
| Burkina Faso | PROMIS CS (38) | 424 | 436 | −0.06 (−0.22, 0.10) | 0.08 |
| Ghana | iLiNS−DYADG (40) | 347 | 693 | 0.09 (−0.03, 0.20) | 0.11 |
| Malawi | iLiNS−DYADM (44) | 220 | 444 | −0.04 (−0.20, 0.12) | 0.08 |
| Malawi | iLiNS−DOSE (45) | 696 | 240 | 0.06 (−0.05, 0.18) | 0.12 |
| Mali | PROMIS (46) | 507 | 505 | 0.19 (0.05, 0.32) | 0.10 |
| Mali | PROMIS CS (46) | 954 | 970 | 0.13 (0.01, 0.25) | 0.12 |
| Zimbabwe | SHINE (HIV−) (47) | 1872 | 1780 | 0.06 (0.00, 0.12) | 0.17 |
| Zimbabwe | SHINE (HIV+) (48) | 337 | 328 | 0.04 (−0.10, 0.17) | 0.10 |
| **I² = 0.08, Tau² = 0.00** | | **6211** | **6308** | **0.07 (0.03, 0.11)** | |

**Average SQ−LNS compliance − High**

| Country | Trial | N | N | MD (95% CI) | W |
|---|---|---|---|---|---|
| Bangladesh | JiVitA−4 (34) | 2897 | 1271 | 0.08 (0.03, 0.13) | 0.29 |
| Bangladesh | RDNS (35) | 1663 | 815 | 0.04 (−0.04, 0.12) | 0.24 |
| Bangladesh | WASH−B (36) | | | | |
| Burkina Faso | iLiNS−Zinc (37) | 1959 | 666 | 0.30 (0.20, 0.39) | 0.21 |
| Ghana | GHANA (39) | | | | |
| Haiti | HAITI (41) | | | | |
| Kenya | WASH−B (42) | 1461 | 5185 | 0.12 (0.05, 0.18) | 0.26 |
| **I² = 0.85, Tau² = 0.01** | | **7980** | **7937** | **0.13 (0.02, 0.24)** | |

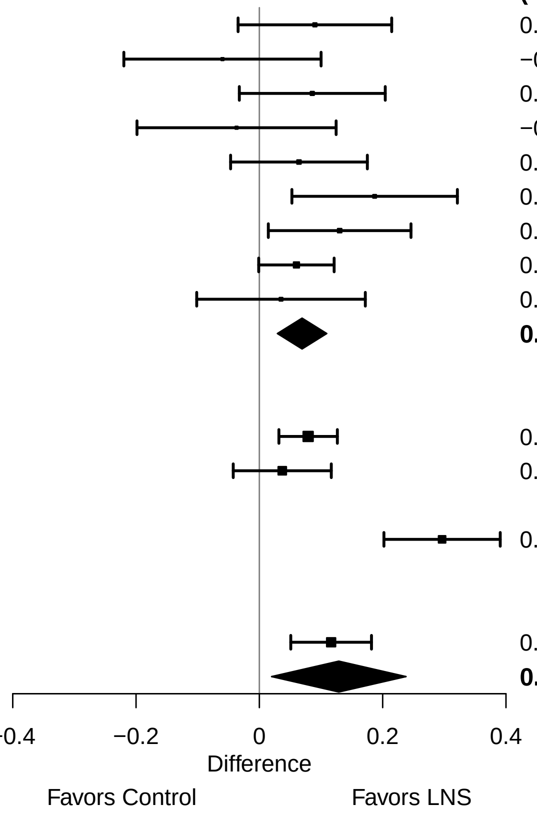

### Supplemental figure 4H: Low MUAC prevalence ratio

#### 4H1: Stratified by Geographic region (insufficient comparisons)

#### Supplemental figure 4H: Low MUAC prevalence ratio

### 4H2: Stratified by Stunting burden

**Stunting burden**
**(p−diff = 0.925)**

| Country | Trial | N | N | PR (95% CI) | W |
|---|---|---|---|---|---|
| **Stunting burden − Less than 35%** | | | | | |
| Burkina Faso | PROMIS (38) | 854 | 912 | 0.94 (0.63, 1.40) | 0.21 |
| Burkina Faso | PROMIS CS (38) | 424 | 436 | 0.92 (0.66, 1.28) | 0.29 |
| Ghana | GHANA (39) | | | | |
| Ghana | iLiNS−DYADG (40) | 347 | 693 | 0.77 (0.37, 1.57) | 0.07 |
| Haiti | HAITI (41) | | | | |
| Kenya | WASH−B (42) | 1461 | 5185 | 0.71 (0.54, 0.94) | 0.39 |
| Malawi | iLiNS−DYADM (44) | 220 | 444 | 0.94 (0.39, 2.28) | 0.05 |
| **I² = 0.00, Tau² = 0.00** | | **3306** | **7670** | **0.82 (0.69, 0.98)** | |
| **Stunting burden − More than 35%** | | | | | |
| Bangladesh | JiVitA−4 (34) | 2897 | 1271 | 0.85 (0.70, 1.03) | 0.26 |
| Bangladesh | RDNS (35) | 1663 | 815 | 1.02 (0.69, 1.49) | 0.09 |
| Bangladesh | WASH−B (36) | | | | |
| Burkina Faso | iLiNS−Zinc (37) | 1959 | 666 | 0.64 (0.51, 0.79) | 0.23 |
| Madagascar | MAHAY (43) | 1680 | 1658 | 0.97 (0.76, 1.26) | 0.18 |
| Malawi | iLiNS−DOSE (45) | 696 | 240 | 1.35 (0.54, 3.36) | 0.02 |
| Mali | PROMIS (46) | 507 | 505 | 0.58 (0.31, 1.07) | 0.04 |
| Mali | PROMIS CS (46) | 954 | 970 | 0.90 (0.68, 1.19) | 0.15 |
| Zimbabwe | SHINE (HIV−) (47) | 1872 | 1780 | 0.51 (0.27, 0.97) | 0.03 |
| Zimbabwe | SHINE (HIV+) (48) | 337 | 328 | 0.97 (0.29, 3.28) | 0.01 |
| **I² = 0.41, Tau² = 0.02** | | **12565** | **8233** | **0.82 (0.70, 0.96)** | |

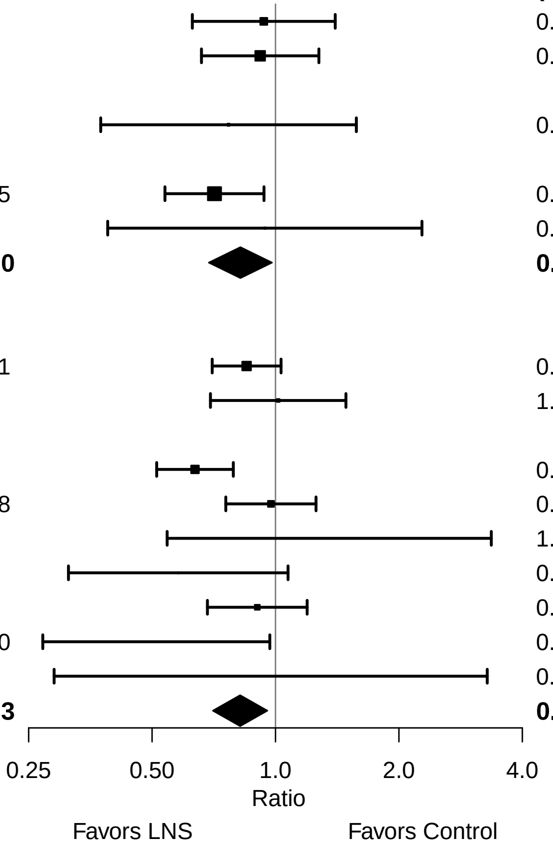

##### Supplemental figure 4H: Low MUAC prevalence ratio

#### 4H3: Stratified by Malaria prevalence

**Malaria prevalence**
**(p−diff = 0.684)**

| Country | Trial | N | N | PR (95% CI) | W |
|---|---|---|---|---|---|
| **Malaria prevalence − Less than 10%** | | | | | |
| Bangladesh | JiVitA−4 (34) | 2897 | 1271 | 0.85 (0.70, 1.03) | 0.36 |
| Bangladesh | RDNS (35) | 1663 | 815 | 1.02 (0.69, 1.49) | 0.12 |
| Bangladesh | WASH−B (36) | | | | |
| Haiti | HAITI (41) | | | | |
| Kenya | WASH−B (42) | 1461 | 5185 | 0.71 (0.54, 0.94) | 0.21 |
| Madagascar | MAHAY (43) | 1680 | 1658 | 0.97 (0.76, 1.26) | 0.24 |
| Zimbabwe | SHINE (HIV−) (47) | 1872 | 1780 | 0.51 (0.27, 0.97) | 0.05 |
| Zimbabwe | SHINE (HIV+) (48) | 337 | 328 | 0.97 (0.29, 3.28) | 0.01 |
| **I² = 0.17, Tau² = 0.01** | | **9910** | **11037** | **0.84 (0.73, 0.98)** | |
| **Malaria prevalence − At least 10%** | | | | | |
| Burkina Faso | iLiNS−Zinc (37) | 1959 | 666 | 0.64 (0.51, 0.79) | 0.34 |
| Burkina Faso | PROMIS (38) | 854 | 912 | 0.94 (0.63, 1.40) | 0.12 |
| Burkina Faso | PROMIS CS (38) | 424 | 436 | 0.92 (0.66, 1.28) | 0.17 |
| Ghana | GHANA (39) | | | | |
| Ghana | iLiNS−DYADG (40) | 347 | 693 | 0.77 (0.37, 1.57) | 0.04 |
| Malawi | iLiNS−DYADM (44) | 220 | 444 | 0.94 (0.39, 2.28) | 0.03 |
| Malawi | iLiNS−DOSE (45) | 696 | 240 | 1.35 (0.54, 3.36) | 0.03 |
| Mali | PROMIS (46) | 507 | 505 | 0.58 (0.31, 1.07) | 0.05 |
| Mali | PROMIS CS (46) | 954 | 970 | 0.90 (0.68, 1.19) | 0.22 |
| **I² = 0.19, Tau² = 0.01** | | **5961** | **4866** | **0.79 (0.68, 0.92)** | |

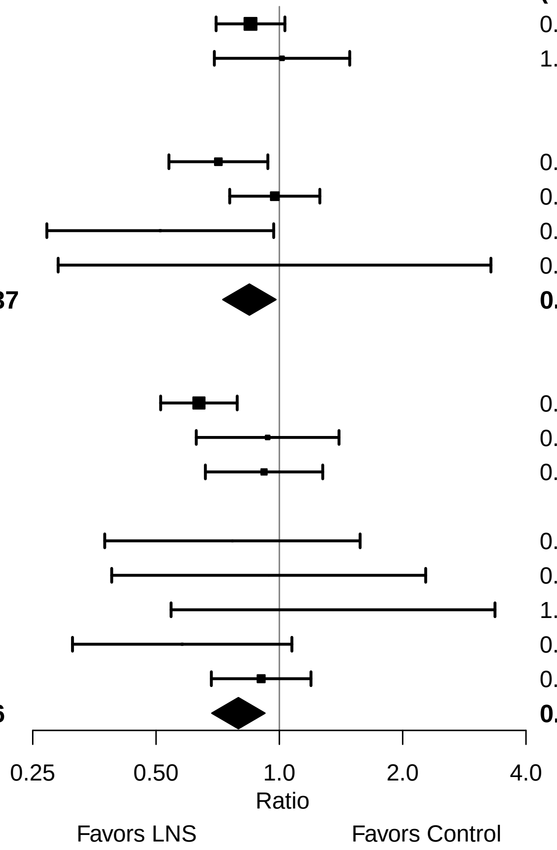

#### Supplemental figure 4H: Low MUAC prevalence ratio

#### 4H4: Stratified by Source water quality

**Source water quality**
**(p−diff = 0.367)**

| Country | Trial | N | N | PR (95% CI) | W |
|---|---|---|---|---|---|
| **Source water quality − Improved** | | | | | |
| Bangladesh | JiVitA−4 (34) | 2897 | 1271 | 0.85 (0.70, 1.03) | 0.69 |
| Bangladesh | RDNS (35) | 1663 | 815 | 1.02 (0.69, 1.49) | 0.19 |
| Bangladesh | WASH−B (36) | | | | |
| Ghana | GHANA (39) | | | | |
| Ghana | iLiNS−DYADG (40) | 347 | 693 | 0.77 (0.37, 1.57) | 0.05 |
| Haiti | HAITI (41) | | | | |
| Malawi | iLiNS−DYADM (44) | 220 | 444 | 0.94 (0.39, 2.28) | 0.03 |
| Malawi | iLiNS−DOSE (45) | 696 | 240 | 1.35 (0.54, 3.36) | 0.03 |
| **I² = 0.00, Tau² = 0.00** | | **5823** | **3463** | **0.89 (0.76, 1.05)** | |
| **Source water quality − Unimproved** | | | | | |
| Burkina Faso | iLiNS−Zinc (37) | 1959 | 666 | 0.64 (0.51, 0.79) | 0.29 |
| Burkina Faso | PROMIS (38) | 854 | 912 | 0.94 (0.63, 1.40) | 0.09 |
| Burkina Faso | PROMIS CS (38) | 424 | 436 | 0.92 (0.66, 1.28) | 0.13 |
| Kenya | WASH−B (42) | 697 | 2270 | 0.67 (0.41, 1.10) | 0.06 |
| Madagascar | MAHAY (43) | 1680 | 1658 | 0.97 (0.76, 1.26) | 0.21 |
| Mali | PROMIS (46) | 507 | 505 | 0.58 (0.31, 1.07) | 0.04 |
| Mali | PROMIS CS (46) | 954 | 970 | 0.90 (0.68, 1.19) | 0.17 |
| Zimbabwe | SHINE (HIV−) (47) | 889 | 871 | 0.65 (0.25, 1.68) | 0.02 |
| Zimbabwe | SHINE (HIV+) (48) | 147 | 146 | 0.50 (0.05, 5.43) | 0.00 |
| **I² = 0.24, Tau² = 0.01** | | **8111** | **8434** | **0.80 (0.70, 0.92)** | |

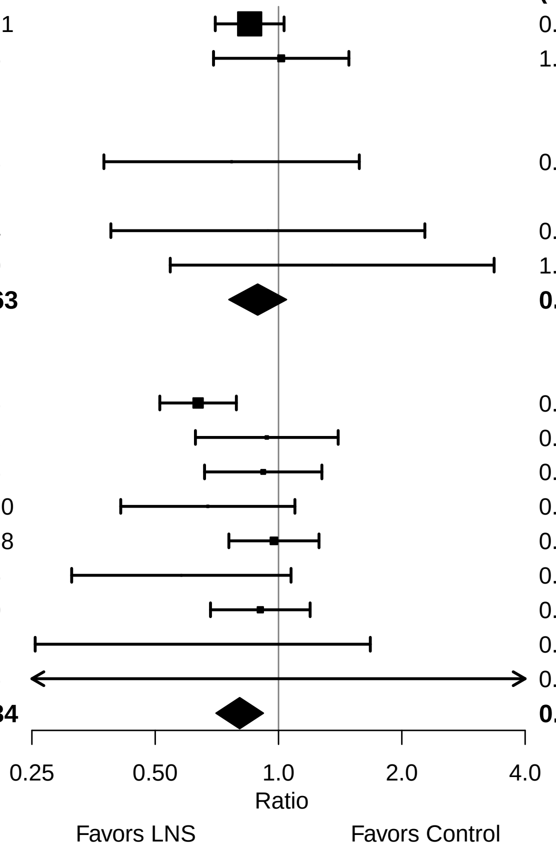

#### Supplemental figure 4H: Low MUAC prevalence ratio

### 4H5: Stratified by Sanitation

**Sanitation**
**(p−diff = 0.291)**

| **Sanitation − Improved** Country | Trial | N | N | PR (95% CI) | W |
|---|---|---|---|---|---|
| Bangladesh | JiVitA−4 (34) | 2897 | 1271 | 0.85 (0.70, 1.03) | 0.40 |
| Bangladesh | RDNS (35) | 1663 | 815 | 1.02 (0.69, 1.49) | 0.11 |
| Bangladesh | WASH−B (36) | | | | |
| Burkina Faso | PROMIS (38) | 854 | 912 | 0.94 (0.63, 1.40) | 0.10 |
| Burkina Faso | PROMIS CS (38) | 424 | 436 | 0.92 (0.66, 1.28) | 0.14 |
| Ghana | GHANA (39) | | | | |
| Ghana | iLiNS−DYADG (40) | 347 | 693 | 0.77 (0.37, 1.57) | 0.03 |
| Haiti | HAITI (41) | | | | |
| Mali | PROMIS (46) | 507 | 505 | 0.58 (0.31, 1.07) | 0.04 |
| Mali | PROMIS CS (46) | 954 | 970 | 0.90 (0.68, 1.19) | 0.19 |
| **I² = 0.00, Tau² = 0.00** | | **7646** | **5602** | **0.88 (0.78, 0.99)** | |
| **Sanitation − Unimproved** | | | | | |
| Burkina Faso | iLiNS−Zinc (37) | 1959 | 666 | 0.64 (0.51, 0.79) | 0.47 |
| Kenya | WASH−B (42) | 697 | 2270 | 0.67 (0.41, 1.10) | 0.09 |
| Madagascar | MAHAY (43) | 1680 | 1658 | 0.97 (0.76, 1.26) | 0.35 |
| Malawi | iLiNS−DYADM (44) | 220 | 444 | 0.94 (0.39, 2.28) | 0.03 |
| Malawi | iLiNS−DOSE (45) | 696 | 240 | 1.35 (0.54, 3.36) | 0.03 |
| Zimbabwe | SHINE (HIV−) (47) | 889 | 871 | 0.65 (0.25, 1.68) | 0.03 |
| Zimbabwe | SHINE (HIV+) (48) | 147 | 146 | 0.50 (0.05, 5.43) | 0.00 |
| **I² = 0.30, Tau² = 0.01** | | **6288** | **6295** | **0.77 (0.65, 0.93)** | |

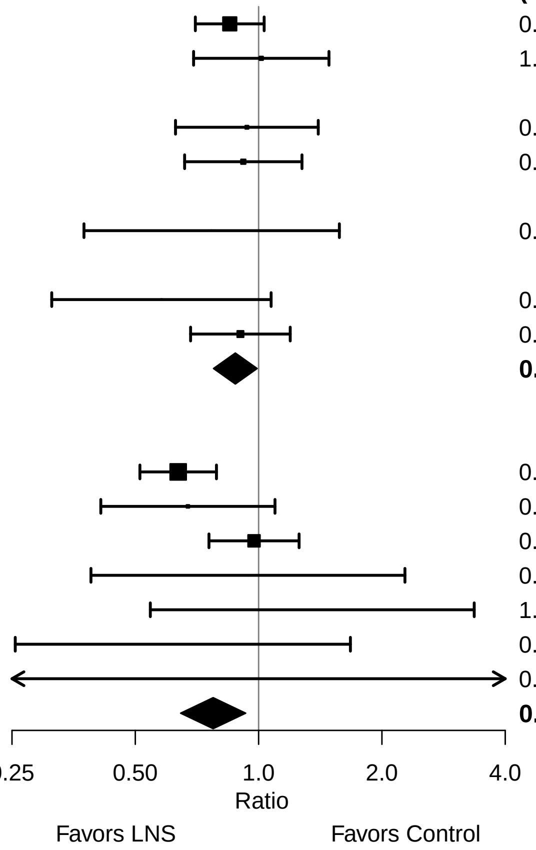

#### Supplemental figure 4H: Low MUAC prevalence ratio

### 4H6: Stratified by Supplement duration

**Supplement duration**
**(p−diff = 0.928)**

| Country | Trial | N | N | PR (95% CI) | W |
|---|---|---|---|---|---|
| **Supplement duration − 12m or less** | | | | | |
| Bangladesh | JiVitA−4 (34) | 2897 | 1271 | 0.85 (0.70, 1.03) | 0.27 |
| Burkina Faso | iLiNS−Zinc (37) | 1959 | 666 | 0.64 (0.51, 0.79) | 0.23 |
| Burkina Faso | PROMIS (38) | 854 | 912 | 0.94 (0.63, 1.40) | 0.08 |
| Burkina Faso | PROMIS CS (38) | 424 | 436 | 0.92 (0.66, 1.28) | 0.12 |
| Ghana | GHANA (39) | | | | |
| Ghana | iLiNS−DYADG (40) | 347 | 693 | 0.77 (0.37, 1.57) | 0.03 |
| Haiti | HAITI (41) | | | | |
| Madagascar | MAHAY (43) | 1680 | 1658 | 0.97 (0.76, 1.26) | 0.18 |
| Malawi | iLiNS−DYADM (44) | 220 | 444 | 0.94 (0.39, 2.28) | 0.02 |
| Malawi | iLiNS−DOSE (45) | 696 | 240 | 1.35 (0.54, 3.36) | 0.02 |
| Zimbabwe | SHINE (HIV−) (47) | 1872 | 1780 | 0.51 (0.27, 0.97) | 0.03 |
| Zimbabwe | SHINE (HIV+) (48) | 337 | 328 | 0.97 (0.29, 3.28) | 0.01 |
| **$I^2$ = 0.22, $Tau^2$ = 0.01** | | **11286** | **8428** | **0.82 (0.72, 0.93)** | |
| **Supplement duration − > 12m** | | | | | |
| Bangladesh | RDNS (35) | 1663 | 815 | 1.02 (0.69, 1.49) | 0.21 |
| Bangladesh | WASH−B (36) | | | | |
| Kenya | WASH−B (42) | 1461 | 5185 | 0.71 (0.54, 0.94) | 0.36 |
| Mali | PROMIS (46) | 507 | 505 | 0.58 (0.31, 1.07) | 0.08 |
| Mali | PROMIS CS (46) | 954 | 970 | 0.90 (0.68, 1.19) | 0.35 |
| **$I^2$ = 0.23, $Tau^2$ = 0.01** | | **4585** | **7475** | **0.82 (0.67, 1.00)** | |

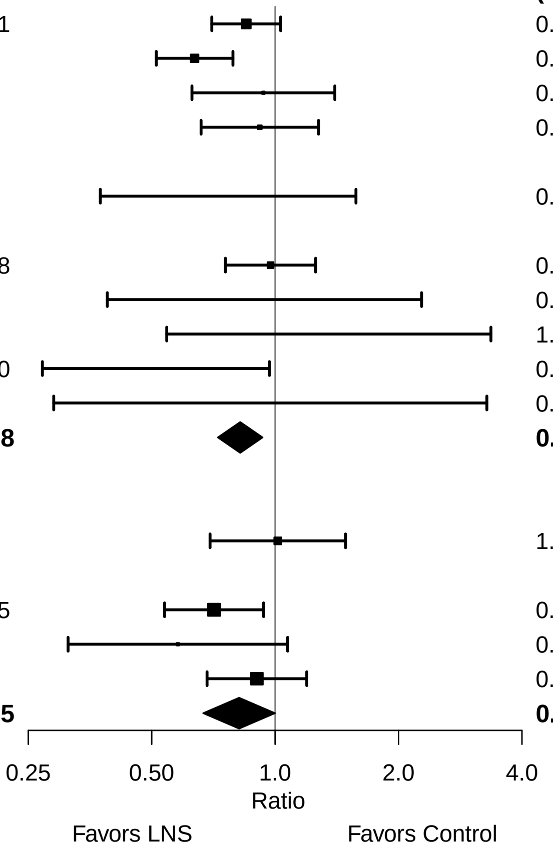

#### Supplemental figure 4H: Low MUAC prevalence ratio

### 4H7: Stratified by Frequency of contact

**Frequency of contact**
**(p−diff = 0.354)**

| Country | Trial | N | N | PR (95% CI) | W |
|---|---|---|---|---|---|
| **Frequency of contact − Monthly** | | | | | |
| Bangladesh | RDNS (35) | 1663 | 815 | 1.02 (0.69, 1.49) | 0.11 |
| Burkina Faso | PROMIS (38) | 854 | 912 | 0.94 (0.63, 1.40) | 0.10 |
| Burkina Faso | PROMIS CS (38) | 424 | 436 | 0.92 (0.66, 1.28) | 0.14 |
| Haiti | HAITI (41) | | | | |
| Kenya | WASH−B (42) | 1461 | 5185 | 0.71 (0.54, 0.94) | 0.18 |
| Madagascar | MAHAY (43) | 1680 | 1658 | 0.97 (0.76, 1.26) | 0.21 |
| Mali | PROMIS (46) | 507 | 505 | 0.58 (0.31, 1.07) | 0.04 |
| Mali | PROMIS CS (46) | 954 | 970 | 0.90 (0.68, 1.19) | 0.18 |
| Zimbabwe | SHINE (HIV−) (47) | 1872 | 1780 | 0.51 (0.27, 0.97) | 0.04 |
| Zimbabwe | SHINE (HIV+) (48) | 337 | 328 | 0.97 (0.29, 3.28) | 0.01 |
| **I² = 0.01, Tau² = 0.00** | | **9752** | **12589** | **0.86 (0.76, 0.97)** | |
| **Frequency of contact − Weekly** | | | | | |
| Bangladesh | JiVitA−4 (34) | 2897 | 1271 | 0.85 (0.70, 1.03) | 0.48 |
| Bangladesh | WASH−B (36) | | | | |
| Burkina Faso | iLiNS−Zinc (37) | 1959 | 666 | 0.64 (0.51, 0.79) | 0.41 |
| Ghana | GHANA (39) | | | | |
| Ghana | iLiNS−DYADG (40) | 347 | 693 | 0.77 (0.37, 1.57) | 0.05 |
| Malawi | iLiNS−DYADM (44) | 220 | 444 | 0.94 (0.39, 2.28) | 0.03 |
| Malawi | iLiNS−DOSE (45) | 696 | 240 | 1.35 (0.54, 3.36) | 0.03 |
| **I² = 0.30, Tau² = 0.01** | | **6119** | **3314** | **0.77 (0.64, 0.91)** | |

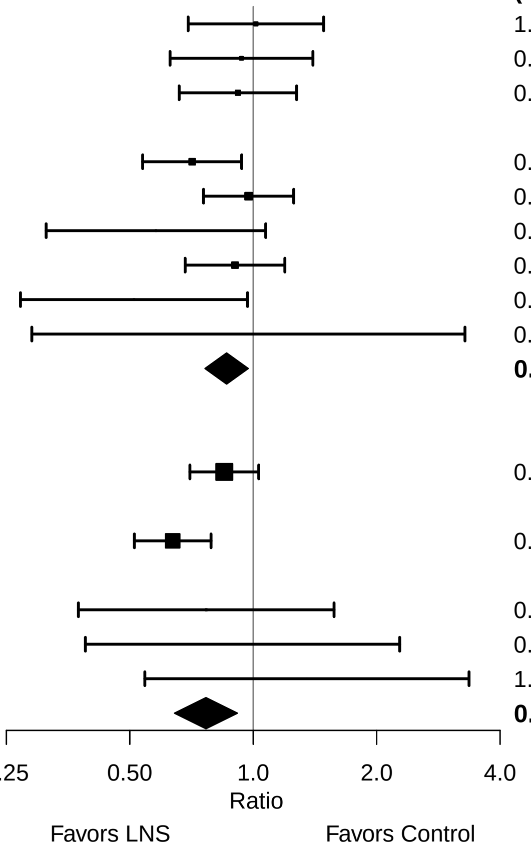

##### Supplemental figure 4H: Low MUAC prevalence ratio

#### 4H8: Stratified by Average SQ-LNS compliance

**Average SQ−LNS compliance**
**(p−diff = 0.377)**

| Country | Trial | N | N | PR (95% CI) | W |
|---|---|---|---|---|---|
| **Average SQ−LNS compliance − Low** | | | | | |
| Burkina Faso | PROMIS (38) | 854 | 912 | 0.94 (0.63, 1.40) | 0.16 |
| Burkina Faso | PROMIS CS (38) | 424 | 436 | 0.92 (0.66, 1.28) | 0.24 |
| Ghana | iLiNS−DYADG (40) | 347 | 693 | 0.77 (0.37, 1.57) | 0.05 |
| Malawi | iLiNS−DYADM (44) | 220 | 444 | 0.94 (0.39, 2.28) | 0.04 |
| Malawi | iLiNS−DOSE (45) | 696 | 240 | 1.35 (0.54, 3.36) | 0.03 |
| Mali | PROMIS (46) | 507 | 505 | 0.58 (0.31, 1.07) | 0.07 |
| Mali | PROMIS CS (46) | 954 | 970 | 0.90 (0.68, 1.19) | 0.32 |
| Zimbabwe | SHINE (HIV−) (47) | 1872 | 1780 | 0.51 (0.27, 0.97) | 0.07 |
| Zimbabwe | SHINE (HIV+) (48) | 337 | 328 | 0.97 (0.29, 3.28) | 0.02 |
| **I² = 0.00, Tau² = 0.00** | | **6211** | **6308** | **0.86 (0.73, 1.01)** | |
| **Average SQ−LNS compliance − High** | | | | | |
| Bangladesh | JiVitA−4 (34) | 2897 | 1271 | 0.85 (0.70, 1.03) | 0.37 |
| Bangladesh | RDNS (35) | 1663 | 815 | 1.02 (0.69, 1.49) | 0.11 |
| Bangladesh | WASH−B (36) | | | | |
| Burkina Faso | iLiNS−Zinc (37) | 1959 | 666 | 0.64 (0.51, 0.79) | 0.31 |
| Ghana | GHANA (39) | | | | |
| Haiti | HAITI (41) | | | | |
| Kenya | WASH−B (42) | 1461 | 5185 | 0.71 (0.54, 0.94) | 0.20 |
| **I² = 0.53, Tau² = 0.02** | | **7980** | **7937** | **0.77 (0.64, 0.93)** | |

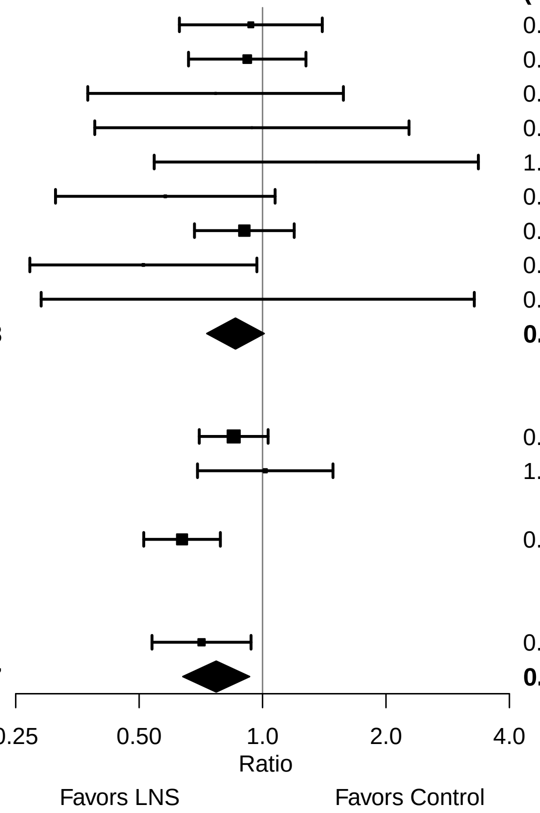

### Supplemental figure 4I: Low MUAC prevalence difference

#### 4I1: Stratified by Geographic region (insufficient comparisons)

#### Supplemental figure 4I: Low MUAC prevalence difference

### 4I2: Stratified by Stunting burden

**Stunting burden**
**(p−diff = 0.829)**

| Country | Trial | N | N | PD (95% CI) | W |
|---|---|---|---|---|---|
| **Stunting burden − Less than 35%** | | | | | |
| Burkina Faso | PROMIS (38) | 854 | 912 | −0.01 (−0.03, 0.02) | 0.20 |
| Burkina Faso | PROMIS CS (38) | 424 | 436 | −0.01 (−0.06, 0.03) | 0.09 |
| Ghana | GHANA (39) | | | | |
| Ghana | iLiNS−DYADG (40) | 347 | 693 | −0.01 (−0.03, 0.01) | 0.22 |
| Haiti | HAITI (41) | | | | |
| Kenya | WASH−B (42) | 1461 | 5185 | −0.02 (−0.03, 0.00) | 0.32 |
| Malawi | iLiNS−DYADM (44) | 220 | 444 | 0.00 (−0.03, 0.03) | 0.17 |
| **I² = 0.00, Tau² = 0.00** | | **3306** | **7670** | **−0.01 (−0.02, 0.00)** | |
| **Stunting burden − More than 35%** | | | | | |
| Bangladesh | JiVitA−4 (34) | 2897 | 1271 | −0.02 (−0.04, 0.00) | 0.11 |
| Bangladesh | RDNS (35) | 1663 | 815 | 0.00 (−0.02, 0.02) | 0.12 |
| Bangladesh | WASH−B (36) | | | | |
| Burkina Faso | iLiNS−Zinc (37) | 1959 | 666 | −0.07 (−0.10, −0.04) | 0.08 |
| Madagascar | MAHAY (43) | 1680 | 1658 | 0.00 (−0.02, 0.02) | 0.11 |
| Malawi | iLiNS−DOSE (45) | 696 | 240 | 0.01 (−0.01, 0.03) | 0.11 |
| Mali | PROMIS (46) | 507 | 505 | −0.02 (−0.05, 0.00) | 0.09 |
| Mali | PROMIS CS (46) | 954 | 970 | −0.01 (−0.04, 0.02) | 0.09 |
| Zimbabwe | SHINE (HIV−) (47) | 1872 | 1780 | −0.01 (−0.01, 0.00) | 0.17 |
| Zimbabwe | SHINE (HIV+) (48) | 337 | 328 | 0.00 (−0.02, 0.02) | 0.12 |
| **I² = 0.67, Tau² = 0.00** | | **12565** | **8233** | **−0.01 (−0.03, 0.00)** | |

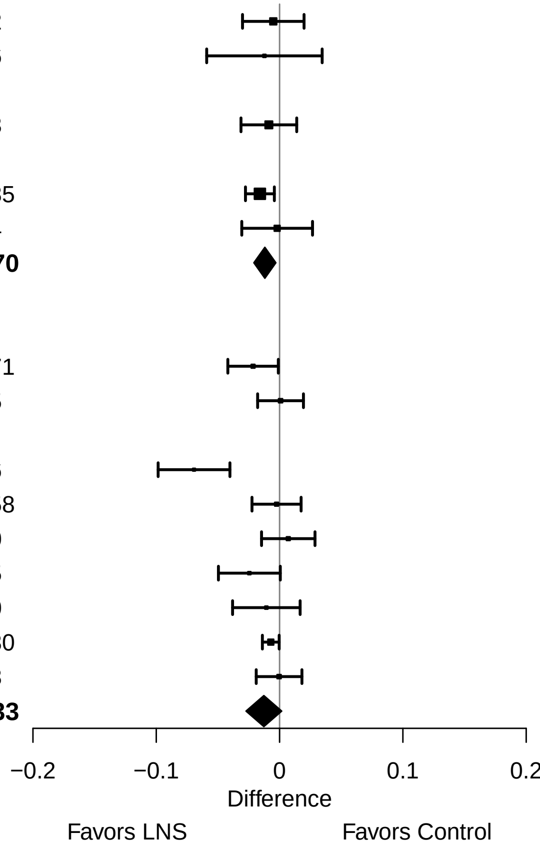

#### Supplemental figure 4I: Low MUAC prevalence difference

### 4I3: Stratified by Malaria prevalence

**Malaria prevalence**
**(p−diff = 0.447)**

**Malaria prevalence − Less than 10%**

| Country | Trial | N | N | PD (95% CI) | W |
|---|---|---|---|---|---|
| Bangladesh | JiVitA−4 (34) | 2897 | 1271 | −0.02 (−0.04, 0.00) | 0.14 |
| Bangladesh | RDNS (35) | 1663 | 815 | 0.00 (−0.02, 0.02) | 0.15 |
| Bangladesh | WASH−B (36) | | | | |
| Haiti | HAITI (41) | | | | |
| Kenya | WASH−B (42) | 1461 | 5185 | −0.02 (−0.03, 0.00) | 0.19 |
| Madagascar | MAHAY (43) | 1680 | 1658 | 0.00 (−0.02, 0.02) | 0.15 |
| Zimbabwe | SHINE (HIV−) (47) | 1872 | 1780 | −0.01 (−0.01, 0.00) | 0.22 |
| Zimbabwe | SHINE (HIV+) (48) | 337 | 328 | 0.00 (−0.02, 0.02) | 0.15 |
| **I² = 0.06, Tau² = 0.00** | | **9910** | **11037** | **−0.01 (−0.01, 0.00)** | |

**Malaria prevalence − At least 10%**

| Country | Trial | N | N | PD (95% CI) | W |
|---|---|---|---|---|---|
| Burkina Faso | iLiNS−Zinc (37) | 1959 | 666 | −0.07 (−0.10, −0.04) | 0.12 |
| Burkina Faso | PROMIS (38) | 854 | 912 | −0.01 (−0.03, 0.02) | 0.14 |
| Burkina Faso | PROMIS CS (38) | 424 | 436 | −0.01 (−0.06, 0.03) | 0.06 |
| Ghana | GHANA (39) | | | | |
| Ghana | iLiNS−DYADG (40) | 347 | 693 | −0.01 (−0.03, 0.01) | 0.15 |
| Malawi | iLiNS−DYADM (44) | 220 | 444 | 0.00 (−0.03, 0.03) | 0.12 |
| Malawi | iLiNS−DOSE (45) | 696 | 240 | 0.01 (−0.01, 0.03) | 0.16 |
| Mali | PROMIS (46) | 507 | 505 | −0.02 (−0.05, 0.00) | 0.14 |
| Mali | PROMIS CS (46) | 954 | 970 | −0.01 (−0.04, 0.02) | 0.12 |
| **I² = 0.64, Tau² = 0.00** | | **5961** | **4866** | **−0.02 (−0.03, 0.00)** | |

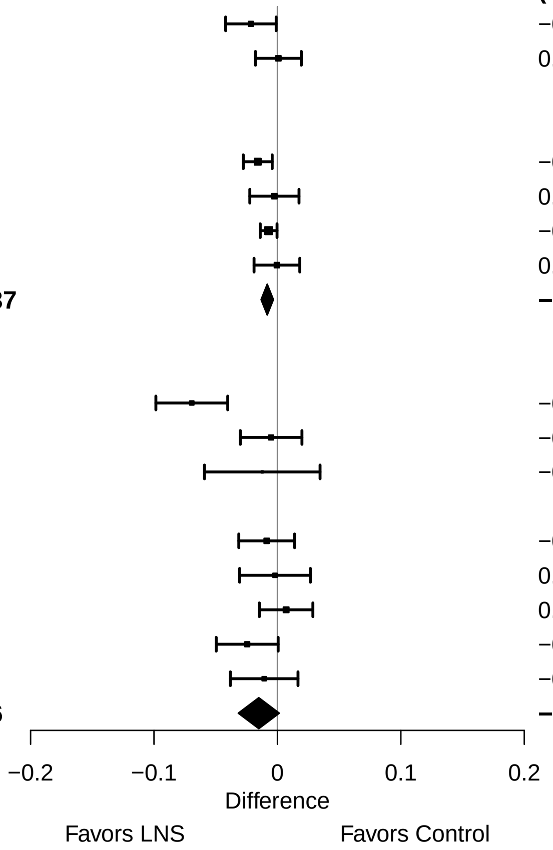

#### Supplemental figure 4I: Low MUAC prevalence difference

#### 4I4: Stratified by Source water quality

**Source water quality**
**(p−diff = 0.262)**

| Country | Trial | N | N | PD (95% CI) | W |
|---|---|---|---|---|---|
| **Source water quality − Improved** | | | | | |
| Bangladesh | JiVitA−4 (34) | 2897 | 1271 | −0.02 (−0.04, 0.00) | 0.22 |
| Bangladesh | RDNS (35) | 1663 | 815 | 0.00 (−0.02, 0.02) | 0.23 |
| Bangladesh | WASH−B (36) | | | | |
| Ghana | GHANA (39) | | | | |
| Ghana | iLiNS−DYADG (40) | 347 | 693 | −0.01 (−0.03, 0.01) | 0.20 |
| Haiti | HAITI (41) | | | | |
| Malawi | iLiNS−DYADM (44) | 220 | 444 | 0.00 (−0.03, 0.03) | 0.15 |
| Malawi | iLiNS−DOSE (45) | 696 | 240 | 0.01 (−0.01, 0.03) | 0.20 |
| **I² = 0.05, Tau² = 0.00** | | **5823** | **3463** | **−0.01 (−0.01, 0.00)** | |
| **Source water quality − Unimproved** | | | | | |
| Burkina Faso | iLiNS−Zinc (37) | 1959 | 666 | −0.07 (−0.10, −0.04) | 0.08 |
| Burkina Faso | PROMIS (38) | 854 | 912 | −0.01 (−0.03, 0.02) | 0.10 |
| Burkina Faso | PROMIS CS (38) | 424 | 436 | −0.01 (−0.06, 0.03) | 0.04 |
| Kenya | WASH−B (42) | 697 | 2270 | −0.02 (−0.04, 0.00) | 0.15 |
| Madagascar | MAHAY (43) | 1680 | 1658 | 0.00 (−0.02, 0.02) | 0.13 |
| Mali | PROMIS (46) | 507 | 505 | −0.02 (−0.05, 0.00) | 0.10 |
| Mali | PROMIS CS (46) | 954 | 970 | −0.01 (−0.04, 0.02) | 0.09 |
| Zimbabwe | SHINE (HIV−) (47) | 889 | 871 | 0.00 (−0.01, 0.01) | 0.19 |
| Zimbabwe | SHINE (HIV+) (48) | 147 | 146 | −0.01 (−0.03, 0.02) | 0.11 |
| **I² = 0.60, Tau² = 0.00** | | **8111** | **8434** | **−0.02 (−0.03, 0.00)** | |

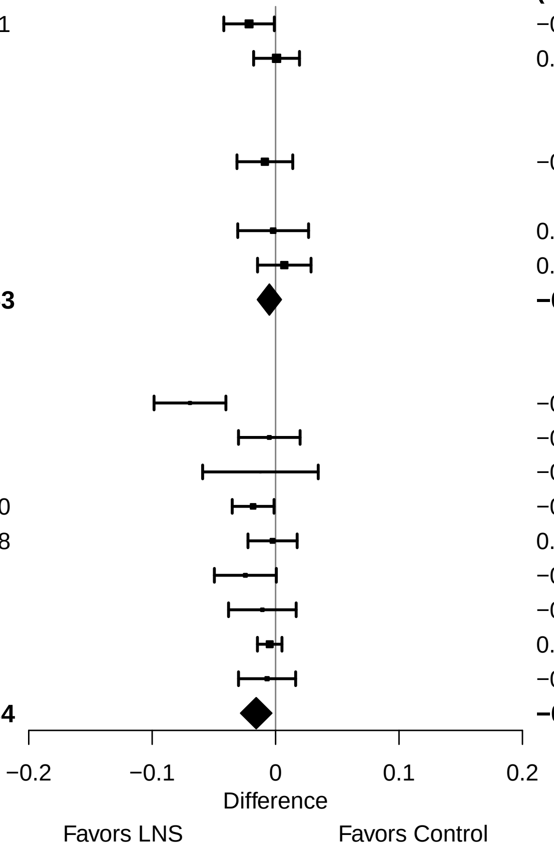

#### Supplemental figure 4I: Low MUAC prevalence difference

### 4I5: Stratified by Sanitation

**Sanitation**
**(p−diff = 0.982)**

| Country | Trial | N | N | PD (95% CI) | W |
|---|---|---|---|---|---|
| **Sanitation − Improved** | | | | | |
| Bangladesh | JiVitA−4 (34) | 2897 | 1271 | −0.02 (−0.04, 0.00) | 0.18 |
| Bangladesh | RDNS (35) | 1663 | 815 | 0.00 (−0.02, 0.02) | 0.19 |
| Bangladesh | WASH−B (36) | | | | |
| Burkina Faso | PROMIS (38) | 854 | 912 | −0.01 (−0.03, 0.02) | 0.14 |
| Burkina Faso | PROMIS CS (38) | 424 | 436 | −0.01 (−0.06, 0.03) | 0.06 |
| Ghana | GHANA (39) | | | | |
| Ghana | iLiNS−DYADG (40) | 347 | 693 | −0.01 (−0.03, 0.01) | 0.16 |
| Haiti | HAITI (41) | | | | |
| Mali | PROMIS (46) | 507 | 505 | −0.02 (−0.05, 0.00) | 0.14 |
| Mali | PROMIS CS (46) | 954 | 970 | −0.01 (−0.04, 0.02) | 0.13 |
| **I² = 0.00, Tau² = 0.00** | | **7646** | **5602** | **−0.01 (−0.02, 0.00)** | |
| **Sanitation − Unimproved** | | | | | |
| Burkina Faso | iLiNS−Zinc (37) | 1959 | 666 | −0.07 (−0.10, −0.04) | 0.10 |
| Kenya | WASH−B (42) | 697 | 2270 | −0.02 (−0.04, 0.00) | 0.17 |
| Madagascar | MAHAY (43) | 1680 | 1658 | 0.00 (−0.02, 0.02) | 0.15 |
| Malawi | iLiNS−DYADM (44) | 220 | 444 | 0.00 (−0.03, 0.03) | 0.10 |
| Malawi | iLiNS−DOSE (45) | 696 | 240 | 0.01 (−0.01, 0.03) | 0.14 |
| Zimbabwe | SHINE (HIV−) (47) | 889 | 871 | 0.00 (−0.01, 0.01) | 0.22 |
| Zimbabwe | SHINE (HIV+) (48) | 147 | 146 | −0.01 (−0.03, 0.02) | 0.13 |
| **I² = 0.72, Tau² = 0.00** | | **6288** | **6295** | **−0.01 (−0.03, 0.01)** | |

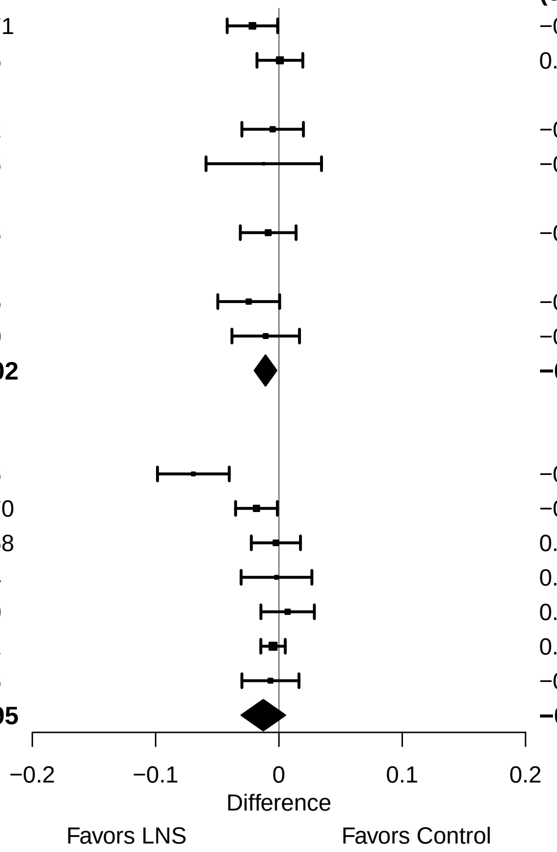

#### Supplemental figure 4I: Low MUAC prevalence difference

### 4I6: Stratified by Supplement duration

**Supplement duration**
**(p−diff = 0.842)**

| Country | Trial | N | N | PD (95% CI) | W |
|---|---|---|---|---|---|
| **Supplement duration − 12m or less** | | | | | |
| Bangladesh | JiVitA−4 (34) | 2897 | 1271 | −0.02 (−0.04, 0.00) | 0.11 |
| Burkina Faso | iLiNS−Zinc (37) | 1959 | 666 | −0.07 (−0.10, −0.04) | 0.08 |
| Burkina Faso | PROMIS (38) | 854 | 912 | −0.01 (−0.03, 0.02) | 0.09 |
| Burkina Faso | PROMIS CS (38) | 424 | 436 | −0.01 (−0.06, 0.03) | 0.04 |
| Ghana | GHANA (39) | | | | |
| Ghana | iLiNS−DYADG (40) | 347 | 693 | −0.01 (−0.03, 0.01) | 0.10 |
| Haiti | HAITI (41) | | | | |
| Madagascar | MAHAY (43) | 1680 | 1658 | 0.00 (−0.02, 0.02) | 0.11 |
| Malawi | iLiNS−DYADM (44) | 220 | 444 | 0.00 (−0.03, 0.03) | 0.08 |
| Malawi | iLiNS−DOSE (45) | 696 | 240 | 0.01 (−0.01, 0.03) | 0.10 |
| Zimbabwe | SHINE (HIV−) (47) | 1872 | 1780 | −0.01 (−0.01, 0.00) | 0.17 |
| Zimbabwe | SHINE (HIV+) (48) | 337 | 328 | 0.00 (−0.02, 0.02) | 0.12 |
| **I² = 0.59, Tau² = 0.00** | | **11286** | **8428** | **−0.01 (−0.02, 0.00)** | |
| **Supplement duration − > 12m** | | | | | |
| Bangladesh | RDNS (35) | 1663 | 815 | 0.00 (−0.02, 0.02) | 0.27 |
| Bangladesh | WASH−B (36) | | | | |
| Kenya | WASH−B (42) | 1461 | 5185 | −0.02 (−0.03, 0.00) | 0.33 |
| Mali | PROMIS (46) | 507 | 505 | −0.02 (−0.05, 0.00) | 0.21 |
| Mali | PROMIS CS (46) | 954 | 970 | −0.01 (−0.04, 0.02) | 0.19 |
| **I² = 0.06, Tau² = 0.00** | | **4585** | **7475** | **−0.01 (−0.02, 0.00)** | |

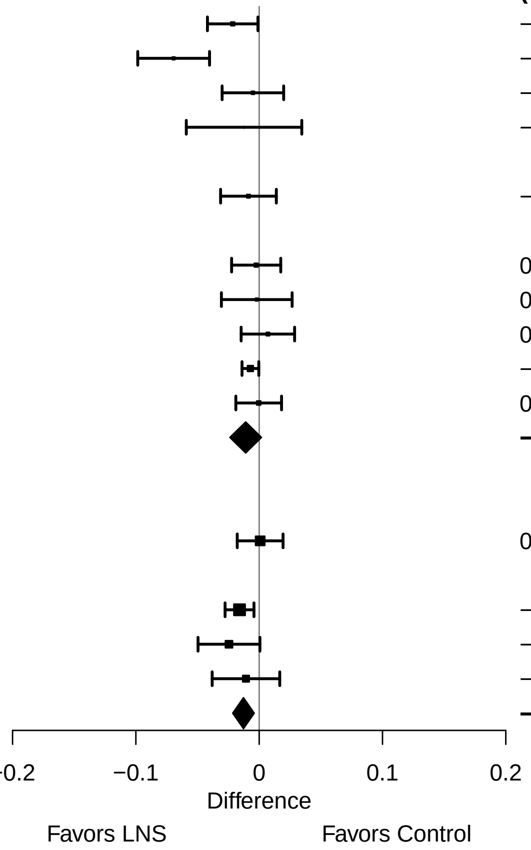

#### Supplemental figure 4I: Low MUAC prevalence difference

### 4I7: Stratified by Frequency of contact

**Frequency of contact**
**(p−diff = 0.331)**

| Country | Trial | N | N | PD (95% CI) | W |
|---|---|---|---|---|---|
| **Frequency of contact − Monthly** | | | | | |
| Bangladesh | RDNS (35) | 1663 | 815 | 0.00 (−0.02, 0.02) | 0.12 |
| Burkina Faso | PROMIS (38) | 854 | 912 | −0.01 (−0.03, 0.02) | 0.10 |
| Burkina Faso | PROMIS CS (38) | 424 | 436 | −0.01 (−0.06, 0.03) | 0.04 |
| Haiti | HAITI (41) | | | | |
| Kenya | WASH−B (42) | 1461 | 5185 | −0.02 (−0.03, 0.00) | 0.15 |
| Madagascar | MAHAY (43) | 1680 | 1658 | 0.00 (−0.02, 0.02) | 0.12 |
| Mali | PROMIS (46) | 507 | 505 | −0.02 (−0.05, 0.00) | 0.09 |
| Mali | PROMIS CS (46) | 954 | 970 | −0.01 (−0.04, 0.02) | 0.09 |
| Zimbabwe | SHINE (HIV−) (47) | 1872 | 1780 | −0.01 (−0.01, 0.00) | 0.17 |
| Zimbabwe | SHINE (HIV+) (48) | 337 | 328 | 0.00 (−0.02, 0.02) | 0.12 |
| **I² = 0.00, Tau² = 0.00** | | **9752** | **12589** | **−0.01 (−0.01, 0.00)** | |
| **Frequency of contact − Weekly** | | | | | |
| Bangladesh | JiVitA−4 (34) | 2897 | 1271 | −0.02 (−0.04, 0.00) | 0.23 |
| Bangladesh | WASH−B (36) | | | | |
| Burkina Faso | iLiNS−Zinc (37) | 1959 | 666 | −0.07 (−0.10, −0.04) | 0.17 |
| Ghana | GHANA (39) | | | | |
| Ghana | iLiNS−DYADG (40) | 347 | 693 | −0.01 (−0.03, 0.01) | 0.21 |
| Malawi | iLiNS−DYADM (44) | 220 | 444 | 0.00 (−0.03, 0.03) | 0.17 |
| Malawi | iLiNS−DOSE (45) | 696 | 240 | 0.01 (−0.01, 0.03) | 0.22 |
| **I² = 0.79, Tau² = 0.00** | | **6119** | **3314** | **−0.02 (−0.04, 0.01)** | |

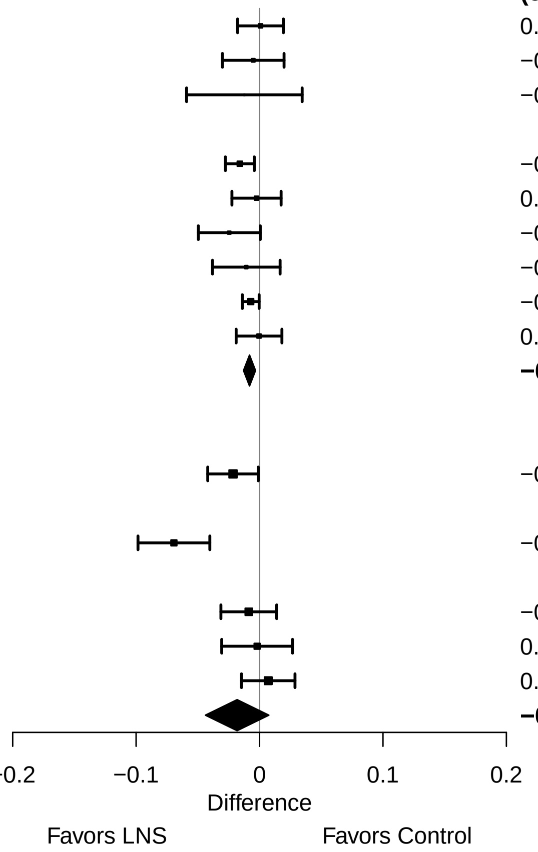

#### Supplemental figure 4I: Low MUAC prevalence difference

##### 4I8: Stratified by Average SQ-LNS compliance

**Average SQ−LNS compliance**
**(p−diff = 0.094)**

| Country | Trial | N | N | PD (95% CI) | W |
|---|---|---|---|---|---|
| **Average SQ−LNS compliance − Low** | | | | | |
| Burkina Faso | PROMIS (38) | 854 | 912 | −0.01 (−0.03, 0.02) | 0.11 |
| Burkina Faso | PROMIS CS (38) | 424 | 436 | −0.01 (−0.06, 0.03) | 0.05 |
| Ghana | iLiNS−DYADG (40) | 347 | 693 | −0.01 (−0.03, 0.01) | 0.12 |
| Malawi | iLiNS−DYADM (44) | 220 | 444 | 0.00 (−0.03, 0.03) | 0.09 |
| Malawi | iLiNS−DOSE (45) | 696 | 240 | 0.01 (−0.01, 0.03) | 0.12 |
| Mali | PROMIS (46) | 507 | 505 | −0.02 (−0.05, 0.00) | 0.11 |
| Mali | PROMIS CS (46) | 954 | 970 | −0.01 (−0.04, 0.02) | 0.10 |
| Zimbabwe | SHINE (HIV−) (47) | 1872 | 1780 | −0.01 (−0.01, 0.00) | 0.18 |
| Zimbabwe | SHINE (HIV+) (48) | 337 | 328 | 0.00 (−0.02, 0.02) | 0.13 |
| **I² = 0.00, Tau² = 0.00** | | **6211** | **6308** | **−0.01 (−0.01, 0.00)** | |
| **Average SQ−LNS compliance − High** | | | | | |
| Bangladesh | JiVitA−4 (34) | 2897 | 1271 | −0.02 (−0.04, 0.00) | 0.24 |
| Bangladesh | RDNS (35) | 1663 | 815 | 0.00 (−0.02, 0.02) | 0.26 |
| Bangladesh | WASH−B (36) | | | | |
| Burkina Faso | iLiNS−Zinc (37) | 1959 | 666 | −0.07 (−0.10, −0.04) | 0.18 |
| Ghana | GHANA (39) | | | | |
| Haiti | HAITI (41) | | | | |
| Kenya | WASH−B (42) | 1461 | 5185 | −0.02 (−0.03, 0.00) | 0.32 |
| **I² = 0.81, Tau² = 0.00** | | **7980** | **7937** | **−0.02 (−0.05, 0.00)** | |

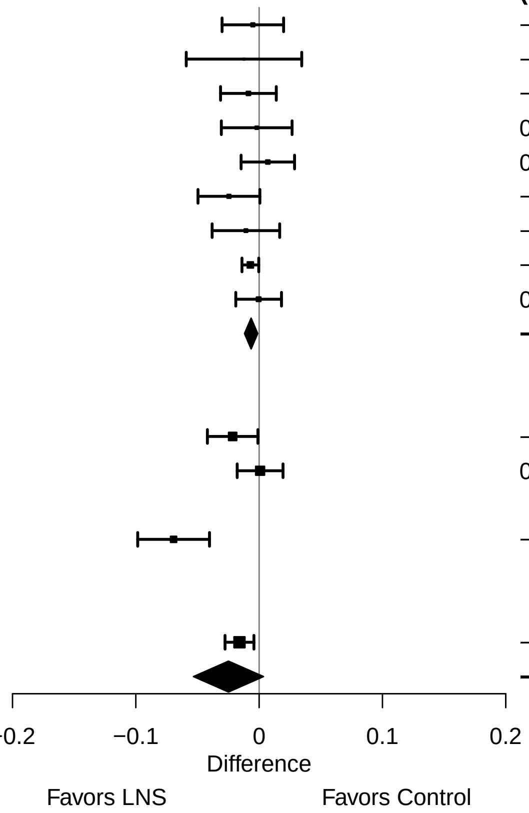

### Supplemental figure 4J: Acute malnutrition prevalence ratio

#### 4J1: Stratified by Geographic region (insufficient comparisons)

**Supplemental figure 4J: Acute malnutrition prevalence ratio**

#### 4J2: Stratified by Stunting burden

**Stunting burden**
**(p−diff = 0.835)**

| Country | Trial | N | N | PR (95% CI) | W |
|---|---|---|---|---|---|
| **Stunting burden − Less than 35%** | | | | | |
| Burkina Faso | PROMIS (38) | 846 | 901 | 0.83 (0.60, 1.14) | 0.26 |
| Burkina Faso | PROMIS CS (38) | 424 | 434 | 0.95 (0.73, 1.23) | 0.38 |
| Ghana | GHANA (39) | | | | |
| Ghana | iLiNS−DYADG (40) | 347 | 692 | 0.85 (0.54, 1.35) | 0.12 |
| Haiti | HAITI (41) | | | | |
| Kenya | WASH−B (42) | 1454 | 5117 | 0.71 (0.49, 1.02) | 0.20 |
| Malawi | iLiNS−DYADM (44) | 220 | 444 | 0.81 (0.36, 1.80) | 0.04 |
| **I² = 0.00, Tau² = 0.00** | | **3291** | **7588** | **0.85 (0.72, 1.00)** | |
| **Stunting burden − More than 35%** | | | | | |
| Bangladesh | JiVitA−4 (34) | 2783 | 1216 | 0.89 (0.78, 1.01) | 0.41 |
| Bangladesh | RDNS (35) | 1661 | 815 | 0.86 (0.70, 1.05) | 0.16 |
| Bangladesh | WASH−B (36) | | | | |
| Burkina Faso | iLiNS−Zinc (37) | 1951 | 664 | 0.67 (0.54, 0.85) | 0.13 |
| Madagascar | MAHAY (43) | 1675 | 1655 | 0.98 (0.75, 1.29) | 0.09 |
| Malawi | iLiNS−DOSE (45) | 690 | 240 | 1.42 (0.83, 2.43) | 0.02 |
| Mali | PROMIS (46) | 499 | 502 | 0.59 (0.23, 1.47) | 0.01 |
| Mali | PROMIS CS (46) | 944 | 959 | 0.89 (0.70, 1.13) | 0.12 |
| Zimbabwe | SHINE (HIV−) (47) | 1867 | 1777 | 0.79 (0.55, 1.14) | 0.05 |
| Zimbabwe | SHINE (HIV+) (48) | 336 | 327 | 1.23 (0.61, 2.48) | 0.01 |
| **I² = 0.27, Tau² = 0.01** | | **12406** | **8155** | **0.87 (0.77, 0.98)** | |

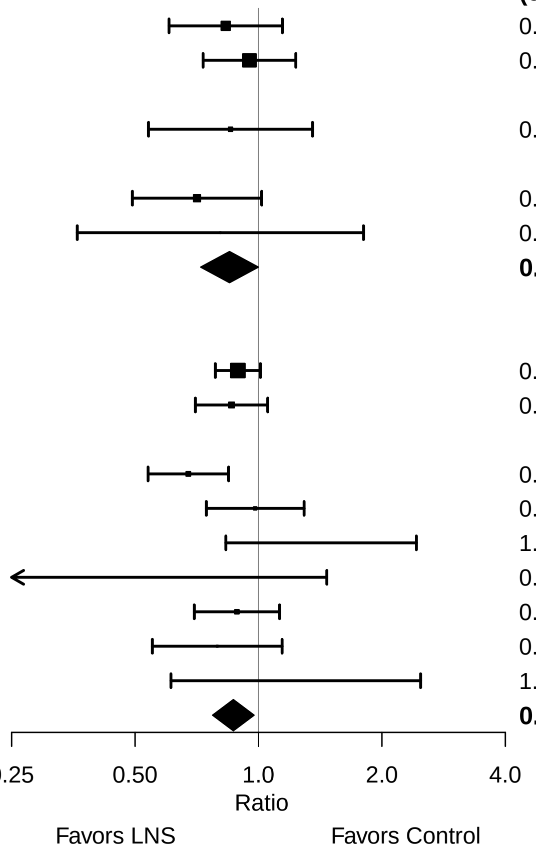

#### Supplemental figure 4J: Acute malnutrition prevalence ratio

#### 4J3: Stratified by Malaria prevalence

**Malaria prevalence**
**(p−diff = 0.526)**

| Country | Trial | N | N | PR (95% CI) | W |
|---|---|---|---|---|---|
| **Malaria prevalence − Less than 10%** | | | | | |
| Bangladesh | JiVitA−4 (34) | 2783 | 1216 | 0.89 (0.78, 1.01) | 0.53 |
| Bangladesh | RDNS (35) | 1661 | 815 | 0.86 (0.70, 1.05) | 0.21 |
| Bangladesh | WASH−B (36) | | | | |
| Haiti | HAITI (41) | | | | |
| Kenya | WASH−B (42) | 1454 | 5117 | 0.71 (0.49, 1.02) | 0.06 |
| Madagascar | MAHAY (43) | 1675 | 1655 | 0.98 (0.75, 1.29) | 0.11 |
| Zimbabwe | SHINE (HIV−) (47) | 1867 | 1777 | 0.79 (0.55, 1.14) | 0.06 |
| Zimbabwe | SHINE (HIV+) (48) | 336 | 327 | 1.23 (0.61, 2.48) | 0.02 |
| **I² = 0.00, Tau² = 0.00** | | **9776** | **10907** | **0.88 (0.80, 0.96)** | |
| **Malaria prevalence − At least 10%** | | | | | |
| Burkina Faso | iLiNS−Zinc (37) | 1951 | 664 | 0.67 (0.54, 0.85) | 0.27 |
| Burkina Faso | PROMIS (38) | 846 | 901 | 0.83 (0.60, 1.14) | 0.14 |
| Burkina Faso | PROMIS CS (38) | 424 | 434 | 0.95 (0.73, 1.23) | 0.20 |
| Ghana | GHANA (39) | | | | |
| Ghana | iLiNS−DYADG (40) | 347 | 692 | 0.85 (0.54, 1.35) | 0.07 |
| Malawi | iLiNS−DYADM (44) | 220 | 444 | 0.81 (0.36, 1.80) | 0.02 |
| Malawi | iLiNS−DOSE (45) | 690 | 240 | 1.42 (0.83, 2.43) | 0.05 |
| Mali | PROMIS (46) | 499 | 502 | 0.59 (0.23, 1.47) | 0.02 |
| Mali | PROMIS CS (46) | 944 | 959 | 0.89 (0.70, 1.13) | 0.24 |
| **I² = 0.22, Tau² = 0.01** | | **5921** | **4836** | **0.85 (0.74, 0.97)** | |

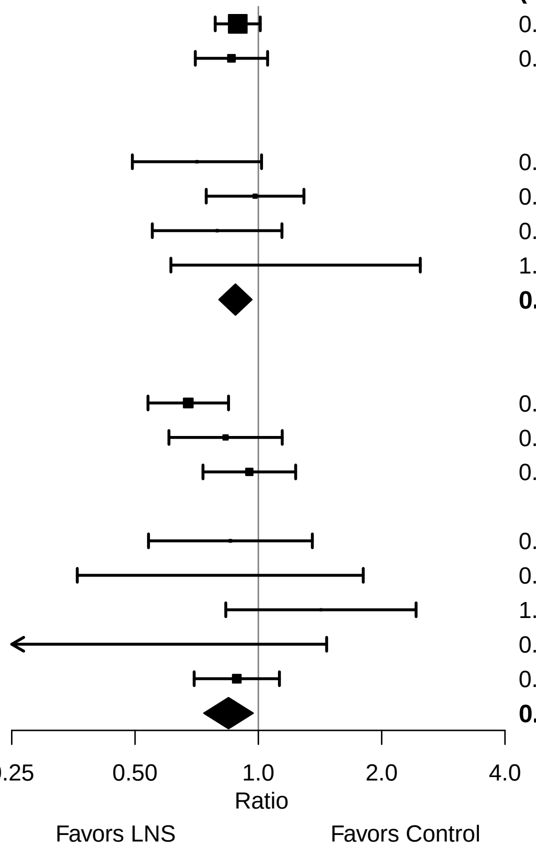

#### Supplemental figure 4J: Acute malnutrition prevalence ratio

##### 4J4: Stratified by Source water quality

**Source water quality**
**(p−diff = 0.378)**

| Country | Trial | N | N | PR (95% CI) | W |
|---|---|---|---|---|---|
| **Source water quality − Improved** | | | | | |
| Bangladesh | JiVitA−4 (34) | 2783 | 1216 | 0.89 (0.78, 1.01) | 0.65 |
| Bangladesh | RDNS (35) | 1661 | 815 | 0.86 (0.70, 1.05) | 0.25 |
| Bangladesh | WASH−B (36) | | | | |
| Ghana | GHANA (39) | | | | |
| Ghana | iLiNS−DYADG (40) | 347 | 692 | 0.85 (0.54, 1.35) | 0.05 |
| Haiti | HAITI (41) | | | | |
| Malawi | iLiNS−DYADM (44) | 220 | 444 | 0.81 (0.36, 1.80) | 0.02 |
| Malawi | iLiNS−DOSE (45) | 690 | 240 | 1.42 (0.83, 2.43) | 0.04 |
| **I² = 0.00, Tau² = 0.00** | | **5701** | **3407** | **0.89 (0.81, 0.99)** | |
| **Source water quality − Unimproved** | | | | | |
| Burkina Faso | iLiNS−Zinc (37) | 1951 | 664 | 0.67 (0.54, 0.85) | 0.23 |
| Burkina Faso | PROMIS (38) | 846 | 901 | 0.83 (0.60, 1.14) | 0.12 |
| Burkina Faso | PROMIS CS (38) | 424 | 434 | 0.95 (0.73, 1.23) | 0.18 |
| Kenya | WASH−B (42) | 694 | 2244 | 0.76 (0.43, 1.34) | 0.04 |
| Madagascar | MAHAY (43) | 1675 | 1655 | 0.98 (0.75, 1.29) | 0.16 |
| Mali | PROMIS (46) | 499 | 502 | 0.59 (0.23, 1.47) | 0.01 |
| Mali | PROMIS CS (46) | 944 | 959 | 0.89 (0.70, 1.13) | 0.21 |
| Zimbabwe | SHINE (HIV−) (47) | 886 | 871 | 0.82 (0.49, 1.37) | 0.05 |
| Zimbabwe | SHINE (HIV+) (48) | 147 | 146 | 0.87 (0.27, 2.79) | 0.01 |
| **I² = 0.00, Tau² = 0.00** | | **8066** | **8376** | **0.84 (0.75, 0.93)** | |

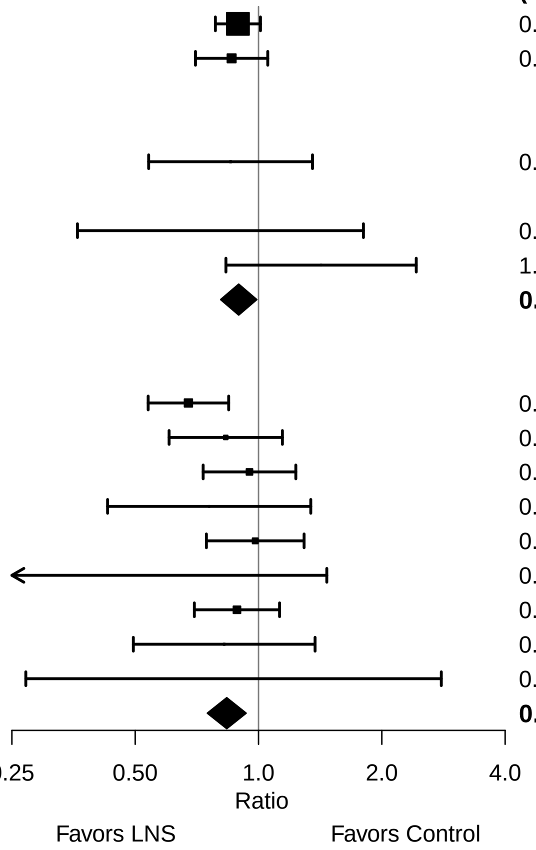

#### Supplemental figure 4J: Acute malnutrition prevalence ratio

### 4J5: Stratified by Sanitation

**Sanitation**
**(p−diff = 0.450)**

| Country | Trial | N | N | PR (95% CI) | W |
|---|---|---|---|---|---|
| **Sanitation − Improved** | | | | | |
| Bangladesh | JiVitA−4 (34) | 2783 | 1216 | 0.89 (0.78, 1.01) | 0.46 |
| Bangladesh | RDNS (35) | 1661 | 815 | 0.86 (0.70, 1.05) | 0.18 |
| Bangladesh | WASH−B (36) | | | | |
| Burkina Faso | PROMIS (38) | 846 | 901 | 0.83 (0.60, 1.14) | 0.07 |
| Burkina Faso | PROMIS CS (38) | 424 | 434 | 0.95 (0.73, 1.23) | 0.11 |
| Ghana | GHANA (39) | | | | |
| Ghana | iLiNS−DYADG (40) | 347 | 692 | 0.85 (0.54, 1.35) | 0.04 |
| Haiti | HAITI (41) | | | | |
| Mali | PROMIS (46) | 499 | 502 | 0.59 (0.23, 1.47) | 0.01 |
| Mali | PROMIS CS (46) | 944 | 959 | 0.89 (0.70, 1.13) | 0.13 |
| **I² = 0.00, Tau² = 0.00** | | **7504** | **5519** | **0.88 (0.81, 0.96)** | |
| **Sanitation − Unimproved** | | | | | |
| Burkina Faso | iLiNS−Zinc (37) | 1951 | 664 | 0.67 (0.54, 0.85) | 0.43 |
| Kenya | WASH−B (42) | 694 | 2244 | 0.76 (0.43, 1.34) | 0.07 |
| Madagascar | MAHAY (43) | 1675 | 1655 | 0.98 (0.75, 1.29) | 0.29 |
| Malawi | iLiNS−DYADM (44) | 220 | 444 | 0.81 (0.36, 1.80) | 0.03 |
| Malawi | iLiNS−DOSE (45) | 690 | 240 | 1.42 (0.83, 2.43) | 0.08 |
| Zimbabwe | SHINE (HIV−) (47) | 886 | 871 | 0.82 (0.49, 1.37) | 0.08 |
| Zimbabwe | SHINE (HIV+) (48) | 147 | 146 | 0.87 (0.27, 2.79) | 0.02 |
| **I² = 0.31, Tau² = 0.01** | | **6263** | **6264** | **0.85 (0.70, 1.02)** | |

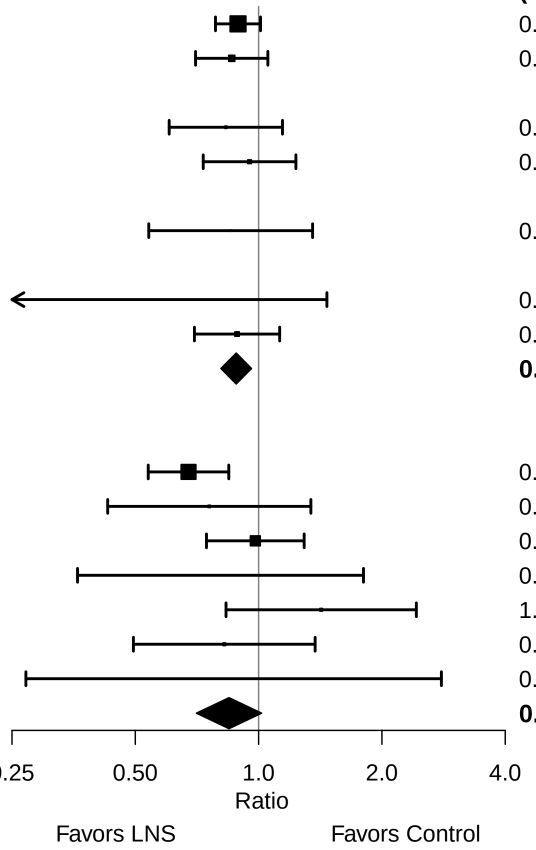

#### Supplemental figure 4J: Acute malnutrition prevalence ratio

#### 4J6: Stratified by Supplement duration

**Supplement duration**
**(p−diff = 0.607)**

| Country | Trial | N | N | PR (95% CI) | W |
|---|---|---|---|---|---|
| **Supplement duration − 12m or less** | | | | | |
| Bangladesh | JiVitA−4 (34) | 2783 | 1216 | 0.89 (0.78, 1.01) | 0.45 |
| Burkina Faso | iLiNS−Zinc (37) | 1951 | 664 | 0.67 (0.54, 0.85) | 0.14 |
| Burkina Faso | PROMIS (38) | 846 | 901 | 0.83 (0.60, 1.14) | 0.07 |
| Burkina Faso | PROMIS CS (38) | 424 | 434 | 0.95 (0.73, 1.23) | 0.11 |
| Ghana | GHANA (39) | | | | |
| Ghana | iLiNS−DYADG (40) | 347 | 692 | 0.85 (0.54, 1.35) | 0.03 |
| Haiti | HAITI (41) | | | | |
| Madagascar | MAHAY (43) | 1675 | 1655 | 0.98 (0.75, 1.29) | 0.10 |
| Malawi | iLiNS−DYADM (44) | 220 | 444 | 0.81 (0.36, 1.80) | 0.01 |
| Malawi | iLiNS−DOSE (45) | 690 | 240 | 1.42 (0.83, 2.43) | 0.03 |
| Zimbabwe | SHINE (HIV−) (47) | 1867 | 1777 | 0.79 (0.55, 1.14) | 0.05 |
| Zimbabwe | SHINE (HIV+) (48) | 336 | 327 | 1.23 (0.61, 2.48) | 0.01 |
| **I² = 0.16, Tau² = 0.00** | | **11139** | **8350** | **0.87 (0.79, 0.97)** | |
| **Supplement duration − > 12m** | | | | | |
| Bangladesh | RDNS (35) | 1661 | 815 | 0.86 (0.70, 1.05) | 0.48 |
| Bangladesh | WASH−B (36) | | | | |
| Kenya | WASH−B (42) | 1454 | 5117 | 0.71 (0.49, 1.02) | 0.15 |
| Mali | PROMIS (46) | 499 | 502 | 0.59 (0.23, 1.47) | 0.02 |
| Mali | PROMIS CS (46) | 944 | 959 | 0.89 (0.70, 1.13) | 0.35 |
| **I² = 0.00, Tau² = 0.00** | | **4558** | **7393** | **0.84 (0.73, 0.96)** | |

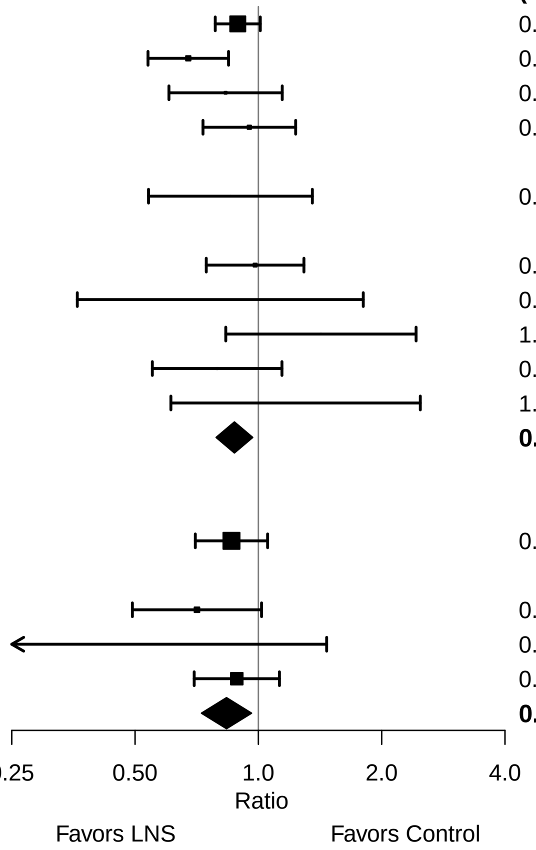

#### Supplemental figure 4J: Acute malnutrition prevalence ratio

#### 4J7: Stratified by Frequency of contact

**Frequency of contact**
**(p−diff = 0.706)**

| Country | Trial | N | N | PR (95% CI) | W |
|---|---|---|---|---|---|
| **Frequency of contact − Monthly** | | | | | |
| Bangladesh | RDNS (35) | 1661 | 815 | 0.86 (0.70, 1.05) | 0.25 |
| Burkina Faso | PROMIS (38) | 846 | 901 | 0.83 (0.60, 1.14) | 0.10 |
| Burkina Faso | PROMIS CS (38) | 424 | 434 | 0.95 (0.73, 1.23) | 0.15 |
| Haiti | HAITI (41) | | | | |
| Kenya | WASH−B (42) | 1454 | 5117 | 0.71 (0.49, 1.02) | 0.08 |
| Madagascar | MAHAY (43) | 1675 | 1655 | 0.98 (0.75, 1.29) | 0.14 |
| Mali | PROMIS (46) | 499 | 502 | 0.59 (0.23, 1.47) | 0.01 |
| Mali | PROMIS CS (46) | 944 | 959 | 0.89 (0.70, 1.13) | 0.18 |
| Zimbabwe | SHINE (HIV−) (47) | 1867 | 1777 | 0.79 (0.55, 1.14) | 0.08 |
| Zimbabwe | SHINE (HIV+) (48) | 336 | 327 | 1.23 (0.61, 2.48) | 0.02 |
| **I² = 0.00, Tau² = 0.00** | | **9706** | **12487** | **0.87 (0.79, 0.97)** | |
| **Frequency of contact − Weekly** | | | | | |
| Bangladesh | JiVitA−4 (34) | 2783 | 1216 | 0.89 (0.78, 1.01) | 0.68 |
| Bangladesh | WASH−B (36) | | | | |
| Burkina Faso | iLiNS−Zinc (37) | 1951 | 664 | 0.67 (0.54, 0.85) | 0.21 |
| Ghana | GHANA (39) | | | | |
| Ghana | iLiNS−DYADG (40) | 347 | 692 | 0.85 (0.54, 1.35) | 0.05 |
| Malawi | iLiNS−DYADM (44) | 220 | 444 | 0.81 (0.36, 1.80) | 0.02 |
| Malawi | iLiNS−DOSE (45) | 690 | 240 | 1.42 (0.83, 2.43) | 0.04 |
| **I² = 0.51, Tau² = 0.03** | | **5991** | **3256** | **0.86 (0.69, 1.06)** | |

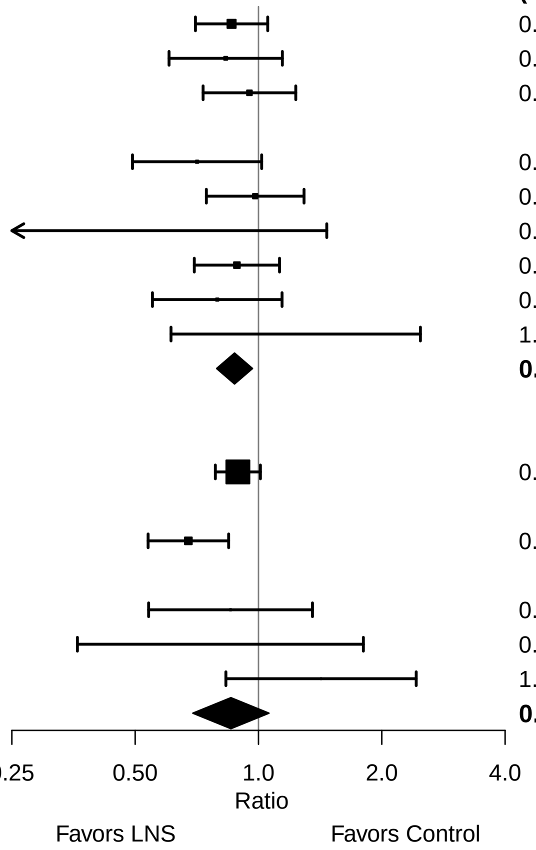

#### Supplemental figure 4J: Acute malnutrition prevalence ratio

#### 4J8: Stratified by Average SQ-LNS compliance

**Average SQ−LNS compliance**
**(p−diff = 0.269)**

**Average SQ−LNS compliance − Low**

| Country | Trial | N | N | PR (95% CI) | W |
|---|---|---|---|---|---|
| Burkina Faso | PROMIS (38) | 846 | 901 | 0.83 (0.60, 1.14) | 0.16 |
| Burkina Faso | PROMIS CS (38) | 424 | 434 | 0.95 (0.73, 1.23) | 0.24 |
| Ghana | iLiNS−DYADG (40) | 347 | 692 | 0.85 (0.54, 1.35) | 0.08 |
| Malawi | iLiNS−DYADM (44) | 220 | 444 | 0.81 (0.36, 1.80) | 0.02 |
| Malawi | iLiNS−DOSE (45) | 690 | 240 | 1.42 (0.83, 2.43) | 0.06 |
| Mali | PROMIS (46) | 499 | 502 | 0.59 (0.23, 1.47) | 0.02 |
| Mali | PROMIS CS (46) | 944 | 959 | 0.89 (0.70, 1.13) | 0.28 |
| Zimbabwe | SHINE (HIV−) (47) | 1867 | 1777 | 0.79 (0.55, 1.14) | 0.12 |
| Zimbabwe | SHINE (HIV+) (48) | 336 | 327 | 1.23 (0.61, 2.48) | 0.03 |
| **I² = 0.00, Tau² = 0.00** | | **6173** | **6276** | **0.90 (0.79, 1.02)** | |

**Average SQ−LNS compliance − High**

| Country | Trial | N | N | PR (95% CI) | W |
|---|---|---|---|---|---|
| Bangladesh | JiVitA−4 (34) | 2783 | 1216 | 0.89 (0.78, 1.01) | 0.55 |
| Bangladesh | RDNS (35) | 1661 | 815 | 0.86 (0.70, 1.05) | 0.21 |
| Bangladesh | WASH−B (36) | | | | |
| Burkina Faso | iLiNS−Zinc (37) | 1951 | 664 | 0.67 (0.54, 0.85) | 0.17 |
| Ghana | GHANA (39) | | | | |
| Haiti | HAITI (41) | | | | |
| Kenya | WASH−B (42) | 1454 | 5117 | 0.71 (0.49, 1.02) | 0.07 |
| **I² = 0.43, Tau² = 0.01** | | **7849** | **7812** | **0.81 (0.71, 0.92)** | |

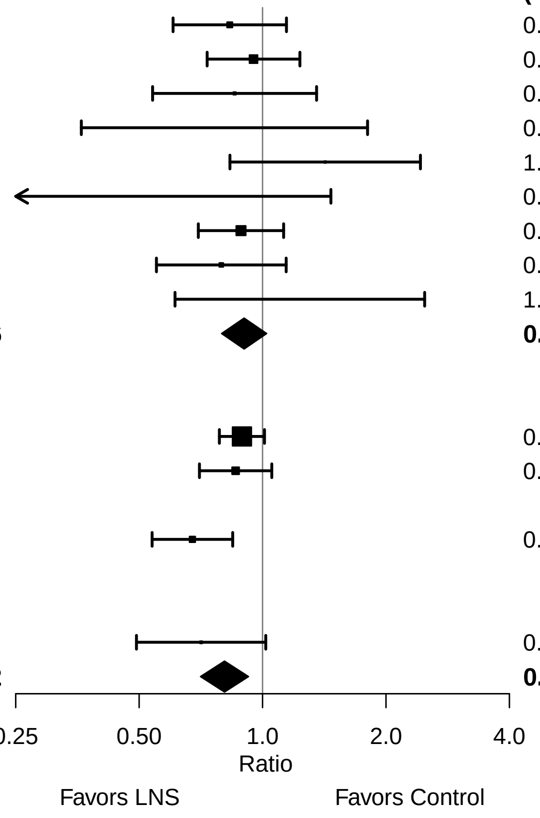

### Supplemental figure 4K: Acute malnutrition prevalence difference

#### 4K1: Stratified by Geographic region (insufficient comparisons)

#### Supplemental figure 4K: Acute malnutrition prevalence difference

### 4K2: Stratified by Stunting burden

**Stunting burden**
**(p−diff = 0.872)**

| Country | Trial | N | N | PD (95% CI) | W |
|---|---|---|---|---|---|
| **Stunting burden − Less than 35%** | | | | | |
| Burkina Faso | PROMIS (38) | 846 | 901 | −0.02 (−0.05, 0.01) | 0.16 |
| Burkina Faso | PROMIS CS (38) | 424 | 434 | −0.01 (−0.06, 0.04) | 0.06 |
| Ghana | GHANA (39) | | | | |
| Ghana | iLiNS−DYADG (40) | 347 | 692 | −0.01 (−0.05, 0.02) | 0.13 |
| Haiti | HAITI (41) | | | | |
| Kenya | WASH−B (42) | 1454 | 5117 | −0.01 (−0.02, 0.00) | 0.51 |
| Malawi | iLiNS−DYADM (44) | 220 | 444 | −0.01 (−0.04, 0.02) | 0.14 |
| **I² = 0.00, Tau² = 0.00** | | **3291** | **7588** | **−0.01 (−0.02, 0.00)** | |
| **Stunting burden − More than 35%** | | | | | |
| Bangladesh | JiVitA−4 (34) | 2783 | 1216 | −0.02 (−0.05, 0.00) | 0.11 |
| Bangladesh | RDNS (35) | 1661 | 815 | −0.02 (−0.05, 0.01) | 0.08 |
| Bangladesh | WASH−B (36) | | | | |
| Burkina Faso | iLiNS−Zinc (37) | 1951 | 664 | −0.06 (−0.09, −0.03) | 0.08 |
| Madagascar | MAHAY (43) | 1675 | 1655 | 0.00 (−0.02, 0.02) | 0.14 |
| Malawi | iLiNS−DOSE (45) | 690 | 240 | 0.01 (−0.03, 0.04) | 0.06 |
| Mali | PROMIS (46) | 499 | 502 | −0.01 (−0.03, 0.01) | 0.15 |
| Mali | PROMIS CS (46) | 944 | 959 | −0.01 (−0.04, 0.01) | 0.08 |
| Zimbabwe | SHINE (HIV−) (47) | 1867 | 1777 | −0.01 (−0.02, 0.00) | 0.24 |
| Zimbabwe | SHINE (HIV+) (48) | 336 | 327 | 0.01 (−0.02, 0.04) | 0.07 |
| **I² = 0.50, Tau² = 0.00** | | **12406** | **8155** | **−0.01 (−0.03, 0.00)** | |

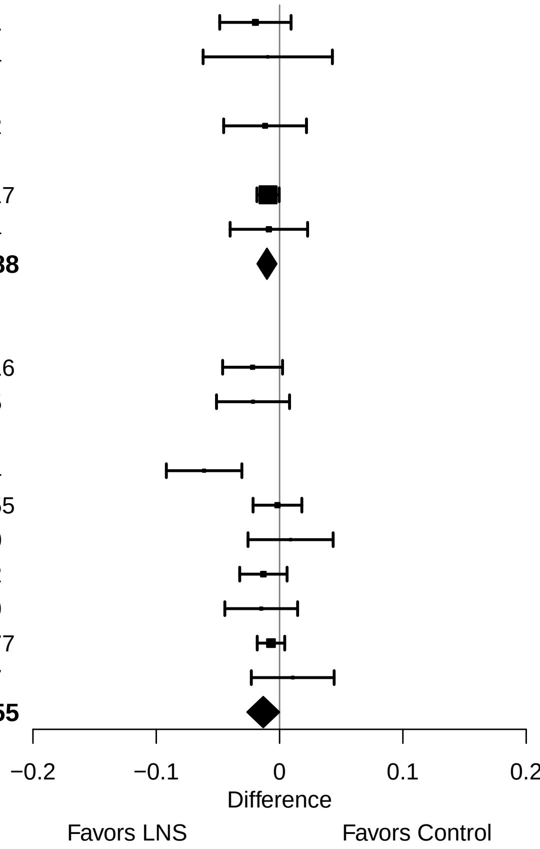

#### Supplemental figure 4K: Acute malnutrition prevalence difference

### 4K3: Stratified by Malaria prevalence

**Malaria prevalence**
**(p−diff = 0.181)**

| Country | Trial | N | N | PD (95% CI) | W |
|---|---|---|---|---|---|
| **Malaria prevalence − Less than 10%** | | | | | |
| Bangladesh | JiVitA−4 (34) | 2783 | 1216 | −0.02 (−0.05, 0.00) | 0.12 |
| Bangladesh | RDNS (35) | 1661 | 815 | −0.02 (−0.05, 0.01) | 0.09 |
| Bangladesh | WASH−B (36) | | | | |
| Haiti | HAITI (41) | | | | |
| Kenya | WASH−B (42) | 1454 | 5117 | −0.01 (−0.02, 0.00) | 0.30 |
| Madagascar | MAHAY (43) | 1675 | 1655 | 0.00 (−0.02, 0.02) | 0.16 |
| Zimbabwe | SHINE (HIV−) (47) | 1867 | 1777 | −0.01 (−0.02, 0.00) | 0.26 |
| Zimbabwe | SHINE (HIV+) (48) | 336 | 327 | 0.01 (−0.02, 0.04) | 0.07 |
| **$I^2$ = 0.00, $Tau^2$ = 0.00** | | **9776** | **10907** | **−0.01 (−0.01, 0.00)** | |
| **Malaria prevalence − At least 10%** | | | | | |
| Burkina Faso | iLiNS−Zinc (37) | 1951 | 664 | −0.06 (−0.09, −0.03) | 0.12 |
| Burkina Faso | PROMIS (38) | 846 | 901 | −0.02 (−0.05, 0.01) | 0.13 |
| Burkina Faso | PROMIS CS (38) | 424 | 434 | −0.01 (−0.06, 0.04) | 0.05 |
| Ghana | GHANA (39) | | | | |
| Ghana | iLiNS−DYADG (40) | 347 | 692 | −0.01 (−0.05, 0.02) | 0.11 |
| Malawi | iLiNS−DYADM (44) | 220 | 444 | −0.01 (−0.04, 0.02) | 0.12 |
| Malawi | iLiNS−DOSE (45) | 690 | 240 | 0.01 (−0.03, 0.04) | 0.10 |
| Mali | PROMIS (46) | 499 | 502 | −0.01 (−0.03, 0.01) | 0.23 |
| Mali | PROMIS CS (46) | 944 | 959 | −0.01 (−0.04, 0.01) | 0.13 |
| **$I^2$ = 0.35, $Tau^2$ = 0.00** | | **5921** | **4836** | **−0.02 (−0.03, 0.00)** | |

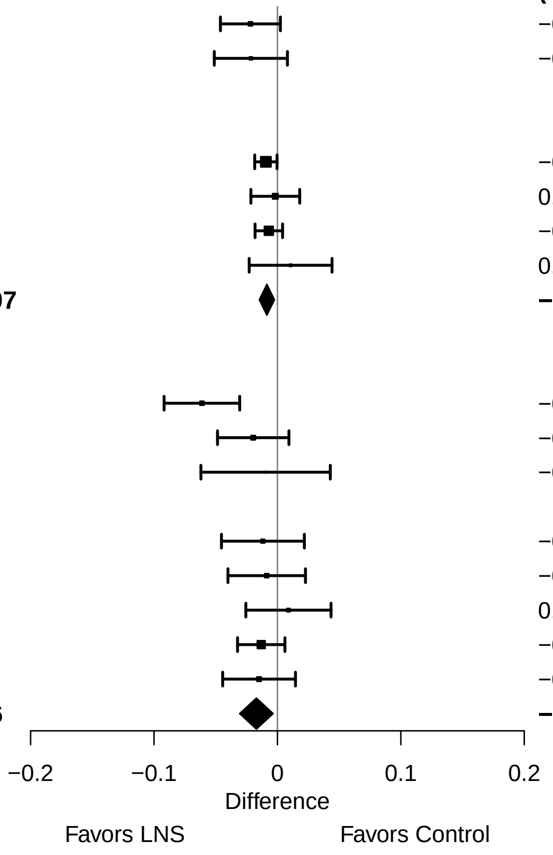

#### Supplemental figure 4K: Acute malnutrition prevalence difference

### 4K4: Stratified by Source water quality

**Source water quality**
**(p−diff = 0.852)**

| Country | Trial | N | N | PD (95% CI) | W |
|---|---|---|---|---|---|
| **Source water quality − Improved** | | | | | |
| Bangladesh | JiVitA−4 (34) | 2783 | 1216 | −0.02 (−0.05, 0.00) | 0.30 |
| Bangladesh | RDNS (35) | 1661 | 815 | −0.02 (−0.05, 0.01) | 0.20 |
| Bangladesh | WASH−B (36) | | | | |
| Ghana | GHANA (39) | | | | |
| Ghana | iLiNS−DYADG (40) | 347 | 692 | −0.01 (−0.05, 0.02) | 0.16 |
| Haiti | HAITI (41) | | | | |
| Malawi | iLiNS−DYADM (44) | 220 | 444 | −0.01 (−0.04, 0.02) | 0.18 |
| Malawi | iLiNS−DOSE (45) | 690 | 240 | 0.01 (−0.03, 0.04) | 0.15 |
| **I² = 0.00, Tau² = 0.00** | | **5701** | **3407** | **−0.01 (−0.03, 0.00)** | |
| **Source water quality − Unimproved** | | | | | |
| Burkina Faso | iLiNS−Zinc (37) | 1951 | 664 | −0.06 (−0.09, −0.03) | 0.06 |
| Burkina Faso | PROMIS (38) | 846 | 901 | −0.02 (−0.05, 0.01) | 0.06 |
| Burkina Faso | PROMIS CS (38) | 424 | 434 | −0.01 (−0.06, 0.04) | 0.02 |
| Kenya | WASH−B (42) | 694 | 2244 | −0.01 (−0.02, 0.01) | 0.32 |
| Madagascar | MAHAY (43) | 1675 | 1655 | 0.00 (−0.02, 0.02) | 0.13 |
| Mali | PROMIS (46) | 499 | 502 | −0.01 (−0.03, 0.01) | 0.14 |
| Mali | PROMIS CS (46) | 944 | 959 | −0.01 (−0.04, 0.01) | 0.06 |
| Zimbabwe | SHINE (HIV−) (47) | 886 | 871 | −0.01 (−0.02, 0.01) | 0.19 |
| Zimbabwe | SHINE (HIV+) (48) | 147 | 146 | −0.01 (−0.06, 0.04) | 0.02 |
| **I² = 0.35, Tau² = 0.00** | | **8066** | **8376** | **−0.01 (−0.02, 0.00)** | |

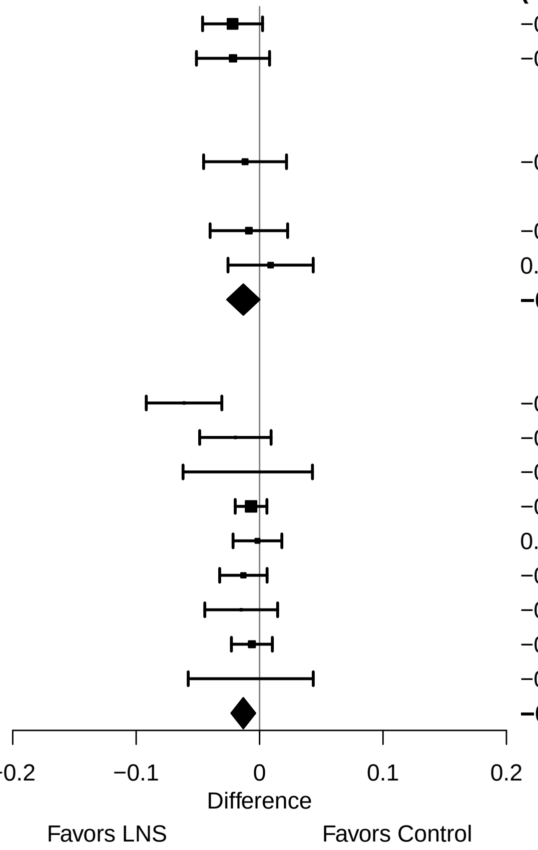

#### Supplemental figure 4K: Acute malnutrition prevalence difference

### 4K5: Stratified by Sanitation

**Sanitation**
**(p−diff = 0.263)**

| Country | Trial | N | N | PD (95% CI) | W |
|---|---|---|---|---|---|
| **Sanitation − Improved** | | | | | |
| Bangladesh | JiVitA−4 (34) | 2783 | 1216 | −0.02 (−0.05, 0.00) | 0.19 |
| Bangladesh | RDNS (35) | 1661 | 815 | −0.02 (−0.05, 0.01) | 0.12 |
| Bangladesh | WASH−B (36) | | | | |
| Burkina Faso | PROMIS (38) | 846 | 901 | −0.02 (−0.05, 0.01) | 0.13 |
| Burkina Faso | PROMIS CS (38) | 424 | 434 | −0.01 (−0.06, 0.04) | 0.04 |
| Ghana | GHANA (39) | | | | |
| Ghana | iLiNS−DYADG (40) | 347 | 692 | −0.01 (−0.05, 0.02) | 0.10 |
| Haiti | HAITI (41) | | | | |
| Mali | PROMIS (46) | 499 | 502 | −0.01 (−0.03, 0.01) | 0.30 |
| Mali | PROMIS CS (46) | 944 | 959 | −0.01 (−0.04, 0.01) | 0.13 |
| **I² = 0.00, Tau² = 0.00** | | **7504** | **5519** | **−0.02 (−0.03, −0.01)** | |
| **Sanitation − Unimproved** | | | | | |
| Burkina Faso | iLiNS−Zinc (37) | 1951 | 664 | −0.06 (−0.09, −0.03) | 0.07 |
| Kenya | WASH−B (42) | 694 | 2244 | −0.01 (−0.02, 0.01) | 0.39 |
| Madagascar | MAHAY (43) | 1675 | 1655 | 0.00 (−0.02, 0.02) | 0.16 |
| Malawi | iLiNS−DYADM (44) | 220 | 444 | −0.01 (−0.04, 0.02) | 0.07 |
| Malawi | iLiNS−DOSE (45) | 690 | 240 | 0.01 (−0.03, 0.04) | 0.05 |
| Zimbabwe | SHINE (HIV−) (47) | 886 | 871 | −0.01 (−0.02, 0.01) | 0.23 |
| Zimbabwe | SHINE (HIV+) (48) | 147 | 146 | −0.01 (−0.06, 0.04) | 0.03 |
| **I² = 0.54, Tau² = 0.00** | | **6263** | **6264** | **−0.01 (−0.03, 0.00)** | |

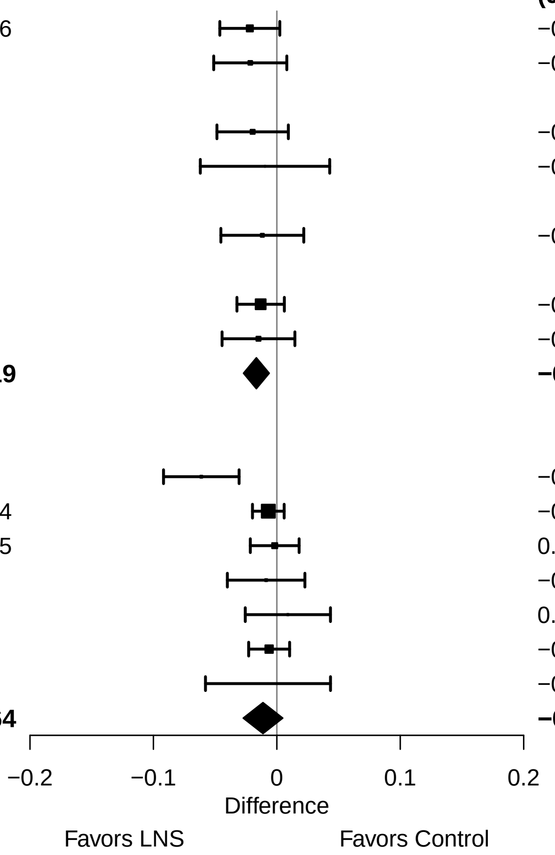

##### Supplemental figure 4K: Acute malnutrition prevalence difference

#### 4K6: Stratified by Supplement duration

**Supplement duration**
**(p−diff = 0.887)**

| Country | Trial | N | N | PD (95% CI) | W |
|---|---|---|---|---|---|
| **Supplement duration − 12m or less** | | | | | |
| Bangladesh | JiVitA−4 (34) | 2783 | 1216 | −0.02 (−0.05, 0.00) | 0.11 |
| Burkina Faso | iLiNS−Zinc (37) | 1951 | 664 | −0.06 (−0.09, −0.03) | 0.08 |
| Burkina Faso | PROMIS (38) | 846 | 901 | −0.02 (−0.05, 0.01) | 0.09 |
| Burkina Faso | PROMIS CS (38) | 424 | 434 | −0.01 (−0.06, 0.04) | 0.03 |
| Ghana | GHANA (39) | | | | |
| Ghana | iLiNS−DYADG (40) | 347 | 692 | −0.01 (−0.05, 0.02) | 0.07 |
| Haiti | HAITI (41) | | | | |
| Madagascar | MAHAY (43) | 1675 | 1655 | 0.00 (−0.02, 0.02) | 0.15 |
| Malawi | iLiNS−DYADM (44) | 220 | 444 | −0.01 (−0.04, 0.02) | 0.08 |
| Malawi | iLiNS−DOSE (45) | 690 | 240 | 0.01 (−0.03, 0.04) | 0.07 |
| Zimbabwe | SHINE (HIV−) (47) | 1867 | 1777 | −0.01 (−0.02, 0.00) | 0.25 |
| Zimbabwe | SHINE (HIV+) (48) | 336 | 327 | 0.01 (−0.02, 0.04) | 0.07 |
| **I² = 0.43, Tau² = 0.00** | | **11139** | **8350** | **−0.01 (−0.02, 0.00)** | |
| **Supplement duration − > 12m** | | | | | |
| Bangladesh | RDNS (35) | 1661 | 815 | −0.02 (−0.05, 0.01) | 0.14 |
| Bangladesh | WASH−B (36) | | | | |
| Kenya | WASH−B (42) | 1454 | 5117 | −0.01 (−0.02, 0.00) | 0.46 |
| Mali | PROMIS (46) | 499 | 502 | −0.01 (−0.03, 0.01) | 0.25 |
| Mali | PROMIS CS (46) | 944 | 959 | −0.01 (−0.04, 0.01) | 0.14 |
| **I² = 0.00, Tau² = 0.00** | | **4558** | **7393** | **−0.01 (−0.02, 0.00)** | |

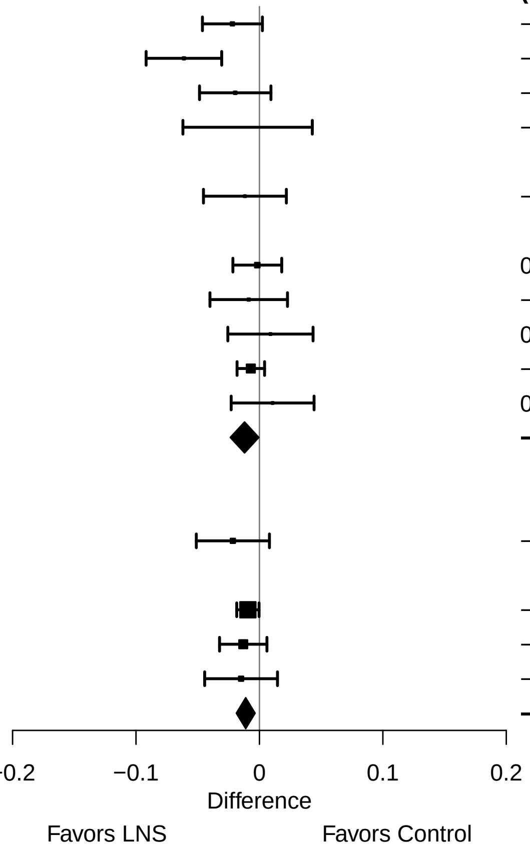

**Supplemental figure 4K: Acute malnutrition prevalence difference**

#### 4K7: Stratified by Frequency of contact

**Frequency of contact**
**(p−diff = 0.113)**

| Country | Trial | N | N | PD (95% CI) | W |
|---|---|---|---|---|---|
| **Frequency of contact − Monthly** | | | | | |
| Bangladesh | RDNS (35) | 1661 | 815 | −0.02 (−0.05, 0.01) | 0.07 |
| Burkina Faso | PROMIS (38) | 846 | 901 | −0.02 (−0.05, 0.01) | 0.07 |
| Burkina Faso | PROMIS CS (38) | 424 | 434 | −0.01 (−0.06, 0.04) | 0.03 |
| Haiti | HAITI (41) | | | | |
| Kenya | WASH−B (42) | 1454 | 5117 | −0.01 (−0.02, 0.00) | 0.23 |
| Madagascar | MAHAY (43) | 1675 | 1655 | 0.00 (−0.02, 0.02) | 0.12 |
| Mali | PROMIS (46) | 499 | 502 | −0.01 (−0.03, 0.01) | 0.13 |
| Mali | PROMIS CS (46) | 944 | 959 | −0.01 (−0.04, 0.01) | 0.07 |
| Zimbabwe | SHINE (HIV−) (47) | 1867 | 1777 | −0.01 (−0.02, 0.00) | 0.21 |
| Zimbabwe | SHINE (HIV+) (48) | 336 | 327 | 0.01 (−0.02, 0.04) | 0.06 |
| **I² = 0.00, Tau² = 0.00** | | **9706** | **12487** | **−0.01 (−0.01, 0.00)** | |
| **Frequency of contact − Weekly** | | | | | |
| Bangladesh | JiVitA−4 (34) | 2783 | 1216 | −0.02 (−0.05, 0.00) | 0.28 |
| Bangladesh | WASH−B (36) | | | | |
| Burkina Faso | iLiNS−Zinc (37) | 1951 | 664 | −0.06 (−0.09, −0.03) | 0.20 |
| Ghana | GHANA (39) | | | | |
| Ghana | iLiNS−DYADG (40) | 347 | 692 | −0.01 (−0.05, 0.02) | 0.17 |
| Malawi | iLiNS−DYADM (44) | 220 | 444 | −0.01 (−0.04, 0.02) | 0.19 |
| Malawi | iLiNS−DOSE (45) | 690 | 240 | 0.01 (−0.03, 0.04) | 0.16 |
| **I² = 0.62, Tau² = 0.00** | | **5991** | **3256** | **−0.02 (−0.04, 0.00)** | |

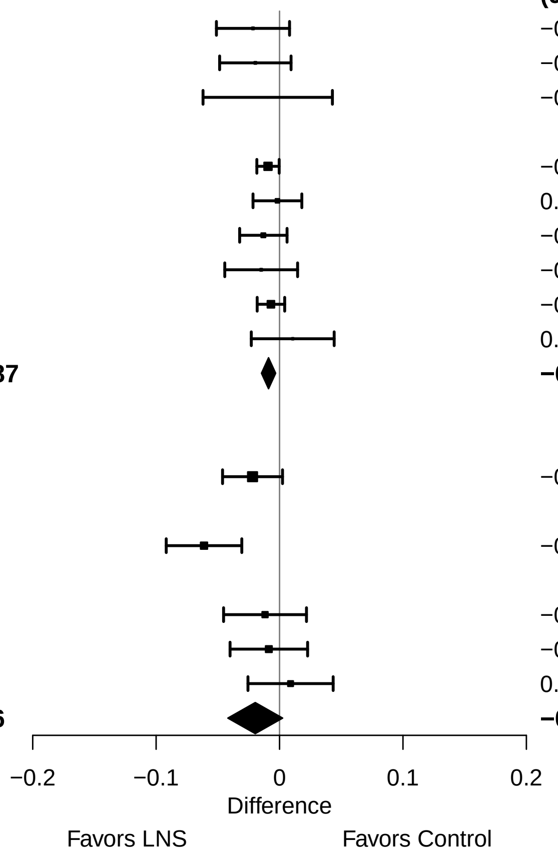

#### Supplemental figure 4K: Acute malnutrition prevalence difference

### 4K8: Stratified by Average SQ-LNS compliance

**Average SQ−LNS compliance**
**(p−diff = 0.129)**

| Country | Trial | N | N | PD (95% CI) | W |
|---|---|---|---|---|---|
| **Average SQ−LNS compliance − Low** | | | | | |
| Burkina Faso | PROMIS (38) | 846 | 901 | −0.02 (−0.05, 0.01) | 0.10 |
| Burkina Faso | PROMIS CS (38) | 424 | 434 | −0.01 (−0.06, 0.04) | 0.04 |
| Ghana | iLiNS−DYADG (40) | 347 | 692 | −0.01 (−0.05, 0.02) | 0.08 |
| Malawi | iLiNS−DYADM (44) | 220 | 444 | −0.01 (−0.04, 0.02) | 0.09 |
| Malawi | iLiNS−DOSE (45) | 690 | 240 | 0.01 (−0.03, 0.04) | 0.08 |
| Mali | PROMIS (46) | 499 | 502 | −0.01 (−0.03, 0.01) | 0.17 |
| Mali | PROMIS CS (46) | 944 | 959 | −0.01 (−0.04, 0.01) | 0.10 |
| Zimbabwe | SHINE (HIV−) (47) | 1867 | 1777 | −0.01 (−0.02, 0.00) | 0.25 |
| Zimbabwe | SHINE (HIV+) (48) | 336 | 327 | 0.01 (−0.02, 0.04) | 0.08 |
| **I² = 0.00, Tau² = 0.00** | | **6173** | **6276** | **−0.01 (−0.02, 0.00)** | |
| **Average SQ−LNS compliance − High** | | | | | |
| Bangladesh | JiVitA−4 (34) | 2783 | 1216 | −0.02 (−0.05, 0.00) | 0.22 |
| Bangladesh | RDNS (35) | 1661 | 815 | −0.02 (−0.05, 0.01) | 0.17 |
| Bangladesh | WASH−B (36) | | | | |
| Burkina Faso | iLiNS−Zinc (37) | 1951 | 664 | −0.06 (−0.09, −0.03) | 0.16 |
| Ghana | GHANA (39) | | | | |
| Haiti | HAITI (41) | | | | |
| Kenya | WASH−B (42) | 1454 | 5117 | −0.01 (−0.02, 0.00) | 0.45 |
| **I² = 0.72, Tau² = 0.00** | | **7849** | **7812** | **−0.03 (−0.05, 0.00)** | |

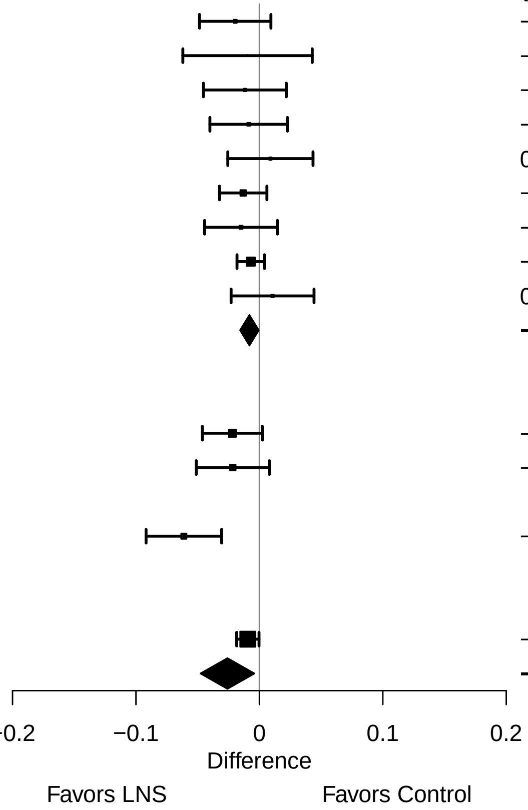

### Supplemental figure 4L: Mean difference in WAZ

#### 4L1: Stratified by Geographic region

**Geographic region**
**(p−diff = 0.791)**

| Country | Trial | N | N | MD (95% CI) | W |
|---|---|---|---|---|---|
| **Geographic region − SEAR** | | | | | |
| Bangladesh | JiVitA−4 (34) | 2814 | 1232 | 0.10 (0.07, 0.14) | 0.38 |
| Bangladesh | RDNS (35) | 1663 | 815 | 0.09 (0.01, 0.18) | 0.30 |
| Bangladesh | WASH−B (36) | 1163 | 3461 | 0.20 (0.13, 0.26) | 0.33 |
| **I² = 0.68, Tau² = 0.00** | | **5640** | **5508** | **0.13 (0.07, 0.19)** | |
| **Geographic region − AFR** | | | | | |
| Burkina Faso | iLiNS−Zinc (37) | 1960 | 666 | 0.28 (0.21, 0.35) | 0.11 |
| Burkina Faso | PROMIS (38) | 857 | 911 | 0.15 (0.03, 0.27) | 0.08 |
| Burkina Faso | PROMIS CS (38) | 430 | 436 | −0.05 (−0.19, 0.09) | 0.07 |
| Ghana | GHANA (39) | 98 | 96 | 0.28 (−0.05, 0.61) | 0.02 |
| Ghana | iLiNS−DYADG (40) | 347 | 693 | 0.16 (0.03, 0.29) | 0.07 |
| Kenya | WASH−B (42) | 1461 | 5165 | 0.13 (0.07, 0.20) | 0.11 |
| Madagascar | MAHAY (43) | 1702 | 1685 | 0.01 (−0.10, 0.13) | 0.08 |
| Malawi | iLiNS−DYADM (44) | 220 | 444 | −0.07 (−0.23, 0.10) | 0.06 |
| Malawi | iLiNS−DOSE (45) | 702 | 241 | 0.05 (−0.05, 0.16) | 0.09 |
| Mali | PROMIS (46) | 500 | 502 | 0.16 (0.05, 0.28) | 0.08 |
| Mali | PROMIS CS (46) | 946 | 960 | 0.23 (0.11, 0.36) | 0.08 |
| Zimbabwe | SHINE (HIV−) (47) | 1869 | 1786 | 0.13 (0.07, 0.20) | 0.11 |
| Zimbabwe | SHINE (HIV+) (48) | 336 | 328 | 0.01 (−0.16, 0.18) | 0.06 |
| **I² = 0.70, Tau² = 0.01** | | **11428** | **13913** | **0.12 (0.06, 0.17)** | |

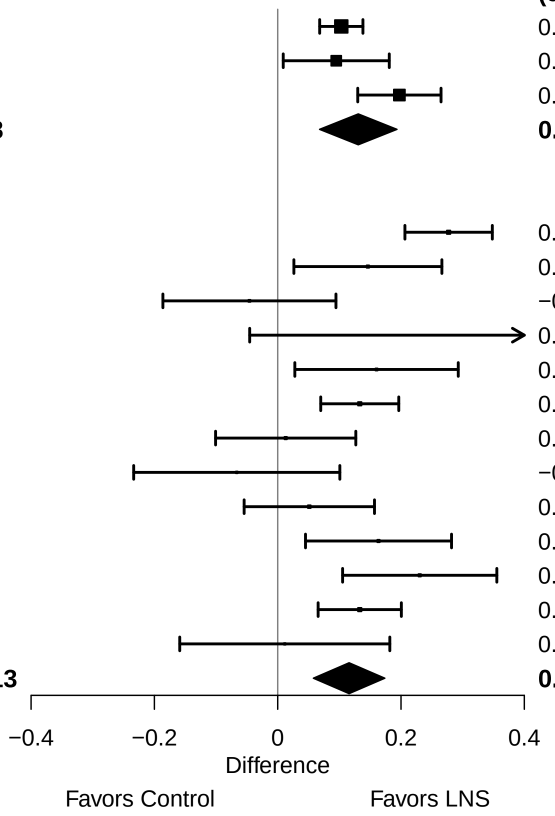

#### Supplemental figure 4L: Mean difference in WAZ

### 4L2: Stratified by Stunting burden

**Stunting burden**
**(p−diff = 0.292)**

| Country | Trial | N | N | MD (95% CI) | W |
|---|---|---|---|---|---|
| **Stunting burden − Less than 35%** | | | | | |
| Burkina Faso | PROMIS (38) | 857 | 911 | 0.15 (0.03, 0.27) | 0.16 |
| Burkina Faso | PROMIS CS (38) | 430 | 436 | −0.05 (−0.19, 0.09) | 0.14 |
| Ghana | GHANA (39) | 98 | 96 | 0.28 (−0.05, 0.61) | 0.04 |
| Ghana | iLiNS−DYADG (40) | 347 | 693 | 0.16 (0.03, 0.29) | 0.15 |
| Haiti | HAITI (41) | 148 | 150 | 0.06 (−0.06, 0.19) | 0.16 |
| Kenya | WASH−B (42) | 1461 | 5165 | 0.13 (0.07, 0.20) | 0.23 |
| Malawi | iLiNS−DYADM (44) | 220 | 444 | −0.07 (−0.23, 0.10) | 0.12 |
| **I² = 0.50, Tau² = 0.01** | | **3561** | **7895** | **0.09 (0.01, 0.16)** | |
| **Stunting burden − More than 35%** | | | | | |
| Bangladesh | JiVitA−4 (34) | 2814 | 1232 | 0.10 (0.07, 0.14) | 0.14 |
| Bangladesh | RDNS (35) | 1663 | 815 | 0.09 (0.01, 0.18) | 0.11 |
| Bangladesh | WASH−B (36) | 1163 | 3461 | 0.20 (0.13, 0.26) | 0.12 |
| Burkina Faso | iLiNS−Zinc (37) | 1960 | 666 | 0.28 (0.21, 0.35) | 0.12 |
| Madagascar | MAHAY (43) | 1702 | 1685 | 0.01 (−0.10, 0.13) | 0.09 |
| Malawi | iLiNS−DOSE (45) | 702 | 241 | 0.05 (−0.05, 0.16) | 0.09 |
| Mali | PROMIS (46) | 500 | 502 | 0.16 (0.05, 0.28) | 0.09 |
| Mali | PROMIS CS (46) | 946 | 960 | 0.23 (0.11, 0.36) | 0.08 |
| Zimbabwe | SHINE (HIV−) (47) | 1869 | 1786 | 0.13 (0.07, 0.20) | 0.12 |
| Zimbabwe | SHINE (HIV+) (48) | 336 | 328 | 0.01 (−0.16, 0.18) | 0.06 |
| **I² = 0.74, Tau² = 0.01** | | **13655** | **11676** | **0.13 (0.08, 0.19)** | |

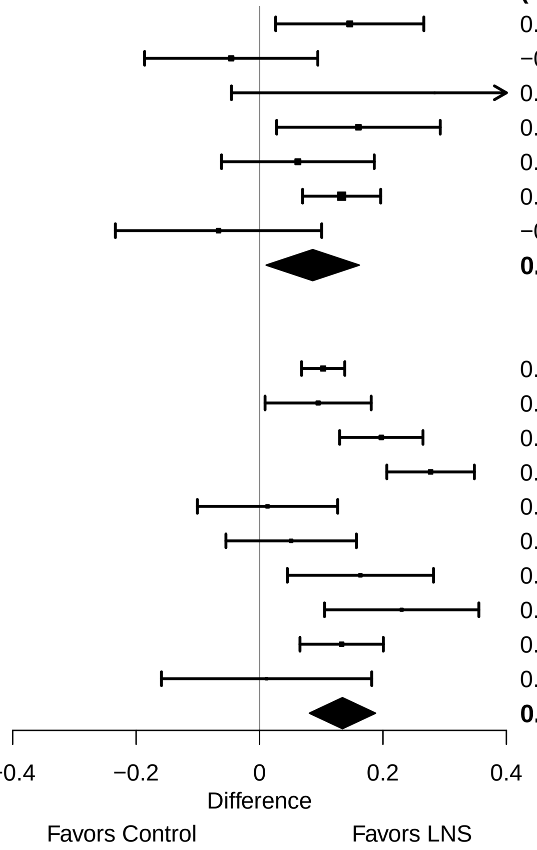

##### Supplemental figure 4L: Mean difference in WAZ

#### 4L3: Stratified by Malaria prevalence

**Malaria prevalence**
**(p−diff = 0.490)**

| Country | Trial | N | N | MD (95% CI) | W |
|---|---|---|---|---|---|
| **Malaria prevalence − Less than 10%** | | | | | |
| Bangladesh | JiVitA−4 (34) | 2814 | 1232 | 0.10 (0.07, 0.14) | 0.16 |
| Bangladesh | RDNS (35) | 1663 | 815 | 0.09 (0.01, 0.18) | 0.13 |
| Bangladesh | WASH−B (36) | 1163 | 3461 | 0.20 (0.13, 0.26) | 0.14 |
| Haiti | HAITI (41) | 148 | 150 | 0.06 (−0.06, 0.19) | 0.10 |
| Kenya | WASH−B (42) | 1461 | 5165 | 0.13 (0.07, 0.20) | 0.15 |
| Madagascar | MAHAY (43) | 1702 | 1685 | 0.01 (−0.10, 0.13) | 0.11 |
| Zimbabwe | SHINE (HIV−) (47) | 1869 | 1786 | 0.13 (0.07, 0.20) | 0.14 |
| Zimbabwe | SHINE (HIV+) (48) | 336 | 328 | 0.01 (−0.16, 0.18) | 0.07 |
| **I² = 0.42, Tau² = 0.00** | | **11156** | **14622** | **0.11 (0.07, 0.15)** | |
| **Malaria prevalence − At least 10%** | | | | | |
| Burkina Faso | iLiNS−Zinc (37) | 1960 | 666 | 0.28 (0.21, 0.35) | 0.17 |
| Burkina Faso | PROMIS (38) | 857 | 911 | 0.15 (0.03, 0.27) | 0.12 |
| Burkina Faso | PROMIS CS (38) | 430 | 436 | −0.05 (−0.19, 0.09) | 0.11 |
| Ghana | GHANA (39) | 98 | 96 | 0.28 (−0.05, 0.61) | 0.03 |
| Ghana | iLiNS−DYADG (40) | 347 | 693 | 0.16 (0.03, 0.29) | 0.11 |
| Malawi | iLiNS−DYADM (44) | 220 | 444 | −0.07 (−0.23, 0.10) | 0.09 |
| Malawi | iLiNS−DOSE (45) | 702 | 241 | 0.05 (−0.05, 0.16) | 0.13 |
| Mali | PROMIS (46) | 500 | 502 | 0.16 (0.05, 0.28) | 0.12 |
| Mali | PROMIS CS (46) | 946 | 960 | 0.23 (0.11, 0.36) | 0.12 |
| **I² = 0.75, Tau² = 0.01** | | **6060** | **4949** | **0.13 (0.05, 0.21)** | |

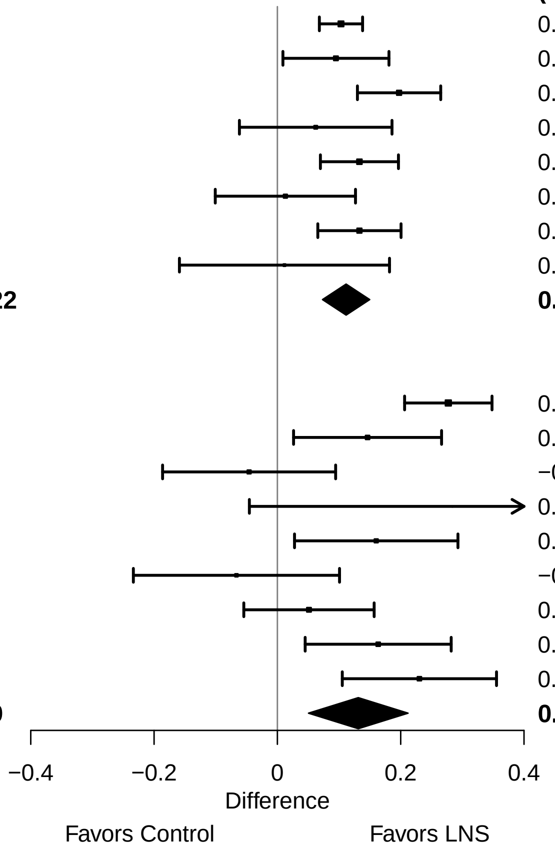

#### Supplemental figure 4L: Mean difference in WAZ

#### 4L4: Stratified by Source water quality

**Source water quality**
**(p−diff = 0.814)**

| Country | Trial | N | N | MD (95% CI) | W |
|---|---|---|---|---|---|
| **Source water quality − Improved** | | | | | |
| Bangladesh | JiVitA−4 (34) | 2814 | 1232 | 0.10 (0.07, 0.14) | 0.20 |
| Bangladesh | RDNS (35) | 1663 | 815 | 0.09 (0.01, 0.18) | 0.16 |
| Bangladesh | WASH−B (36) | 572 | 1119 | 0.25 (0.14, 0.37) | 0.13 |
| Ghana | GHANA (39) | 98 | 96 | 0.28 (−0.05, 0.61) | 0.04 |
| Ghana | iLiNS−DYADG (40) | 347 | 693 | 0.16 (0.03, 0.29) | 0.12 |
| Haiti | HAITI (41) | 148 | 150 | 0.06 (−0.06, 0.19) | 0.12 |
| Malawi | iLiNS−DYADM (44) | 220 | 444 | −0.07 (−0.23, 0.10) | 0.09 |
| Malawi | iLiNS−DOSE (45) | 702 | 241 | 0.05 (−0.05, 0.16) | 0.14 |
| **I² = 0.49, Tau² = 0.00** | | **6564** | **4790** | **0.11 (0.04, 0.17)** | |
| **Source water quality − Unimproved** | | | | | |
| Burkina Faso | iLiNS−Zinc (37) | 1960 | 666 | 0.28 (0.21, 0.35) | 0.15 |
| Burkina Faso | PROMIS (38) | 857 | 911 | 0.15 (0.03, 0.27) | 0.11 |
| Burkina Faso | PROMIS CS (38) | 430 | 436 | −0.05 (−0.19, 0.09) | 0.10 |
| Kenya | WASH−B (42) | 697 | 2261 | 0.09 (−0.01, 0.18) | 0.13 |
| Madagascar | MAHAY (43) | 1702 | 1685 | 0.01 (−0.10, 0.13) | 0.11 |
| Mali | PROMIS (46) | 500 | 502 | 0.16 (0.05, 0.28) | 0.11 |
| Mali | PROMIS CS (46) | 946 | 960 | 0.23 (0.11, 0.36) | 0.11 |
| Zimbabwe | SHINE (HIV−) (47) | 887 | 876 | 0.07 (−0.02, 0.16) | 0.13 |
| Zimbabwe | SHINE (HIV+) (48) | 147 | 146 | 0.03 (−0.21, 0.26) | 0.05 |
| **I² = 0.75, Tau² = 0.01** | | **8126** | **8443** | **0.12 (0.05, 0.19)** | |

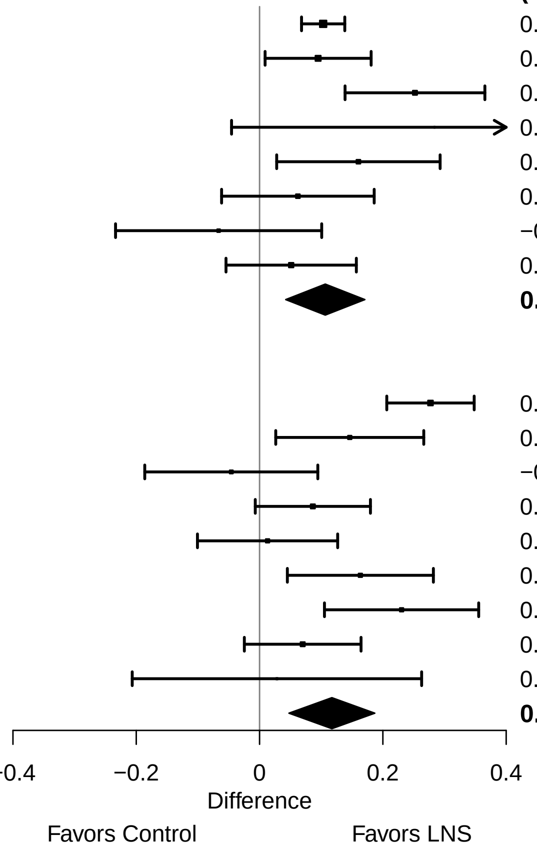

#### Supplemental figure 4L: Mean difference in WAZ

### 4L5: Stratified by Sanitation

**Sanitation**
**(p−diff = 0.301)**

| Country | Trial | N | N | MD (95% CI) | W |
|---|---|---|---|---|---|
| **Sanitation − Improved** | | | | | |
| Bangladesh | JiVitA−4 (34) | 2814 | 1232 | 0.10 (0.07, 0.14) | 0.16 |
| Bangladesh | RDNS (35) | 1663 | 815 | 0.09 (0.01, 0.18) | 0.13 |
| Bangladesh | WASH−B (36) | 572 | 1119 | 0.25 (0.14, 0.37) | 0.11 |
| Burkina Faso | PROMIS (38) | 857 | 911 | 0.15 (0.03, 0.27) | 0.10 |
| Burkina Faso | PROMIS CS (38) | 430 | 436 | −0.05 (−0.19, 0.09) | 0.09 |
| Ghana | GHANA (39) | 98 | 96 | 0.28 (−0.05, 0.61) | 0.03 |
| Ghana | iLiNS−DYADG (40) | 347 | 693 | 0.16 (0.03, 0.29) | 0.09 |
| Haiti | HAITI (41) | 148 | 150 | 0.06 (−0.06, 0.19) | 0.10 |
| Mali | PROMIS (46) | 500 | 502 | 0.16 (0.05, 0.28) | 0.10 |
| Mali | PROMIS CS (46) | 946 | 960 | 0.23 (0.11, 0.36) | 0.10 |
| **I² = 0.49, Tau² = 0.00** | | **8375** | **6914** | **0.13 (0.08, 0.19)** | |
| **Sanitation − Unimproved** | | | | | |
| Burkina Faso | iLiNS−Zinc (37) | 1960 | 666 | 0.28 (0.21, 0.35) | 0.19 |
| Kenya | WASH−B (42) | 697 | 2261 | 0.09 (−0.01, 0.18) | 0.17 |
| Madagascar | MAHAY (43) | 1702 | 1685 | 0.01 (−0.10, 0.13) | 0.15 |
| Malawi | iLiNS−DYADM (44) | 220 | 444 | −0.07 (−0.23, 0.10) | 0.10 |
| Malawi | iLiNS−DOSE (45) | 702 | 241 | 0.05 (−0.05, 0.16) | 0.16 |
| Zimbabwe | SHINE (HIV−) (47) | 887 | 876 | 0.07 (−0.02, 0.16) | 0.17 |
| Zimbabwe | SHINE (HIV+) (48) | 147 | 146 | 0.03 (−0.21, 0.26) | 0.07 |
| **I² = 0.81, Tau² = 0.01** | | **6315** | **6319** | **0.08 (0.00, 0.16)** | |

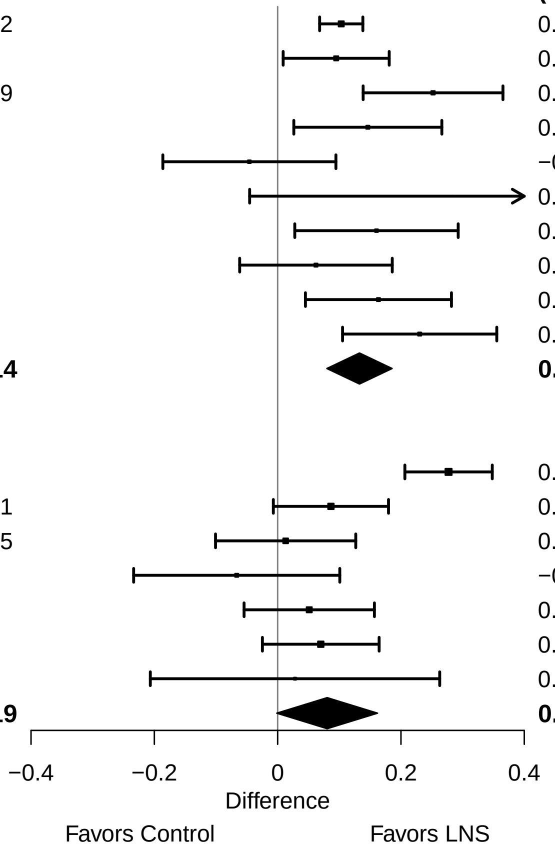

#### Supplemental figure 4L: Mean difference in WAZ

##### 4L6: Stratified by Supplement duration

**Supplement duration**
**(p−diff = 0.165)**

| Country | Trial | N | N | MD (95% CI) | W |
|---|---|---|---|---|---|
| **Supplement duration − 12m or less** | | | | | |
| Bangladesh | JiVitA−4 (34) | 2814 | 1232 | 0.10 (0.07, 0.14) | 0.13 |
| Burkina Faso | iLiNS−Zinc (37) | 1960 | 666 | 0.28 (0.21, 0.35) | 0.11 |
| Burkina Faso | PROMIS (38) | 857 | 911 | 0.15 (0.03, 0.27) | 0.08 |
| Burkina Faso | PROMIS CS (38) | 430 | 436 | −0.05 (−0.19, 0.09) | 0.07 |
| Ghana | GHANA (39) | 98 | 96 | 0.28 (−0.05, 0.61) | 0.02 |
| Ghana | iLiNS−DYADG (40) | 347 | 693 | 0.16 (0.03, 0.29) | 0.08 |
| Haiti | HAITI (41) | 148 | 150 | 0.06 (−0.06, 0.19) | 0.08 |
| Madagascar | MAHAY (43) | 1702 | 1685 | 0.01 (−0.10, 0.13) | 0.09 |
| Malawi | iLiNS−DYADM (44) | 220 | 444 | −0.07 (−0.23, 0.10) | 0.06 |
| Malawi | iLiNS−DOSE (45) | 702 | 241 | 0.05 (−0.05, 0.16) | 0.09 |
| Zimbabwe | SHINE (HIV−) (47) | 1869 | 1786 | 0.13 (0.07, 0.20) | 0.12 |
| Zimbabwe | SHINE (HIV+) (48) | 336 | 328 | 0.01 (−0.16, 0.18) | 0.06 |
| **I² = 0.72, Tau² = 0.01** | | **11483** | **8668** | **0.09 (0.04, 0.15)** | |
| **Supplement duration − > 12m** | | | | | |
| Bangladesh | RDNS (35) | 1663 | 815 | 0.09 (0.01, 0.18) | 0.21 |
| Bangladesh | WASH−B (36) | 1163 | 3461 | 0.20 (0.13, 0.26) | 0.23 |
| Kenya | WASH−B (42) | 1461 | 5165 | 0.13 (0.07, 0.20) | 0.24 |
| Mali | PROMIS (46) | 500 | 502 | 0.16 (0.05, 0.28) | 0.17 |
| Mali | PROMIS CS (46) | 946 | 960 | 0.23 (0.11, 0.36) | 0.16 |
| **I² = 0.24, Tau² = 0.00** | | **5733** | **10903** | **0.16 (0.11, 0.20)** | |

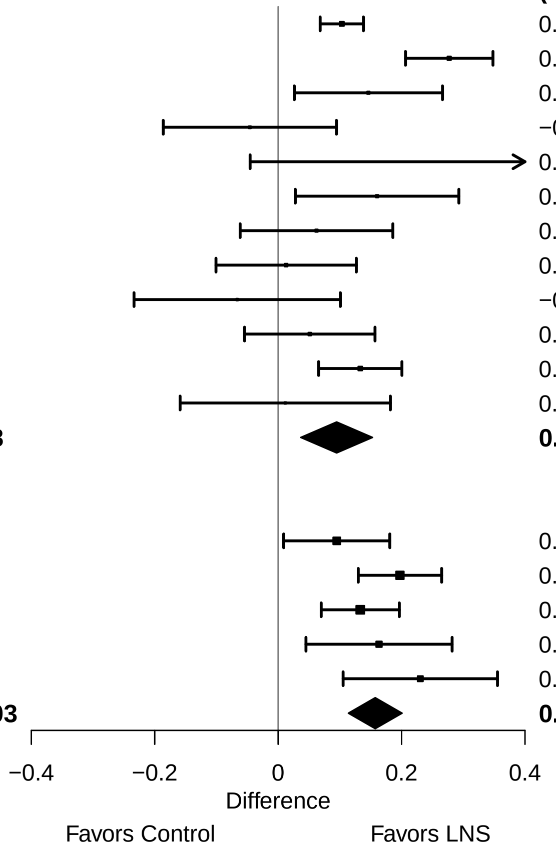

#### Supplemental figure 4L: Mean difference in WAZ

### 4L7: Stratified by Frequency of contact

**Frequency of contact**
**(p−diff = 0.339)**

| Country | Trial | N | N | MD (95% CI) | W |
|---|---|---|---|---|---|
| **Frequency of contact − Monthly** | | | | | |
| Bangladesh | RDNS (35) | 1663 | 815 | 0.09 (0.01, 0.18) | 0.12 |
| Burkina Faso | PROMIS (38) | 857 | 911 | 0.15 (0.03, 0.27) | 0.09 |
| Burkina Faso | PROMIS CS (38) | 430 | 436 | −0.05 (−0.19, 0.09) | 0.08 |
| Haiti | HAITI (41) | 148 | 150 | 0.06 (−0.06, 0.19) | 0.09 |
| Kenya | WASH−B (42) | 1461 | 5165 | 0.13 (0.07, 0.20) | 0.13 |
| Madagascar | MAHAY (43) | 1702 | 1685 | 0.01 (−0.10, 0.13) | 0.10 |
| Mali | PROMIS (46) | 500 | 502 | 0.16 (0.05, 0.28) | 0.09 |
| Mali | PROMIS CS (46) | 946 | 960 | 0.23 (0.11, 0.36) | 0.09 |
| Zimbabwe | SHINE (HIV−) (47) | 1869 | 1786 | 0.13 (0.07, 0.20) | 0.13 |
| Zimbabwe | SHINE (HIV+) (48) | 336 | 328 | 0.01 (−0.16, 0.18) | 0.07 |
| **$I^2$ = 0.41, $Tau^2$ = 0.00** | | **9912** | **12738** | **0.10 (0.06, 0.15)** | |
| **Frequency of contact − Weekly** | | | | | |
| Bangladesh | JiVitA−4 (34) | 2814 | 1232 | 0.10 (0.07, 0.14) | 0.22 |
| Bangladesh | WASH−B (36) | 1163 | 3461 | 0.20 (0.13, 0.26) | 0.19 |
| Burkina Faso | iLiNS−Zinc (37) | 1960 | 666 | 0.28 (0.21, 0.35) | 0.19 |
| Ghana | GHANA (39) | 98 | 96 | 0.28 (−0.05, 0.61) | 0.04 |
| Ghana | iLiNS−DYADG (40) | 347 | 693 | 0.16 (0.03, 0.29) | 0.12 |
| Malawi | iLiNS−DYADM (44) | 220 | 444 | −0.07 (−0.23, 0.10) | 0.10 |
| Malawi | iLiNS−DOSE (45) | 702 | 241 | 0.05 (−0.05, 0.16) | 0.15 |
| **$I^2$ = 0.81, $Tau^2$ = 0.01** | | **7304** | **6833** | **0.14 (0.06, 0.23)** | |

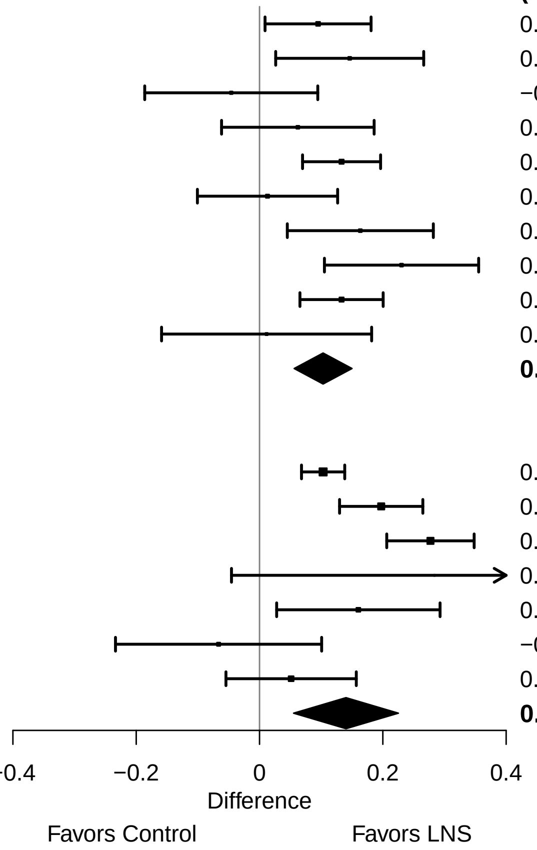

#### Supplemental figure 4L: Mean difference in WAZ

### 4L8: Stratified by Average SQ-LNS compliance

**Average SQ−LNS compliance**
**(p−diff = 0.210)**

**Average SQ−LNS compliance − Low**

| **Country** | **Trial** | **N** | **N** | **MD (95% CI)** | **W** |
|---|---|---|---|---|---|
| Burkina Faso | PROMIS (38) | 857 | 911 | 0.15 (0.03, 0.27) | 0.12 |
| Burkina Faso | PROMIS CS (38) | 430 | 436 | −0.05 (−0.19, 0.09) | 0.10 |
| Ghana | iLiNS−DYADG (40) | 347 | 693 | 0.16 (0.03, 0.29) | 0.11 |
| Malawi | iLiNS−DYADM (44) | 220 | 444 | −0.07 (−0.23, 0.10) | 0.08 |
| Malawi | iLiNS−DOSE (45) | 702 | 241 | 0.05 (−0.05, 0.16) | 0.13 |
| Mali | PROMIS (46) | 500 | 502 | 0.16 (0.05, 0.28) | 0.12 |
| Mali | PROMIS CS (46) | 946 | 960 | 0.23 (0.11, 0.36) | 0.11 |
| Zimbabwe | SHINE (HIV−) (47) | 1869 | 1786 | 0.13 (0.07, 0.20) | 0.16 |
| Zimbabwe | SHINE (HIV+) (48) | 336 | 328 | 0.01 (−0.16, 0.18) | 0.08 |
| **I² = 0.53, Tau² = 0.01** | | **6207** | **6301** | **0.10 (0.03, 0.16)** | |

**Average SQ−LNS compliance − High**

| **Country** | **Trial** | **N** | **N** | **MD (95% CI)** | **W** |
|---|---|---|---|---|---|
| Bangladesh | JiVitA−4 (34) | 2814 | 1232 | 0.10 (0.07, 0.14) | 0.20 |
| Bangladesh | RDNS (35) | 1663 | 815 | 0.09 (0.01, 0.18) | 0.15 |
| Bangladesh | WASH−B (36) | 1163 | 3461 | 0.20 (0.13, 0.26) | 0.17 |
| Burkina Faso | iLiNS−Zinc (37) | 1960 | 666 | 0.28 (0.21, 0.35) | 0.17 |
| Ghana | GHANA (39) | 98 | 96 | 0.28 (−0.05, 0.61) | 0.03 |
| Haiti | HAITI (41) | 148 | 150 | 0.06 (−0.06, 0.19) | 0.12 |
| Kenya | WASH−B (42) | 1461 | 5165 | 0.13 (0.07, 0.20) | 0.17 |
| **I² = 0.76, Tau² = 0.00** | | **9307** | **11585** | **0.15 (0.09, 0.21)** | |

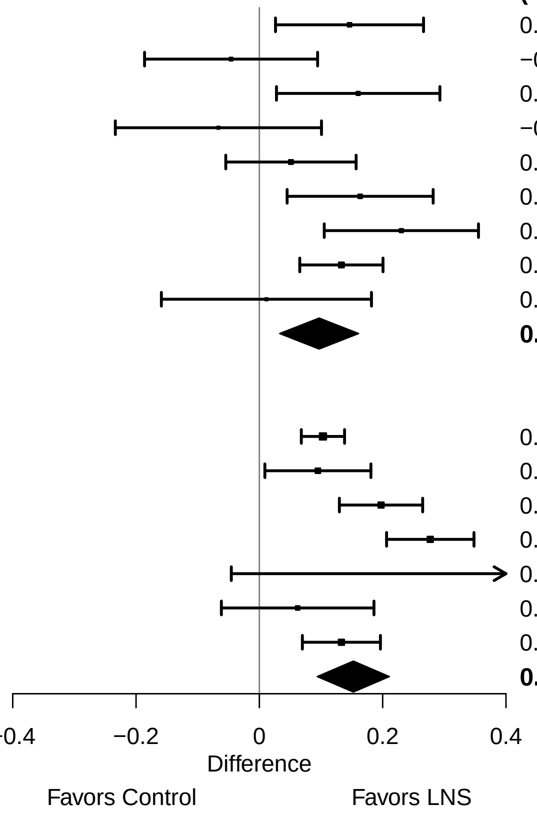

### Supplemental figure 4M: Underweight prevalence ratio

#### 4M1: Stratified by Geographic region

**Geographic region**
**(p−diff = 0.909)**

| Country | Trial | N | N | PR (95% CI) | W |
|---|---|---|---|---|---|
| **Geographic region − SEAR** | | | | | |
| Bangladesh | JiVitA−4 (34) | 2814 | 1232 | 0.91 (0.83, 1.01) | 0.35 |
| Bangladesh | RDNS (35) | 1663 | 815 | 0.92 (0.83, 1.02) | 0.34 |
| Bangladesh | WASH−B (36) | 1163 | 3461 | 0.79 (0.70, 0.89) | 0.31 |
| **I² = 0.57, Tau² = 0.00** | | **5640** | **5508** | **0.87 (0.79, 0.96)** | |
| **Geographic region − AFR** | | | | | |
| Burkina Faso | iLiNS−Zinc (37) | 1960 | 666 | 0.70 (0.59, 0.82) | 0.13 |
| Burkina Faso | PROMIS (38) | 857 | 911 | 0.86 (0.71, 1.05) | 0.10 |
| Burkina Faso | PROMIS CS (38) | 430 | 436 | 0.99 (0.77, 1.26) | 0.08 |
| Ghana | GHANA (39) | 98 | 96 | 0.71 (0.30, 1.69) | 0.01 |
| Ghana | iLiNS−DYADG (40) | 347 | 693 | 0.82 (0.57, 1.18) | 0.04 |
| Kenya | WASH−B (42) | 1461 | 5165 | 0.76 (0.62, 0.93) | 0.10 |
| Madagascar | MAHAY (43) | 1702 | 1685 | 1.04 (0.89, 1.21) | 0.13 |
| Malawi | iLiNS−DYADM (44) | 220 | 444 | 1.13 (0.74, 1.72) | 0.03 |
| Malawi | iLiNS−DOSE (45) | 702 | 241 | 1.01 (0.78, 1.31) | 0.07 |
| Mali | PROMIS (46) | 500 | 502 | 0.85 (0.62, 1.18) | 0.05 |
| Mali | PROMIS CS (46) | 946 | 960 | 0.88 (0.71, 1.10) | 0.09 |
| Zimbabwe | SHINE (HIV−) (47) | 1869 | 1786 | 0.71 (0.57, 0.87) | 0.10 |
| Zimbabwe | SHINE (HIV+) (48) | 336 | 328 | 1.01 (0.73, 1.40) | 0.05 |
| **I² = 0.47, Tau² = 0.01** | | **11428** | **13913** | **0.86 (0.79, 0.94)** | |

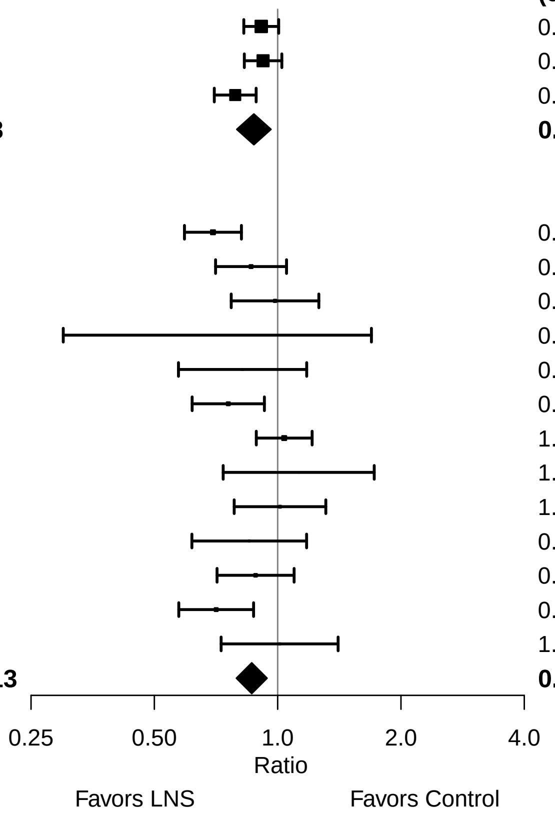

#### Supplemental figure 4M: Underweight prevalence ratio

### 4M2: Stratified by Stunting burden

**Stunting burden**
**(p−diff = 0.979)**

| Country | Trial | N | N | PR (95% CI) | W |
|---|---|---|---|---|---|
| **Stunting burden − Less than 35%** | | | | | |
| Burkina Faso | PROMIS (38) | 857 | 911 | 0.86 (0.71, 1.05) | 0.28 |
| Burkina Faso | PROMIS CS (38) | 430 | 436 | 0.99 (0.77, 1.26) | 0.21 |
| Ghana | GHANA (39) | 98 | 96 | 0.71 (0.30, 1.69) | 0.02 |
| Ghana | iLiNS−DYADG (40) | 347 | 693 | 0.82 (0.57, 1.18) | 0.11 |
| Haiti | HAITI (41) | 148 | 150 | 0.71 (0.28, 1.78) | 0.02 |
| Kenya | WASH−B (42) | 1461 | 5165 | 0.76 (0.62, 0.93) | 0.27 |
| Malawi | iLiNS−DYADM (44) | 220 | 444 | 1.13 (0.74, 1.72) | 0.09 |
| **I² = 0.00, Tau² = 0.00** | | **3561** | **7895** | **0.86 (0.77, 0.96)** | |
| **Stunting burden − More than 35%** | | | | | |
| Bangladesh | JiVitA−4 (34) | 2814 | 1232 | 0.91 (0.83, 1.01) | 0.17 |
| Bangladesh | RDNS (35) | 1663 | 815 | 0.92 (0.83, 1.02) | 0.16 |
| Bangladesh | WASH−B (36) | 1163 | 3461 | 0.79 (0.70, 0.89) | 0.15 |
| Burkina Faso | iLiNS−Zinc (37) | 1960 | 666 | 0.70 (0.59, 0.82) | 0.11 |
| Madagascar | MAHAY (43) | 1702 | 1685 | 1.04 (0.89, 1.21) | 0.11 |
| Malawi | iLiNS−DOSE (45) | 702 | 241 | 1.01 (0.78, 1.31) | 0.06 |
| Mali | PROMIS (46) | 500 | 502 | 0.85 (0.62, 1.18) | 0.04 |
| Mali | PROMIS CS (46) | 946 | 960 | 0.88 (0.71, 1.10) | 0.08 |
| Zimbabwe | SHINE (HIV−) (47) | 1869 | 1786 | 0.71 (0.57, 0.87) | 0.08 |
| Zimbabwe | SHINE (HIV+) (48) | 336 | 328 | 1.01 (0.73, 1.40) | 0.04 |
| **I² = 0.61, Tau² = 0.01** | | **13655** | **11676** | **0.87 (0.79, 0.95)** | |

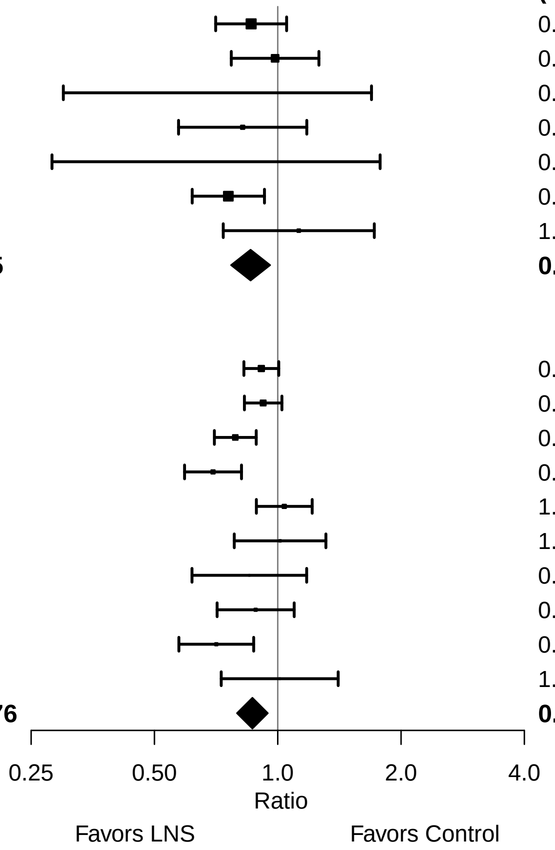

#### Supplemental figure 4M: Underweight prevalence ratio

### 4M3: Stratified by Malaria prevalence

**Malaria prevalence**
**(p−diff = 0.998)**

| Country | Trial | N | N | PR (95% CI) | W |
|---|---|---|---|---|---|
| **Malaria prevalence − Less than 10%** | | | | | |
| Bangladesh | JiVitA−4 (34) | 2814 | 1232 | 0.91 (0.83, 1.01) | 0.21 |
| Bangladesh | RDNS (35) | 1663 | 815 | 0.92 (0.83, 1.02) | 0.20 |
| Bangladesh | WASH−B (36) | 1163 | 3461 | 0.79 (0.70, 0.89) | 0.18 |
| Haiti | HAITI (41) | 148 | 150 | 0.71 (0.28, 1.78) | 0.01 |
| Kenya | WASH−B (42) | 1461 | 5165 | 0.76 (0.62, 0.93) | 0.11 |
| Madagascar | MAHAY (43) | 1702 | 1685 | 1.04 (0.89, 1.21) | 0.14 |
| Zimbabwe | SHINE (HIV−) (47) | 1869 | 1786 | 0.71 (0.57, 0.87) | 0.10 |
| Zimbabwe | SHINE (HIV+) (48) | 336 | 328 | 1.01 (0.73, 1.40) | 0.05 |
| **I² = 0.56, Tau² = 0.01** | | **11156** | **14622** | **0.87 (0.79, 0.95)** | |
| **Malaria prevalence − At least 10%** | | | | | |
| Burkina Faso | iLiNS−Zinc (37) | 1960 | 666 | 0.70 (0.59, 0.82) | 0.22 |
| Burkina Faso | PROMIS (38) | 857 | 911 | 0.86 (0.71, 1.05) | 0.17 |
| Burkina Faso | PROMIS CS (38) | 430 | 436 | 0.99 (0.77, 1.26) | 0.13 |
| Ghana | GHANA (39) | 98 | 96 | 0.71 (0.30, 1.69) | 0.01 |
| Ghana | iLiNS−DYADG (40) | 347 | 693 | 0.82 (0.57, 1.18) | 0.07 |
| Malawi | iLiNS−DYADM (44) | 220 | 444 | 1.13 (0.74, 1.72) | 0.05 |
| Malawi | iLiNS−DOSE (45) | 702 | 241 | 1.01 (0.78, 1.31) | 0.12 |
| Mali | PROMIS (46) | 500 | 502 | 0.85 (0.62, 1.18) | 0.08 |
| Mali | PROMIS CS (46) | 946 | 960 | 0.88 (0.71, 1.10) | 0.15 |
| **I² = 0.29, Tau² = 0.00** | | **6060** | **4949** | **0.86 (0.78, 0.95)** | |

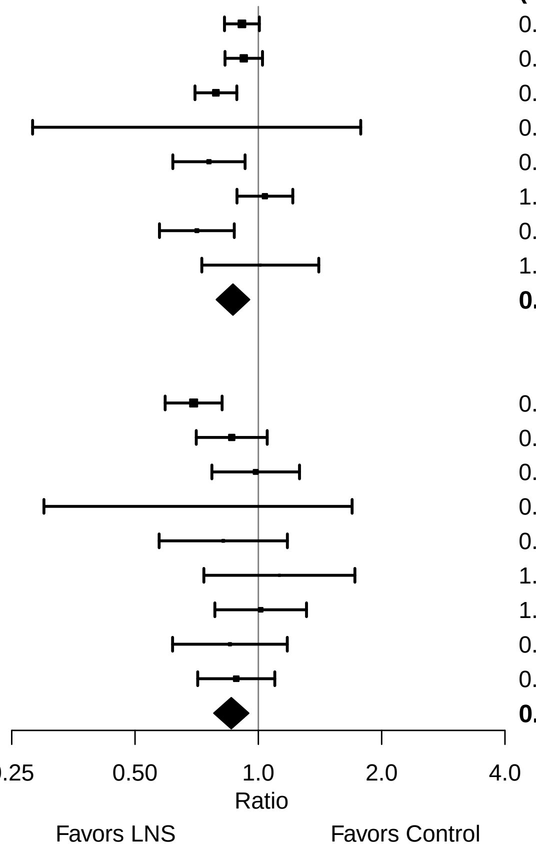

#### Supplemental figure 4M: Underweight prevalence ratio

### 4M4: Stratified by Source water quality

**Source water quality**
**(p−diff = 0.601)**

| Country | Trial | N | N | PR (95% CI) | W |
|---|---|---|---|---|---|
| **Source water quality − Improved** | | | | | |
| Bangladesh | JiVitA−4 (34) | 2814 | 1232 | 0.91 (0.83, 1.01) | 0.36 |
| Bangladesh | RDNS (35) | 1663 | 815 | 0.92 (0.83, 1.02) | 0.33 |
| Bangladesh | WASH−B (36) | 572 | 1119 | 0.85 (0.73, 1.00) | 0.17 |
| Ghana | GHANA (39) | 98 | 96 | 0.71 (0.30, 1.69) | 0.01 |
| Ghana | iLiNS−DYADG (40) | 347 | 693 | 0.82 (0.57, 1.18) | 0.04 |
| Haiti | HAITI (41) | 148 | 150 | 0.71 (0.28, 1.78) | 0.01 |
| Malawi | iLiNS−DYADM (44) | 220 | 444 | 1.13 (0.74, 1.72) | 0.03 |
| Malawi | iLiNS−DOSE (45) | 702 | 241 | 1.01 (0.78, 1.31) | 0.07 |
| **I² = 0.00, Tau² = 0.00** | | **6564** | **4790** | **0.91 (0.86, 0.97)** | |
| **Source water quality − Unimproved** | | | | | |
| Burkina Faso | iLiNS−Zinc (37) | 1960 | 666 | 0.70 (0.59, 0.82) | 0.21 |
| Burkina Faso | PROMIS (38) | 857 | 911 | 0.86 (0.71, 1.05) | 0.14 |
| Burkina Faso | PROMIS CS (38) | 430 | 436 | 0.99 (0.77, 1.26) | 0.10 |
| Kenya | WASH−B (42) | 697 | 2261 | 0.89 (0.65, 1.23) | 0.06 |
| Madagascar | MAHAY (43) | 1702 | 1685 | 1.04 (0.89, 1.21) | 0.22 |
| Mali | PROMIS (46) | 500 | 502 | 0.85 (0.62, 1.18) | 0.06 |
| Mali | PROMIS CS (46) | 946 | 960 | 0.88 (0.71, 1.10) | 0.12 |
| Zimbabwe | SHINE (HIV−) (47) | 887 | 876 | 0.86 (0.64, 1.14) | 0.07 |
| Zimbabwe | SHINE (HIV+) (48) | 147 | 146 | 1.03 (0.64, 1.69) | 0.03 |
| **I² = 0.43, Tau² = 0.01** | | **8126** | **8443** | **0.88 (0.80, 0.96)** | |

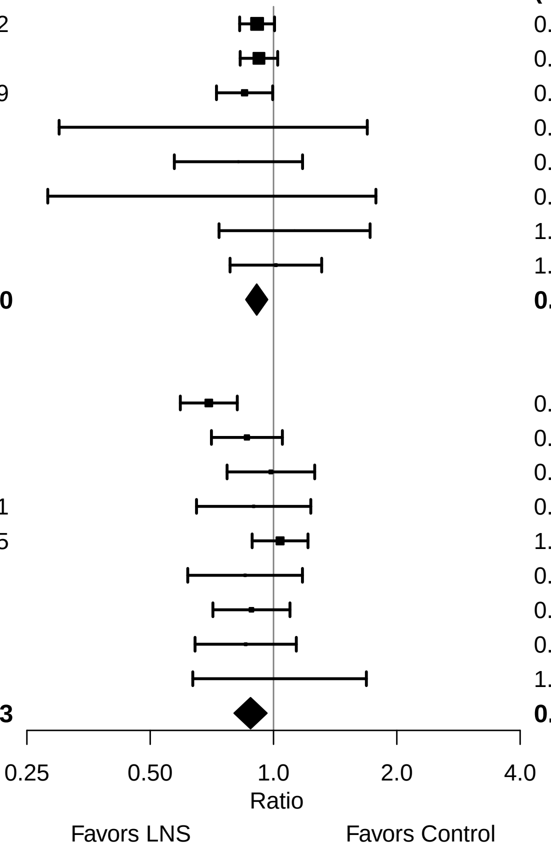

#### Supplemental figure 4M: Underweight prevalence ratio

### 4M5: Stratified by Sanitation

**Sanitation**
**(p−diff = 0.873)**

| Country | Trial | N | N | PR (95% CI) | W |
|---|---|---|---|---|---|
| **Sanitation − Improved** | | | | | |
| Bangladesh | JiVitA−4 (34) | 2814 | 1232 | 0.91 (0.83, 1.01) | 0.30 |
| Bangladesh | RDNS (35) | 1663 | 815 | 0.92 (0.83, 1.02) | 0.27 |
| Bangladesh | WASH−B (36) | 572 | 1119 | 0.85 (0.73, 1.00) | 0.14 |
| Burkina Faso | PROMIS (38) | 857 | 911 | 0.86 (0.71, 1.05) | 0.09 |
| Burkina Faso | PROMIS CS (38) | 430 | 436 | 0.99 (0.77, 1.26) | 0.06 |
| Ghana | GHANA (39) | 98 | 96 | 0.71 (0.30, 1.69) | 0.01 |
| Ghana | iLiNS−DYADG (40) | 347 | 693 | 0.82 (0.57, 1.18) | 0.03 |
| Haiti | HAITI (41) | 148 | 150 | 0.71 (0.28, 1.78) | 0.00 |
| Mali | PROMIS (46) | 500 | 502 | 0.85 (0.62, 1.18) | 0.04 |
| Mali | PROMIS CS (46) | 946 | 960 | 0.88 (0.71, 1.10) | 0.08 |
| **I² = 0.00, Tau² = 0.00** | | **8375** | **6914** | **0.90 (0.85, 0.95)** | |
| **Sanitation − Unimproved** | | | | | |
| Burkina Faso | iLiNS−Zinc (37) | 1960 | 666 | 0.70 (0.59, 0.82) | 0.30 |
| Kenya | WASH−B (42) | 697 | 2261 | 0.89 (0.65, 1.23) | 0.08 |
| Madagascar | MAHAY (43) | 1702 | 1685 | 1.04 (0.89, 1.21) | 0.31 |
| Malawi | iLiNS−DYADM (44) | 220 | 444 | 1.13 (0.74, 1.72) | 0.05 |
| Malawi | iLiNS−DOSE (45) | 702 | 241 | 1.01 (0.78, 1.31) | 0.12 |
| Zimbabwe | SHINE (HIV−) (47) | 887 | 876 | 0.86 (0.64, 1.14) | 0.10 |
| Zimbabwe | SHINE (HIV+) (48) | 147 | 146 | 1.03 (0.64, 1.69) | 0.04 |
| **I² = 0.61, Tau² = 0.01** | | **6315** | **6319** | **0.91 (0.80, 1.04)** | |

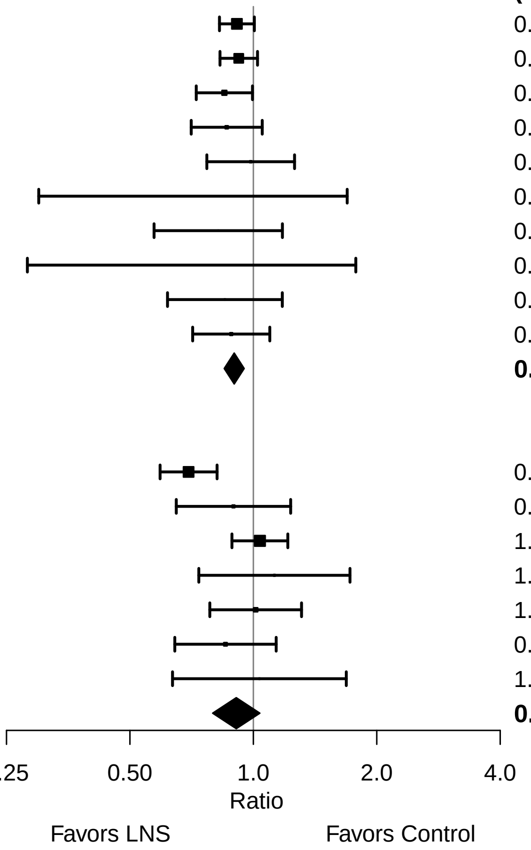

##### Supplemental figure 4M: Underweight prevalence ratio

#### 4M6: Stratified by Supplement duration

**Supplement duration**
**(p−diff = 0.519)**

| Country | Trial | N | N | PR (95% CI) | W |
|---|---|---|---|---|---|
| **Supplement duration − 12m or less** | | | | | |
| Bangladesh | JiVitA−4 (34) | 2814 | 1232 | 0.91 (0.83, 1.01) | 0.21 |
| Burkina Faso | iLiNS−Zinc (37) | 1960 | 666 | 0.70 (0.59, 0.82) | 0.14 |
| Burkina Faso | PROMIS (38) | 857 | 911 | 0.86 (0.71, 1.05) | 0.11 |
| Burkina Faso | PROMIS CS (38) | 430 | 436 | 0.99 (0.77, 1.26) | 0.08 |
| Ghana | GHANA (39) | 98 | 96 | 0.71 (0.30, 1.69) | 0.01 |
| Ghana | iLiNS−DYADG (40) | 347 | 693 | 0.82 (0.57, 1.18) | 0.04 |
| Haiti | HAITI (41) | 148 | 150 | 0.71 (0.28, 1.78) | 0.01 |
| Madagascar | MAHAY (43) | 1702 | 1685 | 1.04 (0.89, 1.21) | 0.14 |
| Malawi | iLiNS−DYADM (44) | 220 | 444 | 1.13 (0.74, 1.72) | 0.03 |
| Malawi | iLiNS−DOSE (45) | 702 | 241 | 1.01 (0.78, 1.31) | 0.08 |
| Zimbabwe | SHINE (HIV−) (47) | 1869 | 1786 | 0.71 (0.57, 0.87) | 0.10 |
| Zimbabwe | SHINE (HIV+) (48) | 336 | 328 | 1.01 (0.73, 1.40) | 0.05 |
| **$I^2$ = 0.49, $Tau^2$ = 0.01** | | **11483** | **8668** | **0.88 (0.81, 0.97)** | |
| **Supplement duration − > 12m** | | | | | |
| Bangladesh | RDNS (35) | 1663 | 815 | 0.92 (0.83, 1.02) | 0.31 |
| Bangladesh | WASH−B (36) | 1163 | 3461 | 0.79 (0.70, 0.89) | 0.29 |
| Kenya | WASH−B (42) | 1461 | 5165 | 0.76 (0.62, 0.93) | 0.16 |
| Mali | PROMIS (46) | 500 | 502 | 0.85 (0.62, 1.18) | 0.08 |
| Mali | PROMIS CS (46) | 946 | 960 | 0.88 (0.71, 1.10) | 0.15 |
| **$I^2$ = 0.23, $Tau^2$ = 0.00** | | **5733** | **10903** | **0.85 (0.78, 0.91)** | |

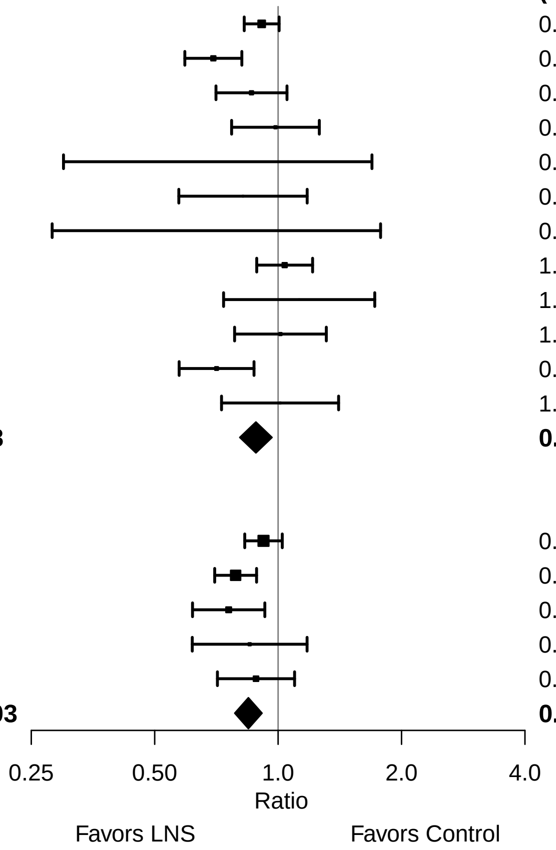

#### Supplemental figure 4M: Underweight prevalence ratio

### 4M7: Stratified by Frequency of contact

**Frequency of contact**
**(p−diff = 0.516)**

**Frequency of contact − Monthly**

| Country | Trial | N | N | PR (95% CI) | W |
|---|---|---|---|---|---|
| Bangladesh | RDNS (35) | 1663 | 815 | 0.92 (0.83, 1.02) | 0.21 |
| Burkina Faso | PROMIS (38) | 857 | 911 | 0.86 (0.71, 1.05) | 0.11 |
| Burkina Faso | PROMIS CS (38) | 430 | 436 | 0.99 (0.77, 1.26) | 0.09 |
| Haiti | HAITI (41) | 148 | 150 | 0.71 (0.28, 1.78) | 0.01 |
| Kenya | WASH−B (42) | 1461 | 5165 | 0.76 (0.62, 0.93) | 0.11 |
| Madagascar | MAHAY (43) | 1702 | 1685 | 1.04 (0.89, 1.21) | 0.15 |
| Mali | PROMIS (46) | 500 | 502 | 0.85 (0.62, 1.18) | 0.06 |
| Mali | PROMIS CS (46) | 946 | 960 | 0.88 (0.71, 1.10) | 0.10 |
| Zimbabwe | SHINE (HIV−) (47) | 1869 | 1786 | 0.71 (0.57, 0.87) | 0.11 |
| Zimbabwe | SHINE (HIV+) (48) | 336 | 328 | 1.01 (0.73, 1.40) | 0.05 |
| **I² = 0.29, Tau² = 0.00** | | **9912** | **12738** | **0.89 (0.82, 0.96)** | |

**Frequency of contact − Weekly**

| Country | Trial | N | N | PR (95% CI) | W |
|---|---|---|---|---|---|
| Bangladesh | JiVitA−4 (34) | 2814 | 1232 | 0.91 (0.83, 1.01) | 0.30 |
| Bangladesh | WASH−B (36) | 1163 | 3461 | 0.79 (0.70, 0.89) | 0.27 |
| Burkina Faso | iLiNS−Zinc (37) | 1960 | 666 | 0.70 (0.59, 0.82) | 0.20 |
| Ghana | GHANA (39) | 98 | 96 | 0.71 (0.30, 1.69) | 0.01 |
| Ghana | iLiNS−DYADG (40) | 347 | 693 | 0.82 (0.57, 1.18) | 0.06 |
| Malawi | iLiNS−DYADM (44) | 220 | 444 | 1.13 (0.74, 1.72) | 0.05 |
| Malawi | iLiNS−DOSE (45) | 702 | 241 | 1.01 (0.78, 1.31) | 0.11 |
| **I² = 0.55, Tau² = 0.01** | | **7304** | **6833** | **0.84 (0.75, 0.94)** | |

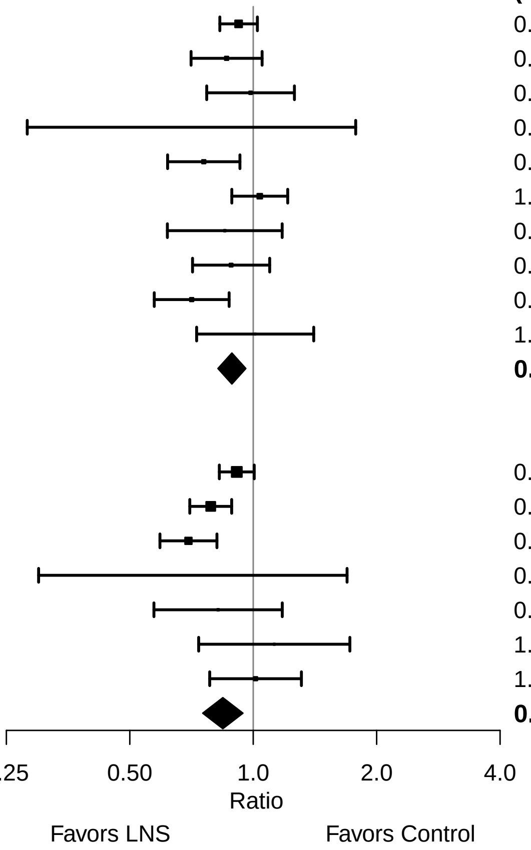

#### Supplemental figure 4M: Underweight prevalence ratio

### 4M8: Stratified by Average SQ-LNS compliance

**Average SQ−LNS compliance**
**(p−diff = 0.272)**

| Country | Trial | N | N | PR (95% CI) | W |
|---|---|---|---|---|---|
| **Average SQ−LNS compliance − Low** | | | | | |
| Burkina Faso | PROMIS (38) | 857 | 911 | 0.86 (0.71, 1.05) | 0.17 |
| Burkina Faso | PROMIS CS (38) | 430 | 436 | 0.99 (0.77, 1.26) | 0.13 |
| Ghana | iLiNS−DYADG (40) | 347 | 693 | 0.82 (0.57, 1.18) | 0.07 |
| Malawi | iLiNS−DYADM (44) | 220 | 444 | 1.13 (0.74, 1.72) | 0.05 |
| Malawi | iLiNS−DOSE (45) | 702 | 241 | 1.01 (0.78, 1.31) | 0.12 |
| Mali | PROMIS (46) | 500 | 502 | 0.85 (0.62, 1.18) | 0.08 |
| Mali | PROMIS CS (46) | 946 | 960 | 0.88 (0.71, 1.10) | 0.15 |
| Zimbabwe | SHINE (HIV−) (47) | 1869 | 1786 | 0.71 (0.57, 0.87) | 0.16 |
| Zimbabwe | SHINE (HIV+) (48) | 336 | 328 | 1.01 (0.73, 1.40) | 0.08 |
| **I² = 0.03, Tau² = 0.00** | | **6207** | **6301** | **0.88 (0.81, 0.97)** | |
| **Average SQ−LNS compliance − High** | | | | | |
| Bangladesh | JiVitA−4 (34) | 2814 | 1232 | 0.91 (0.83, 1.01) | 0.26 |
| Bangladesh | RDNS (35) | 1663 | 815 | 0.92 (0.83, 1.02) | 0.24 |
| Bangladesh | WASH−B (36) | 1163 | 3461 | 0.79 (0.70, 0.89) | 0.22 |
| Burkina Faso | iLiNS−Zinc (37) | 1960 | 666 | 0.70 (0.59, 0.82) | 0.15 |
| Ghana | GHANA (39) | 98 | 96 | 0.71 (0.30, 1.69) | 0.01 |
| Haiti | HAITI (41) | 148 | 150 | 0.71 (0.28, 1.78) | 0.01 |
| Kenya | WASH−B (42) | 1461 | 5165 | 0.76 (0.62, 0.93) | 0.11 |
| **I² = 0.55, Tau² = 0.01** | | **9307** | **11585** | **0.82 (0.76, 0.90)** | |

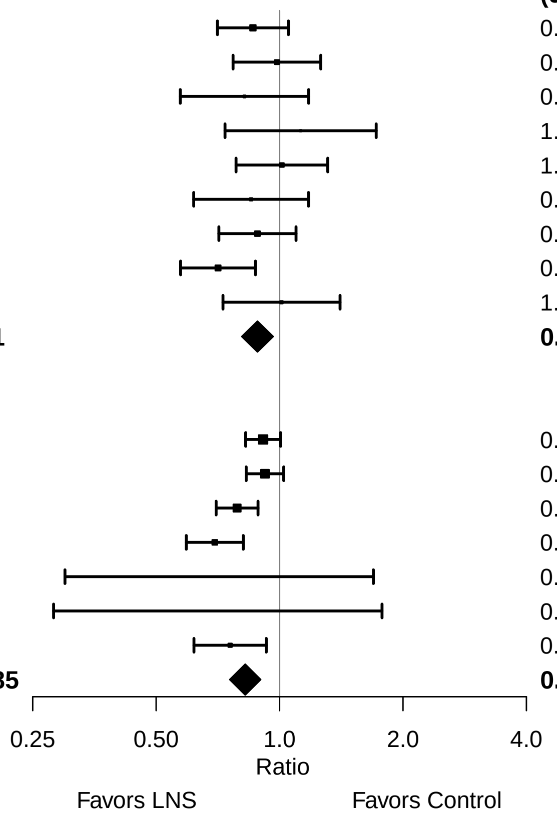

### Supplemental figure 4N: Underweight prevalence difference

#### 4N1: Stratified by Geographic region

**Geographic region**
**(p−diff = 0.198)**

| Country | Trial | N | N | PD (95% CI) | W |
|---|---|---|---|---|---|
| **Geographic region − SEAR** | | | | | |
| Bangladesh | JiVitA−4 (34) | 2814 | 1232 | −0.04 (−0.07, −0.02) | 0.38 |
| Bangladesh | RDNS (35) | 1663 | 815 | −0.03 (−0.07, 0.01) | 0.27 |
| Bangladesh | WASH−B (36) | 1163 | 3461 | −0.07 (−0.10, −0.04) | 0.35 |
| **I² = 0.31, Tau² = 0.00** | | **5640** | **5508** | **−0.05 (−0.07, −0.03)** | |
| **Geographic region − AFR** | | | | | |
| Burkina Faso | iLiNS−Zinc (37) | 1960 | 666 | −0.10 (−0.13, −0.07) | 0.09 |
| Burkina Faso | PROMIS (38) | 857 | 911 | −0.03 (−0.06, 0.01) | 0.08 |
| Burkina Faso | PROMIS CS (38) | 430 | 436 | 0.00 (−0.06, 0.05) | 0.05 |
| Ghana | GHANA (39) | 98 | 96 | −0.03 (−0.12, 0.05) | 0.03 |
| Ghana | iLiNS−DYADG (40) | 347 | 693 | −0.02 (−0.06, 0.02) | 0.07 |
| Kenya | WASH−B (42) | 1461 | 5165 | −0.02 (−0.04, −0.01) | 0.13 |
| Madagascar | MAHAY (43) | 1702 | 1685 | 0.01 (−0.02, 0.04) | 0.10 |
| Malawi | iLiNS−DYADM (44) | 220 | 444 | 0.01 (−0.04, 0.07) | 0.05 |
| Malawi | iLiNS−DOSE (45) | 702 | 241 | −0.02 (−0.07, 0.03) | 0.06 |
| Mali | PROMIS (46) | 500 | 502 | −0.04 (−0.08, 0.00) | 0.08 |
| Mali | PROMIS CS (46) | 946 | 960 | −0.03 (−0.06, 0.01) | 0.08 |
| Zimbabwe | SHINE (HIV−) (47) | 1869 | 1786 | −0.03 (−0.05, −0.01) | 0.12 |
| Zimbabwe | SHINE (HIV+) (48) | 336 | 328 | 0.00 (−0.06, 0.06) | 0.05 |
| **I² = 0.61, Tau² = 0.00** | | **11428** | **13913** | **−0.03 (−0.04, −0.01)** | |

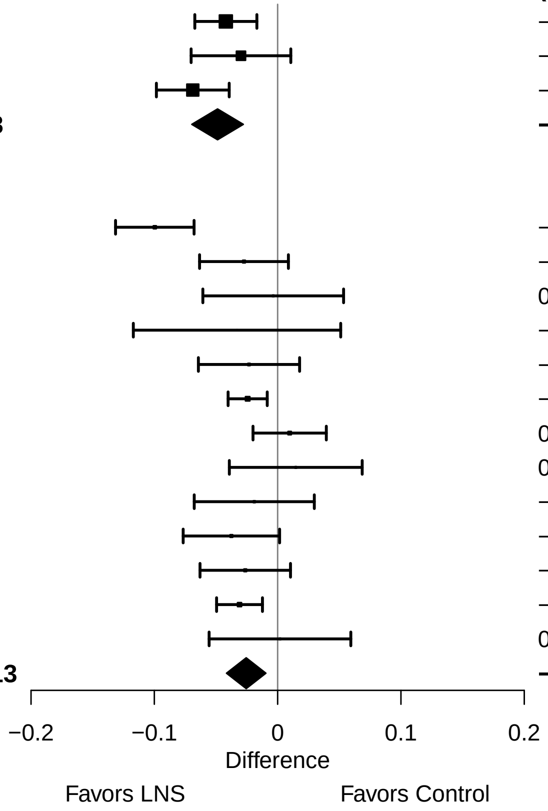

#### Supplemental figure 4N: Underweight prevalence difference

### 4N2: Stratified by Stunting burden

**Stunting burden**
**(p−diff = 0.149)**

| Country | Trial | N | N | PD (95% CI) | W |
|---|---|---|---|---|---|
| **Stunting burden − Less than 35%** | | | | | |
| Burkina Faso | PROMIS (38) | 857 | 911 | −0.03 (−0.06, 0.01) | 0.17 |
| Burkina Faso | PROMIS CS (38) | 430 | 436 | 0.00 (−0.06, 0.05) | 0.10 |
| Ghana | GHANA (39) | 98 | 96 | −0.03 (−0.12, 0.05) | 0.06 |
| Ghana | iLiNS−DYADG (40) | 347 | 693 | −0.02 (−0.06, 0.02) | 0.15 |
| Haiti | HAITI (41) | 148 | 150 | 0.00 (−0.05, 0.04) | 0.14 |
| Kenya | WASH−B (42) | 1461 | 5165 | −0.02 (−0.04, −0.01) | 0.26 |
| Malawi | iLiNS−DYADM (44) | 220 | 444 | 0.01 (−0.04, 0.07) | 0.11 |
| **I² = 0.00, Tau² = 0.00** | | **3561** | **7895** | **−0.02 (−0.03, −0.01)** | |
| **Stunting burden − More than 35%** | | | | | |
| Bangladesh | JiVitA−4 (34) | 2814 | 1232 | −0.04 (−0.07, −0.02) | 0.12 |
| Bangladesh | RDNS (35) | 1663 | 815 | −0.03 (−0.07, 0.01) | 0.09 |
| Bangladesh | WASH−B (36) | 1163 | 3461 | −0.07 (−0.10, −0.04) | 0.11 |
| Burkina Faso | iLiNS−Zinc (37) | 1960 | 666 | −0.10 (−0.13, −0.07) | 0.11 |
| Madagascar | MAHAY (43) | 1702 | 1685 | 0.01 (−0.02, 0.04) | 0.11 |
| Malawi | iLiNS−DOSE (45) | 702 | 241 | −0.02 (−0.07, 0.03) | 0.07 |
| Mali | PROMIS (46) | 500 | 502 | −0.04 (−0.08, 0.00) | 0.09 |
| Mali | PROMIS CS (46) | 946 | 960 | −0.03 (−0.06, 0.01) | 0.10 |
| Zimbabwe | SHINE (HIV−) (47) | 1869 | 1786 | −0.03 (−0.05, −0.01) | 0.14 |
| Zimbabwe | SHINE (HIV+) (48) | 336 | 328 | 0.00 (−0.06, 0.06) | 0.06 |
| **I² = 0.72, Tau² = 0.00** | | **13655** | **11676** | **−0.04 (−0.06, −0.02)** | |

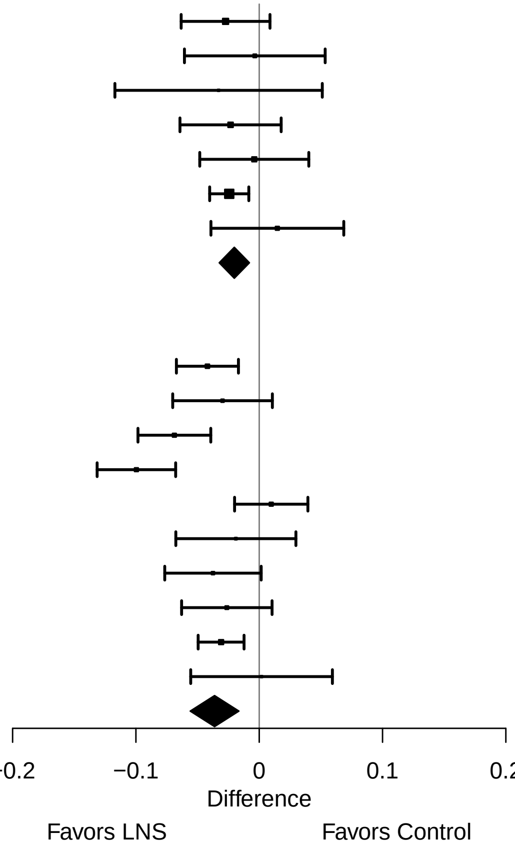

##### Supplemental figure 4N: Underweight prevalence difference

#### 4N3: Stratified by Malaria prevalence

**Malaria prevalence**
**(p−diff = 0.671)**

| Country | Trial | N | N | PD (95% CI) | W |
|---|---|---|---|---|---|
| **Malaria prevalence − Less than 10%** | | | | | |
| Bangladesh | JiVitA−4 (34) | 2814 | 1232 | −0.04 (−0.07, −0.02) | 0.14 |
| Bangladesh | RDNS (35) | 1663 | 815 | −0.03 (−0.07, 0.01) | 0.10 |
| Bangladesh | WASH−B (36) | 1163 | 3461 | −0.07 (−0.10, −0.04) | 0.13 |
| Haiti | HAITI (41) | 148 | 150 | 0.00 (−0.05, 0.04) | 0.09 |
| Kenya | WASH−B (42) | 1461 | 5165 | −0.02 (−0.04, −0.01) | 0.17 |
| Madagascar | MAHAY (43) | 1702 | 1685 | 0.01 (−0.02, 0.04) | 0.13 |
| Zimbabwe | SHINE (HIV−) (47) | 1869 | 1786 | −0.03 (−0.05, −0.01) | 0.16 |
| Zimbabwe | SHINE (HIV+) (48) | 336 | 328 | 0.00 (−0.06, 0.06) | 0.07 |
| **I² = 0.59, Tau² = 0.00** | | **11156** | **14622** | **−0.03 (−0.04, −0.01)** | |
| **Malaria prevalence − At least 10%** | | | | | |
| Burkina Faso | iLiNS−Zinc (37) | 1960 | 666 | −0.10 (−0.13, −0.07) | 0.15 |
| Burkina Faso | PROMIS (38) | 857 | 911 | −0.03 (−0.06, 0.01) | 0.14 |
| Burkina Faso | PROMIS CS (38) | 430 | 436 | 0.00 (−0.06, 0.05) | 0.08 |
| Ghana | GHANA (39) | 98 | 96 | −0.03 (−0.12, 0.05) | 0.05 |
| Ghana | iLiNS−DYADG (40) | 347 | 693 | −0.02 (−0.06, 0.02) | 0.12 |
| Malawi | iLiNS−DYADM (44) | 220 | 444 | 0.01 (−0.04, 0.07) | 0.09 |
| Malawi | iLiNS−DOSE (45) | 702 | 241 | −0.02 (−0.07, 0.03) | 0.10 |
| Mali | PROMIS (46) | 500 | 502 | −0.04 (−0.08, 0.00) | 0.13 |
| Mali | PROMIS CS (46) | 946 | 960 | −0.03 (−0.06, 0.01) | 0.14 |
| **I² = 0.62, Tau² = 0.00** | | **6060** | **4949** | **−0.03 (−0.05, −0.01)** | |

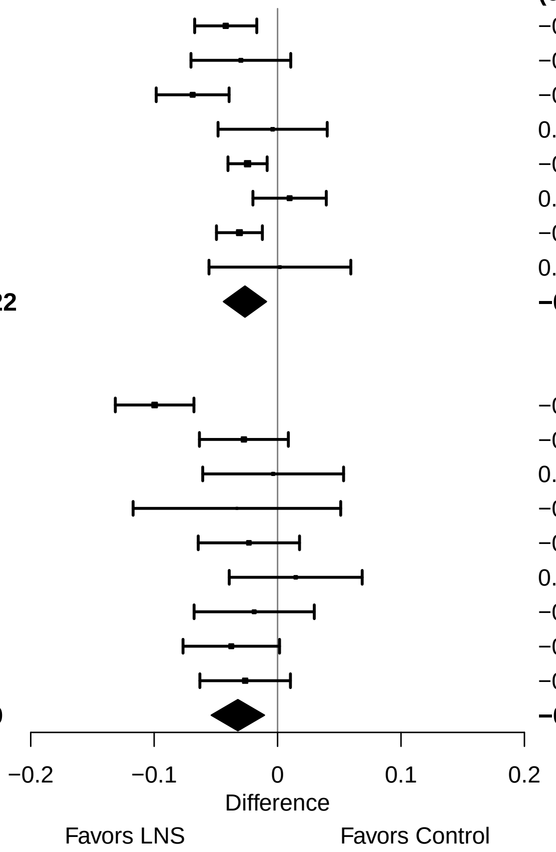

##### Supplemental figure 4N: Underweight prevalence difference

#### 4N4: Stratified by Source water quality

**Source water quality**
**(p−diff = 0.966)**

**Source water quality − Improved**

| Country | Trial | N | N | PD (95% CI) | W |
|---|---|---|---|---|---|
| Bangladesh | JiVitA−4 (34) | 2814 | 1232 | −0.04 (−0.07, −0.02) | 0.22 |
| Bangladesh | RDNS (35) | 1663 | 815 | −0.03 (−0.07, 0.01) | 0.14 |
| Bangladesh | WASH−B (36) | 572 | 1119 | −0.05 (−0.09, 0.00) | 0.12 |
| Ghana | GHANA (39) | 98 | 96 | −0.03 (−0.12, 0.05) | 0.05 |
| Ghana | iLiNS−DYADG (40) | 347 | 693 | −0.02 (−0.06, 0.02) | 0.14 |
| Haiti | HAITI (41) | 148 | 150 | 0.00 (−0.05, 0.04) | 0.13 |
| Malawi | iLiNS−DYADM (44) | 220 | 444 | 0.01 (−0.04, 0.07) | 0.10 |
| Malawi | iLiNS−DOSE (45) | 702 | 241 | −0.02 (−0.07, 0.03) | 0.11 |
| **I² = 0.00, Tau² = 0.00** | | **6564** | **4790** | **−0.03 (−0.04, −0.01)** | |

**Source water quality − Unimproved**

| Country | Trial | N | N | PD (95% CI) | W |
|---|---|---|---|---|---|
| Burkina Faso | iLiNS−Zinc (37) | 1960 | 666 | −0.10 (−0.13, −0.07) | 0.13 |
| Burkina Faso | PROMIS (38) | 857 | 911 | −0.03 (−0.06, 0.01) | 0.11 |
| Burkina Faso | PROMIS CS (38) | 430 | 436 | 0.00 (−0.06, 0.05) | 0.06 |
| Kenya | WASH−B (42) | 697 | 2261 | −0.01 (−0.03, 0.01) | 0.16 |
| Madagascar | MAHAY (43) | 1702 | 1685 | 0.01 (−0.02, 0.04) | 0.14 |
| Mali | PROMIS (46) | 500 | 502 | −0.04 (−0.08, 0.00) | 0.10 |
| Mali | PROMIS CS (46) | 946 | 960 | −0.03 (−0.06, 0.01) | 0.11 |
| Zimbabwe | SHINE (HIV−) (47) | 887 | 876 | −0.01 (−0.04, 0.01) | 0.15 |
| Zimbabwe | SHINE (HIV+) (48) | 147 | 146 | 0.01 (−0.08, 0.09) | 0.03 |
| **I² = 0.73, Tau² = 0.00** | | **8126** | **8443** | **−0.02 (−0.05, 0.00)** | |

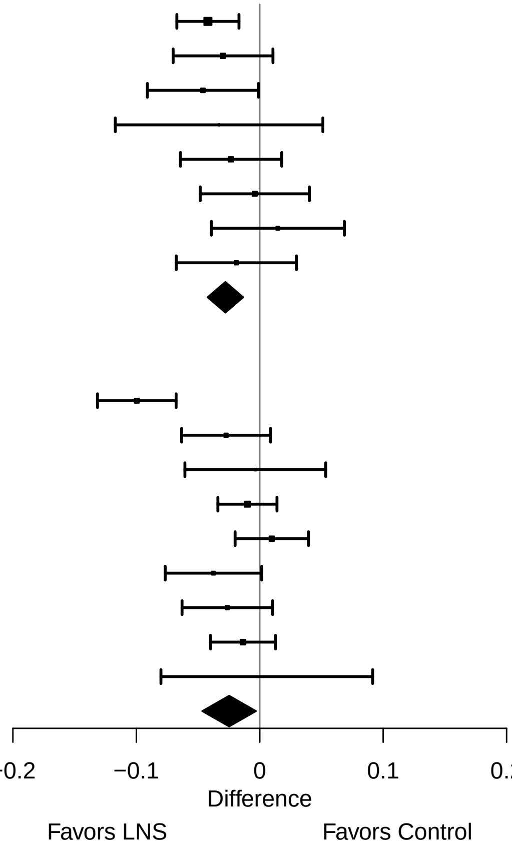

#### Supplemental figure 4N: Underweight prevalence difference

### 4N5: Stratified by Sanitation

**Sanitation**
**(p−diff = 0.573)**

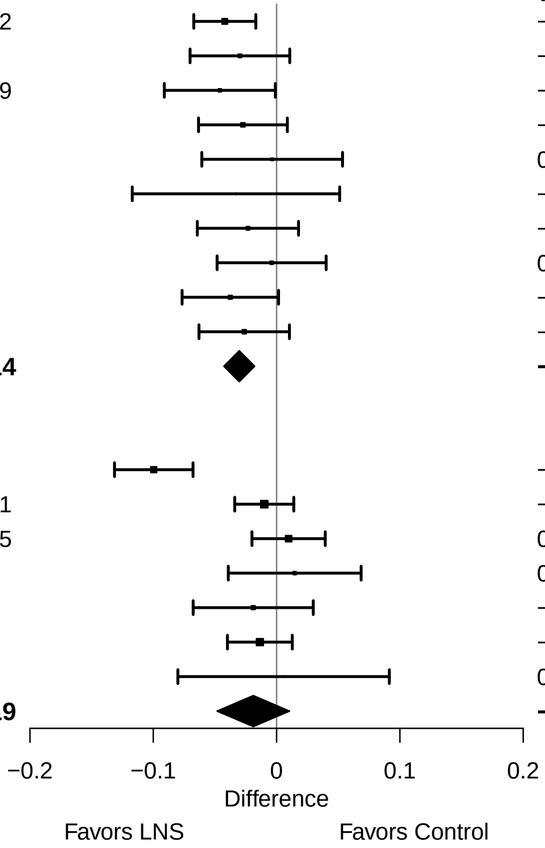

| Country | Trial | N | N | PD (95% CI) | W |
|---|---|---|---|---|---|
| **Sanitation − Improved** | | | | | |
| Bangladesh | JiVitA−4 (34) | 2814 | 1232 | −0.04 (−0.07, −0.02) | 0.16 |
| Bangladesh | RDNS (35) | 1663 | 815 | −0.03 (−0.07, 0.01) | 0.10 |
| Bangladesh | WASH−B (36) | 572 | 1119 | −0.05 (−0.09, 0.00) | 0.09 |
| Burkina Faso | PROMIS (38) | 857 | 911 | −0.03 (−0.06, 0.01) | 0.12 |
| Burkina Faso | PROMIS CS (38) | 430 | 436 | 0.00 (−0.06, 0.05) | 0.07 |
| Ghana | GHANA (39) | 98 | 96 | −0.03 (−0.12, 0.05) | 0.04 |
| Ghana | iLiNS−DYADG (40) | 347 | 693 | −0.02 (−0.06, 0.02) | 0.10 |
| Haiti | HAITI (41) | 148 | 150 | 0.00 (−0.05, 0.04) | 0.09 |
| Mali | PROMIS (46) | 500 | 502 | −0.04 (−0.08, 0.00) | 0.11 |
| Mali | PROMIS CS (46) | 946 | 960 | −0.03 (−0.06, 0.01) | 0.12 |
| **I² = 0.00, Tau² = 0.00** | | **8375** | **6914** | **−0.03 (−0.04, −0.02)** | |
| **Sanitation − Unimproved** | | | | | |
| Burkina Faso | iLiNS−Zinc (37) | 1960 | 666 | −0.10 (−0.13, −0.07) | 0.17 |
| Kenya | WASH−B (42) | 697 | 2261 | −0.01 (−0.03, 0.01) | 0.21 |
| Madagascar | MAHAY (43) | 1702 | 1685 | 0.01 (−0.02, 0.04) | 0.18 |
| Malawi | iLiNS−DYADM (44) | 220 | 444 | 0.01 (−0.04, 0.07) | 0.09 |
| Malawi | iLiNS−DOSE (45) | 702 | 241 | −0.02 (−0.07, 0.03) | 0.11 |
| Zimbabwe | SHINE (HIV−) (47) | 887 | 876 | −0.01 (−0.04, 0.01) | 0.20 |
| Zimbabwe | SHINE (HIV+) (48) | 147 | 146 | 0.01 (−0.08, 0.09) | 0.04 |
| **I² = 0.80, Tau² = 0.00** | | **6315** | **6319** | **−0.02 (−0.05, 0.01)** | |

#### Supplemental figure 4N: Underweight prevalence difference

### 4N6: Stratified by Supplement duration

**Supplement duration**
**(p−diff = 0.415)**

| Country | Trial | N | N | PD (95% CI) | W |
|---|---|---|---|---|---|
| **Supplement duration − 12m or less** | | | | | |
| Bangladesh | JiVitA−4 (34) | 2814 | 1232 | −0.04 (−0.07, −0.02) | 0.12 |
| Burkina Faso | iLiNS−Zinc (37) | 1960 | 666 | −0.10 (−0.13, −0.07) | 0.10 |
| Burkina Faso | PROMIS (38) | 857 | 911 | −0.03 (−0.06, 0.01) | 0.09 |
| Burkina Faso | PROMIS CS (38) | 430 | 436 | 0.00 (−0.06, 0.05) | 0.06 |
| Ghana | GHANA (39) | 98 | 96 | −0.03 (−0.12, 0.05) | 0.03 |
| Ghana | iLiNS−DYADG (40) | 347 | 693 | −0.02 (−0.06, 0.02) | 0.08 |
| Haiti | HAITI (41) | 148 | 150 | 0.00 (−0.05, 0.04) | 0.08 |
| Madagascar | MAHAY (43) | 1702 | 1685 | 0.01 (−0.02, 0.04) | 0.11 |
| Malawi | iLiNS−DYADM (44) | 220 | 444 | 0.01 (−0.04, 0.07) | 0.06 |
| Malawi | iLiNS−DOSE (45) | 702 | 241 | −0.02 (−0.07, 0.03) | 0.07 |
| Zimbabwe | SHINE (HIV−) (47) | 1869 | 1786 | −0.03 (−0.05, −0.01) | 0.14 |
| Zimbabwe | SHINE (HIV+) (48) | 336 | 328 | 0.00 (−0.06, 0.06) | 0.06 |
| **I² = 0.66, Tau² = 0.00** | | **11483** | **8668** | **−0.02 (−0.04, −0.01)** | |
| **Supplement duration − > 12m** | | | | | |
| Bangladesh | RDNS (35) | 1663 | 815 | −0.03 (−0.07, 0.01) | 0.16 |
| Bangladesh | WASH−B (36) | 1163 | 3461 | −0.07 (−0.10, −0.04) | 0.21 |
| Kenya | WASH−B (42) | 1461 | 5165 | −0.02 (−0.04, −0.01) | 0.28 |
| Mali | PROMIS (46) | 500 | 502 | −0.04 (−0.08, 0.00) | 0.17 |
| Mali | PROMIS CS (46) | 946 | 960 | −0.03 (−0.06, 0.01) | 0.18 |
| **I² = 0.43, Tau² = 0.00** | | **5733** | **10903** | **−0.04 (−0.05, −0.02)** | |

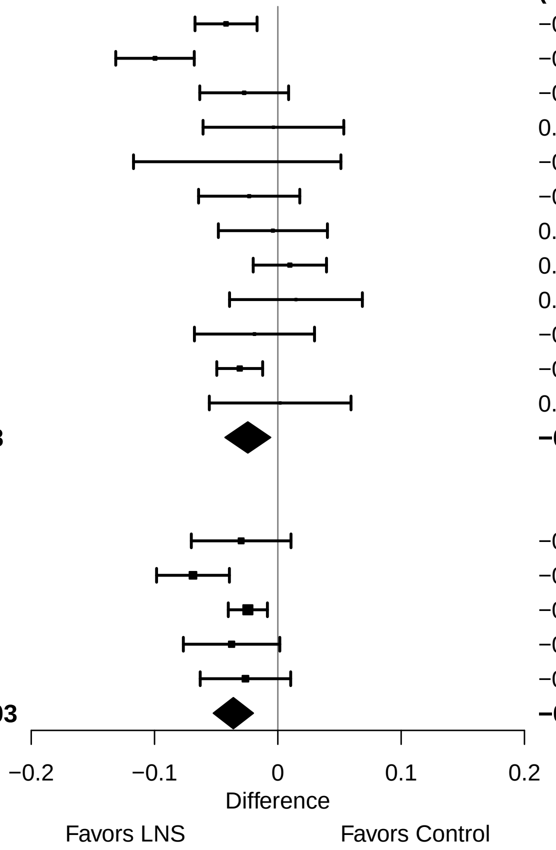

##### Supplemental figure 4N: Underweight prevalence difference

#### 4N7: Stratified by Frequency of contact

**Frequency of contact**
**(p−diff = 0.032)**

**Frequency of contact − Monthly**

| Country | Trial | N | N | PD (95% CI) | W |
|---|---|---|---|---|---|
| Bangladesh | RDNS (35) | 1663 | 815 | −0.03 (−0.07, 0.01) | 0.09 |
| Burkina Faso | PROMIS (38) | 857 | 911 | −0.03 (−0.06, 0.01) | 0.10 |
| Burkina Faso | PROMIS CS (38) | 430 | 436 | 0.00 (−0.06, 0.05) | 0.06 |
| Haiti | HAITI (41) | 148 | 150 | 0.00 (−0.05, 0.04) | 0.08 |
| Kenya | WASH−B (42) | 1461 | 5165 | −0.02 (−0.04, −0.01) | 0.15 |
| Madagascar | MAHAY (43) | 1702 | 1685 | 0.01 (−0.02, 0.04) | 0.12 |
| Mali | PROMIS (46) | 500 | 502 | −0.04 (−0.08, 0.00) | 0.09 |
| Mali | PROMIS CS (46) | 946 | 960 | −0.03 (−0.06, 0.01) | 0.10 |
| Zimbabwe | SHINE (HIV−) (47) | 1869 | 1786 | −0.03 (−0.05, −0.01) | 0.15 |
| Zimbabwe | SHINE (HIV+) (48) | 336 | 328 | 0.00 (−0.06, 0.06) | 0.06 |
| **$I^2$ = 0.00, $Tau^2$ = 0.00** | | **9912** | **12738** | **−0.02 (−0.03, −0.01)** | |

**Frequency of contact − Weekly**

| Country | Trial | N | N | PD (95% CI) | W |
|---|---|---|---|---|---|
| Bangladesh | JiVitA−4 (34) | 2814 | 1232 | −0.04 (−0.07, −0.02) | 0.21 |
| Bangladesh | WASH−B (36) | 1163 | 3461 | −0.07 (−0.10, −0.04) | 0.19 |
| Burkina Faso | iLiNS−Zinc (37) | 1960 | 666 | −0.10 (−0.13, −0.07) | 0.18 |
| Ghana | GHANA (39) | 98 | 96 | −0.03 (−0.12, 0.05) | 0.06 |
| Ghana | iLiNS−DYADG (40) | 347 | 693 | −0.02 (−0.06, 0.02) | 0.14 |
| Malawi | iLiNS−DYADM (44) | 220 | 444 | 0.01 (−0.04, 0.07) | 0.11 |
| Malawi | iLiNS−DOSE (45) | 702 | 241 | −0.02 (−0.07, 0.03) | 0.12 |
| **$I^2$ = 0.70, $Tau^2$ = 0.00** | | **7304** | **6833** | **−0.04 (−0.07, −0.02)** | |

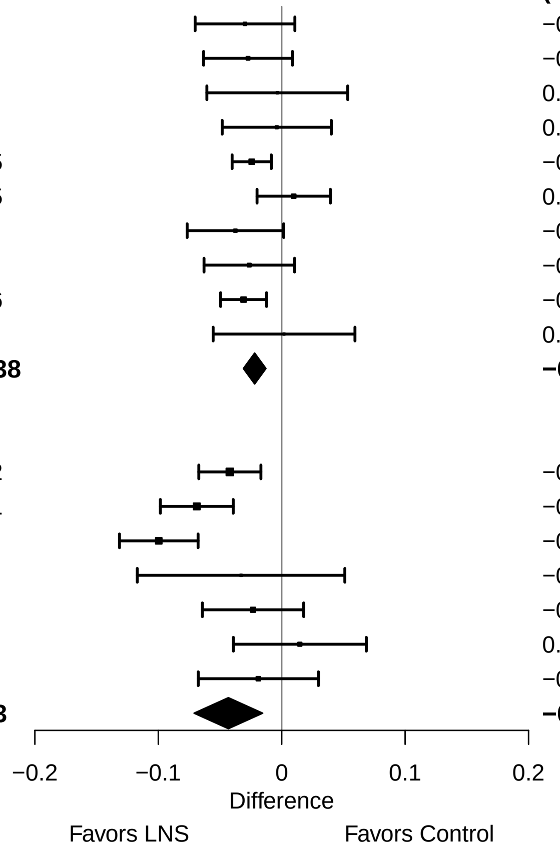

#### Supplemental figure 4N: Underweight prevalence difference

##### 4N8: Stratified by Average SQ-LNS compliance

**Average SQ−LNS compliance**
**(p−diff = 0.062)**

| Country | Trial | N | N | PD (95% CI) | W |
|---|---|---|---|---|---|
| **Average SQ−LNS compliance − Low** | | | | | |
| Burkina Faso | PROMIS (38) | 857 | 911 | −0.03 (−0.06, 0.01) | 0.13 |
| Burkina Faso | PROMIS CS (38) | 430 | 436 | 0.00 (−0.06, 0.05) | 0.07 |
| Ghana | iLiNS−DYADG (40) | 347 | 693 | −0.02 (−0.06, 0.02) | 0.11 |
| Malawi | iLiNS−DYADM (44) | 220 | 444 | 0.01 (−0.04, 0.07) | 0.08 |
| Malawi | iLiNS−DOSE (45) | 702 | 241 | −0.02 (−0.07, 0.03) | 0.09 |
| Mali | PROMIS (46) | 500 | 502 | −0.04 (−0.08, 0.00) | 0.12 |
| Mali | PROMIS CS (46) | 946 | 960 | −0.03 (−0.06, 0.01) | 0.13 |
| Zimbabwe | SHINE (HIV−) (47) | 1869 | 1786 | −0.03 (−0.05, −0.01) | 0.20 |
| Zimbabwe | SHINE (HIV+) (48) | 336 | 328 | 0.00 (−0.06, 0.06) | 0.07 |
| **I² = 0.00, Tau² = 0.00** | | **6207** | **6301** | **−0.02 (−0.04, −0.01)** | |
| **Average SQ−LNS compliance − High** | | | | | |
| Bangladesh | JiVitA−4 (34) | 2814 | 1232 | −0.04 (−0.07, −0.02) | 0.18 |
| Bangladesh | RDNS (35) | 1663 | 815 | −0.03 (−0.07, 0.01) | 0.12 |
| Bangladesh | WASH−B (36) | 1163 | 3461 | −0.07 (−0.10, −0.04) | 0.16 |
| Burkina Faso | iLiNS−Zinc (37) | 1960 | 666 | −0.10 (−0.13, −0.07) | 0.15 |
| Ghana | GHANA (39) | 98 | 96 | −0.03 (−0.12, 0.05) | 0.04 |
| Haiti | HAITI (41) | 148 | 150 | 0.00 (−0.05, 0.04) | 0.11 |
| Kenya | WASH−B (42) | 1461 | 5165 | −0.02 (−0.04, −0.01) | 0.23 |
| **I² = 0.75, Tau² = 0.00** | | **9307** | **11585** | **−0.04 (−0.07, −0.02)** | |

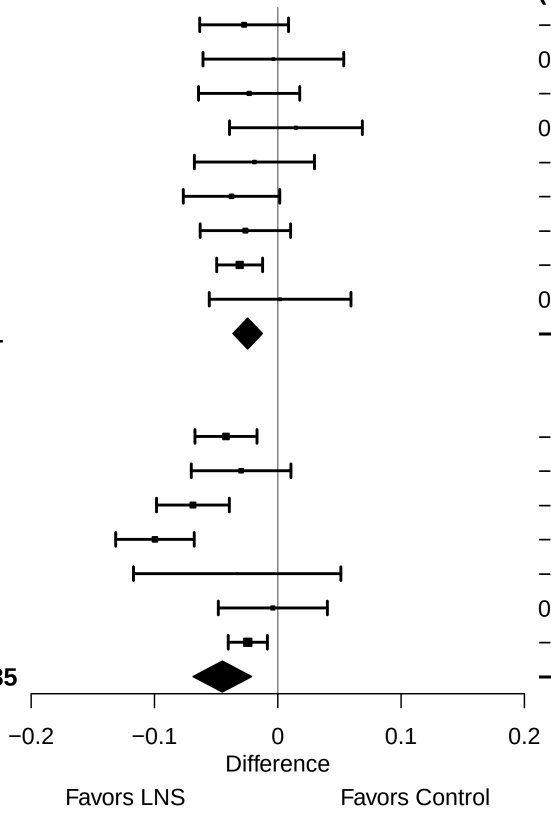

### Supplemental figure 4O: Mean difference in HCZ

#### 4O1: Stratified by Geographic region

**Geographic region**
**(p−diff = 0.840)**

| Country | Trial | N | N | MD (95% CI) | W |
|---|---|---|---|---|---|
| **Geographic region − SEAR** | | | | | |
| Bangladesh | JiVitA−4 (34) | 2885 | 1267 | 0.06 (0.01, 0.11) | 0.38 |
| Bangladesh | RDNS (35) | 1663 | 815 | 0.10 (0.03, 0.17) | 0.30 |
| Bangladesh | WASH−B (36) | 1163 | 3463 | 0.12 (0.06, 0.19) | 0.32 |
| **I² = 0.28, Tau² = 0.00** | | **5711** | **5545** | **0.09 (0.05, 0.13)** | |
| **Geographic region − AFR** | | | | | |
| Burkina Faso | iLiNS−Zinc (37) | 1946 | 664 | 0.16 (0.10, 0.23) | 0.22 |
| Burkina Faso | PROMIS (38) | | | | |
| Burkina Faso | PROMIS CS (38) | | | | |
| Ghana | GHANA (39) | 98 | 96 | 0.20 (−0.06, 0.46) | 0.03 |
| Ghana | iLiNS−DYADG (40) | 347 | 692 | 0.07 (−0.05, 0.18) | 0.11 |
| Kenya | WASH−B (42) | 1461 | 5176 | 0.04 (−0.02, 0.10) | 0.22 |
| Madagascar | MAHAY (43) | | | | |
| Malawi | iLiNS−DYADM (44) | 220 | 444 | −0.13 (−0.28, 0.03) | 0.07 |
| Malawi | iLiNS−DOSE (45) | 692 | 238 | 0.11 (−0.02, 0.23) | 0.10 |
| Mali | PROMIS (46) | | | | |
| Mali | PROMIS CS (46) | | | | |
| Zimbabwe | SHINE (HIV−) (47) | 1871 | 1783 | 0.09 (0.02, 0.16) | 0.19 |
| Zimbabwe | SHINE (HIV+) (48) | 337 | 329 | 0.10 (−0.07, 0.28) | 0.06 |
| **I² = 0.56, Tau² = 0.00** | | **6972** | **9422** | **0.08 (0.02, 0.14)** | |

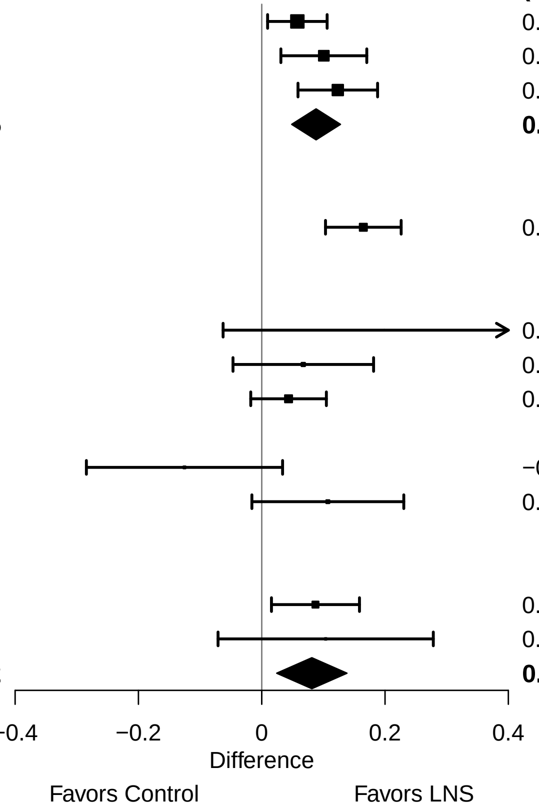

#### Supplemental figure 4O: Mean difference in HCZ

#### 4O2: Stratified by Stunting burden

**Stunting burden**
**(p−diff = 0.053)**

| Country | Trial | N | N | MD (95% CI) | W |
|---|---|---|---|---|---|
| **Stunting burden − Less than 35%** | | | | | |
| Burkina Faso | PROMIS (38) | | | | |
| Burkina Faso | PROMIS CS (38) | | | | |
| Ghana | GHANA (39) | 98 | 96 | 0.20 (−0.06, 0.46) | 0.07 |
| Ghana | iLiNS−DYADG (40) | 347 | 692 | 0.07 (−0.05, 0.18) | 0.27 |
| Haiti | HAITI (41) | | | | |
| Kenya | WASH−B (42) | 1461 | 5176 | 0.04 (−0.02, 0.10) | 0.50 |
| Malawi | iLiNS−DYADM (44) | 220 | 444 | −0.13 (−0.28, 0.03) | 0.16 |
| **I² = 0.48, Tau² = 0.01** | | **2126** | **6408** | **0.03 (−0.08, 0.14)** | |
| **Stunting burden − More than 35%** | | | | | |
| Bangladesh | JiVitA−4 (34) | 2885 | 1267 | 0.06 (0.01, 0.11) | 0.20 |
| Bangladesh | RDNS (35) | 1663 | 815 | 0.10 (0.03, 0.17) | 0.16 |
| Bangladesh | WASH−B (36) | 1163 | 3463 | 0.12 (0.06, 0.19) | 0.17 |
| Burkina Faso | iLiNS−Zinc (37) | 1946 | 664 | 0.16 (0.10, 0.23) | 0.18 |
| Madagascar | MAHAY (43) | | | | |
| Malawi | iLiNS−DOSE (45) | 692 | 238 | 0.11 (−0.02, 0.23) | 0.08 |
| Mali | PROMIS (46) | | | | |
| Mali | PROMIS CS (46) | | | | |
| Zimbabwe | SHINE (HIV−) (47) | 1871 | 1783 | 0.09 (0.02, 0.16) | 0.16 |
| Zimbabwe | SHINE (HIV+) (48) | 337 | 329 | 0.10 (−0.07, 0.28) | 0.05 |
| **I² = 0.23, Tau² = 0.00** | | **10557** | **8559** | **0.10 (0.07, 0.13)** | |

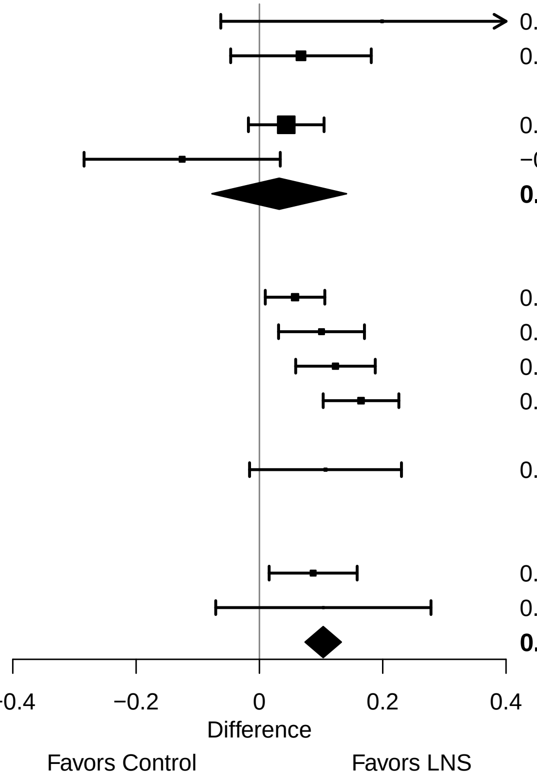

#### Supplemental figure 4O: Mean difference in HCZ

### 4O3: Stratified by Malaria prevalence

**Malaria prevalence**
**(p−diff = 0.621)**

| Country | Trial | N | N | MD (95% CI) | W |
|---|---|---|---|---|---|
| **Malaria prevalence − Less than 10%** | | | | | |
| Bangladesh | JiVitA−4 (34) | 2885 | 1267 | 0.06 (0.01, 0.11) | 0.22 |
| Bangladesh | RDNS (35) | 1663 | 815 | 0.10 (0.03, 0.17) | 0.17 |
| Bangladesh | WASH−B (36) | 1163 | 3463 | 0.12 (0.06, 0.19) | 0.19 |
| Haiti | HAITI (41) | | | | |
| Kenya | WASH−B (42) | 1461 | 5176 | 0.04 (−0.02, 0.10) | 0.19 |
| Madagascar | MAHAY (43) | | | | |
| Zimbabwe | SHINE (HIV−) (47) | 1871 | 1783 | 0.09 (0.02, 0.16) | 0.17 |
| Zimbabwe | SHINE (HIV+) (48) | 337 | 329 | 0.10 (−0.07, 0.28) | 0.05 |
| **I² = 0.00, Tau² = 0.00** | | **9380** | **12833** | **0.08 (0.05, 0.11)** | |
| **Malaria prevalence − At least 10%** | | | | | |
| Burkina Faso | iLiNS−Zinc (37) | 1946 | 664 | 0.16 (0.10, 0.23) | 0.40 |
| Burkina Faso | PROMIS (38) | | | | |
| Burkina Faso | PROMIS CS (38) | | | | |
| Ghana | GHANA (39) | 98 | 96 | 0.20 (−0.06, 0.46) | 0.06 |
| Ghana | iLiNS−DYADG (40) | 347 | 692 | 0.07 (−0.05, 0.18) | 0.21 |
| Malawi | iLiNS−DYADM (44) | 220 | 444 | −0.13 (−0.28, 0.03) | 0.13 |
| Malawi | iLiNS−DOSE (45) | 692 | 238 | 0.11 (−0.02, 0.23) | 0.19 |
| Mali | PROMIS (46) | | | | |
| Mali | PROMIS CS (46) | | | | |
| **I² = 0.68, Tau² = 0.01** | | **3303** | **2134** | **0.08 (−0.02, 0.19)** | |

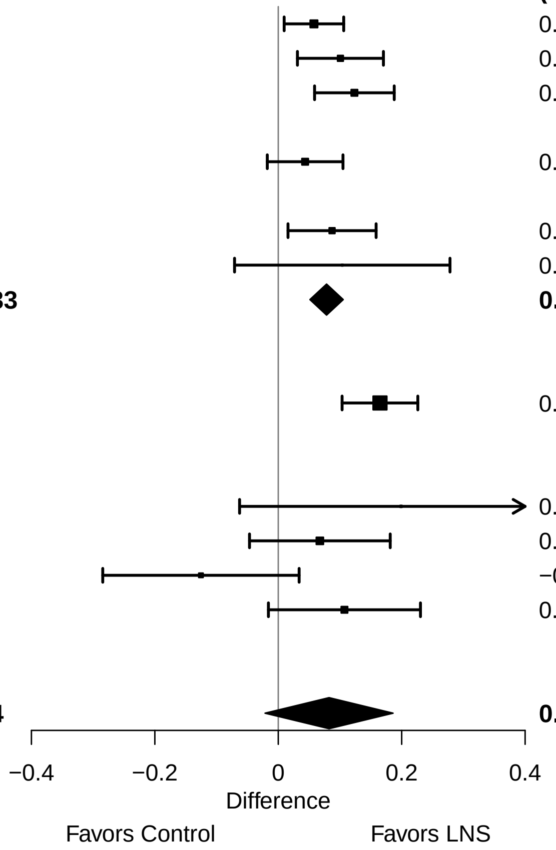

#### Supplemental figure 4O: Mean difference in HCZ

#### 4O4: Stratified by Source water quality

**Source water quality**
**(p−diff = 0.875)**

| Country | Trial | N | N | MD (95% CI) | W |
|---|---|---|---|---|---|
| **Source water quality − Improved** | | | | | |
| Bangladesh | JiVitA−4 (34) | 2885 | 1267 | 0.06 (0.01, 0.11) | 0.26 |
| Bangladesh | RDNS (35) | 1663 | 815 | 0.10 (0.03, 0.17) | 0.21 |
| Bangladesh | WASH−B (36) | 571 | 1121 | 0.16 (0.05, 0.27) | 0.14 |
| Ghana | GHANA (39) | 98 | 96 | 0.20 (−0.06, 0.46) | 0.04 |
| Ghana | iLiNS−DYADG (40) | 347 | 692 | 0.07 (−0.05, 0.18) | 0.13 |
| Haiti | HAITI (41) | | | | |
| Malawi | iLiNS−DYADM (44) | 220 | 444 | −0.13 (−0.28, 0.03) | 0.09 |
| Malawi | iLiNS−DOSE (45) | 692 | 238 | 0.11 (−0.02, 0.23) | 0.12 |
| **I² = 0.43, Tau² = 0.00** | | **6476** | **4673** | **0.08 (0.02, 0.14)** | |
| **Source water quality − Unimproved** | | | | | |
| Burkina Faso | iLiNS−Zinc (37) | 1946 | 664 | 0.16 (0.10, 0.23) | 0.39 |
| Burkina Faso | PROMIS (38) | | | | |
| Burkina Faso | PROMIS CS (38) | | | | |
| Kenya | WASH−B (42) | 696 | 2266 | 0.04 (−0.05, 0.13) | 0.29 |
| Madagascar | MAHAY (43) | | | | |
| Mali | PROMIS (46) | | | | |
| Mali | PROMIS CS (46) | | | | |
| Zimbabwe | SHINE (HIV−) (47) | 887 | 873 | 0.04 (−0.07, 0.14) | 0.24 |
| Zimbabwe | SHINE (HIV+) (48) | 147 | 146 | 0.04 (−0.21, 0.28) | 0.07 |
| **I² = 0.62, Tau² = 0.00** | | **3676** | **3949** | **0.09 (0.01, 0.16)** | |

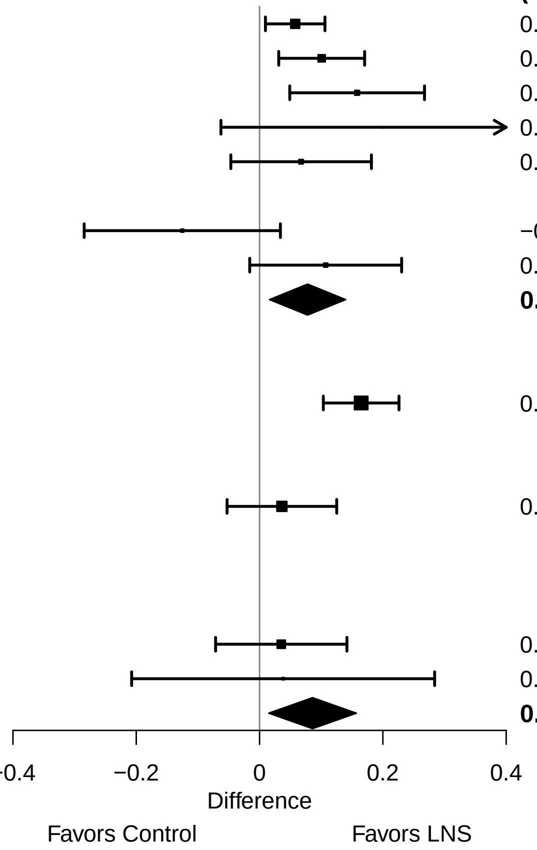

**Supplemental figure 4O: Mean difference in HCZ**

#### 4O5: Stratified by Sanitation

**Sanitation**
**(p−diff = 0.484)**

| Country | Trial | N | N | MD (95% CI) | W |
|---|---|---|---|---|---|
| **Sanitation − Improved** | | | | | |
| Bangladesh | JiVitA−4 (34) | 2885 | 1267 | 0.06 (0.01, 0.11) | 0.33 |
| Bangladesh | RDNS (35) | 1663 | 815 | 0.10 (0.03, 0.17) | 0.27 |
| Bangladesh | WASH−B (36) | 571 | 1121 | 0.16 (0.05, 0.27) | 0.18 |
| Burkina Faso | PROMIS (38) | | | | |
| Burkina Faso | PROMIS CS (38) | | | | |
| Ghana | GHANA (39) | 98 | 96 | 0.20 (−0.06, 0.46) | 0.05 |
| Ghana | iLiNS−DYADG (40) | 347 | 692 | 0.07 (−0.05, 0.18) | 0.17 |
| Haiti | HAITI (41) | | | | |
| Mali | PROMIS (46) | | | | |
| Mali | PROMIS CS (46) | | | | |
| **I² = 0.00, Tau² = 0.00** | | **5564** | **3991** | **0.08 (0.05, 0.12)** | |
| **Sanitation − Unimproved** | | | | | |
| Burkina Faso | iLiNS−Zinc (37) | 1946 | 664 | 0.16 (0.10, 0.23) | 0.29 |
| Kenya | WASH−B (42) | 696 | 2266 | 0.04 (−0.05, 0.13) | 0.22 |
| Madagascar | MAHAY (43) | | | | |
| Malawi | iLiNS−DYADM (44) | 220 | 444 | −0.13 (−0.28, 0.03) | 0.11 |
| Malawi | iLiNS−DOSE (45) | 692 | 238 | 0.11 (−0.02, 0.23) | 0.15 |
| Zimbabwe | SHINE (HIV−) (47) | 887 | 873 | 0.04 (−0.07, 0.14) | 0.18 |
| Zimbabwe | SHINE (HIV+) (48) | 147 | 146 | 0.04 (−0.21, 0.28) | 0.05 |
| **I² = 0.68, Tau² = 0.01** | | **4588** | **4631** | **0.06 (−0.02, 0.14)** | |

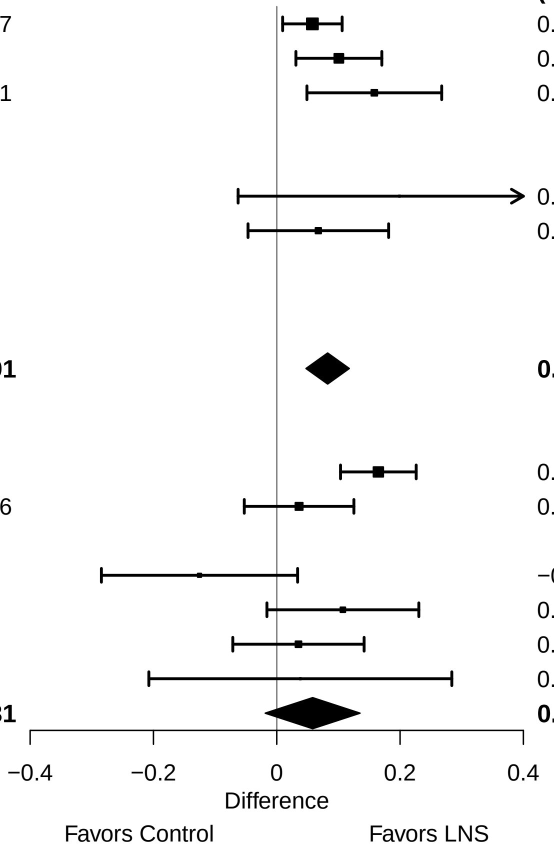

##### Supplemental figure 4O: Mean difference in HCZ

#### 4O6: Stratified by Supplement duration

**Supplement duration**
**(p−diff = 0.949)**

| Country | Trial | N | N | MD (95% CI) | W |
|---|---|---|---|---|---|
| **Supplement duration − 12m or less** | | | | | |
| Bangladesh | JiVitA−4 (34) | 2885 | 1267 | 0.06 (0.01, 0.11) | 0.24 |
| Burkina Faso | iLiNS−Zinc (37) | 1946 | 664 | 0.16 (0.10, 0.23) | 0.21 |
| Burkina Faso | PROMIS (38) | | | | |
| Burkina Faso | PROMIS CS (38) | | | | |
| Ghana | GHANA (39) | 98 | 96 | 0.20 (−0.06, 0.46) | 0.03 |
| Ghana | iLiNS−DYADG (40) | 347 | 692 | 0.07 (−0.05, 0.18) | 0.11 |
| Haiti | HAITI (41) | | | | |
| Madagascar | MAHAY (43) | | | | |
| Malawi | iLiNS−DYADM (44) | 220 | 444 | −0.13 (−0.28, 0.03) | 0.07 |
| Malawi | iLiNS−DOSE (45) | 692 | 238 | 0.11 (−0.02, 0.23) | 0.10 |
| Zimbabwe | SHINE (HIV−) (47) | 1871 | 1783 | 0.09 (0.02, 0.16) | 0.18 |
| Zimbabwe | SHINE (HIV+) (48) | 337 | 329 | 0.10 (−0.07, 0.28) | 0.06 |
| **$I^2$ = 0.54, $Tau^2$ = 0.00** | | **8396** | **5513** | **0.08 (0.03, 0.14)** | |
| **Supplement duration − > 12m** | | | | | |
| Bangladesh | RDNS (35) | 1663 | 815 | 0.10 (0.03, 0.17) | 0.32 |
| Bangladesh | WASH−B (36) | 1163 | 3463 | 0.12 (0.06, 0.19) | 0.34 |
| Kenya | WASH−B (42) | 1461 | 5176 | 0.04 (−0.02, 0.10) | 0.35 |
| Mali | PROMIS (46) | | | | |
| Mali | PROMIS CS (46) | | | | |
| **$I^2$ = 0.39, $Tau^2$ = 0.00** | | **4287** | **9454** | **0.09 (0.04, 0.14)** | |

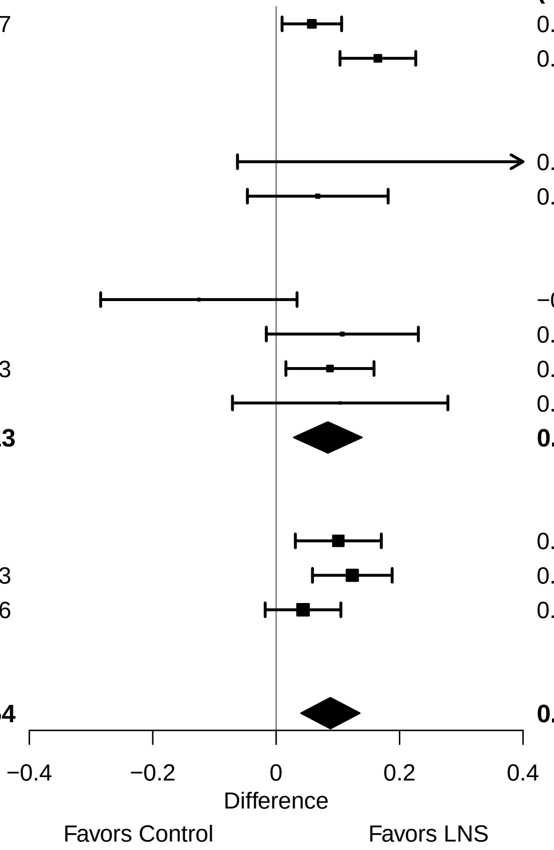

#### Supplemental figure 4O: Mean difference in HCZ

### 4O7: Stratified by Frequency of contact

**Frequency of contact**
**(p−diff = 0.687)**

| Country | Trial | N | N | MD (95% CI) | W |
|---|---|---|---|---|---|
| **Frequency of contact − Monthly** | | | | | |
| Bangladesh | RDNS (35) | 1663 | 815 | 0.10 (0.03, 0.17) | 0.29 |
| Burkina Faso | PROMIS (38) | | | | |
| Burkina Faso | PROMIS CS (38) | | | | |
| Haiti | HAITI (41) | | | | |
| Kenya | WASH−B (42) | 1461 | 5176 | 0.04 (−0.02, 0.10) | 0.33 |
| Madagascar | MAHAY (43) | | | | |
| Mali | PROMIS (46) | | | | |
| Mali | PROMIS CS (46) | | | | |
| Zimbabwe | SHINE (HIV−) (47) | 1871 | 1783 | 0.09 (0.02, 0.16) | 0.29 |
| Zimbabwe | SHINE (HIV+) (48) | 337 | 329 | 0.10 (−0.07, 0.28) | 0.09 |
| **$I^2$ = 0.00, $Tau^2$ = 0.00** | | **5332** | **8103** | **0.08 (0.04, 0.11)** | |
| **Frequency of contact − Weekly** | | | | | |
| Bangladesh | JiVitA−4 (34) | 2885 | 1267 | 0.06 (0.01, 0.11) | 0.25 |
| Bangladesh | WASH−B (36) | 1163 | 3463 | 0.12 (0.06, 0.19) | 0.21 |
| Burkina Faso | iLiNS−Zinc (37) | 1946 | 664 | 0.16 (0.10, 0.23) | 0.22 |
| Ghana | GHANA (39) | 98 | 96 | 0.20 (−0.06, 0.46) | 0.03 |
| Ghana | iLiNS−DYADG (40) | 347 | 692 | 0.07 (−0.05, 0.18) | 0.12 |
| Malawi | iLiNS−DYADM (44) | 220 | 444 | −0.13 (−0.28, 0.03) | 0.07 |
| Malawi | iLiNS−DOSE (45) | 692 | 238 | 0.11 (−0.02, 0.23) | 0.10 |
| **$I^2$ = 0.63, $Tau^2$ = 0.00** | | **7351** | **6864** | **0.09 (0.02, 0.15)** | |

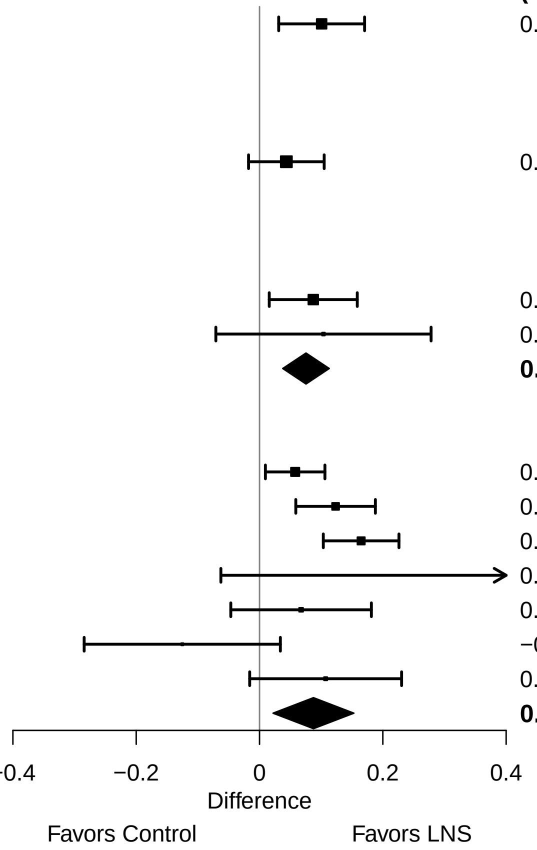

##### Supplemental figure 4O: Mean difference in HCZ

#### 4O8: Stratified by Average SQ-LNS compliance

**Average SQ−LNS compliance**
**(p−diff = 0.336)**

| Country | Trial | N | N | MD (95% CI) | W |
|---|---|---|---|---|---|
| **Average SQ−LNS compliance − Low** | | | | | |
| Burkina Faso | PROMIS (38) | | | | |
| Burkina Faso | PROMIS CS (38) | | | | |
| Ghana | iLiNS−DYADG (40) | 347 | 692 | 0.07 (−0.05, 0.18) | 0.21 |
| Malawi | iLiNS−DYADM (44) | 220 | 444 | −0.13 (−0.28, 0.03) | 0.13 |
| Malawi | iLiNS−DOSE (45) | 692 | 238 | 0.11 (−0.02, 0.23) | 0.19 |
| Mali | PROMIS (46) | | | | |
| Mali | PROMIS CS (46) | | | | |
| Zimbabwe | SHINE (HIV−) (47) | 1871 | 1783 | 0.09 (0.02, 0.16) | 0.35 |
| Zimbabwe | SHINE (HIV+) (48) | 337 | 329 | 0.10 (−0.07, 0.28) | 0.11 |
| **I² = 0.38, Tau² = 0.00** | | **3467** | **3486** | **0.06 (−0.02, 0.13)** | |
| **Average SQ−LNS compliance − High** | | | | | |
| Bangladesh | JiVitA−4 (34) | 2885 | 1267 | 0.06 (0.01, 0.11) | 0.22 |
| Bangladesh | RDNS (35) | 1663 | 815 | 0.10 (0.03, 0.17) | 0.18 |
| Bangladesh | WASH−B (36) | 1163 | 3463 | 0.12 (0.06, 0.19) | 0.19 |
| Burkina Faso | iLiNS−Zinc (37) | 1946 | 664 | 0.16 (0.10, 0.23) | 0.19 |
| Ghana | GHANA (39) | 98 | 96 | 0.20 (−0.06, 0.46) | 0.03 |
| Haiti | HAITI (41) | | | | |
| Kenya | WASH−B (42) | 1461 | 5176 | 0.04 (−0.02, 0.10) | 0.19 |
| **I² = 0.56, Tau² = 0.00** | | **9216** | **11481** | **0.10 (0.06, 0.14)** | |

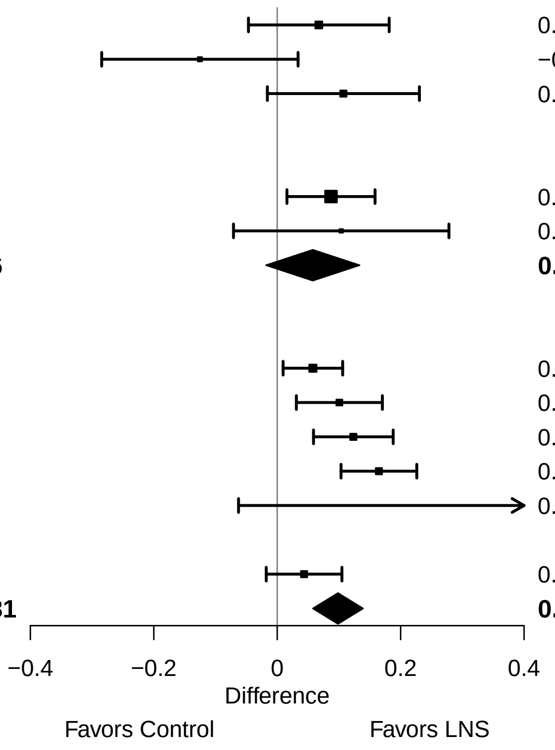

### Supplemental figure 4P: Small head size prevalence ratio

#### 4P1: Stratified by Geographic region

**Geographic region**
**(p−diff = 0.271)**

| Country | Trial | N | N | PR (95% CI) | W |
|---|---|---|---|---|---|
| **Geographic region − SEAR** | | | | | |
| Bangladesh | JiVitA−4 (34) | 2885 | 1267 | 1.03 (0.90, 1.18) | 0.19 |
| Bangladesh | RDNS (35) | 1663 | 815 | 0.90 (0.82, 0.98) | 0.47 |
| Bangladesh | WASH−B (36) | 1163 | 3463 | 0.90 (0.82, 1.00) | 0.34 |
| **I² = 0.36, Tau² = 0.00** | | **5711** | **5545** | **0.93 (0.86, 1.01)** | |
| **Geographic region − AFR** | | | | | |
| Burkina Faso | iLiNS−Zinc (37) | 1946 | 664 | 0.89 (0.78, 1.01) | 0.49 |
| Burkina Faso | PROMIS (38) | | | | |
| Burkina Faso | PROMIS CS (38) | | | | |
| Ghana | GHANA (39) | 98 | 96 | 0.33 (0.07, 1.58) | 0.00 |
| Ghana | iLiNS−DYADG (40) | 347 | 692 | 0.88 (0.65, 1.20) | 0.09 |
| Kenya | WASH−B (42) | 1461 | 5176 | 0.92 (0.67, 1.25) | 0.09 |
| Madagascar | MAHAY (43) | | | | |
| Malawi | iLiNS−DYADM (44) | 220 | 444 | 0.88 (0.54, 1.44) | 0.04 |
| Malawi | iLiNS−DOSE (45) | 692 | 238 | 0.79 (0.60, 1.03) | 0.12 |
| Mali | PROMIS (46) | | | | |
| Mali | PROMIS CS (46) | | | | |
| Zimbabwe | SHINE (HIV−) (47) | 1871 | 1783 | 0.81 (0.63, 1.05) | 0.13 |
| Zimbabwe | SHINE (HIV+) (48) | 337 | 329 | 0.98 (0.61, 1.57) | 0.04 |
| **I² = 0.00, Tau² = 0.00** | | **6972** | **9422** | **0.87 (0.79, 0.95)** | |

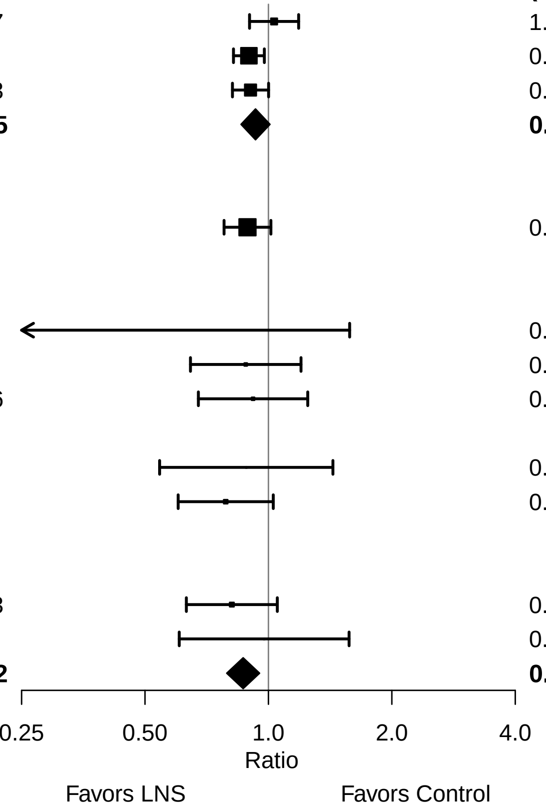

##### Supplemental figure 4P: Small head size prevalence ratio

#### 4P2: Stratified by Stunting burden

**Stunting burden**
**(p−diff = 0.780)**

| Country | Trial | N | N | PR (95% CI) | W |
|---|---|---|---|---|---|
| **Stunting burden − Less than 35%** | | | | | |
| Burkina Faso | PROMIS (38) | | | | |
| Burkina Faso | PROMIS CS (38) | | | | |
| Ghana | GHANA (39) | 98 | 96 | 0.33 (0.07, 1.58) | 0.02 |
| Ghana | iLiNS−DYADG (40) | 347 | 692 | 0.88 (0.65, 1.20) | 0.41 |
| Haiti | HAITI (41) | | | | |
| Kenya | WASH−B (42) | 1461 | 5176 | 0.92 (0.67, 1.25) | 0.41 |
| Malawi | iLiNS−DYADM (44) | 220 | 444 | 0.88 (0.54, 1.44) | 0.16 |
| **I² = 0.00, Tau² = 0.00** | | **2126** | **6408** | **0.88 (0.72, 1.07)** | |
| **Stunting burden − More than 35%** | | | | | |
| Bangladesh | JiVitA−4 (34) | 2885 | 1267 | 1.03 (0.90, 1.18) | 0.14 |
| Bangladesh | RDNS (35) | 1663 | 815 | 0.90 (0.82, 0.98) | 0.36 |
| Bangladesh | WASH−B (36) | 1163 | 3463 | 0.90 (0.82, 1.00) | 0.26 |
| Burkina Faso | iLiNS−Zinc (37) | 1946 | 664 | 0.89 (0.78, 1.01) | 0.15 |
| Madagascar | MAHAY (43) | | | | |
| Malawi | iLiNS−DOSE (45) | 692 | 238 | 0.79 (0.60, 1.03) | 0.04 |
| Mali | PROMIS (46) | | | | |
| Mali | PROMIS CS (46) | | | | |
| Zimbabwe | SHINE (HIV−) (47) | 1871 | 1783 | 0.81 (0.63, 1.05) | 0.04 |
| Zimbabwe | SHINE (HIV+) (48) | 337 | 329 | 0.98 (0.61, 1.57) | 0.01 |
| **I² = 0.00, Tau² = 0.00** | | **10557** | **8559** | **0.91 (0.86, 0.96)** | |

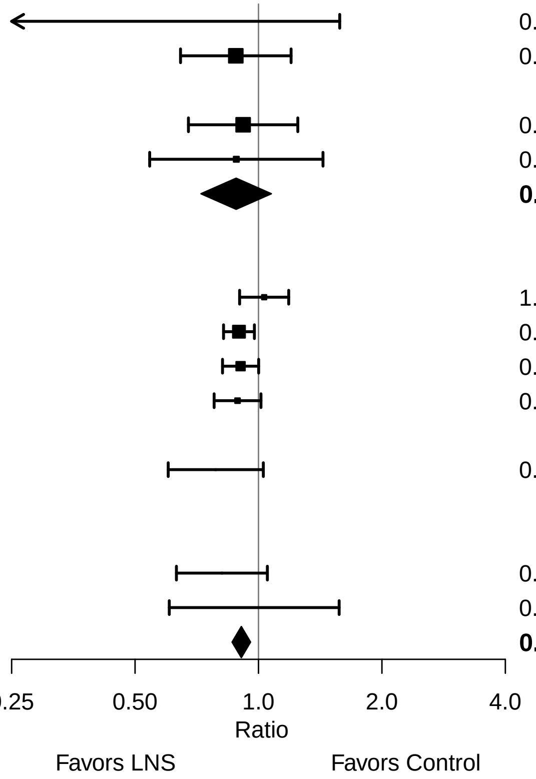

#### Supplemental figure 4P: Small head size prevalence ratio

### 4P3: Stratified by Malaria prevalence

**Malaria prevalence**
**(p−diff = 0.351)**

| Country | Trial | N | N | PR (95% CI) | W |
|---|---|---|---|---|---|
| **Malaria prevalence − Less than 10%** | | | | | |
| Bangladesh | JiVitA−4 (34) | 2885 | 1267 | 1.03 (0.90, 1.18) | 0.17 |
| Bangladesh | RDNS (35) | 1663 | 815 | 0.90 (0.82, 0.98) | 0.43 |
| Bangladesh | WASH−B (36) | 1163 | 3463 | 0.90 (0.82, 1.00) | 0.31 |
| Haiti | HAITI (41) | | | | |
| Kenya | WASH−B (42) | 1461 | 5176 | 0.92 (0.67, 1.25) | 0.03 |
| Madagascar | MAHAY (43) | | | | |
| Zimbabwe | SHINE (HIV−) (47) | 1871 | 1783 | 0.81 (0.63, 1.05) | 0.05 |
| Zimbabwe | SHINE (HIV+) (48) | 337 | 329 | 0.98 (0.61, 1.57) | 0.01 |
| **I² = 0.00, Tau² = 0.00** | | **9380** | **12833** | **0.92 (0.87, 0.97)** | |
| **Malaria prevalence − At least 10%** | | | | | |
| Burkina Faso | iLiNS−Zinc (37) | 1946 | 664 | 0.89 (0.78, 1.01) | 0.67 |
| Burkina Faso | PROMIS (38) | | | | |
| Burkina Faso | PROMIS CS (38) | | | | |
| Ghana | GHANA (39) | 98 | 96 | 0.33 (0.07, 1.58) | 0.00 |
| Ghana | iLiNS−DYADG (40) | 347 | 692 | 0.88 (0.65, 1.20) | 0.12 |
| Malawi | iLiNS−DYADM (44) | 220 | 444 | 0.88 (0.54, 1.44) | 0.05 |
| Malawi | iLiNS−DOSE (45) | 692 | 238 | 0.79 (0.60, 1.03) | 0.16 |
| Mali | PROMIS (46) | | | | |
| Mali | PROMIS CS (46) | | | | |
| **I² = 0.00, Tau² = 0.00** | | **3303** | **2134** | **0.87 (0.78, 0.96)** | |

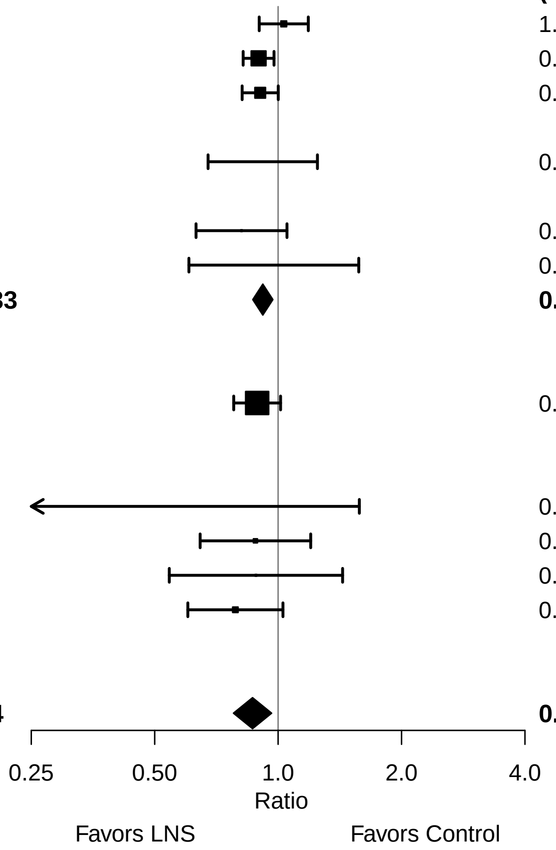

#### Supplemental figure 4P: Small head size prevalence ratio

### 4P4: Stratified by Source water quality

**Source water quality**
**(p−diff = 0.958)**

| Country | Trial | N | N | PR (95% CI) | W |
|---|---|---|---|---|---|
| **Source water quality − Improved** | | | | | |
| Bangladesh | JiVitA−4 (34) | 2885 | 1267 | 1.03 (0.90, 1.18) | 0.21 |
| Bangladesh | RDNS (35) | 1663 | 815 | 0.90 (0.82, 0.98) | 0.54 |
| Bangladesh | WASH−B (36) | 571 | 1121 | 0.86 (0.73, 1.03) | 0.13 |
| Ghana | GHANA (39) | 98 | 96 | 0.33 (0.07, 1.58) | 0.00 |
| Ghana | iLiNS−DYADG (40) | 347 | 692 | 0.88 (0.65, 1.20) | 0.04 |
| Haiti | HAITI (41) | | | | |
| Malawi | iLiNS−DYADM (44) | 220 | 444 | 0.88 (0.54, 1.44) | 0.02 |
| Malawi | iLiNS−DOSE (45) | 692 | 238 | 0.79 (0.60, 1.03) | 0.06 |
| **I² = 0.08, Tau² = 0.00** | | **6476** | **4673** | **0.91 (0.85, 0.98)** | |
| **Source water quality − Unimproved** | | | | | |
| Burkina Faso | iLiNS−Zinc (37) | 1946 | 664 | 0.89 (0.78, 1.01) | 0.79 |
| Burkina Faso | PROMIS (38) | | | | |
| Burkina Faso | PROMIS CS (38) | | | | |
| Kenya | WASH−B (42) | 696 | 2266 | 0.73 (0.45, 1.17) | 0.06 |
| Madagascar | MAHAY (43) | | | | |
| Mali | PROMIS (46) | | | | |
| Mali | PROMIS CS (46) | | | | |
| Zimbabwe | SHINE (HIV−) (47) | 887 | 873 | 1.01 (0.73, 1.39) | 0.13 |
| Zimbabwe | SHINE (HIV+) (48) | 147 | 146 | 1.44 (0.68, 3.06) | 0.02 |
| **I² = 0.00, Tau² = 0.00** | | **3676** | **3949** | **0.90 (0.80, 1.01)** | |

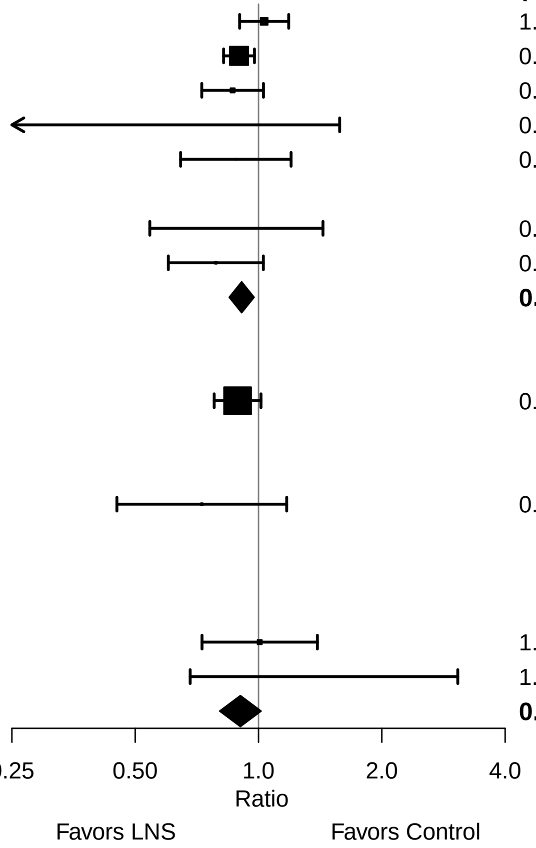

#### Supplemental figure 4P: Small head size prevalence ratio

#### 4P5: Stratified by Sanitation

**Sanitation**
**(p−diff = 0.549)**

| Country | Trial | N | N | PR (95% CI) | W |
|---|---|---|---|---|---|
| **Sanitation − Improved** | | | | | |
| Bangladesh | JiVitA−4 (34) | 2885 | 1267 | 1.03 (0.90, 1.18) | 0.23 |
| Bangladesh | RDNS (35) | 1663 | 815 | 0.90 (0.82, 0.98) | 0.58 |
| Bangladesh | WASH−B (36) | 571 | 1121 | 0.86 (0.73, 1.03) | 0.14 |
| Burkina Faso | PROMIS (38) | | | | |
| Burkina Faso | PROMIS CS (38) | | | | |
| Ghana | GHANA (39) | 98 | 96 | 0.33 (0.07, 1.58) | 0.00 |
| Ghana | iLiNS−DYADG (40) | 347 | 692 | 0.88 (0.65, 1.20) | 0.05 |
| Haiti | HAITI (41) | | | | |
| Mali | PROMIS (46) | | | | |
| Mali | PROMIS CS (46) | | | | |
| **I² = 0.24, Tau² = 0.00** | | **5564** | **3991** | **0.92 (0.84, 1.01)** | |
| **Sanitation − Unimproved** | | | | | |
| Burkina Faso | iLiNS−Zinc (37) | 1946 | 664 | 0.89 (0.78, 1.01) | 0.63 |
| Kenya | WASH−B (42) | 696 | 2266 | 0.73 (0.45, 1.17) | 0.05 |
| Madagascar | MAHAY (43) | | | | |
| Malawi | iLiNS−DYADM (44) | 220 | 444 | 0.88 (0.54, 1.44) | 0.05 |
| Malawi | iLiNS−DOSE (45) | 692 | 238 | 0.79 (0.60, 1.03) | 0.15 |
| Zimbabwe | SHINE (HIV−) (47) | 887 | 873 | 1.01 (0.73, 1.39) | 0.10 |
| Zimbabwe | SHINE (HIV+) (48) | 147 | 146 | 1.44 (0.68, 3.06) | 0.02 |
| **I² = 0.00, Tau² = 0.00** | | **4588** | **4631** | **0.88 (0.80, 0.98)** | |

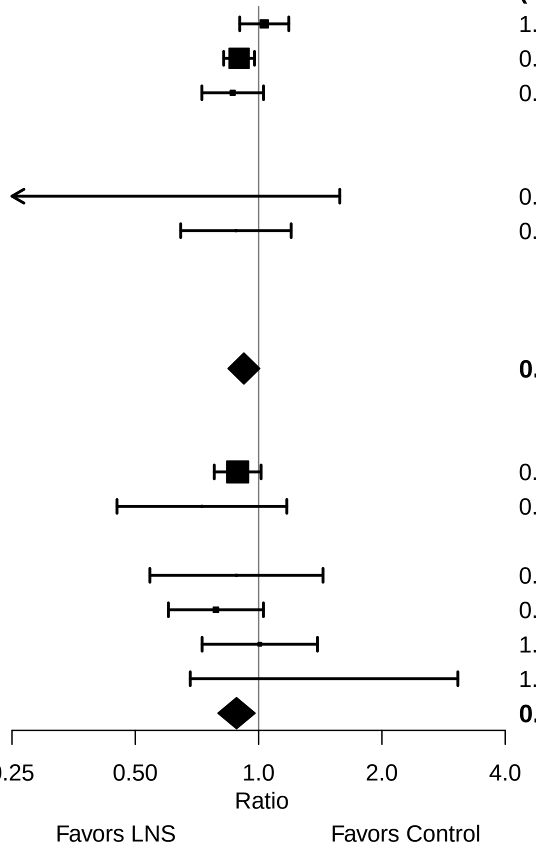

#### Supplemental figure 4P: Small head size prevalence ratio

#### 4P6: Stratified by Supplement duration

**Supplement duration**
**(p−diff = 0.749)**

| Country | Trial | N | N | PR (95% CI) | W |
|---|---|---|---|---|---|
| **Supplement duration − 12m or less** | | | | | |
| Bangladesh | JiVitA−4 (34) | 2885 | 1267 | 1.03 (0.90, 1.18) | 0.33 |
| Burkina Faso | iLiNS−Zinc (37) | 1946 | 664 | 0.89 (0.78, 1.01) | 0.36 |
| Burkina Faso | PROMIS (38) | | | | |
| Burkina Faso | PROMIS CS (38) | | | | |
| Ghana | GHANA (39) | 98 | 96 | 0.33 (0.07, 1.58) | 0.00 |
| Ghana | iLiNS−DYADG (40) | 347 | 692 | 0.88 (0.65, 1.20) | 0.07 |
| Haiti | HAITI (41) | | | | |
| Madagascar | MAHAY (43) | | | | |
| Malawi | iLiNS−DYADM (44) | 220 | 444 | 0.88 (0.54, 1.44) | 0.03 |
| Malawi | iLiNS−DOSE (45) | 692 | 238 | 0.79 (0.60, 1.03) | 0.09 |
| Zimbabwe | SHINE (HIV−) (47) | 1871 | 1783 | 0.81 (0.63, 1.05) | 0.10 |
| Zimbabwe | SHINE (HIV+) (48) | 337 | 329 | 0.98 (0.61, 1.57) | 0.03 |
| **I² = 0.00, Tau² = 0.00** | | **8396** | **5513** | **0.92 (0.85, 0.99)** | |
| **Supplement duration − > 12m** | | | | | |
| Bangladesh | RDNS (35) | 1663 | 815 | 0.90 (0.82, 0.98) | 0.55 |
| Bangladesh | WASH−B (36) | 1163 | 3463 | 0.90 (0.82, 1.00) | 0.40 |
| Kenya | WASH−B (42) | 1461 | 5176 | 0.92 (0.67, 1.25) | 0.04 |
| Mali | PROMIS (46) | | | | |
| Mali | PROMIS CS (46) | | | | |
| **I² = 0.00, Tau² = 0.00** | | **4287** | **9454** | **0.90 (0.84, 0.96)** | |

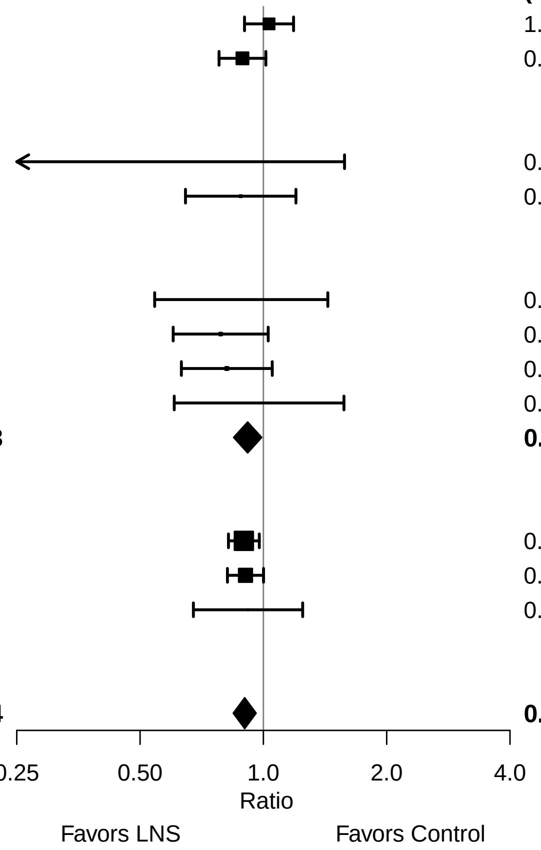

##### Supplemental figure 4P: Small head size prevalence ratio

#### 4P7: Stratified by Frequency of contact

**Frequency of contact**
**(p−diff = 0.591)**

| Country | Trial | N | N | PR (95% CI) | W |
|---|---|---|---|---|---|
| **Frequency of contact − Monthly** | | | | | |
| Bangladesh | RDNS (35) | 1663 | 815 | 0.90 (0.82, 0.98) | 0.81 |
| Burkina Faso | PROMIS (38) | | | | |
| Burkina Faso | PROMIS CS (38) | | | | |
| Haiti | HAITI (41) | | | | |
| Kenya | WASH−B (42) | 1461 | 5176 | 0.92 (0.67, 1.25) | 0.06 |
| Madagascar | MAHAY (43) | | | | |
| Mali | PROMIS (46) | | | | |
| Mali | PROMIS CS (46) | | | | |
| Zimbabwe | SHINE (HIV−) (47) | 1871 | 1783 | 0.81 (0.63, 1.05) | 0.09 |
| Zimbabwe | SHINE (HIV+) (48) | 337 | 329 | 0.98 (0.61, 1.57) | 0.03 |
| **$I^2$ = 0.00, $Tau^2$ = 0.00** | | **5332** | **8103** | **0.89 (0.82, 0.96)** | |
| **Frequency of contact − Weekly** | | | | | |
| Bangladesh | JiVitA−4 (34) | 2885 | 1267 | 1.03 (0.90, 1.18) | 0.22 |
| Bangladesh | WASH−B (36) | 1163 | 3463 | 0.90 (0.82, 1.00) | 0.41 |
| Burkina Faso | iLiNS−Zinc (37) | 1946 | 664 | 0.89 (0.78, 1.01) | 0.24 |
| Ghana | GHANA (39) | 98 | 96 | 0.33 (0.07, 1.58) | 0.00 |
| Ghana | iLiNS−DYADG (40) | 347 | 692 | 0.88 (0.65, 1.20) | 0.04 |
| Malawi | iLiNS−DYADM (44) | 220 | 444 | 0.88 (0.54, 1.44) | 0.02 |
| Malawi | iLiNS−DOSE (45) | 692 | 238 | 0.79 (0.60, 1.03) | 0.06 |
| **$I^2$ = 0.02, $Tau^2$ = 0.00** | | **7351** | **6864** | **0.92 (0.86, 0.98)** | |

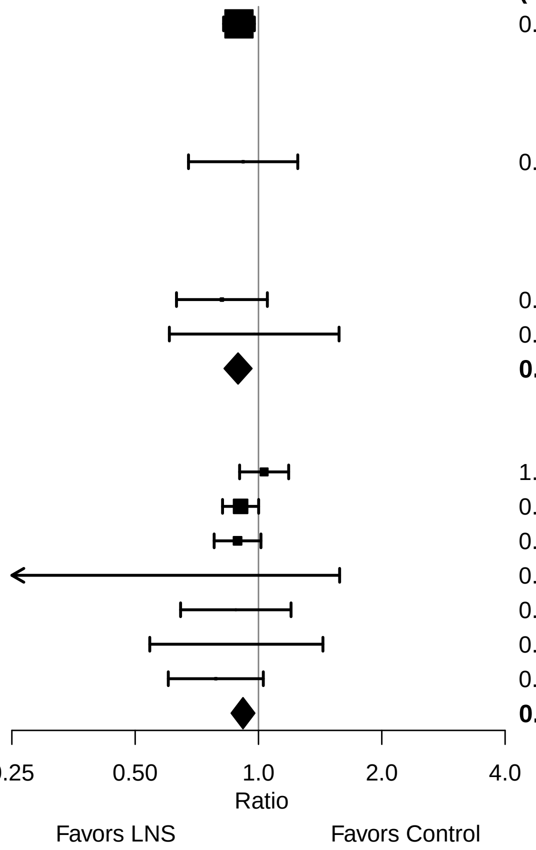

**Supplemental figure 4P: Small head size prevalence ratio**

#### 4P8: Stratified by Average SQ-LNS compliance

**Average SQ−LNS compliance**
**(p−diff = 0.264)**

| Country | Trial | N | N | PR (95% CI) | W |
|---|---|---|---|---|---|
| **Average SQ−LNS compliance − Low** | | | | | |
| Burkina Faso | PROMIS (38) | | | | |
| Burkina Faso | PROMIS CS (38) | | | | |
| Ghana | iLiNS−DYADG (40) | 347 | 692 | 0.88 (0.65, 1.20) | 0.21 |
| Malawi | iLiNS−DYADM (44) | 220 | 444 | 0.88 (0.54, 1.44) | 0.09 |
| Malawi | iLiNS−DOSE (45) | 692 | 238 | 0.79 (0.60, 1.03) | 0.29 |
| Mali | PROMIS (46) | | | | |
| Mali | PROMIS CS (46) | | | | |
| Zimbabwe | SHINE (HIV−) (47) | 1871 | 1783 | 0.81 (0.63, 1.05) | 0.32 |
| Zimbabwe | SHINE (HIV+) (48) | 337 | 329 | 0.98 (0.61, 1.57) | 0.09 |
| **I² = 0.00, Tau² = 0.00** | | **3467** | **3486** | **0.84 (0.73, 0.97)** | |
| **Average SQ−LNS compliance − High** | | | | | |
| Bangladesh | JiVitA−4 (34) | 2885 | 1267 | 1.03 (0.90, 1.18) | 0.15 |
| Bangladesh | RDNS (35) | 1663 | 815 | 0.90 (0.82, 0.98) | 0.38 |
| Bangladesh | WASH−B (36) | 1163 | 3463 | 0.90 (0.82, 1.00) | 0.28 |
| Burkina Faso | iLiNS−Zinc (37) | 1946 | 664 | 0.89 (0.78, 1.01) | 0.16 |
| Ghana | GHANA (39) | 98 | 96 | 0.33 (0.07, 1.58) | 0.00 |
| Haiti | HAITI (41) | | | | |
| Kenya | WASH−B (42) | 1461 | 5176 | 0.92 (0.67, 1.25) | 0.03 |
| **I² = 0.01, Tau² = 0.00** | | **9216** | **11481** | **0.92 (0.87, 0.97)** | |

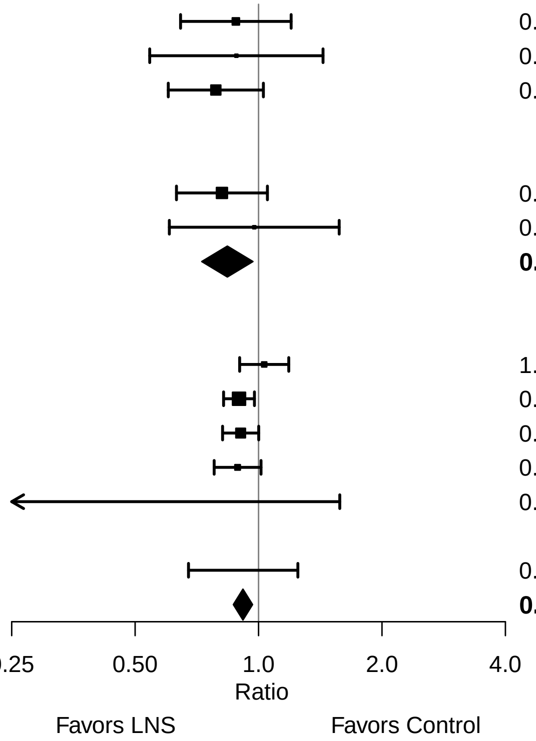

### Supplemental figure 4Q: Small head size prevalence difference

#### 4Q1: Stratified by Geographic region

**Geographic region**
**(p−diff = 0.664)**

| Country | Trial | N | N | PD (95% CI) | W |
|---|---|---|---|---|---|
| **Geographic region − SEAR** | | | | | |
| Bangladesh | JiVitA−4 (34) | 2885 | 1267 | 0.00 (−0.03, 0.02) | 0.48 |
| Bangladesh | RDNS (35) | 1663 | 815 | −0.04 (−0.09, 0.00) | 0.20 |
| Bangladesh | WASH−B (36) | 1163 | 3463 | −0.03 (−0.06, 0.00) | 0.32 |
| **I² = 0.50, Tau² = 0.00** | | **5711** | **5545** | **−0.02 (−0.05, 0.00)** | |
| **Geographic region − AFR** | | | | | |
| Burkina Faso | iLiNS−Zinc (37) | 1946 | 664 | −0.04 (−0.08, −0.01) | 0.11 |
| Burkina Faso | PROMIS (38) | | | | |
| Burkina Faso | PROMIS CS (38) | | | | |
| Ghana | GHANA (39) | 98 | 96 | −0.04 (−0.10, 0.01) | 0.04 |
| Ghana | iLiNS−DYADG (40) | 347 | 692 | −0.02 (−0.06, 0.03) | 0.06 |
| Kenya | WASH−B (42) | 1461 | 5176 | 0.00 (−0.02, 0.01) | 0.34 |
| Madagascar | MAHAY (43) | | | | |
| Malawi | iLiNS−DYADM (44) | 220 | 444 | −0.01 (−0.06, 0.04) | 0.06 |
| Malawi | iLiNS−DOSE (45) | 692 | 238 | −0.04 (−0.10, 0.01) | 0.05 |
| Mali | PROMIS (46) | | | | |
| Mali | PROMIS CS (46) | | | | |
| Zimbabwe | SHINE (HIV−) (47) | 1871 | 1783 | −0.01 (−0.03, 0.00) | 0.29 |
| Zimbabwe | SHINE (HIV+) (48) | 337 | 329 | 0.00 (−0.05, 0.04) | 0.06 |
| **I² = 0.14, Tau² = 0.00** | | **6972** | **9422** | **−0.01 (−0.03, 0.00)** | |

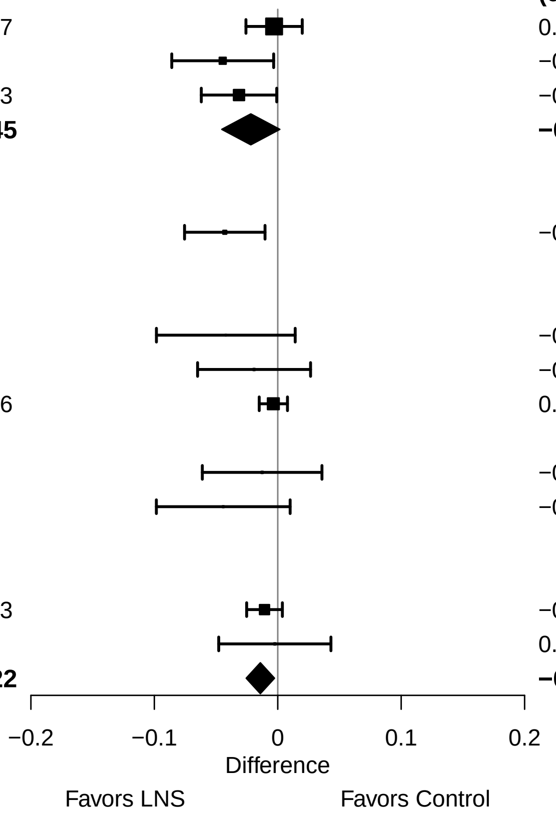

#### Supplemental figure 4Q: Small head size prevalence difference

### 4Q2: Stratified by Stunting burden

**Stunting burden**
**(p−diff = 0.345)**

| Country | Trial | N | N | PD (95% CI) | W |
|---|---|---|---|---|---|
| **Stunting burden − Less than 35%** | | | | | |
| Burkina Faso | PROMIS (38) | | | | |
| Burkina Faso | PROMIS CS (38) | | | | |
| Ghana | GHANA (39) | 98 | 96 | −0.04 (−0.10, 0.01) | 0.09 |
| Ghana | iLiNS−DYADG (40) | 347 | 692 | −0.02 (−0.06, 0.03) | 0.13 |
| Haiti | HAITI (41) | | | | |
| Kenya | WASH−B (42) | 1461 | 5176 | 0.00 (−0.02, 0.01) | 0.67 |
| Malawi | iLiNS−DYADM (44) | 220 | 444 | −0.01 (−0.06, 0.04) | 0.11 |
| **I² = 0.00, Tau² = 0.00** | | **2126** | **6408** | **−0.01 (−0.02, 0.00)** | |
| **Stunting burden − More than 35%** | | | | | |
| Bangladesh | JiVitA−4 (34) | 2885 | 1267 | 0.00 (−0.03, 0.02) | 0.20 |
| Bangladesh | RDNS (35) | 1663 | 815 | −0.04 (−0.09, 0.00) | 0.08 |
| Bangladesh | WASH−B (36) | 1163 | 3463 | −0.03 (−0.06, 0.00) | 0.14 |
| Burkina Faso | iLiNS−Zinc (37) | 1946 | 664 | −0.04 (−0.08, −0.01) | 0.12 |
| Madagascar | MAHAY (43) | | | | |
| Malawi | iLiNS−DOSE (45) | 692 | 238 | −0.04 (−0.10, 0.01) | 0.05 |
| Mali | PROMIS (46) | | | | |
| Mali | PROMIS CS (46) | | | | |
| Zimbabwe | SHINE (HIV−) (47) | 1871 | 1783 | −0.01 (−0.03, 0.00) | 0.33 |
| Zimbabwe | SHINE (HIV+) (48) | 337 | 329 | 0.00 (−0.05, 0.04) | 0.07 |
| **I² = 0.30, Tau² = 0.00** | | **10557** | **8559** | **−0.02 (−0.03, −0.01)** | |

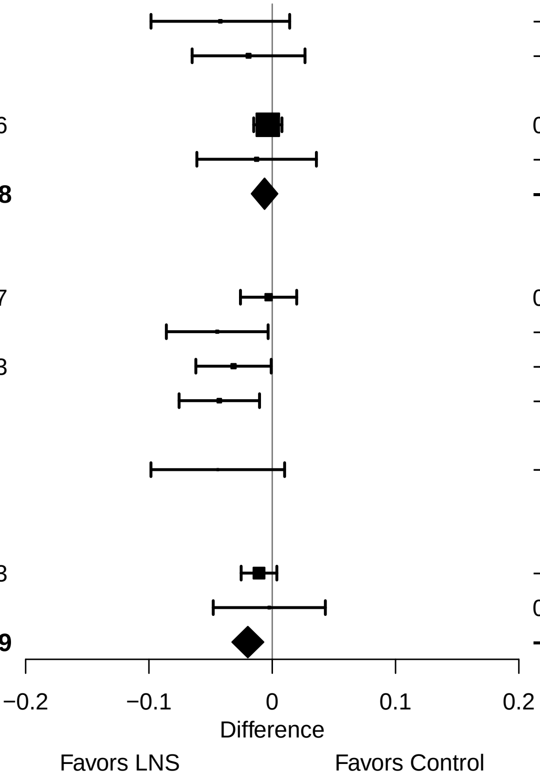

#### Supplemental figure 4Q: Small head size prevalence difference

##### 4Q3: Stratified by Malaria prevalence

**Malaria prevalence**
**(p−diff = 0.025)**

| Country | Trial | N | N | PD (95% CI) | W |
|---|---|---|---|---|---|
| **Malaria prevalence − Less than 10%** | | | | | |
| Bangladesh | JiVitA−4 (34) | 2885 | 1267 | 0.00 (−0.03, 0.02) | 0.17 |
| Bangladesh | RDNS (35) | 1663 | 815 | −0.04 (−0.09, 0.00) | 0.07 |
| Bangladesh | WASH−B (36) | 1163 | 3463 | −0.03 (−0.06, 0.00) | 0.11 |
| Haiti | HAITI (41) | | | | |
| Kenya | WASH−B (42) | 1461 | 5176 | 0.00 (−0.02, 0.01) | 0.32 |
| Madagascar | MAHAY (43) | | | | |
| Zimbabwe | SHINE (HIV−) (47) | 1871 | 1783 | −0.01 (−0.03, 0.00) | 0.27 |
| Zimbabwe | SHINE (HIV+) (48) | 337 | 329 | 0.00 (−0.05, 0.04) | 0.06 |
| **I² = 0.19, Tau² = 0.00** | | **9380** | **12833** | **−0.01 (−0.02, 0.00)** | |
| **Malaria prevalence − At least 10%** | | | | | |
| Burkina Faso | iLiNS−Zinc (37) | 1946 | 664 | −0.04 (−0.08, −0.01) | 0.34 |
| Burkina Faso | PROMIS (38) | | | | |
| Burkina Faso | PROMIS CS (38) | | | | |
| Ghana | GHANA (39) | 98 | 96 | −0.04 (−0.10, 0.01) | 0.14 |
| Ghana | iLiNS−DYADG (40) | 347 | 692 | −0.02 (−0.06, 0.03) | 0.20 |
| Malawi | iLiNS−DYADM (44) | 220 | 444 | −0.01 (−0.06, 0.04) | 0.18 |
| Malawi | iLiNS−DOSE (45) | 692 | 238 | −0.04 (−0.10, 0.01) | 0.15 |
| Mali | PROMIS (46) | | | | |
| Mali | PROMIS CS (46) | | | | |
| **I² = 0.00, Tau² = 0.00** | | **3303** | **2134** | **−0.03 (−0.05, −0.01)** | |

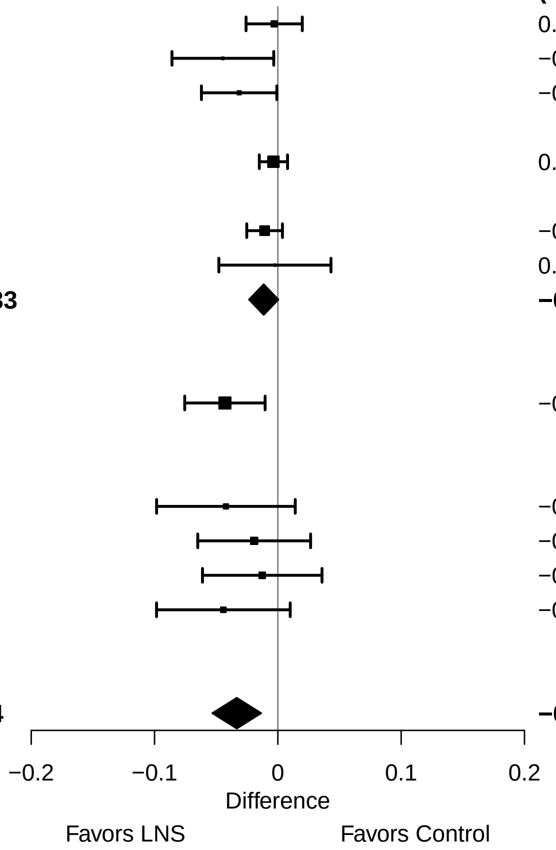

#### Supplemental figure 4Q: Small head size prevalence difference

#### 4Q4: Stratified by Source water quality

**Source water quality**
**(p−diff = 0.301)**

| Country | Trial | N | N | PD (95% CI) | W |
|---|---|---|---|---|---|
| **Source water quality − Improved** | | | | | |
| Bangladesh | JiVitA−4 (34) | 2885 | 1267 | 0.00 (−0.03, 0.02) | 0.30 |
| Bangladesh | RDNS (35) | 1663 | 815 | −0.04 (−0.09, 0.00) | 0.15 |
| Bangladesh | WASH−B (36) | 571 | 1121 | −0.05 (−0.09, 0.00) | 0.12 |
| Ghana | GHANA (39) | 98 | 96 | −0.04 (−0.10, 0.01) | 0.09 |
| Ghana | iLiNS−DYADG (40) | 347 | 692 | −0.02 (−0.06, 0.03) | 0.13 |
| Haiti | HAITI (41) | | | | |
| Malawi | iLiNS−DYADM (44) | 220 | 444 | −0.01 (−0.06, 0.04) | 0.11 |
| Malawi | iLiNS−DOSE (45) | 692 | 238 | −0.04 (−0.10, 0.01) | 0.10 |
| **I² = 0.02, Tau² = 0.00** | | **6476** | **4673** | **−0.02 (−0.04, −0.01)** | |
| **Source water quality − Unimproved** | | | | | |
| Burkina Faso | iLiNS−Zinc (37) | 1946 | 664 | −0.04 (−0.08, −0.01) | 0.20 |
| Burkina Faso | PROMIS (38) | | | | |
| Burkina Faso | PROMIS CS (38) | | | | |
| Kenya | WASH−B (42) | 696 | 2266 | −0.01 (−0.03, 0.00) | 0.40 |
| Madagascar | MAHAY (43) | | | | |
| Mali | PROMIS (46) | | | | |
| Mali | PROMIS CS (46) | | | | |
| Zimbabwe | SHINE (HIV−) (47) | 887 | 873 | 0.00 (−0.02, 0.02) | 0.33 |
| Zimbabwe | SHINE (HIV+) (48) | 147 | 146 | 0.03 (−0.03, 0.10) | 0.07 |
| **I² = 0.55, Tau² = 0.00** | | **3676** | **3949** | **−0.01 (−0.04, 0.01)** | |

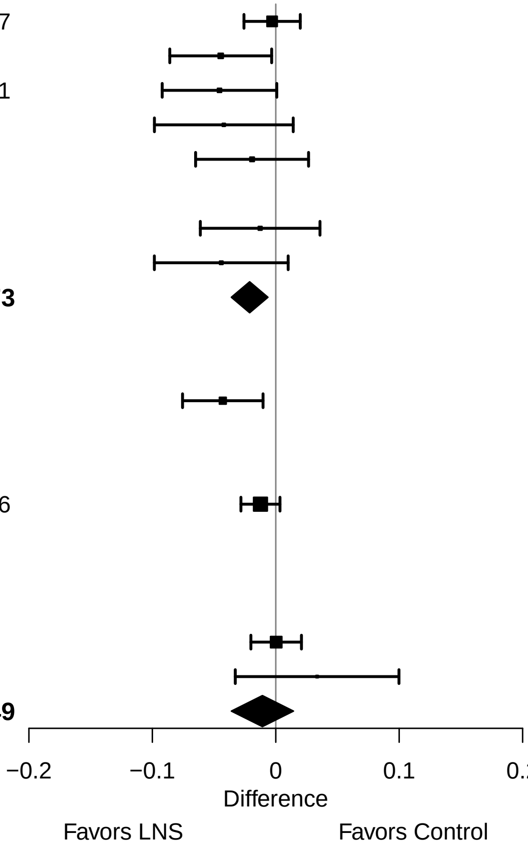

#### Supplemental figure 4Q: Small head size prevalence difference

##### 4Q5: Stratified by Sanitation

**Sanitation**
**(p−diff = 0.442)**

| Country | Trial | N | N | PD (95% CI) | W |
|---|---|---|---|---|---|
| **Sanitation − Improved** | | | | | |
| Bangladesh | JiVitA−4 (34) | 2885 | 1267 | 0.00 (−0.03, 0.02) | 0.38 |
| Bangladesh | RDNS (35) | 1663 | 815 | −0.04 (−0.09, 0.00) | 0.19 |
| Bangladesh | WASH−B (36) | 571 | 1121 | −0.05 (−0.09, 0.00) | 0.16 |
| Burkina Faso | PROMIS (38) | | | | |
| Burkina Faso | PROMIS CS (38) | | | | |
| Ghana | GHANA (39) | 98 | 96 | −0.04 (−0.10, 0.01) | 0.11 |
| Ghana | iLiNS−DYADG (40) | 347 | 692 | −0.02 (−0.06, 0.03) | 0.16 |
| Haiti | HAITI (41) | | | | |
| Mali | PROMIS (46) | | | | |
| Mali | PROMIS CS (46) | | | | |
| **I² = 0.24, Tau² = 0.00** | | **5564** | **3991** | **−0.02 (−0.04, 0.00)** | |
| **Sanitation − Unimproved** | | | | | |
| Burkina Faso | iLiNS−Zinc (37) | 1946 | 664 | −0.04 (−0.08, −0.01) | 0.17 |
| Kenya | WASH−B (42) | 696 | 2266 | −0.01 (−0.03, 0.00) | 0.33 |
| Madagascar | MAHAY (43) | | | | |
| Malawi | iLiNS−DYADM (44) | 220 | 444 | −0.01 (−0.06, 0.04) | 0.09 |
| Malawi | iLiNS−DOSE (45) | 692 | 238 | −0.04 (−0.10, 0.01) | 0.08 |
| Zimbabwe | SHINE (HIV−) (47) | 887 | 873 | 0.00 (−0.02, 0.02) | 0.27 |
| Zimbabwe | SHINE (HIV+) (48) | 147 | 146 | 0.03 (−0.03, 0.10) | 0.06 |
| **I² = 0.38, Tau² = 0.00** | | **4588** | **4631** | **−0.01 (−0.03, 0.00)** | |

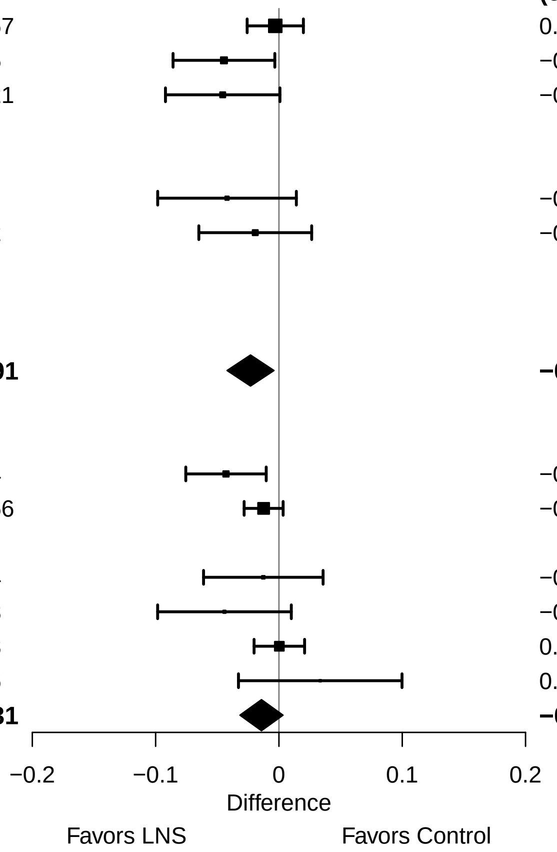

#### Supplemental figure 4Q: Small head size prevalence difference

### 4Q6: Stratified by Supplement duration

**Supplement duration**
**(p−diff = 0.995)**

**Supplement duration − 12m or less**

| Country | Trial | N | N | PD (95% CI) | W |
|---|---|---|---|---|---|
| Bangladesh | JiVitA−4 (34) | 2885 | 1267 | 0.00 (−0.03, 0.02) | 0.21 |
| Burkina Faso | iLiNS−Zinc (37) | 1946 | 664 | −0.04 (−0.08, −0.01) | 0.13 |
| Burkina Faso | PROMIS (38) | | | | |
| Burkina Faso | PROMIS CS (38) | | | | |
| Ghana | GHANA (39) | 98 | 96 | −0.04 (−0.10, 0.01) | 0.05 |
| Ghana | iLiNS−DYADG (40) | 347 | 692 | −0.02 (−0.06, 0.03) | 0.07 |
| Haiti | HAITI (41) | | | | |
| Madagascar | MAHAY (43) | | | | |
| Malawi | iLiNS−DYADM (44) | 220 | 444 | −0.01 (−0.06, 0.04) | 0.07 |
| Malawi | iLiNS−DOSE (45) | 692 | 238 | −0.04 (−0.10, 0.01) | 0.05 |
| Zimbabwe | SHINE (HIV−) (47) | 1871 | 1783 | −0.01 (−0.03, 0.00) | 0.34 |
| Zimbabwe | SHINE (HIV+) (48) | 337 | 329 | 0.00 (−0.05, 0.04) | 0.07 |
| **I² = 0.00, Tau² = 0.00** | | **8396** | **5513** | **−0.01 (−0.02, 0.00)** | |

**Supplement duration − > 12m**

| Country | Trial | N | N | PD (95% CI) | W |
|---|---|---|---|---|---|
| Bangladesh | RDNS (35) | 1663 | 815 | −0.04 (−0.09, 0.00) | 0.14 |
| Bangladesh | WASH−B (36) | 1163 | 3463 | −0.03 (−0.06, 0.00) | 0.23 |
| Kenya | WASH−B (42) | 1461 | 5176 | 0.00 (−0.02, 0.01) | 0.63 |
| Mali | PROMIS (46) | | | | |
| Mali | PROMIS CS (46) | | | | |
| **I² = 0.65, Tau² = 0.00** | | **4287** | **9454** | **−0.02 (−0.04, 0.00)** | |

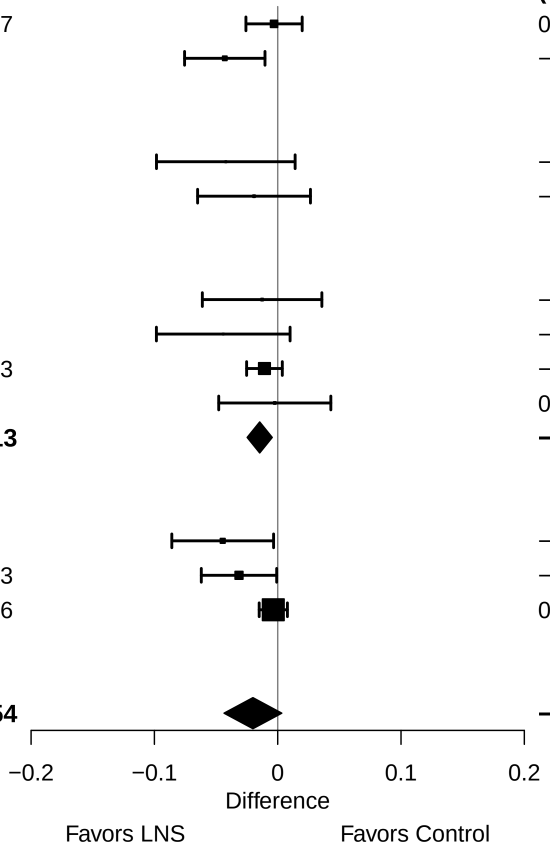

#### Supplemental figure 4Q: Small head size prevalence difference

### 4Q7: Stratified by Frequency of contact

**Frequency of contact**
**(p−diff = 0.091)**

| **Frequency of contact − Monthly** | | | | **PD** | |
|---|---|---|---|---|---|
| **Country** | **Trial** | **N** | **N** | **(95% CI)** | **W** |
| Bangladesh | RDNS (35) | 1663 | 815 | −0.04 (−0.09, 0.00) | 0.10 |
| Burkina Faso | PROMIS (38) | | | | |
| Burkina Faso | PROMIS CS (38) | | | | |
| Haiti | HAITI (41) | | | | |
| Kenya | WASH−B (42) | 1461 | 5176 | 0.00 (−0.02, 0.01) | 0.44 |
| Madagascar | MAHAY (43) | | | | |
| Mali | PROMIS (46) | | | | |
| Mali | PROMIS CS (46) | | | | |
| Zimbabwe | SHINE (HIV−) (47) | 1871 | 1783 | −0.01 (−0.03, 0.00) | 0.38 |
| Zimbabwe | SHINE (HIV+) (48) | 337 | 329 | 0.00 (−0.05, 0.04) | 0.08 |
| **I² = 0.21, Tau² = 0.00** | | **5332** | **8103** | **−0.01 (−0.02, 0.00)** | |
| **Frequency of contact − Weekly** | | | | | |
| Bangladesh | JiVitA−4 (34) | 2885 | 1267 | 0.00 (−0.03, 0.02) | 0.29 |
| Bangladesh | WASH−B (36) | 1163 | 3463 | −0.03 (−0.06, 0.00) | 0.19 |
| Burkina Faso | iLiNS−Zinc (37) | 1946 | 664 | −0.04 (−0.08, −0.01) | 0.18 |
| Ghana | GHANA (39) | 98 | 96 | −0.04 (−0.10, 0.01) | 0.07 |
| Ghana | iLiNS−DYADG (40) | 347 | 692 | −0.02 (−0.06, 0.03) | 0.10 |
| Malawi | iLiNS−DYADM (44) | 220 | 444 | −0.01 (−0.06, 0.04) | 0.09 |
| Malawi | iLiNS−DOSE (45) | 692 | 238 | −0.04 (−0.10, 0.01) | 0.07 |
| **I² = 0.00, Tau² = 0.00** | | **7351** | **6864** | **−0.02 (−0.04, −0.01)** | |

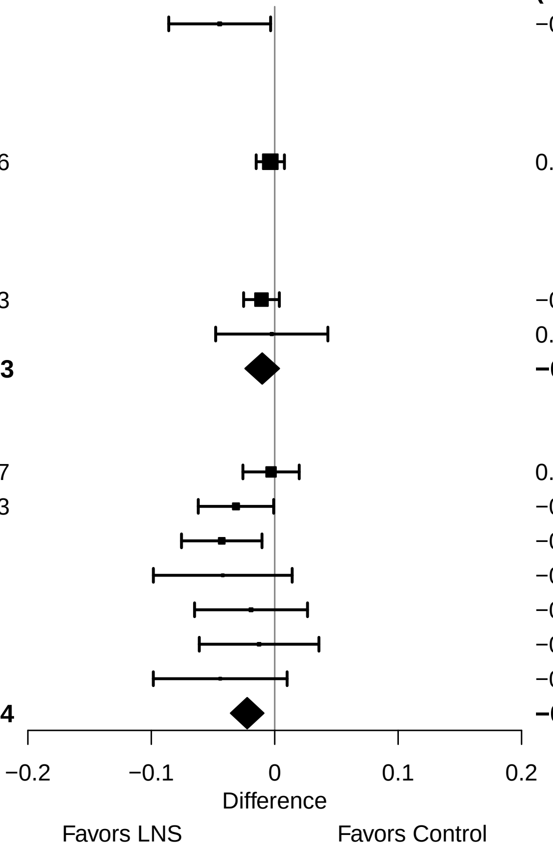

#### Supplemental figure 4Q: Small head size prevalence difference

### 4Q8: Stratified by Average SQ-LNS compliance

**Average SQ−LNS compliance**
**(p−diff = 0.682)**

| Country | Trial | N | N | PD (95% CI) | W |
|---|---|---|---|---|---|
| **Average SQ−LNS compliance − Low** | | | | | |
| Burkina Faso | PROMIS (38) | | | | |
| Burkina Faso | PROMIS CS (38) | | | | |
| Ghana | iLiNS−DYADG (40) | 347 | 692 | −0.02 (−0.06, 0.03) | 0.12 |
| Malawi | iLiNS−DYADM (44) | 220 | 444 | −0.01 (−0.06, 0.04) | 0.11 |
| Malawi | iLiNS−DOSE (45) | 692 | 238 | −0.04 (−0.10, 0.01) | 0.09 |
| Mali | PROMIS (46) | | | | |
| Mali | PROMIS CS (46) | | | | |
| Zimbabwe | SHINE (HIV−) (47) | 1871 | 1783 | −0.01 (−0.03, 0.00) | 0.56 |
| Zimbabwe | SHINE (HIV+) (48) | 337 | 329 | 0.00 (−0.05, 0.04) | 0.12 |
| **I² = 0.00, Tau² = 0.00** | | **3467** | **3486** | **−0.01 (−0.03, 0.00)** | |
| **Average SQ−LNS compliance − High** | | | | | |
| Bangladesh | JiVitA−4 (34) | 2885 | 1267 | 0.00 (−0.03, 0.02) | 0.21 |
| Bangladesh | RDNS (35) | 1663 | 815 | −0.04 (−0.09, 0.00) | 0.09 |
| Bangladesh | WASH−B (36) | 1163 | 3463 | −0.03 (−0.06, 0.00) | 0.14 |
| Burkina Faso | iLiNS−Zinc (37) | 1946 | 664 | −0.04 (−0.08, −0.01) | 0.13 |
| Ghana | GHANA (39) | 98 | 96 | −0.04 (−0.10, 0.01) | 0.05 |
| Haiti | HAITI (41) | | | | |
| Kenya | WASH−B (42) | 1461 | 5176 | 0.00 (−0.02, 0.01) | 0.39 |
| **I² = 0.55, Tau² = 0.00** | | **9216** | **11481** | **−0.02 (−0.04, 0.00)** | |

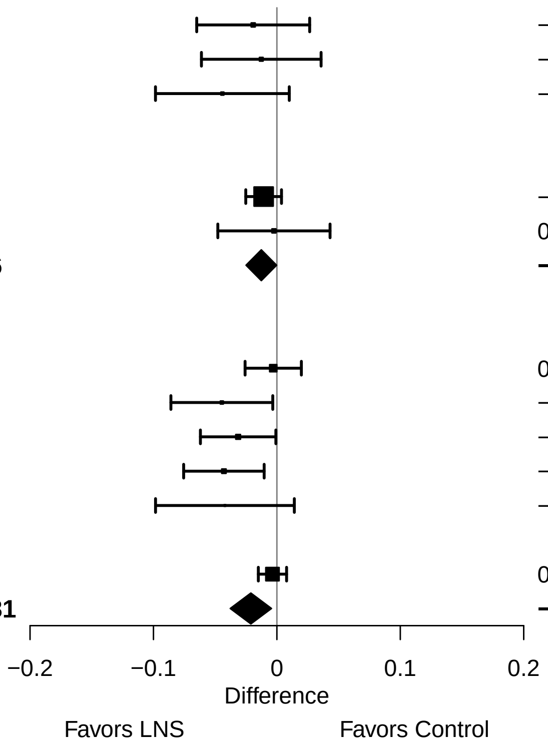

### Supplemental figure 5: Pooled effect of SQ-LNS on prevalence ratios for low MUAC, acute malnutrition, underweight, and small head size stratified by study-level and individual-level characteristics

These figures show pooled effects of SQ-LNS within study-level and individual-level characteristic subgroups along with the p-for-interaction. The labels on the left y-axis correspond to the characteristic subgroups and their sample sizes. The values on the right indicate the pooled prevalence ratio and confidence interval within that subgroup. MUAC, mid-upper arm circumference.

### Supplemental figure 5A: Low MUAC prevalence ratio

#### 5A1: Stratified by study-level characteristics

| Effect modifier (p−diff) | LNS N | Control N | PR (95% CI) |
|---|---|---|---|
| Stunting burden (p=0.925) | | | |
| Less than 35% | 3306 | 7670 | 0.82 (0.69, 0.98) |
| More than 35% | 12565 | 8233 | 0.82 (0.70, 0.96) |
| Malaria prevalence (p=0.684) | | | |
| At least 10% | 5961 | 4866 | 0.79 (0.68, 0.92) |
| Less than 10% | 9910 | 11037 | 0.84 (0.73, 0.98) |
| Source water quality (p=0.367) | | | |
| Improved | 5823 | 3463 | 0.89 (0.76, 1.05) |
| Unimproved | 8111 | 8434 | 0.80 (0.70, 0.92) |
| Sanitation (p=0.291) | | | |
| Improved | 7646 | 5602 | 0.88 (0.78, 0.99) |
| Unimproved | 6288 | 6295 | 0.77 (0.65, 0.93) |
| Supplement duration (p=0.928) | | | |
| 12 mo or less | 11286 | 8428 | 0.82 (0.72, 0.93) |
| > 12 mo | 4585 | 7475 | 0.82 (0.67, 1.00) |
| Frequency of contact (p=0.354) | | | |
| Weekly | 6119 | 3314 | 0.77 (0.64, 0.91) |
| Monthly | 9752 | 12589 | 0.86 (0.76, 0.97) |
| Average SQ−LNS compliance (p=0.377) | | | |
| High | 7980 | 7937 | 0.77 (0.64, 0.93) |
| Low | 6211 | 6308 | 0.86 (0.73, 1.01) |

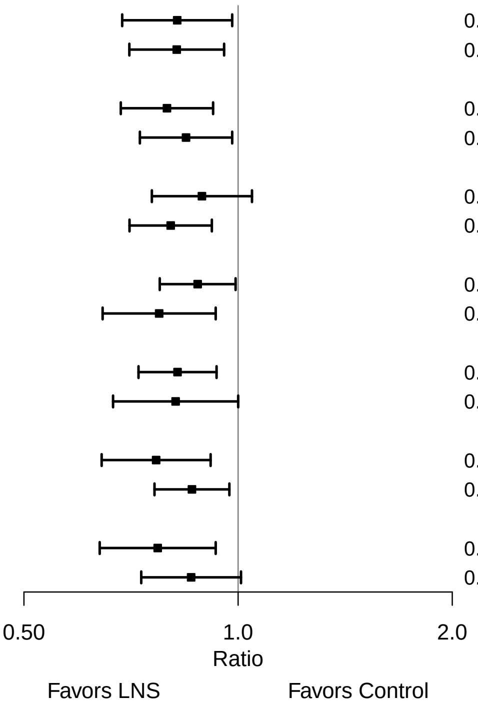

##### Supplemental figure 5A: Low MUAC prevalence ratio

#### 5A2: Stratified by individual-level maternal and child characteristics

| Effect modifier (p−for−interaction) | LNS N | Control N | PR (95% CI) |
|---|---|---|---|
| Maternal BMI (p=0.337) | | | |
| Less than 20 kg/m² | 2643 | 2629 | 0.84 (0.71, 0.99) |
| At least 20 kg/m² | 5015 | 6556 | 0.76 (0.65, 0.89) |
| Maternal age (p=0.741) | | | |
| Less than 25 y | 7068 | 6639 | 0.84 (0.74, 0.95) |
| At least 25 y | 6964 | 7323 | 0.80 (0.70, 0.91) |
| Maternal education (p=0.240) | | | |
| Primary or greater | 4299 | 4381 | 0.78 (0.66, 0.92) |
| Incomplete or no formal | 4773 | 5946 | 0.92 (0.81, 1.04) |
| Maternal depressive symptoms (p=0.798) | | | |
| At least 75th | 1875 | 2484 | 0.85 (0.64, 1.13) |
| Less than 75th | 4430 | 6410 | 0.85 (0.71, 1.02) |
| Child sex (p=0.002) | | | |
| Male | 7368 | 7357 | 0.93 (0.82, 1.05) |
| Female | 7250 | 7534 | 0.73 (0.65, 0.82) |
| Child birth order (p=0.002) | | | |
| Later born | 7377 | 8512 | 0.77 (0.69, 0.86) |
| Firstborn | 4490 | 3376 | 0.95 (0.81, 1.11) |

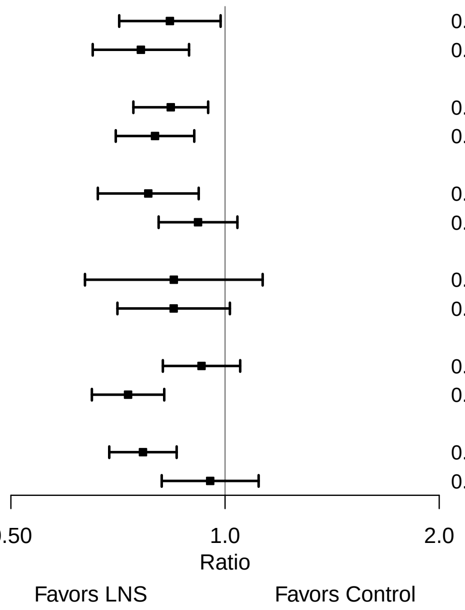

#### Supplemental figure 5A: Low MUAC prevalence ratio

### 5A3: Stratified by individual-level household characteristics

| Effect modifier (p−for−interaction) | LNS N | Control N | PR (95% CI) |
|---|---|---|---|
| Socio−economic status (p=0.129) | | | |
| At least median | 6492 | 6561 | 0.75 (0.66, 0.85) |
| Less than median | 5831 | 5800 | 0.86 (0.76, 0.96) |
| Food security (p=0.182) | | | |
| Mild to secure | 7226 | 8131 | 0.74 (0.66, 0.83) |
| Moderate to severe | 3423 | 2516 | 0.87 (0.74, 1.04) |
| Source Water quality (p=0.130) | | | |
| Improved | 2846 | 3621 | 0.88 (0.74, 1.03) |
| Unimproved | 3946 | 3535 | 0.73 (0.63, 0.84) |
| Sanitation (p=0.239) | | | |
| Improved | 4794 | 3320 | 0.84 (0.72, 0.97) |
| Unimproved | 2660 | 3317 | 0.94 (0.78, 1.13) |
| Home environment (p=0.543) | | | |
| At least median | 3410 | 4452 | 0.76 (0.63, 0.92) |
| Less than median | 2088 | 3353 | 0.89 (0.72, 1.12) |
| Season at time of assessment (p=0.301) | | | |
| Rainy | 4554 | 5979 | 0.78 (0.68, 0.90) |
| Dry | 4704 | 3306 | 0.81 (0.70, 0.95) |

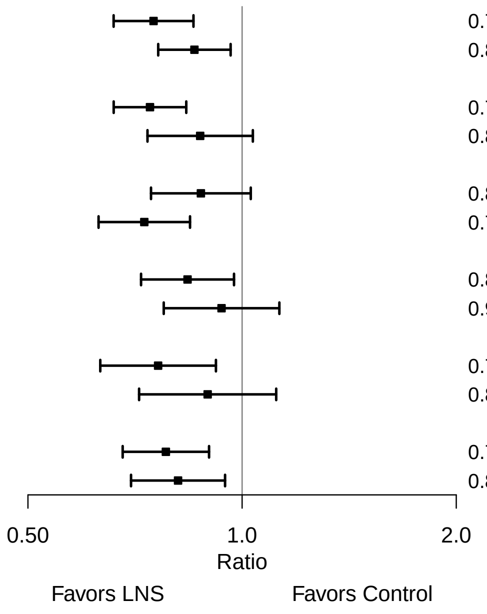

### Supplemental figure 5B: Acute malnutrition prevalence ratio

#### 5B1: Stratified by study-level characteristics

| Effect modifier (p−diff) | LNS N | Control N | PR (95% CI) |
|---|---|---|---|
| Stunting burden (p=0.835) | | | |
| Less than 35% | 3291 | 7588 | 0.85 (0.72, 1.00) |
| More than 35% | 12406 | 8155 | 0.87 (0.77, 0.98) |
| Malaria prevalence (p=0.526) | | | |
| At least 10% | 5921 | 4836 | 0.85 (0.74, 0.97) |
| Less than 10% | 9776 | 10907 | 0.88 (0.80, 0.96) |
| Source water quality (p=0.378) | | | |
| Improved | 5701 | 3407 | 0.89 (0.81, 0.99) |
| Unimproved | 8066 | 8376 | 0.84 (0.75, 0.93) |
| Sanitation (p=0.450) | | | |
| Improved | 7504 | 5519 | 0.88 (0.81, 0.96) |
| Unimproved | 6263 | 6264 | 0.85 (0.70, 1.02) |
| Supplement duration (p=0.607) | | | |
| 12 mo or less | 11139 | 8350 | 0.87 (0.79, 0.97) |
| > 12 mo | 4558 | 7393 | 0.84 (0.73, 0.96) |
| Frequency of contact (p=0.706) | | | |
| Weekly | 5991 | 3256 | 0.86 (0.69, 1.06) |
| Monthly | 9706 | 12487 | 0.87 (0.79, 0.97) |
| Average SQ−LNS compliance (p=0.269) | | | |
| High | 7849 | 7812 | 0.81 (0.71, 0.92) |
| Low | 6173 | 6276 | 0.90 (0.79, 1.02) |

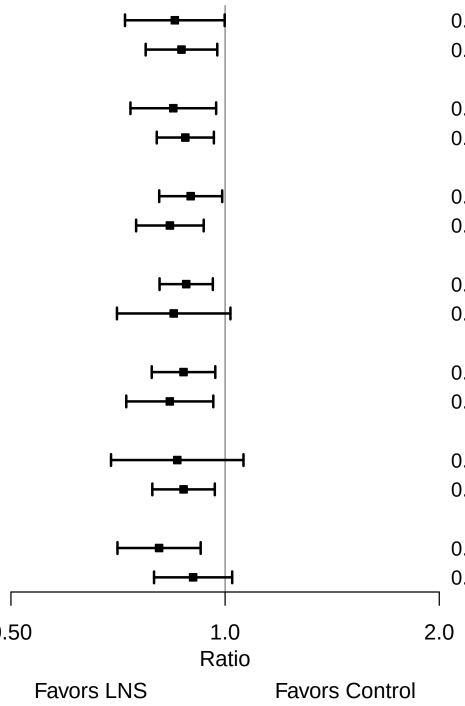

##### Supplemental figure 5B: Acute malnutrition prevalence ratio

#### 5B2: Stratified by individual-level maternal and child characteristics

| Effect modifier (p−for−interaction) | LNS N | Control N | PR (95% CI) |
|---|---|---|---|
| Maternal height (p=0.435) | | | |
| At least 150.1 cm | 3335 | 2574 | 0.93 (0.80, 1.09) |
| Less than 150.1 cm | 1471 | 983 | 0.88 (0.70, 1.09) |
| Maternal BMI (p=0.490) | | | |
| Less than 20 kg/m² | 2655 | 1715 | 0.84 (0.74, 0.94) |
| At least 20 kg/m² | 5380 | 3795 | 0.82 (0.70, 0.95) |
| Maternal age (p=0.429) | | | |
| At least 25 y | 6543 | 6888 | 0.83 (0.74, 0.92) |
| Less than 25 y | 6824 | 6414 | 0.89 (0.80, 0.99) |
| Maternal education (p=0.659) | | | |
| Primary or greater | 4492 | 4857 | 0.87 (0.77, 0.98) |
| Incomplete or no formal | 4789 | 6023 | 0.92 (0.82, 1.02) |
| Maternal depressive symptoms (p=0.532) | | | |
| At least 75th | 2035 | 2770 | 0.81 (0.65, 1.00) |
| Less than 75th | 4903 | 7032 | 0.88 (0.76, 1.03) |
| Child sex (p=0.307) | | | |
| Male | 7013 | 7016 | 0.88 (0.79, 0.97) |
| Female | 6939 | 7214 | 0.84 (0.75, 0.92) |
| Child birth order (p=0.124) | | | |
| Later born | 8620 | 9626 | 0.80 (0.73, 0.88) |
| Firstborn | 4873 | 3805 | 0.93 (0.83, 1.04) |

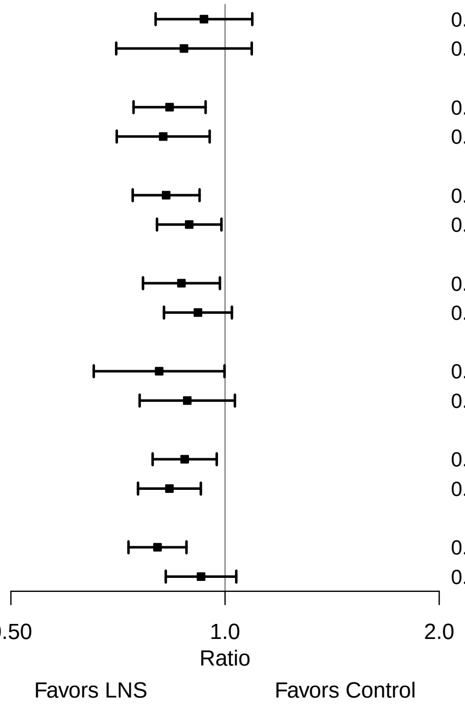

#### Supplemental figure 5B: Acute malnutrition prevalence ratio

### 5B3: Stratified by individual-level household characteristics

| Effect modifier (p−for−interaction) | LNS N | Control N | PR (95% CI) |
|---|---|---|---|
| Socio−economic status (p=0.312) | | | |
| At least median | 7393 | 7535 | 0.82 (0.73, 0.91) |
| Less than median | 6684 | 6835 | 0.89 (0.81, 0.98) |
| Food security (p=0.565) | | | |
| Mild to secure | 7131 | 4863 | 0.89 (0.80, 0.99) |
| Moderate to severe | 3440 | 2345 | 0.78 (0.69, 0.88) |
| Source Water quality (p=0.789) | | | |
| Improved | 2651 | 2273 | 0.83 (0.70, 0.97) |
| Unimproved | 3708 | 2874 | 0.83 (0.72, 0.96) |
| Sanitation (p=0.013) | | | |
| Improved | 4882 | 3145 | 0.82 (0.73, 0.91) |
| Unimproved | 2528 | 1931 | 1.01 (0.88, 1.16) |
| Home environment (p=0.310) | | | |
| At least median | 3614 | 4835 | 0.73 (0.63, 0.85) |
| Less than median | 2201 | 3567 | 0.91 (0.75, 1.10) |
| Season at time of assessment (p=0.172) | | | |
| Rainy | 4757 | 5992 | 0.89 (0.80, 1.00) |
| Dry | 5052 | 3395 | 0.79 (0.70, 0.89) |

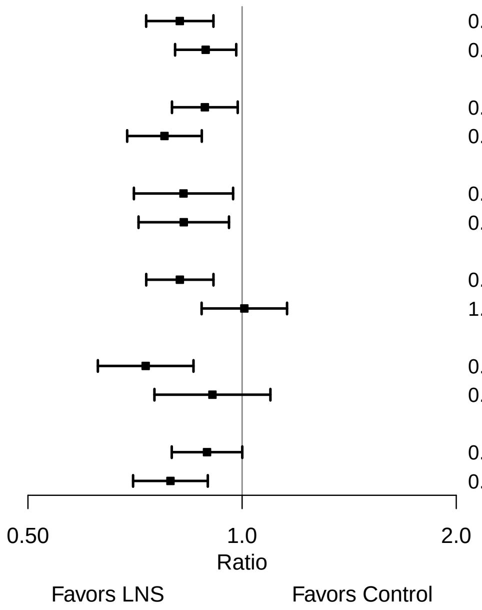

### Supplemental figure 5C: Underweight prevalence ratio

#### 5C1: Stratified by study-level characteristics

| Effect modifier (p−diff) | LNS N | Control N | PR (95% CI) |
|---|---|---|---|
| Geographic region (p=0.909) | | | |
| AFR | 11428 | 13913 | 0.86 (0.79, 0.94) |
| SEAR | 5640 | 5508 | 0.87 (0.79, 0.96) |
| Stunting burden (p=0.979) | | | |
| Less than 35% | 3561 | 7895 | 0.86 (0.77, 0.96) |
| More than 35% | 13655 | 11676 | 0.87 (0.79, 0.95) |
| Malaria prevalence (p=0.998) | | | |
| At least 10% | 6060 | 4949 | 0.86 (0.78, 0.95) |
| Less than 10% | 11156 | 14622 | 0.87 (0.79, 0.95) |
| Source water quality (p=0.601) | | | |
| Improved | 6564 | 4790 | 0.91 (0.86, 0.97) |
| Unimproved | 8126 | 8443 | 0.88 (0.80, 0.96) |
| Sanitation (p=0.873) | | | |
| Improved | 8375 | 6914 | 0.90 (0.85, 0.95) |
| Unimproved | 6315 | 6319 | 0.91 (0.80, 1.04) |
| Supplement duration (p=0.519) | | | |
| 12 mo or less | 11483 | 8668 | 0.88 (0.81, 0.97) |
| > 12 mo | 5733 | 10903 | 0.85 (0.78, 0.91) |
| Frequency of contact (p=0.516) | | | |
| Weekly | 7304 | 6833 | 0.84 (0.75, 0.94) |
| Monthly | 9912 | 12738 | 0.89 (0.82, 0.96) |
| Average SQ−LNS compliance (p=0.272) | | | |
| High | 9307 | 11585 | 0.82 (0.76, 0.90) |
| Low | 6207 | 6301 | 0.88 (0.81, 0.97) |

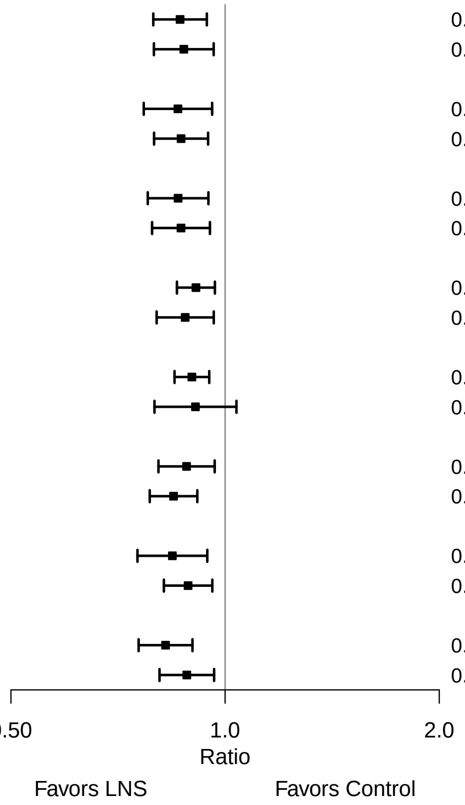

##### Supplemental figure 5C: Underweight prevalence ratio

#### 5C2: Stratified by individual-level maternal and child characteristics

| Effect modifier (p−for−interaction) | LNS N | Control N | PR (95% CI) |
|---|---|---|---|
| Maternal height (p=0.546) | | | |
| At least 150.1 cm | 9992 | 12808 | 0.82 (0.77, 0.88) |
| Less than 150.1 cm | 2242 | 2905 | 0.89 (0.82, 0.96) |
| Maternal BMI (p=0.545) | | | |
| At least 20 kg/m² | 7782 | 10117 | 0.83 (0.76, 0.90) |
| Less than 20 kg/m² | 3854 | 5024 | 0.85 (0.80, 0.91) |
| Maternal age (p=0.245) | | | |
| At least 25 y | 8357 | 9838 | 0.85 (0.80, 0.91) |
| Less than 25 y | 8335 | 9208 | 0.89 (0.85, 0.94) |
| Maternal education (p=0.399) | | | |
| Primary or greater | 7121 | 9070 | 0.85 (0.79, 0.90) |
| Incomplete or no formal | 6029 | 7977 | 0.91 (0.85, 0.97) |
| Maternal depressive symptoms (p=0.490) | | | |
| At least 75th | 2454 | 3925 | 0.90 (0.81, 1.01) |
| Less than 75th | 6364 | 10243 | 0.87 (0.81, 0.93) |
| Child sex (p=0.617) | | | |
| Male | 8523 | 9577 | 0.87 (0.82, 0.92) |
| Female | 8447 | 9748 | 0.86 (0.81, 0.92) |
| Child birth order (p=0.001) | | | |
| Later born | 10986 | 13686 | 0.83 (0.79, 0.88) |
| Firstborn | 5779 | 5491 | 0.94 (0.88, 1.00) |
| Child baseline anthropometric status (p=0.493) | | | |
| WAZ at least −1 Z | 4114 | 2898 | 0.83 (0.65, 1.05) |
| WAZ less than −1 Z | 3585 | 2144 | 0.92 (0.87, 0.96) |

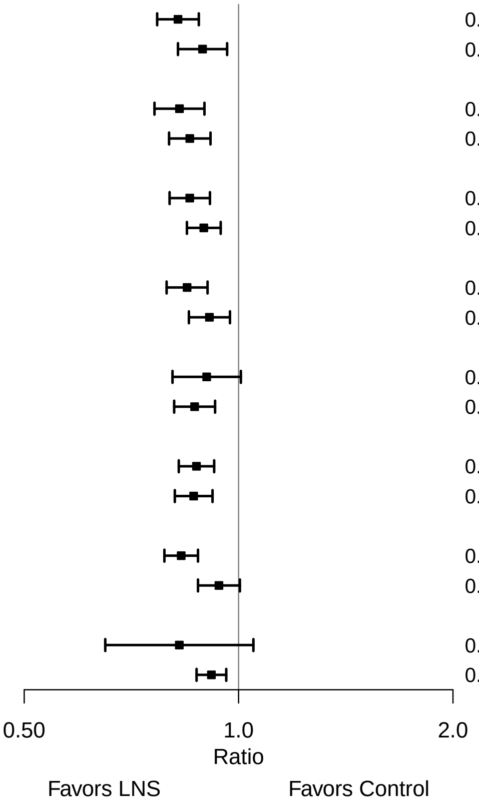

#### Supplemental figure 5C: Underweight prevalence ratio

### 5C3: Stratified by individual-level household characteristics

| **Effect modifier (p−for−interaction)** | **LNS N** | **Control N** | **PR (95% CI)** |
|---|---|---|---|
| Socio−economic status (p=0.877) | | | |
| At least median | 8704 | 9937 | 0.86 (0.81, 0.92) |
| Less than median | 7946 | 9160 | 0.87 (0.82, 0.92) |
| Food security (p=0.661) | | | |
| Mild to secure | 10207 | 12969 | 0.85 (0.81, 0.90) |
| Moderate to severe | 4692 | 4369 | 0.85 (0.79, 0.92) |
| Source Water quality (p=0.762) | | | |
| Improved | 3949 | 5191 | 0.87 (0.79, 0.95) |
| Unimproved | 4360 | 4017 | 0.85 (0.78, 0.93) |
| Sanitation (p=0.840) | | | |
| Improved | 5298 | 3530 | 0.91 (0.85, 0.98) |
| Unimproved | 2657 | 2077 | 0.92 (0.84, 1.01) |
| Home environment (p=0.416) | | | |
| At least median | 4790 | 6967 | 0.88 (0.81, 0.95) |
| Less than median | 3068 | 5535 | 0.92 (0.84, 1.00) |
| Season at time of assessment (p=0.102) | | | |
| Rainy | 5651 | 8113 | 0.88 (0.83, 0.94) |
| Dry | 7835 | 7737 | 0.81 (0.76, 0.87) |

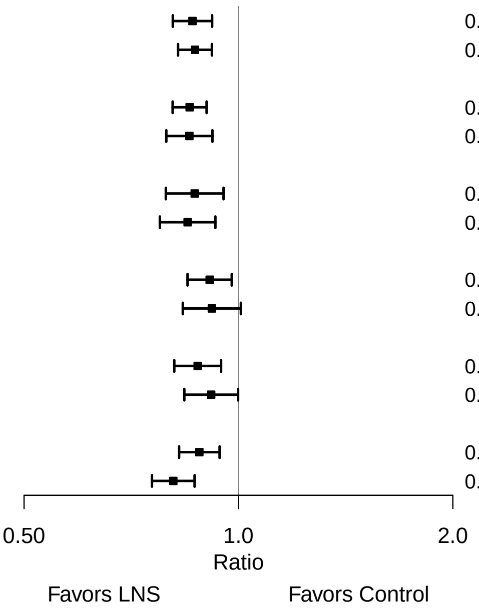

### Supplemental figure 5D: Small head size prevalence ratio

#### 5D1: Stratified by study-level characteristics

| **Effect modifier (p−diff)** | **LNS N** | **Control N** | **PR (95% CI)** |
|---|---|---|---|
| Geographic region (p=0.271) | | | |
| AFR | 6972 | 9422 | 0.87 (0.79, 0.95) |
| SEAR | 5711 | 5545 | 0.93 (0.86, 1.01) |
| Stunting burden (p=0.780) | | | |
| Less than 35% | 2126 | 6408 | 0.88 (0.72, 1.07) |
| More than 35% | 10557 | 8559 | 0.91 (0.86, 0.96) |
| Malaria prevalence (p=0.351) | | | |
| At least 10% | 3303 | 2134 | 0.87 (0.78, 0.96) |
| Less than 10% | 9380 | 12833 | 0.92 (0.87, 0.97) |
| Source water quality (p=0.958) | | | |
| Improved | 6476 | 4673 | 0.91 (0.85, 0.98) |
| Unimproved | 3676 | 3949 | 0.90 (0.80, 1.01) |
| Sanitation (p=0.549) | | | |
| Improved | 5564 | 3991 | 0.92 (0.84, 1.01) |
| Unimproved | 4588 | 4631 | 0.88 (0.80, 0.98) |
| Supplement duration (p=0.749) | | | |
| 12 mo or less | 8396 | 5513 | 0.92 (0.85, 0.99) |
| > 12 mo | 4287 | 9454 | 0.90 (0.84, 0.96) |
| Frequency of contact (p=0.591) | | | |
| Weekly | 7351 | 6864 | 0.92 (0.86, 0.98) |
| Monthly | 5332 | 8103 | 0.89 (0.82, 0.96) |
| Average SQ−LNS compliance (p=0.264) | | | |
| High | 9216 | 11481 | 0.92 (0.87, 0.97) |
| Low | 3467 | 3486 | 0.84 (0.73, 0.97) |

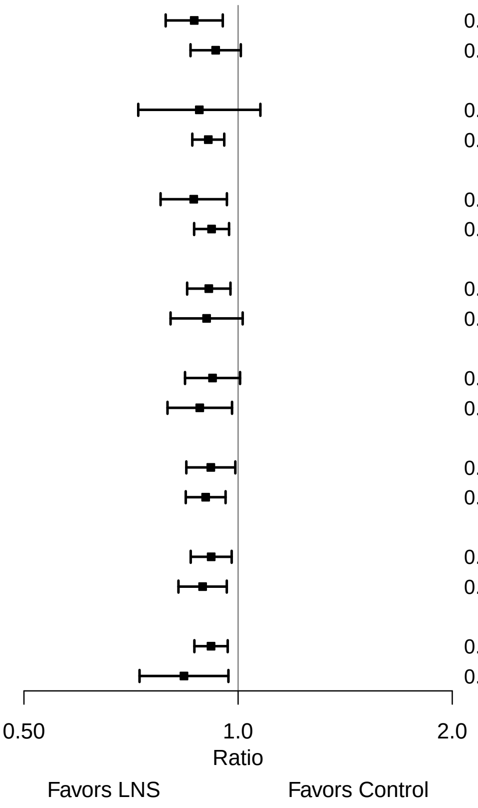

##### Supplemental figure 5D: Small head size prevalence ratio

#### 5D2: Stratified by individual-level maternal and child characteristics

| Effect modifier (p−for−interaction) | LNS N | Control N | PR (95% CI) |
|---|---|---|---|
| Maternal height (p=0.420) | | | |
| At least 150.1 cm | 7041 | 9582 | 0.89 (0.83, 0.96) |
| Less than 150.1 cm | 1556 | 2230 | 0.87 (0.80, 0.94) |
| Maternal BMI (p=0.607) | | | |
| At least 20 kg/m² | 5865 | 8445 | 0.89 (0.83, 0.96) |
| Less than 20 kg/m² | 3066 | 4285 | 0.87 (0.81, 0.94) |
| Maternal age (p=0.907) | | | |
| At least 25 y | 5963 | 7331 | 0.91 (0.84, 0.99) |
| Less than 25 y | 6345 | 7261 | 0.92 (0.86, 0.98) |
| Maternal education (p=0.091) | | | |
| Primary or greater | 5132 | 6916 | 0.88 (0.82, 0.95) |
| Incomplete or no formal | 4996 | 5382 | 0.94 (0.87, 1.02) |
| Maternal depressive symptoms (p=0.095) | | | |
| At least 75th | 1827 | 3333 | 0.84 (0.75, 0.95) |
| Less than 75th | 4816 | 8588 | 0.94 (0.87, 1.01) |
| Child sex (p=0.020) | | | |
| Male | 6321 | 7278 | 0.96 (0.90, 1.03) |
| Female | 6264 | 7593 | 0.85 (0.79, 0.92) |
| Child birth order (p=0.696) | | | |
| Later born | 7141 | 9861 | 0.89 (0.83, 0.95) |
| Firstborn | 4925 | 4553 | 0.93 (0.86, 1.01) |
| Child baseline anthropometric status (p=0.152) | | | |
| HCZ at least −1 Z | 4324 | 2696 | 0.73 (0.55, 0.97) |
| HCZ less than −1 Z | 4720 | 2315 | 0.92 (0.87, 0.97) |

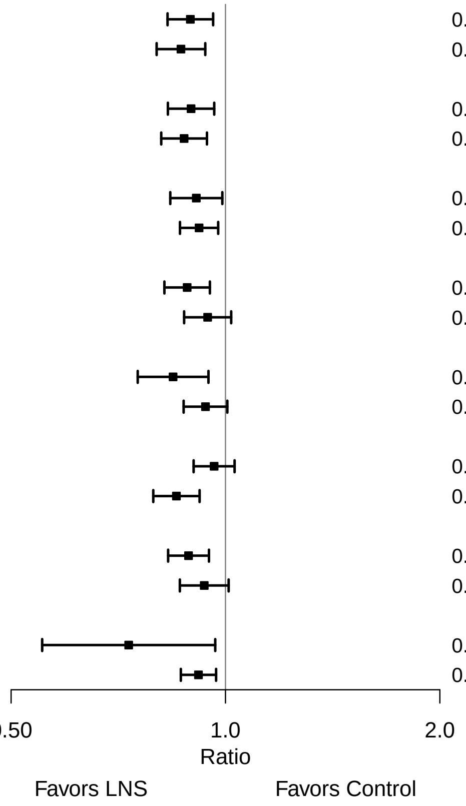

**Supplemental figure 5D: Small head size prevalence ratio**

#### 5D3: Stratified by individual-level household characteristics

| Effect modifier (p−for−interaction) | LNS N | Control N | PR (95% CI) |
|---|---|---|---|
| Socio−economic status (p=0.462) | | | |
| At least median | 6582 | 7678 | 0.89 (0.83, 0.96) |
| Less than median | 5745 | 7014 | 0.93 (0.88, 1.00) |
| Food security (p=0.542) | | | |
| Mild to secure | 8420 | 11192 | 0.90 (0.85, 0.95) |
| Moderate to severe | 3834 | 3435 | 0.94 (0.87, 1.02) |
| Source Water quality (p=0.505) | | | |
| Improved | 2571 | 3382 | 0.88 (0.77, 1.00) |
| Unimproved | 2027 | 1648 | 0.86 (0.77, 0.96) |
| Sanitation (p=0.109) | | | |
| Improved | 4256 | 2821 | 0.90 (0.83, 0.97) |
| Unimproved | 3555 | 1775 | 0.96 (0.87, 1.05) |
| Home environment (p=0.826) | | | |
| At least median | 3870 | 6058 | 0.90 (0.84, 0.98) |
| Less than median | 2275 | 4766 | 0.90 (0.84, 0.97) |
| Season at time of assessment (p=0.434) | | | |
| Rainy | 4914 | 7295 | 0.93 (0.86, 1.00) |
| Dry | 5463 | 5464 | 0.90 (0.84, 0.96) |

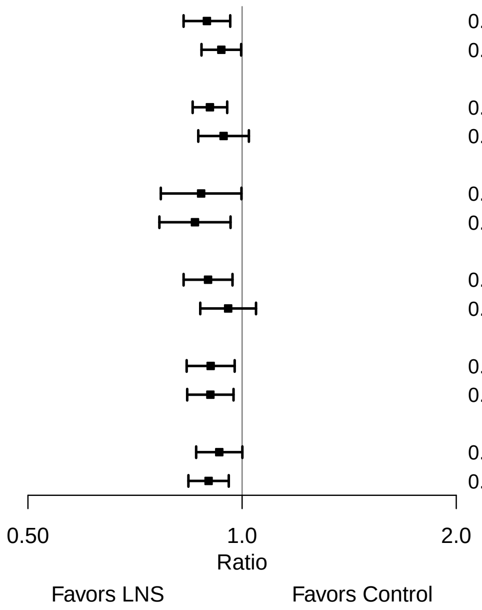

### Supplemental figure 6: Forest plots for effects of SQ-LNS on growth outcomes stratified by individual-level maternal and child effect modifiers

These figures are forest plots showing the individual-level effect modification of intervention effects. Each figure has the estimates of intervention effect stratified within study by individual-level effect modifier category. For continuous outcomes analyzed via mean differences, the effect estimate is the mean in the LNS group minus the mean in the control group. For dichotomous outcomes analyzed via prevalence ratios, the effect estimate is the prevalence in the LNS group divided by the prevalence in the control group. For dichotomous outcomes analyzed via prevalence differences, the effect estimate is the prevalence in the LNS group minus the prevalence in the control group. The labels on the far left correspond to trial level information. In the middle left and on the right the values indicate the study level effect estimate, confidence interval, and weighting for deriving the pooled estimates is shown by subgroup. LAZ, length-for-age z-score; WLZ, weight-for-length z-score; WAZ, weight-for-age z-score; MUACZ, mid-upper arm circumference z-score; HCZ, head circumference-for-age z-score.

### Supplemental figure 6A: Mean difference in LAZ

#### 6A1: Stratified by Maternal height

**P−for−interaction = 0.215**
**Difference in MDs = −0.04 (−0.11, 0.02)**

| | | At least 150.1 cm | | | | | | Less than 150.1 cm | | | | | |
|---|---|---|---|---|---|---|---|---|---|---|---|---|---|
| **Country** | **Trial** | **LNS N** | **Control N** | **Control Mean** | **MD (95% CI)** | **Fixed W** | **Random W** | **LNS N** | **Control N** | **Control Mean** | **MD (95% CI)** | **Fixed W** | **Random W** |
| Bangladesh | JiVitA−4 (34) | | | | | | | | | | | | |
| Bangladesh | RDNS (35) | 868 | 432 | −1.55 | 0.12 (0.01, 0.22) | 0.08 | 0.08 | 735 | 358 | −2.12 | 0.06 (−0.05, 0.16) | 0.26 | 0.26 |
| Bangladesh | WASH−B (36) | 616 | 1883 | −1.56 | 0.24 (0.15, 0.33) | 0.11 | 0.09 | 542 | 1545 | −2.13 | 0.19 (0.11, 0.28) | 0.44 | 0.40 |
| Burkina Faso | iLiNS−Zinc (37) | 1916 | 649 | −1.74 | 0.31 (0.22, 0.39) | 0.12 | 0.09 | 34 | 15 | −2.62 | 0.35 (0.04, 0.66) | 0.03 | 0.04 |
| Burkina Faso | PROMIS (38) | 835 | 884 | −1.46 | 0.15 (0.02, 0.29) | 0.05 | 0.07 | 25 | 25 | −2.21 | 0.02 (−0.76, 0.80) | 0.01 | 0.01 |
| Burkina Faso | PROMIS CS (38) | 419 | 426 | −1.19 | 0.03 (−0.15, 0.22) | 0.03 | 0.05 | 11 | 9 | −2.53 | 0.26 (−0.71, 1.23) | 0.00 | 0.00 |
| Ghana | GHANA (39) | | | | | | | | | | | | |
| Ghana | iLiNS−DYADG (40) | 326 | 645 | −0.86 | 0.20 (0.07, 0.33) | 0.05 | 0.07 | 16 | 36 | −1.51 | 0.10 (−0.46, 0.66) | 0.01 | 0.01 |
| Haiti | HAITI (41) | | | | | | | | | | | | |
| Kenya | WASH−B (42) | 1313 | 4686 | −1.53 | 0.15 (0.08, 0.22) | 0.19 | 0.10 | 57 | 190 | −2.16 | 0.02 (−0.29, 0.33) | 0.03 | 0.04 |
| Madagascar | MAHAY (43) | 1004 | 1037 | −2.11 | 0.09 (−0.06, 0.24) | 0.04 | 0.06 | 598 | 565 | −2.54 | 0.00 (−0.20, 0.19) | 0.08 | 0.09 |
| Malawi | iLiNS−DYADM (44) | 192 | 381 | −1.53 | −0.09 (−0.27, 0.09) | 0.03 | 0.05 | 28 | 60 | −2.21 | −0.11 (−0.59, 0.37) | 0.01 | 0.02 |
| Malawi | iLiNS−DOSE (45) | 580 | 198 | −1.80 | 0.03 (−0.09, 0.15) | 0.07 | 0.08 | 115 | 42 | −2.55 | 0.22 (0.01, 0.43) | 0.07 | 0.08 |
| Mali | PROMIS (46) | 481 | 496 | −1.73 | 0.19 (0.04, 0.34) | 0.04 | 0.06 | 19 | 9 | −1.81 | −0.31 (−0.86, 0.24) | 0.01 | 0.01 |
| Mali | PROMIS CS (46) | 914 | 936 | −1.55 | 0.24 (0.06, 0.41) | 0.03 | 0.05 | 33 | 27 | −2.39 | 0.12 (−0.36, 0.61) | 0.01 | 0.01 |
| Zimbabwe | SHINE (HIV−) (47) | 1755 | 1697 | −1.58 | 0.16 (0.09, 0.24) | 0.15 | 0.10 | 76 | 62 | −2.42 | 0.23 (−0.10, 0.56) | 0.03 | 0.03 |
| Zimbabwe | SHINE (HIV+) (48) | 314 | 312 | −1.96 | 0.23 (0.05, 0.41) | 0.03 | 0.05 | 18 | 12 | −2.81 | 0.63 (−0.30, 1.55) | 0.00 | 0.00 |
| | | **11533** | **14662** | | **$I^2$ = 0.58, $Tau^2$ = 0.01** | | | **2307** | **2955** | | **$I^2$ = 0.06, $Tau^2$ = 0.00** | | |
| **Fixed** | | | | | **0.17 (0.14, 0.20)** | | | | | | **0.13 (0.08, 0.19)** | | |
| **Random** | | | | | **0.16 (0.10, 0.21)** | | | | | | **0.13 (0.07, 0.19)** | | |

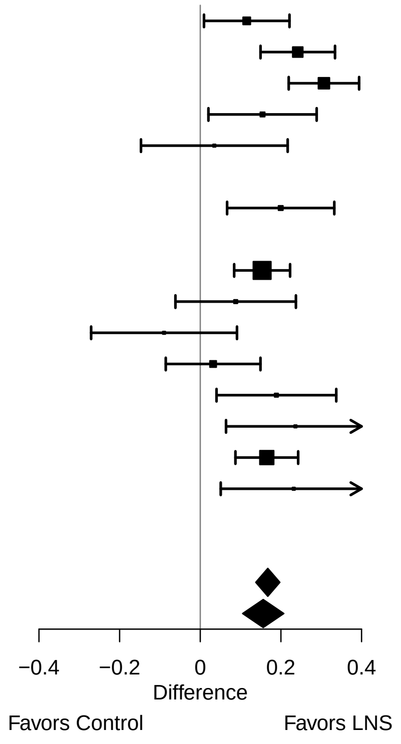

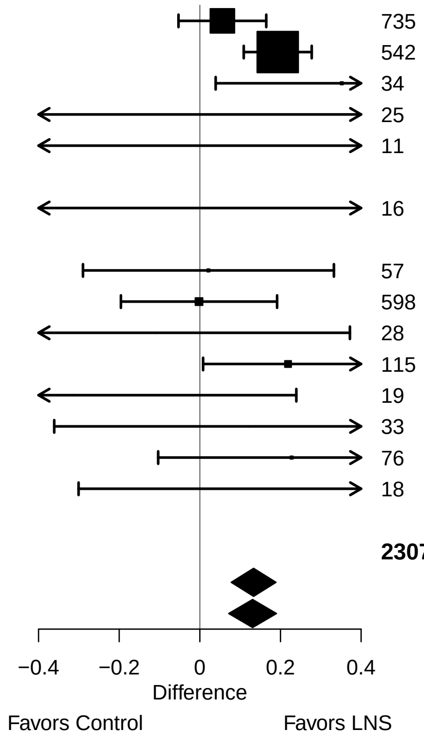

#### Supplemental figure 6A: Mean difference in LAZ

#### 6A2: Stratified by Maternal BMI

**P−for−interaction = 0.676**
**Difference in MDs = −0.01 (−0.06, 0.04)**

| | | At least 20 kg/m² | | | | | | Less than 20 kg/m² | | | | | |
|---|---|---|---|---|---|---|---|---|---|---|---|---|---|
| Country | Trial | LNS N | Control N | Control Mean | MD (95% CI) | Fixed W | Random W | LNS N | Control N | Control Mean | MD (95% CI) | Fixed W | Random W |
| Bangladesh | JiVitA−4 (34) | | | | | | | | | | | | |
| Bangladesh | RDNS (35) | 730 | 335 | −1.69 | 0.10 (−0.04, 0.23) | 0.07 | 0.08 | 873 | 455 | −1.90 | 0.06 (−0.03, 0.16) | 0.18 | 0.11 |
| Bangladesh | WASH−B (36) | 519 | 1576 | −1.69 | 0.21 (0.09, 0.33) | 0.08 | 0.09 | 639 | 1852 | −1.92 | 0.21 (0.13, 0.29) | 0.25 | 0.12 |
| Burkina Faso | iLiNS−Zinc (37) | 1214 | 390 | −1.65 | 0.30 (0.21, 0.40) | 0.13 | 0.11 | 736 | 274 | −1.91 | 0.32 (0.20, 0.43) | 0.14 | 0.11 |
| Burkina Faso | PROMIS (38) | 647 | 675 | −1.47 | 0.22 (0.10, 0.34) | 0.08 | 0.09 | 213 | 233 | −1.52 | −0.07 (−0.30, 0.16) | 0.03 | 0.06 |
| Burkina Faso | PROMIS CS (38) | 246 | 269 | −1.09 | 0.04 (−0.15, 0.23) | 0.03 | 0.05 | 184 | 166 | −1.42 | 0.06 (−0.19, 0.32) | 0.03 | 0.06 |
| Ghana | GHANA (39) | | | | | | | | | | | | |
| Ghana | iLiNS−DYADG (40) | 305 | 562 | −0.87 | 0.17 (0.03, 0.31) | 0.06 | 0.07 | 37 | 119 | −1.01 | 0.31 (−0.05, 0.68) | 0.01 | 0.04 |
| Haiti | HAITI (41) | | | | | | | | | | | | |
| Kenya | WASH−B (42) | 1098 | 3803 | −1.52 | 0.11 (0.03, 0.18) | 0.20 | 0.13 | 272 | 1073 | −1.68 | 0.29 (0.16, 0.42) | 0.10 | 0.10 |
| Madagascar | MAHAY (43) | | | | | | | | | | | | |
| Malawi | iLiNS−DYADM (44) | 131 | 263 | −1.61 | −0.01 (−0.23, 0.22) | 0.02 | 0.04 | 88 | 178 | −1.63 | −0.20 (−0.47, 0.08) | 0.02 | 0.05 |
| Malawi | iLiNS−DOSE (45) | 513 | 174 | −1.88 | 0.06 (−0.06, 0.18) | 0.08 | 0.09 | 180 | 65 | −2.09 | 0.08 (−0.12, 0.28) | 0.04 | 0.07 |
| Mali | PROMIS (46) | 380 | 370 | −1.68 | 0.20 (0.02, 0.38) | 0.04 | 0.05 | 120 | 135 | −1.85 | 0.09 (−0.06, 0.23) | 0.08 | 0.09 |
| Mali | PROMIS CS (46) | 702 | 684 | −1.50 | 0.20 (0.01, 0.38) | 0.03 | 0.05 | 245 | 279 | −1.75 | 0.28 (0.09, 0.47) | 0.05 | 0.08 |
| Zimbabwe | SHINE (HIV−) (47) | 1364 | 1305 | −1.62 | 0.20 (0.11, 0.28) | 0.16 | 0.12 | 240 | 249 | −1.67 | 0.03 (−0.16, 0.22) | 0.05 | 0.08 |
| Zimbabwe | SHINE (HIV+) (48) | 247 | 254 | −1.96 | 0.27 (0.06, 0.47) | 0.03 | 0.04 | 62 | 52 | −2.17 | 0.21 (−0.19, 0.61) | 0.01 | 0.03 |
| | | **8096** | **10660** | | **I² = 0.44, Tau² = 0.00** | | | **3889** | **5130** | | **I² = 0.64, Tau² = 0.01** | | |
| **Fixed** | | | | | **0.17 (0.14, 0.20)** | | | | | | **0.16 (0.12, 0.21)** | | |
| **Random** | | | | | **0.17 (0.12, 0.21)** | | | | | | **0.14 (0.06, 0.22)** | | |

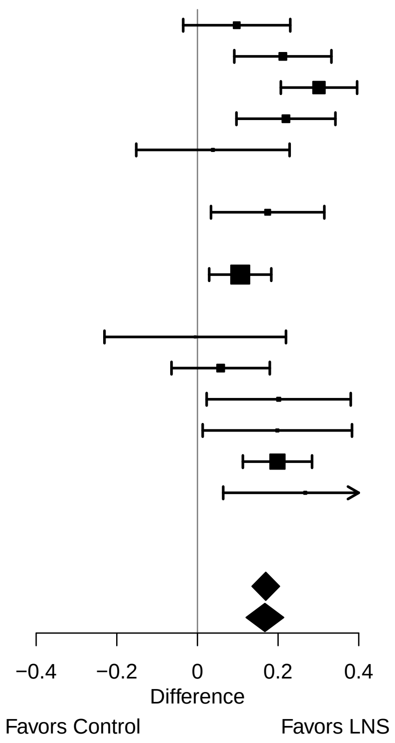

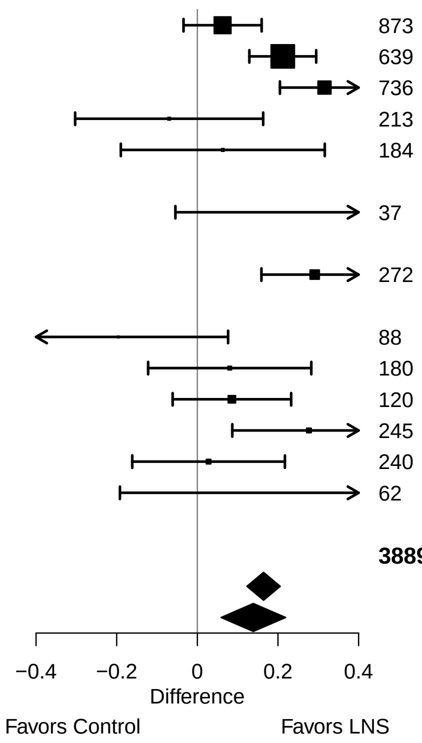

#### Supplemental figure 6A: Mean difference in LAZ

#### 6A3: Stratified by Maternal age

**P−for−interaction = 0.818**
**Difference in MDs = 0.00 (−0.05, 0.04)**

**At least 25 y**

| Country | Trial | LNS N | Control N | Control Mean | MD (95% CI) | Fixed W | Random W |
|---|---|---|---|---|---|---|---|
| Bangladesh | JiVitA−4 (34) | 1218 | 546 | −1.94 | 0.10 (0.03, 0.16) | 0.21 | 0.12 |
| Bangladesh | RDNS (35) | 453 | 221 | −1.80 | 0.19 (0.03, 0.35) | 0.04 | 0.05 |
| Bangladesh | WASH−B (36) | 492 | 1512 | −1.83 | 0.17 (0.09, 0.26) | 0.12 | 0.10 |
| Burkina Faso | iLiNS−Zinc (37) | 1131 | 399 | −1.72 | 0.33 (0.22, 0.45) | 0.07 | 0.08 |
| Burkina Faso | PROMIS (38) | 485 | 526 | −1.47 | 0.15 (−0.01, 0.32) | 0.04 | 0.05 |
| Burkina Faso | PROMIS CS (38) | 202 | 192 | −1.17 | 0.05 (−0.17, 0.27) | 0.02 | 0.03 |
| Ghana | GHANA (39) | 69 | 63 | −0.30 | 0.28 (−0.08, 0.65) | 0.01 | 0.01 |
| Ghana | iLiNS−DYADG (40) | 220 | 416 | −0.84 | 0.11 (−0.05, 0.27) | 0.04 | 0.05 |
| Haiti | HAITI (41) | 101 | 103 | −0.67 | 0.04 (−0.13, 0.21) | 0.03 | 0.05 |
| Kenya | WASH−B (42) | 844 | 2811 | −1.50 | 0.13 (0.05, 0.21) | 0.16 | 0.11 |
| Madagascar | MAHAY (43) | 896 | 949 | −2.26 | 0.03 (−0.13, 0.18) | 0.04 | 0.05 |
| Malawi | iLiNS−DYADM (44) | 106 | 226 | −1.65 | −0.04 (−0.30, 0.21) | 0.02 | 0.03 |
| Malawi | iLiNS−DOSE (45) | 368 | 134 | −1.88 | 0.00 (−0.14, 0.14) | 0.05 | 0.06 |
| Mali | PROMIS (46) | 346 | 359 | −1.72 | 0.17 (0.02, 0.33) | 0.04 | 0.05 |
| Mali | PROMIS CS (46) | 503 | 507 | −1.47 | 0.16 (−0.04, 0.36) | 0.02 | 0.04 |
| Zimbabwe | SHINE (HIV−) (47) | 875 | 800 | −1.55 | 0.20 (0.08, 0.31) | 0.07 | 0.08 |
| Zimbabwe | SHINE (HIV+) (48) | 244 | 231 | −1.97 | 0.24 (0.04, 0.45) | 0.02 | 0.04 |
| | | **8553** | **9995** | | **I² = 0.38, Tau² = 0.00** | | |
| **Fixed** | | | | | **0.14 (0.11, 0.17)** | | |
| **Random** | | | | | **0.14 (0.10, 0.18)** | | |

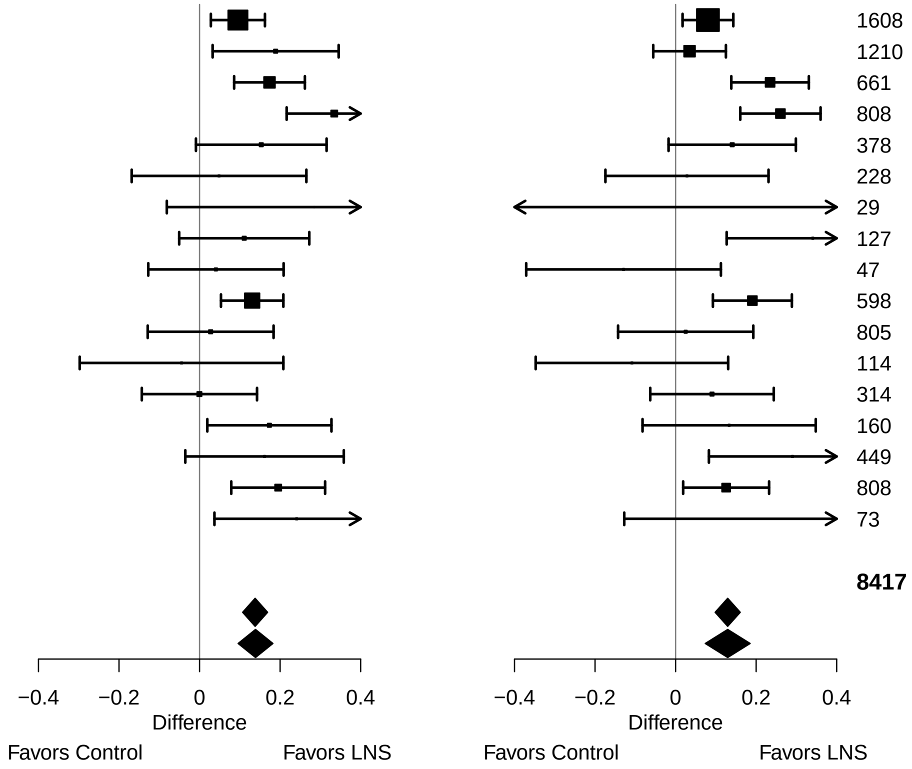

**Less than 25 y**

| LNS N | Control N | Control Mean | MD (95% CI) | Fixed W | Random W |
|---|---|---|---|---|---|
| 1608 | 693 | −1.88 | 0.08 (0.02, 0.14) | 0.24 | 0.10 |
| 1210 | 594 | −1.81 | 0.03 (−0.06, 0.12) | 0.12 | 0.09 |
| 661 | 1903 | −1.80 | 0.23 (0.14, 0.33) | 0.10 | 0.09 |
| 808 | 262 | −1.79 | 0.26 (0.16, 0.36) | 0.10 | 0.09 |
| 378 | 388 | −1.49 | 0.14 (−0.02, 0.30) | 0.04 | 0.06 |
| 228 | 247 | −1.27 | 0.03 (−0.17, 0.23) | 0.02 | 0.05 |
| 29 | 25 | −0.58 | 0.16 (−0.42, 0.75) | 0.00 | 0.01 |
| 127 | 276 | −0.97 | 0.34 (0.13, 0.55) | 0.02 | 0.04 |
| 47 | 45 | −0.88 | −0.13 (−0.37, 0.11) | 0.02 | 0.04 |
| 598 | 2294 | −1.65 | 0.19 (0.09, 0.29) | 0.10 | 0.09 |
| 805 | 733 | −2.22 | 0.03 (−0.14, 0.19) | 0.03 | 0.06 |
| 114 | 218 | −1.60 | −0.11 (−0.35, 0.13) | 0.02 | 0.04 |
| 314 | 100 | −1.99 | 0.09 (−0.06, 0.24) | 0.04 | 0.06 |
| 160 | 147 | −1.75 | 0.13 (−0.08, 0.35) | 0.02 | 0.04 |
| 449 | 462 | −1.68 | 0.29 (0.08, 0.50) | 0.02 | 0.05 |
| 808 | 797 | −1.66 | 0.13 (0.02, 0.23) | 0.09 | 0.08 |
| 73 | 78 | −2.07 | 0.24 (−0.13, 0.60) | 0.01 | 0.02 |
| **8417** | **9262** | | **I² = 0.56, Tau² = 0.01** | | |
| | | | **0.13 (0.10, 0.16)** | | |
| | | | **0.13 (0.07, 0.19)** | | |

#### Supplemental figure 6A: Mean difference in LAZ

#### 6A4: Stratified by Maternal education

**P−for−interaction = 0.699**
**Difference in MDs = 0.01 (−0.04, 0.06)**

**Primary or greater**

| Country | Trial | LNS N | Control N | Control Mean | MD (95% CI) | Fixed W | Random W |
|---|---|---|---|---|---|---|---|
| Bangladesh | JiVitA−4 (34) | 1830 | 756 | −1.78 | 0.08 (0.02, 0.14) | 0.27 | 0.15 |
| Bangladesh | RDNS (35) | 1245 | 589 | −1.77 | 0.10 (0.01, 0.19) | 0.11 | 0.11 |
| Bangladesh | WASH−B (36) | 813 | 2452 | −1.72 | 0.24 (0.16, 0.32) | 0.14 | 0.12 |
| Burkina Faso | iLiNS−Zinc (37) | 82 | 16 | −1.47 | 0.41 (0.11, 0.71) | 0.01 | 0.02 |
| Burkina Faso | PROMIS (38) | 70 | 54 | −1.39 | −0.08 (−0.38, 0.23) | 0.01 | 0.02 |
| Burkina Faso | PROMIS CS (38) | 34 | 39 | −1.26 | 0.00 (−0.49, 0.49) | 0.00 | 0.01 |
| Ghana | GHANA (39) | 91 | 75 | −0.36 | 0.28 (−0.05, 0.62) | 0.01 | 0.02 |
| Ghana | iLiNS−DYADG (40) | 267 | 548 | −0.89 | 0.18 (0.03, 0.32) | 0.04 | 0.07 |
| Haiti | HAITI (41) | 130 | 126 | −0.71 | −0.04 (−0.19, 0.11) | 0.04 | 0.07 |
| Kenya | WASH−B (42) | 700 | 2434 | −1.39 | 0.09 (0.00, 0.18) | 0.11 | 0.11 |
| Madagascar | MAHAY (43) | 351 | 431 | −2.17 | 0.03 (−0.17, 0.23) | 0.02 | 0.04 |
| Malawi | iLiNS−DYADM (44) | 35 | 69 | −1.37 | −0.10 (−0.51, 0.32) | 0.01 | 0.01 |
| Malawi | iLiNS−DOSE (45) | 162 | 56 | −1.73 | 0.12 (−0.08, 0.32) | 0.02 | 0.04 |
| Mali | PROMIS (46) | 41 | 37 | −1.25 | 0.10 (−0.29, 0.49) | 0.01 | 0.01 |
| Mali | PROMIS CS (46) | 107 | 96 | −1.42 | 0.29 (−0.04, 0.63) | 0.01 | 0.02 |
| Zimbabwe | SHINE (HIV−) (47) | 1717 | 1644 | −1.59 | 0.15 (0.07, 0.23) | 0.14 | 0.12 |
| Zimbabwe | SHINE (HIV+) (48) | 301 | 293 | −1.96 | 0.21 (0.03, 0.40) | 0.03 | 0.05 |
| | | **7976** | **9715** | | **I² = 0.40, Tau² = 0.00** | | |
| **Fixed** | | | | | **0.12 (0.09, 0.15)** | | |
| **Random** | | | | | **0.12 (0.08, 0.17)** | | |

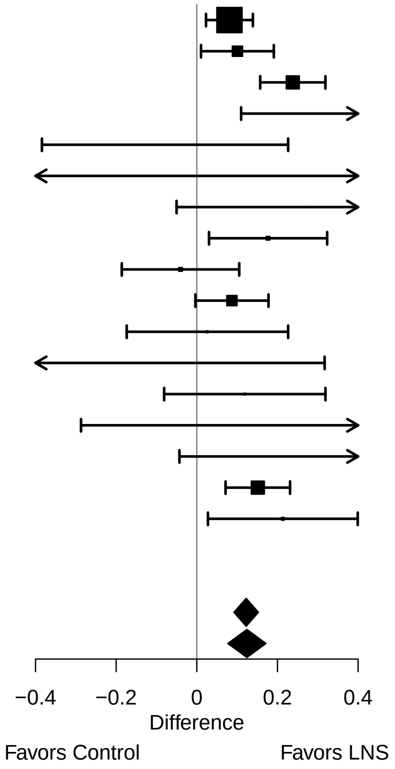

**Incomplete or no formal**

| Country | Trial | LNS N | Control N | Control Mean | MD (95% CI) | Fixed W | Random W |
|---|---|---|---|---|---|---|---|
| Bangladesh | JiVitA−4 (34) | 1006 | 485 | −2.10 | 0.09 (0.01, 0.16) | 0.20 | 0.11 |
| Bangladesh | RDNS (35) | 418 | 226 | −1.89 | −0.01 (−0.14, 0.13) | 0.06 | 0.07 |
| Bangladesh | WASH−B (36) | 345 | 979 | −2.06 | 0.16 (0.02, 0.29) | 0.06 | 0.08 |
| Burkina Faso | iLiNS−Zinc (37) | 1857 | 645 | −1.76 | 0.30 (0.22, 0.39) | 0.15 | 0.10 |
| Burkina Faso | PROMIS (38) | 793 | 860 | −1.49 | 0.17 (0.04, 0.30) | 0.06 | 0.08 |
| Burkina Faso | PROMIS CS (38) | 393 | 399 | −1.22 | 0.04 (−0.15, 0.22) | 0.03 | 0.05 |
| Ghana | GHANA (39) | 7 | 15 | −0.47 | −0.39 (−1.25, 0.46) | 0.00 | 0.00 |
| Ghana | iLiNS−DYADG (40) | 80 | 144 | −0.91 | 0.28 (0.00, 0.55) | 0.01 | 0.03 |
| Haiti | HAITI (41) | 19 | 23 | −0.88 | 0.13 (−0.26, 0.53) | 0.01 | 0.02 |
| Kenya | WASH−B (42) | 755 | 2699 | −1.73 | 0.21 (0.13, 0.29) | 0.17 | 0.10 |
| Madagascar | MAHAY (43) | 1351 | 1251 | −2.26 | 0.03 (−0.12, 0.18) | 0.05 | 0.07 |
| Malawi | iLiNS−DYADM (44) | 184 | 372 | −1.68 | −0.07 (−0.26, 0.12) | 0.03 | 0.05 |
| Malawi | iLiNS−DOSE (45) | 522 | 178 | −1.99 | 0.02 (−0.10, 0.15) | 0.07 | 0.08 |
| Mali | PROMIS (46) | 464 | 469 | −1.76 | 0.17 (0.03, 0.31) | 0.05 | 0.07 |
| Mali | PROMIS CS (46) | 844 | 873 | −1.58 | 0.21 (0.04, 0.39) | 0.04 | 0.06 |
| Zimbabwe | SHINE (HIV−) (47) | 69 | 61 | −1.69 | 0.09 (−0.32, 0.50) | 0.01 | 0.02 |
| Zimbabwe | SHINE (HIV+) (48) | 21 | 18 | −2.40 | 0.54 (−0.17, 1.25) | 0.00 | 0.01 |
| | | **9128** | **9697** | | **I² = 0.60, Tau² = 0.01** | | |
| **Fixed** | | | | | **0.14 (0.11, 0.18)** | | |
| **Random** | | | | | **0.13 (0.07, 0.18)** | | |

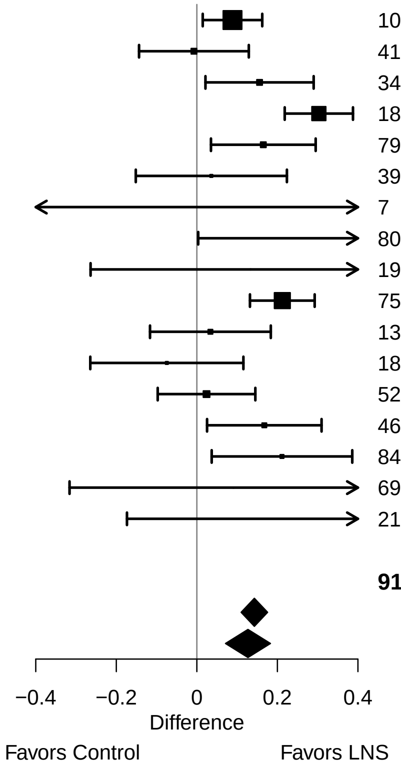

#### Supplemental figure 6A: Mean difference in LAZ

##### 6A5: Stratified by Maternal depressive symptoms

**P−for−interaction = 0.060**
**Difference in MDs = 0.06 (0.00, 0.13)**

| | | Less than 75th percentile | | | | | | At least 75th percentile | | | | | |
|---|---|---|---|---|---|---|---|---|---|---|---|---|---|
| **Country** | **Trial** | **LNS N** | **Control N** | **Control Mean** | **MD (95% CI)** | **Fixed W** | **Random W** | **LNS N** | **Control N** | **Control Mean** | **MD (95% CI)** | **Fixed W** | **Random W** |
| Bangladesh | JiVitA−4 (34) | | | | | | | | | | | | |
| Bangladesh | RDNS (35) | 1081 | 472 | −1.80 | 0.12 (0.02, 0.23) | 0.13 | 0.13 | 520 | 288 | −1.81 | 0.01 (−0.14, 0.15) | 0.14 | 0.13 |
| Bangladesh | WASH−B (36) | 897 | 2462 | −1.79 | 0.23 (0.14, 0.31) | 0.21 | 0.16 | 237 | 878 | −1.88 | 0.15 (0.02, 0.28) | 0.18 | 0.14 |
| Burkina Faso | iLiNS−Zinc (37) | | | | | | | | | | | | |
| Burkina Faso | PROMIS (38) | 508 | 573 | −1.53 | 0.24 (0.08, 0.40) | 0.05 | 0.07 | 223 | 215 | −1.40 | −0.06 (−0.24, 0.12) | 0.10 | 0.11 |
| Burkina Faso | PROMIS CS (38) | | | | | | | | | | | | |
| Ghana | GHANA (39) | | | | | | | | | | | | |
| Ghana | iLiNS−DYADG (40) | 252 | 459 | −0.90 | 0.23 (0.08, 0.38) | 0.06 | 0.08 | 83 | 209 | −0.84 | 0.12 (−0.13, 0.37) | 0.05 | 0.07 |
| Haiti | HAITI (41) | | | | | | | | | | | | |
| Kenya | WASH−B (42) | 996 | 3496 | −1.53 | 0.14 (0.07, 0.22) | 0.25 | 0.18 | 339 | 1235 | −1.63 | 0.12 (−0.02, 0.25) | 0.17 | 0.14 |
| Madagascar | MAHAY (43) | 673 | 721 | −2.28 | 0.04 (−0.15, 0.22) | 0.04 | 0.06 | 284 | 252 | −2.27 | −0.14 (−0.36, 0.08) | 0.06 | 0.09 |
| Malawi | iLiNS−DYADM (44) | 157 | 303 | −1.55 | −0.07 (−0.28, 0.14) | 0.03 | 0.05 | 47 | 113 | −1.77 | −0.15 (−0.49, 0.20) | 0.03 | 0.05 |
| Malawi | iLiNS−DOSE (45) | | | | | | | | | | | | |
| Mali | PROMIS (46) | 379 | 373 | −1.68 | 0.17 (0.01, 0.32) | 0.06 | 0.08 | 123 | 131 | −1.86 | 0.17 (−0.04, 0.37) | 0.07 | 0.09 |
| Mali | PROMIS CS (46) | | | | | | | | | | | | |
| Zimbabwe | SHINE (HIV−) (47) | 1198 | 1145 | −1.56 | 0.12 (0.02, 0.22) | 0.14 | 0.14 | 513 | 489 | −1.73 | 0.27 (0.14, 0.40) | 0.18 | 0.14 |
| Zimbabwe | SHINE (HIV+) (48) | 234 | 215 | −1.96 | 0.25 (0.05, 0.46) | 0.03 | 0.05 | 89 | 100 | −2.13 | 0.21 (−0.14, 0.56) | 0.02 | 0.05 |
| | | **6375** | **10219** | | **I² = 0.30, Tau² = 0.00** | | | **2458** | **3910** | | **I² = 0.53, Tau² = 0.01** | | |
| **Fixed** | | | | | **0.16 (0.12, 0.20)** | | | | | | **0.10 (0.05, 0.16)** | | |
| **Random** | | | | | **0.15 (0.10, 0.21)** | | | | | | **0.08 (0.00, 0.17)** | | |

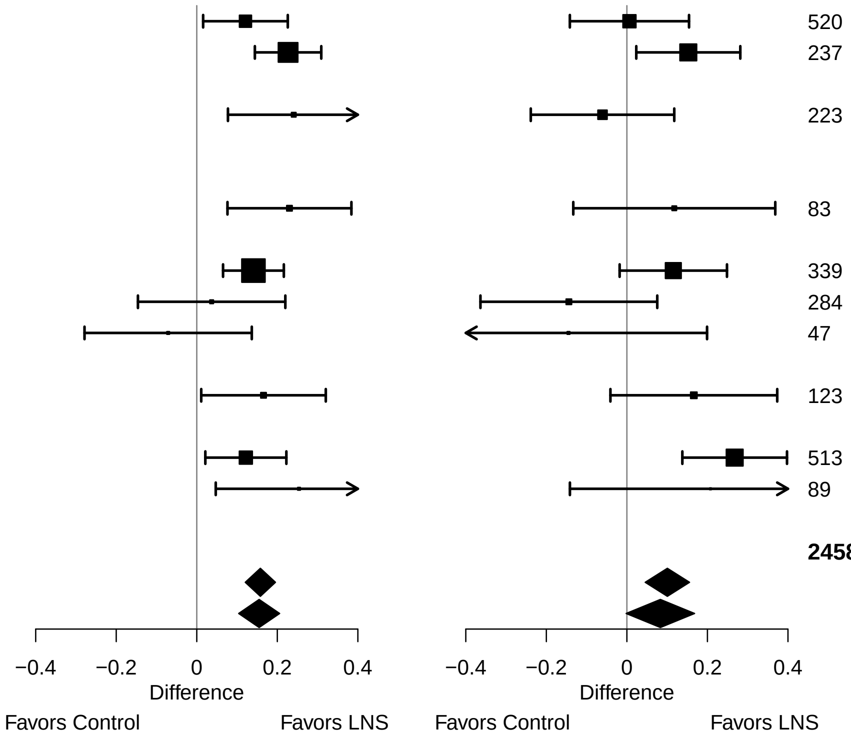

#### Supplemental figure 6A: Mean difference in LAZ

### 6A6: Stratified by Child sex

**P−for−interaction = 0.170**
**Difference in MDs = 0.03 (−0.01, 0.07)**

| Country | Trial | Male LNS N | Male Control N | Male Control Mean | Male MD (95% CI) | Male Fixed W | Male Random W | Female LNS N | Female Control N | Female Control Mean | Female MD (95% CI) | Female Fixed W | Female Random W |
|---|---|---|---|---|---|---|---|---|---|---|---|---|---|
| Bangladesh | JiVitA−4 (34) | 1448 | 600 | −1.91 | 0.07 (0.00, 0.13) | 0.24 | 0.11 | 1390 | 644 | −1.90 | 0.11 (0.04, 0.17) | 0.21 | 0.10 |
| Bangladesh | RDNS (35) | 833 | 411 | −1.82 | 0.00 (−0.12, 0.13) | 0.06 | 0.07 | 830 | 404 | −1.80 | 0.15 (0.04, 0.26) | 0.08 | 0.08 |
| Bangladesh | WASH−B (36) | 565 | 1714 | −1.83 | 0.21 (0.12, 0.30) | 0.13 | 0.10 | 593 | 1717 | −1.80 | 0.21 (0.11, 0.31) | 0.09 | 0.08 |
| Burkina Faso | iLiNS−Zinc (37) | 990 | 336 | −1.89 | 0.31 (0.19, 0.42) | 0.07 | 0.08 | 962 | 328 | −1.63 | 0.31 (0.21, 0.40) | 0.10 | 0.09 |
| Burkina Faso | PROMIS (38) | 438 | 483 | −1.60 | 0.11 (−0.05, 0.28) | 0.04 | 0.05 | 425 | 431 | −1.35 | 0.17 (0.02, 0.32) | 0.04 | 0.06 |
| Burkina Faso | PROMIS CS (38) | 234 | 222 | −1.37 | 0.05 (−0.18, 0.27) | 0.02 | 0.04 | 196 | 217 | −1.07 | 0.06 (−0.13, 0.24) | 0.03 | 0.04 |
| Ghana | GHANA (39) | 60 | 41 | −0.44 | 0.27 (−0.15, 0.69) | 0.01 | 0.01 | 38 | 55 | −0.39 | 0.32 (−0.14, 0.78) | 0.00 | 0.01 |
| Ghana | iLiNS−DYADG (40) | 175 | 323 | −0.92 | 0.19 (−0.02, 0.39) | 0.02 | 0.04 | 172 | 369 | −0.87 | 0.22 (0.05, 0.38) | 0.03 | 0.05 |
| Haiti | HAITI (41) | 70 | 59 | −0.87 | 0.06 (−0.16, 0.27) | 0.02 | 0.04 | 79 | 90 | −0.66 | −0.02 (−0.19, 0.16) | 0.03 | 0.05 |
| Kenya | WASH−B (42) | 734 | 2440 | −1.70 | 0.14 (0.05, 0.23) | 0.12 | 0.10 | 723 | 2697 | −1.45 | 0.18 (0.09, 0.27) | 0.12 | 0.09 |
| Madagascar | MAHAY (43) | 826 | 832 | −2.40 | 0.01 (−0.14, 0.17) | 0.04 | 0.06 | 876 | 850 | −2.08 | 0.03 (−0.13, 0.20) | 0.04 | 0.05 |
| Malawi | iLiNS−DYADM (44) | 107 | 207 | −1.73 | −0.07 (−0.33, 0.19) | 0.01 | 0.03 | 113 | 237 | −1.54 | −0.08 (−0.31, 0.16) | 0.02 | 0.03 |
| Malawi | iLiNS−DOSE (45) | 352 | 124 | −2.14 | 0.01 (−0.14, 0.16) | 0.04 | 0.06 | 344 | 117 | −1.73 | 0.11 (−0.03, 0.26) | 0.05 | 0.06 |
| Mali | PROMIS (46) | 266 | 263 | −1.80 | 0.07 (−0.09, 0.24) | 0.04 | 0.05 | 240 | 243 | −1.64 | 0.26 (0.09, 0.44) | 0.03 | 0.05 |
| Mali | PROMIS CS (46) | 477 | 525 | −1.63 | 0.21 (0.00, 0.42) | 0.02 | 0.04 | 475 | 444 | −1.49 | 0.23 (0.04, 0.42) | 0.02 | 0.04 |
| Zimbabwe | SHINE (HIV−) (47) | 932 | 908 | −1.80 | 0.24 (0.14, 0.34) | 0.10 | 0.09 | 948 | 886 | −1.40 | 0.06 (−0.04, 0.17) | 0.08 | 0.08 |
| Zimbabwe | SHINE (HIV+) (48) | 162 | 169 | −2.14 | 0.19 (−0.07, 0.44) | 0.02 | 0.03 | 175 | 161 | −1.86 | 0.30 (0.10, 0.50) | 0.02 | 0.04 |
| | | **8669** | **9657** | | **I² = 0.53, Tau² = 0.00** | | | **8579** | **9890** | | **I² = 0.51, Tau² = 0.00** | | |
| **Fixed** | | | | | **0.13 (0.10, 0.16)** | | | | | | **0.15 (0.12, 0.18)** | | |
| **Random** | | | | | **0.13 (0.08, 0.17)** | | | | | | **0.15 (0.10, 0.20)** | | |

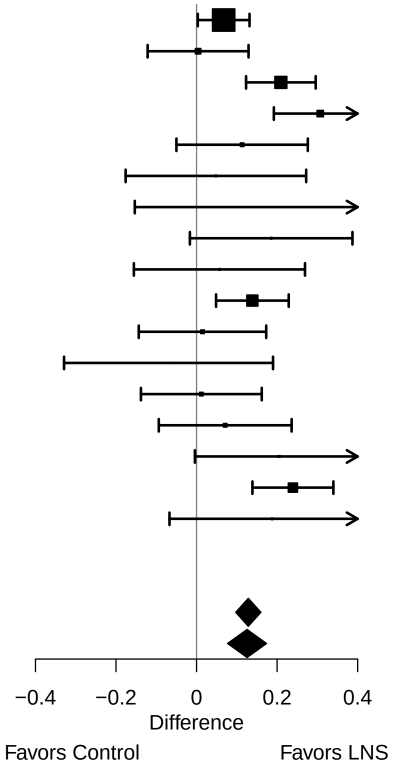

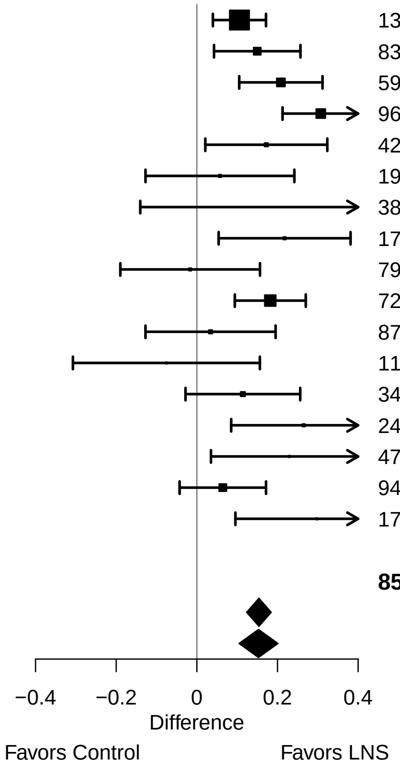

#### Supplemental figure 6A: Mean difference in LAZ

##### 6A7: Stratified by Child birth order

**P−for−interaction = 0.134**
**Difference in MDs = −0.03 (−0.08, 0.01)**

| Country | Trial | Later born LNS N | Later born Control N | Later born Control Mean | Later born MD (95% CI) | Later born Fixed W | Later born Random W | Firstborn LNS N | Firstborn Control N | Firstborn Control Mean | Firstborn MD (95% CI) | Firstborn Fixed W | Firstborn Random W |
|---|---|---|---|---|---|---|---|---|---|---|---|---|---|
| Bangladesh | JiVitA−4 (34) | 611 | 255 | −2.03 | 0.14 (0.04, 0.24) | 0.08 | 0.08 | 2223 | 982 | −1.87 | 0.07 (0.02, 0.13) | 0.44 | 0.10 |
| Bangladesh | RDNS (35) | 974 | 508 | −1.83 | 0.11 (0.01, 0.22) | 0.07 | 0.07 | 688 | 307 | −1.78 | 0.02 (−0.13, 0.17) | 0.06 | 0.08 |
| Bangladesh | WASH−B (36) | 736 | 2290 | −1.85 | 0.20 (0.12, 0.28) | 0.14 | 0.09 | 422 | 1141 | −1.73 | 0.21 (0.09, 0.34) | 0.09 | 0.08 |
| Burkina Faso | iLiNS−Zinc (37) | 1520 | 519 | −1.72 | 0.30 (0.22, 0.39) | 0.11 | 0.08 | 432 | 145 | −1.88 | 0.32 (0.20, 0.44) | 0.09 | 0.08 |
| Burkina Faso | PROMIS (38) | 715 | 754 | −1.46 | 0.16 (0.02, 0.31) | 0.04 | 0.05 | 148 | 158 | −1.59 | 0.06 (−0.18, 0.30) | 0.02 | 0.06 |
| Burkina Faso | PROMIS CS (38) | 366 | 359 | −1.20 | 0.03 (−0.14, 0.20) | 0.03 | 0.05 | 63 | 80 | −1.33 | 0.07 (−0.41, 0.55) | 0.01 | 0.02 |
| Ghana | GHANA (39) | 60 | 54 | −0.39 | 0.40 (0.01, 0.80) | 0.01 | 0.01 | 38 | 38 | −0.36 | −0.01 (−0.49, 0.47) | 0.01 | 0.02 |
| Ghana | iLiNS−DYADG (40) | 237 | 458 | −0.90 | 0.12 (−0.03, 0.28) | 0.03 | 0.05 | 110 | 234 | −0.87 | 0.36 (0.14, 0.58) | 0.03 | 0.06 |
| Haiti | HAITI (41) | 88 | 99 | −0.83 | 0.02 (−0.15, 0.19) | 0.03 | 0.05 | 61 | 50 | −0.56 | −0.10 (−0.33, 0.13) | 0.02 | 0.06 |
| Kenya | WASH−B (42) | 1134 | 4045 | −1.57 | 0.14 (0.07, 0.20) | 0.19 | 0.10 | 321 | 1087 | −1.57 | 0.22 (0.06, 0.37) | 0.05 | 0.08 |
| Madagascar | MAHAY (43) | 1272 | 1217 | −2.28 | 0.09 (−0.06, 0.24) | 0.04 | 0.05 | 430 | 465 | −2.14 | −0.14 (−0.34, 0.06) | 0.03 | 0.06 |
| Malawi | iLiNS−DYADM (44) | 173 | 353 | −1.64 | −0.09 (−0.28, 0.11) | 0.02 | 0.04 | 47 | 89 | −1.57 | −0.05 (−0.44, 0.34) | 0.01 | 0.03 |
| Malawi | iLiNS−DOSE (45) | 458 | 165 | −1.92 | 0.00 (−0.12, 0.13) | 0.05 | 0.06 | 149 | 45 | −1.95 | 0.12 (−0.10, 0.34) | 0.03 | 0.06 |
| Mali | PROMIS (46) | 442 | 445 | −1.74 | 0.21 (0.06, 0.37) | 0.03 | 0.05 | 63 | 59 | −1.61 | −0.20 (−0.47, 0.07) | 0.02 | 0.05 |
| Mali | PROMIS CS (46) | 790 | 831 | −1.54 | 0.23 (0.06, 0.40) | 0.03 | 0.04 | 145 | 126 | −1.74 | 0.27 (−0.02, 0.57) | 0.01 | 0.04 |
| Zimbabwe | SHINE (HIV−) (47) | 1302 | 1223 | −1.59 | 0.18 (0.09, 0.28) | 0.09 | 0.08 | 491 | 487 | −1.64 | 0.10 (−0.02, 0.23) | 0.08 | 0.08 |
| Zimbabwe | SHINE (HIV+) (48) | 280 | 247 | −1.89 | 0.15 (−0.04, 0.33) | 0.02 | 0.04 | 54 | 78 | −2.29 | 0.56 (0.12, 0.99) | 0.01 | 0.03 |
| | | **11158** | **13822** | | **I² = 0.53, Tau² = 0.00** | | | **5885** | **5571** | | **I² = 0.65, Tau² = 0.02** | | |
| **Fixed** | | | | | **0.15 (0.13, 0.18)** | | | | | | **0.11 (0.08, 0.15)** | | |
| **Random** | | | | | **0.14 (0.10, 0.19)** | | | | | | **0.11 (0.03, 0.20)** | | |

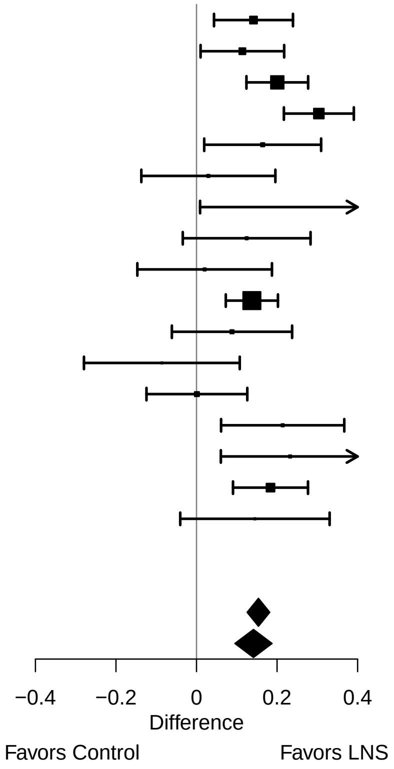

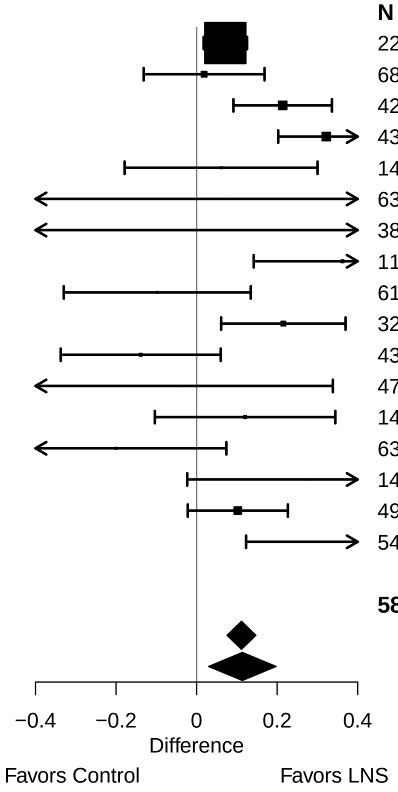

#### Supplemental figure 6A: Mean difference in LAZ

##### 6A8: Stratified by Child baseline anthropometric status

**P−for−interaction = 0.932**
**Difference in MDs = 0.00 (−0.06, 0.05)**

| | | LAZ at least −1 Z | | | | | | LAZ less than −1 Z | | | | | |
|---|---|---|---|---|---|---|---|---|---|---|---|---|---|
| **Country** | **Trial** | **LNS N** | **Control N** | **Control Mean** | **MD (95% CI)** | **Fixed W** | **Random W** | **LNS N** | **Control N** | **Control Mean** | **MD (95% CI)** | **Fixed W** | **Random W** |
| Bangladesh | JiVitA−4 (34) | 1055 | 454 | −1.11 | 0.10 (0.01, 0.19) | 0.21 | 0.20 | 1783 | 790 | −2.36 | 0.07 (−0.01, 0.15) | 0.26 | 0.16 |
| Bangladesh | RDNS (35) | 630 | 306 | −1.08 | 0.09 (−0.01, 0.19) | 0.18 | 0.18 | 971 | 453 | −2.30 | 0.10 (0.00, 0.20) | 0.18 | 0.14 |
| Bangladesh | WASH−B (36) | | | | | | | | | | | | |
| Burkina Faso | iLiNS−Zinc (37) | 839 | 291 | −0.85 | 0.21 (0.04, 0.38) | 0.06 | 0.06 | 1113 | 373 | −2.47 | 0.38 (0.23, 0.52) | 0.08 | 0.10 |
| Burkina Faso | PROMIS (38) | 444 | 484 | −1.04 | 0.16 (0.04, 0.28) | 0.12 | 0.12 | 257 | 274 | −2.17 | 0.07 (−0.10, 0.23) | 0.06 | 0.08 |
| Burkina Faso | PROMIS CS (38) | | | | | | | | | | | | |
| Ghana | GHANA (39) | | | | | | | | | | | | |
| Ghana | iLiNS−DYADG (40) | 203 | 387 | −0.37 | 0.16 (0.03, 0.30) | 0.10 | 0.10 | 120 | 254 | −1.68 | 0.10 (−0.05, 0.26) | 0.07 | 0.09 |
| Haiti | HAITI (41) | 101 | 109 | −0.30 | 0.13 (−0.12, 0.37) | 0.03 | 0.03 | 48 | 40 | −1.95 | 0.12 (−0.23, 0.46) | 0.01 | 0.03 |
| Kenya | WASH−B (42) | | | | | | | | | | | | |
| Madagascar | MAHAY (43) | | | | | | | | | | | | |
| Malawi | iLiNS−DYADM (44) | 82 | 179 | −0.91 | −0.06 (−0.29, 0.17) | 0.03 | 0.03 | 122 | 244 | −2.16 | −0.04 (−0.24, 0.15) | 0.04 | 0.07 |
| Malawi | iLiNS−DOSE (45) | 260 | 79 | −0.94 | −0.06 (−0.27, 0.15) | 0.04 | 0.04 | 436 | 162 | −2.42 | 0.12 (−0.04, 0.27) | 0.07 | 0.09 |
| Mali | PROMIS (46) | 296 | 266 | −1.18 | 0.21 (0.00, 0.42) | 0.04 | 0.04 | 210 | 240 | −2.33 | 0.15 (−0.05, 0.35) | 0.04 | 0.07 |
| Mali | PROMIS CS (46) | | | | | | | | | | | | |
| Zimbabwe | SHINE (HIV−) (47) | 725 | 638 | −1.01 | 0.09 (−0.01, 0.19) | 0.17 | 0.17 | 602 | 571 | −2.11 | 0.08 (−0.03, 0.19) | 0.14 | 0.13 |
| Zimbabwe | SHINE (HIV+) (48) | 103 | 107 | −1.28 | 0.29 (0.03, 0.56) | 0.02 | 0.03 | 165 | 131 | −2.40 | 0.22 (0.00, 0.45) | 0.03 | 0.05 |
| | | **4738** | **3300** | | **I² = 0.03, Tau² = 0.00** | | | **5827** | **3532** | | **I² = 0.43, Tau² = 0.00** | | |
| **Fixed** | | | | | **0.11 (0.07, 0.15)** | | | | | | **0.11 (0.07, 0.15)** | | |
| **Random** | | | | | **0.11 (0.07, 0.16)** | | | | | | **0.12 (0.06, 0.18)** | | |

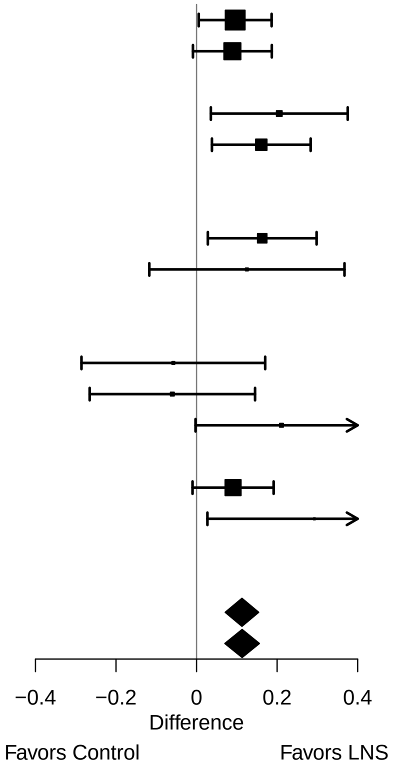

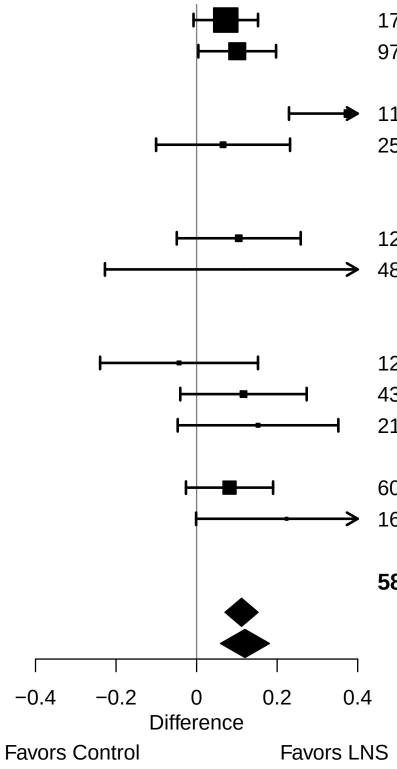

### Supplemental figure 6B: Stunting prevalence ratio

#### 6B1: Stratified by Maternal height

**P−for−interaction = 0.020**
**Ratio of PRs = 1.09 (1.01, 1.18)**

| | | At least 150.1 cm | | | | | | Less than 150.1 cm | | | | | |
|---|---|---|---|---|---|---|---|---|---|---|---|---|---|
| **Country** | **Trial** | **LNS N** | **Control N** | **Control Prevalence** | **PR (95% CI)** | **Fixed W** | **Random W** | **LNS N** | **Control N** | **Control Prevalence** | **PR (95% CI)** | **Fixed W** | **Random W** |
| Bangladesh | JiVitA−4 (34) | | | | | | | | | | | | |
| Bangladesh | RDNS (35) | 868 | 432 | 30.6 | 0.86 (0.70, 1.06) | 0.05 | 0.08 | 735 | 358 | 56.1 | 0.94 (0.84, 1.05) | 0.23 | 0.23 |
| Bangladesh | WASH−B (36) | 616 | 1883 | 32.4 | 0.76 (0.66, 0.88) | 0.10 | 0.11 | 542 | 1545 | 53.5 | 0.88 (0.80, 0.97) | 0.29 | 0.29 |
| Burkina Faso | iLiNS−Zinc (37) | 1916 | 649 | 38.8 | 0.75 (0.64, 0.87) | 0.09 | 0.10 | 34 | 15 | 66.7 | 0.99 (0.65, 1.50) | 0.02 | 0.02 |
| Burkina Faso | PROMIS (38) | 835 | 884 | 29.6 | 0.80 (0.64, 1.00) | 0.04 | 0.07 | 25 | 25 | 68.0 | 0.88 (0.47, 1.65) | 0.01 | 0.01 |
| Burkina Faso | PROMIS CS (38) | | | | | | | | | | | | |
| Ghana | GHANA (39) | | | | | | | | | | | | |
| Ghana | iLiNS−DYADG (40) | | | | | | | | | | | | |
| Haiti | HAITI (41) | | | | | | | | | | | | |
| Kenya | WASH−B (42) | 1313 | 4686 | 30.7 | 0.87 (0.78, 0.97) | 0.18 | 0.13 | 57 | 190 | 58.4 | 0.84 (0.61, 1.15) | 0.03 | 0.03 |
| Madagascar | MAHAY (43) | 1004 | 1037 | 53.9 | 0.94 (0.84, 1.05) | 0.17 | 0.13 | 598 | 565 | 68.8 | 1.01 (0.91, 1.11) | 0.29 | 0.29 |
| Malawi | iLiNS−DYADM (44) | 192 | 381 | 29.9 | 1.13 (0.88, 1.45) | 0.03 | 0.06 | 28 | 60 | 61.7 | 1.10 (0.80, 1.52) | 0.03 | 0.03 |
| Malawi | iLiNS−DOSE (45) | 580 | 198 | 41.9 | 0.98 (0.83, 1.16) | 0.08 | 0.10 | 115 | 42 | 66.7 | 1.03 (0.82, 1.30) | 0.05 | 0.05 |
| Mali | PROMIS (46) | | | | | | | | | | | | |
| Mali | PROMIS CS (46) | 914 | 936 | 32.9 | 0.76 (0.63, 0.93) | 0.06 | 0.08 | 33 | 27 | 66.7 | 0.91 (0.67, 1.23) | 0.03 | 0.03 |
| Zimbabwe | SHINE (HIV−) (47) | 1755 | 1697 | 34.0 | 0.77 (0.69, 0.85) | 0.19 | 0.13 | 76 | 62 | 61.3 | 0.90 (0.70, 1.16) | 0.04 | 0.04 |
| Zimbabwe | SHINE (HIV+) (48) | | | | | | | | | | | | |
| | | **9993** | **12783** | | **I² = 0.57, Tau² = 0.01** | | | **2243** | **2889** | | **I² = 0.00, Tau² = 0.00** | | |
| **Fixed** | | | | | **0.84 (0.81, 0.88)** | | | | | | **0.94 (0.90, 0.99)** | | |
| **Random** | | | | | **0.85 (0.78, 0.92)** | | | | | | **0.94 (0.90, 0.99)** | | |

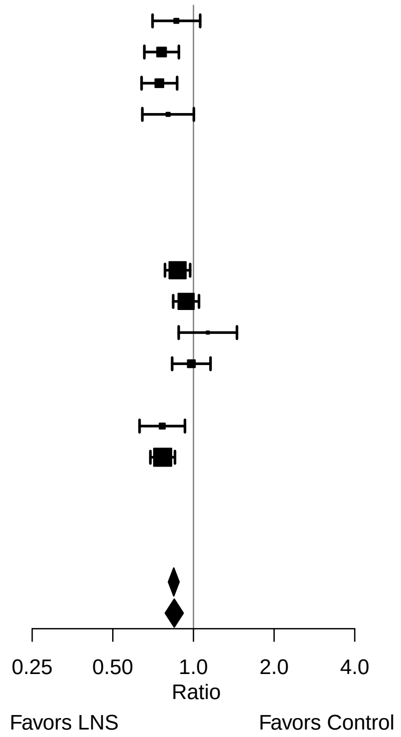

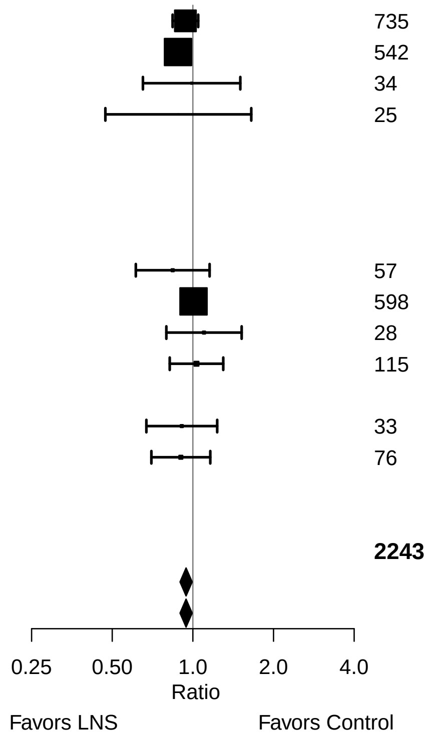

#### Supplemental figure 6B: Stunting prevalence ratio

#### 6B2: Stratified by Maternal BMI

**P−for−interaction = 0.017**
**Ratio of PRs = 1.10 (1.02, 1.19)**

| | | At least 20 $kg/m^2$ | | | | | | Less than 20 $kg/m^2$ | | | | | |
|---|---|---|---|---|---|---|---|---|---|---|---|---|---|
| **Country** | **Trial** | **LNS N** | **Control N** | **Control Prevalence** | **PR (95% CI)** | **Fixed W** | **Random W** | **LNS N** | **Control N** | **Control Prevalence** | **PR (95% CI)** | **Fixed W** | **Random W** |
| Bangladesh | JiVitA−4 (34) | | | | | | | | | | | | |
| Bangladesh | RDNS (35) | 730 | 335 | 38.8 | 0.86 (0.73, 1.01) | 0.09 | 0.10 | 873 | 455 | 44.6 | 0.96 (0.86, 1.06) | 0.30 | 0.22 |
| Bangladesh | WASH−B (36) | 519 | 1576 | 37.3 | 0.84 (0.72, 0.98) | 0.10 | 0.10 | 639 | 1852 | 45.8 | 0.84 (0.75, 0.93) | 0.26 | 0.20 |
| Burkina Faso | iLiNS−Zinc (37) | 1214 | 390 | 37.1 | 0.67 (0.58, 0.77) | 0.11 | 0.10 | 736 | 274 | 42.7 | 0.87 (0.69, 1.10) | 0.06 | 0.08 |
| Burkina Faso | PROMIS (38) | 647 | 675 | 29.9 | 0.71 (0.56, 0.89) | 0.04 | 0.07 | 213 | 233 | 32.6 | 1.11 (0.82, 1.50) | 0.04 | 0.05 |
| Burkina Faso | PROMIS CS (38) | 246 | 269 | 21.6 | 0.89 (0.61, 1.29) | 0.02 | 0.03 | 184 | 166 | 28.3 | 0.84 (0.59, 1.21) | 0.02 | 0.03 |
| Ghana | GHANA (39) | | | | | | | | | | | | |
| Ghana | iLiNS−DYADG (40) | | | | | | | | | | | | |
| Haiti | HAITI (41) | | | | | | | | | | | | |
| Kenya | WASH−B (42) | 1098 | 3803 | 30.6 | 0.90 (0.81, 1.01) | 0.18 | 0.12 | 272 | 1073 | 36.1 | 0.78 (0.63, 0.98) | 0.07 | 0.08 |
| Madagascar | MAHAY (43) | | | | | | | | | | | | |
| Malawi | iLiNS−DYADM (44) | 131 | 263 | 33.8 | 1.11 (0.84, 1.46) | 0.03 | 0.05 | 88 | 178 | 34.8 | 1.11 (0.80, 1.54) | 0.03 | 0.04 |
| Malawi | iLiNS−DOSE (45) | 513 | 174 | 44.8 | 0.98 (0.83, 1.16) | 0.08 | 0.09 | 180 | 65 | 50.8 | 1.04 (0.80, 1.34) | 0.05 | 0.06 |
| Mali | PROMIS (46) | 380 | 370 | 34.9 | 0.88 (0.69, 1.11) | 0.04 | 0.06 | 120 | 135 | 40.7 | 1.01 (0.78, 1.31) | 0.05 | 0.06 |
| Mali | PROMIS CS (46) | 702 | 684 | 31.1 | 0.83 (0.66, 1.03) | 0.05 | 0.07 | 245 | 279 | 40.5 | 0.70 (0.54, 0.90) | 0.05 | 0.06 |
| Zimbabwe | SHINE (HIV−) (47) | 1364 | 1305 | 35.9 | 0.74 (0.66, 0.82) | 0.20 | 0.12 | 240 | 249 | 34.1 | 0.98 (0.77, 1.24) | 0.06 | 0.07 |
| Zimbabwe | SHINE (HIV+) (48) | 247 | 254 | 48.8 | 0.75 (0.62, 0.92) | 0.06 | 0.08 | 62 | 52 | 53.8 | 0.93 (0.65, 1.33) | 0.02 | 0.04 |
| | | **7791** | **10098** | | **$I^2$ = 0.58, $Tau^2$ = 0.01** | | | **3852** | **5011** | | **$I^2$ = 0.22, $Tau^2$ = 0.00** | | |
| **Fixed** | | | | | **0.82 (0.78, 0.86)** | | | | | | **0.90 (0.86, 0.96)** | | |
| **Random** | | | | | **0.83 (0.76, 0.89)** | | | | | | **0.91 (0.85, 0.97)** | | |

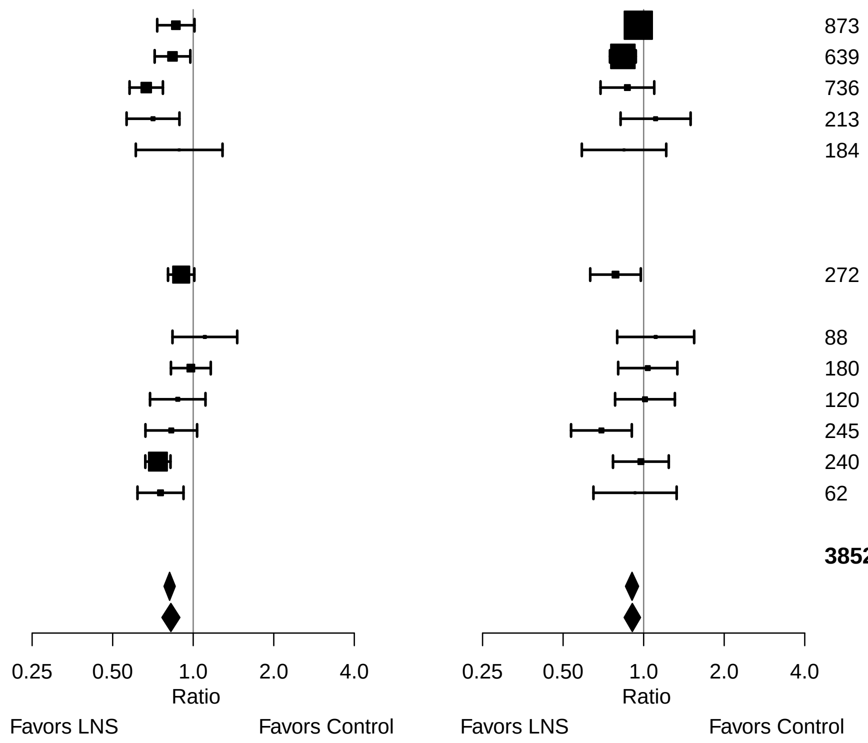

#### Supplemental figure 6B: Stunting prevalence ratio

### 6B3: Stratified by Maternal age

**P−for−interaction = 0.192**
**Ratio of PRs = 1.04 (0.98, 1.10)**

| | | At least 25 y | | | | | | Less than 25 y | | | | | |
|---|---|---|---|---|---|---|---|---|---|---|---|---|---|
| **Country** | **Trial** | **LNS N** | **Control N** | **Control Prevalence** | **PR (95% CI)** | **Fixed W** | **Random W** | **LNS N** | **Control N** | **Control Prevalence** | **PR (95% CI)** | **Fixed W** | **Random W** |
| Bangladesh | JiVitA−4 (34) | 1218 | 546 | 46.3 | 0.91 (0.80, 1.03) | 0.11 | 0.10 | 1608 | 693 | 42.6 | 0.93 (0.83, 1.04) | 0.14 | 0.10 |
| Bangladesh | RDNS (35) | 453 | 221 | 40.7 | 0.80 (0.65, 0.97) | 0.05 | 0.06 | 1210 | 594 | 42.4 | 0.96 (0.88, 1.06) | 0.20 | 0.11 |
| Bangladesh | WASH−B (36) | 492 | 1512 | 42.7 | 0.85 (0.76, 0.95) | 0.15 | 0.11 | 661 | 1903 | 41.4 | 0.82 (0.73, 0.93) | 0.12 | 0.10 |
| Burkina Faso | iLiNS−Zinc (37) | 1131 | 399 | 38.0 | 0.66 (0.55, 0.80) | 0.06 | 0.07 | 808 | 262 | 41.6 | 0.86 (0.73, 1.03) | 0.06 | 0.08 |
| Burkina Faso | PROMIS (38) | 485 | 526 | 30.8 | 0.77 (0.60, 0.99) | 0.03 | 0.04 | 378 | 388 | 30.2 | 0.89 (0.70, 1.13) | 0.03 | 0.05 |
| Burkina Faso | PROMIS CS (38) | 202 | 192 | 22.4 | 0.86 (0.62, 1.20) | 0.02 | 0.03 | 228 | 247 | 25.9 | 0.88 (0.61, 1.26) | 0.01 | 0.03 |
| Ghana | GHANA (39) | | | | | | | | | | | | |
| Ghana | iLiNS−DYADG (40) | 220 | 416 | 11.1 | 0.95 (0.59, 1.52) | 0.01 | 0.01 | 127 | 276 | 15.9 | 0.35 (0.16, 0.75) | 0.00 | 0.01 |
| Haiti | HAITI (41) | 101 | 103 | 9.7 | 1.02 (0.46, 2.23) | 0.00 | 0.01 | 47 | 45 | 22.2 | 1.15 (0.59, 2.23) | 0.00 | 0.01 |
| Kenya | WASH−B (42) | 844 | 2811 | 30.1 | 0.89 (0.79, 1.02) | 0.11 | 0.10 | 598 | 2294 | 34.8 | 0.81 (0.71, 0.92) | 0.10 | 0.09 |
| Madagascar | MAHAY (43) | 896 | 949 | 59.0 | 0.97 (0.88, 1.07) | 0.19 | 0.12 | 805 | 733 | 57.8 | 0.99 (0.89, 1.12) | 0.13 | 0.10 |
| Malawi | iLiNS−DYADM (44) | 106 | 226 | 38.9 | 1.04 (0.79, 1.38) | 0.02 | 0.04 | 114 | 218 | 30.3 | 1.19 (0.87, 1.63) | 0.02 | 0.04 |
| Malawi | iLiNS−DOSE (45) | 368 | 134 | 41.8 | 1.09 (0.90, 1.33) | 0.05 | 0.06 | 314 | 100 | 51.0 | 0.95 (0.77, 1.16) | 0.04 | 0.06 |
| Mali | PROMIS (46) | 346 | 359 | 36.8 | 0.85 (0.70, 1.03) | 0.05 | 0.06 | 160 | 147 | 35.4 | 1.08 (0.75, 1.54) | 0.01 | 0.03 |
| Mali | PROMIS CS (46) | 503 | 507 | 31.0 | 0.82 (0.63, 1.06) | 0.03 | 0.04 | 449 | 462 | 36.8 | 0.75 (0.60, 0.94) | 0.03 | 0.06 |
| Zimbabwe | SHINE (HIV−) (47) | 875 | 800 | 32.0 | 0.79 (0.68, 0.93) | 0.07 | 0.08 | 808 | 797 | 38.5 | 0.79 (0.68, 0.91) | 0.09 | 0.09 |
| Zimbabwe | SHINE (HIV+) (48) | 244 | 231 | 50.2 | 0.78 (0.64, 0.95) | 0.05 | 0.06 | 73 | 78 | 50.0 | 0.93 (0.68, 1.28) | 0.02 | 0.04 |
| | | **8484** | **9932** | | **I² = 0.40, Tau² = 0.01** | | | **8388** | **9237** | | **I² = 0.44, Tau² = 0.01** | | |
| **Fixed** | | | | | **0.87 (0.84, 0.91)** | | | | | | **0.90 (0.86, 0.94)** | | |
| **Random** | | | | | **0.86 (0.81, 0.92)** | | | | | | **0.89 (0.83, 0.95)** | | |

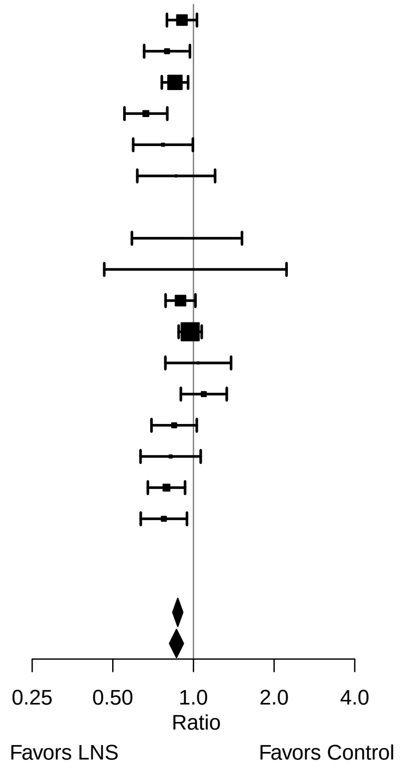

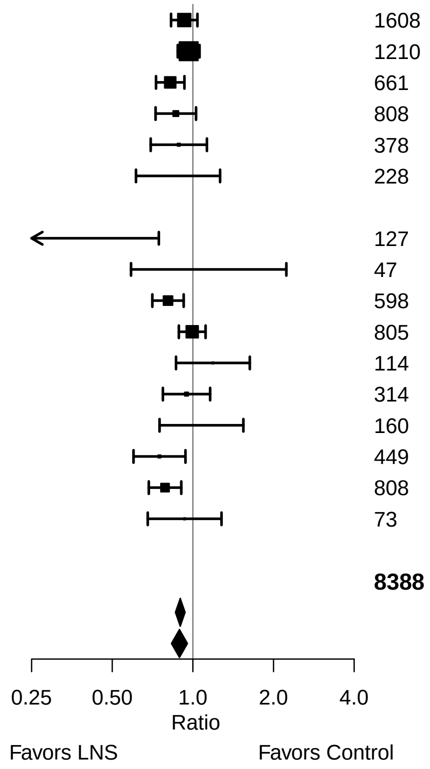

#### Supplemental figure 6B: Stunting prevalence ratio

##### 6B4: Stratified by Maternal education

**P−for−interaction = 0.042**
**Ratio of PRs = 1.07 (1.00, 1.15)**

| | | Primary or greater | | | | | | Incomplete or no formal | | | | | |
|---|---|---|---|---|---|---|---|---|---|---|---|---|---|
| **Country** | **Trial** | **LNS N** | **Control N** | **Control Prevalence** | **PR (95% CI)** | **Fixed W** | **Random W** | **LNS N** | **Control N** | **Control Prevalence** | **PR (95% CI)** | **Fixed W** | **Random W** |
| Bangladesh | JiVitA−4 (34) | 1830 | 756 | 40.1 | 0.87 (0.78, 0.97) | 0.16 | 0.16 | 1006 | 485 | 50.8 | 0.97 (0.86, 1.09) | 0.12 | 0.10 |
| Bangladesh | RDNS (35) | 1245 | 589 | 40.2 | 0.88 (0.79, 0.99) | 0.16 | 0.15 | 418 | 226 | 46.5 | 1.03 (0.90, 1.18) | 0.09 | 0.09 |
| Bangladesh | WASH−B (36) | 813 | 2452 | 38.4 | 0.78 (0.71, 0.87) | 0.19 | 0.17 | 345 | 979 | 50.7 | 0.93 (0.81, 1.06) | 0.09 | 0.09 |
| Burkina Faso | iLiNS−Zinc (37) | 82 | 16 | 31.2 | 0.46 (0.22, 0.97) | 0.00 | 0.00 | 1857 | 645 | 39.5 | 0.77 (0.66, 0.89) | 0.08 | 0.08 |
| Burkina Faso | PROMIS (38) | 70 | 54 | 27.8 | 1.13 (0.62, 2.05) | 0.01 | 0.01 | 793 | 860 | 30.7 | 0.80 (0.65, 0.97) | 0.04 | 0.06 |
| Burkina Faso | PROMIS CS (38) | 34 | 39 | 25.6 | 0.69 (0.35, 1.37) | 0.00 | 0.01 | 393 | 399 | 24.3 | 0.89 (0.65, 1.23) | 0.02 | 0.03 |
| Ghana | GHANA (39) | | | | | | | | | | | | |
| Ghana | iLiNS−DYADG (40) | 267 | 548 | 13.7 | 0.57 (0.36, 0.91) | 0.01 | 0.01 | 80 | 144 | 10.4 | 1.08 (0.50, 2.36) | 0.00 | 0.01 |
| Haiti | HAITI (41) | | | | | | | | | | | | |
| Kenya | WASH−B (42) | 700 | 2434 | 26.0 | 0.91 (0.78, 1.06) | 0.09 | 0.10 | 755 | 2699 | 37.8 | 0.82 (0.74, 0.92) | 0.14 | 0.11 |
| Madagascar | MAHAY (43) | 351 | 431 | 56.8 | 0.97 (0.84, 1.12) | 0.10 | 0.10 | 1351 | 1251 | 59.1 | 0.98 (0.90, 1.08) | 0.19 | 0.12 |
| Malawi | iLiNS−DYADM (44) | 35 | 69 | 29.0 | 0.99 (0.52, 1.87) | 0.00 | 0.01 | 184 | 372 | 35.8 | 1.12 (0.90, 1.41) | 0.03 | 0.06 |
| Malawi | iLiNS−DOSE (45) | 162 | 56 | 44.6 | 0.86 (0.58, 1.26) | 0.01 | 0.02 | 522 | 178 | 46.1 | 1.07 (0.92, 1.25) | 0.07 | 0.08 |
| Mali | PROMIS (46) | 41 | 37 | 24.3 | 1.08 (0.54, 2.17) | 0.00 | 0.01 | 464 | 469 | 37.3 | 0.91 (0.77, 1.09) | 0.06 | 0.07 |
| Mali | PROMIS CS (46) | 107 | 96 | 32.3 | 0.69 (0.43, 1.12) | 0.01 | 0.01 | 844 | 873 | 33.9 | 0.80 (0.66, 0.96) | 0.05 | 0.07 |
| Zimbabwe | SHINE (HIV−) (47) | 1717 | 1644 | 34.8 | 0.79 (0.71, 0.87) | 0.20 | 0.18 | 69 | 61 | 36.1 | 0.88 (0.56, 1.40) | 0.01 | 0.02 |
| Zimbabwe | SHINE (HIV+) (48) | 301 | 293 | 48.5 | 0.84 (0.70, 1.01) | 0.06 | 0.07 | 21 | 18 | 66.7 | 0.57 (0.31, 1.05) | 0.00 | 0.01 |
| | | **7755** | **9514** | | **I² = 0.16, Tau² = 0.00** | | | **9102** | **9659** | | **I² = 0.52, Tau² = 0.01** | | |
| **Fixed** | | | | | **0.84 (0.81, 0.88)** | | | | | | **0.92 (0.89, 0.96)** | | |
| **Random** | | | | | **0.85 (0.80, 0.89)** | | | | | | **0.92 (0.86, 0.98)** | | |

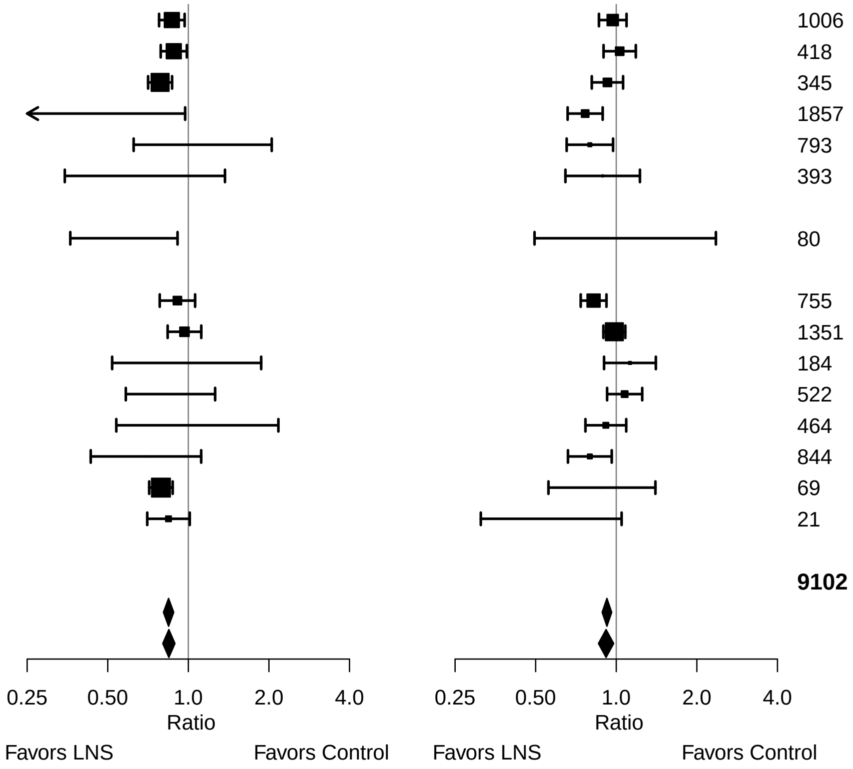

#### Supplemental figure 6B: Stunting prevalence ratio

##### 6B5: Stratified by Maternal depressive symptoms

**P−for−interaction = 0.003**
**Ratio of PRs = 0.89 (0.82, 0.96)**

| | | Less than 75th percentile | | | | | | At least 75th percentile | | | | | |
|---|---|---|---|---|---|---|---|---|---|---|---|---|---|
| **Country** | **Trial** | **LNS N** | **Control N** | **Control Prevalence** | **PR (95% CI)** | **Fixed W** | **Random W** | **LNS N** | **Control N** | **Control Prevalence** | **PR (95% CI)** | **Fixed W** | **Random W** |
| Bangladesh | JiVitA−4 (34) | | | | | | | | | | | | |
| Bangladesh | RDNS (35) | 1081 | 472 | 42.4 | 0.87 (0.79, 0.97) | 0.21 | 0.17 | 520 | 288 | 41.0 | 1.00 (0.81, 1.23) | 0.11 | 0.12 |
| Bangladesh | WASH−B (36) | 897 | 2462 | 41.3 | 0.80 (0.72, 0.89) | 0.21 | 0.17 | 237 | 878 | 43.8 | 0.94 (0.79, 1.12) | 0.15 | 0.13 |
| Burkina Faso | iLiNS−Zinc (37) | | | | | | | | | | | | |
| Burkina Faso | PROMIS (38) | 508 | 573 | 30.7 | 0.77 (0.58, 1.02) | 0.03 | 0.04 | 223 | 215 | 28.8 | 1.03 (0.77, 1.37) | 0.06 | 0.09 |
| Burkina Faso | PROMIS CS (38) | | | | | | | | | | | | |
| Ghana | GHANA (39) | | | | | | | | | | | | |
| Ghana | iLiNS−DYADG (40) | 252 | 459 | 13.3 | 0.51 (0.30, 0.85) | 0.01 | 0.01 | 83 | 209 | 10.5 | 1.37 (0.71, 2.65) | 0.01 | 0.03 |
| Haiti | HAITI (41) | | | | | | | | | | | | |
| Kenya | WASH−B (42) | 996 | 3496 | 31.1 | 0.87 (0.76, 0.98) | 0.14 | 0.14 | 339 | 1235 | 34.3 | 0.94 (0.79, 1.12) | 0.15 | 0.13 |
| Madagascar | MAHAY (43) | 673 | 721 | 60.5 | 0.94 (0.83, 1.05) | 0.16 | 0.15 | 284 | 252 | 60.3 | 1.08 (0.94, 1.23) | 0.25 | 0.15 |
| Malawi | iLiNS−DYADM (44) | 157 | 303 | 31.7 | 1.03 (0.78, 1.36) | 0.03 | 0.04 | 47 | 113 | 39.8 | 1.44 (1.03, 2.02) | 0.04 | 0.07 |
| Malawi | iLiNS−DOSE (45) | | | | | | | | | | | | |
| Mali | PROMIS (46) | 379 | 373 | 35.1 | 0.92 (0.74, 1.13) | 0.05 | 0.07 | 123 | 131 | 40.5 | 0.89 (0.67, 1.18) | 0.06 | 0.09 |
| Mali | PROMIS CS (46) | | | | | | | | | | | | |
| Zimbabwe | SHINE (HIV−) (47) | 1198 | 1145 | 33.8 | 0.82 (0.72, 0.94) | 0.13 | 0.13 | 513 | 489 | 38.2 | 0.71 (0.59, 0.86) | 0.13 | 0.12 |
| Zimbabwe | SHINE (HIV+) (48) | 234 | 215 | 49.3 | 0.75 (0.60, 0.94) | 0.04 | 0.06 | 89 | 100 | 53.0 | 0.98 (0.73, 1.31) | 0.05 | 0.08 |
| | | **6375** | **10219** | | **I² = 0.26, Tau² = 0.00** | | | **2458** | **3910** | | **I² = 0.56, Tau² = 0.02** | | |
| **Fixed** | | | | | **0.85 (0.81, 0.89)** | | | | | | **0.97 (0.91, 1.04)** | | |
| **Random** | | | | | **0.85 (0.80, 0.91)** | | | | | | **0.98 (0.87, 1.09)** | | |

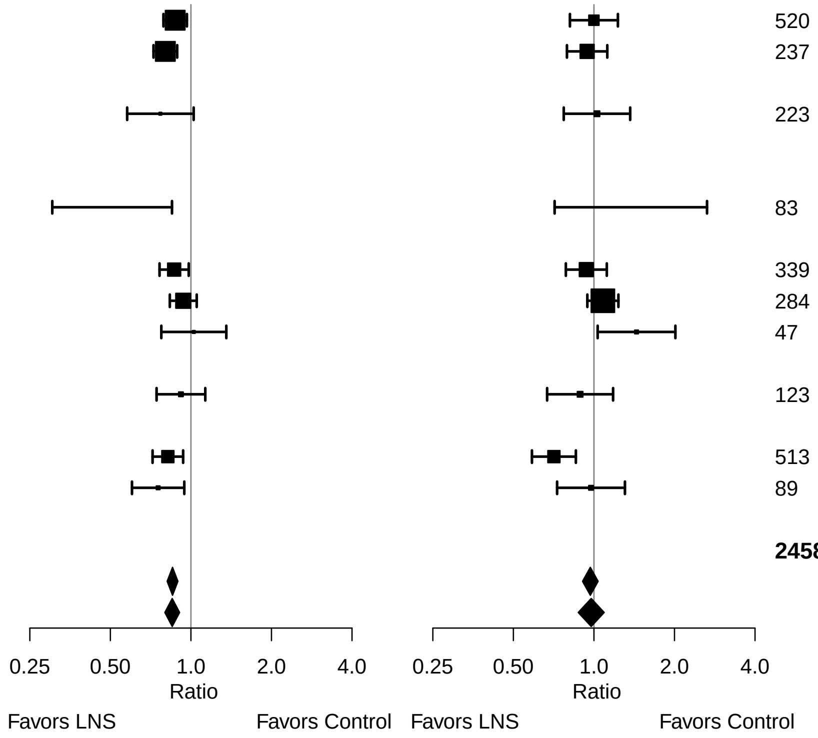

#### Supplemental figure 6B: Stunting prevalence ratio

#### 6B6: Stratified by Child sex

**P−for−interaction = 0.002**
**Ratio of PRs = 0.92 (0.87, 0.97)**

| Country | Trial | Male LNS N | Male Control N | Male Control Prevalence | Male PR (95% CI) | Male Fixed W | Male Random W | Female LNS N | Female Control N | Female Control Prevalence | Female PR (95% CI) | Female Fixed W | Female Random W |
|---|---|---|---|---|---|---|---|---|---|---|---|---|---|
| Bangladesh | JiVitA−4 (34) | 1448 | 600 | 44.5 | 0.94 (0.84, 1.06) | 0.11 | 0.10 | 1390 | 644 | 43.9 | 0.90 (0.81, 1.01) | 0.17 | 0.13 |
| Bangladesh | RDNS (35) | 833 | 411 | 42.6 | 1.00 (0.87, 1.15) | 0.07 | 0.08 | 830 | 404 | 41.3 | 0.84 (0.73, 0.96) | 0.12 | 0.11 |
| Bangladesh | WASH−B (36) | 565 | 1714 | 43.1 | 0.83 (0.75, 0.93) | 0.12 | 0.10 | 593 | 1717 | 40.7 | 0.85 (0.74, 0.97) | 0.12 | 0.11 |
| Burkina Faso | iLiNS−Zinc (37) | 990 | 336 | 43.2 | 0.81 (0.69, 0.94) | 0.06 | 0.07 | 962 | 328 | 35.6 | 0.68 (0.57, 0.81) | 0.07 | 0.08 |
| Burkina Faso | PROMIS (38) | 438 | 483 | 36.2 | 0.79 (0.62, 1.01) | 0.03 | 0.04 | 425 | 431 | 24.1 | 0.88 (0.68, 1.14) | 0.03 | 0.05 |
| Burkina Faso | PROMIS CS (38) | 234 | 222 | 29.3 | 0.95 (0.68, 1.32) | 0.01 | 0.02 | 196 | 217 | 19.4 | 0.69 (0.41, 1.16) | 0.01 | 0.01 |
| Ghana | GHANA (39) | | | | | | | | | | | | |
| Ghana | iLiNS−DYADG (40) | 175 | 323 | 16.7 | 0.75 (0.47, 1.19) | 0.01 | 0.01 | 172 | 369 | 9.8 | 0.48 (0.23, 1.00) | 0.00 | 0.01 |
| Haiti | HAITI (41) | 70 | 59 | 18.6 | 0.78 (0.40, 1.52) | 0.00 | 0.01 | 79 | 90 | 10.0 | 1.19 (0.50, 2.85) | 0.00 | 0.01 |
| Kenya | WASH−B (42) | 734 | 2440 | 36.8 | 0.93 (0.83, 1.04) | 0.11 | 0.10 | 723 | 2697 | 28.1 | 0.75 (0.64, 0.88) | 0.09 | 0.09 |
| Madagascar | MAHAY (43) | 826 | 832 | 63.3 | 1.02 (0.94, 1.12) | 0.19 | 0.12 | 876 | 850 | 53.8 | 0.94 (0.83, 1.07) | 0.14 | 0.11 |
| Malawi | iLiNS−DYADM (44) | 107 | 207 | 38.6 | 1.11 (0.84, 1.47) | 0.02 | 0.03 | 113 | 237 | 31.2 | 1.08 (0.78, 1.48) | 0.02 | 0.03 |
| Malawi | iLiNS−DOSE (45) | 352 | 124 | 54.8 | 1.00 (0.85, 1.18) | 0.06 | 0.07 | 344 | 117 | 37.6 | 0.99 (0.77, 1.28) | 0.03 | 0.05 |
| Mali | PROMIS (46) | 266 | 263 | 38.8 | 1.01 (0.82, 1.25) | 0.03 | 0.05 | 240 | 243 | 33.7 | 0.79 (0.60, 1.04) | 0.03 | 0.04 |
| Mali | PROMIS CS (46) | 477 | 525 | 36.6 | 0.85 (0.69, 1.05) | 0.03 | 0.05 | 475 | 444 | 30.4 | 0.72 (0.56, 0.93) | 0.04 | 0.05 |
| Zimbabwe | SHINE (HIV−) (47) | 932 | 908 | 41.9 | 0.77 (0.68, 0.86) | 0.11 | 0.10 | 948 | 886 | 27.7 | 0.84 (0.71, 0.98) | 0.09 | 0.09 |
| Zimbabwe | SHINE (HIV+) (48) | 162 | 169 | 53.8 | 0.86 (0.70, 1.05) | 0.04 | 0.05 | 175 | 161 | 46.0 | 0.76 (0.59, 0.97) | 0.04 | 0.05 |
| | | **8609** | **9616** | | **$I^2$ = 0.49, $Tau^2$ = 0.00** | | | **8541** | **9835** | | **$I^2$ = 0.31, $Tau^2$ = 0.00** | | |
| **Fixed** | | | | | **0.91 (0.88, 0.95)** | | | | | | **0.84 (0.80, 0.88)** | | |
| **Random** | | | | | **0.91 (0.86, 0.96)** | | | | | | **0.83 (0.78, 0.89)** | | |

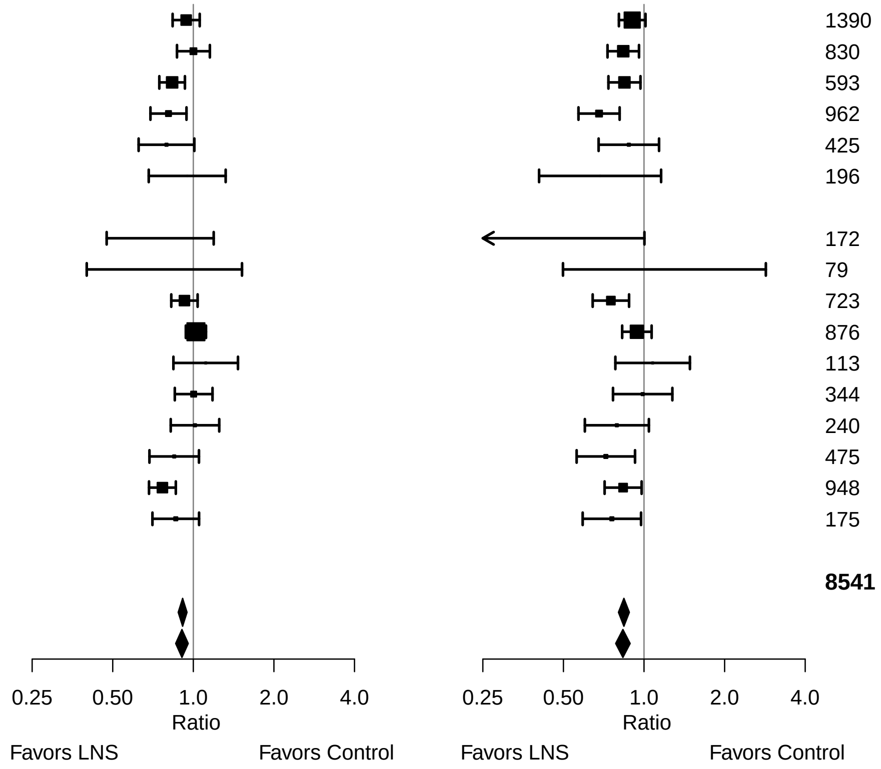

#### Supplemental figure 6B: Stunting prevalence ratio

#### 6B7: Stratified by Child birth order

**P−for−interaction = 0.054**
**Ratio of PRs = 1.06 (1.00, 1.13)**

| Country | Trial | Later born LNS N | Later born Control N | Later born Control Prevalence | Later born PR (95% CI) | Later born Fixed W | Later born Random W | Firstborn LNS N | Firstborn Control N | Firstborn Control Prevalence | Firstborn PR (95% CI) | Firstborn Fixed W | Firstborn Random W |
|---|---|---|---|---|---|---|---|---|---|---|---|---|---|
| Bangladesh | JiVitA−4 (34) | 611 | 255 | 49.0 | 0.84 (0.73, 0.98) | 0.06 | 0.07 | 2223 | 982 | 42.8 | 0.94 (0.85, 1.04) | 0.27 | 0.12 |
| Bangladesh | RDNS (35) | 974 | 508 | 42.7 | 0.89 (0.79, 0.99) | 0.11 | 0.09 | 688 | 307 | 40.7 | 0.97 (0.83, 1.14) | 0.11 | 0.10 |
| Bangladesh | WASH−B (36) | 736 | 2290 | 43.4 | 0.84 (0.76, 0.92) | 0.16 | 0.11 | 422 | 1141 | 38.8 | 0.85 (0.72, 0.99) | 0.11 | 0.10 |
| Burkina Faso | iLiNS−Zinc (37) | 1520 | 519 | 38.8 | 0.71 (0.60, 0.84) | 0.05 | 0.06 | 432 | 145 | 41.4 | 0.88 (0.73, 1.06) | 0.08 | 0.09 |
| Burkina Faso | PROMIS (38) | 715 | 754 | 30.1 | 0.81 (0.65, 1.01) | 0.03 | 0.04 | 148 | 158 | 32.3 | 0.88 (0.62, 1.26) | 0.02 | 0.05 |
| Burkina Faso | PROMIS CS (38) | 366 | 359 | 23.7 | 0.89 (0.65, 1.22) | 0.01 | 0.03 | 63 | 80 | 27.5 | 0.81 (0.44, 1.48) | 0.01 | 0.02 |
| Ghana | GHANA (39) | | | | | | | | | | | | |
| Ghana | iLiNS−DYADG (40) | 237 | 458 | 13.3 | 0.79 (0.51, 1.23) | 0.01 | 0.01 | 110 | 234 | 12.4 | 0.37 (0.15, 0.92) | 0.00 | 0.01 |
| Haiti | HAITI (41) | 88 | 99 | 15.2 | 0.86 (0.46, 1.61) | 0.00 | 0.01 | 61 | 50 | 10.0 | 1.71 (0.65, 4.48) | 0.00 | 0.01 |
| Kenya | WASH−B (42) | 1134 | 4045 | 32.1 | 0.88 (0.80, 0.97) | 0.14 | 0.10 | 321 | 1087 | 32.7 | 0.76 (0.62, 0.94) | 0.06 | 0.08 |
| Madagascar | MAHAY (43) | 1272 | 1217 | 59.7 | 0.94 (0.86, 1.04) | 0.16 | 0.11 | 430 | 465 | 55.3 | 1.09 (0.96, 1.25) | 0.15 | 0.11 |
| Malawi | iLiNS−DYADM (44) | 173 | 353 | 35.7 | 1.12 (0.89, 1.41) | 0.03 | 0.04 | 47 | 89 | 30.3 | 1.05 (0.62, 1.77) | 0.01 | 0.03 |
| Malawi | iLiNS−DOSE (45) | 458 | 165 | 44.2 | 1.12 (0.95, 1.32) | 0.05 | 0.06 | 149 | 45 | 53.3 | 0.84 (0.60, 1.16) | 0.03 | 0.05 |
| Mali | PROMIS (46) | 442 | 445 | 37.1 | 0.86 (0.72, 1.03) | 0.04 | 0.06 | 63 | 59 | 30.5 | 1.43 (0.86, 2.36) | 0.01 | 0.03 |
| Mali | PROMIS CS (46) | 790 | 831 | 32.1 | 0.78 (0.65, 0.95) | 0.04 | 0.05 | 145 | 126 | 42.9 | 0.72 (0.53, 0.98) | 0.03 | 0.06 |
| Zimbabwe | SHINE (HIV−) (47) | 1302 | 1223 | 33.5 | 0.81 (0.71, 0.91) | 0.09 | 0.09 | 491 | 487 | 37.8 | 0.78 (0.65, 0.93) | 0.08 | 0.09 |
| Zimbabwe | SHINE (HIV+) (48) | 280 | 247 | 46.6 | 0.87 (0.71, 1.05) | 0.04 | 0.05 | 54 | 78 | 59.0 | 0.66 (0.45, 0.96) | 0.02 | 0.04 |
| | | **11098** | **13768** | | **$I^2$ = 0.43, $Tau^2$ = 0.01** | | | **5847** | **5533** | | **$I^2$ = 0.48, $Tau^2$ = 0.02** | | |
| **Fixed** | | | | | **0.87 (0.84, 0.91)** | | | | | | **0.91 (0.86, 0.96)** | | |
| **Random** | | | | | **0.87 (0.82, 0.92)** | | | | | | **0.88 (0.80, 0.97)** | | |

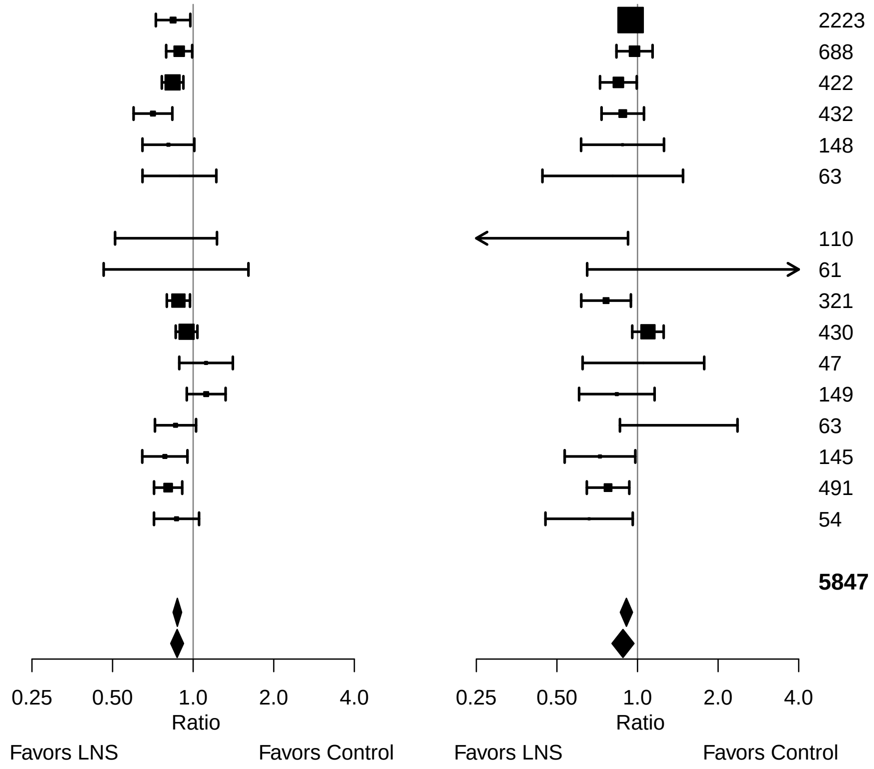

#### Supplemental figure 6B: Stunting prevalence ratio

#### 6B8: Stratified by Child baseline anthropometric status

**P−for−interaction = 0.006**
**Ratio of PRs = 1.25 (1.07, 1.46)**

| | | LAZ at least −1 Z | | | | | | LAZ less than −1 Z | | | | | |
|---|---|---|---|---|---|---|---|---|---|---|---|---|---|
| **Country** | **Trial** | **LNS N** | **Control N** | **Control Prevalence** | **PR (95% CI)** | **Fixed W** | **Random W** | **LNS N** | **Control N** | **Control Prevalence** | **PR (95% CI)** | **Fixed W** | **Random W** |
| Bangladesh | JiVitA−4 (34) | 1055 | 454 | 8.8 | 0.79 (0.52, 1.18) | 0.15 | 0.15 | 1783 | 790 | 64.9 | 0.94 (0.87, 1.01) | 0.30 | 0.19 |
| Bangladesh | RDNS (35) | 630 | 306 | 9.5 | 0.72 (0.47, 1.10) | 0.14 | 0.14 | 971 | 453 | 63.8 | 0.92 (0.85, 1.00) | 0.23 | 0.18 |
| Bangladesh | WASH−B (36) | | | | | | | | | | | | |
| Burkina Faso | iLiNS−Zinc (37) | 839 | 291 | 6.2 | 0.40 (0.19, 0.86) | 0.04 | 0.04 | 1113 | 373 | 65.4 | 0.76 (0.65, 0.88) | 0.07 | 0.10 |
| Burkina Faso | PROMIS (38) | 444 | 484 | 12.4 | 0.69 (0.48, 0.99) | 0.19 | 0.19 | 257 | 274 | 58.4 | 0.93 (0.77, 1.12) | 0.04 | 0.07 |
| Burkina Faso | PROMIS CS (38) | | | | | | | | | | | | |
| Ghana | GHANA (39) | | | | | | | | | | | | |
| Ghana | iLiNS−DYADG (40) | | | | | | | | | | | | |
| Haiti | HAITI (41) | | | | | | | | | | | | |
| Kenya | WASH−B (42) | | | | | | | | | | | | |
| Madagascar | MAHAY (43) | | | | | | | | | | | | |
| Malawi | iLiNS−DYADM (44) | 82 | 179 | 8.4 | 1.31 (0.60, 2.87) | 0.04 | 0.04 | 122 | 244 | 53.7 | 1.10 (0.91, 1.33) | 0.04 | 0.07 |
| Malawi | iLiNS−DOSE (45) | 260 | 79 | 8.9 | 1.13 (0.51, 2.50) | 0.04 | 0.04 | 436 | 162 | 64.8 | 0.96 (0.84, 1.10) | 0.09 | 0.11 |
| Mali | PROMIS (46) | 296 | 266 | 13.2 | 0.87 (0.51, 1.50) | 0.09 | 0.09 | 210 | 240 | 62.1 | 0.88 (0.74, 1.05) | 0.05 | 0.08 |
| Mali | PROMIS CS (46) | | | | | | | | | | | | |
| Zimbabwe | SHINE (HIV−) (47) | 725 | 638 | 12.9 | 0.71 (0.51, 0.98) | 0.24 | 0.24 | 602 | 571 | 54.6 | 0.87 (0.78, 0.97) | 0.14 | 0.14 |
| Zimbabwe | SHINE (HIV+) (48) | 103 | 107 | 25.2 | 0.50 (0.27, 0.91) | 0.07 | 0.07 | 165 | 131 | 65.6 | 0.88 (0.73, 1.05) | 0.05 | 0.07 |
| | | **4434** | **2804** | | **I² = 0.00, Tau² = 0.00** | | | **5659** | **3238** | | **I² = 0.34, Tau² = 0.00** | | |
| **Fixed** | | | | | **0.73 (0.62, 0.85)** | | | | | | **0.91 (0.88, 0.95)** | | |
| **Random** | | | | | **0.73 (0.62, 0.85)** | | | | | | **0.91 (0.86, 0.96)** | | |

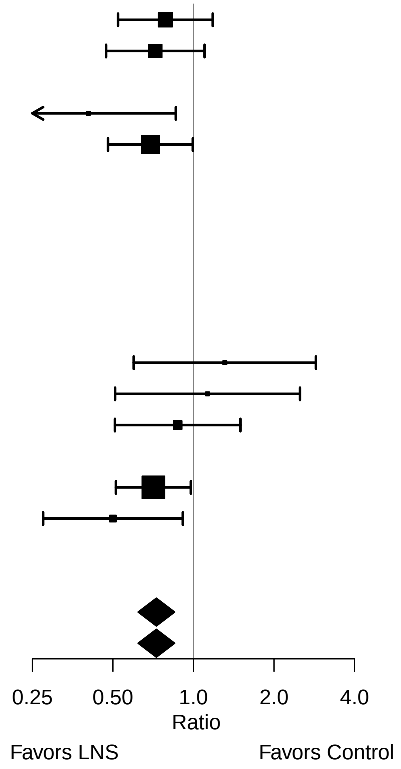

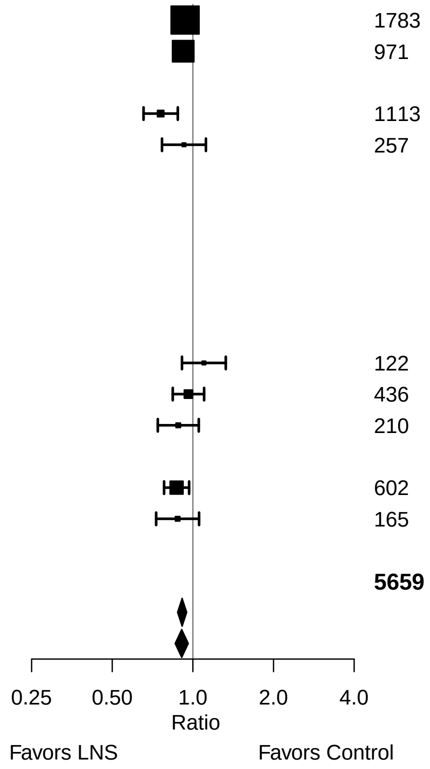

### Supplemental figure 6C: Stunting prevalence difference

#### 6C1: Stratified by Maternal height

**P−for−interaction = 0.339**
**Difference in PDs = 0.02 (−0.02, 0.05)**

| | | At least 150.1 cm | | | | | | Less than 150.1 cm | | | | | |
|---|---|---|---|---|---|---|---|---|---|---|---|---|---|
| **Country** | **Trial** | **LNS N** | **Control N** | **Control Prevalence** | **PD (95% CI)** | **Fixed W** | **Random W** | **LNS N** | **Control N** | **Control Prevalence** | **PD (95% CI)** | **Fixed W** | **Random W** |
| Bangladesh | JiVitA−4 (34) | | | | | | | | | | | | |
| Bangladesh | RDNS (35) | 868 | 432 | 30.6 | −0.04 (−0.10, 0.02) | 0.06 | 0.08 | 735 | 358 | 56.1 | −0.03 (−0.09, 0.03) | 0.24 | 0.24 |
| Bangladesh | WASH−B (36) | 616 | 1883 | 32.4 | −0.08 (−0.12, −0.04) | 0.13 | 0.12 | 542 | 1545 | 53.5 | −0.06 (−0.11, −0.02) | 0.39 | 0.39 |
| Burkina Faso | iLiNS−Zinc (37) | 1916 | 649 | 38.8 | −0.11 (−0.15, −0.06) | 0.10 | 0.11 | 34 | 15 | 66.7 | −0.02 (−0.28, 0.24) | 0.01 | 0.01 |
| Burkina Faso | PROMIS (38) | 835 | 884 | 29.6 | −0.06 (−0.12, 0.00) | 0.06 | 0.08 | 25 | 25 | 68.0 | −0.08 (−0.48, 0.32) | 0.01 | 0.01 |
| Burkina Faso | PROMIS CS (38) | | | | | | | | | | | | |
| Ghana | GHANA (39) | | | | | | | | | | | | |
| Ghana | iLiNS−DYADG (40) | | | | | | | | | | | | |
| Haiti | HAITI (41) | | | | | | | | | | | | |
| Kenya | WASH−B (42) | 1313 | 4686 | 30.7 | −0.04 (−0.07, −0.01) | 0.23 | 0.15 | 57 | 190 | 58.4 | −0.09 (−0.25, 0.07) | 0.03 | 0.03 |
| Madagascar | MAHAY (43) | 1004 | 1037 | 53.9 | −0.03 (−0.09, 0.02) | 0.06 | 0.09 | 598 | 565 | 68.8 | 0.00 (−0.06, 0.07) | 0.19 | 0.19 |
| Malawi | iLiNS−DYADM (44) | 192 | 381 | 29.9 | 0.04 (−0.04, 0.12) | 0.03 | 0.06 | 28 | 60 | 61.7 | 0.06 (−0.16, 0.28) | 0.02 | 0.02 |
| Malawi | iLiNS−DOSE (45) | 580 | 198 | 41.9 | −0.02 (−0.08, 0.04) | 0.05 | 0.08 | 115 | 42 | 66.7 | 0.01 (−0.12, 0.14) | 0.05 | 0.05 |
| Mali | PROMIS (46) | | | | | | | | | | | | |
| Mali | PROMIS CS (46) | 914 | 936 | 32.9 | −0.08 (−0.13, −0.02) | 0.07 | 0.09 | 33 | 27 | 66.7 | −0.06 (−0.26, 0.13) | 0.02 | 0.02 |
| Zimbabwe | SHINE (HIV−) (47) | 1755 | 1697 | 34.0 | −0.08 (−0.11, −0.05) | 0.21 | 0.14 | 76 | 62 | 61.3 | −0.06 (−0.21, 0.09) | 0.04 | 0.04 |
| Zimbabwe | SHINE (HIV+) (48) | | | | | | | | | | | | |
| | | **9993** | **12783** | | **I² = 0.48, Tau² = 0.00** | | | **2243** | **2889** | | **I² = 0.00, Tau² = 0.00** | | |
| **Fixed** | | | | | **−0.06 (−0.07, −0.05)** | | | | | | **−0.04 (−0.07, −0.01)** | | |
| **Random** | | | | | **−0.06 (−0.08, −0.03)** | | | | | | **−0.04 (−0.07, −0.01)** | | |

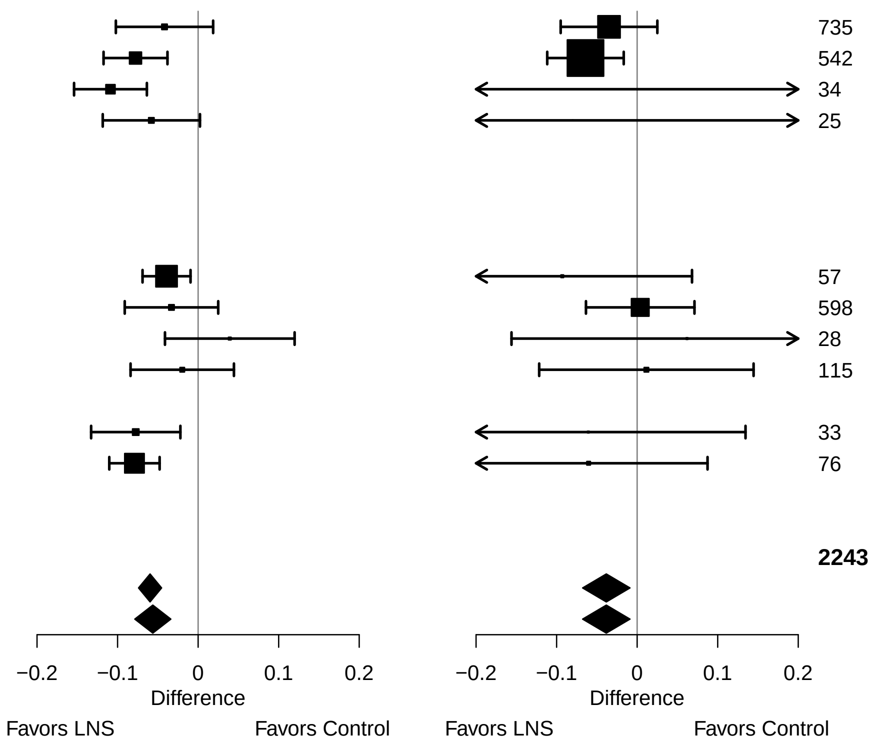

#### Supplemental figure 6C: Stunting prevalence difference

### 6C2: Stratified by Maternal BMI

**P−for−interaction = 0.306**
**Difference in PDs = 0.01 (−0.01, 0.04)**

| | | At least 20 kg/m² | | | | | | Less than 20 kg/m² | | | | | |
|---|---|---|---|---|---|---|---|---|---|---|---|---|---|
| **Country** | **Trial** | **LNS N** | **Control N** | **Control Prevalence** | **PD (95% CI)** | **Fixed W** | **Random W** | **LNS N** | **Control N** | **Control Prevalence** | **PD (95% CI)** | **Fixed W** | **Random W** |
| Bangladesh | JiVitA−4 (34) | | | | | | | | | | | | |
| Bangladesh | RDNS (35) | 730 | 335 | 38.8 | −0.05 (−0.11, 0.01) | 0.06 | 0.08 | 873 | 455 | 44.6 | −0.02 (−0.07, 0.02) | 0.22 | 0.17 |
| Bangladesh | WASH−B (36) | 519 | 1576 | 37.3 | −0.06 (−0.11, −0.01) | 0.08 | 0.09 | 639 | 1852 | 45.8 | −0.08 (−0.12, −0.03) | 0.22 | 0.17 |
| Burkina Faso | iLiNS−Zinc (37) | 1214 | 390 | 37.1 | −0.12 (−0.16, −0.08) | 0.15 | 0.12 | 736 | 274 | 42.7 | −0.08 (−0.15, −0.02) | 0.10 | 0.11 |
| Burkina Faso | PROMIS (38) | 647 | 675 | 29.9 | −0.09 (−0.15, −0.03) | 0.06 | 0.08 | 213 | 233 | 32.6 | 0.04 (−0.07, 0.14) | 0.04 | 0.05 |
| Burkina Faso | PROMIS CS (38) | 246 | 269 | 21.6 | −0.02 (−0.10, 0.05) | 0.04 | 0.06 | 184 | 166 | 28.3 | −0.04 (−0.14, 0.05) | 0.05 | 0.06 |
| Ghana | GHANA (39) | | | | | | | | | | | | |
| Ghana | iLiNS−DYADG (40) | | | | | | | | | | | | |
| Haiti | HAITI (41) | | | | | | | | | | | | |
| Kenya | WASH−B (42) | 1098 | 3803 | 30.6 | −0.03 (−0.06, 0.00) | 0.21 | 0.13 | 272 | 1073 | 36.1 | −0.08 (−0.14, −0.01) | 0.11 | 0.11 |
| Madagascar | MAHAY (43) | | | | | | | | | | | | |
| Malawi | iLiNS−DYADM (44) | 131 | 263 | 33.8 | 0.04 (−0.06, 0.14) | 0.02 | 0.04 | 88 | 178 | 34.8 | 0.04 (−0.08, 0.16) | 0.03 | 0.04 |
| Malawi | iLiNS−DOSE (45) | 513 | 174 | 44.8 | −0.02 (−0.08, 0.05) | 0.05 | 0.07 | 180 | 65 | 50.8 | −0.02 (−0.14, 0.09) | 0.03 | 0.04 |
| Mali | PROMIS (46) | 380 | 370 | 34.9 | −0.05 (−0.12, 0.01) | 0.04 | 0.07 | 120 | 135 | 40.7 | 0.01 (−0.08, 0.10) | 0.05 | 0.07 |
| Mali | PROMIS CS (46) | 702 | 684 | 31.1 | −0.05 (−0.12, 0.01) | 0.05 | 0.08 | 245 | 279 | 40.5 | −0.12 (−0.21, −0.04) | 0.06 | 0.08 |
| Zimbabwe | SHINE (HIV−) (47) | 1364 | 1305 | 35.9 | −0.09 (−0.13, −0.06) | 0.20 | 0.13 | 240 | 249 | 34.1 | −0.01 (−0.09, 0.07) | 0.07 | 0.08 |
| Zimbabwe | SHINE (HIV+) (48) | 247 | 254 | 48.8 | −0.12 (−0.21, −0.03) | 0.03 | 0.05 | 62 | 52 | 53.8 | −0.04 (−0.22, 0.15) | 0.01 | 0.02 |
| | | **7791** | **10098** | | **I² = 0.57, Tau² = 0.00** | | | **3852** | **5011** | | **I² = 0.26, Tau² = 0.00** | | |
| **Fixed** | | | | | **−0.07 (−0.08, −0.05)** | | | | | | **−0.05 (−0.07, −0.03)** | | |
| **Random** | | | | | **−0.06 (−0.09, −0.04)** | | | | | | **−0.04 (−0.07, −0.02)** | | |

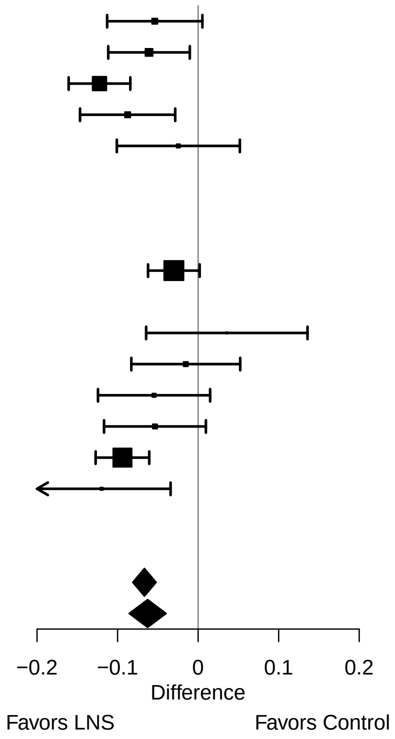

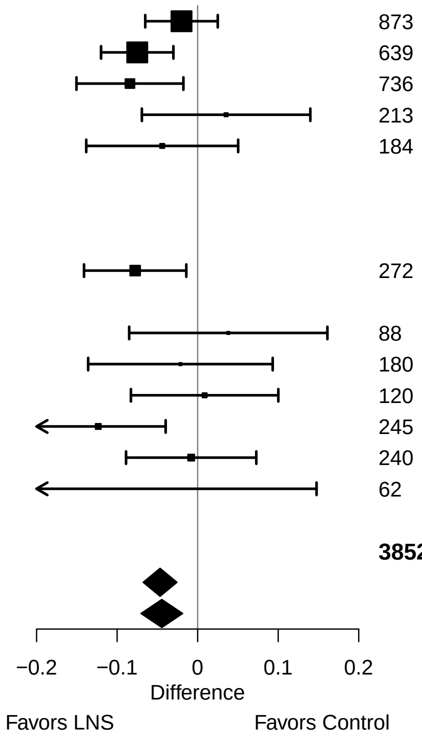

#### Supplemental figure 6C: Stunting prevalence difference

#### 6C3: Stratified by Maternal age

**P−for−interaction = 0.541**
**Difference in PDs = −0.01 (−0.03, 0.01)**

**At least 25 y**

| Country | Trial | LNS N | Control N | Control Prevalence | PD (95% CI) | Fixed W | Random W |
|---|---|---|---|---|---|---|---|
| Bangladesh | JiVitA−4 (34) | 1218 | 546 | 46.3 | −0.04 (−0.09, 0.00) | 0.10 | 0.09 |
| Bangladesh | RDNS (35) | 453 | 221 | 40.7 | −0.08 (−0.16, −0.01) | 0.04 | 0.05 |
| Bangladesh | WASH−B (36) | 492 | 1512 | 42.7 | −0.06 (−0.10, −0.02) | 0.11 | 0.09 |
| Burkina Faso | iLiNS−Zinc (37) | 1131 | 399 | 38.0 | −0.11 (−0.16, −0.06) | 0.08 | 0.08 |
| Burkina Faso | PROMIS (38) | 485 | 526 | 30.8 | −0.07 (−0.14, 0.00) | 0.04 | 0.05 |
| Burkina Faso | PROMIS CS (38) | 202 | 192 | 22.4 | −0.03 (−0.10, 0.04) | 0.04 | 0.05 |
| Ghana | GHANA (39) | | | | | | |
| Ghana | iLiNS−DYADG (40) | 220 | 416 | 11.1 | −0.01 (−0.06, 0.04) | 0.08 | 0.08 |
| Haiti | HAITI (41) | 101 | 103 | 9.7 | −0.01 (−0.08, 0.06) | 0.04 | 0.05 |
| Kenya | WASH−B (42) | 844 | 2811 | 30.1 | −0.03 (−0.07, 0.00) | 0.16 | 0.11 |
| Madagascar | MAHAY (43) | 896 | 949 | 59.0 | −0.02 (−0.07, 0.04) | 0.06 | 0.06 |
| Malawi | iLiNS−DYADM (44) | 106 | 226 | 38.9 | 0.02 (−0.10, 0.13) | 0.02 | 0.02 |
| Malawi | iLiNS−DOSE (45) | 368 | 134 | 41.8 | 0.03 (−0.05, 0.11) | 0.03 | 0.04 |
| Mali | PROMIS (46) | 346 | 359 | 36.8 | −0.05 (−0.11, 0.00) | 0.06 | 0.06 |
| Mali | PROMIS CS (46) | 503 | 507 | 31.0 | −0.06 (−0.13, 0.02) | 0.04 | 0.05 |
| Zimbabwe | SHINE (HIV−) (47) | 875 | 800 | 32.0 | −0.07 (−0.11, −0.02) | 0.10 | 0.09 |
| Zimbabwe | SHINE (HIV+) (48) | 244 | 231 | 50.2 | −0.11 (−0.20, −0.02) | 0.03 | 0.03 |
| | | **8484** | **9932** | | **I² = 0.30, Tau² = 0.00** | | |
| **Fixed** | | | | | **−0.05 (−0.06, −0.03)** | | |
| **Random** | | | | | **−0.05 (−0.06, −0.03)** | | |

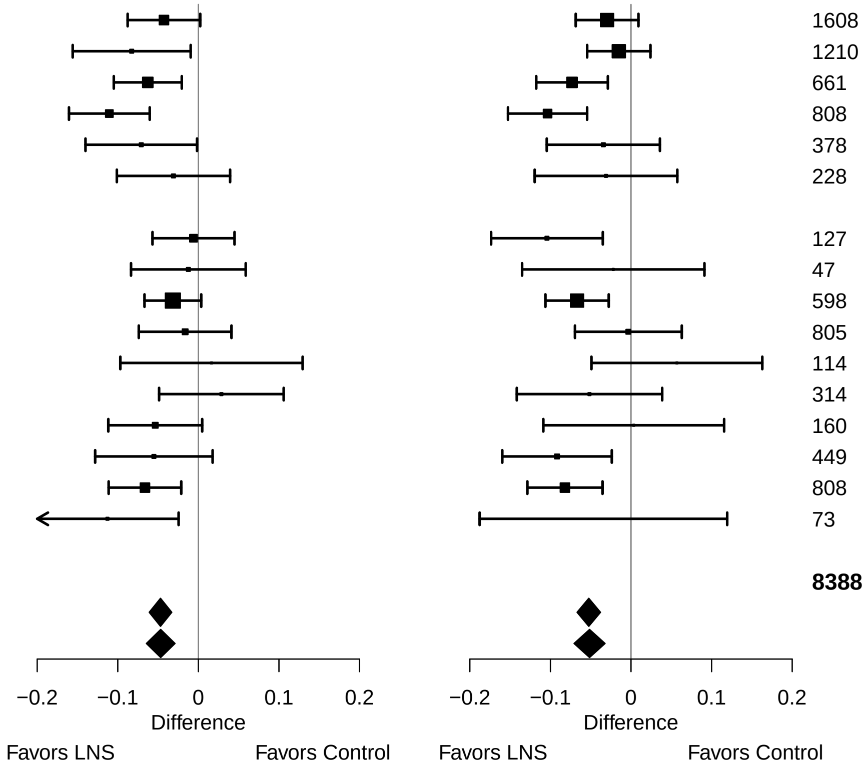

**Less than 25 y**

| Country | Trial | LNS N | Control N | Control Prevalence | PD (95% CI) | Fixed W | Random W |
|---|---|---|---|---|---|---|---|
| Bangladesh | JiVitA−4 (34) | 1608 | 693 | 42.6 | −0.03 (−0.07, 0.01) | 0.14 | 0.11 |
| Bangladesh | RDNS (35) | 1210 | 594 | 42.4 | −0.02 (−0.05, 0.02) | 0.14 | 0.11 |
| Bangladesh | WASH−B (36) | 661 | 1903 | 41.4 | −0.07 (−0.12, −0.03) | 0.11 | 0.10 |
| Burkina Faso | iLiNS−Zinc (37) | 808 | 262 | 41.6 | −0.10 (−0.15, −0.05) | 0.09 | 0.09 |
| Burkina Faso | PROMIS (38) | 378 | 388 | 30.2 | −0.03 (−0.10, 0.04) | 0.04 | 0.05 |
| Burkina Faso | PROMIS CS (38) | 228 | 247 | 25.9 | −0.03 (−0.12, 0.06) | 0.03 | 0.04 |
| Ghana | GHANA (39) | | | | | | |
| Ghana | iLiNS−DYADG (40) | 127 | 276 | 15.9 | −0.10 (−0.17, −0.04) | 0.04 | 0.06 |
| Haiti | HAITI (41) | 47 | 45 | 22.2 | −0.02 (−0.14, 0.09) | 0.02 | 0.03 |
| Kenya | WASH−B (42) | 598 | 2294 | 34.8 | −0.07 (−0.11, −0.03) | 0.14 | 0.11 |
| Madagascar | MAHAY (43) | 805 | 733 | 57.8 | 0.00 (−0.07, 0.06) | 0.05 | 0.06 |
| Malawi | iLiNS−DYADM (44) | 114 | 218 | 30.3 | 0.06 (−0.05, 0.16) | 0.02 | 0.03 |
| Malawi | iLiNS−DOSE (45) | 314 | 100 | 51.0 | −0.05 (−0.14, 0.04) | 0.03 | 0.04 |
| Mali | PROMIS (46) | 160 | 147 | 35.4 | 0.00 (−0.11, 0.12) | 0.02 | 0.03 |
| Mali | PROMIS CS (46) | 449 | 462 | 36.8 | −0.09 (−0.16, −0.02) | 0.05 | 0.06 |
| Zimbabwe | SHINE (HIV−) (47) | 808 | 797 | 38.5 | −0.08 (−0.13, −0.04) | 0.10 | 0.09 |
| Zimbabwe | SHINE (HIV+) (48) | 73 | 78 | 50.0 | −0.03 (−0.19, 0.12) | 0.01 | 0.01 |
| | | **8388** | **9237** | | **I² = 0.35, Tau² = 0.00** | | |
| **Fixed** | | | | | **−0.05 (−0.07, −0.04)** | | |
| **Random** | | | | | **−0.05 (−0.07, −0.03)** | | |

#### Supplemental figure 6C: Stunting prevalence difference

#### 6C4: Stratified by Maternal education

**P−for−interaction = 0.233**
**Difference in PDs = 0.02 (−0.01, 0.04)**

| | | Primary or greater | | | | | | Incomplete or no formal | | | | | |
|---|---|---|---|---|---|---|---|---|---|---|---|---|---|
| Country | Trial | LNS N | Control N | Control Prevalence | PD (95% CI) | Fixed W | Random W | LNS N | Control N | Control Prevalence | PD (95% CI) | Fixed W | Random W |
| Bangladesh | JiVitA−4 (34) | 1830 | 756 | 40.1 | −0.04 (−0.08, −0.01) | 0.16 | 0.16 | 1006 | 485 | 50.8 | −0.02 (−0.07, 0.03) | 0.11 | 0.09 |
| Bangladesh | RDNS (35) | 1245 | 589 | 40.2 | −0.05 (−0.09, 0.00) | 0.10 | 0.10 | 418 | 226 | 46.5 | 0.01 (−0.05, 0.08) | 0.06 | 0.07 |
| Bangladesh | WASH−B (36) | 813 | 2452 | 38.4 | −0.08 (−0.12, −0.05) | 0.18 | 0.18 | 345 | 979 | 50.7 | −0.04 (−0.10, 0.03) | 0.06 | 0.07 |
| Burkina Faso | iLiNS−Zinc (37) | 82 | 16 | 31.2 | −0.15 (−0.39, 0.09) | 0.00 | 0.00 | 1857 | 645 | 39.5 | −0.11 (−0.15, −0.06) | 0.12 | 0.10 |
| Burkina Faso | PROMIS (38) | 70 | 54 | 27.8 | 0.04 (−0.14, 0.21) | 0.01 | 0.01 | 793 | 860 | 30.7 | −0.06 (−0.12, −0.01) | 0.08 | 0.08 |
| Burkina Faso | PROMIS CS (38) | 34 | 39 | 25.6 | −0.08 (−0.23, 0.07) | 0.01 | 0.01 | 393 | 399 | 24.3 | −0.03 (−0.10, 0.05) | 0.04 | 0.06 |
| Ghana | GHANA (39) | | | | | | | | | | | | |
| Ghana | iLiNS−DYADG (40) | 267 | 548 | 13.7 | −0.06 (−0.11, −0.01) | 0.09 | 0.09 | 80 | 144 | 10.4 | 0.01 (−0.08, 0.09) | 0.03 | 0.05 |
| Haiti | HAITI (41) | | | | | | | | | | | | |
| Kenya | WASH−B (42) | 700 | 2434 | 26.0 | −0.02 (−0.06, 0.01) | 0.15 | 0.15 | 755 | 2699 | 37.8 | −0.07 (−0.10, −0.03) | 0.19 | 0.11 |
| Madagascar | MAHAY (43) | 351 | 431 | 56.8 | −0.02 (−0.10, 0.06) | 0.03 | 0.03 | 1351 | 1251 | 59.1 | −0.01 (−0.06, 0.05) | 0.08 | 0.08 |
| Malawi | iLiNS−DYADM (44) | 35 | 69 | 29.0 | 0.00 (−0.19, 0.18) | 0.01 | 0.01 | 184 | 372 | 35.8 | 0.04 (−0.04, 0.13) | 0.03 | 0.05 |
| Malawi | iLiNS−DOSE (45) | 162 | 56 | 44.6 | −0.10 (−0.21, 0.01) | 0.02 | 0.02 | 522 | 178 | 46.1 | 0.02 (−0.05, 0.09) | 0.05 | 0.07 |
| Mali | PROMIS (46) | 41 | 37 | 24.3 | −0.07 (−0.23, 0.09) | 0.01 | 0.01 | 464 | 469 | 37.3 | −0.03 (−0.09, 0.03) | 0.07 | 0.08 |
| Mali | PROMIS CS (46) | 107 | 96 | 32.3 | −0.10 (−0.23, 0.04) | 0.01 | 0.01 | 844 | 873 | 33.9 | −0.07 (−0.13, −0.01) | 0.07 | 0.08 |
| Zimbabwe | SHINE (HIV−) (47) | 1717 | 1644 | 34.8 | −0.07 (−0.10, −0.04) | 0.21 | 0.21 | 69 | 61 | 36.1 | −0.04 (−0.20, 0.11) | 0.01 | 0.02 |
| Zimbabwe | SHINE (HIV+) (48) | 301 | 293 | 48.5 | −0.08 (−0.16, 0.01) | 0.03 | 0.03 | 21 | 18 | 66.7 | −0.29 (−0.58, 0.01) | 0.00 | 0.01 |
| | | **7755** | **9514** | | **I² = 0.00, Tau² = 0.00** | | | **9102** | **9659** | | **I² = 0.49, Tau² = 0.00** | | |
| **Fixed** | | | | | **−0.06 (−0.07, −0.04)** | | | | | | **−0.04 (−0.06, −0.03)** | | |
| **Random** | | | | | **−0.06 (−0.07, −0.04)** | | | | | | **−0.03 (−0.06, −0.01)** | | |

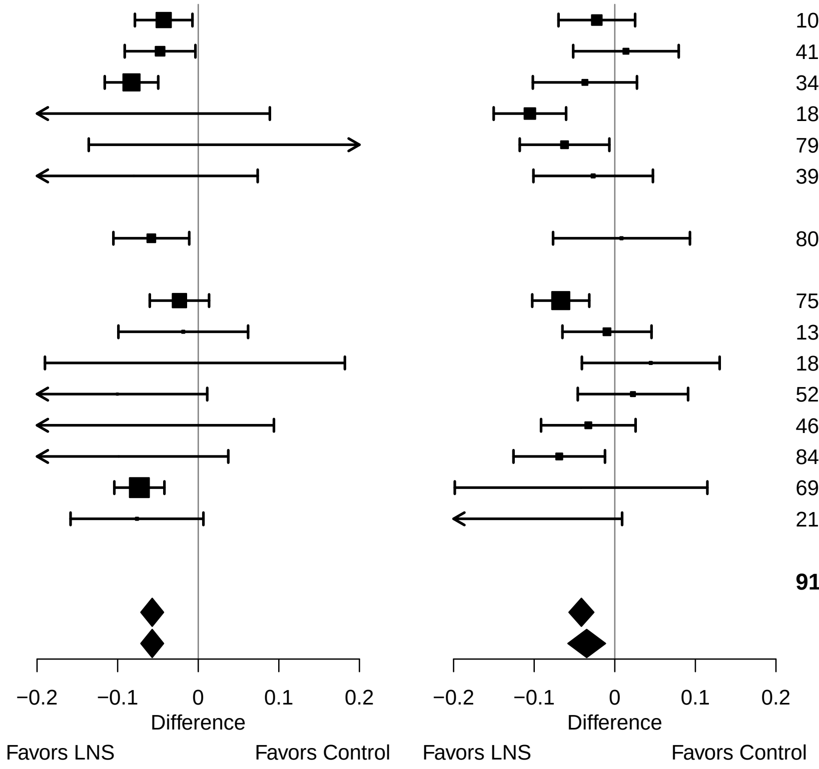

#### Supplemental figure 6C: Stunting prevalence difference

#### 6C5: Stratified by Maternal depressive symptoms

-for−interaction = 0.010
fference in PDs = −0.04 (−0.07, −0.01)

| | | | | | Less than 75th percentile | | | | | | At least 75th percentile | | |
|---|---|---|---|---|---|---|---|---|---|---|---|---|---|
| ountry | Trial | LNS N | Control N | Control Prevalence | PD (95% CI) | Fixed W | Random W | LNS N | Control N | Control Prevalence | PD (95% CI) | Fixed W | Random W |
| ngladesh | JiVitA−4 (34) | | | | | | | | | | | | |
| ngladesh | RDNS (35) | 1081 | 472 | 42.4 | −0.05 (−0.09, −0.01) | 0.15 | 0.15 | 520 | 288 | 41.0 | 0.00 (−0.08, 0.08) | 0.09 | 0.10 |
| ngladesh | WASH−B (36) | 897 | 2462 | 41.3 | −0.08 (−0.12, −0.05) | 0.19 | 0.19 | 237 | 878 | 43.8 | −0.02 (−0.10, 0.05) | 0.12 | 0.12 |
| rkina Faso | iLiNS−Zinc (37) | | | | | | | | | | | | |
| rkina Faso | PROMIS (38) | 508 | 573 | 30.7 | −0.07 (−0.15, 0.01) | 0.04 | 0.04 | 223 | 215 | 28.8 | 0.01 (−0.08, 0.09) | 0.09 | 0.11 |
| rkina Faso | PROMIS CS (38) | | | | | | | | | | | | |
| hana | GHANA (39) | | | | | | | | | | | | |
| hana | iLiNS−DYADG (40) | 252 | 459 | 13.3 | −0.07 (−0.11, −0.02) | 0.11 | 0.11 | 83 | 209 | 10.5 | 0.04 (−0.04, 0.12) | 0.10 | 0.11 |
| iti | HAITI (41) | | | | | | | | | | | | |
| nya | WASH−B (42) | 996 | 3496 | 31.1 | −0.04 (−0.08, −0.01) | 0.20 | 0.20 | 339 | 1235 | 34.3 | −0.02 (−0.08, 0.04) | 0.20 | 0.14 |
| adagascar | MAHAY (43) | 673 | 721 | 60.5 | −0.04 (−0.11, 0.03) | 0.05 | 0.05 | 284 | 252 | 60.3 | 0.05 (−0.04, 0.13) | 0.09 | 0.11 |
| alawi | iLiNS−DYADM (44) | 157 | 303 | 31.7 | 0.01 (−0.08, 0.10) | 0.03 | 0.03 | 47 | 113 | 39.8 | 0.18 (0.01, 0.34) | 0.02 | 0.04 |
| alawi | iLiNS−DOSE (45) | | | | | | | | | | | | |
| ali | PROMIS (46) | 379 | 373 | 35.1 | −0.04 (−0.10, 0.03) | 0.06 | 0.06 | 123 | 131 | 40.5 | −0.04 (−0.15, 0.06) | 0.06 | 0.09 |
| ali | PROMIS CS (46) | | | | | | | | | | | | |
| mbabwe | SHINE (HIV−) (47) | 1198 | 1145 | 33.8 | −0.06 (−0.10, −0.02) | 0.15 | 0.15 | 513 | 489 | 38.2 | −0.11 (−0.17, −0.05) | 0.20 | 0.14 |
| mbabwe | SHINE (HIV+) (48) | 234 | 215 | 49.3 | −0.12 (−0.22, −0.02) | 0.03 | 0.03 | 89 | 100 | 53.0 | −0.01 (−0.17, 0.14) | 0.03 | 0.05 |
| | | **6375** | **10219** | | **I² = 0.00, Tau² = 0.00** | | | **2458** | **3910** | | **I² = 0.56, Tau² = 0.00** | | |
| **xed** | | | | | **−0.06 (−0.07, −0.04)** | | | | | | **−0.02 (−0.04, 0.01)** | | |
| **andom** | | | | | **−0.06 (−0.07, −0.04)** | | | | | | **−0.01 (−0.05, 0.03)** | | |

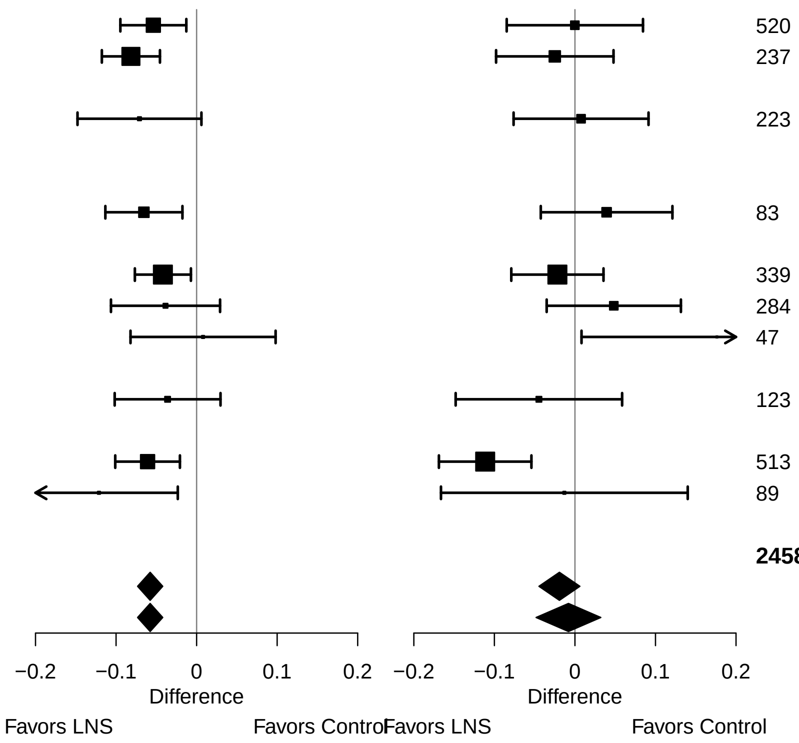

#### Supplemental figure 6C: Stunting prevalence difference

#### 6C6: Stratified by Child sex

**P−for−interaction = 0.187**
**Difference in PDs = −0.01 (−0.03, 0.01)**

**Male**

| Country | Trial | LNS N | Control N | Control Prevalence | PD (95% CI) | Fixed W | Random W |
|---|---|---|---|---|---|---|---|
| Bangladesh | JiVitA−4 (34) | 1448 | 600 | 44.5 | −0.03 (−0.07, 0.01) | 0.13 | 0.10 |
| Bangladesh | RDNS (35) | 833 | 411 | 42.6 | 0.00 (−0.06, 0.06) | 0.06 | 0.07 |
| Bangladesh | WASH−B (36) | 565 | 1714 | 43.1 | −0.07 (−0.11, −0.03) | 0.13 | 0.10 |
| Burkina Faso | iLiNS−Zinc (37) | 990 | 336 | 43.2 | −0.10 (−0.15, −0.04) | 0.07 | 0.08 |
| Burkina Faso | PROMIS (38) | 438 | 483 | 36.2 | −0.07 (−0.15, 0.01) | 0.03 | 0.04 |
| Burkina Faso | PROMIS CS (38) | 234 | 222 | 29.3 | −0.02 (−0.11, 0.08) | 0.02 | 0.03 |
| Ghana | GHANA (39) | | | | | | |
| Ghana | iLiNS−DYADG (40) | 175 | 323 | 16.7 | −0.04 (−0.11, 0.02) | 0.05 | 0.06 |
| Haiti | HAITI (41) | 70 | 59 | 18.6 | −0.05 (−0.15, 0.06) | 0.02 | 0.03 |
| Kenya | WASH−B (42) | 734 | 2440 | 36.8 | −0.03 (−0.07, 0.01) | 0.14 | 0.11 |
| Madagascar | MAHAY (43) | 826 | 832 | 63.3 | 0.01 (−0.04, 0.07) | 0.07 | 0.07 |
| Malawi | iLiNS−DYADM (44) | 107 | 207 | 38.6 | 0.04 (−0.07, 0.16) | 0.02 | 0.02 |
| Malawi | iLiNS−DOSE (45) | 352 | 124 | 54.8 | −0.02 (−0.10, 0.06) | 0.03 | 0.04 |
| Mali | PROMIS (46) | 266 | 263 | 38.8 | −0.02 (−0.09, 0.06) | 0.04 | 0.05 |
| Mali | PROMIS CS (46) | 477 | 525 | 36.6 | −0.06 (−0.13, 0.01) | 0.04 | 0.05 |
| Zimbabwe | SHINE (HIV−) (47) | 932 | 908 | 41.9 | −0.10 (−0.14, −0.06) | 0.12 | 0.10 |
| Zimbabwe | SHINE (HIV+) (48) | 162 | 169 | 53.8 | −0.08 (−0.18, 0.02) | 0.02 | 0.03 |
| | | **8609** | **9616** | | **I² = 0.37, Tau² = 0.00** | | |
| **Fixed** | | | | | **−0.05 (−0.06, −0.03)** | | |
| **Random** | | | | | **−0.04 (−0.06, −0.02)** | | |

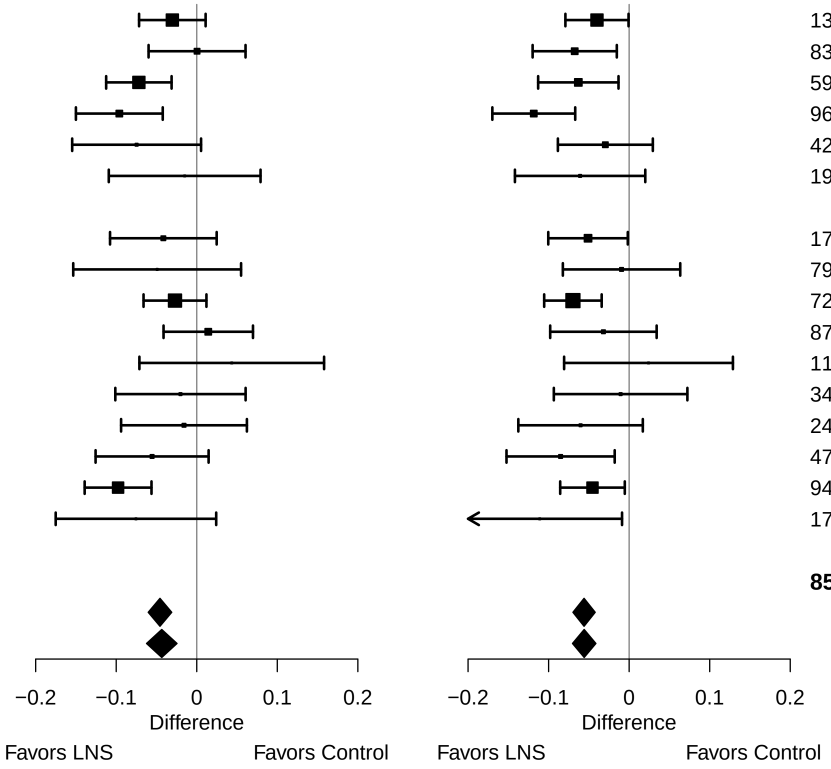

**Female**

| Country | Trial | LNS N | Control N | Control Prevalence | PD (95% CI) | Fixed W | Random W |
|---|---|---|---|---|---|---|---|
| Bangladesh | JiVitA−4 (34) | 1390 | 644 | 43.9 | −0.04 (−0.08, 0.00) | 0.13 | 0.12 |
| Bangladesh | RDNS (35) | 830 | 404 | 41.3 | −0.07 (−0.12, −0.02) | 0.07 | 0.07 |
| Bangladesh | WASH−B (36) | 593 | 1717 | 40.7 | −0.06 (−0.11, −0.01) | 0.08 | 0.08 |
| Burkina Faso | iLiNS−Zinc (37) | 962 | 328 | 35.6 | −0.12 (−0.17, −0.07) | 0.07 | 0.07 |
| Burkina Faso | PROMIS (38) | 425 | 431 | 24.1 | −0.03 (−0.09, 0.03) | 0.06 | 0.06 |
| Burkina Faso | PROMIS CS (38) | 196 | 217 | 19.4 | −0.06 (−0.14, 0.02) | 0.03 | 0.03 |
| Ghana | GHANA (39) | | | | | | |
| Ghana | iLiNS−DYADG (40) | 172 | 369 | 9.8 | −0.05 (−0.10, 0.00) | 0.08 | 0.08 |
| Haiti | HAITI (41) | 79 | 90 | 10.0 | −0.01 (−0.08, 0.06) | 0.04 | 0.04 |
| Kenya | WASH−B (42) | 723 | 2697 | 28.1 | −0.07 (−0.11, −0.03) | 0.15 | 0.14 |
| Madagascar | MAHAY (43) | 876 | 850 | 53.8 | −0.03 (−0.10, 0.03) | 0.04 | 0.05 |
| Malawi | iLiNS−DYADM (44) | 113 | 237 | 31.2 | 0.02 (−0.08, 0.13) | 0.02 | 0.02 |
| Malawi | iLiNS−DOSE (45) | 344 | 117 | 37.6 | −0.01 (−0.09, 0.07) | 0.03 | 0.03 |
| Mali | PROMIS (46) | 240 | 243 | 33.7 | −0.06 (−0.14, 0.02) | 0.03 | 0.03 |
| Mali | PROMIS CS (46) | 475 | 444 | 30.4 | −0.09 (−0.15, −0.02) | 0.04 | 0.04 |
| Zimbabwe | SHINE (HIV−) (47) | 948 | 886 | 27.7 | −0.05 (−0.09, −0.01) | 0.12 | 0.12 |
| Zimbabwe | SHINE (HIV+) (48) | 175 | 161 | 46.0 | −0.11 (−0.21, −0.01) | 0.02 | 0.02 |
| | | **8541** | **9835** | | **I² = 0.04, Tau² = 0.00** | | |
| **Fixed** | | | | | **−0.06 (−0.07, −0.04)** | | |
| **Random** | | | | | **−0.06 (−0.07, −0.04)** | | |

#### Supplemental figure 6C: Stunting prevalence difference

#### 6C7: Stratified by Child birth order

**P−for−interaction = 0.858**
**Difference in PDs = 0.00 (−0.02, 0.02)**

| | | Later born | | | | | | | Firstborn | | | | |
|---|---|---|---|---|---|---|---|---|---|---|---|---|---|
| **Country** | **Trial** | **LNS N** | **Control N** | **Control Prevalence** | **PD (95% CI)** | **Fixed W** | **Random W** | **LNS N** | **Control N** | **Control Prevalence** | **PD (95% CI)** | **Fixed W** | **Random W** |
| Bangladesh | JiVitA−4 (34) | 611 | 255 | 49.0 | −0.05 (−0.12, 0.01) | 0.04 | 0.05 | 2223 | 982 | 42.8 | −0.03 (−0.06, 0.00) | 0.28 | 0.12 |
| Bangladesh | RDNS (35) | 974 | 508 | 42.7 | −0.05 (−0.09, 0.00) | 0.08 | 0.08 | 688 | 307 | 40.7 | −0.01 (−0.07, 0.05) | 0.07 | 0.08 |
| Bangladesh | WASH−B (36) | 736 | 2290 | 43.4 | −0.07 (−0.11, −0.04) | 0.13 | 0.10 | 422 | 1141 | 38.8 | −0.06 (−0.11, 0.00) | 0.10 | 0.09 |
| Burkina Faso | iLiNS−Zinc (37) | 1520 | 519 | 38.8 | −0.11 (−0.16, −0.06) | 0.07 | 0.07 | 432 | 145 | 41.4 | −0.10 (−0.15, −0.04) | 0.10 | 0.09 |
| Burkina Faso | PROMIS (38) | 715 | 754 | 30.1 | −0.06 (−0.12, 0.00) | 0.04 | 0.05 | 148 | 158 | 32.3 | −0.04 (−0.15, 0.07) | 0.03 | 0.05 |
| Burkina Faso | PROMIS CS (38) | 366 | 359 | 23.7 | −0.03 (−0.10, 0.04) | 0.03 | 0.04 | 63 | 80 | 27.5 | −0.05 (−0.20, 0.10) | 0.01 | 0.03 |
| Ghana | GHANA (39) | | | | | | | | | | | | |
| Ghana | iLiNS−DYADG (40) | 237 | 458 | 13.3 | −0.03 (−0.08, 0.02) | 0.06 | 0.07 | 110 | 234 | 12.4 | −0.08 (−0.15, −0.01) | 0.07 | 0.08 |
| Haiti | HAITI (41) | 88 | 99 | 15.2 | −0.03 (−0.11, 0.06) | 0.02 | 0.03 | 61 | 50 | 10.0 | 0.00 (−0.09, 0.09) | 0.04 | 0.06 |
| Kenya | WASH−B (42) | 1134 | 4045 | 32.1 | −0.04 (−0.07, −0.01) | 0.19 | 0.12 | 321 | 1087 | 32.7 | −0.08 (−0.13, −0.02) | 0.10 | 0.09 |
| Madagascar | MAHAY (43) | 1272 | 1217 | 59.7 | −0.03 (−0.09, 0.02) | 0.05 | 0.06 | 430 | 465 | 55.3 | 0.05 (−0.03, 0.13) | 0.05 | 0.07 |
| Malawi | iLiNS−DYADM (44) | 173 | 353 | 35.7 | 0.04 (−0.05, 0.13) | 0.02 | 0.03 | 47 | 89 | 30.3 | 0.02 (−0.15, 0.18) | 0.01 | 0.02 |
| Malawi | iLiNS−DOSE (45) | 458 | 165 | 44.2 | 0.03 (−0.04, 0.10) | 0.03 | 0.04 | 149 | 45 | 53.3 | −0.11 (−0.24, 0.02) | 0.02 | 0.04 |
| Mali | PROMIS (46) | 442 | 445 | 37.1 | −0.06 (−0.11, 0.00) | 0.05 | 0.06 | 63 | 59 | 30.5 | 0.11 (−0.05, 0.26) | 0.01 | 0.03 |
| Mali | PROMIS CS (46) | 790 | 831 | 32.1 | −0.07 (−0.12, −0.01) | 0.05 | 0.06 | 145 | 126 | 42.9 | −0.12 (−0.23, −0.01) | 0.02 | 0.05 |
| Zimbabwe | SHINE (HIV−) (47) | 1302 | 1223 | 33.5 | −0.06 (−0.10, −0.03) | 0.12 | 0.10 | 491 | 487 | 37.8 | −0.08 (−0.14, −0.03) | 0.09 | 0.09 |
| Zimbabwe | SHINE (HIV+) (48) | 280 | 247 | 46.6 | −0.06 (−0.15, 0.02) | 0.02 | 0.03 | 54 | 78 | 59.0 | −0.20 (−0.37, −0.04) | 0.01 | 0.02 |
| | | **11098** | **13768** | | **I² = 0.26, Tau² = 0.00** | | | **5847** | **5533** | | **I² = 0.44, Tau² = 0.00** | | |
| **Fixed** | | | | | **−0.05 (−0.06, −0.04)** | | | | | | **−0.05 (−0.07, −0.03)** | | |
| **Random** | | | | | **−0.05 (−0.06, −0.03)** | | | | | | **−0.05 (−0.08, −0.02)** | | |

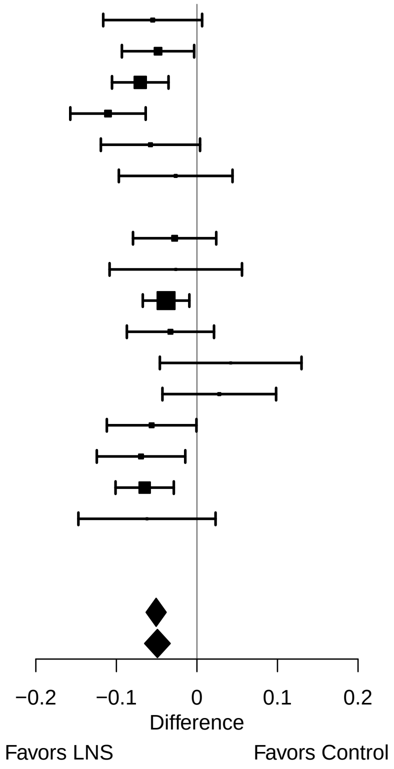

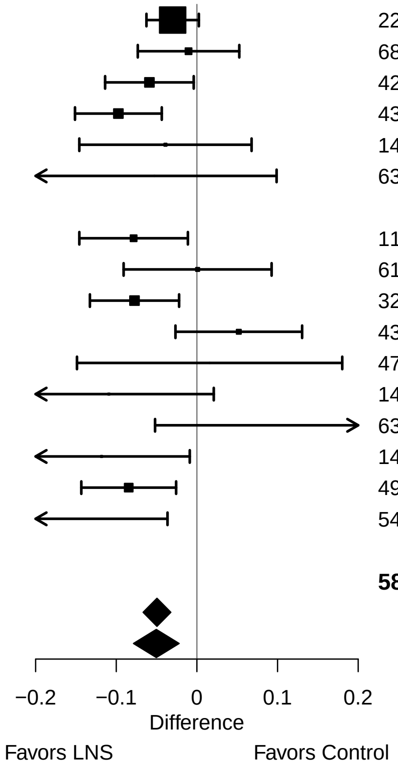

#### Supplemental figure 6C: Stunting prevalence difference

### 6C8: Stratified by Child baseline anthropometric status

**P−for−interaction = 0.004**
**Difference in PDs = −0.04 (−0.06, −0.01)**

**LAZ at least −1 Z**

| Country | Trial | LNS N | Control N | Control Prevalence | PD (95% CI) | Fixed W | Random W |
|---|---|---|---|---|---|---|---|
| Bangladesh | JiVitA−4 (34) | 1055 | 454 | 8.8 | −0.02 (−0.05, 0.01) | 0.19 | 0.19 |
| Bangladesh | RDNS (35) | 630 | 306 | 9.5 | −0.03 (−0.06, 0.01) | 0.16 | 0.16 |
| Bangladesh | WASH−B (36) | | | | | | |
| Burkina Faso | iLiNS−Zinc (37) | 839 | 291 | 6.2 | −0.04 (−0.07, 0.00) | 0.18 | 0.18 |
| Burkina Faso | PROMIS (38) | 444 | 484 | 12.4 | −0.04 (−0.08, 0.00) | 0.15 | 0.15 |
| Burkina Faso | PROMIS CS (38) | | | | | | |
| Ghana | GHANA (39) | | | | | | |
| Ghana | iLiNS−DYADG (40) | | | | | | |
| Haiti | HAITI (41) | | | | | | |
| Kenya | WASH−B (42) | | | | | | |
| Madagascar | MAHAY (43) | | | | | | |
| Malawi | iLiNS−DYADM (44) | 82 | 179 | 8.4 | 0.03 (−0.05, 0.10) | 0.04 | 0.04 |
| Malawi | iLiNS−DOSE (45) | 260 | 79 | 8.9 | 0.01 (−0.06, 0.09) | 0.04 | 0.04 |
| Mali | PROMIS (46) | 296 | 266 | 13.2 | −0.02 (−0.08, 0.05) | 0.05 | 0.05 |
| Mali | PROMIS CS (46) | | | | | | |
| Zimbabwe | SHINE (HIV−) (47) | 725 | 638 | 12.9 | −0.04 (−0.07, 0.00) | 0.17 | 0.17 |
| Zimbabwe | SHINE (HIV+) (48) | 103 | 107 | 25.2 | −0.13 (−0.23, −0.02) | 0.02 | 0.02 |
| | | **4434** | **2804** | | **I² = 0.00, Tau² = 0.00** | | |
| **Fixed** | | | | | **−0.03 (−0.04, −0.01)** | | |
| **Random** | | | | | **−0.03 (−0.04, −0.01)** | | |

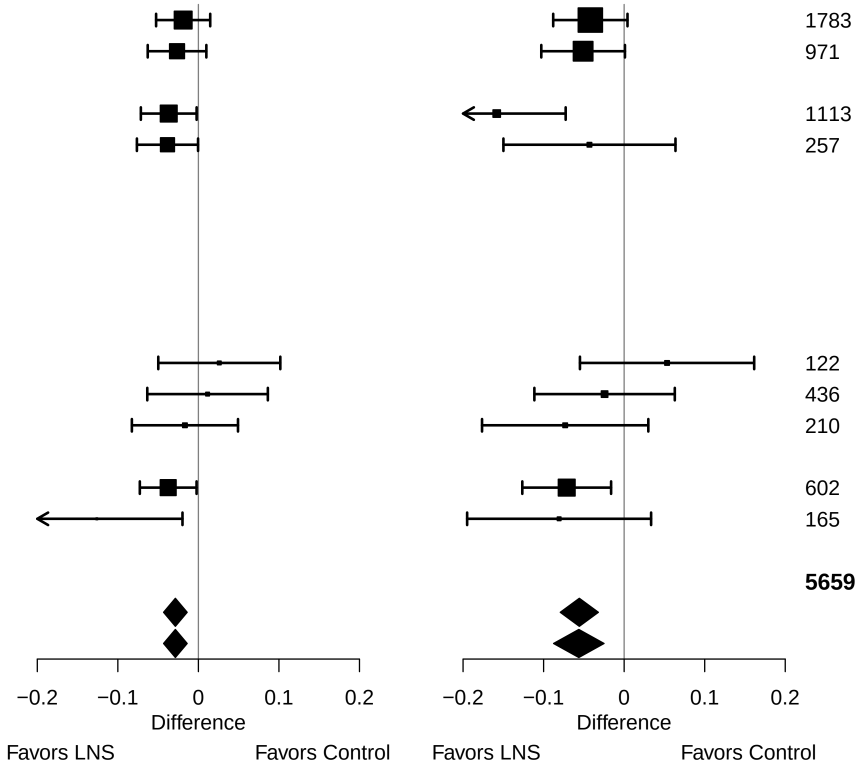

**LAZ less than −1 Z**

| Country | Trial | LNS N | Control N | Control Prevalence | PD (95% CI) | Fixed W | Random W |
|---|---|---|---|---|---|---|---|
| Bangladesh | JiVitA−4 (34) | 1783 | 790 | 64.9 | −0.04 (−0.09, 0.00) | 0.26 | 0.20 |
| Bangladesh | RDNS (35) | 971 | 453 | 63.8 | −0.05 (−0.10, 0.00) | 0.21 | 0.18 |
| Bangladesh | WASH−B (36) | | | | | | |
| Burkina Faso | iLiNS−Zinc (37) | 1113 | 373 | 65.4 | −0.16 (−0.24, −0.07) | 0.08 | 0.10 |
| Burkina Faso | PROMIS (38) | 257 | 274 | 58.4 | −0.04 (−0.15, 0.06) | 0.05 | 0.07 |
| Burkina Faso | PROMIS CS (38) | | | | | | |
| Ghana | GHANA (39) | | | | | | |
| Ghana | iLiNS−DYADG (40) | | | | | | |
| Haiti | HAITI (41) | | | | | | |
| Kenya | WASH−B (42) | | | | | | |
| Madagascar | MAHAY (43) | | | | | | |
| Malawi | iLiNS−DYADM (44) | 122 | 244 | 53.7 | 0.05 (−0.05, 0.16) | 0.05 | 0.07 |
| Malawi | iLiNS−DOSE (45) | 436 | 162 | 64.8 | −0.02 (−0.11, 0.06) | 0.07 | 0.09 |
| Mali | PROMIS (46) | 210 | 240 | 62.1 | −0.07 (−0.18, 0.03) | 0.05 | 0.07 |
| Mali | PROMIS CS (46) | | | | | | |
| Zimbabwe | SHINE (HIV−) (47) | 602 | 571 | 54.6 | −0.07 (−0.13, −0.02) | 0.18 | 0.17 |
| Zimbabwe | SHINE (HIV+) (48) | 165 | 131 | 65.6 | −0.08 (−0.19, 0.03) | 0.04 | 0.06 |
| | | **5659** | **3238** | | **I² = 0.27, Tau² = 0.00** | | |
| **Fixed** | | | | | **−0.06 (−0.08, −0.03)** | | |
| **Random** | | | | | **−0.06 (−0.09, −0.02)** | | |

### Supplemental figure 6D: Mean difference in WLZ

#### 6D1: Stratified by Maternal height

**P−for−interaction = 0.630**
**Difference in MDs = −0.02 (−0.09, 0.05)**

| Country | Trial | At least 150.1 cm: LNS N | Control N | Control Mean | MD (95% CI) | Fixed W | Random W | Less than 150.1 cm: LNS N | Control N | Control Mean | MD (95% CI) | Fixed W | Random W |
|---|---|---|---|---|---|---|---|---|---|---|---|---|---|
| Bangladesh | JiVitA−4 (34) | | | | | | | | | | | | |
| Bangladesh | RDNS (35) | 868 | 432 | −1.07 | 0.09 (−0.01, 0.18) | 0.08 | 0.08 | 733 | 358 | −1.17 | 0.09 (−0.04, 0.21) | 0.27 | 0.27 |
| Bangladesh | WASH−B (36) | 616 | 1880 | −0.80 | 0.13 (0.04, 0.22) | 0.09 | 0.08 | 540 | 1541 | −0.96 | 0.13 (0.02, 0.23) | 0.36 | 0.36 |
| Burkina Faso | iLiNS−Zinc (37) | 1916 | 649 | −0.95 | 0.19 (0.12, 0.26) | 0.14 | 0.09 | 34 | 15 | −0.73 | −0.32 (−0.73, 0.10) | 0.02 | 0.02 |
| Burkina Faso | PROMIS (38) | 828 | 877 | −0.70 | 0.10 (−0.02, 0.23) | 0.05 | 0.07 | 25 | 25 | −1.13 | 0.09 (−0.43, 0.61) | 0.02 | 0.02 |
| Burkina Faso | PROMIS CS (38) | 419 | 423 | −0.88 | −0.11 (−0.24, 0.03) | 0.04 | 0.06 | 11 | 9 | −1.14 | −0.12 (−1.06, 0.82) | 0.00 | 0.00 |
| Ghana | GHANA (39) | | | | | | | | | | | | |
| Ghana | iLiNS−DYADG (40) | 326 | 645 | −0.57 | 0.09 (−0.04, 0.23) | 0.04 | 0.06 | 16 | 36 | −0.73 | 0.06 (−0.50, 0.63) | 0.01 | 0.01 |
| Haiti | HAITI (41) | | | | | | | | | | | | |
| Kenya | WASH−B (42) | 1311 | 4668 | 0.10 | 0.06 (−0.01, 0.12) | 0.17 | 0.09 | 57 | 190 | −0.06 | 0.10 (−0.22, 0.41) | 0.04 | 0.04 |
| Madagascar | MAHAY (43) | 1002 | 1037 | −0.37 | −0.04 (−0.15, 0.07) | 0.06 | 0.07 | 598 | 565 | −0.52 | 0.05 (−0.11, 0.21) | 0.15 | 0.15 |
| Malawi | iLiNS−DYADM (44) | 192 | 381 | −0.12 | −0.03 (−0.20, 0.14) | 0.02 | 0.05 | 28 | 60 | −0.20 | −0.27 (−0.73, 0.19) | 0.02 | 0.02 |
| Malawi | iLiNS−DOSE (45) | 580 | 198 | −0.19 | −0.01 (−0.15, 0.12) | 0.04 | 0.06 | 115 | 42 | −0.40 | 0.14 (−0.22, 0.50) | 0.03 | 0.03 |
| Mali | PROMIS (46) | 474 | 492 | −0.33 | 0.11 (−0.01, 0.24) | 0.05 | 0.07 | 19 | 9 | −0.51 | −0.13 (−0.72, 0.46) | 0.01 | 0.01 |
| Mali | PROMIS CS (46) | 907 | 927 | −0.77 | 0.18 (0.08, 0.29) | 0.07 | 0.08 | 32 | 26 | −0.78 | −0.28 (−0.78, 0.23) | 0.02 | 0.02 |
| Zimbabwe | SHINE (HIV−) (47) | 1745 | 1688 | 0.01 | 0.09 (0.02, 0.17) | 0.14 | 0.09 | 75 | 62 | 0.21 | −0.26 (−0.60, 0.08) | 0.03 | 0.03 |
| Zimbabwe | SHINE (HIV+) (48) | 313 | 311 | 0.03 | −0.15 (−0.34, 0.04) | 0.02 | 0.04 | 18 | 12 | −0.31 | 0.05 (−0.69, 0.78) | 0.01 | 0.01 |
| | | **11497** | **14608** | | **I² = 0.64, Tau² = 0.01** | | | **2301** | **2950** | | **I² = 0.00, Tau² = 0.00** | | |
| **Fixed** | | | | | **0.08 (0.06, 0.11)** | | | | | | **0.06 (0.00, 0.12)** | | |
| **Random** | | | | | **0.06 (0.01, 0.11)** | | | | | | **0.06 (0.00, 0.12)** | | |

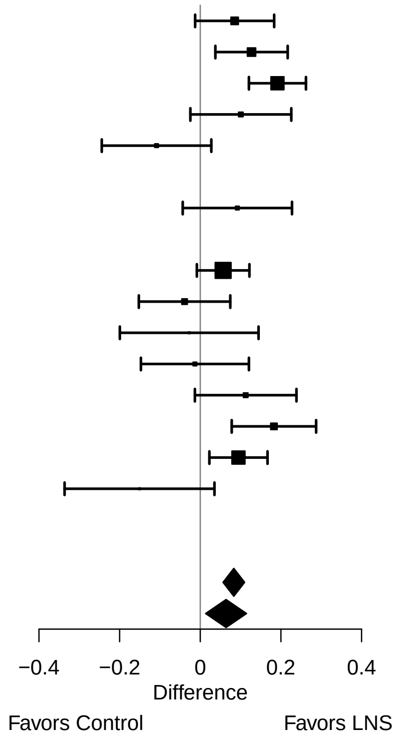

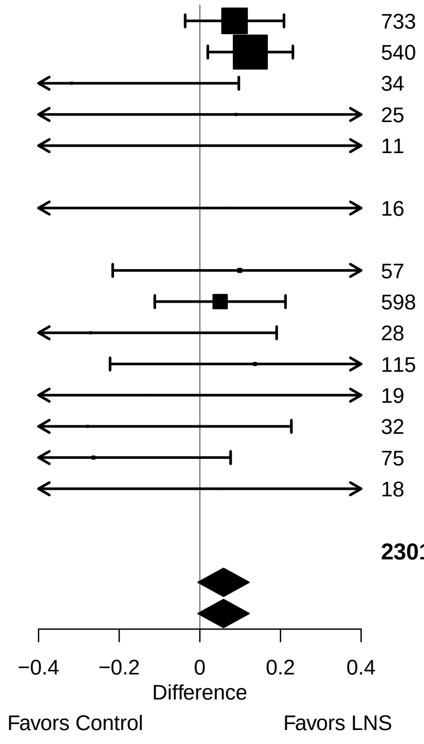

#### Supplemental figure 6D: Mean difference in WLZ

##### 6D2: Stratified by Maternal BMI

**P−for−interaction** = **0.849**

**Difference in MDs** = **0.00 (−0.05, 0.04)**

| | | At least 20 kg/m² | | | | | | Less than 20 kg/m² | | | | | |
|---|---|---|---|---|---|---|---|---|---|---|---|---|---|
| **Country** | **Trial** | **LNS N** | **Control N** | **Control Mean** | **MD (95% CI)** | **Fixed W** | **Random W** | **LNS N** | **Control N** | **Control Mean** | **MD (95% CI)** | **Fixed W** | **Random W** |
| Bangladesh | JiVitA−4 (34) | | | | | | | | | | | | |
| Bangladesh | RDNS (35) | 730 | 335 | −0.90 | 0.04 (−0.07, 0.14) | 0.08 | 0.09 | 871 | 455 | −1.28 | 0.10 (0.00, 0.21) | 0.14 | 0.14 |
| Bangladesh | WASH−B (36) | 517 | 1573 | −0.67 | 0.15 (0.05, 0.26) | 0.09 | 0.09 | 639 | 1848 | −1.05 | 0.11 (0.02, 0.19) | 0.22 | 0.22 |
| Burkina Faso | iLiNS−Zinc (37) | 1214 | 390 | −0.82 | 0.20 (0.12, 0.29) | 0.12 | 0.10 | 736 | 274 | −1.11 | 0.14 (0.06, 0.22) | 0.26 | 0.26 |
| Burkina Faso | PROMIS (38) | 644 | 669 | −0.62 | 0.11 (−0.02, 0.25) | 0.05 | 0.07 | 209 | 232 | −0.96 | 0.04 (−0.13, 0.20) | 0.05 | 0.05 |
| Burkina Faso | PROMIS CS (38) | 246 | 267 | −0.73 | −0.17 (−0.31, −0.03) | 0.05 | 0.07 | 184 | 165 | −1.13 | 0.01 (−0.17, 0.19) | 0.05 | 0.05 |
| Ghana | GHANA (39) | | | | | | | | | | | | |
| Ghana | iLiNS−DYADG (40) | 305 | 562 | −0.54 | 0.07 (−0.07, 0.21) | 0.05 | 0.07 | 37 | 119 | −0.79 | 0.13 (−0.23, 0.48) | 0.01 | 0.01 |
| Haiti | HAITI (41) | | | | | | | | | | | | |
| Kenya | WASH−B (42) | 1096 | 3788 | 0.17 | 0.03 (−0.03, 0.10) | 0.21 | 0.11 | 272 | 1070 | −0.19 | 0.13 (−0.02, 0.27) | 0.07 | 0.07 |
| Madagascar | MAHAY (43) | | | | | | | | | | | | |
| Malawi | iLiNS−DYADM (44) | 131 | 263 | 0.03 | −0.10 (−0.31, 0.11) | 0.02 | 0.04 | 88 | 178 | −0.37 | 0.01 (−0.22, 0.25) | 0.03 | 0.03 |
| Malawi | iLiNS−DOSE (45) | 513 | 174 | −0.09 | 0.01 (−0.14, 0.16) | 0.04 | 0.07 | 180 | 65 | −0.59 | 0.01 (−0.23, 0.25) | 0.03 | 0.03 |
| Mali | PROMIS (46) | 376 | 368 | −0.23 | 0.08 (−0.05, 0.21) | 0.05 | 0.07 | 117 | 133 | −0.61 | 0.15 (−0.05, 0.35) | 0.04 | 0.04 |
| Mali | PROMIS CS (46) | 696 | 675 | −0.70 | 0.18 (0.07, 0.30) | 0.07 | 0.08 | 243 | 278 | −0.96 | 0.09 (−0.10, 0.28) | 0.04 | 0.04 |
| Zimbabwe | SHINE (HIV−) (47) | 1355 | 1299 | 0.06 | 0.09 (0.02, 0.17) | 0.15 | 0.10 | 239 | 248 | −0.29 | 0.15 (−0.03, 0.33) | 0.05 | 0.05 |
| Zimbabwe | SHINE (HIV+) (48) | 246 | 253 | 0.08 | −0.06 (−0.26, 0.15) | 0.02 | 0.05 | 62 | 52 | −0.39 | −0.22 (−0.55, 0.12) | 0.01 | 0.01 |
| | | **8069** | **10616** | | **I² = 0.64, Tau² = 0.01** | | | **3877** | **5117** | | **I² = 0.00, Tau² = 0.00** | | |
| **Fixed** | | | | | **0.08 (0.05, 0.11)** | | | | | | **0.10 (0.06, 0.14)** | | |
| **Random** | | | | | **0.06 (0.01, 0.12)** | | | | | | **0.10 (0.06, 0.14)** | | |

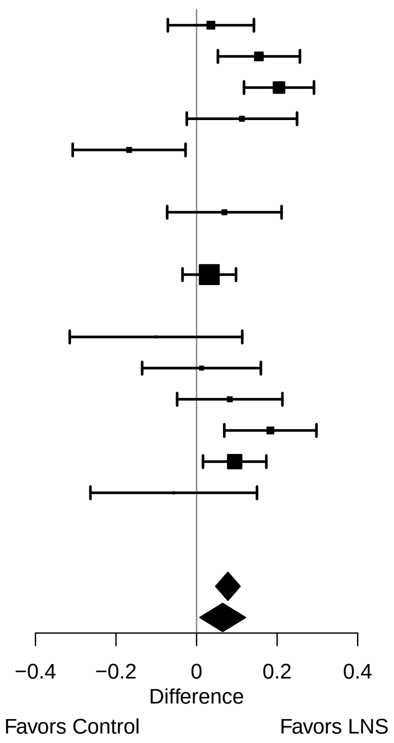

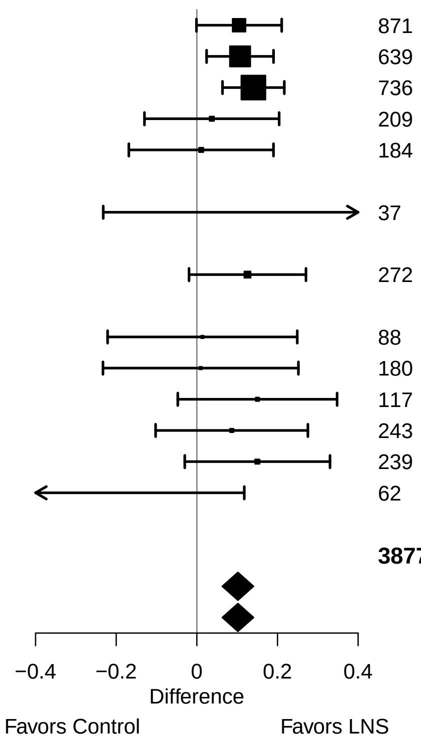

#### Supplemental figure 6D: Mean difference in WLZ

##### 6D3: Stratified by Maternal age

**P−for−interaction = 0.354**
**Difference in MDs = −0.02 (−0.06, 0.02)**

| | | At least 25 y | | | | | | Less than 25 y | | | | | |
|---|---|---|---|---|---|---|---|---|---|---|---|---|---|
| Country | Trial | LNS N | Control N | Control Mean | MD (95% CI) | Fixed W | Random W | LNS N | Control N | Control Mean | MD (95% CI) | Fixed W | Random W |
| Bangladesh | JiVitA−4 (34) | 1191 | 535 | −1.18 | 0.09 (0.02, 0.15) | 0.19 | 0.15 | 1580 | 677 | −1.16 | 0.07 (0.01, 0.14) | 0.22 | 0.11 |
| Bangladesh | RDNS (35) | 453 | 221 | −1.11 | 0.01 (−0.13, 0.16) | 0.04 | 0.05 | 1208 | 594 | −1.11 | 0.10 (0.01, 0.18) | 0.12 | 0.09 |
| Bangladesh | WASH−B (36) | 491 | 1508 | −0.92 | 0.05 (−0.05, 0.16) | 0.07 | 0.08 | 660 | 1900 | −0.84 | 0.17 (0.07, 0.26) | 0.11 | 0.09 |
| Burkina Faso | iLiNS−Zinc (37) | 1131 | 399 | −0.94 | 0.17 (0.10, 0.25) | 0.15 | 0.13 | 808 | 262 | −0.94 | 0.19 (0.06, 0.32) | 0.05 | 0.07 |
| Burkina Faso | PROMIS (38) | 480 | 522 | −0.74 | 0.13 (−0.01, 0.28) | 0.04 | 0.05 | 376 | 385 | −0.68 | 0.05 (−0.12, 0.23) | 0.03 | 0.05 |
| Burkina Faso | PROMIS CS (38) | 202 | 190 | −0.93 | −0.09 (−0.28, 0.11) | 0.02 | 0.03 | 228 | 246 | −0.85 | −0.13 (−0.30, 0.04) | 0.03 | 0.05 |
| Ghana | GHANA (39) | 69 | 63 | −0.72 | 0.37 (−0.04, 0.79) | 0.00 | 0.01 | 29 | 25 | −0.40 | −0.24 (−0.86, 0.37) | 0.00 | 0.01 |
| Ghana | iLiNS−DYADG (40) | 220 | 416 | −0.54 | 0.02 (−0.15, 0.19) | 0.03 | 0.04 | 127 | 276 | −0.66 | 0.21 (0.01, 0.41) | 0.02 | 0.04 |
| Haiti | HAITI (41) | 100 | 103 | −0.03 | 0.07 (−0.12, 0.25) | 0.03 | 0.03 | 47 | 45 | 0.04 | 0.12 (−0.16, 0.41) | 0.01 | 0.03 |
| Kenya | WASH−B (42) | 842 | 2801 | 0.09 | 0.09 (0.01, 0.17) | 0.14 | 0.12 | 598 | 2285 | 0.09 | 0.03 (−0.05, 0.11) | 0.14 | 0.10 |
| Madagascar | MAHAY (43) | 894 | 949 | −0.48 | 0.05 (−0.07, 0.17) | 0.06 | 0.07 | 805 | 733 | −0.34 | −0.07 (−0.19, 0.05) | 0.06 | 0.07 |
| Malawi | iLiNS−DYADM (44) | 106 | 226 | −0.11 | −0.04 (−0.27, 0.19) | 0.02 | 0.02 | 114 | 218 | −0.17 | −0.07 (−0.29, 0.16) | 0.02 | 0.04 |
| Malawi | iLiNS−DOSE (45) | 368 | 134 | −0.28 | 0.01 (−0.15, 0.18) | 0.03 | 0.04 | 314 | 100 | −0.17 | 0.02 (−0.18, 0.22) | 0.02 | 0.04 |
| Mali | PROMIS (46) | 341 | 355 | −0.29 | 0.08 (−0.07, 0.23) | 0.04 | 0.04 | 158 | 147 | −0.43 | 0.18 (−0.02, 0.37) | 0.02 | 0.04 |
| Mali | PROMIS CS (46) | 498 | 501 | −0.78 | 0.15 (0.02, 0.29) | 0.05 | 0.05 | 446 | 458 | −0.75 | 0.17 (0.04, 0.29) | 0.06 | 0.07 |
| Zimbabwe | SHINE (HIV−) (47) | 870 | 796 | −0.01 | 0.12 (0.02, 0.23) | 0.08 | 0.08 | 802 | 792 | 0.05 | 0.04 (−0.07, 0.15) | 0.08 | 0.08 |
| Zimbabwe | SHINE (HIV+) (48) | 243 | 231 | −0.04 | −0.11 (−0.33, 0.10) | 0.02 | 0.02 | 73 | 77 | 0.20 | −0.25 (−0.63, 0.14) | 0.01 | 0.02 |
| | | **8499** | **9950** | | **I² = 0.17, Tau² = 0.00** | | | **8373** | **9220** | | **I² = 0.47, Tau² = 0.01** | | |
| **Fixed** | | | | | **0.09 (0.06, 0.12)** | | | | | | **0.07 (0.04, 0.10)** | | |
| **Random** | | | | | **0.08 (0.05, 0.12)** | | | | | | **0.07 (0.02, 0.12)** | | |

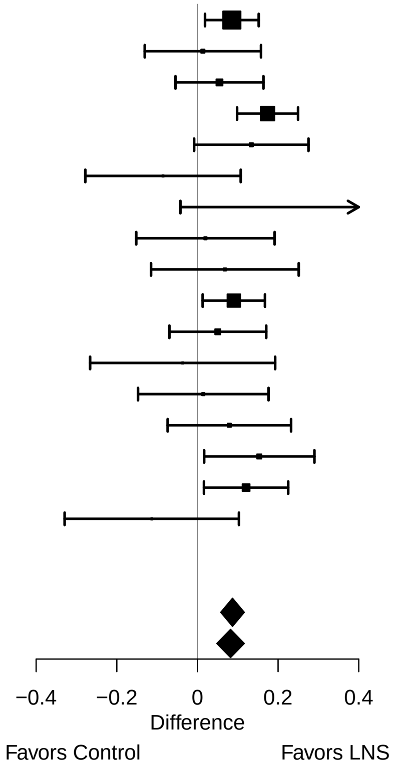

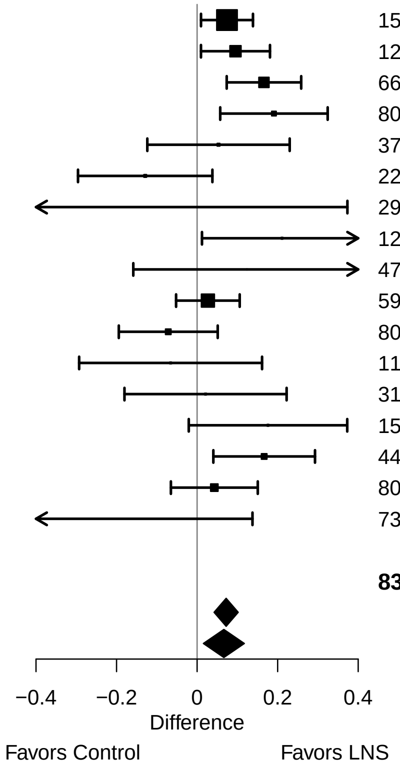

#### Supplemental figure 6D: Mean difference in WLZ

##### 6D4: Stratified by Maternal education

**P−for−interaction = 0.629**
**Difference in MDs = 0.01 (−0.04, 0.06)**

| | | Primary or greater | | | | | | Incomplete or no formal | | | | | |
|---|---|---|---|---|---|---|---|---|---|---|---|---|---|
| **Country** | **Trial** | **LNS N** | **Control N** | **Control Mean** | **MD (95% CI)** | **Fixed W** | **Random W** | **LNS N** | **Control N** | **Control Mean** | **MD (95% CI)** | **Fixed W** | **Random W** |
| Bangladesh | JiVitA−4 (34) | 1797 | 739 | −1.12 | 0.08 (0.02, 0.14) | 0.25 | 0.24 | 984 | 475 | −1.24 | 0.08 (0.01, 0.15) | 0.16 | 0.12 |
| Bangladesh | RDNS (35) | 1245 | 589 | −1.10 | 0.10 (0.00, 0.19) | 0.11 | 0.11 | 416 | 226 | −1.13 | 0.00 (−0.16, 0.17) | 0.03 | 0.05 |
| Bangladesh | WASH−B (36) | 813 | 2447 | −0.80 | 0.13 (0.05, 0.21) | 0.15 | 0.15 | 343 | 977 | −1.06 | 0.12 (0.00, 0.23) | 0.06 | 0.08 |
| Burkina Faso | iLiNS−Zinc (37) | 82 | 16 | −0.64 | −0.01 (−0.41, 0.39) | 0.01 | 0.01 | 1857 | 645 | −0.95 | 0.19 (0.12, 0.26) | 0.20 | 0.12 |
| Burkina Faso | PROMIS (38) | 70 | 54 | −0.77 | 0.08 (−0.19, 0.34) | 0.01 | 0.01 | 786 | 853 | −0.71 | 0.10 (−0.03, 0.23) | 0.05 | 0.07 |
| Burkina Faso | PROMIS CS (38) | 34 | 39 | −0.82 | −0.16 (−0.64, 0.32) | 0.00 | 0.00 | 393 | 396 | −0.89 | −0.10 (−0.23, 0.02) | 0.06 | 0.07 |
| Ghana | GHANA (39) | 91 | 75 | −0.68 | 0.25 (−0.12, 0.62) | 0.01 | 0.01 | 7 | 15 | −0.32 | −0.06 (−1.07, 0.95) | 0.00 | 0.00 |
| Ghana | iLiNS−DYADG (40) | 267 | 548 | −0.56 | 0.06 (−0.09, 0.21) | 0.04 | 0.04 | 80 | 144 | −0.69 | 0.20 (−0.06, 0.46) | 0.01 | 0.02 |
| Haiti | HAITI (41) | 129 | 126 | 0.09 | 0.12 (−0.04, 0.28) | 0.04 | 0.04 | 19 | 23 | −0.44 | −0.19 (−0.71, 0.32) | 0.00 | 0.01 |
| Kenya | WASH−B (42) | 700 | 2426 | 0.13 | 0.04 (−0.05, 0.12) | 0.12 | 0.12 | 753 | 2688 | 0.06 | 0.09 (0.01, 0.16) | 0.15 | 0.11 |
| Madagascar | MAHAY (43) | 351 | 431 | −0.20 | −0.01 (−0.17, 0.14) | 0.04 | 0.04 | 1349 | 1251 | −0.50 | 0.02 (−0.09, 0.14) | 0.07 | 0.08 |
| Malawi | iLiNS−DYADM (44) | 35 | 69 | −0.02 | −0.20 (−0.62, 0.22) | 0.01 | 0.01 | 184 | 372 | −0.16 | −0.03 (−0.21, 0.14) | 0.03 | 0.04 |
| Malawi | iLiNS−DOSE (45) | 162 | 56 | 0.01 | 0.02 (−0.25, 0.28) | 0.01 | 0.01 | 522 | 178 | −0.31 | 0.02 (−0.13, 0.16) | 0.04 | 0.06 |
| Mali | PROMIS (46) | 39 | 37 | −0.38 | −0.08 (−0.38, 0.21) | 0.01 | 0.01 | 459 | 465 | −0.33 | 0.12 (−0.02, 0.25) | 0.05 | 0.07 |
| Mali | PROMIS CS (46) | 107 | 96 | −0.69 | 0.31 (−0.02, 0.64) | 0.01 | 0.01 | 836 | 863 | −0.78 | 0.14 (0.03, 0.24) | 0.08 | 0.09 |
| Zimbabwe | SHINE (HIV−) (47) | 1707 | 1634 | 0.03 | 0.06 (−0.02, 0.13) | 0.16 | 0.16 | 68 | 61 | 0.01 | 0.15 (−0.23, 0.54) | 0.01 | 0.01 |
| Zimbabwe | SHINE (HIV+) (48) | 300 | 292 | 0.03 | −0.14 (−0.33, 0.05) | 0.02 | 0.03 | 21 | 18 | 0.00 | −0.34 (−0.93, 0.25) | 0.00 | 0.01 |
| | | **7929** | **9674** | | **I² = 0.03, Tau² = 0.00** | | | **9077** | **9650** | | **I² = 0.44, Tau² = 0.00** | | |
| **Fixed** | | | | | **0.07 (0.04, 0.10)** | | | | | | **0.09 (0.06, 0.12)** | | |
| **Random** | | | | | **0.07 (0.04, 0.10)** | | | | | | **0.08 (0.03, 0.12)** | | |

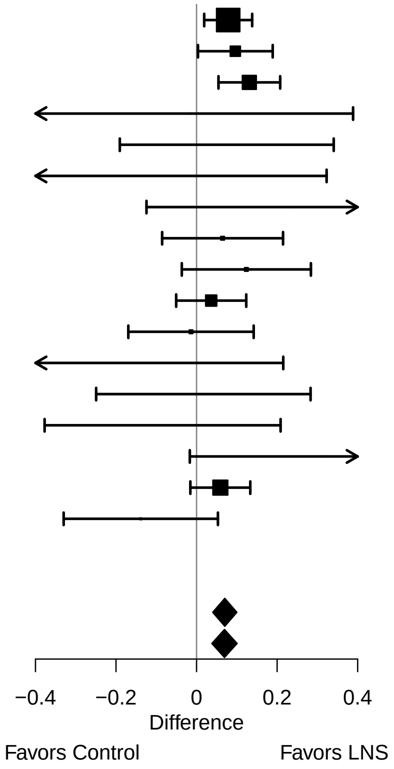

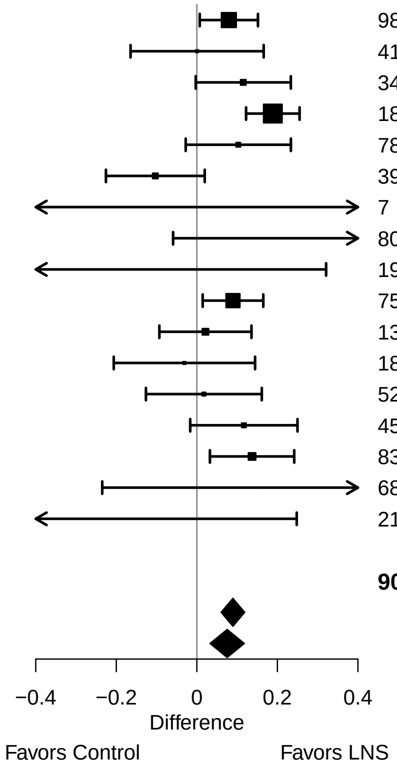

#### Supplemental figure 6D: Mean difference in WLZ

### 6D5: Stratified by Maternal depressive symptoms

**P−for−interaction = 0.863**
**Difference in MDs = −0.01 (−0.07, 0.05)**

| | | Less than 75th percentile | | | | | | At least 75th percentile | | | | | |
|---|---|---|---|---|---|---|---|---|---|---|---|---|---|
| **Country** | **Trial** | **LNS N** | **Control N** | **Control Mean** | **MD (95% CI)** | **Fixed W** | **Random W** | **LNS N** | **Control N** | **Control Mean** | **MD (95% CI)** | **Fixed W** | **Random W** |
| Bangladesh | JiVitA−4 (34) | | | | | | | | | | | | |
| Bangladesh | RDNS (35) | 1081 | 472 | −1.08 | 0.04 (−0.06, 0.14) | 0.12 | 0.12 | 519 | 288 | −1.14 | 0.09 (−0.04, 0.23) | 0.14 | 0.14 |
| Bangladesh | WASH−B (36) | 895 | 2457 | −0.86 | 0.13 (0.05, 0.21) | 0.19 | 0.17 | 237 | 876 | −0.90 | 0.10 (−0.04, 0.23) | 0.14 | 0.14 |
| Burkina Faso | iLiNS−Zinc (37) | | | | | | | | | | | | |
| Burkina Faso | PROMIS (38) | 505 | 569 | −0.75 | 0.15 (0.00, 0.30) | 0.05 | 0.06 | 219 | 212 | −0.74 | 0.06 (−0.09, 0.21) | 0.12 | 0.12 |
| Burkina Faso | PROMIS CS (38) | | | | | | | | | | | | |
| Ghana | GHANA (39) | | | | | | | | | | | | |
| Ghana | iLiNS−DYADG (40) | 252 | 459 | −0.55 | 0.08 (−0.07, 0.24) | 0.05 | 0.06 | 83 | 209 | −0.67 | 0.15 (−0.12, 0.41) | 0.04 | 0.04 |
| Haiti | HAITI (41) | | | | | | | | | | | | |
| Kenya | WASH−B (42) | 994 | 3480 | 0.10 | 0.07 (−0.01, 0.14) | 0.23 | 0.19 | 339 | 1233 | 0.09 | 0.04 (−0.07, 0.15) | 0.21 | 0.21 |
| Madagascar | MAHAY (43) | 672 | 721 | −0.45 | 0.08 (−0.06, 0.21) | 0.07 | 0.08 | 283 | 252 | −0.38 | −0.08 (−0.26, 0.10) | 0.08 | 0.08 |
| Malawi | iLiNS−DYADM (44) | 157 | 303 | −0.09 | −0.08 (−0.27, 0.11) | 0.03 | 0.04 | 47 | 113 | −0.26 | 0.12 (−0.22, 0.47) | 0.02 | 0.02 |
| Malawi | iLiNS−DOSE (45) | | | | | | | | | | | | |
| Mali | PROMIS (46) | 374 | 372 | −0.31 | 0.06 (−0.06, 0.18) | 0.08 | 0.09 | 121 | 129 | −0.42 | 0.24 (0.02, 0.45) | 0.06 | 0.06 |
| Mali | PROMIS CS (46) | | | | | | | | | | | | |
| Zimbabwe | SHINE (HIV−) (47) | 1192 | 1136 | 0.02 | 0.09 (0.00, 0.17) | 0.16 | 0.16 | 509 | 489 | −0.02 | 0.13 (0.00, 0.26) | 0.16 | 0.16 |
| Zimbabwe | SHINE (HIV+) (48) | 234 | 214 | 0.04 | −0.18 (−0.41, 0.05) | 0.02 | 0.03 | 88 | 100 | −0.07 | −0.08 (−0.38, 0.23) | 0.03 | 0.03 |
| | | **6356** | **10183** | | **I² = 0.13, Tau² = 0.00** | | | **2445** | **3901** | | **I² = 0.00, Tau² = 0.00** | | |
| **Fixed** | | | | | **0.07 (0.04, 0.11)** | | | | | | **0.08 (0.02, 0.13)** | | |
| **Random** | | | | | **0.07 (0.03, 0.11)** | | | | | | **0.08 (0.02, 0.13)** | | |

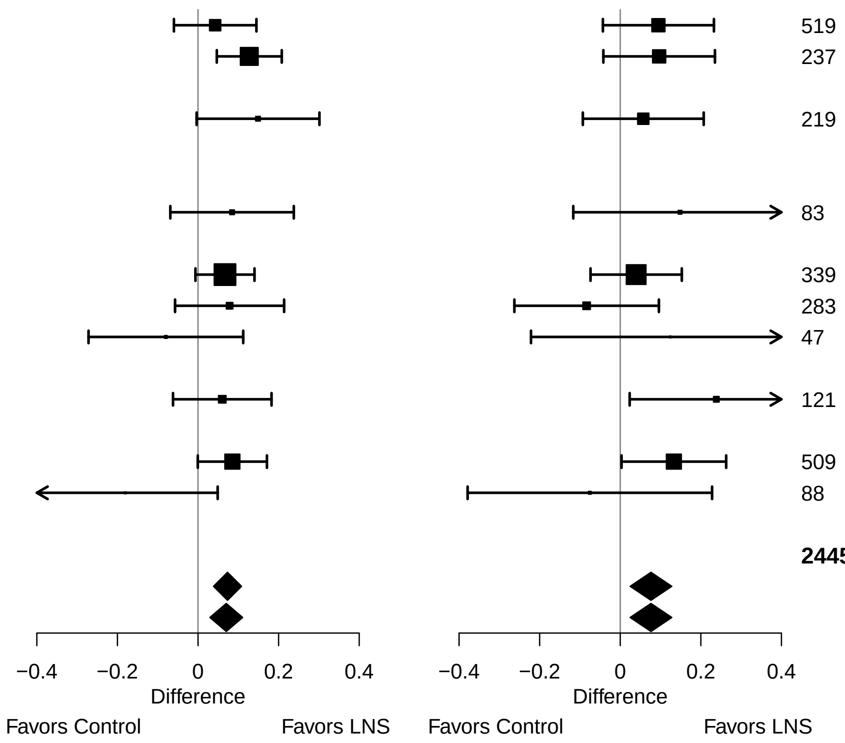

#### Supplemental figure 6D: Mean difference in WLZ

##### 6D6: Stratified by Child sex

**P−for−interaction = 0.647**
**Difference in MDs = 0.01 (−0.03, 0.05)**

**Male**

| Country | Trial | LNS N | Control N | Control Mean | MD (95% CI) | Fixed W | Random W |
|---|---|---|---|---|---|---|---|
| Bangladesh | JiVitA−4 (34) | 1420 | 586 | −1.16 | 0.07 (0.00, 0.14) | 0.21 | 0.12 |
| Bangladesh | RDNS (35) | 832 | 411 | −1.08 | 0.00 (−0.12, 0.12) | 0.07 | 0.07 |
| Bangladesh | WASH−B (36) | 564 | 1711 | −0.88 | 0.13 (0.03, 0.23) | 0.10 | 0.09 |
| Burkina Faso | iLiNS−Zinc (37) | 990 | 336 | −1.04 | 0.21 (0.10, 0.32) | 0.08 | 0.08 |
| Burkina Faso | PROMIS (38) | 434 | 480 | −0.82 | 0.17 (0.01, 0.32) | 0.04 | 0.06 |
| Burkina Faso | PROMIS CS (38) | 234 | 221 | −0.89 | −0.17 (−0.36, 0.02) | 0.03 | 0.04 |
| Ghana | GHANA (39) | 60 | 41 | −0.69 | 0.13 (−0.31, 0.56) | 0.01 | 0.01 |
| Ghana | iLiNS−DYADG (40) | 175 | 323 | −0.69 | 0.09 (−0.10, 0.28) | 0.03 | 0.04 |
| Haiti | HAITI (41) | 70 | 59 | 0.11 | −0.06 (−0.29, 0.18) | 0.02 | 0.03 |
| Kenya | WASH−B (42) | 732 | 2431 | 0.07 | 0.05 (−0.03, 0.14) | 0.14 | 0.10 |
| Madagascar | MAHAY (43) | 824 | 832 | −0.53 | 0.01 (−0.13, 0.14) | 0.05 | 0.07 |
| Malawi | iLiNS−DYADM (44) | 107 | 207 | −0.15 | −0.09 (−0.33, 0.15) | 0.02 | 0.03 |
| Malawi | iLiNS−DOSE (45) | 352 | 124 | −0.37 | −0.01 (−0.19, 0.17) | 0.03 | 0.04 |
| Mali | PROMIS (46) | 264 | 262 | −0.36 | 0.13 (−0.02, 0.28) | 0.04 | 0.06 |
| Mali | PROMIS CS (46) | 472 | 520 | −0.81 | 0.14 (−0.02, 0.30) | 0.04 | 0.05 |
| Zimbabwe | SHINE (HIV−) (47) | 924 | 905 | −0.04 | 0.09 (−0.01, 0.19) | 0.09 | 0.09 |
| Zimbabwe | SHINE (HIV+) (48) | 161 | 167 | −0.12 | −0.08 (−0.32, 0.16) | 0.02 | 0.03 |
| | | **8615** | **9616** | | **I² = 0.35, Tau² = 0.00** | | |
| **Fixed** | | | | | **0.07 (0.04, 0.10)** | | |
| **Random** | | | | | **0.07 (0.02, 0.11)** | | |

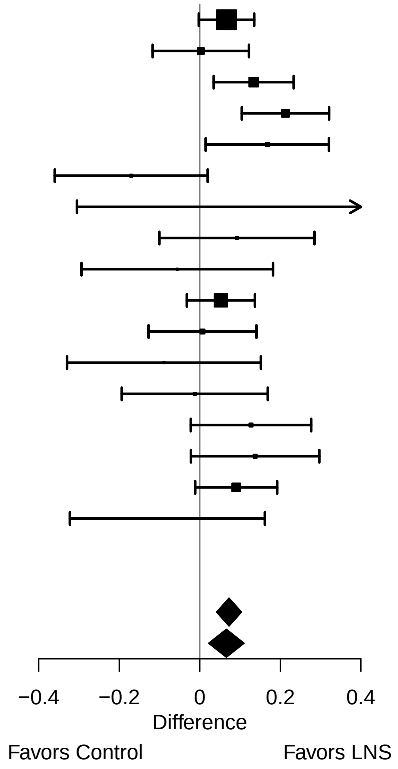

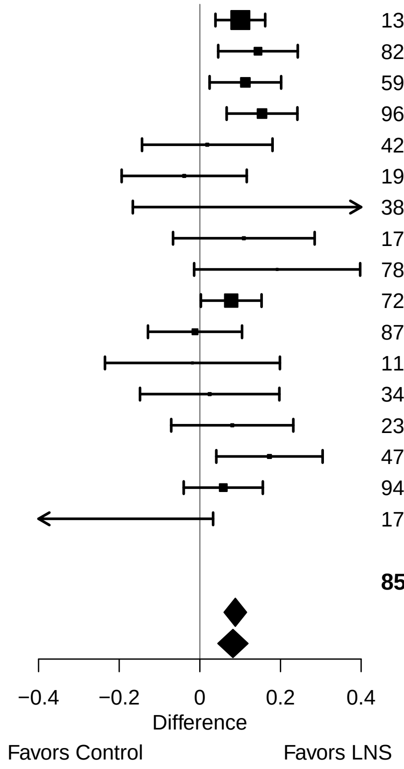

**Female**

| Country | Trial | LNS N | Control N | Control Mean | MD (95% CI) | Fixed W | Random W |
|---|---|---|---|---|---|---|---|
| Bangladesh | JiVitA−4 (34) | 1363 | 631 | −1.18 | 0.10 (0.04, 0.16) | 0.20 | 0.13 |
| Bangladesh | RDNS (35) | 829 | 404 | −1.14 | 0.14 (0.05, 0.24) | 0.08 | 0.08 |
| Bangladesh | WASH−B (36) | 592 | 1713 | −0.86 | 0.11 (0.02, 0.20) | 0.10 | 0.09 |
| Burkina Faso | iLiNS−Zinc (37) | 962 | 328 | −0.85 | 0.15 (0.07, 0.24) | 0.10 | 0.09 |
| Burkina Faso | PROMIS (38) | 422 | 427 | −0.59 | 0.02 (−0.14, 0.18) | 0.03 | 0.04 |
| Burkina Faso | PROMIS CS (38) | 196 | 215 | −0.87 | −0.04 (−0.19, 0.12) | 0.03 | 0.04 |
| Ghana | GHANA (39) | 38 | 55 | −0.61 | 0.37 (−0.17, 0.90) | 0.00 | 0.00 |
| Ghana | iLiNS−DYADG (40) | 172 | 369 | −0.50 | 0.11 (−0.07, 0.28) | 0.03 | 0.04 |
| Haiti | HAITI (41) | 78 | 90 | −0.06 | 0.19 (−0.01, 0.40) | 0.02 | 0.03 |
| Kenya | WASH−B (42) | 723 | 2687 | 0.11 | 0.08 (0.00, 0.15) | 0.14 | 0.11 |
| Madagascar | MAHAY (43) | 876 | 850 | −0.31 | −0.01 (−0.13, 0.10) | 0.06 | 0.07 |
| Malawi | iLiNS−DYADM (44) | 113 | 237 | −0.12 | −0.02 (−0.24, 0.20) | 0.02 | 0.03 |
| Malawi | iLiNS−DOSE (45) | 344 | 117 | −0.08 | 0.02 (−0.15, 0.20) | 0.03 | 0.04 |
| Mali | PROMIS (46) | 235 | 240 | −0.30 | 0.08 (−0.07, 0.23) | 0.03 | 0.05 |
| Mali | PROMIS CS (46) | 472 | 439 | −0.71 | 0.17 (0.04, 0.30) | 0.04 | 0.06 |
| Zimbabwe | SHINE (HIV−) (47) | 945 | 879 | 0.08 | 0.06 (−0.04, 0.16) | 0.08 | 0.08 |
| Zimbabwe | SHINE (HIV+) (48) | 175 | 161 | 0.14 | −0.22 (−0.46, 0.03) | 0.01 | 0.02 |
| | | **8535** | **9842** | | **I² = 0.25, Tau² = 0.00** | | |
| **Fixed** | | | | | **0.09 (0.06, 0.12)** | | |
| **Random** | | | | | **0.08 (0.04, 0.12)** | | |

#### Supplemental figure 6D: Mean difference in WLZ

##### 6D7: Stratified by Child birth order

**P−for−interaction = 0.250**
**Difference in MDs = −0.03 (−0.07, 0.02)**

| | | Later born | | | | | | Firstborn | | | | | |
|---|---|---|---|---|---|---|---|---|---|---|---|---|---|
| **Country** | **Trial** | **LNS N** | **Control N** | **Control Mean** | **MD (95% CI)** | **Fixed W** | **Random W** | **LNS N** | **Control N** | **Control Mean** | **MD (95% CI)** | **Fixed W** | **Random W** |
| Bangladesh | JiVitA−4 (34) | 598 | 247 | −1.24 | 0.11 (0.02, 0.21) | 0.08 | 0.07 | 2181 | 964 | −1.15 | 0.07 (0.02, 0.13) | 0.41 | 0.20 |
| Bangladesh | RDNS (35) | 972 | 508 | −1.13 | 0.08 (−0.01, 0.17) | 0.09 | 0.08 | 688 | 307 | −1.08 | 0.06 (−0.04, 0.17) | 0.09 | 0.11 |
| Bangladesh | WASH−B (36) | 735 | 2285 | −0.92 | 0.11 (0.03, 0.20) | 0.11 | 0.08 | 421 | 1139 | −0.78 | 0.13 (0.01, 0.24) | 0.08 | 0.10 |
| Burkina Faso | iLiNS−Zinc (37) | 1520 | 519 | −0.94 | 0.19 (0.12, 0.26) | 0.14 | 0.08 | 432 | 145 | −0.95 | 0.16 (0.01, 0.32) | 0.05 | 0.07 |
| Burkina Faso | PROMIS (38) | 708 | 747 | −0.73 | 0.12 (0.00, 0.25) | 0.05 | 0.06 | 148 | 158 | −0.65 | −0.02 (−0.24, 0.21) | 0.02 | 0.04 |
| Burkina Faso | PROMIS CS (38) | 366 | 357 | −0.87 | −0.13 (−0.28, 0.02) | 0.03 | 0.05 | 63 | 79 | −0.92 | −0.03 (−0.27, 0.21) | 0.02 | 0.03 |
| Ghana | GHANA (39) | 60 | 54 | −0.64 | 0.27 (−0.21, 0.75) | 0.00 | 0.01 | 38 | 38 | −0.59 | 0.06 (−0.41, 0.53) | 0.00 | 0.01 |
| Ghana | iLiNS−DYADG (40) | 237 | 458 | −0.56 | 0.02 (−0.14, 0.18) | 0.03 | 0.05 | 110 | 234 | −0.65 | 0.24 (0.02, 0.47) | 0.02 | 0.04 |
| Haiti | HAITI (41) | 88 | 99 | −0.05 | 0.16 (−0.04, 0.36) | 0.02 | 0.04 | 60 | 50 | 0.13 | −0.03 (−0.28, 0.22) | 0.02 | 0.03 |
| Kenya | WASH−B (42) | 1132 | 4027 | 0.08 | 0.07 (−0.01, 0.14) | 0.12 | 0.08 | 321 | 1086 | 0.12 | 0.06 (−0.04, 0.15) | 0.12 | 0.12 |
| Madagascar | MAHAY (43) | 1270 | 1217 | −0.46 | 0.02 (−0.10, 0.14) | 0.05 | 0.06 | 430 | 465 | −0.32 | −0.05 (−0.21, 0.11) | 0.04 | 0.06 |
| Malawi | iLiNS−DYADM (44) | 173 | 353 | −0.12 | −0.05 (−0.23, 0.12) | 0.02 | 0.04 | 47 | 89 | −0.20 | −0.06 (−0.45, 0.33) | 0.01 | 0.01 |
| Malawi | iLiNS−DOSE (45) | 458 | 165 | −0.29 | 0.06 (−0.09, 0.21) | 0.03 | 0.05 | 149 | 45 | −0.11 | −0.06 (−0.37, 0.25) | 0.01 | 0.02 |
| Mali | PROMIS (46) | 436 | 441 | −0.32 | 0.10 (−0.03, 0.24) | 0.04 | 0.06 | 62 | 59 | −0.43 | 0.07 (−0.17, 0.31) | 0.02 | 0.03 |
| Mali | PROMIS CS (46) | 783 | 821 | −0.76 | 0.14 (0.03, 0.24) | 0.07 | 0.07 | 144 | 126 | −0.81 | 0.32 (0.12, 0.52) | 0.03 | 0.04 |
| Zimbabwe | SHINE (HIV−) (47) | 1295 | 1218 | −0.02 | 0.15 (0.07, 0.23) | 0.12 | 0.08 | 487 | 483 | 0.10 | −0.09 (−0.23, 0.06) | 0.05 | 0.08 |
| Zimbabwe | SHINE (HIV+) (48) | 279 | 245 | 0.07 | −0.22 (−0.43, −0.02) | 0.02 | 0.03 | 54 | 78 | −0.17 | 0.16 (−0.19, 0.51) | 0.01 | 0.02 |
| | | **11110** | **13761** | | **$I^2$ = 0.54, $Tau^2$ = 0.01** | | | **5835** | **5545** | | **$I^2$ = 0.21, $Tau^2$ = 0.00** | | |
| **Fixed** | | | | | **0.10 (0.07, 0.12)** | | | | | | **0.07 (0.03, 0.10)** | | |
| **Random** | | | | | **0.08 (0.03, 0.12)** | | | | | | **0.07 (0.02, 0.11)** | | |

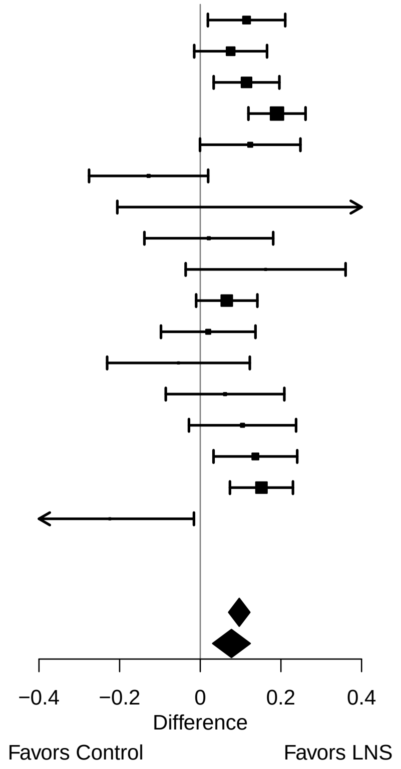

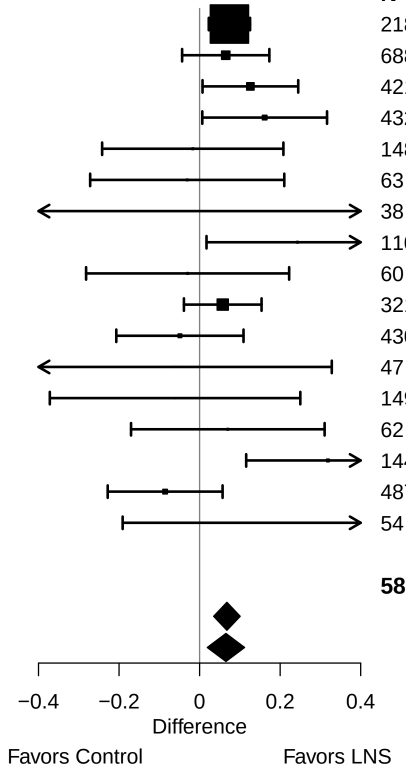

#### Supplemental figure 6D: Mean difference in WLZ

##### 6D8: Stratified by Child baseline anthropometric status

**P−for−interaction = 0.325**
**Difference in MDs = 0.03 (−0.03, 0.09)**

| Country | Trial | WLZ at least 0 Z: LNS N | Control N | Control Mean | MD (95% CI) | Fixed W | Random W | WLZ less than 0 Z: LNS N | Control N | Control Mean | MD (95% CI) | Fixed W | Random W |
|---|---|---|---|---|---|---|---|---|---|---|---|---|---|
| Bangladesh | JiVitA−4 (34) | 961 | 423 | −0.51 | 0.10 (0.01, 0.19) | 0.26 | 0.26 | 1822 | 794 | −1.51 | 0.08 (0.02, 0.15) | 0.31 | 0.14 |
| Bangladesh | RDNS (35) | 592 | 270 | −0.45 | −0.01 (−0.13, 0.11) | 0.14 | 0.14 | 1008 | 489 | −1.46 | 0.08 (−0.01, 0.18) | 0.15 | 0.13 |
| Bangladesh | WASH−B (36) | | | | | | | | | | | | |
| Burkina Faso | iLiNS−Zinc (37) | 349 | 103 | 0.17 | 0.07 (−0.17, 0.30) | 0.04 | 0.04 | 1603 | 561 | −1.15 | 0.22 (0.14, 0.30) | 0.20 | 0.13 |
| Burkina Faso | PROMIS (38) | 279 | 350 | −0.22 | 0.12 (−0.05, 0.30) | 0.07 | 0.07 | 413 | 393 | −1.15 | 0.15 (0.00, 0.30) | 0.06 | 0.09 |
| Burkina Faso | PROMIS CS (38) | | | | | | | | | | | | |
| Ghana | GHANA (39) | | | | | | | | | | | | |
| Ghana | iLiNS−DYADG (40) | 154 | 310 | 0.02 | 0.11 (−0.06, 0.27) | 0.07 | 0.07 | 169 | 328 | −1.14 | 0.10 (−0.05, 0.25) | 0.05 | 0.09 |
| Haiti | HAITI (41) | 76 | 76 | 0.66 | −0.10 (−0.37, 0.16) | 0.03 | 0.03 | 72 | 73 | −0.67 | 0.24 (0.00, 0.48) | 0.02 | 0.05 |
| Kenya | WASH−B (42) | | | | | | | | | | | | |
| Madagascar | MAHAY (43) | | | | | | | | | | | | |
| Malawi | iLiNS−DYADM (44) | 131 | 274 | 0.21 | 0.01 (−0.18, 0.20) | 0.06 | 0.06 | 73 | 148 | −0.80 | −0.12 (−0.35, 0.11) | 0.02 | 0.06 |
| Malawi | iLiNS−DOSE (45) | 398 | 141 | 0.23 | −0.07 (−0.26, 0.12) | 0.06 | 0.06 | 298 | 100 | −0.88 | −0.02 (−0.24, 0.20) | 0.03 | 0.06 |
| Mali | PROMIS (46) | 166 | 152 | 0.21 | 0.03 (−0.15, 0.20) | 0.06 | 0.06 | 333 | 350 | −0.56 | 0.13 (−0.02, 0.29) | 0.05 | 0.09 |
| Mali | PROMIS CS (46) | | | | | | | | | | | | |
| Zimbabwe | SHINE (HIV−) (47) | 743 | 702 | 0.35 | 0.09 (−0.02, 0.19) | 0.19 | 0.19 | 555 | 479 | −0.49 | 0.12 (0.00, 0.24) | 0.08 | 0.11 |
| Zimbabwe | SHINE (HIV+) (48) | 129 | 135 | 0.26 | 0.10 (−0.17, 0.38) | 0.03 | 0.03 | 132 | 99 | −0.34 | −0.21 (−0.46, 0.04) | 0.02 | 0.05 |
| | | **3978** | **2936** | | **I² = 0.00, Tau² = 0.00** | | | **6478** | **3814** | | **I² = 0.54, Tau² = 0.01** | | |
| **Fixed** | | | | | **0.06 (0.01, 0.10)** | | | | | | **0.11 (0.08, 0.15)** | | |
| **Random** | | | | | **0.06 (0.01, 0.10)** | | | | | | **0.09 (0.03, 0.16)** | | |

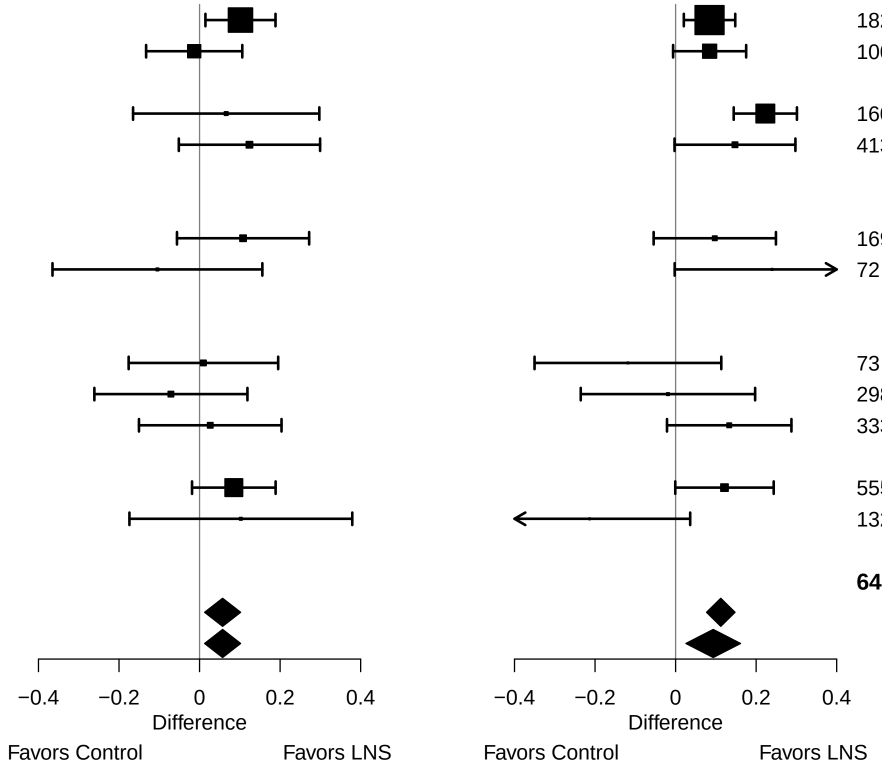

### Supplemental figure 6E: Wasting prevalence ratio

#### 6E1: Stratified by Maternal height

**P−for−interaction = 0.883**
**Ratio of PRs = 1.02 (0.81, 1.28)**

| | | At least 150.1 cm | | | | | | Less than 150.1 cm | | | | | |
|---|---|---|---|---|---|---|---|---|---|---|---|---|---|
| **Country** | **Trial** | **LNS N** | **Control N** | **Control Prevalence** | **PR (95% CI)** | **Fixed W** | **Random W** | **LNS N** | **Control N** | **Control Prevalence** | **PR (95% CI)** | **Fixed W** | **Random W** |
| Bangladesh | JiVitA−4 (34) | | | | | | | | | | | | |
| Bangladesh | RDNS (35) | 868 | 432 | 13.9 | 0.87 (0.67, 1.14) | 0.32 | 0.25 | 733 | 358 | 16.8 | 0.83 (0.60, 1.15) | 0.27 | 0.27 |
| Bangladesh | WASH−B (36) | 616 | 1880 | 10.2 | 0.65 (0.49, 0.87) | 0.26 | 0.24 | 540 | 1541 | 12.5 | 0.91 (0.71, 1.15) | 0.50 | 0.50 |
| Burkina Faso | iLiNS−Zinc (37) | | | | | | | | | | | | |
| Burkina Faso | PROMIS (38) | | | | | | | | | | | | |
| Burkina Faso | PROMIS CS (38) | | | | | | | | | | | | |
| Ghana | GHANA (39) | | | | | | | | | | | | |
| Ghana | iLiNS−DYADG (40) | | | | | | | | | | | | |
| Haiti | HAITI (41) | | | | | | | | | | | | |
| Kenya | WASH−B (42) | | | | | | | | | | | | |
| Madagascar | MAHAY (43) | 1002 | 1037 | 4.6 | 1.19 (0.74, 1.89) | 0.11 | 0.17 | 598 | 565 | 8.1 | 0.72 (0.46, 1.11) | 0.15 | 0.15 |
| Malawi | iLiNS−DYADM (44) | | | | | | | | | | | | |
| Malawi | iLiNS−DOSE (45) | 580 | 198 | 4.0 | 1.65 (0.81, 3.36) | 0.05 | 0.10 | 115 | 42 | 11.9 | 0.78 (0.30, 2.04) | 0.03 | 0.03 |
| Mali | PROMIS (46) | | | | | | | | | | | | |
| Mali | PROMIS CS (46) | 907 | 927 | 10.0 | 0.80 (0.60, 1.08) | 0.27 | 0.24 | 32 | 26 | 23.1 | 0.95 (0.44, 2.03) | 0.05 | 0.05 |
| Zimbabwe | SHINE (HIV−) (47) | | | | | | | | | | | | |
| Zimbabwe | SHINE (HIV+) (48) | | | | | | | | | | | | |
| | | **3973** | **4474** | | **I² = 0.54, Tau² = 0.06** | | | **2018** | **2532** | | **I² = 0.00, Tau² = 0.00** | | |
| **Fixed** | | | | | **0.84 (0.72, 0.98)** | | | | | | **0.85 (0.72, 1.01)** | | |
| **Random** | | | | | **0.90 (0.68, 1.18)** | | | | | | **0.85 (0.72, 1.01)** | | |

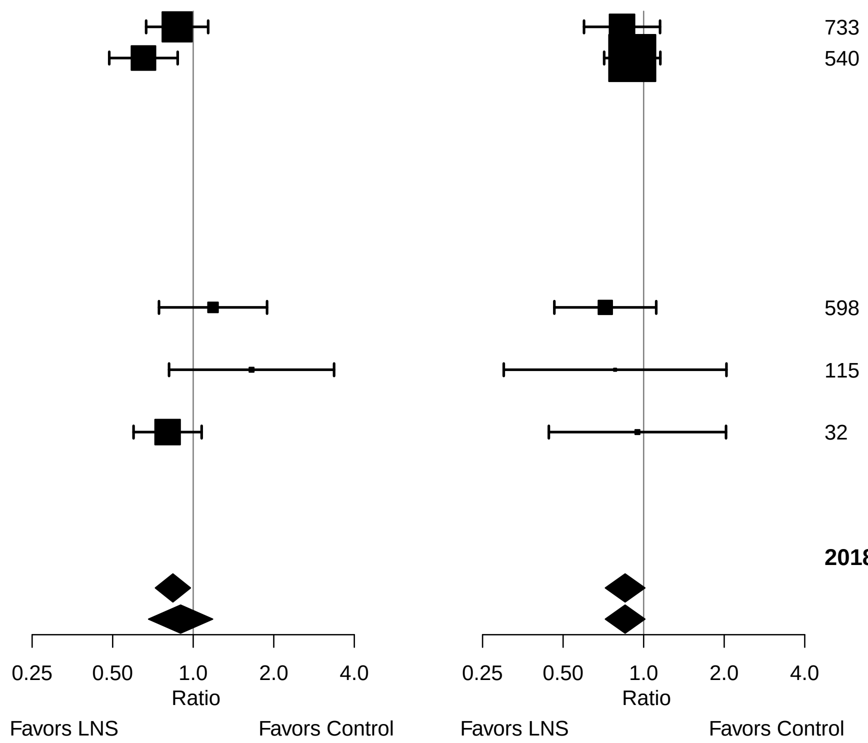

#### Supplemental figure 6E: Wasting prevalence ratio

##### 6E2: Stratified by Maternal BMI

**P−for−interaction = 0.848**
**Ratio of PRs = 1.02 (0.84, 1.23)**

| | | At least 20 kg/m² | | | | | | Less than 20 kg/m² | | | | | |
|---|---|---|---|---|---|---|---|---|---|---|---|---|---|
| **Country** | **Trial** | **LNS N** | **Control N** | **Control Prevalence** | **PR (95% CI)** | **Fixed W** | **Random W** | **LNS N** | **Control N** | **Control Prevalence** | **PR (95% CI)** | **Fixed W** | **Random W** |
| Bangladesh | JiVitA−4 (34) | | | | | | | | | | | | |
| Bangladesh | RDNS (35) | 730 | 335 | 9.6 | 0.99 (0.68, 1.44) | 0.16 | 0.16 | 871 | 455 | 19.3 | 0.82 (0.65, 1.04) | 0.25 | 0.25 |
| Bangladesh | WASH−B (36) | 517 | 1573 | 7.1 | 0.79 (0.55, 1.14) | 0.17 | 0.17 | 639 | 1848 | 14.8 | 0.77 (0.62, 0.96) | 0.30 | 0.30 |
| Burkina Faso | iLiNS−Zinc (37) | 1214 | 390 | 10.5 | 0.60 (0.41, 0.88) | 0.16 | 0.16 | 736 | 274 | 17.9 | 0.78 (0.60, 1.00) | 0.22 | 0.22 |
| Burkina Faso | PROMIS (38) | 644 | 669 | 7.3 | 0.89 (0.56, 1.43) | 0.10 | 0.10 | 209 | 232 | 13.8 | 0.80 (0.47, 1.35) | 0.05 | 0.05 |
| Burkina Faso | PROMIS CS (38) | 246 | 267 | 8.2 | 1.18 (0.70, 2.00) | 0.08 | 0.08 | 184 | 165 | 19.4 | 0.87 (0.57, 1.33) | 0.08 | 0.08 |
| Ghana | GHANA (39) | | | | | | | | | | | | |
| Ghana | iLiNS−DYADG (40) | | | | | | | | | | | | |
| Haiti | HAITI (41) | | | | | | | | | | | | |
| Kenya | WASH−B (42) | | | | | | | | | | | | |
| Madagascar | MAHAY (43) | | | | | | | | | | | | |
| Malawi | iLiNS−DYADM (44) | | | | | | | | | | | | |
| Malawi | iLiNS−DOSE (45) | 513 | 174 | 4.6 | 1.07 (0.51, 2.25) | 0.04 | 0.04 | 180 | 65 | 7.7 | 1.49 (0.60, 3.66) | 0.02 | 0.02 |
| Mali | PROMIS (46) | | | | | | | | | | | | |
| Mali | PROMIS CS (46) | 696 | 675 | 9.8 | 0.71 (0.50, 0.99) | 0.20 | 0.20 | 243 | 278 | 11.9 | 1.11 (0.71, 1.73) | 0.07 | 0.07 |
| Zimbabwe | SHINE (HIV−) (47) | 1355 | 1299 | 2.4 | 0.90 (0.54, 1.49) | 0.09 | 0.09 | 239 | 248 | 5.2 | 0.64 (0.28, 1.44) | 0.02 | 0.02 |
| Zimbabwe | SHINE (HIV+) (48) | | | | | | | | | | | | |
| | | **5915** | **5382** | | **I² = 0.00, Tau² = 0.00** | | | **3301** | **3565** | | **I² = 0.00, Tau² = 0.00** | | |
| **Fixed** | | | | | **0.82 (0.71, 0.96)** | | | | | | **0.82 (0.73, 0.92)** | | |
| **Random** | | | | | **0.82 (0.71, 0.96)** | | | | | | **0.82 (0.73, 0.92)** | | |

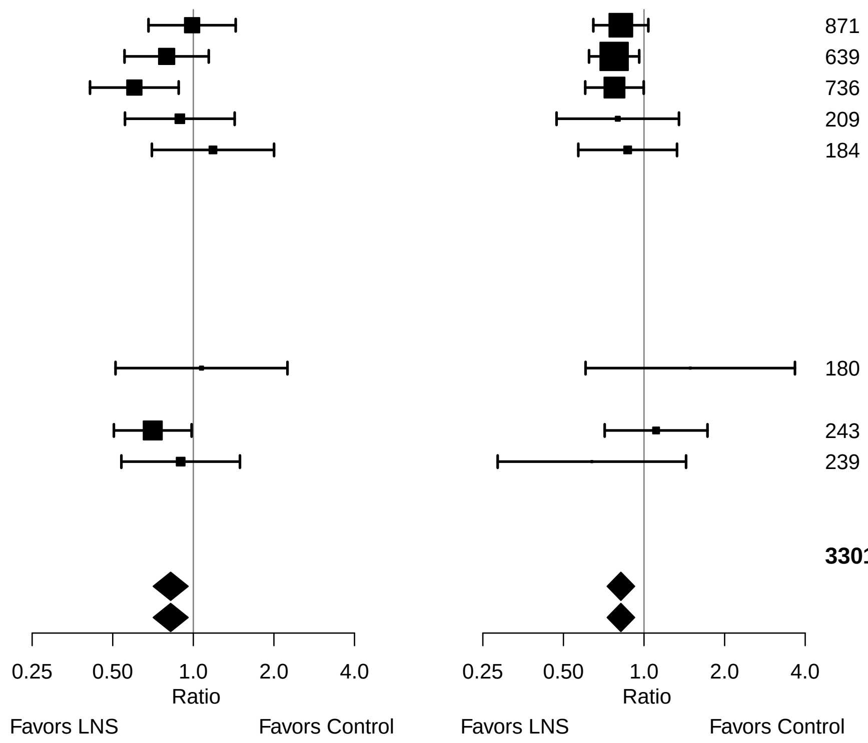

#### Supplemental figure 6E: Wasting prevalence ratio

### 6E3: Stratified by Maternal age

**P−for−interaction = 0.707**
**Ratio of PRs = 1.03 (0.88, 1.22)**

| Country | Trial | At least 25 y: LNS N | Control N | Control Prevalence | PR (95% CI) | Fixed W | Random W | Less than 25 y: LNS N | Control N | Control Prevalence | PR (95% CI) | Fixed W | Random W |
|---|---|---|---|---|---|---|---|---|---|---|---|---|---|
| Bangladesh | JiVitA−4 (34) | 1191 | 535 | 17.3 | 0.88 (0.70, 1.11) | 0.26 | 0.25 | 1580 | 677 | 15.5 | 0.94 (0.76, 1.15) | 0.30 | 0.30 |
| Bangladesh | RDNS (35) | 453 | 221 | 14.0 | 1.16 (0.82, 1.66) | 0.11 | 0.11 | 1208 | 594 | 15.5 | 0.76 (0.58, 1.01) | 0.16 | 0.16 |
| Bangladesh | WASH−B (36) | 491 | 1508 | 11.6 | 0.95 (0.69, 1.29) | 0.14 | 0.14 | 660 | 1900 | 10.9 | 0.65 (0.48, 0.88) | 0.14 | 0.14 |
| Burkina Faso | iLiNS−Zinc (37) | 1131 | 399 | 14.0 | 0.62 (0.46, 0.84) | 0.14 | 0.14 | 808 | 262 | 12.6 | 0.80 (0.56, 1.14) | 0.10 | 0.10 |
| Burkina Faso | PROMIS (38) | 480 | 522 | 8.6 | 0.92 (0.63, 1.34) | 0.09 | 0.09 | 376 | 385 | 9.4 | 0.80 (0.42, 1.51) | 0.03 | 0.03 |
| Burkina Faso | PROMIS CS (38) | 202 | 190 | 13.2 | 0.94 (0.52, 1.69) | 0.04 | 0.04 | 228 | 246 | 11.8 | 1.12 (0.75, 1.67) | 0.08 | 0.08 |
| Ghana | GHANA (39) | | | | | | | | | | | | |
| Ghana | iLiNS−DYADG (40) | | | | | | | | | | | | |
| Haiti | HAITI (41) | | | | | | | | | | | | |
| Kenya | WASH−B (42) | 842 | 2801 | 1.7 | 0.62 (0.31, 1.24) | 0.03 | 0.03 | 598 | 2285 | 1.2 | 1.23 (0.57, 2.63) | 0.02 | 0.02 |
| Madagascar | MAHAY (43) | 894 | 949 | 6.8 | 0.85 (0.58, 1.23) | 0.10 | 0.10 | 805 | 733 | 4.5 | 1.19 (0.71, 1.98) | 0.05 | 0.05 |
| Malawi | iLiNS−DYADM (44) | | | | | | | | | | | | |
| Malawi | iLiNS−DOSE (45) | | | | | | | | | | | | |
| Mali | PROMIS (46) | | | | | | | | | | | | |
| Mali | PROMIS CS (46) | 498 | 501 | 11.0 | 0.79 (0.51, 1.22) | 0.07 | 0.07 | 446 | 458 | 9.6 | 0.89 (0.60, 1.31) | 0.08 | 0.08 |
| Zimbabwe | SHINE (HIV−) (47) | 870 | 796 | 3.1 | 0.66 (0.33, 1.31) | 0.03 | 0.03 | 802 | 792 | 2.1 | 0.99 (0.53, 1.86) | 0.03 | 0.03 |
| Zimbabwe | SHINE (HIV+) (48) | | | | | | | | | | | | |
| | | **7052** | **8422** | | **I² = 0.05, Tau² = 0.00** | | | **7511** | **8332** | | **I² = 0.00, Tau² = 0.00** | | |
| **Fixed** | | | | | **0.85 (0.76, 0.96)** | | | | | | **0.87 (0.78, 0.97)** | | |
| **Random** | | | | | **0.85 (0.76, 0.96)** | | | | | | **0.87 (0.78, 0.97)** | | |

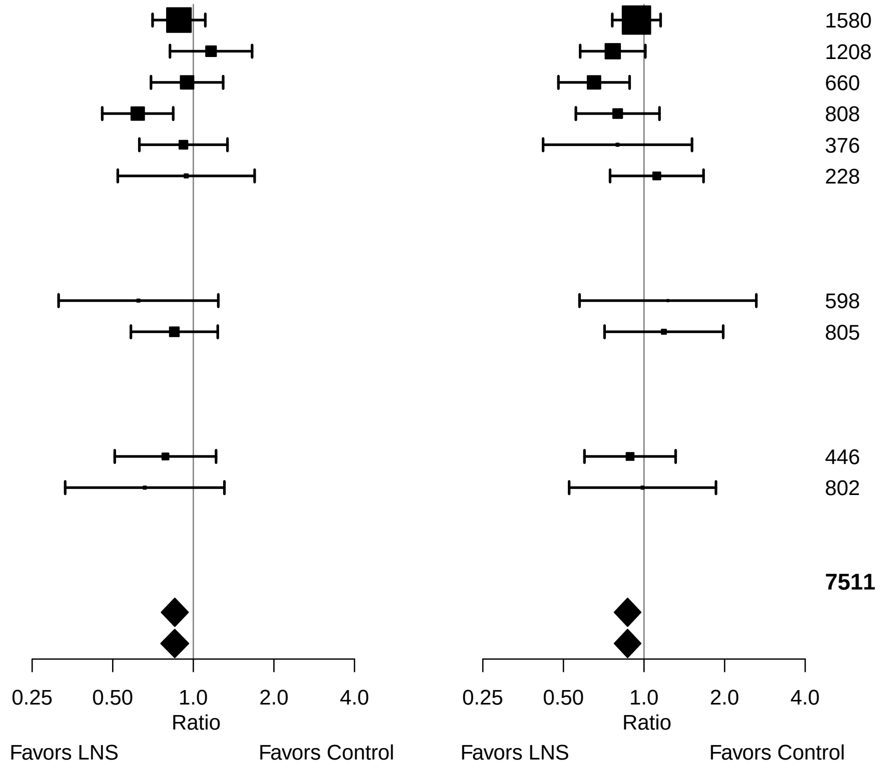

#### Supplemental figure 6E: Wasting prevalence ratio

#### 6E4: Stratified by Maternal education

**P−for−interaction = 0.890**
**Ratio of PRs = 1.01 (0.83, 1.23)**

| Country | Trial | Primary or greater: LNS N | Control N | Control Prevalence | PR (95% CI) | Fixed W | Random W | Incomplete or no formal: LNS N | Control N | Control Prevalence | PR (95% CI) | Fixed W | Random W |
|---|---|---|---|---|---|---|---|---|---|---|---|---|---|
| Bangladesh | JiVitA−4 (34) | 1797 | 739 | 15.2 | 0.94 (0.77, 1.16) | 0.38 | 0.38 | 984 | 475 | 18.3 | 0.88 (0.70, 1.09) | 0.34 | 0.34 |
| Bangladesh | RDNS (35) | 1245 | 589 | 14.9 | 0.81 (0.63, 1.05) | 0.23 | 0.23 | 416 | 226 | 15.5 | 1.02 (0.74, 1.42) | 0.15 | 0.15 |
| Bangladesh | WASH−B (36) | 813 | 2447 | 9.9 | 0.77 (0.61, 0.97) | 0.28 | 0.28 | 343 | 977 | 14.4 | 0.81 (0.58, 1.13) | 0.15 | 0.15 |
| Burkina Faso | iLiNS−Zinc (37) | | | | | | | | | | | | |
| Burkina Faso | PROMIS (38) | | | | | | | | | | | | |
| Burkina Faso | PROMIS CS (38) | | | | | | | | | | | | |
| Ghana | GHANA (39) | | | | | | | | | | | | |
| Ghana | iLiNS−DYADG (40) | 267 | 548 | 6.6 | 0.97 (0.55, 1.69) | 0.05 | 0.05 | 80 | 144 | 11.1 | 0.56 (0.21, 1.48) | 0.02 | 0.02 |
| Haiti | HAITI (41) | | | | | | | | | | | | |
| Kenya | WASH−B (42) | 700 | 2426 | 1.2 | 1.11 (0.51, 2.44) | 0.03 | 0.03 | 753 | 2688 | 1.8 | 0.74 (0.35, 1.56) | 0.03 | 0.03 |
| Madagascar | MAHAY (43) | 351 | 431 | 3.2 | 1.05 (0.49, 2.24) | 0.03 | 0.03 | 1349 | 1251 | 6.7 | 0.92 (0.62, 1.36) | 0.11 | 0.11 |
| Malawi | iLiNS−DYADM (44) | | | | | | | | | | | | |
| Malawi | iLiNS−DOSE (45) | | | | | | | | | | | | |
| Mali | PROMIS (46) | | | | | | | | | | | | |
| Mali | PROMIS CS (46) | 107 | 96 | 9.4 | 0.50 (0.17, 1.49) | 0.01 | 0.01 | 836 | 863 | 10.4 | 0.87 (0.65, 1.16) | 0.20 | 0.20 |
| Zimbabwe | SHINE (HIV−) (47) | | | | | | | | | | | | |
| Zimbabwe | SHINE (HIV+) (48) | | | | | | | | | | | | |
| | | **5280** | **7276** | | **I² = 0.00, Tau² = 0.00** | | | **4761** | **6624** | | **I² = 0.00, Tau² = 0.00** | | |
| **Fixed** | | | | | **0.86 (0.76, 0.97)** | | | | | | **0.88 (0.77, 1.00)** | | |
| **Random** | | | | | **0.86 (0.76, 0.97)** | | | | | | **0.88 (0.77, 1.00)** | | |

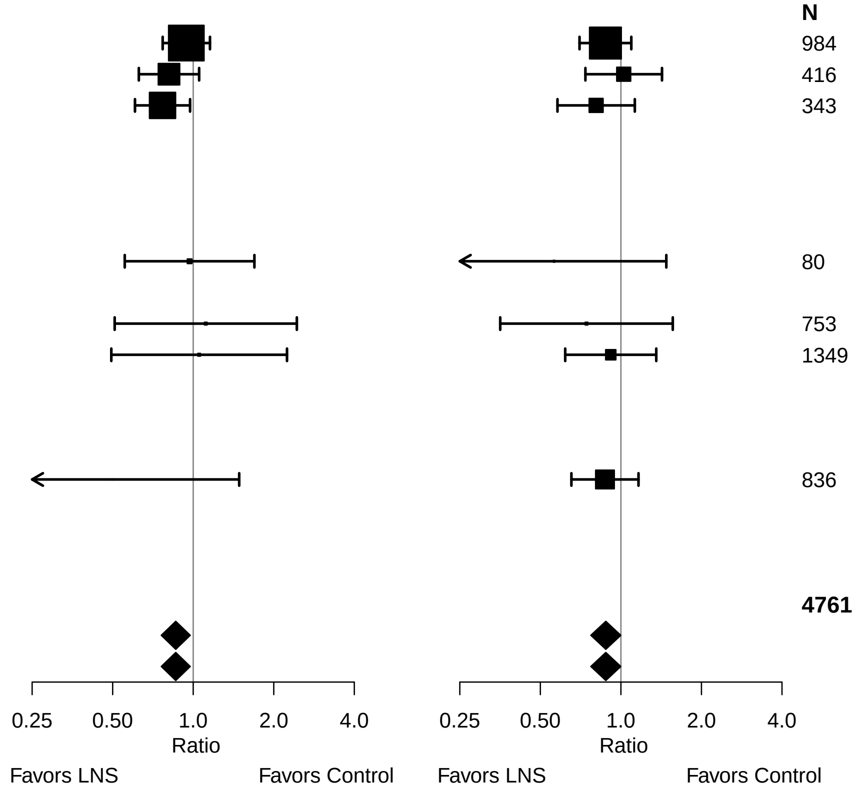

#### Supplemental figure 6E: Wasting prevalence ratio

##### 6E5: Stratified by Maternal depressive symptoms

**P−for−interaction = 0.265**
**Ratio of PRs = 1.15 (0.90, 1.48)**

| | | Less than 75th percentile | | | | | | At least 75th percentile | | | | | |
|---|---|---|---|---|---|---|---|---|---|---|---|---|---|
| **Country** | **Trial** | **LNS N** | **Control N** | **Control Prevalence** | **PR (95% CI)** | **Fixed W** | **Random W** | **LNS N** | **Control N** | **Control Prevalence** | **PR (95% CI)** | **Fixed W** | **Random W** |
| Bangladesh | JiVitA−4 (34) | | | | | | | | | | | | |
| Bangladesh | RDNS (35) | 1081 | 472 | 13.6 | 1.00 (0.76, 1.32) | 0.27 | 0.27 | 519 | 288 | 16.7 | 0.72 (0.48, 1.07) | 0.30 | 0.30 |
| Bangladesh | WASH−B (36) | 895 | 2457 | 10.9 | 0.81 (0.65, 1.01) | 0.41 | 0.41 | 237 | 876 | 12.0 | 0.74 (0.49, 1.10) | 0.29 | 0.29 |
| Burkina Faso | iLiNS−Zinc (37) | | | | | | | | | | | | |
| Burkina Faso | PROMIS (38) | 505 | 569 | 8.3 | 0.86 (0.56, 1.34) | 0.10 | 0.10 | 219 | 212 | 12.7 | 0.72 (0.41, 1.26) | 0.15 | 0.15 |
| Burkina Faso | PROMIS CS (38) | | | | | | | | | | | | |
| Ghana | GHANA (39) | | | | | | | | | | | | |
| Ghana | iLiNS−DYADG (40) | 252 | 459 | 6.8 | 0.88 (0.49, 1.60) | 0.05 | 0.05 | 83 | 209 | 8.1 | 1.04 (0.45, 2.41) | 0.07 | 0.07 |
| Haiti | HAITI (41) | | | | | | | | | | | | |
| Kenya | WASH−B (42) | | | | | | | | | | | | |
| Madagascar | MAHAY (43) | 672 | 721 | 5.8 | 0.95 (0.58, 1.54) | 0.08 | 0.08 | 283 | 252 | 4.0 | 1.16 (0.54, 2.50) | 0.08 | 0.08 |
| Malawi | iLiNS−DYADM (44) | | | | | | | | | | | | |
| Malawi | iLiNS−DOSE (45) | | | | | | | | | | | | |
| Mali | PROMIS (46) | | | | | | | | | | | | |
| Mali | PROMIS CS (46) | | | | | | | | | | | | |
| Zimbabwe | SHINE (HIV−) (47) | 1192 | 1136 | 2.7 | 0.83 (0.49, 1.41) | 0.07 | 0.07 | 509 | 489 | 2.7 | 0.81 (0.39, 1.69) | 0.09 | 0.09 |
| Zimbabwe | SHINE (HIV+) (48) | 234 | 214 | 2.3 | 2.01 (0.74, 5.44) | 0.02 | 0.02 | 88 | 100 | 6.0 | 0.95 (0.28, 3.16) | 0.03 | 0.03 |
| | | **4831** | **6028** | | **I² = 0.00, Tau² = 0.00** | | | **1938** | **2426** | | **I² = 0.00, Tau² = 0.00** | | |
| **Fixed** | | | | | **0.90 (0.78, 1.03)** | | | | | | **0.79 (0.63, 0.98)** | | |
| **Random** | | | | | **0.90 (0.78, 1.03)** | | | | | | **0.79 (0.63, 0.98)** | | |

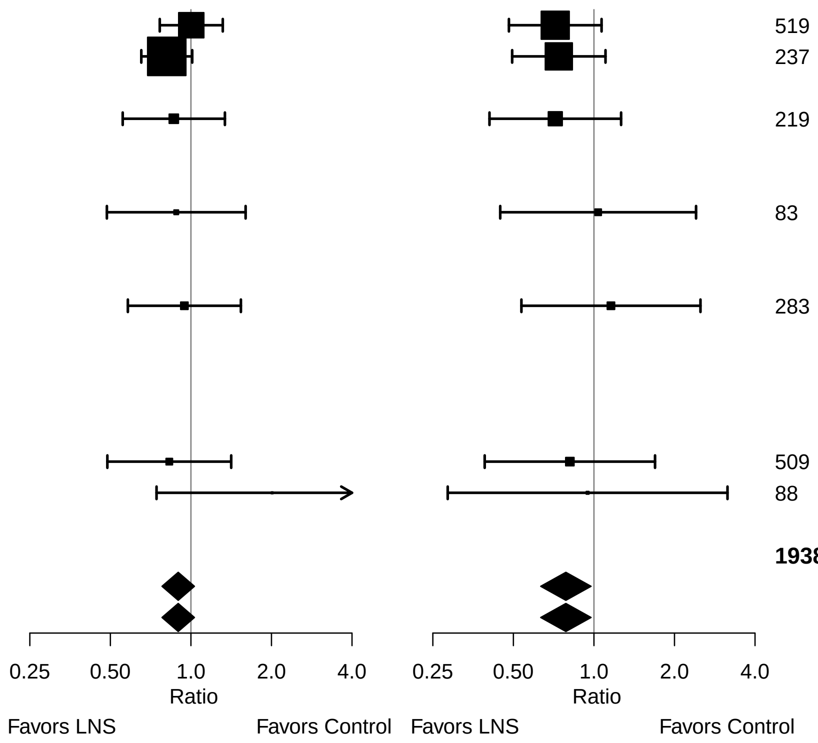

#### Supplemental figure 6E: Wasting prevalence ratio

### 6E6: Stratified by Child sex

**P−for−interaction = 0.074**
**Ratio of PRs = 0.87 (0.75, 1.01)**

| | | Male | | | | | | Female | | | | | |
|---|---|---|---|---|---|---|---|---|---|---|---|---|---|
| **Country** | **Trial** | **LNS N** | **Control N** | **Control Prevalence** | **PR (95% CI)** | **Fixed W** | **Random W** | **LNS N** | **Control N** | **Control Prevalence** | **PR (95% CI)** | **Fixed W** | **Random W** |
| Bangladesh | JiVitA−4 (34) | 1420 | 586 | 15.6 | 1.04 (0.85, 1.28) | 0.24 | 0.24 | 1363 | 631 | 17.1 | 0.77 (0.63, 0.95) | 0.33 | 0.28 |
| Bangladesh | RDNS (35) | 832 | 411 | 15.6 | 0.90 (0.68, 1.18) | 0.14 | 0.14 | 829 | 404 | 14.6 | 0.83 (0.61, 1.15) | 0.13 | 0.14 |
| Bangladesh | WASH−B (36) | 564 | 1711 | 12.2 | 0.90 (0.71, 1.16) | 0.17 | 0.17 | 592 | 1713 | 10.3 | 0.66 (0.48, 0.90) | 0.14 | 0.14 |
| Burkina Faso | iLiNS−Zinc (37) | 990 | 336 | 14.9 | 0.75 (0.57, 0.98) | 0.15 | 0.15 | 962 | 328 | 12.2 | 0.62 (0.44, 0.87) | 0.12 | 0.13 |
| Burkina Faso | PROMIS (38) | 434 | 480 | 12.5 | 0.72 (0.43, 1.20) | 0.04 | 0.04 | 422 | 427 | 4.9 | 1.30 (0.76, 2.23) | 0.05 | 0.05 |
| Burkina Faso | PROMIS CS (38) | 234 | 221 | 13.6 | 1.16 (0.70, 1.94) | 0.04 | 0.04 | 196 | 215 | 11.2 | 0.82 (0.44, 1.52) | 0.04 | 0.04 |
| Ghana | GHANA (39) | | | | | | | | | | | | |
| Ghana | iLiNS−DYADG (40) | 175 | 323 | 9.9 | 0.63 (0.33, 1.23) | 0.02 | 0.02 | 172 | 369 | 5.4 | 1.18 (0.58, 2.41) | 0.03 | 0.03 |
| Haiti | HAITI (41) | | | | | | | | | | | | |
| Kenya | WASH−B (42) | 732 | 2431 | 2.0 | 0.97 (0.50, 1.89) | 0.02 | 0.02 | 723 | 2687 | 1.0 | 0.66 (0.25, 1.75) | 0.01 | 0.02 |
| Madagascar | MAHAY (43) | 824 | 832 | 8.1 | 0.90 (0.59, 1.39) | 0.06 | 0.06 | 876 | 850 | 3.6 | 1.10 (0.67, 1.80) | 0.06 | 0.06 |
| Malawi | iLiNS−DYADM (44) | | | | | | | | | | | | |
| Malawi | iLiNS−DOSE (45) | | | | | | | | | | | | |
| Mali | PROMIS (46) | | | | | | | | | | | | |
| Mali | PROMIS CS (46) | 472 | 520 | 11.5 | 0.94 (0.63, 1.40) | 0.07 | 0.07 | 472 | 439 | 8.9 | 0.72 (0.45, 1.14) | 0.06 | 0.07 |
| Zimbabwe | SHINE (HIV−) (47) | 924 | 905 | 3.3 | 0.78 (0.47, 1.32) | 0.04 | 0.04 | 945 | 879 | 1.8 | 1.10 (0.57, 2.16) | 0.03 | 0.04 |
| Zimbabwe | SHINE (HIV+) (48) | | | | | | | | | | | | |
| | | **7601** | **8756** | | **I² = 0.00, Tau² = 0.00** | | | **7552** | **8942** | | **I² = 0.08, Tau² = 0.00** | | |
| **Fixed** | | | | | **0.90 (0.81, 1.00)** | | | | | | **0.79 (0.70, 0.89)** | | |
| **Random** | | | | | **0.90 (0.81, 1.00)** | | | | | | **0.80 (0.70, 0.91)** | | |

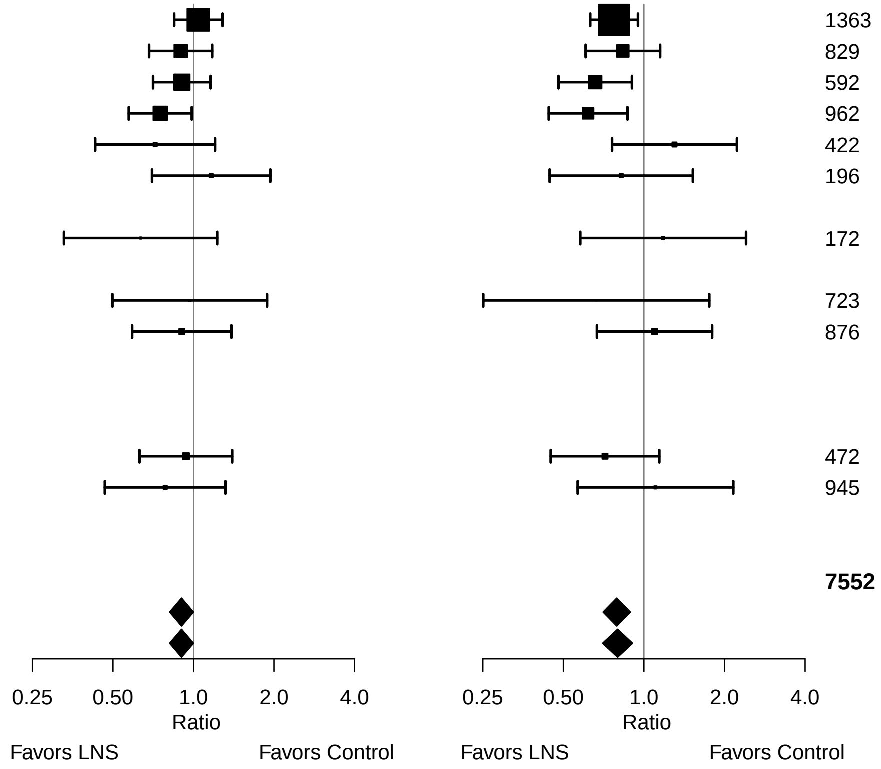

#### Supplemental figure 6E: Wasting prevalence ratio

#### 6E7: Stratified by Child birth order

**P−for−interaction = 0.609**
**Ratio of PRs = 1.05 (0.88, 1.25)**

**Later born**

| Country | Trial | LNS N | Control N | Control Prevalence | PR (95% CI) | Fixed W | Random W |
|---|---|---|---|---|---|---|---|
| Bangladesh | JiVitA−4 (34) | 598 | 247 | 17.1 | 0.87 (0.64, 1.19) | 0.11 | 0.11 |
| Bangladesh | RDNS (35) | 972 | 508 | 15.7 | 0.86 (0.66, 1.10) | 0.16 | 0.16 |
| Bangladesh | WASH−B (36) | 735 | 2285 | 11.3 | 0.78 (0.59, 1.02) | 0.14 | 0.14 |
| Burkina Faso | iLiNS−Zinc (37) | 1520 | 519 | 13.7 | 0.64 (0.51, 0.80) | 0.21 | 0.21 |
| Burkina Faso | PROMIS (38) | 708 | 747 | 9.4 | 0.87 (0.61, 1.26) | 0.08 | 0.08 |
| Burkina Faso | PROMIS CS (38) | 366 | 357 | 11.5 | 1.14 (0.74, 1.76) | 0.05 | 0.05 |
| Ghana | GHANA (39) | | | | | | |
| Ghana | iLiNS−DYADG (40) | | | | | | |
| Haiti | HAITI (41) | | | | | | |
| Kenya | WASH−B (42) | 1132 | 4027 | 1.5 | 0.83 (0.45, 1.52) | 0.03 | 0.03 |
| Madagascar | MAHAY (43) | 1270 | 1217 | 6.4 | 0.91 (0.62, 1.34) | 0.07 | 0.07 |
| Malawi | iLiNS−DYADM (44) | | | | | | |
| Malawi | iLiNS−DOSE (45) | | | | | | |
| Mali | PROMIS (46) | | | | | | |
| Mali | PROMIS CS (46) | 783 | 821 | 10.5 | 0.85 (0.64, 1.14) | 0.12 | 0.12 |
| Zimbabwe | SHINE (HIV−) (47) | 1295 | 1218 | 2.5 | 0.82 (0.49, 1.38) | 0.04 | 0.04 |
| Zimbabwe | SHINE (HIV+) (48) | | | | | | |
| | | **9379** | **11946** | | **I² = 0.00, Tau² = 0.00** | | |
| **Fixed** | | | | | **0.81 (0.73, 0.90)** | | |
| **Random** | | | | | **0.81 (0.73, 0.90)** | | |

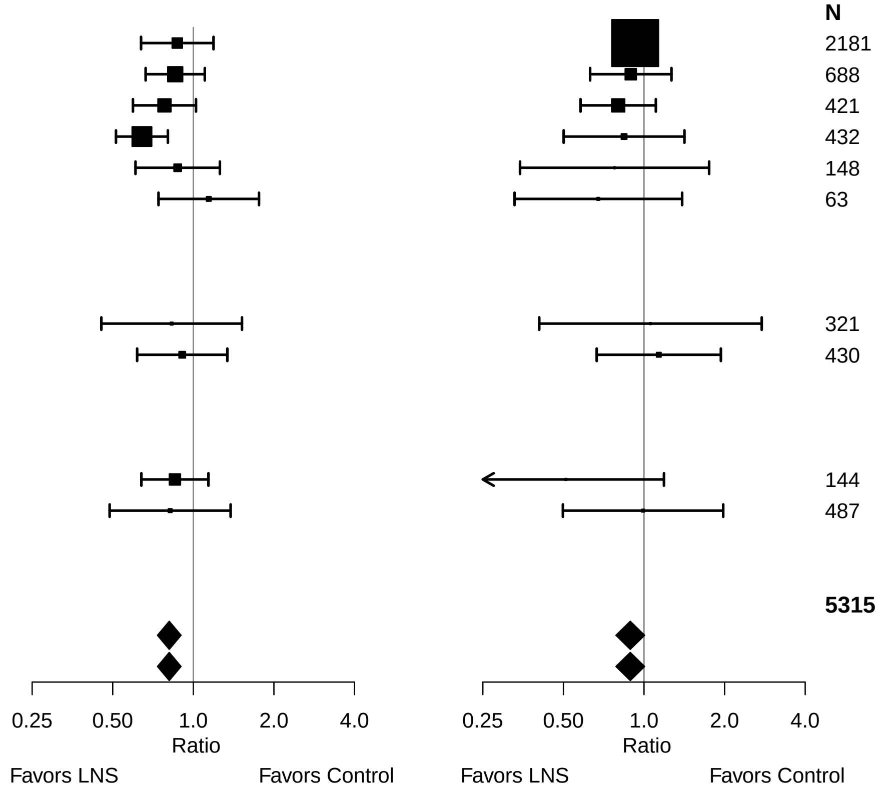

**Firstborn**

| Country | Trial | LNS N | Control N | Control Prevalence | PR (95% CI) | Fixed W | Random W |
|---|---|---|---|---|---|---|---|
| Bangladesh | JiVitA−4 (34) | 2181 | 964 | 16.2 | 0.93 (0.78, 1.10) | 0.50 | 0.50 |
| Bangladesh | RDNS (35) | 688 | 307 | 14.0 | 0.89 (0.63, 1.27) | 0.12 | 0.12 |
| Bangladesh | WASH−B (36) | 421 | 1139 | 11.0 | 0.80 (0.58, 1.11) | 0.14 | 0.14 |
| Burkina Faso | iLiNS−Zinc (37) | 432 | 145 | 13.1 | 0.84 (0.50, 1.42) | 0.06 | 0.06 |
| Burkina Faso | PROMIS (38) | 148 | 158 | 7.0 | 0.78 (0.34, 1.75) | 0.02 | 0.02 |
| Burkina Faso | PROMIS CS (38) | 63 | 79 | 16.5 | 0.68 (0.33, 1.39) | 0.03 | 0.03 |
| Ghana | GHANA (39) | | | | | | |
| Ghana | iLiNS−DYADG (40) | | | | | | |
| Haiti | HAITI (41) | | | | | | |
| Kenya | WASH−B (42) | 321 | 1086 | 1.5 | 1.06 (0.41, 2.75) | 0.02 | 0.02 |
| Madagascar | MAHAY (43) | 430 | 465 | 4.3 | 1.14 (0.67, 1.94) | 0.05 | 0.05 |
| Malawi | iLiNS−DYADM (44) | | | | | | |
| Malawi | iLiNS−DOSE (45) | | | | | | |
| Mali | PROMIS (46) | | | | | | |
| Mali | PROMIS CS (46) | 144 | 126 | 9.5 | 0.51 (0.22, 1.19) | 0.02 | 0.02 |
| Zimbabwe | SHINE (HIV−) (47) | 487 | 483 | 2.9 | 0.99 (0.50, 1.98) | 0.03 | 0.03 |
| Zimbabwe | SHINE (HIV+) (48) | | | | | | |
| | | **5315** | **4952** | | **I² = 0.00, Tau² = 0.00** | | |
| **Fixed** | | | | | **0.89 (0.79, 1.01)** | | |
| **Random** | | | | | **0.89 (0.79, 1.01)** | | |

##### Supplemental figure 6E: Wasting prevalence ratio

#### 6E8: Stratified by Child baseline anthropometric status

**P−for−interaction = 0.682**
**Ratio of PRs = 0.89 (0.50, 1.58)**

| Country | Trial | WLZ at least 0 Z LNS N | Control N | Control Prevalence | PR (95% CI) | Fixed W | Random W | WLZ less than 0 Z LNS N | Control N | Control Prevalence | PR (95% CI) | Fixed W | Random W |
|---|---|---|---|---|---|---|---|---|---|---|---|---|---|
| Bangladesh | JiVitA−4 (34) | 961 | 423 | 2.4 | 0.88 (0.41, 1.89) | 0.49 | 0.49 | 1822 | 794 | 23.9 | 0.90 (0.78, 1.04) | 0.84 | 0.84 |
| Bangladesh | RDNS (35) | | | | | | | | | | | | |
| Bangladesh | WASH−B (36) | | | | | | | | | | | | |
| Burkina Faso | iLiNS−Zinc (37) | | | | | | | | | | | | |
| Burkina Faso | PROMIS (38) | 279 | 350 | 2.0 | 0.90 (0.25, 3.19) | 0.18 | 0.18 | 413 | 393 | 14.2 | 0.80 (0.54, 1.18) | 0.12 | 0.12 |
| Burkina Faso | PROMIS CS (38) | | | | | | | | | | | | |
| Ghana | GHANA (39) | | | | | | | | | | | | |
| Ghana | iLiNS−DYADG (40) | | | | | | | | | | | | |
| Haiti | HAITI (41) | | | | | | | | | | | | |
| Kenya | WASH−B (42) | | | | | | | | | | | | |
| Madagascar | MAHAY (43) | | | | | | | | | | | | |
| Malawi | iLiNS−DYADM (44) | | | | | | | | | | | | |
| Malawi | iLiNS−DOSE (45) | | | | | | | | | | | | |
| Mali | PROMIS (46) | | | | | | | | | | | | |
| Mali | PROMIS CS (46) | | | | | | | | | | | | |
| Zimbabwe | SHINE (HIV−) (47) | 743 | 702 | 1.0 | 1.08 (0.42, 2.76) | 0.33 | 0.33 | 555 | 479 | 4.8 | 0.68 (0.34, 1.33) | 0.04 | 0.04 |
| Zimbabwe | SHINE (HIV+) (48) | | | | | | | | | | | | |
| | | **1983** | **1475** | | **I² = 0.00, Tau² = 0.00** | | | **2790** | **1666** | | **I² = 0.00, Tau² = 0.00** | | |
| **Fixed** | | | | | **0.94 (0.55, 1.62)** | | | | | | **0.88 (0.77, 1.01)** | | |
| **Random** | | | | | **0.94 (0.55, 1.62)** | | | | | | **0.88 (0.77, 1.01)** | | |

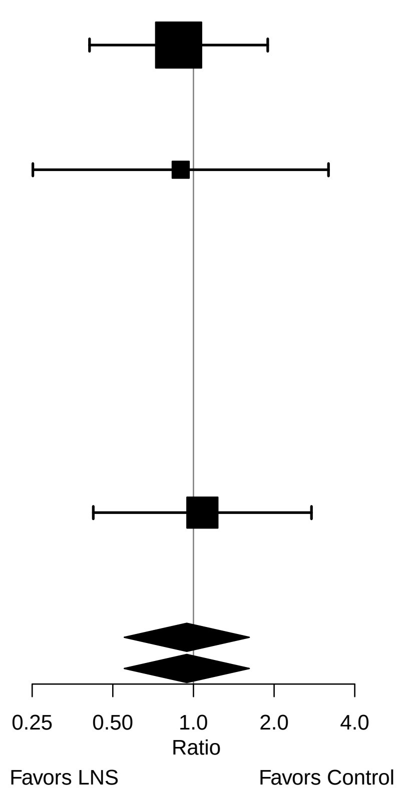

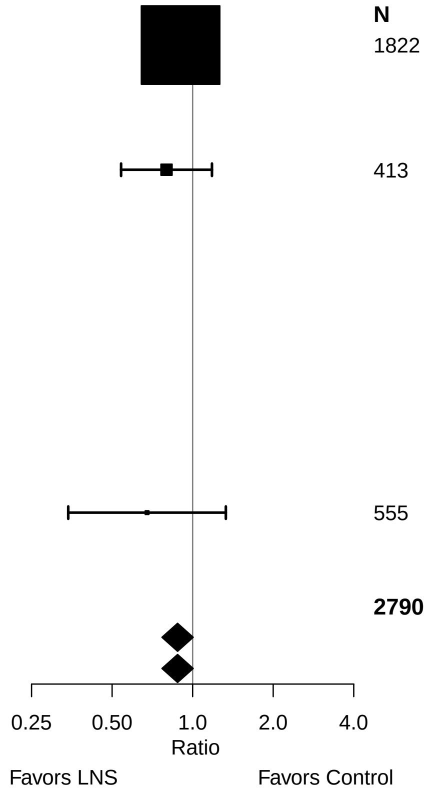

### Supplemental figure 6F: Wasting prevalence difference

#### 6F1: Stratified by Maternal height

**P−for−interaction = 0.438**
**Difference in PDs = −0.01 (−0.03, 0.01)**

| | | At least 150.1 cm | | | | | | Less than 150.1 cm | | | | | |
|---|---|---|---|---|---|---|---|---|---|---|---|---|---|
| Country | Trial | LNS N | Control N | Control Prevalence | PD (95% CI) | Fixed W | Random W | LNS N | Control N | Control Prevalence | PD (95% CI) | Fixed W | Random W |
| Bangladesh | JiVitA−4 (34) | | | | | | | | | | | | |
| Bangladesh | RDNS (35) | 868 | 432 | 13.9 | −0.02 (−0.05, 0.02) | 0.12 | 0.16 | 733 | 358 | 16.8 | −0.03 (−0.08, 0.02) | 0.13 | 0.13 |
| Bangladesh | WASH−B (36) | 616 | 1880 | 10.2 | −0.04 (−0.06, −0.01) | 0.30 | 0.25 | 540 | 1541 | 12.5 | −0.01 (−0.04, 0.02) | 0.46 | 0.46 |
| Burkina Faso | iLiNS−Zinc (37) | | | | | | | | | | | | |
| Burkina Faso | PROMIS (38) | | | | | | | | | | | | |
| Burkina Faso | PROMIS CS (38) | | | | | | | | | | | | |
| Ghana | GHANA (39) | | | | | | | | | | | | |
| Ghana | iLiNS−DYADG (40) | | | | | | | | | | | | |
| Haiti | HAITI (41) | | | | | | | | | | | | |
| Kenya | WASH−B (42) | | | | | | | | | | | | |
| Madagascar | MAHAY (43) | 1002 | 1037 | 4.6 | 0.01 (−0.01, 0.03) | 0.27 | 0.23 | 598 | 565 | 8.1 | −0.02 (−0.05, 0.01) | 0.37 | 0.37 |
| Malawi | iLiNS−DYADM (44) | | | | | | | | | | | | |
| Malawi | iLiNS−DOSE (45) | 580 | 198 | 4.0 | 0.01 (−0.02, 0.05) | 0.11 | 0.15 | 115 | 42 | 11.9 | −0.03 (−0.14, 0.07) | 0.03 | 0.03 |
| Mali | PROMIS (46) | | | | | | | | | | | | |
| Mali | PROMIS CS (46) | 907 | 927 | 10.0 | −0.02 (−0.05, 0.01) | 0.20 | 0.21 | 32 | 26 | 23.1 | −0.01 (−0.18, 0.16) | 0.01 | 0.01 |
| Zimbabwe | SHINE (HIV−) (47) | | | | | | | | | | | | |
| Zimbabwe | SHINE (HIV+) (48) | | | | | | | | | | | | |
| | | **3973** | **4474** | | **I² = 0.60, Tau² = 0.00** | | | **2018** | **2532** | | **I² = 0.00, Tau² = 0.00** | | |
| **Fixed** | | | | | **−0.01 (−0.03, 0.00)** | | | | | | **−0.02 (−0.04, 0.00)** | | |
| **Random** | | | | | **−0.01 (−0.03, 0.01)** | | | | | | **−0.02 (−0.04, 0.00)** | | |

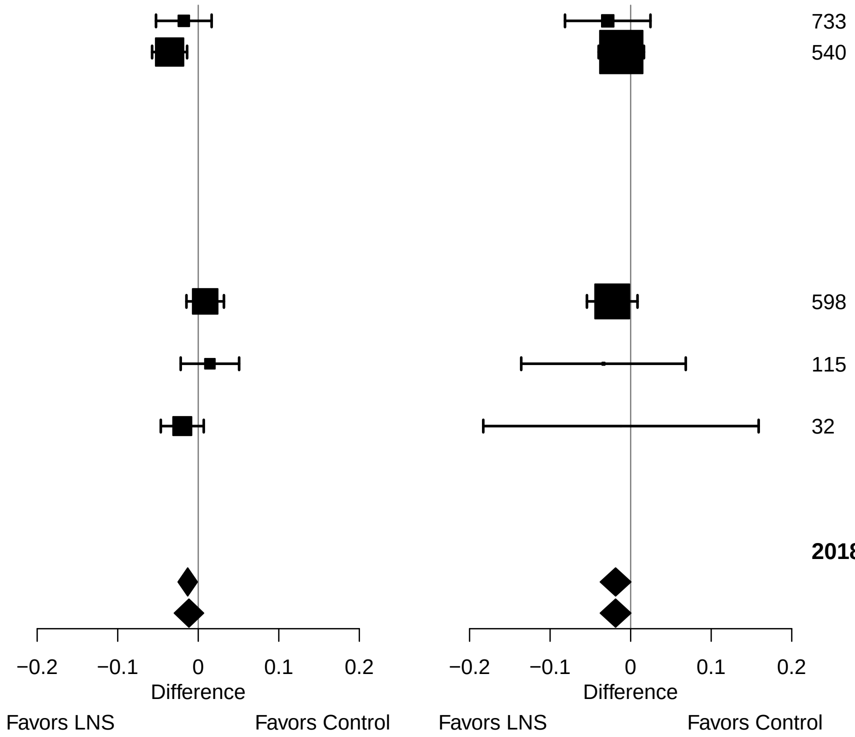

#### Supplemental figure 6F: Wasting prevalence difference

#### 6F2: Stratified by Maternal BMI

**P−for−interaction = 0.228**
**Difference in PDs = −0.01 (−0.03, 0.01)**

| | | At least 20 kg/m² | | | | | | Less than 20 kg/m² | | | | | |
|---|---|---|---|---|---|---|---|---|---|---|---|---|---|
| **Country** | **Trial** | **LNS N** | **Control N** | **Control Prevalence** | **PD (95% CI)** | **Fixed W** | **Random W** | **LNS N** | **Control N** | **Control Prevalence** | **PD (95% CI)** | **Fixed W** | **Random W** |
| Bangladesh | JiVitA−4 (34) | | | | | | | | | | | | |
| Bangladesh | RDNS (35) | 730 | 335 | 9.6 | 0.00 (−0.04, 0.03) | 0.05 | 0.09 | 871 | 455 | 19.3 | −0.03 (−0.08, 0.01) | 0.12 | 0.12 |
| Bangladesh | WASH−B (36) | 517 | 1573 | 7.1 | −0.01 (−0.04, 0.01) | 0.13 | 0.16 | 639 | 1848 | 14.8 | −0.03 (−0.06, −0.01) | 0.33 | 0.33 |
| Burkina Faso | iLiNS−Zinc (37) | 1214 | 390 | 10.5 | −0.04 (−0.06, −0.01) | 0.11 | 0.14 | 736 | 274 | 17.9 | −0.04 (−0.09, 0.00) | 0.14 | 0.14 |
| Burkina Faso | PROMIS (38) | 644 | 669 | 7.3 | −0.01 (−0.04, 0.02) | 0.06 | 0.10 | 209 | 232 | 13.8 | −0.03 (−0.09, 0.04) | 0.05 | 0.05 |
| Burkina Faso | PROMIS CS (38) | 246 | 267 | 8.2 | 0.02 (−0.03, 0.06) | 0.03 | 0.06 | 184 | 165 | 19.4 | −0.03 (−0.10, 0.05) | 0.04 | 0.04 |
| Ghana | GHANA (39) | | | | | | | | | | | | |
| Ghana | iLiNS−DYADG (40) | | | | | | | | | | | | |
| Haiti | HAITI (41) | | | | | | | | | | | | |
| Kenya | WASH−B (42) | | | | | | | | | | | | |
| Madagascar | MAHAY (43) | | | | | | | | | | | | |
| Malawi | iLiNS−DYADM (44) | | | | | | | | | | | | |
| Malawi | iLiNS−DOSE (45) | 513 | 174 | 4.6 | 0.00 (−0.04, 0.03) | 0.05 | 0.08 | 180 | 65 | 7.7 | 0.02 (−0.06, 0.11) | 0.03 | 0.03 |
| Mali | PROMIS (46) | | | | | | | | | | | | |
| Mali | PROMIS CS (46) | 696 | 675 | 9.8 | −0.03 (−0.06, 0.00) | 0.08 | 0.11 | 243 | 278 | 11.9 | 0.01 (−0.04, 0.07) | 0.08 | 0.08 |
| Zimbabwe | SHINE (HIV−) (47) | 1355 | 1299 | 2.4 | 0.00 (−0.01, 0.01) | 0.49 | 0.26 | 239 | 248 | 5.2 | −0.02 (−0.05, 0.01) | 0.21 | 0.21 |
| Zimbabwe | SHINE (HIV+) (48) | | | | | | | | | | | | |
| | | **5915** | **5382** | | **I² = 0.32, Tau² = 0.00** | | | **3301** | **3565** | | **I² = 0.00, Tau² = 0.00** | | |
| **Fixed** | | | | | **−0.01 (−0.02, 0.00)** | | | | | | **−0.03 (−0.04, −0.01)** | | |
| **Random** | | | | | **−0.01 (−0.02, 0.00)** | | | | | | **−0.03 (−0.04, −0.01)** | | |

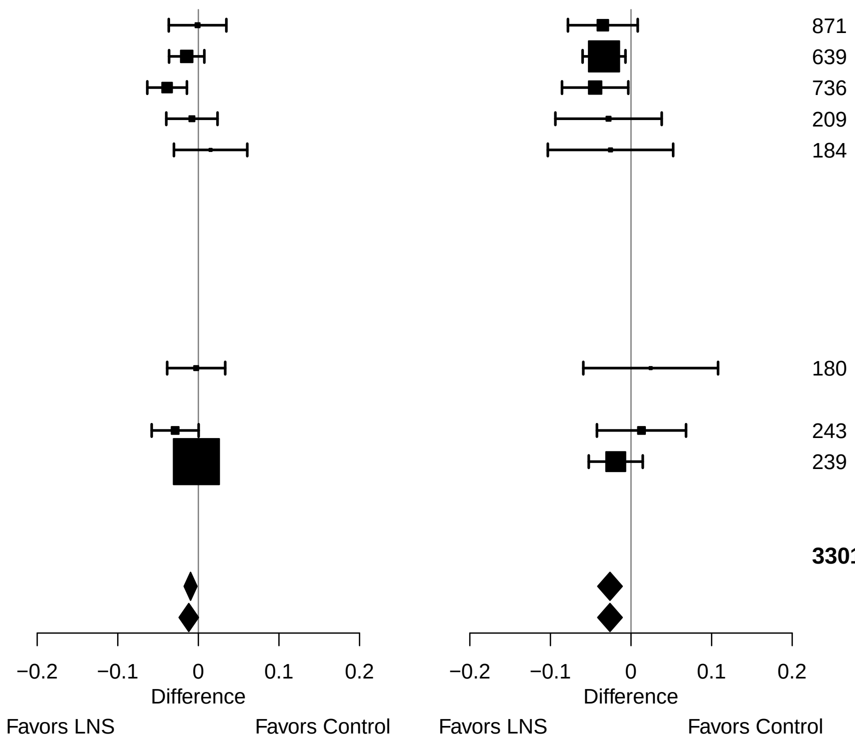

#### Supplemental figure 6F: Wasting prevalence difference

#### 6F3: Stratified by Maternal age

**P−for−interaction = 0.171**
**Difference in PDs = 0.01 (0.00, 0.02)**

| | | At least 25 y | | | | | | Less than 25 y | | | | | |
|---|---|---|---|---|---|---|---|---|---|---|---|---|---|
| **Country** | **Trial** | **LNS N** | **Control N** | **Control Prevalence** | **PD (95% CI)** | **Fixed W** | **Random W** | **LNS N** | **Control N** | **Control Prevalence** | **PD (95% CI)** | **Fixed W** | **Random W** |
| Bangladesh | JiVitA−4 (34) | 1191 | 535 | 17.3 | −0.02 (−0.05, 0.02) | 0.03 | 0.03 | 1580 | 677 | 15.5 | −0.01 (−0.04, 0.02) | 0.05 | 0.09 |
| Bangladesh | RDNS (35) | 453 | 221 | 14.0 | 0.02 (−0.03, 0.08) | 0.01 | 0.01 | 1208 | 594 | 15.5 | −0.04 (−0.08, 0.00) | 0.03 | 0.05 |
| Bangladesh | WASH−B (36) | 491 | 1508 | 11.6 | −0.01 (−0.04, 0.03) | 0.03 | 0.03 | 660 | 1900 | 10.9 | −0.04 (−0.06, −0.01) | 0.08 | 0.11 |
| Burkina Faso | iLiNS−Zinc (37) | 1131 | 399 | 14.0 | −0.05 (−0.09, −0.01) | 0.03 | 0.03 | 808 | 262 | 12.6 | −0.03 (−0.07, 0.02) | 0.03 | 0.05 |
| Burkina Faso | PROMIS (38) | 480 | 522 | 8.6 | −0.01 (−0.04, 0.02) | 0.04 | 0.04 | 376 | 385 | 9.4 | −0.02 (−0.07, 0.03) | 0.02 | 0.03 |
| Burkina Faso | PROMIS CS (38) | 202 | 190 | 13.2 | −0.01 (−0.08, 0.07) | 0.01 | 0.01 | 228 | 246 | 11.8 | 0.01 (−0.04, 0.06) | 0.02 | 0.04 |
| Ghana | GHANA (39) | | | | | | | | | | | | |
| Ghana | iLiNS−DYADG (40) | | | | | | | | | | | | |
| Haiti | HAITI (41) | | | | | | | | | | | | |
| Kenya | WASH−B (42) | 842 | 2801 | 1.7 | −0.01 (−0.01, 0.00) | 0.62 | 0.62 | 598 | 2285 | 1.2 | 0.00 (−0.01, 0.01) | 0.40 | 0.24 |
| Madagascar | MAHAY (43) | 894 | 949 | 6.8 | −0.01 (−0.03, 0.01) | 0.07 | 0.07 | 805 | 733 | 4.5 | 0.01 (−0.02, 0.03) | 0.08 | 0.11 |
| Malawi | iLiNS−DYADM (44) | | | | | | | | | | | | |
| Malawi | iLiNS−DOSE (45) | | | | | | | | | | | | |
| Mali | PROMIS (46) | | | | | | | | | | | | |
| Mali | PROMIS CS (46) | 498 | 501 | 11.0 | −0.02 (−0.07, 0.02) | 0.02 | 0.02 | 446 | 458 | 9.6 | −0.01 (−0.05, 0.02) | 0.04 | 0.07 |
| Zimbabwe | SHINE (HIV−) (47) | 870 | 796 | 3.1 | −0.01 (−0.03, 0.01) | 0.14 | 0.14 | 802 | 792 | 2.1 | 0.00 (−0.01, 0.01) | 0.26 | 0.21 |
| Zimbabwe | SHINE (HIV+) (48) | | | | | | | | | | | | |
| | | **7052** | **8422** | | **I² = 0.00, Tau² = 0.00** | | | **7511** | **8332** | | **I² = 0.39, Tau² = 0.00** | | |
| **Fixed** | | | | | **−0.01 (−0.02, 0.00)** | | | | | | **0.00 (−0.01, 0.00)** | | |
| **Random** | | | | | **−0.01 (−0.02, 0.00)** | | | | | | **−0.01 (−0.02, 0.00)** | | |

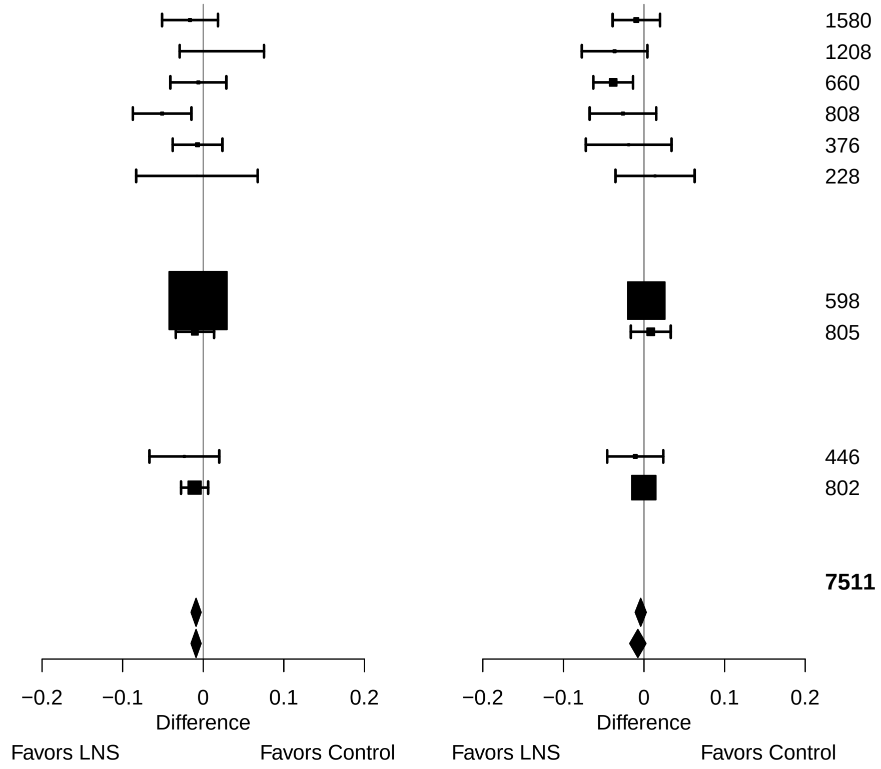

#### Supplemental figure 6F: Wasting prevalence difference

##### 6F4: Stratified by Maternal education

**P−for−interaction = 0.380**
**Difference in PDs = −0.01 (−0.02, 0.01)**

| | | Primary or greater | | | | | | Incomplete or no formal | | | | | |
|---|---|---|---|---|---|---|---|---|---|---|---|---|---|
| **Country** | **Trial** | **LNS N** | **Control N** | **Control Prevalence** | **PD (95% CI)** | **Fixed W** | **Random W** | **LNS N** | **Control N** | **Control Prevalence** | **PD (95% CI)** | **Fixed W** | **Random W** |
| Bangladesh | JiVitA−4 (34) | 1797 | 739 | 15.2 | −0.01 (−0.04, 0.02) | 0.08 | 0.13 | 984 | 475 | 18.3 | −0.02 (−0.06, 0.02) | 0.05 | 0.05 |
| Bangladesh | RDNS (35) | 1245 | 589 | 14.9 | −0.03 (−0.06, 0.01) | 0.04 | 0.09 | 416 | 226 | 15.5 | 0.00 (−0.05, 0.06) | 0.03 | 0.03 |
| Bangladesh | WASH−B (36) | 813 | 2447 | 9.9 | −0.02 (−0.04, 0.00) | 0.15 | 0.20 | 343 | 977 | 14.4 | −0.03 (−0.07, 0.01) | 0.04 | 0.04 |
| Burkina Faso | iLiNS−Zinc (37) | | | | | | | | | | | | |
| Burkina Faso | PROMIS (38) | | | | | | | | | | | | |
| Burkina Faso | PROMIS CS (38) | | | | | | | | | | | | |
| Ghana | GHANA (39) | | | | | | | | | | | | |
| Ghana | iLiNS−DYADG (40) | 267 | 548 | 6.6 | 0.00 (−0.04, 0.03) | 0.04 | 0.09 | 80 | 144 | 11.1 | −0.05 (−0.13, 0.03) | 0.01 | 0.01 |
| Haiti | HAITI (41) | | | | | | | | | | | | |
| Kenya | WASH−B (42) | 700 | 2426 | 1.2 | 0.00 (−0.01, 0.01) | 0.59 | 0.32 | 753 | 2688 | 1.8 | 0.00 (−0.01, 0.01) | 0.68 | 0.68 |
| Madagascar | MAHAY (43) | 351 | 431 | 3.2 | 0.00 (−0.02, 0.03) | 0.09 | 0.15 | 1349 | 1251 | 6.7 | −0.01 (−0.03, 0.02) | 0.11 | 0.11 |
| Malawi | iLiNS−DYADM (44) | | | | | | | | | | | | |
| Malawi | iLiNS−DOSE (45) | | | | | | | | | | | | |
| Mali | PROMIS (46) | | | | | | | | | | | | |
| Mali | PROMIS CS (46) | 107 | 96 | 9.4 | −0.05 (−0.12, 0.02) | 0.01 | 0.03 | 836 | 863 | 10.4 | −0.01 (−0.04, 0.01) | 0.09 | 0.09 |
| Zimbabwe | SHINE (HIV−) (47) | | | | | | | | | | | | |
| Zimbabwe | SHINE (HIV+) (48) | | | | | | | | | | | | |
| | | **5280** | **7276** | | **I² = 0.27, Tau² = 0.00** | | | **4761** | **6624** | | **I² = 0.00, Tau² = 0.00** | | |
| **Fixed** | | | | | **−0.01 (−0.01, 0.00)** | | | | | | **−0.01 (−0.02, 0.00)** | | |
| **Random** | | | | | **−0.01 (−0.02, 0.00)** | | | | | | **−0.01 (−0.02, 0.00)** | | |

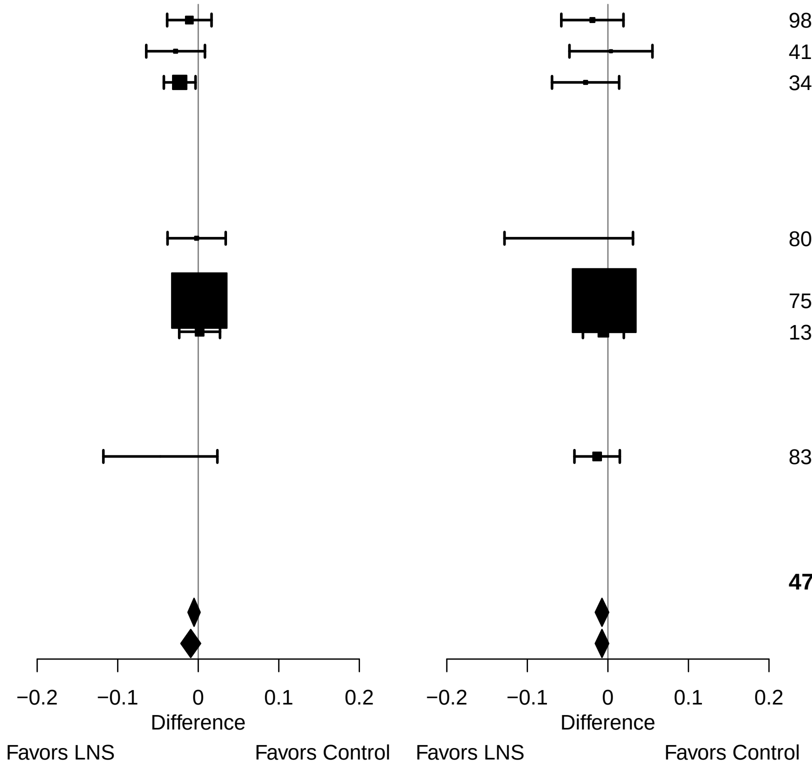

#### Supplemental figure 6F: Wasting prevalence difference

#### 6F5: Stratified by Maternal depressive symptoms

**−for−interaction = 0.511**
**ifference in PDs = 0.01 (−0.01, 0.02)**

| | | Less than 75th percentile | | | | | | At least 75th percentile | | | | | |
|---|---|---|---|---|---|---|---|---|---|---|---|---|---|
| **ountry** | **Trial** | **LNS N** | **Control N** | **Control Prevalence** | **PD (95% CI)** | **Fixed W** | **Random W** | **LNS N** | **Control N** | **Control Prevalence** | **PD (95% CI)** | **Fixed W** | **Random W** |
| angladesh | JiVitA−4 (34) | | | | | | | | | | | | |
| angladesh | RDNS (35) | 1081 | 472 | 13.6 | 0.00 (−0.04, 0.04) | 0.06 | 0.06 | 519 | 288 | 16.7 | −0.05 (−0.11, 0.01) | 0.05 | 0.05 |
| angladesh | WASH−B (36) | 895 | 2457 | 10.9 | −0.02 (−0.04, 0.00) | 0.18 | 0.18 | 237 | 876 | 12.0 | −0.03 (−0.07, 0.01) | 0.12 | 0.12 |
| urkina Faso | iLiNS−Zinc (37) | | | | | | | | | | | | |
| urkina Faso | PROMIS (38) | 505 | 569 | 8.3 | −0.01 (−0.05, 0.02) | 0.07 | 0.07 | 219 | 212 | 12.7 | −0.04 (−0.10, 0.03) | 0.04 | 0.04 |
| urkina Faso | PROMIS CS (38) | | | | | | | | | | | | |
| hana | GHANA (39) | | | | | | | | | | | | |
| hana | iLiNS−DYADG (40) | 252 | 459 | 6.8 | −0.01 (−0.05, 0.03) | 0.05 | 0.05 | 83 | 209 | 8.1 | 0.00 (−0.07, 0.07) | 0.04 | 0.04 |
| aiti | HAITI (41) | | | | | | | | | | | | |
| enya | WASH−B (42) | | | | | | | | | | | | |
| adagascar | MAHAY (43) | 672 | 721 | 5.8 | 0.00 (−0.03, 0.02) | 0.10 | 0.10 | 283 | 252 | 4.0 | 0.01 (−0.03, 0.04) | 0.16 | 0.16 |
| alawi | iLiNS−DYADM (44) | | | | | | | | | | | | |
| alawi | iLiNS−DOSE (45) | | | | | | | | | | | | |
| ali | PROMIS (46) | | | | | | | | | | | | |
| ali | PROMIS CS (46) | | | | | | | | | | | | |
| imbabwe | SHINE (HIV−) (47) | 1192 | 1136 | 2.7 | 0.00 (−0.02, 0.01) | 0.45 | 0.45 | 509 | 489 | 2.7 | 0.00 (−0.02, 0.01) | 0.56 | 0.56 |
| imbabwe | SHINE (HIV+) (48) | 234 | 214 | 2.3 | 0.02 (−0.01, 0.05) | 0.08 | 0.08 | 88 | 100 | 6.0 | 0.00 (−0.07, 0.07) | 0.03 | 0.03 |
| | | **4831** | **6028** | | **I² = 0.00, Tau² = 0.00** | | | **1938** | **2426** | | **I² = 0.00, Tau² = 0.00** | | |
| **ixed** | | | | | **−0.01 (−0.01, 0.00)** | | | | | | **−0.01 (−0.02, 0.00)** | | |
| **andom** | | | | | **−0.01 (−0.01, 0.00)** | | | | | | **−0.01 (−0.02, 0.00)** | | |

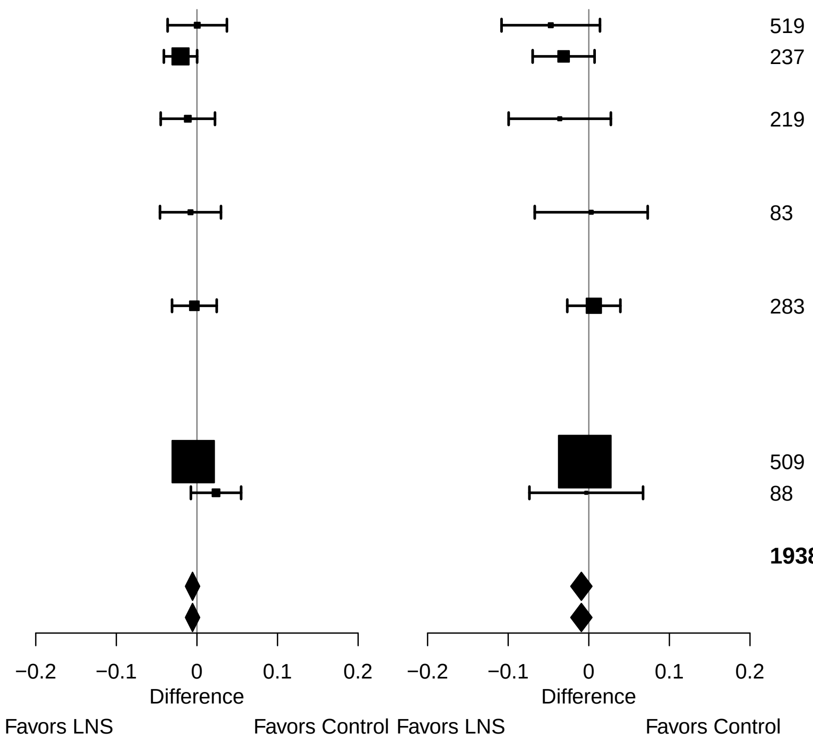

#### Supplemental figure 6F: Wasting prevalence difference

#### 6F6: Stratified by Child sex

**P−for−interaction = 0.876**
**Difference in PDs = 0.00 (−0.01, 0.01)**

| Country | Trial | Male LNS N | Male Control N | Male Control Prevalence | Male PD (95% CI) | Male Fixed W | Male Random W | Female LNS N | Female Control N | Female Control Prevalence | Female PD (95% CI) | Female Fixed W | Female Random W |
|---|---|---|---|---|---|---|---|---|---|---|---|---|---|
| Bangladesh | JiVitA−4 (34) | 1420 | 586 | 15.6 | 0.01 (−0.03, 0.04) | 0.06 | 0.06 | 1363 | 631 | 17.1 | −0.04 (−0.07, −0.01) | 0.03 | 0.09 |
| Bangladesh | RDNS (35) | 832 | 411 | 15.6 | −0.02 (−0.06, 0.02) | 0.04 | 0.04 | 829 | 404 | 14.6 | −0.02 (−0.07, 0.02) | 0.02 | 0.05 |
| Bangladesh | WASH−B (36) | 564 | 1711 | 12.2 | −0.01 (−0.04, 0.02) | 0.08 | 0.08 | 592 | 1713 | 10.3 | −0.04 (−0.06, −0.01) | 0.05 | 0.10 |
| Burkina Faso | iLiNS−Zinc (37) | 990 | 336 | 14.9 | −0.03 (−0.06, 0.00) | 0.06 | 0.06 | 962 | 328 | 12.2 | −0.05 (−0.09, −0.02) | 0.02 | 0.07 |
| Burkina Faso | PROMIS (38) | 434 | 480 | 12.5 | −0.04 (−0.09, 0.02) | 0.02 | 0.02 | 422 | 427 | 4.9 | 0.01 (−0.01, 0.04) | 0.04 | 0.09 |
| Burkina Faso | PROMIS CS (38) | 234 | 221 | 13.6 | 0.02 (−0.05, 0.10) | 0.01 | 0.01 | 196 | 215 | 11.2 | −0.02 (−0.08, 0.04) | 0.01 | 0.03 |
| Ghana | GHANA (39) | | | | | | | | | | | | |
| Ghana | iLiNS−DYADG (40) | 175 | 323 | 9.9 | −0.04 (−0.09, 0.02) | 0.02 | 0.02 | 172 | 369 | 5.4 | 0.01 (−0.03, 0.05) | 0.02 | 0.06 |
| Haiti | HAITI (41) | | | | | | | | | | | | |
| Kenya | WASH−B (42) | 732 | 2431 | 2.0 | 0.00 (−0.01, 0.01) | 0.37 | 0.37 | 723 | 2687 | 1.0 | 0.00 (−0.01, 0.00) | 0.53 | 0.17 |
| Madagascar | MAHAY (43) | 824 | 832 | 8.1 | −0.01 (−0.04, 0.03) | 0.06 | 0.06 | 876 | 850 | 3.6 | 0.00 (−0.02, 0.02) | 0.08 | 0.12 |
| Malawi | iLiNS−DYADM (44) | | | | | | | | | | | | |
| Malawi | iLiNS−DOSE (45) | | | | | | | | | | | | |
| Mali | PROMIS (46) | | | | | | | | | | | | |
| Mali | PROMIS CS (46) | 472 | 520 | 11.5 | −0.01 (−0.05, 0.04) | 0.03 | 0.03 | 472 | 439 | 8.9 | −0.03 (−0.06, 0.01) | 0.02 | 0.07 |
| Zimbabwe | SHINE (HIV−) (47) | 924 | 905 | 3.3 | −0.01 (−0.02, 0.01) | 0.26 | 0.26 | 945 | 879 | 1.8 | 0.00 (−0.01, 0.01) | 0.18 | 0.15 |
| Zimbabwe | SHINE (HIV+) (48) | | | | | | | | | | | | |
| | | **7601** | **8756** | | **I² = 0.00, Tau² = 0.00** | | | **7552** | **8942** | | **I² = 0.57, Tau² = 0.00** | | |
| **Fixed** | | | | | **−0.01 (−0.01, 0.00)** | | | | | | **−0.01 (−0.01, 0.00)** | | |
| **Random** | | | | | **−0.01 (−0.01, 0.00)** | | | | | | **−0.01 (−0.02, 0.00)** | | |

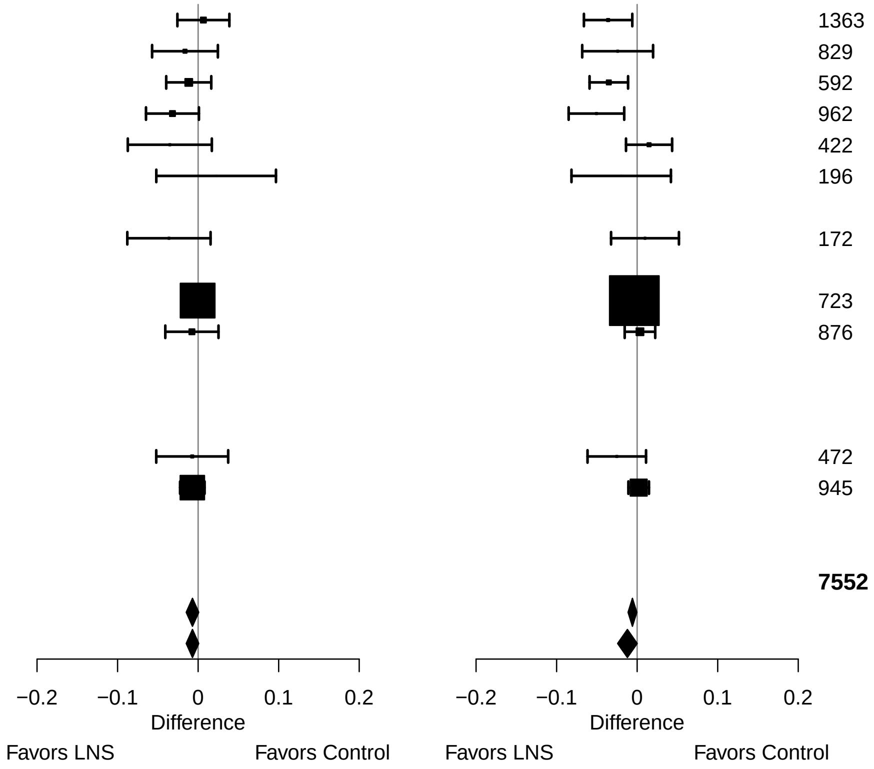

#### Supplemental figure 6F: Wasting prevalence difference

### 6F7: Stratified by Child birth order

**P−for−interaction = 0.549**
**Difference in PDs = 0.00 (−0.01, 0.01)**

| | | Later born | | | | | | Firstborn | | | | | |
|---|---|---|---|---|---|---|---|---|---|---|---|---|---|
| **Country** | **Trial** | **LNS N** | **Control N** | **Control Prevalence** | **PD (95% CI)** | **Fixed W** | **Random W** | **LNS N** | **Control N** | **Control Prevalence** | **PD (95% CI)** | **Fixed W** | **Random W** |
| Bangladesh | JiVitA−4 (34) | 598 | 247 | 17.1 | −0.02 (−0.06, 0.03) | 0.01 | 0.03 | 2181 | 964 | 16.2 | −0.01 (−0.04, 0.01) | 0.12 | 0.12 |
| Bangladesh | RDNS (35) | 972 | 508 | 15.7 | −0.02 (−0.06, 0.02) | 0.02 | 0.05 | 688 | 307 | 14.0 | −0.02 (−0.06, 0.03) | 0.03 | 0.03 |
| Bangladesh | WASH−B (36) | 735 | 2285 | 11.3 | −0.02 (−0.05, 0.00) | 0.05 | 0.09 | 421 | 1139 | 11.0 | −0.02 (−0.05, 0.01) | 0.08 | 0.08 |
| Burkina Faso | iLiNS−Zinc (37) | 1520 | 519 | 13.7 | −0.05 (−0.07, −0.02) | 0.05 | 0.09 | 432 | 145 | 13.1 | −0.02 (−0.07, 0.03) | 0.03 | 0.03 |
| Burkina Faso | PROMIS (38) | 708 | 747 | 9.4 | −0.01 (−0.04, 0.02) | 0.03 | 0.07 | 148 | 158 | 7.0 | −0.02 (−0.06, 0.03) | 0.03 | 0.03 |
| Burkina Faso | PROMIS CS (38) | 366 | 357 | 11.5 | 0.02 (−0.04, 0.07) | 0.01 | 0.03 | 63 | 79 | 16.5 | −0.05 (−0.15, 0.04) | 0.01 | 0.01 |
| Ghana | GHANA (39) | | | | | | | | | | | | |
| Ghana | iLiNS−DYADG (40) | | | | | | | | | | | | |
| Haiti | HAITI (41) | | | | | | | | | | | | |
| Kenya | WASH−B (42) | 1132 | 4027 | 1.5 | 0.00 (−0.01, 0.01) | 0.52 | 0.27 | 321 | 1086 | 1.5 | 0.00 (−0.01, 0.02) | 0.35 | 0.35 |
| Madagascar | MAHAY (43) | 1270 | 1217 | 6.4 | −0.01 (−0.03, 0.02) | 0.05 | 0.10 | 430 | 465 | 4.3 | 0.01 (−0.02, 0.03) | 0.13 | 0.13 |
| Malawi | iLiNS−DYADM (44) | | | | | | | | | | | | |
| Malawi | iLiNS−DOSE (45) | | | | | | | | | | | | |
| Mali | PROMIS (46) | | | | | | | | | | | | |
| Mali | PROMIS CS (46) | 783 | 821 | 10.5 | −0.02 (−0.04, 0.01) | 0.04 | 0.08 | 144 | 126 | 9.5 | −0.05 (−0.11, 0.01) | 0.02 | 0.02 |
| Zimbabwe | SHINE (HIV−) (47) | 1295 | 1218 | 2.5 | 0.00 (−0.02, 0.01) | 0.21 | 0.21 | 487 | 483 | 2.9 | 0.00 (−0.02, 0.02) | 0.19 | 0.19 |
| Zimbabwe | SHINE (HIV+) (48) | | | | | | | | | | | | |
| | | **9379** | **11946** | | **I² = 0.38, Tau² = 0.00** | | | **5315** | **4952** | | **I² = 0.00, Tau² = 0.00** | | |
| **Fixed** | | | | | **−0.01 (−0.01, 0.00)** | | | | | | **−0.01 (−0.01, 0.00)** | | |
| **Random** | | | | | **−0.01 (−0.02, 0.00)** | | | | | | **−0.01 (−0.01, 0.00)** | | |

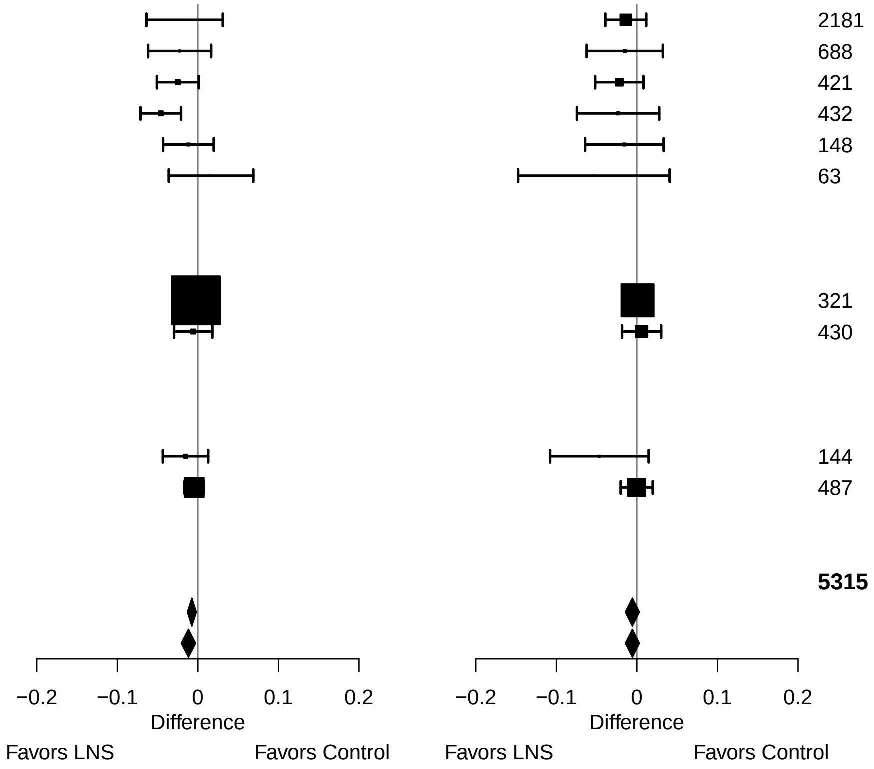

#### Supplemental figure 6F: Wasting prevalence difference

### 6F8: Stratified by Child baseline anthropometric status

**P−for−interaction = 0.062**
**Difference in PDs = −0.02 (−0.04, 0.00)**

| | | WLZ at least 0 Z | | | | | | WLZ less than 0 Z | | | | | |
|---|---|---|---|---|---|---|---|---|---|---|---|---|---|
| **Country** | **Trial** | **LNS N** | **Control N** | **Control Prevalence** | **PD (95% CI)** | **Fixed W** | **Random W** | **LNS N** | **Control N** | **Control Prevalence** | **PD (95% CI)** | **Fixed W** | **Random W** |
| Bangladesh | JiVitA−4 (34) | 961 | 423 | 2.4 | 0.00 (−0.02, 0.01) | 0.21 | 0.21 | 1822 | 794 | 23.9 | −0.02 (−0.06, 0.01) | 0.32 | 0.32 |
| Bangladesh | RDNS (35) | | | | | | | | | | | | |
| Bangladesh | WASH−B (36) | | | | | | | | | | | | |
| Burkina Faso | iLiNS−Zinc (37) | | | | | | | | | | | | |
| Burkina Faso | PROMIS (38) | 279 | 350 | 2.0 | 0.00 (−0.03, 0.02) | 0.12 | 0.12 | 413 | 393 | 14.2 | −0.03 (−0.08, 0.02) | 0.16 | 0.16 |
| Burkina Faso | PROMIS CS (38) | | | | | | | | | | | | |
| Ghana | GHANA (39) | | | | | | | | | | | | |
| Ghana | iLiNS−DYADG (40) | | | | | | | | | | | | |
| Haiti | HAITI (41) | | | | | | | | | | | | |
| Kenya | WASH−B (42) | | | | | | | | | | | | |
| Madagascar | MAHAY (43) | | | | | | | | | | | | |
| Malawi | iLiNS−DYADM (44) | | | | | | | | | | | | |
| Malawi | iLiNS−DOSE (45) | | | | | | | | | | | | |
| Mali | PROMIS (46) | | | | | | | | | | | | |
| Mali | PROMIS CS (46) | | | | | | | | | | | | |
| Zimbabwe | SHINE (HIV−) (47) | 743 | 702 | 1.0 | 0.00 (−0.01, 0.01) | 0.68 | 0.68 | 555 | 479 | 4.8 | −0.02 (−0.04, 0.01) | 0.52 | 0.52 |
| Zimbabwe | SHINE (HIV+) (48) | | | | | | | | | | | | |
| | | **1983** | **1475** | | **I² = 0.00, Tau² = 0.00** | | | **2790** | **1666** | | **I² = 0.00, Tau² = 0.00** | | |
| **Fixed** | | | | | **0.00 (−0.01, 0.01)** | | | | | | **−0.02 (−0.04, 0.00)** | | |
| **Random** | | | | | **0.00 (−0.01, 0.01)** | | | | | | **−0.02 (−0.04, 0.00)** | | |

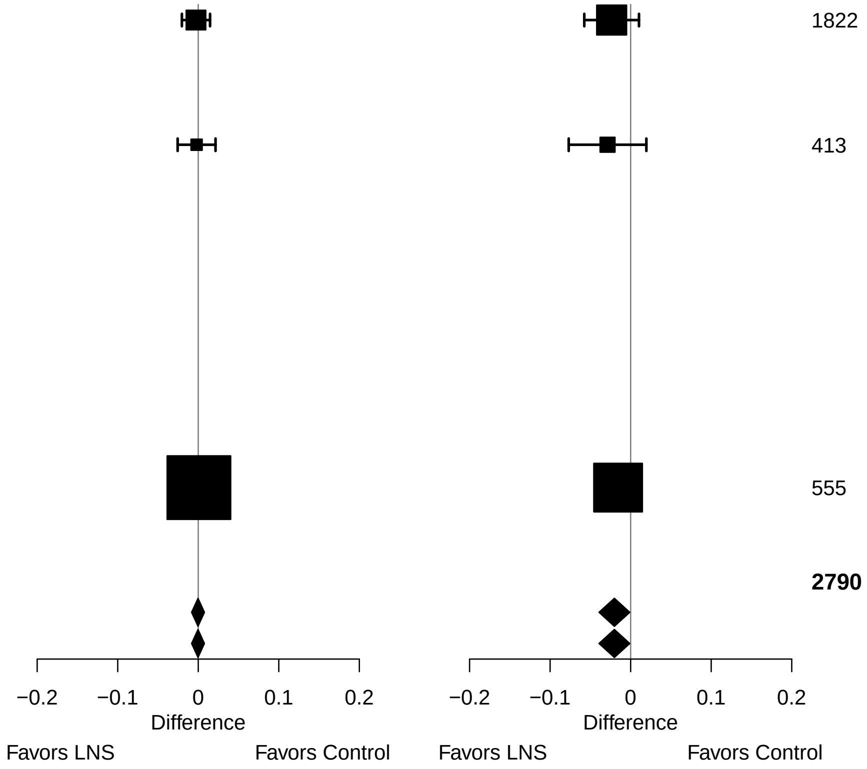

### Supplemental figure 6G: Mean difference in MUACZ

#### 6G1: Stratified by Maternal height

**P−for−interaction = 0.016**
**Difference in MDs = −0.09 (−0.17, −0.02)**

| | | At least 150.1 cm | | | | | | | Less than 150.1 cm | | | | |
|---|---|---|---|---|---|---|---|---|---|---|---|---|---|
| **Country** | **Trial** | **LNS N** | **Control N** | **Control Mean** | **MD (95% CI)** | **Fixed W** | **Random W** | **LNS N** | **Control N** | **Control Mean** | **MD (95% CI)** | **Fixed W** | **Random W** |
| Bangladesh | JiVitA−4 (34) | | | | | | | | | | | | |
| Bangladesh | RDNS (35) | 868 | 432 | −0.65 | 0.06 (−0.02, 0.14) | 0.12 | 0.10 | 735 | 358 | −0.79 | 0.05 (−0.08, 0.17) | 0.38 | 0.28 |
| Bangladesh | WASH−B (36) | | | | | | | | | | | | |
| Burkina Faso | iLiNS−Zinc (37) | 1923 | 651 | −1.13 | 0.30 (0.21, 0.39) | 0.10 | 0.10 | 34 | 15 | −1.22 | 0.04 (−0.35, 0.44) | 0.04 | 0.05 |
| Burkina Faso | PROMIS (38) | 826 | 883 | −0.73 | 0.10 (−0.03, 0.23) | 0.05 | 0.07 | 25 | 24 | −0.98 | −0.13 (−0.56, 0.29) | 0.03 | 0.04 |
| Burkina Faso | PROMIS CS (38) | | | | | | | | | | | | |
| Ghana | GHANA (39) | | | | | | | | | | | | |
| Ghana | iLiNS−DYADG (40) | 326 | 646 | −0.46 | 0.09 (−0.03, 0.22) | 0.06 | 0.08 | 16 | 36 | −0.58 | −0.12 (−0.61, 0.36) | 0.02 | 0.03 |
| Haiti | HAITI (41) | | | | | | | | | | | | |
| Kenya | WASH−B (42) | 1316 | 4728 | −0.45 | 0.12 (0.05, 0.19) | 0.17 | 0.11 | 57 | 195 | −0.56 | −0.04 (−0.35, 0.27) | 0.06 | 0.08 |
| Madagascar | MAHAY (43) | 987 | 1021 | −0.71 | 0.00 (−0.13, 0.13) | 0.05 | 0.07 | 594 | 557 | −0.78 | −0.07 (−0.23, 0.09) | 0.22 | 0.21 |
| Malawi | iLiNS−DYADM (44) | 192 | 381 | 0.09 | 0.01 (−0.16, 0.18) | 0.03 | 0.05 | 28 | 60 | −0.02 | −0.39 (−0.88, 0.09) | 0.02 | 0.03 |
| Malawi | iLiNS−DOSE (45) | 583 | 197 | 0.06 | 0.06 (−0.07, 0.18) | 0.06 | 0.08 | 112 | 42 | −0.15 | 0.13 (−0.15, 0.41) | 0.07 | 0.09 |
| Mali | PROMIS (46) | 482 | 495 | −0.78 | 0.20 (0.07, 0.34) | 0.05 | 0.07 | 19 | 9 | −0.79 | −0.22 (−0.65, 0.21) | 0.03 | 0.04 |
| Mali | PROMIS CS (46) | 916 | 937 | −0.83 | 0.15 (0.04, 0.27) | 0.06 | 0.08 | 33 | 27 | −1.05 | −0.30 (−0.75, 0.14) | 0.03 | 0.04 |
| Zimbabwe | SHINE (HIV−) (47) | 1747 | 1684 | 0.00 | 0.07 (0.01, 0.13) | 0.21 | 0.12 | 76 | 62 | 0.13 | −0.16 (−0.43, 0.11) | 0.08 | 0.09 |
| Zimbabwe | SHINE (HIV+) (48) | 314 | 311 | −0.16 | 0.00 (−0.14, 0.14) | 0.04 | 0.07 | 18 | 12 | −0.85 | 0.68 (−0.02, 1.37) | 0.01 | 0.02 |
| | | **10480** | **12366** | | **I² = 0.63, Tau² = 0.00** | | | **1747** | **1397** | | **I² = 0.13, Tau² = 0.00** | | |
| **Fixed** | | | | | **0.10 (0.08, 0.13)** | | | | | | **−0.03 (−0.10, 0.05)** | | |
| **Random** | | | | | **0.10 (0.05, 0.15)** | | | | | | **−0.04 (−0.13, 0.05)** | | |

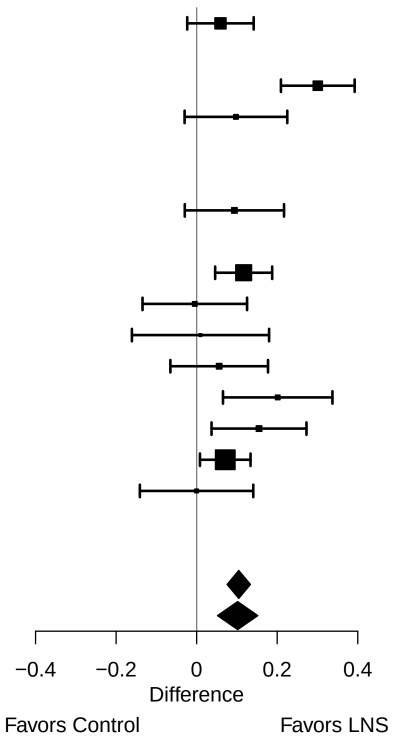

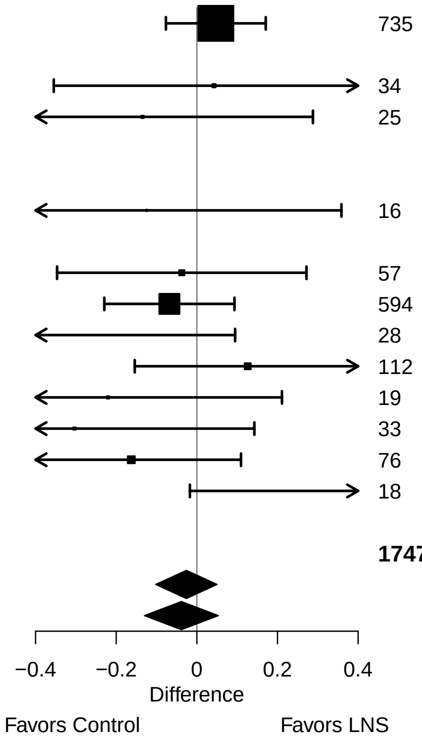

#### Supplemental figure 6G: Mean difference in MUACZ

##### 6G2: Stratified by Maternal BMI

**P−for−interaction = 0.580**
**Difference in MDs = −0.01 (−0.07, 0.04)**

| | | At least 20 kg/m² | | | | | | Less than 20 kg/m² | | | | | |
|---|---|---|---|---|---|---|---|---|---|---|---|---|---|
| **Country** | **Trial** | **LNS N** | **Control N** | **Control Mean** | **MD (95% CI)** | **Fixed W** | **Random W** | **LNS N** | **Control N** | **Control Mean** | **MD (95% CI)** | **Fixed W** | **Random W** |
| Bangladesh | JiVitA−4 (34) | | | | | | | | | | | | |
| Bangladesh | RDNS (35) | 730 | 335 | −0.57 | 0.07 (−0.03, 0.18) | 0.11 | 0.10 | 873 | 455 | −0.82 | 0.02 (−0.08, 0.12) | 0.19 | 0.15 |
| Bangladesh | WASH−B (36) | | | | | | | | | | | | |
| Burkina Faso | iLiNS−Zinc (37) | 1218 | 391 | −1.01 | 0.31 (0.19, 0.42) | 0.08 | 0.09 | 739 | 275 | −1.31 | 0.27 (0.17, 0.36) | 0.23 | 0.16 |
| Burkina Faso | PROMIS (38) | 641 | 673 | −0.66 | 0.11 (−0.02, 0.24) | 0.06 | 0.08 | 210 | 233 | −0.93 | 0.01 (−0.16, 0.18) | 0.07 | 0.09 |
| Burkina Faso | PROMIS CS (38) | 241 | 266 | −0.69 | −0.12 (−0.30, 0.06) | 0.03 | 0.06 | 183 | 166 | −1.00 | 0.05 (−0.15, 0.24) | 0.05 | 0.07 |
| Ghana | GHANA (39) | | | | | | | | | | | | |
| Ghana | iLiNS−DYADG (40) | 305 | 563 | −0.44 | 0.07 (−0.06, 0.20) | 0.06 | 0.08 | 37 | 119 | −0.61 | 0.12 (−0.21, 0.45) | 0.02 | 0.03 |
| Haiti | HAITI (41) | | | | | | | | | | | | |
| Kenya | WASH−B (42) | 1101 | 3837 | −0.38 | 0.07 (0.00, 0.15) | 0.19 | 0.11 | 272 | 1086 | −0.73 | 0.22 (0.09, 0.35) | 0.12 | 0.11 |
| Madagascar | MAHAY (43) | | | | | | | | | | | | |
| Malawi | iLiNS−DYADM (44) | 131 | 263 | 0.21 | −0.06 (−0.28, 0.15) | 0.02 | 0.05 | 88 | 178 | −0.12 | 0.00 (−0.24, 0.24) | 0.03 | 0.05 |
| Malawi | iLiNS−DOSE (45) | 515 | 174 | 0.17 | 0.05 (−0.08, 0.17) | 0.07 | 0.08 | 178 | 64 | −0.38 | 0.11 (−0.12, 0.33) | 0.04 | 0.06 |
| Mali | PROMIS (46) | 381 | 369 | −0.71 | 0.20 (0.06, 0.35) | 0.05 | 0.08 | 120 | 135 | −0.96 | 0.14 (−0.07, 0.34) | 0.05 | 0.06 |
| Mali | PROMIS CS (46) | 703 | 685 | −0.78 | 0.16 (0.04, 0.29) | 0.07 | 0.09 | 246 | 279 | −0.97 | 0.05 (−0.10, 0.20) | 0.09 | 0.10 |
| Zimbabwe | SHINE (HIV−) (47) | 1357 | 1294 | 0.03 | 0.09 (0.02, 0.16) | 0.21 | 0.12 | 240 | 249 | −0.19 | 0.06 (−0.10, 0.22) | 0.08 | 0.09 |
| Zimbabwe | SHINE (HIV+) (48) | 247 | 253 | −0.16 | 0.13 (−0.04, 0.30) | 0.04 | 0.07 | 62 | 52 | −0.38 | −0.06 (−0.32, 0.21) | 0.03 | 0.04 |
| | | **7570** | **9103** | | **I² = 0.56, Tau² = 0.01** | | | **3248** | **3291** | | **I² = 0.49, Tau² = 0.00** | | |
| **Fixed** | | | | | **0.10 (0.07, 0.13)** | | | | | | **0.12 (0.07, 0.16)** | | |
| **Random** | | | | | **0.10 (0.04, 0.16)** | | | | | | **0.10 (0.04, 0.16)** | | |

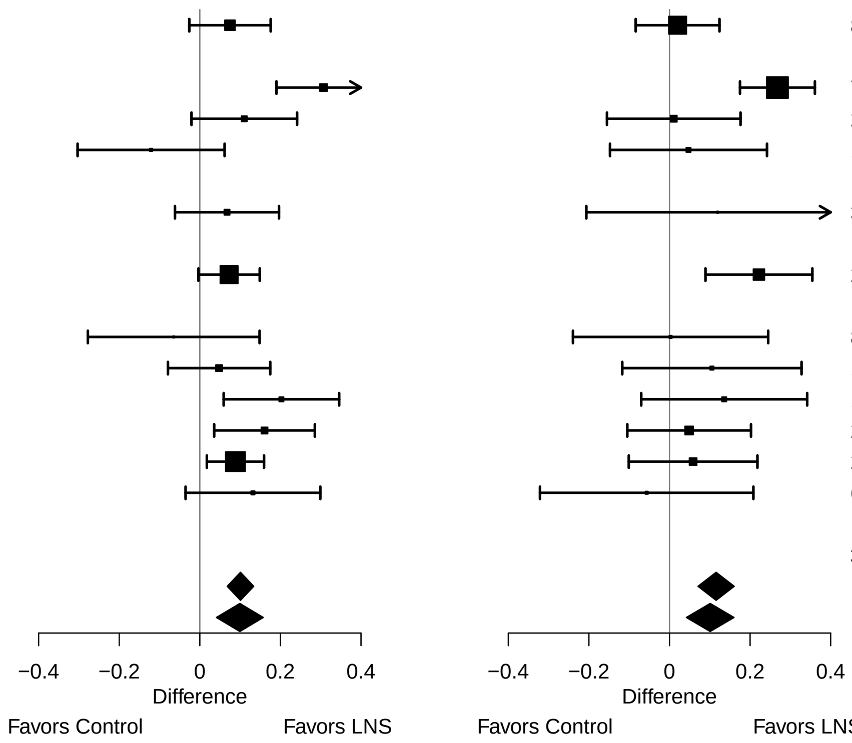

#### Supplemental figure 6G: Mean difference in MUACZ

#### 6G3: Stratified by Maternal age

**P−for−interaction = 0.395**
**Difference in MDs = −0.02 (−0.06, 0.02)**

| | | At least 25 y | | | | | | Less than 25 y | | | | | |
|---|---|---|---|---|---|---|---|---|---|---|---|---|---|
| **Country** | **Trial** | **LNS N** | **Control N** | **Control Mean** | **MD (95% CI)** | **Fixed W** | **Random W** | **LNS N** | **Control N** | **Control Mean** | **MD (95% CI)** | **Fixed W** | **Random W** |
| Bangladesh | JiVitA−4 (34) | 1244 | 555 | −1.07 | 0.09 (0.02, 0.15) | 0.25 | 0.12 | 1640 | 711 | −1.06 | 0.07 (0.01, 0.13) | 0.26 | 0.14 |
| Bangladesh | RDNS (35) | 453 | 221 | −0.70 | 0.02 (−0.13, 0.16) | 0.05 | 0.07 | 1210 | 594 | −0.70 | 0.05 (−0.03, 0.12) | 0.16 | 0.12 |
| Bangladesh | WASH−B (36) | | | | | | | | | | | | |
| Burkina Faso | iLiNS−Zinc (37) | 1134 | 401 | −1.13 | 0.31 (0.20, 0.42) | 0.08 | 0.08 | 812 | 262 | −1.12 | 0.26 (0.13, 0.40) | 0.05 | 0.07 |
| Burkina Faso | PROMIS (38) | 481 | 525 | −0.77 | 0.12 (−0.02, 0.26) | 0.05 | 0.07 | 373 | 387 | −0.69 | 0.05 (−0.13, 0.23) | 0.03 | 0.05 |
| Burkina Faso | PROMIS CS (38) | 200 | 190 | −0.89 | −0.03 (−0.20, 0.15) | 0.03 | 0.05 | 224 | 246 | −0.76 | −0.08 (−0.28, 0.12) | 0.02 | 0.04 |
| Ghana | GHANA (39) | | | | | | | | | | | | |
| Ghana | iLiNS−DYADG (40) | 220 | 417 | −0.43 | 0.03 (−0.13, 0.18) | 0.04 | 0.06 | 127 | 276 | −0.54 | 0.17 (0.00, 0.35) | 0.03 | 0.05 |
| Haiti | HAITI (41) | | | | | | | | | | | | |
| Kenya | WASH−B (42) | 845 | 2838 | −0.44 | 0.13 (0.05, 0.22) | 0.15 | 0.10 | 601 | 2315 | −0.48 | 0.09 (0.00, 0.18) | 0.12 | 0.11 |
| Madagascar | MAHAY (43) | 887 | 933 | −0.77 | −0.02 (−0.15, 0.11) | 0.06 | 0.07 | 792 | 725 | −0.69 | −0.05 (−0.20, 0.10) | 0.04 | 0.06 |
| Malawi | iLiNS−DYADM (44) | 106 | 226 | 0.10 | −0.04 (−0.28, 0.19) | 0.02 | 0.03 | 114 | 218 | 0.05 | −0.03 (−0.25, 0.19) | 0.02 | 0.04 |
| Malawi | iLiNS−DOSE (45) | 369 | 134 | 0.00 | 0.07 (−0.07, 0.21) | 0.05 | 0.07 | 313 | 99 | 0.08 | 0.01 (−0.17, 0.18) | 0.03 | 0.05 |
| Mali | PROMIS (46) | 347 | 358 | −0.74 | 0.17 (0.02, 0.33) | 0.04 | 0.06 | 160 | 147 | −0.86 | 0.22 (0.05, 0.40) | 0.03 | 0.05 |
| Mali | PROMIS CS (46) | 503 | 508 | −0.84 | 0.13 (−0.02, 0.28) | 0.04 | 0.06 | 451 | 462 | −0.81 | 0.13 (−0.01, 0.26) | 0.05 | 0.07 |
| Zimbabwe | SHINE (HIV−) (47) | 870 | 794 | 0.01 | 0.09 (0.00, 0.18) | 0.12 | 0.10 | 805 | 790 | 0.01 | 0.04 (−0.05, 0.13) | 0.13 | 0.11 |
| Zimbabwe | SHINE (HIV+) (48) | 244 | 230 | −0.17 | 0.00 (−0.17, 0.16) | 0.04 | 0.06 | 73 | 78 | −0.22 | 0.12 (−0.16, 0.40) | 0.01 | 0.02 |
| | | **7903** | **8330** | | **I² = 0.52, Tau² = 0.00** | | | **7695** | **7310** | | **I² = 0.35, Tau² = 0.00** | | |
| **Fixed** | | | | | **0.10 (0.07, 0.13)** | | | | | | **0.07 (0.04, 0.10)** | | |
| **Random** | | | | | **0.09 (0.04, 0.14)** | | | | | | **0.08 (0.03, 0.12)** | | |

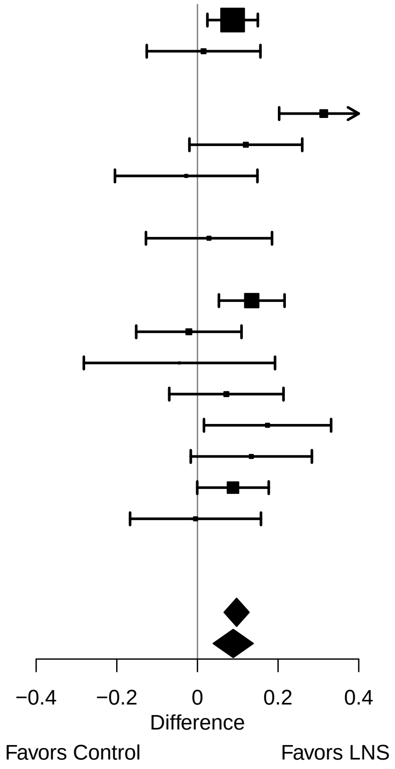

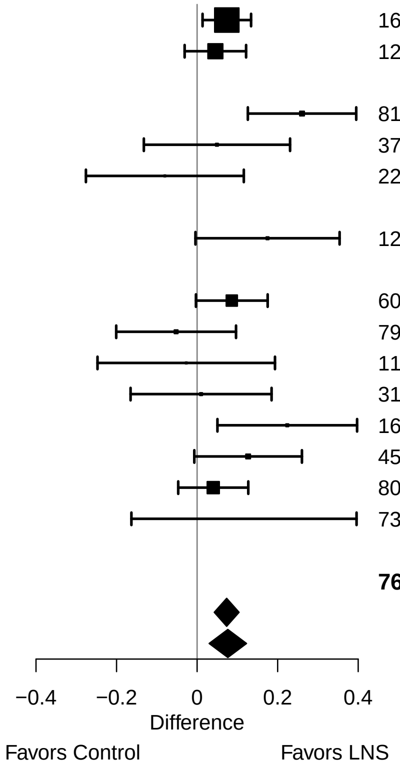

#### Supplemental figure 6G: Mean difference in MUACZ

#### 6G4: Stratified by Maternal education

**P−for−interaction = 0.688**
**Difference in MDs = 0.01 (−0.04, 0.06)**

| | | Primary or greater | | | | | | Incomplete or no formal | | | | | |
|---|---|---|---|---|---|---|---|---|---|---|---|---|---|
| Country | Trial | LNS N | Control N | Control Mean | MD (95% CI) | Fixed W | Random W | LNS N | Control N | Control Mean | MD (95% CI) | Fixed W | Random W |
| Bangladesh | JiVitA−4 (34) | 1865 | 777 | −0.98 | 0.08 (0.02, 0.13) | 0.30 | 0.21 | 1030 | 491 | −1.20 | 0.08 (0.01, 0.15) | 0.20 | 0.11 |
| Bangladesh | RDNS (35) | 1245 | 589 | −0.68 | 0.05 (−0.03, 0.14) | 0.13 | 0.14 | 418 | 226 | −0.76 | −0.02 (−0.16, 0.12) | 0.05 | 0.07 |
| Bangladesh | WASH−B (36) | | | | | | | | | | | | |
| Burkina Faso | iLiNS−Zinc (37) | 82 | 16 | −0.76 | 0.23 (−0.19, 0.66) | 0.01 | 0.01 | 1864 | 647 | −1.14 | 0.30 (0.21, 0.38) | 0.14 | 0.10 |
| Burkina Faso | PROMIS (38) | 69 | 54 | −0.78 | 0.09 (−0.19, 0.36) | 0.01 | 0.02 | 785 | 858 | −0.73 | 0.09 (−0.04, 0.22) | 0.06 | 0.08 |
| Burkina Faso | PROMIS CS (38) | 34 | 39 | −0.83 | −0.03 (−0.53, 0.48) | 0.00 | 0.01 | 388 | 396 | −0.82 | −0.06 (−0.22, 0.09) | 0.04 | 0.07 |
| Ghana | GHANA (39) | | | | | | | | | | | | |
| Ghana | iLiNS−DYADG (40) | 267 | 549 | −0.44 | 0.04 (−0.09, 0.18) | 0.05 | 0.08 | 80 | 144 | −0.60 | 0.24 (0.01, 0.47) | 0.02 | 0.04 |
| Haiti | HAITI (41) | | | | | | | | | | | | |
| Kenya | WASH−B (42) | 704 | 2460 | −0.36 | 0.10 (0.00, 0.19) | 0.11 | 0.13 | 755 | 2721 | −0.54 | 0.13 (0.06, 0.21) | 0.20 | 0.11 |
| Madagascar | MAHAY (43) | 344 | 420 | −0.48 | −0.08 (−0.28, 0.13) | 0.02 | 0.04 | 1336 | 1238 | −0.82 | 0.00 (−0.12, 0.12) | 0.07 | 0.08 |
| Malawi | iLiNS−DYADM (44) | 35 | 69 | 0.39 | −0.21 (−0.61, 0.19) | 0.01 | 0.01 | 184 | 372 | 0.01 | −0.01 (−0.18, 0.17) | 0.03 | 0.06 |
| Malawi | iLiNS−DOSE (45) | 165 | 56 | 0.34 | 0.13 (−0.09, 0.35) | 0.02 | 0.04 | 519 | 177 | −0.07 | 0.02 (−0.11, 0.15) | 0.06 | 0.08 |
| Mali | PROMIS (46) | 41 | 37 | −0.59 | −0.02 (−0.33, 0.30) | 0.01 | 0.02 | 465 | 468 | −0.79 | 0.21 (0.07, 0.34) | 0.05 | 0.07 |
| Mali | PROMIS CS (46) | 107 | 96 | −0.75 | 0.41 (0.17, 0.65) | 0.02 | 0.03 | 846 | 874 | −0.83 | 0.09 (−0.02, 0.21) | 0.07 | 0.09 |
| Zimbabwe | SHINE (HIV−) (47) | 1709 | 1630 | 0.02 | 0.05 (−0.01, 0.11) | 0.25 | 0.20 | 69 | 61 | −0.05 | 0.15 (−0.19, 0.49) | 0.01 | 0.02 |
| Zimbabwe | SHINE (HIV+) (48) | 301 | 292 | −0.16 | 0.01 (−0.13, 0.15) | 0.05 | 0.07 | 21 | 18 | −0.38 | 0.27 (−0.35, 0.88) | 0.00 | 0.01 |
| | | **6968** | **7084** | | **I² = 0.12, Tau² = 0.00** | | | **8760** | **8691** | | **I² = 0.66, Tau² = 0.01** | | |
| **Fixed** | | | | | **0.07 (0.03, 0.10)** | | | | | | **0.11 (0.08, 0.14)** | | |
| **Random** | | | | | **0.07 (0.02, 0.11)** | | | | | | **0.10 (0.04, 0.15)** | | |

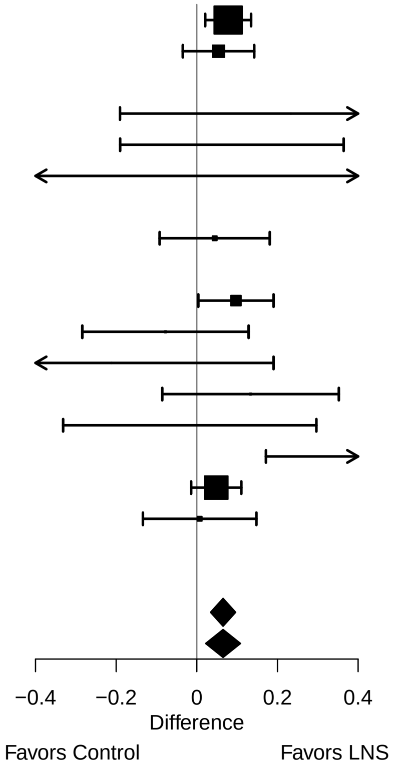

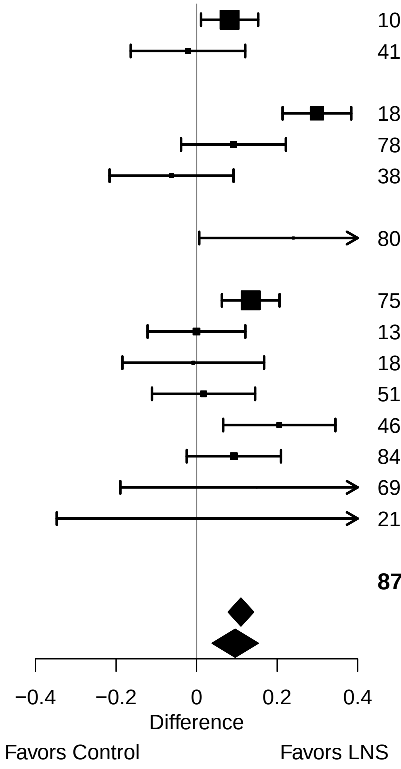

#### Supplemental figure 6G: Mean difference in MUACZ

##### 6G5: Stratified by Maternal depressive symptoms

**P−for−interaction** = **0.348**
**Difference in MDs =** **−0.03 (−0.09, 0.03)**

| | | Less than 75th percentile | | | | | | At least 75th percentile | | | | | |
|---|---|---|---|---|---|---|---|---|---|---|---|---|---|
| **Country** | **Trial** | **LNS N** | **Control N** | **Control Mean** | **MD (95% CI)** | **Fixed W** | **Random W** | **LNS N** | **Control N** | **Control Mean** | **MD (95% CI)** | **Fixed W** | **Random W** |
| Bangladesh | JiVitA−4 (34) | | | | | | | | | | | | |
| Bangladesh | RDNS (35) | 1081 | 472 | −0.68 | 0.03 (−0.07, 0.14) | 0.13 | 0.13 | 520 | 288 | −0.72 | 0.01 (−0.10, 0.13) | 0.21 | 0.14 |
| Bangladesh | WASH−B (36) | | | | | | | | | | | | |
| Burkina Faso | iLiNS−Zinc (37) | | | | | | | | | | | | |
| Burkina Faso | PROMIS (38) | 500 | 573 | −0.76 | 0.11 (−0.01, 0.24) | 0.09 | 0.09 | 224 | 215 | −0.77 | 0.07 (−0.10, 0.23) | 0.11 | 0.12 |
| Burkina Faso | PROMIS CS (38) | | | | | | | | | | | | |
| Ghana | GHANA (39) | | | | | | | | | | | | |
| Ghana | iLiNS−DYADG (40) | 252 | 460 | −0.43 | 0.08 (−0.06, 0.22) | 0.07 | 0.07 | 83 | 209 | −0.56 | 0.12 (−0.11, 0.36) | 0.05 | 0.09 |
| Haiti | HAITI (41) | | | | | | | | | | | | |
| Kenya | WASH−B (42) | 999 | 3523 | −0.43 | 0.10 (0.03, 0.18) | 0.27 | 0.27 | 340 | 1249 | −0.49 | 0.11 (−0.02, 0.23) | 0.20 | 0.14 |
| Madagascar | MAHAY (43) | 656 | 707 | −0.74 | 0.00 (−0.16, 0.17) | 0.05 | 0.05 | 282 | 245 | −0.63 | −0.17 (−0.35, 0.01) | 0.09 | 0.12 |
| Malawi | iLiNS−DYADM (44) | 157 | 303 | 0.13 | −0.08 (−0.27, 0.11) | 0.04 | 0.04 | 47 | 113 | −0.05 | 0.17 (−0.18, 0.51) | 0.02 | 0.06 |
| Malawi | iLiNS−DOSE (45) | | | | | | | | | | | | |
| Mali | PROMIS (46) | 379 | 373 | −0.72 | 0.11 (−0.04, 0.26) | 0.07 | 0.07 | 124 | 131 | −0.96 | 0.42 (0.20, 0.64) | 0.06 | 0.10 |
| Mali | PROMIS CS (46) | | | | | | | | | | | | |
| Zimbabwe | SHINE (HIV−) (47) | 1194 | 1135 | 0.03 | 0.03 (−0.05, 0.11) | 0.24 | 0.24 | 509 | 487 | −0.07 | 0.19 (0.07, 0.30) | 0.22 | 0.15 |
| Zimbabwe | SHINE (HIV+) (48) | 234 | 215 | −0.20 | 0.04 (−0.14, 0.21) | 0.05 | 0.05 | 89 | 99 | −0.17 | 0.02 (−0.26, 0.30) | 0.04 | 0.08 |
| | | **5452** | **7761** | | **I² = 0.00, Tau² = 0.00** | | | **2218** | **3036** | | **I² = 0.63, Tau² = 0.02** | | |
| **Fixed** | | | | | **0.06 (0.02, 0.10)** | | | | | | **0.09 (0.04, 0.15)** | | |
| **Random** | | | | | **0.06 (0.02, 0.10)** | | | | | | **0.10 (−0.01, 0.20)** | | |

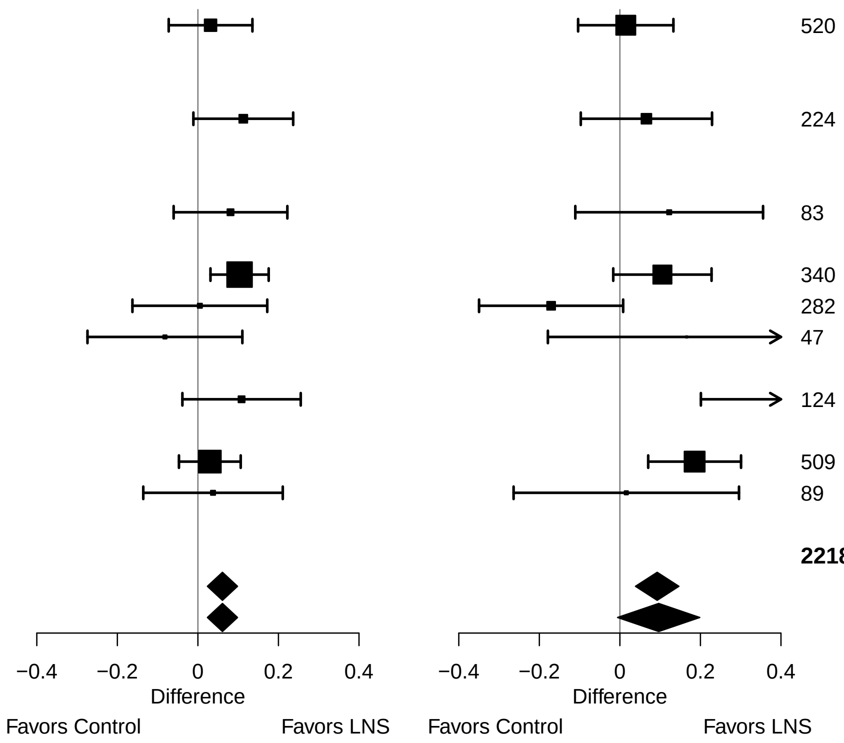

#### Supplemental figure 6G: Mean difference in MUACZ

#### 6G6: Stratified by Child sex

**P−for−interaction = 0.131**
**Difference in MDs = 0.03 (−0.01, 0.07)**

| | | Male | | | | | | Female | | | | | |
|---|---|---|---|---|---|---|---|---|---|---|---|---|---|
| **Country** | **Trial** | **LNS N** | **Control N** | **Control Mean** | **MD (95% CI)** | **Fixed W** | **Random W** | **LNS N** | **Control N** | **Control Mean** | **MD (95% CI)** | **Fixed W** | **Random W** |
| Bangladesh | JiVitA−4 (34) | 1477 | 612 | −1.02 | 0.05 (−0.01, 0.12) | 0.25 | 0.11 | 1420 | 659 | −1.10 | 0.11 (0.05, 0.17) | 0.25 | 0.13 |
| Bangladesh | RDNS (35) | 833 | 411 | −0.66 | −0.03 (−0.13, 0.07) | 0.10 | 0.09 | 830 | 404 | −0.75 | 0.10 (−0.01, 0.21) | 0.08 | 0.09 |
| Bangladesh | WASH−B (36) | | | | | | | | | | | | |
| Burkina Faso | iLiNS−Zinc (37) | 995 | 336 | −1.24 | 0.33 (0.22, 0.45) | 0.08 | 0.08 | 964 | 330 | −1.02 | 0.26 (0.14, 0.37) | 0.07 | 0.08 |
| Burkina Faso | PROMIS (38) | 432 | 482 | −0.80 | 0.11 (−0.03, 0.25) | 0.05 | 0.07 | 422 | 430 | −0.66 | 0.06 (−0.09, 0.22) | 0.04 | 0.06 |
| Burkina Faso | PROMIS CS (38) | 229 | 219 | −0.84 | −0.08 (−0.25, 0.09) | 0.03 | 0.06 | 195 | 217 | −0.79 | −0.03 (−0.25, 0.19) | 0.02 | 0.03 |
| Ghana | GHANA (39) | | | | | | | | | | | | |
| Ghana | iLiNS−DYADG (40) | 175 | 324 | −0.55 | 0.09 (−0.08, 0.26) | 0.03 | 0.06 | 172 | 369 | −0.40 | 0.09 (−0.07, 0.25) | 0.03 | 0.05 |
| Haiti | HAITI (41) | | | | | | | | | | | | |
| Kenya | WASH−B (42) | 736 | 2467 | −0.47 | 0.08 (−0.02, 0.18) | 0.11 | 0.09 | 725 | 2718 | −0.44 | 0.16 (0.08, 0.23) | 0.16 | 0.12 |
| Madagascar | MAHAY (43) | 817 | 817 | −0.80 | −0.07 (−0.21, 0.07) | 0.05 | 0.07 | 863 | 841 | −0.67 | 0.00 (−0.13, 0.14) | 0.05 | 0.07 |
| Malawi | iLiNS−DYADM (44) | 107 | 207 | 0.00 | 0.07 (−0.16, 0.30) | 0.02 | 0.04 | 113 | 237 | 0.14 | −0.13 (−0.36, 0.10) | 0.02 | 0.03 |
| Malawi | iLiNS−DOSE (45) | 351 | 123 | −0.12 | 0.02 (−0.14, 0.18) | 0.04 | 0.06 | 345 | 117 | 0.16 | 0.11 (−0.05, 0.26) | 0.04 | 0.06 |
| Mali | PROMIS (46) | 267 | 263 | −0.80 | 0.16 (0.00, 0.33) | 0.04 | 0.06 | 240 | 242 | −0.75 | 0.21 (0.06, 0.37) | 0.04 | 0.06 |
| Mali | PROMIS CS (46) | 479 | 526 | −0.90 | 0.11 (−0.06, 0.29) | 0.03 | 0.06 | 475 | 444 | −0.74 | 0.13 (0.01, 0.26) | 0.06 | 0.07 |
| Zimbabwe | SHINE (HIV−) (47) | 928 | 900 | −0.05 | 0.09 (0.01, 0.18) | 0.13 | 0.10 | 944 | 880 | 0.07 | 0.03 (−0.06, 0.11) | 0.12 | 0.10 |
| Zimbabwe | SHINE (HIV+) (48) | 162 | 167 | −0.25 | −0.01 (−0.19, 0.17) | 0.03 | 0.06 | 175 | 161 | −0.13 | 0.07 (−0.11, 0.25) | 0.03 | 0.05 |
| | | **7988** | **7854** | | **I² = 0.62, Tau² = 0.01** | | | **7883** | **8049** | | **I² = 0.41, Tau² = 0.00** | | |
| **Fixed** | | | | | **0.07 (0.04, 0.10)** | | | | | | **0.11 (0.08, 0.14)** | | |
| **Random** | | | | | **0.07 (0.01, 0.13)** | | | | | | **0.10 (0.05, 0.14)** | | |

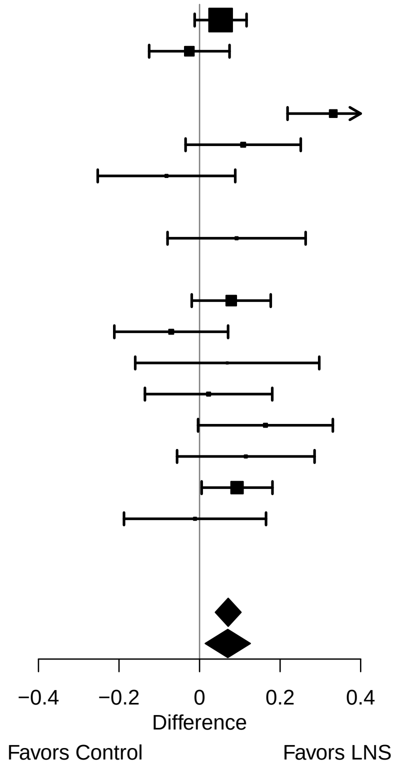

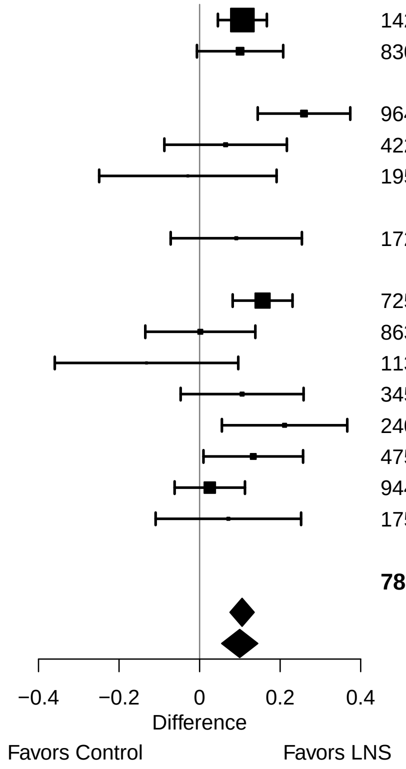

##### Supplemental figure 6G: Mean difference in MUACZ

#### 6G7: Stratified by Child birth order

**P−for−interaction = 0.004**
**Difference in MDs = −0.07 (−0.11, −0.02)**

**Later born**

| Country | Trial | LNS N | Control N | Control Mean | MD (95% CI) | Fixed W | Random W |
|---|---|---|---|---|---|---|---|
| Bangladesh | JiVitA−4 (34) | 617 | 260 | −1.16 | 0.18 (0.09, 0.27) | 0.11 | 0.09 |
| Bangladesh | RDNS (35) | 974 | 508 | −0.73 | 0.04 (−0.05, 0.14) | 0.09 | 0.08 |
| Bangladesh | WASH−B (36) | | | | | | |
| Burkina Faso | iLiNS−Zinc (37) | 1524 | 521 | −1.13 | 0.31 (0.21, 0.40) | 0.10 | 0.08 |
| Burkina Faso | PROMIS (38) | 709 | 753 | −0.74 | 0.11 (−0.03, 0.25) | 0.04 | 0.06 |
| Burkina Faso | PROMIS CS (38) | 362 | 356 | −0.84 | −0.03 (−0.20, 0.14) | 0.03 | 0.05 |
| Ghana | GHANA (39) | | | | | | |
| Ghana | iLiNS−DYADG (40) | 237 | 459 | −0.45 | 0.03 (−0.11, 0.18) | 0.04 | 0.06 |
| Haiti | HAITI (41) | | | | | | |
| Kenya | WASH−B (42) | 1137 | 4083 | −0.47 | 0.12 (0.04, 0.20) | 0.15 | 0.09 |
| Madagascar | MAHAY (43) | 1263 | 1199 | −0.78 | −0.01 (−0.13, 0.11) | 0.06 | 0.07 |
| Malawi | iLiNS−DYADM (44) | 173 | 353 | 0.10 | −0.03 (−0.21, 0.15) | 0.03 | 0.05 |
| Malawi | iLiNS−DOSE (45) | 459 | 164 | 0.00 | 0.06 (−0.07, 0.19) | 0.05 | 0.07 |
| Mali | PROMIS (46) | 443 | 444 | −0.77 | 0.20 (0.06, 0.34) | 0.04 | 0.06 |
| Mali | PROMIS CS (46) | 791 | 832 | −0.82 | 0.12 (0.01, 0.24) | 0.06 | 0.07 |
| Zimbabwe | SHINE (HIV−) (47) | 1296 | 1211 | −0.02 | 0.11 (0.04, 0.18) | 0.16 | 0.10 |
| Zimbabwe | SHINE (HIV+) (48) | 280 | 245 | −0.15 | −0.03 (−0.19, 0.13) | 0.03 | 0.06 |
| | | **10265** | **11388** | | **$I^2$ = 0.63, $Tau^2$ = 0.01** | | |
| **Fixed** | | | | | **0.11 (0.08, 0.14)** | | |
| **Random** | | | | | **0.10 (0.05, 0.15)** | | |

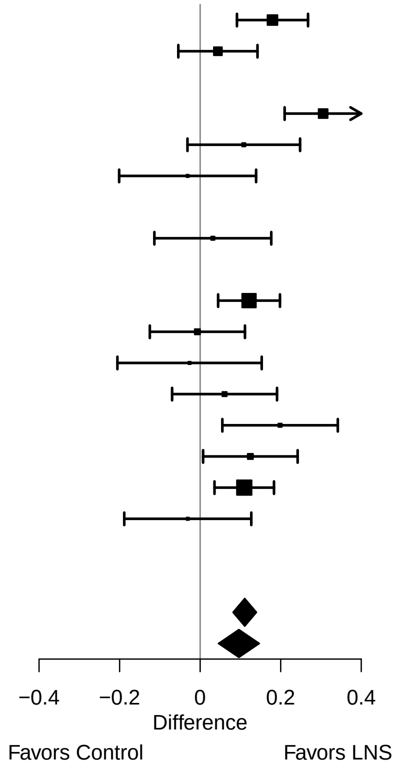

**Firstborn**

| Country | Trial | LNS N | Control N | Control Mean | MD (95% CI) | Fixed W | Random W |
|---|---|---|---|---|---|---|---|
| Bangladesh | JiVitA−4 (34) | 2276 | 1004 | −1.04 | 0.05 (0.00, 0.10) | 0.46 | 0.13 |
| Bangladesh | RDNS (35) | 688 | 307 | −0.66 | 0.02 (−0.08, 0.12) | 0.11 | 0.11 |
| Bangladesh | WASH−B (36) | | | | | | |
| Burkina Faso | iLiNS−Zinc (37) | 435 | 145 | −1.16 | 0.27 (0.10, 0.43) | 0.04 | 0.08 |
| Burkina Faso | PROMIS (38) | 145 | 158 | −0.69 | 0.00 (−0.20, 0.20) | 0.03 | 0.06 |
| Burkina Faso | PROMIS CS (38) | 61 | 80 | −0.70 | −0.20 (−0.48, 0.07) | 0.02 | 0.04 |
| Ghana | GHANA (39) | | | | | | |
| Ghana | iLiNS−DYADG (40) | 110 | 234 | −0.51 | 0.20 (0.00, 0.40) | 0.03 | 0.06 |
| Haiti | HAITI (41) | | | | | | |
| Kenya | WASH−B (42) | 322 | 1097 | −0.39 | 0.10 (−0.02, 0.22) | 0.08 | 0.10 |
| Madagascar | MAHAY (43) | 417 | 459 | −0.63 | −0.09 (−0.28, 0.10) | 0.03 | 0.07 |
| Malawi | iLiNS−DYADM (44) | 47 | 89 | −0.06 | −0.06 (−0.43, 0.31) | 0.01 | 0.03 |
| Malawi | iLiNS−DOSE (45) | 148 | 45 | 0.24 | −0.07 (−0.33, 0.19) | 0.02 | 0.05 |
| Mali | PROMIS (46) | 63 | 59 | −0.84 | 0.12 (−0.09, 0.32) | 0.03 | 0.06 |
| Mali | PROMIS CS (46) | 146 | 126 | −0.86 | 0.23 (0.03, 0.42) | 0.03 | 0.07 |
| Zimbabwe | SHINE (HIV−) (47) | 489 | 486 | 0.07 | −0.06 (−0.17, 0.05) | 0.10 | 0.10 |
| Zimbabwe | SHINE (HIV+) (48) | 54 | 78 | −0.29 | 0.27 (−0.03, 0.56) | 0.01 | 0.04 |
| | | **5401** | **4367** | | **$I^2$ = 0.50, $Tau^2$ = 0.01** | | |
| **Fixed** | | | | | **0.05 (0.02, 0.09)** | | |
| **Random** | | | | | **0.06 (−0.01, 0.13)** | | |

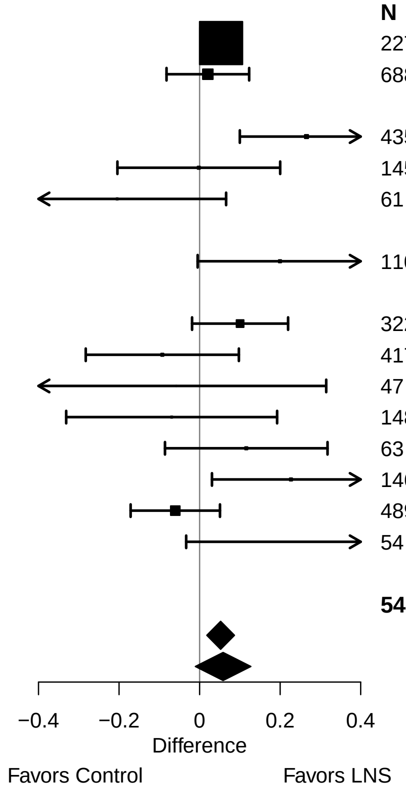

#### Supplemental figure 6G: Mean difference in MUACZ

#### 6G8: Stratified by Child baseline anthropometric status

**P−for−interaction = 0.177**
**Difference in MDs = 0.04 (−0.02, 0.10)**

| | | | | | MUACZ at least 0 Z | | | | | | MUACZ less than 0 Z | | |
|---|---|---|---|---|---|---|---|---|---|---|---|---|---|
| **Country** | **Trial** | **LNS N** | **Control N** | **Control Mean** | **MD (95% CI)** | **Fixed W** | **Random W** | **LNS N** | **Control N** | **Control Mean** | **MD (95% CI)** | **Fixed W** | **Random W** |
| Bangladesh | JiVitA−4 (34) | 696 | 295 | −0.26 | 0.08 (−0.02, 0.19) | 0.17 | 0.17 | 2201 | 976 | −1.31 | 0.08 (0.02, 0.14) | 0.34 | 0.14 |
| Bangladesh | RDNS (35) | 702 | 321 | −0.14 | −0.02 (−0.10, 0.06) | 0.28 | 0.28 | 899 | 439 | −1.10 | 0.04 (−0.05, 0.12) | 0.18 | 0.13 |
| Bangladesh | WASH−B (36) | | | | | | | | | | | | |
| Burkina Faso | iLiNS−Zinc (37) | 349 | 128 | −0.05 | 0.24 (0.04, 0.44) | 0.05 | 0.05 | 1610 | 538 | −1.39 | 0.36 (0.24, 0.48) | 0.10 | 0.12 |
| Burkina Faso | PROMIS (38) | | | | | | | | | | | | |
| Burkina Faso | PROMIS CS (38) | | | | | | | | | | | | |
| Ghana | GHANA (39) | | | | | | | | | | | | |
| Ghana | iLiNS−DYADG (40) | 149 | 290 | 0.10 | 0.08 (−0.08, 0.24) | 0.07 | 0.07 | 174 | 352 | −0.92 | 0.06 (−0.07, 0.20) | 0.07 | 0.11 |
| Haiti | HAITI (41) | | | | | | | | | | | | |
| Kenya | WASH−B (42) | | | | | | | | | | | | |
| Madagascar | MAHAY (43) | | | | | | | | | | | | |
| Malawi | iLiNS−DYADM (44) | 111 | 233 | 0.54 | −0.01 (−0.19, 0.17) | 0.06 | 0.06 | 93 | 193 | −0.50 | −0.07 (−0.29, 0.16) | 0.03 | 0.08 |
| Malawi | iLiNS−DOSE (45) | 397 | 141 | 0.46 | 0.01 (−0.16, 0.17) | 0.07 | 0.07 | 299 | 99 | −0.61 | 0.04 (−0.15, 0.23) | 0.04 | 0.09 |
| Mali | PROMIS (46) | 185 | 156 | −0.18 | 0.08 (−0.10, 0.25) | 0.06 | 0.06 | 322 | 349 | −1.05 | 0.20 (0.04, 0.37) | 0.05 | 0.10 |
| Mali | PROMIS CS (46) | | | | | | | | | | | | |
| Zimbabwe | SHINE (HIV−) (47) | 678 | 616 | 0.40 | 0.05 (−0.05, 0.14) | 0.21 | 0.21 | 640 | 578 | −0.46 | 0.14 (0.05, 0.23) | 0.17 | 0.13 |
| Zimbabwe | SHINE (HIV+) (48) | 123 | 96 | 0.29 | 0.10 (−0.12, 0.31) | 0.04 | 0.04 | 144 | 139 | −0.45 | −0.08 (−0.26, 0.11) | 0.04 | 0.09 |
| | | **3390** | **2276** | | **I² = 0.00, Tau² = 0.00** | | | **6382** | **3663** | | **I² = 0.73, Tau² = 0.01** | | |
| **Fixed** | | | | | **0.04 (0.00, 0.09)** | | | | | | **0.10 (0.07, 0.14)** | | |
| **Random** | | | | | **0.04 (0.00, 0.09)** | | | | | | **0.10 (0.01, 0.18)** | | |

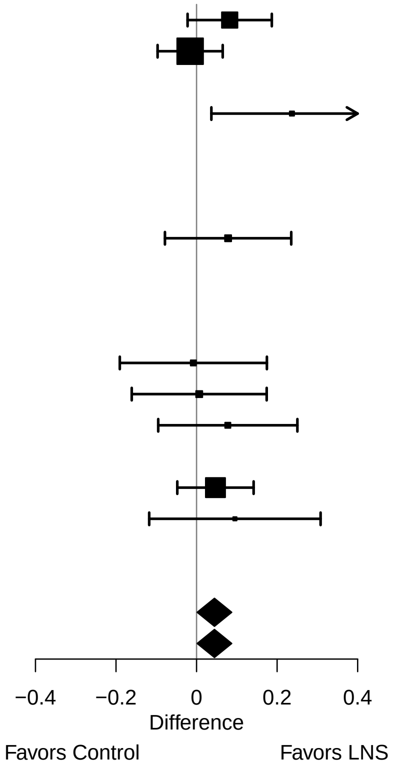

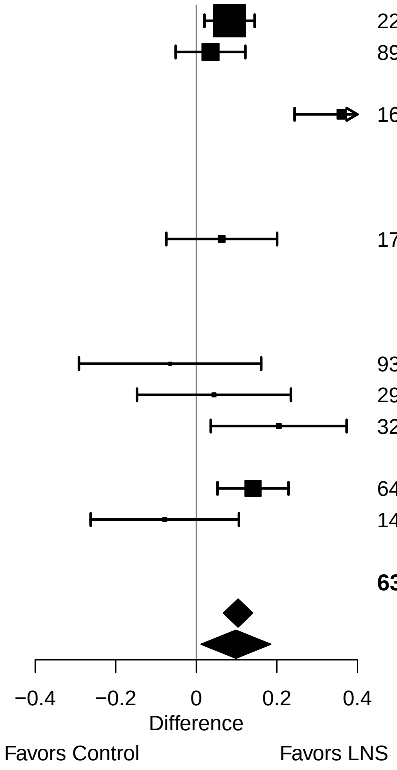

### Supplemental figure 6H: Low MUAC prevalence ratio

#### 6H1: Stratified by Maternal height (insufficient comparisons)

#### Supplemental figure 6H: Low MUAC prevalence ratio

##### 6H2: Stratified by Maternal BMI

**P−for−interaction = 0.337**
**Ratio of PRs = 1.12 (0.89, 1.40)**

| | | At least 20 kg/m² | | | | | | Less than 20 kg/m² | | | | | |
|---|---|---|---|---|---|---|---|---|---|---|---|---|---|
| **Country** | **Trial** | **LNS N** | **Control N** | **Control Prevalence** | **PR (95% CI)** | **Fixed W** | **Random W** | **LNS N** | **Control N** | **Control Prevalence** | **PR (95% CI)** | **Fixed W** | **Random W** |
| Bangladesh | JiVitA−4 (34) | | | | | | | | | | | | |
| Bangladesh | RDNS (35) | 730 | 335 | 3.0 | 0.78 (0.39, 1.54) | 0.05 | 0.07 | 873 | 455 | 7.0 | 1.11 (0.72, 1.71) | 0.14 | 0.14 |
| Bangladesh | WASH−B (36) | | | | | | | | | | | | |
| Burkina Faso | iLiNS−Zinc (37) | 1218 | 391 | 14.6 | 0.55 (0.42, 0.74) | 0.30 | 0.24 | 739 | 275 | 23.6 | 0.75 (0.57, 0.99) | 0.36 | 0.36 |
| Burkina Faso | PROMIS (38) | 641 | 673 | 7.1 | 0.92 (0.56, 1.50) | 0.10 | 0.12 | 210 | 233 | 10.3 | 1.02 (0.62, 1.67) | 0.11 | 0.11 |
| Burkina Faso | PROMIS CS (38) | 241 | 266 | 12.0 | 0.93 (0.54, 1.60) | 0.08 | 0.10 | 183 | 166 | 19.3 | 0.88 (0.58, 1.32) | 0.16 | 0.16 |
| Ghana | GHANA (39) | | | | | | | | | | | | |
| Ghana | iLiNS−DYADG (40) | | | | | | | | | | | | |
| Haiti | HAITI (41) | | | | | | | | | | | | |
| Kenya | WASH−B (42) | 1101 | 3837 | 4.4 | 0.89 (0.62, 1.29) | 0.18 | 0.18 | 272 | 1086 | 8.5 | 0.48 (0.25, 0.92) | 0.06 | 0.06 |
| Madagascar | MAHAY (43) | | | | | | | | | | | | |
| Malawi | iLiNS−DYADM (44) | | | | | | | | | | | | |
| Malawi | iLiNS−DOSE (45) | | | | | | | | | | | | |
| Mali | PROMIS (46) | 381 | 369 | 5.7 | 0.47 (0.23, 0.97) | 0.05 | 0.07 | 120 | 135 | 5.9 | 0.95 (0.39, 2.35) | 0.03 | 0.03 |
| Mali | PROMIS CS (46) | 703 | 685 | 10.1 | 0.95 (0.69, 1.30) | 0.24 | 0.21 | 246 | 279 | 13.6 | 0.84 (0.53, 1.32) | 0.13 | 0.13 |
| Zimbabwe | SHINE (HIV−) (47) | | | | | | | | | | | | |
| Zimbabwe | SHINE (HIV+) (48) | | | | | | | | | | | | |
| | | **5015** | **6556** | | **I² = 0.40, Tau² = 0.02** | | | **2643** | **2629** | | **I² = 0.00, Tau² = 0.00** | | |
| **Fixed** | | | | | **0.76 (0.65, 0.89)** | | | | | | **0.84 (0.71, 0.99)** | | |
| **Random** | | | | | **0.77 (0.63, 0.94)** | | | | | | **0.84 (0.71, 0.99)** | | |

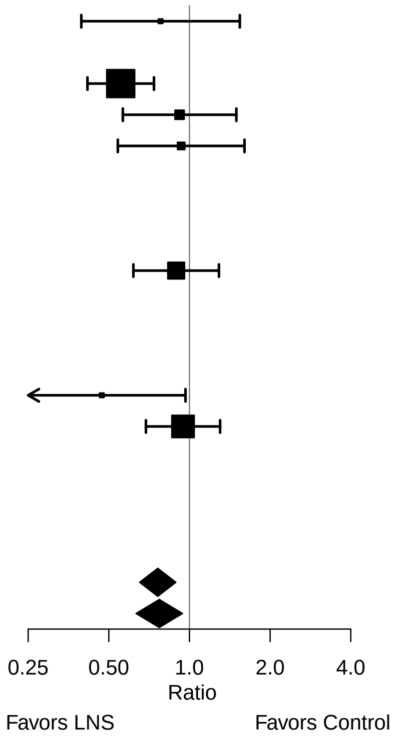

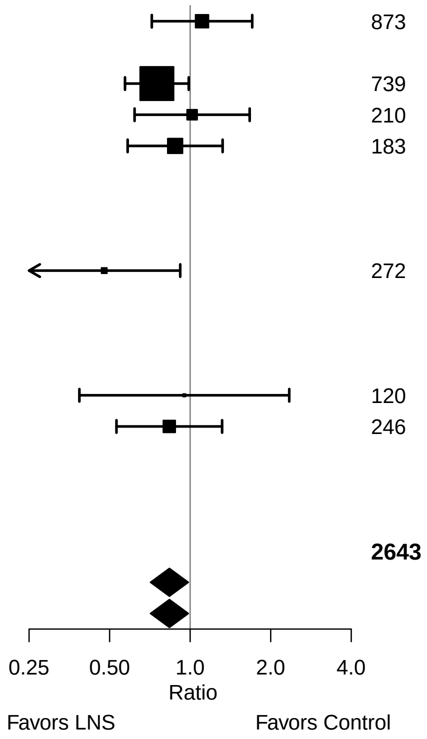

#### Supplemental figure 6H: Low MUAC prevalence ratio

##### 6H3: Stratified by Maternal age

**P−for−interaction = 0.741**
**Ratio of PRs = 1.03 (0.87, 1.22)**

| | | At least 25 y | | | | | | Less than 25 y | | | | | |
|---|---|---|---|---|---|---|---|---|---|---|---|---|---|
| **Country** | **Trial** | **LNS N** | **Control N** | **Control Prevalence** | **PR (95% CI)** | **Fixed W** | **Random W** | **LNS N** | **Control N** | **Control Prevalence** | **PR (95% CI)** | **Fixed W** | **Random W** |
| Bangladesh | JiVitA−4 (34) | 1244 | 555 | 15.7 | 0.82 (0.63, 1.07) | 0.23 | 0.15 | 1640 | 711 | 14.7 | 0.84 (0.67, 1.05) | 0.30 | 0.30 |
| Bangladesh | RDNS (35) | 453 | 221 | 5.0 | 1.24 (0.66, 2.34) | 0.04 | 0.07 | 1210 | 594 | 5.2 | 0.93 (0.61, 1.44) | 0.08 | 0.08 |
| Bangladesh | WASH−B (36) | | | | | | | | | | | | |
| Burkina Faso | iLiNS−Zinc (37) | 1134 | 401 | 18.5 | 0.59 (0.44, 0.81) | 0.17 | 0.14 | 812 | 262 | 17.9 | 0.69 (0.51, 0.94) | 0.15 | 0.15 |
| Burkina Faso | PROMIS (38) | 481 | 525 | 8.8 | 0.78 (0.51, 1.21) | 0.08 | 0.11 | 373 | 387 | 7.0 | 1.19 (0.65, 2.20) | 0.04 | 0.04 |
| Burkina Faso | PROMIS CS (38) | 200 | 190 | 15.8 | 0.95 (0.59, 1.54) | 0.07 | 0.10 | 224 | 246 | 14.2 | 0.88 (0.55, 1.41) | 0.06 | 0.06 |
| Ghana | GHANA (39) | | | | | | | | | | | | |
| Ghana | iLiNS−DYADG (40) | | | | | | | | | | | | |
| Haiti | HAITI (41) | | | | | | | | | | | | |
| Kenya | WASH−B (42) | 845 | 2838 | 5.0 | 0.69 (0.46, 1.02) | 0.10 | 0.12 | 601 | 2315 | 6.1 | 0.71 (0.47, 1.08) | 0.08 | 0.08 |
| Madagascar | MAHAY (43) | 887 | 933 | 10.1 | 1.07 (0.80, 1.45) | 0.18 | 0.14 | 792 | 725 | 9.1 | 0.86 (0.62, 1.19) | 0.14 | 0.14 |
| Malawi | iLiNS−DYADM (44) | | | | | | | | | | | | |
| Malawi | iLiNS−DOSE (45) | | | | | | | | | | | | |
| Mali | PROMIS (46) | 347 | 358 | 5.8 | 0.41 (0.17, 0.99) | 0.02 | 0.04 | 160 | 147 | 5.4 | 0.89 (0.38, 2.09) | 0.02 | 0.02 |
| Mali | PROMIS CS (46) | 503 | 508 | 10.6 | 0.86 (0.56, 1.32) | 0.09 | 0.11 | 451 | 462 | 11.5 | 0.95 (0.65, 1.38) | 0.10 | 0.10 |
| Zimbabwe | SHINE (HIV−) (47) | 870 | 794 | 2.0 | 0.29 (0.11, 0.77) | 0.02 | 0.04 | 805 | 790 | 1.0 | 0.86 (0.33, 2.24) | 0.02 | 0.02 |
| Zimbabwe | SHINE (HIV+) (48) | | | | | | | | | | | | |
| | | **6964** | **7323** | | **$I^2$ = 0.46, $Tau^2$ = 0.06** | | | **7068** | **6639** | | **$I^2$ = 0.00, $Tau^2$ = 0.00** | | |
| **Fixed** | | | | | **0.80 (0.70, 0.91)** | | | | | | **0.84 (0.74, 0.95)** | | |
| **Random** | | | | | **0.78 (0.63, 0.96)** | | | | | | **0.84 (0.74, 0.95)** | | |

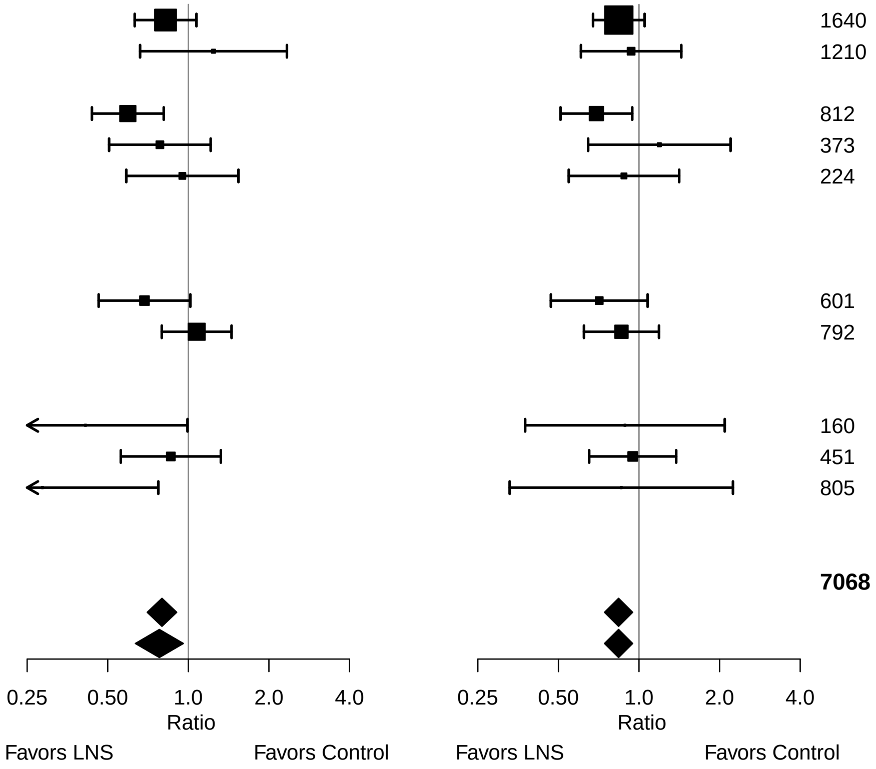

#### Supplemental figure 6H: Low MUAC prevalence ratio

#### 6H4: Stratified by Maternal education

**P−for−interaction = 0.240**
**Ratio of PRs = 1.16 (0.91, 1.48)**

| Country | Trial | Primary or greater: LNS N | Control N | Control Prevalence | PR (95% CI) | Fixed W | Random W | Incomplete or no formal: LNS N | Control N | Control Prevalence | PR (95% CI) | Fixed W | Random W |
|---|---|---|---|---|---|---|---|---|---|---|---|---|---|
| Bangladesh | JiVitA−4 (34) | 1865 | 777 | 14.5 | 0.72 (0.59, 0.88) | 0.64 | 0.42 | 1030 | 491 | 16.2 | 0.98 (0.75, 1.27) | 0.23 | 0.23 |
| Bangladesh | RDNS (35) | 1245 | 589 | 4.1 | 1.18 (0.75, 1.86) | 0.13 | 0.19 | 418 | 226 | 8.0 | 0.81 (0.46, 1.44) | 0.05 | 0.05 |
| Bangladesh | WASH−B (36) | | | | | | | | | | | | |
| Burkina Faso | iLiNS−Zinc (37) | | | | | | | | | | | | |
| Burkina Faso | PROMIS (38) | | | | | | | | | | | | |
| Burkina Faso | PROMIS CS (38) | 34 | 39 | 15.4 | 0.96 (0.31, 2.92) | 0.02 | 0.04 | 388 | 396 | 14.9 | 0.92 (0.65, 1.29) | 0.14 | 0.14 |
| Ghana | GHANA (39) | | | | | | | | | | | | |
| Ghana | iLiNS−DYADG (40) | | | | | | | | | | | | |
| Haiti | HAITI (41) | | | | | | | | | | | | |
| Kenya | WASH−B (42) | 704 | 2460 | 4.2 | 0.57 (0.32, 1.01) | 0.08 | 0.13 | 755 | 2721 | 6.6 | 0.80 (0.57, 1.12) | 0.15 | 0.15 |
| Madagascar | MAHAY (43) | 344 | 420 | 5.7 | 1.07 (0.63, 1.81) | 0.10 | 0.15 | 1336 | 1238 | 11.0 | 0.93 (0.71, 1.22) | 0.22 | 0.22 |
| Malawi | iLiNS−DYADM (44) | | | | | | | | | | | | |
| Malawi | iLiNS−DOSE (45) | | | | | | | | | | | | |
| Mali | PROMIS (46) | | | | | | | | | | | | |
| Mali | PROMIS CS (46) | 107 | 96 | 10.4 | 0.54 (0.22, 1.33) | 0.03 | 0.06 | 846 | 874 | 11.1 | 0.95 (0.72, 1.25) | 0.21 | 0.21 |
| Zimbabwe | SHINE (HIV−) (47) | | | | | | | | | | | | |
| Zimbabwe | SHINE (HIV+) (48) | | | | | | | | | | | | |
| | | **4299** | **4381** | | **I² = 0.30, Tau² = 0.02** | | | **4773** | **5946** | | **I² = 0.00, Tau² = 0.00** | | |
| **Fixed** | | | | | **0.78 (0.66, 0.92)** | | | | | | **0.92 (0.81, 1.04)** | | |
| **Random** | | | | | **0.81 (0.64, 1.02)** | | | | | | **0.92 (0.81, 1.04)** | | |

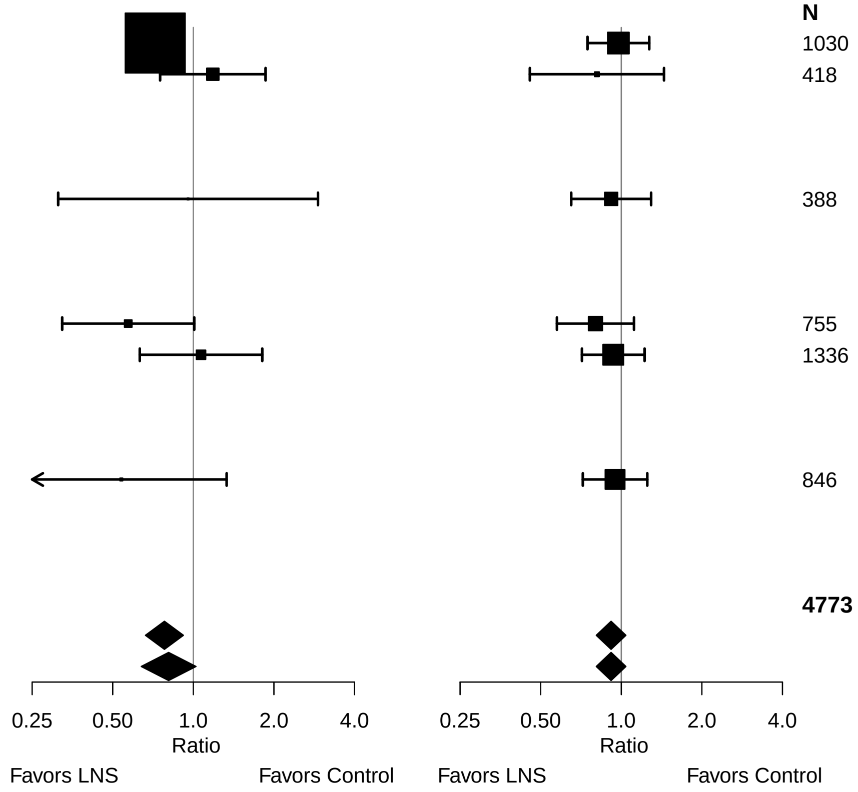

#### Supplemental figure 6H: Low MUAC prevalence ratio

#### 6H5: Stratified by Maternal depressive symptoms

**P−for−interaction = 0.798**
**Ratio of PRs = 1.04 (0.77, 1.40)**

| | | Less than 75th percentile | | | | | | At least 75th percentile | | | | | |
|---|---|---|---|---|---|---|---|---|---|---|---|---|---|
| **Country** | **Trial** | **LNS N** | **Control N** | **Control Prevalence** | **PR (95% CI)** | **Fixed W** | **Random W** | **LNS N** | **Control N** | **Control Prevalence** | **PR (95% CI)** | **Fixed W** | **Random W** |
| Bangladesh | JiVitA−4 (34) | | | | | | | | | | | | |
| Bangladesh | RDNS (35) | 1081 | 472 | 5.3 | 0.94 (0.61, 1.45) | 0.18 | 0.19 | 520 | 288 | 4.5 | 1.28 (0.67, 2.44) | 0.20 | 0.20 |
| Bangladesh | WASH−B (36) | | | | | | | | | | | | |
| Burkina Faso | iLiNS−Zinc (37) | | | | | | | | | | | | |
| Burkina Faso | PROMIS (38) | 500 | 573 | 7.9 | 0.99 (0.64, 1.54) | 0.17 | 0.19 | 224 | 215 | 9.3 | 0.72 (0.37, 1.42) | 0.18 | 0.18 |
| Burkina Faso | PROMIS CS (38) | | | | | | | | | | | | |
| Ghana | GHANA (39) | | | | | | | | | | | | |
| Ghana | iLiNS−DYADG (40) | | | | | | | | | | | | |
| Haiti | HAITI (41) | | | | | | | | | | | | |
| Kenya | WASH−B (42) | 999 | 3523 | 5.4 | 0.76 (0.57, 1.01) | 0.40 | 0.36 | 340 | 1249 | 4.9 | 0.72 (0.37, 1.40) | 0.19 | 0.19 |
| Madagascar | MAHAY (43) | 656 | 707 | 8.5 | 0.99 (0.66, 1.49) | 0.20 | 0.21 | 282 | 245 | 10.6 | 0.87 (0.54, 1.40) | 0.36 | 0.36 |
| Malawi | iLiNS−DYADM (44) | | | | | | | | | | | | |
| Malawi | iLiNS−DOSE (45) | | | | | | | | | | | | |
| Mali | PROMIS (46) | | | | | | | | | | | | |
| Mali | PROMIS CS (46) | | | | | | | | | | | | |
| Zimbabwe | SHINE (HIV−) (47) | 1194 | 1135 | 1.4 | 0.42 (0.18, 0.98) | 0.05 | 0.06 | 509 | 487 | 1.8 | 0.53 (0.17, 1.62) | 0.07 | 0.07 |
| Zimbabwe | SHINE (HIV+) (48) | | | | | | | | | | | | |
| | | **4430** | **6410** | | **I² = 0.12, Tau² = 0.01** | | | **1875** | **2484** | | **I² = 0.00, Tau² = 0.00** | | |
| **Fixed** | | | | | **0.85 (0.71, 1.02)** | | | | | | **0.85 (0.64, 1.13)** | | |
| **Random** | | | | | **0.85 (0.69, 1.04)** | | | | | | **0.85 (0.64, 1.13)** | | |

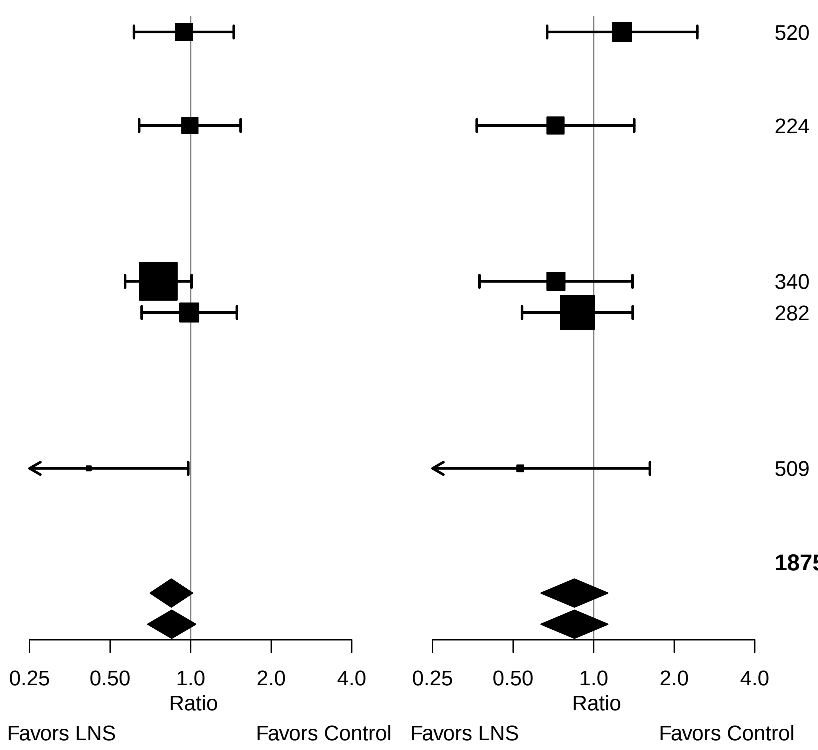

#### Supplemental figure 6H: Low MUAC prevalence ratio

#### 6H6: Stratified by Child sex

**P−for−interaction = 0.002**
**Ratio of PRs = 0.77 (0.66, 0.91)**

| | | Male | | | | | | Female | | | | | |
|---|---|---|---|---|---|---|---|---|---|---|---|---|---|
| **Country** | **Trial** | **LNS N** | **Control N** | **Control Prevalence** | **PR (95% CI)** | **Fixed W** | **Random W** | **LNS N** | **Control N** | **Control Prevalence** | **PR (95% CI)** | **Fixed W** | **Random W** |
| Bangladesh | JiVitA−4 (34) | 1477 | 612 | 12.1 | 1.00 (0.73, 1.36) | 0.16 | 0.16 | 1420 | 659 | 18.1 | 0.76 (0.62, 0.94) | 0.32 | 0.22 |
| Bangladesh | RDNS (35) | 833 | 411 | 4.1 | 1.45 (0.87, 2.42) | 0.06 | 0.06 | 830 | 404 | 6.2 | 0.72 (0.43, 1.20) | 0.05 | 0.07 |
| Bangladesh | WASH−B (36) | | | | | | | | | | | | |
| Burkina Faso | iLiNS−Zinc (37) | 995 | 336 | 17.9 | 0.70 (0.52, 0.95) | 0.17 | 0.17 | 964 | 330 | 18.8 | 0.57 (0.43, 0.76) | 0.17 | 0.16 |
| Burkina Faso | PROMIS (38) | 432 | 482 | 8.1 | 1.00 (0.58, 1.73) | 0.05 | 0.05 | 422 | 430 | 7.9 | 0.87 (0.52, 1.44) | 0.05 | 0.07 |
| Burkina Faso | PROMIS CS (38) | 229 | 219 | 10.5 | 1.12 (0.71, 1.77) | 0.07 | 0.07 | 195 | 217 | 19.4 | 0.82 (0.52, 1.30) | 0.07 | 0.08 |
| Ghana | GHANA (39) | | | | | | | | | | | | |
| Ghana | iLiNS−DYADG (40) | 175 | 324 | 4.3 | 0.66 (0.24, 1.81) | 0.02 | 0.02 | 172 | 369 | 3.3 | 0.89 (0.32, 2.50) | 0.01 | 0.02 |
| Haiti | HAITI (41) | | | | | | | | | | | | |
| Kenya | WASH−B (42) | 736 | 2467 | 5.9 | 0.92 (0.66, 1.28) | 0.14 | 0.14 | 725 | 2718 | 5.1 | 0.46 (0.29, 0.72) | 0.07 | 0.09 |
| Madagascar | MAHAY (43) | 817 | 817 | 10.5 | 0.99 (0.73, 1.34) | 0.17 | 0.17 | 863 | 841 | 8.8 | 0.96 (0.70, 1.31) | 0.14 | 0.14 |
| Malawi | iLiNS−DYADM (44) | | | | | | | | | | | | |
| Malawi | iLiNS−DOSE (45) | | | | | | | | | | | | |
| Mali | PROMIS (46) | 267 | 263 | 5.3 | 0.79 (0.40, 1.59) | 0.03 | 0.03 | 240 | 242 | 6.2 | 0.37 (0.12, 1.14) | 0.01 | 0.02 |
| Mali | PROMIS CS (46) | 479 | 526 | 11.0 | 0.95 (0.63, 1.42) | 0.10 | 0.10 | 475 | 444 | 11.0 | 0.86 (0.58, 1.26) | 0.09 | 0.11 |
| Zimbabwe | SHINE (HIV−) (47) | 928 | 900 | 1.6 | 0.55 (0.24, 1.30) | 0.02 | 0.02 | 944 | 880 | 1.4 | 0.47 (0.18, 1.21) | 0.02 | 0.02 |
| Zimbabwe | SHINE (HIV+) (48) | | | | | | | | | | | | |
| | | **7368** | **7357** | | **I² = 0.00, Tau² = 0.00** | | | **7250** | **7534** | | **I² = 0.28, Tau² = 0.02** | | |
| **Fixed** | | | | | **0.93 (0.82, 1.05)** | | | | | | **0.73 (0.65, 0.82)** | | |
| **Random** | | | | | **0.93 (0.82, 1.05)** | | | | | | **0.72 (0.62, 0.84)** | | |

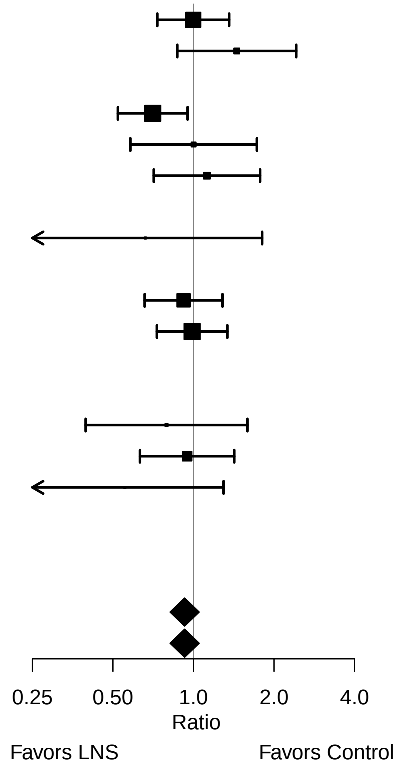

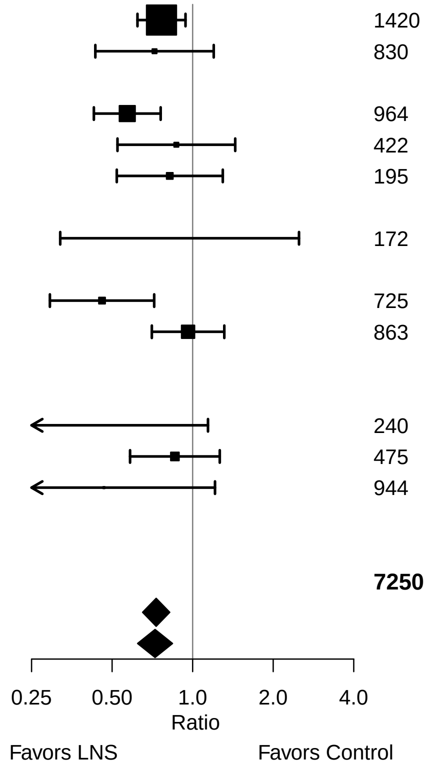

#### Supplemental figure 6H: Low MUAC prevalence ratio

#### 6H7: Stratified by Child birth order

**P−for−interaction = 0.002**
**Ratio of PRs = 1.37 (1.12, 1.67)**

| | | Later born | | | | | | Firstborn | | | | | |
|---|---|---|---|---|---|---|---|---|---|---|---|---|---|
| Country | Trial | LNS N | Control N | Control Prevalence | PR (95% CI) | Fixed W | Random W | LNS N | Control N | Control Prevalence | PR (95% CI) | Fixed W | Random W |
| Bangladesh | JiVitA−4 (34) | 617 | 260 | 17.0 | 0.58 (0.42, 0.80) | 0.11 | 0.12 | 2276 | 1004 | 14.6 | 0.94 (0.76, 1.17) | 0.53 | 0.32 |
| Bangladesh | RDNS (35) | 974 | 508 | 6.9 | 0.82 (0.54, 1.25) | 0.07 | 0.08 | 688 | 307 | 2.3 | 2.04 (0.83, 5.04) | 0.03 | 0.05 |
| Bangladesh | WASH−B (36) | | | | | | | | | | | | |
| Burkina Faso | iLiNS−Zinc (37) | 1524 | 521 | 18.2 | 0.60 (0.46, 0.76) | 0.19 | 0.17 | 435 | 145 | 18.6 | 0.78 (0.53, 1.13) | 0.17 | 0.20 |
| Burkina Faso | PROMIS (38) | 709 | 753 | 8.6 | 0.87 (0.58, 1.29) | 0.07 | 0.09 | 145 | 158 | 5.1 | 1.50 (0.62, 3.61) | 0.03 | 0.06 |
| Burkina Faso | PROMIS CS (38) | 362 | 356 | 16.0 | 0.81 (0.57, 1.16) | 0.09 | 0.11 | 61 | 80 | 10.0 | 1.80 (0.82, 3.98) | 0.04 | 0.07 |
| Ghana | GHANA (39) | | | | | | | | | | | | |
| Ghana | iLiNS−DYADG (40) | | | | | | | | | | | | |
| Haiti | HAITI (41) | | | | | | | | | | | | |
| Kenya | WASH−B (42) | 1137 | 4083 | 5.5 | 0.72 (0.52, 0.99) | 0.12 | 0.12 | 322 | 1097 | 5.4 | 0.69 (0.37, 1.30) | 0.06 | 0.10 |
| Madagascar | MAHAY (43) | 1263 | 1199 | 10.4 | 0.96 (0.75, 1.22) | 0.20 | 0.17 | 417 | 459 | 7.6 | 1.01 (0.60, 1.68) | 0.09 | 0.13 |
| Malawi | iLiNS−DYADM (44) | | | | | | | | | | | | |
| Malawi | iLiNS−DOSE (45) | | | | | | | | | | | | |
| Mali | PROMIS (46) | | | | | | | | | | | | |
| Mali | PROMIS CS (46) | 791 | 832 | 10.8 | 0.91 (0.68, 1.22) | 0.14 | 0.14 | 146 | 126 | 10.3 | 0.80 (0.37, 1.72) | 0.04 | 0.07 |
| Zimbabwe | SHINE (HIV−) (47) | | | | | | | | | | | | |
| Zimbabwe | SHINE (HIV+) (48) | | | | | | | | | | | | |
| | | **7377** | **8512** | | **$I^2$ = 0.42, $Tau^2$ = 0.01** | | | **4490** | **3376** | | **$I^2$ = 0.19, $Tau^2$ = 0.03** | | |
| **Fixed** | | | | | **0.77 (0.69, 0.86)** | | | | | | **0.95 (0.81, 1.11)** | | |
| **Random** | | | | | **0.77 (0.67, 0.88)** | | | | | | **0.98 (0.79, 1.22)** | | |

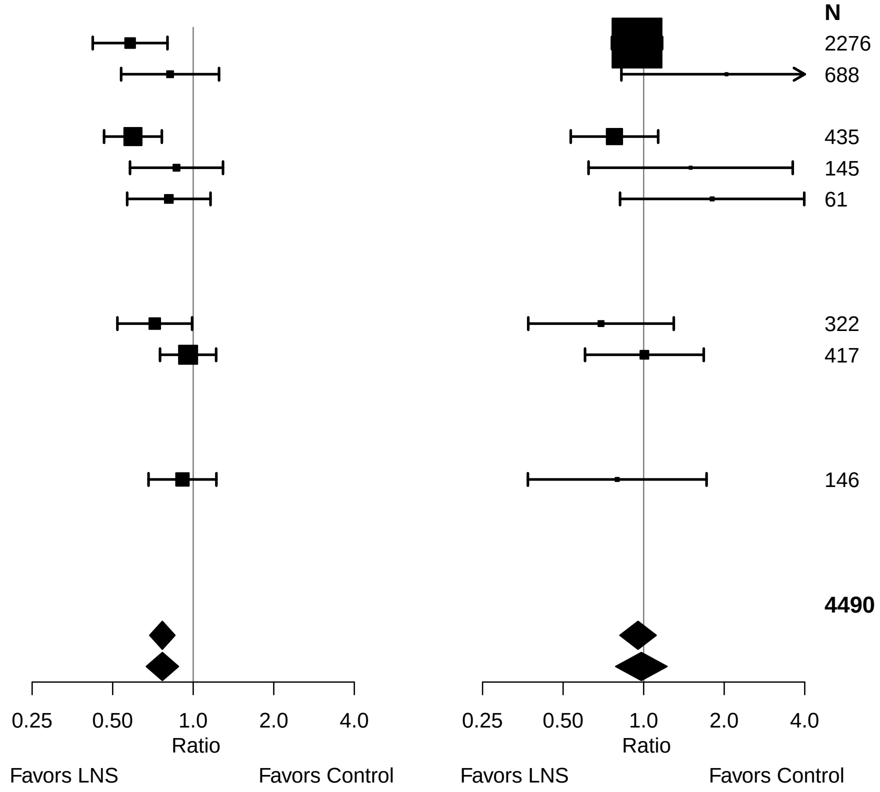

#### 6H8: Stratified by Child baseline anthropometric status (insufficient comparisons)

### Supplemental figure 6I: Low MUAC prevalence difference

#### 6I1: Stratified by Maternal height (insufficient comparisons)

#### Supplemental figure 6I: Low MUAC prevalence difference

#### 6I2: Stratified by Maternal BMI

**P−for−interaction = 0.260**
**Difference in PDs = −0.01 (−0.03, 0.01)**

| | | At least 20 kg/m² | | | | | | Less than 20 kg/m² | | | | | |
|---|---|---|---|---|---|---|---|---|---|---|---|---|---|
| **Country** | **Trial** | **LNS N** | **Control N** | **Control Prevalence** | **PD (95% CI)** | **Fixed W** | **Random W** | **LNS N** | **Control N** | **Control Prevalence** | **PD (95% CI)** | **Fixed W** | **Random W** |
| Bangladesh | JiVitA−4 (34) | | | | | | | | | | | | |
| Bangladesh | RDNS (35) | 730 | 335 | 3.0 | −0.01 (−0.03, 0.01) | 0.24 | 0.20 | 873 | 455 | 7.0 | 0.01 (−0.02, 0.04) | 0.26 | 0.21 |
| Bangladesh | WASH−B (36) | | | | | | | | | | | | |
| Burkina Faso | iLiNS−Zinc (37) | 1218 | 391 | 14.6 | −0.06 (−0.09, −0.03) | 0.08 | 0.13 | 739 | 275 | 23.6 | −0.08 (−0.12, −0.03) | 0.12 | 0.14 |
| Burkina Faso | PROMIS (38) | 641 | 673 | 7.1 | −0.01 (−0.04, 0.03) | 0.08 | 0.12 | 210 | 233 | 10.3 | 0.00 (−0.05, 0.05) | 0.10 | 0.12 |
| Burkina Faso | PROMIS CS (38) | 241 | 266 | 12.0 | −0.01 (−0.07, 0.05) | 0.02 | 0.05 | 183 | 166 | 19.3 | −0.02 (−0.10, 0.05) | 0.05 | 0.07 |
| Ghana | GHANA (39) | | | | | | | | | | | | |
| Ghana | iLiNS−DYADG (40) | | | | | | | | | | | | |
| Haiti | HAITI (41) | | | | | | | | | | | | |
| Kenya | WASH−B (42) | 1101 | 3837 | 4.4 | 0.00 (−0.02, 0.01) | 0.40 | 0.23 | 272 | 1086 | 8.5 | −0.04 (−0.07, −0.02) | 0.31 | 0.22 |
| Madagascar | MAHAY (43) | | | | | | | | | | | | |
| Malawi | iLiNS−DYADM (44) | | | | | | | | | | | | |
| Malawi | iLiNS−DOSE (45) | | | | | | | | | | | | |
| Mali | PROMIS (46) | 381 | 369 | 5.7 | −0.03 (−0.06, 0.00) | 0.09 | 0.13 | 120 | 135 | 5.9 | 0.00 (−0.05, 0.05) | 0.10 | 0.13 |
| Mali | PROMIS CS (46) | 703 | 685 | 10.1 | −0.01 (−0.04, 0.03) | 0.09 | 0.13 | 246 | 279 | 13.6 | −0.02 (−0.08, 0.03) | 0.08 | 0.11 |
| Zimbabwe | SHINE (HIV−) (47) | | | | | | | | | | | | |
| Zimbabwe | SHINE (HIV+) (48) | | | | | | | | | | | | |
| | | **5015** | **6556** | | **I² = 0.49, Tau² = 0.00** | | | **2643** | **2629** | | **I² = 0.51, Tau² = 0.00** | | |
| **Fixed** | | | | | **−0.01 (−0.02, 0.00)** | | | | | | **−0.02 (−0.04, −0.01)** | | |
| **Random** | | | | | **−0.02 (−0.03, 0.00)** | | | | | | **−0.02 (−0.05, 0.00)** | | |

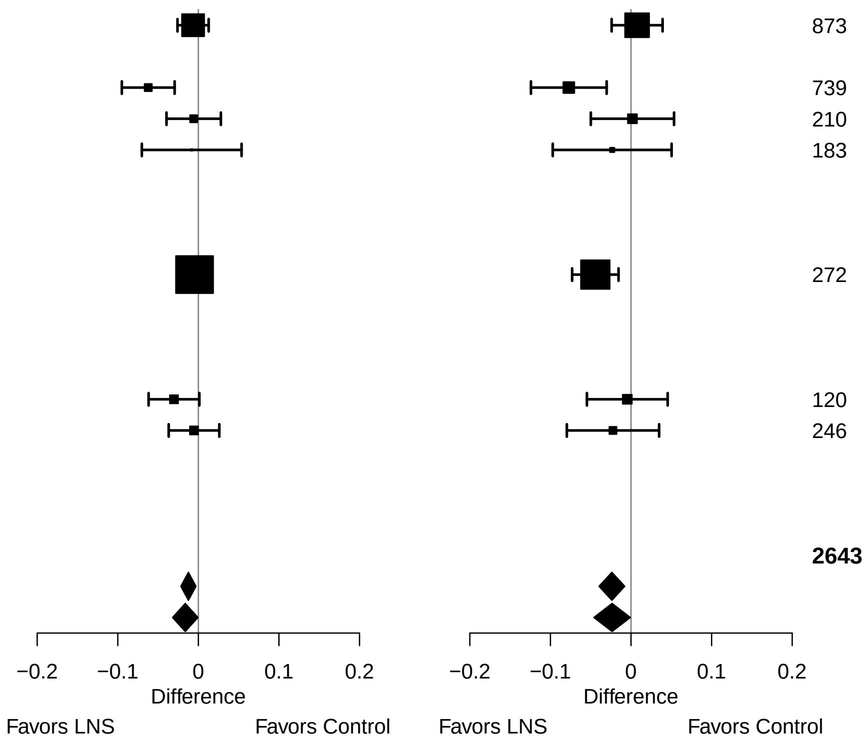

#### Supplemental figure 6I: Low MUAC prevalence difference

#### 6I3: Stratified by Maternal age

**P−for−interaction = 0.160**
**Difference in PDs = 0.01 (0.00, 0.02)**

| Country | Trial | At least 25 y: LNS N | At least 25 y: Control N | At least 25 y: Control Prevalence | At least 25 y: PD (95% CI) | At least 25 y: Fixed W | At least 25 y: Random W | Less than 25 y: LNS N | Less than 25 y: Control N | Less than 25 y: Control Prevalence | Less than 25 y: PD (95% CI) | Less than 25 y: Fixed W | Less than 25 y: Random W |
|---|---|---|---|---|---|---|---|---|---|---|---|---|---|
| Bangladesh | JiVitA−4 (34) | 1244 | 555 | 15.7 | −0.03 (−0.06, 0.01) | 0.05 | 0.09 | 1640 | 711 | 14.7 | −0.02 (−0.04, 0.01) | 0.06 | 0.06 |
| Bangladesh | RDNS (35) | 453 | 221 | 5.0 | 0.01 (−0.02, 0.05) | 0.05 | 0.09 | 1210 | 594 | 5.2 | 0.00 (−0.03, 0.02) | 0.10 | 0.10 |
| Bangladesh | WASH−B (36) | | | | | | | | | | | | |
| Burkina Faso | iLiNS−Zinc (37) | 1134 | 401 | 18.5 | −0.08 (−0.13, −0.03) | 0.02 | 0.05 | 812 | 262 | 17.9 | −0.06 (−0.11, −0.01) | 0.02 | 0.02 |
| Burkina Faso | PROMIS (38) | 481 | 525 | 8.8 | −0.02 (−0.05, 0.02) | 0.04 | 0.08 | 373 | 387 | 7.0 | 0.01 (−0.03, 0.06) | 0.02 | 0.02 |
| Burkina Faso | PROMIS CS (38) | 200 | 190 | 15.8 | −0.01 (−0.08, 0.07) | 0.01 | 0.02 | 224 | 246 | 14.2 | −0.02 (−0.08, 0.05) | 0.01 | 0.01 |
| Ghana | GHANA (39) | | | | | | | | | | | | |
| Ghana | iLiNS−DYADG (40) | | | | | | | | | | | | |
| Haiti | HAITI (41) | | | | | | | | | | | | |
| Kenya | WASH−B (42) | 845 | 2838 | 5.0 | −0.02 (−0.03, 0.00) | 0.26 | 0.20 | 601 | 2315 | 6.1 | −0.02 (−0.04, 0.00) | 0.13 | 0.13 |
| Madagascar | MAHAY (43) | 887 | 933 | 10.1 | 0.01 (−0.02, 0.04) | 0.06 | 0.10 | 792 | 725 | 9.1 | −0.01 (−0.04, 0.01) | 0.06 | 0.06 |
| Malawi | iLiNS−DYADM (44) | | | | | | | | | | | | |
| Malawi | iLiNS−DOSE (45) | | | | | | | | | | | | |
| Mali | PROMIS (46) | 347 | 358 | 5.8 | −0.03 (−0.07, 0.00) | 0.06 | 0.10 | 160 | 147 | 5.4 | 0.00 (−0.05, 0.04) | 0.02 | 0.02 |
| Mali | PROMIS CS (46) | 503 | 508 | 10.6 | −0.01 (−0.06, 0.03) | 0.03 | 0.06 | 451 | 462 | 11.5 | −0.01 (−0.05, 0.04) | 0.03 | 0.03 |
| Zimbabwe | SHINE (HIV−) (47) | 870 | 794 | 2.0 | −0.01 (−0.03, 0.00) | 0.41 | 0.22 | 805 | 790 | 1.0 | 0.00 (−0.01, 0.01) | 0.56 | 0.56 |
| Zimbabwe | SHINE (HIV+) (48) | | | | | | | | | | | | |
| | | **6964** | **7323** | | **I² = 0.28, Tau² = 0.00** | | | **7068** | **6639** | | **I² = 0.00, Tau² = 0.00** | | |
| **Fixed** | | | | | **−0.02 (−0.02, −0.01)** | | | | | | **−0.01 (−0.01, 0.00)** | | |
| **Random** | | | | | **−0.02 (−0.03, 0.00)** | | | | | | **−0.01 (−0.01, 0.00)** | | |

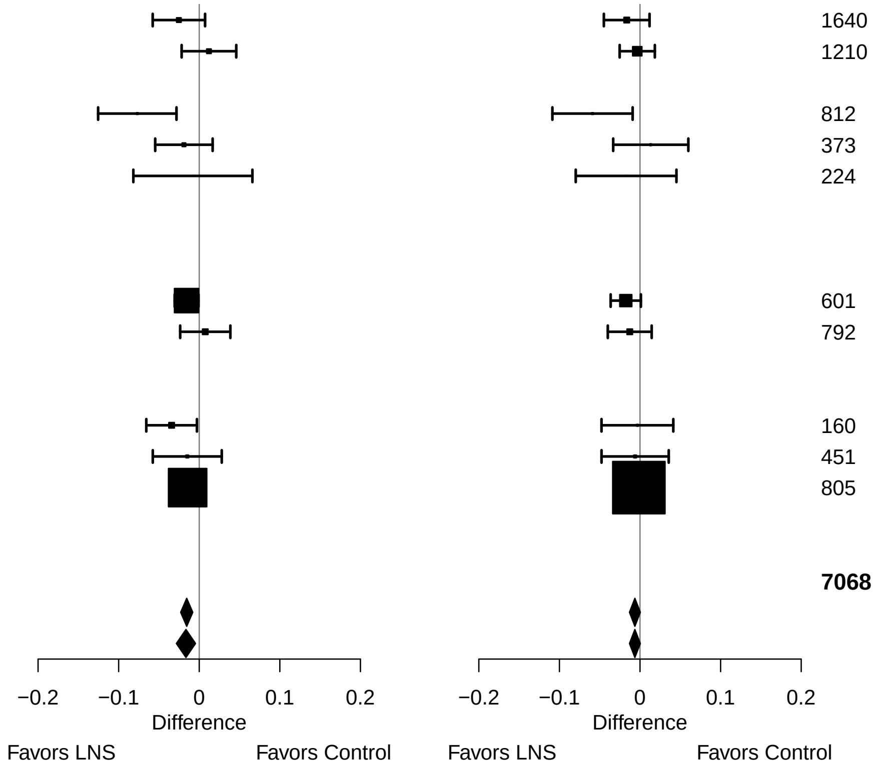

##### Supplemental figure 6I: Low MUAC prevalence difference

#### 6I4: Stratified by Maternal education

**P−for−interaction = 0.676**
**Difference in PDs = 0.00 (−0.01, 0.02)**

| | | Primary or greater | | | | | | Incomplete or no formal | | | | | |
|---|---|---|---|---|---|---|---|---|---|---|---|---|---|
| **Country** | **Trial** | **LNS N** | **Control N** | **Control Prevalence** | **PD (95% CI)** | **Fixed W** | **Random W** | **LNS N** | **Control N** | **Control Prevalence** | **PD (95% CI)** | **Fixed W** | **Random W** |
| Bangladesh | JiVitA−4 (34) | 1865 | 777 | 14.5 | −0.03 (−0.06, −0.01) | 0.15 | 0.20 | 1030 | 491 | 16.2 | −0.01 (−0.04, 0.03) | 0.11 | 0.11 |
| Bangladesh | RDNS (35) | 1245 | 589 | 4.1 | 0.01 (−0.01, 0.03) | 0.25 | 0.26 | 418 | 226 | 8.0 | −0.02 (−0.06, 0.03) | 0.08 | 0.08 |
| Bangladesh | WASH−B (36) | | | | | | | | | | | | |
| Burkina Faso | iLiNS−Zinc (37) | | | | | | | | | | | | |
| Burkina Faso | PROMIS (38) | | | | | | | | | | | | |
| Burkina Faso | PROMIS CS (38) | 34 | 39 | 15.4 | −0.01 (−0.17, 0.16) | 0.00 | 0.01 | 388 | 396 | 14.9 | −0.01 (−0.06, 0.04) | 0.06 | 0.06 |
| Ghana | GHANA (39) | | | | | | | | | | | | |
| Ghana | iLiNS−DYADG (40) | | | | | | | | | | | | |
| Haiti | HAITI (41) | | | | | | | | | | | | |
| Kenya | WASH−B (42) | 704 | 2460 | 4.2 | −0.02 (−0.03, 0.00) | 0.46 | 0.34 | 755 | 2721 | 6.6 | −0.01 (−0.03, 0.00) | 0.43 | 0.43 |
| Madagascar | MAHAY (43) | 344 | 420 | 5.7 | 0.00 (−0.03, 0.04) | 0.10 | 0.15 | 1336 | 1238 | 11.0 | −0.01 (−0.04, 0.02) | 0.17 | 0.17 |
| Malawi | iLiNS−DYADM (44) | | | | | | | | | | | | |
| Malawi | iLiNS−DOSE (45) | | | | | | | | | | | | |
| Mali | PROMIS (46) | | | | | | | | | | | | |
| Mali | PROMIS CS (46) | 107 | 96 | 10.4 | −0.05 (−0.12, 0.02) | 0.02 | 0.04 | 846 | 874 | 11.1 | −0.01 (−0.04, 0.02) | 0.16 | 0.16 |
| Zimbabwe | SHINE (HIV−) (47) | | | | | | | | | | | | |
| Zimbabwe | SHINE (HIV+) (48) | | | | | | | | | | | | |
| | | **4299** | **4381** | | **I² = 0.43, Tau² = 0.00** | | | **4773** | **5946** | | **I² = 0.00, Tau² = 0.00** | | |
| **Fixed** | | | | | **−0.01 (−0.02, 0.00)** | | | | | | **−0.01 (−0.02, 0.00)** | | |
| **Random** | | | | | **−0.01 (−0.03, 0.00)** | | | | | | **−0.01 (−0.02, 0.00)** | | |

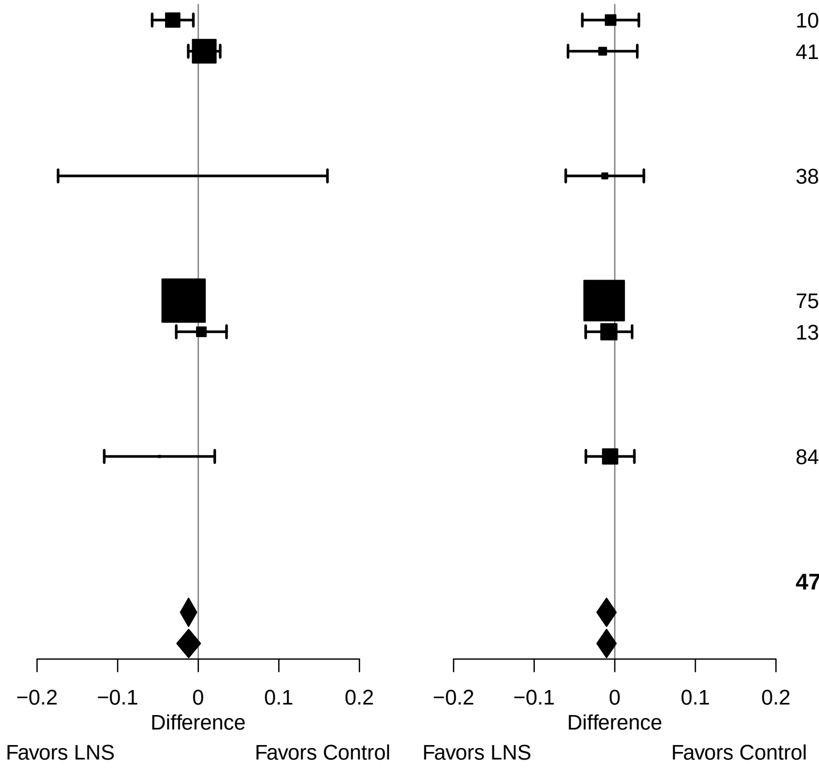

#### Supplemental figure 6I: Low MUAC prevalence difference

##### 6I5: Stratified by Maternal depressive symptoms

**P−for−interaction = 0.946**
**Difference in PDs = 0.00 (−0.01, 0.01)**

| | | Less than 75th percentile | | | | | | At least 75th percentile | | | | | |
|---|---|---|---|---|---|---|---|---|---|---|---|---|---|
| **Country** | **Trial** | **LNS N** | **Control N** | **Control Prevalence** | **PD (95% CI)** | **Fixed W** | **Random W** | **LNS N** | **Control N** | **Control Prevalence** | **PD (95% CI)** | **Fixed W** | **Random W** |
| Bangladesh | JiVitA−4 (34) | | | | | | | | | | | | |
| Bangladesh | RDNS (35) | 1081 | 472 | 5.3 | 0.00 (−0.03, 0.02) | 0.08 | 0.08 | 520 | 288 | 4.5 | 0.01 (−0.02, 0.04) | 0.13 | 0.13 |
| Bangladesh | WASH−B (36) | | | | | | | | | | | | |
| Burkina Faso | iLiNS−Zinc (37) | | | | | | | | | | | | |
| Burkina Faso | PROMIS (38) | 500 | 573 | 7.9 | 0.00 (−0.03, 0.03) | 0.03 | 0.03 | 224 | 215 | 9.3 | −0.03 (−0.08, 0.03) | 0.05 | 0.05 |
| Burkina Faso | PROMIS CS (38) | | | | | | | | | | | | |
| Ghana | GHANA (39) | | | | | | | | | | | | |
| Ghana | iLiNS−DYADG (40) | | | | | | | | | | | | |
| Haiti | HAITI (41) | | | | | | | | | | | | |
| Kenya | WASH−B (42) | 999 | 3523 | 5.4 | −0.01 (−0.03, 0.00) | 0.25 | 0.25 | 340 | 1249 | 4.9 | −0.01 (−0.04, 0.01) | 0.22 | 0.22 |
| Madagascar | MAHAY (43) | 656 | 707 | 8.5 | 0.00 (−0.04, 0.03) | 0.03 | 0.03 | 282 | 245 | 10.6 | −0.01 (−0.06, 0.03) | 0.06 | 0.06 |
| Malawi | iLiNS−DYADM (44) | | | | | | | | | | | | |
| Malawi | iLiNS−DOSE (45) | | | | | | | | | | | | |
| Mali | PROMIS (46) | | | | | | | | | | | | |
| Mali | PROMIS CS (46) | | | | | | | | | | | | |
| Zimbabwe | SHINE (HIV−) (47) | 1194 | 1135 | 1.4 | −0.01 (−0.02, 0.00) | 0.60 | 0.60 | 509 | 487 | 1.8 | −0.01 (−0.02, 0.01) | 0.55 | 0.55 |
| Zimbabwe | SHINE (HIV+) (48) | | | | | | | | | | | | |
| | | **4430** | **6410** | | **I² = 0.00, Tau² = 0.00** | | | **1875** | **2484** | | **I² = 0.00, Tau² = 0.00** | | |
| **Fixed** | | | | | **−0.01 (−0.01, 0.00)** | | | | | | **−0.01 (−0.02, 0.00)** | | |
| **Random** | | | | | **−0.01 (−0.01, 0.00)** | | | | | | **−0.01 (−0.02, 0.00)** | | |

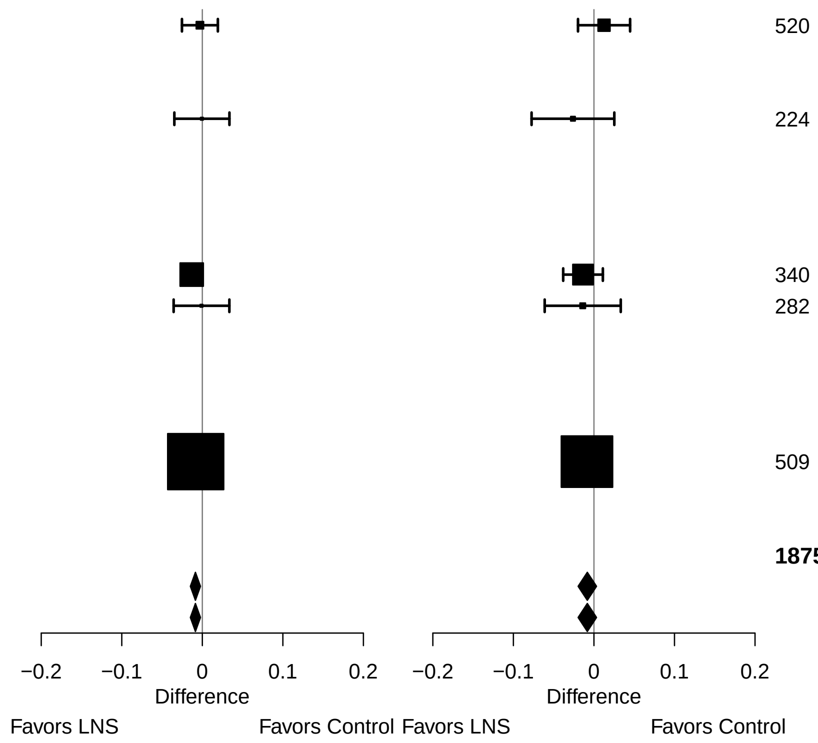

#### Supplemental figure 6I: Low MUAC prevalence difference

#### 6I6: Stratified by Child sex

**P−for−interaction = 0.032**
**Difference in PDs = −0.01 (−0.02, 0.00)**

| | | | | | Male | | | | | | Female | | |
|---|---|---|---|---|---|---|---|---|---|---|---|---|---|
| **Country** | **Trial** | **LNS N** | **Control N** | **Control Prevalence** | **PD (95% CI)** | **Fixed W** | **Random W** | **LNS N** | **Control N** | **Control Prevalence** | **PD (95% CI)** | **Fixed W** | **Random W** |
| Bangladesh | JiVitA−4 (34) | 1477 | 612 | 12.1 | 0.00 (−0.03, 0.03) | 0.06 | 0.06 | 1420 | 659 | 18.1 | −0.04 (−0.06, −0.01) | 0.05 | 0.09 |
| Bangladesh | RDNS (35) | 833 | 411 | 4.1 | 0.02 (−0.01, 0.04) | 0.08 | 0.08 | 830 | 404 | 6.2 | −0.02 (−0.04, 0.01) | 0.05 | 0.09 |
| Bangladesh | WASH−B (36) | | | | | | | | | | | | |
| Burkina Faso | iLiNS−Zinc (37) | 995 | 336 | 17.9 | −0.06 (−0.10, −0.02) | 0.03 | 0.03 | 964 | 330 | 18.8 | −0.08 (−0.13, −0.03) | 0.01 | 0.03 |
| Burkina Faso | PROMIS (38) | 432 | 482 | 8.1 | 0.00 (−0.04, 0.04) | 0.03 | 0.03 | 422 | 430 | 7.9 | −0.01 (−0.05, 0.03) | 0.03 | 0.06 |
| Burkina Faso | PROMIS CS (38) | 229 | 219 | 10.5 | 0.01 (−0.04, 0.06) | 0.02 | 0.02 | 195 | 217 | 19.4 | −0.03 (−0.11, 0.04) | 0.01 | 0.02 |
| Ghana | GHANA (39) | | | | | | | | | | | | |
| Ghana | iLiNS−DYADG (40) | 175 | 324 | 4.3 | −0.01 (−0.05, 0.02) | 0.04 | 0.04 | 172 | 369 | 3.3 | 0.00 (−0.04, 0.03) | 0.04 | 0.07 |
| Haiti | HAITI (41) | | | | | | | | | | | | |
| Kenya | WASH−B (42) | 736 | 2467 | 5.9 | 0.00 (−0.02, 0.01) | 0.14 | 0.14 | 725 | 2718 | 5.1 | −0.03 (−0.04, −0.02) | 0.25 | 0.20 |
| Madagascar | MAHAY (43) | 817 | 817 | 10.5 | 0.00 (−0.03, 0.03) | 0.05 | 0.05 | 863 | 841 | 8.8 | 0.00 (−0.03, 0.02) | 0.05 | 0.09 |
| Malawi | iLiNS−DYADM (44) | | | | | | | | | | | | |
| Malawi | iLiNS−DOSE (45) | | | | | | | | | | | | |
| Mali | PROMIS (46) | 267 | 263 | 5.3 | −0.01 (−0.04, 0.02) | 0.05 | 0.05 | 240 | 242 | 6.2 | −0.04 (−0.08, 0.00) | 0.02 | 0.05 |
| Mali | PROMIS CS (46) | 479 | 526 | 11.0 | −0.01 (−0.05, 0.04) | 0.03 | 0.03 | 475 | 444 | 11.0 | −0.02 (−0.06, 0.02) | 0.03 | 0.05 |
| Zimbabwe | SHINE (HIV−) (47) | 928 | 900 | 1.6 | −0.01 (−0.02, 0.00) | 0.49 | 0.49 | 944 | 880 | 1.4 | −0.01 (−0.02, 0.00) | 0.46 | 0.24 |
| Zimbabwe | SHINE (HIV+) (48) | | | | | | | | | | | | |
| | | **7368** | **7357** | | **I² = 0.06, Tau² = 0.00** | | | **7250** | **7534** | | **I² = 0.44, Tau² = 0.00** | | |
| **Fixed** | | | | | **−0.01 (−0.01, 0.00)** | | | | | | **−0.02 (−0.02, −0.01)** | | |
| **Random** | | | | | **−0.01 (−0.01, 0.00)** | | | | | | **−0.02 (−0.03, −0.01)** | | |

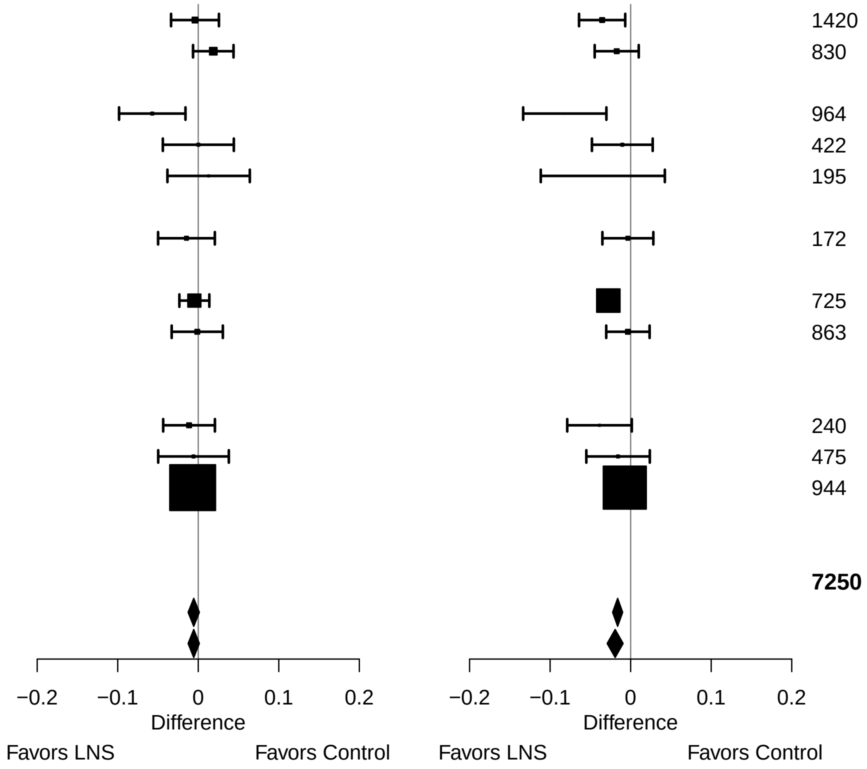

##### Supplemental figure 6I: Low MUAC prevalence difference

#### 6I7: Stratified by Child birth order

**P−for−interaction = 0.032**
**Difference in PDs = 0.02 (0.00, 0.03)**

| | | Later born | | | | | | Firstborn | | | | | |
|---|---|---|---|---|---|---|---|---|---|---|---|---|---|
| **Country** | **Trial** | **LNS N** | **Control N** | **Control Prevalence** | **PD (95% CI)** | **Fixed W** | **Random W** | **LNS N** | **Control N** | **Control Prevalence** | **PD (95% CI)** | **Fixed W** | **Random W** |
| Bangladesh | JiVitA−4 (34) | 617 | 260 | 17.0 | −0.06 (−0.10, −0.01) | 0.04 | 0.09 | 2276 | 1004 | 14.6 | −0.01 (−0.04, 0.01) | 0.28 | 0.21 |
| Bangladesh | RDNS (35) | 974 | 508 | 6.9 | −0.01 (−0.04, 0.01) | 0.12 | 0.14 | 688 | 307 | 2.3 | 0.02 (0.00, 0.05) | 0.21 | 0.19 |
| Bangladesh | WASH−B (36) | | | | | | | | | | | | |
| Burkina Faso | iLiNS−Zinc (37) | 1524 | 521 | 18.2 | −0.07 (−0.11, −0.03) | 0.06 | 0.10 | 435 | 145 | 18.6 | −0.05 (−0.11, 0.00) | 0.06 | 0.09 |
| Burkina Faso | PROMIS (38) | 709 | 753 | 8.6 | −0.01 (−0.04, 0.02) | 0.08 | 0.12 | 145 | 158 | 5.1 | 0.03 (−0.03, 0.08) | 0.05 | 0.08 |
| Burkina Faso | PROMIS CS (38) | 362 | 356 | 16.0 | −0.03 (−0.08, 0.02) | 0.03 | 0.07 | 61 | 80 | 10.0 | 0.08 (−0.04, 0.20) | 0.01 | 0.02 |
| Ghana | GHANA (39) | | | | | | | | | | | | |
| Ghana | iLiNS−DYADG (40) | | | | | | | | | | | | |
| Haiti | HAITI (41) | | | | | | | | | | | | |
| Kenya | WASH−B (42) | 1137 | 4083 | 5.5 | −0.02 (−0.03, 0.00) | 0.45 | 0.20 | 322 | 1097 | 5.4 | −0.02 (−0.04, 0.01) | 0.26 | 0.21 |
| Madagascar | MAHAY (43) | 1263 | 1199 | 10.4 | 0.00 (−0.03, 0.02) | 0.14 | 0.15 | 417 | 459 | 7.6 | 0.00 (−0.04, 0.04) | 0.10 | 0.14 |
| Malawi | iLiNS−DYADM (44) | | | | | | | | | | | | |
| Malawi | iLiNS−DOSE (45) | | | | | | | | | | | | |
| Mali | PROMIS (46) | | | | | | | | | | | | |
| Mali | PROMIS CS (46) | 791 | 832 | 10.8 | −0.01 (−0.04, 0.02) | 0.09 | 0.13 | 146 | 126 | 10.3 | −0.02 (−0.09, 0.05) | 0.03 | 0.06 |
| Zimbabwe | SHINE (HIV−) (47) | | | | | | | | | | | | |
| Zimbabwe | SHINE (HIV+) (48) | | | | | | | | | | | | |
| | | **7377** | **8512** | | **I² = 0.46, Tau² = 0.00** | | | **4490** | **3376** | | **I² = 0.41, Tau² = 0.00** | | |
| **Fixed** | | | | | **−0.02 (−0.03, −0.01)** | | | | | | **0.00 (−0.02, 0.01)** | | |
| **Random** | | | | | **−0.02 (−0.04, −0.01)** | | | | | | **0.00 (−0.02, 0.02)** | | |

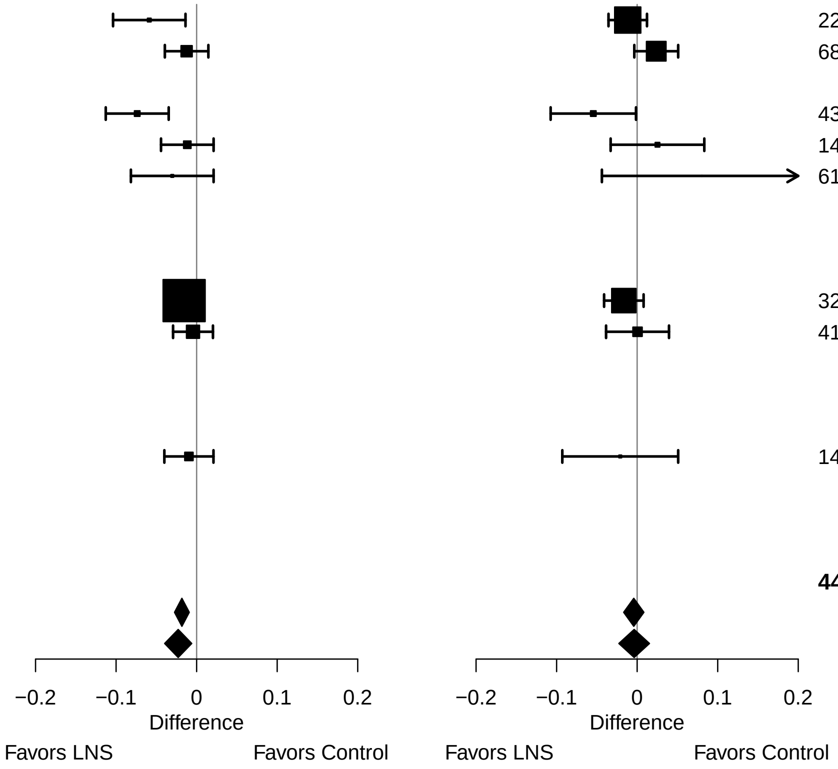

#### 6I8: Stratified by Child baseline anthropometric status (insufficient comparisons)

### Supplemental figure 6J: Acute malnutrition prevalence ratio

#### 6J1: Stratified by Maternal height

**P−for−interaction = 0.435**
**Ratio of PRs = 0.90 (0.68, 1.18)**

| | | At least 150.1 cm | | | | | | Less than 150.1 cm | | | | | |
|---|---|---|---|---|---|---|---|---|---|---|---|---|---|
| **Country** | **Trial** | **LNS N** | **Control N** | **Control Prevalence** | **PR (95% CI)** | **Fixed W** | **Random W** | **LNS N** | **Control N** | **Control Prevalence** | **PR (95% CI)** | **Fixed W** | **Random W** |
| Bangladesh | JiVitA−4 (34) | | | | | | | | | | | | |
| Bangladesh | RDNS (35) | 868 | 432 | 14.1 | 0.86 (0.66, 1.11) | 0.36 | 0.32 | 733 | 358 | 17.0 | 0.83 (0.60, 1.15) | 0.47 | 0.47 |
| Bangladesh | WASH−B (36) | | | | | | | | | | | | |
| Burkina Faso | iLiNS−Zinc (37) | | | | | | | | | | | | |
| Burkina Faso | PROMIS (38) | | | | | | | | | | | | |
| Burkina Faso | PROMIS CS (38) | | | | | | | | | | | | |
| Ghana | GHANA (39) | | | | | | | | | | | | |
| Ghana | iLiNS−DYADG (40) | | | | | | | | | | | | |
| Haiti | HAITI (41) | | | | | | | | | | | | |
| Kenya | WASH−B (42) | | | | | | | | | | | | |
| Madagascar | MAHAY (43) | 983 | 1018 | 7.6 | 1.10 (0.77, 1.57) | 0.19 | 0.24 | 594 | 557 | 12.0 | 0.88 (0.62, 1.25) | 0.39 | 0.39 |
| Malawi | iLiNS−DYADM (44) | | | | | | | | | | | | |
| Malawi | iLiNS−DOSE (45) | 577 | 197 | 4.6 | 1.64 (0.85, 3.19) | 0.06 | 0.10 | 112 | 42 | 11.9 | 0.90 (0.36, 2.28) | 0.06 | 0.06 |
| Mali | PROMIS (46) | | | | | | | | | | | | |
| Mali | PROMIS CS (46) | 907 | 927 | 12.8 | 0.86 (0.67, 1.10) | 0.39 | 0.34 | 32 | 26 | 23.1 | 1.08 (0.52, 2.26) | 0.09 | 0.09 |
| Zimbabwe | SHINE (HIV−) (47) | | | | | | | | | | | | |
| Zimbabwe | SHINE (HIV+) (48) | | | | | | | | | | | | |
| | | **3335** | **2574** | | **I² = 0.33, Tau² = 0.02** | | | **1471** | **983** | | **I² = 0.00, Tau² = 0.00** | | |
| **Fixed** | | | | | **0.93 (0.80, 1.09)** | | | | | | **0.88 (0.70, 1.09)** | | |
| **Random** | | | | | **0.97 (0.77, 1.22)** | | | | | | **0.88 (0.70, 1.09)** | | |

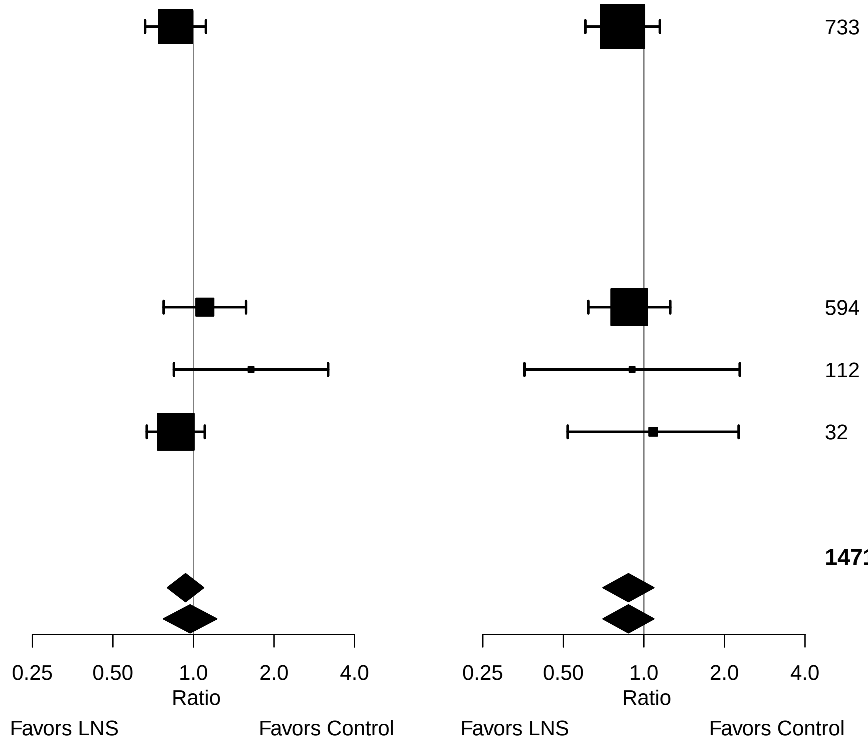

#### Supplemental figure 6J: Acute malnutrition prevalence ratio

##### 6J2: Stratified by Maternal BMI

**P−for−interaction = 0.490**
**Ratio of PRs = 1.07 (0.88, 1.30)**

| | | At least 20 kg/m² | | | | | | Less than 20 kg/m² | | | | | |
|---|---|---|---|---|---|---|---|---|---|---|---|---|---|
| **Country** | **Trial** | **LNS N** | **Control N** | **Control Prevalence** | **PR (95% CI)** | **Fixed W** | **Random W** | **LNS N** | **Control N** | **Control Prevalence** | **PR (95% CI)** | **Fixed W** | **Random W** |
| Bangladesh | JiVitA−4 (34) | | | | | | | | | | | | |
| Bangladesh | RDNS (35) | 730 | 335 | 9.6 | 0.99 (0.68, 1.44) | 0.16 | 0.16 | 871 | 455 | 19.8 | 0.81 (0.65, 1.02) | 0.27 | 0.27 |
| Bangladesh | WASH−B (36) | | | | | | | | | | | | |
| Burkina Faso | iLiNS−Zinc (37) | 1214 | 390 | 15.3 | 0.59 (0.40, 0.87) | 0.15 | 0.15 | 735 | 274 | 24.8 | 0.77 (0.63, 0.93) | 0.36 | 0.34 |
| Burkina Faso | PROMIS (38) | 637 | 663 | 9.5 | 0.89 (0.58, 1.36) | 0.13 | 0.13 | 206 | 232 | 17.7 | 0.74 (0.49, 1.11) | 0.08 | 0.09 |
| Burkina Faso | PROMIS CS (38) | 241 | 265 | 15.5 | 0.89 (0.58, 1.34) | 0.13 | 0.13 | 183 | 165 | 25.5 | 0.97 (0.71, 1.32) | 0.14 | 0.15 |
| Ghana | GHANA (39) | | | | | | | | | | | | |
| Ghana | iLiNS−DYADG (40) | | | | | | | | | | | | |
| Haiti | HAITI (41) | | | | | | | | | | | | |
| Kenya | WASH−B (42) | | | | | | | | | | | | |
| Madagascar | MAHAY (43) | | | | | | | | | | | | |
| Malawi | iLiNS−DYADM (44) | | | | | | | | | | | | |
| Malawi | iLiNS−DOSE (45) | 509 | 174 | 5.2 | 1.00 (0.49, 2.02) | 0.05 | 0.05 | 178 | 64 | 7.8 | 1.77 (0.72, 4.31) | 0.02 | 0.02 |
| Mali | PROMIS (46) | | | | | | | | | | | | |
| Mali | PROMIS CS (46) | 696 | 675 | 12.3 | 0.82 (0.61, 1.09) | 0.27 | 0.27 | 243 | 278 | 15.1 | 1.04 (0.72, 1.49) | 0.10 | 0.11 |
| Zimbabwe | SHINE (HIV−) (47) | 1353 | 1293 | 3.4 | 0.74 (0.47, 1.16) | 0.11 | 0.11 | 239 | 247 | 5.7 | 0.66 (0.32, 1.40) | 0.02 | 0.03 |
| Zimbabwe | SHINE (HIV+) (48) | | | | | | | | | | | | |
| | | **5380** | **3795** | | **I² = 0.00, Tau² = 0.00** | | | **2655** | **1715** | | **I² = 0.06, Tau² = 0.00** | | |
| **Fixed** | | | | | **0.82 (0.70, 0.95)** | | | | | | **0.84 (0.74, 0.94)** | | |
| **Random** | | | | | **0.82 (0.70, 0.95)** | | | | | | **0.84 (0.74, 0.95)** | | |

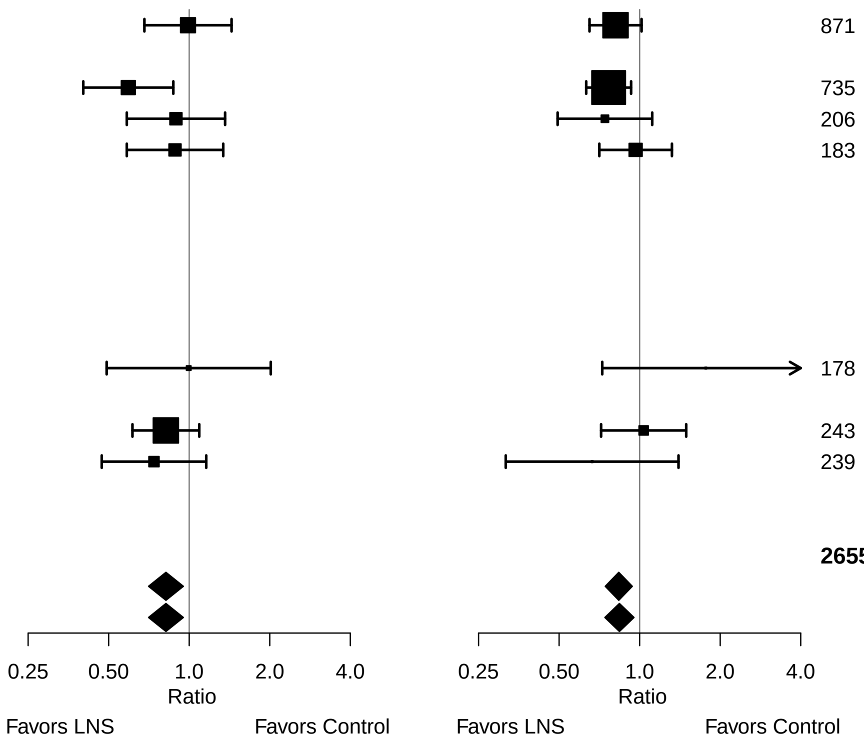

#### Supplemental figure 6J: Acute malnutrition prevalence ratio

#### 6J3: Stratified by Maternal age

**P−for−interaction = 0.429**
**Ratio of PRs = 1.06 (0.91, 1.23)**

| | | At least 25 y | | | | | | Less than 25 y | | | | | |
|---|---|---|---|---|---|---|---|---|---|---|---|---|---|
| **Country** | **Trial** | **LNS N** | **Control N** | **Control Prevalence** | **PR (95% CI)** | **Fixed W** | **Random W** | **LNS N** | **Control N** | **Control Prevalence** | **PR (95% CI)** | **Fixed W** | **Random W** |
| Bangladesh | JiVitA−4 (34) | 1191 | 534 | 21.5 | 0.88 (0.72, 1.07) | 0.30 | 0.19 | 1580 | 677 | 20.5 | 0.91 (0.76, 1.08) | 0.35 | 0.35 |
| Bangladesh | RDNS (35) | 453 | 221 | 14.0 | 1.18 (0.83, 1.67) | 0.10 | 0.11 | 1208 | 594 | 15.8 | 0.75 (0.58, 0.98) | 0.16 | 0.16 |
| Bangladesh | WASH−B (36) | | | | | | | | | | | | |
| Burkina Faso | iLiNS−Zinc (37) | 1131 | 399 | 19.5 | 0.62 (0.47, 0.81) | 0.16 | 0.15 | 807 | 262 | 18.7 | 0.75 (0.53, 1.07) | 0.09 | 0.09 |
| Burkina Faso | PROMIS (38) | 476 | 519 | 11.4 | 0.81 (0.55, 1.20) | 0.08 | 0.10 | 370 | 382 | 12.0 | 0.85 (0.52, 1.40) | 0.04 | 0.04 |
| Burkina Faso | PROMIS CS (38) | 200 | 189 | 20.6 | 0.90 (0.58, 1.38) | 0.06 | 0.08 | 224 | 245 | 18.4 | 1.00 (0.73, 1.37) | 0.11 | 0.11 |
| Ghana | GHANA (39) | | | | | | | | | | | | |
| Ghana | iLiNS−DYADG (40) | | | | | | | | | | | | |
| Haiti | HAITI (41) | | | | | | | | | | | | |
| Kenya | WASH−B (42) | 841 | 2801 | 3.2 | 0.62 (0.38, 1.03) | 0.05 | 0.07 | 598 | 2284 | 3.2 | 0.80 (0.46, 1.39) | 0.04 | 0.04 |
| Madagascar | MAHAY (43) | 883 | 930 | 10.5 | 0.96 (0.71, 1.28) | 0.14 | 0.14 | 791 | 725 | 8.0 | 1.04 (0.72, 1.50) | 0.08 | 0.08 |
| Malawi | iLiNS−DYADM (44) | | | | | | | | | | | | |
| Malawi | iLiNS−DOSE (45) | | | | | | | | | | | | |
| Mali | PROMIS (46) | | | | | | | | | | | | |
| Mali | PROMIS CS (46) | 498 | 501 | 13.8 | 0.82 (0.57, 1.18) | 0.09 | 0.11 | 446 | 458 | 12.2 | 0.97 (0.70, 1.36) | 0.10 | 0.10 |
| Zimbabwe | SHINE (HIV−) (47) | 870 | 794 | 4.5 | 0.53 (0.29, 0.97) | 0.03 | 0.05 | 800 | 787 | 2.4 | 1.09 (0.62, 1.89) | 0.04 | 0.04 |
| Zimbabwe | SHINE (HIV+) (48) | | | | | | | | | | | | |
| | | **6543** | **6888** | | **I² = 0.39, Tau² = 0.02** | | | **6824** | **6414** | | **I² = 0.00, Tau² = 0.00** | | |
| **Fixed** | | | | | **0.83 (0.74, 0.92)** | | | | | | **0.89 (0.80, 0.99)** | | |
| **Random** | | | | | **0.82 (0.71, 0.95)** | | | | | | **0.89 (0.80, 0.99)** | | |

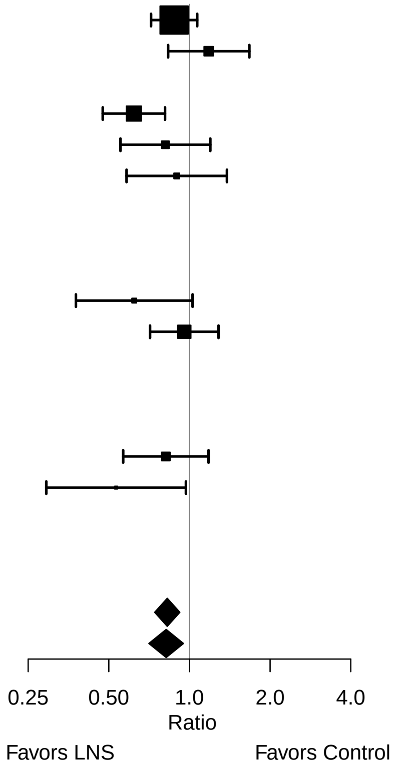

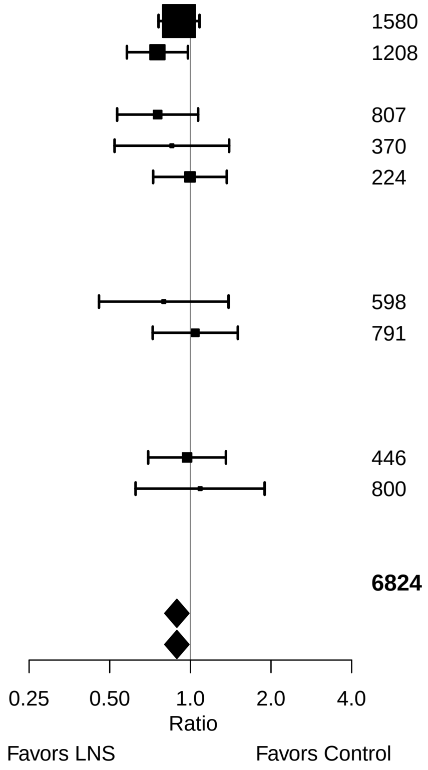

#### Supplemental figure 6J: Acute malnutrition prevalence ratio

#### 6J4: Stratified by Maternal education

**P−for−interaction = 0.659**
**Ratio of PRs = 1.04 (0.86, 1.26)**

| | | Primary or greater | | | | | | Incomplete or no formal | | | | | |
|---|---|---|---|---|---|---|---|---|---|---|---|---|---|
| Country | Trial | LNS N | Control N | Control Prevalence | PR (95% CI) | Fixed W | Random W | LNS N | Control N | Control Prevalence | PR (95% CI) | Fixed W | Random W |
| Bangladesh | JiVitA−4 (34) | 1797 | 739 | 20.2 | 0.89 (0.75, 1.05) | 0.57 | 0.57 | 984 | 474 | 22.3 | 0.91 (0.74, 1.11) | 0.30 | 0.30 |
| Bangladesh | RDNS (35) | 1245 | 589 | 15.1 | 0.81 (0.63, 1.04) | 0.25 | 0.25 | 416 | 226 | 15.9 | 1.01 (0.73, 1.40) | 0.11 | 0.11 |
| Bangladesh | WASH−B (36) | | | | | | | | | | | | |
| Burkina Faso | iLiNS−Zinc (37) | | | | | | | | | | | | |
| Burkina Faso | PROMIS (38) | | | | | | | | | | | | |
| Burkina Faso | PROMIS CS (38) | 34 | 39 | 20.5 | 1.15 (0.41, 3.24) | 0.01 | 0.01 | 388 | 394 | 19.3 | 0.94 (0.71, 1.23) | 0.16 | 0.16 |
| Ghana | GHANA (39) | | | | | | | | | | | | |
| Ghana | iLiNS−DYADG (40) | 267 | 548 | 7.3 | 0.92 (0.54, 1.58) | 0.05 | 0.05 | 80 | 144 | 11.1 | 0.68 (0.28, 1.66) | 0.01 | 0.01 |
| Haiti | HAITI (41) | | | | | | | | | | | | |
| Kenya | WASH−B (42) | 700 | 2426 | 2.5 | 0.75 (0.41, 1.39) | 0.04 | 0.04 | 752 | 2687 | 3.8 | 0.69 (0.44, 1.11) | 0.06 | 0.06 |
| Madagascar | MAHAY (43) | 342 | 420 | 5.5 | 1.23 (0.72, 2.08) | 0.06 | 0.06 | 1333 | 1235 | 10.8 | 0.92 (0.68, 1.24) | 0.14 | 0.14 |
| Malawi | iLiNS−DYADM (44) | | | | | | | | | | | | |
| Malawi | iLiNS−DOSE (45) | | | | | | | | | | | | |
| Mali | PROMIS (46) | | | | | | | | | | | | |
| Mali | PROMIS CS (46) | 107 | 96 | 13.5 | 0.41 (0.17, 1.02) | 0.02 | 0.02 | 836 | 863 | 13.0 | 0.95 (0.75, 1.20) | 0.22 | 0.22 |
| Zimbabwe | SHINE (HIV−) (47) | | | | | | | | | | | | |
| Zimbabwe | SHINE (HIV+) (48) | | | | | | | | | | | | |
| | | **4492** | **4857** | | **I² = 0.00, Tau² = 0.00** | | | **4789** | **6023** | | **I² = 0.00, Tau² = 0.00** | | |
| **Fixed** | | | | | **0.87 (0.77, 0.98)** | | | | | | **0.92 (0.82, 1.02)** | | |
| **Random** | | | | | **0.87 (0.77, 0.98)** | | | | | | **0.92 (0.82, 1.02)** | | |

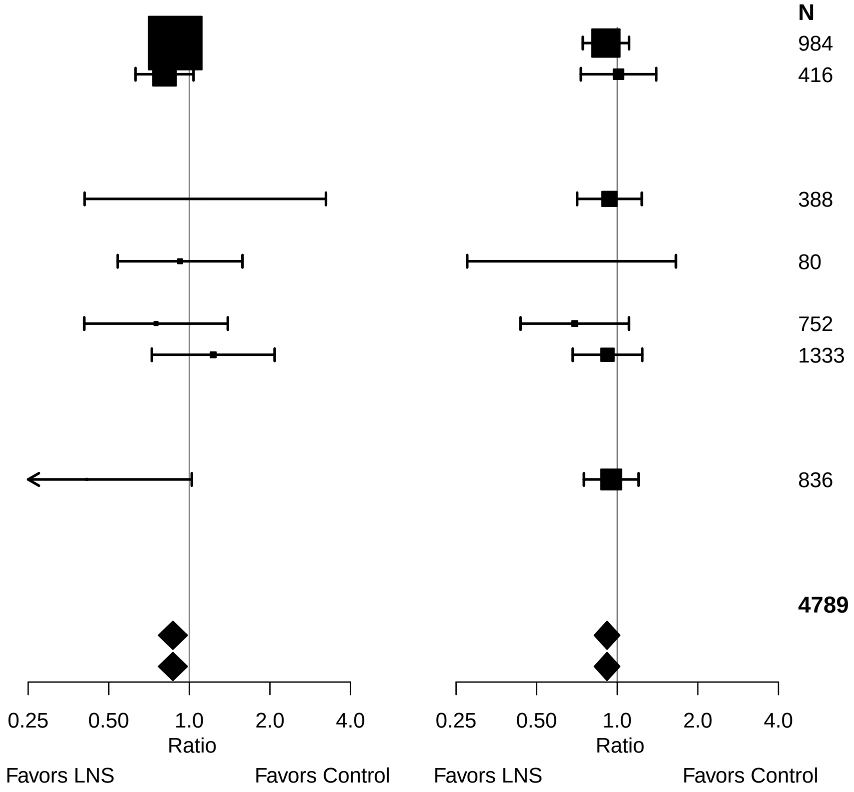

#### Supplemental figure 6J: Acute malnutrition prevalence ratio

#### 6J5: Stratified by Maternal depressive symptoms

**P−for−interaction = 0.532**
**Ratio of PRs = 1.08 (0.84, 1.39)**

| | | Less than 75th percentile | | | | | | At least 75th percentile | | | | | |
|---|---|---|---|---|---|---|---|---|---|---|---|---|---|
| **Country** | **Trial** | **LNS N** | **Control N** | **Control Prevalence** | **PR (95% CI)** | **Fixed W** | **Random W** | **LNS N** | **Control N** | **Control Prevalence** | **PR (95% CI)** | **Fixed W** | **Random W** |
| Bangladesh | JiVitA−4 (34) | | | | | | | | | | | | |
| Bangladesh | RDNS (35) | 1081 | 472 | 13.8 | 0.99 (0.77, 1.29) | 0.36 | 0.36 | 519 | 288 | 17.0 | 0.71 (0.49, 1.04) | 0.31 | 0.31 |
| Bangladesh | WASH−B (36) | | | | | | | | | | | | |
| Burkina Faso | iLiNS−Zinc (37) | | | | | | | | | | | | |
| Burkina Faso | PROMIS (38) | 497 | 567 | 11.6 | 0.78 (0.54, 1.12) | 0.18 | 0.18 | 219 | 211 | 14.7 | 0.75 (0.47, 1.19) | 0.20 | 0.20 |
| Burkina Faso | PROMIS CS (38) | | | | | | | | | | | | |
| Ghana | GHANA (39) | | | | | | | | | | | | |
| Ghana | iLiNS−DYADG (40) | 252 | 459 | 7.4 | 0.86 (0.48, 1.52) | 0.07 | 0.07 | 83 | 209 | 8.6 | 1.12 (0.51, 2.47) | 0.07 | 0.07 |
| Haiti | HAITI (41) | | | | | | | | | | | | |
| Kenya | WASH−B (42) | 994 | 3480 | 3.0 | 0.64 (0.39, 1.04) | 0.10 | 0.10 | 338 | 1232 | 3.5 | 0.85 (0.44, 1.62) | 0.11 | 0.11 |
| Madagascar | MAHAY (43) | 654 | 707 | 9.1 | 0.96 (0.65, 1.42) | 0.16 | 0.16 | 280 | 245 | 9.0 | 0.99 (0.60, 1.65) | 0.17 | 0.17 |
| Malawi | iLiNS−DYADM (44) | | | | | | | | | | | | |
| Malawi | iLiNS−DOSE (45) | | | | | | | | | | | | |
| Mali | PROMIS (46) | | | | | | | | | | | | |
| Mali | PROMIS CS (46) | | | | | | | | | | | | |
| Zimbabwe | SHINE (HIV−) (47) | 1191 | 1133 | 3.4 | 0.78 (0.48, 1.24) | 0.11 | 0.11 | 508 | 486 | 3.9 | 0.70 (0.36, 1.37) | 0.10 | 0.10 |
| Zimbabwe | SHINE (HIV+) (48) | 234 | 214 | 3.7 | 1.49 (0.62, 3.54) | 0.03 | 0.03 | 88 | 99 | 6.1 | 0.94 (0.28, 3.12) | 0.03 | 0.03 |
| | | **4903** | **7032** | | **I² = 0.00, Tau² = 0.00** | | | **2035** | **2770** | | **I² = 0.00, Tau² = 0.00** | | |
| **Fixed** | | | | | **0.88 (0.76, 1.03)** | | | | | | **0.81 (0.65, 1.00)** | | |
| **Random** | | | | | **0.88 (0.76, 1.03)** | | | | | | **0.81 (0.65, 1.00)** | | |

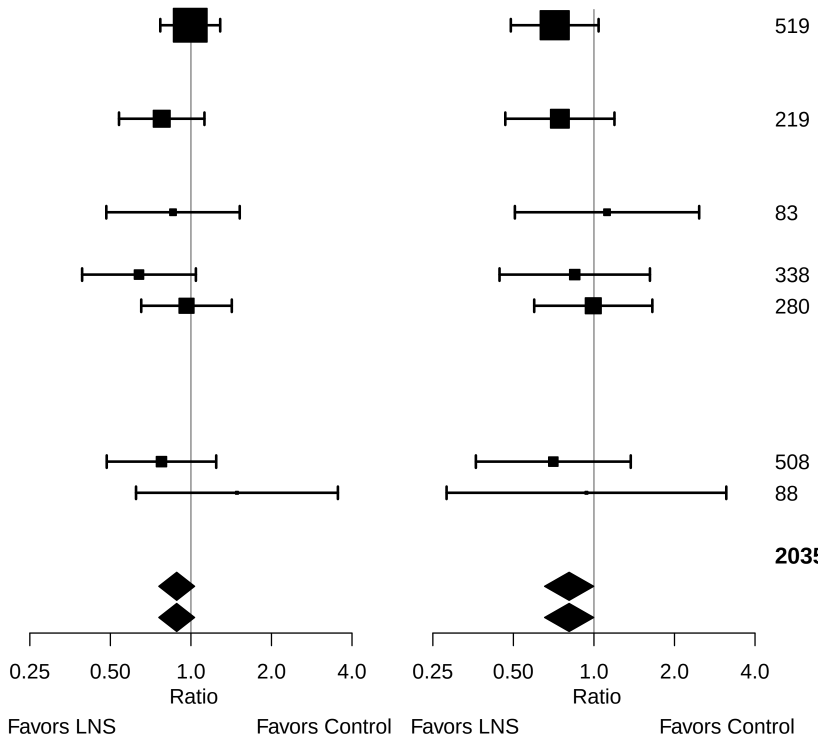

#### Supplemental figure 6J: Acute malnutrition prevalence ratio

#### 6J6: Stratified by Child sex

**P−for−interaction = 0.307**
**Ratio of PRs = 0.93 (0.81, 1.07)**

| Country | Trial | Male LNS N | Male Control N | Male Control Prevalence | Male PR (95% CI) | Male Fixed W | Male Random W | Female LNS N | Female Control N | Female Control Prevalence | Female PR (95% CI) | Female Fixed W | Female Random W |
|---|---|---|---|---|---|---|---|---|---|---|---|---|---|
| Bangladesh | JiVitA−4 (34) | 1420 | 586 | 17.8 | 1.01 (0.83, 1.22) | 0.30 | 0.30 | 1363 | 630 | 24.1 | 0.81 (0.69, 0.96) | 0.37 | 0.30 |
| Bangladesh | RDNS (35) | 832 | 411 | 15.6 | 0.90 (0.68, 1.18) | 0.15 | 0.15 | 829 | 404 | 15.1 | 0.82 (0.61, 1.11) | 0.12 | 0.12 |
| Bangladesh | WASH−B (36) | | | | | | | | | | | | |
| Burkina Faso | iLiNS−Zinc (37) | 990 | 336 | 17.3 | 0.75 (0.59, 0.96) | 0.18 | 0.18 | 961 | 328 | 21.3 | 0.62 (0.45, 0.86) | 0.10 | 0.11 |
| Burkina Faso | PROMIS (38) | 427 | 477 | 13.0 | 0.74 (0.46, 1.19) | 0.05 | 0.05 | 419 | 424 | 10.1 | 0.96 (0.62, 1.51) | 0.05 | 0.06 |
| Burkina Faso | PROMIS CS (38) | 229 | 219 | 17.8 | 1.01 (0.66, 1.52) | 0.06 | 0.06 | 195 | 215 | 20.9 | 0.91 (0.61, 1.34) | 0.07 | 0.08 |
| Ghana | GHANA (39) | | | | | | | | | | | | |
| Ghana | iLiNS−DYADG (40) | 175 | 323 | 10.5 | 0.60 (0.31, 1.15) | 0.03 | 0.03 | 172 | 369 | 6.0 | 1.27 (0.65, 2.46) | 0.02 | 0.03 |
| Haiti | HAITI (41) | | | | | | | | | | | | |
| Kenya | WASH−B (42) | 732 | 2430 | 3.1 | 0.83 (0.49, 1.40) | 0.04 | 0.04 | 722 | 2687 | 3.3 | 0.59 (0.35, 1.01) | 0.04 | 0.04 |
| Madagascar | MAHAY (43) | 812 | 814 | 10.3 | 0.88 (0.61, 1.28) | 0.08 | 0.08 | 863 | 841 | 8.6 | 1.10 (0.80, 1.50) | 0.11 | 0.11 |
| Malawi | iLiNS−DYADM (44) | | | | | | | | | | | | |
| Malawi | iLiNS−DOSE (45) | | | | | | | | | | | | |
| Mali | PROMIS (46) | | | | | | | | | | | | |
| Mali | PROMIS CS (46) | 472 | 520 | 13.3 | 0.91 (0.63, 1.31) | 0.08 | 0.08 | 472 | 439 | 12.8 | 0.86 (0.61, 1.22) | 0.09 | 0.10 |
| Zimbabwe | SHINE (HIV−) (47) | 924 | 900 | 3.9 | 0.75 (0.47, 1.21) | 0.05 | 0.05 | 943 | 877 | 2.9 | 0.86 (0.49, 1.48) | 0.03 | 0.04 |
| Zimbabwe | SHINE (HIV+) (48) | | | | | | | | | | | | |
| | | **7013** | **7016** | | **I² = 0.00, Tau² = 0.00** | | | **6939** | **7214** | | **I² = 0.10, Tau² = 0.00** | | |
| **Fixed** | | | | | **0.88 (0.79, 0.97)** | | | | | | **0.84 (0.75, 0.92)** | | |
| **Random** | | | | | **0.88 (0.79, 0.97)** | | | | | | **0.84 (0.75, 0.94)** | | |

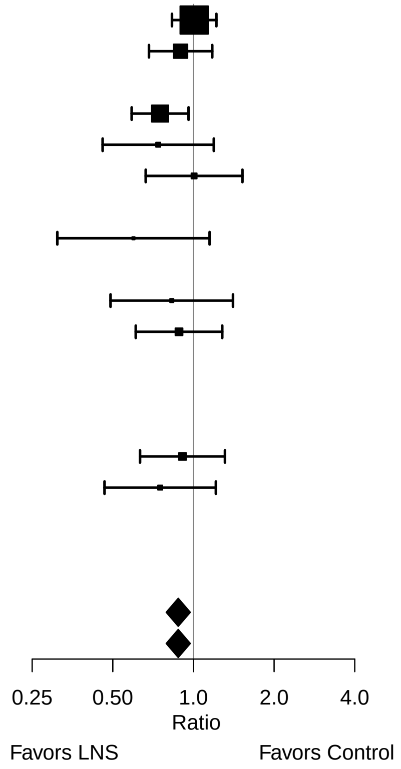

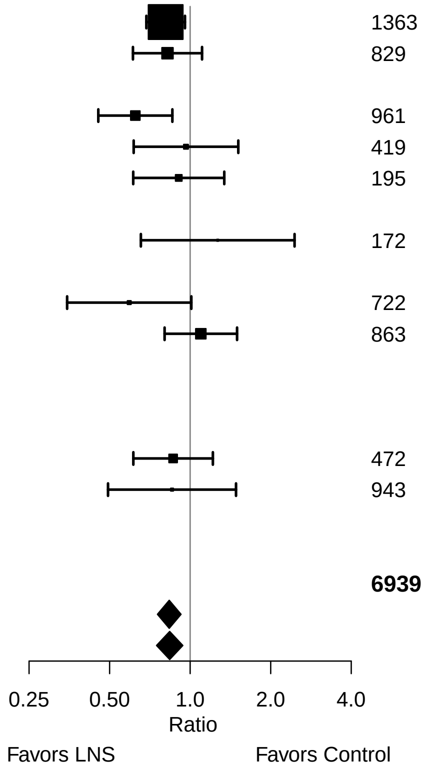

#### Supplemental figure 6J: Acute malnutrition prevalence ratio

#### 6J7: Stratified by Child birth order

**P−for−interaction = 0.124**
**Ratio of PRs = 1.14 (0.96, 1.35)**

| Country | Trial | Later born LNS N | Later born Control N | Later born Control Prevalence | Later born PR (95% CI) | Later born Fixed W | Later born Random W | Firstborn LNS N | Firstborn Control N | Firstborn Control Prevalence | Firstborn PR (95% CI) | Firstborn Fixed W | Firstborn Random W |
|---|---|---|---|---|---|---|---|---|---|---|---|---|---|
| Bangladesh | JiVitA−4 (34) | 598 | 247 | 23.1 | 0.79 (0.61, 1.03) | 0.12 | 0.12 | 2181 | 963 | 20.4 | 0.93 (0.81, 1.08) | 0.62 | 0.62 |
| Bangladesh | RDNS (35) | 972 | 508 | 16.1 | 0.85 (0.66, 1.08) | 0.15 | 0.15 | 688 | 307 | 14.0 | 0.89 (0.63, 1.27) | 0.11 | 0.11 |
| Bangladesh | WASH−B (36) | | | | | | | | | | | | |
| Burkina Faso | iLiNS−Zinc (37) | 1520 | 519 | 18.5 | 0.66 (0.53, 0.81) | 0.20 | 0.20 | 431 | 145 | 22.1 | 0.72 (0.46, 1.13) | 0.06 | 0.06 |
| Burkina Faso | PROMIS (38) | 701 | 743 | 12.0 | 0.83 (0.61, 1.15) | 0.09 | 0.09 | 145 | 157 | 10.2 | 0.81 (0.42, 1.58) | 0.03 | 0.03 |
| Burkina Faso | PROMIS CS (38) | 362 | 355 | 19.7 | 0.90 (0.67, 1.20) | 0.11 | 0.11 | 61 | 79 | 17.7 | 1.30 (0.70, 2.41) | 0.03 | 0.03 |
| Ghana | GHANA (39) | | | | | | | | | | | | |
| Ghana | iLiNS−DYADG (40) | | | | | | | | | | | | |
| Haiti | HAITI (41) | | | | | | | | | | | | |
| Kenya | WASH−B (42) | 1131 | 4026 | 3.4 | 0.69 (0.45, 1.05) | 0.05 | 0.05 | 321 | 1086 | 2.6 | 0.85 (0.39, 1.85) | 0.02 | 0.02 |
| Madagascar | MAHAY (43) | 1259 | 1196 | 10.4 | 0.93 (0.70, 1.25) | 0.11 | 0.11 | 416 | 459 | 7.0 | 1.14 (0.74, 1.75) | 0.07 | 0.07 |
| Malawi | iLiNS−DYADM (44) | | | | | | | | | | | | |
| Malawi | iLiNS−DOSE (45) | | | | | | | | | | | | |
| Mali | PROMIS (46) | | | | | | | | | | | | |
| Mali | PROMIS CS (46) | 783 | 821 | 13.0 | 0.90 (0.70, 1.16) | 0.14 | 0.14 | 144 | 126 | 11.1 | 0.75 (0.36, 1.55) | 0.02 | 0.02 |
| Zimbabwe | SHINE (HIV−) (47) | 1294 | 1211 | 3.5 | 0.71 (0.45, 1.14) | 0.04 | 0.04 | 486 | 483 | 3.5 | 0.94 (0.51, 1.73) | 0.03 | 0.03 |
| Zimbabwe | SHINE (HIV+) (48) | | | | | | | | | | | | |
| | | **8620** | **9626** | | **I² = 0.00, Tau² = 0.00** | | | **4873** | **3805** | | **I² = 0.00, Tau² = 0.00** | | |
| **Fixed** | | | | | **0.80 (0.73, 0.88)** | | | | | | **0.93 (0.83, 1.04)** | | |
| **Random** | | | | | **0.80 (0.73, 0.88)** | | | | | | **0.93 (0.83, 1.04)** | | |

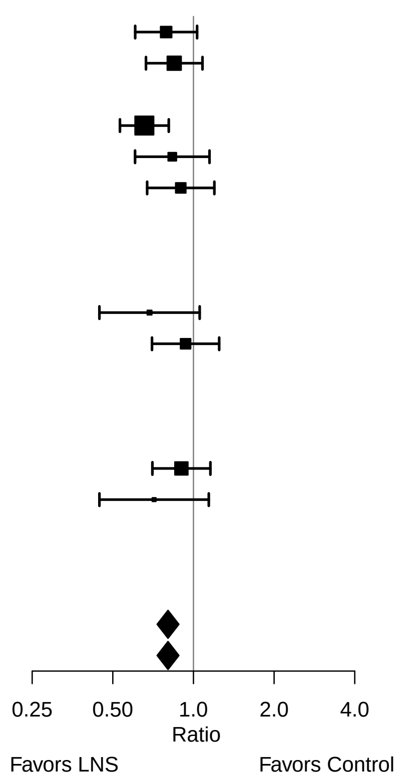

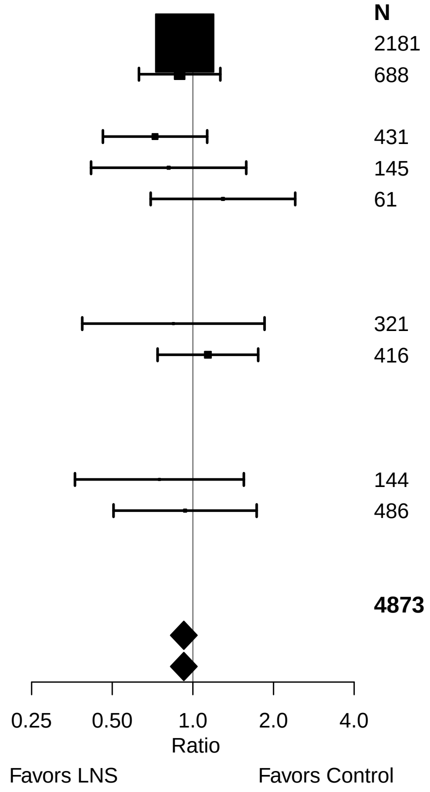

#### Supplemental figure 6J: Acute malnutrition prevalence ratio

#### 6J8: Stratified by Child baseline anthropometric status

**P−for−interaction = 0.490**
**Ratio of PRs = 0.85 (0.53, 1.36)**

| | | WLZ at least 0 Z | | | | | | WLZ less than 0 Z | | | | | |
|---|---|---|---|---|---|---|---|---|---|---|---|---|---|
| **Country** | **Trial** | **LNS N** | **Control N** | **Control Prevalence** | **PR (95% CI)** | **Fixed W** | **Random W** | **LNS N** | **Control N** | **Control Prevalence** | **PR (95% CI)** | **Fixed W** | **Random W** |
| Bangladesh | JiVitA−4 (34) | 961 | 423 | 4.0 | 0.93 (0.51, 1.71) | 0.55 | 0.55 | 1822 | 793 | 30.0 | 0.88 (0.78, 1.00) | 0.84 | 0.84 |
| Bangladesh | RDNS (35) | | | | | | | | | | | | |
| Bangladesh | WASH−B (36) | | | | | | | | | | | | |
| Burkina Faso | iLiNS−Zinc (37) | | | | | | | | | | | | |
| Burkina Faso | PROMIS (38) | 275 | 346 | 3.5 | 0.73 (0.25, 2.19) | 0.17 | 0.17 | 408 | 392 | 18.6 | 0.78 (0.56, 1.08) | 0.12 | 0.12 |
| Burkina Faso | PROMIS CS (38) | | | | | | | | | | | | |
| Ghana | GHANA (39) | | | | | | | | | | | | |
| Ghana | iLiNS−DYADG (40) | | | | | | | | | | | | |
| Haiti | HAITI (41) | | | | | | | | | | | | |
| Kenya | WASH−B (42) | | | | | | | | | | | | |
| Madagascar | MAHAY (43) | | | | | | | | | | | | |
| Malawi | iLiNS−DYADM (44) | | | | | | | | | | | | |
| Malawi | iLiNS−DOSE (45) | | | | | | | | | | | | |
| Mali | PROMIS (46) | | | | | | | | | | | | |
| Mali | PROMIS CS (46) | | | | | | | | | | | | |
| Zimbabwe | SHINE (HIV−) (47) | 742 | 699 | 1.1 | 1.18 (0.50, 2.78) | 0.28 | 0.28 | 555 | 476 | 6.5 | 0.61 (0.34, 1.09) | 0.04 | 0.04 |
| Zimbabwe | SHINE (HIV+) (48) | | | | | | | | | | | | |
| | | **1978** | **1468** | | **I² = 0.00, Tau² = 0.00** | | | **2785** | **1661** | | **I² = 0.00, Tau² = 0.00** | | |
| **Fixed** | | | | | **0.95 (0.61, 1.50)** | | | | | | **0.85 (0.76, 0.96)** | | |
| **Random** | | | | | **0.95 (0.61, 1.50)** | | | | | | **0.85 (0.76, 0.96)** | | |

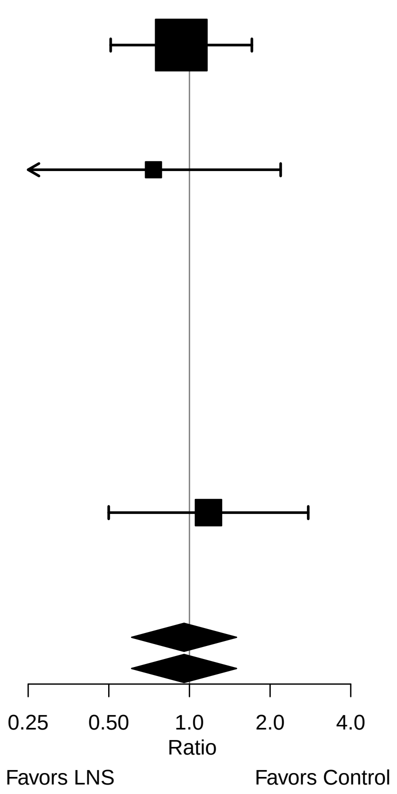

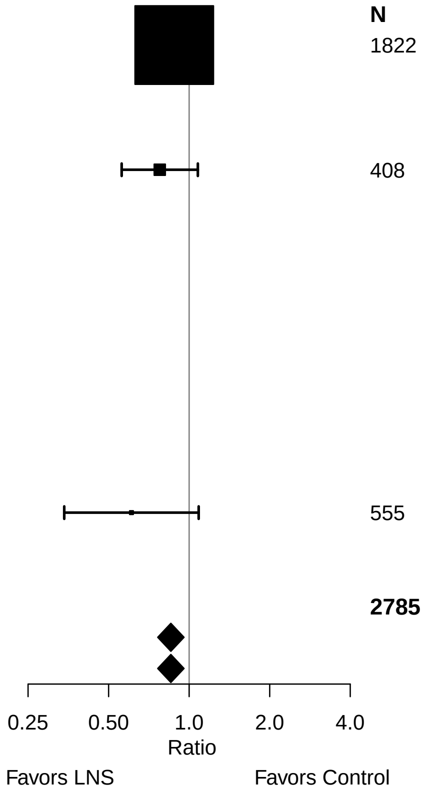

### Supplemental figure 6K: Acute malnutrition prevalence difference

#### 6K1: Stratified by Maternal height

**P−for−interaction = 0.297**
**Difference in PDs = −0.02 (−0.05, 0.02)**

| Country | Trial | LNS N | Control N | Control Prevalence | At least 150.1 cm PD (95% CI) | Fixed W | Random W | LNS N | Control N | Control Prevalence | Less than 150.1 cm PD (95% CI) | Fixed W | Random W |
|---|---|---|---|---|---|---|---|---|---|---|---|---|---|
| Bangladesh | JiVitA−4 (34) | | | | | | | | | | | | |
| Bangladesh | RDNS (35) | 868 | 432 | 14.1 | −0.02 (−0.05, 0.01) | 0.22 | 0.22 | 733 | 358 | 17.0 | −0.03 (−0.08, 0.02) | 0.33 | 0.33 |
| Bangladesh | WASH−B (36) | | | | | | | | | | | | |
| Burkina Faso | iLiNS−Zinc (37) | | | | | | | | | | | | |
| Burkina Faso | PROMIS (38) | | | | | | | | | | | | |
| Burkina Faso | PROMIS CS (38) | | | | | | | | | | | | |
| Ghana | GHANA (39) | | | | | | | | | | | | |
| Ghana | iLiNS−DYADG (40) | | | | | | | | | | | | |
| Haiti | HAITI (41) | | | | | | | | | | | | |
| Kenya | WASH−B (42) | | | | | | | | | | | | |
| Madagascar | MAHAY (43) | 983 | 1018 | 7.6 | 0.01 (−0.02, 0.04) | 0.32 | 0.32 | 594 | 557 | 12.0 | −0.01 (−0.05, 0.03) | 0.56 | 0.56 |
| Malawi | iLiNS−DYADM (44) | | | | | | | | | | | | |
| Malawi | iLiNS−DOSE (45) | 577 | 197 | 4.6 | 0.01 (−0.02, 0.05) | 0.18 | 0.18 | 112 | 42 | 11.9 | −0.02 (−0.13, 0.08) | 0.08 | 0.08 |
| Mali | PROMIS (46) | | | | | | | | | | | | |
| Mali | PROMIS CS (46) | 907 | 927 | 12.8 | −0.02 (−0.05, 0.01) | 0.28 | 0.28 | 32 | 26 | 23.1 | 0.02 (−0.16, 0.20) | 0.03 | 0.03 |
| Zimbabwe | SHINE (HIV−) (47) | | | | | | | | | | | | |
| Zimbabwe | SHINE (HIV+) (48) | | | | | | | | | | | | |
| | | **3335** | **2574** | | **I² = 0.08, Tau² = 0.00** | | | **1471** | **983** | | **I² = 0.00, Tau² = 0.00** | | |
| **Fixed** | | | | | **0.00 (−0.02, 0.01)** | | | | | | **−0.02 (−0.05, 0.01)** | | |
| **Random** | | | | | **0.00 (−0.02, 0.01)** | | | | | | **−0.02 (−0.05, 0.01)** | | |

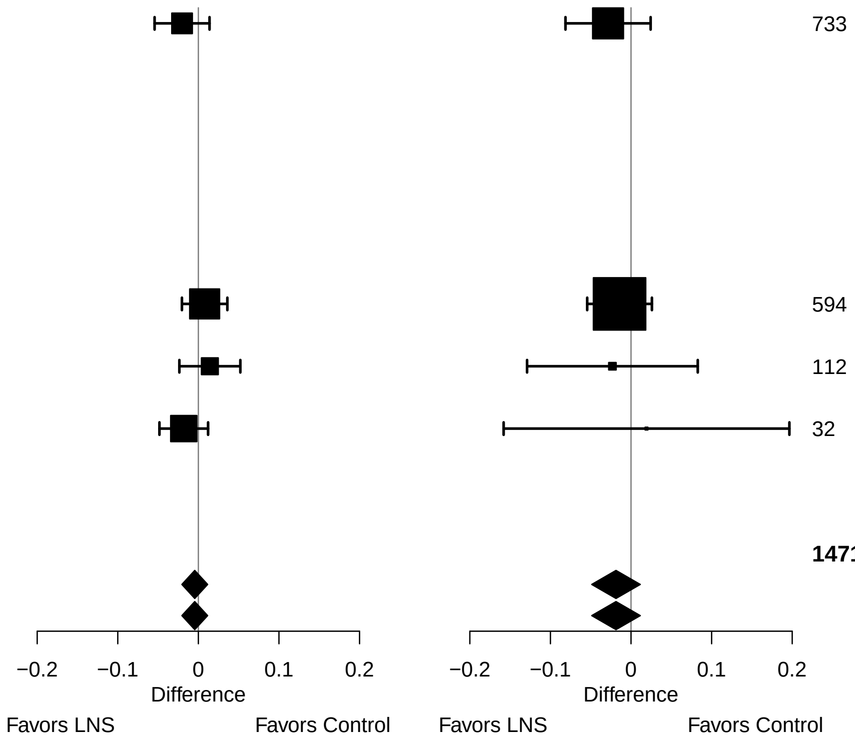

#### Supplemental figure 6K: Acute malnutrition prevalence difference

#### 6K2: Stratified by Maternal BMI

**P−for−interaction = 0.580**
**Difference in PDs = −0.01 (−0.03, 0.02)**

| Country | Trial | At least 20 kg/m²: LNS N | Control N | Control Prevalence | PD (95% CI) | Fixed W | Random W | Less than 20 kg/m²: LNS N | Control N | Control Prevalence | PD (95% CI) | Fixed W | Random W |
|---|---|---|---|---|---|---|---|---|---|---|---|---|---|
| Bangladesh | JiVitA−4 (34) | | | | | | | | | | | | |
| Bangladesh | RDNS (35) | 730 | 335 | 9.6 | 0.00 (−0.04, 0.03) | 0.08 | 0.08 | 871 | 455 | 19.8 | −0.04 (−0.08, 0.00) | 0.21 | 0.20 |
| Bangladesh | WASH−B (36) | | | | | | | | | | | | |
| Burkina Faso | iLiNS−Zinc (37) | 1214 | 390 | 15.3 | −0.06 (−0.10, −0.02) | 0.06 | 0.06 | 735 | 274 | 24.8 | −0.06 (−0.11, −0.02) | 0.18 | 0.18 |
| Burkina Faso | PROMIS (38) | 637 | 663 | 9.5 | −0.01 (−0.05, 0.03) | 0.07 | 0.07 | 206 | 232 | 17.7 | −0.05 (−0.11, 0.02) | 0.09 | 0.10 |
| Burkina Faso | PROMIS CS (38) | 241 | 265 | 15.5 | −0.02 (−0.08, 0.04) | 0.03 | 0.03 | 183 | 165 | 25.5 | −0.01 (−0.09, 0.07) | 0.06 | 0.07 |
| Ghana | GHANA (39) | | | | | | | | | | | | |
| Ghana | iLiNS−DYADG (40) | | | | | | | | | | | | |
| Haiti | HAITI (41) | | | | | | | | | | | | |
| Kenya | WASH−B (42) | | | | | | | | | | | | |
| Madagascar | MAHAY (43) | | | | | | | | | | | | |
| Malawi | iLiNS−DYADM (44) | | | | | | | | | | | | |
| Malawi | iLiNS−DOSE (45) | 509 | 174 | 5.2 | −0.01 (−0.04, 0.03) | 0.07 | 0.07 | 178 | 64 | 7.8 | 0.04 (−0.05, 0.13) | 0.05 | 0.06 |
| Mali | PROMIS (46) | | | | | | | | | | | | |
| Mali | PROMIS CS (46) | 696 | 675 | 12.3 | −0.02 (−0.06, 0.01) | 0.09 | 0.09 | 243 | 278 | 15.1 | 0.01 (−0.05, 0.06) | 0.11 | 0.12 |
| Zimbabwe | SHINE (HIV−) (47) | 1353 | 1293 | 3.4 | −0.01 (−0.02, 0.00) | 0.59 | 0.59 | 239 | 247 | 5.7 | −0.02 (−0.05, 0.02) | 0.31 | 0.27 |
| Zimbabwe | SHINE (HIV+) (48) | | | | | | | | | | | | |
| | | **5380** | **3795** | | **I² = 0.00, Tau² = 0.00** | | | **2655** | **1715** | | **I² = 0.16, Tau² = 0.00** | | |
| **Fixed** | | | | | **−0.01 (−0.02, 0.00)** | | | | | | **−0.03 (−0.05, −0.01)** | | |
| **Random** | | | | | **−0.01 (−0.02, 0.00)** | | | | | | **−0.03 (−0.05, 0.00)** | | |

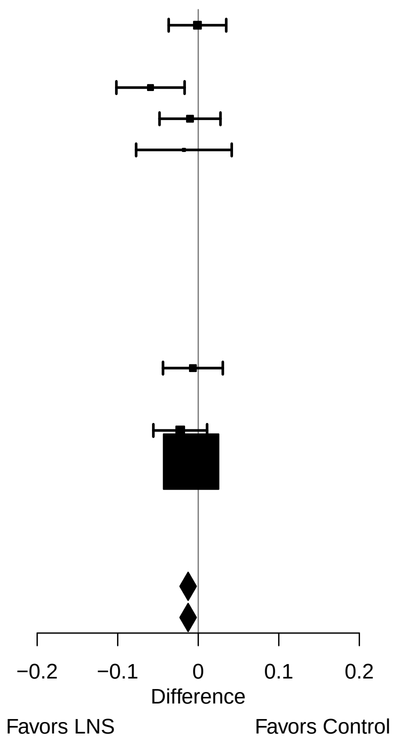

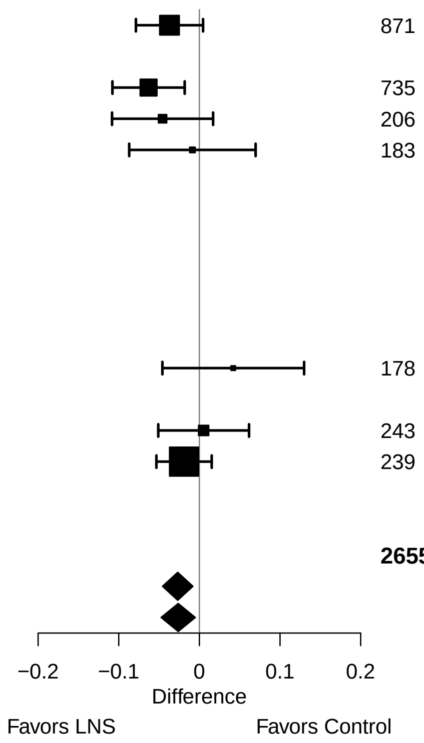

#### Supplemental figure 6K: Acute malnutrition prevalence difference

### 6K3: Stratified by Maternal age

**P−for−interaction = 0.115**
**Difference in PDs = 0.01 (0.00, 0.02)**

| | | At least 25 y | | | | | | Less than 25 y | | | | | |
|---|---|---|---|---|---|---|---|---|---|---|---|---|---|
| **Country** | **Trial** | **LNS N** | **Control N** | **Control Prevalence** | **PD (95% CI)** | **Fixed W** | **Random W** | **LNS N** | **Control N** | **Control Prevalence** | **PD (95% CI)** | **Fixed W** | **Random W** |
| Bangladesh | JiVitA−4 (34) | 1191 | 534 | 21.5 | −0.02 (−0.06, 0.01) | 0.05 | 0.09 | 1580 | 677 | 20.5 | −0.02 (−0.05, 0.01) | 0.07 | 0.07 |
| Bangladesh | RDNS (35) | 453 | 221 | 14.0 | 0.03 (−0.03, 0.08) | 0.03 | 0.05 | 1208 | 594 | 15.8 | −0.04 (−0.08, 0.00) | 0.05 | 0.05 |
| Bangladesh | WASH−B (36) | | | | | | | | | | | | |
| Burkina Faso | iLiNS−Zinc (37) | 1131 | 399 | 19.5 | −0.07 (−0.12, −0.03) | 0.04 | 0.07 | 807 | 262 | 18.7 | −0.05 (−0.10, 0.01) | 0.02 | 0.02 |
| Burkina Faso | PROMIS (38) | 476 | 519 | 11.4 | −0.02 (−0.06, 0.02) | 0.04 | 0.09 | 370 | 382 | 12.0 | −0.02 (−0.07, 0.04) | 0.02 | 0.02 |
| Burkina Faso | PROMIS CS (38) | 200 | 189 | 20.6 | −0.02 (−0.11, 0.06) | 0.01 | 0.02 | 224 | 245 | 18.4 | 0.00 (−0.06, 0.06) | 0.02 | 0.02 |
| Ghana | GHANA (39) | | | | | | | | | | | | |
| Ghana | iLiNS−DYADG (40) | | | | | | | | | | | | |
| Haiti | HAITI (41) | | | | | | | | | | | | |
| Kenya | WASH−B (42) | 841 | 2801 | 3.2 | −0.01 (−0.02, 0.00) | 0.55 | 0.28 | 598 | 2284 | 3.2 | −0.01 (−0.02, 0.01) | 0.34 | 0.34 |
| Madagascar | MAHAY (43) | 883 | 930 | 10.5 | 0.00 (−0.03, 0.03) | 0.07 | 0.12 | 791 | 725 | 8.0 | 0.00 (−0.03, 0.03) | 0.08 | 0.08 |
| Malawi | iLiNS−DYADM (44) | | | | | | | | | | | | |
| Malawi | iLiNS−DOSE (45) | | | | | | | | | | | | |
| Mali | PROMIS (46) | | | | | | | | | | | | |
| Mali | PROMIS CS (46) | 498 | 501 | 13.8 | −0.03 (−0.07, 0.02) | 0.03 | 0.07 | 446 | 458 | 12.2 | 0.00 (−0.04, 0.04) | 0.04 | 0.04 |
| Zimbabwe | SHINE (HIV−) (47) | 870 | 794 | 4.5 | −0.02 (−0.04, 0.00) | 0.18 | 0.20 | 800 | 787 | 2.4 | 0.00 (−0.01, 0.02) | 0.36 | 0.36 |
| Zimbabwe | SHINE (HIV+) (48) | | | | | | | | | | | | |
| | | **6543** | **6888** | | **I² = 0.21, Tau² = 0.00** | | | **6824** | **6414** | | **I² = 0.00, Tau² = 0.00** | | |
| **Fixed** | | | | | **−0.02 (−0.02, −0.01)** | | | | | | **−0.01 (−0.01, 0.00)** | | |
| **Random** | | | | | **−0.02 (−0.03, 0.00)** | | | | | | **−0.01 (−0.01, 0.00)** | | |

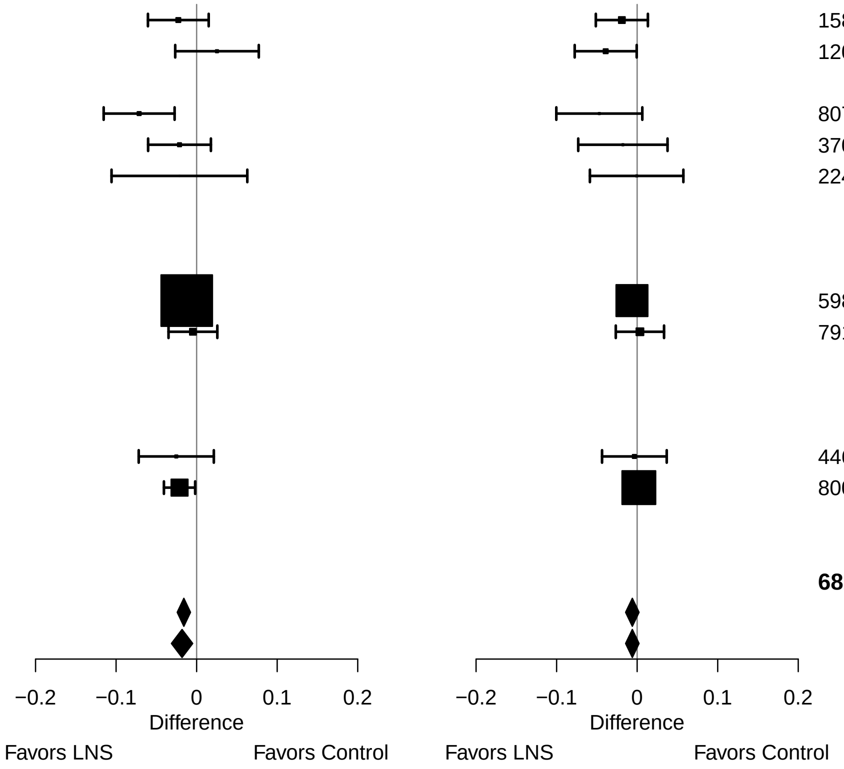

#### Supplemental figure 6K: Acute malnutrition prevalence difference

#### 6K4: Stratified by Maternal education

**P−for−interaction = 0.659**
**Difference in PDs = 0.00 (−0.02, 0.01)**

| | | Primary or greater | | | | | | Incomplete or no formal | | | | | |
|---|---|---|---|---|---|---|---|---|---|---|---|---|---|
| Country | Trial | LNS N | Control N | Control Prevalence | PD (95% CI) | Fixed W | Random W | LNS N | Control N | Control Prevalence | PD (95% CI) | Fixed W | Random W |
| Bangladesh | JiVitA−4 (34) | 1797 | 739 | 20.2 | −0.03 (−0.05, 0.00) | 0.11 | 0.16 | 984 | 474 | 22.3 | −0.02 (−0.06, 0.03) | 0.06 | 0.06 |
| Bangladesh | RDNS (35) | 1245 | 589 | 15.1 | −0.03 (−0.06, 0.01) | 0.07 | 0.12 | 416 | 226 | 15.9 | 0.00 (−0.05, 0.05) | 0.04 | 0.04 |
| Bangladesh | WASH−B (36) | | | | | | | | | | | | |
| Burkina Faso | iLiNS−Zinc (37) | | | | | | | | | | | | |
| Burkina Faso | PROMIS (38) | | | | | | | | | | | | |
| Burkina Faso | PROMIS CS (38) | 34 | 39 | 20.5 | 0.03 (−0.20, 0.26) | 0.00 | 0.00 | 388 | 394 | 19.3 | −0.01 (−0.06, 0.04) | 0.04 | 0.04 |
| Ghana | GHANA (39) | | | | | | | | | | | | |
| Ghana | iLiNS−DYADG (40) | 267 | 548 | 7.3 | −0.01 (−0.04, 0.03) | 0.06 | 0.10 | 80 | 144 | 11.1 | −0.04 (−0.12, 0.05) | 0.02 | 0.02 |
| Haiti | HAITI (41) | | | | | | | | | | | | |
| Kenya | WASH−B (42) | 700 | 2426 | 2.5 | −0.01 (−0.02, 0.01) | 0.65 | 0.46 | 752 | 2687 | 3.8 | −0.01 (−0.02, 0.00) | 0.62 | 0.62 |
| Madagascar | MAHAY (43) | 342 | 420 | 5.5 | 0.01 (−0.02, 0.05) | 0.08 | 0.13 | 1333 | 1235 | 10.8 | −0.01 (−0.04, 0.02) | 0.11 | 0.11 |
| Malawi | iLiNS−DYADM (44) | | | | | | | | | | | | |
| Malawi | iLiNS−DOSE (45) | | | | | | | | | | | | |
| Mali | PROMIS (46) | | | | | | | | | | | | |
| Mali | PROMIS CS (46) | 107 | 96 | 13.5 | −0.08 (−0.16, 0.00) | 0.01 | 0.03 | 836 | 863 | 13.0 | −0.01 (−0.04, 0.02) | 0.12 | 0.12 |
| Zimbabwe | SHINE (HIV−) (47) | | | | | | | | | | | | |
| Zimbabwe | SHINE (HIV+) (48) | | | | | | | | | | | | |
| | | **4492** | **4857** | | **I² = 0.18, Tau² = 0.00** | | | **4789** | **6023** | | **I² = 0.00, Tau² = 0.00** | | |
| **Fixed** | | | | | **−0.01 (−0.02, 0.00)** | | | | | | **−0.01 (−0.02, 0.00)** | | |
| **Random** | | | | | **−0.01 (−0.02, 0.00)** | | | | | | **−0.01 (−0.02, 0.00)** | | |

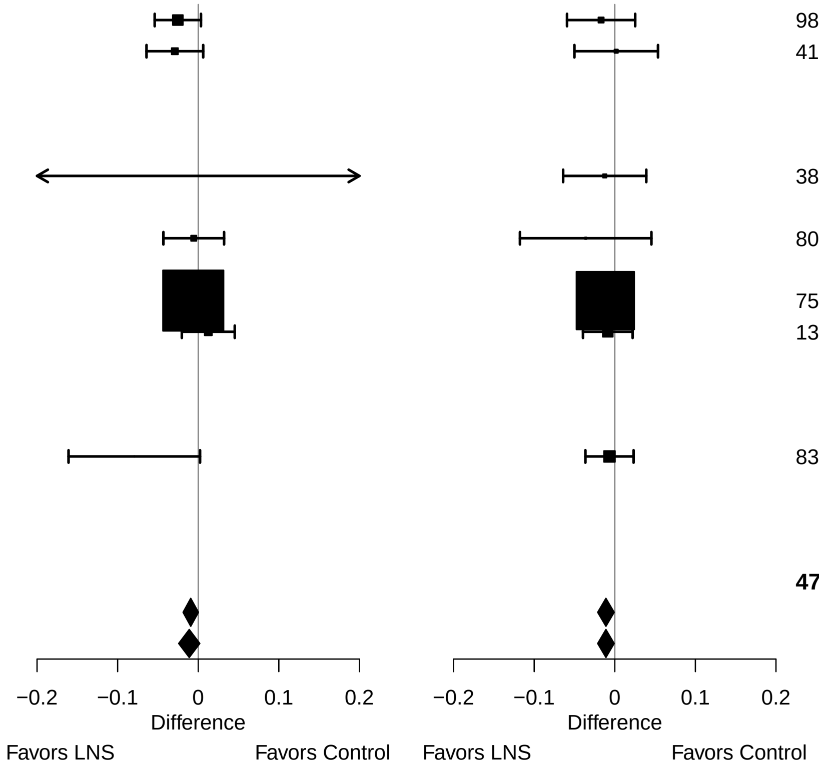

#### Supplemental figure 6K: Acute malnutrition prevalence difference

##### 6K5: Stratified by Maternal depressive symptoms

**P−for−interaction = 0.781**
**Difference in PDs = 0.00 (−0.01, 0.02)**

| Country | Trial | LNS N | Control N | Control Prevalence | Less than 75th percentile PD (95% CI) | Fixed W | Random W | LNS N | Control N | Control Prevalence | At least 75th percentile PD (95% CI) | Fixed W | Random W |
|---|---|---|---|---|---|---|---|---|---|---|---|---|---|
| Bangladesh | JiVitA−4 (34) | | | | | | | | | | | | |
| Bangladesh | RDNS (35) | 1081 | 472 | 13.8 | 0.00 (−0.04, 0.03) | 0.04 | 0.04 | 519 | 288 | 17.0 | −0.05 (−0.11, 0.01) | 0.05 | 0.05 |
| Bangladesh | WASH−B (36) | | | | | | | | | | | | |
| Burkina Faso | iLiNS−Zinc (37) | | | | | | | | | | | | |
| Burkina Faso | PROMIS (38) | 497 | 567 | 11.6 | −0.03 (−0.06, 0.01) | 0.04 | 0.04 | 219 | 211 | 14.7 | −0.04 (−0.10, 0.02) | 0.04 | 0.04 |
| Burkina Faso | PROMIS CS (38) | | | | | | | | | | | | |
| Ghana | GHANA (39) | | | | | | | | | | | | |
| Ghana | iLiNS−DYADG (40) | 252 | 459 | 7.4 | −0.01 (−0.05, 0.03) | 0.03 | 0.03 | 83 | 209 | 8.6 | 0.01 (−0.06, 0.08) | 0.03 | 0.03 |
| Haiti | HAITI (41) | | | | | | | | | | | | |
| Kenya | WASH−B (42) | 994 | 3480 | 3.0 | −0.01 (−0.02, 0.00) | 0.52 | 0.52 | 338 | 1232 | 3.5 | −0.01 (−0.02, 0.01) | 0.42 | 0.42 |
| Madagascar | MAHAY (43) | 654 | 707 | 9.1 | 0.00 (−0.04, 0.03) | 0.04 | 0.04 | 280 | 245 | 9.0 | 0.00 (−0.05, 0.05) | 0.08 | 0.08 |
| Malawi | iLiNS−DYADM (44) | | | | | | | | | | | | |
| Malawi | iLiNS−DOSE (45) | | | | | | | | | | | | |
| Mali | PROMIS (46) | | | | | | | | | | | | |
| Mali | PROMIS CS (46) | | | | | | | | | | | | |
| Zimbabwe | SHINE (HIV−) (47) | 1191 | 1133 | 3.4 | −0.01 (−0.02, 0.01) | 0.28 | 0.28 | 508 | 486 | 3.9 | −0.01 (−0.03, 0.01) | 0.35 | 0.35 |
| Zimbabwe | SHINE (HIV+) (48) | 234 | 214 | 3.7 | 0.02 (−0.02, 0.06) | 0.03 | 0.03 | 88 | 99 | 6.1 | 0.00 (−0.07, 0.07) | 0.03 | 0.03 |
| | | **4903** | **7032** | | **I² = 0.00, Tau² = 0.00** | | | **2035** | **2770** | | **I² = 0.00, Tau² = 0.00** | | |
| **Fixed** | | | | | **−0.01 (−0.02, 0.00)** | | | | | | **−0.01 (−0.02, 0.00)** | | |
| **Random** | | | | | **−0.01 (−0.02, 0.00)** | | | | | | **−0.01 (−0.02, 0.00)** | | |

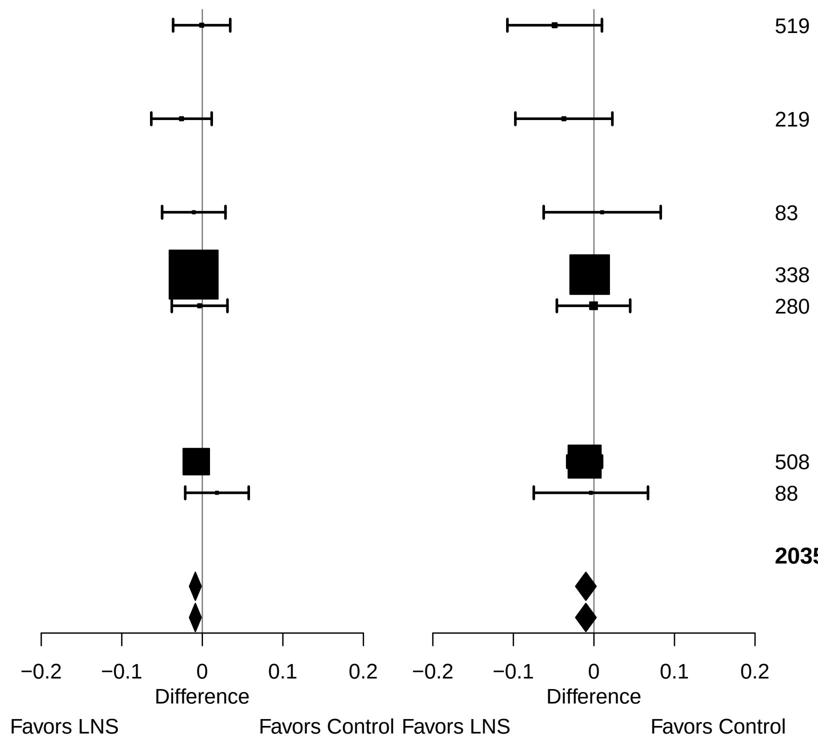

#### Supplemental figure 6K: Acute malnutrition prevalence difference

#### 6K6: Stratified by Child sex

**P−for−interaction = 0.708**
**Difference in PDs = 0.00 (−0.01, 0.01)**

| Country | Trial | Male LNS N | Male Control N | Male Control Prevalence | Male PD (95% CI) | Male Fixed W | Male Random W | Female LNS N | Female Control N | Female Control Prevalence | Female PD (95% CI) | Female Fixed W | Female Random W |
|---|---|---|---|---|---|---|---|---|---|---|---|---|---|
| Bangladesh | JiVitA−4 (34) | 1420 | 586 | 17.8 | 0.00 (−0.03, 0.03) | 0.07 | 0.07 | 1363 | 630 | 24.1 | −0.04 (−0.08, −0.01) | 0.05 | 0.10 |
| Bangladesh | RDNS (35) | 832 | 411 | 15.6 | −0.02 (−0.06, 0.02) | 0.04 | 0.04 | 829 | 404 | 15.1 | −0.03 (−0.07, 0.01) | 0.03 | 0.08 |
| Bangladesh | WASH−B (36) | | | | | | | | | | | | |
| Burkina Faso | iLiNS−Zinc (37) | 990 | 336 | 17.3 | −0.04 (−0.07, −0.01) | 0.07 | 0.07 | 961 | 328 | 21.3 | −0.08 (−0.14, −0.03) | 0.02 | 0.05 |
| Burkina Faso | PROMIS (38) | 427 | 477 | 13.0 | −0.03 (−0.09, 0.02) | 0.03 | 0.03 | 419 | 424 | 10.1 | 0.00 (−0.05, 0.04) | 0.03 | 0.07 |
| Burkina Faso | PROMIS CS (38) | 229 | 219 | 17.8 | 0.00 (−0.07, 0.08) | 0.01 | 0.01 | 195 | 215 | 20.9 | −0.02 (−0.10, 0.06) | 0.01 | 0.03 |
| Ghana | GHANA (39) | | | | | | | | | | | | |
| Ghana | iLiNS−DYADG (40) | 175 | 323 | 10.5 | −0.04 (−0.10, 0.01) | 0.03 | 0.03 | 172 | 369 | 6.0 | 0.02 (−0.03, 0.06) | 0.03 | 0.07 |
| Haiti | HAITI (41) | | | | | | | | | | | | |
| Kenya | WASH−B (42) | 732 | 2430 | 3.1 | −0.01 (−0.02, 0.01) | 0.37 | 0.37 | 722 | 2687 | 3.3 | −0.01 (−0.03, 0.00) | 0.43 | 0.20 |
| Madagascar | MAHAY (43) | 812 | 814 | 10.3 | −0.01 (−0.05, 0.02) | 0.06 | 0.06 | 863 | 841 | 8.6 | 0.01 (−0.02, 0.04) | 0.08 | 0.12 |
| Malawi | iLiNS−DYADM (44) | | | | | | | | | | | | |
| Malawi | iLiNS−DOSE (45) | | | | | | | | | | | | |
| Mali | PROMIS (46) | | | | | | | | | | | | |
| Mali | PROMIS CS (46) | 472 | 520 | 13.3 | −0.01 (−0.06, 0.03) | 0.03 | 0.03 | 472 | 439 | 12.8 | −0.02 (−0.06, 0.02) | 0.04 | 0.08 |
| Zimbabwe | SHINE (HIV−) (47) | 924 | 900 | 3.9 | −0.01 (−0.03, 0.01) | 0.28 | 0.28 | 943 | 877 | 2.9 | 0.00 (−0.02, 0.01) | 0.28 | 0.18 |
| Zimbabwe | SHINE (HIV+) (48) | | | | | | | | | | | | |
| | | **7013** | **7016** | | **I² = 0.00, Tau² = 0.00** | | | **6939** | **7214** | | **I² = 0.41, Tau² = 0.00** | | |
| **Fixed** | | | | | **−0.01 (−0.02, 0.00)** | | | | | | **−0.01 (−0.02, 0.00)** | | |
| **Random** | | | | | **−0.01 (−0.02, 0.00)** | | | | | | **−0.01 (−0.03, 0.00)** | | |

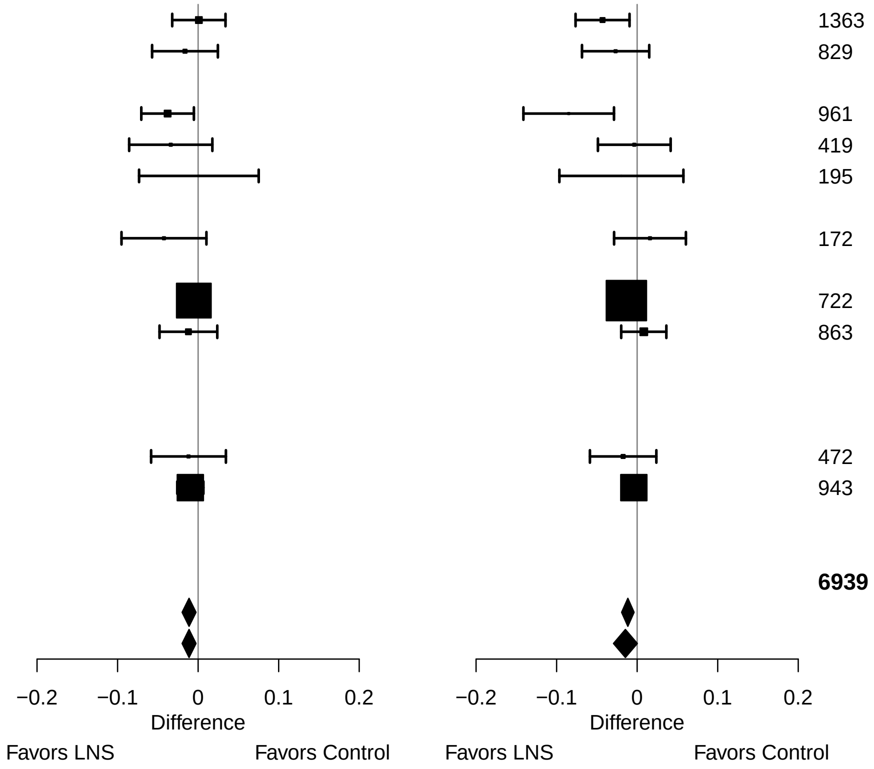

#### Supplemental figure 6K: Acute malnutrition prevalence difference

#### 6K7: Stratified by Child birth order

**P−for−interaction = 0.221**
**Difference in PDs = 0.01 (−0.01, 0.02)**

| | | Later born | | | | | | Firstborn | | | | | |
|---|---|---|---|---|---|---|---|---|---|---|---|---|---|
| **Country** | **Trial** | **LNS N** | **Control N** | **Control Prevalence** | **PD (95% CI)** | **Fixed W** | **Random W** | **LNS N** | **Control N** | **Control Prevalence** | **PD (95% CI)** | **Fixed W** | **Random W** |
| Bangladesh | JiVitA−4 (34) | 598 | 247 | 23.1 | −0.04 (−0.10, 0.01) | 0.02 | 0.04 | 2181 | 963 | 20.4 | −0.02 (−0.04, 0.01) | 0.15 | 0.15 |
| Bangladesh | RDNS (35) | 972 | 508 | 16.1 | −0.02 (−0.06, 0.01) | 0.04 | 0.07 | 688 | 307 | 14.0 | −0.02 (−0.06, 0.03) | 0.05 | 0.05 |
| Bangladesh | WASH−B (36) | | | | | | | | | | | | |
| Burkina Faso | iLiNS−Zinc (37) | 1520 | 519 | 18.5 | −0.06 (−0.09, −0.03) | 0.05 | 0.08 | 431 | 145 | 22.1 | −0.06 (−0.14, 0.01) | 0.02 | 0.02 |
| Burkina Faso | PROMIS (38) | 701 | 743 | 12.0 | −0.02 (−0.06, 0.02) | 0.04 | 0.08 | 145 | 157 | 10.2 | −0.02 (−0.08, 0.04) | 0.03 | 0.03 |
| Burkina Faso | PROMIS CS (38) | 362 | 355 | 19.7 | −0.02 (−0.07, 0.03) | 0.02 | 0.04 | 61 | 79 | 17.7 | 0.05 (−0.08, 0.19) | 0.01 | 0.01 |
| Ghana | GHANA (39) | | | | | | | | | | | | |
| Ghana | iLiNS−DYADG (40) | | | | | | | | | | | | |
| Haiti | HAITI (41) | | | | | | | | | | | | |
| Kenya | WASH−B (42) | 1131 | 4026 | 3.4 | −0.01 (−0.02, 0.00) | 0.47 | 0.28 | 321 | 1086 | 2.6 | 0.00 (−0.02, 0.01) | 0.36 | 0.36 |
| Madagascar | MAHAY (43) | 1259 | 1196 | 10.4 | −0.01 (−0.04, 0.02) | 0.06 | 0.10 | 416 | 459 | 7.0 | 0.01 (−0.02, 0.04) | 0.11 | 0.11 |
| Malawi | iLiNS−DYADM (44) | | | | | | | | | | | | |
| Malawi | iLiNS−DOSE (45) | | | | | | | | | | | | |
| Mali | PROMIS (46) | | | | | | | | | | | | |
| Mali | PROMIS CS (46) | 783 | 821 | 13.0 | −0.01 (−0.04, 0.02) | 0.05 | 0.09 | 144 | 126 | 11.1 | −0.03 (−0.10, 0.05) | 0.02 | 0.02 |
| Zimbabwe | SHINE (HIV−) (47) | 1294 | 1211 | 3.5 | −0.01 (−0.02, 0.00) | 0.26 | 0.23 | 486 | 483 | 3.5 | 0.00 (−0.02, 0.02) | 0.25 | 0.25 |
| Zimbabwe | SHINE (HIV+) (48) | | | | | | | | | | | | |
| | | **8620** | **9626** | | **I² = 0.21, Tau² = 0.00** | | | **4873** | **3805** | | **I² = 0.00, Tau² = 0.00** | | |
| **Fixed** | | | | | **−0.01 (−0.02, −0.01)** | | | | | | **−0.01 (−0.02, 0.00)** | | |
| **Random** | | | | | **−0.02 (−0.03, −0.01)** | | | | | | **−0.01 (−0.02, 0.00)** | | |

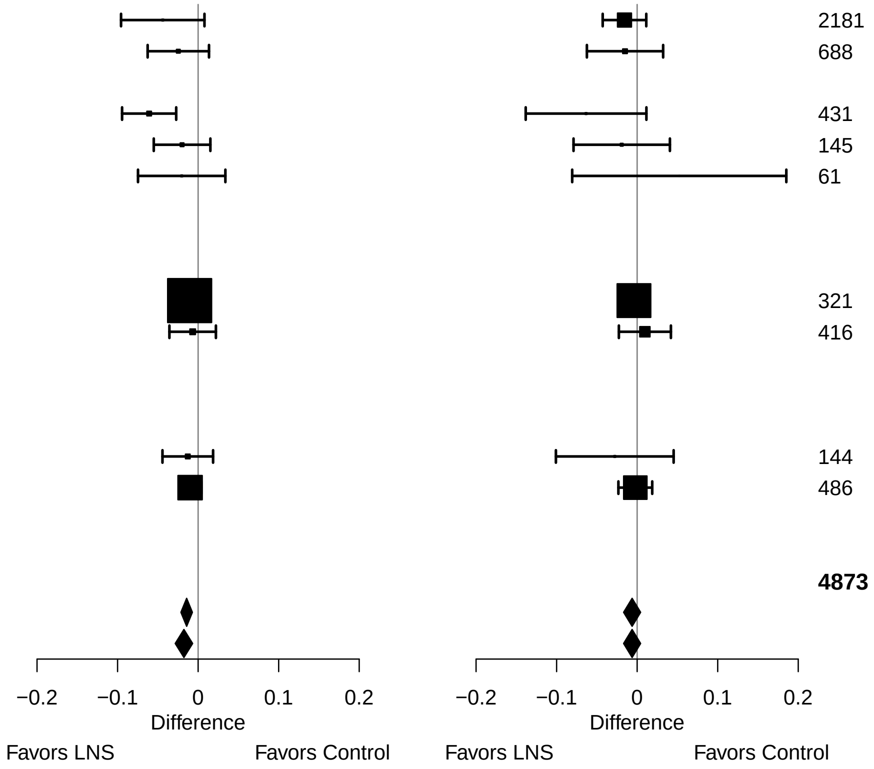

#### Supplemental figure 6K: Acute malnutrition prevalence difference

#### 6K8: Stratified by Child baseline anthropometric status

**P−for−interaction = 0.010**
**Difference in PDs = −0.03 (−0.05, −0.01)**

| | | WLZ at least 0 Z | | | | | | WLZ less than 0 Z | | | | | |
|---|---|---|---|---|---|---|---|---|---|---|---|---|---|
| **Country** | **Trial** | **LNS N** | **Control N** | **Control Prevalence** | **PD (95% CI)** | **Fixed W** | **Random W** | **LNS N** | **Control N** | **Control Prevalence** | **PD (95% CI)** | **Fixed W** | **Random W** |
| Bangladesh | JiVitA−4 (34) | 961 | 423 | 4.0 | 0.00 (−0.03, 0.02) | 0.15 | 0.15 | 1822 | 793 | 30.0 | −0.04 (−0.07, 0.00) | 0.34 | 0.34 |
| Bangladesh | RDNS (35) | | | | | | | | | | | | |
| Bangladesh | WASH−B (36) | | | | | | | | | | | | |
| Burkina Faso | iLiNS−Zinc (37) | | | | | | | | | | | | |
| Burkina Faso | PROMIS (38) | 275 | 346 | 3.5 | −0.01 (−0.04, 0.02) | 0.08 | 0.08 | 408 | 392 | 18.6 | −0.04 (−0.09, 0.01) | 0.16 | 0.16 |
| Burkina Faso | PROMIS CS (38) | | | | | | | | | | | | |
| Ghana | GHANA (39) | | | | | | | | | | | | |
| Ghana | iLiNS−DYADG (40) | | | | | | | | | | | | |
| Haiti | HAITI (41) | | | | | | | | | | | | |
| Kenya | WASH−B (42) | | | | | | | | | | | | |
| Madagascar | MAHAY (43) | | | | | | | | | | | | |
| Malawi | iLiNS−DYADM (44) | | | | | | | | | | | | |
| Malawi | iLiNS−DOSE (45) | | | | | | | | | | | | |
| Mali | PROMIS (46) | | | | | | | | | | | | |
| Mali | PROMIS CS (46) | | | | | | | | | | | | |
| Zimbabwe | SHINE (HIV−) (47) | 742 | 699 | 1.1 | 0.00 (−0.01, 0.01) | 0.77 | 0.77 | 555 | 476 | 6.5 | −0.03 (−0.05, 0.00) | 0.50 | 0.50 |
| Zimbabwe | SHINE (HIV+) (48) | | | | | | | | | | | | |
| | | **1978** | **1468** | | **I² = 0.00, Tau² = 0.00** | | | **2785** | **1661** | | **I² = 0.00, Tau² = 0.00** | | |
| **Fixed** | | | | | **0.00 (−0.01, 0.01)** | | | | | | **−0.03 (−0.05, −0.01)** | | |
| **Random** | | | | | **0.00 (−0.01, 0.01)** | | | | | | **−0.03 (−0.05, −0.01)** | | |

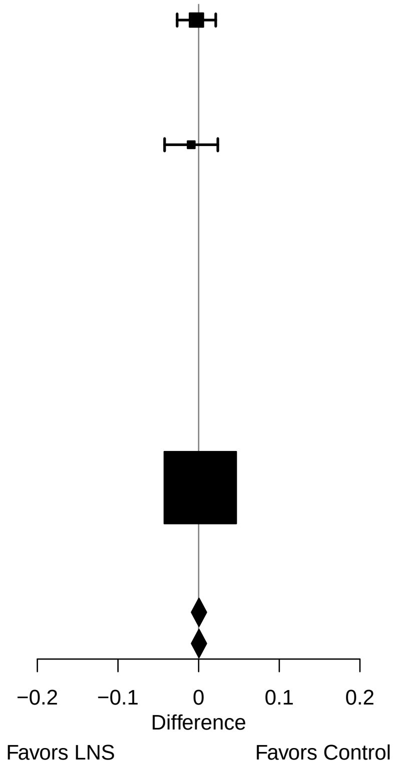

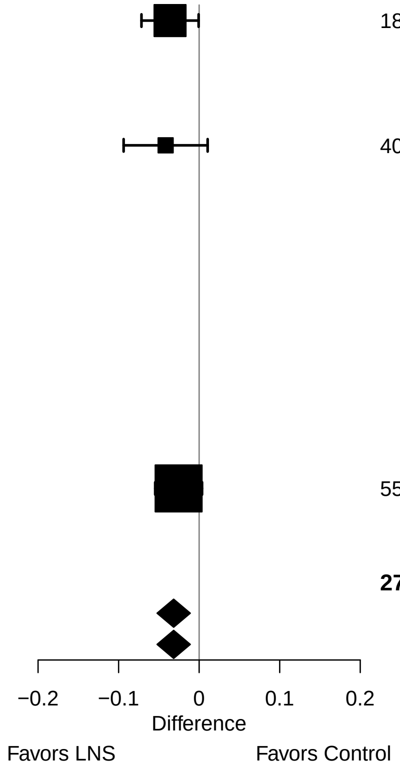

### Supplemental figure 6L: Mean difference in WAZ

#### 6L1: Stratified by Maternal height

**P−for−interaction = 0.234**
**Difference in MDs = −0.04 (−0.11, 0.03)**

| Country | Trial | LNS N | Control N | Control Mean | At least 150.1 cm MD (95% CI) | Fixed W | Random W | LNS N | Control N | Control Mean | Less than 150.1 cm MD (95% CI) | Fixed W | Random W |
|---|---|---|---|---|---|---|---|---|---|---|---|---|---|
| Bangladesh | JiVitA−4 (34) | | | | | | | | | | | | |
| Bangladesh | RDNS (35) | 868 | 432 | −1.58 | 0.13 (0.02, 0.24) | 0.06 | 0.07 | 735 | 358 | −1.97 | 0.09 (−0.02, 0.21) | 0.26 | 0.26 |
| Bangladesh | WASH−B (36) | 620 | 1898 | −1.37 | 0.21 (0.12, 0.30) | 0.09 | 0.08 | 543 | 1557 | −1.78 | 0.19 (0.10, 0.29) | 0.39 | 0.39 |
| Burkina Faso | iLiNS−Zinc (37) | 1924 | 651 | −1.53 | 0.28 (0.21, 0.35) | 0.16 | 0.09 | 34 | 15 | −1.84 | 0.00 (−0.40, 0.40) | 0.02 | 0.02 |
| Burkina Faso | PROMIS (38) | 829 | 881 | −1.20 | 0.15 (0.03, 0.28) | 0.05 | 0.07 | 25 | 25 | −1.86 | 0.04 (−0.43, 0.51) | 0.02 | 0.02 |
| Burkina Faso | PROMIS CS (38) | 419 | 423 | −1.24 | −0.05 (−0.19, 0.10) | 0.03 | 0.06 | 11 | 9 | −2.14 | 0.05 (−0.92, 1.02) | 0.00 | 0.00 |
| Ghana | GHANA (39) | | | | | | | | | | | | |
| Ghana | iLiNS−DYADG (40) | 326 | 646 | −0.83 | 0.16 (0.02, 0.30) | 0.04 | 0.06 | 16 | 36 | −1.26 | 0.10 (−0.46, 0.66) | 0.01 | 0.01 |
| Haiti | HAITI (41) | | | | | | | | | | | | |
| Kenya | WASH−B (42) | 1316 | 4709 | −0.72 | 0.13 (0.06, 0.20) | 0.15 | 0.09 | 57 | 195 | −1.17 | 0.09 (−0.23, 0.41) | 0.04 | 0.04 |
| Madagascar | MAHAY (43) | 1003 | 1040 | −1.33 | 0.02 (−0.10, 0.14) | 0.05 | 0.07 | 598 | 565 | −1.67 | 0.04 (−0.13, 0.20) | 0.13 | 0.13 |
| Malawi | iLiNS−DYADM (44) | 192 | 381 | −0.82 | −0.05 (−0.23, 0.12) | 0.02 | 0.05 | 28 | 60 | −1.21 | −0.24 (−0.72, 0.23) | 0.02 | 0.02 |
| Malawi | iLiNS−DOSE (45) | 586 | 198 | −0.99 | 0.03 (−0.09, 0.14) | 0.06 | 0.07 | 115 | 42 | −1.51 | 0.20 (−0.07, 0.47) | 0.05 | 0.05 |
| Mali | PROMIS (46) | 475 | 492 | −1.14 | 0.18 (0.06, 0.31) | 0.05 | 0.07 | 19 | 9 | −1.32 | −0.29 (−0.83, 0.25) | 0.01 | 0.01 |
| Mali | PROMIS CS (46) | 909 | 928 | −1.33 | 0.25 (0.13, 0.38) | 0.05 | 0.07 | 32 | 26 | −1.75 | −0.17 (−0.69, 0.35) | 0.01 | 0.01 |
| Zimbabwe | SHINE (HIV−) (47) | 1745 | 1690 | −0.75 | 0.15 (0.08, 0.22) | 0.16 | 0.09 | 75 | 62 | −1.04 | −0.04 (−0.33, 0.25) | 0.04 | 0.04 |
| Zimbabwe | SHINE (HIV+) (48) | 313 | 311 | −0.92 | 0.00 (−0.18, 0.17) | 0.02 | 0.05 | 18 | 12 | −1.60 | 0.33 (−0.49, 1.15) | 0.01 | 0.01 |
| | | **11525** | **14680** | | **I² = 0.70, Tau² = 0.01** | | | **2306** | **2971** | | **I² = 0.00, Tau² = 0.00** | | |
| **Fixed** | | | | | **0.15 (0.12, 0.18)** | | | | | | **0.11 (0.05, 0.17)** | | |
| **Random** | | | | | **0.13 (0.07, 0.18)** | | | | | | **0.11 (0.05, 0.17)** | | |

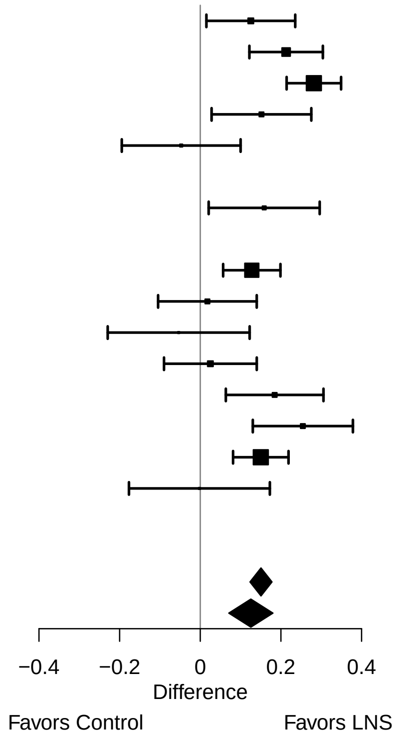

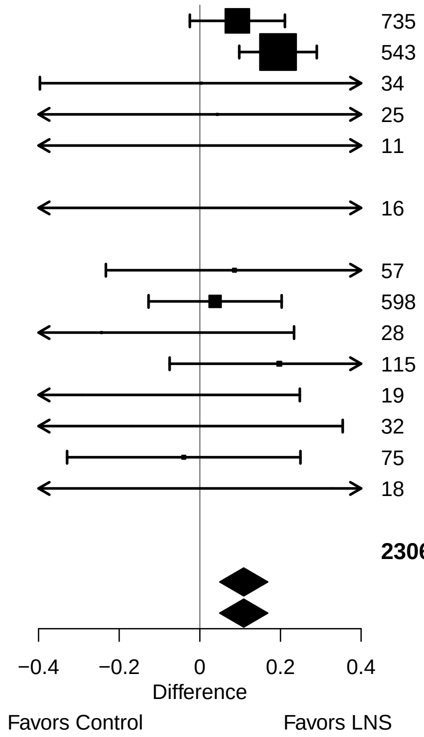

#### Supplemental figure 6L: Mean difference in WAZ

### 6L2: Stratified by Maternal BMI

**P−for−interaction = 0.999**
**Difference in MDs = 0.00 (−0.05, 0.05)**

| | | At least 20 kg/m² | | | | | | Less than 20 kg/m² | | | | | |
|---|---|---|---|---|---|---|---|---|---|---|---|---|---|
| **Country** | **Trial** | **LNS N** | **Control N** | **Control Mean** | **MD (95% CI)** | **Fixed W** | **Random W** | **LNS N** | **Control N** | **Control Mean** | **MD (95% CI)** | **Fixed W** | **Random W** |
| Bangladesh | JiVitA−4 (34) | | | | | | | | | | | | |
| Bangladesh | RDNS (35) | 730 | 335 | −1.53 | 0.08 (−0.04, 0.19) | 0.08 | 0.08 | 873 | 455 | −1.92 | 0.11 (0.01, 0.22) | 0.13 | 0.12 |
| Bangladesh | WASH−B (36) | 518 | 1595 | −1.34 | 0.22 (0.10, 0.34) | 0.07 | 0.08 | 645 | 1860 | −1.74 | 0.19 (0.12, 0.27) | 0.26 | 0.15 |
| Burkina Faso | iLiNS−Zinc (37) | 1218 | 391 | −1.39 | 0.28 (0.20, 0.37) | 0.13 | 0.10 | 740 | 275 | −1.74 | 0.26 (0.18, 0.34) | 0.20 | 0.14 |
| Burkina Faso | PROMIS (38) | 645 | 673 | −1.15 | 0.19 (0.06, 0.32) | 0.06 | 0.08 | 209 | 232 | −1.42 | −0.01 (−0.19, 0.17) | 0.04 | 0.07 |
| Burkina Faso | PROMIS CS (38) | 246 | 267 | −1.09 | −0.08 (−0.24, 0.07) | 0.04 | 0.07 | 184 | 165 | −1.54 | 0.05 (−0.12, 0.21) | 0.05 | 0.07 |
| Ghana | GHANA (39) | | | | | | | | | | | | |
| Ghana | iLiNS−DYADG (40) | 305 | 563 | −0.80 | 0.13 (−0.02, 0.27) | 0.05 | 0.07 | 37 | 119 | −1.06 | 0.24 (−0.12, 0.60) | 0.01 | 0.02 |
| Haiti | HAITI (41) | | | | | | | | | | | | |
| Kenya | WASH−B (42) | 1100 | 3822 | −0.66 | 0.08 (0.01, 0.16) | 0.18 | 0.10 | 273 | 1082 | −1.02 | 0.26 (0.12, 0.39) | 0.08 | 0.09 |
| Madagascar | MAHAY (43) | | | | | | | | | | | | |
| Malawi | iLiNS−DYADM (44) | 131 | 263 | −0.75 | −0.06 (−0.28, 0.16) | 0.02 | 0.04 | 88 | 178 | −1.05 | −0.08 (−0.33, 0.17) | 0.02 | 0.04 |
| Malawi | iLiNS−DOSE (45) | 519 | 174 | −0.96 | 0.05 (−0.07, 0.18) | 0.06 | 0.08 | 180 | 65 | −1.43 | 0.05 (−0.15, 0.25) | 0.03 | 0.06 |
| Mali | PROMIS (46) | 377 | 368 | −1.05 | 0.17 (0.04, 0.30) | 0.06 | 0.07 | 117 | 133 | −1.41 | 0.14 (−0.02, 0.30) | 0.06 | 0.08 |
| Mali | PROMIS CS (46) | 697 | 676 | −1.25 | 0.23 (0.09, 0.37) | 0.05 | 0.07 | 244 | 278 | −1.57 | 0.22 (0.05, 0.39) | 0.05 | 0.07 |
| Zimbabwe | SHINE (HIV−) (47) | 1355 | 1300 | −0.73 | 0.17 (0.09, 0.24) | 0.18 | 0.10 | 239 | 249 | −1.02 | 0.12 (−0.05, 0.30) | 0.04 | 0.07 |
| Zimbabwe | SHINE (HIV+) (48) | 246 | 253 | −0.87 | 0.08 (−0.11, 0.27) | 0.03 | 0.05 | 62 | 52 | −1.34 | −0.05 (−0.40, 0.31) | 0.01 | 0.02 |
| | | **8087** | **10680** | | **I² = 0.62, Tau² = 0.01** | | | **3891** | **5143** | | **I² = 0.44, Tau² = 0.00** | | |
| **Fixed** | | | | | **0.14 (0.11, 0.17)** | | | | | | **0.17 (0.13, 0.20)** | | |
| **Random** | | | | | **0.13 (0.08, 0.19)** | | | | | | **0.14 (0.09, 0.20)** | | |

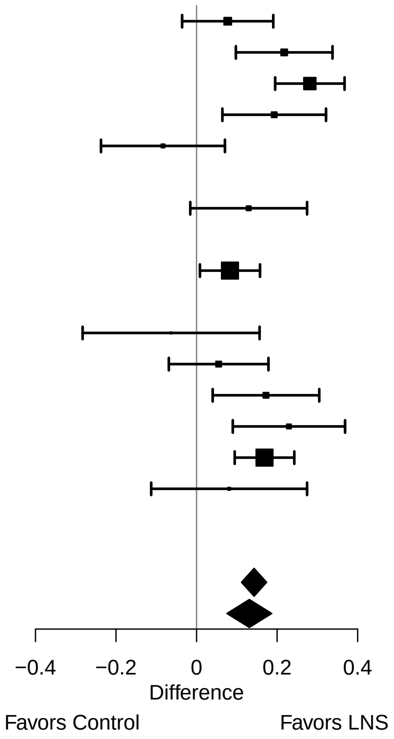

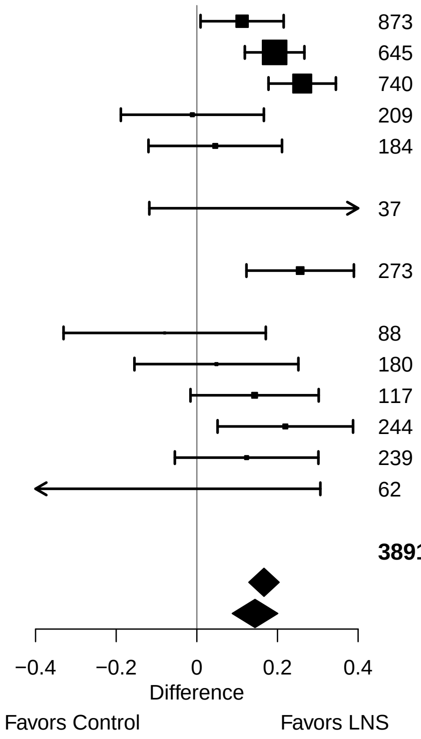

#### Supplemental figure 6L: Mean difference in WAZ

##### 6L3: Stratified by Maternal age

**P−for−interaction = 0.880**
**Difference in MDs = 0.00 (−0.04, 0.04)**

| | | At least 25 y | | | | | | Less than 25 y | | | | | |
|---|---|---|---|---|---|---|---|---|---|---|---|---|---|
| **Country** | **Trial** | **LNS N** | **Control N** | **Control Mean** | **MD (95% CI)** | **Fixed W** | **Random W** | **LNS N** | **Control N** | **Control Mean** | **MD (95% CI)** | **Fixed W** | **Random W** |
| Bangladesh | JiVitA−4 (34) | 1204 | 540 | −1.79 | 0.10 (0.05, 0.16) | 0.25 | 0.12 | 1597 | 687 | −1.75 | 0.11 (0.06, 0.15) | 0.33 | 0.10 |
| Bangladesh | RDNS (35) | 453 | 221 | −1.74 | 0.11 (−0.04, 0.27) | 0.03 | 0.05 | 1210 | 594 | −1.75 | 0.09 (0.01, 0.17) | 0.11 | 0.09 |
| Bangladesh | WASH−B (36) | 493 | 1521 | −1.60 | 0.13 (0.03, 0.23) | 0.08 | 0.08 | 665 | 1924 | −1.52 | 0.24 (0.15, 0.34) | 0.09 | 0.08 |
| Burkina Faso | iLiNS−Zinc (37) | 1134 | 401 | −1.51 | 0.28 (0.19, 0.37) | 0.10 | 0.09 | 813 | 262 | −1.55 | 0.26 (0.14, 0.38) | 0.05 | 0.07 |
| Burkina Faso | PROMIS (38) | 480 | 524 | −1.24 | 0.18 (0.06, 0.30) | 0.05 | 0.07 | 377 | 387 | −1.20 | 0.11 (−0.08, 0.29) | 0.02 | 0.05 |
| Burkina Faso | PROMIS CS (38) | 202 | 190 | −1.28 | −0.02 (−0.21, 0.17) | 0.02 | 0.04 | 228 | 246 | −1.26 | −0.07 (−0.24, 0.11) | 0.02 | 0.05 |
| Ghana | GHANA (39) | 69 | 63 | −0.68 | 0.40 (0.00, 0.81) | 0.00 | 0.01 | 29 | 25 | −0.59 | −0.09 (−0.70, 0.52) | 0.00 | 0.01 |
| Ghana | iLiNS−DYADG (40) | 220 | 417 | −0.79 | 0.06 (−0.11, 0.24) | 0.03 | 0.04 | 127 | 276 | −0.94 | 0.31 (0.11, 0.52) | 0.02 | 0.04 |
| Haiti | HAITI (41) | 100 | 104 | −0.37 | 0.08 (−0.07, 0.22) | 0.04 | 0.05 | 47 | 45 | −0.41 | 0.04 (−0.20, 0.28) | 0.01 | 0.04 |
| Kenya | WASH−B (42) | 845 | 2827 | −0.71 | 0.14 (0.06, 0.22) | 0.12 | 0.10 | 601 | 2306 | −0.80 | 0.13 (0.04, 0.21) | 0.11 | 0.09 |
| Madagascar | MAHAY (43) | 895 | 952 | −1.50 | 0.04 (−0.09, 0.18) | 0.04 | 0.06 | 806 | 733 | −1.37 | −0.03 (−0.17, 0.10) | 0.04 | 0.07 |
| Malawi | iLiNS−DYADM (44) | 106 | 226 | −0.87 | −0.04 (−0.28, 0.20) | 0.01 | 0.02 | 114 | 218 | −0.89 | −0.09 (−0.33, 0.14) | 0.01 | 0.04 |
| Malawi | iLiNS−DOSE (45) | 372 | 134 | −1.10 | 0.01 (−0.13, 0.14) | 0.04 | 0.06 | 316 | 100 | −1.07 | 0.09 (−0.08, 0.25) | 0.03 | 0.06 |
| Mali | PROMIS (46) | 342 | 355 | −1.11 | 0.15 (0.01, 0.29) | 0.04 | 0.06 | 158 | 147 | −1.23 | 0.21 (0.03, 0.39) | 0.02 | 0.05 |
| Mali | PROMIS CS (46) | 498 | 502 | −1.30 | 0.20 (0.05, 0.35) | 0.03 | 0.05 | 448 | 458 | −1.38 | 0.26 (0.11, 0.42) | 0.03 | 0.06 |
| Zimbabwe | SHINE (HIV−) (47) | 870 | 797 | −0.75 | 0.19 (0.09, 0.28) | 0.08 | 0.08 | 802 | 793 | −0.76 | 0.09 (0.00, 0.19) | 0.08 | 0.08 |
| Zimbabwe | SHINE (HIV+) (48) | 243 | 231 | −0.97 | 0.04 (−0.17, 0.25) | 0.02 | 0.03 | 73 | 77 | −0.82 | −0.11 (−0.46, 0.25) | 0.01 | 0.02 |
| | | **8526** | **10005** | | **I² = 0.41, Tau² = 0.00** | | | **8411** | **9278** | | **I² = 0.55, Tau² = 0.01** | | |
| **Fixed** | | | | | **0.13 (0.10, 0.16)** | | | | | | **0.12 (0.09, 0.15)** | | |
| **Random** | | | | | **0.12 (0.08, 0.17)** | | | | | | **0.12 (0.06, 0.17)** | | |

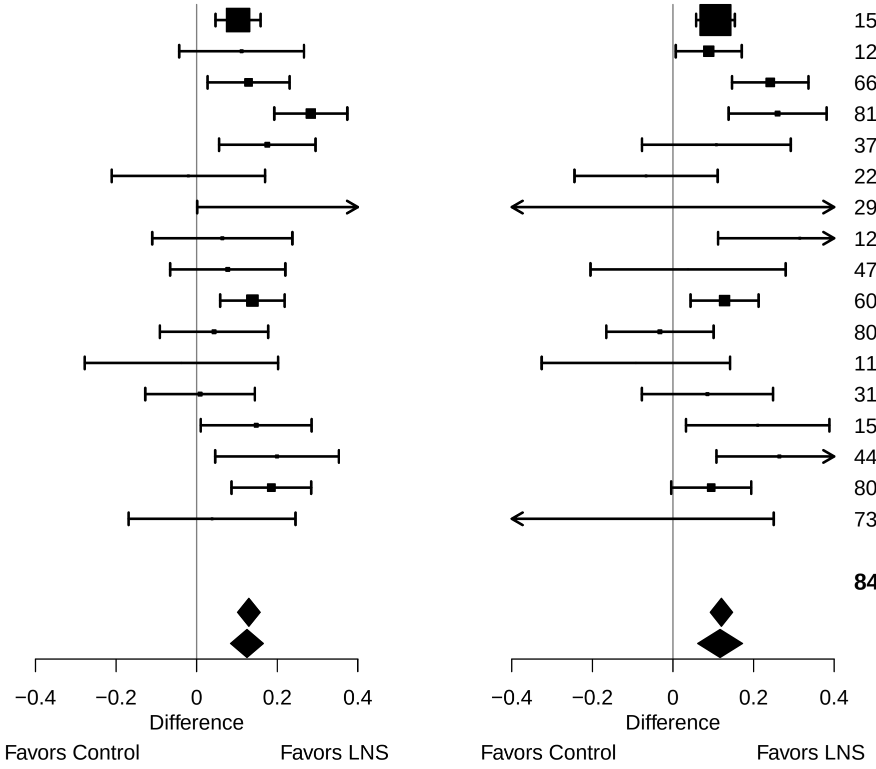

#### Supplemental figure 6L: Mean difference in WAZ

#### 6L4: Stratified by Maternal education

**P−for−interaction = 0.600**
**Difference in MDs = 0.01 (−0.03, 0.06)**

| | | Primary or greater | | | | | | Incomplete or no formal | | | | | |
|---|---|---|---|---|---|---|---|---|---|---|---|---|---|
| **Country** | **Trial** | **LNS N** | **Control N** | **Control Mean** | **MD (95% CI)** | **Fixed W** | **Random W** | **LNS N** | **Control N** | **Control Mean** | **MD (95% CI)** | **Fixed W** | **Random W** |
| Bangladesh | JiVitA−4 (34) | 1816 | 749 | −1.67 | 0.10 (0.06, 0.15) | 0.36 | 0.22 | 996 | 480 | −1.93 | 0.10 (0.04, 0.16) | 0.24 | 0.11 |
| Bangladesh | RDNS (35) | 1245 | 589 | −1.72 | 0.12 (0.03, 0.22) | 0.08 | 0.10 | 418 | 226 | −1.81 | 0.00 (−0.16, 0.15) | 0.03 | 0.06 |
| Bangladesh | WASH−B (36) | 818 | 2476 | −1.45 | 0.21 (0.14, 0.29) | 0.12 | 0.13 | 345 | 985 | −1.83 | 0.18 (0.06, 0.29) | 0.06 | 0.08 |
| Burkina Faso | iLiNS−Zinc (37) | 82 | 16 | −1.16 | 0.17 (−0.20, 0.54) | 0.01 | 0.01 | 1865 | 647 | −1.54 | 0.28 (0.21, 0.35) | 0.18 | 0.10 |
| Burkina Faso | PROMIS (38) | 70 | 54 | −1.23 | 0.03 (−0.22, 0.28) | 0.01 | 0.02 | 787 | 857 | −1.22 | 0.16 (0.03, 0.28) | 0.05 | 0.07 |
| Burkina Faso | PROMIS CS (38) | 34 | 39 | −1.24 | −0.10 (−0.63, 0.42) | 0.00 | 0.00 | 393 | 396 | −1.27 | −0.04 (−0.18, 0.09) | 0.04 | 0.07 |
| Ghana | GHANA (39) | 91 | 75 | −0.69 | 0.31 (−0.05, 0.68) | 0.01 | 0.01 | 7 | 15 | −0.47 | −0.23 (−1.22, 0.76) | 0.00 | 0.00 |
| Ghana | iLiNS−DYADG (40) | 267 | 549 | −0.83 | 0.13 (−0.03, 0.28) | 0.03 | 0.05 | 80 | 144 | −0.94 | 0.28 (0.01, 0.55) | 0.01 | 0.03 |
| Haiti | HAITI (41) | 129 | 126 | −0.29 | 0.08 (−0.05, 0.21) | 0.04 | 0.06 | 19 | 24 | −0.82 | −0.08 (−0.44, 0.27) | 0.01 | 0.02 |
| Kenya | WASH−B (42) | 705 | 2451 | −0.63 | 0.07 (−0.01, 0.16) | 0.10 | 0.11 | 754 | 2710 | −0.86 | 0.19 (0.11, 0.26) | 0.13 | 0.10 |
| Madagascar | MAHAY (43) | 352 | 431 | −1.25 | 0.01 (−0.16, 0.18) | 0.03 | 0.04 | 1350 | 1254 | −1.51 | 0.03 (−0.09, 0.15) | 0.05 | 0.07 |
| Malawi | iLiNS−DYADM (44) | 35 | 69 | −0.66 | −0.20 (−0.60, 0.21) | 0.00 | 0.01 | 184 | 372 | −0.92 | −0.05 (−0.23, 0.14) | 0.02 | 0.05 |
| Malawi | iLiNS−DOSE (45) | 165 | 56 | −0.81 | 0.09 (−0.12, 0.29) | 0.02 | 0.03 | 525 | 178 | −1.17 | 0.04 (−0.08, 0.16) | 0.06 | 0.07 |
| Mali | PROMIS (46) | 39 | 37 | −0.91 | −0.03 (−0.34, 0.28) | 0.01 | 0.01 | 460 | 465 | −1.16 | 0.18 (0.05, 0.31) | 0.05 | 0.07 |
| Mali | PROMIS CS (46) | 107 | 96 | −1.21 | 0.38 (0.10, 0.66) | 0.01 | 0.02 | 838 | 864 | −1.35 | 0.21 (0.08, 0.33) | 0.05 | 0.07 |
| Zimbabwe | SHINE (HIV−) (47) | 1707 | 1636 | −0.74 | 0.12 (0.05, 0.19) | 0.14 | 0.14 | 68 | 61 | −0.80 | 0.16 (−0.26, 0.59) | 0.00 | 0.01 |
| Zimbabwe | SHINE (HIV+) (48) | 300 | 292 | −0.91 | −0.01 (−0.19, 0.17) | 0.02 | 0.04 | 21 | 18 | −1.20 | 0.05 (−0.61, 0.70) | 0.00 | 0.01 |
| | | **7962** | **9741** | | **I² = 0.18, Tau² = 0.00** | | | **9110** | **9696** | | **I² = 0.64, Tau² = 0.01** | | |
| **Fixed** | | | | | **0.11 (0.09, 0.14)** | | | | | | **0.14 (0.11, 0.17)** | | |
| **Random** | | | | | **0.11 (0.08, 0.15)** | | | | | | **0.12 (0.07, 0.17)** | | |

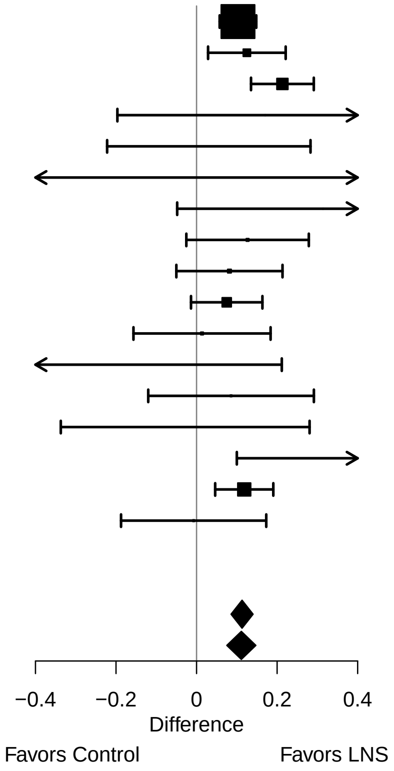

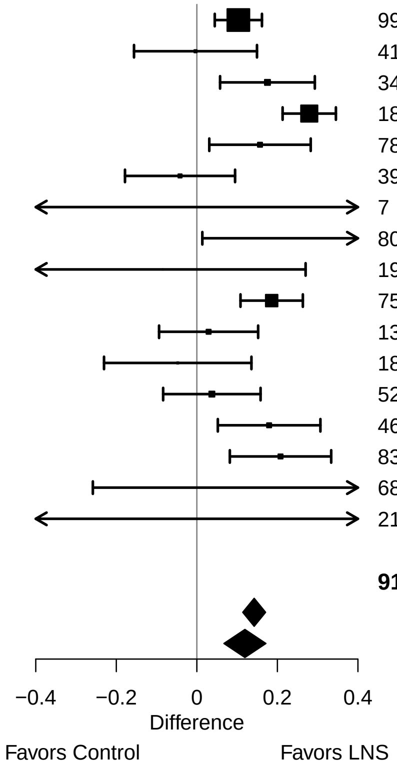

#### Supplemental figure 6L: Mean difference in WAZ

#### 6L5: Stratified by Maternal depressive symptoms

**P−for−interaction = 0.345**
**Difference in MDs = 0.03 (−0.03, 0.09)**

| | | Less than 75th percentile | | | | | | At least 75th percentile | | | | | |
|---|---|---|---|---|---|---|---|---|---|---|---|---|---|
| **Country** | **Trial** | **LNS N** | **Control N** | **Control Mean** | **MD (95% CI)** | **Fixed W** | **Random W** | **LNS N** | **Control N** | **Control Mean** | **MD (95% CI)** | **Fixed W** | **Random W** |
| Bangladesh | JiVitA−4 (34) | | | | | | | | | | | | |
| Bangladesh | RDNS (35) | 1081 | 472 | −1.73 | 0.10 (0.00, 0.20) | 0.13 | 0.13 | 520 | 288 | −1.77 | 0.07 (−0.07, 0.21) | 0.13 | 0.13 |
| Bangladesh | WASH−B (36) | 899 | 2487 | −1.53 | 0.21 (0.13, 0.29) | 0.18 | 0.15 | 240 | 883 | −1.61 | 0.16 (0.03, 0.29) | 0.16 | 0.14 |
| Burkina Faso | iLiNS−Zinc (37) | | | | | | | | | | | | |
| Burkina Faso | PROMIS (38) | 505 | 571 | −1.27 | 0.22 (0.08, 0.37) | 0.06 | 0.07 | 220 | 213 | −1.20 | 0.01 (−0.15, 0.16) | 0.11 | 0.12 |
| Burkina Faso | PROMIS CS (38) | | | | | | | | | | | | |
| Ghana | GHANA (39) | | | | | | | | | | | | |
| Ghana | iLiNS−DYADG (40) | 252 | 460 | −0.83 | 0.17 (0.01, 0.32) | 0.05 | 0.07 | 83 | 209 | −0.89 | 0.17 (−0.10, 0.44) | 0.04 | 0.06 |
| Haiti | HAITI (41) | | | | | | | | | | | | |
| Kenya | WASH−B (42) | 997 | 3506 | −0.72 | 0.13 (0.06, 0.20) | 0.23 | 0.17 | 342 | 1248 | −0.79 | 0.09 (−0.04, 0.22) | 0.16 | 0.14 |
| Madagascar | MAHAY (43) | 673 | 721 | −1.47 | 0.08 (−0.05, 0.21) | 0.07 | 0.09 | 283 | 252 | −1.42 | −0.15 (−0.34, 0.04) | 0.07 | 0.09 |
| Malawi | iLiNS−DYADM (44) | 157 | 303 | −0.80 | −0.09 (−0.29, 0.11) | 0.03 | 0.05 | 47 | 113 | −1.05 | 0.05 (−0.30, 0.40) | 0.02 | 0.04 |
| Malawi | iLiNS−DOSE (45) | | | | | | | | | | | | |
| Mali | PROMIS (46) | 374 | 372 | −1.10 | 0.14 (0.01, 0.27) | 0.07 | 0.09 | 122 | 129 | −1.28 | 0.25 (0.09, 0.42) | 0.09 | 0.11 |
| Mali | PROMIS CS (46) | | | | | | | | | | | | |
| Zimbabwe | SHINE (HIV−) (47) | 1192 | 1137 | −0.73 | 0.13 (0.04, 0.21) | 0.16 | 0.14 | 509 | 490 | −0.85 | 0.22 (0.10, 0.34) | 0.18 | 0.15 |
| Zimbabwe | SHINE (HIV+) (48) | 234 | 214 | −0.90 | −0.02 (−0.24, 0.20) | 0.03 | 0.04 | 88 | 100 | −1.08 | 0.05 (−0.26, 0.36) | 0.03 | 0.05 |
| | | **6364** | **10243** | | **I² = 0.29, Tau² = 0.00** | | | **2454** | **3925** | | **I² = 0.46, Tau² = 0.01** | | |
| **Fixed** | | | | | **0.13 (0.10, 0.17)** | | | | | | **0.11 (0.06, 0.16)** | | |
| **Random** | | | | | **0.13 (0.08, 0.17)** | | | | | | **0.10 (0.03, 0.18)** | | |

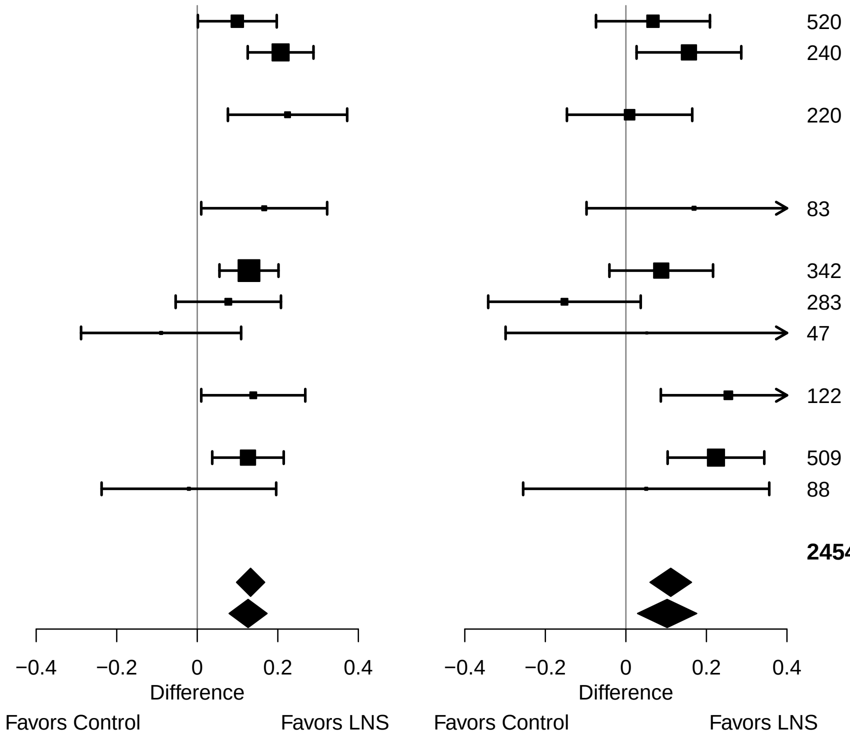

#### Supplemental figure 6L: Mean difference in WAZ

##### 6L6: Stratified by Child sex

**P−for−interaction = 0.292**
**Difference in MDs = 0.02 (−0.02, 0.06)**

| Country | Trial | Male LNS N | Male Control N | Male Control Mean | Male MD (95% CI) | Male Fixed W | Male Random W | Female LNS N | Female Control N | Female Control Mean | Female MD (95% CI) | Female Fixed W | Female Random W |
|---|---|---|---|---|---|---|---|---|---|---|---|---|---|
| Bangladesh | JiVitA−4 (34) | 1440 | 594 | −1.74 | 0.09 (0.04, 0.15) | 0.27 | 0.10 | 1374 | 638 | −1.80 | 0.12 (0.07, 0.17) | 0.31 | 0.13 |
| Bangladesh | RDNS (35) | 833 | 411 | −1.70 | 0.01 (−0.12, 0.13) | 0.05 | 0.07 | 830 | 404 | −1.79 | 0.18 (0.07, 0.30) | 0.06 | 0.07 |
| Bangladesh | WASH−B (36) | 567 | 1730 | −1.55 | 0.20 (0.11, 0.29) | 0.10 | 0.08 | 596 | 1731 | −1.56 | 0.19 (0.10, 0.29) | 0.08 | 0.09 |
| Burkina Faso | iLiNS−Zinc (37) | 995 | 336 | −1.65 | 0.29 (0.18, 0.39) | 0.08 | 0.08 | 965 | 330 | −1.42 | 0.27 (0.18, 0.36) | 0.08 | 0.09 |
| Burkina Faso | PROMIS (38) | 435 | 482 | −1.35 | 0.19 (0.04, 0.33) | 0.04 | 0.06 | 422 | 429 | −1.08 | 0.09 (−0.07, 0.26) | 0.03 | 0.05 |
| Burkina Faso | PROMIS CS (38) | 234 | 221 | −1.35 | −0.09 (−0.28, 0.11) | 0.02 | 0.04 | 196 | 215 | −1.18 | 0.01 (−0.15, 0.18) | 0.03 | 0.05 |
| Ghana | GHANA (39) | 60 | 41 | −0.72 | 0.22 (−0.21, 0.65) | 0.00 | 0.01 | 38 | 55 | −0.64 | 0.42 (−0.11, 0.94) | 0.00 | 0.01 |
| Ghana | iLiNS−DYADG (40) | 175 | 324 | −0.94 | 0.15 (−0.05, 0.35) | 0.02 | 0.04 | 172 | 369 | −0.78 | 0.18 (0.01, 0.36) | 0.02 | 0.04 |
| Haiti | HAITI (41) | 70 | 60 | −0.39 | 0.02 (−0.17, 0.21) | 0.02 | 0.04 | 78 | 90 | −0.37 | 0.12 (−0.05, 0.28) | 0.03 | 0.04 |
| Kenya | WASH−B (42) | 734 | 2457 | −0.83 | 0.11 (0.02, 0.20) | 0.10 | 0.08 | 727 | 2708 | −0.68 | 0.16 (0.08, 0.24) | 0.12 | 0.10 |
| Madagascar | MAHAY (43) | 826 | 835 | −1.59 | 0.01 (−0.12, 0.14) | 0.05 | 0.07 | 876 | 850 | −1.30 | 0.01 (−0.12, 0.14) | 0.04 | 0.06 |
| Malawi | iLiNS−DYADM (44) | 107 | 207 | −0.93 | −0.09 (−0.34, 0.15) | 0.01 | 0.03 | 113 | 237 | −0.84 | −0.04 (−0.26, 0.19) | 0.01 | 0.03 |
| Malawi | iLiNS−DOSE (45) | 353 | 124 | −1.29 | −0.02 (−0.17, 0.13) | 0.04 | 0.06 | 349 | 117 | −0.88 | 0.11 (−0.03, 0.26) | 0.03 | 0.05 |
| Mali | PROMIS (46) | 265 | 262 | −1.19 | 0.13 (0.00, 0.25) | 0.05 | 0.07 | 235 | 240 | −1.09 | 0.21 (0.04, 0.37) | 0.03 | 0.05 |
| Mali | PROMIS CS (46) | 474 | 521 | −1.40 | 0.20 (0.04, 0.37) | 0.03 | 0.05 | 472 | 439 | −1.27 | 0.25 (0.09, 0.40) | 0.03 | 0.05 |
| Zimbabwe | SHINE (HIV−) (47) | 924 | 906 | −0.88 | 0.19 (0.10, 0.27) | 0.10 | 0.08 | 945 | 880 | −0.62 | 0.08 (−0.02, 0.17) | 0.08 | 0.09 |
| Zimbabwe | SHINE (HIV+) (48) | 161 | 167 | −1.09 | 0.02 (−0.21, 0.24) | 0.02 | 0.03 | 175 | 161 | −0.80 | −0.01 (−0.23, 0.22) | 0.01 | 0.03 |
| | | **8653** | **9678** | | **$I^2$ = 0.56, $Tau^2$ = 0.01** | | | **8563** | **9893** | | **$I^2$ = 0.40, $Tau^2$ = 0.00** | | |
| **Fixed** | | | | | **0.12 (0.09, 0.15)** | | | | | | **0.14 (0.11, 0.16)** | | |
| **Random** | | | | | **0.11 (0.06, 0.16)** | | | | | | **0.14 (0.10, 0.18)** | | |

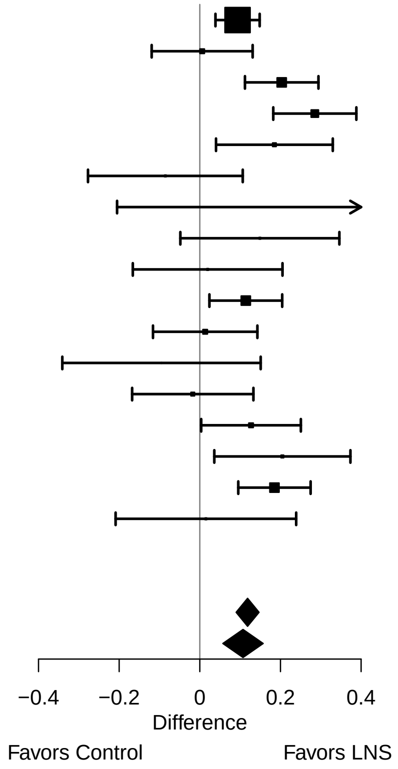

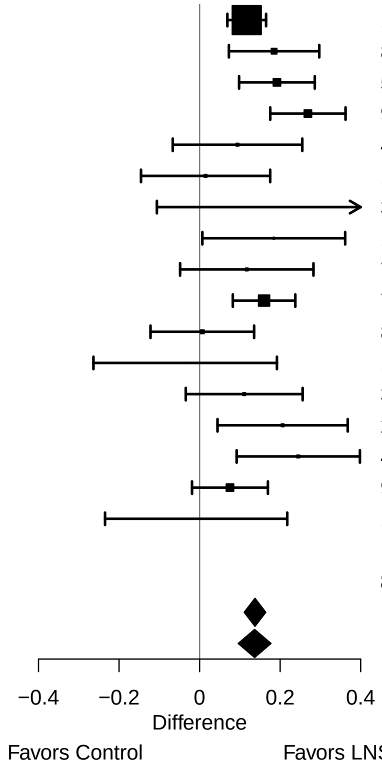

#### Supplemental figure 6L: Mean difference in WAZ

#### 6L7: Stratified by Child birth order

**P−for−interaction = 0.021**
**Difference in MDs = −0.05 (−0.09, −0.01)**

**Later born**

| Country | Trial | LNS N | Control N | Control Mean | MD (95% CI) | Fixed W | Random W |
|---|---|---|---|---|---|---|---|
| Bangladesh | JiVitA−4 (34) | 600 | 250 | −1.89 | 0.16 (0.09, 0.24) | 0.12 | 0.08 |
| Bangladesh | RDNS (35) | 974 | 508 | −1.77 | 0.11 (0.01, 0.22) | 0.06 | 0.07 |
| Bangladesh | WASH−B (36) | 737 | 2310 | −1.61 | 0.19 (0.11, 0.26) | 0.11 | 0.08 |
| Burkina Faso | iLiNS−Zinc (37) | 1524 | 521 | −1.52 | 0.28 (0.21, 0.35) | 0.14 | 0.08 |
| Burkina Faso | PROMIS (38) | 709 | 750 | −1.22 | 0.17 (0.05, 0.30) | 0.05 | 0.06 |
| Burkina Faso | PROMIS CS (38) | 366 | 357 | −1.25 | −0.07 (−0.21, 0.07) | 0.03 | 0.05 |
| Ghana | GHANA (39) | 60 | 54 | −0.67 | 0.39 (−0.07, 0.85) | 0.00 | 0.01 |
| Ghana | iLiNS−DYADG (40) | 237 | 459 | −0.83 | 0.07 (−0.09, 0.23) | 0.03 | 0.05 |
| Haiti | HAITI (41) | 88 | 100 | −0.47 | 0.14 (−0.02, 0.30) | 0.03 | 0.05 |
| Kenya | WASH−B (42) | 1137 | 4065 | −0.75 | 0.12 (0.05, 0.20) | 0.11 | 0.08 |
| Madagascar | MAHAY (43) | 1271 | 1220 | −1.49 | 0.06 (−0.06, 0.18) | 0.05 | 0.06 |
| Malawi | iLiNS−DYADM (44) | 173 | 353 | −0.87 | −0.07 (−0.26, 0.12) | 0.02 | 0.04 |
| Malawi | iLiNS−DOSE (45) | 463 | 165 | −1.13 | 0.06 (−0.06, 0.18) | 0.04 | 0.06 |
| Mali | PROMIS (46) | 437 | 441 | −1.14 | 0.19 (0.07, 0.32) | 0.04 | 0.06 |
| Mali | PROMIS CS (46) | 784 | 822 | −1.31 | 0.22 (0.10, 0.35) | 0.04 | 0.06 |
| Zimbabwe | SHINE (HIV−) (47) | 1295 | 1220 | −0.77 | 0.21 (0.13, 0.28) | 0.11 | 0.08 |
| Zimbabwe | SHINE (HIV+) (48) | 279 | 245 | −0.84 | −0.11 (−0.31, 0.09) | 0.02 | 0.04 |
| | | **11134** | **13840** | | **I² = 0.66, Tau² = 0.01** | | |
| **Fixed** | | | | | **0.16 (0.13, 0.18)** | | |
| **Random** | | | | | **0.13 (0.08, 0.18)** | | |

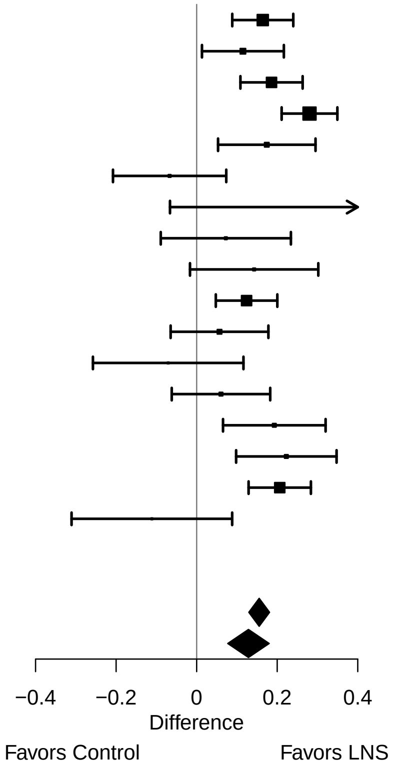

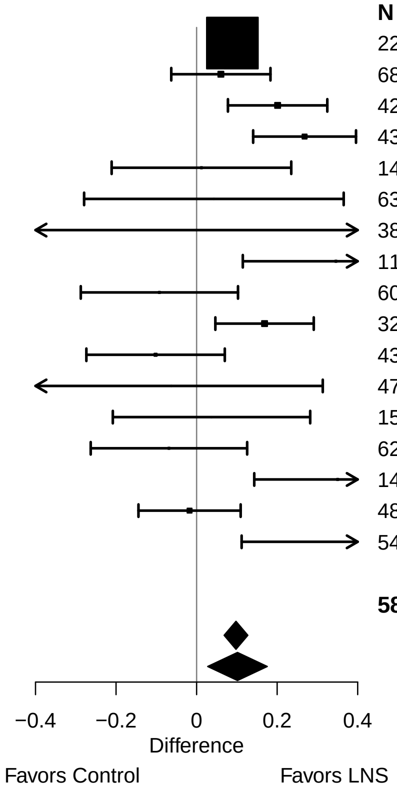

**Firstborn**

| Country | Trial | LNS N | Control N | Control Mean | MD (95% CI) | Fixed W | Random W |
|---|---|---|---|---|---|---|---|
| Bangladesh | JiVitA−4 (34) | 2210 | 976 | −1.74 | 0.09 (0.05, 0.13) | 0.55 | 0.10 |
| Bangladesh | RDNS (35) | 688 | 307 | −1.71 | 0.06 (−0.06, 0.18) | 0.06 | 0.08 |
| Bangladesh | WASH−B (36) | 426 | 1151 | −1.44 | 0.20 (0.08, 0.32) | 0.06 | 0.08 |
| Burkina Faso | iLiNS−Zinc (37) | 436 | 145 | −1.60 | 0.27 (0.14, 0.40) | 0.05 | 0.08 |
| Burkina Faso | PROMIS (38) | 148 | 159 | −1.22 | 0.01 (−0.21, 0.24) | 0.02 | 0.05 |
| Burkina Faso | PROMIS CS (38) | 63 | 79 | −1.36 | 0.04 (−0.28, 0.37) | 0.01 | 0.03 |
| Ghana | GHANA (39) | 38 | 38 | −0.62 | 0.04 (−0.44, 0.51) | 0.00 | 0.02 |
| Ghana | iLiNS−DYADG (40) | 110 | 234 | −0.89 | 0.35 (0.11, 0.58) | 0.02 | 0.05 |
| Haiti | HAITI (41) | 60 | 50 | −0.19 | −0.09 (−0.29, 0.10) | 0.02 | 0.06 |
| Kenya | WASH−B (42) | 322 | 1095 | −0.73 | 0.17 (0.05, 0.29) | 0.06 | 0.08 |
| Madagascar | MAHAY (43) | 431 | 465 | −1.33 | −0.10 (−0.27, 0.07) | 0.03 | 0.07 |
| Malawi | iLiNS−DYADM (44) | 47 | 89 | −0.89 | −0.06 (−0.44, 0.31) | 0.01 | 0.03 |
| Malawi | iLiNS−DOSE (45) | 150 | 45 | −1.01 | 0.04 (−0.21, 0.28) | 0.01 | 0.05 |
| Mali | PROMIS (46) | 62 | 59 | −1.15 | −0.07 (−0.26, 0.13) | 0.02 | 0.06 |
| Mali | PROMIS CS (46) | 145 | 126 | −1.46 | 0.35 (0.14, 0.56) | 0.02 | 0.06 |
| Zimbabwe | SHINE (HIV−) (47) | 487 | 483 | −0.71 | −0.02 (−0.14, 0.11) | 0.05 | 0.08 |
| Zimbabwe | SHINE (HIV+) (48) | 54 | 78 | −1.25 | 0.40 (0.11, 0.69) | 0.01 | 0.04 |
| | | **5877** | **5579** | | **I² = 0.62, Tau² = 0.01** | | |
| **Fixed** | | | | | **0.10 (0.07, 0.13)** | | |
| **Random** | | | | | **0.10 (0.03, 0.18)** | | |

#### Supplemental figure 6L: Mean difference in WAZ

#### 6L8: Stratified by Child baseline anthropometric status

**P−for−interaction = 0.078**
**Difference in MDs = 0.05 (−0.01, 0.10)**

| | | WAZ at least −1 Z | | | | | | WAZ less than −1 Z | | | | | |
|---|---|---|---|---|---|---|---|---|---|---|---|---|---|
| **Country** | **Trial** | **LNS N** | **Control N** | **Control Mean** | **MD (95% CI)** | **Fixed W** | **Random W** | **LNS N** | **Control N** | **Control Mean** | **MD (95% CI)** | **Fixed W** | **Random W** |
| Bangladesh | JiVitA−4 (34) | 1198 | 531 | −1.05 | 0.14 (0.06, 0.21) | 0.24 | 0.18 | 1616 | 701 | −2.31 | 0.11 (0.05, 0.18) | 0.33 | 0.18 |
| Bangladesh | RDNS (35) | 780 | 338 | −1.05 | 0.01 (−0.08, 0.10) | 0.17 | 0.15 | 821 | 422 | −2.29 | 0.07 (−0.01, 0.14) | 0.22 | 0.16 |
| Bangladesh | WASH−B (36) | | | | | | | | | | | | |
| Burkina Faso | iLiNS−Zinc (37) | 718 | 235 | −0.57 | 0.14 (−0.08, 0.37) | 0.03 | 0.04 | 1242 | 431 | −2.06 | 0.37 (0.25, 0.48) | 0.11 | 0.12 |
| Burkina Faso | PROMIS (38) | 427 | 482 | −0.82 | 0.15 (0.03, 0.28) | 0.09 | 0.10 | 268 | 267 | −1.92 | 0.14 (0.01, 0.27) | 0.08 | 0.11 |
| Burkina Faso | PROMIS CS (38) | | | | | | | | | | | | |
| Ghana | GHANA (39) | | | | | | | | | | | | |
| Ghana | iLiNS−DYADG (40) | 222 | 424 | −0.40 | 0.13 (−0.01, 0.27) | 0.07 | 0.08 | 101 | 215 | −1.71 | 0.08 (−0.10, 0.25) | 0.04 | 0.07 |
| Haiti | HAITI (41) | 113 | 110 | 0.02 | 0.00 (−0.23, 0.23) | 0.03 | 0.04 | 35 | 40 | −1.48 | 0.20 (−0.09, 0.48) | 0.02 | 0.04 |
| Kenya | WASH−B (42) | | | | | | | | | | | | |
| Madagascar | MAHAY (43) | | | | | | | | | | | | |
| Malawi | iLiNS−DYADM (44) | 129 | 292 | −0.44 | −0.10 (−0.27, 0.07) | 0.05 | 0.06 | 75 | 133 | −1.86 | 0.22 (−0.03, 0.47) | 0.02 | 0.04 |
| Malawi | iLiNS−DOSE (45) | 419 | 143 | −0.50 | −0.03 (−0.19, 0.14) | 0.05 | 0.06 | 283 | 98 | −1.95 | 0.08 (−0.13, 0.28) | 0.03 | 0.06 |
| Mali | PROMIS (46) | 279 | 243 | −0.65 | 0.12 (−0.01, 0.26) | 0.07 | 0.09 | 221 | 259 | −1.61 | 0.21 (0.02, 0.39) | 0.04 | 0.07 |
| Mali | PROMIS CS (46) | | | | | | | | | | | | |
| Zimbabwe | SHINE (HIV−) (47) | 917 | 836 | −0.37 | 0.09 (0.01, 0.18) | 0.19 | 0.16 | 383 | 349 | −1.57 | 0.18 (0.05, 0.31) | 0.08 | 0.11 |
| Zimbabwe | SHINE (HIV+) (48) | 151 | 144 | −0.42 | 0.06 (−0.16, 0.28) | 0.03 | 0.04 | 113 | 92 | −1.68 | 0.07 (−0.17, 0.32) | 0.02 | 0.04 |
| | | **5353** | **3778** | | **I² = 0.27, Tau² = 0.00** | | | **5158** | **3007** | | **I² = 0.55, Tau² = 0.00** | | |
| **Fixed** | | | | | **0.08 (0.05, 0.12)** | | | | | | **0.14 (0.10, 0.18)** | | |
| **Random** | | | | | **0.08 (0.03, 0.12)** | | | | | | **0.15 (0.10, 0.21)** | | |

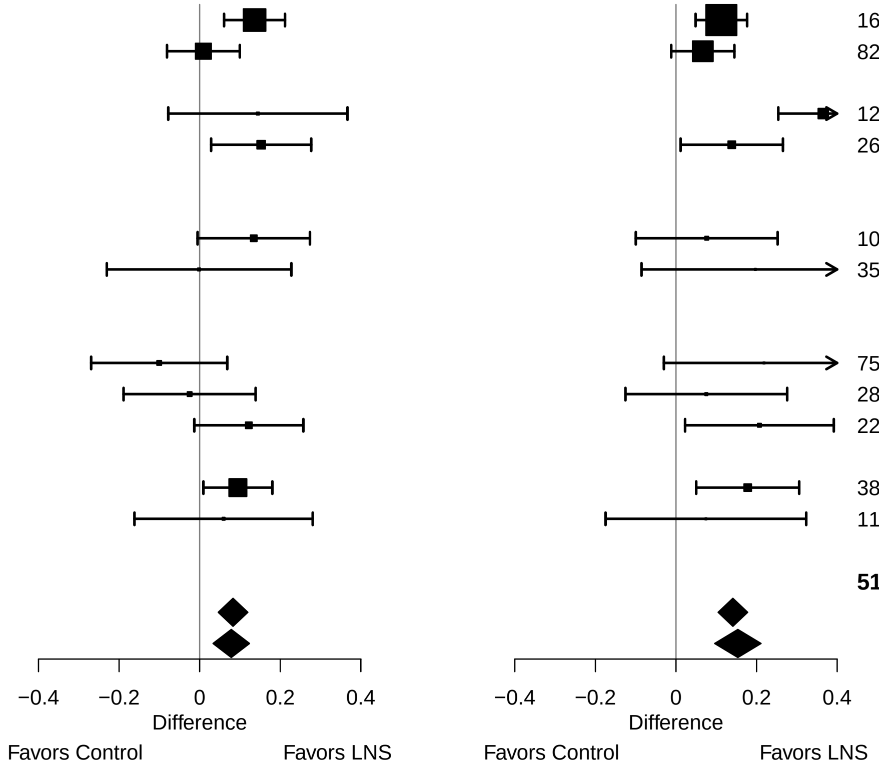

### Supplemental figure 6M: Underweight prevalence ratio

#### 6M1: Stratified by Maternal height

**P−for−interaction = 0.546**
**Ratio of PRs = 1.04 (0.92, 1.16)**

| | | At least 150.1 cm | | | | | | Less than 150.1 cm | | | | | |
|---|---|---|---|---|---|---|---|---|---|---|---|---|---|
| **Country** | **Trial** | **LNS N** | **Control N** | **Control Prevalence** | **PR (95% CI)** | **Fixed W** | **Random W** | **LNS N** | **Control N** | **Control Prevalence** | **PR (95% CI)** | **Fixed W** | **Random W** |
| Bangladesh | JiVitA−4 (34) | | | | | | | | | | | | |
| Bangladesh | RDNS (35) | 868 | 432 | 31.7 | 0.81 (0.67, 0.96) | 0.14 | 0.12 | 735 | 358 | 45.8 | 0.99 (0.87, 1.12) | 0.38 | 0.27 |
| Bangladesh | WASH−B (36) | 620 | 1898 | 25.7 | 0.80 (0.67, 0.96) | 0.14 | 0.12 | 543 | 1557 | 40.7 | 0.76 (0.67, 0.87) | 0.36 | 0.26 |
| Burkina Faso | iLiNS−Zinc (37) | 1924 | 651 | 30.9 | 0.68 (0.58, 0.81) | 0.17 | 0.13 | 34 | 15 | 33.3 | 1.25 (0.76, 2.07) | 0.02 | 0.06 |
| Burkina Faso | PROMIS (38) | 829 | 881 | 19.3 | 0.84 (0.67, 1.04) | 0.10 | 0.10 | 25 | 25 | 36.0 | 1.00 (0.49, 2.04) | 0.01 | 0.03 |
| Burkina Faso | PROMIS CS (38) | | | | | | | | | | | | |
| Ghana | GHANA (39) | | | | | | | | | | | | |
| Ghana | iLiNS−DYADG (40) | | | | | | | | | | | | |
| Haiti | HAITI (41) | | | | | | | | | | | | |
| Kenya | WASH−B (42) | 1316 | 4709 | 9.0 | 0.81 (0.65, 1.02) | 0.09 | 0.10 | 57 | 195 | 20.0 | 0.53 (0.25, 1.10) | 0.01 | 0.03 |
| Madagascar | MAHAY (43) | 1003 | 1040 | 22.2 | 1.07 (0.88, 1.30) | 0.11 | 0.11 | 598 | 565 | 33.8 | 0.93 (0.76, 1.15) | 0.14 | 0.18 |
| Malawi | iLiNS−DYADM (44) | 192 | 381 | 9.7 | 1.07 (0.64, 1.80) | 0.02 | 0.03 | 28 | 60 | 21.7 | 1.48 (0.72, 3.05) | 0.01 | 0.03 |
| Malawi | iLiNS−DOSE (45) | 586 | 198 | 16.7 | 1.09 (0.79, 1.50) | 0.04 | 0.07 | 115 | 42 | 33.3 | 0.82 (0.53, 1.27) | 0.03 | 0.07 |
| Mali | PROMIS (46) | | | | | | | | | | | | |
| Mali | PROMIS CS (46) | 909 | 928 | 22.3 | 0.86 (0.69, 1.06) | 0.10 | 0.10 | 32 | 26 | 34.6 | 1.17 (0.67, 2.07) | 0.02 | 0.05 |
| Zimbabwe | SHINE (HIV−) (47) | 1745 | 1690 | 10.4 | 0.69 (0.55, 0.86) | 0.09 | 0.10 | 75 | 62 | 19.4 | 0.62 (0.29, 1.30) | 0.01 | 0.03 |
| Zimbabwe | SHINE (HIV+) (48) | | | | | | | | | | | | |
| | | **9992** | **12808** | | **I² = 0.51, Tau² = 0.01** | | | **2242** | **2905** | | **I² = 0.41, Tau² = 0.01** | | |
| **Fixed** | | | | | **0.82 (0.77, 0.88)** | | | | | | **0.89 (0.82, 0.96)** | | |
| **Random** | | | | | **0.83 (0.76, 0.92)** | | | | | | **0.90 (0.79, 1.03)** | | |

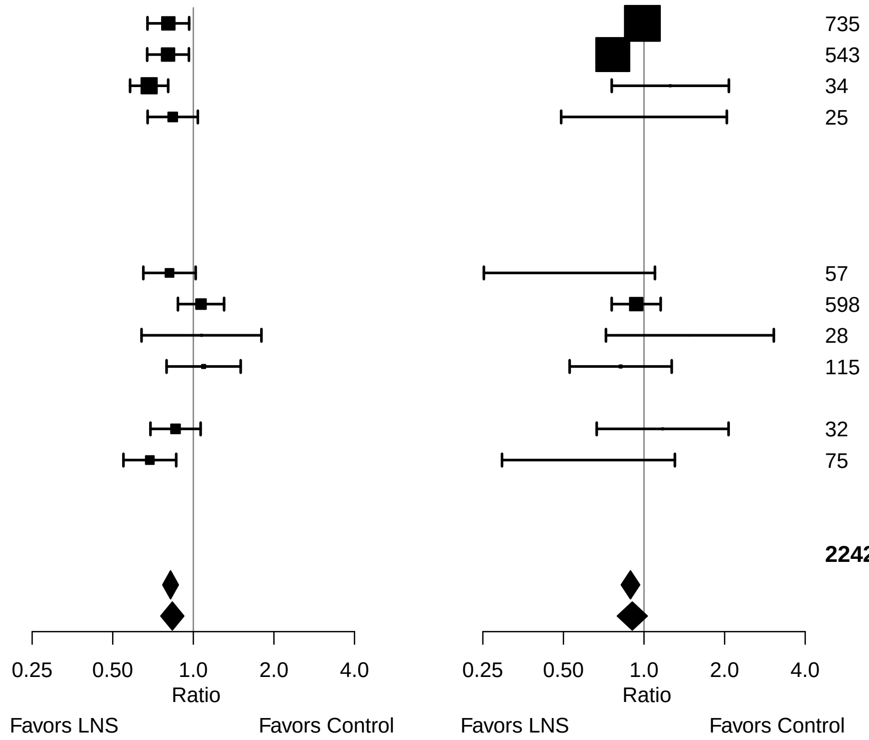

#### Supplemental figure 6M: Underweight prevalence ratio

##### 6M2: Stratified by Maternal BMI

**P−for−interaction = 0.545**
**Ratio of PRs = 1.03 (0.93, 1.15)**

| | | At least 20 kg/m² | | | | | | Less than 20 kg/m² | | | | | |
|---|---|---|---|---|---|---|---|---|---|---|---|---|---|
| **Country** | **Trial** | **LNS N** | **Control N** | **Control Prevalence** | **PR (95% CI)** | **Fixed W** | **Random W** | **LNS N** | **Control N** | **Control Prevalence** | **PR (95% CI)** | **Fixed W** | **Random W** |
| Bangladesh | JiVitA−4 (34) | | | | | | | | | | | | |
| Bangladesh | RDNS (35) | 730 | 335 | 29.3 | 0.92 (0.76, 1.10) | 0.19 | 0.19 | 873 | 455 | 44.6 | 0.92 (0.82, 1.04) | 0.31 | 0.21 |
| Bangladesh | WASH−B (36) | 518 | 1595 | 24.6 | 0.82 (0.66, 1.02) | 0.13 | 0.13 | 645 | 1860 | 39.2 | 0.76 (0.67, 0.87) | 0.26 | 0.20 |
| Burkina Faso | iLiNS−Zinc (37) | 1218 | 391 | 24.8 | 0.68 (0.53, 0.88) | 0.10 | 0.10 | 740 | 275 | 39.6 | 0.72 (0.60, 0.86) | 0.13 | 0.14 |
| Burkina Faso | PROMIS (38) | 645 | 673 | 17.8 | 0.73 (0.55, 0.97) | 0.08 | 0.08 | 209 | 232 | 25.0 | 1.13 (0.85, 1.51) | 0.05 | 0.08 |
| Burkina Faso | PROMIS CS (38) | 246 | 267 | 18.4 | 1.02 (0.72, 1.44) | 0.05 | 0.05 | 184 | 165 | 33.9 | 0.91 (0.70, 1.18) | 0.07 | 0.09 |
| Ghana | GHANA (39) | | | | | | | | | | | | |
| Ghana | iLiNS−DYADG (40) | | | | | | | | | | | | |
| Haiti | HAITI (41) | | | | | | | | | | | | |
| Kenya | WASH−B (42) | 1100 | 3822 | 8.4 | 0.81 (0.63, 1.06) | 0.09 | 0.09 | 273 | 1082 | 13.4 | 0.77 (0.54, 1.09) | 0.03 | 0.05 |
| Madagascar | MAHAY (43) | | | | | | | | | | | | |
| Malawi | iLiNS−DYADM (44) | 131 | 263 | 11.4 | 0.87 (0.47, 1.61) | 0.02 | 0.02 | 88 | 178 | 11.2 | 1.52 (0.82, 2.82) | 0.01 | 0.02 |
| Malawi | iLiNS−DOSE (45) | 519 | 174 | 15.5 | 0.99 (0.70, 1.40) | 0.05 | 0.05 | 180 | 65 | 30.8 | 0.99 (0.67, 1.47) | 0.03 | 0.05 |
| Mali | PROMIS (46) | 377 | 368 | 14.3 | 0.86 (0.57, 1.29) | 0.04 | 0.04 | 117 | 133 | 22.2 | 0.85 (0.50, 1.44) | 0.02 | 0.03 |
| Mali | PROMIS CS (46) | 697 | 676 | 20.4 | 0.91 (0.70, 1.17) | 0.10 | 0.10 | 244 | 278 | 28.1 | 0.85 (0.63, 1.15) | 0.05 | 0.07 |
| Zimbabwe | SHINE (HIV−) (47) | 1355 | 1300 | 10.2 | 0.64 (0.49, 0.83) | 0.10 | 0.10 | 239 | 249 | 14.9 | 0.84 (0.52, 1.37) | 0.02 | 0.03 |
| Zimbabwe | SHINE (HIV+) (48) | 246 | 253 | 15.4 | 0.90 (0.60, 1.34) | 0.04 | 0.04 | 62 | 52 | 26.9 | 1.08 (0.62, 1.87) | 0.01 | 0.03 |
| | | **7782** | **10117** | | **I² = 0.00, Tau² = 0.00** | | | **3854** | **5024** | | **I² = 0.33, Tau² = 0.01** | | |
| **Fixed** | | | | | **0.83 (0.76, 0.90)** | | | | | | **0.85 (0.80, 0.91)** | | |
| **Random** | | | | | **0.83 (0.76, 0.90)** | | | | | | **0.87 (0.79, 0.95)** | | |

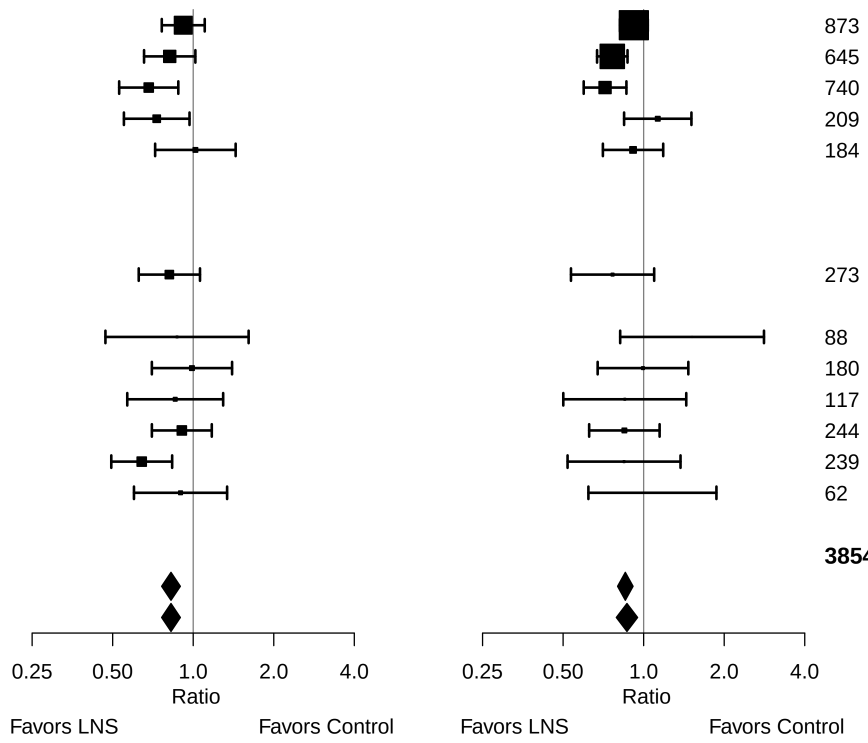

#### Supplemental figure 6M: Underweight prevalence ratio

#### 6M3: Stratified by Maternal age

**P−for−interaction = 0.245**
**Ratio of PRs = 1.05 (0.97, 1.15)**

| Country | Trial | At least 25 y LNS N | Control N | Control Prevalence | PR (95% CI) | Fixed W | Random W | Less than 25 y LNS N | Control N | Control Prevalence | PR (95% CI) | Fixed W | Random W |
|---|---|---|---|---|---|---|---|---|---|---|---|---|---|
| Bangladesh | JiVitA−4 (34) | 1204 | 540 | 40.6 | 0.88 (0.76, 1.01) | 0.20 | 0.13 | 1597 | 687 | 38.1 | 0.92 (0.82, 1.03) | 0.24 | 0.21 |
| Bangladesh | RDNS (35) | 453 | 221 | 36.7 | 0.92 (0.75, 1.14) | 0.10 | 0.09 | 1210 | 594 | 38.2 | 0.92 (0.83, 1.02) | 0.27 | 0.22 |
| Bangladesh | WASH−B (36) | 493 | 1521 | 34.8 | 0.83 (0.69, 0.99) | 0.13 | 0.11 | 665 | 1924 | 30.6 | 0.76 (0.64, 0.90) | 0.11 | 0.11 |
| Burkina Faso | iLiNS−Zinc (37) | 1134 | 401 | 31.7 | 0.61 (0.49, 0.77) | 0.09 | 0.09 | 813 | 262 | 29.8 | 0.85 (0.72, 1.01) | 0.10 | 0.11 |
| Burkina Faso | PROMIS (38) | 480 | 524 | 20.2 | 0.75 (0.59, 0.96) | 0.07 | 0.08 | 377 | 387 | 18.9 | 1.01 (0.73, 1.40) | 0.03 | 0.04 |
| Burkina Faso | PROMIS CS (38) | 202 | 190 | 24.2 | 0.94 (0.62, 1.43) | 0.02 | 0.04 | 228 | 246 | 24.4 | 1.02 (0.74, 1.42) | 0.03 | 0.04 |
| Ghana | GHANA (39) | | | | | | | | | | | | |
| Ghana | iLiNS−DYADG (40) | 220 | 417 | 12.0 | 0.99 (0.63, 1.54) | 0.02 | 0.03 | 127 | 276 | 14.5 | 0.60 (0.32, 1.13) | 0.01 | 0.01 |
| Haiti | HAITI (41) | | | | | | | | | | | | |
| Kenya | WASH−B (42) | 845 | 2827 | 9.3 | 0.79 (0.60, 1.04) | 0.06 | 0.07 | 601 | 2306 | 10.8 | 0.72 (0.53, 0.98) | 0.03 | 0.04 |
| Madagascar | MAHAY (43) | 895 | 952 | 27.4 | 1.01 (0.84, 1.22) | 0.12 | 0.10 | 806 | 733 | 24.1 | 1.08 (0.87, 1.34) | 0.06 | 0.07 |
| Malawi | iLiNS−DYADM (44) | 106 | 226 | 11.1 | 1.11 (0.59, 2.08) | 0.01 | 0.02 | 114 | 218 | 12.4 | 1.13 (0.64, 2.01) | 0.01 | 0.01 |
| Malawi | iLiNS−DOSE (45) | 372 | 134 | 20.1 | 1.13 (0.81, 1.57) | 0.04 | 0.05 | 316 | 100 | 18.0 | 0.97 (0.63, 1.51) | 0.02 | 0.02 |
| Mali | PROMIS (46) | 342 | 355 | 15.6 | 0.95 (0.63, 1.43) | 0.03 | 0.04 | 158 | 147 | 18.4 | 0.67 (0.37, 1.20) | 0.01 | 0.01 |
| Mali | PROMIS CS (46) | 498 | 502 | 21.5 | 0.91 (0.68, 1.21) | 0.05 | 0.06 | 448 | 458 | 23.6 | 0.86 (0.66, 1.12) | 0.04 | 0.05 |
| Zimbabwe | SHINE (HIV−) (47) | 870 | 797 | 10.9 | 0.59 (0.42, 0.83) | 0.04 | 0.05 | 802 | 793 | 10.3 | 0.80 (0.59, 1.07) | 0.03 | 0.04 |
| Zimbabwe | SHINE (HIV+) (48) | 243 | 231 | 17.7 | 0.95 (0.63, 1.44) | 0.02 | 0.04 | 73 | 77 | 14.3 | 1.25 (0.60, 2.60) | 0.01 | 0.01 |
| | | **8357** | **9838** | | **I² = 0.38, Tau² = 0.01** | | | **8335** | **9208** | | **I² = 0.11, Tau² = 0.00** | | |
| **Fixed** | | | | | **0.85 (0.80, 0.91)** | | | | | | **0.89 (0.85, 0.94)** | | |
| **Random** | | | | | **0.85 (0.78, 0.93)** | | | | | | **0.89 (0.84, 0.95)** | | |

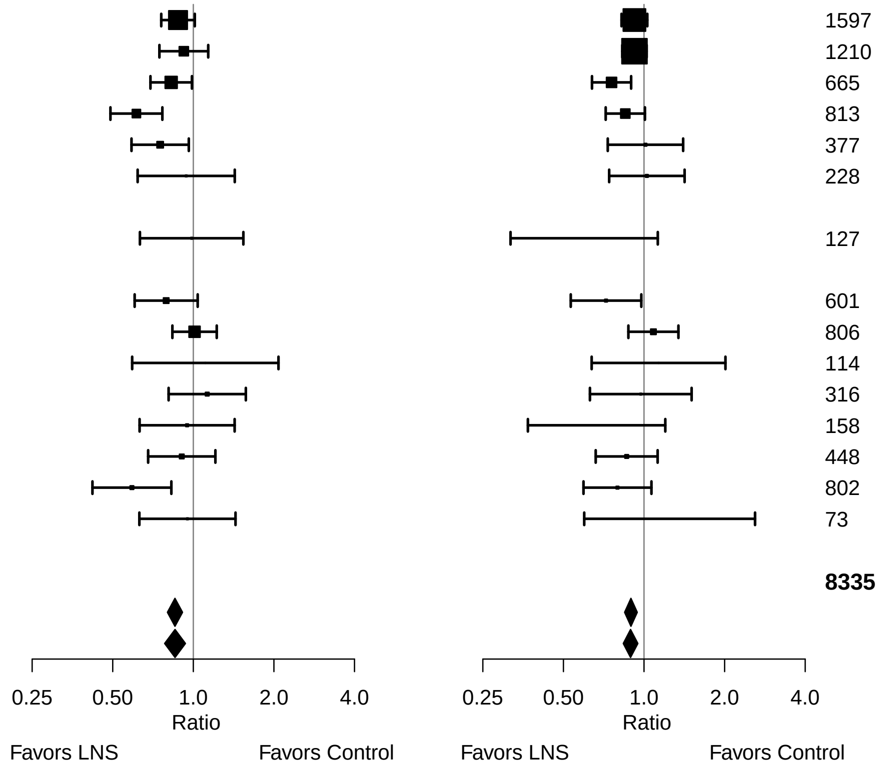

#### Supplemental figure 6M: Underweight prevalence ratio

#### 6M4: Stratified by Maternal education

**P−for−interaction = 0.399**
**Ratio of PRs = 1.05 (0.94, 1.17)**

| | | Primary or greater | | | | | | Incomplete or no formal | | | | | |
|---|---|---|---|---|---|---|---|---|---|---|---|---|---|
| **Country** | **Trial** | **LNS N** | **Control N** | **Control Prevalence** | **PR (95% CI)** | **Fixed W** | **Random W** | **LNS N** | **Control N** | **Control Prevalence** | **PR (95% CI)** | **Fixed W** | **Random W** |
| Bangladesh | JiVitA−4 (34) | 1816 | 749 | 36.6 | 0.84 (0.74, 0.95) | 0.29 | 0.22 | 996 | 480 | 43.3 | 0.98 (0.86, 1.12) | 0.26 | 0.19 |
| Bangladesh | RDNS (35) | 1245 | 589 | 35.8 | 0.93 (0.82, 1.06) | 0.26 | 0.21 | 418 | 226 | 42.9 | 0.92 (0.74, 1.14) | 0.10 | 0.11 |
| Bangladesh | WASH−B (36) | 818 | 2476 | 28.7 | 0.75 (0.66, 0.87) | 0.22 | 0.20 | 345 | 985 | 41.8 | 0.83 (0.69, 1.00) | 0.13 | 0.13 |
| Burkina Faso | iLiNS−Zinc (37) | | | | | | | | | | | | |
| Burkina Faso | PROMIS (38) | 70 | 54 | 13.0 | 1.65 (0.69, 3.98) | 0.01 | 0.01 | 787 | 857 | 20.1 | 0.82 (0.67, 1.01) | 0.10 | 0.12 |
| Burkina Faso | PROMIS CS (38) | 34 | 39 | 28.2 | 0.94 (0.41, 2.13) | 0.01 | 0.01 | 393 | 396 | 24.0 | 1.00 (0.77, 1.28) | 0.07 | 0.09 |
| Ghana | GHANA (39) | | | | | | | | | | | | |
| Ghana | iLiNS−DYADG (40) | 267 | 549 | 12.4 | 0.94 (0.63, 1.40) | 0.03 | 0.05 | 80 | 144 | 15.3 | 0.49 (0.21, 1.16) | 0.01 | 0.01 |
| Haiti | HAITI (41) | | | | | | | | | | | | |
| Kenya | WASH−B (42) | 705 | 2451 | 6.7 | 0.96 (0.68, 1.36) | 0.04 | 0.07 | 754 | 2710 | 13.1 | 0.67 (0.52, 0.87) | 0.06 | 0.08 |
| Madagascar | MAHAY (43) | 352 | 431 | 21.1 | 1.05 (0.79, 1.39) | 0.05 | 0.09 | 1350 | 1254 | 27.7 | 1.02 (0.86, 1.21) | 0.15 | 0.15 |
| Malawi | iLiNS−DYADM (44) | | | | | | | | | | | | |
| Malawi | iLiNS−DOSE (45) | | | | | | | | | | | | |
| Mali | PROMIS (46) | | | | | | | | | | | | |
| Mali | PROMIS CS (46) | 107 | 96 | 19.8 | 0.57 (0.28, 1.14) | 0.01 | 0.02 | 838 | 864 | 22.8 | 0.92 (0.75, 1.13) | 0.10 | 0.11 |
| Zimbabwe | SHINE (HIV−) (47) | 1707 | 1636 | 10.5 | 0.70 (0.56, 0.88) | 0.09 | 0.12 | 68 | 61 | 13.1 | 0.90 (0.37, 2.20) | 0.01 | 0.01 |
| Zimbabwe | SHINE (HIV+) (48) | | | | | | | | | | | | |
| | | **7121** | **9070** | | **I² = 0.34, Tau² = 0.01** | | | **6029** | **7977** | | **I² = 0.29, Tau² = 0.01** | | |
| **Fixed** | | | | | **0.85 (0.79, 0.90)** | | | | | | **0.91 (0.85, 0.97)** | | |
| **Random** | | | | | **0.85 (0.77, 0.94)** | | | | | | **0.90 (0.82, 0.98)** | | |

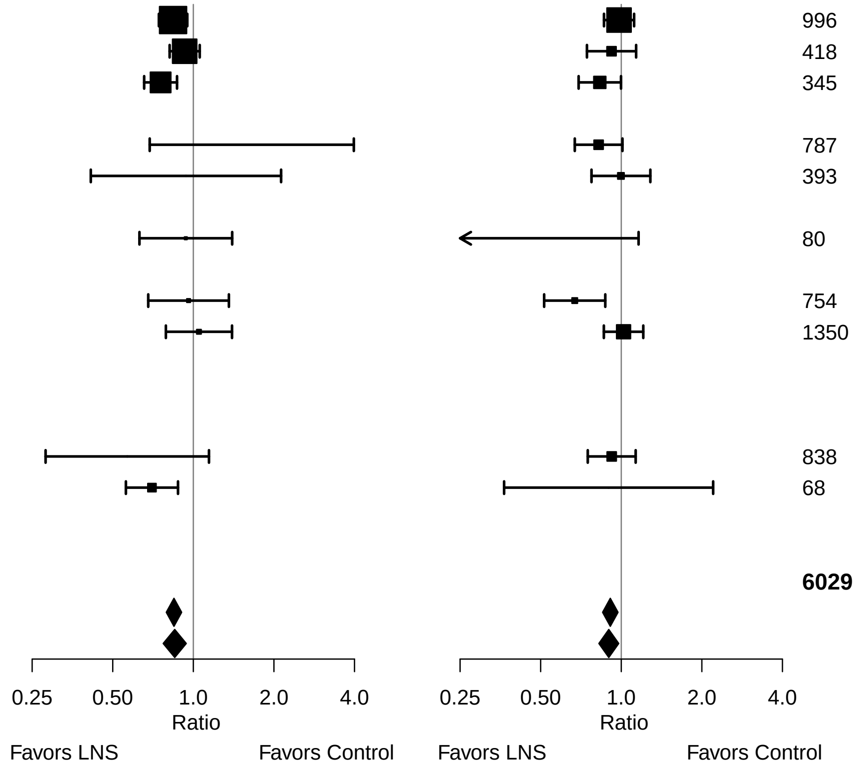

#### Supplemental figure 6M: Underweight prevalence ratio

### 6M5: Stratified by Maternal depressive symptoms

**P−for−interaction = 0.490**
**Ratio of PRs = 0.96 (0.84, 1.09)**

| | | Less than 75th percentile | | | | | | At least 75th percentile | | | | | |
|---|---|---|---|---|---|---|---|---|---|---|---|---|---|
| **Country** | **Trial** | **LNS N** | **Control N** | **Control Prevalence** | **PR (95% CI)** | **Fixed W** | **Random W** | **LNS N** | **Control N** | **Control Prevalence** | **PR (95% CI)** | **Fixed W** | **Random W** |
| Bangladesh | JiVitA−4 (34) | | | | | | | | | | | | |
| Bangladesh | RDNS (35) | 1081 | 472 | 36.9 | 0.93 (0.83, 1.04) | 0.35 | 0.34 | 520 | 288 | 38.2 | 0.93 (0.75, 1.16) | 0.26 | 0.18 |
| Bangladesh | WASH−B (36) | 899 | 2487 | 31.6 | 0.79 (0.69, 0.90) | 0.26 | 0.25 | 240 | 883 | 34.7 | 0.78 (0.61, 1.01) | 0.19 | 0.16 |
| Burkina Faso | iLiNS−Zinc (37) | | | | | | | | | | | | |
| Burkina Faso | PROMIS (38) | 505 | 571 | 19.4 | 0.79 (0.59, 1.08) | 0.05 | 0.05 | 220 | 213 | 21.1 | 0.95 (0.71, 1.26) | 0.15 | 0.14 |
| Burkina Faso | PROMIS CS (38) | | | | | | | | | | | | |
| Ghana | GHANA (39) | | | | | | | | | | | | |
| Ghana | iLiNS−DYADG (40) | 252 | 460 | 12.4 | 0.70 (0.44, 1.12) | 0.02 | 0.02 | 83 | 209 | 12.9 | 1.31 (0.72, 2.36) | 0.03 | 0.06 |
| Haiti | HAITI (41) | | | | | | | | | | | | |
| Kenya | WASH−B (42) | 997 | 3506 | 9.6 | 0.82 (0.66, 1.02) | 0.09 | 0.09 | 342 | 1248 | 10.5 | 0.72 (0.46, 1.15) | 0.06 | 0.08 |
| Madagascar | MAHAY (43) | 673 | 721 | 27.2 | 0.99 (0.81, 1.21) | 0.11 | 0.11 | 283 | 252 | 24.2 | 1.23 (0.90, 1.68) | 0.12 | 0.13 |
| Malawi | iLiNS−DYADM (44) | 157 | 303 | 11.2 | 1.02 (0.60, 1.75) | 0.02 | 0.02 | 47 | 113 | 12.4 | 1.20 (0.52, 2.79) | 0.02 | 0.03 |
| Malawi | iLiNS−DOSE (45) | | | | | | | | | | | | |
| Mali | PROMIS (46) | 374 | 372 | 15.8 | 0.79 (0.52, 1.18) | 0.03 | 0.03 | 122 | 129 | 18.3 | 1.06 (0.66, 1.70) | 0.05 | 0.08 |
| Mali | PROMIS CS (46) | | | | | | | | | | | | |
| Zimbabwe | SHINE (HIV−) (47) | 1192 | 1137 | 9.9 | 0.76 (0.57, 1.02) | 0.05 | 0.05 | 509 | 490 | 12.7 | 0.54 (0.37, 0.81) | 0.08 | 0.10 |
| Zimbabwe | SHINE (HIV+) (48) | 234 | 214 | 16.4 | 1.10 (0.74, 1.63) | 0.03 | 0.03 | 88 | 100 | 20.0 | 0.85 (0.46, 1.59) | 0.03 | 0.05 |
| | | **6364** | **10243** | | **I² = 0.03, Tau² = 0.00** | | | **2454** | **3925** | | **I² = 0.38, Tau² = 0.02** | | |
| **Fixed** | | | | | **0.87 (0.81, 0.93)** | | | | | | **0.90 (0.81, 1.01)** | | |
| **Random** | | | | | **0.87 (0.81, 0.93)** | | | | | | **0.90 (0.77, 1.06)** | | |

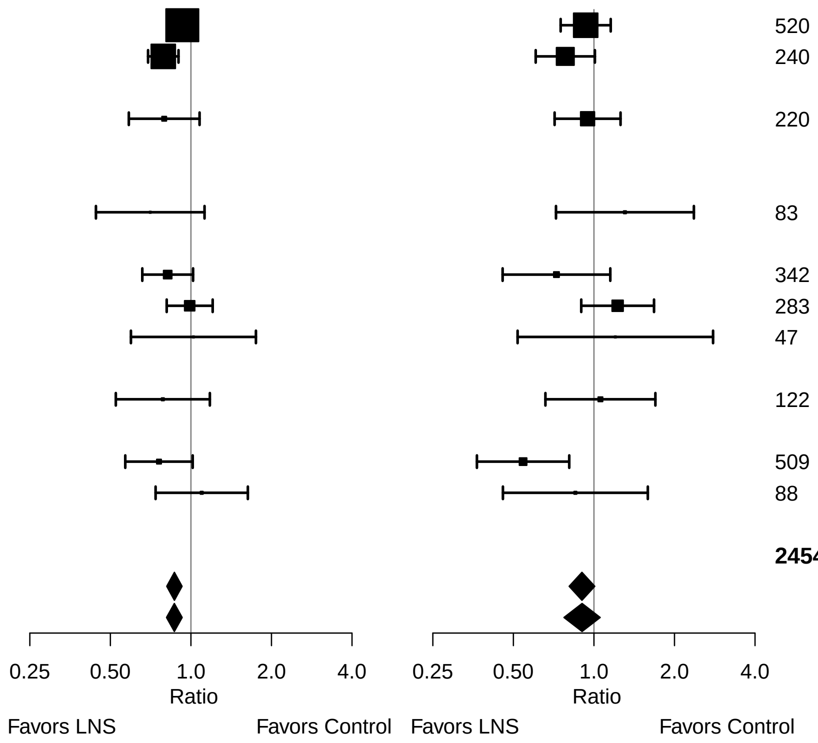

#### Supplemental figure 6M: Underweight prevalence ratio

### 6M6: Stratified by Child sex

**P−for−interaction = 0.617**
**Ratio of PRs = 0.98 (0.91, 1.06)**

| | | Male | | | | | | Female | | | | | |
|---|---|---|---|---|---|---|---|---|---|---|---|---|---|
| **Country** | **Trial** | **LNS N** | **Control N** | **Control Prevalence** | **PR (95% CI)** | **Fixed W** | **Random W** | **LNS N** | **Control N** | **Control Prevalence** | **PR (95% CI)** | **Fixed W** | **Random W** |
| Bangladesh | JiVitA−4 (34) | 1440 | 594 | 38.2 | 0.95 (0.83, 1.09) | 0.19 | 0.13 | 1374 | 638 | 40.1 | 0.87 (0.77, 0.98) | 0.25 | 0.14 |
| Bangladesh | RDNS (35) | 833 | 411 | 38.0 | 0.95 (0.82, 1.10) | 0.14 | 0.12 | 830 | 404 | 37.6 | 0.89 (0.78, 1.02) | 0.21 | 0.13 |
| Bangladesh | WASH−B (36) | 567 | 1730 | 33.2 | 0.78 (0.67, 0.91) | 0.14 | 0.12 | 596 | 1731 | 31.7 | 0.79 (0.67, 0.94) | 0.13 | 0.12 |
| Burkina Faso | iLiNS−Zinc (37) | 995 | 336 | 32.4 | 0.78 (0.64, 0.95) | 0.08 | 0.09 | 965 | 330 | 29.4 | 0.60 (0.48, 0.75) | 0.08 | 0.09 |
| Burkina Faso | PROMIS (38) | 435 | 482 | 24.9 | 0.81 (0.64, 1.04) | 0.05 | 0.07 | 422 | 429 | 13.8 | 0.98 (0.67, 1.43) | 0.03 | 0.05 |
| Burkina Faso | PROMIS CS (38) | 234 | 221 | 27.6 | 1.04 (0.75, 1.43) | 0.03 | 0.04 | 196 | 215 | 20.9 | 0.88 (0.59, 1.30) | 0.02 | 0.04 |
| Ghana | GHANA (39) | | | | | | | | | | | | |
| Ghana | iLiNS−DYADG (40) | 175 | 324 | 16.4 | 0.73 (0.46, 1.17) | 0.01 | 0.02 | 172 | 369 | 10.0 | 0.93 (0.53, 1.62) | 0.01 | 0.03 |
| Haiti | HAITI (41) | | | | | | | | | | | | |
| Kenya | WASH−B (42) | 734 | 2457 | 11.8 | 0.74 (0.57, 0.96) | 0.05 | 0.06 | 727 | 2708 | 8.4 | 0.77 (0.59, 1.00) | 0.05 | 0.08 |
| Madagascar | MAHAY (43) | 826 | 835 | 31.4 | 1.00 (0.84, 1.19) | 0.11 | 0.10 | 876 | 850 | 20.7 | 1.10 (0.90, 1.35) | 0.09 | 0.10 |
| Malawi | iLiNS−DYADM (44) | 107 | 207 | 12.6 | 1.41 (0.82, 2.43) | 0.01 | 0.02 | 113 | 237 | 11.0 | 0.81 (0.40, 1.61) | 0.01 | 0.02 |
| Malawi | iLiNS−DOSE (45) | 353 | 124 | 29.0 | 0.97 (0.71, 1.31) | 0.04 | 0.05 | 349 | 117 | 10.3 | 1.19 (0.73, 1.93) | 0.02 | 0.03 |
| Mali | PROMIS (46) | 265 | 262 | 18.6 | 0.83 (0.60, 1.16) | 0.03 | 0.04 | 235 | 240 | 14.0 | 0.87 (0.50, 1.50) | 0.01 | 0.03 |
| Mali | PROMIS CS (46) | 474 | 521 | 25.1 | 0.88 (0.66, 1.17) | 0.04 | 0.05 | 472 | 439 | 19.4 | 0.91 (0.68, 1.21) | 0.05 | 0.07 |
| Zimbabwe | SHINE (HIV−) (47) | 924 | 906 | 13.4 | 0.66 (0.51, 0.84) | 0.05 | 0.06 | 945 | 880 | 7.7 | 0.81 (0.57, 1.14) | 0.03 | 0.05 |
| Zimbabwe | SHINE (HIV+) (48) | 161 | 167 | 22.8 | 0.79 (0.53, 1.19) | 0.02 | 0.03 | 175 | 161 | 11.2 | 1.48 (0.88, 2.49) | 0.01 | 0.03 |
| | | **8523** | **9577** | | **I² = 0.32, Tau² = 0.01** | | | **8447** | **9748** | | **I² = 0.44, Tau² = 0.01** | | |
| **Fixed** | | | | | **0.87 (0.82, 0.92)** | | | | | | **0.86 (0.81, 0.92)** | | |
| **Random** | | | | | **0.87 (0.80, 0.93)** | | | | | | **0.87 (0.79, 0.96)** | | |

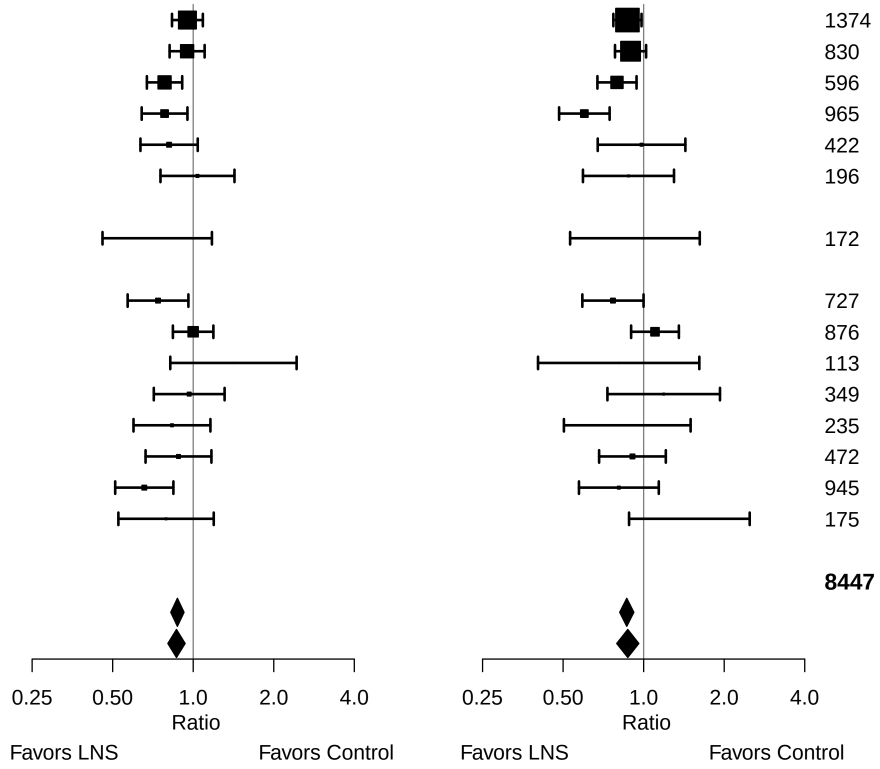

#### Supplemental figure 6M: Underweight prevalence ratio

### 6M7: Stratified by Child birth order

**P−for−interaction = 0.001**
**Ratio of PRs = 1.16 (1.06, 1.27)**

**Later born**

| Country | Trial | LNS N | Control N | Control Prevalence | PR (95% CI) | Fixed W | Random W |
|---|---|---|---|---|---|---|---|
| Bangladesh | JiVitA−4 (34) | 600 | 250 | 43.3 | 0.80 (0.69, 0.92) | 0.13 | 0.09 |
| Bangladesh | RDNS (35) | 974 | 508 | 38.0 | 0.91 (0.79, 1.05) | 0.15 | 0.09 |
| Bangladesh | WASH−B (36) | 737 | 2310 | 34.3 | 0.78 (0.67, 0.90) | 0.14 | 0.09 |
| Burkina Faso | iLiNS−Zinc (37) | 1524 | 521 | 30.9 | 0.63 (0.53, 0.75) | 0.10 | 0.09 |
| Burkina Faso | PROMIS (38) | 709 | 750 | 19.9 | 0.80 (0.66, 0.98) | 0.07 | 0.08 |
| Burkina Faso | PROMIS CS (38) | 366 | 357 | 23.8 | 0.99 (0.76, 1.29) | 0.04 | 0.06 |
| Ghana | GHANA (39) | | | | | | |
| Ghana | iLiNS−DYADG (40) | 237 | 459 | 12.2 | 0.97 (0.63, 1.48) | 0.02 | 0.03 |
| Haiti | HAITI (41) | | | | | | |
| Kenya | WASH−B (42) | 1137 | 4065 | 10.2 | 0.76 (0.60, 0.95) | 0.06 | 0.07 |
| Madagascar | MAHAY (43) | 1271 | 1220 | 27.2 | 0.98 (0.83, 1.16) | 0.10 | 0.09 |
| Malawi | iLiNS−DYADM (44) | 173 | 353 | 11.3 | 1.17 (0.73, 1.90) | 0.01 | 0.03 |
| Malawi | iLiNS−DOSE (45) | 463 | 165 | 19.4 | 1.06 (0.79, 1.42) | 0.03 | 0.05 |
| Mali | PROMIS (46) | 437 | 441 | 16.9 | 0.79 (0.57, 1.09) | 0.03 | 0.05 |
| Mali | PROMIS CS (46) | 784 | 822 | 22.5 | 0.86 (0.69, 1.06) | 0.06 | 0.07 |
| Zimbabwe | SHINE (HIV−) (47) | 1295 | 1220 | 10.4 | 0.61 (0.45, 0.82) | 0.03 | 0.05 |
| Zimbabwe | SHINE (HIV+) (48) | 279 | 245 | 14.3 | 1.25 (0.84, 1.88) | 0.02 | 0.04 |
| | | **10986** | **13686** | | **I² = 0.56, Tau² = 0.02** | | |
| **Fixed** | | | | | **0.83 (0.79, 0.88)** | | |
| **Random** | | | | | **0.84 (0.77, 0.93)** | | |

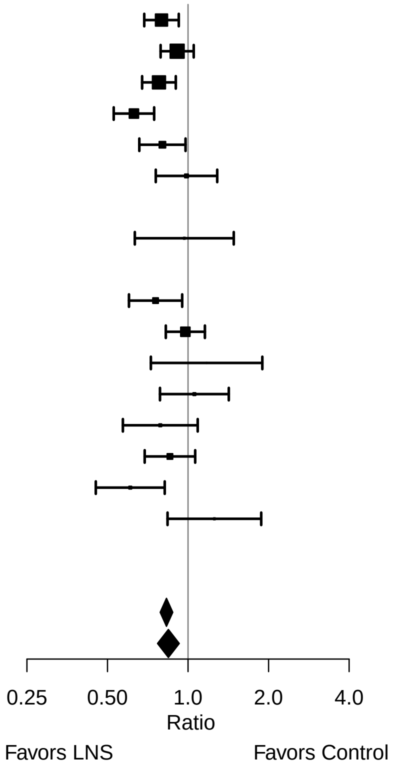

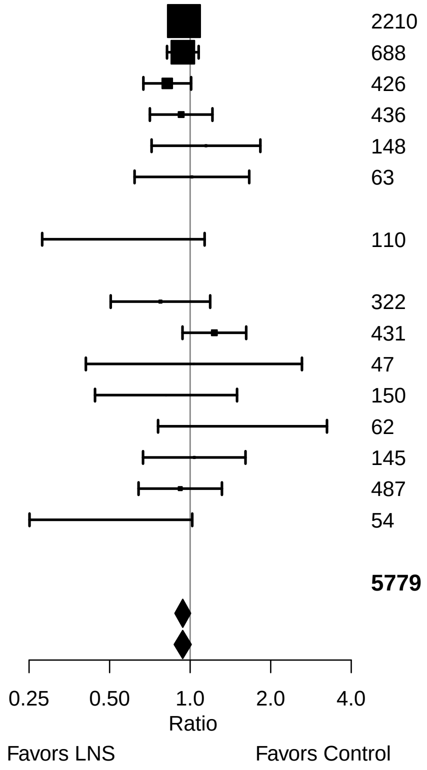

**Firstborn**

| Country | Trial | LNS N | Control N | Control Prevalence | PR (95% CI) | Fixed W | Random W |
|---|---|---|---|---|---|---|---|
| Bangladesh | JiVitA−4 (34) | 2210 | 976 | 38.0 | 0.95 (0.85, 1.06) | 0.35 | 0.31 |
| Bangladesh | RDNS (35) | 688 | 307 | 37.5 | 0.94 (0.82, 1.08) | 0.25 | 0.24 |
| Bangladesh | WASH−B (36) | 426 | 1151 | 28.6 | 0.82 (0.67, 1.01) | 0.11 | 0.12 |
| Burkina Faso | iLiNS−Zinc (37) | 436 | 145 | 31.0 | 0.93 (0.71, 1.21) | 0.06 | 0.07 |
| Burkina Faso | PROMIS (38) | 148 | 159 | 18.9 | 1.15 (0.72, 1.83) | 0.02 | 0.02 |
| Burkina Faso | PROMIS CS (38) | 63 | 79 | 26.6 | 1.02 (0.62, 1.66) | 0.02 | 0.02 |
| Ghana | GHANA (39) | | | | | | |
| Ghana | iLiNS−DYADG (40) | 110 | 234 | 14.5 | 0.56 (0.28, 1.13) | 0.01 | 0.01 |
| Haiti | HAITI (41) | | | | | | |
| Kenya | WASH−B (42) | 322 | 1095 | 9.2 | 0.77 (0.50, 1.19) | 0.03 | 0.03 |
| Madagascar | MAHAY (43) | 431 | 465 | 22.8 | 1.23 (0.94, 1.62) | 0.06 | 0.07 |
| Malawi | iLiNS−DYADM (44) | 47 | 89 | 12.4 | 1.03 (0.41, 2.62) | 0.01 | 0.01 |
| Malawi | iLiNS−DOSE (45) | 150 | 45 | 22.2 | 0.81 (0.44, 1.50) | 0.01 | 0.01 |
| Mali | PROMIS (46) | 62 | 59 | 11.9 | 1.57 (0.76, 3.25) | 0.01 | 0.01 |
| Mali | PROMIS CS (46) | 145 | 126 | 20.6 | 1.04 (0.67, 1.61) | 0.02 | 0.03 |
| Zimbabwe | SHINE (HIV−) (47) | 487 | 483 | 11.2 | 0.92 (0.64, 1.31) | 0.04 | 0.04 |
| Zimbabwe | SHINE (HIV+) (48) | 54 | 78 | 25.6 | 0.51 (0.25, 1.02) | 0.01 | 0.01 |
| | | **5779** | **5491** | | **I² = 0.03, Tau² = 0.00** | | |
| **Fixed** | | | | | **0.94 (0.88, 1.00)** | | |
| **Random** | | | | | **0.94 (0.87, 1.01)** | | |

#### Supplemental figure 6M: Underweight prevalence ratio

#### 6M8: Stratified by Child baseline anthropometric status

**P−for−interaction = 0.493**
**Ratio of PRs = 1.09 (0.85, 1.39)**

| | | WAZ at least −1 Z | | | | | | WAZ less than −1 Z | | | | | |
|---|---|---|---|---|---|---|---|---|---|---|---|---|---|
| **Country** | **Trial** | **LNS N** | **Control N** | **Control Prevalence** | **PR (95% CI)** | **Fixed W** | **Random W** | **LNS N** | **Control N** | **Control Prevalence** | **PR (95% CI)** | **Fixed W** | **Random W** |
| Bangladesh | JiVitA−4 (34) | 1198 | 531 | 6.4 | 0.61 (0.40, 0.95) | 0.30 | 0.30 | 1616 | 701 | 64.3 | 0.89 (0.84, 0.96) | 0.50 | 0.36 |
| Bangladesh | RDNS (35) | 780 | 338 | 5.6 | 1.12 (0.69, 1.81) | 0.24 | 0.24 | 821 | 422 | 62.8 | 0.98 (0.90, 1.07) | 0.33 | 0.31 |
| Bangladesh | WASH−B (36) | | | | | | | | | | | | |
| Burkina Faso | iLiNS−Zinc (37) | | | | | | | | | | | | |
| Burkina Faso | PROMIS (38) | 427 | 482 | 5.8 | 0.77 (0.44, 1.32) | 0.19 | 0.19 | 268 | 267 | 42.7 | 0.87 (0.72, 1.06) | 0.06 | 0.11 |
| Burkina Faso | PROMIS CS (38) | | | | | | | | | | | | |
| Ghana | GHANA (39) | | | | | | | | | | | | |
| Ghana | iLiNS−DYADG (40) | 222 | 424 | 1.7 | 1.36 (0.44, 4.25) | 0.04 | 0.04 | 101 | 215 | 34.0 | 0.93 (0.66, 1.31) | 0.02 | 0.04 |
| Haiti | HAITI (41) | | | | | | | | | | | | |
| Kenya | WASH−B (42) | | | | | | | | | | | | |
| Madagascar | MAHAY (43) | | | | | | | | | | | | |
| Malawi | iLiNS−DYADM (44) | | | | | | | | | | | | |
| Malawi | iLiNS−DOSE (45) | 419 | 143 | 3.5 | 0.89 (0.32, 2.45) | 0.06 | 0.06 | 283 | 98 | 43.9 | 0.96 (0.74, 1.25) | 0.03 | 0.07 |
| Mali | PROMIS (46) | | | | | | | | | | | | |
| Mali | PROMIS CS (46) | | | | | | | | | | | | |
| Zimbabwe | SHINE (HIV−) (47) | 917 | 836 | 1.9 | 0.85 (0.41, 1.77) | 0.11 | 0.11 | 383 | 349 | 30.1 | 0.70 (0.56, 0.89) | 0.04 | 0.08 |
| Zimbabwe | SHINE (HIV+) (48) | 151 | 144 | 5.6 | 0.83 (0.29, 2.36) | 0.05 | 0.05 | 113 | 92 | 32.6 | 1.03 (0.70, 1.51) | 0.02 | 0.03 |
| | | **4114** | **2898** | | **$I^2$ = 0.00, $Tau^2$ = 0.00** | | | **3585** | **2144** | | **$I^2$ = 0.31, $Tau^2$ = 0.00** | | |
| **Fixed** | | | | | **0.83 (0.65, 1.05)** | | | | | | **0.92 (0.87, 0.96)** | | |
| **Random** | | | | | **0.83 (0.65, 1.05)** | | | | | | **0.91 (0.85, 0.98)** | | |

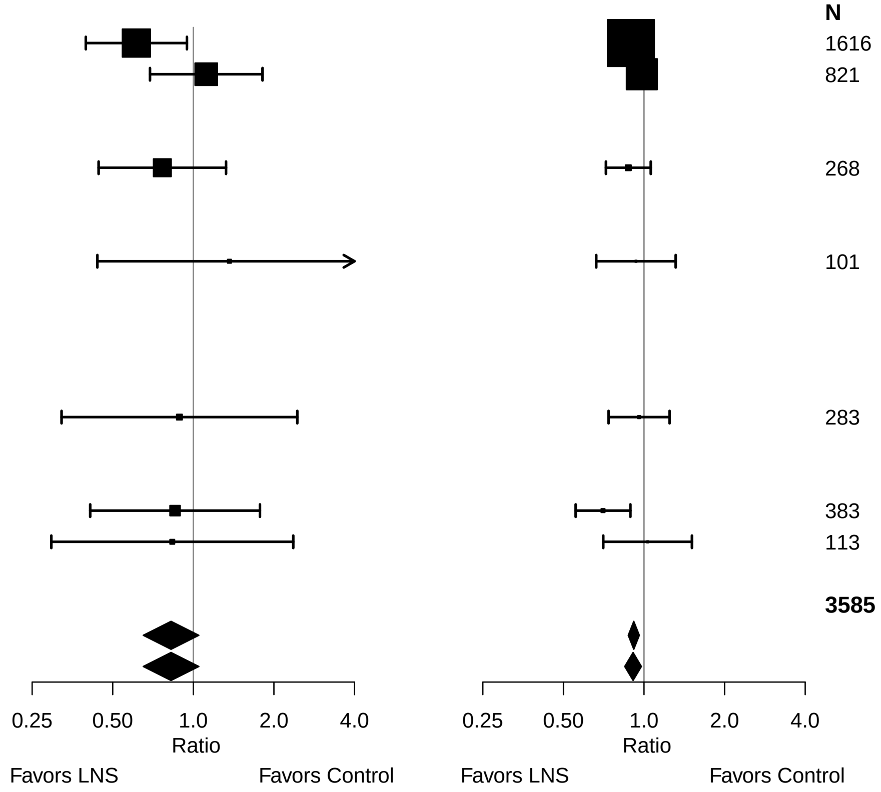

### Supplemental figure 6N: Underweight prevalence difference

#### 6N1: Stratified by Maternal height

**P−for−interaction = 0.211**
**Difference in PDs = −0.02 (−0.05, 0.01)**

| | | At least 150.1 cm | | | | | | Less than 150.1 cm | | | | | |
|---|---|---|---|---|---|---|---|---|---|---|---|---|---|
| **Country** | **Trial** | **LNS N** | **Control N** | **Control Prevalence** | **PD (95% CI)** | **Fixed W** | **Random W** | **LNS N** | **Control N** | **Control Prevalence** | **PD (95% CI)** | **Fixed W** | **Random W** |
| Bangladesh | JiVitA−4 (34) | | | | | | | | | | | | |
| Bangladesh | RDNS (35) | 868 | 432 | 31.7 | −0.06 (−0.11, −0.01) | 0.04 | 0.08 | 735 | 358 | 45.8 | −0.01 (−0.07, 0.05) | 0.22 | 0.20 |
| Bangladesh | WASH−B (36) | 620 | 1898 | 25.7 | −0.05 (−0.09, −0.01) | 0.07 | 0.10 | 543 | 1557 | 40.7 | −0.10 (−0.14, −0.05) | 0.37 | 0.25 |
| Burkina Faso | iLiNS−Zinc (37) | 1924 | 651 | 30.9 | −0.10 (−0.14, −0.07) | 0.08 | 0.11 | 34 | 15 | 33.3 | 0.11 (−0.09, 0.30) | 0.02 | 0.03 |
| Burkina Faso | PROMIS (38) | 829 | 881 | 19.3 | −0.03 (−0.07, 0.01) | 0.06 | 0.10 | 25 | 25 | 36.0 | 0.00 (−0.26, 0.26) | 0.01 | 0.02 |
| Burkina Faso | PROMIS CS (38) | | | | | | | | | | | | |
| Ghana | GHANA (39) | | | | | | | | | | | | |
| Ghana | iLiNS−DYADG (40) | | | | | | | | | | | | |
| Haiti | HAITI (41) | | | | | | | | | | | | |
| Kenya | WASH−B (42) | 1316 | 4709 | 9.0 | −0.02 (−0.03, 0.00) | 0.33 | 0.14 | 57 | 195 | 20.0 | −0.09 (−0.19, 0.00) | 0.09 | 0.12 |
| Madagascar | MAHAY (43) | 1003 | 1040 | 22.2 | 0.02 (−0.03, 0.06) | 0.05 | 0.09 | 598 | 565 | 33.8 | −0.02 (−0.09, 0.05) | 0.16 | 0.17 |
| Malawi | iLiNS−DYADM (44) | 192 | 381 | 9.7 | 0.01 (−0.04, 0.06) | 0.04 | 0.08 | 28 | 60 | 21.7 | 0.10 (−0.09, 0.30) | 0.02 | 0.03 |
| Malawi | iLiNS−DOSE (45) | 586 | 198 | 16.7 | −0.01 (−0.06, 0.05) | 0.04 | 0.08 | 115 | 42 | 33.3 | −0.06 (−0.19, 0.07) | 0.04 | 0.07 |
| Mali | PROMIS (46) | | | | | | | | | | | | |
| Mali | PROMIS CS (46) | 909 | 928 | 22.3 | −0.03 (−0.08, 0.01) | 0.05 | 0.09 | 32 | 26 | 34.6 | 0.06 (−0.15, 0.27) | 0.02 | 0.03 |
| Zimbabwe | SHINE (HIV−) (47) | 1745 | 1690 | 10.4 | −0.03 (−0.05, −0.01) | 0.26 | 0.14 | 75 | 62 | 19.4 | −0.07 (−0.19, 0.04) | 0.06 | 0.08 |
| Zimbabwe | SHINE (HIV+) (48) | | | | | | | | | | | | |
| | | **9992** | **12808** | | **I² = 0.68, Tau² = 0.00** | | | **2242** | **2905** | | **I² = 0.35, Tau² = 0.00** | | |
| **Fixed** | | | | | **−0.03 (−0.04, −0.02)** | | | | | | **−0.05 (−0.08, −0.02)** | | |
| **Random** | | | | | **−0.03 (−0.05, −0.01)** | | | | | | **−0.04 (−0.08, 0.00)** | | |

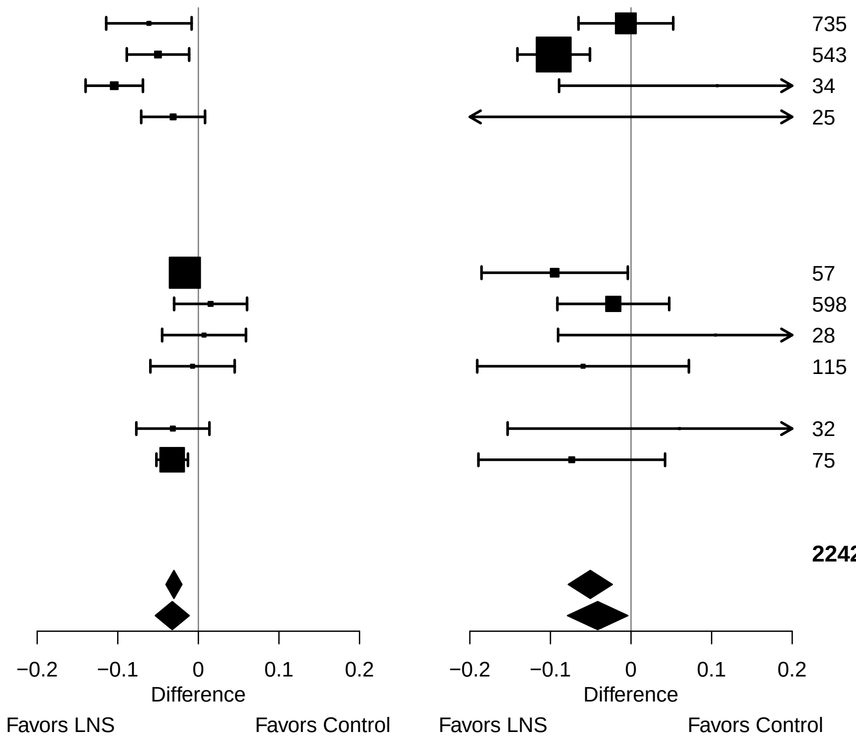

#### Supplemental figure 6N: Underweight prevalence difference

#### 6N2: Stratified by Maternal BMI

**P−for−interaction = 0.263**
**Difference in PDs = −0.01 (−0.03, 0.01)**

| | | At least 20 kg/m² | | | | | | Less than 20 kg/m² | | | | | |
|---|---|---|---|---|---|---|---|---|---|---|---|---|---|
| Country | Trial | LNS N | Control N | Control Prevalence | PD (95% CI) | Fixed W | Random W | LNS N | Control N | Control Prevalence | PD (95% CI) | Fixed W | Random W |
| Bangladesh | JiVitA−4 (34) | | | | | | | | | | | | |
| Bangladesh | RDNS (35) | 730 | 335 | 29.3 | −0.02 (−0.08, 0.03) | 0.04 | 0.04 | 873 | 455 | 44.6 | −0.04 (−0.09, 0.02) | 0.12 | 0.11 |
| Bangladesh | WASH−B (36) | 518 | 1595 | 24.6 | −0.04 (−0.09, 0.00) | 0.05 | 0.05 | 645 | 1860 | 39.2 | −0.09 (−0.14, −0.05) | 0.18 | 0.12 |
| Burkina Faso | iLiNS−Zinc (37) | 1218 | 391 | 24.8 | −0.08 (−0.13, −0.03) | 0.05 | 0.05 | 740 | 275 | 39.6 | −0.13 (−0.18, −0.08) | 0.14 | 0.11 |
| Burkina Faso | PROMIS (38) | 645 | 673 | 17.8 | −0.05 (−0.09, 0.00) | 0.06 | 0.06 | 209 | 232 | 25.0 | 0.03 (−0.04, 0.11) | 0.05 | 0.08 |
| Burkina Faso | PROMIS CS (38) | 246 | 267 | 18.4 | 0.00 (−0.06, 0.07) | 0.03 | 0.03 | 184 | 165 | 33.9 | −0.03 (−0.11, 0.05) | 0.05 | 0.07 |
| Ghana | GHANA (39) | | | | | | | | | | | | |
| Ghana | iLiNS−DYADG (40) | | | | | | | | | | | | |
| Haiti | HAITI (41) | | | | | | | | | | | | |
| Kenya | WASH−B (42) | 1100 | 3822 | 8.4 | −0.02 (−0.03, 0.00) | 0.32 | 0.32 | 273 | 1082 | 13.4 | −0.03 (−0.07, 0.01) | 0.21 | 0.13 |
| Madagascar | MAHAY (43) | | | | | | | | | | | | |
| Malawi | iLiNS−DYADM (44) | 131 | 263 | 11.4 | −0.01 (−0.08, 0.05) | 0.03 | 0.03 | 88 | 178 | 11.2 | 0.06 (−0.03, 0.14) | 0.04 | 0.07 |
| Malawi | iLiNS−DOSE (45) | 519 | 174 | 15.5 | −0.01 (−0.06, 0.05) | 0.04 | 0.04 | 180 | 65 | 30.8 | −0.05 (−0.15, 0.06) | 0.03 | 0.05 |
| Mali | PROMIS (46) | 377 | 368 | 14.3 | −0.04 (−0.09, 0.02) | 0.04 | 0.04 | 117 | 133 | 22.2 | −0.04 (−0.13, 0.05) | 0.04 | 0.06 |
| Mali | PROMIS CS (46) | 697 | 676 | 20.4 | −0.02 (−0.07, 0.03) | 0.05 | 0.05 | 244 | 278 | 28.1 | −0.04 (−0.12, 0.04) | 0.05 | 0.08 |
| Zimbabwe | SHINE (HIV−) (47) | 1355 | 1300 | 10.2 | −0.04 (−0.06, −0.02) | 0.25 | 0.25 | 239 | 249 | 14.9 | −0.02 (−0.09, 0.04) | 0.07 | 0.09 |
| Zimbabwe | SHINE (HIV+) (48) | 246 | 253 | 15.4 | −0.02 (−0.07, 0.04) | 0.03 | 0.03 | 62 | 52 | 26.9 | 0.02 (−0.13, 0.18) | 0.01 | 0.03 |
| | | **7782** | **10117** | | **I² = 0.00, Tau² = 0.00** | | | **3854** | **5024** | | **I² = 0.60, Tau² = 0.00** | | |
| **Fixed** | | | | | **−0.03 (−0.04, −0.02)** | | | | | | **−0.05 (−0.07, −0.03)** | | |
| **Random** | | | | | **−0.03 (−0.04, −0.02)** | | | | | | **−0.04 (−0.07, −0.01)** | | |

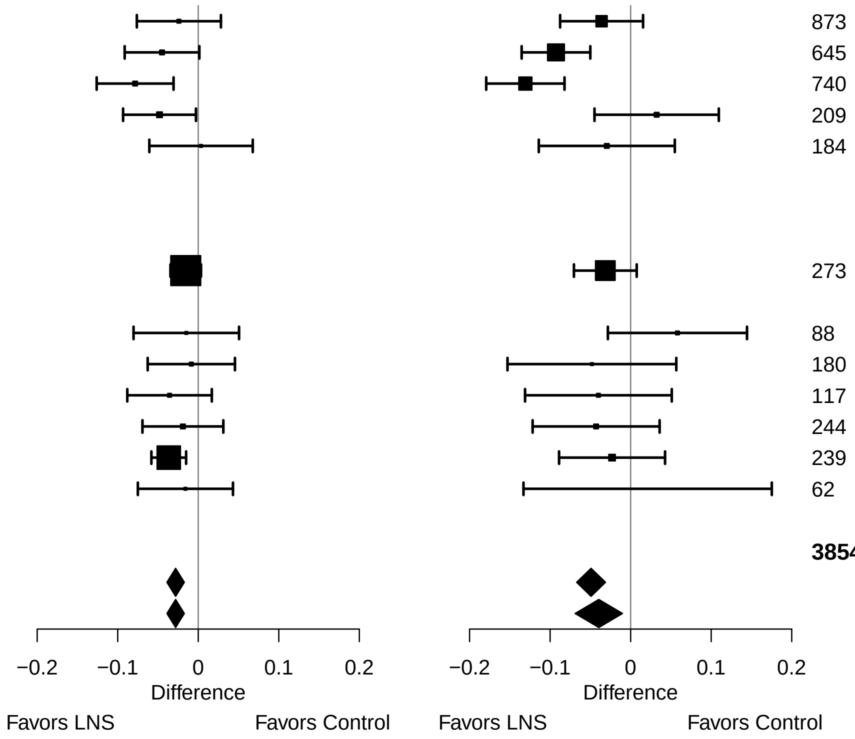

#### Supplemental figure 6N: Underweight prevalence difference

##### 6N3: Stratified by Maternal age

**P−for−interaction = 0.628**
**Difference in PDs = 0.00 (−0.01, 0.02)**

| | | At least 25 y | | | | | | Less than 25 y | | | | | |
|---|---|---|---|---|---|---|---|---|---|---|---|---|---|
| **Country** | **Trial** | **LNS N** | **Control N** | **Control Prevalence** | **PD (95% CI)** | **Fixed W** | **Random W** | **LNS N** | **Control N** | **Control Prevalence** | **PD (95% CI)** | **Fixed W** | **Random W** |
| Bangladesh | JiVitA−4 (34) | 1204 | 540 | 40.6 | −0.05 (−0.09, −0.01) | 0.08 | 0.09 | 1597 | 687 | 38.1 | −0.04 (−0.07, 0.00) | 0.12 | 0.11 |
| Bangladesh | RDNS (35) | 453 | 221 | 36.7 | −0.03 (−0.10, 0.05) | 0.02 | 0.04 | 1210 | 594 | 38.2 | −0.03 (−0.07, 0.01) | 0.09 | 0.09 |
| Bangladesh | WASH−B (36) | 493 | 1521 | 34.8 | −0.06 (−0.11, −0.01) | 0.04 | 0.06 | 665 | 1924 | 30.6 | −0.07 (−0.12, −0.03) | 0.07 | 0.08 |
| Burkina Faso | iLiNS−Zinc (37) | 1134 | 401 | 31.7 | −0.12 (−0.17, −0.07) | 0.05 | 0.07 | 813 | 262 | 29.8 | −0.07 (−0.12, −0.03) | 0.07 | 0.08 |
| Burkina Faso | PROMIS (38) | 480 | 524 | 20.2 | −0.05 (−0.09, −0.01) | 0.07 | 0.08 | 377 | 387 | 18.9 | 0.00 (−0.06, 0.06) | 0.04 | 0.05 |
| Burkina Faso | PROMIS CS (38) | 202 | 190 | 24.2 | −0.01 (−0.11, 0.08) | 0.01 | 0.02 | 228 | 246 | 24.4 | 0.01 (−0.07, 0.09) | 0.02 | 0.03 |
| Ghana | GHANA (39) | | | | | | | | | | | | |
| Ghana | iLiNS−DYADG (40) | 220 | 417 | 12.0 | 0.00 (−0.05, 0.05) | 0.05 | 0.06 | 127 | 276 | 14.5 | −0.06 (−0.13, 0.01) | 0.03 | 0.04 |
| Haiti | HAITI (41) | | | | | | | | | | | | |
| Kenya | WASH−B (42) | 845 | 2827 | 9.3 | −0.02 (−0.04, 0.00) | 0.30 | 0.15 | 601 | 2306 | 10.8 | −0.03 (−0.06, 0.00) | 0.21 | 0.15 |
| Madagascar | MAHAY (43) | 895 | 952 | 27.4 | 0.00 (−0.05, 0.06) | 0.05 | 0.06 | 806 | 733 | 24.1 | 0.02 (−0.03, 0.07) | 0.05 | 0.06 |
| Malawi | iLiNS−DYADM (44) | 106 | 226 | 11.1 | 0.01 (−0.06, 0.09) | 0.02 | 0.04 | 114 | 218 | 12.4 | 0.02 (−0.06, 0.09) | 0.02 | 0.03 |
| Malawi | iLiNS−DOSE (45) | 372 | 134 | 20.1 | 0.00 (−0.06, 0.07) | 0.03 | 0.05 | 316 | 100 | 18.0 | −0.03 (−0.10, 0.05) | 0.02 | 0.03 |
| Mali | PROMIS (46) | 342 | 355 | 15.6 | −0.02 (−0.08, 0.03) | 0.05 | 0.06 | 158 | 147 | 18.4 | −0.07 (−0.15, 0.00) | 0.02 | 0.03 |
| Mali | PROMIS CS (46) | 498 | 502 | 21.5 | −0.02 (−0.08, 0.04) | 0.04 | 0.05 | 448 | 458 | 23.6 | −0.03 (−0.09, 0.02) | 0.04 | 0.05 |
| Zimbabwe | SHINE (HIV−) (47) | 870 | 797 | 10.9 | −0.04 (−0.07, −0.02) | 0.16 | 0.12 | 802 | 793 | 10.3 | −0.02 (−0.05, 0.01) | 0.19 | 0.14 |
| Zimbabwe | SHINE (HIV+) (48) | 243 | 231 | 17.7 | −0.01 (−0.08, 0.06) | 0.03 | 0.04 | 73 | 77 | 14.3 | 0.04 (−0.08, 0.15) | 0.01 | 0.01 |
| | | **8357** | **9838** | | **I² = 0.39, Tau² = 0.00** | | | **8335** | **9208** | | **I² = 0.23, Tau² = 0.00** | | |
| **Fixed** | | | | | **−0.03 (−0.04, −0.02)** | | | | | | **−0.03 (−0.04, −0.02)** | | |
| **Random** | | | | | **−0.03 (−0.05, −0.02)** | | | | | | **−0.03 (−0.05, −0.02)** | | |

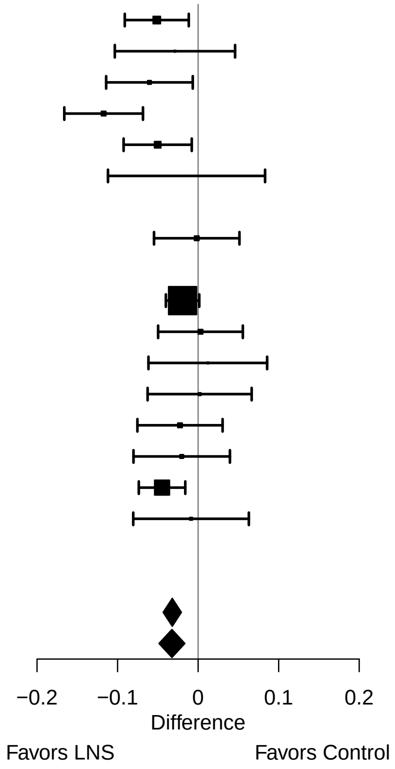

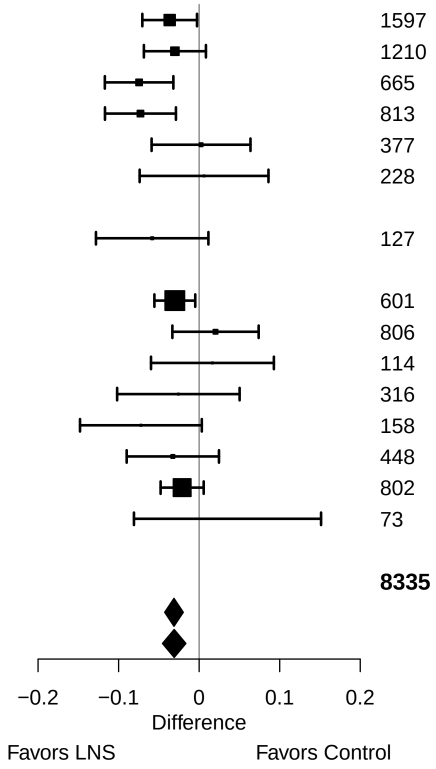

#### Supplemental figure 6N: Underweight prevalence difference

##### 6N4: Stratified by Maternal education

**P−for−interaction = 0.136**
**Difference in PDs = −0.02 (−0.04, 0.01)**

| | | Primary or greater | | | | | | Incomplete or no formal | | | | | |
|---|---|---|---|---|---|---|---|---|---|---|---|---|---|
| **Country** | **Trial** | **LNS N** | **Control N** | **Control Prevalence** | **PD (95% CI)** | **Fixed W** | **Random W** | **LNS N** | **Control N** | **Control Prevalence** | **PD (95% CI)** | **Fixed W** | **Random W** |
| Bangladesh | JiVitA−4 (34) | 1816 | 749 | 36.6 | −0.06 (−0.09, −0.02) | 0.11 | 0.14 | 996 | 480 | 43.3 | −0.02 (−0.06, 0.02) | 0.13 | 0.13 |
| Bangladesh | RDNS (35) | 1245 | 589 | 35.8 | −0.03 (−0.07, 0.02) | 0.06 | 0.11 | 418 | 226 | 42.9 | −0.03 (−0.12, 0.05) | 0.03 | 0.03 |
| Bangladesh | WASH−B (36) | 818 | 2476 | 28.7 | −0.07 (−0.10, −0.04) | 0.12 | 0.14 | 345 | 985 | 41.8 | −0.07 (−0.14, 0.00) | 0.05 | 0.05 |
| Burkina Faso | iLiNS−Zinc (37) | | | | | | | | | | | | |
| Burkina Faso | PROMIS (38) | 70 | 54 | 13.0 | 0.08 (−0.04, 0.21) | 0.01 | 0.02 | 787 | 857 | 20.1 | −0.04 (−0.07, 0.00) | 0.15 | 0.15 |
| Burkina Faso | PROMIS CS (38) | 34 | 39 | 28.2 | −0.02 (−0.24, 0.20) | 0.00 | 0.01 | 393 | 396 | 24.0 | 0.00 (−0.06, 0.06) | 0.06 | 0.06 |
| Ghana | GHANA (39) | | | | | | | | | | | | |
| Ghana | iLiNS−DYADG (40) | 267 | 549 | 12.4 | −0.01 (−0.06, 0.04) | 0.06 | 0.10 | 80 | 144 | 15.3 | −0.08 (−0.17, 0.01) | 0.03 | 0.03 |
| Haiti | HAITI (41) | | | | | | | | | | | | |
| Kenya | WASH−B (42) | 705 | 2451 | 6.7 | 0.00 (−0.03, 0.02) | 0.26 | 0.18 | 754 | 2710 | 13.1 | −0.04 (−0.07, −0.02) | 0.35 | 0.35 |
| Madagascar | MAHAY (43) | 352 | 431 | 21.1 | 0.01 (−0.05, 0.07) | 0.03 | 0.08 | 1350 | 1254 | 27.7 | 0.01 (−0.04, 0.05) | 0.10 | 0.10 |
| Malawi | iLiNS−DYADM (44) | | | | | | | | | | | | |
| Malawi | iLiNS−DOSE (45) | | | | | | | | | | | | |
| Mali | PROMIS (46) | | | | | | | | | | | | |
| Mali | PROMIS CS (46) | 107 | 96 | 19.8 | −0.09 (−0.20, 0.03) | 0.01 | 0.03 | 838 | 864 | 22.8 | −0.02 (−0.06, 0.03) | 0.10 | 0.10 |
| Zimbabwe | SHINE (HIV−) (47) | 1707 | 1636 | 10.5 | −0.03 (−0.05, −0.01) | 0.34 | 0.19 | 68 | 61 | 13.1 | −0.01 (−0.12, 0.10) | 0.02 | 0.02 |
| Zimbabwe | SHINE (HIV+) (48) | | | | | | | | | | | | |
| | | **7121** | **9070** | | **I² = 0.56, Tau² = 0.00** | | | **6029** | **7977** | | **I² = 0.00, Tau² = 0.00** | | |
| **Fixed** | | | | | **−0.03 (−0.04, −0.02)** | | | | | | **−0.03 (−0.05, −0.02)** | | |
| **Random** | | | | | **−0.03 (−0.05, −0.01)** | | | | | | **−0.03 (−0.05, −0.02)** | | |

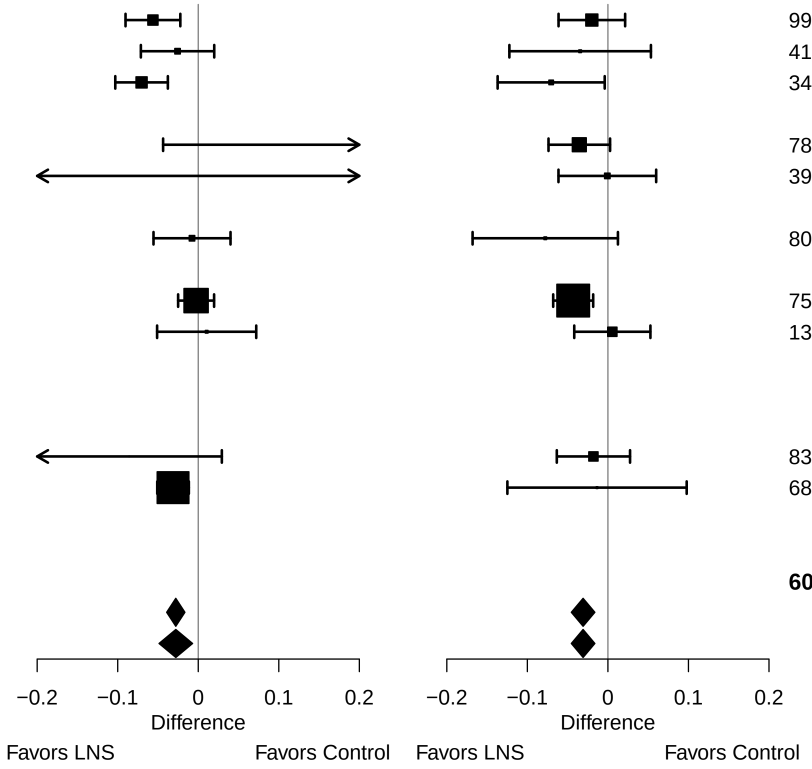

#### Supplemental figure 6N: Underweight prevalence difference

#### 6N5: Stratified by Maternal depressive symptoms

**-for-interaction = 0.717**
**ifference in PDs = 0.00 (−0.02, 0.03)**

| | | Less than 75th percentile | | | | | | At least 75th percentile | | | | | |
|---|---|---|---|---|---|---|---|---|---|---|---|---|---|
| **ountry** | **Trial** | **LNS N** | **Control N** | **Control Prevalence** | **PD (95% CI)** | **Fixed W** | **Random W** | **LNS N** | **Control N** | **Control Prevalence** | **PD (95% CI)** | **Fixed W** | **Random W** |
| angladesh | JiVitA−4 (34) | | | | | | | | | | | | |
| angladesh | RDNS (35) | 1081 | 472 | 36.9 | −0.03 (−0.07, 0.01) | 0.08 | 0.09 | 520 | 288 | 38.2 | −0.03 (−0.11, 0.05) | 0.05 | 0.07 |
| angladesh | WASH−B (36) | 899 | 2487 | 31.6 | −0.07 (−0.10, −0.03) | 0.10 | 0.11 | 240 | 883 | 34.7 | −0.08 (−0.15, 0.00) | 0.07 | 0.08 |
| urkina Faso | iLiNS−Zinc (37) | | | | | | | | | | | | |
| urkina Faso | PROMIS (38) | 505 | 571 | 19.4 | −0.04 (−0.09, 0.01) | 0.04 | 0.06 | 220 | 213 | 21.1 | −0.01 (−0.07, 0.05) | 0.11 | 0.12 |
| urkina Faso | PROMIS CS (38) | | | | | | | | | | | | |
| hana | GHANA (39) | | | | | | | | | | | | |
| hana | iLiNS−DYADG (40) | 252 | 460 | 12.4 | −0.04 (−0.08, 0.01) | 0.05 | 0.07 | 83 | 209 | 12.9 | 0.04 (−0.05, 0.13) | 0.05 | 0.06 |
| aiti | HAITI (41) | | | | | | | | | | | | |
| enya | WASH−B (42) | 997 | 3506 | 9.6 | −0.02 (−0.03, 0.00) | 0.39 | 0.29 | 342 | 1248 | 10.5 | −0.03 (−0.07, 0.01) | 0.25 | 0.21 |
| adagascar | MAHAY (43) | 673 | 721 | 27.2 | 0.00 (−0.06, 0.05) | 0.04 | 0.05 | 283 | 252 | 24.2 | 0.05 (−0.03, 0.14) | 0.05 | 0.07 |
| alawi | iLiNS−DYADM (44) | 157 | 303 | 11.2 | 0.00 (−0.06, 0.06) | 0.03 | 0.04 | 47 | 113 | 12.4 | 0.03 (−0.09, 0.14) | 0.03 | 0.04 |
| alawi | iLiNS−DOSE (45) | | | | | | | | | | | | |
| ali | PROMIS (46) | 374 | 372 | 15.8 | −0.04 (−0.10, 0.01) | 0.04 | 0.06 | 122 | 129 | 18.3 | −0.01 (−0.09, 0.06) | 0.06 | 0.08 |
| ali | PROMIS CS (46) | | | | | | | | | | | | |
| imbabwe | SHINE (HIV−) (47) | 1192 | 1137 | 9.9 | −0.02 (−0.05, 0.00) | 0.19 | 0.19 | 509 | 490 | 12.7 | −0.06 (−0.09, −0.02) | 0.30 | 0.23 |
| imbabwe | SHINE (HIV+) (48) | 234 | 214 | 16.4 | 0.02 (−0.05, 0.08) | 0.03 | 0.04 | 88 | 100 | 20.0 | −0.03 (−0.14, 0.08) | 0.03 | 0.04 |
| | | **6364** | **10243** | | **I² = 0.11, Tau² = 0.00** | | | **2454** | **3925** | | **I² = 0.25, Tau² = 0.00** | | |
| **ixed** | | | | | **−0.03 (−0.04, −0.01)** | | | | | | **−0.03 (−0.05, −0.01)** | | |
| **andom** | | | | | **−0.03 (−0.04, −0.01)** | | | | | | **−0.02 (−0.05, 0.00)** | | |

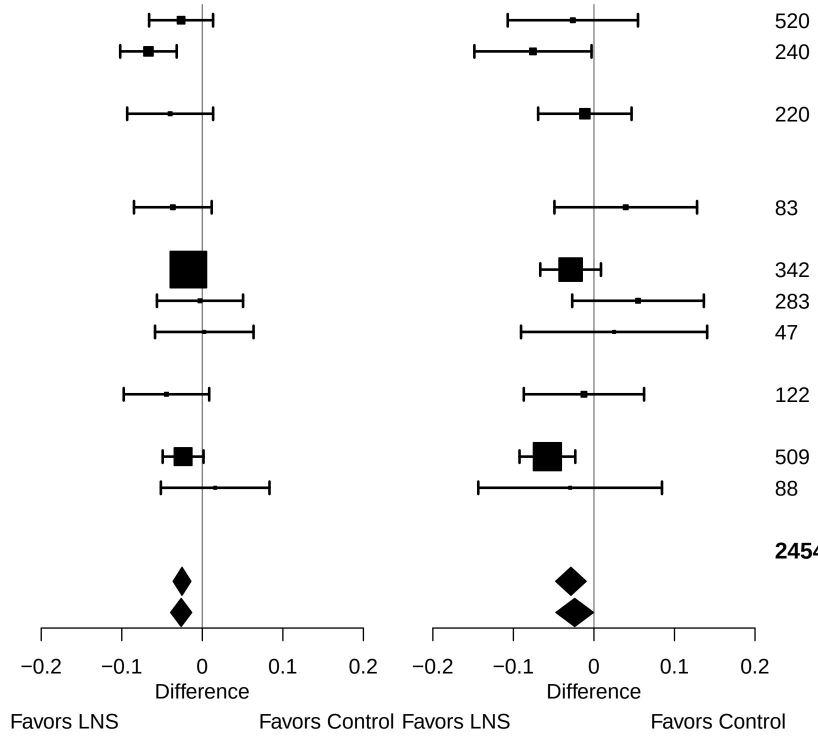

#### Supplemental figure 6N: Underweight prevalence difference

#### 6N6: Stratified by Child sex

**P−for−interaction = 0.231**
**Difference in PDs = 0.01 (−0.01, 0.03)**

| Country | Trial | Male LNS N | Male Control N | Male Control Prevalence | Male PD (95% CI) | Male Fixed W | Male Random W | Female LNS N | Female Control N | Female Control Prevalence | Female PD (95% CI) | Female Fixed W | Female Random W |
|---|---|---|---|---|---|---|---|---|---|---|---|---|---|
| Bangladesh | JiVitA−4 (34) | 1440 | 594 | 38.2 | −0.03 (−0.06, 0.01) | 0.11 | 0.11 | 1374 | 638 | 40.1 | −0.06 (−0.09, −0.02) | 0.09 | 0.08 |
| Bangladesh | RDNS (35) | 833 | 411 | 38.0 | −0.02 (−0.08, 0.04) | 0.04 | 0.04 | 830 | 404 | 37.6 | −0.04 (−0.09, 0.01) | 0.05 | 0.07 |
| Bangladesh | WASH−B (36) | 567 | 1730 | 33.2 | −0.07 (−0.11, −0.03) | 0.08 | 0.08 | 596 | 1731 | 31.7 | −0.06 (−0.11, −0.02) | 0.05 | 0.07 |
| Burkina Faso | iLiNS−Zinc (37) | 995 | 336 | 32.4 | −0.08 (−0.12, −0.03) | 0.06 | 0.06 | 965 | 330 | 29.4 | −0.12 (−0.17, −0.08) | 0.05 | 0.07 |
| Burkina Faso | PROMIS (38) | 435 | 482 | 24.9 | −0.05 (−0.10, 0.01) | 0.05 | 0.05 | 422 | 429 | 13.8 | 0.00 (−0.05, 0.05) | 0.04 | 0.06 |
| Burkina Faso | PROMIS CS (38) | 234 | 221 | 27.6 | 0.01 (−0.08, 0.10) | 0.02 | 0.02 | 196 | 215 | 20.9 | −0.03 (−0.10, 0.05) | 0.02 | 0.04 |
| Ghana | GHANA (39) | | | | | | | | | | | | |
| Ghana | iLiNS−DYADG (40) | 175 | 324 | 16.4 | −0.04 (−0.11, 0.02) | 0.03 | 0.03 | 172 | 369 | 10.0 | −0.01 (−0.06, 0.05) | 0.04 | 0.06 |
| Haiti | HAITI (41) | | | | | | | | | | | | |
| Kenya | WASH−B (42) | 734 | 2457 | 11.8 | −0.03 (−0.05, −0.01) | 0.24 | 0.24 | 727 | 2708 | 8.4 | −0.02 (−0.04, 0.00) | 0.31 | 0.10 |
| Madagascar | MAHAY (43) | 826 | 835 | 31.4 | 0.00 (−0.05, 0.05) | 0.05 | 0.05 | 876 | 850 | 20.7 | 0.02 (−0.02, 0.07) | 0.05 | 0.07 |
| Malawi | iLiNS−DYADM (44) | 107 | 207 | 12.6 | 0.05 (−0.03, 0.13) | 0.02 | 0.02 | 113 | 237 | 11.0 | −0.02 (−0.09, 0.05) | 0.02 | 0.05 |
| Malawi | iLiNS−DOSE (45) | 353 | 124 | 29.0 | −0.05 (−0.12, 0.03) | 0.03 | 0.03 | 349 | 117 | 10.3 | 0.02 (−0.05, 0.08) | 0.03 | 0.05 |
| Mali | PROMIS (46) | 265 | 262 | 18.6 | −0.04 (−0.10, 0.01) | 0.05 | 0.05 | 235 | 240 | 14.0 | −0.03 (−0.09, 0.03) | 0.03 | 0.06 |
| Mali | PROMIS CS (46) | 474 | 521 | 25.1 | −0.03 (−0.10, 0.04) | 0.03 | 0.03 | 472 | 439 | 19.4 | −0.02 (−0.07, 0.04) | 0.04 | 0.06 |
| Zimbabwe | SHINE (HIV−) (47) | 924 | 906 | 13.4 | −0.05 (−0.07, −0.02) | 0.18 | 0.18 | 945 | 880 | 7.7 | −0.01 (−0.04, 0.01) | 0.18 | 0.09 |
| Zimbabwe | SHINE (HIV+) (48) | 161 | 167 | 22.8 | −0.05 (−0.13, 0.03) | 0.02 | 0.02 | 175 | 161 | 11.2 | 0.05 (−0.02, 0.13) | 0.02 | 0.05 |
| | | **8523** | **9577** | | **I² = 0.01, Tau² = 0.00** | | | **8447** | **9748** | | **I² = 0.63, Tau² = 0.00** | | |
| **Fixed** | | | | | **−0.04 (−0.05, −0.03)** | | | | | | **−0.03 (−0.04, −0.01)** | | |
| **Random** | | | | | **−0.04 (−0.05, −0.03)** | | | | | | **−0.02 (−0.05, 0.00)** | | |

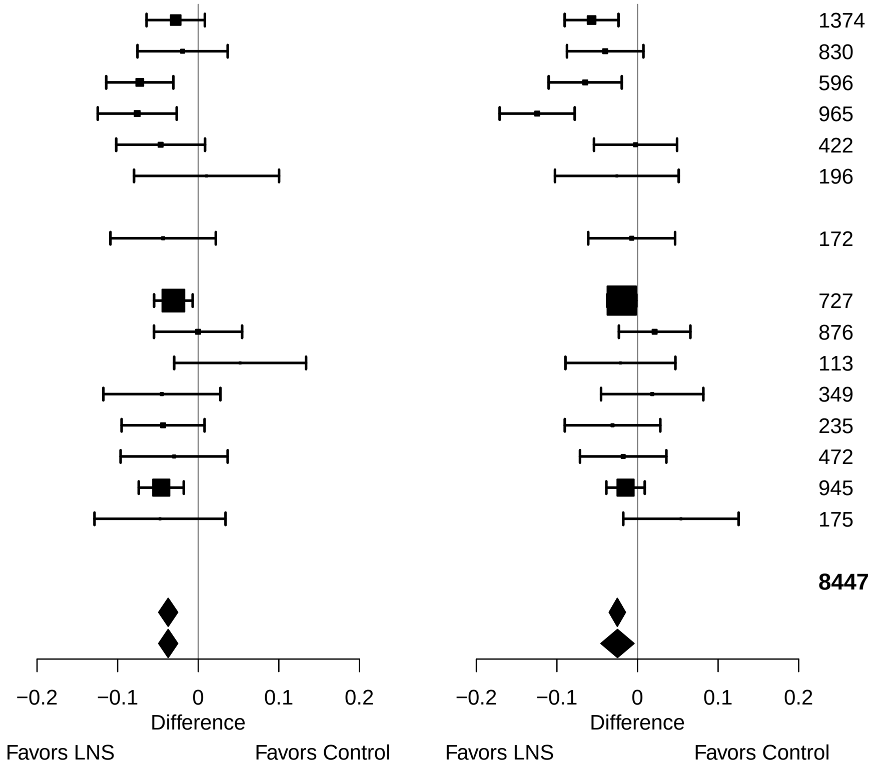

#### Supplemental figure 6N: Underweight prevalence difference

### 6N7: Stratified by Child birth order

**P−for−interaction = 0.043**
**Difference in PDs = 0.02 (0.00, 0.04)**

| | | Later born | | | | | | Firstborn | | | | | |
|---|---|---|---|---|---|---|---|---|---|---|---|---|---|
| **Country** | **Trial** | **LNS N** | **Control N** | **Control Prevalence** | **PD (95% CI)** | **Fixed W** | **Random W** | **LNS N** | **Control N** | **Control Prevalence** | **PD (95% CI)** | **Fixed W** | **Random W** |
| Bangladesh | JiVitA−4 (34) | 600 | 250 | 43.3 | −0.07 (−0.12, −0.02) | 0.04 | 0.06 | 2210 | 976 | 38.0 | −0.04 (−0.06, −0.01) | 0.26 | 0.18 |
| Bangladesh | RDNS (35) | 974 | 508 | 38.0 | −0.03 (−0.09, 0.02) | 0.04 | 0.06 | 688 | 307 | 37.5 | −0.02 (−0.07, 0.03) | 0.08 | 0.10 |
| Bangladesh | WASH−B (36) | 737 | 2310 | 34.3 | −0.08 (−0.12, −0.04) | 0.06 | 0.07 | 426 | 1151 | 28.6 | −0.05 (−0.10, 0.00) | 0.08 | 0.09 |
| Burkina Faso | iLiNS−Zinc (37) | 1524 | 521 | 30.9 | −0.11 (−0.15, −0.08) | 0.07 | 0.08 | 436 | 145 | 31.0 | −0.06 (−0.13, 0.02) | 0.04 | 0.05 |
| Burkina Faso | PROMIS (38) | 709 | 750 | 19.9 | −0.04 (−0.08, 0.00) | 0.07 | 0.08 | 148 | 159 | 18.9 | 0.03 (−0.07, 0.12) | 0.02 | 0.03 |
| Burkina Faso | PROMIS CS (38) | 366 | 357 | 23.8 | 0.00 (−0.07, 0.06) | 0.02 | 0.05 | 63 | 79 | 26.6 | 0.00 (−0.13, 0.14) | 0.01 | 0.02 |
| Ghana | GHANA (39) | | | | | | | | | | | | |
| Ghana | iLiNS−DYADG (40) | 237 | 459 | 12.2 | 0.00 (−0.06, 0.05) | 0.04 | 0.06 | 110 | 234 | 14.5 | −0.06 (−0.14, 0.01) | 0.04 | 0.05 |
| Haiti | HAITI (41) | | | | | | | | | | | | |
| Kenya | WASH−B (42) | 1137 | 4065 | 10.2 | −0.02 (−0.04, −0.01) | 0.28 | 0.10 | 322 | 1095 | 9.2 | −0.02 (−0.05, 0.01) | 0.20 | 0.16 |
| Madagascar | MAHAY (43) | 1271 | 1220 | 27.2 | −0.01 (−0.05, 0.04) | 0.05 | 0.07 | 431 | 465 | 22.8 | 0.05 (−0.02, 0.12) | 0.04 | 0.06 |
| Malawi | iLiNS−DYADM (44) | 173 | 353 | 11.3 | 0.02 (−0.04, 0.08) | 0.03 | 0.05 | 47 | 89 | 12.4 | 0.00 (−0.11, 0.12) | 0.01 | 0.02 |
| Malawi | iLiNS−DOSE (45) | 463 | 165 | 19.4 | −0.01 (−0.07, 0.05) | 0.03 | 0.05 | 150 | 45 | 22.2 | −0.07 (−0.19, 0.04) | 0.02 | 0.02 |
| Mali | PROMIS (46) | 437 | 441 | 16.9 | −0.05 (−0.09, 0.00) | 0.05 | 0.07 | 62 | 59 | 11.9 | 0.06 (−0.05, 0.16) | 0.02 | 0.03 |
| Mali | PROMIS CS (46) | 784 | 822 | 22.5 | −0.03 (−0.08, 0.01) | 0.05 | 0.07 | 145 | 126 | 20.6 | 0.01 (−0.08, 0.10) | 0.02 | 0.04 |
| Zimbabwe | SHINE (HIV−) (47) | 1295 | 1220 | 10.4 | −0.04 (−0.07, −0.02) | 0.16 | 0.09 | 487 | 483 | 11.2 | −0.01 (−0.05, 0.03) | 0.14 | 0.13 |
| Zimbabwe | SHINE (HIV+) (48) | 279 | 245 | 14.3 | 0.04 (−0.03, 0.10) | 0.02 | 0.05 | 54 | 78 | 25.6 | −0.13 (−0.24, −0.01) | 0.02 | 0.02 |
| | | **10986** | **13686** | | **I² = 0.62, Tau² = 0.00** | | | **5779** | **5491** | | **I² = 0.19, Tau² = 0.00** | | |
| **Fixed** | | | | | **−0.04 (−0.05, −0.03)** | | | | | | **−0.02 (−0.04, −0.01)** | | |
| **Random** | | | | | **−0.03 (−0.05, −0.01)** | | | | | | **−0.02 (−0.04, 0.00)** | | |

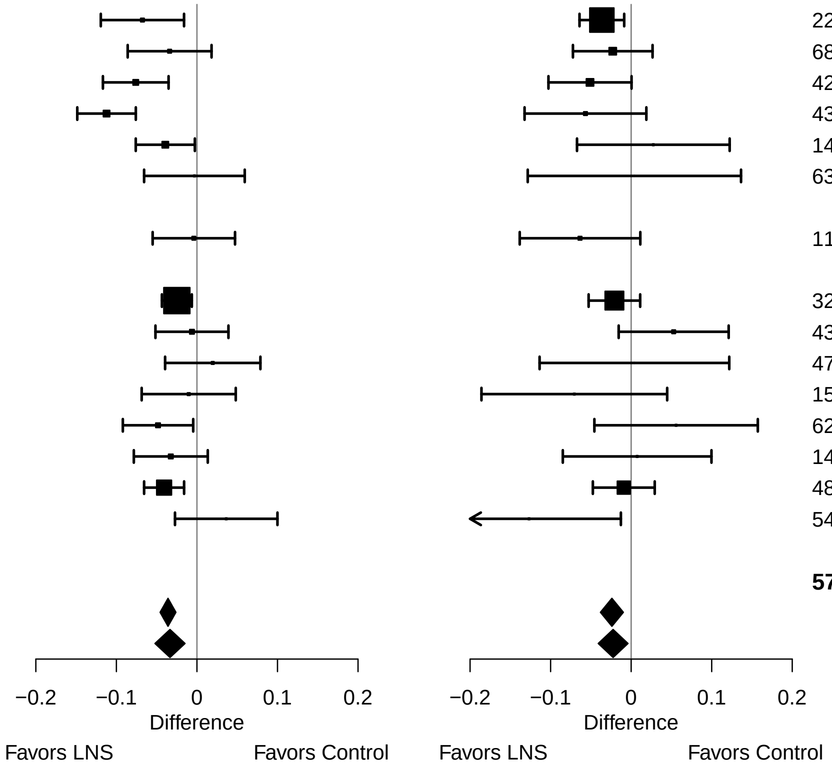

#### Supplemental figure 6N: Underweight prevalence difference

#### 6N8: Stratified by Child baseline anthropometric status

**P−for−interaction = 0.004**
**Difference in PDs = −0.04 (−0.07, −0.01)**

| | | WAZ at least −1 Z | | | | | | WAZ less than −1 Z | | | | | |
|---|---|---|---|---|---|---|---|---|---|---|---|---|---|
| **Country** | **Trial** | **LNS N** | **Control N** | **Control Prevalence** | **PD (95% CI)** | **Fixed W** | **Random W** | **LNS N** | **Control N** | **Control Prevalence** | **PD (95% CI)** | **Fixed W** | **Random W** |
| Bangladesh | JiVitA−4 (34) | 1198 | 531 | 6.4 | −0.02 (−0.05, 0.00) | 0.13 | 0.13 | 1616 | 701 | 64.3 | −0.07 (−0.11, −0.03) | 0.35 | 0.35 |
| Bangladesh | RDNS (35) | 780 | 338 | 5.6 | 0.01 (−0.02, 0.03) | 0.09 | 0.09 | 821 | 422 | 62.8 | −0.01 (−0.06, 0.04) | 0.23 | 0.23 |
| Bangladesh | WASH−B (36) | | | | | | | | | | | | |
| Burkina Faso | iLiNS−Zinc (37) | | | | | | | | | | | | |
| Burkina Faso | PROMIS (38) | 427 | 482 | 5.8 | −0.01 (−0.04, 0.01) | 0.10 | 0.10 | 268 | 267 | 42.7 | −0.05 (−0.13, 0.02) | 0.11 | 0.11 |
| Burkina Faso | PROMIS CS (38) | | | | | | | | | | | | |
| Ghana | GHANA (39) | | | | | | | | | | | | |
| Ghana | iLiNS−DYADG (40) | 222 | 424 | 1.7 | 0.01 (−0.02, 0.03) | 0.15 | 0.15 | 101 | 215 | 34.0 | −0.02 (−0.13, 0.09) | 0.05 | 0.05 |
| Haiti | HAITI (41) | | | | | | | | | | | | |
| Kenya | WASH−B (42) | | | | | | | | | | | | |
| Madagascar | MAHAY (43) | | | | | | | | | | | | |
| Malawi | iLiNS−DYADM (44) | | | | | | | | | | | | |
| Malawi | iLiNS−DOSE (45) | 419 | 143 | 3.5 | 0.00 (−0.04, 0.03) | 0.06 | 0.06 | 283 | 98 | 43.9 | −0.02 (−0.13, 0.10) | 0.05 | 0.05 |
| Mali | PROMIS (46) | | | | | | | | | | | | |
| Mali | PROMIS CS (46) | | | | | | | | | | | | |
| Zimbabwe | SHINE (HIV−) (47) | 917 | 836 | 1.9 | 0.00 (−0.02, 0.01) | 0.45 | 0.45 | 383 | 349 | 30.1 | −0.09 (−0.15, −0.03) | 0.18 | 0.18 |
| Zimbabwe | SHINE (HIV+) (48) | 151 | 144 | 5.6 | −0.01 (−0.06, 0.04) | 0.03 | 0.03 | 113 | 92 | 32.6 | 0.01 (−0.12, 0.14) | 0.04 | 0.04 |
| | | **4114** | **2898** | | **I² = 0.00, Tau² = 0.00** | | | **3585** | **2144** | | **I² = 0.00, Tau² = 0.00** | | |
| **Fixed** | | | | | **0.00 (−0.01, 0.00)** | | | | | | **−0.05 (−0.08, −0.03)** | | |
| **Random** | | | | | **0.00 (−0.01, 0.00)** | | | | | | **−0.05 (−0.08, −0.03)** | | |

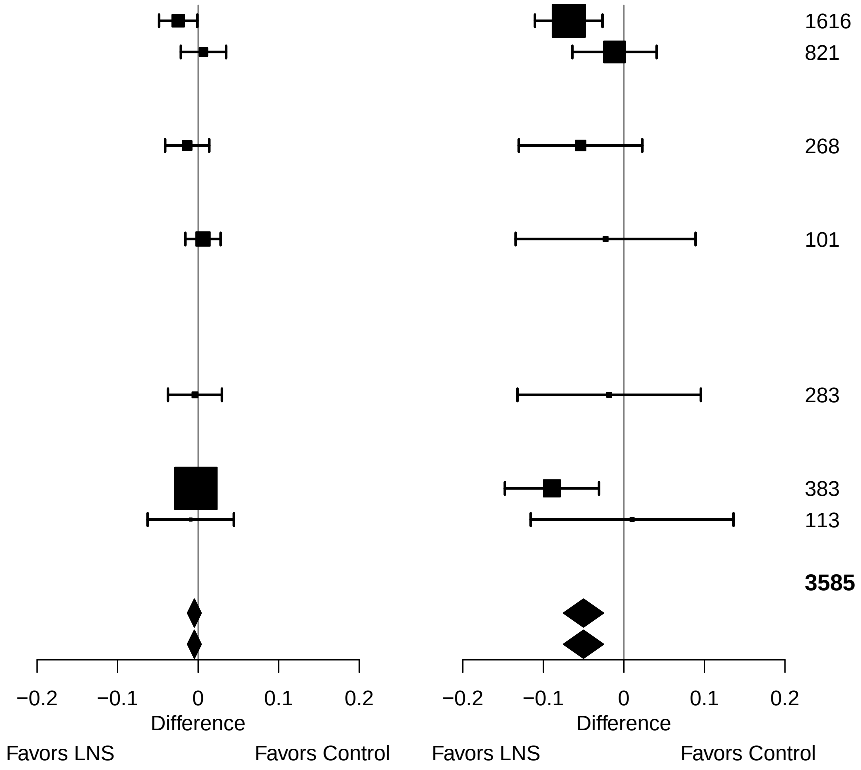

### Supplemental figure 6O: Mean difference in HCZ

#### 6O1: Stratified by Maternal height

**P−for−interaction = 0.185**
**Difference in MDs = 0.05 (−0.03, 0.13)**

| | | At least 150.1 cm | | | | | | Less than 150.1 cm | | | | | |
|---|---|---|---|---|---|---|---|---|---|---|---|---|---|
| **Country** | **Trial** | **LNS N** | **Control N** | **Control Mean** | **MD (95% CI)** | **Fixed W** | **Random W** | **LNS N** | **Control N** | **Control Mean** | **MD (95% CI)** | **Fixed W** | **Random W** |
| Bangladesh | JiVitA−4 (34) | | | | | | | | | | | | |
| Bangladesh | RDNS (35) | 868 | 432 | −1.74 | 0.09 (−0.01, 0.18) | 0.09 | 0.11 | 735 | 358 | −2.01 | 0.13 (0.03, 0.23) | 0.37 | 0.20 |
| Bangladesh | WASH−B (36) | 619 | 1899 | −1.48 | 0.09 (0.00, 0.17) | 0.12 | 0.13 | 544 | 1559 | −1.78 | 0.17 (0.09, 0.26) | 0.45 | 0.20 |
| Burkina Faso | iLiNS−Zinc (37) | 1911 | 649 | −1.83 | 0.16 (0.10, 0.23) | 0.22 | 0.18 | 33 | 15 | −2.10 | 0.23 (−0.07, 0.53) | 0.04 | 0.12 |
| Burkina Faso | PROMIS (38) | | | | | | | | | | | | |
| Burkina Faso | PROMIS CS (38) | | | | | | | | | | | | |
| Ghana | GHANA (39) | | | | | | | | | | | | |
| Ghana | iLiNS−DYADG (40) | 326 | 645 | −1.13 | 0.07 (−0.05, 0.18) | 0.06 | 0.08 | 16 | 36 | −1.44 | 0.22 (−0.35, 0.78) | 0.01 | 0.05 |
| Haiti | HAITI (41) | | | | | | | | | | | | |
| Kenya | WASH−B (42) | 1316 | 4720 | −0.26 | 0.04 (−0.02, 0.10) | 0.24 | 0.19 | 57 | 194 | −0.52 | −0.02 (−0.36, 0.32) | 0.03 | 0.10 |
| Madagascar | MAHAY (43) | | | | | | | | | | | | |
| Malawi | iLiNS−DYADM (44) | 192 | 381 | −0.75 | −0.09 (−0.27, 0.08) | 0.03 | 0.04 | 28 | 60 | −0.91 | −0.41 (−0.78, −0.04) | 0.03 | 0.09 |
| Malawi | iLiNS−DOSE (45) | 581 | 195 | −1.03 | 0.07 (−0.07, 0.20) | 0.05 | 0.07 | 111 | 42 | −1.60 | 0.29 (−0.03, 0.60) | 0.04 | 0.11 |
| Mali | PROMIS (46) | | | | | | | | | | | | |
| Mali | PROMIS CS (46) | | | | | | | | | | | | |
| Zimbabwe | SHINE (HIV−) (47) | 1746 | 1687 | −0.26 | 0.12 (0.04, 0.19) | 0.16 | 0.15 | 76 | 62 | −0.65 | −0.20 (−0.59, 0.20) | 0.02 | 0.09 |
| Zimbabwe | SHINE (HIV+) (48) | 314 | 312 | −0.52 | 0.10 (−0.09, 0.28) | 0.03 | 0.04 | 18 | 12 | −1.28 | 0.53 (−0.27, 1.33) | 0.01 | 0.03 |
| | | **7873** | **10920** | | **I² = 0.39, Tau² = 0.00** | | | **1618** | **2338** | | **I² = 0.46, Tau² = 0.03** | | |
| **Fixed** | | | | | **0.09 (0.06, 0.12)** | | | | | | **0.14 (0.08, 0.20)** | | |
| **Random** | | | | | **0.08 (0.05, 0.12)** | | | | | | **0.09 (−0.06, 0.24)** | | |

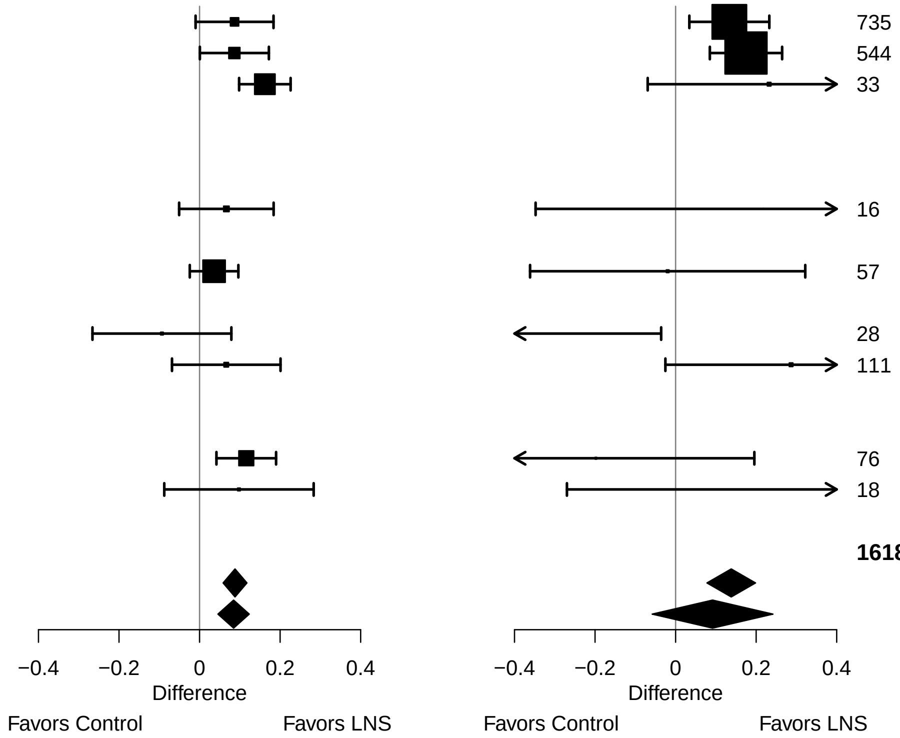

#### Supplemental figure 6O: Mean difference in HCZ

##### 6O2: Stratified by Maternal BMI

**P−for−interaction = 0.407**
**Difference in MDs = 0.02 (−0.03, 0.07)**

| | | At least 20 kg/m² | | | | | | Less than 20 kg/m² | | | | | |
|---|---|---|---|---|---|---|---|---|---|---|---|---|---|
| **Country** | **Trial** | **LNS N** | **Control N** | **Control Mean** | **MD (95% CI)** | **Fixed W** | **Random W** | **LNS N** | **Control N** | **Control Mean** | **MD (95% CI)** | **Fixed W** | **Random W** |
| Bangladesh | JiVitA−4 (34) | | | | | | | | | | | | |
| Bangladesh | RDNS (35) | 730 | 335 | −1.76 | 0.09 (−0.02, 0.20) | 0.09 | 0.12 | 873 | 455 | −1.94 | 0.11 (0.03, 0.19) | 0.24 | 0.24 |
| Bangladesh | WASH−B (36) | 519 | 1595 | −1.50 | 0.09 (−0.01, 0.19) | 0.10 | 0.12 | 644 | 1863 | −1.70 | 0.15 (0.08, 0.22) | 0.31 | 0.31 |
| Burkina Faso | iLiNS−Zinc (37) | 1210 | 389 | −1.81 | 0.17 (0.11, 0.23) | 0.29 | 0.16 | 734 | 275 | −1.88 | 0.16 (0.07, 0.24) | 0.24 | 0.24 |
| Burkina Faso | PROMIS (38) | | | | | | | | | | | | |
| Burkina Faso | PROMIS CS (38) | | | | | | | | | | | | |
| Ghana | GHANA (39) | | | | | | | | | | | | |
| Ghana | iLiNS−DYADG (40) | 305 | 562 | −1.12 | 0.03 (−0.09, 0.16) | 0.07 | 0.11 | 37 | 119 | −1.26 | 0.34 (0.01, 0.67) | 0.02 | 0.02 |
| Haiti | HAITI (41) | | | | | | | | | | | | |
| Kenya | WASH−B (42) | 1100 | 3831 | −0.22 | 0.02 (−0.05, 0.09) | 0.21 | 0.15 | 273 | 1083 | −0.46 | 0.07 (−0.08, 0.21) | 0.08 | 0.08 |
| Madagascar | MAHAY (43) | | | | | | | | | | | | |
| Malawi | iLiNS−DYADM (44) | 131 | 263 | −0.67 | −0.21 (−0.41, −0.01) | 0.03 | 0.06 | 88 | 178 | −0.92 | 0.01 (−0.26, 0.27) | 0.02 | 0.02 |
| Malawi | iLiNS−DOSE (45) | 512 | 172 | −1.03 | 0.10 (−0.04, 0.24) | 0.05 | 0.09 | 178 | 64 | −1.41 | 0.14 (−0.10, 0.39) | 0.03 | 0.03 |
| Mali | PROMIS (46) | | | | | | | | | | | | |
| Mali | PROMIS CS (46) | | | | | | | | | | | | |
| Zimbabwe | SHINE (HIV−) (47) | 1358 | 1298 | −0.25 | 0.09 (0.01, 0.18) | 0.15 | 0.14 | 239 | 248 | −0.44 | 0.08 (−0.11, 0.26) | 0.05 | 0.05 |
| Zimbabwe | SHINE (HIV+) (48) | 247 | 254 | −0.53 | 0.16 (−0.06, 0.38) | 0.02 | 0.05 | 62 | 52 | −0.60 | −0.05 (−0.38, 0.28) | 0.02 | 0.02 |
| | | **6112** | **8699** | | **I² = 0.62, Tau² = 0.01** | | | **3128** | **4337** | | **I² = 0.00, Tau² = 0.00** | | |
| **Fixed** | | | | | **0.09 (0.06, 0.12)** | | | | | | **0.13 (0.09, 0.17)** | | |
| **Random** | | | | | **0.07 (0.01, 0.13)** | | | | | | **0.13 (0.09, 0.17)** | | |

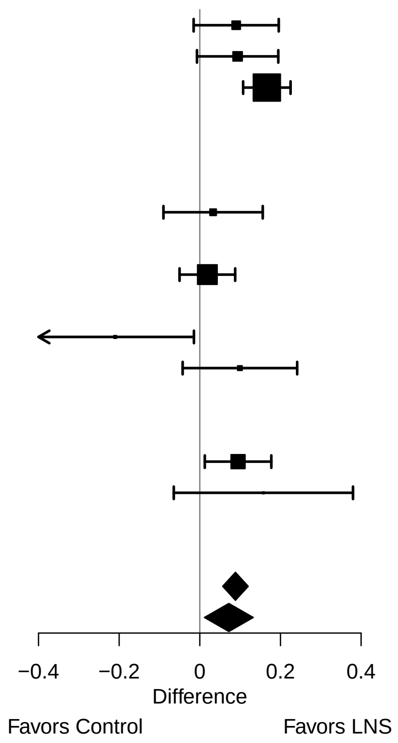

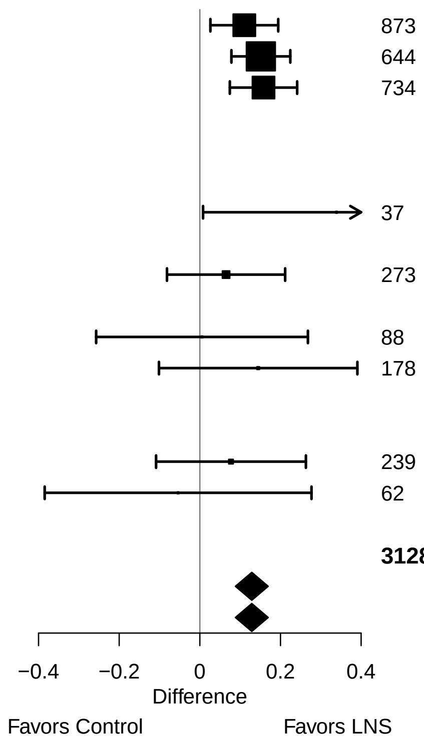

#### Supplemental figure 6O: Mean difference in HCZ

#### 6O3: Stratified by Maternal age

**P−for−interaction = 0.901**
**Difference in MDs = 0.00 (−0.04, 0.04)**

| | | At least 25 y | | | | | | Less than 25 y | | | | | |
|---|---|---|---|---|---|---|---|---|---|---|---|---|---|
| Country | Trial | LNS N | Control N | Control Mean | MD (95% CI) | Fixed W | Random W | LNS N | Control N | Control Mean | MD (95% CI) | Fixed W | Random W |
| Bangladesh | JiVitA−4 (34) | 1238 | 554 | −1.43 | 0.06 (−0.01, 0.12) | 0.27 | 0.27 | 1634 | 708 | −1.45 | 0.06 (0.00, 0.12) | 0.25 | 0.15 |
| Bangladesh | RDNS (35) | 453 | 221 | −1.83 | 0.13 (0.00, 0.26) | 0.06 | 0.06 | 1210 | 594 | −1.88 | 0.09 (0.00, 0.18) | 0.11 | 0.13 |
| Bangladesh | WASH−B (36) | 492 | 1523 | −1.60 | 0.09 (−0.01, 0.19) | 0.10 | 0.10 | 666 | 1924 | −1.63 | 0.15 (0.06, 0.23) | 0.10 | 0.12 |
| Burkina Faso | iLiNS−Zinc (37) | 1128 | 399 | −1.78 | 0.16 (0.07, 0.24) | 0.15 | 0.15 | 805 | 262 | −1.92 | 0.17 (0.12, 0.23) | 0.28 | 0.15 |
| Burkina Faso | PROMIS (38) | | | | | | | | | | | | |
| Burkina Faso | PROMIS CS (38) | | | | | | | | | | | | |
| Ghana | GHANA (39) | 69 | 63 | −0.67 | 0.29 (−0.03, 0.61) | 0.01 | 0.01 | 29 | 25 | −0.38 | −0.09 (−0.58, 0.41) | 0.00 | 0.01 |
| Ghana | iLiNS−DYADG (40) | 220 | 417 | −1.12 | 0.02 (−0.12, 0.16) | 0.05 | 0.05 | 127 | 275 | −1.18 | 0.15 (−0.05, 0.34) | 0.02 | 0.06 |
| Haiti | HAITI (41) | | | | | | | | | | | | |
| Kenya | WASH−B (42) | 845 | 2831 | −0.23 | 0.06 (−0.02, 0.13) | 0.18 | 0.18 | 601 | 2313 | −0.35 | 0.02 (−0.06, 0.11) | 0.11 | 0.12 |
| Madagascar | MAHAY (43) | | | | | | | | | | | | |
| Malawi | iLiNS−DYADM (44) | 106 | 226 | −0.72 | 0.03 (−0.20, 0.25) | 0.02 | 0.02 | 114 | 218 | −0.83 | −0.26 (−0.49, −0.04) | 0.02 | 0.05 |
| Malawi | iLiNS−DOSE (45) | 368 | 134 | −1.11 | 0.05 (−0.12, 0.22) | 0.04 | 0.04 | 310 | 97 | −1.16 | 0.19 (0.01, 0.38) | 0.02 | 0.07 |
| Mali | PROMIS (46) | | | | | | | | | | | | |
| Mali | PROMIS CS (46) | | | | | | | | | | | | |
| Zimbabwe | SHINE (HIV−) (47) | 869 | 795 | −0.19 | 0.10 (−0.01, 0.21) | 0.09 | 0.09 | 805 | 792 | −0.36 | 0.09 (−0.02, 0.19) | 0.08 | 0.11 |
| Zimbabwe | SHINE (HIV+) (48) | 244 | 231 | −0.49 | 0.05 (−0.16, 0.26) | 0.02 | 0.02 | 73 | 78 | −0.69 | 0.10 (−0.23, 0.44) | 0.01 | 0.03 |
| | | **6032** | **7394** | | **I² = 0.00, Tau² = 0.00** | | | **6374** | **7286** | | **I² = 0.59, Tau² = 0.01** | | |
| **Fixed** | | | | | **0.08 (0.05, 0.12)** | | | | | | **0.10 (0.07, 0.13)** | | |
| **Random** | | | | | **0.08 (0.05, 0.12)** | | | | | | **0.09 (0.02, 0.15)** | | |

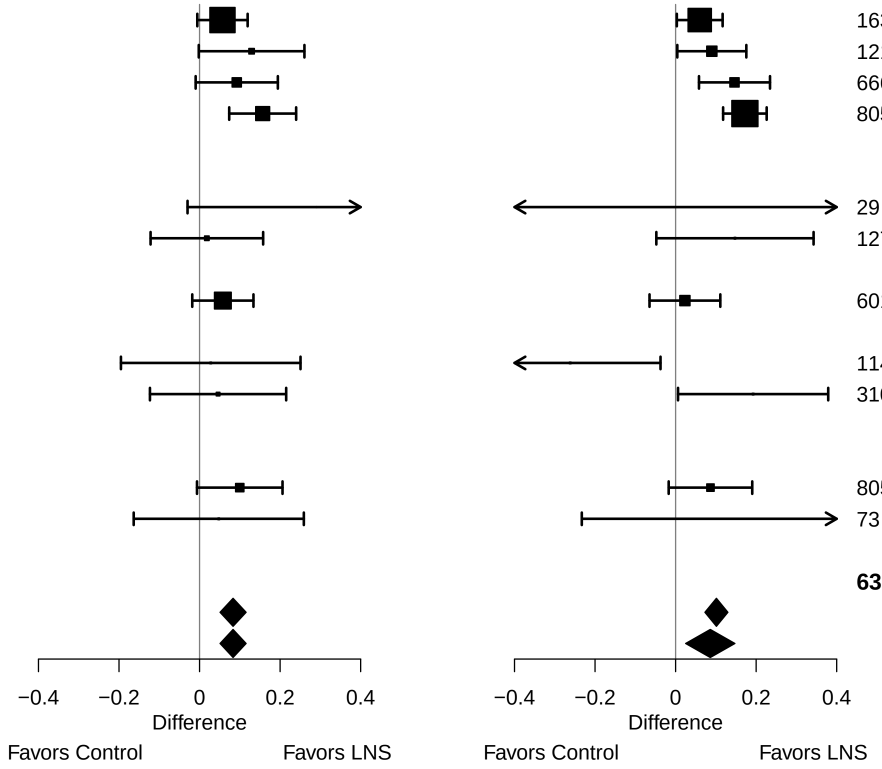

##### Supplemental figure 6O: Mean difference in HCZ

#### 6O4: Stratified by Maternal education

**P−for−interaction = 0.711**
**Difference in MDs = 0.01 (−0.04, 0.06)**

| | | Primary or greater | | | | | | Incomplete or no formal | | | | | |
|---|---|---|---|---|---|---|---|---|---|---|---|---|---|
| Country | Trial | LNS N | Control N | Control Mean | MD (95% CI) | Fixed W | Random W | LNS N | Control N | Control Mean | MD (95% CI) | Fixed W | Random W |
| Bangladesh | JiVitA−4 (34) | 1855 | 775 | −1.38 | 0.08 (0.02, 0.13) | 0.29 | 0.12 | 1028 | 489 | −1.54 | 0.03 (−0.04, 0.09) | 0.25 | 0.17 |
| Bangladesh | RDNS (35) | 1245 | 589 | −1.85 | 0.12 (0.04, 0.20) | 0.14 | 0.11 | 418 | 226 | −1.89 | 0.04 (−0.10, 0.18) | 0.06 | 0.10 |
| Bangladesh | WASH−B (36) | 817 | 2477 | −1.55 | 0.13 (0.05, 0.20) | 0.16 | 0.11 | 346 | 986 | −1.76 | 0.12 (0.00, 0.24) | 0.08 | 0.12 |
| Burkina Faso | iLiNS−Zinc (37) | 82 | 16 | −1.65 | 0.03 (−0.16, 0.22) | 0.03 | 0.08 | 1851 | 645 | −1.84 | 0.17 (0.11, 0.23) | 0.31 | 0.18 |
| Burkina Faso | PROMIS (38) | | | | | | | | | | | | |
| Burkina Faso | PROMIS CS (38) | | | | | | | | | | | | |
| Ghana | GHANA (39) | 91 | 75 | −0.64 | 0.29 (0.01, 0.57) | 0.01 | 0.06 | 7 | 15 | −0.31 | −0.81 (−1.71, 0.10) | 0.00 | 0.00 |
| Ghana | iLiNS−DYADG (40) | 267 | 548 | −1.12 | 0.05 (−0.08, 0.18) | 0.06 | 0.10 | 80 | 144 | −1.22 | 0.13 (−0.11, 0.37) | 0.02 | 0.05 |
| Haiti | HAITI (41) | | | | | | | | | | | | |
| Kenya | WASH−B (42) | 703 | 2456 | −0.15 | −0.05 (−0.14, 0.04) | 0.12 | 0.11 | 756 | 2716 | −0.40 | 0.12 (0.05, 0.20) | 0.18 | 0.16 |
| Madagascar | MAHAY (43) | | | | | | | | | | | | |
| Malawi | iLiNS−DYADM (44) | 35 | 69 | −0.56 | −0.55 (−0.91, −0.18) | 0.01 | 0.04 | 184 | 372 | −0.81 | −0.04 (−0.22, 0.13) | 0.04 | 0.08 |
| Malawi | iLiNS−DOSE (45) | 163 | 55 | −1.02 | 0.25 (−0.01, 0.50) | 0.01 | 0.07 | 517 | 176 | −1.16 | 0.07 (−0.07, 0.21) | 0.06 | 0.10 |
| Mali | PROMIS (46) | | | | | | | | | | | | |
| Mali | PROMIS CS (46) | | | | | | | | | | | | |
| Zimbabwe | SHINE (HIV−) (47) | 1708 | 1633 | −0.28 | 0.11 (0.03, 0.18) | 0.16 | 0.11 | 69 | 61 | −0.32 | −0.15 (−0.52, 0.23) | 0.01 | 0.02 |
| Zimbabwe | SHINE (HIV+) (48) | 301 | 293 | −0.51 | 0.06 (−0.13, 0.25) | 0.03 | 0.08 | 21 | 18 | −1.17 | 0.47 (−0.20, 1.14) | 0.00 | 0.01 |
| | | **7267** | **8986** | | **I² = 0.62, Tau² = 0.02** | | | **5277** | **5848** | | **I² = 0.51, Tau² = 0.00** | | |
| **Fixed** | | | | | **0.08 (0.05, 0.11)** | | | | | | **0.10 (0.06, 0.13)** | | |
| **Random** | | | | | **0.07 (−0.03, 0.16)** | | | | | | **0.08 (0.02, 0.14)** | | |

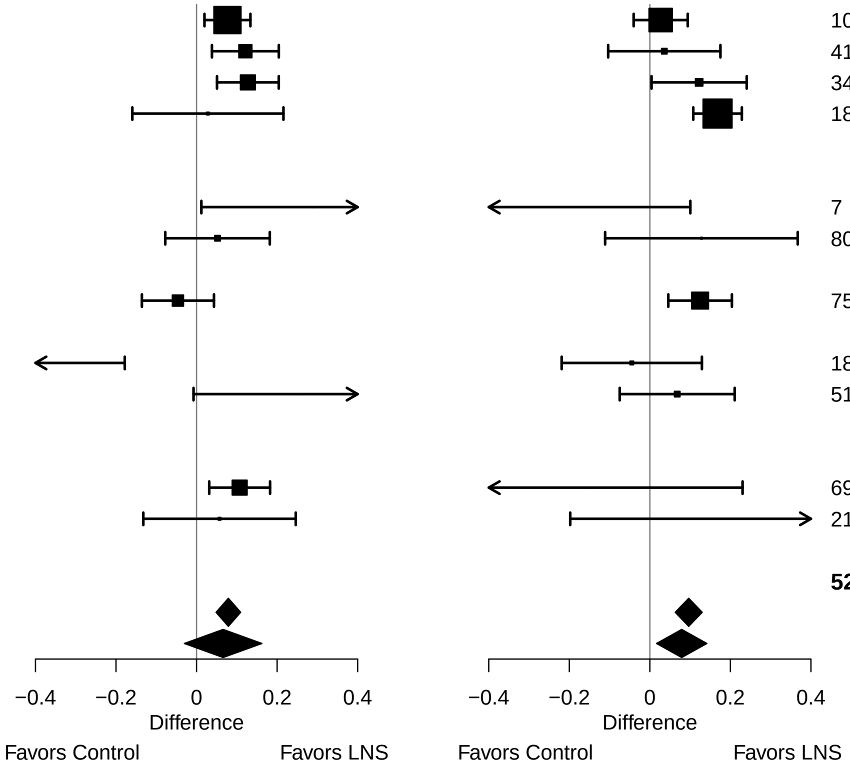

#### Supplemental figure 6O: Mean difference in HCZ

##### 6O5: Stratified by Maternal depressive symptoms

**P−for−interaction = 0.386**

**Difference in MDs = −0.03 (−0.10, 0.04)**

| | | Less than 75th percentile | | | | | | At least 75th percentile | | | | | |
|---|---|---|---|---|---|---|---|---|---|---|---|---|---|
| **Country** | **Trial** | **LNS N** | **Control N** | **Control Mean** | **MD (95% CI)** | **Fixed W** | **Random W** | **LNS N** | **Control N** | **Control Mean** | **MD (95% CI)** | **Fixed W** | **Random W** |
| Bangladesh | JiVitA−4 (34) | | | | | | | | | | | | |
| Bangladesh | RDNS (35) | 1081 | 472 | −1.81 | 0.04 (−0.03, 0.12) | 0.22 | 0.20 | 520 | 288 | −1.95 | 0.18 (0.04, 0.31) | 0.19 | 0.18 |
| Bangladesh | WASH−B (36) | 901 | 2487 | −1.60 | 0.13 (0.06, 0.20) | 0.27 | 0.22 | 238 | 885 | −1.61 | 0.09 (−0.04, 0.23) | 0.20 | 0.19 |
| Burkina Faso | iLiNS−Zinc (37) | | | | | | | | | | | | |
| Burkina Faso | PROMIS (38) | | | | | | | | | | | | |
| Burkina Faso | PROMIS CS (38) | | | | | | | | | | | | |
| Ghana | GHANA (39) | | | | | | | | | | | | |
| Ghana | iLiNS−DYADG (40) | 252 | 459 | −1.13 | 0.07 (−0.07, 0.20) | 0.07 | 0.10 | 83 | 209 | −1.15 | 0.02 (−0.20, 0.24) | 0.08 | 0.11 |
| Haiti | HAITI (41) | | | | | | | | | | | | |
| Kenya | WASH−B (42) | 998 | 3517 | −0.27 | 0.05 (−0.02, 0.13) | 0.23 | 0.21 | 341 | 1248 | −0.29 | −0.04 (−0.15, 0.08) | 0.27 | 0.21 |
| Madagascar | MAHAY (43) | | | | | | | | | | | | |
| Malawi | iLiNS−DYADM (44) | 157 | 303 | −0.74 | −0.15 (−0.33, 0.04) | 0.04 | 0.06 | 47 | 113 | −0.84 | −0.07 (−0.41, 0.28) | 0.03 | 0.06 |
| Malawi | iLiNS−DOSE (45) | | | | | | | | | | | | |
| Mali | PROMIS (46) | | | | | | | | | | | | |
| Mali | PROMIS CS (46) | | | | | | | | | | | | |
| Zimbabwe | SHINE (HIV−) (47) | 1193 | 1135 | −0.24 | 0.06 (−0.03, 0.15) | 0.15 | 0.16 | 509 | 490 | −0.35 | 0.19 (0.05, 0.32) | 0.19 | 0.18 |
| Zimbabwe | SHINE (HIV+) (48) | 234 | 215 | −0.52 | 0.01 (−0.20, 0.23) | 0.03 | 0.05 | 89 | 100 | −0.64 | 0.34 (0.03, 0.66) | 0.04 | 0.06 |
| | | **4816** | **8588** | | **I² = 0.36, Tau² = 0.00** | | | **1827** | **3333** | | **I² = 0.49, Tau² = 0.01** | | |
| **Fixed** | | | | | **0.07 (0.03, 0.10)** | | | | | | **0.09 (0.03, 0.15)** | | |
| **Random** | | | | | **0.06 (0.01, 0.11)** | | | | | | **0.10 (0.01, 0.19)** | | |

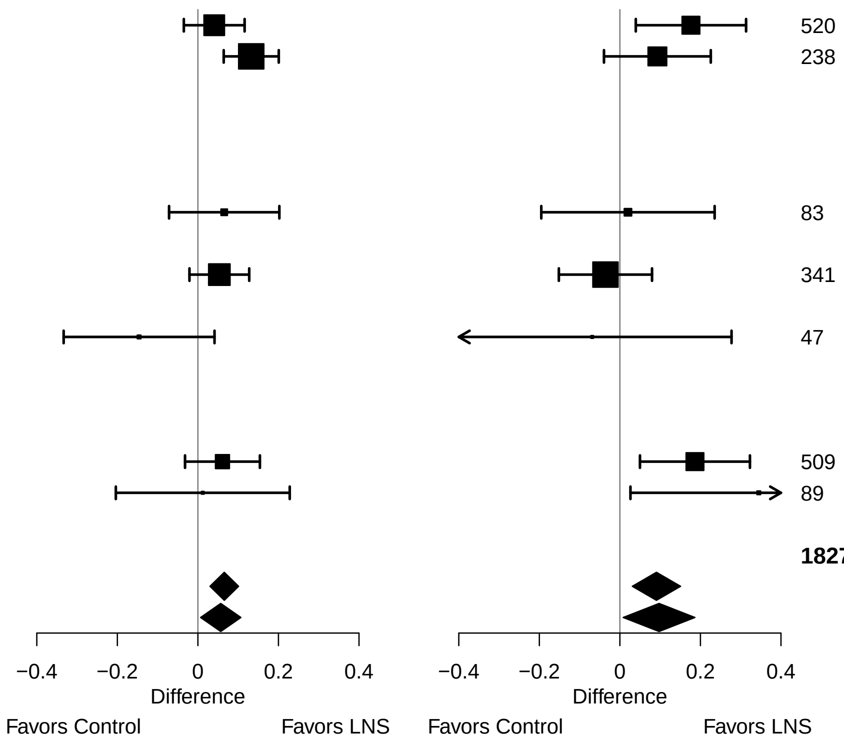

#### Supplemental figure 6O: Mean difference in HCZ

#### 6O6: Stratified by Child sex

**P−for−interaction = 0.307**
**Difference in MDs = 0.02 (−0.02, 0.06)**

| | | Male | | | | | | Female | | | | | |
|---|---|---|---|---|---|---|---|---|---|---|---|---|---|
| **Country** | **Trial** | **LNS N** | **Control N** | **Control Mean** | **MD (95% CI)** | **Fixed W** | **Random W** | **LNS N** | **Control N** | **Control Mean** | **MD (95% CI)** | **Fixed W** | **Random W** |
| Bangladesh | JiVitA−4 (34) | 1475 | 612 | −1.45 | 0.07 (0.02, 0.13) | 0.30 | 0.24 | 1410 | 655 | −1.44 | 0.04 (−0.02, 0.10) | 0.25 | 0.14 |
| Bangladesh | RDNS (35) | 833 | 411 | −1.78 | 0.00 (−0.11, 0.11) | 0.09 | 0.10 | 830 | 404 | −1.96 | 0.20 (0.10, 0.30) | 0.08 | 0.11 |
| Bangladesh | WASH−B (36) | 566 | 1730 | −1.60 | 0.09 (0.00, 0.19) | 0.11 | 0.12 | 597 | 1733 | −1.63 | 0.15 (0.07, 0.24) | 0.12 | 0.12 |
| Burkina Faso | iLiNS−Zinc (37) | 991 | 335 | −1.89 | 0.14 (0.05, 0.23) | 0.12 | 0.13 | 955 | 329 | −1.79 | 0.19 (0.13, 0.25) | 0.24 | 0.14 |
| Burkina Faso | PROMIS (38) | | | | | | | | | | | | |
| Burkina Faso | PROMIS CS (38) | | | | | | | | | | | | |
| Ghana | GHANA (39) | 60 | 41 | −0.70 | 0.24 (−0.13, 0.61) | 0.01 | 0.01 | 38 | 55 | −0.53 | 0.21 (−0.17, 0.59) | 0.01 | 0.02 |
| Ghana | iLiNS−DYADG (40) | 175 | 323 | −1.24 | 0.18 (0.01, 0.35) | 0.04 | 0.04 | 172 | 369 | −1.06 | −0.03 (−0.18, 0.12) | 0.04 | 0.08 |
| Haiti | HAITI (41) | | | | | | | | | | | | |
| Kenya | WASH−B (42) | 736 | 2466 | −0.29 | 0.02 (−0.06, 0.10) | 0.15 | 0.15 | 725 | 2710 | −0.27 | 0.07 (−0.02, 0.15) | 0.12 | 0.12 |
| Madagascar | MAHAY (43) | | | | | | | | | | | | |
| Malawi | iLiNS−DYADM (44) | 107 | 207 | −0.82 | −0.13 (−0.37, 0.11) | 0.02 | 0.02 | 113 | 237 | −0.73 | −0.12 (−0.33, 0.09) | 0.02 | 0.05 |
| Malawi | iLiNS−DOSE (45) | 349 | 123 | −1.25 | 0.00 (−0.16, 0.17) | 0.04 | 0.04 | 343 | 115 | −1.00 | 0.22 (0.04, 0.40) | 0.03 | 0.06 |
| Mali | PROMIS (46) | | | | | | | | | | | | |
| Mali | PROMIS CS (46) | | | | | | | | | | | | |
| Zimbabwe | SHINE (HIV−) (47) | 927 | 903 | −0.32 | 0.08 (−0.02, 0.18) | 0.11 | 0.12 | 944 | 880 | −0.23 | 0.09 (−0.01, 0.19) | 0.09 | 0.11 |
| Zimbabwe | SHINE (HIV+) (48) | 162 | 168 | −0.63 | 0.15 (−0.12, 0.42) | 0.01 | 0.02 | 175 | 161 | −0.48 | 0.05 (−0.16, 0.26) | 0.02 | 0.05 |
| | | **6381** | **7319** | | **$I^2$ = 0.12, $Tau^2$ = 0.00** | | | **6302** | **7648** | | **$I^2$ = 0.65, $Tau^2$ = 0.00** | | |
| **Fixed** | | | | | **0.07 (0.04, 0.10)** | | | | | | **0.11 (0.08, 0.14)** | | |
| **Random** | | | | | **0.07 (0.03, 0.11)** | | | | | | **0.10 (0.05, 0.16)** | | |

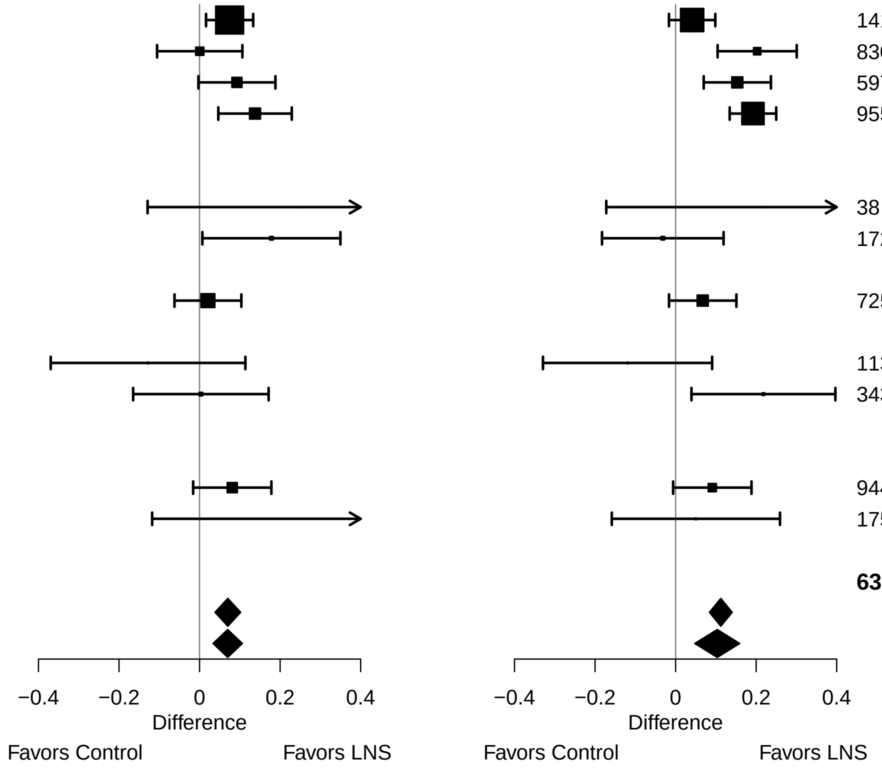

#### Supplemental figure 6O: Mean difference in HCZ

#### 6O7: Stratified by Child birth order

**P−for−interaction = 0.966**
**Difference in MDs = 0.00 (−0.04, 0.04)**

| Country | Trial | Later born LNS N | Later born Control N | Later born Control Mean | Later born MD (95% CI) | Later born Fixed W | Later born Random W | Firstborn LNS N | Firstborn Control N | Firstborn Control Mean | Firstborn MD (95% CI) | Firstborn Fixed W | Firstborn Random W |
|---|---|---|---|---|---|---|---|---|---|---|---|---|---|
| Bangladesh | JiVitA−4 (34) | 616 | 260 | −1.50 | 0.10 (0.02, 0.17) | 0.15 | 0.14 | 2265 | 1000 | −1.43 | 0.05 (0.00, 0.10) | 0.41 | 0.14 |
| Bangladesh | RDNS (35) | 974 | 508 | −1.85 | 0.12 (0.03, 0.22) | 0.09 | 0.10 | 688 | 307 | −1.89 | 0.07 (−0.05, 0.19) | 0.08 | 0.12 |
| Bangladesh | WASH−B (36) | 736 | 2313 | −1.60 | 0.14 (0.06, 0.21) | 0.15 | 0.14 | 427 | 1150 | −1.63 | 0.11 (−0.01, 0.22) | 0.09 | 0.12 |
| Burkina Faso | iLiNS−Zinc (37) | 1515 | 519 | −1.82 | 0.16 (0.09, 0.22) | 0.20 | 0.18 | 431 | 145 | −1.92 | 0.20 (0.12, 0.27) | 0.21 | 0.14 |
| Burkina Faso | PROMIS (38) | | | | | | | | | | | | |
| Burkina Faso | PROMIS CS (38) | | | | | | | | | | | | |
| Ghana | GHANA (39) | 60 | 54 | −0.60 | 0.25 (−0.11, 0.61) | 0.01 | 0.01 | 38 | 38 | −0.57 | 0.08 (−0.32, 0.47) | 0.01 | 0.04 |
| Ghana | iLiNS−DYADG (40) | 237 | 458 | −1.11 | 0.03 (−0.11, 0.17) | 0.04 | 0.05 | 110 | 234 | −1.20 | 0.14 (−0.06, 0.35) | 0.03 | 0.08 |
| Haiti | HAITI (41) | | | | | | | | | | | | |
| Kenya | WASH−B (42) | 1137 | 4074 | −0.26 | 0.05 (−0.02, 0.12) | 0.17 | 0.16 | 322 | 1097 | −0.35 | 0.02 (−0.11, 0.14) | 0.08 | 0.12 |
| Madagascar | MAHAY (43) | | | | | | | | | | | | |
| Malawi | iLiNS−DYADM (44) | 173 | 353 | −0.77 | −0.06 (−0.24, 0.11) | 0.03 | 0.03 | 47 | 89 | −0.76 | −0.38 (−0.74, −0.03) | 0.01 | 0.04 |
| Malawi | iLiNS−DOSE (45) | 458 | 162 | −1.13 | 0.03 (−0.13, 0.18) | 0.04 | 0.04 | 146 | 45 | −1.19 | 0.38 (0.11, 0.64) | 0.02 | 0.06 |
| Mali | PROMIS (46) | | | | | | | | | | | | |
| Mali | PROMIS CS (46) | | | | | | | | | | | | |
| Zimbabwe | SHINE (HIV−) (47) | 1295 | 1214 | −0.24 | 0.11 (0.02, 0.19) | 0.11 | 0.12 | 489 | 486 | −0.37 | 0.07 (−0.06, 0.21) | 0.06 | 0.11 |
| Zimbabwe | SHINE (HIV+) (48) | 280 | 246 | −0.49 | 0.03 (−0.18, 0.24) | 0.02 | 0.02 | 54 | 78 | −0.69 | 0.30 (−0.05, 0.65) | 0.01 | 0.04 |
| | | **7481** | **10161** | | **I² = 0.18, Tau² = 0.00** | | | **5017** | **4669** | | **I² = 0.60, Tau² = 0.01** | | |
| **Fixed** | | | | | **0.10 (0.07, 0.13)** | | | | | | **0.09 (0.06, 0.13)** | | |
| **Random** | | | | | **0.10 (0.06, 0.13)** | | | | | | **0.10 (0.01, 0.18)** | | |

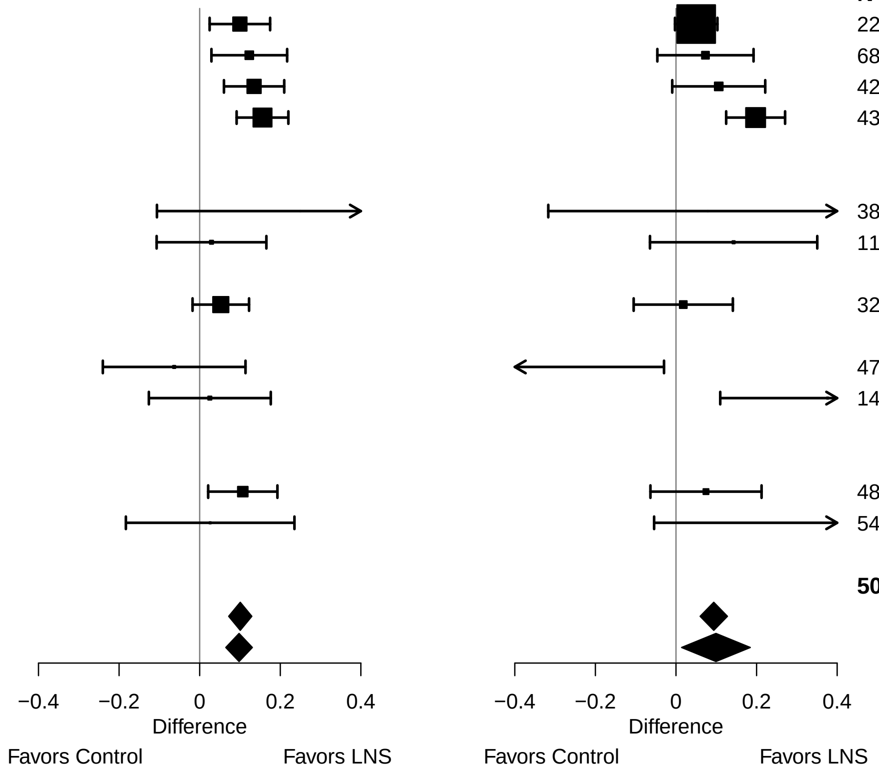

#### Supplemental figure 6O: Mean difference in HCZ

#### 6O8: Stratified by Child baseline anthropometric status

**P−for−interaction = 0.980**
**Difference in MDs = 0.00 (−0.05, 0.05)**

| Country | Trial | HCZ at least −1 Z: LNS N | Control N | Control Mean | MD (95% CI) | Fixed W | Random W | HCZ less than −1 Z: LNS N | Control N | Control Mean | MD (95% CI) | Fixed W | Random W |
|---|---|---|---|---|---|---|---|---|---|---|---|---|---|
| Bangladesh | JiVitA−4 (34) | 1411 | 630 | −0.88 | 0.09 (0.02, 0.16) | 0.26 | 0.16 | 1474 | 637 | −2.01 | 0.07 (0.00, 0.15) | 0.30 | 0.30 |
| Bangladesh | RDNS (35) | 521 | 239 | −1.06 | 0.02 (−0.06, 0.11) | 0.18 | 0.15 | 1080 | 521 | −2.23 | 0.11 (0.04, 0.17) | 0.35 | 0.35 |
| Bangladesh | WASH−B (36) | | | | | | | | | | | | |
| Burkina Faso | iLiNS−Zinc (37) | 550 | 176 | −0.96 | 0.26 (0.16, 0.36) | 0.13 | 0.14 | 1396 | 488 | −2.16 | 0.16 (0.06, 0.26) | 0.16 | 0.16 |
| Burkina Faso | PROMIS (38) | | | | | | | | | | | | |
| Burkina Faso | PROMIS CS (38) | | | | | | | | | | | | |
| Ghana | GHANA (39) | | | | | | | | | | | | |
| Ghana | iLiNS−DYADG (40) | 189 | 384 | −0.70 | 0.05 (−0.08, 0.17) | 0.09 | 0.12 | 134 | 257 | −1.81 | 0.12 (−0.01, 0.26) | 0.08 | 0.08 |
| Haiti | HAITI (41) | | | | | | | | | | | | |
| Kenya | WASH−B (42) | | | | | | | | | | | | |
| Madagascar | MAHAY (43) | | | | | | | | | | | | |
| Malawi | iLiNS−DYADM (44) | 155 | 323 | −0.43 | −0.15 (−0.30, 0.00) | 0.06 | 0.11 | 49 | 101 | −1.90 | 0.01 (−0.26, 0.28) | 0.02 | 0.02 |
| Malawi | iLiNS−DOSE (45) | 381 | 132 | −0.65 | 0.08 (−0.10, 0.27) | 0.04 | 0.09 | 311 | 106 | −1.73 | 0.16 (−0.05, 0.36) | 0.04 | 0.04 |
| Mali | PROMIS (46) | | | | | | | | | | | | |
| Mali | PROMIS CS (46) | | | | | | | | | | | | |
| Zimbabwe | SHINE (HIV−) (47) | 1072 | 967 | −0.02 | 0.06 (−0.02, 0.14) | 0.21 | 0.15 | 254 | 239 | −1.07 | −0.03 (−0.21, 0.16) | 0.04 | 0.04 |
| Zimbabwe | SHINE (HIV+) (48) | 200 | 168 | −0.23 | 0.11 (−0.11, 0.32) | 0.03 | 0.08 | 71 | 67 | −1.30 | 0.16 (−0.25, 0.57) | 0.01 | 0.01 |
| | | **4479** | **3019** | | **$I^2$ = 0.69, $Tau^2$ = 0.01** | | | **4769** | **2416** | | **$I^2$ = 0.00, $Tau^2$ = 0.00** | | |
| **Fixed** | | | | | **0.08 (0.04, 0.11)** | | | | | | **0.10 (0.06, 0.14)** | | |
| **Random** | | | | | **0.07 (−0.01, 0.15)** | | | | | | **0.10 (0.06, 0.14)** | | |

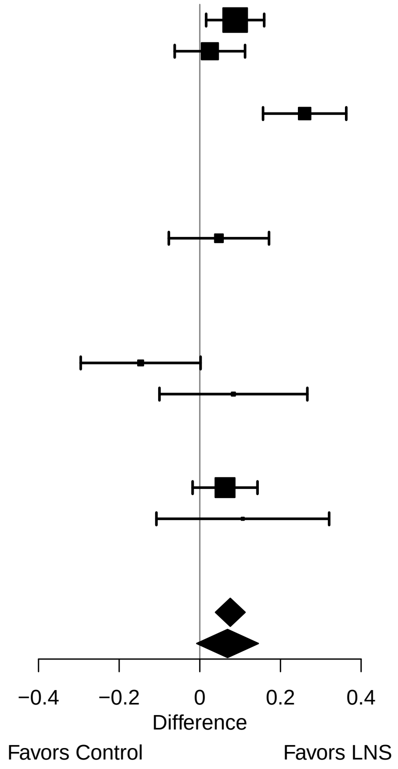

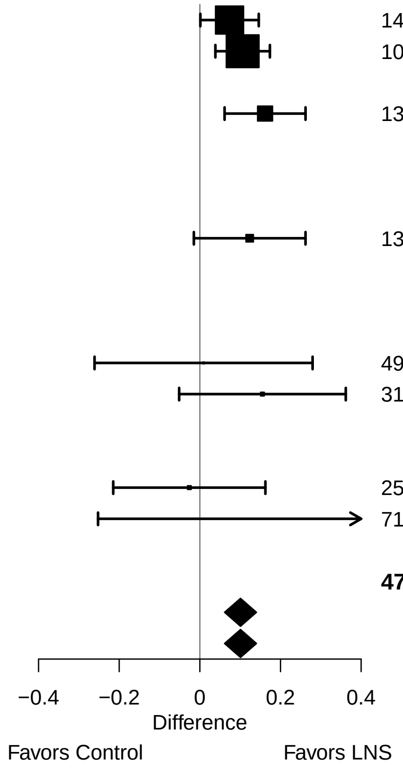

### Supplemental figure 6P: Small head size prevalence ratio

#### 6P1: Stratified by Maternal height

**P−for−interaction = 0.420**
**Ratio of PRs = 0.95 (0.85, 1.07)**

| | | At least 150.1 cm | | | | | | Less than 150.1 cm | | | | | |
|---|---|---|---|---|---|---|---|---|---|---|---|---|---|
| **Country** | **Trial** | **LNS N** | **Control N** | **Control Prevalence** | **PR (95% CI)** | **Fixed W** | **Random W** | **LNS N** | **Control N** | **Control Prevalence** | **PR (95% CI)** | **Fixed W** | **Random W** |
| Bangladesh | JiVitA−4 (34) | | | | | | | | | | | | |
| Bangladesh | RDNS (35) | 868 | 432 | 37.7 | 0.88 (0.77, 1.01) | 0.30 | 0.30 | 735 | 358 | 50.0 | 0.88 (0.79, 0.97) | 0.56 | 0.56 |
| Bangladesh | WASH−B (36) | 619 | 1899 | 27.3 | 0.96 (0.83, 1.12) | 0.24 | 0.24 | 544 | 1559 | 39.4 | 0.85 (0.75, 0.97) | 0.38 | 0.38 |
| Burkina Faso | iLiNS−Zinc (37) | 1911 | 649 | 40.9 | 0.89 (0.78, 1.02) | 0.28 | 0.28 | 33 | 15 | 46.7 | 0.92 (0.53, 1.60) | 0.02 | 0.02 |
| Burkina Faso | PROMIS (38) | | | | | | | | | | | | |
| Burkina Faso | PROMIS CS (38) | | | | | | | | | | | | |
| Ghana | GHANA (39) | | | | | | | | | | | | |
| Ghana | iLiNS−DYADG (40) | | | | | | | | | | | | |
| Haiti | HAITI (41) | | | | | | | | | | | | |
| Kenya | WASH−B (42) | 1316 | 4720 | 4.2 | 0.93 (0.68, 1.28) | 0.05 | 0.05 | 57 | 194 | 7.7 | 1.13 (0.42, 3.05) | 0.01 | 0.01 |
| Madagascar | MAHAY (43) | | | | | | | | | | | | |
| Malawi | iLiNS−DYADM (44) | | | | | | | | | | | | |
| Malawi | iLiNS−DOSE (45) | 581 | 195 | 18.9 | 0.82 (0.60, 1.13) | 0.05 | 0.05 | 111 | 42 | 35.7 | 0.72 (0.44, 1.17) | 0.03 | 0.03 |
| Mali | PROMIS (46) | | | | | | | | | | | | |
| Mali | PROMIS CS (46) | | | | | | | | | | | | |
| Zimbabwe | SHINE (HIV−) (47) | 1746 | 1687 | 5.6 | 0.78 (0.60, 1.02) | 0.08 | 0.08 | 76 | 62 | 12.9 | 0.92 (0.38, 2.22) | 0.01 | 0.01 |
| Zimbabwe | SHINE (HIV+) (48) | | | | | | | | | | | | |
| | | **7041** | **9582** | | **I² = 0.00, Tau² = 0.00** | | | **1556** | **2230** | | **I² = 0.00, Tau² = 0.00** | | |
| **Fixed** | | | | | **0.89 (0.83, 0.96)** | | | | | | **0.87 (0.80, 0.94)** | | |
| **Random** | | | | | **0.89 (0.83, 0.96)** | | | | | | **0.87 (0.80, 0.94)** | | |

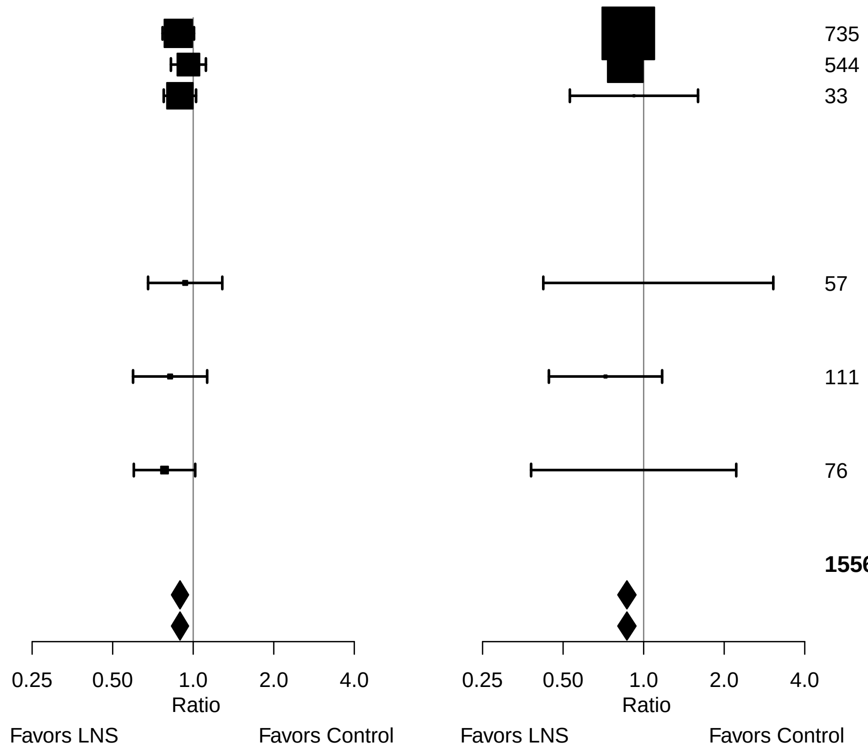

#### Supplemental figure 6P: Small head size prevalence ratio

#### 6P2: Stratified by Maternal BMI

**P−for−interaction = 0.607**
**Ratio of PRs = 0.97 (0.88, 1.08)**

| | | At least 20 kg/m² | | | | | | Less than 20 kg/m² | | | | | |
|---|---|---|---|---|---|---|---|---|---|---|---|---|---|
| **Country** | **Trial** | **LNS N** | **Control N** | **Control Prevalence** | **PR (95% CI)** | **Fixed W** | **Random W** | **LNS N** | **Control N** | **Control Prevalence** | **PR (95% CI)** | **Fixed W** | **Random W** |
| Bangladesh | JiVitA−4 (34) | | | | | | | | | | | | |
| Bangladesh | RDNS (35) | 730 | 335 | 39.7 | 0.85 (0.72, 1.00) | 0.22 | 0.22 | 873 | 455 | 45.9 | 0.91 (0.80, 1.03) | 0.37 | 0.37 |
| Bangladesh | WASH−B (36) | 519 | 1595 | 28.2 | 0.99 (0.84, 1.17) | 0.20 | 0.20 | 644 | 1863 | 36.6 | 0.85 (0.75, 0.96) | 0.36 | 0.36 |
| Burkina Faso | iLiNS−Zinc (37) | 1210 | 389 | 38.9 | 0.90 (0.79, 1.01) | 0.38 | 0.38 | 734 | 275 | 44.0 | 0.89 (0.74, 1.06) | 0.18 | 0.18 |
| Burkina Faso | PROMIS (38) | | | | | | | | | | | | |
| Burkina Faso | PROMIS CS (38) | | | | | | | | | | | | |
| Ghana | GHANA (39) | | | | | | | | | | | | |
| Ghana | iLiNS−DYADG (40) | 305 | 562 | 15.3 | 0.90 (0.64, 1.27) | 0.05 | 0.05 | 37 | 119 | 20.2 | 0.80 (0.36, 1.82) | 0.01 | 0.01 |
| Haiti | HAITI (41) | | | | | | | | | | | | |
| Kenya | WASH−B (42) | 1100 | 3831 | 4.0 | 0.90 (0.64, 1.28) | 0.05 | 0.05 | 273 | 1083 | 5.3 | 1.11 (0.65, 1.91) | 0.02 | 0.02 |
| Madagascar | MAHAY (43) | | | | | | | | | | | | |
| Malawi | iLiNS−DYADM (44) | 131 | 263 | 7.6 | 1.00 (0.48, 2.08) | 0.01 | 0.01 | 88 | 178 | 14.6 | 0.78 (0.39, 1.54) | 0.01 | 0.01 |
| Malawi | iLiNS−DOSE (45) | 512 | 172 | 17.3 | 0.87 (0.61, 1.25) | 0.04 | 0.04 | 178 | 64 | 34.4 | 0.66 (0.45, 0.95) | 0.04 | 0.04 |
| Mali | PROMIS (46) | | | | | | | | | | | | |
| Mali | PROMIS CS (46) | | | | | | | | | | | | |
| Zimbabwe | SHINE (HIV−) (47) | 1358 | 1298 | 5.9 | 0.76 (0.56, 1.03) | 0.06 | 0.06 | 239 | 248 | 6.0 | 1.18 (0.61, 2.26) | 0.01 | 0.01 |
| Zimbabwe | SHINE (HIV+) (48) | | | | | | | | | | | | |
| | | **5865** | **8445** | | **I² = 0.00, Tau² = 0.00** | | | **3066** | **4285** | | **I² = 0.00, Tau² = 0.00** | | |
| **Fixed** | | | | | **0.89 (0.83, 0.96)** | | | | | | **0.87 (0.81, 0.94)** | | |
| **Random** | | | | | **0.89 (0.83, 0.96)** | | | | | | **0.87 (0.81, 0.94)** | | |

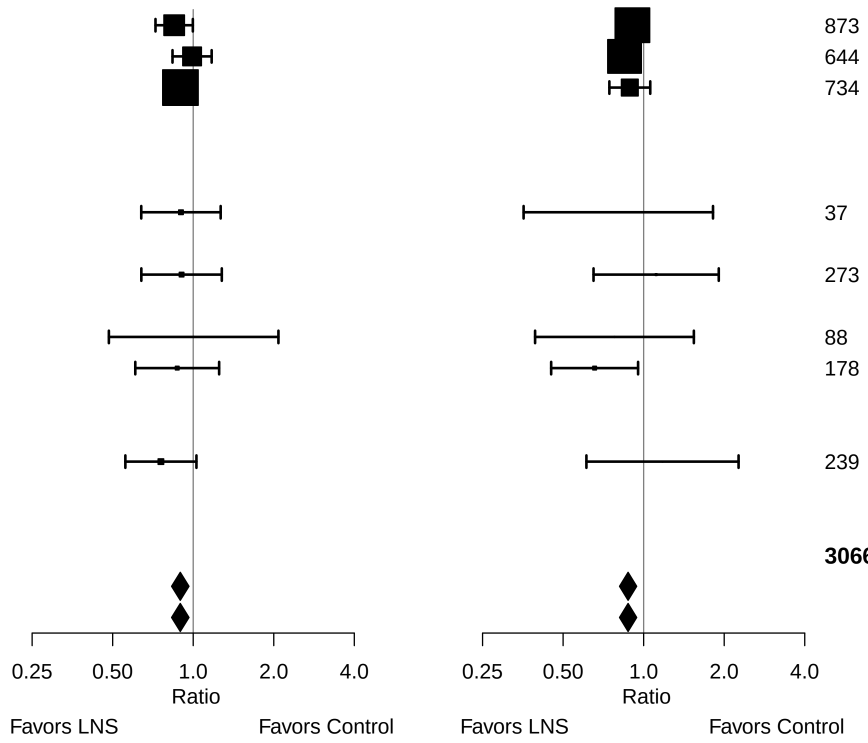

#### Supplemental figure 6P: Small head size prevalence ratio

#### 6P3: Stratified by Maternal age

**P−for−interaction = 0.907**
**Ratio of PRs = 0.99 (0.89, 1.11)**

| Country | Trial | At least 25 y LNS N | Control N | Control Prevalence | PR (95% CI) | Fixed W | Random W | Less than 25 y LNS N | Control N | Control Prevalence | PR (95% CI) | Fixed W | Random W |
|---|---|---|---|---|---|---|---|---|---|---|---|---|---|
| Bangladesh | JiVitA−4 (34) | 1238 | 554 | 22.1 | 1.12 (0.91, 1.39) | 0.16 | 0.16 | 1634 | 708 | 25.6 | 0.96 (0.82, 1.13) | 0.14 | 0.16 |
| Bangladesh | RDNS (35) | 453 | 221 | 42.5 | 0.82 (0.69, 0.98) | 0.23 | 0.23 | 1210 | 594 | 43.1 | 0.92 (0.82, 1.04) | 0.29 | 0.22 |
| Bangladesh | WASH−B (36) | 492 | 1523 | 32.2 | 0.91 (0.77, 1.07) | 0.26 | 0.26 | 666 | 1924 | 33.4 | 0.90 (0.79, 1.03) | 0.22 | 0.20 |
| Burkina Faso | iLiNS−Zinc (37) | 1128 | 399 | 39.4 | 0.84 (0.67, 1.04) | 0.14 | 0.14 | 805 | 262 | 43.1 | 0.98 (0.86, 1.11) | 0.24 | 0.21 |
| Burkina Faso | PROMIS (38) | | | | | | | | | | | | |
| Burkina Faso | PROMIS CS (38) | | | | | | | | | | | | |
| Ghana | GHANA (39) | | | | | | | | | | | | |
| Ghana | iLiNS−DYADG (40) | 220 | 417 | 14.6 | 0.87 (0.57, 1.32) | 0.04 | 0.04 | 127 | 275 | 18.2 | 0.91 (0.57, 1.45) | 0.02 | 0.03 |
| Haiti | HAITI (41) | | | | | | | | | | | | |
| Kenya | WASH−B (42) | 845 | 2831 | 4.0 | 0.94 (0.64, 1.38) | 0.05 | 0.05 | 601 | 2313 | 4.7 | 0.92 (0.59, 1.42) | 0.02 | 0.04 |
| Madagascar | MAHAY (43) | | | | | | | | | | | | |
| Malawi | iLiNS−DYADM (44) | 106 | 226 | 9.3 | 0.71 (0.31, 1.62) | 0.01 | 0.01 | 114 | 218 | 12.4 | 0.99 (0.54, 1.81) | 0.01 | 0.02 |
| Malawi | iLiNS−DOSE (45) | 368 | 134 | 16.4 | 1.09 (0.73, 1.63) | 0.04 | 0.04 | 310 | 97 | 29.6 | 0.53 (0.37, 0.75) | 0.03 | 0.05 |
| Mali | PROMIS (46) | | | | | | | | | | | | |
| Mali | PROMIS CS (46) | | | | | | | | | | | | |
| Zimbabwe | SHINE (HIV−) (47) | 869 | 795 | 5.4 | 0.79 (0.52, 1.18) | 0.04 | 0.04 | 805 | 792 | 6.3 | 0.81 (0.54, 1.21) | 0.02 | 0.04 |
| Zimbabwe | SHINE (HIV+) (48) | 244 | 231 | 8.2 | 1.20 (0.64, 2.22) | 0.02 | 0.02 | 73 | 78 | 14.1 | 0.78 (0.37, 1.65) | 0.01 | 0.01 |
| | | **5963** | **7331** | | **I² = 0.00, Tau² = 0.00** | | | **6345** | **7261** | | **I² = 0.21, Tau² = 0.01** | | |
| **Fixed** | | | | | **0.91 (0.84, 0.99)** | | | | | | **0.92 (0.86, 0.98)** | | |
| **Random** | | | | | **0.91 (0.84, 0.99)** | | | | | | **0.90 (0.82, 0.99)** | | |

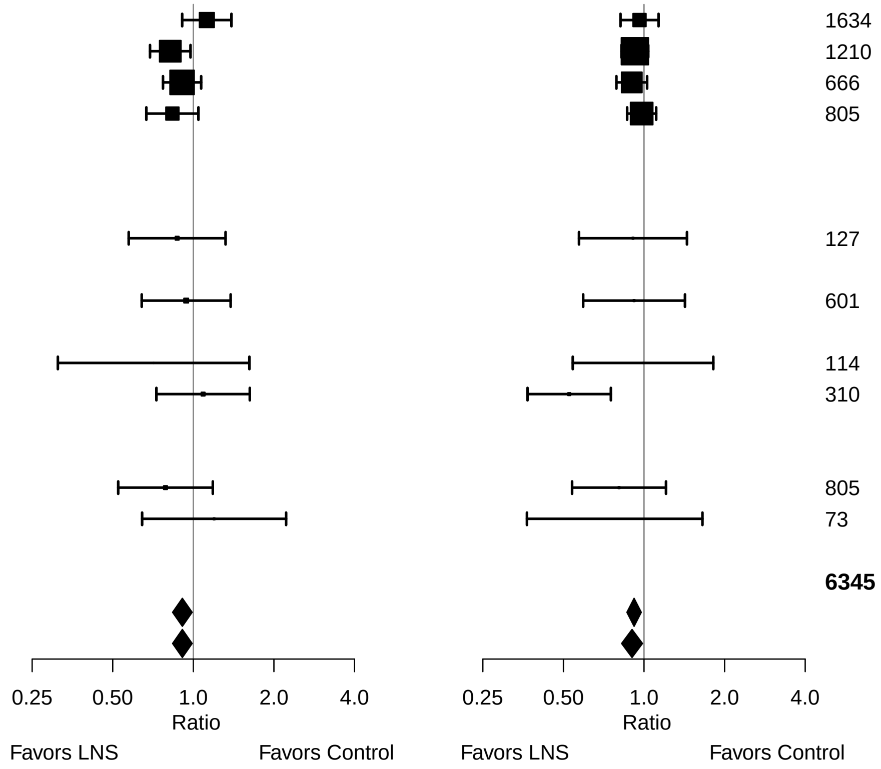

#### Supplemental figure 6P: Small head size prevalence ratio

#### 6P4: Stratified by Maternal education

**P−for−interaction = 0.091**
**Ratio of PRs = 1.11 (0.98, 1.26)**

| | | Primary or greater | | | | | | Incomplete or no formal | | | | | |
|---|---|---|---|---|---|---|---|---|---|---|---|---|---|
| **Country** | **Trial** | **LNS N** | **Control N** | **Control Prevalence** | **PR (95% CI)** | **Fixed W** | **Random W** | **LNS N** | **Control N** | **Control Prevalence** | **PR (95% CI)** | **Fixed W** | **Random W** |
| Bangladesh | JiVitA−4 (34) | 1855 | 775 | 23.3 | 0.91 (0.77, 1.08) | 0.18 | 0.18 | 1028 | 489 | 25.5 | 1.21 (0.98, 1.48) | 0.14 | 0.16 |
| Bangladesh | RDNS (35) | 1245 | 589 | 41.6 | 0.88 (0.78, 0.98) | 0.41 | 0.41 | 418 | 226 | 46.5 | 0.96 (0.80, 1.14) | 0.18 | 0.19 |
| Bangladesh | WASH−B (36) | 817 | 2477 | 31.2 | 0.85 (0.75, 0.97) | 0.32 | 0.32 | 346 | 986 | 36.7 | 1.00 (0.84, 1.19) | 0.20 | 0.19 |
| Burkina Faso | iLiNS−Zinc (37) | 82 | 16 | 31.2 | 1.20 (0.67, 2.14) | 0.02 | 0.02 | 1851 | 645 | 41.1 | 0.88 (0.78, 1.01) | 0.34 | 0.24 |
| Burkina Faso | PROMIS (38) | | | | | | | | | | | | |
| Burkina Faso | PROMIS CS (38) | | | | | | | | | | | | |
| Ghana | GHANA (39) | | | | | | | | | | | | |
| Ghana | iLiNS−DYADG (40) | 267 | 548 | 15.3 | 0.86 (0.59, 1.23) | 0.04 | 0.04 | 80 | 144 | 18.8 | 0.93 (0.52, 1.67) | 0.02 | 0.03 |
| Haiti | HAITI (41) | | | | | | | | | | | | |
| Kenya | WASH−B (42) | 703 | 2456 | 2.8 | 1.16 (0.68, 1.99) | 0.02 | 0.02 | 756 | 2716 | 5.7 | 0.81 (0.59, 1.12) | 0.06 | 0.09 |
| Madagascar | MAHAY (43) | | | | | | | | | | | | |
| Malawi | iLiNS−DYADM (44) | | | | | | | | | | | | |
| Malawi | iLiNS−DOSE (45) | 163 | 55 | 16.1 | 0.86 (0.45, 1.65) | 0.01 | 0.01 | 517 | 176 | 23.9 | 0.74 (0.55, 0.99) | 0.07 | 0.10 |
| Mali | PROMIS (46) | | | | | | | | | | | | |
| Mali | PROMIS CS (46) | | | | | | | | | | | | |
| Zimbabwe | SHINE (HIV−) (47) | | | | | | | | | | | | |
| Zimbabwe | SHINE (HIV+) (48) | | | | | | | | | | | | |
| | | **5132** | **6916** | | **I² = 0.00, Tau² = 0.00** | | | **4996** | **5382** | | **I² = 0.43, Tau² = 0.01** | | |
| **Fixed** | | | | | **0.88 (0.82, 0.95)** | | | | | | **0.94 (0.87, 1.02)** | | |
| **Random** | | | | | **0.88 (0.82, 0.95)** | | | | | | **0.94 (0.84, 1.05)** | | |

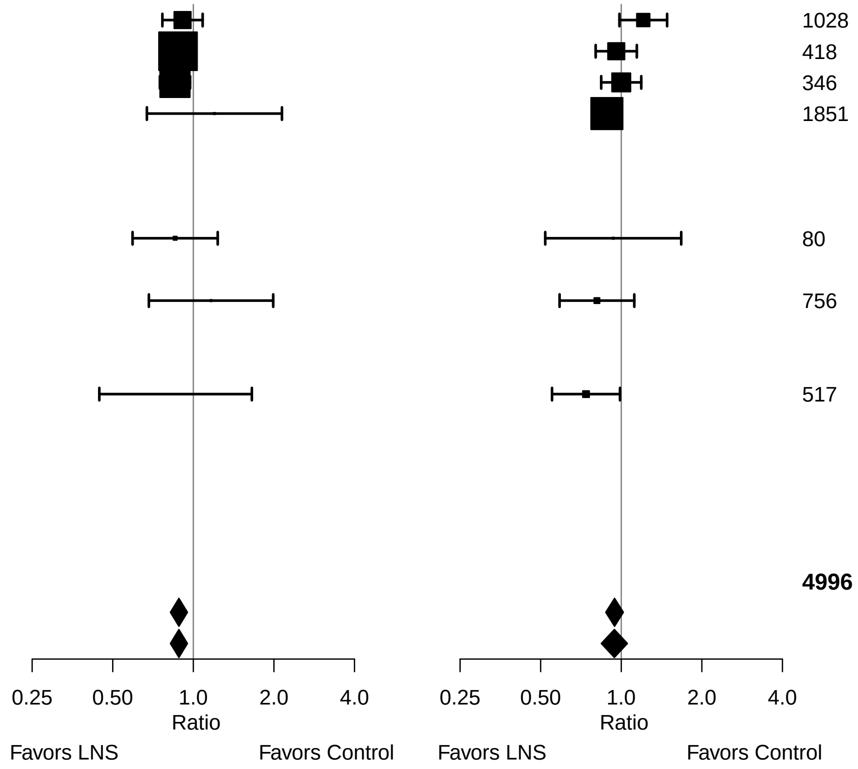

#### Supplemental figure 6P: Small head size prevalence ratio

### 6P5: Stratified by Maternal depressive symptoms

**P−for−interaction = 0.095**
**Ratio of PRs = 1.12 (0.98, 1.27)**

| | | Less than 75th percentile | | | | | | At least 75th percentile | | | | | |
|---|---|---|---|---|---|---|---|---|---|---|---|---|---|
| **Country** | **Trial** | **LNS N** | **Control N** | **Control Prevalence** | **PR (95% CI)** | **Fixed W** | **Random W** | **LNS N** | **Control N** | **Control Prevalence** | **PR (95% CI)** | **Fixed W** | **Random W** |
| Bangladesh | JiVitA−4 (34) | | | | | | | | | | | | |
| Bangladesh | RDNS (35) | 1081 | 472 | 39.0 | 1.01 (0.91, 1.11) | 0.48 | 0.48 | 520 | 288 | 49.3 | 0.74 (0.63, 0.88) | 0.50 | 0.42 |
| Bangladesh | WASH−B (36) | 901 | 2487 | 33.0 | 0.85 (0.76, 0.96) | 0.38 | 0.38 | 238 | 885 | 31.9 | 1.05 (0.87, 1.28) | 0.35 | 0.34 |
| Burkina Faso | iLiNS−Zinc (37) | | | | | | | | | | | | |
| Burkina Faso | PROMIS (38) | | | | | | | | | | | | |
| Burkina Faso | PROMIS CS (38) | | | | | | | | | | | | |
| Ghana | GHANA (39) | | | | | | | | | | | | |
| Ghana | iLiNS−DYADG (40) | 252 | 459 | 15.0 | 1.00 (0.70, 1.44) | 0.04 | 0.04 | 83 | 209 | 17.7 | 0.68 (0.36, 1.30) | 0.03 | 0.05 |
| Haiti | HAITI (41) | | | | | | | | | | | | |
| Kenya | WASH−B (42) | 998 | 3517 | 4.0 | 0.92 (0.64, 1.31) | 0.04 | 0.04 | 341 | 1248 | 5.4 | 0.71 (0.32, 1.56) | 0.02 | 0.04 |
| Madagascar | MAHAY (43) | | | | | | | | | | | | |
| Malawi | iLiNS−DYADM (44) | 157 | 303 | 10.2 | 0.81 (0.44, 1.50) | 0.01 | 0.01 | 47 | 113 | 11.5 | 1.29 (0.55, 3.04) | 0.02 | 0.03 |
| Malawi | iLiNS−DOSE (45) | | | | | | | | | | | | |
| Mali | PROMIS (46) | | | | | | | | | | | | |
| Mali | PROMIS CS (46) | | | | | | | | | | | | |
| Zimbabwe | SHINE (HIV−) (47) | 1193 | 1135 | 5.0 | 0.88 (0.60, 1.30) | 0.03 | 0.03 | 509 | 490 | 7.1 | 0.72 (0.45, 1.14) | 0.06 | 0.09 |
| Zimbabwe | SHINE (HIV+) (48) | 234 | 215 | 8.8 | 1.16 (0.68, 1.99) | 0.02 | 0.02 | 89 | 100 | 12.0 | 0.75 (0.32, 1.75) | 0.02 | 0.03 |
| | | **4816** | **8588** | | **I² = 0.00, Tau² = 0.00** | | | **1827** | **3333** | | **I² = 0.37, Tau² = 0.01** | | |
| **Fixed** | | | | | **0.94 (0.87, 1.01)** | | | | | | **0.84 (0.75, 0.95)** | | |
| **Random** | | | | | **0.94 (0.87, 1.01)** | | | | | | **0.85 (0.73, 0.98)** | | |

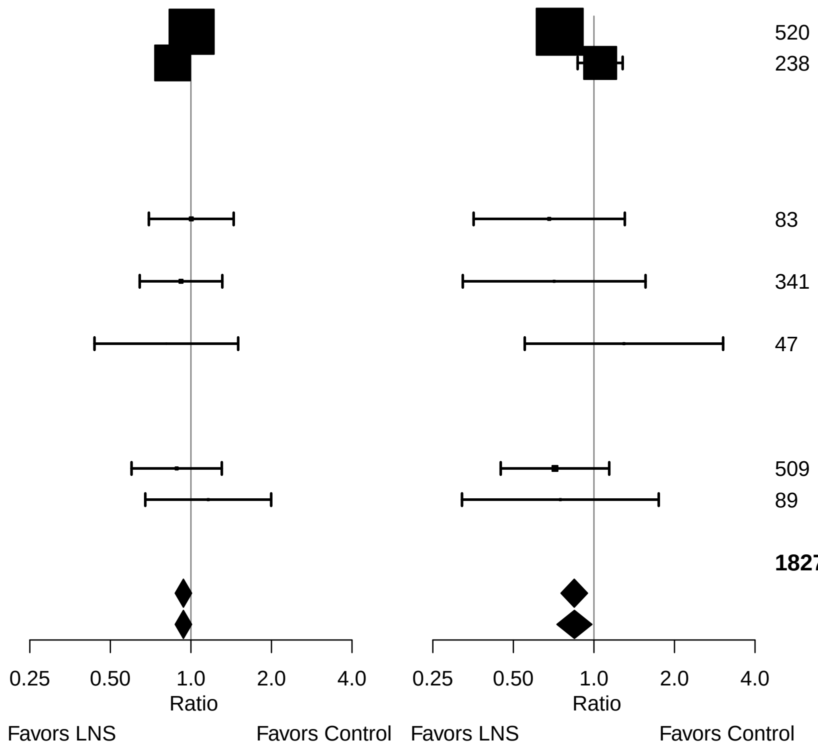

#### Supplemental figure 6P: Small head size prevalence ratio

#### 6P6: Stratified by Child sex

**P−for−interaction = 0.020**
**Ratio of PRs = 0.89 (0.81, 0.98)**

| Country | Trial | Male LNS N | Male Control N | Male Control Prevalence | Male PR (95% CI) | Male Fixed W | Male Random W | Female LNS N | Female Control N | Female Control Prevalence | Female PR (95% CI) | Female Fixed W | Female Random W |
|---|---|---|---|---|---|---|---|---|---|---|---|---|---|
| Bangladesh | JiVitA−4 (34) | 1475 | 612 | 25.8 | 0.92 (0.79, 1.08) | 0.18 | 0.18 | 1410 | 655 | 22.4 | 1.22 (0.99, 1.49) | 0.13 | 0.15 |
| Bangladesh | RDNS (35) | 833 | 411 | 37.7 | 1.07 (0.94, 1.21) | 0.28 | 0.28 | 830 | 404 | 48.3 | 0.76 (0.65, 0.89) | 0.24 | 0.17 |
| Bangladesh | WASH−B (36) | 566 | 1730 | 32.4 | 0.93 (0.81, 1.07) | 0.22 | 0.22 | 597 | 1733 | 33.2 | 0.88 (0.76, 1.02) | 0.26 | 0.17 |
| Burkina Faso | iLiNS−Zinc (37) | 991 | 335 | 42.3 | 0.97 (0.82, 1.14) | 0.16 | 0.16 | 955 | 329 | 39.7 | 0.78 (0.67, 0.92) | 0.23 | 0.17 |
| Burkina Faso | PROMIS (38) | | | | | | | | | | | | |
| Burkina Faso | PROMIS CS (38) | | | | | | | | | | | | |
| Ghana | GHANA (39) | | | | | | | | | | | | |
| Ghana | iLiNS−DYADG (40) | 175 | 323 | 19.2 | 0.86 (0.58, 1.29) | 0.03 | 0.03 | 172 | 369 | 13.3 | 0.88 (0.54, 1.43) | 0.02 | 0.06 |
| Haiti | HAITI (41) | | | | | | | | | | | | |
| Kenya | WASH−B (42) | 736 | 2466 | 4.9 | 0.87 (0.58, 1.28) | 0.03 | 0.03 | 725 | 2710 | 3.8 | 0.97 (0.62, 1.51) | 0.03 | 0.07 |
| Madagascar | MAHAY (43) | | | | | | | | | | | | |
| Malawi | iLiNS−DYADM (44) | 107 | 207 | 13.0 | 0.86 (0.45, 1.63) | 0.01 | 0.01 | 113 | 237 | 8.9 | 0.90 (0.43, 1.90) | 0.01 | 0.03 |
| Malawi | iLiNS−DOSE (45) | 349 | 123 | 23.6 | 0.97 (0.69, 1.36) | 0.04 | 0.04 | 343 | 115 | 19.8 | 0.57 (0.37, 0.87) | 0.03 | 0.07 |
| Mali | PROMIS (46) | | | | | | | | | | | | |
| Mali | PROMIS CS (46) | | | | | | | | | | | | |
| Zimbabwe | SHINE (HIV−) (47) | 927 | 903 | 6.4 | 0.87 (0.63, 1.21) | 0.04 | 0.04 | 944 | 880 | 5.1 | 0.75 (0.50, 1.11) | 0.04 | 0.08 |
| Zimbabwe | SHINE (HIV+) (48) | 162 | 168 | 14.3 | 0.73 (0.40, 1.33) | 0.01 | 0.01 | 175 | 161 | 5.6 | 1.64 (0.68, 3.93) | 0.01 | 0.02 |
| | | **6321** | **7278** | | **I² = 0.00, Tau² = 0.00** | | | **6264** | **7593** | | **I² = 0.58, Tau² = 0.02** | | |
| **Fixed** | | | | | **0.96 (0.90, 1.03)** | | | | | | **0.85 (0.79, 0.92)** | | |
| **Random** | | | | | **0.96 (0.90, 1.03)** | | | | | | **0.86 (0.75, 0.99)** | | |

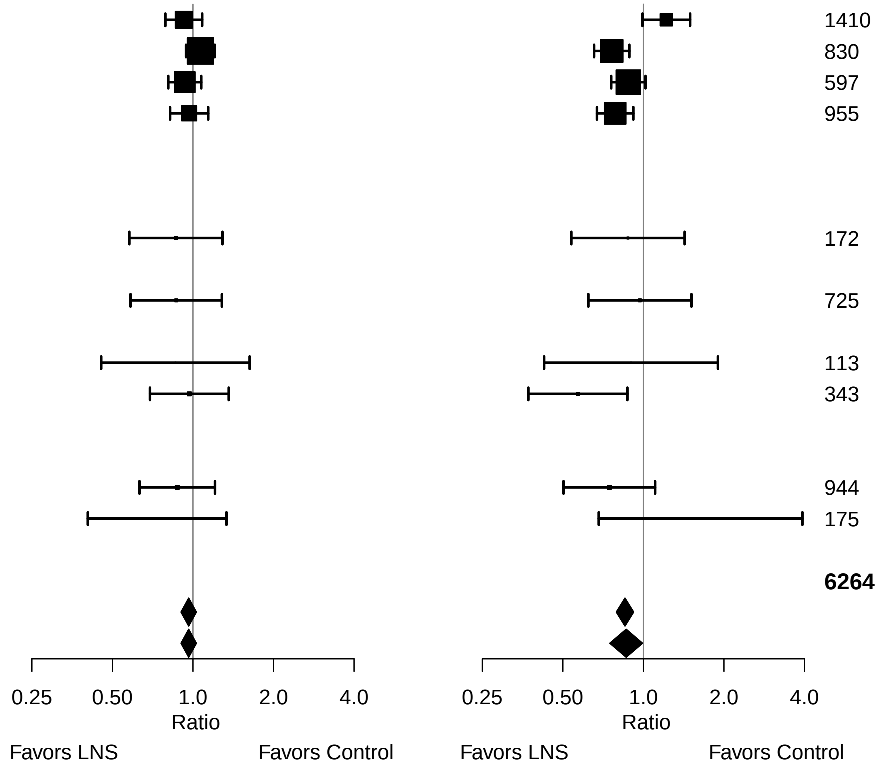

#### Supplemental figure 6P: Small head size prevalence ratio

#### 6P7: Stratified by Child birth order

**P−for−interaction = 0.696**
**Ratio of PRs = 1.02 (0.92, 1.14)**

| | | Later born | | | | | | Firstborn | | | | | |
|---|---|---|---|---|---|---|---|---|---|---|---|---|---|
| **Country** | **Trial** | **LNS N** | **Control N** | **Control Prevalence** | **PR (95% CI)** | **Fixed W** | **Random W** | **LNS N** | **Control N** | **Control Prevalence** | **PR (95% CI)** | **Fixed W** | **Random W** |
| Bangladesh | JiVitA−4 (34) | 616 | 260 | 25.0 | 0.99 (0.78, 1.26) | 0.08 | 0.08 | 2265 | 1000 | 23.9 | 1.03 (0.88, 1.21) | 0.25 | 0.18 |
| Bangladesh | RDNS (35) | 974 | 508 | 43.1 | 0.87 (0.77, 0.97) | 0.34 | 0.34 | 688 | 307 | 42.7 | 0.94 (0.80, 1.09) | 0.27 | 0.18 |
| Bangladesh | WASH−B (36) | 736 | 2313 | 31.9 | 0.90 (0.79, 1.02) | 0.27 | 0.27 | 427 | 1150 | 34.6 | 0.91 (0.77, 1.07) | 0.22 | 0.18 |
| Burkina Faso | iLiNS−Zinc (37) | 1515 | 519 | 40.3 | 0.86 (0.72, 1.02) | 0.14 | 0.14 | 431 | 145 | 43.4 | 0.99 (0.81, 1.19) | 0.17 | 0.17 |
| Burkina Faso | PROMIS (38) | | | | | | | | | | | | |
| Burkina Faso | PROMIS CS (38) | | | | | | | | | | | | |
| Ghana | GHANA (39) | | | | | | | | | | | | |
| Ghana | iLiNS−DYADG (40) | 237 | 458 | 14.4 | 0.97 (0.66, 1.42) | 0.03 | 0.03 | 110 | 234 | 19.2 | 0.76 (0.45, 1.28) | 0.02 | 0.07 |
| Haiti | HAITI (41) | | | | | | | | | | | | |
| Kenya | WASH−B (42) | 1137 | 4074 | 4.3 | 0.97 (0.70, 1.35) | 0.04 | 0.04 | 322 | 1097 | 4.3 | 0.72 (0.40, 1.30) | 0.02 | 0.06 |
| Madagascar | MAHAY (43) | | | | | | | | | | | | |
| Malawi | iLiNS−DYADM (44) | 173 | 353 | 10.8 | 0.75 (0.42, 1.35) | 0.01 | 0.01 | 47 | 89 | 10.1 | 1.47 (0.59, 3.70) | 0.01 | 0.03 |
| Malawi | iLiNS−DOSE (45) | 458 | 162 | 20.2 | 0.91 (0.65, 1.27) | 0.04 | 0.04 | 146 | 45 | 26.7 | 0.42 (0.25, 0.71) | 0.02 | 0.07 |
| Mali | PROMIS (46) | | | | | | | | | | | | |
| Mali | PROMIS CS (46) | | | | | | | | | | | | |
| Zimbabwe | SHINE (HIV−) (47) | 1295 | 1214 | 5.0 | 0.80 (0.58, 1.10) | 0.04 | 0.04 | 489 | 486 | 7.8 | 0.76 (0.46, 1.25) | 0.03 | 0.07 |
| Zimbabwe | SHINE (HIV+) (48) | | | | | | | | | | | | |
| | | **7141** | **9861** | | **I² = 0.00, Tau² = 0.00** | | | **4925** | **4553** | | **I² = 0.42, Tau² = 0.03** | | |
| **Fixed** | | | | | **0.89 (0.83, 0.95)** | | | | | | **0.93 (0.86, 1.01)** | | |
| **Random** | | | | | **0.89 (0.83, 0.95)** | | | | | | **0.88 (0.74, 1.03)** | | |

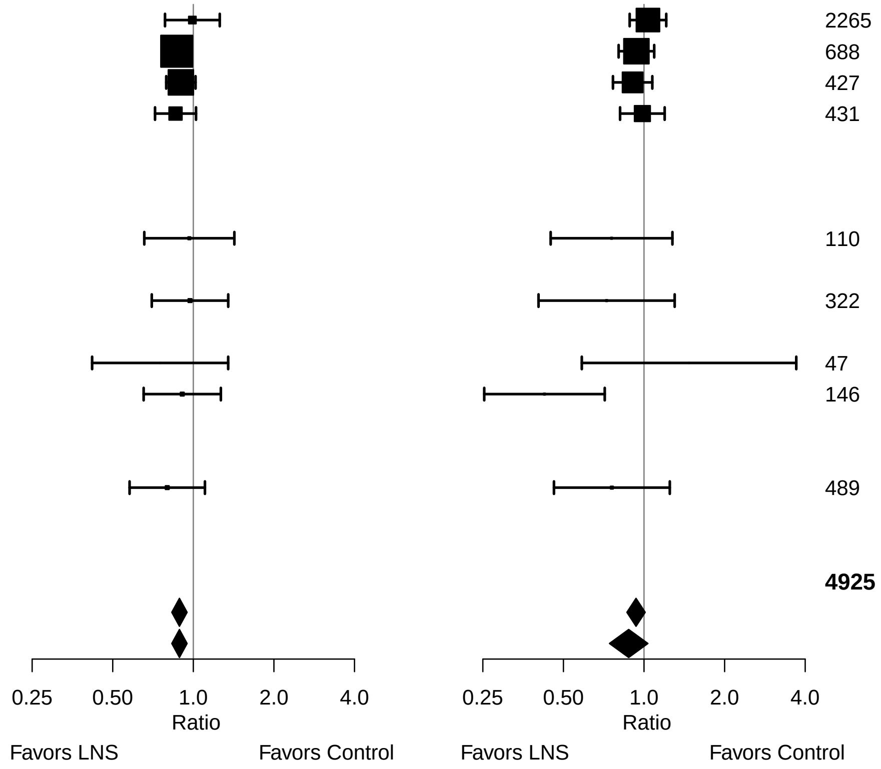

#### Supplemental figure 6P: Small head size prevalence ratio

#### 6P8: Stratified by Child baseline anthropometric status

**P−for−interaction = 0.152**
**Ratio of PRs = 1.25 (0.92, 1.70)**

| | | HCZ at least −1 Z | | | | | | HCZ less than −1 Z | | | | | |
|---|---|---|---|---|---|---|---|---|---|---|---|---|---|
| **Country** | **Trial** | **LNS N** | **Control N** | **Control Prevalence** | **PR (95% CI)** | **Fixed W** | **Random W** | **LNS N** | **Control N** | **Control Prevalence** | **PR (95% CI)** | **Fixed W** | **Random W** |
| Bangladesh | JiVitA−4 (34) | 1411 | 630 | 2.7 | 1.05 (0.54, 2.04) | 0.18 | 0.16 | 1474 | 637 | 45.4 | 0.95 (0.85, 1.06) | 0.26 | 0.26 |
| Bangladesh | RDNS (35) | 521 | 239 | 5.0 | 0.50 (0.23, 1.07) | 0.13 | 0.14 | 1080 | 521 | 60.3 | 0.92 (0.85, 1.00) | 0.48 | 0.48 |
| Bangladesh | WASH−B (36) | | | | | | | | | | | | |
| Burkina Faso | iLiNS−Zinc (37) | 550 | 176 | 5.1 | 0.21 (0.09, 0.53) | 0.10 | 0.12 | 1396 | 488 | 54.1 | 0.89 (0.77, 1.03) | 0.16 | 0.16 |
| Burkina Faso | PROMIS (38) | | | | | | | | | | | | |
| Burkina Faso | PROMIS CS (38) | | | | | | | | | | | | |
| Ghana | GHANA (39) | | | | | | | | | | | | |
| Ghana | iLiNS−DYADG (40) | 189 | 384 | 2.3 | 1.35 (0.49, 3.75) | 0.08 | 0.11 | 134 | 257 | 35.4 | 0.89 (0.66, 1.20) | 0.04 | 0.04 |
| Haiti | HAITI (41) | | | | | | | | | | | | |
| Kenya | WASH−B (42) | | | | | | | | | | | | |
| Madagascar | MAHAY (43) | | | | | | | | | | | | |
| Malawi | iLiNS−DYADM (44) | | | | | | | | | | | | |
| Malawi | iLiNS−DOSE (45) | 381 | 132 | 7.6 | 0.90 (0.45, 1.82) | 0.16 | 0.16 | 311 | 106 | 39.6 | 0.76 (0.57, 1.02) | 0.04 | 0.04 |
| Mali | PROMIS (46) | | | | | | | | | | | | |
| Mali | PROMIS CS (46) | | | | | | | | | | | | |
| Zimbabwe | SHINE (HIV−) (47) | 1072 | 967 | 2.8 | 0.77 (0.45, 1.31) | 0.27 | 0.19 | 254 | 239 | 15.5 | 0.94 (0.62, 1.44) | 0.02 | 0.02 |
| Zimbabwe | SHINE (HIV+) (48) | 200 | 168 | 4.2 | 0.84 (0.33, 2.17) | 0.09 | 0.12 | 71 | 67 | 28.4 | 0.74 (0.37, 1.48) | 0.01 | 0.01 |
| | | **4324** | **2696** | | **I² = 0.46, Tau² = 0.17** | | | **4720** | **2315** | | **I² = 0.00, Tau² = 0.00** | | |
| **Fixed** | | | | | **0.73 (0.55, 0.97)** | | | | | | **0.92 (0.87, 0.97)** | | |
| **Random** | | | | | **0.71 (0.47, 1.09)** | | | | | | **0.92 (0.87, 0.97)** | | |

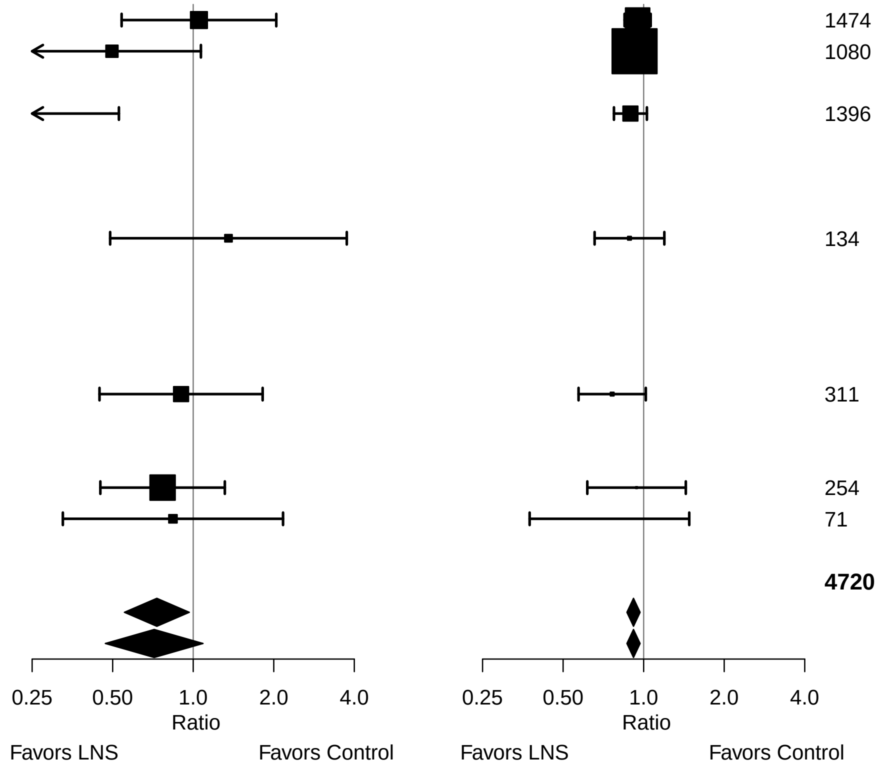

### Supplemental figure 6Q: Small head size prevalence difference

#### 6Q1: Stratified by Maternal height

**P−for−interaction = 0.301**
**Difference in PDs = −0.02 (−0.06, 0.02)**

| | | At least 150.1 cm | | | | | | Less than 150.1 cm | | | | | |
|---|---|---|---|---|---|---|---|---|---|---|---|---|---|
| **Country** | **Trial** | **LNS N** | **Control N** | **Control Prevalence** | **PD (95% CI)** | **Fixed W** | **Random W** | **LNS N** | **Control N** | **Control Prevalence** | **PD (95% CI)** | **Fixed W** | **Random W** |
| Bangladesh | JiVitA−4 (34) | | | | | | | | | | | | |
| Bangladesh | RDNS (35) | 868 | 432 | 37.7 | −0.05 (−0.09, 0.00) | 0.03 | 0.03 | 735 | 358 | 50.0 | −0.06 (−0.11, −0.01) | 0.34 | 0.34 |
| Bangladesh | WASH−B (36) | 619 | 1899 | 27.3 | −0.01 (−0.05, 0.03) | 0.05 | 0.05 | 544 | 1559 | 39.4 | −0.06 (−0.10, −0.01) | 0.42 | 0.42 |
| Burkina Faso | iLiNS−Zinc (37) | 1911 | 649 | 40.9 | −0.04 (−0.10, 0.01) | 0.02 | 0.02 | 33 | 15 | 46.7 | −0.01 (−0.26, 0.23) | 0.01 | 0.01 |
| Burkina Faso | PROMIS (38) | | | | | | | | | | | | |
| Burkina Faso | PROMIS CS (38) | | | | | | | | | | | | |
| Ghana | GHANA (39) | | | | | | | | | | | | |
| Ghana | iLiNS−DYADG (40) | | | | | | | | | | | | |
| Haiti | HAITI (41) | | | | | | | | | | | | |
| Kenya | WASH−B (42) | 1316 | 4720 | 4.2 | 0.00 (−0.02, 0.01) | 0.47 | 0.47 | 57 | 194 | 7.7 | 0.01 (−0.07, 0.09) | 0.12 | 0.12 |
| Madagascar | MAHAY (43) | | | | | | | | | | | | |
| Malawi | iLiNS−DYADM (44) | | | | | | | | | | | | |
| Malawi | iLiNS−DOSE (45) | 581 | 195 | 18.9 | −0.04 (−0.09, 0.02) | 0.02 | 0.02 | 111 | 42 | 35.7 | −0.07 (−0.22, 0.07) | 0.04 | 0.04 |
| Mali | PROMIS (46) | | | | | | | | | | | | |
| Mali | PROMIS CS (46) | | | | | | | | | | | | |
| Zimbabwe | SHINE (HIV−) (47) | 1746 | 1687 | 5.6 | −0.01 (−0.03, 0.00) | 0.41 | 0.41 | 76 | 62 | 12.9 | −0.01 (−0.12, 0.10) | 0.07 | 0.07 |
| Zimbabwe | SHINE (HIV+) (48) | | | | | | | | | | | | |
| | | **7041** | **9582** | | **I² = 0.12, Tau² = 0.00** | | | **1556** | **2230** | | **I² = 0.00, Tau² = 0.00** | | |
| **Fixed** | | | | | **−0.01 (−0.02, 0.00)** | | | | | | **−0.05 (−0.08, −0.02)** | | |
| **Random** | | | | | **−0.01 (−0.02, 0.00)** | | | | | | **−0.05 (−0.08, −0.02)** | | |

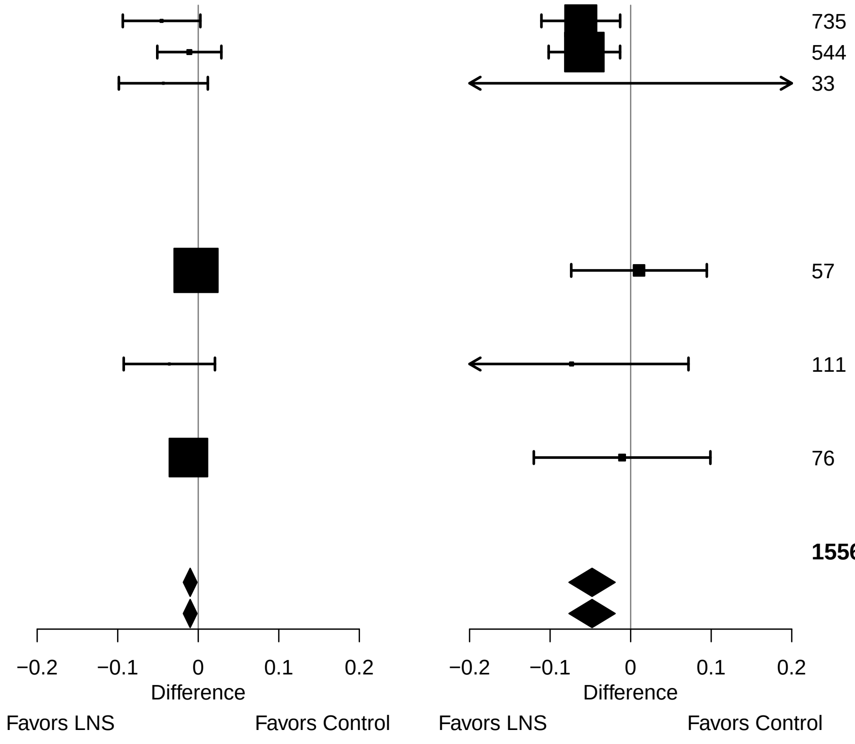

#### Supplemental figure 6Q: Small head size prevalence difference

#### 6Q2: Stratified by Maternal BMI

**P−for−interaction = 0.881**
**Difference in PDs = 0.00 (−0.02, 0.02)**

| | | At least 20 kg/m² | | | | | | Less than 20 kg/m² | | | | | |
|---|---|---|---|---|---|---|---|---|---|---|---|---|---|
| **Country** | **Trial** | **LNS N** | **Control N** | **Control Prevalence** | **PD (95% CI)** | **Fixed W** | **Random W** | **LNS N** | **Control N** | **Control Prevalence** | **PD (95% CI)** | **Fixed W** | **Random W** |
| Bangladesh | JiVitA−4 (34) | | | | | | | | | | | | |
| Bangladesh | RDNS (35) | 730 | 335 | 39.7 | −0.06 (−0.12, 0.00) | 0.02 | 0.02 | 873 | 455 | 45.9 | −0.04 (−0.10, 0.01) | 0.11 | 0.14 |
| Bangladesh | WASH−B (36) | 519 | 1595 | 28.2 | 0.00 (−0.05, 0.04) | 0.04 | 0.04 | 644 | 1863 | 36.6 | −0.06 (−0.09, −0.02) | 0.21 | 0.20 |
| Burkina Faso | iLiNS−Zinc (37) | 1210 | 389 | 38.9 | −0.04 (−0.08, 0.00) | 0.05 | 0.05 | 734 | 275 | 44.0 | −0.05 (−0.12, 0.02) | 0.06 | 0.09 |
| Burkina Faso | PROMIS (38) | | | | | | | | | | | | |
| Burkina Faso | PROMIS CS (38) | | | | | | | | | | | | |
| Ghana | GHANA (39) | | | | | | | | | | | | |
| Ghana | iLiNS−DYADG (40) | 305 | 562 | 15.3 | −0.02 (−0.06, 0.03) | 0.03 | 0.03 | 37 | 119 | 20.2 | −0.04 (−0.19, 0.11) | 0.02 | 0.03 |
| Haiti | HAITI (41) | | | | | | | | | | | | |
| Kenya | WASH−B (42) | 1100 | 3831 | 4.0 | 0.00 (−0.02, 0.01) | 0.50 | 0.50 | 273 | 1083 | 5.3 | 0.01 (−0.02, 0.04) | 0.34 | 0.25 |
| Madagascar | MAHAY (43) | | | | | | | | | | | | |
| Malawi | iLiNS−DYADM (44) | 131 | 263 | 7.6 | 0.00 (−0.06, 0.06) | 0.03 | 0.03 | 88 | 178 | 14.6 | −0.03 (−0.12, 0.06) | 0.04 | 0.07 |
| Malawi | iLiNS−DOSE (45) | 512 | 172 | 17.3 | −0.02 (−0.08, 0.04) | 0.02 | 0.02 | 178 | 64 | 34.4 | −0.11 (−0.22, 0.00) | 0.03 | 0.04 |
| Mali | PROMIS (46) | | | | | | | | | | | | |
| Mali | PROMIS CS (46) | | | | | | | | | | | | |
| Zimbabwe | SHINE (HIV−) (47) | 1358 | 1298 | 5.9 | −0.01 (−0.03, 0.00) | 0.31 | 0.31 | 239 | 248 | 6.0 | 0.01 (−0.03, 0.05) | 0.18 | 0.18 |
| Zimbabwe | SHINE (HIV+) (48) | | | | | | | | | | | | |
| | | **5865** | **8445** | | **I² = 0.00, Tau² = 0.00** | | | **3066** | **4285** | | **I² = 0.41, Tau² = 0.00** | | |
| **Fixed** | | | | | **−0.01 (−0.02, 0.00)** | | | | | | **−0.02 (−0.04, 0.00)** | | |
| **Random** | | | | | **−0.01 (−0.02, 0.00)** | | | | | | **−0.03 (−0.05, 0.00)** | | |

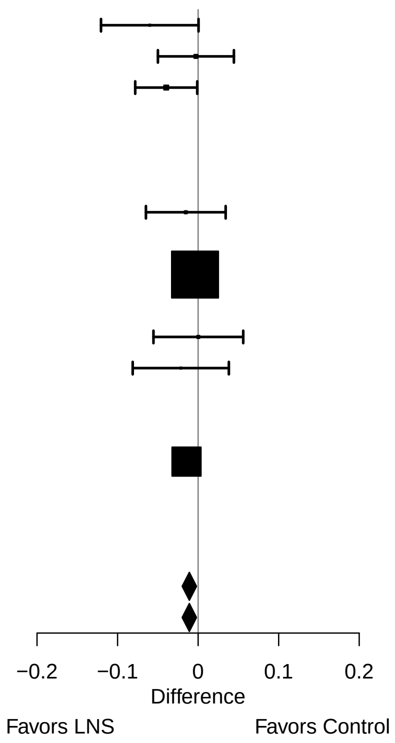

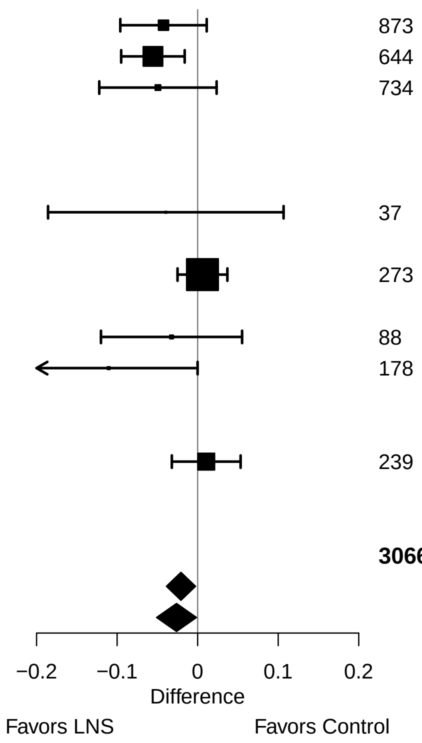

#### Supplemental figure 6Q: Small head size prevalence difference

#### 6Q3: Stratified by Maternal age

**P−for−interaction = 0.536**
**Difference in PDs = 0.00 (−0.02, 0.01)**

| | | At least 25 y | | | | | | Less than 25 y | | | | | |
|---|---|---|---|---|---|---|---|---|---|---|---|---|---|
| **Country** | **Trial** | **LNS N** | **Control N** | **Control Prevalence** | **PD (95% CI)** | **Fixed W** | **Random W** | **LNS N** | **Control N** | **Control Prevalence** | **PD (95% CI)** | **Fixed W** | **Random W** |
| Bangladesh | JiVitA−4 (34) | 1238 | 554 | 22.1 | 0.02 (−0.02, 0.05) | 0.07 | 0.11 | 1634 | 708 | 25.6 | −0.02 (−0.05, 0.02) | 0.12 | 0.14 |
| Bangladesh | RDNS (35) | 453 | 221 | 42.5 | −0.08 (−0.14, −0.01) | 0.02 | 0.05 | 1210 | 594 | 43.1 | −0.03 (−0.08, 0.02) | 0.06 | 0.10 |
| Bangladesh | WASH−B (36) | 492 | 1523 | 32.2 | −0.03 (−0.08, 0.02) | 0.04 | 0.08 | 666 | 1924 | 33.4 | −0.03 (−0.07, 0.01) | 0.08 | 0.12 |
| Burkina Faso | iLiNS−Zinc (37) | 1128 | 399 | 39.4 | −0.06 (−0.13, 0.01) | 0.02 | 0.05 | 805 | 262 | 43.1 | −0.01 (−0.06, 0.04) | 0.05 | 0.09 |
| Burkina Faso | PROMIS (38) | | | | | | | | | | | | |
| Burkina Faso | PROMIS CS (38) | | | | | | | | | | | | |
| Ghana | GHANA (39) | | | | | | | | | | | | |
| Ghana | iLiNS−DYADG (40) | 220 | 417 | 14.6 | −0.02 (−0.08, 0.04) | 0.03 | 0.06 | 127 | 275 | 18.2 | −0.02 (−0.10, 0.06) | 0.02 | 0.05 |
| Haiti | HAITI (41) | | | | | | | | | | | | |
| Kenya | WASH−B (42) | 845 | 2831 | 4.0 | 0.00 (−0.02, 0.01) | 0.48 | 0.26 | 601 | 2313 | 4.7 | 0.00 (−0.02, 0.02) | 0.37 | 0.19 |
| Madagascar | MAHAY (43) | | | | | | | | | | | | |
| Malawi | iLiNS−DYADM (44) | 106 | 226 | 9.3 | −0.03 (−0.09, 0.04) | 0.02 | 0.05 | 114 | 218 | 12.4 | 0.00 (−0.08, 0.07) | 0.02 | 0.05 |
| Malawi | iLiNS−DOSE (45) | 368 | 134 | 16.4 | 0.03 (−0.04, 0.10) | 0.02 | 0.05 | 310 | 97 | 29.6 | −0.16 (−0.24, −0.07) | 0.02 | 0.05 |
| Mali | PROMIS (46) | | | | | | | | | | | | |
| Mali | PROMIS CS (46) | | | | | | | | | | | | |
| Zimbabwe | SHINE (HIV−) (47) | 869 | 795 | 5.4 | −0.01 (−0.03, 0.01) | 0.26 | 0.22 | 805 | 792 | 6.3 | −0.01 (−0.04, 0.01) | 0.24 | 0.17 |
| Zimbabwe | SHINE (HIV+) (48) | 244 | 231 | 8.2 | 0.02 (−0.04, 0.07) | 0.03 | 0.07 | 73 | 78 | 14.1 | −0.03 (−0.12, 0.06) | 0.02 | 0.04 |
| | | **5963** | **7331** | | **I² = 0.21, Tau² = 0.00** | | | **6345** | **7261** | | **I² = 0.35, Tau² = 0.00** | | |
| **Fixed** | | | | | **−0.01 (−0.02, 0.00)** | | | | | | **−0.02 (−0.03, 0.00)** | | |
| **Random** | | | | | **−0.01 (−0.03, 0.01)** | | | | | | **−0.02 (−0.04, 0.00)** | | |

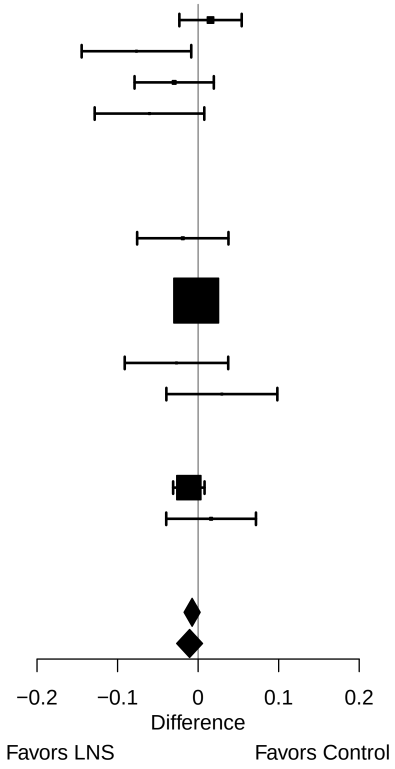

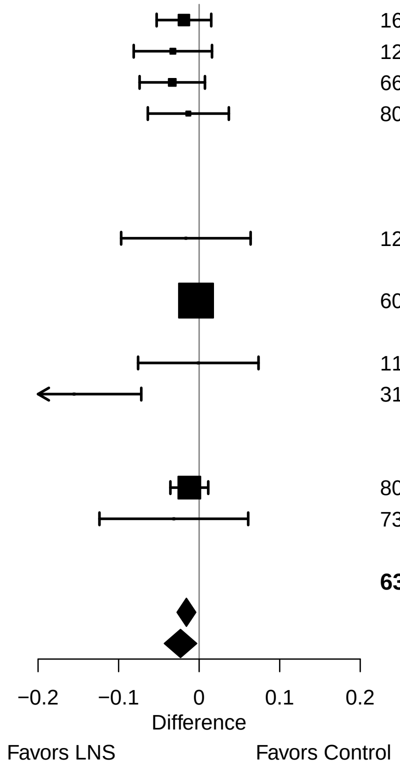

#### Supplemental figure 6Q: Small head size prevalence difference

#### 6Q4: Stratified by Maternal education

**P−for−interaction = 0.801**
**Difference in PDs = 0.00 (−0.02, 0.02)**

| Country | Trial | LNS N | Control N | Control Prevalence | Primary or greater PD (95% CI) | Fixed W | Random W | LNS N | Control N | Control Prevalence | Incomplete or no formal PD (95% CI) | Fixed W | Random W |
|---|---|---|---|---|---|---|---|---|---|---|---|---|---|
| Bangladesh | JiVitA−4 (34) | 1855 | 775 | 23.3 | −0.03 (−0.06, 0.00) | 0.15 | 0.20 | 1028 | 489 | 25.5 | 0.04 (−0.01, 0.08) | 0.08 | 0.17 |
| Bangladesh | RDNS (35) | 1245 | 589 | 41.6 | −0.05 (−0.10, −0.01) | 0.08 | 0.12 | 418 | 226 | 46.5 | −0.02 (−0.10, 0.06) | 0.03 | 0.07 |
| Bangladesh | WASH−B (36) | 817 | 2477 | 31.2 | −0.05 (−0.08, −0.01) | 0.13 | 0.17 | 346 | 986 | 36.7 | 0.00 (−0.06, 0.06) | 0.04 | 0.10 |
| Burkina Faso | iLiNS−Zinc (37) | 82 | 16 | 31.2 | 0.07 (−0.11, 0.24) | 0.01 | 0.01 | 1851 | 645 | 41.1 | −0.05 (−0.10, 0.00) | 0.07 | 0.15 |
| Burkina Faso | PROMIS (38) | | | | | | | | | | | | |
| Burkina Faso | PROMIS CS (38) | | | | | | | | | | | | |
| Ghana | GHANA (39) | | | | | | | | | | | | |
| Ghana | iLiNS−DYADG (40) | 267 | 548 | 15.3 | −0.02 (−0.07, 0.03) | 0.06 | 0.10 | 80 | 144 | 18.8 | −0.01 (−0.12, 0.09) | 0.01 | 0.04 |
| Haiti | HAITI (41) | | | | | | | | | | | | |
| Kenya | WASH−B (42) | 703 | 2456 | 2.8 | 0.00 (−0.01, 0.02) | 0.56 | 0.36 | 756 | 2716 | 5.7 | −0.01 (−0.03, 0.00) | 0.73 | 0.37 |
| Madagascar | MAHAY (43) | | | | | | | | | | | | |
| Malawi | iLiNS−DYADM (44) | | | | | | | | | | | | |
| Malawi | iLiNS−DOSE (45) | 163 | 55 | 16.1 | −0.02 (−0.12, 0.09) | 0.01 | 0.03 | 517 | 176 | 23.9 | −0.06 (−0.12, 0.00) | 0.04 | 0.11 |
| Mali | PROMIS (46) | | | | | | | | | | | | |
| Mali | PROMIS CS (46) | | | | | | | | | | | | |
| Zimbabwe | SHINE (HIV−) (47) | | | | | | | | | | | | |
| Zimbabwe | SHINE (HIV+) (48) | | | | | | | | | | | | |
| | | **5132** | **6916** | | **I² = 0.49, Tau² = 0.00** | | | **4996** | **5382** | | **I² = 0.33, Tau² = 0.00** | | |
| **Fixed** | | | | | **−0.01 (−0.03, 0.00)** | | | | | | **−0.01 (−0.02, 0.00)** | | |
| **Random** | | | | | **−0.02 (−0.04, 0.00)** | | | | | | **−0.01 (−0.04, 0.01)** | | |

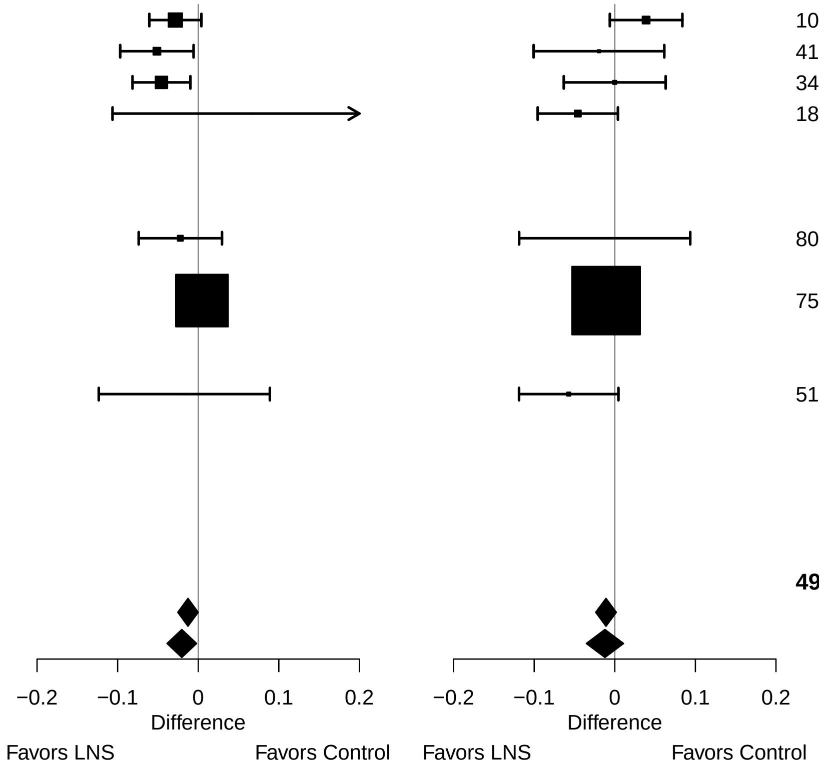

#### Supplemental figure 6Q: Small head size prevalence difference

#### 6Q5: Stratified by Maternal depressive symptoms

**P−for−interaction = 0.134**
**Difference in PDs = 0.01 (0.00, 0.03)**

| Country | Trial | Less than 75th percentile LNS N | Control N | Control Prevalence | PD (95% CI) | Fixed W | Random W | At least 75th percentile LNS N | Control N | Control Prevalence | PD (95% CI) | Fixed W | Random W |
|---|---|---|---|---|---|---|---|---|---|---|---|---|---|
| Bangladesh | JiVitA−4 (34) | | | | | | | | | | | | |
| Bangladesh | RDNS (35) | 1081 | 472 | 39.0 | 0.00 (−0.04, 0.04) | 0.06 | 0.09 | 520 | 288 | 49.3 | −0.13 (−0.20, −0.06) | 0.07 | 0.13 |
| Bangladesh | WASH−B (36) | 901 | 2487 | 33.0 | −0.05 (−0.08, −0.01) | 0.08 | 0.12 | 238 | 885 | 31.9 | 0.02 (−0.05, 0.08) | 0.08 | 0.14 |
| Burkina Faso | iLiNS−Zinc (37) | | | | | | | | | | | | |
| Burkina Faso | PROMIS (38) | | | | | | | | | | | | |
| Burkina Faso | PROMIS CS (38) | | | | | | | | | | | | |
| Ghana | GHANA (39) | | | | | | | | | | | | |
| Ghana | iLiNS−DYADG (40) | 252 | 459 | 15.0 | 0.00 (−0.05, 0.06) | 0.03 | 0.05 | 83 | 209 | 17.7 | −0.06 (−0.15, 0.04) | 0.04 | 0.09 |
| Haiti | HAITI (41) | | | | | | | | | | | | |
| Kenya | WASH−B (42) | 998 | 3517 | 4.0 | 0.00 (−0.02, 0.01) | 0.51 | 0.37 | 341 | 1248 | 5.4 | −0.02 (−0.05, 0.01) | 0.36 | 0.23 |
| Madagascar | MAHAY (43) | | | | | | | | | | | | |
| Malawi | iLiNS−DYADM (44) | 157 | 303 | 10.2 | −0.02 (−0.08, 0.04) | 0.03 | 0.05 | 47 | 113 | 11.5 | 0.03 (−0.08, 0.15) | 0.03 | 0.07 |
| Malawi | iLiNS−DOSE (45) | | | | | | | | | | | | |
| Mali | PROMIS (46) | | | | | | | | | | | | |
| Mali | PROMIS CS (46) | | | | | | | | | | | | |
| Zimbabwe | SHINE (HIV−) (47) | 1193 | 1135 | 5.0 | −0.01 (−0.02, 0.01) | 0.26 | 0.27 | 509 | 490 | 7.1 | −0.02 (−0.05, 0.01) | 0.40 | 0.23 |
| Zimbabwe | SHINE (HIV+) (48) | 234 | 215 | 8.8 | 0.01 (−0.04, 0.07) | 0.03 | 0.06 | 89 | 100 | 12.0 | −0.03 (−0.12, 0.06) | 0.04 | 0.10 |
| | | **4816** | **8588** | | **I² = 0.17, Tau² = 0.00** | | | **1827** | **3333** | | **I² = 0.48, Tau² = 0.00** | | |
| **Fixed** | | | | | **−0.01 (−0.02, 0.00)** | | | | | | **−0.02 (−0.04, −0.01)** | | |
| **Random** | | | | | **−0.01 (−0.02, 0.00)** | | | | | | **−0.03 (−0.06, 0.01)** | | |

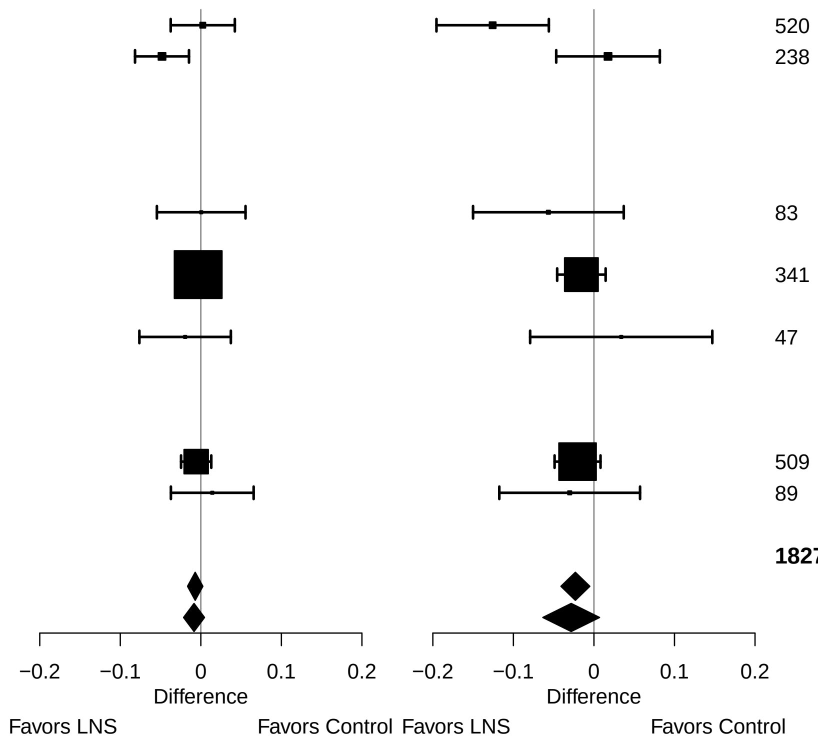

#### Supplemental figure 6Q: Small head size prevalence difference

### 6Q6: Stratified by Child sex

**P−for−interaction = 0.624**
**Difference in PDs = 0.00 (−0.02, 0.01)**

| Country | Trial | Male LNS N | Male Control N | Male Control Prevalence | Male PD (95% CI) | Male Fixed W | Male Random W | Female LNS N | Female Control N | Female Control Prevalence | Female PD (95% CI) | Female Fixed W | Female Random W |
|---|---|---|---|---|---|---|---|---|---|---|---|---|---|
| Bangladesh | JiVitA−4 (34) | 1475 | 612 | 25.8 | −0.02 (−0.05, 0.02) | 0.09 | 0.09 | 1410 | 655 | 22.4 | 0.01 (−0.03, 0.04) | 0.08 | 0.12 |
| Bangladesh | RDNS (35) | 833 | 411 | 37.7 | 0.03 (−0.02, 0.07) | 0.05 | 0.05 | 830 | 404 | 48.3 | −0.12 (−0.18, −0.05) | 0.02 | 0.08 |
| Bangladesh | WASH−B (36) | 566 | 1730 | 32.4 | −0.02 (−0.07, 0.02) | 0.06 | 0.06 | 597 | 1733 | 33.2 | −0.04 (−0.08, 0.00) | 0.05 | 0.11 |
| Burkina Faso | iLiNS−Zinc (37) | 991 | 335 | 42.3 | −0.02 (−0.09, 0.05) | 0.02 | 0.02 | 955 | 329 | 39.7 | −0.07 (−0.12, −0.02) | 0.04 | 0.10 |
| Burkina Faso | PROMIS (38) | | | | | | | | | | | | |
| Burkina Faso | PROMIS CS (38) | | | | | | | | | | | | |
| Ghana | GHANA (39) | | | | | | | | | | | | |
| Ghana | iLiNS−DYADG (40) | 175 | 323 | 19.2 | −0.03 (−0.10, 0.04) | 0.02 | 0.02 | 172 | 369 | 13.3 | −0.02 (−0.08, 0.04) | 0.03 | 0.08 |
| Haiti | HAITI (41) | | | | | | | | | | | | |
| Kenya | WASH−B (42) | 736 | 2466 | 4.9 | −0.01 (−0.02, 0.01) | 0.40 | 0.40 | 725 | 2710 | 3.8 | 0.00 (−0.02, 0.02) | 0.37 | 0.14 |
| Madagascar | MAHAY (43) | | | | | | | | | | | | |
| Malawi | iLiNS−DYADM (44) | 107 | 207 | 13.0 | −0.02 (−0.10, 0.06) | 0.02 | 0.02 | 113 | 237 | 8.9 | −0.01 (−0.07, 0.05) | 0.03 | 0.08 |
| Malawi | iLiNS−DOSE (45) | 349 | 123 | 23.6 | −0.01 (−0.09, 0.07) | 0.02 | 0.02 | 343 | 115 | 19.8 | −0.08 (−0.15, −0.01) | 0.02 | 0.07 |
| Mali | PROMIS (46) | | | | | | | | | | | | |
| Mali | PROMIS CS (46) | | | | | | | | | | | | |
| Zimbabwe | SHINE (HIV−) (47) | 927 | 903 | 6.4 | −0.01 (−0.03, 0.01) | 0.30 | 0.30 | 944 | 880 | 5.1 | −0.01 (−0.03, 0.00) | 0.34 | 0.14 |
| Zimbabwe | SHINE (HIV+) (48) | 162 | 168 | 14.3 | −0.04 (−0.11, 0.03) | 0.02 | 0.02 | 175 | 161 | 5.6 | 0.04 (−0.02, 0.09) | 0.03 | 0.09 |
| | | **6321** | **7278** | | **$I^2$ = 0.00, $Tau^2$ = 0.00** | | | **6264** | **7593** | | **$I^2$ = 0.64, $Tau^2$ = 0.00** | | |
| **Fixed** | | | | | **−0.01 (−0.02, 0.00)** | | | | | | **−0.01 (−0.02, 0.00)** | | |
| **Random** | | | | | **−0.01 (−0.02, 0.00)** | | | | | | **−0.03 (−0.05, 0.00)** | | |

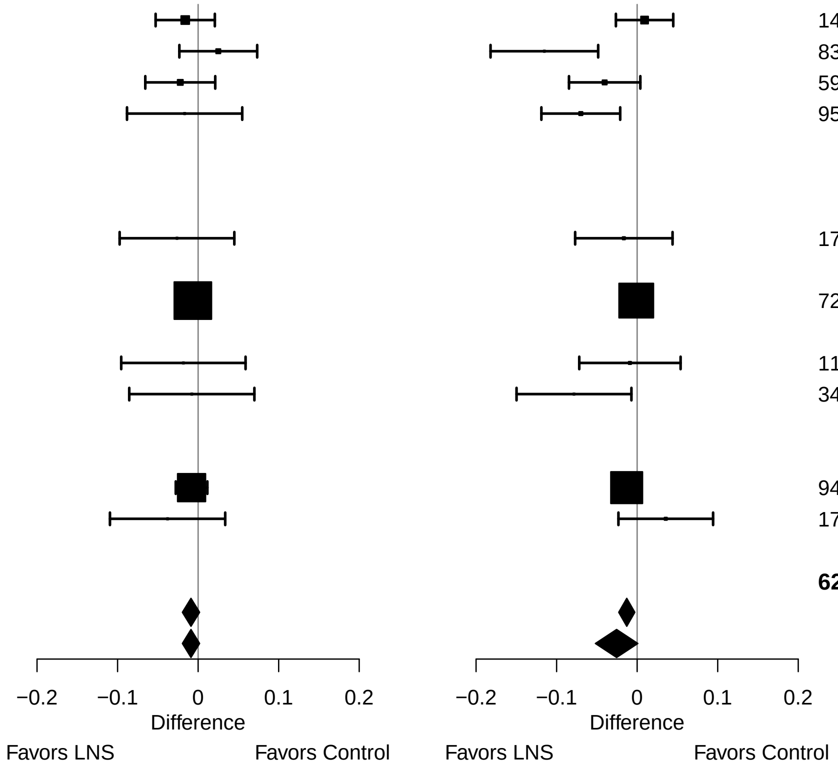

#### Supplemental figure 6Q: Small head size prevalence difference

#### 6Q7: Stratified by Child birth order

**P−for−interaction = 0.402**
**Difference in PDs = −0.01 (−0.02, 0.01)**

| Country | Trial | Later born LNS N | Later born Control N | Later born Control Prevalence | Later born PD (95% CI) | Later born Fixed W | Later born Random W | Firstborn LNS N | Firstborn Control N | Firstborn Control Prevalence | Firstborn PD (95% CI) | Firstborn Fixed W | Firstborn Random W |
|---|---|---|---|---|---|---|---|---|---|---|---|---|---|
| Bangladesh | JiVitA−4 (34) | 616 | 260 | 25.0 | 0.00 (−0.05, 0.05) | 0.03 | 0.05 | 2265 | 1000 | 23.9 | 0.00 (−0.04, 0.03) | 0.18 | 0.18 |
| Bangladesh | RDNS (35) | 974 | 508 | 43.1 | −0.06 (−0.10, −0.01) | 0.04 | 0.06 | 688 | 307 | 42.7 | −0.03 (−0.09, 0.04) | 0.04 | 0.04 |
| Bangladesh | WASH−B (36) | 736 | 2313 | 31.9 | −0.03 (−0.07, 0.00) | 0.06 | 0.09 | 427 | 1150 | 34.6 | −0.03 (−0.09, 0.02) | 0.06 | 0.06 |
| Burkina Faso | iLiNS−Zinc (37) | 1515 | 519 | 40.3 | −0.05 (−0.10, 0.01) | 0.02 | 0.04 | 431 | 145 | 43.4 | −0.04 (−0.11, 0.04) | 0.03 | 0.03 |
| Burkina Faso | PROMIS (38) | | | | | | | | | | | | |
| Burkina Faso | PROMIS CS (38) | | | | | | | | | | | | |
| Ghana | GHANA (39) | | | | | | | | | | | | |
| Ghana | iLiNS−DYADG (40) | 237 | 458 | 14.4 | 0.00 (−0.06, 0.05) | 0.03 | 0.04 | 110 | 234 | 19.2 | −0.05 (−0.13, 0.04) | 0.02 | 0.02 |
| Haiti | HAITI (41) | | | | | | | | | | | | |
| Kenya | WASH−B (42) | 1137 | 4074 | 4.3 | 0.00 (−0.02, 0.01) | 0.41 | 0.33 | 322 | 1097 | 4.3 | −0.01 (−0.03, 0.01) | 0.48 | 0.48 |
| Madagascar | MAHAY (43) | | | | | | | | | | | | |
| Malawi | iLiNS−DYADM (44) | 173 | 353 | 10.8 | −0.03 (−0.08, 0.03) | 0.03 | 0.04 | 47 | 89 | 10.1 | 0.05 (−0.07, 0.16) | 0.01 | 0.01 |
| Malawi | iLiNS−DOSE (45) | 458 | 162 | 20.2 | −0.01 (−0.08, 0.05) | 0.02 | 0.03 | 146 | 45 | 26.7 | −0.13 (−0.25, −0.01) | 0.01 | 0.01 |
| Mali | PROMIS (46) | | | | | | | | | | | | |
| Mali | PROMIS CS (46) | | | | | | | | | | | | |
| Zimbabwe | SHINE (HIV−) (47) | 1295 | 1214 | 5.0 | −0.01 (−0.02, 0.00) | 0.37 | 0.31 | 489 | 486 | 7.8 | −0.02 (−0.05, 0.02) | 0.15 | 0.15 |
| Zimbabwe | SHINE (HIV+) (48) | | | | | | | | | | | | |
| | | **7141** | **9861** | | **I² = 0.14, Tau² = 0.00** | | | **4925** | **4553** | | **I² = 0.00, Tau² = 0.00** | | |
| **Fixed** | | | | | **−0.01 (−0.02, 0.00)** | | | | | | **−0.02 (−0.03, 0.00)** | | |
| **Random** | | | | | **−0.01 (−0.03, 0.00)** | | | | | | **−0.02 (−0.03, 0.00)** | | |

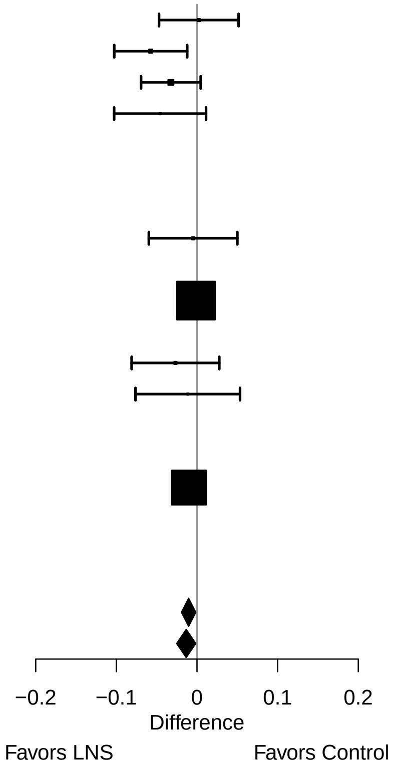

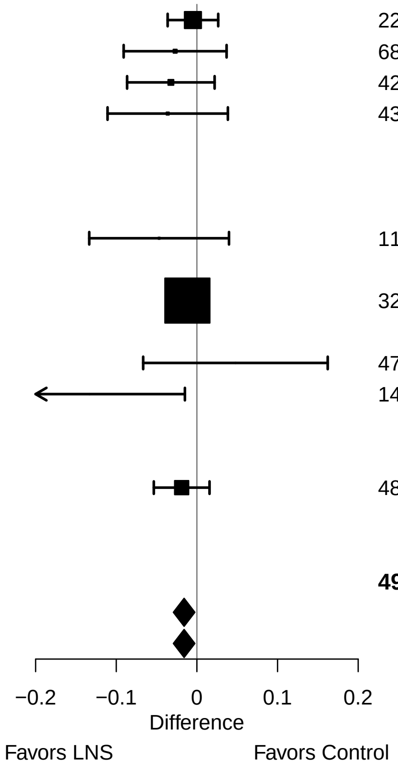

#### Supplemental figure 6Q: Small head size prevalence difference

#### 6Q8: Stratified by Child baseline anthropometric status

**P−for−interaction = 0.069**
**Difference in PDs = −0.02 (−0.05, 0.00)**

| Country | Trial | HCZ at least −1 Z: LNS N | Control N | Control Prevalence | PD (95% CI) | Fixed W | Random W | HCZ less than −1 Z: LNS N | Control N | Control Prevalence | PD (95% CI) | Fixed W | Random W |
|---|---|---|---|---|---|---|---|---|---|---|---|---|---|
| Bangladesh | JiVitA−4 (34) | 1411 | 630 | 2.7 | 0.00 (−0.02, 0.02) | 0.21 | 0.21 | 1474 | 637 | 45.4 | −0.02 (−0.07, 0.03) | 0.27 | 0.27 |
| Bangladesh | RDNS (35) | 521 | 239 | 5.0 | −0.03 (−0.05, 0.00) | 0.10 | 0.14 | 1080 | 521 | 60.3 | −0.05 (−0.09, 0.00) | 0.29 | 0.29 |
| Bangladesh | WASH−B (36) | | | | | | | | | | | | |
| Burkina Faso | iLiNS−Zinc (37) | 550 | 176 | 5.1 | −0.04 (−0.07, −0.01) | 0.08 | 0.12 | 1396 | 488 | 54.1 | −0.06 (−0.13, 0.02) | 0.12 | 0.12 |
| Burkina Faso | PROMIS (38) | | | | | | | | | | | | |
| Burkina Faso | PROMIS CS (38) | | | | | | | | | | | | |
| Ghana | GHANA (39) | | | | | | | | | | | | |
| Ghana | iLiNS−DYADG (40) | 189 | 384 | 2.3 | 0.01 (−0.02, 0.04) | 0.09 | 0.13 | 134 | 257 | 35.4 | −0.04 (−0.14, 0.06) | 0.07 | 0.07 |
| Haiti | HAITI (41) | | | | | | | | | | | | |
| Kenya | WASH−B (42) | | | | | | | | | | | | |
| Madagascar | MAHAY (43) | | | | | | | | | | | | |
| Malawi | iLiNS−DYADM (44) | | | | | | | | | | | | |
| Malawi | iLiNS−DOSE (45) | 381 | 132 | 7.6 | −0.01 (−0.06, 0.04) | 0.03 | 0.05 | 311 | 106 | 39.6 | −0.09 (−0.20, 0.01) | 0.06 | 0.06 |
| Mali | PROMIS (46) | | | | | | | | | | | | |
| Mali | PROMIS CS (46) | | | | | | | | | | | | |
| Zimbabwe | SHINE (HIV−) (47) | 1072 | 967 | 2.8 | −0.01 (−0.02, 0.01) | 0.43 | 0.26 | 254 | 239 | 15.5 | −0.01 (−0.07, 0.05) | 0.17 | 0.17 |
| Zimbabwe | SHINE (HIV+) (48) | 200 | 168 | 4.2 | −0.01 (−0.04, 0.03) | 0.05 | 0.09 | 71 | 67 | 28.4 | −0.07 (−0.24, 0.09) | 0.02 | 0.02 |
| | | **4324** | **2696** | | **I² = 0.31, Tau² = 0.00** | | | **4720** | **2315** | | **I² = 0.00, Tau² = 0.00** | | |
| **Fixed** | | | | | **−0.01 (−0.02, 0.00)** | | | | | | **−0.04 (−0.06, −0.01)** | | |
| **Random** | | | | | **−0.01 (−0.02, 0.00)** | | | | | | **−0.04 (−0.06, −0.01)** | | |

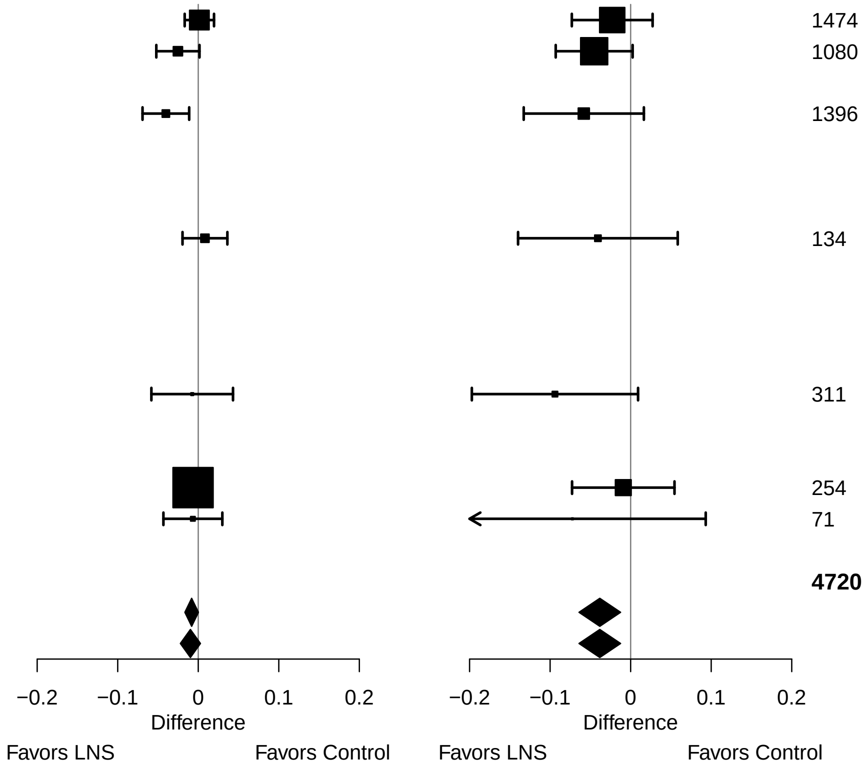

### Supplemental figure 7: Forest plots for effects of SQ-LNS on growth outcomes stratified by individual-level household effect modifiers

These figures are forest plots showing the individual-level effect modification of intervention effects. Each figure has the estimates of intervention effect stratified within study by individual-level effect modifier category. For continuous outcomes analyzed via mean differences, the effect estimate is the mean in the LNS group minus the mean in the control group. For dichotomous outcomes analyzed via prevalence ratios, the effect estimate is the prevalence in the LNS group divided by the prevalence in the control group. For dichotomous outcomes analyzed via prevalence differences, the effect estimate is the prevalence in the LNS group minus the prevalence in the control group. The labels on the far left correspond to trial level information. In the middle left and on the right the values indicate the study level effect estimate, confidence interval, and weighting for deriving the pooled estimates is shown by subgroup. LAZ, length-for-age z-score; WLZ, weight-for-length z-score; WAZ, weight-for-age z-score; MUACZ, mid-upper arm circumference z-score; HCZ, head circumference-for-age z-score.

### Supplemental figure 7A: Mean difference in LAZ

#### 7A1: Stratified by Household socio-economic status

**P−for−interaction = 0.805**
**Difference in MDs = 0.01 (−0.03, 0.04)**

| | | At least median | | | | | | Less than median | | | | | |
|---|---|---|---|---|---|---|---|---|---|---|---|---|---|
| **Country** | **Trial** | **LNS N** | **Control N** | **Control Mean** | **MD (95% CI)** | **Fixed W** | **Random W** | **LNS N** | **Control N** | **Control Mean** | **MD (95% CI)** | **Fixed W** | **Random W** |
| Bangladesh | JiVitA−4 (34) | 1429 | 621 | −1.74 | 0.08 (0.02, 0.15) | 0.23 | 0.10 | 1408 | 623 | −2.07 | 0.09 (0.02, 0.15) | 0.22 | 0.11 |
| Bangladesh | RDNS (35) | 834 | 407 | −1.69 | 0.11 (0.00, 0.22) | 0.08 | 0.08 | 829 | 408 | −1.93 | 0.04 (−0.05, 0.13) | 0.11 | 0.09 |
| Bangladesh | WASH−B (36) | 579 | 1716 | −1.60 | 0.19 (0.09, 0.30) | 0.08 | 0.08 | 579 | 1715 | −2.03 | 0.23 (0.14, 0.32) | 0.13 | 0.09 |
| Burkina Faso | iLiNS−Zinc (37) | 1225 | 299 | −1.77 | 0.28 (0.18, 0.37) | 0.10 | 0.09 | 720 | 364 | −1.74 | 0.33 (0.23, 0.43) | 0.10 | 0.09 |
| Burkina Faso | PROMIS (38) | 407 | 482 | −1.50 | 0.22 (0.03, 0.41) | 0.03 | 0.05 | 456 | 432 | −1.47 | 0.08 (−0.06, 0.22) | 0.05 | 0.06 |
| Burkina Faso | PROMIS CS (38) | 220 | 210 | −1.15 | −0.04 (−0.29, 0.20) | 0.02 | 0.03 | 210 | 229 | −1.29 | 0.12 (−0.13, 0.36) | 0.02 | 0.03 |
| Ghana | GHANA (39) | 57 | 44 | −0.18 | 0.21 (−0.21, 0.63) | 0.01 | 0.01 | 41 | 46 | −0.51 | 0.16 (−0.28, 0.60) | 0.00 | 0.01 |
| Ghana | iLiNS−DYADG (40) | 157 | 356 | −0.76 | 0.08 (−0.11, 0.26) | 0.03 | 0.05 | 190 | 334 | −1.03 | 0.33 (0.14, 0.51) | 0.03 | 0.05 |
| Haiti | HAITI (41) | 75 | 60 | −0.68 | −0.04 (−0.23, 0.16) | 0.02 | 0.04 | 61 | 72 | −0.90 | 0.04 (−0.15, 0.24) | 0.03 | 0.04 |
| Kenya | WASH−B (42) | 825 | 2921 | −1.47 | 0.20 (0.12, 0.28) | 0.15 | 0.09 | 630 | 2212 | −1.70 | 0.09 (−0.02, 0.20) | 0.08 | 0.08 |
| Madagascar | MAHAY (43) | 771 | 884 | −2.22 | −0.04 (−0.22, 0.13) | 0.03 | 0.05 | 878 | 755 | −2.26 | 0.09 (−0.06, 0.24) | 0.04 | 0.06 |
| Malawi | iLiNS−DYADM (44) | 121 | 234 | −1.49 | −0.05 (−0.28, 0.18) | 0.02 | 0.04 | 99 | 209 | −1.77 | −0.14 (−0.39, 0.12) | 0.01 | 0.03 |
| Malawi | iLiNS−DOSE (45) | 301 | 104 | −1.72 | −0.01 (−0.16, 0.15) | 0.04 | 0.06 | 294 | 103 | −2.14 | 0.07 (−0.09, 0.23) | 0.04 | 0.06 |
| Mali | PROMIS (46) | 247 | 251 | −1.72 | 0.22 (0.07, 0.37) | 0.04 | 0.06 | 246 | 246 | −1.74 | 0.12 (−0.09, 0.34) | 0.02 | 0.04 |
| Mali | PROMIS CS (46) | 505 | 463 | −1.57 | 0.26 (0.09, 0.44) | 0.03 | 0.05 | 447 | 506 | −1.57 | 0.18 (−0.05, 0.41) | 0.02 | 0.04 |
| Zimbabwe | SHINE (HIV−) (47) | 908 | 802 | −1.52 | 0.15 (0.05, 0.26) | 0.08 | 0.08 | 832 | 860 | −1.70 | 0.18 (0.08, 0.28) | 0.09 | 0.09 |
| Zimbabwe | SHINE (HIV+) (48) | 171 | 156 | −1.93 | 0.23 (−0.02, 0.48) | 0.01 | 0.03 | 158 | 167 | −2.06 | 0.25 (−0.02, 0.53) | 0.01 | 0.03 |
| | | **8832** | **10010** | | **I² = 0.55, Tau² = 0.01** | | | **8078** | **9281** | | **I² = 0.58, Tau² = 0.01** | | |
| **Fixed** | | | | | **0.14 (0.11, 0.17)** | | | | | | **0.14 (0.11, 0.17)** | | |
| **Random** | | | | | **0.13 (0.08, 0.18)** | | | | | | **0.14 (0.09, 0.19)** | | |

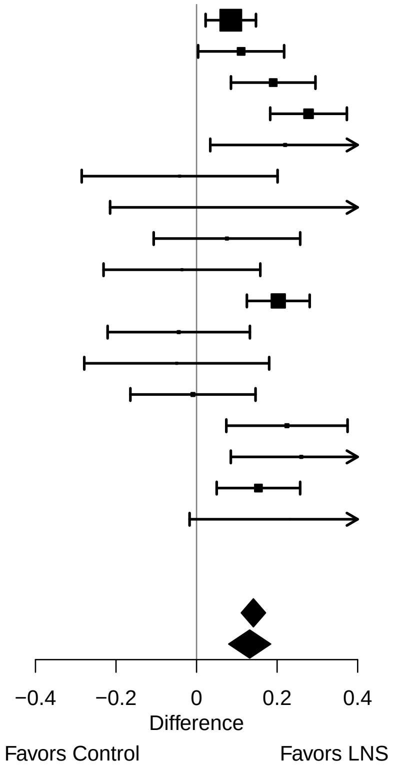

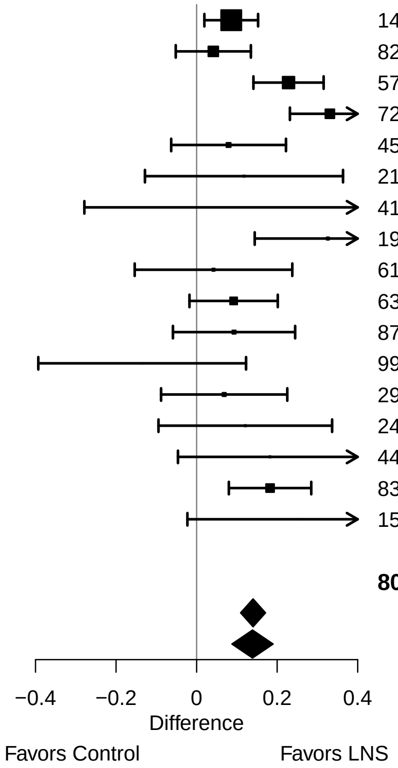

#### Supplemental figure 7A: Mean difference in LAZ

#### 7A2: Stratified by Household food insecurity

**P−for−interaction = 0.943**
**Difference in MDs = 0.00 (−0.04, 0.05)**

| | | Mild to secure | | | | | | Moderate to severe | | | | | |
|---|---|---|---|---|---|---|---|---|---|---|---|---|---|
| **Country** | **Trial** | **LNS N** | **Control N** | **Control Mean** | **MD (95% CI)** | **Fixed W** | **Random W** | **LNS N** | **Control N** | **Control Mean** | **MD (95% CI)** | **Fixed W** | **Random W** |
| Bangladesh | JiVitA−4 (34) | 2012 | 863 | −1.82 | 0.09 (0.03, 0.14) | 0.24 | 0.11 | 825 | 381 | −2.09 | 0.08 (−0.01, 0.16) | 0.23 | 0.13 |
| Bangladesh | RDNS (35) | 1058 | 493 | −1.71 | 0.03 (−0.07, 0.13) | 0.08 | 0.09 | 605 | 322 | −1.96 | 0.14 (0.02, 0.25) | 0.13 | 0.11 |
| Bangladesh | WASH−B (36) | 918 | 2652 | −1.76 | 0.23 (0.16, 0.31) | 0.13 | 0.10 | 240 | 779 | −2.01 | 0.09 (−0.05, 0.23) | 0.09 | 0.09 |
| Burkina Faso | iLiNS−Zinc (37) | 1013 | 320 | −1.77 | 0.34 (0.25, 0.44) | 0.09 | 0.09 | 932 | 343 | −1.74 | 0.27 (0.17, 0.37) | 0.16 | 0.12 |
| Burkina Faso | PROMIS (38) | 502 | 550 | −1.43 | 0.11 (−0.03, 0.25) | 0.04 | 0.07 | 361 | 364 | −1.56 | 0.21 (0.02, 0.39) | 0.05 | 0.07 |
| Burkina Faso | PROMIS CS (38) | | | | | | | | | | | | |
| Ghana | GHANA (39) | | | | | | | | | | | | |
| Ghana | iLiNS−DYADG (40) | 244 | 470 | −0.87 | 0.27 (0.12, 0.42) | 0.03 | 0.07 | 102 | 219 | −0.95 | 0.04 (−0.20, 0.28) | 0.03 | 0.05 |
| Haiti | HAITI (41) | | | | | | | | | | | | |
| Kenya | WASH−B (42) | 1299 | 4595 | −1.55 | 0.15 (0.09, 0.22) | 0.19 | 0.11 | 156 | 525 | −1.75 | 0.17 (−0.05, 0.40) | 0.03 | 0.05 |
| Madagascar | MAHAY (43) | 988 | 927 | −2.25 | 0.02 (−0.15, 0.18) | 0.03 | 0.06 | 355 | 410 | −2.24 | 0.04 (−0.17, 0.25) | 0.04 | 0.06 |
| Malawi | iLiNS−DYADM (44) | 70 | 122 | −1.45 | −0.04 (−0.37, 0.29) | 0.01 | 0.03 | 148 | 316 | −1.68 | −0.12 (−0.32, 0.08) | 0.04 | 0.06 |
| Malawi | iLiNS−DOSE (45) | 151 | 55 | −1.73 | −0.03 (−0.22, 0.17) | 0.02 | 0.05 | 431 | 146 | −2.03 | 0.05 (−0.08, 0.19) | 0.09 | 0.10 |
| Mali | PROMIS (46) | 329 | 341 | −1.69 | 0.13 (−0.03, 0.29) | 0.03 | 0.06 | 174 | 163 | −1.81 | 0.24 (0.06, 0.43) | 0.05 | 0.07 |
| Mali | PROMIS CS (46) | | | | | | | | | | | | |
| Zimbabwe | SHINE (HIV−) (47) | 1393 | 1305 | −1.59 | 0.15 (0.06, 0.23) | 0.11 | 0.10 | 296 | 325 | −1.74 | 0.28 (0.10, 0.45) | 0.06 | 0.07 |
| Zimbabwe | SHINE (HIV+) (48) | 236 | 239 | −1.96 | 0.25 (0.05, 0.46) | 0.02 | 0.05 | 86 | 76 | −2.17 | 0.27 (−0.10, 0.64) | 0.01 | 0.02 |
| | | **10213** | **12932** | | **I² = 0.72, Tau² = 0.01** | | | **4711** | **4369** | | **I² = 0.47, Tau² = 0.01** | | |
| **Fixed** | | | | | **0.15 (0.12, 0.18)** | | | | | | **0.13 (0.09, 0.18)** | | |
| **Random** | | | | | **0.14 (0.08, 0.20)** | | | | | | **0.13 (0.07, 0.19)** | | |

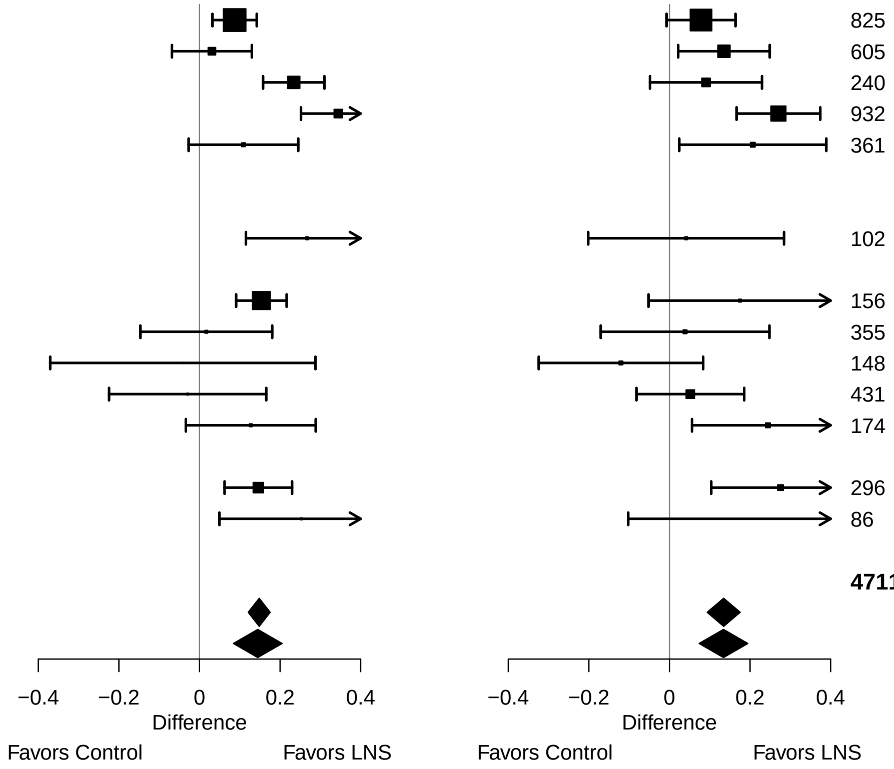

#### Supplemental figure 7A: Mean difference in LAZ

##### 7A3: Stratified by Household source water quality

**P−for−interaction = 0.169**
**Difference in MDs = 0.04 (−0.02, 0.11)**

| Country | Trial | LNS N | Control N | Control Mean | Improved MD (95% CI) | Fixed W | Random W | LNS N | Control N | Control Mean | Unimproved MD (95% CI) | Fixed W | Random W |
|---|---|---|---|---|---|---|---|---|---|---|---|---|---|
| Bangladesh | JiVitA−4 (34) | | | | | | | | | | | | |
| Bangladesh | RDNS (35) | | | | | | | | | | | | |
| Bangladesh | WASH−B (36) | 496 | 989 | −1.77 | 0.28 (0.18, 0.39) | 0.16 | 0.12 | 70 | 114 | −1.96 | 0.11 (−0.20, 0.42) | 0.03 | 0.04 |
| Burkina Faso | iLiNS−Zinc (37) | 550 | 141 | −1.68 | 0.21 (0.12, 0.30) | 0.21 | 0.13 | 1394 | 519 | −1.77 | 0.33 (0.24, 0.43) | 0.33 | 0.21 |
| Burkina Faso | PROMIS (38) | 355 | 379 | −1.49 | 0.12 (−0.03, 0.26) | 0.08 | 0.09 | 361 | 355 | −1.50 | 0.25 (0.01, 0.48) | 0.05 | 0.07 |
| Burkina Faso | PROMIS CS (38) | 280 | 268 | −1.25 | 0.09 (−0.13, 0.31) | 0.04 | 0.06 | 149 | 171 | −1.18 | −0.06 (−0.32, 0.21) | 0.04 | 0.06 |
| Ghana | GHANA (39) | | | | | | | | | | | | |
| Ghana | iLiNS−DYADG (40) | | | | | | | | | | | | |
| Haiti | HAITI (41) | | | | | | | | | | | | |
| Kenya | WASH−B (42) | 447 | 1549 | −1.52 | 0.10 (−0.03, 0.22) | 0.11 | 0.11 | 248 | 697 | −1.58 | 0.13 (−0.03, 0.29) | 0.12 | 0.13 |
| Madagascar | MAHAY (43) | 419 | 453 | −2.05 | 0.03 (−0.19, 0.24) | 0.04 | 0.06 | 1203 | 1180 | −2.31 | 0.04 (−0.11, 0.18) | 0.14 | 0.14 |
| Malawi | iLiNS−DYADM (44) | 200 | 406 | −1.62 | −0.10 (−0.28, 0.08) | 0.05 | 0.07 | 20 | 36 | −1.63 | 0.08 (−0.59, 0.76) | 0.01 | 0.01 |
| Malawi | iLiNS−DOSE (45) | 555 | 193 | −1.89 | 0.03 (−0.08, 0.14) | 0.13 | 0.11 | 52 | 17 | −2.29 | 0.08 (−0.33, 0.48) | 0.02 | 0.03 |
| Mali | PROMIS (46) | 249 | 258 | −1.68 | 0.17 (0.00, 0.34) | 0.06 | 0.08 | 247 | 243 | −1.76 | 0.16 (−0.02, 0.33) | 0.10 | 0.11 |
| Mali | PROMIS CS (46) | 549 | 566 | −1.51 | 0.17 (−0.03, 0.38) | 0.04 | 0.06 | 362 | 388 | −1.66 | 0.27 (0.04, 0.50) | 0.06 | 0.07 |
| Zimbabwe | SHINE (HIV−) (47) | 521 | 489 | −1.62 | 0.15 (0.00, 0.30) | 0.08 | 0.09 | 274 | 302 | −1.56 | 0.07 (−0.11, 0.25) | 0.09 | 0.11 |
| Zimbabwe | SHINE (HIV+) (48) | 85 | 87 | −2.05 | 0.34 (−0.03, 0.71) | 0.01 | 0.02 | 55 | 56 | −2.03 | 0.18 (−0.24, 0.61) | 0.02 | 0.02 |
| | | **4706** | **5778** | | **I² = 0.52, Tau² = 0.01** | | | **4435** | **4078** | | **I² = 0.45, Tau² = 0.00** | | |
| **Fixed** | | | | | **0.14 (0.10, 0.18)** | | | | | | **0.19 (0.13, 0.24)** | | |
| **Random** | | | | | **0.13 (0.07, 0.19)** | | | | | | **0.16 (0.09, 0.23)** | | |

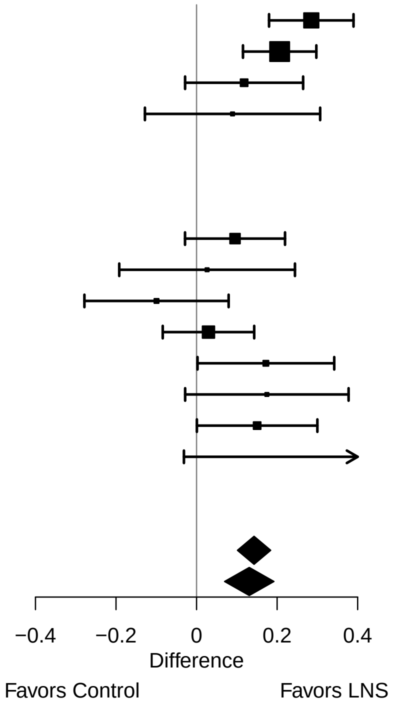

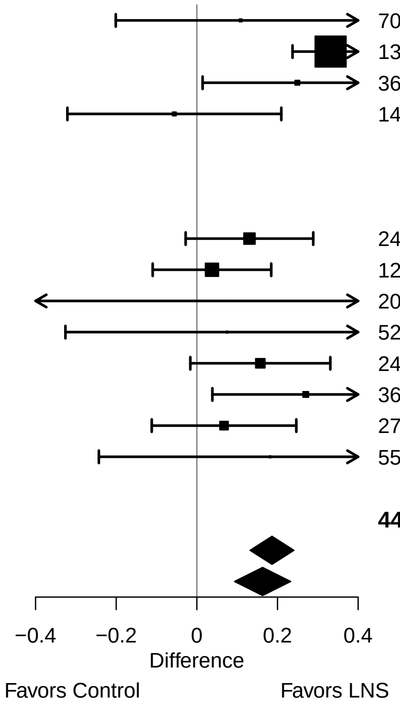

#### Supplemental figure 7A: Mean difference in LAZ

#### 7A4: Stratified by Household sanitation

**P−for−interaction = 0.135**
**Difference in MDs = −0.04 (−0.10, 0.01)**

| | | Improved | | | | | | Unimproved | | | | | |
|---|---|---|---|---|---|---|---|---|---|---|---|---|---|
| **Country** | **Trial** | **LNS N** | **Control N** | **Control Mean** | **MD (95% CI)** | **Fixed W** | **Random W** | **LNS N** | **Control N** | **Control Mean** | **MD (95% CI)** | **Fixed W** | **Random W** |
| Bangladesh | JiVitA−4 (34) | 2189 | 950 | −1.84 | 0.08 (0.03, 0.14) | 0.42 | 0.36 | 648 | 294 | −2.10 | 0.09 (0.00, 0.19) | 0.17 | 0.10 |
| Bangladesh | RDNS (35) | 1179 | 573 | −1.75 | 0.11 (0.02, 0.19) | 0.17 | 0.17 | 482 | 241 | −1.95 | 0.01 (−0.14, 0.15) | 0.06 | 0.08 |
| Bangladesh | WASH−B (36) | 506 | 1001 | −1.76 | 0.27 (0.15, 0.39) | 0.09 | 0.10 | 31 | 54 | −2.03 | 0.37 (0.00, 0.73) | 0.01 | 0.03 |
| Burkina Faso | iLiNS−Zinc (37) | 43 | 16 | −1.84 | 0.18 (−0.11, 0.46) | 0.02 | 0.02 | 1901 | 644 | −1.75 | 0.31 (0.23, 0.39) | 0.21 | 0.11 |
| Burkina Faso | PROMIS (38) | 398 | 550 | −1.49 | 0.20 (0.05, 0.35) | 0.05 | 0.06 | 465 | 359 | −1.47 | 0.10 (−0.04, 0.24) | 0.07 | 0.09 |
| Burkina Faso | PROMIS CS (38) | 176 | 165 | −1.29 | 0.18 (−0.05, 0.42) | 0.02 | 0.03 | 252 | 274 | −1.19 | −0.06 (−0.27, 0.14) | 0.03 | 0.07 |
| Ghana | GHANA (39) | | | | | | | | | | | | |
| Ghana | iLiNS−DYADG (40) | 336 | 671 | −0.88 | 0.17 (0.04, 0.30) | 0.07 | 0.08 | 10 | 19 | −1.29 | 0.96 (0.17, 1.75) | 0.00 | 0.01 |
| Haiti | HAITI (41) | | | | | | | | | | | | |
| Kenya | WASH−B (42) | 100 | 364 | −1.33 | 0.14 (−0.09, 0.36) | 0.02 | 0.03 | 595 | 1883 | −1.58 | 0.10 (−0.01, 0.22) | 0.11 | 0.09 |
| Madagascar | MAHAY (43) | | | | | | | | | | | | |
| Malawi | iLiNS−DYADM (44) | 20 | 42 | −1.35 | 0.20 (−0.37, 0.77) | 0.00 | 0.00 | 200 | 400 | −1.65 | −0.11 (−0.29, 0.07) | 0.04 | 0.07 |
| Malawi | iLiNS−DOSE (45) | 13 | 9 | −1.54 | 0.06 (−0.55, 0.66) | 0.00 | 0.00 | 593 | 201 | −1.94 | 0.03 (−0.08, 0.15) | 0.12 | 0.10 |
| Mali | PROMIS (46) | 387 | 366 | −1.69 | 0.20 (0.03, 0.36) | 0.05 | 0.05 | 112 | 135 | −1.82 | 0.09 (−0.10, 0.28) | 0.04 | 0.07 |
| Mali | PROMIS CS (46) | 701 | 696 | −1.56 | 0.26 (0.08, 0.45) | 0.04 | 0.04 | 220 | 248 | −1.60 | 0.18 (−0.05, 0.41) | 0.03 | 0.06 |
| Zimbabwe | SHINE (HIV−) (47) | 295 | 244 | −1.51 | 0.14 (−0.03, 0.30) | 0.05 | 0.05 | 497 | 546 | −1.64 | 0.10 (−0.03, 0.22) | 0.09 | 0.09 |
| Zimbabwe | SHINE (HIV+) (48) | 50 | 35 | −2.11 | 0.46 (−0.02, 0.93) | 0.01 | 0.01 | 90 | 107 | −2.02 | 0.19 (−0.16, 0.53) | 0.01 | 0.04 |
| | | **6393** | **5682** | | **I² = 0.13, Tau² = 0.00** | | | **6096** | **5405** | | **I² = 0.69, Tau² = 0.01** | | |
| **Fixed** | | | | | **0.14 (0.10, 0.17)** | | | | | | **0.12 (0.08, 0.16)** | | |
| **Random** | | | | | **0.14 (0.11, 0.18)** | | | | | | **0.11 (0.03, 0.18)** | | |

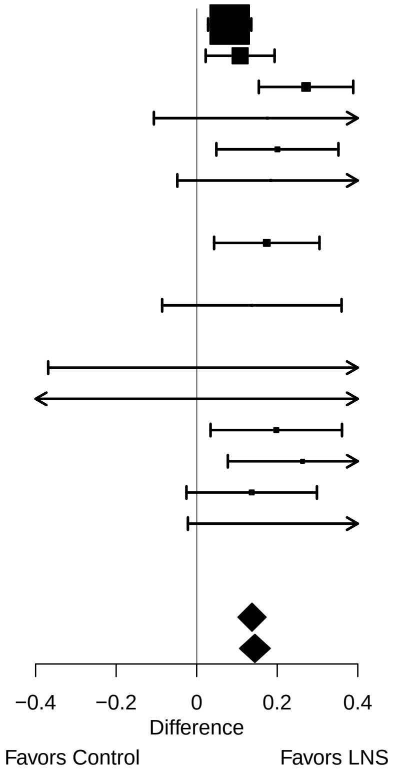

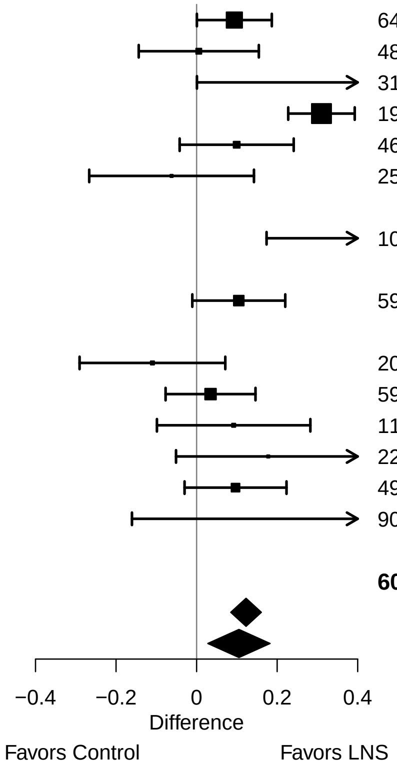

#### Supplemental figure 7A: Mean difference in LAZ

#### 7A5: Stratified by Home environment

**P−for−interaction = 0.394**
**Difference in MDs = 0.03 (−0.04, 0.09)**

| | | At least median | | | | | | Less than median | | | | | |
|---|---|---|---|---|---|---|---|---|---|---|---|---|---|
| Country | Trial | LNS N | Control N | Control Mean | MD (95% CI) | Fixed W | Random W | LNS N | Control N | Control Mean | MD (95% CI) | Fixed W | Random W |
| Bangladesh | JiVitA−4 (34) | | | | | | | | | | | | |
| Bangladesh | RDNS (35) | 1192 | 582 | −1.75 | 0.07 (−0.03, 0.17) | 0.18 | 0.16 | 468 | 230 | −1.93 | 0.09 (−0.03, 0.21) | 0.17 | 0.15 |
| Bangladesh | WASH−B (36) | 637 | 1653 | −1.67 | 0.19 (0.09, 0.29) | 0.18 | 0.16 | 493 | 1690 | −1.96 | 0.18 (0.07, 0.29) | 0.21 | 0.16 |
| Burkina Faso | iLiNS−Zinc (37) | 473 | 199 | −1.62 | 0.24 (0.12, 0.35) | 0.13 | 0.14 | 271 | 174 | −1.87 | 0.25 (0.10, 0.41) | 0.10 | 0.13 |
| Burkina Faso | PROMIS (38) | | | | | | | | | | | | |
| Burkina Faso | PROMIS CS (38) | | | | | | | | | | | | |
| Ghana | GHANA (39) | | | | | | | | | | | | |
| Ghana | iLiNS−DYADG (40) | 213 | 409 | −0.88 | 0.16 (−0.01, 0.32) | 0.06 | 0.10 | 119 | 248 | −0.93 | 0.27 (0.05, 0.49) | 0.05 | 0.09 |
| Haiti | HAITI (41) | | | | | | | | | | | | |
| Kenya | WASH−B (42) | 835 | 2764 | −1.47 | 0.10 (0.02, 0.18) | 0.27 | 0.18 | 574 | 2160 | −1.65 | 0.18 (0.09, 0.28) | 0.26 | 0.17 |
| Madagascar | MAHAY (43) | 915 | 907 | −2.24 | −0.05 (−0.22, 0.13) | 0.06 | 0.09 | 787 | 775 | −2.24 | 0.11 (−0.05, 0.28) | 0.09 | 0.12 |
| Malawi | iLiNS−DYADM (44) | 137 | 289 | −1.56 | −0.06 (−0.28, 0.16) | 0.04 | 0.07 | 78 | 149 | −1.76 | −0.11 (−0.40, 0.18) | 0.03 | 0.06 |
| Malawi | iLiNS−DOSE (45) | 375 | 136 | −2.05 | 0.16 (0.02, 0.30) | 0.09 | 0.12 | 275 | 86 | −1.75 | −0.08 (−0.24, 0.09) | 0.09 | 0.12 |
| Mali | PROMIS (46) | | | | | | | | | | | | |
| Mali | PROMIS CS (46) | | | | | | | | | | | | |
| Zimbabwe | SHINE (HIV−) (47) | | | | | | | | | | | | |
| Zimbabwe | SHINE (HIV+) (48) | | | | | | | | | | | | |
| | | **4777** | **6939** | | **I² = 0.49, Tau² = 0.00** | | | **3065** | **5512** | | **I² = 0.53, Tau² = 0.01** | | |
| **Fixed** | | | | | **0.12 (0.08, 0.16)** | | | | | | **0.14 (0.09, 0.19)** | | |
| **Random** | | | | | **0.12 (0.05, 0.18)** | | | | | | **0.13 (0.04, 0.21)** | | |

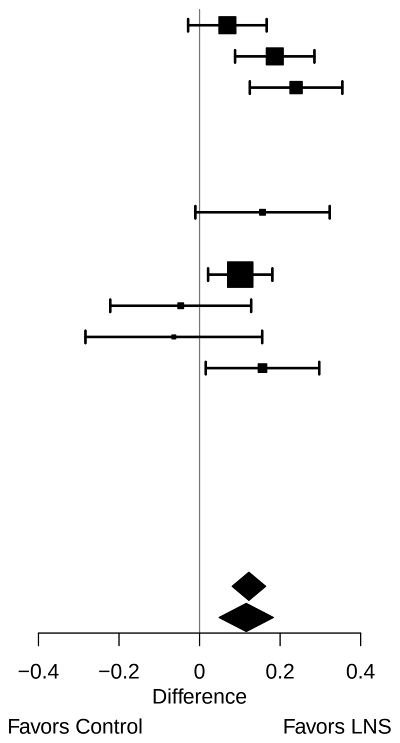

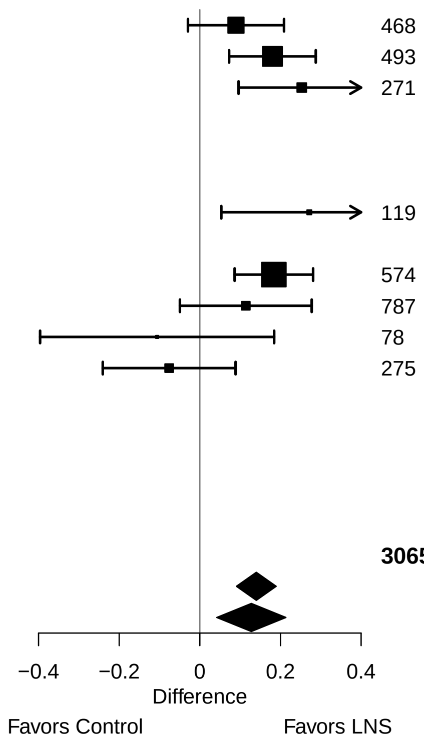

#### Supplemental figure 7A: Mean difference in LAZ

#### 7A6: Stratified by Season at the time of assessment

**P−for−interaction = 0.834**
**Difference in MDs = 0.00 (−0.04, 0.05)**

| Country | Trial | Dry LNS N | Dry Control N | Dry Control Mean | Dry MD (95% CI) | Dry Fixed W | Dry Random W | Rainy LNS N | Rainy Control N | Rainy Control Mean | Rainy MD (95% CI) | Rainy Fixed W | Rainy Random W |
|---|---|---|---|---|---|---|---|---|---|---|---|---|---|
| Bangladesh | JiVitA−4 (34) | 1550 | 663 | −1.90 | 0.08 (0.02, 0.14) | 0.26 | 0.11 | 1288 | 581 | −1.91 | 0.10 (0.02, 0.17) | 0.22 | 0.15 |
| Bangladesh | RDNS (35) | 739 | 359 | −1.82 | 0.05 (−0.07, 0.18) | 0.06 | 0.08 | 924 | 456 | −1.79 | 0.09 (−0.01, 0.20) | 0.11 | 0.12 |
| Bangladesh | WASH−B (36) | 640 | 1913 | −1.74 | 0.19 (0.09, 0.29) | 0.11 | 0.10 | 518 | 1518 | −1.91 | 0.23 (0.15, 0.32) | 0.15 | 0.13 |
| Burkina Faso | iLiNS−Zinc (37) | 1394 | 477 | −1.80 | 0.32 (0.22, 0.41) | 0.11 | 0.10 | 558 | 187 | −1.64 | 0.29 (0.18, 0.39) | 0.10 | 0.12 |
| Burkina Faso | PROMIS (38) | 434 | 446 | −1.41 | 0.09 (−0.06, 0.23) | 0.05 | 0.08 | 429 | 468 | −1.55 | 0.21 (0.04, 0.37) | 0.04 | 0.08 |
| Burkina Faso | PROMIS CS (38) | 224 | 190 | −1.30 | 0.16 (−0.10, 0.41) | 0.02 | 0.04 | 206 | 249 | −1.17 | −0.06 (−0.33, 0.21) | 0.02 | 0.04 |
| Ghana | GHANA (39) | 57 | 55 | −0.43 | 0.48 (0.08, 0.88) | 0.01 | 0.02 | 41 | 41 | −0.38 | 0.00 (−0.46, 0.47) | 0.01 | 0.01 |
| Ghana | iLiNS−DYADG (40) | 204 | 407 | −0.88 | 0.14 (−0.03, 0.30) | 0.04 | 0.07 | 143 | 285 | −0.90 | 0.29 (0.09, 0.50) | 0.03 | 0.06 |
| Haiti | HAITI (41) | | | | | | | | | | | | |
| Kenya | WASH−B (42) | 347 | 1151 | −1.56 | 0.24 (0.13, 0.35) | 0.08 | 0.09 | 1110 | 3986 | −1.57 | 0.13 (0.06, 0.20) | 0.24 | 0.15 |
| Madagascar | MAHAY (43) | | | | | | | | | | | | |
| Malawi | iLiNS−DYADM (44) | 156 | 316 | −1.60 | −0.06 (−0.27, 0.14) | 0.02 | 0.06 | 64 | 128 | −1.68 | −0.11 (−0.44, 0.22) | 0.01 | 0.03 |
| Malawi | iLiNS−DOSE (45) | 415 | 134 | −1.90 | −0.03 (−0.17, 0.10) | 0.05 | 0.08 | 281 | 107 | −1.99 | 0.20 (0.04, 0.36) | 0.05 | 0.08 |
| Mali | PROMIS (46) | | | | | | | | | | | | |
| Mali | PROMIS CS (46) | | | | | | | | | | | | |
| Zimbabwe | SHINE (HIV−) (47) | 1747 | 1662 | −1.62 | 0.17 (0.09, 0.25) | 0.16 | 0.10 | 133 | 132 | −1.40 | −0.03 (−0.32, 0.26) | 0.01 | 0.03 |
| Zimbabwe | SHINE (HIV+) (48) | 319 | 306 | −2.02 | 0.26 (0.08, 0.44) | 0.03 | 0.07 | 18 | 24 | −1.78 | 0.23 (−0.35, 0.81) | 0.00 | 0.01 |
| | | **8226** | **8079** | | **I² = 0.69, Tau² = 0.01** | | | **5713** | **8162** | | **I² = 0.48, Tau² = 0.00** | | |
| **Fixed** | | | | | **0.15 (0.11, 0.18)** | | | | | | **0.15 (0.12, 0.19)** | | |
| **Random** | | | | | **0.15 (0.08, 0.21)** | | | | | | **0.15 (0.09, 0.21)** | | |

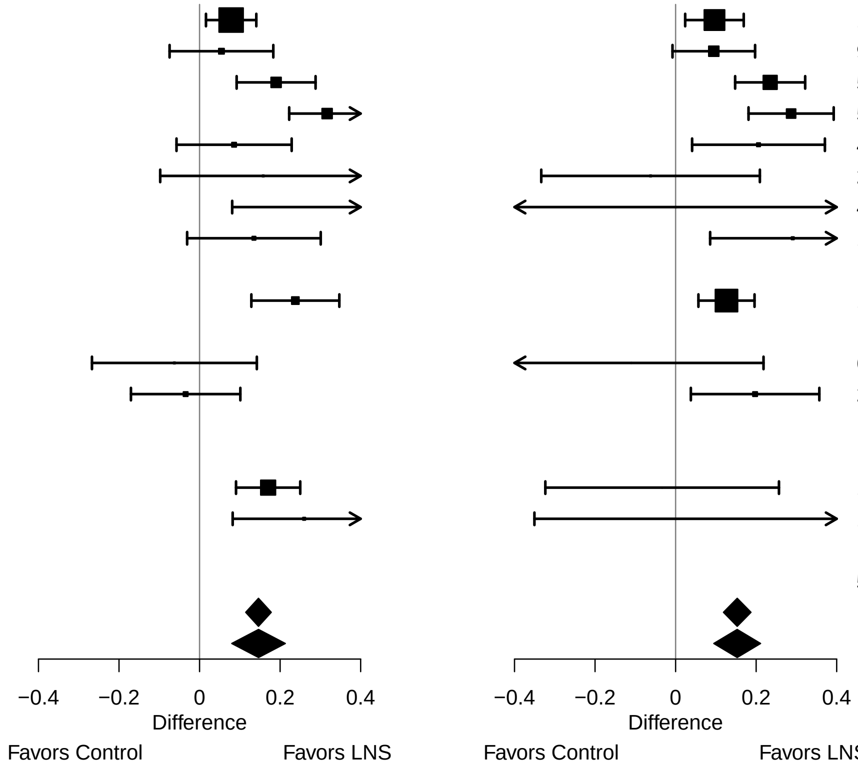

### Supplemental figure 7B: Stunting prevalence ratio

#### 7B1: Stratified by Household socio-economic status

**P−for−interaction = 0.476**
**Ratio of PRs = 1.02 (0.96, 1.08)**

| | | At least median | | | | | | | Less than median | | | | |
|---|---|---|---|---|---|---|---|---|---|---|---|---|---|
| **Country** | **Trial** | **LNS N** | **Control N** | **Control Prevalence** | **PR (95% CI)** | **Fixed W** | **Random W** | **LNS N** | **Control N** | **Control Prevalence** | **PR (95% CI)** | **Fixed W** | **Random W** |
| Bangladesh | JiVitA−4 (34) | 1429 | 621 | 39.0 | 0.87 (0.78, 0.98) | 0.16 | 0.12 | 1408 | 623 | 49.4 | 0.94 (0.85, 1.05) | 0.13 | 0.11 |
| Bangladesh | RDNS (35) | 834 | 407 | 35.9 | 0.86 (0.74, 1.01) | 0.08 | 0.09 | 829 | 408 | 48.0 | 0.96 (0.89, 1.05) | 0.21 | 0.13 |
| Bangladesh | WASH−B (36) | 579 | 1716 | 33.1 | 0.85 (0.74, 0.98) | 0.10 | 0.09 | 579 | 1715 | 50.7 | 0.83 (0.75, 0.92) | 0.15 | 0.12 |
| Burkina Faso | iLiNS−Zinc (37) | 1225 | 299 | 38.1 | 0.82 (0.71, 0.95) | 0.09 | 0.09 | 720 | 364 | 40.3 | 0.70 (0.56, 0.87) | 0.03 | 0.05 |
| Burkina Faso | PROMIS (38) | 407 | 482 | 32.0 | 0.75 (0.58, 0.98) | 0.03 | 0.04 | 456 | 432 | 28.9 | 0.89 (0.68, 1.17) | 0.02 | 0.04 |
| Burkina Faso | PROMIS CS (38) | 220 | 210 | 21.0 | 1.11 (0.76, 1.60) | 0.01 | 0.02 | 210 | 229 | 27.5 | 0.69 (0.41, 1.16) | 0.01 | 0.01 |
| Ghana | GHANA (39) | | | | | | | | | | | | |
| Ghana | iLiNS−DYADG (40) | 157 | 356 | 10.4 | 0.80 (0.44, 1.46) | 0.01 | 0.01 | 190 | 334 | 15.3 | 0.59 (0.35, 0.98) | 0.01 | 0.01 |
| Haiti | HAITI (41) | 75 | 60 | 18.3 | 0.73 (0.33, 1.62) | 0.00 | 0.01 | 61 | 72 | 9.7 | 0.99 (0.47, 2.08) | 0.00 | 0.01 |
| Kenya | WASH−B (42) | 825 | 2921 | 28.8 | 0.80 (0.69, 0.93) | 0.09 | 0.09 | 630 | 2212 | 36.8 | 0.91 (0.80, 1.03) | 0.09 | 0.10 |
| Madagascar | MAHAY (43) | 771 | 884 | 58.5 | 1.02 (0.91, 1.15) | 0.16 | 0.12 | 878 | 755 | 58.5 | 0.94 (0.86, 1.04) | 0.15 | 0.12 |
| Malawi | iLiNS−DYADM (44) | 121 | 234 | 31.2 | 0.98 (0.71, 1.36) | 0.02 | 0.03 | 99 | 209 | 38.3 | 1.24 (0.95, 1.62) | 0.02 | 0.04 |
| Malawi | iLiNS−DOSE (45) | 301 | 104 | 37.5 | 1.11 (0.88, 1.40) | 0.04 | 0.05 | 294 | 103 | 55.3 | 0.96 (0.80, 1.17) | 0.04 | 0.06 |
| Mali | PROMIS (46) | 247 | 251 | 37.5 | 0.88 (0.71, 1.08) | 0.05 | 0.06 | 246 | 246 | 36.2 | 0.93 (0.66, 1.29) | 0.01 | 0.03 |
| Mali | PROMIS CS (46) | 505 | 463 | 33.3 | 0.71 (0.57, 0.90) | 0.04 | 0.05 | 447 | 506 | 34.2 | 0.86 (0.68, 1.10) | 0.02 | 0.04 |
| Zimbabwe | SHINE (HIV−) (47) | 908 | 802 | 33.4 | 0.73 (0.63, 0.85) | 0.09 | 0.09 | 832 | 860 | 37.3 | 0.82 (0.72, 0.94) | 0.08 | 0.09 |
| Zimbabwe | SHINE (HIV+) (48) | 171 | 156 | 45.5 | 0.84 (0.64, 1.10) | 0.03 | 0.04 | 158 | 167 | 53.9 | 0.81 (0.64, 1.03) | 0.03 | 0.05 |
| | | **8775** | **9966** | | **I² = 0.41, Tau² = 0.00** | | | **8037** | **9235** | | **I² = 0.38, Tau² = 0.01** | | |
| **Fixed** | | | | | **0.87 (0.83, 0.91)** | | | | | | **0.90 (0.87, 0.94)** | | |
| **Random** | | | | | **0.86 (0.81, 0.92)** | | | | | | **0.89 (0.84, 0.95)** | | |

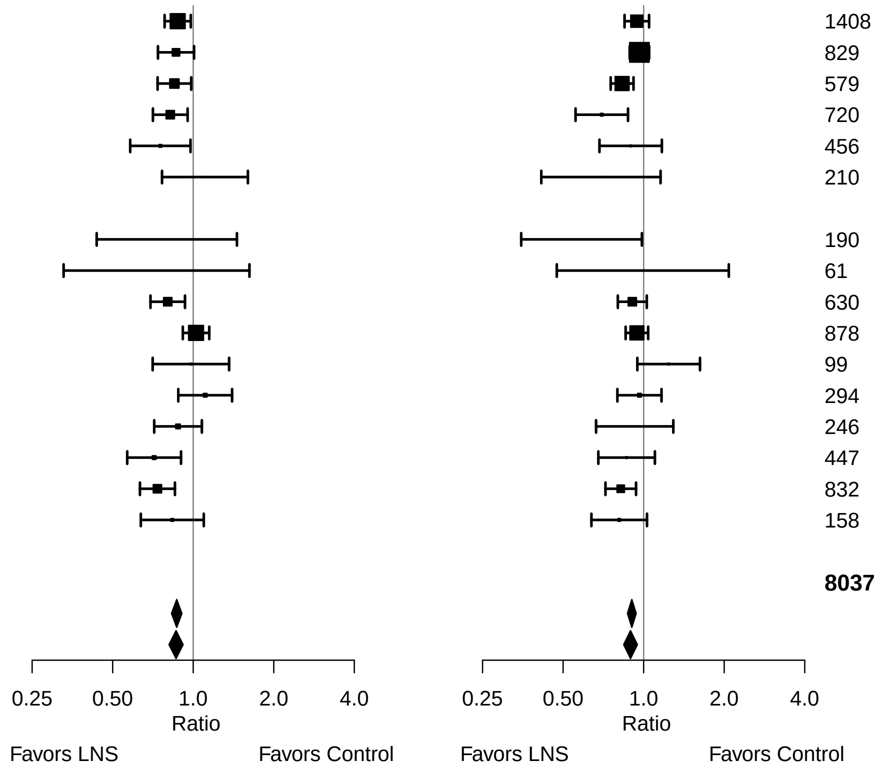

#### Supplemental figure 7B: Stunting prevalence ratio

#### 7B2: Stratified by Household food insecurity

**P−for−interaction = 0.827**
**Ratio of PRs = 1.01 (0.94, 1.08)**

| | | Mild to secure | | | | | | Moderate to severe | | | | | |
|---|---|---|---|---|---|---|---|---|---|---|---|---|---|
| Country | Trial | LNS N | Control N | Control Prevalence | PR (95% CI) | Fixed W | Random W | LNS N | Control N | Control Prevalence | PR (95% CI) | Fixed W | Random W |
| Bangladesh | JiVitA−4 (34) | 2012 | 863 | 41.2 | 0.86 (0.78, 0.95) | 0.16 | 0.14 | 825 | 381 | 50.9 | 1.00 (0.87, 1.14) | 0.15 | 0.11 |
| Bangladesh | RDNS (35) | 1058 | 493 | 36.3 | 0.99 (0.88, 1.12) | 0.11 | 0.11 | 605 | 322 | 50.6 | 0.85 (0.75, 0.96) | 0.18 | 0.12 |
| Bangladesh | WASH−B (36) | 918 | 2652 | 39.4 | 0.81 (0.73, 0.90) | 0.15 | 0.13 | 240 | 779 | 50.3 | 0.93 (0.80, 1.07) | 0.13 | 0.11 |
| Burkina Faso | iLiNS−Zinc (37) | 1013 | 320 | 40.5 | 0.76 (0.60, 0.96) | 0.03 | 0.04 | 932 | 343 | 38.2 | 0.74 (0.64, 0.86) | 0.12 | 0.10 |
| Burkina Faso | PROMIS (38) | 502 | 550 | 29.6 | 0.88 (0.71, 1.10) | 0.03 | 0.05 | 361 | 364 | 31.9 | 0.74 (0.56, 0.98) | 0.03 | 0.06 |
| Burkina Faso | PROMIS CS (38) | | | | | | | | | | | | |
| Ghana | GHANA (39) | | | | | | | | | | | | |
| Ghana | iLiNS−DYADG (40) | 244 | 470 | 12.6 | 0.62 (0.38, 1.02) | 0.01 | 0.01 | 102 | 219 | 14.2 | 0.76 (0.40, 1.45) | 0.01 | 0.02 |
| Haiti | HAITI (41) | | | | | | | | | | | | |
| Kenya | WASH−B (42) | 1299 | 4595 | 31.3 | 0.86 (0.78, 0.95) | 0.16 | 0.14 | 156 | 525 | 41.0 | 0.80 (0.59, 1.07) | 0.03 | 0.05 |
| Madagascar | MAHAY (43) | 988 | 927 | 59.4 | 0.96 (0.86, 1.06) | 0.15 | 0.13 | 355 | 410 | 58.5 | 1.02 (0.88, 1.18) | 0.12 | 0.11 |
| Malawi | iLiNS−DYADM (44) | 70 | 122 | 27.0 | 1.16 (0.74, 1.83) | 0.01 | 0.01 | 148 | 316 | 36.7 | 1.12 (0.88, 1.43) | 0.05 | 0.07 |
| Malawi | iLiNS−DOSE (45) | 151 | 55 | 38.2 | 0.99 (0.73, 1.35) | 0.02 | 0.02 | 431 | 146 | 48.6 | 1.06 (0.89, 1.27) | 0.09 | 0.09 |
| Mali | PROMIS (46) | 329 | 341 | 35.8 | 0.92 (0.73, 1.14) | 0.03 | 0.04 | 174 | 163 | 38.0 | 0.90 (0.66, 1.23) | 0.03 | 0.05 |
| Mali | PROMIS CS (46) | | | | | | | | | | | | |
| Zimbabwe | SHINE (HIV−) (47) | 1393 | 1305 | 34.6 | 0.80 (0.72, 0.90) | 0.13 | 0.12 | 296 | 325 | 38.8 | 0.69 (0.54, 0.88) | 0.04 | 0.07 |
| Zimbabwe | SHINE (HIV+) (48) | 236 | 239 | 46.4 | 0.85 (0.68, 1.05) | 0.03 | 0.05 | 86 | 76 | 59.2 | 0.77 (0.57, 1.03) | 0.03 | 0.05 |
| | | **10213** | **12932** | | **I² = 0.30, Tau² = 0.00** | | | **4711** | **4369** | | **I² = 0.56, Tau² = 0.01** | | |
| **Fixed** | | | | | **0.87 (0.84, 0.91)** | | | | | | **0.90 (0.85, 0.95)** | | |
| **Random** | | | | | **0.88 (0.83, 0.92)** | | | | | | **0.89 (0.82, 0.97)** | | |

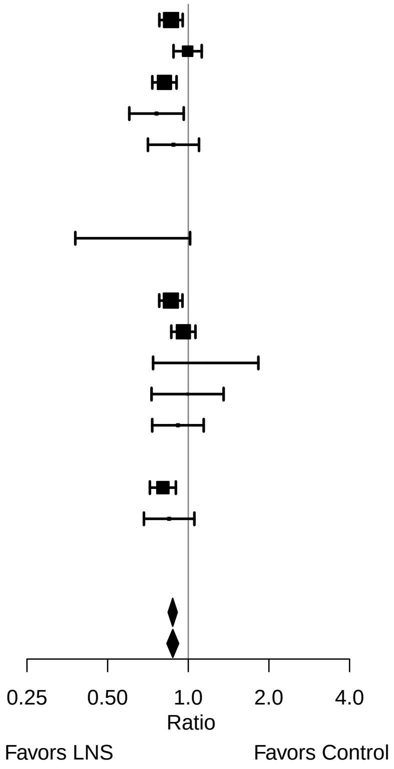

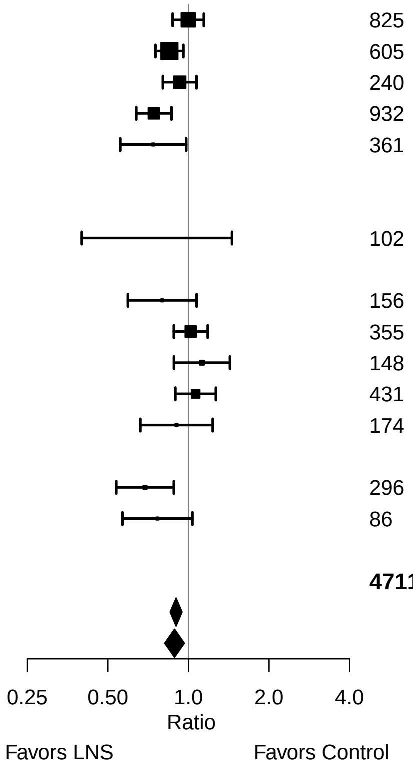

#### Supplemental figure 7B: Stunting prevalence ratio

#### 7B3: Stratified by Household source water quality

**P−for−interaction = 0.782**
**Ratio of PRs = 1.01 (0.93, 1.10)**

| Country | Trial | Improved LNS N | Improved Control N | Improved Control Prevalence | Improved PR (95% CI) | Improved Fixed W | Improved Random W | Unimproved LNS N | Unimproved Control N | Unimproved Control Prevalence | Unimproved PR (95% CI) | Unimproved Fixed W | Unimproved Random W |
|---|---|---|---|---|---|---|---|---|---|---|---|---|---|
| Bangladesh | JiVitA−4 (34) | | | | | | | | | | | | |
| Bangladesh | RDNS (35) | | | | | | | | | | | | |
| Bangladesh | WASH−B (36) | 496 | 989 | 40.7 | 0.77 (0.67, 0.88) | 0.18 | 0.15 | 70 | 114 | 43.0 | 1.00 (0.71, 1.40) | 0.03 | 0.07 |
| Burkina Faso | iLiNS−Zinc (37) | 550 | 141 | 35.5 | 0.82 (0.68, 0.99) | 0.10 | 0.10 | 1394 | 519 | 40.2 | 0.74 (0.63, 0.86) | 0.15 | 0.14 |
| Burkina Faso | PROMIS (38) | 355 | 379 | 31.1 | 0.92 (0.74, 1.15) | 0.07 | 0.08 | 361 | 355 | 31.5 | 0.62 (0.42, 0.92) | 0.03 | 0.06 |
| Burkina Faso | PROMIS CS (38) | 280 | 268 | 25.4 | 0.77 (0.54, 1.11) | 0.03 | 0.03 | 149 | 171 | 22.8 | 1.06 (0.71, 1.58) | 0.02 | 0.05 |
| Ghana | GHANA (39) | | | | | | | | | | | | |
| Ghana | iLiNS−DYADG (40) | | | | | | | | | | | | |
| Haiti | HAITI (41) | | | | | | | | | | | | |
| Kenya | WASH−B (42) | 447 | 1549 | 30.6 | 0.96 (0.80, 1.14) | 0.11 | 0.11 | 248 | 697 | 33.1 | 0.85 (0.66, 1.09) | 0.06 | 0.10 |
| Madagascar | MAHAY (43) | 419 | 453 | 51.4 | 1.02 (0.86, 1.20) | 0.13 | 0.12 | 1203 | 1180 | 61.2 | 0.96 (0.88, 1.05) | 0.47 | 0.18 |
| Malawi | iLiNS−DYADM (44) | | | | | | | | | | | | |
| Malawi | iLiNS−DOSE (45) | 555 | 193 | 46.1 | 1.01 (0.86, 1.19) | 0.13 | 0.12 | 52 | 17 | 47.1 | 1.33 (0.85, 2.07) | 0.02 | 0.05 |
| Mali | PROMIS (46) | 249 | 258 | 36.4 | 0.80 (0.63, 1.01) | 0.06 | 0.07 | 247 | 243 | 36.2 | 1.04 (0.82, 1.32) | 0.07 | 0.10 |
| Mali | PROMIS CS (46) | 549 | 566 | 31.4 | 0.81 (0.64, 1.02) | 0.06 | 0.07 | 362 | 388 | 37.6 | 0.78 (0.62, 0.98) | 0.07 | 0.11 |
| Zimbabwe | SHINE (HIV−) (47) | 521 | 489 | 35.2 | 0.77 (0.65, 0.93) | 0.11 | 0.11 | 274 | 302 | 32.5 | 0.97 (0.74, 1.26) | 0.05 | 0.09 |
| Zimbabwe | SHINE (HIV+) (48) | 85 | 87 | 49.4 | 0.76 (0.55, 1.05) | 0.03 | 0.04 | 55 | 56 | 57.1 | 0.76 (0.50, 1.16) | 0.02 | 0.05 |
| | | **4506** | **5372** | | **I² = 0.36, Tau² = 0.00** | | | **4415** | **4042** | | **I² = 0.50, Tau² = 0.01** | | |
| **Fixed** | | | | | **0.87 (0.82, 0.92)** | | | | | | **0.90 (0.85, 0.96)** | | |
| **Random** | | | | | **0.87 (0.81, 0.93)** | | | | | | **0.89 (0.80, 0.99)** | | |

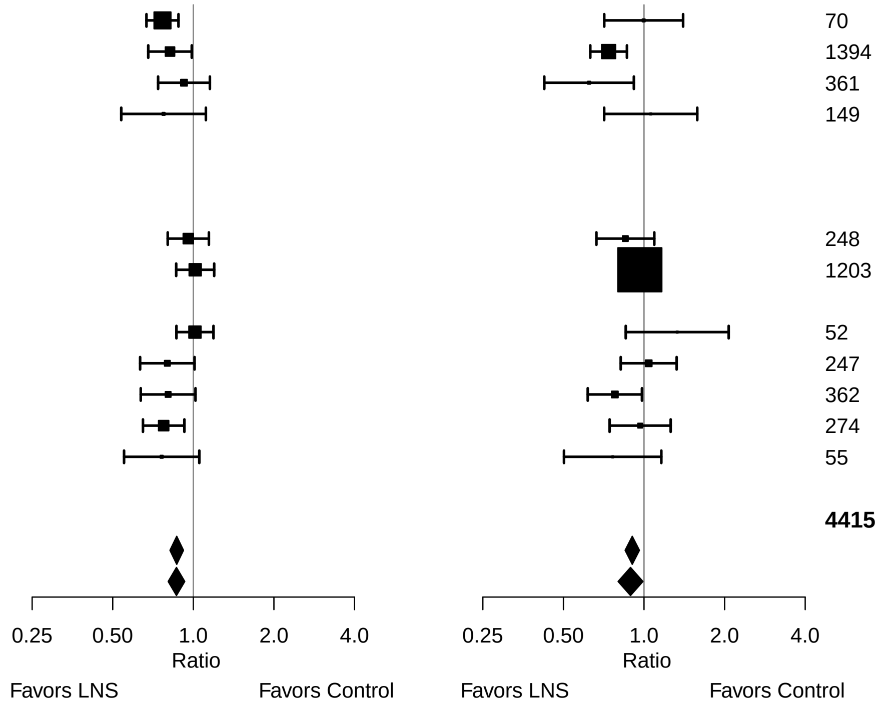

#### Supplemental figure 7B: Stunting prevalence ratio

#### 7B4: Stratified by Household sanitation

**P−for−interaction = 0.200**
**Ratio of PRs = 1.06 (0.97, 1.17)**

| Country | Trial | Improved LNS N | Improved Control N | Improved Control Prevalence | Improved PR (95% CI) | Improved Fixed W | Improved Random W | Unimproved LNS N | Unimproved Control N | Unimproved Control Prevalence | Unimproved PR (95% CI) | Unimproved Fixed W | Unimproved Random W |
|---|---|---|---|---|---|---|---|---|---|---|---|---|---|
| Bangladesh | JiVitA−4 (34) | 2189 | 950 | 42.5 | 0.92 (0.82, 1.02) | 0.27 | 0.27 | 648 | 294 | 49.7 | 0.94 (0.82, 1.07) | 0.19 | 0.18 |
| Bangladesh | RDNS (35) | 1179 | 573 | 39.6 | 0.88 (0.79, 0.97) | 0.30 | 0.30 | 482 | 241 | 47.7 | 1.01 (0.87, 1.17) | 0.16 | 0.16 |
| Bangladesh | WASH−B (36) | 506 | 1001 | 39.7 | 0.78 (0.66, 0.91) | 0.13 | 0.13 | 31 | 54 | 51.9 | 0.75 (0.45, 1.23) | 0.01 | 0.02 |
| Burkina Faso | iLiNS−Zinc (37) | 43 | 16 | 37.5 | 1.37 (0.61, 3.07) | 0.00 | 0.00 | 1901 | 644 | 39.2 | 0.75 (0.65, 0.87) | 0.16 | 0.16 |
| Burkina Faso | PROMIS (38) | 398 | 550 | 31.3 | 0.79 (0.63, 0.99) | 0.06 | 0.06 | 465 | 359 | 29.2 | 0.87 (0.69, 1.10) | 0.06 | 0.07 |
| Burkina Faso | PROMIS CS (38) | 176 | 165 | 24.2 | 0.87 (0.56, 1.35) | 0.02 | 0.02 | 252 | 274 | 24.5 | 0.88 (0.62, 1.24) | 0.03 | 0.03 |
| Ghana | GHANA (39) | | | | | | | | | | | | |
| Ghana | iLiNS−DYADG (40) | | | | | | | | | | | | |
| Haiti | HAITI (41) | | | | | | | | | | | | |
| Kenya | WASH−B (42) | 100 | 364 | 24.7 | 0.89 (0.61, 1.30) | 0.02 | 0.02 | 595 | 1883 | 32.7 | 0.92 (0.78, 1.09) | 0.13 | 0.13 |
| Madagascar | MAHAY (43) | | | | | | | | | | | | |
| Malawi | iLiNS−DYADM (44) | | | | | | | | | | | | |
| Malawi | iLiNS−DOSE (45) | | | | | | | | | | | | |
| Mali | PROMIS (46) | 387 | 366 | 36.1 | 0.87 (0.70, 1.08) | 0.07 | 0.07 | 112 | 135 | 37.8 | 1.03 (0.76, 1.39) | 0.04 | 0.04 |
| Mali | PROMIS CS (46) | 701 | 696 | 32.6 | 0.76 (0.60, 0.96) | 0.06 | 0.06 | 220 | 248 | 35.9 | 0.80 (0.60, 1.07) | 0.04 | 0.04 |
| Zimbabwe | SHINE (HIV−) (47) | 295 | 244 | 32.0 | 0.82 (0.63, 1.05) | 0.05 | 0.05 | 497 | 546 | 35.2 | 0.85 (0.73, 1.00) | 0.14 | 0.14 |
| Zimbabwe | SHINE (HIV+) (48) | 50 | 35 | 54.3 | 0.70 (0.46, 1.08) | 0.02 | 0.02 | 90 | 107 | 51.4 | 0.80 (0.57, 1.11) | 0.03 | 0.03 |
| | | **6024** | **4960** | | **$I^2$ = 0.00, $Tau^2$ = 0.00** | | | **5293** | **4785** | | **$I^2$ = 0.09, $Tau^2$ = 0.00** | | |
| **Fixed** | | | | | **0.86 (0.81, 0.91)** | | | | | | **0.88 (0.83, 0.94)** | | |
| **Random** | | | | | **0.86 (0.81, 0.91)** | | | | | | **0.88 (0.83, 0.94)** | | |

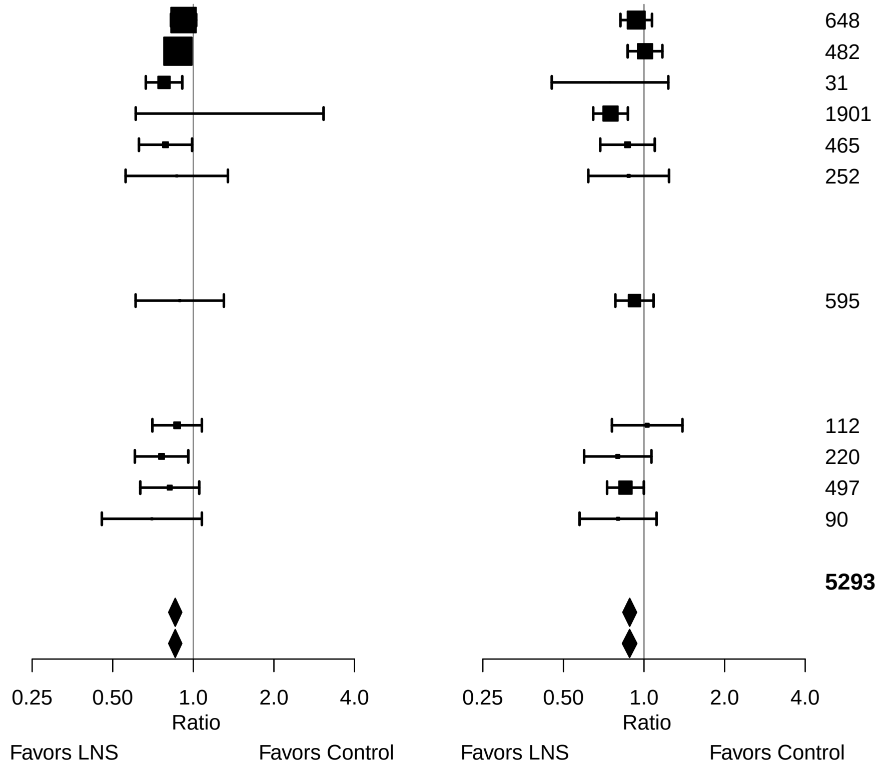

#### Supplemental figure 7B: Stunting prevalence ratio

#### 7B5: Stratified by Home environment

**P−for−interaction = 0.280**
**Ratio of PRs = 0.96 (0.89, 1.03)**

| | | At least median | | | | | | Less than median | | | | | |
|---|---|---|---|---|---|---|---|---|---|---|---|---|---|
| **Country** | **Trial** | **LNS N** | **Control N** | **Control Prevalence** | **PR (95% CI)** | **Fixed W** | **Random W** | **LNS N** | **Control N** | **Control Prevalence** | **PR (95% CI)** | **Fixed W** | **Random W** |
| Bangladesh | JiVitA−4 (34) | | | | | | | | | | | | |
| Bangladesh | RDNS (35) | 1192 | 582 | 39.2 | 0.94 (0.84, 1.06) | 0.21 | 0.16 | 468 | 230 | 48.7 | 0.88 (0.78, 0.99) | 0.21 | 0.19 |
| Bangladesh | WASH−B (36) | 637 | 1653 | 36.9 | 0.84 (0.74, 0.96) | 0.17 | 0.15 | 493 | 1690 | 46.9 | 0.87 (0.77, 0.98) | 0.23 | 0.20 |
| Burkina Faso | iLiNS−Zinc (37) | 473 | 199 | 37.7 | 0.66 (0.54, 0.81) | 0.07 | 0.11 | 271 | 174 | 41.7 | 0.80 (0.60, 1.05) | 0.04 | 0.07 |
| Burkina Faso | PROMIS (38) | | | | | | | | | | | | |
| Burkina Faso | PROMIS CS (38) | | | | | | | | | | | | |
| Ghana | GHANA (39) | | | | | | | | | | | | |
| Ghana | iLiNS−DYADG (40) | 213 | 409 | 12.0 | 0.78 (0.48, 1.28) | 0.01 | 0.04 | 119 | 248 | 15.3 | 0.55 (0.28, 1.06) | 0.01 | 0.02 |
| Haiti | HAITI (41) | | | | | | | | | | | | |
| Kenya | WASH−B (42) | 835 | 2764 | 29.6 | 0.89 (0.78, 1.01) | 0.17 | 0.16 | 574 | 2160 | 34.3 | 0.86 (0.75, 1.00) | 0.15 | 0.17 |
| Madagascar | MAHAY (43) | 915 | 907 | 57.9 | 1.04 (0.94, 1.16) | 0.25 | 0.17 | 787 | 775 | 59.2 | 0.91 (0.82, 1.02) | 0.27 | 0.21 |
| Malawi | iLiNS−DYADM (44) | 137 | 289 | 33.6 | 1.07 (0.81, 1.41) | 0.04 | 0.08 | 78 | 149 | 36.9 | 1.18 (0.85, 1.64) | 0.03 | 0.06 |
| Malawi | iLiNS−DOSE (45) | 375 | 136 | 49.3 | 0.95 (0.80, 1.13) | 0.09 | 0.13 | 275 | 86 | 40.7 | 1.10 (0.86, 1.41) | 0.05 | 0.09 |
| Mali | PROMIS (46) | | | | | | | | | | | | |
| Mali | PROMIS CS (46) | | | | | | | | | | | | |
| Zimbabwe | SHINE (HIV−) (47) | | | | | | | | | | | | |
| Zimbabwe | SHINE (HIV+) (48) | | | | | | | | | | | | |
| | | **4777** | **6939** | | **I² = 0.64, Tau² = 0.01** | | | **3065** | **5512** | | **I² = 0.22, Tau² = 0.01** | | |
| **Fixed** | | | | | **0.92 (0.87, 0.97)** | | | | | | **0.90 (0.85, 0.95)** | | |
| **Random** | | | | | **0.90 (0.81, 1.00)** | | | | | | **0.90 (0.83, 0.98)** | | |

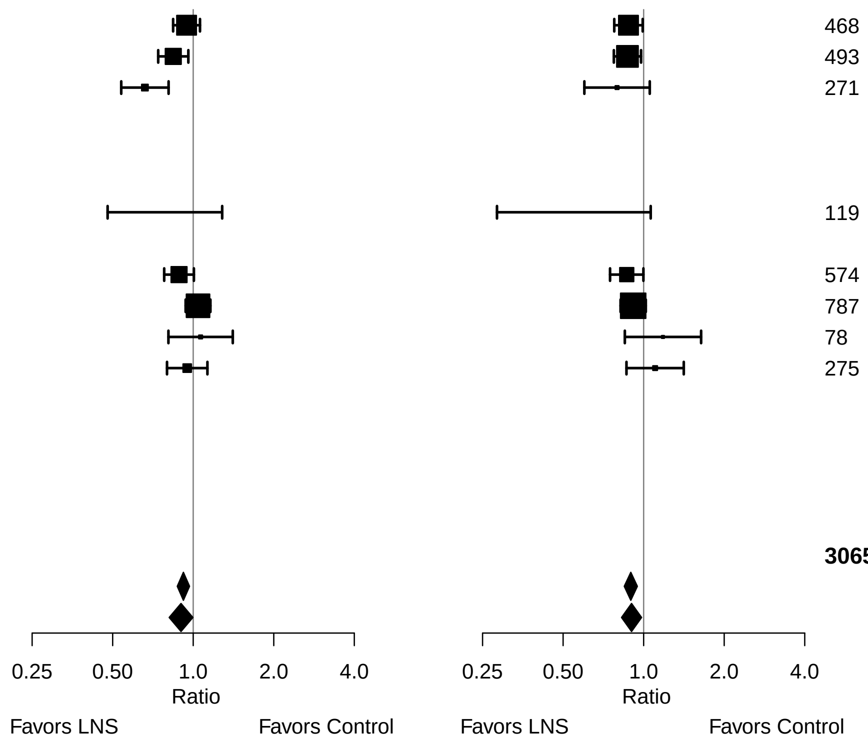

#### Supplemental figure 7B: Stunting prevalence ratio

#### 7B6: Stratified by Season at the time of assessment

**P−for−interaction = 0.389**
**Ratio of PRs = 0.97 (0.90, 1.04)**

| Country | Trial | LNS N | Control N | Control Prevalence | Dry PR (95% CI) | Fixed W | Random W | LNS N | Control N | Control Prevalence | Rainy PR (95% CI) | Fixed W | Random W |
|---|---|---|---|---|---|---|---|---|---|---|---|---|---|
| Bangladesh | JiVitA−4 (34) | 1550 | 663 | 44.9 | 0.96 (0.87, 1.07) | 0.17 | 0.13 | 1288 | 581 | 43.4 | 0.87 (0.75, 1.00) | 0.13 | 0.13 |
| Bangladesh | RDNS (35) | 739 | 359 | 43.7 | 0.90 (0.78, 1.04) | 0.10 | 0.11 | 924 | 456 | 40.6 | 0.94 (0.85, 1.04) | 0.25 | 0.25 |
| Bangladesh | WASH−B (36) | 640 | 1913 | 39.1 | 0.86 (0.77, 0.96) | 0.15 | 0.12 | 518 | 1518 | 45.5 | 0.82 (0.73, 0.91) | 0.20 | 0.20 |
| Burkina Faso | iLiNS−Zinc (37) | 1394 | 477 | 40.3 | 0.76 (0.67, 0.86) | 0.13 | 0.12 | 558 | 187 | 37.2 | 0.72 (0.54, 0.96) | 0.03 | 0.03 |
| Burkina Faso | PROMIS (38) | 434 | 446 | 27.1 | 0.84 (0.65, 1.09) | 0.03 | 0.05 | 429 | 468 | 33.8 | 0.81 (0.64, 1.01) | 0.05 | 0.05 |
| Burkina Faso | PROMIS CS (38) | 224 | 190 | 25.3 | 0.80 (0.52, 1.21) | 0.01 | 0.02 | 206 | 249 | 23.7 | 0.94 (0.58, 1.52) | 0.01 | 0.01 |
| Ghana | GHANA (39) | | | | | | | | | | | | |
| Ghana | iLiNS−DYADG (40) | 204 | 407 | 12.8 | 0.77 (0.47, 1.25) | 0.01 | 0.02 | 143 | 285 | 13.3 | 0.52 (0.27, 1.02) | 0.01 | 0.01 |
| Haiti | HAITI (41) | | | | | | | | | | | | |
| Kenya | WASH−B (42) | 347 | 1151 | 30.1 | 0.81 (0.66, 1.00) | 0.04 | 0.07 | 1110 | 3986 | 32.8 | 0.87 (0.79, 0.97) | 0.24 | 0.24 |
| Madagascar | MAHAY (43) | | | | | | | | | | | | |
| Malawi | iLiNS−DYADM (44) | 156 | 316 | 32.9 | 1.13 (0.87, 1.46) | 0.03 | 0.05 | 64 | 128 | 39.1 | 1.04 (0.72, 1.50) | 0.02 | 0.02 |
| Malawi | iLiNS−DOSE (45) | 415 | 134 | 44.0 | 1.08 (0.90, 1.28) | 0.06 | 0.09 | 281 | 107 | 49.5 | 0.88 (0.71, 1.10) | 0.05 | 0.05 |
| Mali | PROMIS (46) | | | | | | | | | | | | |
| Mali | PROMIS CS (46) | | | | | | | | | | | | |
| Zimbabwe | SHINE (HIV−) (47) | 1747 | 1662 | 35.4 | 0.77 (0.70, 0.86) | 0.20 | 0.13 | 133 | 132 | 28.0 | 1.05 (0.71, 1.55) | 0.02 | 0.02 |
| Zimbabwe | SHINE (HIV+) (48) | 319 | 306 | 50.7 | 0.81 (0.68, 0.97) | 0.06 | 0.08 | 18 | 24 | 41.7 | 0.67 (0.28, 1.60) | 0.00 | 0.00 |
| | | **8169** | **8024** | | **$I^2$ = 0.56, $Tau^2$ = 0.01** | | | **5672** | **8121** | | **$I^2$ = 0.00, $Tau^2$ = 0.00** | | |
| **Fixed** | | | | | **0.86 (0.82, 0.90)** | | | | | | **0.87 (0.83, 0.92)** | | |
| **Random** | | | | | **0.87 (0.81, 0.93)** | | | | | | **0.87 (0.83, 0.92)** | | |

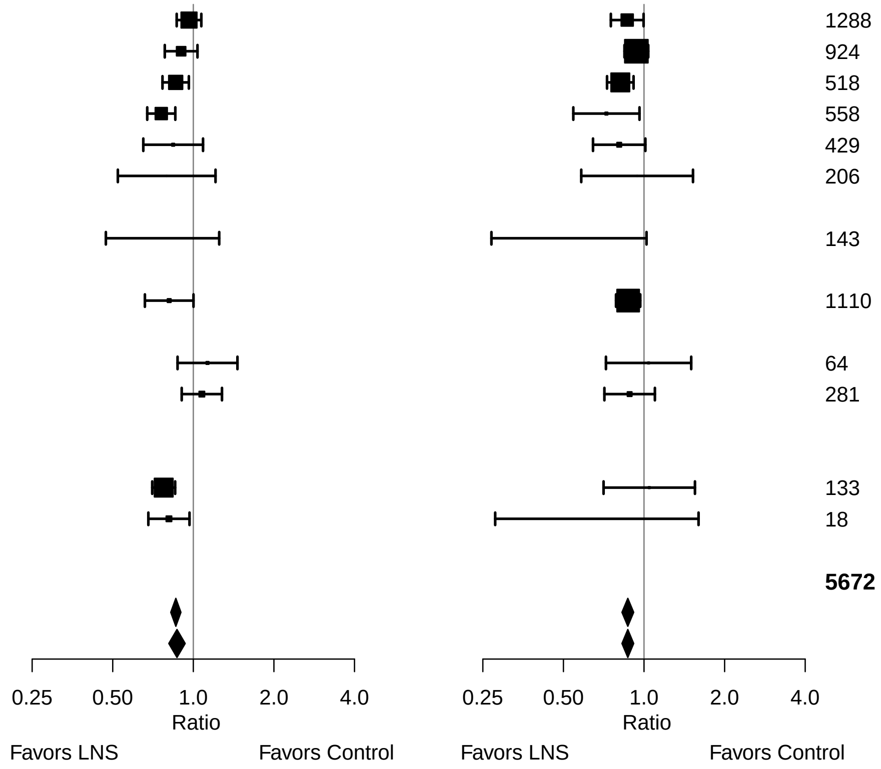

### Supplemental figure 7C: Stunting prevalence difference

#### 7C1: Stratified by Household socio-economic status

**P−for−interaction = 0.724**
**Difference in PDs = 0.00 (−0.02, 0.02)**

| Country | Trial | LNS N | Control N | Control Prevalence | At least median PD (95% CI) | Fixed W | Random W | LNS N | Control N | Control Prevalence | Less than median PD (95% CI) | Fixed W | Random W |
|---|---|---|---|---|---|---|---|---|---|---|---|---|---|
| Bangladesh | JiVitA−4 (34) | 1429 | 621 | 39.0 | −0.04 (−0.08, 0.00) | 0.13 | 0.10 | 1408 | 623 | 49.4 | −0.03 (−0.07, 0.01) | 0.13 | 0.10 |
| Bangladesh | RDNS (35) | 834 | 407 | 35.9 | −0.05 (−0.10, 0.00) | 0.07 | 0.07 | 829 | 408 | 48.0 | −0.02 (−0.06, 0.02) | 0.14 | 0.10 |
| Bangladesh | WASH−B (36) | 579 | 1716 | 33.1 | −0.05 (−0.09, −0.01) | 0.10 | 0.09 | 579 | 1715 | 50.7 | −0.09 (−0.13, −0.04) | 0.11 | 0.09 |
| Burkina Faso | iLiNS−Zinc (37) | 1225 | 299 | 38.1 | −0.08 (−0.12, −0.03) | 0.11 | 0.09 | 720 | 364 | 40.3 | −0.14 (−0.20, −0.08) | 0.06 | 0.07 |
| Burkina Faso | PROMIS (38) | 407 | 482 | 32.0 | −0.08 (−0.15, 0.00) | 0.03 | 0.04 | 456 | 432 | 28.9 | −0.03 (−0.10, 0.04) | 0.04 | 0.05 |
| Burkina Faso | PROMIS CS (38) | 220 | 210 | 21.0 | 0.02 (−0.06, 0.10) | 0.03 | 0.04 | 210 | 229 | 27.5 | −0.08 (−0.20, 0.03) | 0.02 | 0.03 |
| Ghana | GHANA (39) | | | | | | | | | | | | |
| Ghana | iLiNS−DYADG (40) | 157 | 356 | 10.4 | −0.02 (−0.08, 0.03) | 0.06 | 0.07 | 190 | 334 | 15.3 | −0.06 (−0.12, 0.00) | 0.06 | 0.07 |
| Haiti | HAITI (41) | 75 | 60 | 18.3 | −0.08 (−0.17, 0.02) | 0.02 | 0.03 | 61 | 72 | 9.7 | 0.01 (−0.08, 0.09) | 0.03 | 0.04 |
| Kenya | WASH−B (42) | 825 | 2921 | 28.8 | −0.06 (−0.09, −0.02) | 0.15 | 0.11 | 630 | 2212 | 36.8 | −0.03 (−0.08, 0.01) | 0.12 | 0.10 |
| Madagascar | MAHAY (43) | 771 | 884 | 58.5 | 0.01 (−0.05, 0.08) | 0.04 | 0.05 | 878 | 755 | 58.5 | −0.03 (−0.09, 0.02) | 0.07 | 0.08 |
| Malawi | iLiNS−DYADM (44) | 121 | 234 | 31.2 | −0.01 (−0.11, 0.10) | 0.02 | 0.03 | 99 | 209 | 38.3 | 0.09 (−0.03, 0.21) | 0.02 | 0.03 |
| Malawi | iLiNS−DOSE (45) | 301 | 104 | 37.5 | 0.05 (−0.04, 0.13) | 0.03 | 0.04 | 294 | 103 | 55.3 | −0.05 (−0.14, 0.04) | 0.03 | 0.04 |
| Mali | PROMIS (46) | 247 | 251 | 37.5 | −0.06 (−0.13, 0.01) | 0.04 | 0.05 | 246 | 246 | 36.2 | −0.03 (−0.13, 0.07) | 0.02 | 0.03 |
| Mali | PROMIS CS (46) | 505 | 463 | 33.3 | −0.09 (−0.16, −0.03) | 0.05 | 0.06 | 447 | 506 | 34.2 | −0.05 (−0.12, 0.03) | 0.04 | 0.05 |
| Zimbabwe | SHINE (HIV−) (47) | 908 | 802 | 33.4 | −0.09 (−0.13, −0.05) | 0.11 | 0.09 | 832 | 860 | 37.3 | −0.07 (−0.11, −0.02) | 0.11 | 0.09 |
| Zimbabwe | SHINE (HIV+) (48) | 171 | 156 | 45.5 | −0.08 (−0.19, 0.04) | 0.01 | 0.02 | 158 | 167 | 53.9 | −0.10 (−0.22, 0.02) | 0.02 | 0.03 |
| | | **8775** | **9966** | | **I² = 0.29, Tau² = 0.00** | | | **8037** | **9235** | | **I² = 0.39, Tau² = 0.00** | | |
| **Fixed** | | | | | **−0.05 (−0.06, −0.04)** | | | | | | **−0.05 (−0.06, −0.03)** | | |
| **Random** | | | | | **−0.05 (−0.07, −0.03)** | | | | | | **−0.05 (−0.07, −0.03)** | | |

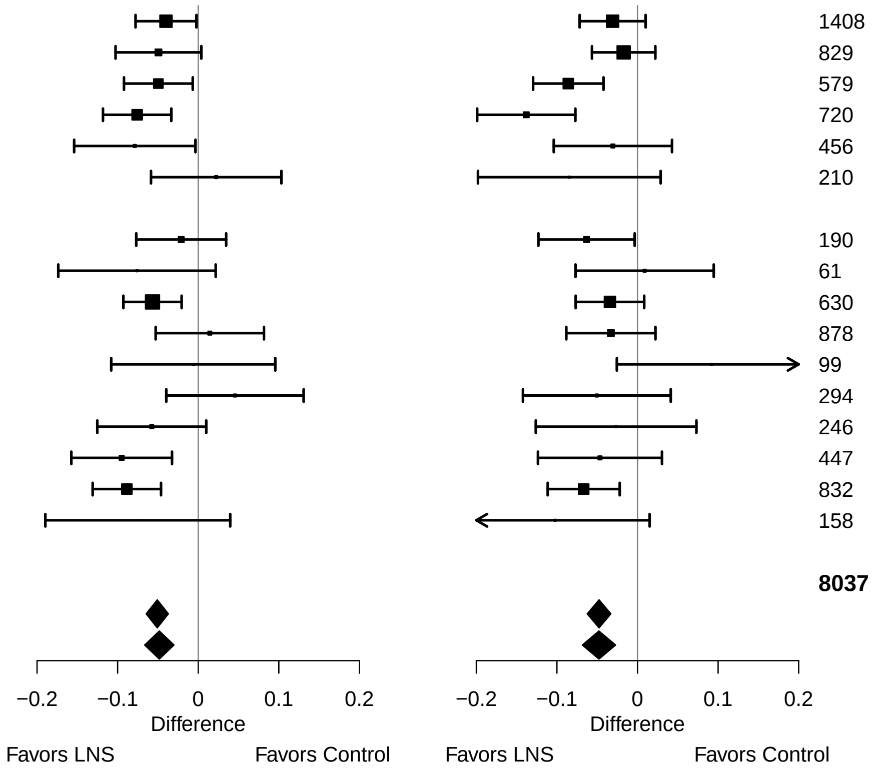

#### Supplemental figure 7C: Stunting prevalence difference

#### 7C2: Stratified by Household food insecurity

**P−for−interaction = 0.663**
**Difference in PDs = −0.01 (−0.03, 0.02)**

**Mild to secure**

| Country | Trial | LNS N | Control N | Control Prevalence | PD (95% CI) | Fixed W | Random W |
|---|---|---|---|---|---|---|---|
| Bangladesh | JiVitA−4 (34) | 2012 | 863 | 41.2 | −0.04 (−0.08, −0.01) | 0.14 | 0.13 |
| Bangladesh | RDNS (35) | 1058 | 493 | 36.3 | 0.00 (−0.05, 0.04) | 0.09 | 0.10 |
| Bangladesh | WASH−B (36) | 918 | 2652 | 39.4 | −0.07 (−0.11, −0.04) | 0.13 | 0.12 |
| Burkina Faso | iLiNS−Zinc (37) | 1013 | 320 | 40.5 | −0.11 (−0.19, −0.04) | 0.03 | 0.04 |
| Burkina Faso | PROMIS (38) | 502 | 550 | 29.6 | −0.04 (−0.10, 0.03) | 0.05 | 0.06 |
| Burkina Faso | PROMIS CS (38) | | | | | | |
| Ghana | GHANA (39) | | | | | | |
| Ghana | iLiNS−DYADG (40) | 244 | 470 | 12.6 | −0.05 (−0.10, 0.00) | 0.07 | 0.08 |
| Haiti | HAITI (41) | | | | | | |
| Kenya | WASH−B (42) | 1299 | 4595 | 31.3 | −0.04 (−0.07, −0.02) | 0.22 | 0.17 |
| Madagascar | MAHAY (43) | 988 | 927 | 59.4 | −0.02 (−0.08, 0.04) | 0.05 | 0.06 |
| Malawi | iLiNS−DYADM (44) | 70 | 122 | 27.0 | 0.04 (−0.09, 0.18) | 0.01 | 0.01 |
| Malawi | iLiNS−DOSE (45) | 151 | 55 | 38.2 | 0.04 (−0.07, 0.15) | 0.01 | 0.02 |
| Mali | PROMIS (46) | 329 | 341 | 35.8 | −0.04 (−0.11, 0.03) | 0.04 | 0.05 |
| Mali | PROMIS CS (46) | | | | | | |
| Zimbabwe | SHINE (HIV−) (47) | 1393 | 1305 | 34.6 | −0.07 (−0.10, −0.03) | 0.14 | 0.13 |
| Zimbabwe | SHINE (HIV+) (48) | 236 | 239 | 46.4 | −0.07 (−0.16, 0.02) | 0.02 | 0.03 |
| | | **10213** | **12932** | | **I² = 0.23, Tau² = 0.00** | | |
| **Fixed** | | | | | **−0.05 (−0.06, −0.03)** | | |
| **Random** | | | | | **−0.05 (−0.06, −0.03)** | | |

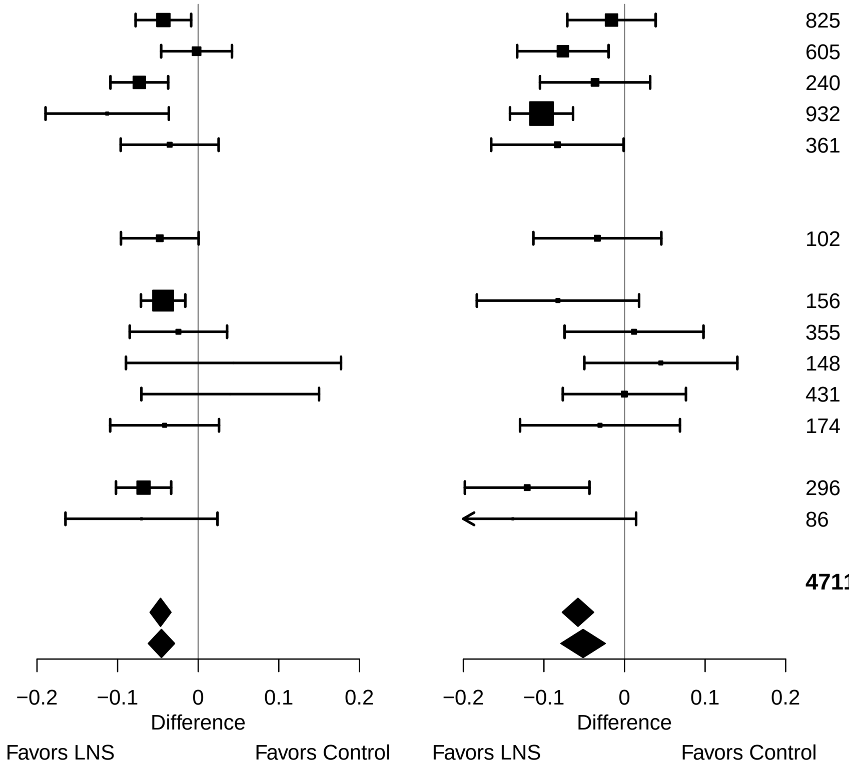

**Moderate to severe**

| Country | Trial | LNS N | Control N | Control Prevalence | PD (95% CI) | Fixed W | Random W |
|---|---|---|---|---|---|---|---|
| Bangladesh | JiVitA−4 (34) | 825 | 381 | 50.9 | −0.02 (−0.07, 0.04) | 0.13 | 0.11 |
| Bangladesh | RDNS (35) | 605 | 322 | 50.6 | −0.08 (−0.13, −0.02) | 0.12 | 0.11 |
| Bangladesh | WASH−B (36) | 240 | 779 | 50.3 | −0.04 (−0.10, 0.03) | 0.08 | 0.09 |
| Burkina Faso | iLiNS−Zinc (37) | 932 | 343 | 38.2 | −0.10 (−0.14, −0.06) | 0.25 | 0.14 |
| Burkina Faso | PROMIS (38) | 361 | 364 | 31.9 | −0.08 (−0.17, 0.00) | 0.06 | 0.07 |
| Burkina Faso | PROMIS CS (38) | | | | | | |
| Ghana | GHANA (39) | | | | | | |
| Ghana | iLiNS−DYADG (40) | 102 | 219 | 14.2 | −0.03 (−0.11, 0.05) | 0.06 | 0.07 |
| Haiti | HAITI (41) | | | | | | |
| Kenya | WASH−B (42) | 156 | 525 | 41.0 | −0.08 (−0.18, 0.02) | 0.04 | 0.05 |
| Madagascar | MAHAY (43) | 355 | 410 | 58.5 | 0.01 (−0.07, 0.10) | 0.05 | 0.07 |
| Malawi | iLiNS−DYADM (44) | 148 | 316 | 36.7 | 0.05 (−0.05, 0.14) | 0.04 | 0.06 |
| Malawi | iLiNS−DOSE (45) | 431 | 146 | 48.6 | 0.00 (−0.08, 0.08) | 0.06 | 0.08 |
| Mali | PROMIS (46) | 174 | 163 | 38.0 | −0.03 (−0.13, 0.07) | 0.04 | 0.05 |
| Mali | PROMIS CS (46) | | | | | | |
| Zimbabwe | SHINE (HIV−) (47) | 296 | 325 | 38.8 | −0.12 (−0.20, −0.04) | 0.06 | 0.08 |
| Zimbabwe | SHINE (HIV+) (48) | 86 | 76 | 59.2 | −0.14 (−0.29, 0.01) | 0.02 | 0.03 |
| | | **4711** | **4369** | | **I² = 0.46, Tau² = 0.00** | | |
| **Fixed** | | | | | **−0.06 (−0.08, −0.04)** | | |
| **Random** | | | | | **−0.05 (−0.08, −0.02)** | | |

#### Supplemental figure 7C: Stunting prevalence difference

#### 7C3: Stratified by Household source water quality

**P−for−interaction = 0.469**
**Difference in PDs = −0.01 (−0.04, 0.02)**

| Country | Trial | Improved LNS N | Improved Control N | Improved Control Prevalence | Improved PD (95% CI) | Improved Fixed W | Improved Random W | Unimproved LNS N | Unimproved Control N | Unimproved Control Prevalence | Unimproved PD (95% CI) | Unimproved Fixed W | Unimproved Random W |
|---|---|---|---|---|---|---|---|---|---|---|---|---|---|
| Bangladesh | JiVitA−4 (34) | | | | | | | | | | | | |
| Bangladesh | RDNS (35) | | | | | | | | | | | | |
| Bangladesh | WASH−B (36) | 496 | 989 | 40.7 | −0.09 (−0.14, −0.05) | 0.17 | 0.16 | 70 | 114 | 43.0 | 0.00 (−0.15, 0.14) | 0.03 | 0.05 |
| Burkina Faso | iLiNS−Zinc (37) | 550 | 141 | 35.5 | −0.04 (−0.11, 0.02) | 0.09 | 0.10 | 1394 | 519 | 40.2 | −0.13 (−0.17, −0.08) | 0.29 | 0.16 |
| Burkina Faso | PROMIS (38) | 355 | 379 | 31.1 | −0.02 (−0.09, 0.04) | 0.09 | 0.09 | 361 | 355 | 31.5 | −0.12 (−0.22, −0.02) | 0.05 | 0.08 |
| Burkina Faso | PROMIS CS (38) | 280 | 268 | 25.4 | −0.06 (−0.14, 0.02) | 0.06 | 0.06 | 149 | 171 | 22.8 | 0.01 (−0.08, 0.11) | 0.06 | 0.09 |
| Ghana | GHANA (39) | | | | | | | | | | | | |
| Ghana | iLiNS−DYADG (40) | | | | | | | | | | | | |
| Haiti | HAITI (41) | | | | | | | | | | | | |
| Kenya | WASH−B (42) | 447 | 1549 | 30.6 | −0.01 (−0.07, 0.04) | 0.14 | 0.13 | 248 | 697 | 33.1 | −0.05 (−0.12, 0.02) | 0.10 | 0.11 |
| Madagascar | MAHAY (43) | 419 | 453 | 51.4 | 0.01 (−0.08, 0.09) | 0.05 | 0.06 | 1203 | 1180 | 61.2 | −0.02 (−0.08, 0.03) | 0.19 | 0.14 |
| Malawi | iLiNS−DYADM (44) | | | | | | | | | | | | |
| Malawi | iLiNS−DOSE (45) | 555 | 193 | 46.1 | −0.02 (−0.08, 0.05) | 0.09 | 0.09 | 52 | 17 | 47.1 | 0.15 (−0.07, 0.38) | 0.01 | 0.02 |
| Mali | PROMIS (46) | 249 | 258 | 36.4 | −0.08 (−0.15, −0.01) | 0.08 | 0.08 | 247 | 243 | 36.2 | 0.01 (−0.07, 0.09) | 0.09 | 0.11 |
| Mali | PROMIS CS (46) | 549 | 566 | 31.4 | −0.06 (−0.13, 0.00) | 0.09 | 0.09 | 362 | 388 | 37.6 | −0.08 (−0.16, −0.01) | 0.09 | 0.11 |
| Zimbabwe | SHINE (HIV−) (47) | 521 | 489 | 35.2 | −0.08 (−0.13, −0.02) | 0.12 | 0.12 | 274 | 302 | 32.5 | −0.01 (−0.09, 0.07) | 0.08 | 0.10 |
| Zimbabwe | SHINE (HIV+) (48) | 85 | 87 | 49.4 | −0.12 (−0.26, 0.02) | 0.02 | 0.02 | 55 | 56 | 57.1 | −0.14 (−0.34, 0.07) | 0.01 | 0.03 |
| | | **4506** | **5372** | | **I² = 0.12, Tau² = 0.00** | | | **4415** | **4042** | | **I² = 0.59, Tau² = 0.00** | | |
| **Fixed** | | | | | **−0.05 (−0.07, −0.03)** | | | | | | **−0.06 (−0.08, −0.04)** | | |
| **Random** | | | | | **−0.05 (−0.07, −0.03)** | | | | | | **−0.05 (−0.08, −0.01)** | | |

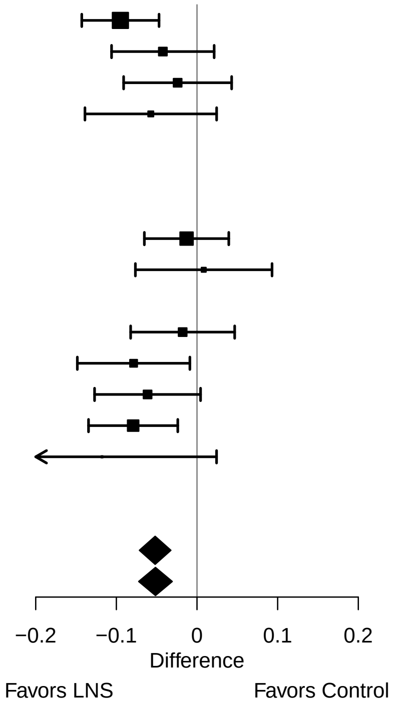

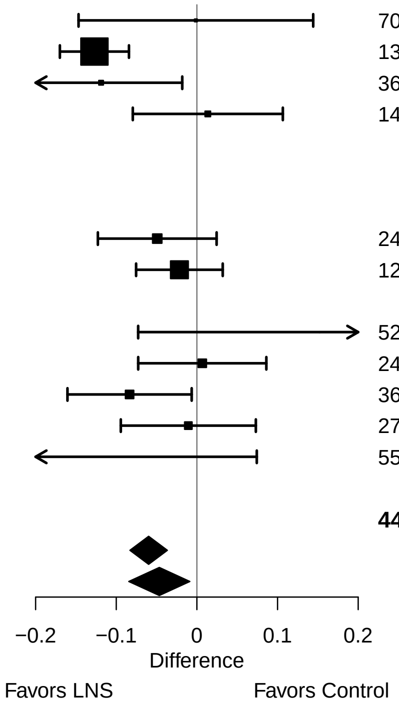

#### Supplemental figure 7C: Stunting prevalence difference

#### 7C4: Stratified by Household sanitation

**P−for−interaction = 0.310**
**Difference in PDs = 0.02 (−0.01, 0.05)**

| Country | Trial | Improved LNS N | Improved Control N | Improved Control Prevalence | Improved PD (95% CI) | Improved Fixed W | Improved Random W | Unimproved LNS N | Unimproved Control N | Unimproved Control Prevalence | Unimproved PD (95% CI) | Unimproved Fixed W | Unimproved Random W |
|---|---|---|---|---|---|---|---|---|---|---|---|---|---|
| Bangladesh | JiVitA−4 (34) | 2189 | 950 | 42.5 | −0.04 (−0.07, 0.00) | 0.28 | 0.28 | 648 | 294 | 49.7 | −0.03 (−0.09, 0.03) | 0.13 | 0.13 |
| Bangladesh | RDNS (35) | 1179 | 573 | 39.6 | −0.05 (−0.09, −0.01) | 0.21 | 0.21 | 482 | 241 | 47.7 | 0.00 (−0.07, 0.08) | 0.08 | 0.09 |
| Bangladesh | WASH−B (36) | 506 | 1001 | 39.7 | −0.09 (−0.14, −0.03) | 0.11 | 0.11 | 31 | 54 | 51.9 | −0.13 (−0.34, 0.08) | 0.01 | 0.01 |
| Burkina Faso | iLiNS−Zinc (37) | 43 | 16 | 37.5 | −0.05 (−0.20, 0.10) | 0.01 | 0.01 | 1901 | 644 | 39.2 | −0.11 (−0.15, −0.06) | 0.20 | 0.18 |
| Burkina Faso | PROMIS (38) | 398 | 550 | 31.3 | −0.07 (−0.14, 0.00) | 0.07 | 0.07 | 465 | 359 | 29.2 | −0.04 (−0.10, 0.02) | 0.10 | 0.11 |
| Burkina Faso | PROMIS CS (38) | 176 | 165 | 24.2 | −0.03 (−0.13, 0.07) | 0.03 | 0.03 | 252 | 274 | 24.5 | −0.03 (−0.11, 0.05) | 0.06 | 0.07 |
| Ghana | GHANA (39) | | | | | | | | | | | | |
| Ghana | iLiNS−DYADG (40) | | | | | | | | | | | | |
| Haiti | HAITI (41) | | | | | | | | | | | | |
| Kenya | WASH−B (42) | 100 | 364 | 24.7 | −0.03 (−0.11, 0.06) | 0.05 | 0.05 | 595 | 1883 | 32.7 | −0.03 (−0.08, 0.02) | 0.16 | 0.15 |
| Madagascar | MAHAY (43) | | | | | | | | | | | | |
| Malawi | iLiNS−DYADM (44) | | | | | | | | | | | | |
| Malawi | iLiNS−DOSE (45) | | | | | | | | | | | | |
| Mali | PROMIS (46) | 387 | 366 | 36.1 | −0.06 (−0.12, 0.01) | 0.07 | 0.07 | 112 | 135 | 37.8 | 0.01 (−0.10, 0.12) | 0.03 | 0.04 |
| Mali | PROMIS CS (46) | 701 | 696 | 32.6 | −0.08 (−0.14, −0.01) | 0.08 | 0.08 | 220 | 248 | 35.9 | −0.07 (−0.16, 0.02) | 0.05 | 0.06 |
| Zimbabwe | SHINE (HIV−) (47) | 295 | 244 | 32.0 | −0.06 (−0.13, 0.01) | 0.06 | 0.06 | 497 | 546 | 35.2 | −0.05 (−0.10, 0.00) | 0.16 | 0.15 |
| Zimbabwe | SHINE (HIV+) (48) | 50 | 35 | 54.3 | −0.16 (−0.36, 0.04) | 0.01 | 0.01 | 90 | 107 | 51.4 | −0.10 (−0.26, 0.05) | 0.02 | 0.02 |
| | | **6024** | **4960** | | **I² = 0.00, Tau² = 0.00** | | | **5293** | **4785** | | **I² = 0.21, Tau² = 0.00** | | |
| **Fixed** | | | | | **−0.05 (−0.07, −0.04)** | | | | | | **−0.05 (−0.07, −0.03)** | | |
| **Random** | | | | | **−0.05 (−0.07, −0.04)** | | | | | | **−0.05 (−0.07, −0.03)** | | |

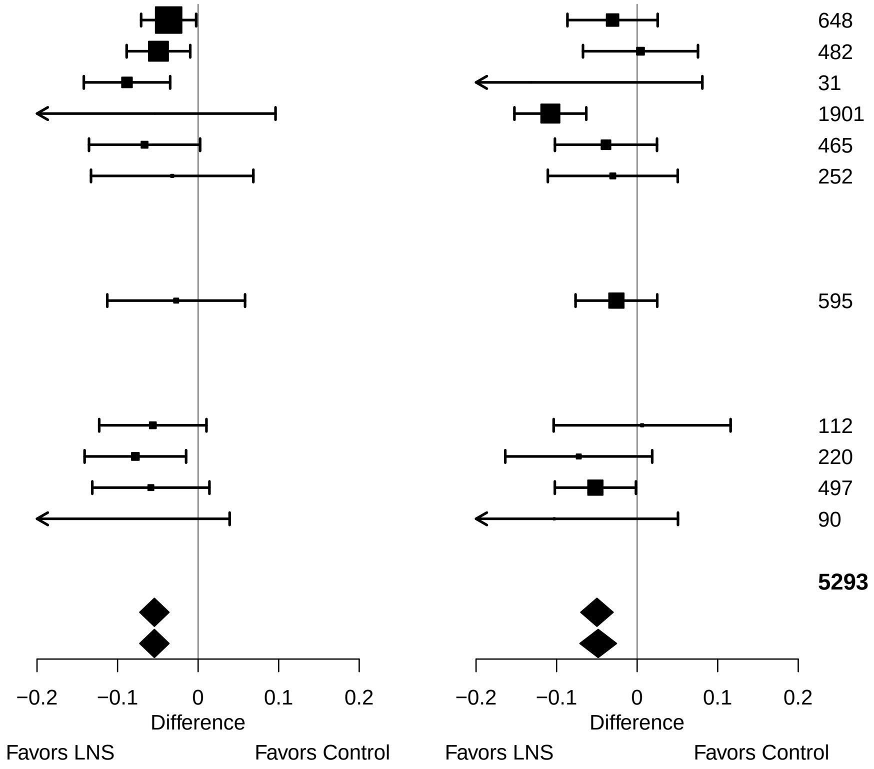

#### Supplemental figure 7C: Stunting prevalence difference

#### 7C5: Stratified by Home environment

**P−for−interaction = 0.228**
**Difference in PDs = −0.02 (−0.05, 0.01)**

| Country | Trial | LNS N | Control N | Control Prevalence | At least median PD (95% CI) | Fixed W | Random W | LNS N | Control N | Control Prevalence | Less than median PD (95% CI) | Fixed W | Random W |
|---|---|---|---|---|---|---|---|---|---|---|---|---|---|
| Bangladesh | JiVitA−4 (34) | | | | | | | | | | | | |
| Bangladesh | RDNS (35) | 1192 | 582 | 39.2 | −0.02 (−0.07, 0.02) | 0.17 | 0.15 | 468 | 230 | 48.7 | −0.06 (−0.12, 0.00) | 0.16 | 0.16 |
| Bangladesh | WASH−B (36) | 637 | 1653 | 36.9 | −0.06 (−0.10, −0.02) | 0.18 | 0.16 | 493 | 1690 | 46.9 | −0.06 (−0.11, −0.01) | 0.20 | 0.20 |
| Burkina Faso | iLiNS−Zinc (37) | 473 | 199 | 37.7 | −0.12 (−0.18, −0.06) | 0.09 | 0.11 | 271 | 174 | 41.7 | −0.08 (−0.17, 0.00) | 0.07 | 0.07 |
| Burkina Faso | PROMIS (38) | | | | | | | | | | | | |
| Burkina Faso | PROMIS CS (38) | | | | | | | | | | | | |
| Ghana | GHANA (39) | | | | | | | | | | | | |
| Ghana | iLiNS−DYADG (40) | 213 | 409 | 12.0 | −0.03 (−0.08, 0.03) | 0.12 | 0.13 | 119 | 248 | 15.3 | −0.07 (−0.14, 0.00) | 0.09 | 0.09 |
| Haiti | HAITI (41) | | | | | | | | | | | | |
| Kenya | WASH−B (42) | 835 | 2764 | 29.6 | −0.03 (−0.07, 0.00) | 0.27 | 0.18 | 574 | 2160 | 34.3 | −0.05 (−0.09, 0.00) | 0.26 | 0.26 |
| Madagascar | MAHAY (43) | 915 | 907 | 57.9 | 0.02 (−0.04, 0.09) | 0.08 | 0.11 | 787 | 775 | 59.2 | −0.05 (−0.11, 0.01) | 0.13 | 0.13 |
| Malawi | iLiNS−DYADM (44) | 137 | 289 | 33.6 | 0.02 (−0.07, 0.12) | 0.04 | 0.07 | 78 | 149 | 36.9 | 0.07 (−0.07, 0.20) | 0.03 | 0.03 |
| Malawi | iLiNS−DOSE (45) | 375 | 136 | 49.3 | −0.03 (−0.11, 0.04) | 0.06 | 0.09 | 275 | 86 | 40.7 | 0.03 (−0.07, 0.12) | 0.05 | 0.05 |
| Mali | PROMIS (46) | | | | | | | | | | | | |
| Mali | PROMIS CS (46) | | | | | | | | | | | | |
| Zimbabwe | SHINE (HIV−) (47) | | | | | | | | | | | | |
| Zimbabwe | SHINE (HIV+) (48) | | | | | | | | | | | | |
| | | **4777** | **6939** | | **I² = 0.50, Tau² = 0.00** | | | **3065** | **5512** | | **I² = 0.00, Tau² = 0.00** | | |
| **Fixed** | | | | | **−0.04 (−0.05, −0.02)** | | | | | | **−0.05 (−0.07, −0.03)** | | |
| **Random** | | | | | **−0.03 (−0.06, −0.01)** | | | | | | **−0.05 (−0.07, −0.03)** | | |

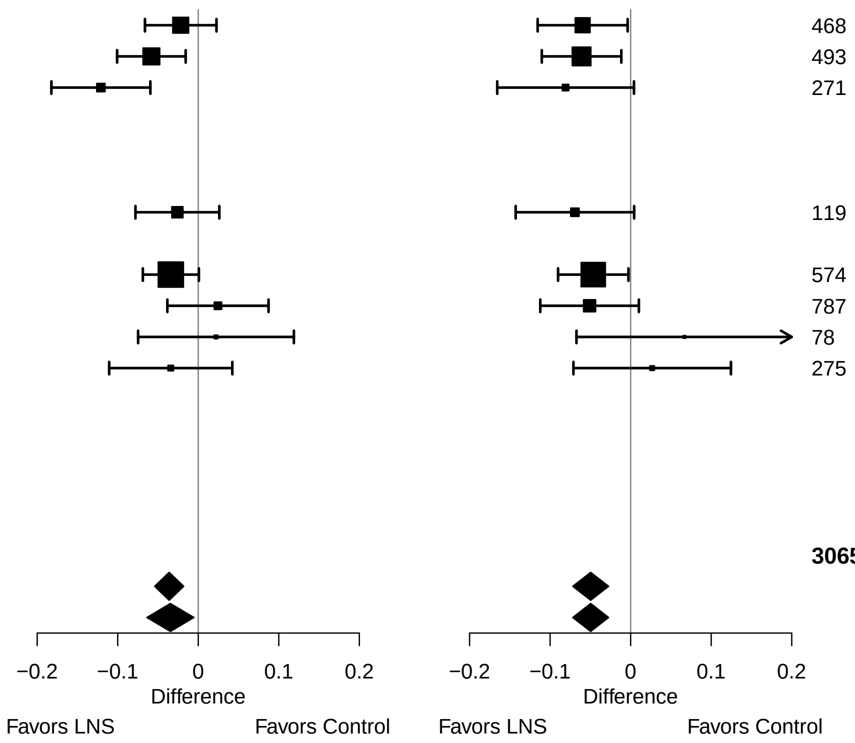

#### Supplemental figure 7C: Stunting prevalence difference

#### 7C6: Stratified by Season at the time of assessment

**P−for−interaction = 0.493**
**Difference in PDs = −0.01 (−0.03, 0.02)**

| Country | Trial | Dry LNS N | Dry Control N | Dry Control Prevalence | Dry PD (95% CI) | Dry Fixed W | Dry Random W | Rainy LNS N | Rainy Control N | Rainy Control Prevalence | Rainy PD (95% CI) | Rainy Fixed W | Rainy Random W |
|---|---|---|---|---|---|---|---|---|---|---|---|---|---|
| Bangladesh | JiVitA−4 (34) | 1550 | 663 | 44.9 | −0.03 (−0.07, 0.01) | 0.15 | 0.12 | 1288 | 581 | 43.4 | −0.04 (−0.09, 0.00) | 0.13 | 0.13 |
| Bangladesh | RDNS (35) | 739 | 359 | 43.7 | −0.04 (−0.10, 0.02) | 0.06 | 0.08 | 924 | 456 | 40.6 | −0.03 (−0.07, 0.01) | 0.17 | 0.17 |
| Bangladesh | WASH−B (36) | 640 | 1913 | 39.1 | −0.06 (−0.10, −0.01) | 0.13 | 0.11 | 518 | 1518 | 45.5 | −0.08 (−0.13, −0.04) | 0.14 | 0.14 |
| Burkina Faso | iLiNS−Zinc (37) | 1394 | 477 | 40.3 | −0.10 (−0.14, −0.07) | 0.14 | 0.11 | 558 | 187 | 37.2 | −0.11 (−0.20, −0.03) | 0.04 | 0.04 |
| Burkina Faso | PROMIS (38) | 434 | 446 | 27.1 | −0.04 (−0.11, 0.02) | 0.05 | 0.07 | 429 | 468 | 33.8 | −0.06 (−0.13, 0.00) | 0.06 | 0.06 |
| Burkina Faso | PROMIS CS (38) | 224 | 190 | 25.3 | −0.05 (−0.15, 0.05) | 0.02 | 0.04 | 206 | 249 | 23.7 | −0.01 (−0.12, 0.10) | 0.02 | 0.02 |
| Ghana | GHANA (39) | | | | | | | | | | | | |
| Ghana | iLiNS−DYADG (40) | 204 | 407 | 12.8 | −0.03 (−0.08, 0.02) | 0.07 | 0.09 | 143 | 285 | 13.3 | −0.06 (−0.13, 0.00) | 0.07 | 0.07 |
| Haiti | HAITI (41) | | | | | | | | | | | | |
| Kenya | WASH−B (42) | 347 | 1151 | 30.1 | −0.06 (−0.11, 0.00) | 0.07 | 0.09 | 1110 | 3986 | 32.8 | −0.04 (−0.07, −0.01) | 0.29 | 0.29 |
| Madagascar | MAHAY (43) | | | | | | | | | | | | |
| Malawi | iLiNS−DYADM (44) | 156 | 316 | 32.9 | 0.04 (−0.05, 0.13) | 0.02 | 0.05 | 64 | 128 | 39.1 | 0.02 (−0.13, 0.16) | 0.01 | 0.01 |
| Malawi | iLiNS−DOSE (45) | 415 | 134 | 44.0 | 0.03 (−0.04, 0.11) | 0.04 | 0.06 | 281 | 107 | 49.5 | −0.08 (−0.17, 0.01) | 0.03 | 0.03 |
| Mali | PROMIS (46) | | | | | | | | | | | | |
| Mali | PROMIS CS (46) | | | | | | | | | | | | |
| Zimbabwe | SHINE (HIV−) (47) | 1747 | 1662 | 35.4 | −0.08 (−0.11, −0.05) | 0.22 | 0.13 | 133 | 132 | 28.0 | 0.01 (−0.10, 0.13) | 0.02 | 0.02 |
| Zimbabwe | SHINE (HIV+) (48) | 319 | 306 | 50.7 | −0.10 (−0.18, −0.01) | 0.03 | 0.05 | 18 | 24 | 41.7 | −0.14 (−0.42, 0.14) | 0.00 | 0.00 |
| | | **8169** | **8024** | | **I² = 0.51, Tau² = 0.00** | | | **5672** | **8121** | | **I² = 0.00, Tau² = 0.00** | | |
| **Fixed** | | | | | **−0.06 (−0.07, −0.04)** | | | | | | **−0.05 (−0.07, −0.03)** | | |
| **Random** | | | | | **−0.05 (−0.07, −0.03)** | | | | | | **−0.05 (−0.07, −0.03)** | | |

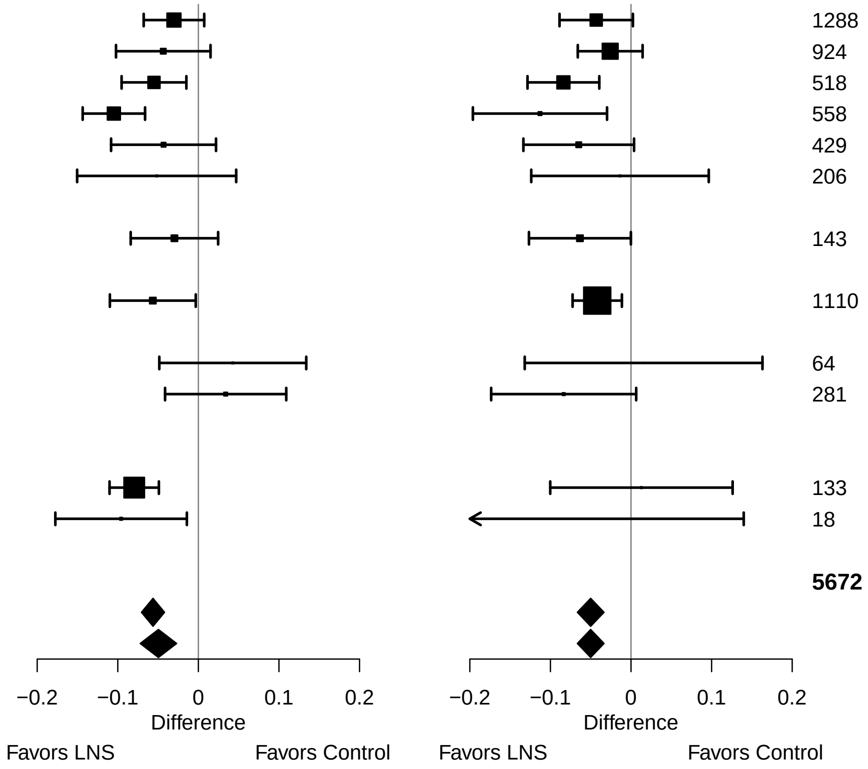

### Supplemental figure 7D: Mean difference in WLZ

#### 7D1: Stratified by Household socio-economic status

**P−for−interaction = 0.814**
**Difference in MDs = 0.00 (−0.03, 0.04)**

| Country | Trial | At least median: LNS N | Control N | Control Mean | MD (95% CI) | Fixed W | Random W | Less than median: LNS N | Control N | Control Mean | MD (95% CI) | Fixed W | Random W |
|---|---|---|---|---|---|---|---|---|---|---|---|---|---|
| Bangladesh | JiVitA−4 (34) | 1401 | 607 | −1.10 | 0.10 (0.04, 0.16) | 0.22 | 0.15 | 1381 | 610 | −1.24 | 0.06 (−0.01, 0.12) | 0.20 | 0.13 |
| Bangladesh | RDNS (35) | 834 | 407 | −1.01 | 0.04 (−0.07, 0.15) | 0.07 | 0.08 | 827 | 408 | −1.21 | 0.10 (0.01, 0.19) | 0.10 | 0.10 |
| Bangladesh | WASH−B (36) | 579 | 1714 | −0.72 | 0.12 (0.02, 0.21) | 0.09 | 0.09 | 577 | 1710 | −1.02 | 0.13 (0.04, 0.22) | 0.10 | 0.10 |
| Burkina Faso | iLiNS−Zinc (37) | 1225 | 299 | −0.90 | 0.19 (0.09, 0.29) | 0.08 | 0.09 | 720 | 364 | −0.97 | 0.16 (0.09, 0.23) | 0.15 | 0.12 |
| Burkina Faso | PROMIS (38) | 405 | 479 | −0.66 | 0.05 (−0.07, 0.17) | 0.06 | 0.06 | 451 | 428 | −0.77 | 0.16 (−0.04, 0.35) | 0.02 | 0.03 |
| Burkina Faso | PROMIS CS (38) | 220 | 208 | −0.91 | −0.06 (−0.25, 0.13) | 0.02 | 0.03 | 210 | 228 | −0.86 | −0.16 (−0.34, 0.02) | 0.03 | 0.04 |
| Ghana | GHANA (39) | 57 | 44 | −0.41 | 0.06 (−0.46, 0.57) | 0.00 | 0.00 | 41 | 46 | −0.84 | 0.30 (−0.15, 0.74) | 0.00 | 0.01 |
| Ghana | iLiNS−DYADG (40) | 157 | 356 | −0.53 | 0.10 (−0.09, 0.29) | 0.02 | 0.03 | 190 | 334 | −0.65 | 0.10 (−0.08, 0.28) | 0.03 | 0.04 |
| Haiti | HAITI (41) | 74 | 60 | 0.23 | −0.04 (−0.28, 0.20) | 0.01 | 0.02 | 61 | 72 | −0.22 | 0.15 (−0.07, 0.37) | 0.02 | 0.03 |
| Kenya | WASH−B (42) | 825 | 2914 | 0.13 | 0.07 (−0.01, 0.15) | 0.13 | 0.11 | 628 | 2200 | 0.05 | 0.06 (−0.03, 0.15) | 0.11 | 0.10 |
| Madagascar | MAHAY (43) | 771 | 884 | −0.31 | 0.01 (−0.12, 0.13) | 0.05 | 0.06 | 876 | 755 | −0.56 | 0.04 (−0.10, 0.19) | 0.04 | 0.05 |
| Malawi | iLiNS−DYADM (44) | 121 | 234 | −0.12 | −0.04 (−0.24, 0.17) | 0.02 | 0.03 | 99 | 209 | −0.14 | −0.08 (−0.33, 0.18) | 0.01 | 0.02 |
| Malawi | iLiNS−DOSE (45) | 301 | 104 | −0.13 | −0.04 (−0.21, 0.13) | 0.03 | 0.04 | 294 | 103 | −0.38 | 0.11 (−0.10, 0.32) | 0.02 | 0.03 |
| Mali | PROMIS (46) | 246 | 250 | −0.32 | 0.13 (−0.01, 0.27) | 0.04 | 0.05 | 245 | 246 | −0.32 | 0.06 (−0.09, 0.22) | 0.03 | 0.05 |
| Mali | PROMIS CS (46) | 501 | 458 | −0.77 | 0.17 (0.04, 0.30) | 0.05 | 0.06 | 443 | 501 | −0.77 | 0.15 (0.01, 0.29) | 0.04 | 0.05 |
| Zimbabwe | SHINE (HIV−) (47) | 903 | 797 | 0.08 | 0.04 (−0.07, 0.14) | 0.08 | 0.08 | 826 | 856 | −0.06 | 0.15 (0.06, 0.25) | 0.09 | 0.09 |
| Zimbabwe | SHINE (HIV+) (48) | 171 | 156 | 0.06 | −0.16 (−0.42, 0.09) | 0.01 | 0.02 | 157 | 166 | −0.03 | −0.11 (−0.38, 0.15) | 0.01 | 0.02 |
| | | **8791** | **9971** | | **I² = 0.23, Tau² = 0.00** | | | **8026** | **9236** | | **I² = 0.27, Tau² = 0.00** | | |
| **Fixed** | | | | | **0.08 (0.05, 0.11)** | | | | | | **0.10 (0.07, 0.12)** | | |
| **Random** | | | | | **0.07 (0.03, 0.11)** | | | | | | **0.09 (0.05, 0.13)** | | |

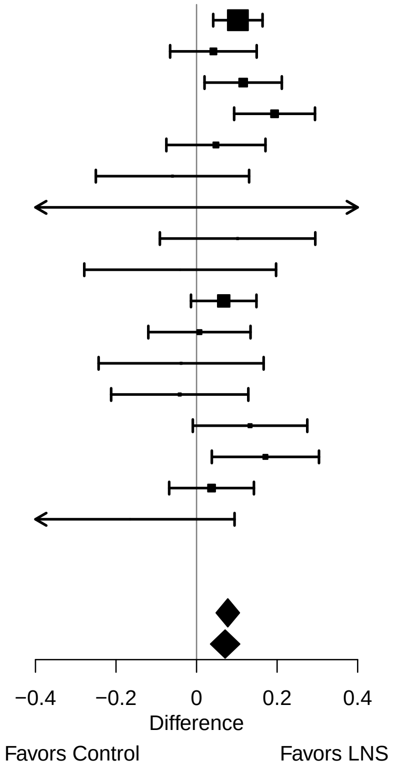

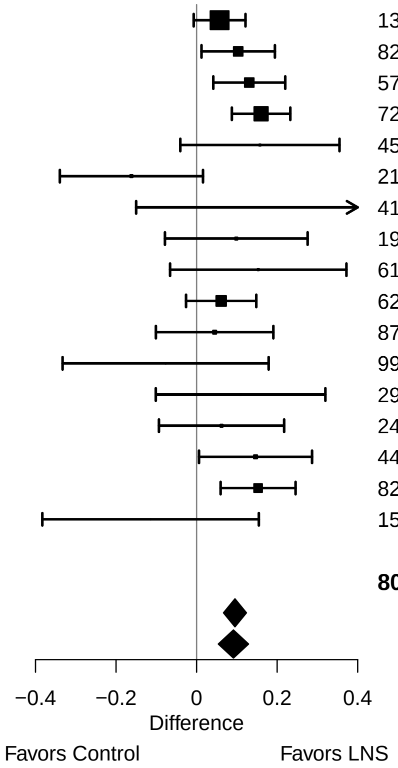

#### Supplemental figure 7D: Mean difference in WLZ

##### 7D2: Stratified by Household food insecurity

P−for−interaction = 0.813
Difference in MDs = 0.01 (−0.04, 0.05)

| Country | Trial | Mild to secure: LNS N | Control N | Control Mean | MD (95% CI) | Fixed W | Random W | Moderate to severe: LNS N | Control N | Control Mean | MD (95% CI) | Fixed W | Random W |
|---|---|---|---|---|---|---|---|---|---|---|---|---|---|
| Bangladesh | JiVitA−4 (34) | 1977 | 841 | −1.14 | 0.09 (0.03, 0.14) | 0.23 | 0.18 | 805 | 376 | −1.22 | 0.06 (−0.02, 0.14) | 0.25 | 0.19 |
| Bangladesh | RDNS (35) | 1057 | 493 | −1.02 | 0.01 (−0.09, 0.11) | 0.08 | 0.09 | 604 | 322 | −1.24 | 0.16 (0.04, 0.28) | 0.11 | 0.11 |
| Bangladesh | WASH−B (36) | 917 | 2647 | −0.84 | 0.13 (0.06, 0.21) | 0.13 | 0.13 | 239 | 777 | −1.01 | 0.07 (−0.06, 0.19) | 0.10 | 0.10 |
| Burkina Faso | iLiNS−Zinc (37) | 1013 | 320 | −0.87 | 0.16 (0.07, 0.25) | 0.09 | 0.10 | 932 | 343 | −1.00 | 0.21 (0.12, 0.30) | 0.19 | 0.17 |
| Burkina Faso | PROMIS (38) | 501 | 548 | −0.72 | 0.14 (0.00, 0.29) | 0.04 | 0.05 | 355 | 359 | −0.70 | 0.04 (−0.17, 0.24) | 0.03 | 0.04 |
| Burkina Faso | PROMIS CS (38) | | | | | | | | | | | | |
| Ghana | GHANA (39) | | | | | | | | | | | | |
| Ghana | iLiNS−DYADG (40) | 244 | 470 | −0.57 | 0.09 (−0.07, 0.25) | 0.03 | 0.04 | 102 | 219 | −0.61 | 0.09 (−0.13, 0.31) | 0.03 | 0.04 |
| Haiti | HAITI (41) | | | | | | | | | | | | |
| Kenya | WASH−B (42) | 1297 | 4577 | 0.09 | 0.07 (0.01, 0.14) | 0.18 | 0.15 | 156 | 524 | 0.10 | −0.02 (−0.21, 0.17) | 0.04 | 0.05 |
| Madagascar | MAHAY (43) | 987 | 927 | −0.41 | 0.03 (−0.10, 0.15) | 0.04 | 0.06 | 354 | 410 | −0.48 | 0.02 (−0.16, 0.19) | 0.05 | 0.06 |
| Malawi | iLiNS−DYADM (44) | 70 | 122 | 0.02 | −0.16 (−0.45, 0.13) | 0.01 | 0.01 | 148 | 316 | −0.19 | −0.01 (−0.21, 0.18) | 0.04 | 0.05 |
| Malawi | iLiNS−DOSE (45) | 151 | 55 | −0.26 | 0.05 (−0.21, 0.31) | 0.01 | 0.02 | 431 | 146 | −0.28 | 0.06 (−0.10, 0.22) | 0.06 | 0.07 |
| Mali | PROMIS (46) | 326 | 338 | −0.33 | 0.12 (−0.01, 0.25) | 0.04 | 0.05 | 170 | 163 | −0.34 | 0.08 (−0.13, 0.29) | 0.03 | 0.04 |
| Mali | PROMIS CS (46) | | | | | | | | | | | | |
| Zimbabwe | SHINE (HIV−) (47) | 1384 | 1297 | 0.03 | 0.08 (0.00, 0.17) | 0.10 | 0.11 | 294 | 324 | −0.07 | 0.17 (0.01, 0.33) | 0.06 | 0.07 |
| Zimbabwe | SHINE (HIV+) (48) | 236 | 238 | −0.02 | −0.12 (−0.33, 0.09) | 0.02 | 0.02 | 85 | 76 | 0.09 | −0.18 (−0.54, 0.17) | 0.01 | 0.02 |
| | | **10160** | **12873** | | **I² = 0.18, Tau² = 0.00** | | | **4675** | **4355** | | **I² = 0.23, Tau² = 0.00** | | |
| **Fixed** | | | | | **0.09 (0.06, 0.11)** | | | | | | **0.10 (0.06, 0.13)** | | |
| **Random** | | | | | **0.08 (0.05, 0.12)** | | | | | | **0.09 (0.05, 0.14)** | | |

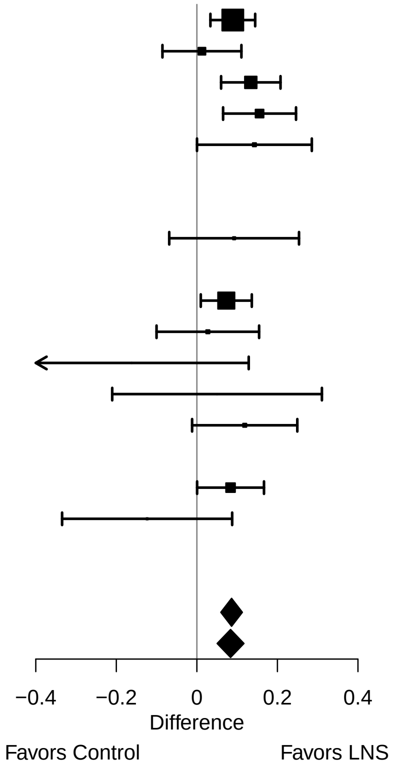

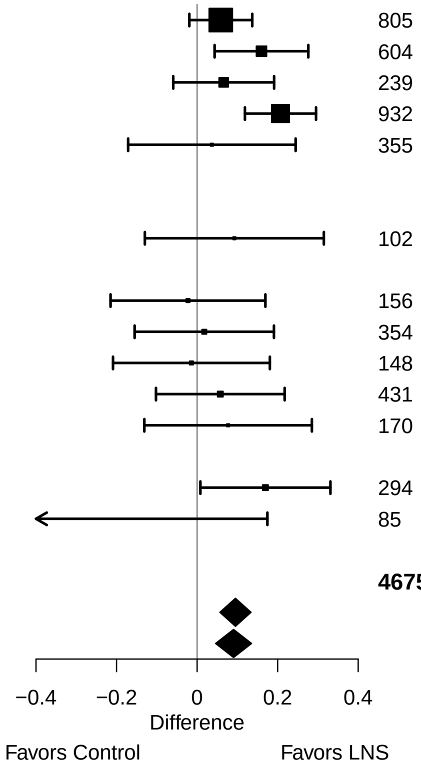

#### Supplemental figure 7D: Mean difference in WLZ

##### 7D3: Stratified by Household source water quality

**P−for−interaction = 0.860**
**Difference in MDs = 0.01 (−0.05, 0.07)**

| | | Improved | | | | | | Unimproved | | | | | |
|---|---|---|---|---|---|---|---|---|---|---|---|---|---|
| **Country** | **Trial** | **LNS N** | **Control N** | **Control Mean** | **MD (95% CI)** | **Fixed W** | **Random W** | **LNS N** | **Control N** | **Control Mean** | **MD (95% CI)** | **Fixed W** | **Random W** |
| Bangladesh | JiVitA−4 (34) | | | | | | | | | | | | |
| Bangladesh | RDNS (35) | | | | | | | | | | | | |
| Bangladesh | WASH−B (36) | 495 | 986 | −0.87 | 0.19 (0.07, 0.31) | 0.11 | 0.10 | 70 | 114 | −0.90 | −0.06 (−0.36, 0.25) | 0.02 | 0.04 |
| Burkina Faso | iLiNS−Zinc (37) | 550 | 141 | −0.89 | 0.16 (0.05, 0.26) | 0.15 | 0.12 | 1394 | 519 | −0.96 | 0.19 (0.12, 0.27) | 0.36 | 0.21 |
| Burkina Faso | PROMIS (38) | 352 | 377 | −0.71 | 0.03 (−0.13, 0.19) | 0.06 | 0.07 | 357 | 350 | −0.70 | 0.16 (−0.02, 0.35) | 0.06 | 0.08 |
| Burkina Faso | PROMIS CS (38) | 280 | 267 | −0.91 | −0.09 (−0.30, 0.11) | 0.04 | 0.05 | 149 | 169 | −0.83 | −0.13 (−0.31, 0.04) | 0.07 | 0.09 |
| Ghana | GHANA (39) | | | | | | | | | | | | |
| Ghana | iLiNS−DYADG (40) | | | | | | | | | | | | |
| Haiti | HAITI (41) | | | | | | | | | | | | |
| Kenya | WASH−B (42) | 447 | 1542 | 0.12 | 0.04 (−0.06, 0.14) | 0.15 | 0.12 | 247 | 695 | 0.06 | 0.03 (−0.11, 0.17) | 0.11 | 0.12 |
| Madagascar | MAHAY (43) | 418 | 453 | −0.36 | 0.00 (−0.17, 0.18) | 0.05 | 0.06 | 1202 | 1180 | −0.45 | 0.01 (−0.11, 0.12) | 0.15 | 0.15 |
| Malawi | iLiNS−DYADM (44) | 200 | 406 | −0.12 | −0.06 (−0.23, 0.10) | 0.06 | 0.07 | 20 | 36 | −0.28 | 0.07 (−0.61, 0.74) | 0.00 | 0.01 |
| Malawi | iLiNS−DOSE (45) | 555 | 193 | −0.24 | 0.04 (−0.10, 0.18) | 0.08 | 0.09 | 52 | 17 | −0.42 | −0.03 (−0.45, 0.39) | 0.01 | 0.02 |
| Mali | PROMIS (46) | 247 | 257 | −0.26 | 0.13 (−0.01, 0.27) | 0.08 | 0.09 | 242 | 240 | −0.41 | 0.08 (−0.11, 0.26) | 0.06 | 0.08 |
| Mali | PROMIS CS (46) | 544 | 562 | −0.74 | 0.15 (0.04, 0.25) | 0.14 | 0.12 | 361 | 382 | −0.79 | 0.17 (0.00, 0.34) | 0.07 | 0.09 |
| Zimbabwe | SHINE (HIV−) (47) | 518 | 486 | 0.01 | 0.08 (−0.06, 0.22) | 0.08 | 0.09 | 273 | 301 | 0.10 | 0.00 (−0.17, 0.17) | 0.07 | 0.09 |
| Zimbabwe | SHINE (HIV+) (48) | 85 | 87 | −0.01 | −0.13 (−0.45, 0.18) | 0.02 | 0.02 | 55 | 56 | −0.02 | −0.11 (−0.55, 0.34) | 0.01 | 0.02 |
| | | **4691** | **5757** | | **I² = 0.35, Tau² = 0.00** | | | **4422** | **4059** | | **I² = 0.47, Tau² = 0.00** | | |
| **Fixed** | | | | | **0.08 (0.04, 0.12)** | | | | | | **0.09 (0.04, 0.13)** | | |
| **Random** | | | | | **0.07 (0.02, 0.12)** | | | | | | **0.06 (0.00, 0.13)** | | |

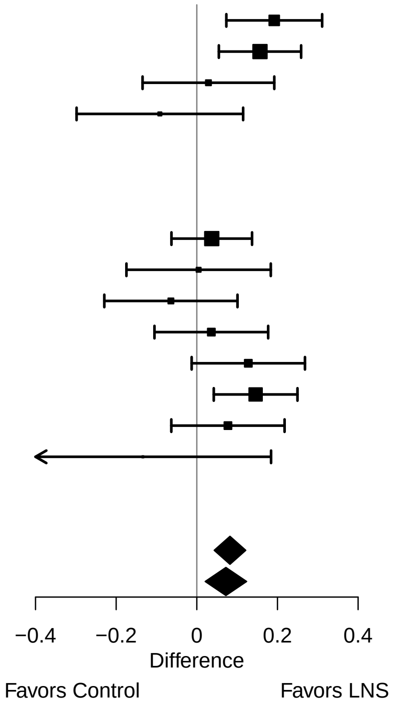

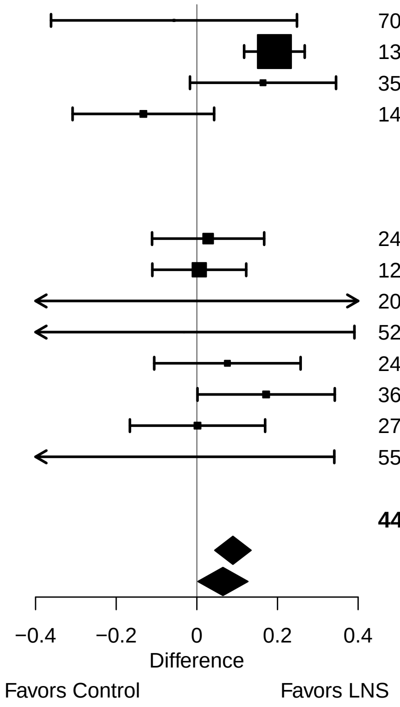

#### Supplemental figure 7D: Mean difference in WLZ

#### 7D4: Stratified by Household sanitation

**P−for−interaction = 0.062**
**Difference in MDs = −0.05 (−0.11, 0.00)**

| | | Improved | | | | | | Unimproved | | | | | |
|---|---|---|---|---|---|---|---|---|---|---|---|---|---|
| **Country** | **Trial** | **LNS N** | **Control N** | **Control Mean** | **MD (95% CI)** | **Fixed W** | **Random W** | **LNS N** | **Control N** | **Control Mean** | **MD (95% CI)** | **Fixed W** | **Random W** |
| Bangladesh | JiVitA−4 (34) | 2151 | 932 | −1.15 | 0.10 (0.05, 0.15) | 0.41 | 0.24 | 631 | 285 | −1.23 | 0.01 (−0.08, 0.09) | 0.15 | 0.13 |
| Bangladesh | RDNS (35) | 1179 | 573 | −1.10 | 0.09 (0.00, 0.18) | 0.13 | 0.14 | 480 | 241 | −1.15 | 0.03 (−0.11, 0.16) | 0.07 | 0.08 |
| Bangladesh | WASH−B (36) | 506 | 998 | −0.85 | 0.15 (0.03, 0.27) | 0.08 | 0.10 | 30 | 54 | −0.98 | 0.31 (−0.08, 0.71) | 0.01 | 0.02 |
| Burkina Faso | iLiNS−Zinc (37) | 43 | 16 | −1.00 | 0.25 (−0.06, 0.56) | 0.01 | 0.02 | 1901 | 644 | −0.94 | 0.18 (0.11, 0.26) | 0.24 | 0.15 |
| Burkina Faso | PROMIS (38) | 392 | 548 | −0.74 | 0.12 (−0.01, 0.24) | 0.07 | 0.09 | 464 | 355 | −0.67 | 0.07 (−0.11, 0.24) | 0.04 | 0.06 |
| Burkina Faso | PROMIS CS (38) | 176 | 165 | −0.90 | −0.09 (−0.32, 0.13) | 0.02 | 0.03 | 252 | 271 | −0.87 | −0.12 (−0.28, 0.04) | 0.05 | 0.07 |
| Ghana | GHANA (39) | | | | | | | | | | | | |
| Ghana | iLiNS−DYADG (40) | 336 | 671 | −0.58 | 0.09 (−0.05, 0.22) | 0.06 | 0.08 | 10 | 19 | −0.72 | 0.33 (−0.57, 1.23) | 0.00 | 0.00 |
| Haiti | HAITI (41) | | | | | | | | | | | | |
| Kenya | WASH−B (42) | 100 | 364 | 0.18 | 0.12 (−0.05, 0.30) | 0.04 | 0.05 | 594 | 1874 | 0.08 | 0.02 (−0.07, 0.11) | 0.17 | 0.13 |
| Madagascar | MAHAY (43) | | | | | | | | | | | | |
| Malawi | iLiNS−DYADM (44) | 20 | 42 | −0.16 | 0.22 (−0.20, 0.65) | 0.01 | 0.01 | 200 | 400 | −0.13 | −0.08 (−0.25, 0.09) | 0.04 | 0.06 |
| Malawi | iLiNS−DOSE (45) | 13 | 9 | −0.42 | −0.45 (−1.07, 0.16) | 0.00 | 0.00 | 593 | 201 | −0.24 | 0.04 (−0.09, 0.18) | 0.07 | 0.08 |
| Mali | PROMIS (46) | 380 | 363 | −0.32 | 0.08 (−0.06, 0.21) | 0.06 | 0.08 | 112 | 134 | −0.38 | 0.18 (−0.02, 0.38) | 0.03 | 0.05 |
| Mali | PROMIS CS (46) | 696 | 692 | −0.78 | 0.18 (0.07, 0.30) | 0.08 | 0.10 | 217 | 242 | −0.73 | 0.11 (−0.05, 0.26) | 0.05 | 0.07 |
| Zimbabwe | SHINE (HIV−) (47) | 293 | 243 | 0.05 | 0.10 (−0.09, 0.29) | 0.03 | 0.04 | 495 | 543 | 0.04 | 0.02 (−0.11, 0.15) | 0.07 | 0.08 |
| Zimbabwe | SHINE (HIV+) (48) | 50 | 35 | 0.10 | −0.43 (−0.95, 0.09) | 0.00 | 0.01 | 90 | 107 | −0.05 | 0.02 (−0.28, 0.32) | 0.01 | 0.02 |
| | | **6335** | **5651** | | **I² = 0.08, Tau² = 0.00** | | | **6069** | **5370** | | **I² = 0.50, Tau² = 0.00** | | |
| **Fixed** | | | | | **0.11 (0.07, 0.14)** | | | | | | **0.06 (0.03, 0.10)** | | |
| **Random** | | | | | **0.10 (0.06, 0.15)** | | | | | | **0.05 (0.00, 0.10)** | | |

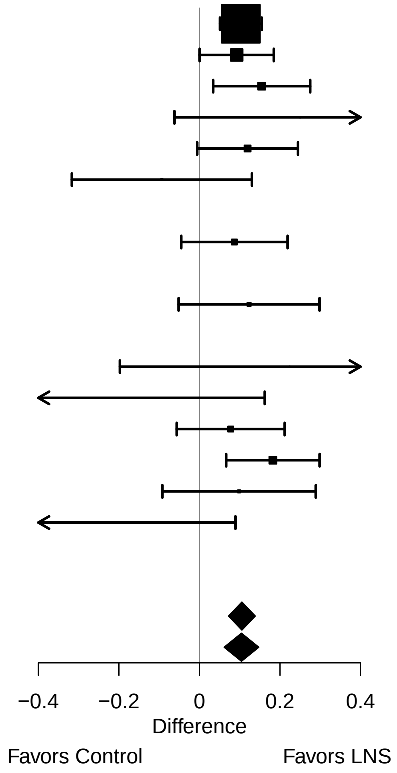

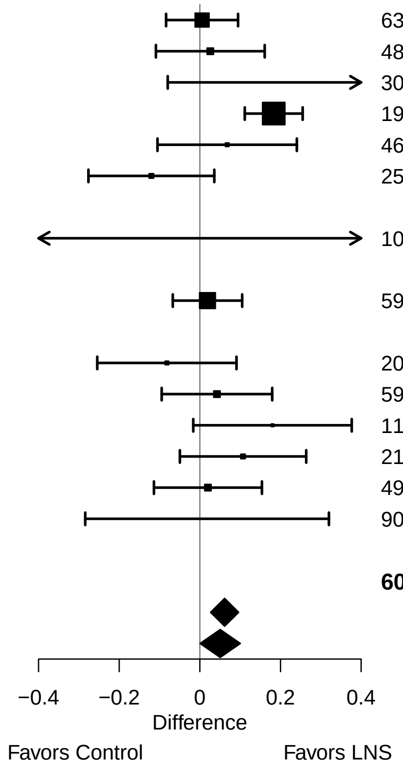

#### Supplemental figure 7D: Mean difference in WLZ

### 7D5: Stratified by Home environment

**P−for−interaction = 0.043**

**Difference in MDs = 0.06 (0.00, 0.11)**

**At least median**

| Country | Trial | LNS N | Control N | Control Mean | MD (95% CI) | Fixed W | Random W |
|---|---|---|---|---|---|---|---|
| Bangladesh | JiVitA−4 (34) | | | | | | |
| Bangladesh | RDNS (35) | 1192 | 582 | −1.08 | 0.07 (−0.02, 0.17) | 0.15 | 0.15 |
| Bangladesh | WASH−B (36) | 636 | 1652 | −0.79 | 0.13 (0.03, 0.23) | 0.15 | 0.15 |
| Burkina Faso | iLiNS−Zinc (37) | 473 | 199 | −0.91 | 0.20 (0.09, 0.30) | 0.13 | 0.14 |
| Burkina Faso | PROMIS (38) | | | | | | |
| Burkina Faso | PROMIS CS (38) | | | | | | |
| Ghana | GHANA (39) | | | | | | |
| Ghana | iLiNS−DYADG (40) | 213 | 409 | −0.56 | 0.06 (−0.10, 0.23) | 0.05 | 0.08 |
| Haiti | HAITI (41) | | | | | | |
| Kenya | WASH−B (42) | 834 | 2757 | 0.13 | −0.02 (−0.09, 0.05) | 0.30 | 0.19 |
| Madagascar | MAHAY (43) | 914 | 907 | −0.36 | 0.00 (−0.11, 0.11) | 0.12 | 0.14 |
| Malawi | iLiNS−DYADM (44) | 137 | 289 | −0.08 | −0.07 (−0.27, 0.13) | 0.03 | 0.06 |
| Malawi | iLiNS−DOSE (45) | 375 | 136 | −0.27 | 0.10 (−0.06, 0.25) | 0.06 | 0.09 |
| Mali | PROMIS (46) | | | | | | |
| Mali | PROMIS CS (46) | | | | | | |
| Zimbabwe | SHINE (HIV−) (47) | | | | | | |
| Zimbabwe | SHINE (HIV+) (48) | | | | | | |
| | | **4774** | **6931** | | **I² = 0.59, Tau² = 0.00** | | |
| **Fixed** | | | | | **0.06 (0.02, 0.09)** | | |
| **Random** | | | | | **0.06 (0.01, 0.12)** | | |

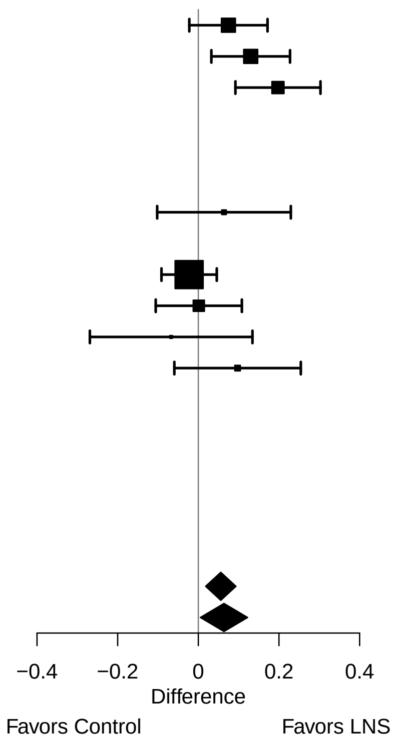

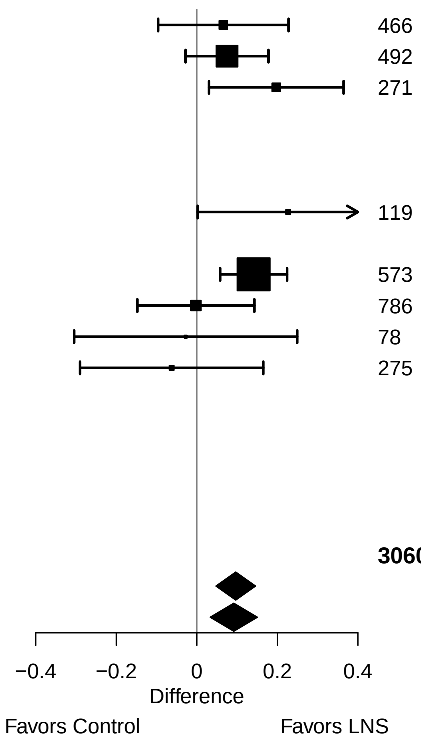

**Less than median**

| Country | Trial | LNS N | Control N | Control Mean | MD (95% CI) | Fixed W | Random W |
|---|---|---|---|---|---|---|---|
| Bangladesh | JiVitA−4 (34) | | | | | | |
| Bangladesh | RDNS (35) | 466 | 230 | −1.17 | 0.07 (−0.10, 0.23) | 0.09 | 0.11 |
| Bangladesh | WASH−B (36) | 492 | 1684 | −0.95 | 0.07 (−0.03, 0.18) | 0.23 | 0.22 |
| Burkina Faso | iLiNS−Zinc (37) | 271 | 174 | −1.08 | 0.20 (0.03, 0.36) | 0.09 | 0.10 |
| Burkina Faso | PROMIS (38) | | | | | | |
| Burkina Faso | PROMIS CS (38) | | | | | | |
| Ghana | GHANA (39) | | | | | | |
| Ghana | iLiNS−DYADG (40) | 119 | 248 | −0.67 | 0.23 (0.00, 0.45) | 0.05 | 0.06 |
| Haiti | HAITI (41) | | | | | | |
| Kenya | WASH−B (42) | 573 | 2149 | 0.07 | 0.14 (0.06, 0.22) | 0.35 | 0.28 |
| Madagascar | MAHAY (43) | 786 | 775 | −0.49 | 0.00 (−0.15, 0.14) | 0.11 | 0.13 |
| Malawi | iLiNS−DYADM (44) | 78 | 149 | −0.25 | −0.03 (−0.30, 0.25) | 0.03 | 0.04 |
| Malawi | iLiNS−DOSE (45) | 275 | 86 | −0.23 | −0.06 (−0.29, 0.16) | 0.05 | 0.06 |
| Mali | PROMIS (46) | | | | | | |
| Mali | PROMIS CS (46) | | | | | | |
| Zimbabwe | SHINE (HIV−) (47) | | | | | | |
| Zimbabwe | SHINE (HIV+) (48) | | | | | | |
| | | **3060** | **5495** | | **I² = 0.18, Tau² = 0.00** | | |
| **Fixed** | | | | | **0.10 (0.05, 0.15)** | | |
| **Random** | | | | | **0.09 (0.03, 0.15)** | | |

#### Supplemental figure 7D: Mean difference in WLZ

#### 7D6: Stratified by Season at the time of assessment

**P−for−interaction = 0.197**
**Difference in MDs = −0.03 (−0.08, 0.02)**

| Country | Trial | Dry LNS N | Dry Control N | Dry Control Mean | Dry MD (95% CI) | Dry Fixed W | Dry Random W | Rainy LNS N | Rainy Control N | Rainy Control Mean | Rainy MD (95% CI) | Rainy Fixed W | Rainy Random W |
|---|---|---|---|---|---|---|---|---|---|---|---|---|---|
| Bangladesh | JiVitA−4 (34) | 1520 | 649 | −1.09 | 0.10 (0.03, 0.16) | 0.27 | 0.11 | 1263 | 568 | −1.26 | 0.06 (−0.01, 0.13) | 0.25 | 0.17 |
| Bangladesh | RDNS (35) | 738 | 359 | −1.05 | 0.11 (−0.02, 0.23) | 0.06 | 0.09 | 923 | 456 | −1.16 | 0.04 (−0.05, 0.14) | 0.12 | 0.13 |
| Bangladesh | WASH−B (36) | 639 | 1912 | −0.75 | 0.13 (0.03, 0.23) | 0.10 | 0.09 | 517 | 1512 | −1.03 | 0.12 (0.03, 0.21) | 0.14 | 0.14 |
| Burkina Faso | iLiNS−Zinc (37) | 1394 | 477 | −0.97 | 0.24 (0.14, 0.34) | 0.11 | 0.10 | 558 | 187 | −0.88 | 0.05 (−0.07, 0.17) | 0.08 | 0.10 |
| Burkina Faso | PROMIS (38) | 428 | 445 | −0.64 | 0.06 (−0.11, 0.23) | 0.04 | 0.07 | 428 | 462 | −0.78 | 0.13 (−0.02, 0.28) | 0.05 | 0.07 |
| Burkina Faso | PROMIS CS (38) | 224 | 190 | −0.80 | −0.14 (−0.36, 0.08) | 0.02 | 0.06 | 206 | 246 | −0.95 | −0.11 (−0.29, 0.07) | 0.03 | 0.06 |
| Ghana | GHANA (39) | 57 | 55 | −0.84 | 0.52 (0.12, 0.92) | 0.01 | 0.03 | 41 | 41 | −0.37 | −0.22 (−0.78, 0.35) | 0.00 | 0.01 |
| Ghana | iLiNS−DYADG (40) | 204 | 407 | −0.62 | −0.03 (−0.20, 0.13) | 0.04 | 0.07 | 143 | 285 | −0.54 | 0.27 (0.06, 0.49) | 0.02 | 0.04 |
| Haiti | HAITI (41) | | | | | | | | | | | | |
| Kenya | WASH−B (42) | 346 | 1146 | 0.03 | 0.07 (−0.06, 0.19) | 0.07 | 0.09 | 1109 | 3972 | 0.11 | 0.07 (−0.01, 0.14) | 0.23 | 0.16 |
| Madagascar | MAHAY (43) | | | | | | | | | | | | |
| Malawi | iLiNS−DYADM (44) | 156 | 316 | −0.07 | −0.03 (−0.22, 0.16) | 0.03 | 0.06 | 64 | 128 | −0.29 | −0.12 (−0.42, 0.18) | 0.01 | 0.02 |
| Malawi | iLiNS−DOSE (45) | 415 | 134 | −0.17 | 0.00 (−0.17, 0.17) | 0.03 | 0.07 | 281 | 107 | −0.30 | 0.01 (−0.17, 0.20) | 0.03 | 0.06 |
| Mali | PROMIS (46) | | | | | | | | | | | | |
| Mali | PROMIS CS (46) | | | | | | | | | | | | |
| Zimbabwe | SHINE (HIV−) (47) | 1736 | 1655 | 0.00 | 0.10 (0.02, 0.17) | 0.19 | 0.10 | 133 | 129 | 0.24 | −0.19 (−0.47, 0.10) | 0.01 | 0.03 |
| Zimbabwe | SHINE (HIV+) (48) | 318 | 304 | 0.03 | −0.16 (−0.34, 0.03) | 0.03 | 0.07 | 18 | 24 | −0.23 | 0.02 (−0.59, 0.63) | 0.00 | 0.01 |
| | | **8175** | **8049** | | **I² = 0.60, Tau² = 0.01** | | | **5684** | **8117** | | **I² = 0.22, Tau² = 0.00** | | |
| **Fixed** | | | | | **0.09 (0.06, 0.12)** | | | | | | **0.06 (0.03, 0.09)** | | |
| **Random** | | | | | **0.07 (−0.01, 0.14)** | | | | | | **0.05 (0.01, 0.10)** | | |

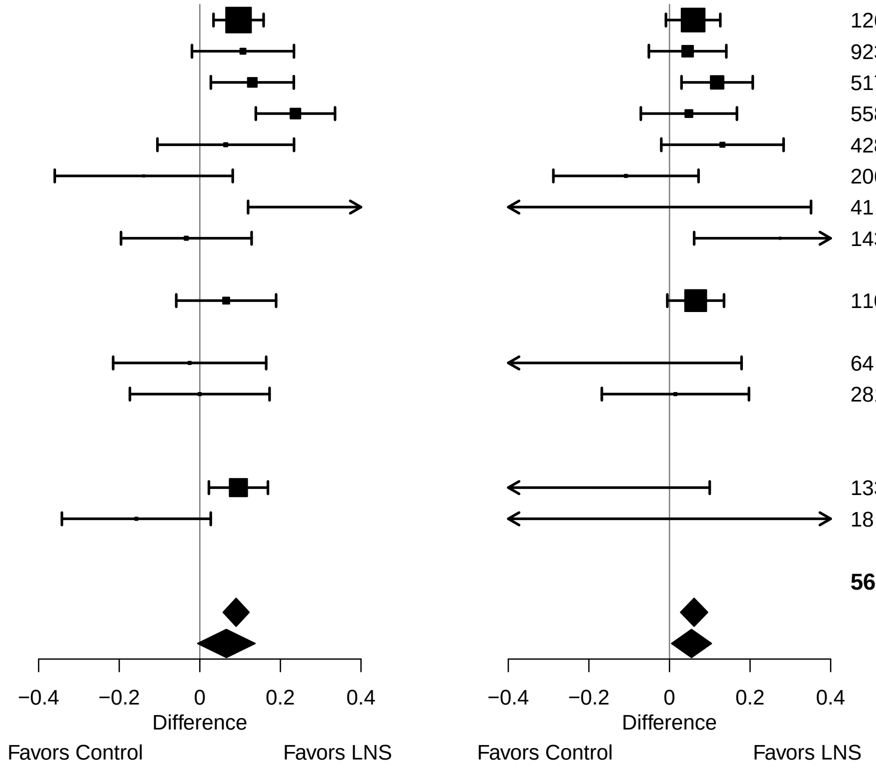

### Supplemental figure 7E: Wasting prevalence ratio

#### 7E1: Stratified by Household socio-economic status

**P−for−interaction = 0.492**
**Ratio of PRs = 1.06 (0.90, 1.24)**

| Country | Trial | At least median: LNS N | Control N | Control Prevalence | PR (95% CI) | Fixed W | Random W | Less than median: LNS N | Control N | Control Prevalence | PR (95% CI) | Fixed W | Random W |
|---|---|---|---|---|---|---|---|---|---|---|---|---|---|
| Bangladesh | JiVitA−4 (34) | 1401 | 607 | 13.6 | 0.84 (0.67, 1.06) | 0.29 | 0.29 | 1381 | 610 | 19.2 | 0.95 (0.78, 1.15) | 0.28 | 0.28 |
| Bangladesh | RDNS (35) | 834 | 407 | 13.3 | 0.88 (0.63, 1.22) | 0.13 | 0.13 | 827 | 408 | 16.9 | 0.86 (0.65, 1.13) | 0.14 | 0.14 |
| Bangladesh | WASH−B (36) | 579 | 1714 | 8.7 | 0.75 (0.53, 1.08) | 0.12 | 0.12 | 577 | 1710 | 13.7 | 0.81 (0.63, 1.04) | 0.17 | 0.17 |
| Burkina Faso | iLiNS−Zinc (37) | 1225 | 299 | 12.4 | 0.58 (0.40, 0.84) | 0.11 | 0.11 | 720 | 364 | 14.2 | 0.86 (0.62, 1.17) | 0.11 | 0.11 |
| Burkina Faso | PROMIS (38) | 405 | 479 | 7.7 | 0.86 (0.47, 1.58) | 0.04 | 0.04 | 451 | 428 | 10.3 | 0.84 (0.51, 1.38) | 0.04 | 0.04 |
| Burkina Faso | PROMIS CS (38) | 220 | 208 | 13.0 | 0.77 (0.45, 1.31) | 0.05 | 0.05 | 210 | 228 | 11.8 | 1.33 (0.83, 2.11) | 0.05 | 0.05 |
| Ghana | GHANA (39) | | | | | | | | | | | | |
| Ghana | iLiNS−DYADG (40) | 157 | 356 | 6.7 | 0.94 (0.46, 1.93) | 0.03 | 0.03 | 190 | 334 | 8.4 | 0.75 (0.39, 1.45) | 0.03 | 0.03 |
| Haiti | HAITI (41) | | | | | | | | | | | | |
| Kenya | WASH−B (42) | 825 | 2914 | 1.2 | 1.01 (0.52, 1.96) | 0.03 | 0.03 | 628 | 2200 | 1.9 | 0.77 (0.40, 1.49) | 0.02 | 0.02 |
| Madagascar | MAHAY (43) | 771 | 884 | 4.1 | 1.05 (0.60, 1.84) | 0.05 | 0.05 | 876 | 755 | 7.9 | 0.85 (0.54, 1.34) | 0.05 | 0.05 |
| Malawi | iLiNS−DYADM (44) | | | | | | | | | | | | |
| Malawi | iLiNS−DOSE (45) | | | | | | | | | | | | |
| Mali | PROMIS (46) | | | | | | | | | | | | |
| Mali | PROMIS CS (46) | 501 | 458 | 10.5 | 0.84 (0.56, 1.25) | 0.09 | 0.09 | 443 | 501 | 10.2 | 0.82 (0.57, 1.18) | 0.08 | 0.08 |
| Zimbabwe | SHINE (HIV−) (47) | 903 | 797 | 2.4 | 1.07 (0.62, 1.85) | 0.05 | 0.05 | 826 | 856 | 2.9 | 0.66 (0.34, 1.30) | 0.02 | 0.02 |
| Zimbabwe | SHINE (HIV+) (48) | | | | | | | | | | | | |
| | | **7821** | **9123** | | **I² = 0.00, Tau² = 0.00** | | | **7129** | **8394** | | **I² = 0.00, Tau² = 0.00** | | |
| **Fixed** | | | | | **0.83 (0.73, 0.93)** | | | | | | **0.88 (0.79, 0.97)** | | |
| **Random** | | | | | **0.83 (0.73, 0.93)** | | | | | | **0.88 (0.79, 0.97)** | | |

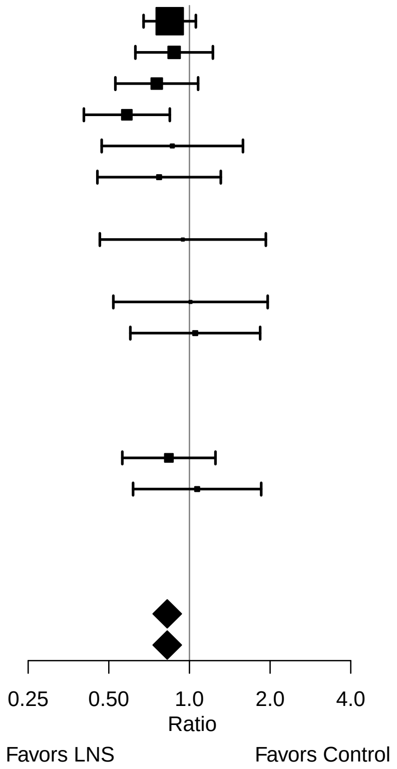

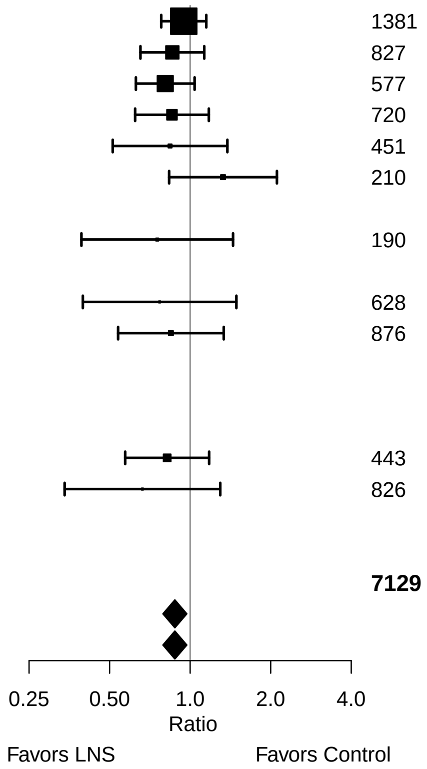

#### Supplemental figure 7E: Wasting prevalence ratio

#### 7E2: Stratified by Household food insecurity

**P−for−interaction = 0.729**
**Ratio of PRs = 0.97 (0.81, 1.16)**

| Country | Trial | Mild to secure: LNS N | Control N | Control Prevalence | PR (95% CI) | Fixed W | Random W | Moderate to severe: LNS N | Control N | Control Prevalence | PR (95% CI) | Fixed W | Random W |
|---|---|---|---|---|---|---|---|---|---|---|---|---|---|
| Bangladesh | JiVitA−4 (34) | 1977 | 841 | 16.2 | 0.89 (0.74, 1.07) | 0.34 | 0.34 | 805 | 376 | 16.8 | 0.96 (0.73, 1.27) | 0.26 | 0.24 |
| Bangladesh | RDNS (35) | 1057 | 493 | 13.8 | 0.90 (0.68, 1.18) | 0.15 | 0.15 | 604 | 322 | 17.1 | 0.83 (0.62, 1.13) | 0.21 | 0.21 |
| Bangladesh | WASH−B (36) | 917 | 2647 | 10.2 | 0.79 (0.63, 0.98) | 0.23 | 0.23 | 239 | 777 | 14.5 | 0.81 (0.54, 1.19) | 0.13 | 0.14 |
| Burkina Faso | iLiNS−Zinc (37) | 1013 | 320 | 10.9 | 0.75 (0.50, 1.13) | 0.07 | 0.07 | 932 | 343 | 15.7 | 0.63 (0.48, 0.82) | 0.27 | 0.25 |
| Burkina Faso | PROMIS (38) | 501 | 548 | 8.4 | 0.97 (0.64, 1.48) | 0.06 | 0.06 | 355 | 359 | 9.7 | 0.72 (0.39, 1.32) | 0.05 | 0.07 |
| Burkina Faso | PROMIS CS (38) | | | | | | | | | | | | |
| Ghana | GHANA (39) | | | | | | | | | | | | |
| Ghana | iLiNS−DYADG (40) | 244 | 470 | 8.1 | 0.81 (0.46, 1.42) | 0.04 | 0.04 | 102 | 219 | 6.4 | 0.92 (0.36, 2.33) | 0.02 | 0.03 |
| Haiti | HAITI (41) | | | | | | | | | | | | |
| Kenya | WASH−B (42) | | | | | | | | | | | | |
| Madagascar | MAHAY (43) | 987 | 927 | 6.8 | 0.78 (0.51, 1.17) | 0.07 | 0.07 | 354 | 410 | 4.6 | 1.40 (0.66, 2.96) | 0.04 | 0.04 |
| Malawi | iLiNS−DYADM (44) | | | | | | | | | | | | |
| Malawi | iLiNS−DOSE (45) | | | | | | | | | | | | |
| Mali | PROMIS (46) | | | | | | | | | | | | |
| Mali | PROMIS CS (46) | | | | | | | | | | | | |
| Zimbabwe | SHINE (HIV−) (47) | 1384 | 1297 | 2.3 | 1.03 (0.62, 1.71) | 0.04 | 0.04 | 294 | 324 | 4.0 | 0.42 (0.15, 1.19) | 0.02 | 0.02 |
| Zimbabwe | SHINE (HIV+) (48) | | | | | | | | | | | | |
| | | **8080** | **7543** | | **I² = 0.00, Tau² = 0.00** | | | **3685** | **3130** | | **I² = 0.19, Tau² = 0.01** | | |
| **Fixed** | | | | | **0.86 (0.77, 0.95)** | | | | | | **0.80 (0.69, 0.92)** | | |
| **Random** | | | | | **0.86 (0.77, 0.95)** | | | | | | **0.80 (0.68, 0.94)** | | |

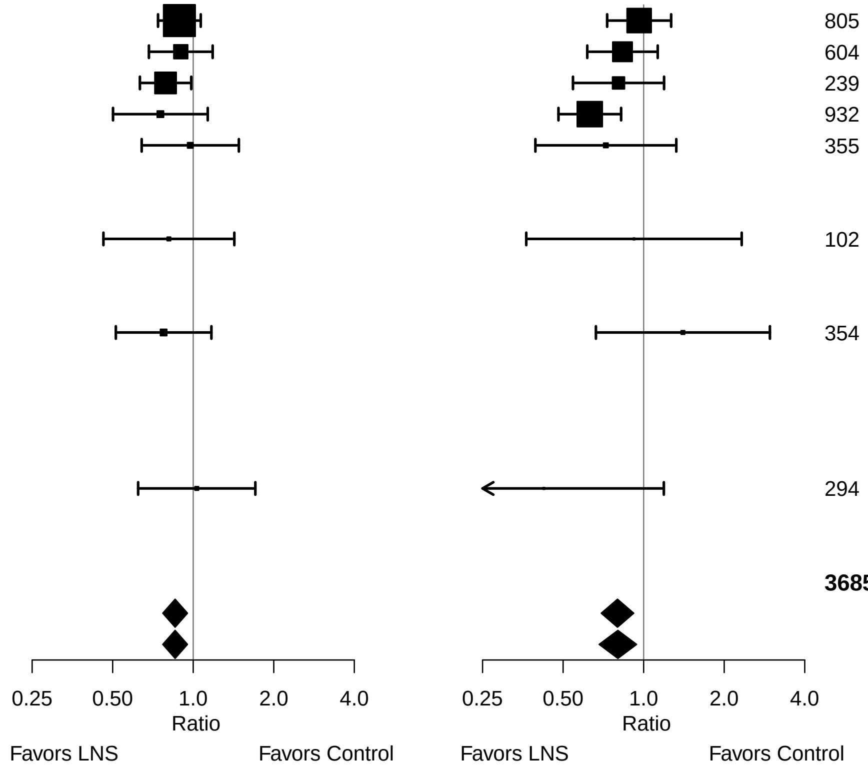

#### Supplemental figure 7E: Wasting prevalence ratio

##### 7E3: Stratified by Household source water quality

**P−for−interaction = 0.075**
**Ratio of PRs = 1.27 (0.98, 1.65)**

| Country | Trial | Improved LNS N | Improved Control N | Improved Control Prevalence | Improved PR (95% CI) | Improved Fixed W | Improved Random W | Unimproved LNS N | Unimproved Control N | Unimproved Control Prevalence | Unimproved PR (95% CI) | Unimproved Fixed W | Unimproved Random W |
|---|---|---|---|---|---|---|---|---|---|---|---|---|---|
| Bangladesh | JiVitA−4 (34) | | | | | | | | | | | | |
| Bangladesh | RDNS (35) | | | | | | | | | | | | |
| Bangladesh | WASH−B (36) | 495 | 986 | 10.6 | 0.76 (0.55, 1.04) | 0.27 | 0.27 | 70 | 114 | 10.5 | 1.36 (0.58, 3.19) | 0.03 | 0.09 |
| Burkina Faso | iLiNS−Zinc (37) | 550 | 141 | 12.8 | 0.52 (0.31, 0.89) | 0.10 | 0.10 | 1394 | 519 | 13.7 | 0.72 (0.58, 0.90) | 0.53 | 0.24 |
| Burkina Faso | PROMIS (38) | 352 | 377 | 9.3 | 0.73 (0.48, 1.12) | 0.15 | 0.15 | 357 | 350 | 9.1 | 0.83 (0.44, 1.54) | 0.07 | 0.13 |
| Burkina Faso | PROMIS CS (38) | 280 | 267 | 16.1 | 0.69 (0.43, 1.09) | 0.13 | 0.13 | 149 | 169 | 6.5 | 2.47 (1.25, 4.88) | 0.06 | 0.11 |
| Ghana | GHANA (39) | | | | | | | | | | | | |
| Ghana | iLiNS−DYADG (40) | | | | | | | | | | | | |
| Haiti | HAITI (41) | | | | | | | | | | | | |
| Kenya | WASH−B (42) | | | | | | | | | | | | |
| Madagascar | MAHAY (43) | 418 | 453 | 5.5 | 1.13 (0.60, 2.11) | 0.07 | 0.07 | 1202 | 1180 | 6.0 | 0.90 (0.60, 1.35) | 0.16 | 0.18 |
| Malawi | iLiNS−DYADM (44) | | | | | | | | | | | | |
| Malawi | iLiNS−DOSE (45) | | | | | | | | | | | | |
| Mali | PROMIS (46) | | | | | | | | | | | | |
| Mali | PROMIS CS (46) | 544 | 562 | 9.8 | 0.77 (0.54, 1.10) | 0.22 | 0.22 | 361 | 382 | 10.7 | 0.93 (0.60, 1.44) | 0.13 | 0.17 |
| Zimbabwe | SHINE (HIV−) (47) | 518 | 486 | 2.9 | 1.01 (0.50, 2.00) | 0.06 | 0.06 | 273 | 301 | 3.0 | 0.74 (0.29, 1.87) | 0.03 | 0.08 |
| Zimbabwe | SHINE (HIV+) (48) | | | | | | | | | | | | |
| | | **3157** | **3272** | | **I² = 0.00, Tau² = 0.00** | | | **3806** | **3015** | | **I² = 0.54, Tau² = 0.09** | | |
| **Fixed** | | | | | **0.75 (0.64, 0.89)** | | | | | | **0.85 (0.73, 1.00)** | | |
| **Random** | | | | | **0.75 (0.64, 0.89)** | | | | | | **0.97 (0.72, 1.31)** | | |

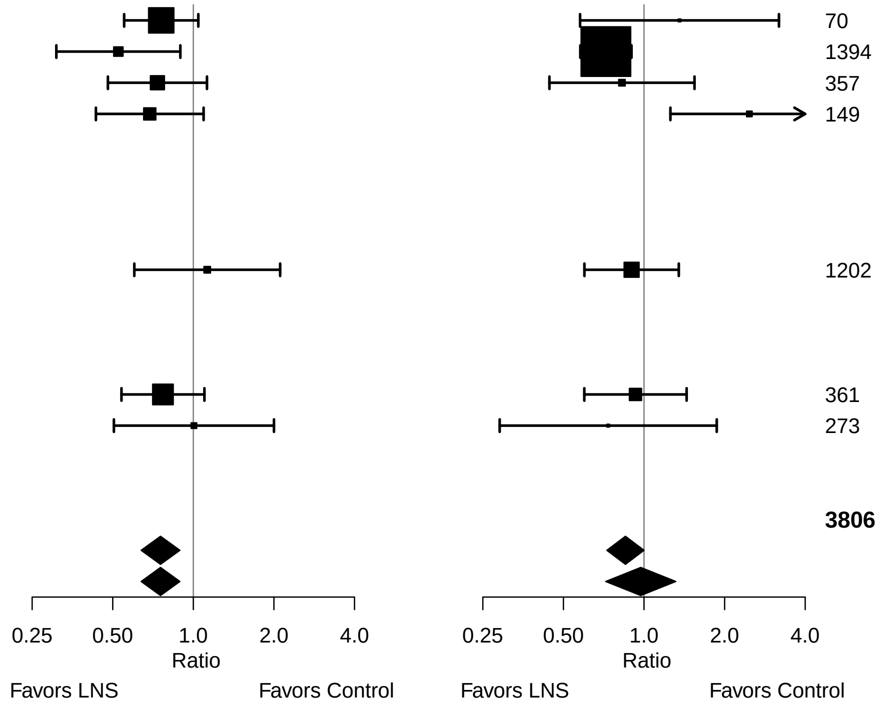

#### Supplemental figure 7E: Wasting prevalence ratio

#### 7E4: Stratified by Household sanitation

**P−for−interaction = 0.080**
**Ratio of PRs = 1.20 (0.98, 1.48)**

| Country | Trial | Improved LNS N | Improved Control N | Improved Control Prevalence | Improved PR (95% CI) | Improved Fixed W | Improved Random W | Unimproved LNS N | Unimproved Control N | Unimproved Control Prevalence | Unimproved PR (95% CI) | Unimproved Fixed W | Unimproved Random W |
|---|---|---|---|---|---|---|---|---|---|---|---|---|---|
| Bangladesh | JiVitA−4 (34) | 2151 | 932 | 16.1 | 0.87 (0.72, 1.04) | 0.48 | 0.48 | 631 | 285 | 17.4 | 1.06 (0.80, 1.39) | 0.39 | 0.27 |
| Bangladesh | RDNS (35) | 1179 | 573 | 15.2 | 0.74 (0.58, 0.96) | 0.25 | 0.25 | 480 | 241 | 14.9 | 1.17 (0.84, 1.63) | 0.27 | 0.24 |
| Bangladesh | WASH−B (36) | | | | | | | | | | | | |
| Burkina Faso | iLiNS−Zinc (37) | | | | | | | | | | | | |
| Burkina Faso | PROMIS (38) | 392 | 548 | 9.1 | 0.87 (0.52, 1.44) | 0.06 | 0.06 | 464 | 355 | 8.7 | 0.86 (0.55, 1.36) | 0.14 | 0.18 |
| Burkina Faso | PROMIS CS (38) | 176 | 165 | 13.3 | 0.77 (0.43, 1.37) | 0.05 | 0.05 | 252 | 271 | 11.8 | 1.24 (0.76, 2.04) | 0.12 | 0.16 |
| Ghana | GHANA (39) | | | | | | | | | | | | |
| Ghana | iLiNS−DYADG (40) | | | | | | | | | | | | |
| Haiti | HAITI (41) | | | | | | | | | | | | |
| Kenya | WASH−B (42) | | | | | | | | | | | | |
| Madagascar | MAHAY (43) | | | | | | | | | | | | |
| Malawi | iLiNS−DYADM (44) | | | | | | | | | | | | |
| Malawi | iLiNS−DOSE (45) | | | | | | | | | | | | |
| Mali | PROMIS (46) | | | | | | | | | | | | |
| Mali | PROMIS CS (46) | 696 | 692 | 11.0 | 0.82 (0.61, 1.12) | 0.17 | 0.17 | 217 | 242 | 8.7 | 0.53 (0.30, 0.93) | 0.09 | 0.14 |
| Zimbabwe | SHINE (HIV−) (47) | | | | | | | | | | | | |
| Zimbabwe | SHINE (HIV+) (48) | | | | | | | | | | | | |
| | | **4594** | **2910** | | **I² = 0.00, Tau² = 0.00** | | | **2044** | **1394** | | **I² = 0.43, Tau² = 0.05** | | |
| **Fixed** | | | | | **0.82 (0.73, 0.93)** | | | | | | **1.01 (0.85, 1.20)** | | |
| **Random** | | | | | **0.82 (0.73, 0.93)** | | | | | | **0.97 (0.74, 1.27)** | | |

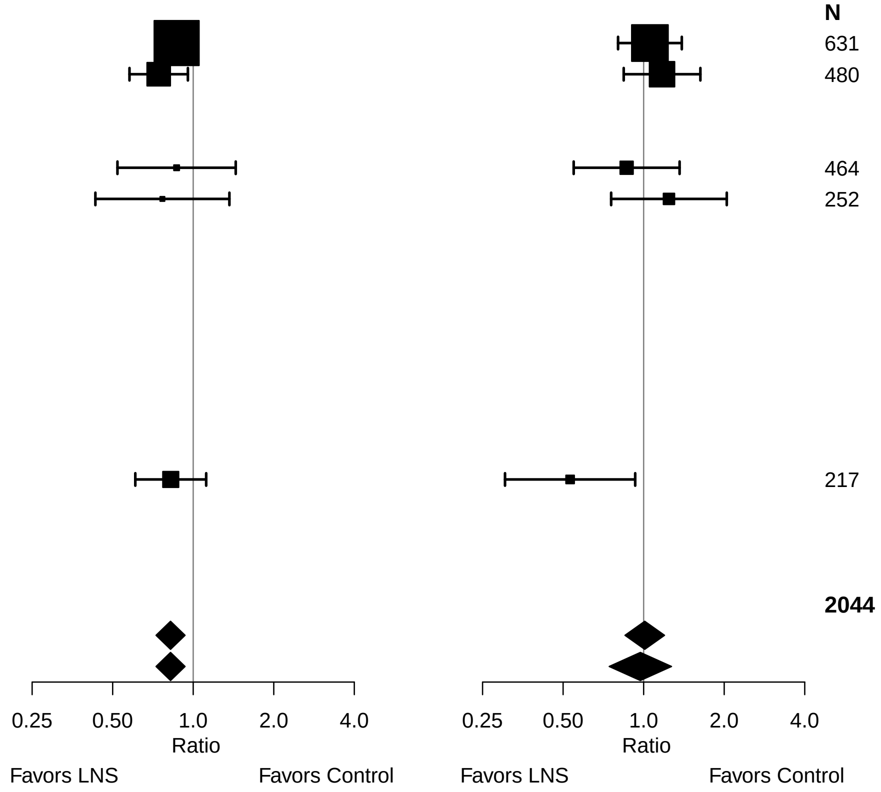

#### Supplemental figure 7E: Wasting prevalence ratio

#### 7E5: Stratified by Home environment

**P−for−interaction = 0.478**
**Ratio of PRs = 1.09 (0.86, 1.37)**

| Country | Trial | LNS N | Control N | Control Prevalence | At least median PR (95% CI) | Fixed W | Random W | LNS N | Control N | Control Prevalence | Less than median PR (95% CI) | Fixed W | Random W |
|---|---|---|---|---|---|---|---|---|---|---|---|---|---|
| Bangladesh | JiVitA−4 (34) | | | | | | | | | | | | |
| Bangladesh | RDNS (35) | 1192 | 582 | 14.6 | 0.80 (0.61, 1.05) | 0.33 | 0.26 | 466 | 230 | 15.7 | 1.06 (0.77, 1.46) | 0.28 | 0.28 |
| Bangladesh | WASH−B (36) | 636 | 1652 | 10.4 | 0.68 (0.50, 0.92) | 0.27 | 0.24 | 492 | 1684 | 11.9 | 0.94 (0.71, 1.25) | 0.35 | 0.35 |
| Burkina Faso | iLiNS−Zinc (37) | 473 | 199 | 11.6 | 0.61 (0.42, 0.87) | 0.18 | 0.19 | 271 | 174 | 17.7 | 0.72 (0.45, 1.16) | 0.12 | 0.12 |
| Burkina Faso | PROMIS (38) | | | | | | | | | | | | |
| Burkina Faso | PROMIS CS (38) | | | | | | | | | | | | |
| Ghana | GHANA (39) | | | | | | | | | | | | |
| Ghana | iLiNS−DYADG (40) | 213 | 409 | 6.4 | 1.03 (0.55, 1.94) | 0.06 | 0.09 | 119 | 248 | 9.7 | 0.43 (0.17, 1.11) | 0.03 | 0.03 |
| Haiti | HAITI (41) | | | | | | | | | | | | |
| Kenya | WASH−B (42) | 834 | 2757 | 1.1 | 1.21 (0.61, 2.41) | 0.05 | 0.08 | 573 | 2149 | 1.5 | 0.80 (0.36, 1.74) | 0.05 | 0.05 |
| Madagascar | MAHAY (43) | 914 | 907 | 3.7 | 1.14 (0.71, 1.83) | 0.11 | 0.14 | 786 | 775 | 8.3 | 0.86 (0.57, 1.30) | 0.17 | 0.17 |
| Malawi | iLiNS−DYADM (44) | | | | | | | | | | | | |
| Malawi | iLiNS−DOSE (45) | | | | | | | | | | | | |
| Mali | PROMIS (46) | | | | | | | | | | | | |
| Mali | PROMIS CS (46) | | | | | | | | | | | | |
| Zimbabwe | SHINE (HIV−) (47) | | | | | | | | | | | | |
| Zimbabwe | SHINE (HIV+) (48) | | | | | | | | | | | | |
| | | **4262** | **6506** | | **I² = 0.33, Tau² = 0.02** | | | **2707** | **5260** | | **I² = 0.00, Tau² = 0.00** | | |
| **Fixed** | | | | | **0.78 (0.67, 0.92)** | | | | | | **0.90 (0.76, 1.06)** | | |
| **Random** | | | | | **0.81 (0.66, 1.00)** | | | | | | **0.90 (0.76, 1.06)** | | |

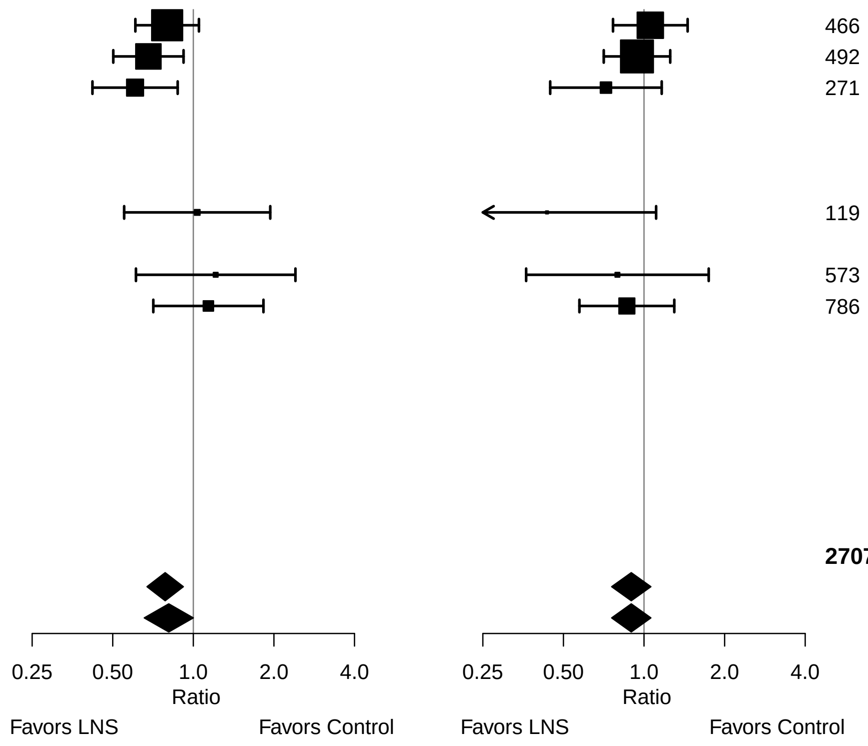

#### Supplemental figure 7E: Wasting prevalence ratio

##### 7E6: Stratified by Season at the time of assessment

**P−for−interaction = 0.088**
**Ratio of PRs = 1.16 (0.98, 1.39)**

| Country | Trial | LNS N | Control N | Control Prevalence | Dry PR (95% CI) | Fixed W | Random W | LNS N | Control N | Control Prevalence | Rainy PR (95% CI) | Fixed W | Random W |
|---|---|---|---|---|---|---|---|---|---|---|---|---|---|
| Bangladesh | JiVitA−4 (34) | 1520 | 649 | 13.7 | 0.87 (0.69, 1.09) | 0.29 | 0.28 | 1263 | 568 | 19.4 | 0.95 (0.77, 1.17) | 0.33 | 0.33 |
| Bangladesh | RDNS (35) | 738 | 359 | 13.6 | 0.81 (0.59, 1.12) | 0.15 | 0.15 | 923 | 456 | 16.2 | 0.90 (0.68, 1.19) | 0.19 | 0.19 |
| Bangladesh | WASH−B (36) | 639 | 1912 | 8.9 | 0.77 (0.57, 1.05) | 0.17 | 0.17 | 517 | 1512 | 14.2 | 0.79 (0.61, 1.02) | 0.22 | 0.22 |
| Burkina Faso | iLiNS−Zinc (37) | 1394 | 477 | 13.8 | 0.60 (0.47, 0.78) | 0.24 | 0.24 | 558 | 187 | 12.8 | 0.95 (0.62, 1.44) | 0.08 | 0.08 |
| Burkina Faso | PROMIS (38) | 428 | 445 | 8.5 | 0.85 (0.51, 1.41) | 0.06 | 0.06 | 428 | 462 | 9.3 | 0.88 (0.54, 1.42) | 0.06 | 0.06 |
| Burkina Faso | PROMIS CS (38) | 224 | 190 | 13.2 | 0.81 (0.45, 1.46) | 0.05 | 0.05 | 206 | 246 | 11.8 | 1.28 (0.80, 2.04) | 0.06 | 0.06 |
| Ghana | GHANA (39) | | | | | | | | | | | | |
| Ghana | iLiNS−DYADG (40) | | | | | | | | | | | | |
| Haiti | HAITI (41) | | | | | | | | | | | | |
| Kenya | WASH−B (42) | 346 | 1146 | 1.9 | 0.75 (0.30, 1.88) | 0.02 | 0.02 | 1109 | 3972 | 1.4 | 0.93 (0.51, 1.68) | 0.04 | 0.04 |
| Madagascar | MAHAY (43) | | | | | | | | | | | | |
| Malawi | iLiNS−DYADM (44) | | | | | | | | | | | | |
| Malawi | iLiNS−DOSE (45) | 415 | 134 | 4.5 | 1.52 (0.68, 3.41) | 0.02 | 0.03 | 281 | 107 | 6.5 | 1.24 (0.56, 2.74) | 0.02 | 0.02 |
| Mali | PROMIS (46) | | | | | | | | | | | | |
| Mali | PROMIS CS (46) | | | | | | | | | | | | |
| Zimbabwe | SHINE (HIV−) (47) | | | | | | | | | | | | |
| Zimbabwe | SHINE (HIV+) (48) | | | | | | | | | | | | |
| | | **5704** | **5312** | | **I² = 0.08, Tau² = 0.00** | | | **5285** | **7510** | | **I² = 0.00, Tau² = 0.00** | | |
| **Fixed** | | | | | **0.78 (0.69, 0.88)** | | | | | | **0.92 (0.82, 1.04)** | | |
| **Random** | | | | | **0.78 (0.68, 0.89)** | | | | | | **0.92 (0.82, 1.04)** | | |

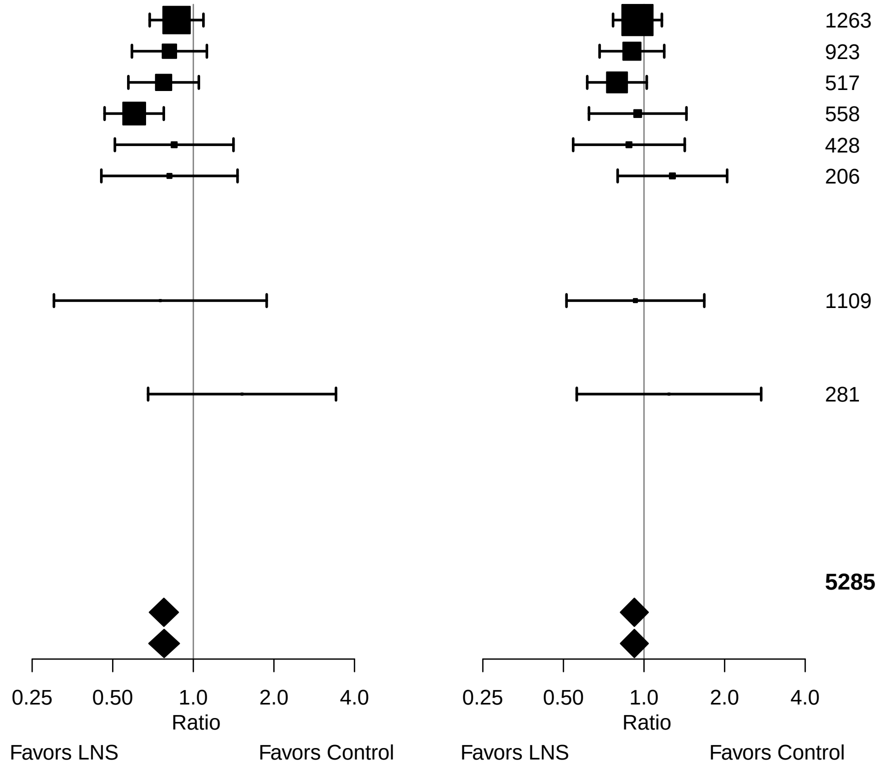

### Supplemental figure 7F: Wasting prevalence difference

#### 7F1: Stratified by Household socio-economic status

**P−for−interaction = 0.223**
**Difference in PDs = −0.01 (−0.02, 0.00)**

**At least median**

| Country | Trial | LNS N | Control N | Control Prevalence | PD (95% CI) | Fixed W | Random W |
|---|---|---|---|---|---|---|---|
| Bangladesh | JiVitA−4 (34) | 1401 | 607 | 13.6 | −0.01 (−0.04, 0.02) | 0.04 | 0.04 |
| Bangladesh | RDNS (35) | 834 | 407 | 13.3 | −0.02 (−0.06, 0.03) | 0.02 | 0.02 |
| Bangladesh | WASH−B (36) | 579 | 1714 | 8.7 | −0.02 (−0.05, 0.00) | 0.05 | 0.05 |
| Burkina Faso | iLiNS−Zinc (37) | 1225 | 299 | 12.4 | −0.05 (−0.08, −0.01) | 0.03 | 0.03 |
| Burkina Faso | PROMIS (38) | 405 | 479 | 7.7 | −0.01 (−0.05, 0.03) | 0.02 | 0.02 |
| Burkina Faso | PROMIS CS (38) | 220 | 208 | 13.0 | −0.03 (−0.09, 0.03) | 0.01 | 0.01 |
| Ghana | GHANA (39) | | | | | | |
| Ghana | iLiNS−DYADG (40) | 157 | 356 | 6.7 | 0.00 (−0.05, 0.04) | 0.02 | 0.02 |
| Haiti | HAITI (41) | | | | | | |
| Kenya | WASH−B (42) | 825 | 2914 | 1.2 | 0.00 (−0.01, 0.01) | 0.54 | 0.54 |
| Madagascar | MAHAY (43) | 771 | 884 | 4.1 | 0.00 (−0.02, 0.03) | 0.06 | 0.06 |
| Malawi | iLiNS−DYADM (44) | | | | | | |
| Malawi | iLiNS−DOSE (45) | | | | | | |
| Mali | PROMIS (46) | | | | | | |
| Mali | PROMIS CS (46) | 501 | 458 | 10.5 | −0.02 (−0.05, 0.02) | 0.02 | 0.02 |
| Zimbabwe | SHINE (HIV−) (47) | 903 | 797 | 2.4 | 0.00 (−0.01, 0.02) | 0.19 | 0.19 |
| Zimbabwe | SHINE (HIV+) (48) | | | | | | |
| | | **7821** | **9123** | | **I² = 0.14, Tau² = 0.00** | | |
| **Fixed** | | | | | **0.00 (−0.01, 0.00)** | | |
| **Random** | | | | | **0.00 (−0.01, 0.00)** | | |

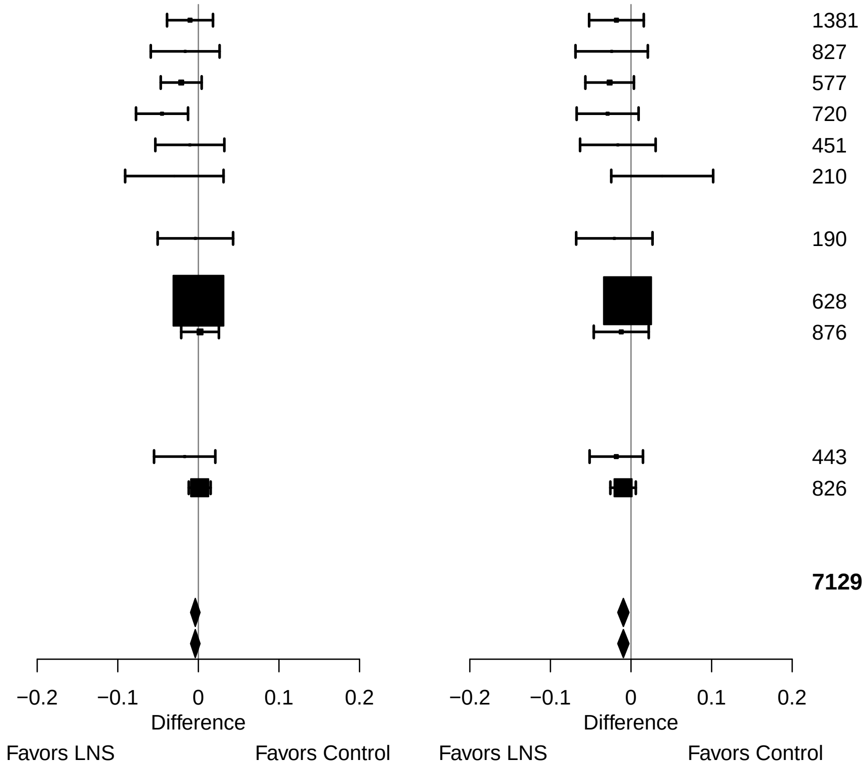

**Less than median**

| Country | Trial | LNS N | Control N | Control Prevalence | PD (95% CI) | Fixed W | Random W |
|---|---|---|---|---|---|---|---|
| Bangladesh | JiVitA−4 (34) | 1381 | 610 | 19.2 | −0.02 (−0.05, 0.02) | 0.04 | 0.04 |
| Bangladesh | RDNS (35) | 827 | 408 | 16.9 | −0.02 (−0.07, 0.02) | 0.02 | 0.02 |
| Bangladesh | WASH−B (36) | 577 | 1710 | 13.7 | −0.03 (−0.06, 0.00) | 0.05 | 0.05 |
| Burkina Faso | iLiNS−Zinc (37) | 720 | 364 | 14.2 | −0.03 (−0.07, 0.01) | 0.03 | 0.03 |
| Burkina Faso | PROMIS (38) | 451 | 428 | 10.3 | −0.02 (−0.06, 0.03) | 0.02 | 0.02 |
| Burkina Faso | PROMIS CS (38) | 210 | 228 | 11.8 | 0.04 (−0.02, 0.10) | 0.01 | 0.01 |
| Ghana | GHANA (39) | | | | | | |
| Ghana | iLiNS−DYADG (40) | 190 | 334 | 8.4 | −0.02 (−0.07, 0.03) | 0.02 | 0.02 |
| Haiti | HAITI (41) | | | | | | |
| Kenya | WASH−B (42) | 628 | 2200 | 1.9 | 0.00 (−0.01, 0.01) | 0.51 | 0.51 |
| Madagascar | MAHAY (43) | 876 | 755 | 7.9 | −0.01 (−0.05, 0.02) | 0.04 | 0.04 |
| Malawi | iLiNS−DYADM (44) | | | | | | |
| Malawi | iLiNS−DOSE (45) | | | | | | |
| Mali | PROMIS (46) | | | | | | |
| Mali | PROMIS CS (46) | 443 | 501 | 10.2 | −0.02 (−0.05, 0.01) | 0.04 | 0.04 |
| Zimbabwe | SHINE (HIV−) (47) | 826 | 856 | 2.9 | −0.01 (−0.03, 0.01) | 0.19 | 0.19 |
| Zimbabwe | SHINE (HIV+) (48) | | | | | | |
| | | **7129** | **8394** | | **I² = 0.00, Tau² = 0.00** | | |
| **Fixed** | | | | | **−0.01 (−0.02, 0.00)** | | |
| **Random** | | | | | **−0.01 (−0.02, 0.00)** | | |

#### Supplemental figure 7F: Wasting prevalence difference

#### 7F2: Stratified by Household food insecurity

**P−for−interaction = 0.433**
**Difference in PDs = −0.01 (−0.02, 0.01)**

| Country | Trial | Mild to secure: LNS N | Mild to secure: Control N | Mild to secure: Control Prevalence | Mild to secure: PD (95% CI) | Mild to secure: Fixed W | Mild to secure: Random W | Moderate to severe: LNS N | Moderate to severe: Control N | Moderate to severe: Control Prevalence | Moderate to severe: PD (95% CI) | Moderate to severe: Fixed W | Moderate to severe: Random W |
|---|---|---|---|---|---|---|---|---|---|---|---|---|---|
| Bangladesh | JiVitA−4 (34) | 1977 | 841 | 16.2 | −0.02 (−0.05, 0.01) | 0.09 | 0.09 | 805 | 376 | 16.8 | 0.00 (−0.04, 0.04) | 0.09 | 0.11 |
| Bangladesh | RDNS (35) | 1057 | 493 | 13.8 | −0.01 (−0.05, 0.02) | 0.05 | 0.05 | 604 | 322 | 17.1 | −0.03 (−0.08, 0.02) | 0.08 | 0.10 |
| Bangladesh | WASH−B (36) | 917 | 2647 | 10.2 | −0.02 (−0.04, 0.00) | 0.17 | 0.17 | 239 | 777 | 14.5 | −0.03 (−0.08, 0.02) | 0.07 | 0.10 |
| Burkina Faso | iLiNS−Zinc (37) | 1013 | 320 | 10.9 | −0.02 (−0.07, 0.02) | 0.03 | 0.03 | 932 | 343 | 15.7 | −0.06 (−0.08, −0.03) | 0.27 | 0.21 |
| Burkina Faso | PROMIS (38) | 501 | 548 | 8.4 | 0.00 (−0.04, 0.03) | 0.05 | 0.05 | 355 | 359 | 9.7 | −0.03 (−0.08, 0.02) | 0.07 | 0.09 |
| Burkina Faso | PROMIS CS (38) | | | | | | | | | | | | |
| Ghana | GHANA (39) | | | | | | | | | | | | |
| Ghana | iLiNS−DYADG (40) | 244 | 470 | 8.1 | −0.02 (−0.06, 0.03) | 0.04 | 0.04 | 102 | 219 | 6.4 | −0.01 (−0.06, 0.05) | 0.05 | 0.08 |
| Haiti | HAITI (41) | | | | | | | | | | | | |
| Kenya | WASH−B (42) | | | | | | | | | | | | |
| Madagascar | MAHAY (43) | 987 | 927 | 6.8 | −0.02 (−0.04, 0.01) | 0.10 | 0.10 | 354 | 410 | 4.6 | 0.02 (−0.02, 0.06) | 0.12 | 0.13 |
| Malawi | iLiNS−DYADM (44) | | | | | | | | | | | | |
| Malawi | iLiNS−DOSE (45) | | | | | | | | | | | | |
| Mali | PROMIS (46) | | | | | | | | | | | | |
| Mali | PROMIS CS (46) | | | | | | | | | | | | |
| Zimbabwe | SHINE (HIV−) (47) | 1384 | 1297 | 2.3 | 0.00 (−0.01, 0.01) | 0.46 | 0.46 | 294 | 324 | 4.0 | −0.02 (−0.05, 0.00) | 0.24 | 0.19 |
| Zimbabwe | SHINE (HIV+) (48) | | | | | | | | | | | | |
| | | **8080** | **7543** | | **I² = 0.00, Tau² = 0.00** | | | **3685** | **3130** | | **I² = 0.44, Tau² = 0.00** | | |
| **Fixed** | | | | | **−0.01 (−0.02, 0.00)** | | | | | | **−0.03 (−0.04, −0.01)** | | |
| **Random** | | | | | **−0.01 (−0.02, 0.00)** | | | | | | **−0.02 (−0.04, 0.00)** | | |

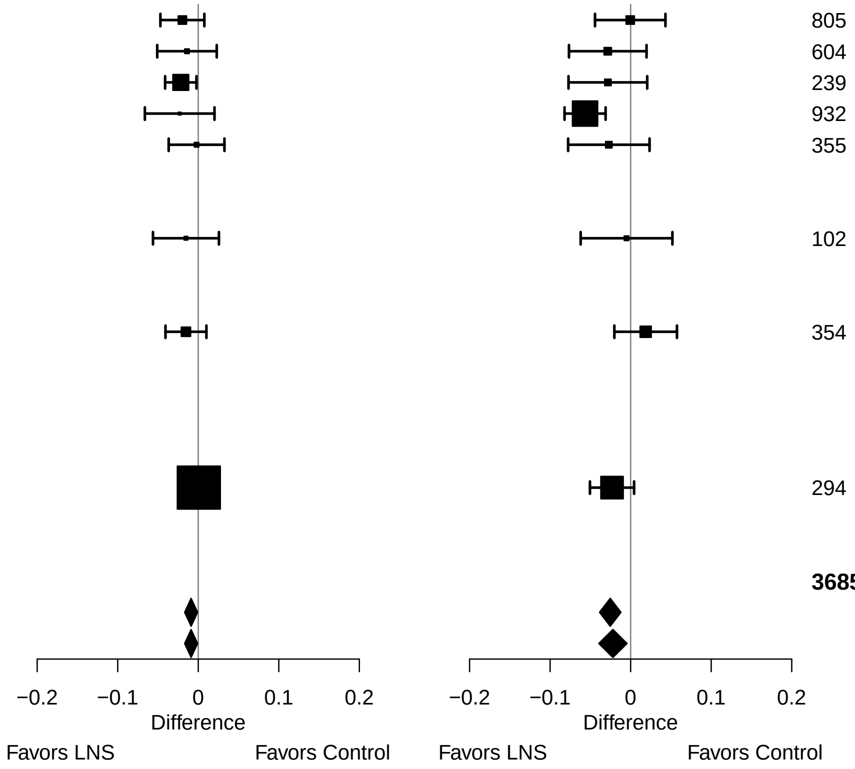

#### Supplemental figure 7F: Wasting prevalence difference

##### 7F3: Stratified by Household source water quality

**P−for−interaction = 0.612**
**Difference in PDs = 0.00 (−0.01, 0.02)**

| Country | Trial | Improved LNS N | Improved Control N | Improved Control Prevalence | Improved PD (95% CI) | Improved Fixed W | Improved Random W | Unimproved LNS N | Unimproved Control N | Unimproved Control Prevalence | Unimproved PD (95% CI) | Unimproved Fixed W | Unimproved Random W |
|---|---|---|---|---|---|---|---|---|---|---|---|---|---|
| Bangladesh | JiVitA−4 (34) | | | | | | | | | | | | |
| Bangladesh | RDNS (35) | | | | | | | | | | | | |
| Bangladesh | WASH−B (36) | 495 | 986 | 10.6 | −0.03 (−0.05, 0.00) | 0.17 | 0.17 | 70 | 114 | 10.5 | 0.04 (−0.08, 0.15) | 0.01 | 0.05 |
| Burkina Faso | iLiNS−Zinc (37) | 550 | 141 | 12.8 | −0.04 (−0.09, 0.01) | 0.05 | 0.05 | 1394 | 519 | 13.7 | −0.04 (−0.07, −0.02) | 0.24 | 0.19 |
| Burkina Faso | PROMIS (38) | 352 | 377 | 9.3 | −0.02 (−0.06, 0.01) | 0.14 | 0.14 | 357 | 350 | 9.1 | −0.02 (−0.07, 0.04) | 0.06 | 0.13 |
| Burkina Faso | PROMIS CS (38) | 280 | 267 | 16.1 | −0.05 (−0.12, 0.02) | 0.03 | 0.03 | 149 | 169 | 6.5 | 0.10 (0.03, 0.16) | 0.03 | 0.10 |
| Ghana | GHANA (39) | | | | | | | | | | | | |
| Ghana | iLiNS−DYADG (40) | | | | | | | | | | | | |
| Haiti | HAITI (41) | | | | | | | | | | | | |
| Kenya | WASH−B (42) | | | | | | | | | | | | |
| Madagascar | MAHAY (43) | 418 | 453 | 5.5 | 0.01 (−0.03, 0.04) | 0.11 | 0.11 | 1202 | 1180 | 6.0 | −0.01 (−0.03, 0.02) | 0.30 | 0.19 |
| Malawi | iLiNS−DYADM (44) | | | | | | | | | | | | |
| Malawi | iLiNS−DOSE (45) | | | | | | | | | | | | |
| Mali | PROMIS (46) | | | | | | | | | | | | |
| Mali | PROMIS CS (46) | 544 | 562 | 9.8 | −0.02 (−0.05, 0.01) | 0.15 | 0.15 | 361 | 382 | 10.7 | −0.01 (−0.05, 0.04) | 0.08 | 0.14 |
| Zimbabwe | SHINE (HIV−) (47) | 518 | 486 | 2.9 | 0.00 (−0.02, 0.02) | 0.36 | 0.36 | 273 | 301 | 3.0 | −0.01 (−0.03, 0.02) | 0.29 | 0.19 |
| Zimbabwe | SHINE (HIV+) (48) | | | | | | | | | | | | |
| | | **3157** | **3272** | | **I² = 0.10, Tau² = 0.00** | | | **3806** | **3015** | | **I² = 0.62, Tau² = 0.00** | | |
| **Fixed** | | | | | **−0.01 (−0.03, 0.00)** | | | | | | **−0.01 (−0.02, 0.00)** | | |
| **Random** | | | | | **−0.01 (−0.03, 0.00)** | | | | | | **0.00 (−0.03, 0.03)** | | |

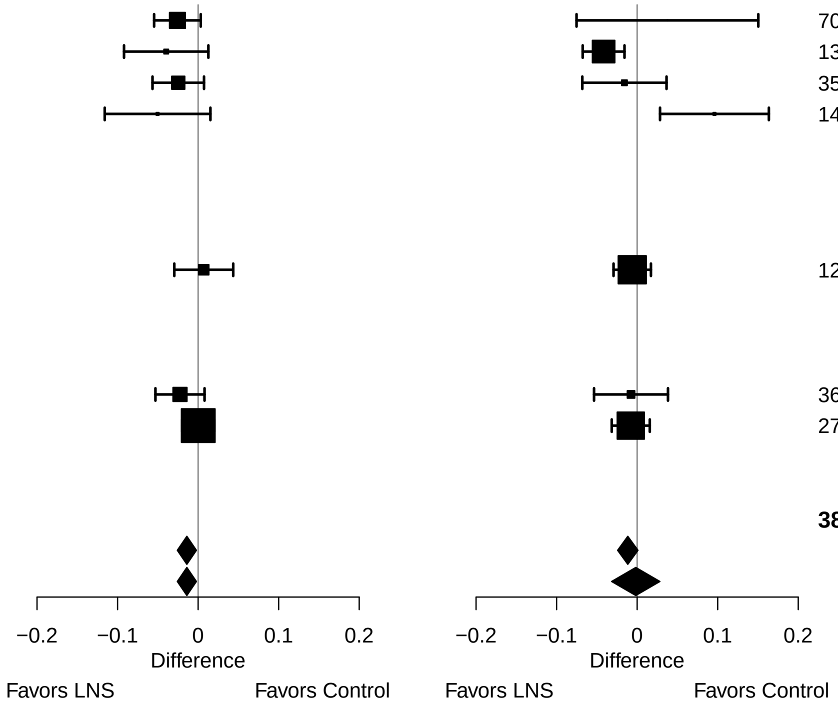

#### Supplemental figure 7F: Wasting prevalence difference

#### 7F4: Stratified by Household sanitation

**P−for−interaction = 0.134**
**Difference in PDs = 0.02 (−0.01, 0.05)**

| Country | Trial | Improved LNS N | Improved Control N | Improved Control Prevalence | Improved PD (95% CI) | Improved Fixed W | Improved Random W | Unimproved LNS N | Unimproved Control N | Unimproved Control Prevalence | Unimproved PD (95% CI) | Unimproved Fixed W | Unimproved Random W |
|---|---|---|---|---|---|---|---|---|---|---|---|---|---|
| Bangladesh | JiVitA−4 (34) | 2151 | 932 | 16.1 | −0.02 (−0.05, 0.00) | 0.37 | 0.37 | 631 | 285 | 17.4 | 0.02 (−0.03, 0.06) | 0.18 | 0.19 |
| Bangladesh | RDNS (35) | 1179 | 573 | 15.2 | −0.04 (−0.07, 0.00) | 0.19 | 0.19 | 480 | 241 | 14.9 | 0.03 (−0.03, 0.08) | 0.15 | 0.17 |
| Bangladesh | WASH−B (36) | | | | | | | | | | | | |
| Burkina Faso | iLiNS−Zinc (37) | | | | | | | | | | | | |
| Burkina Faso | PROMIS (38) | 392 | 548 | 9.1 | −0.01 (−0.05, 0.03) | 0.14 | 0.14 | 464 | 355 | 8.7 | −0.01 (−0.05, 0.03) | 0.28 | 0.25 |
| Burkina Faso | PROMIS CS (38) | 176 | 165 | 13.3 | −0.03 (−0.10, 0.04) | 0.05 | 0.05 | 252 | 271 | 11.8 | 0.03 (−0.04, 0.09) | 0.10 | 0.13 |
| Ghana | GHANA (39) | | | | | | | | | | | | |
| Ghana | iLiNS−DYADG (40) | | | | | | | | | | | | |
| Haiti | HAITI (41) | | | | | | | | | | | | |
| Kenya | WASH−B (42) | | | | | | | | | | | | |
| Madagascar | MAHAY (43) | | | | | | | | | | | | |
| Malawi | iLiNS−DYADM (44) | | | | | | | | | | | | |
| Malawi | iLiNS−DOSE (45) | | | | | | | | | | | | |
| Mali | PROMIS (46) | | | | | | | | | | | | |
| Mali | PROMIS CS (46) | 696 | 692 | 11.0 | −0.02 (−0.05, 0.01) | 0.26 | 0.26 | 217 | 242 | 8.7 | −0.04 (−0.08, 0.00) | 0.29 | 0.26 |
| Zimbabwe | SHINE (HIV−) (47) | | | | | | | | | | | | |
| Zimbabwe | SHINE (HIV+) (48) | | | | | | | | | | | | |
| | | **4594** | **2910** | | **I² = 0.00, Tau² = 0.00** | | | **2044** | **1394** | | **I² = 0.42, Tau² = 0.00** | | |
| **Fixed** | | | | | **−0.02 (−0.04, −0.01)** | | | | | | **−0.01 (−0.03, 0.01)** | | |
| **Random** | | | | | **−0.02 (−0.04, −0.01)** | | | | | | **0.00 (−0.03, 0.02)** | | |

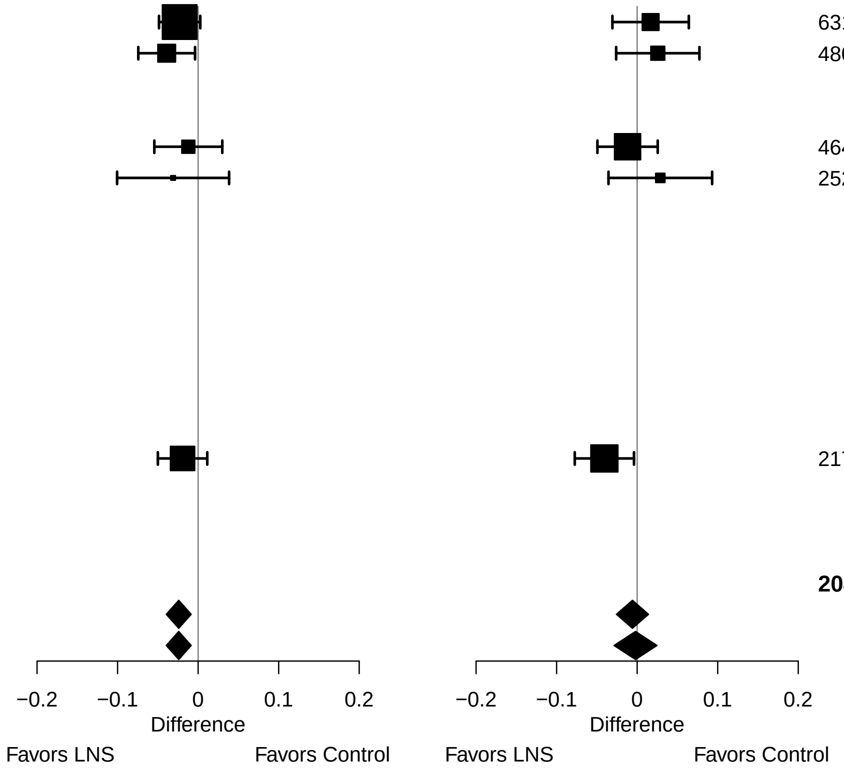

#### Supplemental figure 7F: Wasting prevalence difference

#### 7F5: Stratified by Home environment

**P−for−interaction = 0.413**
**Difference in PDs = 0.00 (−0.02, 0.01)**

| Country | Trial | At least median: LNS N | Control N | Control Prevalence | PD (95% CI) | Fixed W | Random W | Less than median: LNS N | Control N | Control Prevalence | PD (95% CI) | Fixed W | Random W |
|---|---|---|---|---|---|---|---|---|---|---|---|---|---|
| Bangladesh | JiVitA−4 (34) | | | | | | | | | | | | |
| Bangladesh | RDNS (35) | 1192 | 582 | 14.6 | −0.03 (−0.07, 0.01) | 0.04 | 0.12 | 466 | 230 | 15.7 | 0.01 (−0.04, 0.06) | 0.03 | 0.03 |
| Bangladesh | WASH−B (36) | 636 | 1652 | 10.4 | −0.03 (−0.06, −0.01) | 0.09 | 0.18 | 492 | 1684 | 11.9 | −0.01 (−0.04, 0.03) | 0.08 | 0.08 |
| Burkina Faso | iLiNS−Zinc (37) | 473 | 199 | 11.6 | −0.04 (−0.08, 0.00) | 0.03 | 0.11 | 271 | 174 | 17.7 | −0.04 (−0.11, 0.02) | 0.02 | 0.02 |
| Burkina Faso | PROMIS (38) | | | | | | | | | | | | |
| Burkina Faso | PROMIS CS (38) | | | | | | | | | | | | |
| Ghana | GHANA (39) | | | | | | | | | | | | |
| Ghana | iLiNS−DYADG (40) | 213 | 409 | 6.4 | 0.00 (−0.04, 0.04) | 0.03 | 0.10 | 119 | 248 | 9.7 | −0.05 (−0.11, 0.00) | 0.02 | 0.02 |
| Haiti | HAITI (41) | | | | | | | | | | | | |
| Kenya | WASH−B (42) | 834 | 2757 | 1.1 | 0.00 (−0.01, 0.01) | 0.66 | 0.28 | 573 | 2149 | 1.5 | 0.00 (−0.01, 0.01) | 0.77 | 0.77 |
| Madagascar | MAHAY (43) | 914 | 907 | 3.7 | 0.01 (−0.01, 0.02) | 0.14 | 0.21 | 786 | 775 | 8.3 | −0.01 (−0.04, 0.02) | 0.08 | 0.08 |
| Malawi | iLiNS−DYADM (44) | | | | | | | | | | | | |
| Malawi | iLiNS−DOSE (45) | | | | | | | | | | | | |
| Mali | PROMIS (46) | | | | | | | | | | | | |
| Mali | PROMIS CS (46) | | | | | | | | | | | | |
| Zimbabwe | SHINE (HIV−) (47) | | | | | | | | | | | | |
| Zimbabwe | SHINE (HIV+) (48) | | | | | | | | | | | | |
| | | **4262** | **6506** | | **I² = 0.60, Tau² = 0.00** | | | **2707** | **5260** | | **I² = 0.00, Tau² = 0.00** | | |
| **Fixed** | | | | | **0.00 (−0.01, 0.00)** | | | | | | **−0.01 (−0.01, 0.00)** | | |
| **Random** | | | | | **−0.01 (−0.03, 0.00)** | | | | | | **−0.01 (−0.01, 0.00)** | | |

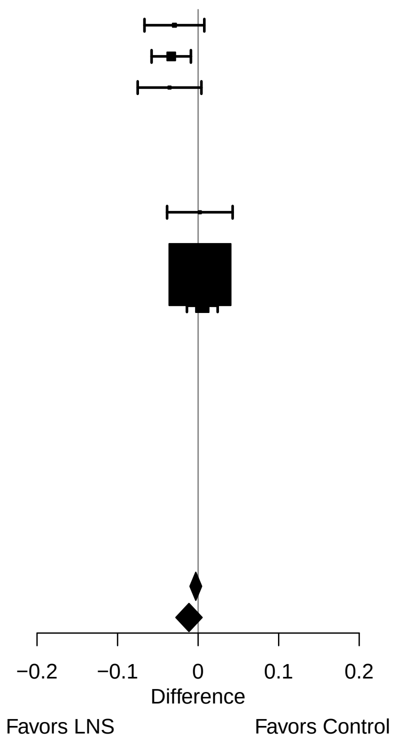

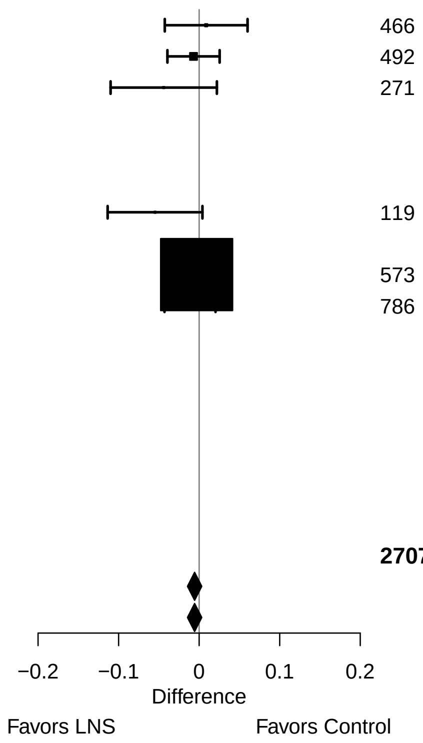

#### Supplemental figure 7F: Wasting prevalence difference

##### 7F6: Stratified by Season at the time of assessment

**P−for−interaction = 0.536**
**Difference in PDs = 0.00 (−0.01, 0.02)**

| | | Dry | | | | | | Rainy | | | | | |
|---|---|---|---|---|---|---|---|---|---|---|---|---|---|
| **Country** | **Trial** | **LNS N** | **Control N** | **Control Prevalence** | **PD (95% CI)** | **Fixed W** | **Random W** | **LNS N** | **Control N** | **Control Prevalence** | **PD (95% CI)** | **Fixed W** | **Random W** |
| Bangladesh | JiVitA−4 (34) | 1520 | 649 | 13.7 | −0.02 (−0.04, 0.01) | 0.10 | 0.13 | 1263 | 568 | 19.4 | −0.01 (−0.05, 0.03) | 0.03 | 0.03 |
| Bangladesh | RDNS (35) | 738 | 359 | 13.6 | −0.03 (−0.07, 0.02) | 0.05 | 0.07 | 923 | 456 | 16.2 | −0.02 (−0.06, 0.03) | 0.02 | 0.02 |
| Bangladesh | WASH−B (36) | 639 | 1912 | 8.9 | −0.02 (−0.04, 0.00) | 0.16 | 0.18 | 517 | 1512 | 14.2 | −0.03 (−0.06, 0.00) | 0.05 | 0.05 |
| Burkina Faso | iLiNS−Zinc (37) | 1394 | 477 | 13.8 | −0.05 (−0.07, −0.03) | 0.18 | 0.18 | 558 | 187 | 12.8 | −0.03 (−0.08, 0.03) | 0.02 | 0.02 |
| Burkina Faso | PROMIS (38) | 428 | 445 | 8.5 | −0.01 (−0.05, 0.03) | 0.05 | 0.08 | 428 | 462 | 9.3 | −0.01 (−0.05, 0.03) | 0.03 | 0.03 |
| Burkina Faso | PROMIS CS (38) | 224 | 190 | 13.2 | −0.02 (−0.10, 0.05) | 0.02 | 0.03 | 206 | 246 | 11.8 | 0.03 (−0.03, 0.10) | 0.01 | 0.01 |
| Ghana | GHANA (39) | | | | | | | | | | | | |
| Ghana | iLiNS−DYADG (40) | | | | | | | | | | | | |
| Haiti | HAITI (41) | | | | | | | | | | | | |
| Kenya | WASH−B (42) | 346 | 1146 | 1.9 | 0.00 (−0.02, 0.01) | 0.40 | 0.27 | 1109 | 3972 | 1.4 | 0.00 (−0.01, 0.01) | 0.83 | 0.83 |
| Madagascar | MAHAY (43) | | | | | | | | | | | | |
| Malawi | iLiNS−DYADM (44) | | | | | | | | | | | | |
| Malawi | iLiNS−DOSE (45) | 415 | 134 | 4.5 | 0.01 (−0.03, 0.06) | 0.04 | 0.06 | 281 | 107 | 6.5 | 0.00 (−0.05, 0.06) | 0.01 | 0.01 |
| Mali | PROMIS (46) | | | | | | | | | | | | |
| Mali | PROMIS CS (46) | | | | | | | | | | | | |
| Zimbabwe | SHINE (HIV−) (47) | | | | | | | | | | | | |
| Zimbabwe | SHINE (HIV+) (48) | | | | | | | | | | | | |
| | | **5704** | **5312** | | **I² = 0.42, Tau² = 0.00** | | | **5285** | **7510** | | **I² = 0.00, Tau² = 0.00** | | |
| **Fixed** | | | | | **−0.02 (−0.03, −0.01)** | | | | | | **0.00 (−0.01, 0.00)** | | |
| **Random** | | | | | **−0.02 (−0.03, −0.01)** | | | | | | **0.00 (−0.01, 0.00)** | | |

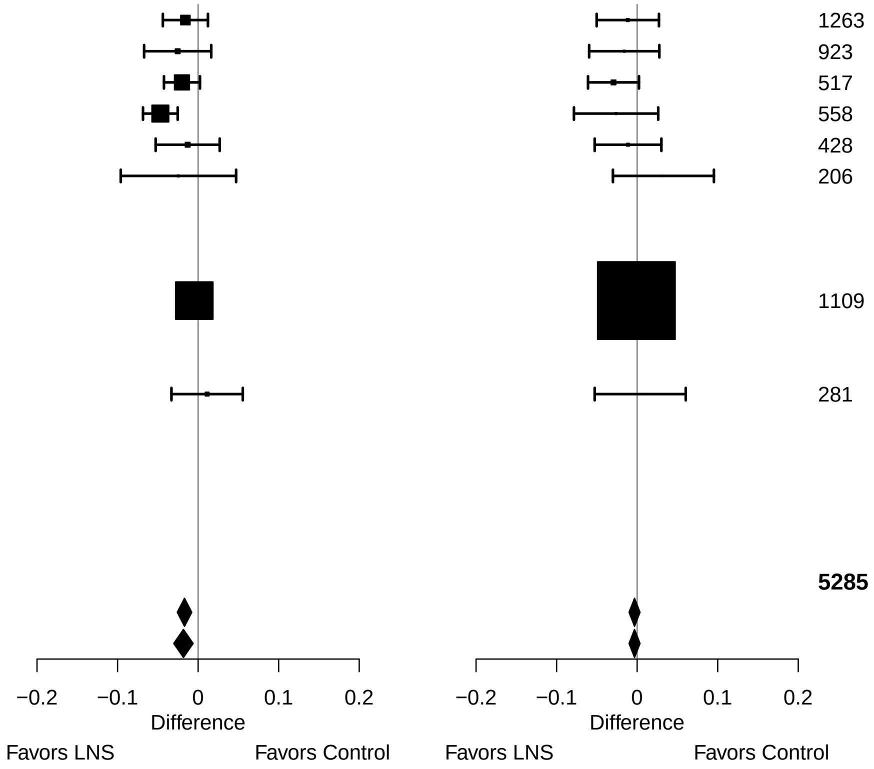

### Supplemental figure 7G: Mean difference in MUACZ

#### 7G1: Stratified by Household socio-economic status

**P−for−interaction = 0.768**
**Difference in MDs = 0.01 (−0.03, 0.04)**

| | | At least median | | | | | | Less than median | | | | | |
|---|---|---|---|---|---|---|---|---|---|---|---|---|---|
| **Country** | **Trial** | **LNS N** | **Control N** | **Control Mean** | **MD (95% CI)** | **Fixed W** | **Random W** | **LNS N** | **Control N** | **Control Mean** | **MD (95% CI)** | **Fixed W** | **Random W** |
| Bangladesh | JiVitA−4 (34) | 1463 | 641 | −0.95 | 0.09 (0.03, 0.15) | 0.29 | 0.12 | 1433 | 630 | −1.18 | 0.07 (0.01, 0.13) | 0.24 | 0.13 |
| Bangladesh | RDNS (35) | 834 | 407 | −0.60 | 0.05 (−0.06, 0.15) | 0.09 | 0.09 | 829 | 408 | −0.80 | 0.03 (−0.07, 0.12) | 0.11 | 0.10 |
| Bangladesh | WASH−B (36) | | | | | | | | | | | | |
| Burkina Faso | iLiNS−Zinc (37) | 1232 | 300 | −1.09 | 0.30 (0.18, 0.42) | 0.06 | 0.08 | 720 | 365 | −1.16 | 0.29 (0.20, 0.38) | 0.11 | 0.10 |
| Burkina Faso | PROMIS (38) | 403 | 479 | −0.71 | 0.08 (−0.09, 0.25) | 0.03 | 0.05 | 451 | 433 | −0.76 | 0.10 (−0.02, 0.22) | 0.07 | 0.08 |
| Burkina Faso | PROMIS CS (38) | 219 | 207 | −0.82 | −0.06 (−0.26, 0.13) | 0.02 | 0.05 | 205 | 229 | −0.81 | −0.06 (−0.27, 0.15) | 0.02 | 0.04 |
| Ghana | GHANA (39) | | | | | | | | | | | | |
| Ghana | iLiNS−DYADG (40) | 157 | 356 | −0.40 | 0.07 (−0.10, 0.25) | 0.03 | 0.05 | 190 | 335 | −0.54 | 0.11 (−0.05, 0.27) | 0.04 | 0.06 |
| Haiti | HAITI (41) | | | | | | | | | | | | |
| Kenya | WASH−B (42) | 827 | 2949 | −0.39 | 0.14 (0.06, 0.22) | 0.15 | 0.11 | 632 | 2232 | −0.55 | 0.09 (−0.01, 0.19) | 0.10 | 0.10 |
| Madagascar | MAHAY (43) | 761 | 865 | −0.61 | −0.05 (−0.22, 0.12) | 0.03 | 0.06 | 866 | 750 | −0.87 | 0.02 (−0.12, 0.17) | 0.05 | 0.07 |
| Malawi | iLiNS−DYADM (44) | 121 | 234 | 0.14 | −0.06 (−0.26, 0.15) | 0.02 | 0.04 | 99 | 209 | 0.00 | −0.02 (−0.27, 0.23) | 0.02 | 0.03 |
| Malawi | iLiNS−DOSE (45) | 303 | 104 | 0.18 | 0.05 (−0.11, 0.21) | 0.03 | 0.06 | 292 | 102 | −0.06 | 0.00 (−0.17, 0.18) | 0.03 | 0.05 |
| Mali | PROMIS (46) | 248 | 250 | −0.78 | 0.17 (0.02, 0.31) | 0.04 | 0.07 | 246 | 246 | −0.76 | 0.19 (0.01, 0.37) | 0.03 | 0.05 |
| Mali | PROMIS CS (46) | 505 | 463 | −0.86 | 0.16 (0.01, 0.31) | 0.04 | 0.07 | 449 | 507 | −0.80 | 0.10 (−0.04, 0.24) | 0.05 | 0.06 |
| Zimbabwe | SHINE (HIV−) (47) | 905 | 797 | 0.11 | 0.00 (−0.09, 0.08) | 0.13 | 0.10 | 827 | 852 | −0.11 | 0.15 (0.06, 0.24) | 0.12 | 0.10 |
| Zimbabwe | SHINE (HIV+) (48) | 171 | 156 | −0.14 | 0.02 (−0.17, 0.21) | 0.02 | 0.05 | 158 | 166 | −0.23 | 0.04 (−0.17, 0.25) | 0.02 | 0.04 |
| | | **8149** | **8208** | | **I² = 0.54, Tau² = 0.00** | | | **7397** | **7464** | | **I² = 0.52, Tau² = 0.00** | | |
| **Fixed** | | | | | **0.08 (0.05, 0.11)** | | | | | | **0.10 (0.07, 0.13)** | | |
| **Random** | | | | | **0.08 (0.03, 0.13)** | | | | | | **0.10 (0.05, 0.14)** | | |

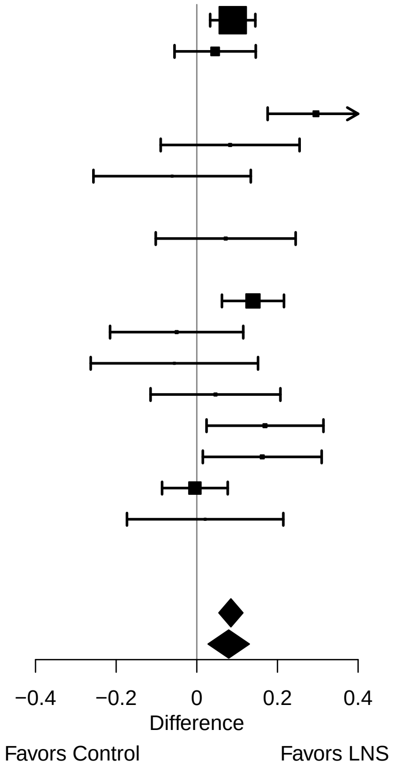

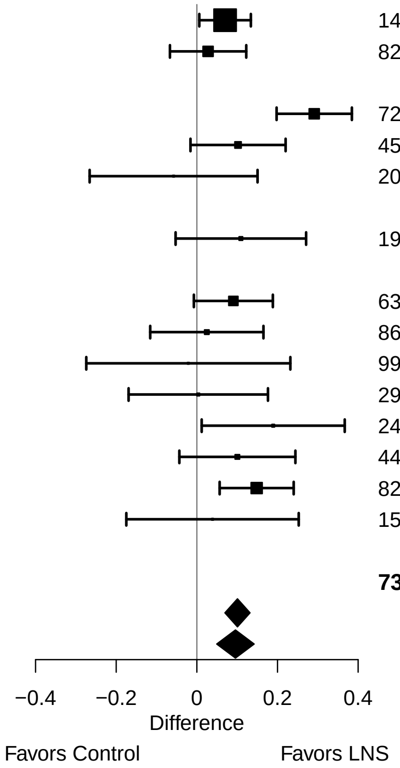

#### Supplemental figure 7G: Mean difference in MUACZ

#### 7G2: Stratified by Household food insecurity

P−for−interaction = 0.595
Difference in MDs = −0.01 (−0.06, 0.03)

| | | Mild to secure | | | | | | Moderate to severe | | | | | |
|---|---|---|---|---|---|---|---|---|---|---|---|---|---|
| Country | Trial | LNS N | Control N | Control Mean | MD (95% CI) | Fixed W | Random W | LNS N | Control N | Control Mean | MD (95% CI) | Fixed W | Random W |
| Bangladesh | JiVitA−4 (34) | 2054 | 882 | −1.03 | 0.11 (0.06, 0.16) | 0.29 | 0.14 | 842 | 389 | −1.14 | 0.01 (−0.07, 0.09) | 0.26 | 0.13 |
| Bangladesh | RDNS (35) | 1058 | 493 | −0.62 | 0.00 (−0.09, 0.09) | 0.09 | 0.11 | 605 | 322 | −0.83 | 0.09 (−0.02, 0.20) | 0.12 | 0.11 |
| Bangladesh | WASH−B (36) | | | | | | | | | | | | |
| Burkina Faso | iLiNS−Zinc (37) | 1017 | 321 | −1.08 | 0.29 (0.17, 0.41) | 0.06 | 0.09 | 935 | 344 | −1.17 | 0.30 (0.19, 0.40) | 0.14 | 0.12 |
| Burkina Faso | PROMIS (38) | 496 | 547 | −0.70 | 0.12 (−0.04, 0.28) | 0.03 | 0.06 | 358 | 365 | −0.78 | 0.05 (−0.09, 0.19) | 0.08 | 0.09 |
| Burkina Faso | PROMIS CS (38) | | | | | | | | | | | | |
| Ghana | GHANA (39) | | | | | | | | | | | | |
| Ghana | iLiNS−DYADG (40) | 244 | 471 | −0.45 | 0.09 (−0.05, 0.24) | 0.04 | 0.07 | 102 | 219 | −0.50 | 0.07 (−0.13, 0.28) | 0.04 | 0.06 |
| Haiti | HAITI (41) | | | | | | | | | | | | |
| Kenya | WASH−B (42) | 1303 | 4640 | −0.44 | 0.12 (0.05, 0.18) | 0.19 | 0.13 | 156 | 528 | −0.54 | 0.12 (−0.08, 0.32) | 0.04 | 0.06 |
| Madagascar | MAHAY (43) | 968 | 907 | −0.67 | −0.06 (−0.21, 0.10) | 0.03 | 0.07 | 353 | 405 | −0.81 | −0.02 (−0.17, 0.14) | 0.06 | 0.08 |
| Malawi | iLiNS−DYADM (44) | 70 | 122 | 0.28 | −0.07 (−0.35, 0.20) | 0.01 | 0.03 | 148 | 316 | 0.00 | −0.04 (−0.24, 0.15) | 0.04 | 0.06 |
| Malawi | iLiNS−DOSE (45) | 151 | 55 | 0.11 | 0.15 (−0.07, 0.38) | 0.02 | 0.04 | 431 | 145 | 0.00 | 0.00 (−0.14, 0.14) | 0.08 | 0.09 |
| Mali | PROMIS (46) | 330 | 341 | −0.75 | 0.15 (0.00, 0.31) | 0.03 | 0.07 | 174 | 163 | −0.85 | 0.25 (0.05, 0.45) | 0.04 | 0.06 |
| Mali | PROMIS CS (46) | | | | | | | | | | | | |
| Zimbabwe | SHINE (HIV−) (47) | 1386 | 1295 | 0.04 | 0.06 (−0.01, 0.12) | 0.18 | 0.13 | 295 | 322 | −0.16 | 0.15 (0.01, 0.29) | 0.08 | 0.09 |
| Zimbabwe | SHINE (HIV+) (48) | 236 | 238 | −0.20 | 0.06 (−0.11, 0.22) | 0.03 | 0.06 | 86 | 76 | −0.15 | 0.01 (−0.24, 0.26) | 0.02 | 0.04 |
| | | **9313** | **10312** | | **I² = 0.52, Tau² = 0.00** | | | **4485** | **3594** | | **I² = 0.61, Tau² = 0.01** | | |
| **Fixed** | | | | | **0.09 (0.07, 0.12)** | | | | | | **0.09 (0.05, 0.13)** | | |
| **Random** | | | | | **0.09 (0.04, 0.14)** | | | | | | **0.09 (0.03, 0.15)** | | |

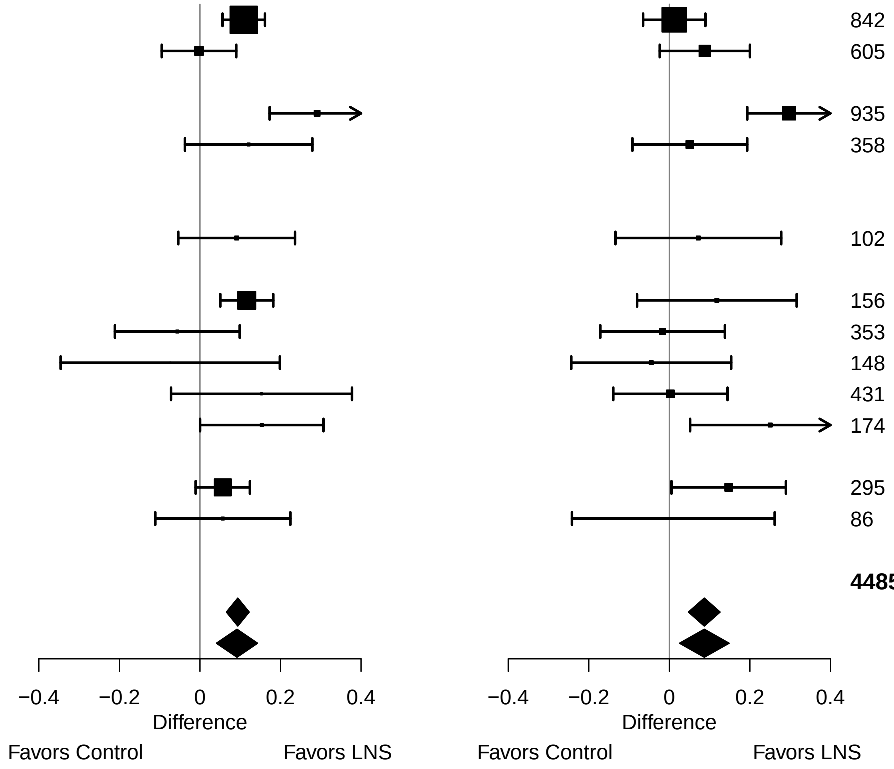

#### Supplemental figure 7G: Mean difference in MUACZ

#### 7G3: Stratified by Household source water quality

**P−for−interaction = 0.924**
**Difference in MDs = 0.00 (−0.06, 0.06)**

| Country | Trial | Improved LNS N | Improved Control N | Improved Control Mean | Improved MD (95% CI) | Improved Fixed W | Improved Random W | Unimproved LNS N | Unimproved Control N | Unimproved Control Mean | Unimproved MD (95% CI) | Unimproved Fixed W | Unimproved Random W |
|---|---|---|---|---|---|---|---|---|---|---|---|---|---|
| Bangladesh | JiVitA−4 (34) | | | | | | | | | | | | |
| Bangladesh | RDNS (35) | | | | | | | | | | | | |
| Bangladesh | WASH−B (36) | | | | | | | | | | | | |
| Burkina Faso | iLiNS−Zinc (37) | 555 | 141 | −1.08 | 0.27 (0.15, 0.38) | 0.14 | 0.12 | 1396 | 521 | −1.15 | 0.31 (0.21, 0.40) | 0.28 | 0.17 |
| Burkina Faso | PROMIS (38) | 352 | 376 | −0.77 | 0.10 (−0.06, 0.25) | 0.08 | 0.09 | 358 | 355 | −0.72 | 0.12 (−0.07, 0.31) | 0.07 | 0.09 |
| Burkina Faso | PROMIS CS (38) | 277 | 265 | −0.81 | −0.06 (−0.26, 0.13) | 0.05 | 0.07 | 147 | 171 | −0.82 | −0.05 (−0.32, 0.21) | 0.04 | 0.06 |
| Ghana | GHANA (39) | | | | | | | | | | | | |
| Ghana | iLiNS−DYADG (40) | | | | | | | | | | | | |
| Haiti | HAITI (41) | | | | | | | | | | | | |
| Kenya | WASH−B (42) | 449 | 1562 | −0.43 | 0.08 (−0.02, 0.19) | 0.17 | 0.13 | 248 | 701 | −0.49 | 0.13 (−0.02, 0.27) | 0.12 | 0.12 |
| Madagascar | MAHAY (43) | 415 | 452 | −0.65 | −0.08 (−0.34, 0.18) | 0.03 | 0.04 | 1185 | 1157 | −0.76 | −0.01 (−0.13, 0.12) | 0.17 | 0.14 |
| Malawi | iLiNS−DYADM (44) | 200 | 406 | 0.09 | −0.04 (−0.21, 0.13) | 0.06 | 0.08 | 20 | 36 | −0.07 | 0.06 (−0.49, 0.60) | 0.01 | 0.02 |
| Malawi | iLiNS−DOSE (45) | 556 | 192 | 0.08 | 0.03 (−0.09, 0.15) | 0.12 | 0.11 | 51 | 17 | −0.27 | 0.05 (−0.37, 0.47) | 0.01 | 0.03 |
| Mali | PROMIS (46) | 249 | 258 | −0.73 | 0.21 (0.04, 0.38) | 0.06 | 0.08 | 248 | 242 | −0.84 | 0.16 (−0.02, 0.34) | 0.08 | 0.10 |
| Mali | PROMIS CS (46) | 549 | 567 | −0.82 | 0.13 (0.00, 0.26) | 0.11 | 0.11 | 364 | 388 | −0.84 | 0.13 (−0.03, 0.29) | 0.10 | 0.11 |
| Zimbabwe | SHINE (HIV−) (47) | 519 | 482 | 0.00 | 0.09 (−0.02, 0.20) | 0.16 | 0.13 | 273 | 300 | 0.07 | −0.06 (−0.23, 0.10) | 0.10 | 0.11 |
| Zimbabwe | SHINE (HIV+) (48) | 85 | 87 | −0.23 | 0.12 (−0.14, 0.39) | 0.03 | 0.04 | 55 | 56 | −0.17 | −0.11 (−0.49, 0.27) | 0.02 | 0.03 |
| | | **4206** | **4788** | | **I² = 0.46, Tau² = 0.00** | | | **4345** | **3944** | | **I² = 0.63, Tau² = 0.01** | | |
| **Fixed** | | | | | **0.10 (0.06, 0.14)** | | | | | | **0.13 (0.08, 0.18)** | | |
| **Random** | | | | | **0.09 (0.03, 0.15)** | | | | | | **0.10 (0.02, 0.17)** | | |

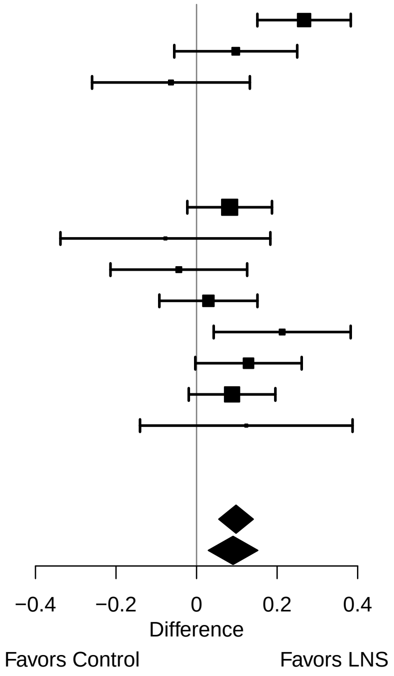

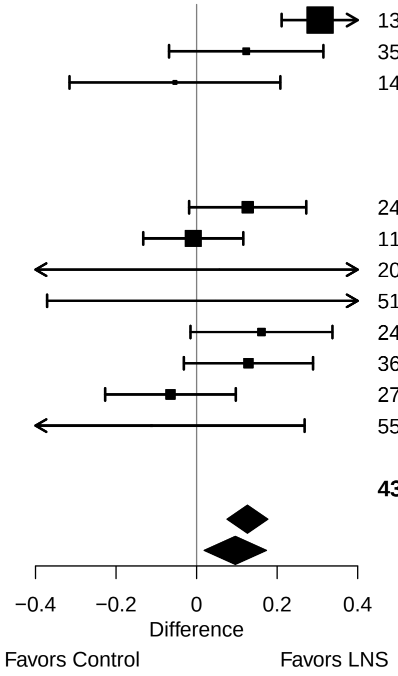

#### Supplemental figure 7G: Mean difference in MUACZ

#### 7G4: Stratified by Household sanitation

**P−for−interaction = 0.026**
**Difference in MDs = −0.06 (−0.12, −0.01)**

| Country | Trial | LNS N (Improved) | Control N (Improved) | Control Mean (Improved) | Improved MD (95% CI) | Fixed W (Improved) | Random W (Improved) | LNS N (Unimproved) | Control N (Unimproved) | Control Mean (Unimproved) | Unimproved MD (95% CI) | Fixed W (Unimproved) | Random W (Unimproved) |
|---|---|---|---|---|---|---|---|---|---|---|---|---|---|
| Bangladesh | JiVitA−4 (34) | 2241 | 974 | −1.01 | 0.09 (0.04, 0.14) | 0.46 | 0.46 | 655 | 297 | −1.23 | 0.05 (−0.03, 0.13) | 0.19 | 0.11 |
| Bangladesh | RDNS (35) | 1179 | 573 | −0.69 | 0.08 (0.00, 0.16) | 0.16 | 0.16 | 482 | 241 | −0.75 | −0.06 (−0.20, 0.07) | 0.07 | 0.09 |
| Bangladesh | WASH−B (36) | | | | | | | | | | | | |
| Burkina Faso | iLiNS−Zinc (37) | 43 | 16 | −1.33 | 0.33 (−0.14, 0.81) | 0.01 | 0.01 | 1908 | 646 | −1.13 | 0.30 (0.21, 0.39) | 0.17 | 0.11 |
| Burkina Faso | PROMIS (38) | 395 | 548 | −0.73 | 0.15 (0.00, 0.29) | 0.05 | 0.05 | 459 | 359 | −0.74 | 0.04 (−0.10, 0.18) | 0.07 | 0.08 |
| Burkina Faso | PROMIS CS (38) | 175 | 163 | −0.86 | −0.03 (−0.24, 0.18) | 0.03 | 0.03 | 248 | 273 | −0.79 | −0.08 (−0.26, 0.11) | 0.04 | 0.07 |
| Ghana | GHANA (39) | | | | | | | | | | | | |
| Ghana | iLiNS−DYADG (40) | 336 | 672 | −0.46 | 0.07 (−0.05, 0.19) | 0.08 | 0.08 | 10 | 19 | −0.64 | 0.53 (−0.32, 1.37) | 0.00 | 0.01 |
| Haiti | HAITI (41) | | | | | | | | | | | | |
| Kenya | WASH−B (42) | 101 | 366 | −0.34 | 0.15 (−0.04, 0.35) | 0.03 | 0.03 | 596 | 1898 | −0.47 | 0.09 (−0.01, 0.19) | 0.14 | 0.11 |
| Madagascar | MAHAY (43) | | | | | | | | | | | | |
| Malawi | iLiNS−DYADM (44) | 20 | 42 | 0.04 | 0.49 (0.01, 0.96) | 0.01 | 0.01 | 200 | 400 | 0.08 | −0.09 (−0.26, 0.08) | 0.05 | 0.07 |
| Malawi | iLiNS−DOSE (45) | 15 | 9 | 0.12 | 0.27 (−0.28, 0.83) | 0.00 | 0.00 | 591 | 200 | 0.05 | 0.02 (−0.10, 0.14) | 0.09 | 0.10 |
| Mali | PROMIS (46) | 388 | 366 | −0.77 | 0.18 (0.03, 0.34) | 0.05 | 0.05 | 112 | 134 | −0.81 | 0.18 (−0.02, 0.38) | 0.03 | 0.06 |
| Mali | PROMIS CS (46) | 703 | 696 | −0.81 | 0.15 (0.02, 0.28) | 0.07 | 0.07 | 220 | 249 | −0.87 | 0.12 (−0.07, 0.30) | 0.04 | 0.06 |
| Zimbabwe | SHINE (HIV−) (47) | 293 | 241 | 0.11 | 0.02 (−0.13, 0.18) | 0.05 | 0.05 | 496 | 540 | −0.01 | 0.03 (−0.09, 0.16) | 0.09 | 0.09 |
| Zimbabwe | SHINE (HIV+) (48) | 50 | 35 | −0.13 | −0.02 (−0.43, 0.40) | 0.01 | 0.01 | 90 | 107 | −0.23 | 0.04 (−0.23, 0.31) | 0.02 | 0.04 |
| | | **5939** | **4701** | | **I² = 0.00, Tau² = 0.00** | | | **6067** | **5363** | | **I² = 0.68, Tau² = 0.01** | | |
| **Fixed** | | | | | **0.10 (0.06, 0.13)** | | | | | | **0.08 (0.04, 0.12)** | | |
| **Random** | | | | | **0.10 (0.06, 0.13)** | | | | | | **0.06 (0.00, 0.13)** | | |

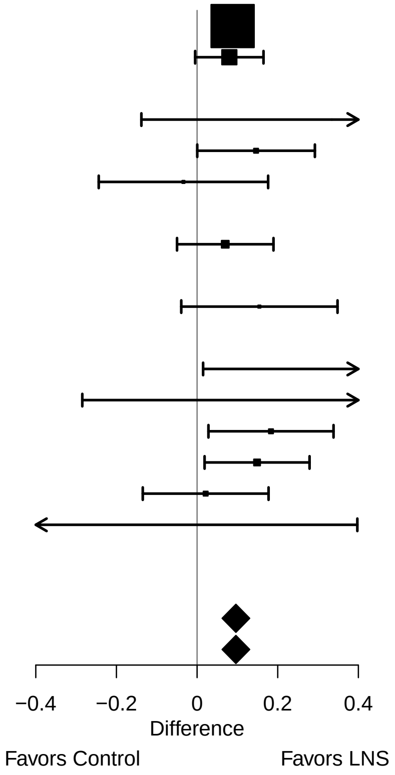

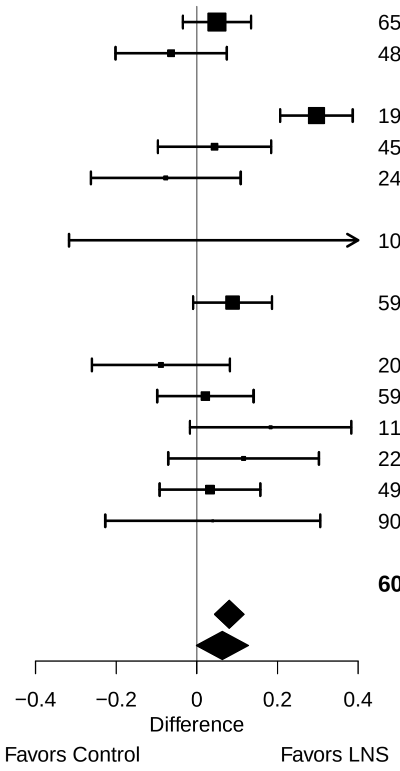

#### Supplemental figure 7G: Mean difference in MUACZ

##### 7G5: Stratified by Home environment

P−for−interaction = 0.048
Difference in MDs = 0.07 (0.00, 0.13)

| | | At least median | | | | | | Less than median | | | | | |
|---|---|---|---|---|---|---|---|---|---|---|---|---|---|
| Country | Trial | LNS N | Control N | Control Mean | MD (95% CI) | Fixed W | Random W | LNS N | Control N | Control Mean | MD (95% CI) | Fixed W | Random W |
| Bangladesh | JiVitA−4 (34) | | | | | | | | | | | | |
| Bangladesh | RDNS (35) | 1192 | 582 | −0.68 | 0.06 (−0.03, 0.16) | 0.22 | 0.16 | 468 | 230 | −0.75 | −0.03 (−0.19, 0.13) | 0.13 | 0.15 |
| Bangladesh | WASH−B (36) | | | | | | | | | | | | |
| Burkina Faso | iLiNS−Zinc (37) | 473 | 199 | −1.08 | 0.32 (0.20, 0.44) | 0.13 | 0.15 | 272 | 175 | −1.22 | 0.32 (0.12, 0.52) | 0.08 | 0.13 |
| Burkina Faso | PROMIS (38) | | | | | | | | | | | | |
| Burkina Faso | PROMIS CS (38) | | | | | | | | | | | | |
| Ghana | GHANA (39) | | | | | | | | | | | | |
| Ghana | iLiNS−DYADG (40) | 213 | 410 | −0.45 | 0.04 (−0.11, 0.20) | 0.08 | 0.13 | 119 | 248 | −0.52 | 0.19 (−0.01, 0.38) | 0.08 | 0.13 |
| Haiti | HAITI (41) | | | | | | | | | | | | |
| Kenya | WASH−B (42) | 840 | 2782 | −0.39 | 0.03 (−0.05, 0.11) | 0.33 | 0.17 | 573 | 2179 | −0.50 | 0.19 (0.10, 0.28) | 0.43 | 0.21 |
| Madagascar | MAHAY (43) | 905 | 889 | −0.68 | −0.04 (−0.19, 0.10) | 0.09 | 0.14 | 775 | 769 | −0.79 | −0.02 (−0.16, 0.13) | 0.15 | 0.17 |
| Malawi | iLiNS−DYADM (44) | 137 | 289 | 0.15 | −0.09 (−0.28, 0.10) | 0.05 | 0.11 | 78 | 149 | −0.07 | 0.05 (−0.25, 0.35) | 0.04 | 0.08 |
| Malawi | iLiNS−DOSE (45) | 378 | 136 | −0.02 | 0.13 (−0.02, 0.27) | 0.09 | 0.14 | 275 | 86 | 0.07 | −0.02 (−0.21, 0.16) | 0.09 | 0.13 |
| Mali | PROMIS (46) | | | | | | | | | | | | |
| Mali | PROMIS CS (46) | | | | | | | | | | | | |
| Zimbabwe | SHINE (HIV−) (47) | | | | | | | | | | | | |
| Zimbabwe | SHINE (HIV+) (48) | | | | | | | | | | | | |
| | | **4138** | **5287** | | **I² = 0.75, Tau² = 0.01** | | | **2560** | **3836** | | **I² = 0.63, Tau² = 0.01** | | |
| **Fixed** | | | | | **0.07 (0.03, 0.12)** | | | | | | **0.12 (0.06, 0.17)** | | |
| **Random** | | | | | **0.07 (−0.03, 0.17)** | | | | | | **0.10 (0.00, 0.20)** | | |

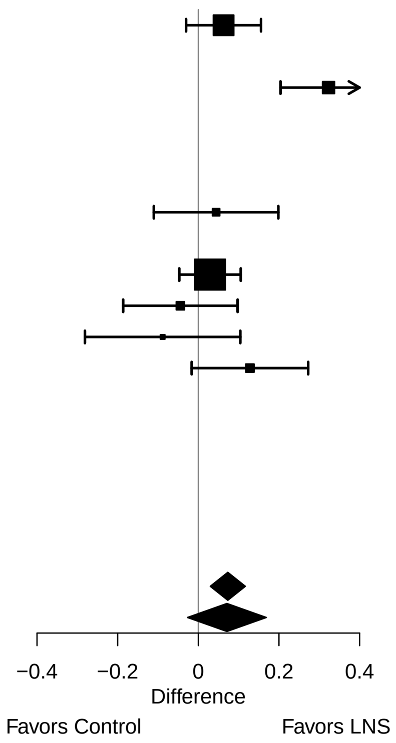

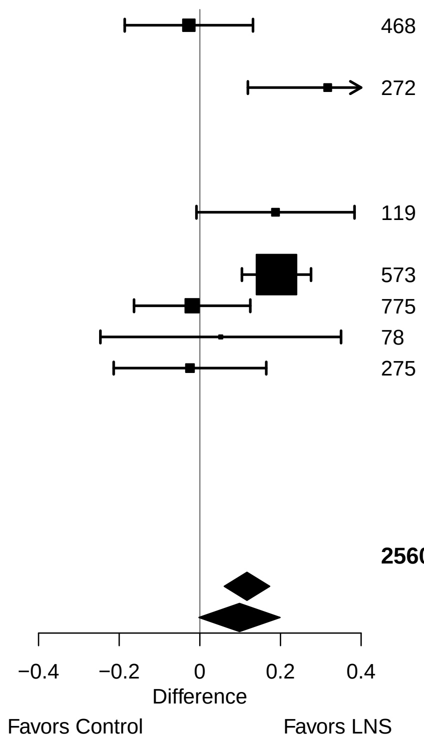

#### Supplemental figure 7G: Mean difference in MUACZ

#### 7G6: Stratified by Season at the time of assessment

**P−for−interaction = 0.977**
**Difference in MDs = 0.00 (−0.05, 0.05)**

| Country | Trial | LNS N | Control N | Control Mean | Dry MD (95% CI) | Fixed W | Random W | LNS N | Control N | Control Mean | Rainy MD (95% CI) | Fixed W | Random W |
|---|---|---|---|---|---|---|---|---|---|---|---|---|---|
| Bangladesh | JiVitA−4 (34) | 1570 | 673 | −1.08 | 0.08 (0.02, 0.14) | 0.28 | 0.13 | 1327 | 598 | −1.05 | 0.08 (0.01, 0.14) | 0.28 | 0.18 |
| Bangladesh | RDNS (35) | 739 | 359 | −0.59 | 0.03 (−0.09, 0.16) | 0.07 | 0.10 | 924 | 456 | −0.79 | 0.04 (−0.04, 0.12) | 0.19 | 0.16 |
| Bangladesh | WASH−B (36) | | | | | | | | | | | | |
| Burkina Faso | iLiNS−Zinc (37) | 1399 | 477 | −1.17 | 0.31 (0.21, 0.42) | 0.09 | 0.11 | 560 | 189 | −1.05 | 0.25 (0.14, 0.36) | 0.09 | 0.12 |
| Burkina Faso | PROMIS (38) | 425 | 444 | −0.74 | 0.04 (−0.10, 0.18) | 0.05 | 0.09 | 429 | 468 | −0.72 | 0.14 (−0.01, 0.29) | 0.05 | 0.08 |
| Burkina Faso | PROMIS CS (38) | 222 | 190 | −0.82 | −0.09 (−0.39, 0.20) | 0.01 | 0.04 | 202 | 246 | −0.81 | −0.02 (−0.19, 0.15) | 0.04 | 0.07 |
| Ghana | GHANA (39) | | | | | | | | | | | | |
| Ghana | iLiNS−DYADG (40) | 204 | 408 | −0.52 | −0.02 (−0.17, 0.13) | 0.05 | 0.08 | 143 | 285 | −0.41 | 0.24 (0.05, 0.43) | 0.03 | 0.06 |
| Haiti | HAITI (41) | | | | | | | | | | | | |
| Kenya | WASH−B (42) | 349 | 1163 | −0.50 | 0.18 (0.05, 0.31) | 0.06 | 0.09 | 1112 | 4022 | −0.44 | 0.10 (0.02, 0.17) | 0.22 | 0.17 |
| Madagascar | MAHAY (43) | | | | | | | | | | | | |
| Malawi | iLiNS−DYADM (44) | 156 | 316 | 0.15 | −0.06 (−0.24, 0.13) | 0.03 | 0.07 | 64 | 128 | −0.11 | 0.01 (−0.30, 0.33) | 0.01 | 0.03 |
| Malawi | iLiNS−DOSE (45) | 421 | 134 | 0.03 | 0.03 (−0.12, 0.18) | 0.05 | 0.08 | 275 | 106 | 0.00 | 0.11 (−0.06, 0.27) | 0.04 | 0.07 |
| Mali | PROMIS (46) | | | | | | | | | | | | |
| Mali | PROMIS CS (46) | | | | | | | | | | | | |
| Zimbabwe | SHINE (HIV−) (47) | 1740 | 1652 | −0.01 | 0.07 (0.01, 0.13) | 0.26 | 0.13 | 132 | 128 | 0.24 | −0.08 (−0.30, 0.14) | 0.02 | 0.05 |
| Zimbabwe | SHINE (HIV+) (48) | 319 | 305 | −0.20 | 0.04 (−0.10, 0.18) | 0.05 | 0.09 | 18 | 23 | −0.03 | 0.06 (−0.45, 0.57) | 0.00 | 0.01 |
| | | **7544** | **6121** | | **$I^2$ = 0.64, $Tau^2$ = 0.01** | | | **5186** | **6649** | | **$I^2$ = 0.39, $Tau^2$ = 0.00** | | |
| **Fixed** | | | | | **0.08 (0.05, 0.12)** | | | | | | **0.09 (0.06, 0.13)** | | |
| **Random** | | | | | **0.07 (0.01, 0.14)** | | | | | | **0.10 (0.04, 0.15)** | | |

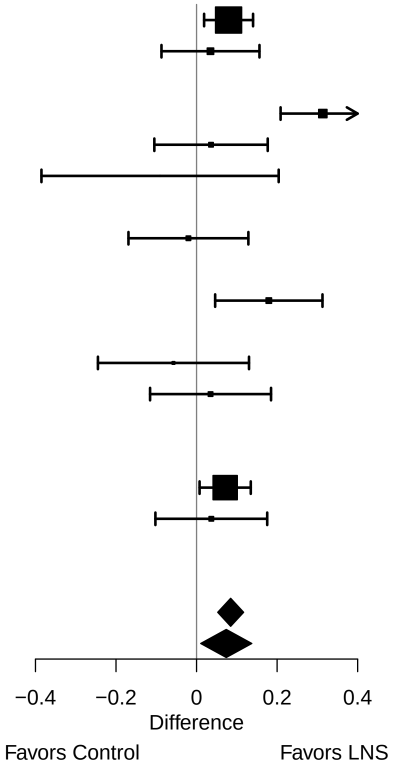

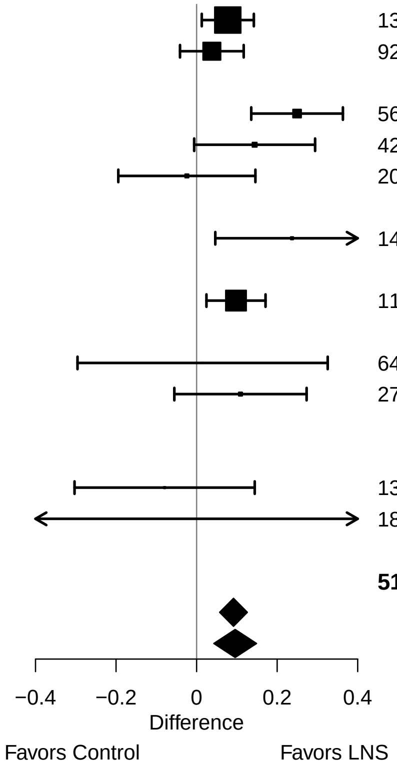

### Supplemental figure 7H: Low MUAC prevalence ratio

#### 7H1: Stratified by Household socio-economic status

**P−for−interaction = 0.129**
**Ratio of PRs = 1.14 (0.96, 1.35)**

| | | At least median | | | | | | Less than median | | | | | |
|---|---|---|---|---|---|---|---|---|---|---|---|---|---|
| **Country** | **Trial** | **LNS N** | **Control N** | **Control Prevalence** | **PR (95% CI)** | **Fixed W** | **Random W** | **LNS N** | **Control N** | **Control Prevalence** | **PR (95% CI)** | **Fixed W** | **Random W** |
| Bangladesh | JiVitA−4 (34) | 1463 | 641 | 11.6 | 0.76 (0.59, 0.96) | 0.29 | 0.29 | 1433 | 630 | 18.8 | 0.87 (0.69, 1.10) | 0.25 | 0.17 |
| Bangladesh | RDNS (35) | 834 | 407 | 4.4 | 0.84 (0.50, 1.40) | 0.06 | 0.06 | 829 | 408 | 5.9 | 1.15 (0.70, 1.89) | 0.06 | 0.08 |
| Bangladesh | WASH−B (36) | | | | | | | | | | | | |
| Burkina Faso | iLiNS−Zinc (37) | 1232 | 300 | 16.0 | 0.60 (0.44, 0.80) | 0.19 | 0.19 | 720 | 365 | 20.0 | 0.68 (0.53, 0.87) | 0.23 | 0.17 |
| Burkina Faso | PROMIS (38) | 403 | 479 | 9.4 | 0.79 (0.45, 1.39) | 0.05 | 0.05 | 451 | 433 | 6.5 | 1.17 (0.70, 1.95) | 0.05 | 0.08 |
| Burkina Faso | PROMIS CS (38) | 219 | 207 | 14.5 | 0.82 (0.50, 1.33) | 0.07 | 0.07 | 205 | 229 | 15.3 | 1.02 (0.69, 1.51) | 0.09 | 0.11 |
| Ghana | GHANA (39) | | | | | | | | | | | | |
| Ghana | iLiNS−DYADG (40) | | | | | | | | | | | | |
| Haiti | HAITI (41) | | | | | | | | | | | | |
| Kenya | WASH−B (42) | 827 | 2949 | 3.7 | 0.72 (0.46, 1.12) | 0.08 | 0.08 | 632 | 2232 | 7.8 | 0.71 (0.48, 1.04) | 0.09 | 0.11 |
| Madagascar | MAHAY (43) | 761 | 865 | 8.1 | 0.97 (0.66, 1.44) | 0.11 | 0.11 | 866 | 750 | 11.2 | 0.91 (0.66, 1.25) | 0.13 | 0.13 |
| Malawi | iLiNS−DYADM (44) | | | | | | | | | | | | |
| Malawi | iLiNS−DOSE (45) | | | | | | | | | | | | |
| Mali | PROMIS (46) | 248 | 250 | 4.8 | 0.90 (0.39, 2.10) | 0.02 | 0.02 | 246 | 246 | 6.9 | 0.33 (0.12, 0.88) | 0.01 | 0.03 |
| Mali | PROMIS CS (46) | 505 | 463 | 12.7 | 0.73 (0.50, 1.07) | 0.12 | 0.12 | 449 | 507 | 9.5 | 1.13 (0.76, 1.67) | 0.09 | 0.11 |
| Zimbabwe | SHINE (HIV−) (47) | | | | | | | | | | | | |
| Zimbabwe | SHINE (HIV+) (48) | | | | | | | | | | | | |
| | | **6492** | **6561** | | **I² = 0.00, Tau² = 0.00** | | | **5831** | **5800** | | **I² = 0.41, Tau² = 0.03** | | |
| **Fixed** | | | | | **0.75 (0.66, 0.85)** | | | | | | **0.86 (0.76, 0.96)** | | |
| **Random** | | | | | **0.75 (0.66, 0.85)** | | | | | | **0.88 (0.73, 1.05)** | | |

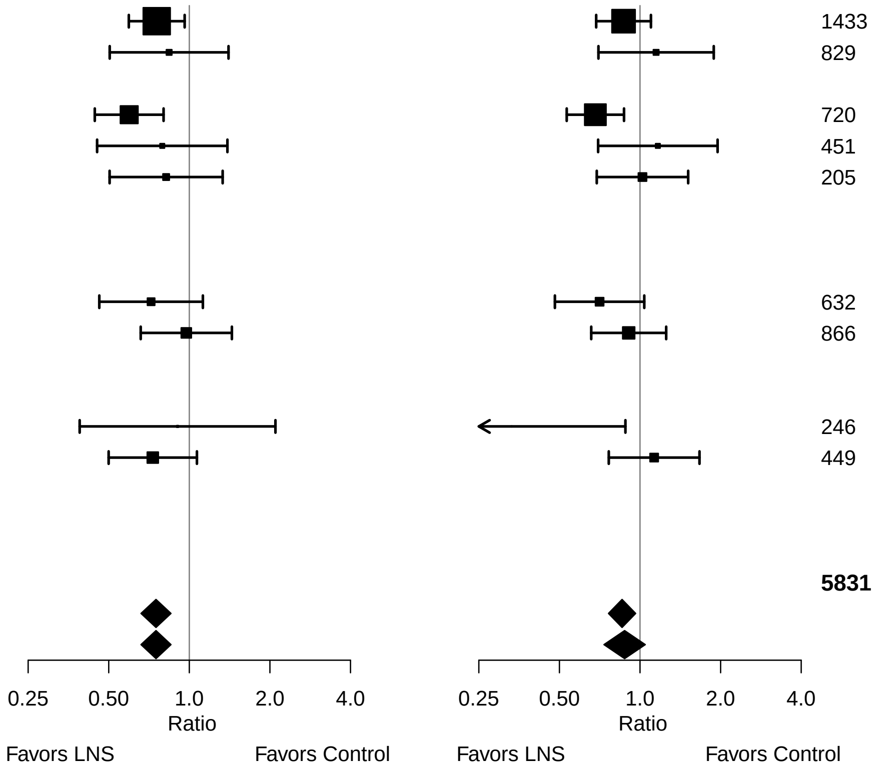

#### Supplemental figure 7H: Low MUAC prevalence ratio

#### 7H2: Stratified by Household food insecurity

**P−for−interaction = 0.182**
**Ratio of PRs = 1.15 (0.93, 1.42)**

| | | Mild to secure | | | | | | Moderate to severe | | | | | |
|---|---|---|---|---|---|---|---|---|---|---|---|---|---|
| **Country** | **Trial** | **LNS N** | **Control N** | **Control Prevalence** | **PR (95% CI)** | **Fixed W** | **Random W** | **LNS N** | **Control N** | **Control Prevalence** | **PR (95% CI)** | **Fixed W** | **Random W** |
| Bangladesh | JiVitA−4 (34) | 2054 | 882 | 14.6 | 0.69 (0.57, 0.84) | 0.39 | 0.25 | 842 | 389 | 16.3 | 1.09 (0.80, 1.51) | 0.29 | 0.27 |
| Bangladesh | RDNS (35) | 1058 | 493 | 4.9 | 1.01 (0.59, 1.74) | 0.05 | 0.08 | 605 | 322 | 5.6 | 1.03 (0.57, 1.88) | 0.08 | 0.09 |
| Bangladesh | WASH−B (36) | | | | | | | | | | | | |
| Burkina Faso | iLiNS−Zinc (37) | 1017 | 321 | 16.2 | 0.62 (0.47, 0.81) | 0.18 | 0.19 | 935 | 344 | 20.1 | 0.65 (0.48, 0.88) | 0.31 | 0.29 |
| Burkina Faso | PROMIS (38) | 496 | 547 | 7.3 | 1.10 (0.73, 1.66) | 0.08 | 0.12 | 358 | 365 | 9.0 | 0.74 (0.41, 1.34) | 0.08 | 0.09 |
| Burkina Faso | PROMIS CS (38) | | | | | | | | | | | | |
| Ghana | GHANA (39) | | | | | | | | | | | | |
| Ghana | iLiNS−DYADG (40) | | | | | | | | | | | | |
| Haiti | HAITI (41) | | | | | | | | | | | | |
| Kenya | WASH−B (42) | 1303 | 4640 | 5.4 | 0.67 (0.50, 0.91) | 0.15 | 0.17 | 156 | 528 | 6.2 | 1.03 (0.52, 2.03) | 0.06 | 0.07 |
| Madagascar | MAHAY (43) | 968 | 907 | 9.5 | 0.96 (0.69, 1.33) | 0.13 | 0.16 | 353 | 405 | 9.1 | 1.05 (0.67, 1.65) | 0.14 | 0.15 |
| Malawi | iLiNS−DYADM (44) | | | | | | | | | | | | |
| Malawi | iLiNS−DOSE (45) | | | | | | | | | | | | |
| Mali | PROMIS (46) | 330 | 341 | 5.0 | 0.57 (0.24, 1.31) | 0.02 | 0.04 | 174 | 163 | 7.4 | 0.63 (0.24, 1.66) | 0.03 | 0.04 |
| Mali | PROMIS CS (46) | | | | | | | | | | | | |
| Zimbabwe | SHINE (HIV−) (47) | | | | | | | | | | | | |
| Zimbabwe | SHINE (HIV+) (48) | | | | | | | | | | | | |
| | | **7226** | **8131** | | **I² = 0.41, Tau² = 0.02** | | | **3423** | **2516** | | **I² = 0.20, Tau² = 0.01** | | |
| **Fixed** | | | | | **0.74 (0.66, 0.83)** | | | | | | **0.87 (0.74, 1.04)** | | |
| **Random** | | | | | **0.77 (0.65, 0.91)** | | | | | | **0.88 (0.73, 1.06)** | | |

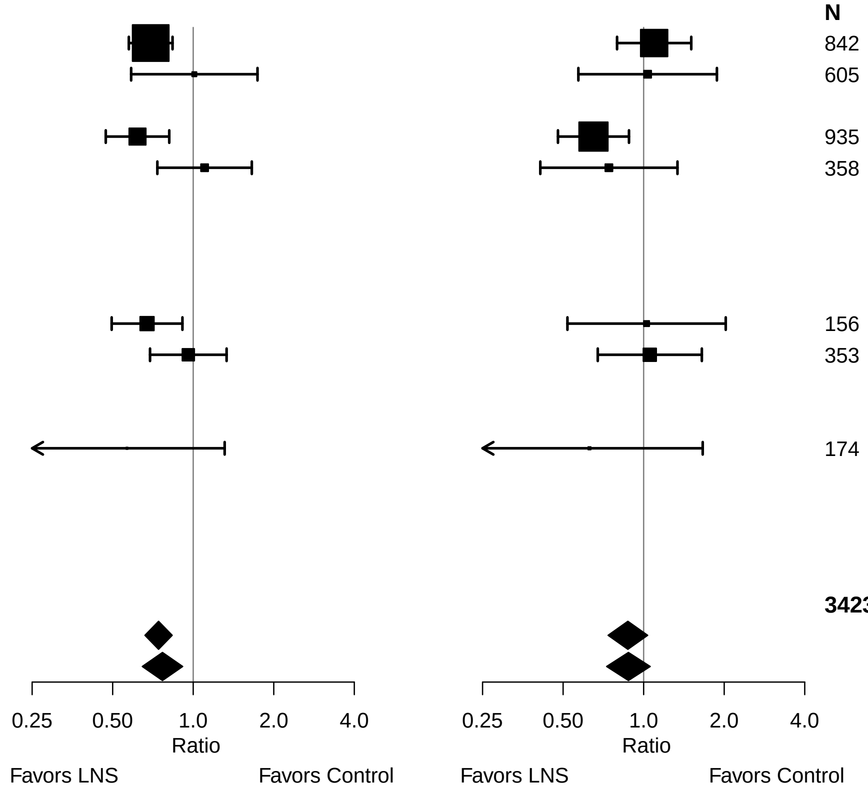

#### Supplemental figure 7H: Low MUAC prevalence ratio

#### 7H3: Stratified by Household source water quality

**P−for−interaction = 0.130**
**Ratio of PRs = 0.85 (0.69, 1.05)**

| Country | Trial | Improved LNS N | Improved Control N | Improved Control Prevalence | Improved PR (95% CI) | Improved Fixed W | Improved Random W | Unimproved LNS N | Unimproved Control N | Unimproved Control Prevalence | Unimproved PR (95% CI) | Unimproved Fixed W | Unimproved Random W |
|---|---|---|---|---|---|---|---|---|---|---|---|---|---|
| Bangladesh | JiVitA−4 (34) | | | | | | | | | | | | |
| Bangladesh | RDNS (35) | | | | | | | | | | | | |
| Bangladesh | WASH−B (36) | | | | | | | | | | | | |
| Burkina Faso | iLiNS−Zinc (37) | 555 | 141 | 14.2 | 0.66 (0.47, 0.91) | 0.24 | 0.21 | 1396 | 521 | 19.4 | 0.63 (0.50, 0.79) | 0.43 | 0.41 |
| Burkina Faso | PROMIS (38) | 352 | 376 | 7.7 | 1.14 (0.69, 1.88) | 0.10 | 0.12 | 358 | 355 | 9.3 | 0.72 (0.43, 1.21) | 0.08 | 0.09 |
| Burkina Faso | PROMIS CS (38) | 277 | 265 | 17.0 | 0.79 (0.52, 1.18) | 0.16 | 0.16 | 147 | 171 | 11.7 | 1.22 (0.63, 2.37) | 0.05 | 0.05 |
| Ghana | GHANA (39) | | | | | | | | | | | | |
| Ghana | iLiNS−DYADG (40) | | | | | | | | | | | | |
| Haiti | HAITI (41) | | | | | | | | | | | | |
| Kenya | WASH−B (42) | 449 | 1562 | 5.3 | 0.75 (0.44, 1.29) | 0.09 | 0.11 | 248 | 701 | 5.8 | 0.55 (0.24, 1.25) | 0.03 | 0.04 |
| Madagascar | MAHAY (43) | 415 | 452 | 7.7 | 1.34 (0.86, 2.08) | 0.13 | 0.15 | 1185 | 1157 | 10.1 | 0.83 (0.62, 1.12) | 0.25 | 0.25 |
| Malawi | iLiNS−DYADM (44) | | | | | | | | | | | | |
| Malawi | iLiNS−DOSE (45) | | | | | | | | | | | | |
| Mali | PROMIS (46) | 249 | 258 | 5.0 | 0.58 (0.22, 1.56) | 0.03 | 0.04 | 248 | 242 | 6.6 | 0.54 (0.25, 1.20) | 0.03 | 0.04 |
| Mali | PROMIS CS (46) | 549 | 567 | 10.4 | 0.96 (0.70, 1.33) | 0.25 | 0.22 | 364 | 388 | 11.6 | 0.88 (0.57, 1.34) | 0.12 | 0.13 |
| Zimbabwe | SHINE (HIV−) (47) | | | | | | | | | | | | |
| Zimbabwe | SHINE (HIV+) (48) | | | | | | | | | | | | |
| | | **2846** | **3621** | | **I² = 0.34, Tau² = 0.02** | | | **3946** | **3535** | | **I² = 0.06, Tau² = 0.00** | | |
| **Fixed** | | | | | **0.88 (0.74, 1.03)** | | | | | | **0.73 (0.63, 0.84)** | | |
| **Random** | | | | | **0.88 (0.72, 1.08)** | | | | | | **0.73 (0.63, 0.86)** | | |

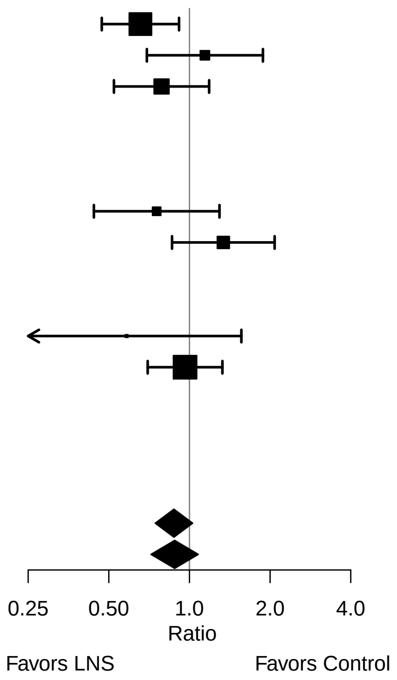

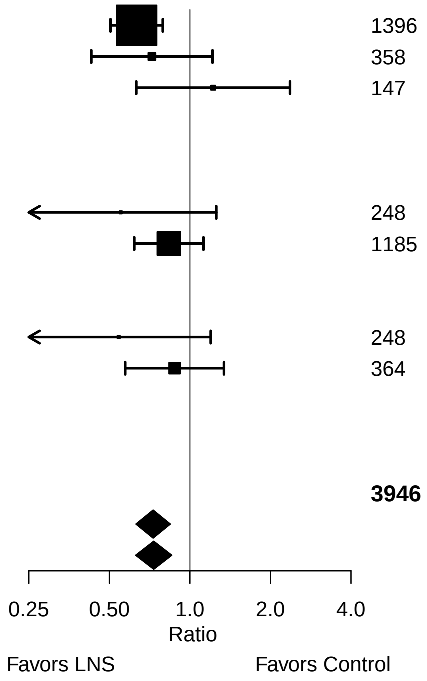

#### Supplemental figure 7H: Low MUAC prevalence ratio

##### 7H4: Stratified by Household sanitation

**P−for−interaction = 0.239**
**Ratio of PRs = 1.15 (0.91, 1.46)**

| Country | Trial | Improved LNS N | Improved Control N | Improved Control Prevalence | Improved PR (95% CI) | Improved Fixed W | Improved Random W | Unimproved LNS N | Unimproved Control N | Unimproved Control Prevalence | Unimproved PR (95% CI) | Unimproved Fixed W | Unimproved Random W |
|---|---|---|---|---|---|---|---|---|---|---|---|---|---|
| Bangladesh | JiVitA−4 (34) | 2241 | 974 | 13.6 | 0.83 (0.66, 1.05) | 0.41 | 0.41 | 655 | 297 | 20.1 | 0.91 (0.67, 1.25) | 0.37 | 0.28 |
| Bangladesh | RDNS (35) | 1179 | 573 | 5.8 | 0.72 (0.50, 1.05) | 0.16 | 0.16 | 482 | 241 | 3.7 | 2.11 (0.90, 4.94) | 0.05 | 0.07 |
| Bangladesh | WASH−B (36) | | | | | | | | | | | | |
| Burkina Faso | iLiNS−Zinc (37) | | | | | | | | | | | | |
| Burkina Faso | PROMIS (38) | 395 | 548 | 8.8 | 0.87 (0.54, 1.40) | 0.10 | 0.10 | 459 | 359 | 7.0 | 1.06 (0.65, 1.74) | 0.14 | 0.16 |
| Burkina Faso | PROMIS CS (38) | 175 | 163 | 16.0 | 0.90 (0.57, 1.41) | 0.11 | 0.11 | 248 | 273 | 14.3 | 0.93 (0.63, 1.38) | 0.23 | 0.22 |
| Ghana | GHANA (39) | | | | | | | | | | | | |
| Ghana | iLiNS−DYADG (40) | | | | | | | | | | | | |
| Haiti | HAITI (41) | | | | | | | | | | | | |
| Kenya | WASH−B (42) | 101 | 366 | 3.6 | 1.39 (0.54, 3.57) | 0.03 | 0.03 | 596 | 1898 | 5.8 | 0.60 (0.34, 1.06) | 0.11 | 0.13 |
| Madagascar | MAHAY (43) | | | | | | | | | | | | |
| Malawi | iLiNS−DYADM (44) | | | | | | | | | | | | |
| Malawi | iLiNS−DOSE (45) | | | | | | | | | | | | |
| Mali | PROMIS (46) | | | | | | | | | | | | |
| Mali | PROMIS CS (46) | 703 | 696 | 10.8 | 0.86 (0.61, 1.21) | 0.19 | 0.19 | 220 | 249 | 11.2 | 0.93 (0.52, 1.65) | 0.11 | 0.13 |
| Zimbabwe | SHINE (HIV−) (47) | | | | | | | | | | | | |
| Zimbabwe | SHINE (HIV+) (48) | | | | | | | | | | | | |
| | | **4794** | **3320** | | **I² = 0.00, Tau² = 0.00** | | | **2660** | **3317** | | **I² = 0.18, Tau² = 0.03** | | |
| **Fixed** | | | | | **0.84 (0.72, 0.97)** | | | | | | **0.94 (0.78, 1.13)** | | |
| **Random** | | | | | **0.84 (0.72, 0.97)** | | | | | | **0.95 (0.74, 1.20)** | | |

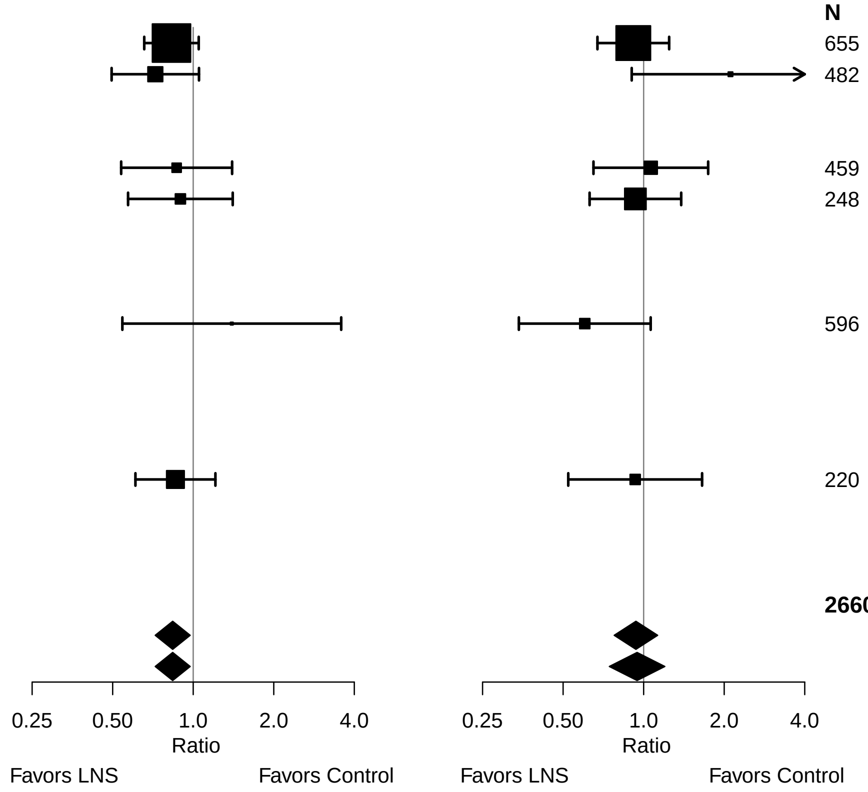

#### Supplemental figure 7H: Low MUAC prevalence ratio

#### 7H5: Stratified by Home environment

**P−for−interaction = 0.543**
**Ratio of PRs = 1.10 (0.81, 1.48)**

| | | At least median | | | | | | Less than median | | | | | |
|---|---|---|---|---|---|---|---|---|---|---|---|---|---|
| **Country** | **Trial** | **LNS N** | **Control N** | **Control Prevalence** | **PR (95% CI)** | **Fixed W** | **Random W** | **LNS N** | **Control N** | **Control Prevalence** | **PR (95% CI)** | **Fixed W** | **Random W** |
| Bangladesh | JiVitA−4 (34) | | | | | | | | | | | | |
| Bangladesh | RDNS (35) | 1192 | 582 | 4.1 | 1.06 (0.66, 1.70) | 0.16 | 0.21 | 468 | 230 | 7.8 | 0.96 (0.49, 1.87) | 0.11 | 0.13 |
| Bangladesh | WASH−B (36) | | | | | | | | | | | | |
| Burkina Faso | iLiNS−Zinc (37) | 473 | 199 | 19.1 | 0.51 (0.37, 0.72) | 0.31 | 0.27 | 272 | 175 | 16.6 | 0.77 (0.42, 1.41) | 0.14 | 0.16 |
| Burkina Faso | PROMIS (38) | | | | | | | | | | | | |
| Burkina Faso | PROMIS CS (38) | | | | | | | | | | | | |
| Ghana | GHANA (39) | | | | | | | | | | | | |
| Ghana | iLiNS−DYADG (40) | | | | | | | | | | | | |
| Haiti | HAITI (41) | | | | | | | | | | | | |
| Kenya | WASH−B (42) | 840 | 2782 | 4.3 | 0.89 (0.61, 1.31) | 0.24 | 0.25 | 573 | 2179 | 6.0 | 0.64 (0.41, 0.99) | 0.25 | 0.27 |
| Madagascar | MAHAY (43) | 905 | 889 | 8.8 | 0.86 (0.61, 1.21) | 0.30 | 0.27 | 775 | 769 | 10.7 | 1.09 (0.80, 1.49) | 0.50 | 0.44 |
| Malawi | iLiNS−DYADM (44) | | | | | | | | | | | | |
| Malawi | iLiNS−DOSE (45) | | | | | | | | | | | | |
| Mali | PROMIS (46) | | | | | | | | | | | | |
| Mali | PROMIS CS (46) | | | | | | | | | | | | |
| Zimbabwe | SHINE (HIV−) (47) | | | | | | | | | | | | |
| Zimbabwe | SHINE (HIV+) (48) | | | | | | | | | | | | |
| | | **3410** | **4452** | | **I² = 0.64, Tau² = 0.06** | | | **2088** | **3353** | | **I² = 0.25, Tau² = 0.01** | | |
| **Fixed** | | | | | **0.76 (0.63, 0.92)** | | | | | | **0.89 (0.72, 1.12)** | | |
| **Random** | | | | | **0.79 (0.58, 1.07)** | | | | | | **0.88 (0.68, 1.13)** | | |

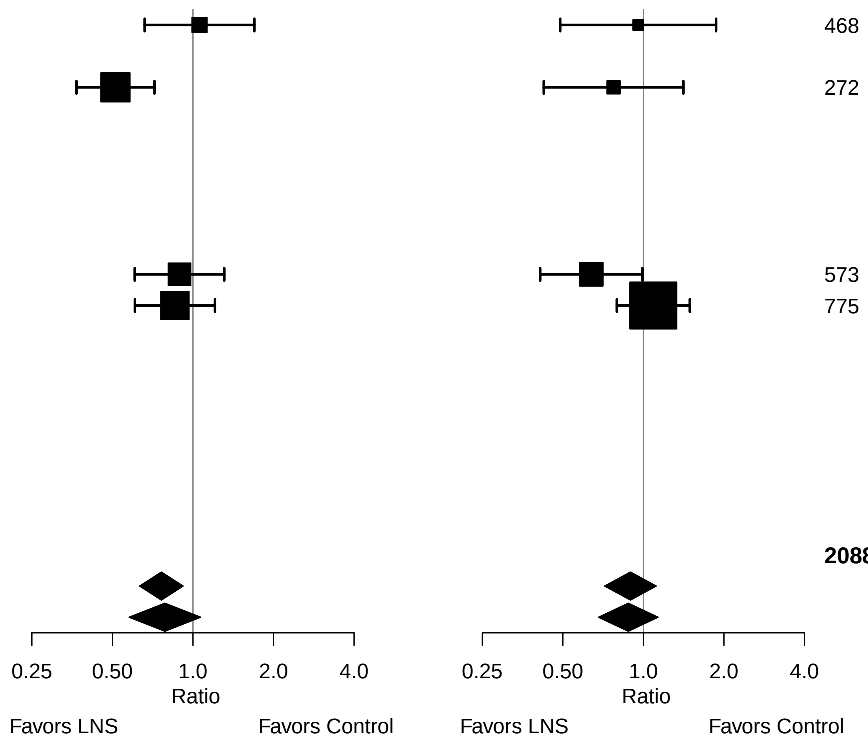

#### Supplemental figure 7H: Low MUAC prevalence ratio

### 7H6: Stratified by Season at the time of assessment

**P−for−interaction = 0.301**
**Ratio of PRs = 0.89 (0.72, 1.11)**

| Country | Trial | Dry LNS N | Dry Control N | Dry Control Prevalence | Dry PR (95% CI) | Dry Fixed W | Dry Random W | Rainy LNS N | Rainy Control N | Rainy Control Prevalence | Rainy PR (95% CI) | Rainy Fixed W | Rainy Random W |
|---|---|---|---|---|---|---|---|---|---|---|---|---|---|
| Bangladesh | JiVitA−4 (34) | 1570 | 673 | 14.8 | 0.91 (0.69, 1.20) | 0.30 | 0.25 | 1327 | 598 | 15.7 | 0.81 (0.63, 1.04) | 0.30 | 0.30 |
| Bangladesh | RDNS (35) | 739 | 359 | 2.5 | 1.30 (0.63, 2.67) | 0.04 | 0.07 | 924 | 456 | 7.2 | 0.94 (0.63, 1.41) | 0.12 | 0.12 |
| Bangladesh | WASH−B (36) | | | | | | | | | | | | |
| Burkina Faso | iLiNS−Zinc (37) | 1399 | 477 | 19.3 | 0.61 (0.47, 0.79) | 0.35 | 0.27 | 560 | 189 | 15.9 | 0.71 (0.48, 1.05) | 0.13 | 0.13 |
| Burkina Faso | PROMIS (38) | 425 | 444 | 9.2 | 1.02 (0.67, 1.55) | 0.13 | 0.16 | 429 | 468 | 6.8 | 0.82 (0.45, 1.48) | 0.06 | 0.06 |
| Burkina Faso | PROMIS CS (38) | 222 | 190 | 14.2 | 1.08 (0.63, 1.85) | 0.08 | 0.11 | 202 | 246 | 15.4 | 0.77 (0.57, 1.03) | 0.23 | 0.23 |
| Ghana | GHANA (39) | | | | | | | | | | | | |
| Ghana | iLiNS−DYADG (40) | | | | | | | | | | | | |
| Haiti | HAITI (41) | | | | | | | | | | | | |
| Kenya | WASH−B (42) | 349 | 1163 | 5.7 | 0.76 (0.47, 1.23) | 0.10 | 0.13 | 1112 | 4022 | 5.4 | 0.69 (0.49, 0.98) | 0.17 | 0.17 |
| Madagascar | MAHAY (43) | | | | | | | | | | | | |
| Malawi | iLiNS−DYADM (44) | | | | | | | | | | | | |
| Malawi | iLiNS−DOSE (45) | | | | | | | | | | | | |
| Mali | PROMIS (46) | | | | | | | | | | | | |
| Mali | PROMIS CS (46) | | | | | | | | | | | | |
| Zimbabwe | SHINE (HIV−) (47) | | | | | | | | | | | | |
| Zimbabwe | SHINE (HIV+) (48) | | | | | | | | | | | | |
| | | **4704** | **3306** | | **I² = 0.45, Tau² = 0.02** | | | **4554** | **5979** | | **I² = 0.00, Tau² = 0.00** | | |
| **Fixed** | | | | | **0.81 (0.70, 0.95)** | | | | | | **0.78 (0.68, 0.90)** | | |
| **Random** | | | | | **0.85 (0.69, 1.04)** | | | | | | **0.78 (0.68, 0.90)** | | |

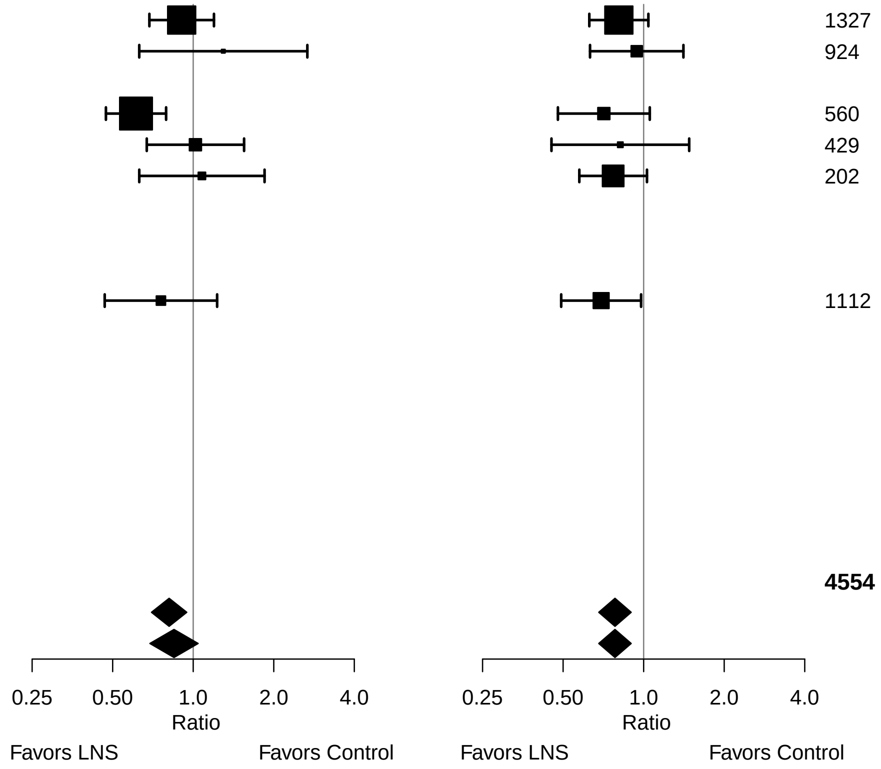

### Supplemental figure 7I: Low MUAC prevalence difference

#### 7I1: Stratified by Household socio-economic status

**P−for−interaction = 0.735**
**Difference in PDs = 0.00 (−0.02, 0.01)**

| Country | Trial | At least median: LNS N | Control N | Control Prevalence | PD (95% CI) | Fixed W | Random W | Less than median: LNS N | Control N | Control Prevalence | PD (95% CI) | Fixed W | Random W |
|---|---|---|---|---|---|---|---|---|---|---|---|---|---|
| Bangladesh | JiVitA−4 (34) | 1463 | 641 | 11.6 | −0.01 (−0.04, 0.01) | 0.11 | 0.11 | 1433 | 630 | 18.8 | −0.03 (−0.06, 0.00) | 0.12 | 0.12 |
| Bangladesh | RDNS (35) | 834 | 407 | 4.4 | −0.01 (−0.03, 0.01) | 0.16 | 0.16 | 829 | 408 | 5.9 | 0.01 (−0.02, 0.04) | 0.14 | 0.13 |
| Bangladesh | WASH−B (36) | | | | | | | | | | | | |
| Burkina Faso | iLiNS−Zinc (37) | 1232 | 300 | 16.0 | −0.06 (−0.10, −0.02) | 0.05 | 0.05 | 720 | 365 | 20.0 | −0.07 (−0.11, −0.03) | 0.08 | 0.10 |
| Burkina Faso | PROMIS (38) | 403 | 479 | 9.4 | −0.02 (−0.07, 0.03) | 0.03 | 0.03 | 451 | 433 | 6.5 | 0.01 (−0.02, 0.05) | 0.11 | 0.11 |
| Burkina Faso | PROMIS CS (38) | 219 | 207 | 14.5 | −0.03 (−0.09, 0.04) | 0.02 | 0.02 | 205 | 229 | 15.3 | 0.00 (−0.06, 0.06) | 0.04 | 0.06 |
| Ghana | GHANA (39) | | | | | | | | | | | | |
| Ghana | iLiNS−DYADG (40) | | | | | | | | | | | | |
| Haiti | HAITI (41) | | | | | | | | | | | | |
| Kenya | WASH−B (42) | 827 | 2949 | 3.7 | −0.01 (−0.02, 0.00) | 0.46 | 0.46 | 632 | 2232 | 7.8 | −0.02 (−0.05, 0.00) | 0.24 | 0.15 |
| Madagascar | MAHAY (43) | 761 | 865 | 8.1 | 0.00 (−0.03, 0.03) | 0.07 | 0.07 | 866 | 750 | 11.2 | −0.01 (−0.05, 0.02) | 0.11 | 0.12 |
| Malawi | iLiNS−DYADM (44) | | | | | | | | | | | | |
| Malawi | iLiNS−DOSE (45) | | | | | | | | | | | | |
| Mali | PROMIS (46) | 248 | 250 | 4.8 | −0.01 (−0.05, 0.03) | 0.04 | 0.04 | 246 | 246 | 6.9 | −0.04 (−0.08, 0.00) | 0.08 | 0.10 |
| Mali | PROMIS CS (46) | 505 | 463 | 12.7 | −0.03 (−0.08, 0.01) | 0.04 | 0.04 | 449 | 507 | 9.5 | 0.01 (−0.03, 0.05) | 0.09 | 0.10 |
| Zimbabwe | SHINE (HIV−) (47) | | | | | | | | | | | | |
| Zimbabwe | SHINE (HIV+) (48) | | | | | | | | | | | | |
| | | **6492** | **6561** | | **I² = 0.03, Tau² = 0.00** | | | **5831** | **5800** | | **I² = 0.55, Tau² = 0.00** | | |
| **Fixed** | | | | | **−0.01 (−0.02, −0.01)** | | | | | | **−0.02 (−0.03, 0.00)** | | |
| **Random** | | | | | **−0.01 (−0.02, −0.01)** | | | | | | **−0.02 (−0.03, 0.00)** | | |

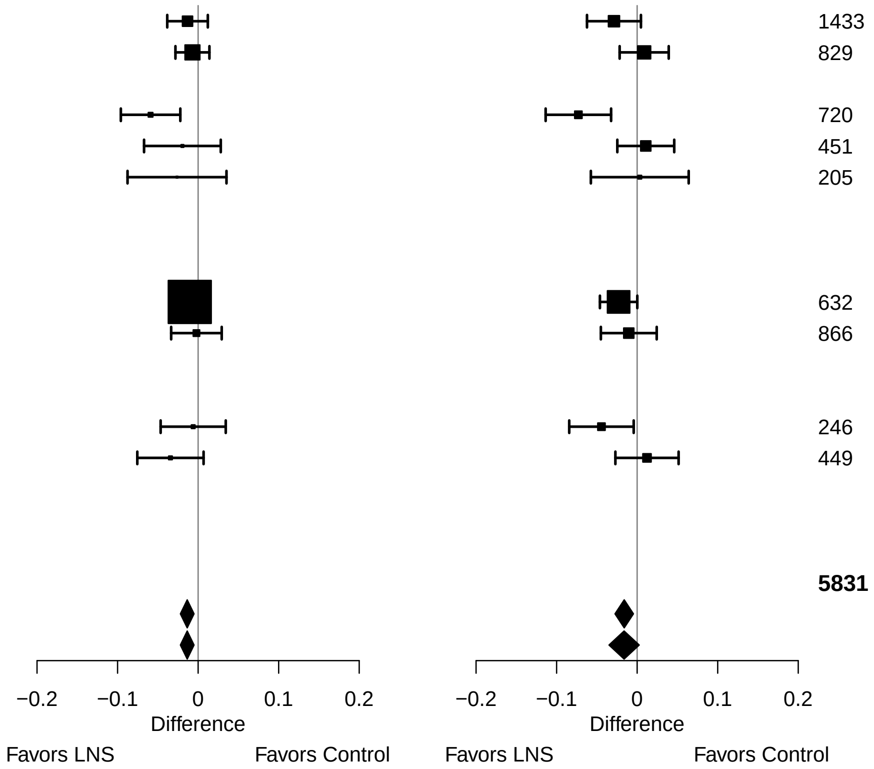

#### Supplemental figure 7I: Low MUAC prevalence difference

#### 7I2: Stratified by Household food insecurity

**P−for−interaction = 0.630**
**Difference in PDs = 0.00 (−0.01, 0.02)**

| Country | Trial | LNS N | Control N | Control Prevalence | Mild to secure PD (95% CI) | Fixed W | Random W | LNS N | Control N | Control Prevalence | Moderate to severe PD (95% CI) | Fixed W | Random W |
|---|---|---|---|---|---|---|---|---|---|---|---|---|---|
| Bangladesh | JiVitA−4 (34) | 2054 | 882 | 14.6 | −0.03 (−0.06, −0.01) | 0.12 | 0.16 | 842 | 389 | 16.3 | 0.01 (−0.03, 0.05) | 0.15 | 0.15 |
| Bangladesh | RDNS (35) | 1058 | 493 | 4.9 | 0.00 (−0.03, 0.03) | 0.10 | 0.15 | 605 | 322 | 5.6 | 0.00 (−0.03, 0.04) | 0.22 | 0.18 |
| Bangladesh | WASH−B (36) | | | | | | | | | | | | |
| Burkina Faso | iLiNS−Zinc (37) | 1017 | 321 | 16.2 | −0.06 (−0.10, −0.02) | 0.04 | 0.08 | 935 | 344 | 20.1 | −0.07 (−0.12, −0.03) | 0.12 | 0.13 |
| Burkina Faso | PROMIS (38) | 496 | 547 | 7.3 | 0.01 (−0.02, 0.04) | 0.07 | 0.12 | 358 | 365 | 9.0 | −0.02 (−0.07, 0.02) | 0.12 | 0.13 |
| Burkina Faso | PROMIS CS (38) | | | | | | | | | | | | |
| Ghana | GHANA (39) | | | | | | | | | | | | |
| Ghana | iLiNS−DYADG (40) | | | | | | | | | | | | |
| Haiti | HAITI (41) | | | | | | | | | | | | |
| Kenya | WASH−B (42) | 1303 | 4640 | 5.4 | −0.02 (−0.03, −0.01) | 0.51 | 0.24 | 156 | 528 | 6.2 | 0.00 (−0.04, 0.05) | 0.14 | 0.14 |
| Madagascar | MAHAY (43) | 968 | 907 | 9.5 | 0.00 (−0.03, 0.03) | 0.08 | 0.13 | 353 | 405 | 9.1 | 0.00 (−0.04, 0.05) | 0.14 | 0.15 |
| Malawi | iLiNS−DYADM (44) | | | | | | | | | | | | |
| Malawi | iLiNS−DOSE (45) | | | | | | | | | | | | |
| Mali | PROMIS (46) | 330 | 341 | 5.0 | −0.02 (−0.05, 0.01) | 0.08 | 0.13 | 174 | 163 | 7.4 | −0.03 (−0.08, 0.02) | 0.11 | 0.12 |
| Mali | PROMIS CS (46) | | | | | | | | | | | | |
| Zimbabwe | SHINE (HIV−) (47) | | | | | | | | | | | | |
| Zimbabwe | SHINE (HIV+) (48) | | | | | | | | | | | | |
| | | **7226** | **8131** | | **I² = 0.43, Tau² = 0.00** | | | **3423** | **2516** | | **I² = 0.40, Tau² = 0.00** | | |
| **Fixed** | | | | | **−0.02 (−0.03, −0.01)** | | | | | | **−0.01 (−0.03, 0.00)** | | |
| **Random** | | | | | **−0.02 (−0.03, 0.00)** | | | | | | **−0.01 (−0.04, 0.01)** | | |

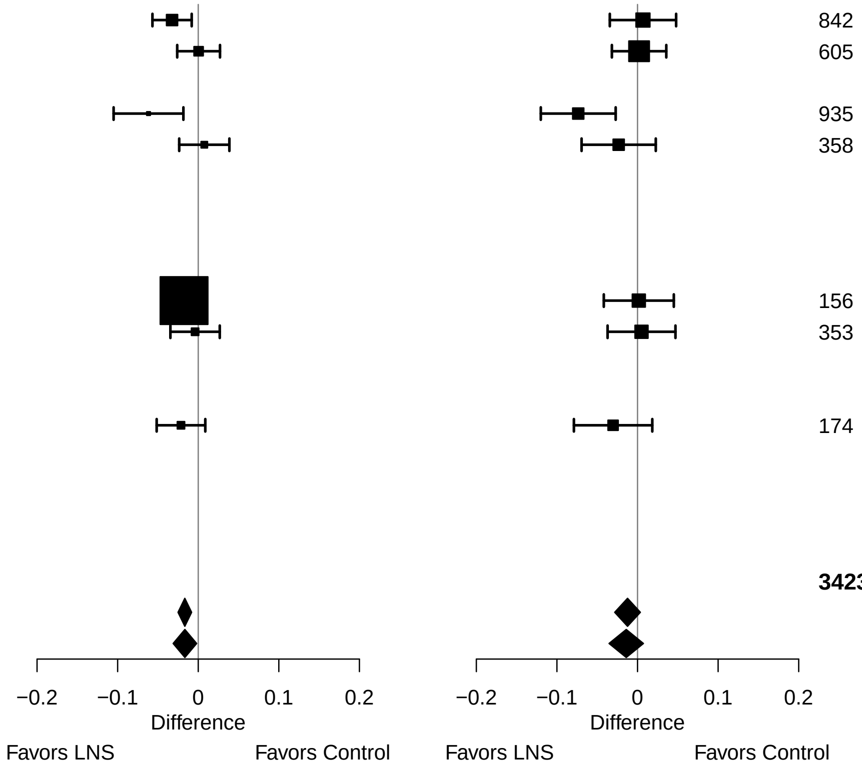

**Supplemental figure 7I: Low MUAC prevalence difference**

#### 7I3: Stratified by Household source water quality

**P−for−interaction = 0.039**
**Difference in PDs = −0.02 (−0.04, 0.00)**

| Country | Trial | Improved LNS N | Improved Control N | Improved Control Prevalence | Improved PD (95% CI) | Improved Fixed W | Improved Random W | Unimproved LNS N | Unimproved Control N | Unimproved Control Prevalence | Unimproved PD (95% CI) | Unimproved Fixed W | Unimproved Random W |
|---|---|---|---|---|---|---|---|---|---|---|---|---|---|
| Bangladesh | JiVitA−4 (34) | | | | | | | | | | | | |
| Bangladesh | RDNS (35) | | | | | | | | | | | | |
| Bangladesh | WASH−B (36) | | | | | | | | | | | | |
| Burkina Faso | iLiNS−Zinc (37) | 555 | 141 | 14.2 | −0.03 (−0.07, 0.00) | 0.12 | 0.13 | 1396 | 521 | 19.4 | −0.08 (−0.11, −0.05) | 0.18 | 0.17 |
| Burkina Faso | PROMIS (38) | 352 | 376 | 7.7 | 0.01 (−0.03, 0.05) | 0.10 | 0.12 | 358 | 355 | 9.3 | −0.03 (−0.07, 0.01) | 0.11 | 0.13 |
| Burkina Faso | PROMIS CS (38) | 277 | 265 | 17.0 | −0.04 (−0.10, 0.02) | 0.05 | 0.06 | 147 | 171 | 11.7 | 0.03 (−0.06, 0.11) | 0.03 | 0.05 |
| Ghana | GHANA (39) | | | | | | | | | | | | |
| Ghana | iLiNS−DYADG (40) | | | | | | | | | | | | |
| Haiti | HAITI (41) | | | | | | | | | | | | |
| Kenya | WASH−B (42) | 449 | 1562 | 5.3 | −0.01 (−0.04, 0.01) | 0.33 | 0.26 | 248 | 701 | 5.8 | −0.03 (−0.06, 0.01) | 0.19 | 0.17 |
| Madagascar | MAHAY (43) | 415 | 452 | 7.7 | 0.03 (−0.01, 0.06) | 0.12 | 0.13 | 1185 | 1157 | 10.1 | −0.02 (−0.04, 0.01) | 0.24 | 0.19 |
| Malawi | iLiNS−DYADM (44) | | | | | | | | | | | | |
| Malawi | iLiNS−DOSE (45) | | | | | | | | | | | | |
| Mali | PROMIS (46) | 249 | 258 | 5.0 | −0.02 (−0.06, 0.02) | 0.11 | 0.13 | 248 | 242 | 6.6 | −0.03 (−0.07, 0.00) | 0.16 | 0.16 |
| Mali | PROMIS CS (46) | 549 | 567 | 10.4 | 0.00 (−0.04, 0.03) | 0.16 | 0.17 | 364 | 388 | 11.6 | −0.01 (−0.06, 0.03) | 0.09 | 0.12 |
| Zimbabwe | SHINE (HIV−) (47) | | | | | | | | | | | | |
| Zimbabwe | SHINE (HIV+) (48) | | | | | | | | | | | | |
| | | **2846** | **3621** | | **I² = 0.18, Tau² = 0.00** | | | **3946** | **3535** | | **I² = 0.50, Tau² = 0.00** | | |
| **Fixed** | | | | | **−0.01 (−0.02, 0.00)** | | | | | | **−0.03 (−0.05, −0.02)** | | |
| **Random** | | | | | **−0.01 (−0.02, 0.01)** | | | | | | **−0.03 (−0.05, −0.01)** | | |

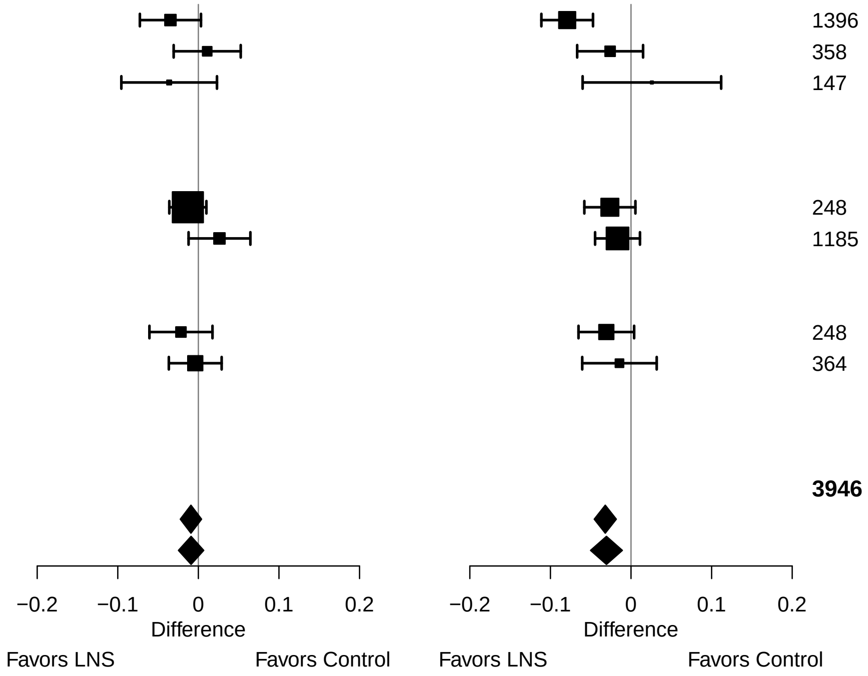

#### Supplemental figure 7I: Low MUAC prevalence difference

#### 7I4: Stratified by Household sanitation

**P−for−interaction = 0.165**
**Difference in PDs = 0.02 (−0.01, 0.04)**

| Country | Trial | Improved LNS N | Improved Control N | Improved Control Prevalence | Improved PD (95% CI) | Improved Fixed W | Improved Random W | Unimproved LNS N | Unimproved Control N | Unimproved Control Prevalence | Unimproved PD (95% CI) | Unimproved Fixed W | Unimproved Random W |
|---|---|---|---|---|---|---|---|---|---|---|---|---|---|
| Bangladesh | JiVitA−4 (34) | 2241 | 974 | 13.6 | −0.02 (−0.05, 0.00) | 0.27 | 0.27 | 655 | 297 | 20.1 | −0.01 (−0.06, 0.04) | 0.10 | 0.12 |
| Bangladesh | RDNS (35) | 1179 | 573 | 5.8 | −0.02 (−0.03, 0.00) | 0.41 | 0.41 | 482 | 241 | 3.7 | 0.04 (0.00, 0.08) | 0.15 | 0.17 |
| Bangladesh | WASH−B (36) | | | | | | | | | | | | |
| Burkina Faso | iLiNS−Zinc (37) | | | | | | | | | | | | |
| Burkina Faso | PROMIS (38) | 395 | 548 | 8.8 | −0.01 (−0.05, 0.03) | 0.10 | 0.10 | 459 | 359 | 7.0 | 0.00 (−0.03, 0.04) | 0.18 | 0.20 |
| Burkina Faso | PROMIS CS (38) | 175 | 163 | 16.0 | −0.02 (−0.08, 0.05) | 0.03 | 0.03 | 248 | 273 | 14.3 | −0.01 (−0.06, 0.04) | 0.08 | 0.10 |
| Ghana | GHANA (39) | | | | | | | | | | | | |
| Ghana | iLiNS−DYADG (40) | | | | | | | | | | | | |
| Haiti | HAITI (41) | | | | | | | | | | | | |
| Kenya | WASH−B (42) | 101 | 366 | 3.6 | 0.01 (−0.03, 0.06) | 0.08 | 0.08 | 596 | 1898 | 5.8 | −0.02 (−0.05, 0.00) | 0.44 | 0.32 |
| Madagascar | MAHAY (43) | | | | | | | | | | | | |
| Malawi | iLiNS−DYADM (44) | | | | | | | | | | | | |
| Malawi | iLiNS−DOSE (45) | | | | | | | | | | | | |
| Mali | PROMIS (46) | | | | | | | | | | | | |
| Mali | PROMIS CS (46) | 703 | 696 | 10.8 | −0.02 (−0.05, 0.02) | 0.12 | 0.12 | 220 | 249 | 11.2 | −0.01 (−0.07, 0.06) | 0.06 | 0.08 |
| Zimbabwe | SHINE (HIV−) (47) | | | | | | | | | | | | |
| Zimbabwe | SHINE (HIV+) (48) | | | | | | | | | | | | |
| | | **4794** | **3320** | | **I² = 0.00, Tau² = 0.00** | | | **2660** | **3317** | | **I² = 0.42, Tau² = 0.00** | | |
| **Fixed** | | | | | **−0.02 (−0.03, 0.00)** | | | | | | **−0.01 (−0.02, 0.01)** | | |
| **Random** | | | | | **−0.02 (−0.03, 0.00)** | | | | | | **0.00 (−0.02, 0.02)** | | |

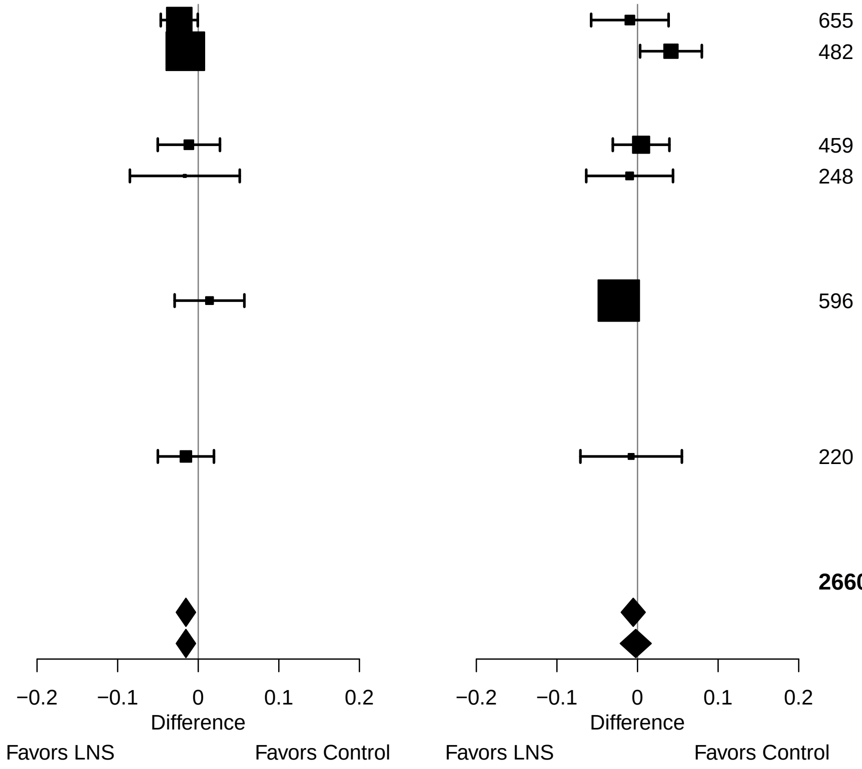

#### Supplemental figure 7I: Low MUAC prevalence difference

#### 7I5: Stratified by Home environment

**P−for−interaction = 0.688**
**Difference in PDs = 0.00 (−0.02, 0.01)**

| Country | Trial | LNS N | Control N | Control Prevalence | At least median PD (95% CI) | Fixed W | Random W | LNS N | Control N | Control Prevalence | Less than median PD (95% CI) | Fixed W | Random W |
|---|---|---|---|---|---|---|---|---|---|---|---|---|---|
| Bangladesh | JiVitA−4 (34) | | | | | | | | | | | | |
| Bangladesh | RDNS (35) | 1192 | 582 | 4.1 | 0.00 (−0.02, 0.02) | 0.29 | 0.26 | 468 | 230 | 7.8 | 0.00 (−0.06, 0.05) | 0.08 | 0.08 |
| Bangladesh | WASH−B (36) | | | | | | | | | | | | |
| Burkina Faso | iLiNS−Zinc (37) | 473 | 199 | 19.1 | −0.09 (−0.13, −0.05) | 0.07 | 0.22 | 272 | 175 | 16.6 | −0.04 (−0.13, 0.05) | 0.03 | 0.03 |
| Burkina Faso | PROMIS (38) | | | | | | | | | | | | |
| Burkina Faso | PROMIS CS (38) | | | | | | | | | | | | |
| Ghana | GHANA (39) | | | | | | | | | | | | |
| Ghana | iLiNS−DYADG (40) | | | | | | | | | | | | |
| Haiti | HAITI (41) | | | | | | | | | | | | |
| Kenya | WASH−B (42) | 840 | 2782 | 4.3 | 0.00 (−0.02, 0.01) | 0.51 | 0.27 | 573 | 2179 | 6.0 | −0.02 (−0.04, 0.00) | 0.70 | 0.70 |
| Madagascar | MAHAY (43) | 905 | 889 | 8.8 | −0.01 (−0.04, 0.02) | 0.14 | 0.25 | 775 | 769 | 10.7 | 0.01 (−0.03, 0.04) | 0.19 | 0.19 |
| Malawi | iLiNS−DYADM (44) | | | | | | | | | | | | |
| Malawi | iLiNS−DOSE (45) | | | | | | | | | | | | |
| Mali | PROMIS (46) | | | | | | | | | | | | |
| Mali | PROMIS CS (46) | | | | | | | | | | | | |
| Zimbabwe | SHINE (HIV−) (47) | | | | | | | | | | | | |
| Zimbabwe | SHINE (HIV+) (48) | | | | | | | | | | | | |
| | | **3410** | **4452** | | **I² = 0.82, Tau² = 0.00** | | | **2088** | **3353** | | **I² = 0.00, Tau² = 0.00** | | |
| **Fixed** | | | | | **−0.01 (−0.02, 0.00)** | | | | | | **−0.01 (−0.03, 0.00)** | | |
| **Random** | | | | | **−0.02 (−0.06, 0.02)** | | | | | | **−0.01 (−0.03, 0.00)** | | |

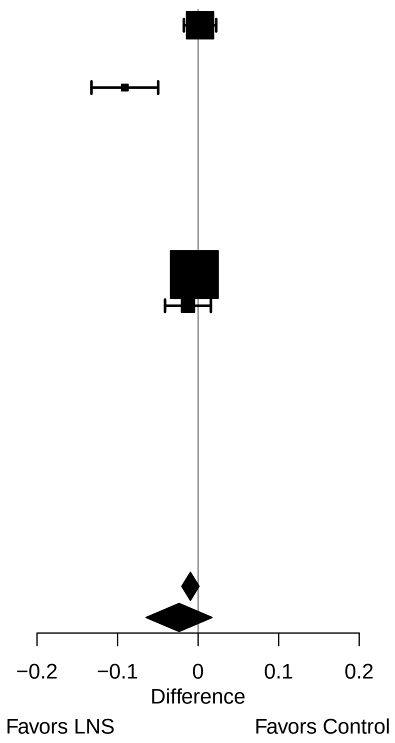

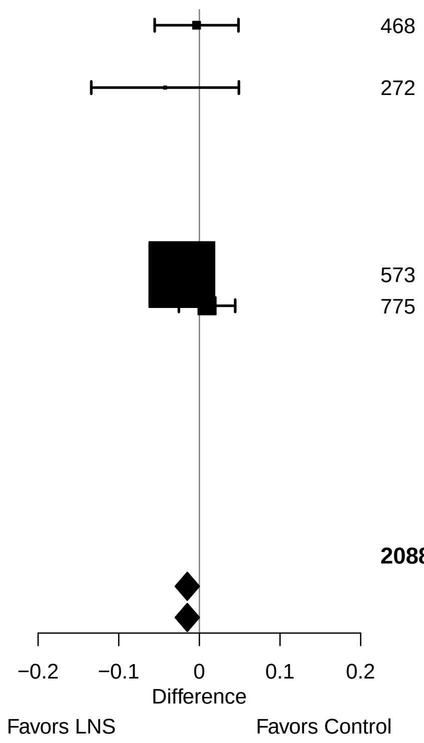

#### Supplemental figure 7I: Low MUAC prevalence difference

#### 7I6: Stratified by Season at the time of assessment

**P−for−interaction = 0.322**
**Difference in PDs = −0.01 (−0.03, 0.01)**

| Country | Trial | Dry LNS N | Dry Control N | Dry Control Prevalence | Dry PD (95% CI) | Dry Fixed W | Dry Random W | Rainy LNS N | Rainy Control N | Rainy Control Prevalence | Rainy PD (95% CI) | Rainy Fixed W | Rainy Random W |
|---|---|---|---|---|---|---|---|---|---|---|---|---|---|
| Bangladesh | JiVitA−4 (34) | 1570 | 673 | 14.8 | −0.01 (−0.04, 0.01) | 0.17 | 0.19 | 1327 | 598 | 15.7 | −0.03 (−0.06, 0.00) | 0.11 | 0.11 |
| Bangladesh | RDNS (35) | 739 | 359 | 2.5 | 0.01 (−0.01, 0.03) | 0.35 | 0.22 | 924 | 456 | 7.2 | 0.00 (−0.03, 0.02) | 0.13 | 0.13 |
| Bangladesh | WASH−B (36) | | | | | | | | | | | | |
| Burkina Faso | iLiNS−Zinc (37) | 1399 | 477 | 19.3 | −0.07 (−0.11, −0.04) | 0.11 | 0.16 | 560 | 189 | 15.9 | −0.06 (−0.11, −0.01) | 0.04 | 0.04 |
| Burkina Faso | PROMIS (38) | 425 | 444 | 9.2 | 0.00 (−0.04, 0.04) | 0.09 | 0.15 | 429 | 468 | 6.8 | −0.01 (−0.05, 0.02) | 0.08 | 0.08 |
| Burkina Faso | PROMIS CS (38) | 222 | 190 | 14.2 | 0.01 (−0.07, 0.09) | 0.02 | 0.07 | 202 | 246 | 15.4 | −0.04 (−0.08, 0.01) | 0.06 | 0.06 |
| Ghana | GHANA (39) | | | | | | | | | | | | |
| Ghana | iLiNS−DYADG (40) | | | | | | | | | | | | |
| Haiti | HAITI (41) | | | | | | | | | | | | |
| Kenya | WASH−B (42) | 349 | 1163 | 5.7 | −0.01 (−0.04, 0.01) | 0.27 | 0.21 | 1112 | 4022 | 5.4 | −0.02 (−0.03, 0.00) | 0.58 | 0.58 |
| Madagascar | MAHAY (43) | | | | | | | | | | | | |
| Malawi | iLiNS−DYADM (44) | | | | | | | | | | | | |
| Malawi | iLiNS−DOSE (45) | | | | | | | | | | | | |
| Mali | PROMIS (46) | | | | | | | | | | | | |
| Mali | PROMIS CS (46) | | | | | | | | | | | | |
| Zimbabwe | SHINE (HIV−) (47) | | | | | | | | | | | | |
| Zimbabwe | SHINE (HIV+) (48) | | | | | | | | | | | | |
| | | **4704** | **3306** | | **I² = 0.67, Tau² = 0.00** | | | **4554** | **5979** | | **I² = 0.00, Tau² = 0.00** | | |
| **Fixed** | | | | | **−0.01 (−0.02, 0.00)** | | | | | | **−0.02 (−0.03, −0.01)** | | |
| **Random** | | | | | **−0.01 (−0.04, 0.01)** | | | | | | **−0.02 (−0.03, −0.01)** | | |

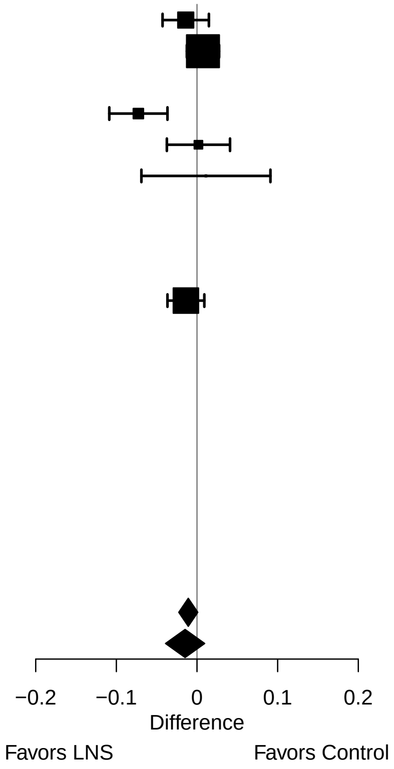

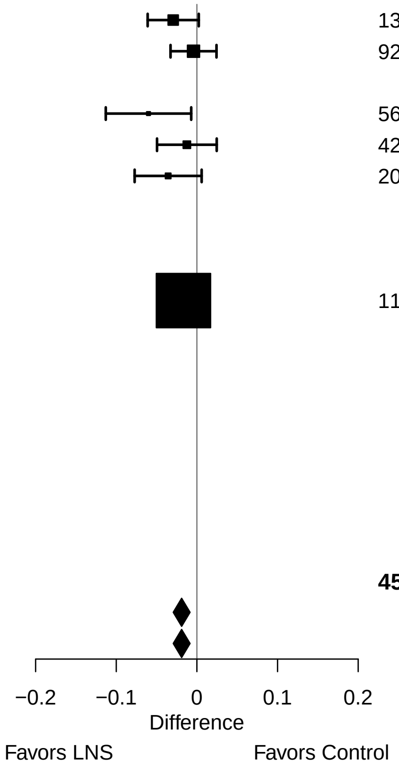

### Supplemental figure 7J: Acute malnutrition prevalence ratio

#### 7J1: Stratified by Household socio-economic status

**P−for−interaction = 0.312**
**Ratio of PRs = 1.08 (0.93, 1.25)**

| | | At least median | | | | | | | Less than median | | | | |
|---|---|---|---|---|---|---|---|---|---|---|---|---|---|
| **Country** | **Trial** | **LNS N** | **Control N** | **Control Prevalence** | **PR (95% CI)** | **Fixed W** | **Random W** | **LNS N** | **Control N** | **Control Prevalence** | **PR (95% CI)** | **Fixed W** | **Random W** |
| Bangladesh | JiVitA−4 (34) | 1401 | 606 | 17.7 | 0.82 (0.68, 0.98) | 0.34 | 0.34 | 1381 | 610 | 24.4 | 0.94 (0.79, 1.12) | 0.34 | 0.34 |
| Bangladesh | RDNS (35) | 834 | 407 | 13.5 | 0.86 (0.62, 1.19) | 0.11 | 0.11 | 827 | 408 | 17.2 | 0.86 (0.66, 1.12) | 0.14 | 0.14 |
| Bangladesh | WASH−B (36) | | | | | | | | | | | | |
| Burkina Faso | iLiNS−Zinc (37) | 1224 | 299 | 18.4 | 0.58 (0.42, 0.79) | 0.12 | 0.12 | 720 | 364 | 19.7 | 0.81 (0.60, 1.09) | 0.11 | 0.11 |
| Burkina Faso | PROMIS (38) | 401 | 473 | 11.6 | 0.81 (0.53, 1.26) | 0.06 | 0.06 | 445 | 428 | 11.7 | 0.85 (0.53, 1.34) | 0.05 | 0.05 |
| Burkina Faso | PROMIS CS (38) | 219 | 206 | 19.4 | 0.80 (0.53, 1.20) | 0.07 | 0.07 | 205 | 228 | 19.3 | 1.11 (0.80, 1.55) | 0.09 | 0.09 |
| Ghana | GHANA (39) | | | | | | | | | | | | |
| Ghana | iLiNS−DYADG (40) | 157 | 356 | 7.6 | 1.01 (0.52, 1.94) | 0.03 | 0.03 | 190 | 334 | 8.7 | 0.73 (0.38, 1.39) | 0.02 | 0.02 |
| Haiti | HAITI (41) | | | | | | | | | | | | |
| Kenya | WASH−B (42) | 824 | 2914 | 2.3 | 0.75 (0.44, 1.27) | 0.04 | 0.04 | 628 | 2199 | 4.4 | 0.69 (0.44, 1.07) | 0.05 | 0.05 |
| Madagascar | MAHAY (43) | 760 | 865 | 7.5 | 1.09 (0.74, 1.60) | 0.08 | 0.08 | 862 | 747 | 11.5 | 0.88 (0.60, 1.28) | 0.07 | 0.07 |
| Malawi | iLiNS−DYADM (44) | | | | | | | | | | | | |
| Malawi | iLiNS−DOSE (45) | | | | | | | | | | | | |
| Mali | PROMIS (46) | | | | | | | | | | | | |
| Mali | PROMIS CS (46) | 501 | 458 | 13.5 | 0.81 (0.57, 1.16) | 0.09 | 0.09 | 443 | 501 | 12.6 | 0.97 (0.70, 1.34) | 0.09 | 0.09 |
| Zimbabwe | SHINE (HIV−) (47) | 901 | 795 | 2.9 | 0.96 (0.57, 1.62) | 0.04 | 0.04 | 826 | 851 | 4.1 | 0.62 (0.36, 1.07) | 0.03 | 0.03 |
| Zimbabwe | SHINE (HIV+) (48) | 171 | 156 | 3.8 | 1.52 (0.53, 4.33) | 0.01 | 0.01 | 157 | 165 | 5.5 | 0.93 (0.36, 2.42) | 0.01 | 0.01 |
| | | **7393** | **7535** | | **I² = 0.00, Tau² = 0.00** | | | **6684** | **6835** | | **I² = 0.00, Tau² = 0.00** | | |
| **Fixed** | | | | | **0.82 (0.73, 0.91)** | | | | | | **0.89 (0.81, 0.98)** | | |
| **Random** | | | | | **0.82 (0.73, 0.91)** | | | | | | **0.89 (0.81, 0.98)** | | |

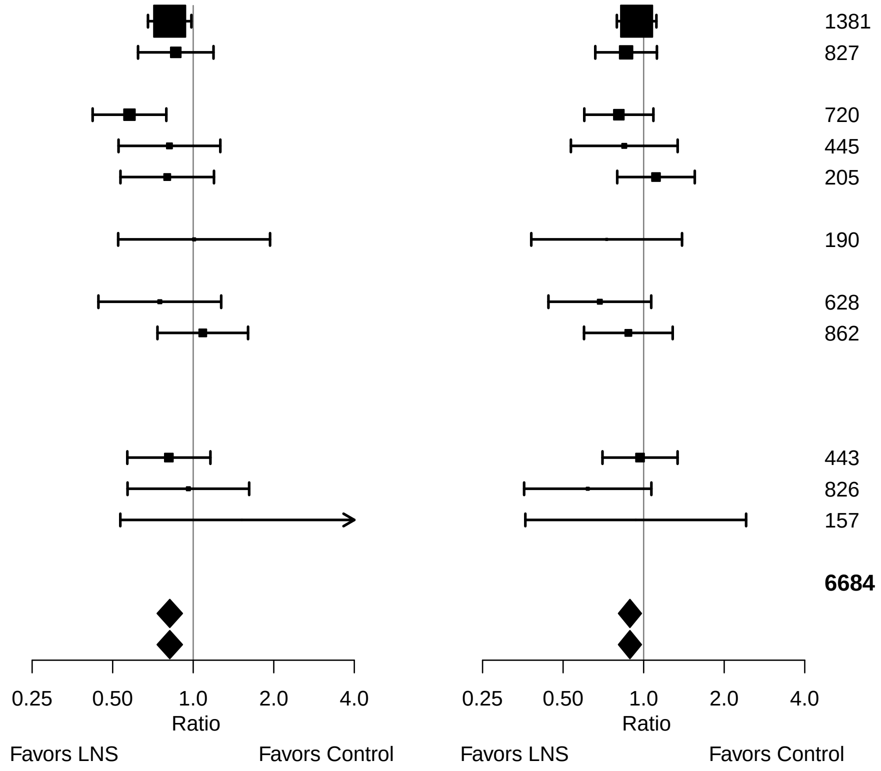

#### Supplemental figure 7J: Acute malnutrition prevalence ratio

#### 7J2: Stratified by Household food insecurity

**P−for−interaction = 0.565**
**Ratio of PRs = 0.95 (0.80, 1.13)**

| | | Mild to secure | | | | | | Moderate to severe | | | | | |
|---|---|---|---|---|---|---|---|---|---|---|---|---|---|
| **Country** | **Trial** | **LNS N** | **Control N** | **Control Prevalence** | **PR (95% CI)** | **Fixed W** | **Random W** | **LNS N** | **Control N** | **Control Prevalence** | **PR (95% CI)** | **Fixed W** | **Random W** |
| Bangladesh | JiVitA−4 (34) | 1977 | 840 | 20.8 | 0.87 (0.74, 1.01) | 0.46 | 0.46 | 805 | 376 | 21.5 | 0.95 (0.76, 1.20) | 0.27 | 0.23 |
| Bangladesh | RDNS (35) | 1057 | 493 | 13.8 | 0.91 (0.69, 1.19) | 0.15 | 0.15 | 604 | 322 | 17.7 | 0.81 (0.61, 1.08) | 0.18 | 0.20 |
| Bangladesh | WASH−B (36) | | | | | | | | | | | | |
| Burkina Faso | iLiNS−Zinc (37) | 1013 | 320 | 16.5 | 0.70 (0.46, 1.04) | 0.07 | 0.07 | 931 | 343 | 21.6 | 0.66 (0.54, 0.80) | 0.40 | 0.25 |
| Burkina Faso | PROMIS (38) | 494 | 543 | 10.9 | 0.99 (0.73, 1.33) | 0.13 | 0.13 | 352 | 358 | 12.8 | 0.64 (0.36, 1.14) | 0.04 | 0.10 |
| Burkina Faso | PROMIS CS (38) | | | | | | | | | | | | |
| Ghana | GHANA (39) | | | | | | | | | | | | |
| Ghana | iLiNS−DYADG (40) | 244 | 470 | 8.5 | 0.87 (0.51, 1.48) | 0.04 | 0.04 | 102 | 219 | 7.3 | 0.81 (0.32, 2.00) | 0.02 | 0.05 |
| Haiti | HAITI (41) | | | | | | | | | | | | |
| Kenya | WASH−B (42) | | | | | | | | | | | | |
| Madagascar | MAHAY (43) | 964 | 905 | 10.1 | 0.94 (0.67, 1.32) | 0.10 | 0.10 | 352 | 405 | 8.6 | 1.12 (0.71, 1.76) | 0.07 | 0.13 |
| Malawi | iLiNS−DYADM (44) | | | | | | | | | | | | |
| Malawi | iLiNS−DOSE (45) | | | | | | | | | | | | |
| Mali | PROMIS (46) | | | | | | | | | | | | |
| Mali | PROMIS CS (46) | | | | | | | | | | | | |
| Zimbabwe | SHINE (HIV−) (47) | 1382 | 1292 | 2.9 | 0.99 (0.63, 1.54) | 0.06 | 0.06 | 294 | 322 | 6.2 | 0.33 (0.13, 0.81) | 0.02 | 0.05 |
| Zimbabwe | SHINE (HIV+) (48) | | | | | | | | | | | | |
| | | **7131** | **4863** | | **I² = 0.00, Tau² = 0.00** | | | **3440** | **2345** | | **I² = 0.52, Tau² = 0.04** | | |
| **Fixed** | | | | | **0.89 (0.80, 0.99)** | | | | | | **0.78 (0.69, 0.88)** | | |
| **Random** | | | | | **0.89 (0.80, 0.99)** | | | | | | **0.78 (0.63, 0.97)** | | |

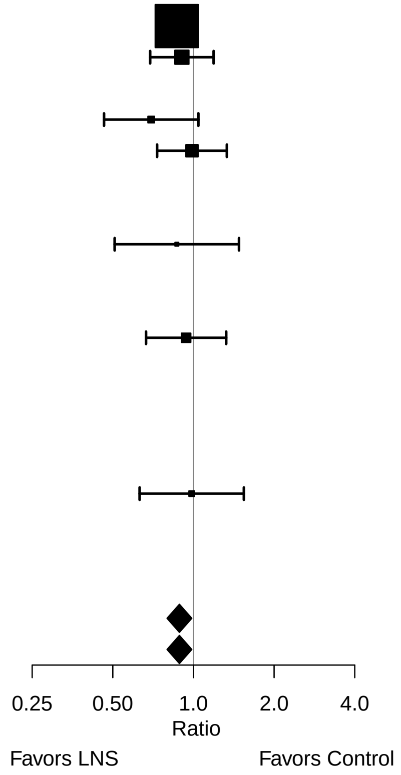

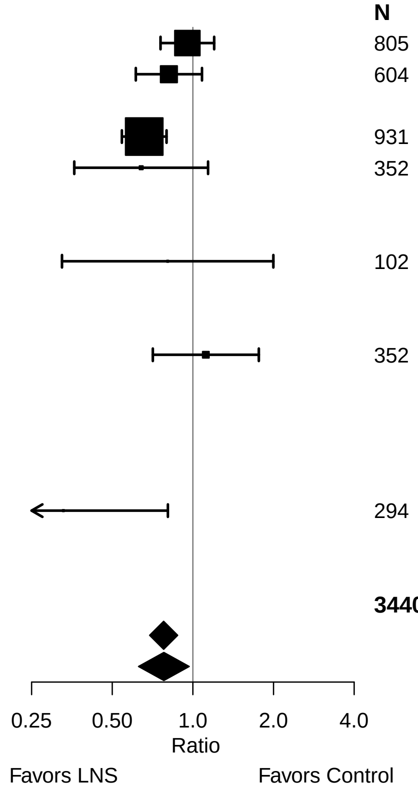

#### Supplemental figure 7J: Acute malnutrition prevalence ratio

##### 7J3: Stratified by Household source water quality

**P−for−interaction = 0.789**
**Ratio of PRs = 1.03 (0.82, 1.30)**

| Country | Trial | Improved LNS N | Improved Control N | Improved Control Prevalence | Improved PR (95% CI) | Improved Fixed W | Improved Random W | Unimproved LNS N | Unimproved Control N | Unimproved Control Prevalence | Unimproved PR (95% CI) | Unimproved Fixed W | Unimproved Random W |
|---|---|---|---|---|---|---|---|---|---|---|---|---|---|
| Bangladesh | JiVitA−4 (34) | | | | | | | | | | | | |
| Bangladesh | RDNS (35) | | | | | | | | | | | | |
| Bangladesh | WASH−B (36) | | | | | | | | | | | | |
| Burkina Faso | iLiNS−Zinc (37) | 550 | 141 | 17.7 | 0.53 (0.34, 0.82) | 0.13 | 0.14 | 1393 | 519 | 19.6 | 0.71 (0.56, 0.90) | 0.38 | 0.34 |
| Burkina Faso | PROMIS (38) | 349 | 372 | 11.3 | 0.89 (0.65, 1.22) | 0.26 | 0.24 | 353 | 349 | 12.9 | 0.68 (0.42, 1.11) | 0.09 | 0.10 |
| Burkina Faso | PROMIS CS (38) | 277 | 265 | 21.1 | 0.82 (0.55, 1.22) | 0.16 | 0.17 | 147 | 169 | 16.6 | 1.23 (0.77, 1.98) | 0.10 | 0.11 |
| Ghana | GHANA (39) | | | | | | | | | | | | |
| Ghana | iLiNS−DYADG (40) | | | | | | | | | | | | |
| Haiti | HAITI (41) | | | | | | | | | | | | |
| Kenya | WASH−B (42) | | | | | | | | | | | | |
| Madagascar | MAHAY (43) | 414 | 451 | 8.0 | 1.18 (0.69, 2.01) | 0.09 | 0.11 | 1181 | 1155 | 9.9 | 0.91 (0.67, 1.24) | 0.22 | 0.23 |
| Malawi | iLiNS−DYADM (44) | | | | | | | | | | | | |
| Malawi | iLiNS−DOSE (45) | | | | | | | | | | | | |
| Mali | PROMIS (46) | | | | | | | | | | | | |
| Mali | PROMIS CS (46) | 544 | 562 | 12.5 | 0.86 (0.64, 1.15) | 0.30 | 0.27 | 361 | 382 | 13.6 | 0.94 (0.66, 1.33) | 0.18 | 0.19 |
| Zimbabwe | SHINE (HIV−) (47) | 517 | 482 | 3.7 | 0.83 (0.42, 1.63) | 0.06 | 0.07 | 273 | 300 | 3.7 | 0.70 (0.30, 1.64) | 0.03 | 0.04 |
| Zimbabwe | SHINE (HIV+) (48) | | | | | | | | | | | | |
| | | **2651** | **2273** | | **I² = 0.16, Tau² = 0.01** | | | **3708** | **2874** | | **I² = 0.14, Tau² = 0.01** | | |
| **Fixed** | | | | | **0.83 (0.70, 0.97)** | | | | | | **0.83 (0.72, 0.96)** | | |
| **Random** | | | | | **0.82 (0.69, 0.99)** | | | | | | **0.84 (0.71, 0.98)** | | |

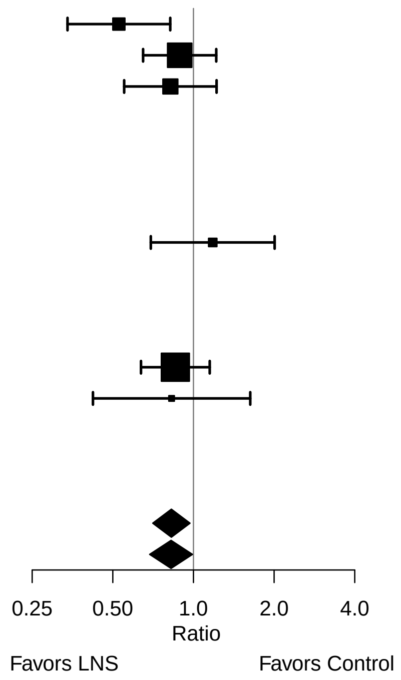

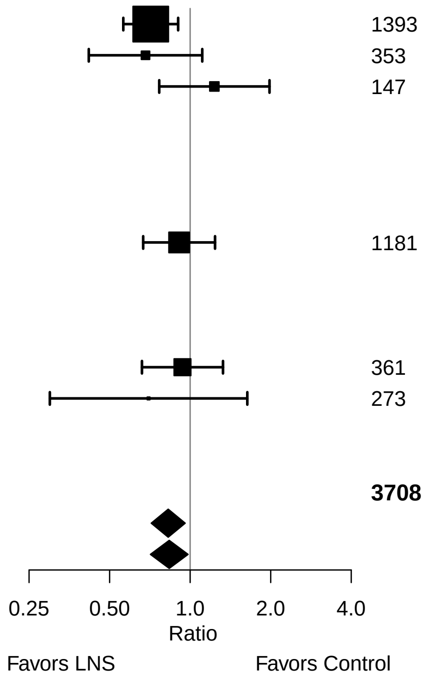

#### Supplemental figure 7J: Acute malnutrition prevalence ratio

### 7J4: Stratified by Household sanitation

**P−for−interaction = 0.013**
**Ratio of PRs = 1.25 (1.05, 1.49)**

| | | Improved | | | | | | Unimproved | | | | | |
|---|---|---|---|---|---|---|---|---|---|---|---|---|---|
| **Country** | **Trial** | **LNS N** | **Control N** | **Control Prevalence** | **PR (95% CI)** | **Fixed W** | **Random W** | **LNS N** | **Control N** | **Control Prevalence** | **PR (95% CI)** | **Fixed W** | **Random W** |
| Bangladesh | JiVitA−4 (34) | 2151 | 931 | 20.1 | 0.86 (0.74, 1.01) | 0.50 | 0.50 | 631 | 285 | 24.0 | 0.99 (0.79, 1.24) | 0.37 | 0.37 |
| Bangladesh | RDNS (35) | 1179 | 573 | 15.5 | 0.73 (0.57, 0.92) | 0.21 | 0.21 | 480 | 241 | 14.9 | 1.20 (0.87, 1.66) | 0.18 | 0.18 |
| Bangladesh | WASH−B (36) | | | | | | | | | | | | |
| Burkina Faso | iLiNS−Zinc (37) | | | | | | | | | | | | |
| Burkina Faso | PROMIS (38) | 389 | 544 | 12.5 | 0.78 (0.53, 1.16) | 0.08 | 0.08 | 457 | 353 | 10.5 | 0.92 (0.59, 1.42) | 0.10 | 0.10 |
| Burkina Faso | PROMIS CS (38) | 175 | 163 | 19.6 | 0.87 (0.58, 1.32) | 0.07 | 0.07 | 248 | 271 | 19.2 | 1.01 (0.74, 1.37) | 0.21 | 0.21 |
| Ghana | GHANA (39) | | | | | | | | | | | | |
| Ghana | iLiNS−DYADG (40) | | | | | | | | | | | | |
| Haiti | HAITI (41) | | | | | | | | | | | | |
| Kenya | WASH−B (42) | | | | | | | | | | | | |
| Madagascar | MAHAY (43) | | | | | | | | | | | | |
| Malawi | iLiNS−DYADM (44) | | | | | | | | | | | | |
| Malawi | iLiNS−DOSE (45) | | | | | | | | | | | | |
| Mali | PROMIS (46) | | | | | | | | | | | | |
| Mali | PROMIS CS (46) | 696 | 692 | 13.6 | 0.85 (0.63, 1.14) | 0.13 | 0.13 | 217 | 242 | 10.7 | 0.86 (0.55, 1.33) | 0.10 | 0.10 |
| Zimbabwe | SHINE (HIV−) (47) | 292 | 242 | 4.1 | 0.41 (0.16, 1.09) | 0.01 | 0.01 | 495 | 539 | 3.5 | 1.03 (0.52, 2.05) | 0.04 | 0.04 |
| Zimbabwe | SHINE (HIV+) (48) | | | | | | | | | | | | |
| | | **4882** | **3145** | | **I² = 0.00, Tau² = 0.00** | | | **2528** | **1931** | | **I² = 0.00, Tau² = 0.00** | | |
| **Fixed** | | | | | **0.82 (0.73, 0.91)** | | | | | | **1.01 (0.88, 1.16)** | | |
| **Random** | | | | | **0.82 (0.73, 0.91)** | | | | | | **1.01 (0.88, 1.16)** | | |

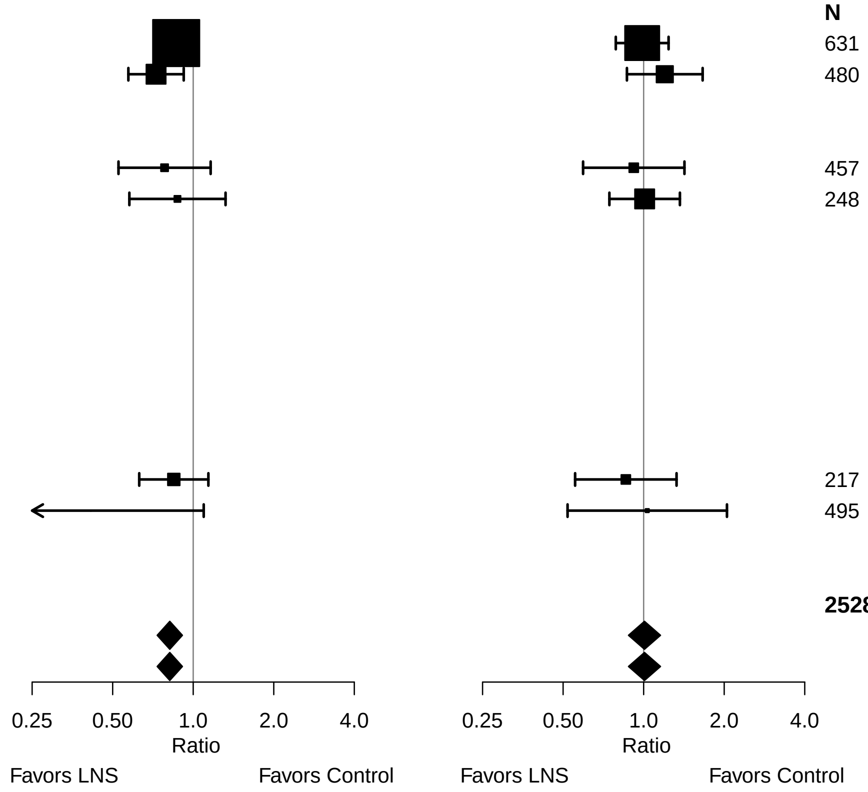

#### Supplemental figure 7J: Acute malnutrition prevalence ratio

#### 7J5: Stratified by Home environment

**P−for−interaction = 0.310**
**Ratio of PRs = 1.14 (0.89, 1.45)**

| Country | Trial | LNS N | Control N | Control Prevalence | At least median PR (95% CI) | Fixed W | Random W | LNS N | Control N | Control Prevalence | Less than median PR (95% CI) | Fixed W | Random W |
|---|---|---|---|---|---|---|---|---|---|---|---|---|---|
| Bangladesh | JiVitA−4 (34) | | | | | | | | | | | | |
| Bangladesh | RDNS (35) | 1192 | 582 | 14.9 | 0.78 (0.60, 1.01) | 0.36 | 0.26 | 466 | 230 | 15.7 | 1.08 (0.79, 1.49) | 0.35 | 0.30 |
| Bangladesh | WASH−B (36) | | | | | | | | | | | | |
| Burkina Faso | iLiNS−Zinc (37) | 473 | 199 | 18.6 | 0.50 (0.38, 0.66) | 0.30 | 0.25 | 271 | 174 | 21.7 | 0.79 (0.50, 1.25) | 0.17 | 0.20 |
| Burkina Faso | PROMIS (38) | | | | | | | | | | | | |
| Burkina Faso | PROMIS CS (38) | | | | | | | | | | | | |
| Ghana | GHANA (39) | | | | | | | | | | | | |
| Ghana | iLiNS−DYADG (40) | 213 | 409 | 7.1 | 1.06 (0.59, 1.91) | 0.07 | 0.13 | 119 | 248 | 9.7 | 0.43 (0.17, 1.11) | 0.04 | 0.07 |
| Haiti | HAITI (41) | | | | | | | | | | | | |
| Kenya | WASH−B (42) | 834 | 2756 | 2.4 | 0.90 (0.56, 1.45) | 0.10 | 0.16 | 572 | 2149 | 3.7 | 0.61 (0.34, 1.11) | 0.10 | 0.14 |
| Madagascar | MAHAY (43) | 902 | 889 | 7.3 | 0.97 (0.66, 1.43) | 0.16 | 0.20 | 773 | 766 | 11.9 | 0.99 (0.72, 1.37) | 0.34 | 0.29 |
| Malawi | iLiNS−DYADM (44) | | | | | | | | | | | | |
| Malawi | iLiNS−DOSE (45) | | | | | | | | | | | | |
| Mali | PROMIS (46) | | | | | | | | | | | | |
| Mali | PROMIS CS (46) | | | | | | | | | | | | |
| Zimbabwe | SHINE (HIV−) (47) | | | | | | | | | | | | |
| Zimbabwe | SHINE (HIV+) (48) | | | | | | | | | | | | |
| | | **3614** | **4835** | | **I² = 0.66, Tau² = 0.05** | | | **2201** | **3567** | | **I² = 0.32, Tau² = 0.03** | | |
| **Fixed** | | | | | **0.73 (0.63, 0.85)** | | | | | | **0.91 (0.75, 1.10)** | | |
| **Random** | | | | | **0.78 (0.59, 1.02)** | | | | | | **0.86 (0.66, 1.12)** | | |

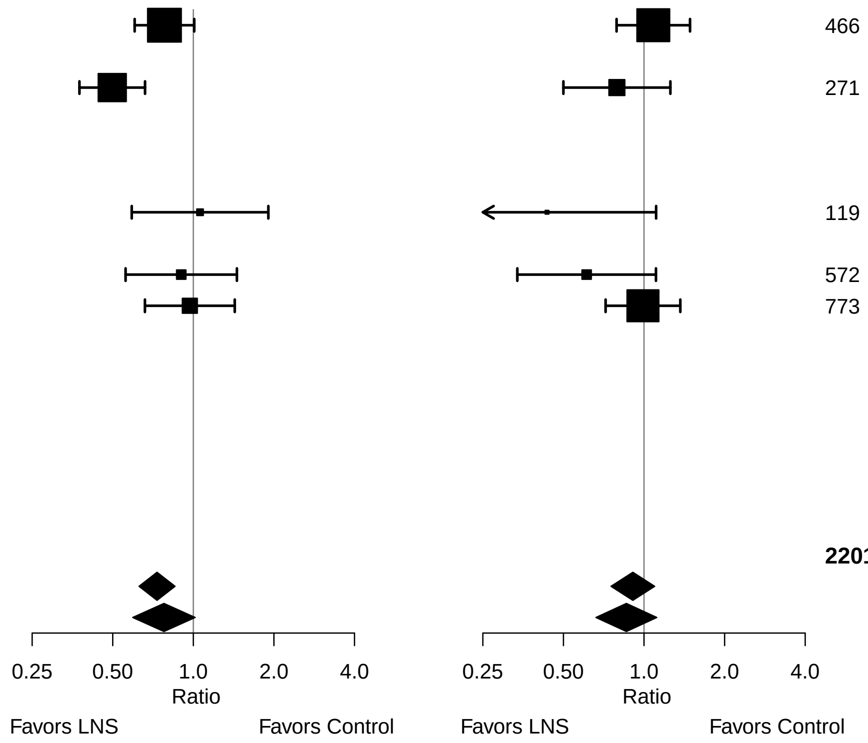

#### Supplemental figure 7J: Acute malnutrition prevalence ratio

#### 7J6: Stratified by Season at the time of assessment

**P−for−interaction = 0.172**
**Ratio of PRs = 1.12 (0.95, 1.32)**

| Country | Trial | Dry: LNS N | Dry: Control N | Dry: Control Prevalence | Dry: PR (95% CI) | Dry: Fixed W | Dry: Random W | Rainy: LNS N | Rainy: Control N | Rainy: Control Prevalence | Rainy: PR (95% CI) | Rainy: Fixed W | Rainy: Random W |
|---|---|---|---|---|---|---|---|---|---|---|---|---|---|
| Bangladesh | JiVitA−4 (34) | 1520 | 648 | 18.7 | 0.86 (0.71, 1.05) | 0.39 | 0.26 | 1263 | 568 | 23.7 | 0.92 (0.77, 1.10) | 0.40 | 0.40 |
| Bangladesh | RDNS (35) | 738 | 359 | 13.6 | 0.81 (0.59, 1.12) | 0.14 | 0.17 | 923 | 456 | 16.7 | 0.89 (0.69, 1.15) | 0.20 | 0.20 |
| Bangladesh | WASH−B (36) | | | | | | | | | | | | |
| Burkina Faso | iLiNS−Zinc (37) | 1393 | 477 | 20.1 | 0.62 (0.48, 0.78) | 0.25 | 0.22 | 558 | 187 | 17.0 | 0.87 (0.62, 1.22) | 0.11 | 0.11 |
| Burkina Faso | PROMIS (38) | 418 | 441 | 12.0 | 0.82 (0.53, 1.25) | 0.08 | 0.12 | 428 | 460 | 11.3 | 0.85 (0.56, 1.28) | 0.08 | 0.08 |
| Burkina Faso | PROMIS CS (38) | 222 | 190 | 18.4 | 1.00 (0.64, 1.57) | 0.07 | 0.11 | 202 | 244 | 20.1 | 0.91 (0.65, 1.27) | 0.12 | 0.12 |
| Ghana | GHANA (39) | | | | | | | | | | | | |
| Ghana | iLiNS−DYADG (40) | | | | | | | | | | | | |
| Haiti | HAITI (41) | | | | | | | | | | | | |
| Kenya | WASH−B (42) | 346 | 1146 | 3.8 | 0.62 (0.33, 1.16) | 0.04 | 0.07 | 1108 | 3971 | 3.0 | 0.74 (0.48, 1.13) | 0.07 | 0.07 |
| Madagascar | MAHAY (43) | | | | | | | | | | | | |
| Malawi | iLiNS−DYADM (44) | | | | | | | | | | | | |
| Malawi | iLiNS−DOSE (45) | 415 | 134 | 4.5 | 1.64 (0.74, 3.64) | 0.02 | 0.04 | 275 | 106 | 7.5 | 1.18 (0.56, 2.48) | 0.02 | 0.02 |
| Mali | PROMIS (46) | | | | | | | | | | | | |
| Mali | PROMIS CS (46) | | | | | | | | | | | | |
| Zimbabwe | SHINE (HIV−) (47) | | | | | | | | | | | | |
| Zimbabwe | SHINE (HIV+) (48) | | | | | | | | | | | | |
| | | **5052** | **3395** | | **I² = 0.39, Tau² = 0.02** | | | **4757** | **5992** | | **I² = 0.00, Tau² = 0.00** | | |
| **Fixed** | | | | | **0.79 (0.70, 0.89)** | | | | | | **0.89 (0.80, 1.00)** | | |
| **Random** | | | | | **0.81 (0.67, 0.96)** | | | | | | **0.89 (0.80, 1.00)** | | |

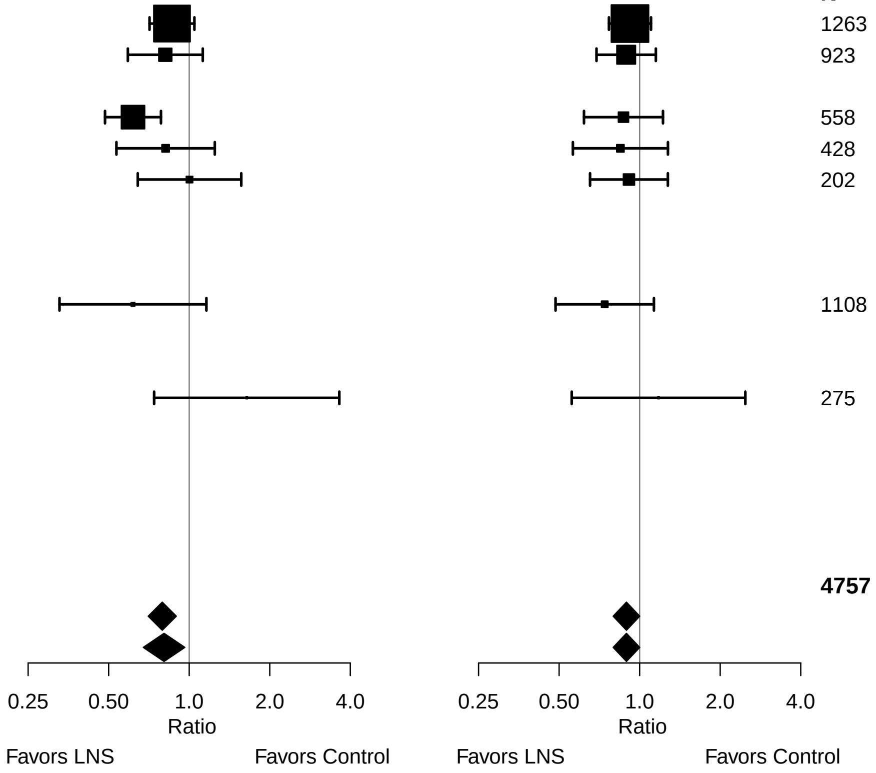

### Supplemental figure 7K: Acute malnutrition prevalence difference

#### 7K1: Stratified by Household socio-economic status

**P−for−interaction = 0.197**
**Difference in PDs = −0.01 (−0.02, 0.00)**

| | | At least median | | | | | | Less than median | | | | | |
|---|---|---|---|---|---|---|---|---|---|---|---|---|---|
| **Country** | **Trial** | **LNS N** | **Control N** | **Control Prevalence** | **PD (95% CI)** | **Fixed W** | **Random W** | **LNS N** | **Control N** | **Control Prevalence** | **PD (95% CI)** | **Fixed W** | **Random W** |
| Bangladesh | JiVitA−4 (34) | 1401 | 606 | 17.7 | −0.02 (−0.05, 0.01) | 0.06 | 0.10 | 1381 | 610 | 24.4 | −0.02 (−0.06, 0.01) | 0.06 | 0.06 |
| Bangladesh | RDNS (35) | 834 | 407 | 13.5 | −0.02 (−0.06, 0.02) | 0.03 | 0.06 | 827 | 408 | 17.2 | −0.02 (−0.07, 0.02) | 0.04 | 0.04 |
| Bangladesh | WASH−B (36) | | | | | | | | | | | | |
| Burkina Faso | iLiNS−Zinc (37) | 1224 | 299 | 18.4 | −0.07 (−0.12, −0.02) | 0.02 | 0.05 | 720 | 364 | 19.7 | −0.05 (−0.10, 0.00) | 0.03 | 0.03 |
| Burkina Faso | PROMIS (38) | 401 | 473 | 11.6 | −0.02 (−0.07, 0.02) | 0.02 | 0.05 | 445 | 428 | 11.7 | −0.02 (−0.07, 0.03) | 0.03 | 0.03 |
| Burkina Faso | PROMIS CS (38) | 219 | 206 | 19.4 | −0.04 (−0.11, 0.03) | 0.01 | 0.03 | 205 | 228 | 19.3 | 0.02 (−0.05, 0.09) | 0.02 | 0.02 |
| Ghana | GHANA (39) | | | | | | | | | | | | |
| Ghana | iLiNS−DYADG (40) | 157 | 356 | 7.6 | 0.00 (−0.05, 0.05) | 0.02 | 0.04 | 190 | 334 | 8.7 | −0.02 (−0.07, 0.02) | 0.04 | 0.04 |
| Haiti | HAITI (41) | | | | | | | | | | | | |
| Kenya | WASH−B (42) | 824 | 2914 | 2.3 | −0.01 (−0.02, 0.00) | 0.52 | 0.27 | 628 | 2199 | 4.4 | −0.01 (−0.03, 0.00) | 0.39 | 0.39 |
| Madagascar | MAHAY (43) | 760 | 865 | 7.5 | 0.01 (−0.02, 0.04) | 0.05 | 0.10 | 862 | 747 | 11.5 | −0.01 (−0.06, 0.03) | 0.05 | 0.05 |
| Malawi | iLiNS−DYADM (44) | | | | | | | | | | | | |
| Malawi | iLiNS−DOSE (45) | | | | | | | | | | | | |
| Mali | PROMIS (46) | | | | | | | | | | | | |
| Mali | PROMIS CS (46) | 501 | 458 | 13.5 | −0.03 (−0.07, 0.02) | 0.03 | 0.06 | 443 | 501 | 12.6 | 0.00 (−0.04, 0.04) | 0.05 | 0.05 |
| Zimbabwe | SHINE (HIV−) (47) | 901 | 795 | 2.9 | 0.00 (−0.02, 0.01) | 0.22 | 0.21 | 826 | 851 | 4.1 | −0.02 (−0.03, 0.00) | 0.26 | 0.26 |
| Zimbabwe | SHINE (HIV+) (48) | 171 | 156 | 3.8 | 0.02 (−0.03, 0.07) | 0.02 | 0.04 | 157 | 165 | 5.5 | 0.00 (−0.05, 0.05) | 0.03 | 0.03 |
| | | **7393** | **7535** | | **I² = 0.20, Tau² = 0.00** | | | **6684** | **6835** | | **I² = 0.00, Tau² = 0.00** | | |
| **Fixed** | | | | | **−0.01 (−0.01, 0.00)** | | | | | | **−0.02 (−0.02, −0.01)** | | |
| **Random** | | | | | **−0.01 (−0.02, 0.00)** | | | | | | **−0.02 (−0.02, −0.01)** | | |

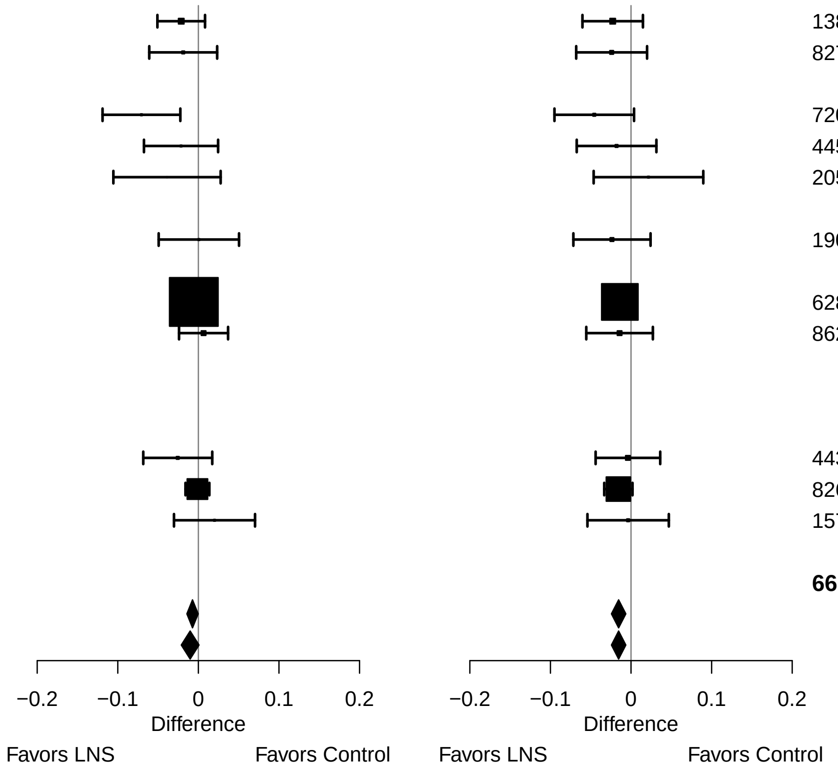

#### Supplemental figure 7K: Acute malnutrition prevalence difference

#### 7K2: Stratified by Household food insecurity

**P−for−interaction = 0.074**
**Difference in PDs = −0.02 (−0.04, 0.00)**

| | | Mild to secure | | | | | | Moderate to severe | | | | | |
|---|---|---|---|---|---|---|---|---|---|---|---|---|---|
| **Country** | **Trial** | **LNS N** | **Control N** | **Control Prevalence** | **PD (95% CI)** | **Fixed W** | **Random W** | **LNS N** | **Control N** | **Control Prevalence** | **PD (95% CI)** | **Fixed W** | **Random W** |
| Bangladesh | JiVitA−4 (34) | 1977 | 840 | 20.8 | −0.03 (−0.06, 0.00) | 0.11 | 0.11 | 805 | 376 | 21.5 | 0.00 (−0.05, 0.04) | 0.10 | 0.13 |
| Bangladesh | RDNS (35) | 1057 | 493 | 13.8 | −0.01 (−0.05, 0.02) | 0.07 | 0.07 | 604 | 322 | 17.7 | −0.03 (−0.08, 0.01) | 0.10 | 0.13 |
| Bangladesh | WASH−B (36) | | | | | | | | | | | | |
| Burkina Faso | iLiNS−Zinc (37) | 1013 | 320 | 16.5 | −0.05 (−0.11, 0.01) | 0.02 | 0.02 | 931 | 343 | 21.6 | −0.07 (−0.10, −0.05) | 0.35 | 0.21 |
| Burkina Faso | PROMIS (38) | 494 | 543 | 10.9 | 0.00 (−0.03, 0.03) | 0.09 | 0.09 | 352 | 358 | 12.8 | −0.05 (−0.11, 0.01) | 0.06 | 0.10 |
| Burkina Faso | PROMIS CS (38) | | | | | | | | | | | | |
| Ghana | GHANA (39) | | | | | | | | | | | | |
| Ghana | iLiNS−DYADG (40) | 244 | 470 | 8.5 | −0.01 (−0.05, 0.03) | 0.05 | 0.05 | 102 | 219 | 7.3 | −0.01 (−0.07, 0.05) | 0.06 | 0.10 |
| Haiti | HAITI (41) | | | | | | | | | | | | |
| Kenya | WASH−B (42) | | | | | | | | | | | | |
| Madagascar | MAHAY (43) | 964 | 905 | 10.1 | −0.01 (−0.04, 0.03) | 0.08 | 0.08 | 352 | 405 | 8.6 | 0.01 (−0.03, 0.05) | 0.12 | 0.15 |
| Malawi | iLiNS−DYADM (44) | | | | | | | | | | | | |
| Malawi | iLiNS−DOSE (45) | | | | | | | | | | | | |
| Mali | PROMIS (46) | | | | | | | | | | | | |
| Mali | PROMIS CS (46) | | | | | | | | | | | | |
| Zimbabwe | SHINE (HIV−) (47) | 1382 | 1292 | 2.9 | 0.00 (−0.01, 0.01) | 0.58 | 0.58 | 294 | 322 | 6.2 | −0.04 (−0.07, −0.01) | 0.21 | 0.18 |
| Zimbabwe | SHINE (HIV+) (48) | | | | | | | | | | | | |
| | | **7131** | **4863** | | **I² = 0.00, Tau² = 0.00** | | | **3440** | **2345** | | **I² = 0.60, Tau² = 0.00** | | |
| **Fixed** | | | | | **−0.01 (−0.02, 0.00)** | | | | | | **−0.04 (−0.05, −0.03)** | | |
| **Random** | | | | | **−0.01 (−0.02, 0.00)** | | | | | | **−0.03 (−0.05, −0.01)** | | |

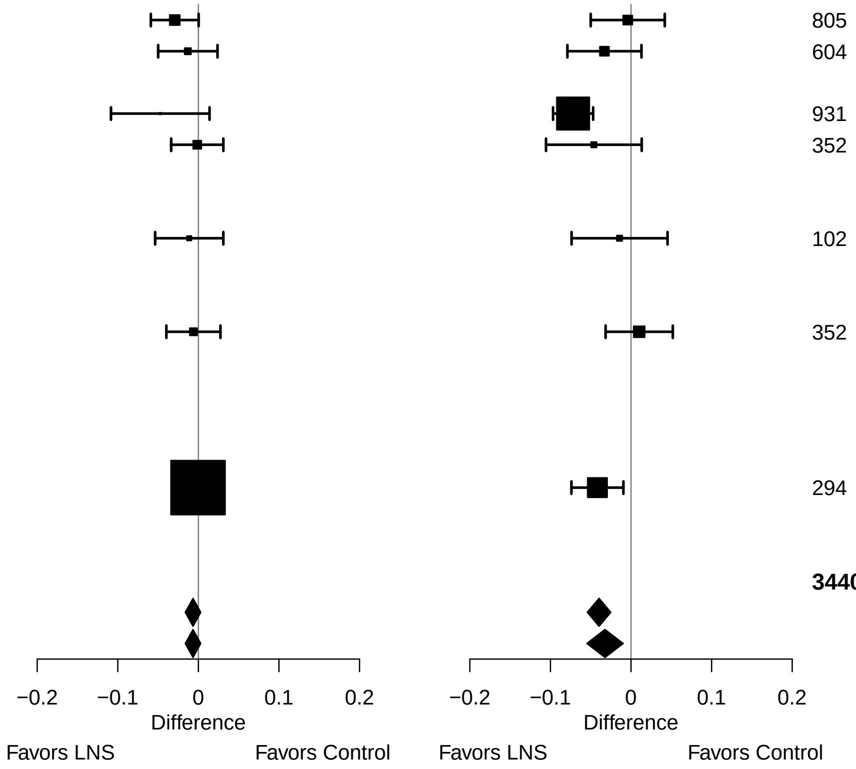

#### Supplemental figure 7K: Acute malnutrition prevalence difference

#### 7K3: Stratified by Household source water quality

**P−for−interaction = 0.574**
**Difference in PDs = −0.01 (−0.03, 0.02)**

| Country | Trial | LNS N | Control N | Control Prevalence | Improved PD (95% CI) | Fixed W | Random W | LNS N | Control N | Control Prevalence | Unimproved PD (95% CI) | Fixed W | Random W |
|---|---|---|---|---|---|---|---|---|---|---|---|---|---|
| Bangladesh | JiVitA−4 (34) | | | | | | | | | | | | |
| Bangladesh | RDNS (35) | | | | | | | | | | | | |
| Bangladesh | WASH−B (36) | | | | | | | | | | | | |
| Burkina Faso | iLiNS−Zinc (37) | 550 | 141 | 17.7 | −0.06 (−0.12, −0.01) | 0.07 | 0.07 | 1393 | 519 | 19.6 | −0.06 (−0.10, −0.02) | 0.16 | 0.18 |
| Burkina Faso | PROMIS (38) | 349 | 372 | 11.3 | −0.01 (−0.05, 0.02) | 0.19 | 0.19 | 353 | 349 | 12.9 | −0.04 (−0.09, 0.01) | 0.08 | 0.12 |
| Burkina Faso | PROMIS CS (38) | 277 | 265 | 21.1 | −0.04 (−0.11, 0.04) | 0.04 | 0.04 | 147 | 169 | 16.6 | 0.04 (−0.05, 0.12) | 0.03 | 0.05 |
| Ghana | GHANA (39) | | | | | | | | | | | | |
| Ghana | iLiNS−DYADG (40) | | | | | | | | | | | | |
| Haiti | HAITI (41) | | | | | | | | | | | | |
| Kenya | WASH−B (42) | | | | | | | | | | | | |
| Madagascar | MAHAY (43) | 414 | 451 | 8.0 | 0.01 (−0.03, 0.06) | 0.10 | 0.10 | 1181 | 1155 | 9.9 | −0.01 (−0.04, 0.02) | 0.28 | 0.24 |
| Malawi | iLiNS−DYADM (44) | | | | | | | | | | | | |
| Malawi | iLiNS−DOSE (45) | | | | | | | | | | | | |
| Mali | PROMIS (46) | | | | | | | | | | | | |
| Mali | PROMIS CS (46) | 544 | 562 | 12.5 | −0.02 (−0.05, 0.02) | 0.19 | 0.19 | 361 | 382 | 13.6 | −0.01 (−0.06, 0.04) | 0.11 | 0.14 |
| Zimbabwe | SHINE (HIV−) (47) | 517 | 482 | 3.7 | −0.01 (−0.03, 0.02) | 0.41 | 0.41 | 273 | 300 | 3.7 | −0.01 (−0.04, 0.02) | 0.34 | 0.26 |
| Zimbabwe | SHINE (HIV+) (48) | | | | | | | | | | | | |
| | | **2651** | **2273** | | **I² = 0.05, Tau² = 0.00** | | | **3708** | **2874** | | **I² = 0.38, Tau² = 0.00** | | |
| **Fixed** | | | | | **−0.01 (−0.03, 0.00)** | | | | | | **−0.02 (−0.03, 0.00)** | | |
| **Random** | | | | | **−0.01 (−0.03, 0.00)** | | | | | | **−0.02 (−0.04, 0.00)** | | |

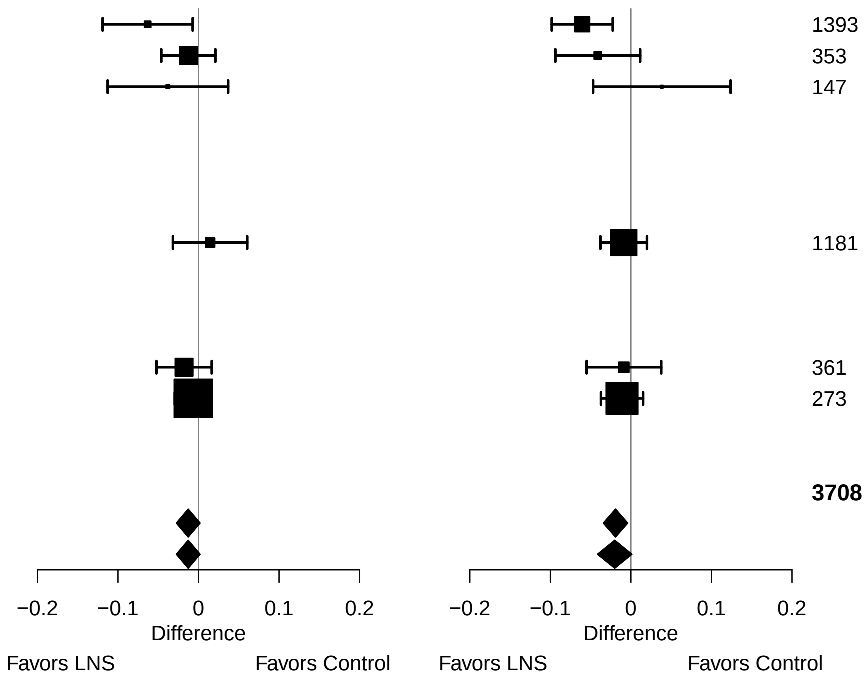

#### Supplemental figure 7K: Acute malnutrition prevalence difference

#### 7K4: Stratified by Household sanitation

**P−for−interaction = 0.016**
**Difference in PDs = 0.03 (0.01, 0.05)**

| Country | Trial | Improved LNS N | Improved Control N | Improved Control Prevalence | Improved PD (95% CI) | Improved Fixed W | Improved Random W | Unimproved LNS N | Unimproved Control N | Unimproved Control Prevalence | Unimproved PD (95% CI) | Unimproved Fixed W | Unimproved Random W |
|---|---|---|---|---|---|---|---|---|---|---|---|---|---|
| Bangladesh | JiVitA−4 (34) | 2151 | 931 | 20.1 | −0.03 (−0.06, 0.00) | 0.27 | 0.27 | 631 | 285 | 24.0 | 0.00 (−0.05, 0.06) | 0.10 | 0.10 |
| Bangladesh | RDNS (35) | 1179 | 573 | 15.5 | −0.04 (−0.08, −0.01) | 0.18 | 0.18 | 480 | 241 | 14.9 | 0.03 (−0.02, 0.08) | 0.10 | 0.10 |
| Bangladesh | WASH−B (36) | | | | | | | | | | | | |
| Burkina Faso | iLiNS−Zinc (37) | | | | | | | | | | | | |
| Burkina Faso | PROMIS (38) | 389 | 544 | 12.5 | −0.03 (−0.07, 0.02) | 0.11 | 0.11 | 457 | 353 | 10.5 | −0.01 (−0.05, 0.04) | 0.14 | 0.14 |
| Burkina Faso | PROMIS CS (38) | 175 | 163 | 19.6 | −0.02 (−0.10, 0.05) | 0.03 | 0.03 | 248 | 271 | 19.2 | 0.00 (−0.06, 0.06) | 0.08 | 0.08 |
| Ghana | GHANA (39) | | | | | | | | | | | | |
| Ghana | iLiNS−DYADG (40) | | | | | | | | | | | | |
| Haiti | HAITI (41) | | | | | | | | | | | | |
| Kenya | WASH−B (42) | | | | | | | | | | | | |
| Madagascar | MAHAY (43) | | | | | | | | | | | | |
| Malawi | iLiNS−DYADM (44) | | | | | | | | | | | | |
| Malawi | iLiNS−DOSE (45) | | | | | | | | | | | | |
| Mali | PROMIS (46) | | | | | | | | | | | | |
| Mali | PROMIS CS (46) | 696 | 692 | 13.6 | −0.02 (−0.06, 0.02) | 0.14 | 0.14 | 217 | 242 | 10.7 | −0.02 (−0.06, 0.03) | 0.13 | 0.13 |
| Zimbabwe | SHINE (HIV−) (47) | 292 | 242 | 4.1 | −0.02 (−0.05, 0.00) | 0.27 | 0.27 | 495 | 539 | 3.5 | 0.00 (−0.02, 0.03) | 0.45 | 0.45 |
| Zimbabwe | SHINE (HIV+) (48) | | | | | | | | | | | | |
| | | **4882** | **3145** | | **I² = 0.00, Tau² = 0.00** | | | **2528** | **1931** | | **I² = 0.00, Tau² = 0.00** | | |
| **Fixed** | | | | | **−0.03 (−0.04, −0.01)** | | | | | | **0.00 (−0.02, 0.02)** | | |
| **Random** | | | | | **−0.03 (−0.04, −0.01)** | | | | | | **0.00 (−0.02, 0.02)** | | |

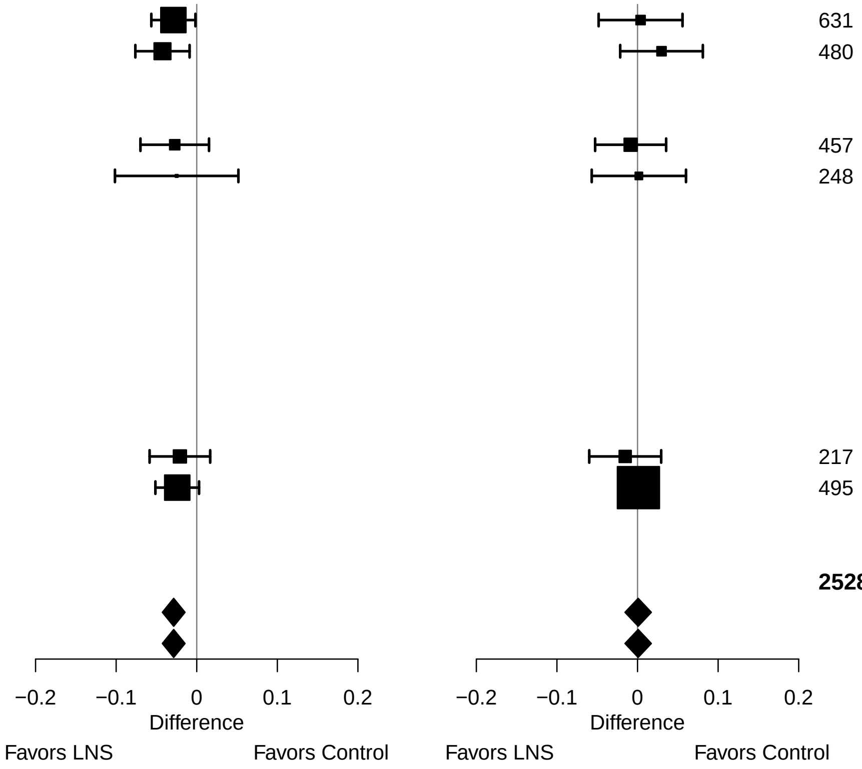

#### Supplemental figure 7K: Acute malnutrition prevalence difference

##### 7K5: Stratified by Home environment

**P−for−interaction = 0.318**
**Difference in PDs = −0.01 (−0.02, 0.01)**

| Country | Trial | LNS N | Control N | Control Prevalence | At least median PD (95% CI) | Fixed W | Random W | LNS N | Control N | Control Prevalence | Less than median PD (95% CI) | Fixed W | Random W |
|---|---|---|---|---|---|---|---|---|---|---|---|---|---|
| Bangladesh | JiVitA−4 (34) | | | | | | | | | | | | |
| Bangladesh | RDNS (35) | 1192 | 582 | 14.9 | −0.03 (−0.07, 0.00) | 0.06 | 0.19 | 466 | 230 | 15.7 | 0.01 (−0.04, 0.06) | 0.07 | 0.07 |
| Bangladesh | WASH−B (36) | | | | | | | | | | | | |
| Burkina Faso | iLiNS−Zinc (37) | 473 | 199 | 18.6 | −0.08 (−0.12, −0.05) | 0.07 | 0.19 | 271 | 174 | 21.7 | −0.04 (−0.13, 0.05) | 0.02 | 0.02 |
| Burkina Faso | PROMIS (38) | | | | | | | | | | | | |
| Burkina Faso | PROMIS CS (38) | | | | | | | | | | | | |
| Ghana | GHANA (39) | | | | | | | | | | | | |
| Ghana | iLiNS−DYADG (40) | 213 | 409 | 7.1 | 0.00 (−0.04, 0.05) | 0.04 | 0.17 | 119 | 248 | 9.7 | −0.05 (−0.11, 0.00) | 0.05 | 0.05 |
| Haiti | HAITI (41) | | | | | | | | | | | | |
| Kenya | WASH−B (42) | 834 | 2756 | 2.4 | 0.00 (−0.01, 0.01) | 0.72 | 0.24 | 572 | 2149 | 3.7 | −0.01 (−0.03, 0.00) | 0.75 | 0.75 |
| Madagascar | MAHAY (43) | 902 | 889 | 7.3 | 0.00 (−0.03, 0.03) | 0.10 | 0.21 | 773 | 766 | 11.9 | 0.00 (−0.04, 0.04) | 0.12 | 0.12 |
| Malawi | iLiNS−DYADM (44) | | | | | | | | | | | | |
| Malawi | iLiNS−DOSE (45) | | | | | | | | | | | | |
| Mali | PROMIS (46) | | | | | | | | | | | | |
| Mali | PROMIS CS (46) | | | | | | | | | | | | |
| Zimbabwe | SHINE (HIV−) (47) | | | | | | | | | | | | |
| Zimbabwe | SHINE (HIV+) (48) | | | | | | | | | | | | |
| | | **3614** | **4835** | | **I² = 0.83, Tau² = 0.00** | | | **2201** | **3567** | | **I² = 0.00, Tau² = 0.00** | | |
| **Fixed** | | | | | **−0.01 (−0.02, 0.00)** | | | | | | **−0.01 (−0.03, 0.00)** | | |
| **Random** | | | | | **−0.02 (−0.05, 0.01)** | | | | | | **−0.01 (−0.03, 0.00)** | | |

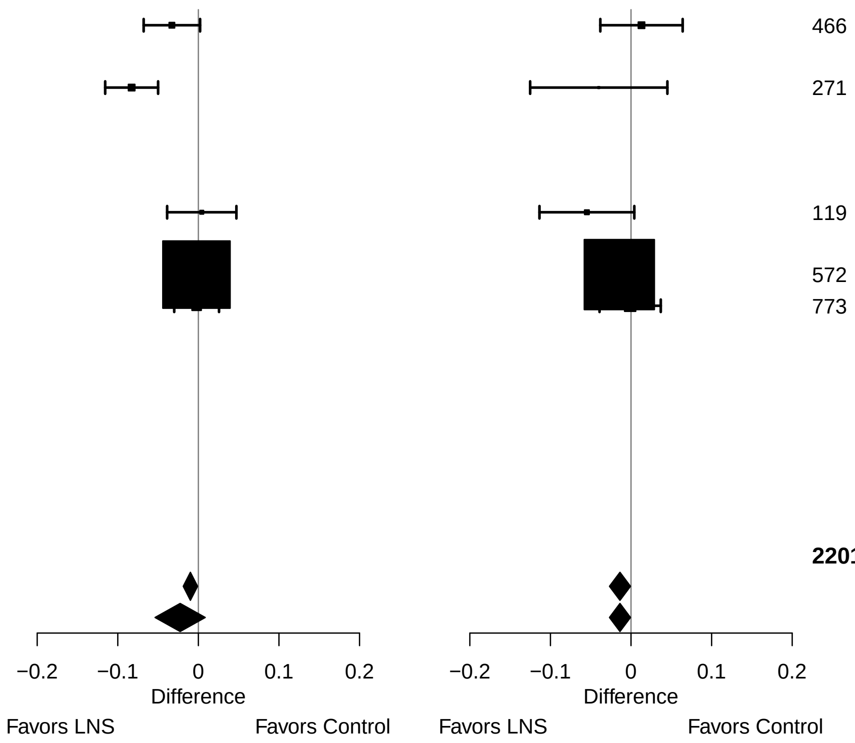

#### Supplemental figure 7K: Acute malnutrition prevalence difference

#### 7K6: Stratified by Season at the time of assessment

**P−for−interaction = 0.534**
**Difference in PDs = 0.01 (−0.01, 0.02)**

| Country | Trial | Dry LNS N | Dry Control N | Dry Control Prevalence | Dry PD (95% CI) | Dry Fixed W | Dry Random W | Rainy LNS N | Rainy Control N | Rainy Control Prevalence | Rainy PD (95% CI) | Rainy Fixed W | Rainy Random W |
|---|---|---|---|---|---|---|---|---|---|---|---|---|---|
| Bangladesh | JiVitA−4 (34) | 1520 | 648 | 18.7 | −0.02 (−0.05, 0.01) | 0.14 | 0.17 | 1263 | 568 | 23.7 | −0.02 (−0.06, 0.02) | 0.05 | 0.05 |
| Bangladesh | RDNS (35) | 738 | 359 | 13.6 | −0.03 (−0.07, 0.02) | 0.08 | 0.13 | 923 | 456 | 16.7 | −0.02 (−0.06, 0.02) | 0.05 | 0.05 |
| Bangladesh | WASH−B (36) | | | | | | | | | | | | |
| Burkina Faso | iLiNS−Zinc (37) | 1393 | 477 | 20.1 | −0.07 (−0.10, −0.03) | 0.12 | 0.16 | 558 | 187 | 17.0 | −0.04 (−0.10, 0.01) | 0.03 | 0.03 |
| Burkina Faso | PROMIS (38) | 418 | 441 | 12.0 | −0.02 (−0.07, 0.02) | 0.07 | 0.12 | 428 | 460 | 11.3 | −0.02 (−0.06, 0.03) | 0.04 | 0.04 |
| Burkina Faso | PROMIS CS (38) | 222 | 190 | 18.4 | 0.00 (−0.08, 0.08) | 0.02 | 0.05 | 202 | 244 | 20.1 | −0.02 (−0.08, 0.05) | 0.02 | 0.02 |
| Ghana | GHANA (39) | | | | | | | | | | | | |
| Ghana | iLiNS−DYADG (40) | | | | | | | | | | | | |
| Haiti | HAITI (41) | | | | | | | | | | | | |
| Kenya | WASH−B (42) | 346 | 1146 | 3.8 | −0.01 (−0.03, 0.00) | 0.50 | 0.26 | 1108 | 3971 | 3.0 | −0.01 (−0.02, 0.00) | 0.79 | 0.79 |
| Madagascar | MAHAY (43) | | | | | | | | | | | | |
| Malawi | iLiNS−DYADM (44) | | | | | | | | | | | | |
| Malawi | iLiNS−DOSE (45) | 415 | 134 | 4.5 | 0.02 (−0.03, 0.06) | 0.07 | 0.11 | 275 | 106 | 7.5 | 0.00 (−0.06, 0.06) | 0.02 | 0.02 |
| Mali | PROMIS (46) | | | | | | | | | | | | |
| Mali | PROMIS CS (46) | | | | | | | | | | | | |
| Zimbabwe | SHINE (HIV−) (47) | | | | | | | | | | | | |
| Zimbabwe | SHINE (HIV+) (48) | | | | | | | | | | | | |
| | | **5052** | **3395** | | **I² = 0.44, Tau² = 0.00** | | | **4757** | **5992** | | **I² = 0.00, Tau² = 0.00** | | |
| **Fixed** | | | | | **−0.02 (−0.03, −0.01)** | | | | | | **−0.01 (−0.02, 0.00)** | | |
| **Random** | | | | | **−0.02 (−0.04, 0.00)** | | | | | | **−0.01 (−0.02, 0.00)** | | |

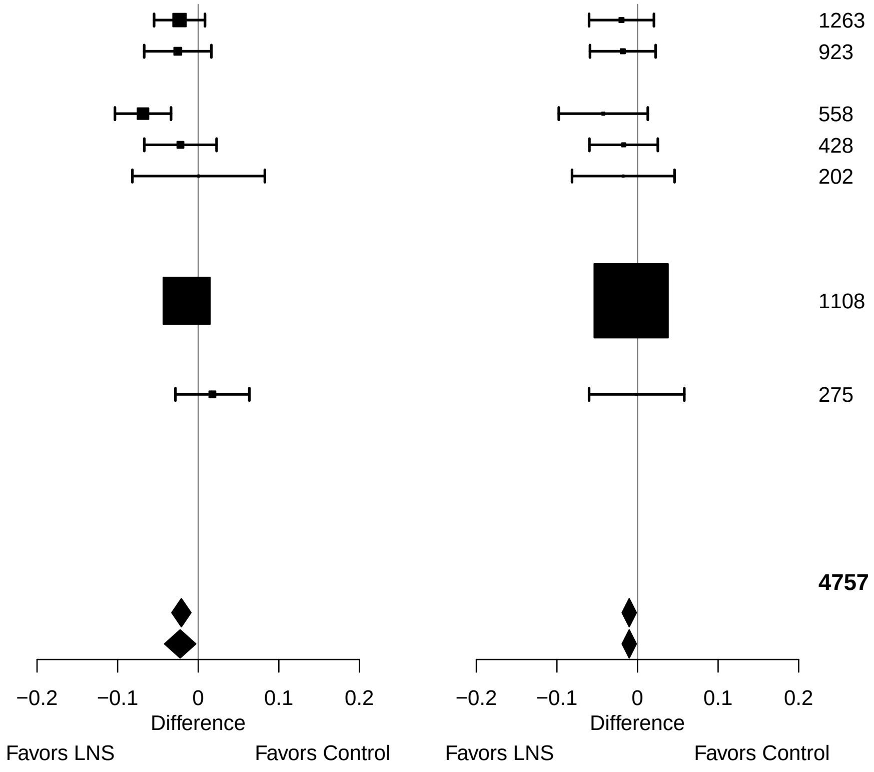

### Supplemental figure 7L: Mean difference in WAZ

#### 7L1: Stratified by Household socio-economic status

**P−for−interaction = 0.954**
**Difference in MDs = 0.00 (−0.03, 0.04)**

| | | At least median | | | | | | Less than median | | | | | |
|---|---|---|---|---|---|---|---|---|---|---|---|---|---|
| **Country** | **Trial** | **LNS N** | **Control N** | **Control Mean** | **MD (95% CI)** | **Fixed W** | **Random W** | **LNS N** | **Control N** | **Control Mean** | **MD (95% CI)** | **Fixed W** | **Random W** |
| Bangladesh | JiVitA−4 (34) | 1421 | 619 | −1.64 | 0.12 (0.07, 0.17) | 0.32 | 0.11 | 1392 | 613 | −1.91 | 0.09 (0.04, 0.14) | 0.29 | 0.13 |
| Bangladesh | RDNS (35) | 834 | 407 | −1.61 | 0.09 (−0.02, 0.20) | 0.06 | 0.07 | 829 | 408 | −1.88 | 0.10 (0.01, 0.19) | 0.09 | 0.09 |
| Bangladesh | WASH−B (36) | 584 | 1741 | −1.33 | 0.18 (0.07, 0.28) | 0.07 | 0.08 | 579 | 1720 | −1.78 | 0.22 (0.14, 0.30) | 0.10 | 0.09 |
| Burkina Faso | iLiNS−Zinc (37) | 1233 | 300 | −1.51 | 0.28 (0.19, 0.37) | 0.09 | 0.08 | 720 | 365 | −1.54 | 0.27 (0.20, 0.34) | 0.15 | 0.11 |
| Burkina Faso | PROMIS (38) | 405 | 482 | −1.19 | 0.14 (−0.02, 0.29) | 0.03 | 0.05 | 452 | 429 | −1.27 | 0.16 (0.00, 0.33) | 0.02 | 0.04 |
| Burkina Faso | PROMIS CS (38) | 220 | 208 | −1.25 | −0.05 (−0.26, 0.17) | 0.02 | 0.04 | 210 | 228 | −1.28 | −0.05 (−0.23, 0.14) | 0.02 | 0.04 |
| Ghana | GHANA (39) | 57 | 44 | −0.40 | 0.14 (−0.35, 0.62) | 0.00 | 0.01 | 41 | 46 | −0.87 | 0.29 (−0.16, 0.74) | 0.00 | 0.01 |
| Ghana | iLiNS−DYADG (40) | 157 | 356 | −0.75 | 0.11 (−0.09, 0.30) | 0.02 | 0.04 | 190 | 335 | −0.96 | 0.22 (0.04, 0.41) | 0.02 | 0.04 |
| Haiti | HAITI (41) | 74 | 61 | −0.21 | −0.03 (−0.21, 0.15) | 0.02 | 0.04 | 61 | 72 | −0.60 | 0.12 (−0.06, 0.30) | 0.02 | 0.04 |
| Kenya | WASH−B (42) | 828 | 2941 | −0.67 | 0.16 (0.09, 0.24) | 0.13 | 0.09 | 631 | 2220 | −0.85 | 0.10 (−0.01, 0.20) | 0.07 | 0.08 |
| Madagascar | MAHAY (43) | 772 | 884 | −1.35 | −0.02 (−0.16, 0.13) | 0.04 | 0.06 | 877 | 758 | −1.56 | 0.08 (−0.07, 0.23) | 0.03 | 0.05 |
| Malawi | iLiNS−DYADM (44) | 121 | 234 | −0.80 | −0.05 (−0.27, 0.16) | 0.02 | 0.04 | 99 | 209 | −0.96 | −0.10 (−0.36, 0.16) | 0.01 | 0.02 |
| Malawi | iLiNS−DOSE (45) | 307 | 104 | −0.91 | 0.00 (−0.15, 0.15) | 0.03 | 0.06 | 294 | 103 | −1.30 | 0.11 (−0.06, 0.27) | 0.03 | 0.05 |
| Mali | PROMIS (46) | 247 | 250 | −1.13 | 0.23 (0.09, 0.36) | 0.04 | 0.06 | 245 | 246 | −1.14 | 0.10 (−0.05, 0.26) | 0.03 | 0.05 |
| Mali | PROMIS CS (46) | 501 | 458 | −1.33 | 0.25 (0.11, 0.40) | 0.04 | 0.06 | 445 | 502 | −1.34 | 0.20 (0.04, 0.37) | 0.03 | 0.04 |
| Zimbabwe | SHINE (HIV−) (47) | 903 | 797 | −0.66 | 0.10 (0.00, 0.20) | 0.07 | 0.08 | 826 | 858 | −0.87 | 0.21 (0.11, 0.30) | 0.09 | 0.09 |
| Zimbabwe | SHINE (HIV+) (48) | 171 | 156 | −0.87 | −0.02 (−0.27, 0.22) | 0.01 | 0.03 | 157 | 166 | −1.01 | 0.04 (−0.21, 0.29) | 0.01 | 0.02 |
| | | **8835** | **10042** | | **I² = 0.53, Tau² = 0.01** | | | **8048** | **9278** | | **I² = 0.55, Tau² = 0.00** | | |
| **Fixed** | | | | | **0.13 (0.10, 0.16)** | | | | | | **0.14 (0.12, 0.17)** | | |
| **Random** | | | | | **0.11 (0.07, 0.16)** | | | | | | **0.14 (0.10, 0.18)** | | |

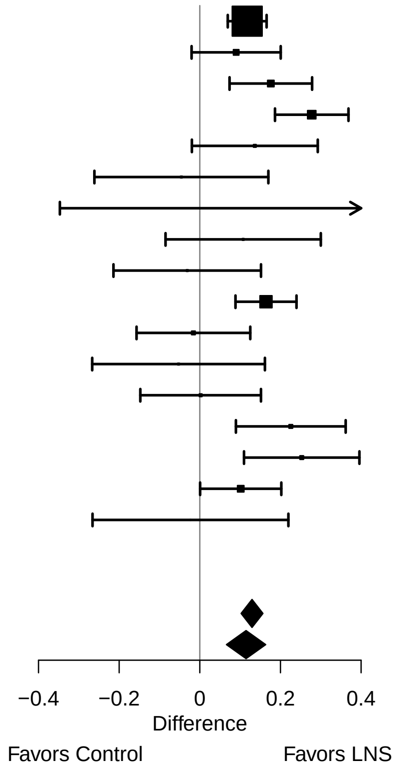

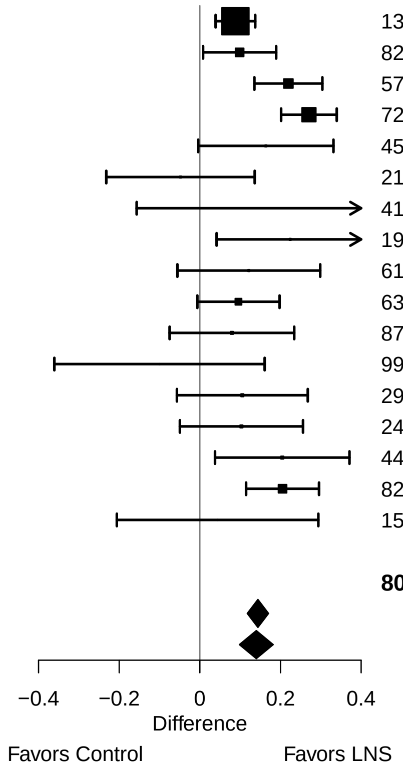

#### Supplemental figure 7L: Mean difference in WAZ

#### 7L2: Stratified by Household food insecurity

**P−for−interaction = 0.751**
**Difference in MDs = −0.01 (−0.05, 0.04)**

| | | Mild to secure | | | | | | | Moderate to severe | | | | | |
|---|---|---|---|---|---|---|---|---|---|---|---|---|---|---|
| **Country** | **Trial** | **LNS N** | **Control N** | **Control Mean** | **MD (95% CI)** | **Fixed W** | **Random W** | | **LNS N** | **Control N** | **Control Mean** | **MD (95% CI)** | **Fixed W** | **Random W** |
| Bangladesh | JiVitA−4 (34) | 2000 | 851 | −1.71 | 0.12 (0.08, 0.16) | 0.36 | 0.13 | | 813 | 381 | −1.90 | 0.07 (0.01, 0.14) | 0.28 | 0.16 |
| Bangladesh | RDNS (35) | 1058 | 493 | −1.63 | 0.02 (−0.07, 0.12) | 0.06 | 0.09 | | 605 | 322 | −1.93 | 0.19 (0.07, 0.31) | 0.09 | 0.10 |
| Bangladesh | WASH−B (36) | 923 | 2681 | −1.49 | 0.22 (0.14, 0.29) | 0.10 | 0.11 | | 240 | 780 | −1.76 | 0.10 (−0.02, 0.22) | 0.08 | 0.10 |
| Burkina Faso | iLiNS−Zinc (37) | 1017 | 321 | −1.48 | 0.28 (0.19, 0.37) | 0.08 | 0.10 | | 936 | 344 | −1.57 | 0.27 (0.19, 0.35) | 0.21 | 0.15 |
| Burkina Faso | PROMIS (38) | 502 | 550 | −1.21 | 0.16 (0.02, 0.31) | 0.03 | 0.06 | | 355 | 361 | −1.25 | 0.12 (−0.05, 0.29) | 0.04 | 0.06 |
| Burkina Faso | PROMIS CS (38) | | | | | | | | | | | | | |
| Ghana | GHANA (39) | | | | | | | | | | | | | |
| Ghana | iLiNS−DYADG (40) | 244 | 471 | −0.83 | 0.19 (0.03, 0.35) | 0.02 | 0.06 | | 102 | 219 | −0.89 | 0.08 (−0.15, 0.31) | 0.02 | 0.04 |
| Haiti | HAITI (41) | | | | | | | | | | | | | |
| Kenya | WASH−B (42) | 1303 | 4621 | −0.74 | 0.14 (0.07, 0.20) | 0.15 | 0.11 | | 156 | 527 | −0.84 | 0.09 (−0.12, 0.30) | 0.03 | 0.05 |
| Madagascar | MAHAY (43) | 989 | 929 | −1.43 | 0.03 (−0.10, 0.16) | 0.04 | 0.07 | | 354 | 410 | −1.49 | 0.04 (−0.15, 0.23) | 0.04 | 0.06 |
| Malawi | iLiNS−DYADM (44) | 70 | 122 | −0.68 | −0.13 (−0.42, 0.16) | 0.01 | 0.02 | | 148 | 316 | −0.94 | −0.06 (−0.26, 0.15) | 0.03 | 0.05 |
| Malawi | iLiNS−DOSE (45) | 154 | 55 | −1.02 | 0.05 (−0.16, 0.25) | 0.01 | 0.04 | | 434 | 146 | −1.17 | 0.07 (−0.06, 0.21) | 0.07 | 0.09 |
| Mali | PROMIS (46) | 327 | 338 | −1.13 | 0.16 (0.02, 0.30) | 0.03 | 0.07 | | 170 | 163 | −1.19 | 0.18 (0.00, 0.36) | 0.04 | 0.06 |
| Mali | PROMIS CS (46) | | | | | | | | | | | | | |
| Zimbabwe | SHINE (HIV−) (47) | 1384 | 1299 | −0.74 | 0.14 (0.06, 0.21) | 0.10 | 0.10 | | 294 | 324 | −0.89 | 0.26 (0.10, 0.42) | 0.05 | 0.07 |
| Zimbabwe | SHINE (HIV+) (48) | 236 | 238 | −0.94 | 0.02 (−0.19, 0.23) | 0.01 | 0.04 | | 85 | 76 | −0.99 | 0.01 (−0.30, 0.32) | 0.01 | 0.02 |
| | | **10207** | **12969** | | **I² = 0.58, Tau² = 0.00** | | | | **4692** | **4369** | | **I² = 0.52, Tau² = 0.00** | | |
| **Fixed** | | | | | **0.14 (0.11, 0.16)** | | | | | | | **0.14 (0.10, 0.17)** | | |
| **Random** | | | | | **0.13 (0.08, 0.18)** | | | | | | | **0.13 (0.08, 0.18)** | | |

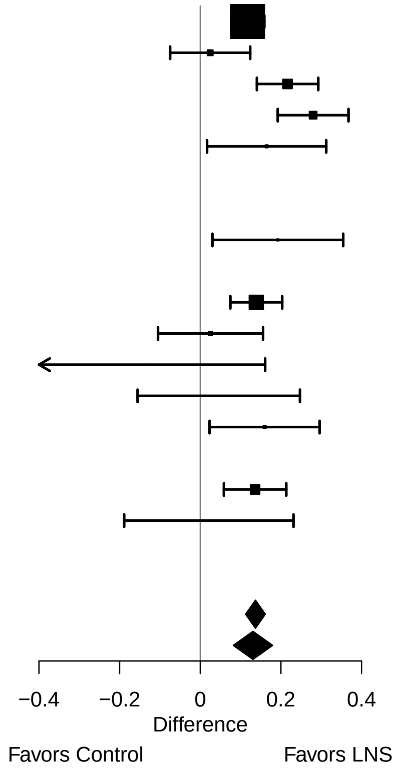

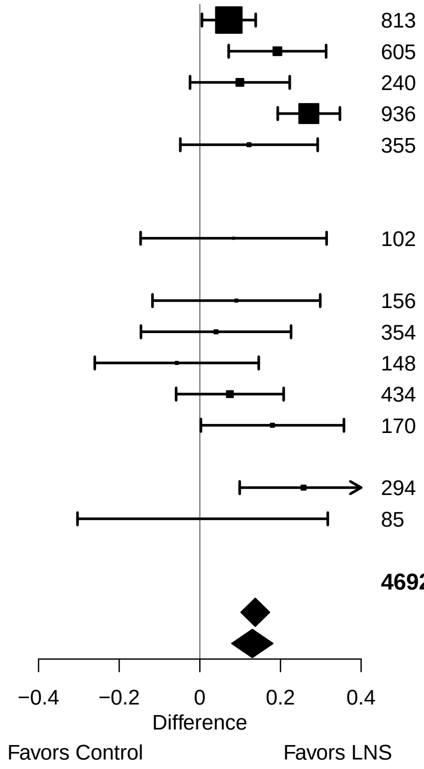

#### Supplemental figure 7L: Mean difference in WAZ

#### 7L3: Stratified by Household source water quality

**P−for−interaction = 0.359**
**Difference in MDs = 0.03 (−0.03, 0.08)**

| | | Improved | | | | | | Unimproved | | | | | |
|---|---|---|---|---|---|---|---|---|---|---|---|---|---|
| Country | Trial | LNS N | Control N | Control Mean | MD (95% CI) | Fixed W | Random W | LNS N | Control N | Control Mean | MD (95% CI) | Fixed W | Random W |
| Bangladesh | JiVitA−4 (34) | | | | | | | | | | | | |
| Bangladesh | RDNS (35) | | | | | | | | | | | | |
| Bangladesh | WASH−B (36) | 501 | 1002 | −1.53 | 0.29 (0.17, 0.41) | 0.10 | 0.10 | 70 | 116 | −1.67 | 0.03 (−0.28, 0.34) | 0.02 | 0.05 |
| Burkina Faso | iLiNS−Zinc (37) | 555 | 141 | −1.45 | 0.21 (0.12, 0.29) | 0.21 | 0.13 | 1397 | 521 | −1.55 | 0.30 (0.23, 0.37) | 0.39 | 0.17 |
| Burkina Faso | PROMIS (38) | 352 | 379 | −1.21 | 0.07 (−0.08, 0.22) | 0.06 | 0.08 | 358 | 351 | −1.23 | 0.25 (0.06, 0.45) | 0.05 | 0.09 |
| Burkina Faso | PROMIS CS (38) | 280 | 267 | −1.30 | −0.01 (−0.21, 0.20) | 0.03 | 0.05 | 149 | 169 | −1.21 | −0.11 (−0.31, 0.08) | 0.05 | 0.09 |
| Ghana | GHANA (39) | | | | | | | | | | | | |
| Ghana | iLiNS−DYADG (40) | | | | | | | | | | | | |
| Haiti | HAITI (41) | | | | | | | | | | | | |
| Kenya | WASH−B (42) | 449 | 1555 | −0.71 | 0.09 (−0.02, 0.19) | 0.13 | 0.11 | 248 | 699 | −0.78 | 0.09 (−0.07, 0.25) | 0.08 | 0.10 |
| Madagascar | MAHAY (43) | 418 | 454 | −1.30 | 0.02 (−0.18, 0.22) | 0.04 | 0.06 | 1204 | 1182 | −1.50 | 0.02 (−0.09, 0.14) | 0.16 | 0.14 |
| Malawi | iLiNS−DYADM (44) | 200 | 406 | −0.86 | −0.09 (−0.26, 0.09) | 0.05 | 0.07 | 20 | 36 | −1.00 | 0.12 (−0.49, 0.73) | 0.01 | 0.01 |
| Malawi | iLiNS−DOSE (45) | 561 | 193 | −1.08 | 0.06 (−0.05, 0.18) | 0.11 | 0.10 | 52 | 17 | −1.41 | 0.02 (−0.37, 0.41) | 0.01 | 0.03 |
| Mali | PROMIS (46) | 247 | 257 | −1.07 | 0.18 (0.05, 0.32) | 0.08 | 0.09 | 243 | 240 | −1.22 | 0.14 (−0.03, 0.30) | 0.07 | 0.10 |
| Mali | PROMIS CS (46) | 544 | 563 | −1.30 | 0.20 (0.07, 0.33) | 0.09 | 0.09 | 363 | 382 | −1.40 | 0.25 (0.05, 0.45) | 0.05 | 0.08 |
| Zimbabwe | SHINE (HIV−) (47) | 518 | 486 | −0.77 | 0.14 (0.01, 0.27) | 0.09 | 0.09 | 273 | 301 | −0.67 | 0.03 (−0.11, 0.17) | 0.10 | 0.12 |
| Zimbabwe | SHINE (HIV+) (48) | 85 | 87 | −1.00 | 0.07 (−0.23, 0.38) | 0.02 | 0.03 | 55 | 56 | −1.00 | 0.02 (−0.37, 0.40) | 0.01 | 0.03 |
| | | **4710** | **5790** | | **I² = 0.52, Tau² = 0.01** | | | **4432** | **4070** | | **I² = 0.69, Tau² = 0.01** | | |
| **Fixed** | | | | | **0.14 (0.10, 0.17)** | | | | | | **0.16 (0.12, 0.20)** | | |
| **Random** | | | | | **0.12 (0.06, 0.18)** | | | | | | **0.12 (0.04, 0.19)** | | |

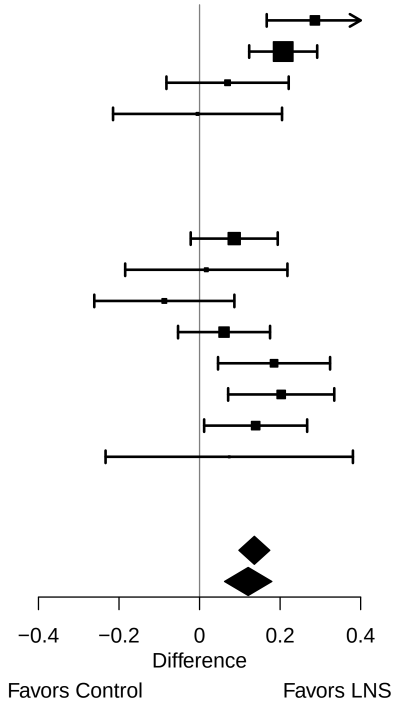

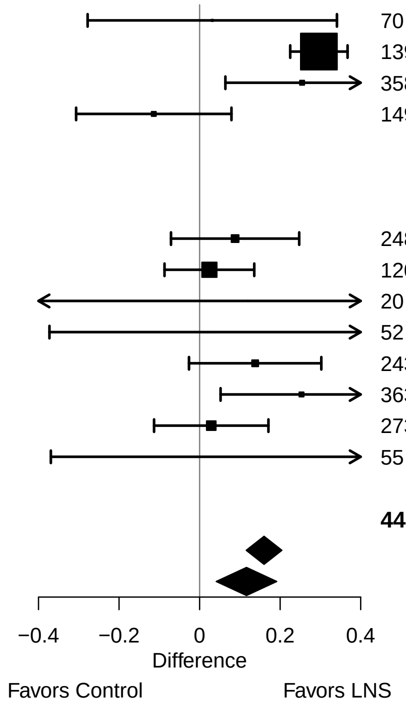

#### Supplemental figure 7L: Mean difference in WAZ

#### 7L4: Stratified by Household sanitation

**P−for−interaction = 0.035**
**Difference in MDs = −0.06 (−0.11, 0.00)**

| Country | Trial | Improved LNS N | Improved Control N | Improved Control Mean | Improved MD (95% CI) | Improved Fixed W | Improved Random W | Unimproved LNS N | Unimproved Control N | Unimproved Control Mean | Unimproved MD (95% CI) | Unimproved Fixed W | Unimproved Random W |
|---|---|---|---|---|---|---|---|---|---|---|---|---|---|
| Bangladesh | JiVitA−4 (34) | 2179 | 944 | −1.72 | 0.12 (0.08, 0.16) | 0.54 | 0.54 | 634 | 288 | −1.92 | 0.06 (−0.01, 0.13) | 0.23 | 0.11 |
| Bangladesh | RDNS (35) | 1179 | 573 | −1.71 | 0.13 (0.03, 0.22) | 0.10 | 0.10 | 482 | 241 | −1.85 | 0.02 (−0.13, 0.17) | 0.05 | 0.08 |
| Bangladesh | WASH−B (36) | 512 | 1015 | −1.51 | 0.25 (0.13, 0.38) | 0.05 | 0.05 | 30 | 55 | −1.73 | 0.42 (0.07, 0.77) | 0.01 | 0.03 |
| Burkina Faso | iLiNS−Zinc (37) | 43 | 16 | −1.62 | 0.23 (−0.14, 0.60) | 0.01 | 0.01 | 1909 | 646 | −1.53 | 0.28 (0.21, 0.35) | 0.25 | 0.11 |
| Burkina Faso | PROMIS (38) | 392 | 550 | −1.24 | 0.18 (0.06, 0.30) | 0.06 | 0.06 | 465 | 357 | −1.19 | 0.11 (−0.06, 0.27) | 0.04 | 0.07 |
| Burkina Faso | PROMIS CS (38) | 176 | 165 | −1.31 | 0.03 (−0.18, 0.25) | 0.02 | 0.02 | 252 | 271 | −1.24 | −0.10 (−0.26, 0.07) | 0.04 | 0.07 |
| Ghana | GHANA (39) | | | | | | | | | | | | |
| Ghana | iLiNS−DYADG (40) | 336 | 672 | −0.84 | 0.14 (0.01, 0.28) | 0.05 | 0.05 | 10 | 19 | −1.13 | 0.66 (−0.24, 1.56) | 0.00 | 0.01 |
| Haiti | HAITI (41) | | | | | | | | | | | | |
| Kenya | WASH−B (42) | 101 | 366 | −0.55 | 0.16 (−0.02, 0.34) | 0.03 | 0.03 | 596 | 1889 | −0.76 | 0.07 (−0.03, 0.18) | 0.11 | 0.10 |
| Madagascar | MAHAY (43) | | | | | | | | | | | | |
| Malawi | iLiNS−DYADM (44) | 20 | 42 | −0.76 | 0.27 (−0.21, 0.75) | 0.00 | 0.00 | 200 | 400 | −0.89 | −0.10 (−0.28, 0.07) | 0.03 | 0.07 |
| Malawi | iLiNS−DOSE (45) | 15 | 9 | −1.02 | −0.20 (−0.76, 0.36) | 0.00 | 0.00 | 597 | 201 | −1.11 | 0.06 (−0.05, 0.17) | 0.09 | 0.09 |
| Mali | PROMIS (46) | 381 | 363 | −1.12 | 0.17 (0.03, 0.30) | 0.05 | 0.05 | 112 | 134 | −1.23 | 0.16 (−0.01, 0.33) | 0.04 | 0.07 |
| Mali | PROMIS CS (46) | 698 | 692 | −1.34 | 0.26 (0.12, 0.40) | 0.04 | 0.04 | 217 | 243 | −1.34 | 0.19 (0.01, 0.37) | 0.03 | 0.07 |
| Zimbabwe | SHINE (HIV−) (47) | 293 | 243 | −0.68 | 0.14 (−0.03, 0.32) | 0.03 | 0.03 | 495 | 543 | −0.76 | 0.07 (−0.05, 0.19) | 0.08 | 0.09 |
| Zimbabwe | SHINE (HIV+) (48) | 50 | 35 | −0.95 | −0.08 (−0.50, 0.34) | 0.00 | 0.00 | 90 | 107 | −1.01 | 0.11 (−0.18, 0.40) | 0.01 | 0.04 |
| | | **6375** | **5685** | | **$I^2$ = 0.00, $Tau^2$ = 0.00** | | | **6089** | **5394** | | **$I^2$ = 0.72, $Tau^2$ = 0.01** | | |
| **Fixed** | | | | | **0.14 (0.11, 0.17)** | | | | | | **0.12 (0.08, 0.15)** | | |
| **Random** | | | | | **0.14 (0.11, 0.17)** | | | | | | **0.09 (0.03, 0.16)** | | |

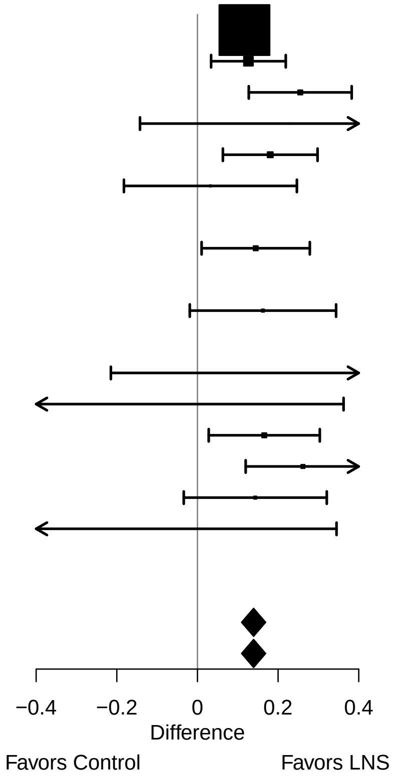

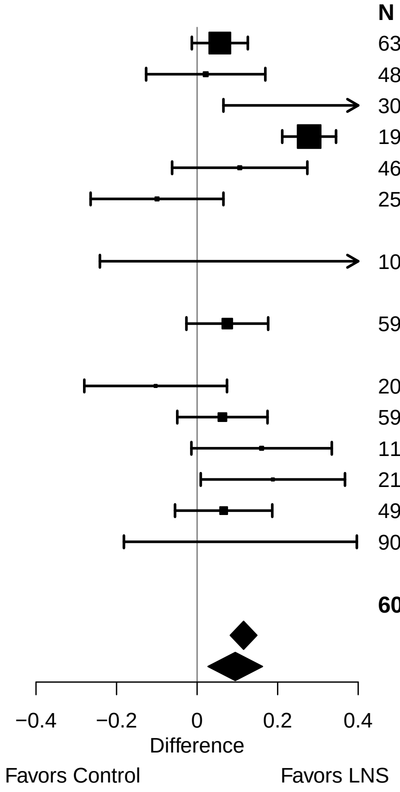

#### Supplemental figure 7L: Mean difference in WAZ

#### 7L5: Stratified by Home environment

**P−for−interaction = 0.076**
**Difference in MDs = 0.06 (−0.01, 0.12)**

| | | At least median | | | | | | Less than median | | | | | |
|---|---|---|---|---|---|---|---|---|---|---|---|---|---|
| **Country** | **Trial** | **LNS N** | **Control N** | **Control Mean** | **MD (95% CI)** | **Fixed W** | **Random W** | **LNS N** | **Control N** | **Control Mean** | **MD (95% CI)** | **Fixed W** | **Random W** |
| Bangladesh | JiVitA−4 (34) | | | | | | | | | | | | |
| Bangladesh | RDNS (35) | 1192 | 582 | −1.70 | 0.09 (−0.01, 0.20) | 0.13 | 0.14 | 468 | 230 | −1.86 | 0.09 (−0.06, 0.25) | 0.11 | 0.13 |
| Bangladesh | WASH−B (36) | 641 | 1668 | −1.41 | 0.19 (0.09, 0.28) | 0.17 | 0.15 | 494 | 1702 | −1.69 | 0.15 (0.04, 0.26) | 0.22 | 0.17 |
| Burkina Faso | iLiNS−Zinc (37) | 473 | 199 | −1.44 | 0.26 (0.15, 0.36) | 0.14 | 0.14 | 272 | 175 | −1.68 | 0.24 (0.05, 0.42) | 0.08 | 0.11 |
| Burkina Faso | PROMIS (38) | | | | | | | | | | | | |
| Burkina Faso | PROMIS CS (38) | | | | | | | | | | | | |
| Ghana | GHANA (39) | | | | | | | | | | | | |
| Ghana | iLiNS−DYADG (40) | 213 | 410 | −0.83 | 0.12 (−0.05, 0.29) | 0.05 | 0.10 | 119 | 248 | −0.93 | 0.29 (0.06, 0.52) | 0.05 | 0.08 |
| Haiti | HAITI (41) | | | | | | | | | | | | |
| Kenya | WASH−B (42) | 840 | 2776 | −0.67 | 0.04 (−0.03, 0.11) | 0.30 | 0.16 | 573 | 2167 | −0.80 | 0.20 (0.11, 0.29) | 0.32 | 0.20 |
| Madagascar | MAHAY (43) | 915 | 907 | −1.40 | −0.03 (−0.15, 0.10) | 0.09 | 0.12 | 787 | 778 | −1.49 | 0.06 (−0.09, 0.21) | 0.12 | 0.14 |
| Malawi | iLiNS−DYADM (44) | 137 | 289 | −0.80 | −0.08 (−0.28, 0.13) | 0.04 | 0.08 | 78 | 149 | −1.03 | −0.05 (−0.35, 0.24) | 0.03 | 0.06 |
| Malawi | iLiNS−DOSE (45) | 379 | 136 | −1.18 | 0.17 (0.03, 0.30) | 0.08 | 0.12 | 277 | 86 | −1.00 | −0.07 (−0.26, 0.11) | 0.08 | 0.11 |
| Mali | PROMIS (46) | | | | | | | | | | | | |
| Mali | PROMIS CS (46) | | | | | | | | | | | | |
| Zimbabwe | SHINE (HIV−) (47) | | | | | | | | | | | | |
| Zimbabwe | SHINE (HIV+) (48) | | | | | | | | | | | | |
| | | **4790** | **6967** | | **I² = 0.69, Tau² = 0.01** | | | **3068** | **5535** | | **I² = 0.46, Tau² = 0.01** | | |
| **Fixed** | | | | | **0.11 (0.07, 0.15)** | | | | | | **0.14 (0.09, 0.19)** | | |
| **Random** | | | | | **0.10 (0.03, 0.18)** | | | | | | **0.12 (0.04, 0.20)** | | |

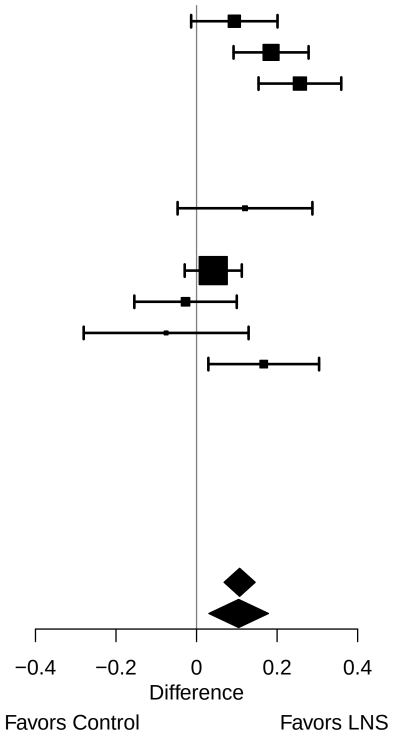

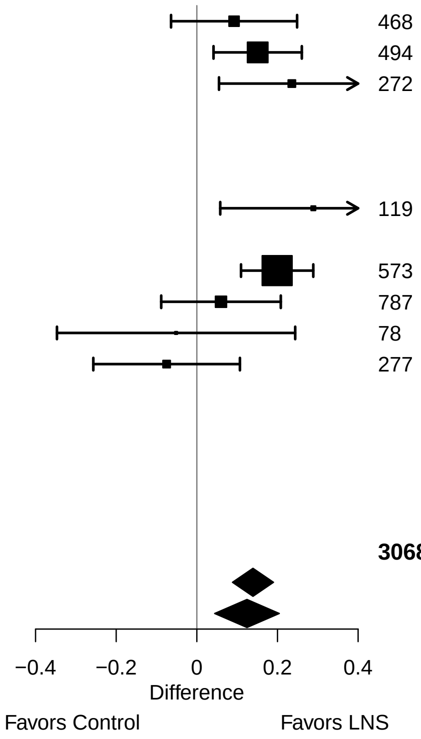

#### Supplemental figure 7L: Mean difference in WAZ

#### 7L6: Stratified by Season at the time of assessment

**P−for−interaction = 0.212**
**Difference in MDs = −0.03 (−0.07, 0.02)**

| Country | Trial | Dry LNS N | Dry Control N | Dry Control Mean | Dry MD (95% CI) | Dry Fixed W | Dry Random W | Rainy LNS N | Rainy Control N | Rainy Control Mean | Rainy MD (95% CI) | Rainy Fixed W | Rainy Random W |
|---|---|---|---|---|---|---|---|---|---|---|---|---|---|
| Bangladesh | JiVitA−4 (34) | 1533 | 656 | −1.71 | 0.11 (0.06, 0.16) | 0.33 | 0.11 | 1281 | 576 | −1.84 | 0.10 (0.04, 0.15) | 0.34 | 0.14 |
| Bangladesh | RDNS (35) | 739 | 359 | −1.71 | 0.11 (−0.04, 0.25) | 0.04 | 0.08 | 924 | 456 | −1.78 | 0.09 (−0.01, 0.18) | 0.10 | 0.12 |
| Bangladesh | WASH−B (36) | 645 | 1938 | −1.42 | 0.19 (0.09, 0.30) | 0.08 | 0.09 | 518 | 1523 | −1.72 | 0.21 (0.13, 0.28) | 0.15 | 0.13 |
| Burkina Faso | iLiNS−Zinc (37) | 1400 | 477 | −1.57 | 0.32 (0.24, 0.40) | 0.12 | 0.10 | 560 | 189 | −1.43 | 0.17 (0.07, 0.28) | 0.09 | 0.11 |
| Burkina Faso | PROMIS (38) | 429 | 447 | −1.14 | 0.10 (−0.04, 0.23) | 0.05 | 0.08 | 428 | 464 | −1.30 | 0.19 (0.03, 0.36) | 0.03 | 0.08 |
| Burkina Faso | PROMIS CS (38) | 224 | 190 | −1.23 | −0.02 (−0.27, 0.24) | 0.01 | 0.05 | 206 | 246 | −1.29 | −0.09 (−0.29, 0.11) | 0.02 | 0.06 |
| Ghana | GHANA (39) | 57 | 55 | −0.84 | 0.61 (0.20, 1.01) | 0.01 | 0.02 | 41 | 41 | −0.46 | −0.16 (−0.70, 0.39) | 0.00 | 0.01 |
| Ghana | iLiNS−DYADG (40) | 204 | 408 | −0.87 | 0.04 (−0.13, 0.20) | 0.03 | 0.07 | 143 | 285 | −0.83 | 0.34 (0.12, 0.56) | 0.02 | 0.06 |
| Haiti | HAITI (41) | | | | | | | | | | | | |
| Kenya | WASH−B (42) | 348 | 1157 | −0.80 | 0.19 (0.08, 0.30) | 0.07 | 0.09 | 1113 | 4008 | −0.74 | 0.12 (0.04, 0.19) | 0.17 | 0.13 |
| Madagascar | MAHAY (43) | | | | | | | | | | | | |
| Malawi | iLiNS−DYADM (44) | 156 | 316 | −0.82 | −0.04 (−0.23, 0.16) | 0.02 | 0.06 | 64 | 128 | −1.02 | −0.14 (−0.45, 0.18) | 0.01 | 0.03 |
| Malawi | iLiNS−DOSE (45) | 421 | 134 | −1.02 | 0.00 (−0.14, 0.15) | 0.04 | 0.08 | 281 | 107 | −1.17 | 0.11 (−0.04, 0.26) | 0.04 | 0.08 |
| Mali | PROMIS (46) | | | | | | | | | | | | |
| Mali | PROMIS CS (46) | | | | | | | | | | | | |
| Zimbabwe | SHINE (HIV−) (47) | 1736 | 1655 | −0.77 | 0.15 (0.09, 0.22) | 0.17 | 0.10 | 133 | 131 | −0.51 | −0.14 (−0.43, 0.15) | 0.01 | 0.04 |
| Zimbabwe | SHINE (HIV+) (48) | 318 | 304 | −0.94 | 0.00 (−0.17, 0.18) | 0.03 | 0.07 | 18 | 24 | −1.01 | 0.13 (−0.42, 0.68) | 0.00 | 0.01 |
| | | **8210** | **8096** | | **I² = 0.70, Tau² = 0.01** | | | **5710** | **8178** | | **I² = 0.47, Tau² = 0.01** | | |
| **Fixed** | | | | | **0.14 (0.11, 0.17)** | | | | | | **0.12 (0.09, 0.15)** | | |
| **Random** | | | | | **0.13 (0.05, 0.20)** | | | | | | **0.11 (0.05, 0.17)** | | |

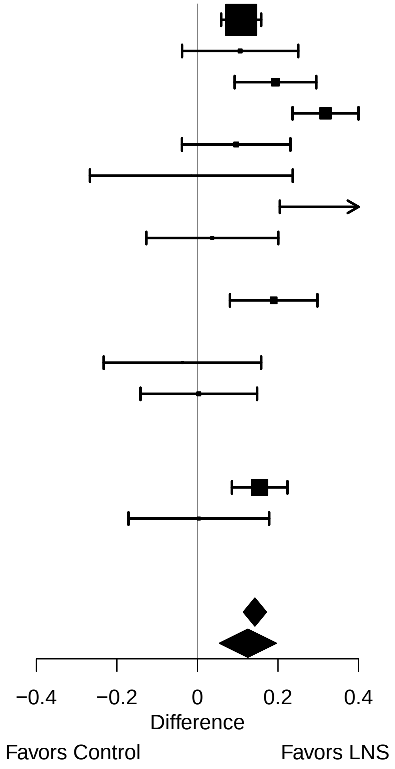

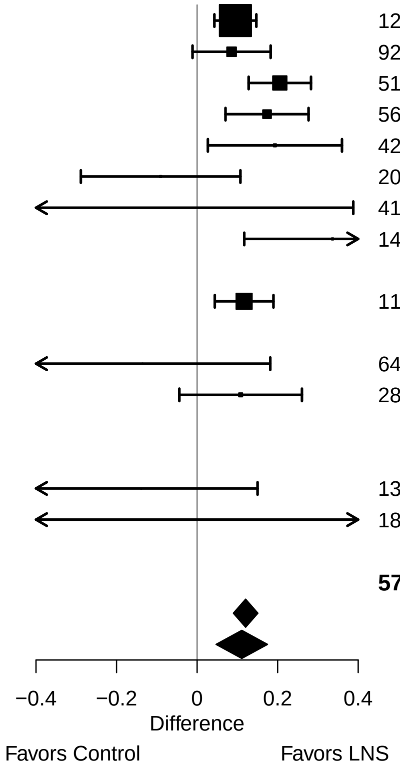

### Supplemental figure 7M: Underweight prevalence ratio

#### 7M1: Stratified by Household socio-economic status

**P−for−interaction = 0.877**
**Ratio of PRs = 1.01 (0.93, 1.09)**

| | | At least median | | | | | | Less than median | | | | | |
|---|---|---|---|---|---|---|---|---|---|---|---|---|---|
| **Country** | **Trial** | **LNS N** | **Control N** | **Control Prevalence** | **PR (95% CI)** | **Fixed W** | **Random W** | **LNS N** | **Control N** | **Control Prevalence** | **PR (95% CI)** | **Fixed W** | **Random W** |
| Bangladesh | JiVitA−4 (34) | 1421 | 619 | 35.4 | 0.81 (0.71, 0.93) | 0.24 | 0.16 | 1392 | 613 | 42.9 | 0.97 (0.86, 1.08) | 0.22 | 0.16 |
| Bangladesh | RDNS (35) | 834 | 407 | 32.2 | 0.91 (0.77, 1.06) | 0.16 | 0.13 | 829 | 408 | 43.4 | 0.93 (0.83, 1.05) | 0.23 | 0.17 |
| Bangladesh | WASH−B (36) | 584 | 1741 | 23.9 | 0.85 (0.70, 1.03) | 0.11 | 0.11 | 579 | 1720 | 41.0 | 0.75 (0.65, 0.86) | 0.15 | 0.13 |
| Burkina Faso | iLiNS−Zinc (37) | 1233 | 300 | 28.0 | 0.72 (0.57, 0.90) | 0.08 | 0.09 | 720 | 365 | 33.2 | 0.69 (0.57, 0.83) | 0.08 | 0.09 |
| Burkina Faso | PROMIS (38) | 405 | 482 | 19.9 | 0.81 (0.59, 1.11) | 0.04 | 0.05 | 452 | 429 | 19.3 | 0.91 (0.67, 1.24) | 0.03 | 0.04 |
| Burkina Faso | PROMIS CS (38) | 220 | 208 | 21.6 | 1.07 (0.79, 1.45) | 0.04 | 0.06 | 210 | 228 | 26.8 | 0.93 (0.68, 1.26) | 0.03 | 0.04 |
| Ghana | GHANA (39) | | | | | | | | | | | | |
| Ghana | iLiNS−DYADG (40) | 157 | 356 | 11.0 | 0.81 (0.46, 1.46) | 0.01 | 0.02 | 190 | 335 | 14.9 | 0.81 (0.51, 1.29) | 0.01 | 0.02 |
| Haiti | HAITI (41) | | | | | | | | | | | | |
| Kenya | WASH−B (42) | 828 | 2941 | 8.0 | 0.68 (0.50, 0.91) | 0.05 | 0.06 | 631 | 2220 | 12.7 | 0.83 (0.63, 1.08) | 0.04 | 0.05 |
| Madagascar | MAHAY (43) | 772 | 884 | 23.2 | 1.07 (0.87, 1.30) | 0.10 | 0.10 | 877 | 758 | 29.6 | 0.97 (0.79, 1.20) | 0.07 | 0.08 |
| Malawi | iLiNS−DYADM (44) | 121 | 234 | 9.0 | 1.38 (0.74, 2.58) | 0.01 | 0.02 | 99 | 209 | 14.4 | 0.99 (0.55, 1.77) | 0.01 | 0.01 |
| Malawi | iLiNS−DOSE (45) | 307 | 104 | 10.6 | 1.28 (0.73, 2.25) | 0.01 | 0.02 | 294 | 103 | 30.1 | 0.91 (0.67, 1.24) | 0.03 | 0.05 |
| Mali | PROMIS (46) | 247 | 250 | 15.5 | 0.75 (0.53, 1.06) | 0.03 | 0.05 | 245 | 246 | 17.1 | 0.88 (0.52, 1.49) | 0.01 | 0.02 |
| Mali | PROMIS CS (46) | 501 | 458 | 21.4 | 0.90 (0.69, 1.16) | 0.06 | 0.07 | 445 | 502 | 23.5 | 0.88 (0.67, 1.16) | 0.04 | 0.05 |
| Zimbabwe | SHINE (HIV−) (47) | 903 | 797 | 9.0 | 0.76 (0.55, 1.05) | 0.04 | 0.05 | 826 | 858 | 12.5 | 0.62 (0.46, 0.84) | 0.03 | 0.05 |
| Zimbabwe | SHINE (HIV+) (48) | 171 | 156 | 14.7 | 1.03 (0.61, 1.74) | 0.01 | 0.02 | 157 | 166 | 19.9 | 0.96 (0.59, 1.56) | 0.01 | 0.02 |
| | | **8704** | **9937** | | **I² = 0.25, Tau² = 0.01** | | | **7946** | **9160** | | **I² = 0.35, Tau² = 0.00** | | |
| **Fixed** | | | | | **0.86 (0.81, 0.92)** | | | | | | **0.87 (0.82, 0.92)** | | |
| **Random** | | | | | **0.87 (0.80, 0.94)** | | | | | | **0.86 (0.80, 0.92)** | | |

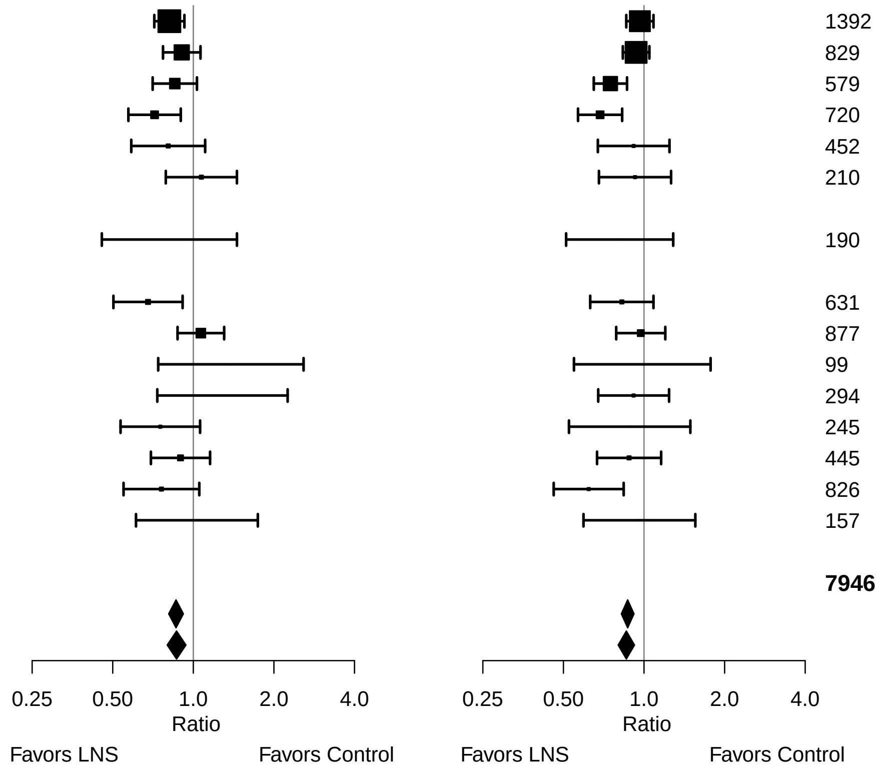

#### Supplemental figure 7M: Underweight prevalence ratio

#### 7M2: Stratified by Household food insecurity

**P−for−interaction = 0.661**
**Ratio of PRs = 0.98 (0.89, 1.08)**

| Country | Trial | LNS N | Control N | Control Prevalence | Mild to secure PR (95% CI) | Fixed W | Random W | LNS N | Control N | Control Prevalence | Moderate to severe PR (95% CI) | Fixed W | Random W |
|---|---|---|---|---|---|---|---|---|---|---|---|---|---|
| Bangladesh | JiVitA−4 (34) | 2000 | 851 | 37.0 | 0.86 (0.77, 0.95) | 0.27 | 0.16 | 813 | 381 | 43.9 | 0.97 (0.83, 1.13) | 0.24 | 0.16 |
| Bangladesh | RDNS (35) | 1058 | 493 | 32.9 | 1.00 (0.86, 1.15) | 0.14 | 0.13 | 605 | 322 | 45.3 | 0.85 (0.73, 0.99) | 0.22 | 0.16 |
| Bangladesh | WASH−B (36) | 923 | 2681 | 30.8 | 0.76 (0.67, 0.87) | 0.18 | 0.14 | 240 | 780 | 38.1 | 0.89 (0.72, 1.09) | 0.13 | 0.13 |
| Burkina Faso | iLiNS−Zinc (37) | 1017 | 321 | 27.4 | 0.71 (0.60, 0.85) | 0.10 | 0.11 | 936 | 344 | 34.0 | 0.67 (0.55, 0.81) | 0.15 | 0.14 |
| Burkina Faso | PROMIS (38) | 502 | 550 | 19.3 | 0.95 (0.74, 1.22) | 0.05 | 0.07 | 355 | 361 | 20.2 | 0.74 (0.51, 1.07) | 0.04 | 0.06 |
| Burkina Faso | PROMIS CS (38) | | | | | | | | | | | | |
| Ghana | GHANA (39) | | | | | | | | | | | | |
| Ghana | iLiNS−DYADG (40) | 244 | 471 | 12.7 | 0.77 (0.49, 1.21) | 0.02 | 0.03 | 102 | 219 | 13.7 | 0.93 (0.51, 1.71) | 0.02 | 0.03 |
| Haiti | HAITI (41) | | | | | | | | | | | | |
| Kenya | WASH−B (42) | 1303 | 4621 | 9.8 | 0.77 (0.62, 0.95) | 0.07 | 0.09 | 156 | 527 | 12.1 | 0.69 (0.34, 1.37) | 0.01 | 0.02 |
| Madagascar | MAHAY (43) | 989 | 929 | 25.6 | 1.03 (0.86, 1.24) | 0.09 | 0.10 | 354 | 410 | 27.3 | 1.03 (0.75, 1.43) | 0.05 | 0.08 |
| Malawi | iLiNS−DYADM (44) | 70 | 122 | 8.2 | 1.57 (0.67, 3.67) | 0.00 | 0.01 | 148 | 316 | 12.7 | 1.07 (0.65, 1.76) | 0.02 | 0.04 |
| Malawi | iLiNS−DOSE (45) | 154 | 55 | 14.5 | 1.00 (0.55, 1.85) | 0.01 | 0.02 | 434 | 146 | 21.9 | 1.03 (0.75, 1.42) | 0.05 | 0.08 |
| Mali | PROMIS (46) | 327 | 338 | 15.2 | 0.88 (0.59, 1.32) | 0.02 | 0.03 | 170 | 163 | 19.0 | 0.80 (0.50, 1.29) | 0.02 | 0.04 |
| Mali | PROMIS CS (46) | | | | | | | | | | | | |
| Zimbabwe | SHINE (HIV−) (47) | 1384 | 1299 | 9.8 | 0.75 (0.57, 0.97) | 0.04 | 0.07 | 294 | 324 | 15.4 | 0.48 (0.31, 0.77) | 0.03 | 0.04 |
| Zimbabwe | SHINE (HIV+) (48) | 236 | 238 | 17.6 | 1.03 (0.71, 1.50) | 0.02 | 0.04 | 85 | 76 | 17.1 | 0.89 (0.45, 1.76) | 0.01 | 0.02 |
| | | **10207** | **12969** | | **$I^2$ = 0.45, $Tau^2$ = 0.01** | | | **4692** | **4369** | | **$I^2$ = 0.38, $Tau^2$ = 0.01** | | |
| **Fixed** | | | | | **0.85 (0.81, 0.90)** | | | | | | **0.85 (0.79, 0.92)** | | |
| **Random** | | | | | **0.86 (0.79, 0.93)** | | | | | | **0.85 (0.76, 0.94)** | | |

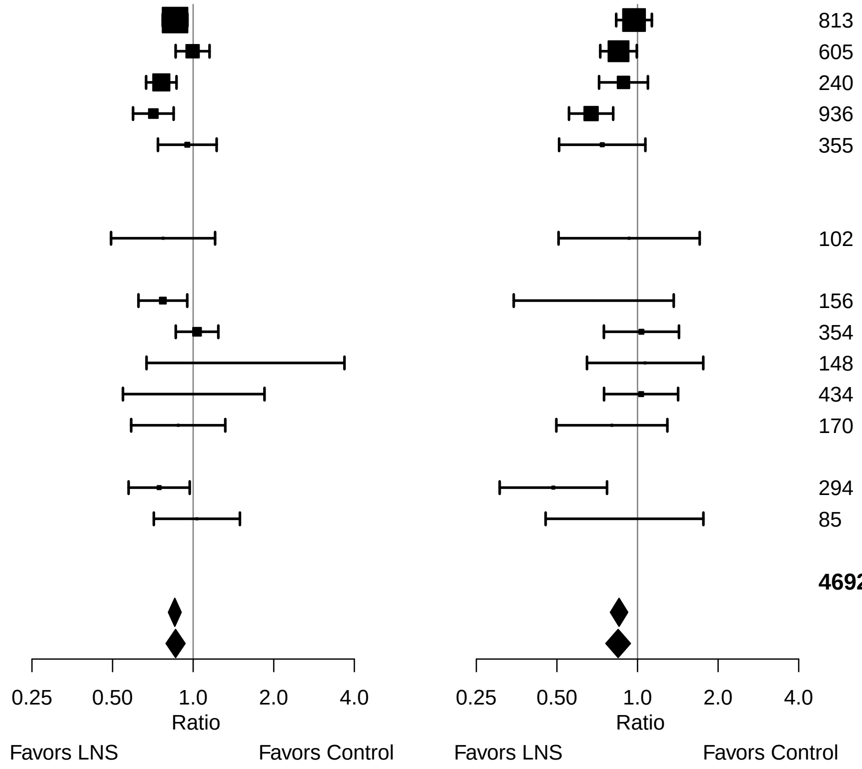

#### Supplemental figure 7M: Underweight prevalence ratio

### 7M3: Stratified by Household source water quality

P−for−interaction = 0.762
Ratio of PRs = 0.98 (0.86, 1.12)

| Country | Trial | Improved LNS N | Improved Control N | Improved Control Prevalence | Improved PR (95% CI) | Improved Fixed W | Improved Random W | Unimproved LNS N | Unimproved Control N | Unimproved Control Prevalence | Unimproved PR (95% CI) | Unimproved Fixed W | Unimproved Random W |
|---|---|---|---|---|---|---|---|---|---|---|---|---|---|
| Bangladesh | JiVitA−4 (34) | | | | | | | | | | | | |
| Bangladesh | RDNS (35) | | | | | | | | | | | | |
| Bangladesh | WASH−B (36) | 501 | 1002 | 30.7 | 0.81 (0.68, 0.97) | 0.28 | 0.19 | 70 | 116 | 29.3 | 1.12 (0.72, 1.74) | 0.04 | 0.08 |
| Burkina Faso | iLiNS−Zinc (37) | 555 | 141 | 27.7 | 0.66 (0.52, 0.85) | 0.14 | 0.13 | 1397 | 521 | 31.9 | 0.69 (0.59, 0.82) | 0.30 | 0.17 |
| Burkina Faso | PROMIS (38) | 352 | 379 | 17.9 | 1.01 (0.75, 1.36) | 0.10 | 0.11 | 358 | 351 | 23.4 | 0.57 (0.42, 0.79) | 0.08 | 0.11 |
| Burkina Faso | PROMIS CS (38) | 280 | 267 | 27.3 | 0.82 (0.61, 1.11) | 0.10 | 0.11 | 149 | 169 | 19.5 | 1.37 (0.95, 1.99) | 0.06 | 0.10 |
| Ghana | GHANA (39) | | | | | | | | | | | | |
| Ghana | iLiNS−DYADG (40) | | | | | | | | | | | | |
| Haiti | HAITI (41) | | | | | | | | | | | | |
| Kenya | WASH−B (42) | 449 | 1555 | 8.8 | 0.94 (0.65, 1.34) | 0.07 | 0.08 | 248 | 699 | 10.7 | 0.83 (0.50, 1.36) | 0.03 | 0.07 |
| Madagascar | MAHAY (43) | 418 | 454 | 20.5 | 1.26 (0.95, 1.67) | 0.11 | 0.12 | 1204 | 1182 | 28.1 | 0.96 (0.82, 1.13) | 0.31 | 0.17 |
| Malawi | iLiNS−DYADM (44) | | | | | | | | | | | | |
| Malawi | iLiNS−DOSE (45) | | | | | | | | | | | | |
| Mali | PROMIS (46) | 247 | 257 | 14.3 | 0.80 (0.47, 1.36) | 0.03 | 0.05 | 243 | 240 | 18.5 | 0.88 (0.57, 1.36) | 0.04 | 0.08 |
| Mali | PROMIS CS (46) | 544 | 563 | 21.5 | 0.88 (0.66, 1.17) | 0.11 | 0.12 | 363 | 382 | 23.6 | 0.91 (0.67, 1.24) | 0.09 | 0.12 |
| Zimbabwe | SHINE (HIV−) (47) | 518 | 486 | 9.7 | 0.76 (0.50, 1.14) | 0.05 | 0.07 | 273 | 301 | 8.6 | 1.02 (0.62, 1.68) | 0.03 | 0.07 |
| Zimbabwe | SHINE (HIV+) (48) | 85 | 87 | 13.8 | 1.28 (0.64, 2.54) | 0.02 | 0.03 | 55 | 56 | 21.4 | 0.85 (0.42, 1.73) | 0.02 | 0.04 |
| | | **3949** | **5191** | | **I² = 0.39, Tau² = 0.01** | | | **4360** | **4017** | | **I² = 0.61, Tau² = 0.03** | | |
| **Fixed** | | | | | **0.87 (0.79, 0.95)** | | | | | | **0.85 (0.78, 0.93)** | | |
| **Random** | | | | | **0.88 (0.78, 1.00)** | | | | | | **0.88 (0.75, 1.03)** | | |

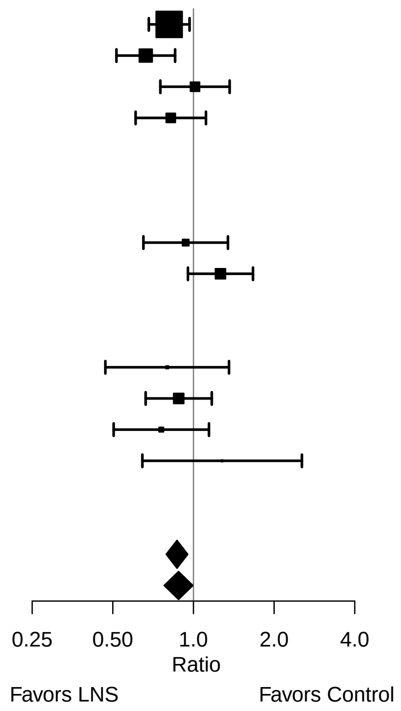

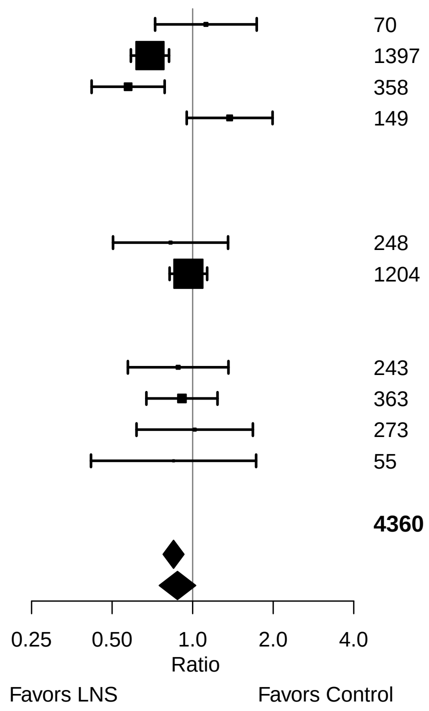

#### Supplemental figure 7M: Underweight prevalence ratio

#### 7M4: Stratified by Household sanitation

**P−for−interaction = 0.840**
**Ratio of PRs = 1.01 (0.89, 1.15)**

| Country | Trial | Improved LNS N | Improved Control N | Improved Control Prevalence | Improved PR (95% CI) | Improved Fixed W | Improved Random W | Unimproved LNS N | Unimproved Control N | Unimproved Control Prevalence | Unimproved PR (95% CI) | Unimproved Fixed W | Unimproved Random W |
|---|---|---|---|---|---|---|---|---|---|---|---|---|---|
| Bangladesh | JiVitA−4 (34) | 2179 | 944 | 37.7 | 0.91 (0.81, 1.02) | 0.37 | 0.37 | 634 | 288 | 43.9 | 0.94 (0.80, 1.09) | 0.37 | 0.37 |
| Bangladesh | RDNS (35) | 1179 | 573 | 34.9 | 0.93 (0.83, 1.04) | 0.41 | 0.41 | 482 | 241 | 44.8 | 0.90 (0.74, 1.10) | 0.22 | 0.22 |
| Bangladesh | WASH−B (36) | | | | | | | | | | | | |
| Burkina Faso | iLiNS−Zinc (37) | | | | | | | | | | | | |
| Burkina Faso | PROMIS (38) | 392 | 550 | 20.0 | 0.85 (0.65, 1.13) | 0.07 | 0.07 | 465 | 357 | 19.3 | 0.87 (0.64, 1.17) | 0.10 | 0.10 |
| Burkina Faso | PROMIS CS (38) | 176 | 165 | 23.0 | 1.04 (0.69, 1.55) | 0.03 | 0.03 | 252 | 271 | 25.1 | 0.96 (0.73, 1.28) | 0.11 | 0.11 |
| Ghana | GHANA (39) | | | | | | | | | | | | |
| Ghana | iLiNS−DYADG (40) | | | | | | | | | | | | |
| Haiti | HAITI (41) | | | | | | | | | | | | |
| Kenya | WASH−B (42) | | | | | | | | | | | | |
| Madagascar | MAHAY (43) | | | | | | | | | | | | |
| Malawi | iLiNS−DYADM (44) | | | | | | | | | | | | |
| Malawi | iLiNS−DOSE (45) | | | | | | | | | | | | |
| Mali | PROMIS (46) | 381 | 363 | 16.1 | 0.86 (0.60, 1.23) | 0.04 | 0.04 | 112 | 134 | 17.0 | 0.90 (0.55, 1.47) | 0.04 | 0.04 |
| Mali | PROMIS CS (46) | 698 | 692 | 22.5 | 0.87 (0.67, 1.14) | 0.07 | 0.07 | 217 | 243 | 21.4 | 0.88 (0.66, 1.18) | 0.10 | 0.10 |
| Zimbabwe | SHINE (HIV−) (47) | 293 | 243 | 7.8 | 0.70 (0.35, 1.38) | 0.01 | 0.01 | 495 | 543 | 9.9 | 0.93 (0.64, 1.36) | 0.06 | 0.06 |
| Zimbabwe | SHINE (HIV+) (48) | | | | | | | | | | | | |
| | | **5298** | **3530** | | **I² = 0.00, Tau² = 0.00** | | | **2657** | **2077** | | **I² = 0.00, Tau² = 0.00** | | |
| **Fixed** | | | | | **0.91 (0.85, 0.98)** | | | | | | **0.92 (0.84, 1.01)** | | |
| **Random** | | | | | **0.91 (0.85, 0.98)** | | | | | | **0.92 (0.84, 1.01)** | | |

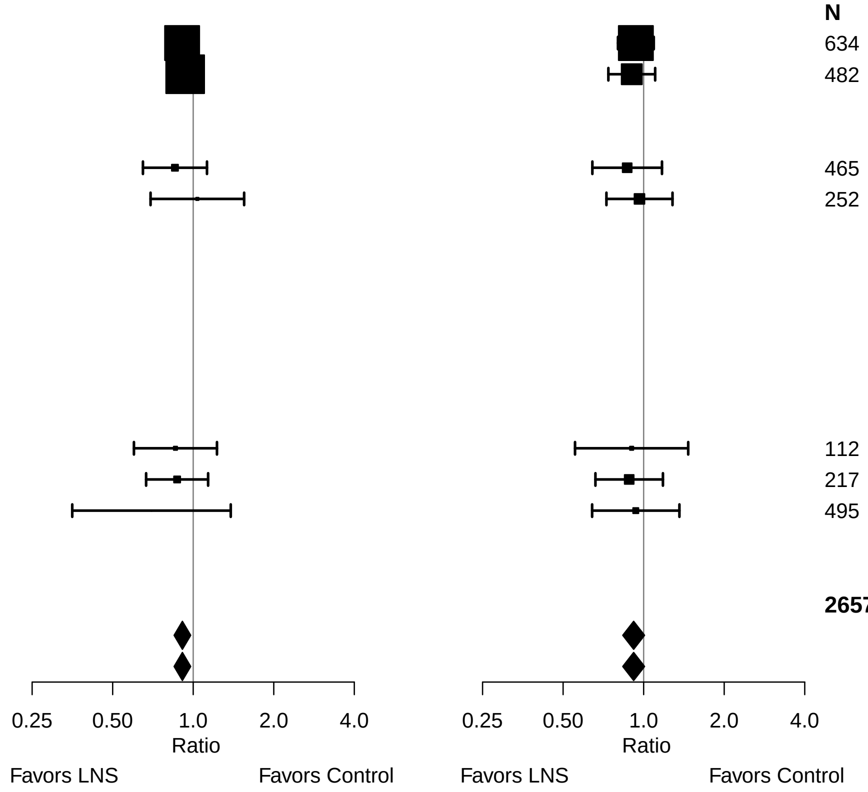

#### Supplemental figure 7M: Underweight prevalence ratio

#### 7M5: Stratified by Home environment

**P−for−interaction = 0.416**
**Ratio of PRs = 1.05 (0.94, 1.17)**

| | | At least median | | | | | | Less than median | | | | | |
|---|---|---|---|---|---|---|---|---|---|---|---|---|---|
| **Country** | **Trial** | **LNS N** | **Control N** | **Control Prevalence** | **PR (95% CI)** | **Fixed W** | **Random W** | **LNS N** | **Control N** | **Control Prevalence** | **PR (95% CI)** | **Fixed W** | **Random W** |
| Bangladesh | JiVitA−4 (34) | | | | | | | | | | | | |
| Bangladesh | RDNS (35) | 1192 | 582 | 36.1 | 0.89 (0.78, 1.01) | 0.35 | 0.25 | 468 | 230 | 41.7 | 1.00 (0.85, 1.18) | 0.28 | 0.22 |
| Bangladesh | WASH−B (36) | 641 | 1668 | 28.1 | 0.80 (0.68, 0.94) | 0.23 | 0.21 | 494 | 1702 | 36.5 | 0.82 (0.69, 0.97) | 0.25 | 0.21 |
| Burkina Faso | iLiNS−Zinc (37) | 473 | 199 | 27.6 | 0.64 (0.49, 0.84) | 0.08 | 0.11 | 272 | 175 | 31.4 | 0.86 (0.67, 1.09) | 0.13 | 0.15 |
| Burkina Faso | PROMIS (38) | | | | | | | | | | | | |
| Burkina Faso | PROMIS CS (38) | | | | | | | | | | | | |
| Ghana | GHANA (39) | | | | | | | | | | | | |
| Ghana | iLiNS−DYADG (40) | 213 | 410 | 12.0 | 0.98 (0.62, 1.54) | 0.03 | 0.05 | 119 | 248 | 15.3 | 0.55 (0.28, 1.06) | 0.02 | 0.03 |
| Haiti | HAITI (41) | | | | | | | | | | | | |
| Kenya | WASH−B (42) | 840 | 2776 | 8.0 | 0.88 (0.67, 1.16) | 0.08 | 0.11 | 573 | 2167 | 10.8 | 0.79 (0.57, 1.09) | 0.07 | 0.10 |
| Madagascar | MAHAY (43) | 915 | 907 | 24.8 | 1.07 (0.89, 1.29) | 0.17 | 0.18 | 787 | 778 | 27.4 | 1.00 (0.82, 1.23) | 0.19 | 0.18 |
| Malawi | iLiNS−DYADM (44) | 137 | 289 | 10.4 | 0.91 (0.49, 1.70) | 0.01 | 0.03 | 78 | 149 | 14.1 | 1.46 (0.81, 2.63) | 0.02 | 0.04 |
| Malawi | iLiNS−DOSE (45) | 379 | 136 | 22.8 | 0.90 (0.64, 1.25) | 0.05 | 0.08 | 277 | 86 | 18.6 | 1.04 (0.67, 1.60) | 0.04 | 0.06 |
| Mali | PROMIS (46) | | | | | | | | | | | | |
| Mali | PROMIS CS (46) | | | | | | | | | | | | |
| Zimbabwe | SHINE (HIV−) (47) | | | | | | | | | | | | |
| Zimbabwe | SHINE (HIV+) (48) | | | | | | | | | | | | |
| | | **4790** | **6967** | | **I² = 0.38, Tau² = 0.01** | | | **3068** | **5535** | | **I² = 0.26, Tau² = 0.01** | | |
| **Fixed** | | | | | **0.88 (0.81, 0.95)** | | | | | | **0.92 (0.84, 1.00)** | | |
| **Random** | | | | | **0.87 (0.79, 0.97)** | | | | | | **0.91 (0.81, 1.03)** | | |

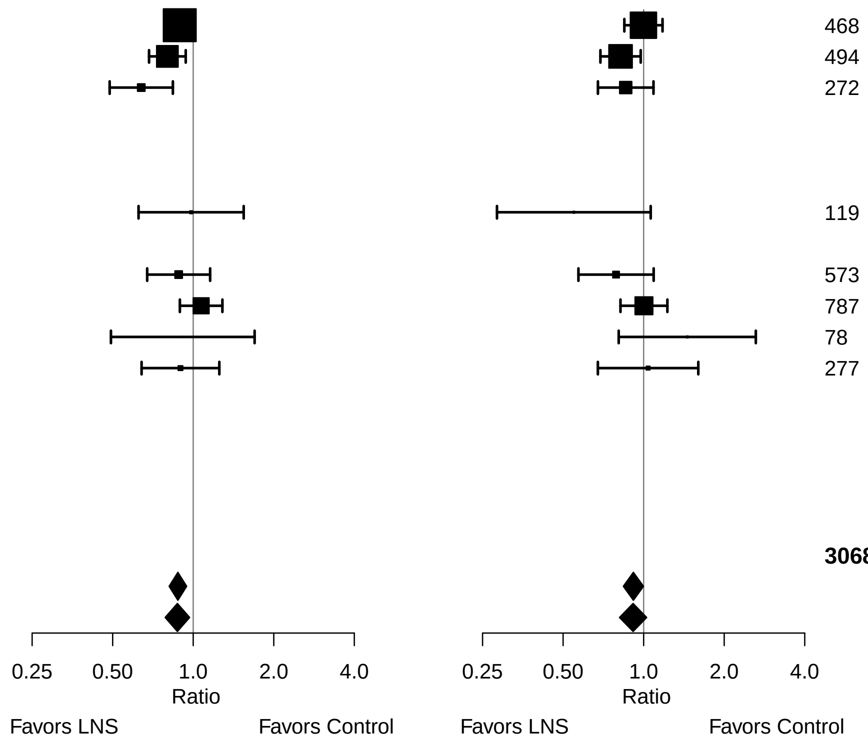

#### Supplemental figure 7M: Underweight prevalence ratio

### 7M6: Stratified by Season at the time of assessment

**P−for−interaction = 0.102**
**Ratio of PRs = 1.08 (0.98, 1.19)**

| | | Dry | | | | | | Rainy | | | | | |
|---|---|---|---|---|---|---|---|---|---|---|---|---|---|
| **Country** | **Trial** | **LNS N** | **Control N** | **Control Prevalence** | **PR (95% CI)** | **Fixed W** | **Random W** | **LNS N** | **Control N** | **Control Prevalence** | **PR (95% CI)** | **Fixed W** | **Random W** |
| Bangladesh | JiVitA−4 (34) | 1533 | 656 | 37.5 | 0.92 (0.80, 1.05) | 0.26 | 0.17 | 1281 | 576 | 41.1 | 0.91 (0.80, 1.04) | 0.24 | 0.21 |
| Bangladesh | RDNS (35) | 739 | 359 | 37.0 | 0.91 (0.75, 1.10) | 0.13 | 0.13 | 924 | 456 | 38.4 | 0.93 (0.81, 1.07) | 0.24 | 0.21 |
| Bangladesh | WASH−B (36) | 645 | 1938 | 28.3 | 0.76 (0.64, 0.91) | 0.14 | 0.13 | 518 | 1523 | 37.7 | 0.81 (0.70, 0.93) | 0.21 | 0.20 |
| Burkina Faso | iLiNS−Zinc (37) | 1400 | 477 | 31.7 | 0.66 (0.55, 0.78) | 0.16 | 0.14 | 560 | 189 | 29.1 | 0.81 (0.65, 1.01) | 0.09 | 0.10 |
| Burkina Faso | PROMIS (38) | 429 | 447 | 18.3 | 0.76 (0.59, 0.98) | 0.07 | 0.09 | 428 | 464 | 20.9 | 0.95 (0.71, 1.27) | 0.05 | 0.06 |
| Burkina Faso | PROMIS CS (38) | 224 | 190 | 23.2 | 0.91 (0.62, 1.32) | 0.03 | 0.05 | 206 | 246 | 25.2 | 1.08 (0.74, 1.58) | 0.03 | 0.04 |
| Ghana | GHANA (39) | | | | | | | | | | | | |
| Ghana | iLiNS−DYADG (40) | 204 | 408 | 11.5 | 1.06 (0.67, 1.68) | 0.02 | 0.04 | 143 | 285 | 15.1 | 0.56 (0.30, 1.02) | 0.01 | 0.02 |
| Haiti | HAITI (41) | | | | | | | | | | | | |
| Kenya | WASH−B (42) | 348 | 1157 | 10.1 | 0.65 (0.40, 1.06) | 0.02 | 0.04 | 1113 | 4008 | 10.0 | 0.79 (0.63, 0.99) | 0.09 | 0.10 |
| Madagascar | MAHAY (43) | | | | | | | | | | | | |
| Malawi | iLiNS−DYADM (44) | 156 | 316 | 11.1 | 0.93 (0.53, 1.62) | 0.02 | 0.03 | 64 | 128 | 13.3 | 1.53 (0.79, 2.95) | 0.01 | 0.01 |
| Malawi | iLiNS−DOSE (45) | 421 | 134 | 20.1 | 1.06 (0.74, 1.51) | 0.04 | 0.06 | 281 | 107 | 19.6 | 0.95 (0.65, 1.39) | 0.03 | 0.04 |
| Mali | PROMIS (46) | | | | | | | | | | | | |
| Mali | PROMIS CS (46) | | | | | | | | | | | | |
| Zimbabwe | SHINE (HIV−) (47) | 1736 | 1655 | 10.8 | 0.67 (0.54, 0.83) | 0.10 | 0.11 | 133 | 131 | 8.4 | 1.34 (0.60, 3.03) | 0.01 | 0.01 |
| Zimbabwe | SHINE (HIV+) (48) | | | | | | | | | | | | |
| | | **7835** | **7737** | | **I² = 0.47, Tau² = 0.01** | | | **5651** | **8113** | | **I² = 0.10, Tau² = 0.00** | | |
| **Fixed** | | | | | **0.81 (0.76, 0.87)** | | | | | | **0.88 (0.83, 0.94)** | | |
| **Random** | | | | | **0.81 (0.73, 0.89)** | | | | | | **0.88 (0.82, 0.95)** | | |

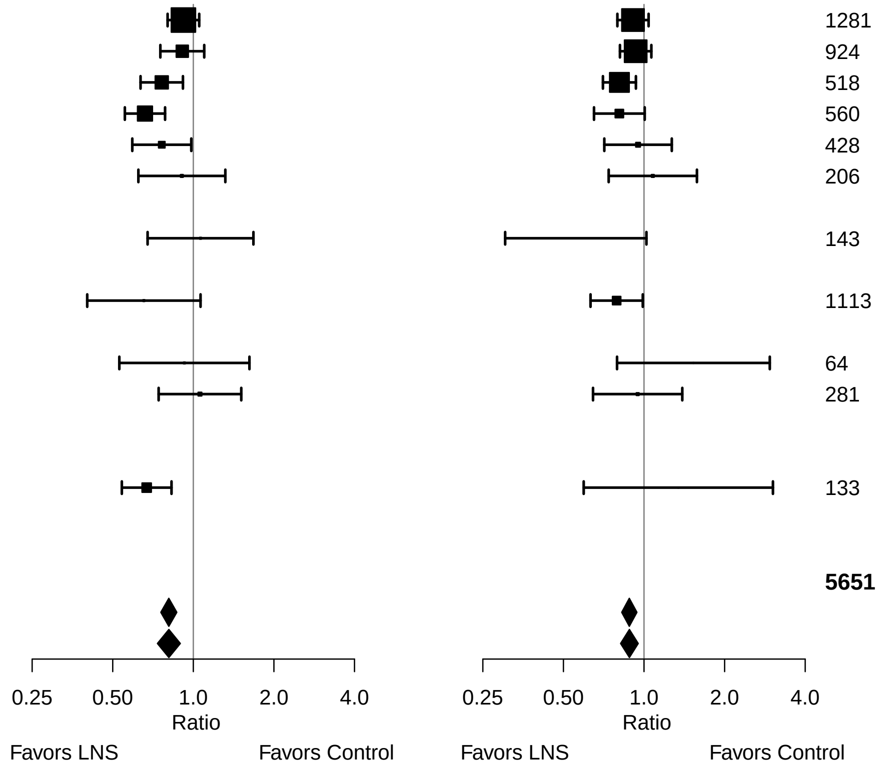

### Supplemental figure 7N: Underweight prevalence difference

#### 7N1: Stratified by Household socio-economic status

**P−for−interaction = 0.123**
**Difference in PDs = −0.01 (−0.03, 0.00)**

| | | At least median | | | | | | Less than median | | | | | |
|---|---|---|---|---|---|---|---|---|---|---|---|---|---|
| **Country** | **Trial** | **LNS N** | **Control N** | **Control Prevalence** | **PD (95% CI)** | **Fixed W** | **Random W** | **LNS N** | **Control N** | **Control Prevalence** | **PD (95% CI)** | **Fixed W** | **Random W** |
| Bangladesh | JiVitA−4 (34) | 1421 | 619 | 35.4 | −0.05 (−0.09, −0.02) | 0.08 | 0.10 | 1392 | 613 | 42.9 | −0.03 (−0.07, 0.01) | 0.12 | 0.10 |
| Bangladesh | RDNS (35) | 834 | 407 | 32.2 | −0.03 (−0.08, 0.02) | 0.04 | 0.06 | 829 | 408 | 43.4 | −0.03 (−0.08, 0.02) | 0.07 | 0.08 |
| Bangladesh | WASH−B (36) | 584 | 1741 | 23.9 | −0.04 (−0.08, 0.01) | 0.06 | 0.08 | 579 | 1720 | 41.0 | −0.10 (−0.15, −0.06) | 0.07 | 0.08 |
| Burkina Faso | iLiNS−Zinc (37) | 1233 | 300 | 28.0 | −0.08 (−0.12, −0.03) | 0.05 | 0.07 | 720 | 365 | 33.2 | −0.12 (−0.15, −0.08) | 0.11 | 0.10 |
| Burkina Faso | PROMIS (38) | 405 | 482 | 19.9 | −0.04 (−0.10, 0.02) | 0.03 | 0.04 | 452 | 429 | 19.3 | −0.02 (−0.07, 0.04) | 0.05 | 0.06 |
| Burkina Faso | PROMIS CS (38) | 220 | 208 | 21.6 | 0.02 (−0.05, 0.08) | 0.02 | 0.03 | 210 | 228 | 26.8 | −0.02 (−0.10, 0.06) | 0.02 | 0.04 |
| Ghana | GHANA (39) | | | | | | | | | | | | |
| Ghana | iLiNS−DYADG (40) | 157 | 356 | 11.0 | −0.02 (−0.08, 0.04) | 0.03 | 0.05 | 190 | 335 | 14.9 | −0.03 (−0.09, 0.03) | 0.04 | 0.06 |
| Haiti | HAITI (41) | | | | | | | | | | | | |
| Kenya | WASH−B (42) | 828 | 2941 | 8.0 | −0.03 (−0.04, −0.01) | 0.35 | 0.18 | 631 | 2220 | 12.7 | −0.02 (−0.05, 0.01) | 0.17 | 0.11 |
| Madagascar | MAHAY (43) | 772 | 884 | 23.2 | 0.02 (−0.03, 0.06) | 0.04 | 0.06 | 877 | 758 | 29.6 | −0.01 (−0.07, 0.05) | 0.04 | 0.06 |
| Malawi | iLiNS−DYADM (44) | 121 | 234 | 9.0 | 0.03 (−0.03, 0.10) | 0.02 | 0.04 | 99 | 209 | 14.4 | 0.00 (−0.09, 0.08) | 0.02 | 0.04 |
| Malawi | iLiNS−DOSE (45) | 307 | 104 | 10.6 | 0.02 (−0.05, 0.09) | 0.02 | 0.03 | 294 | 103 | 30.1 | −0.06 (−0.14, 0.02) | 0.02 | 0.04 |
| Mali | PROMIS (46) | 247 | 250 | 15.5 | −0.05 (−0.10, 0.00) | 0.04 | 0.06 | 245 | 246 | 17.1 | −0.03 (−0.10, 0.03) | 0.04 | 0.05 |
| Mali | PROMIS CS (46) | 501 | 458 | 21.4 | −0.02 (−0.07, 0.03) | 0.04 | 0.05 | 445 | 502 | 23.5 | −0.03 (−0.09, 0.03) | 0.04 | 0.06 |
| Zimbabwe | SHINE (HIV−) (47) | 903 | 797 | 9.0 | −0.02 (−0.05, 0.00) | 0.16 | 0.13 | 826 | 858 | 12.5 | −0.05 (−0.08, −0.02) | 0.17 | 0.11 |
| Zimbabwe | SHINE (HIV+) (48) | 171 | 156 | 14.7 | 0.00 (−0.07, 0.08) | 0.02 | 0.03 | 157 | 166 | 19.9 | −0.01 (−0.10, 0.09) | 0.02 | 0.03 |
| | | **8704** | **9937** | | **I² = 0.24, Tau² = 0.00** | | | **7946** | **9160** | | **I² = 0.49, Tau² = 0.00** | | |
| **Fixed** | | | | | **−0.03 (−0.04, −0.02)** | | | | | | **−0.04 (−0.06, −0.03)** | | |
| **Random** | | | | | **−0.03 (−0.04, −0.01)** | | | | | | **−0.04 (−0.06, −0.02)** | | |

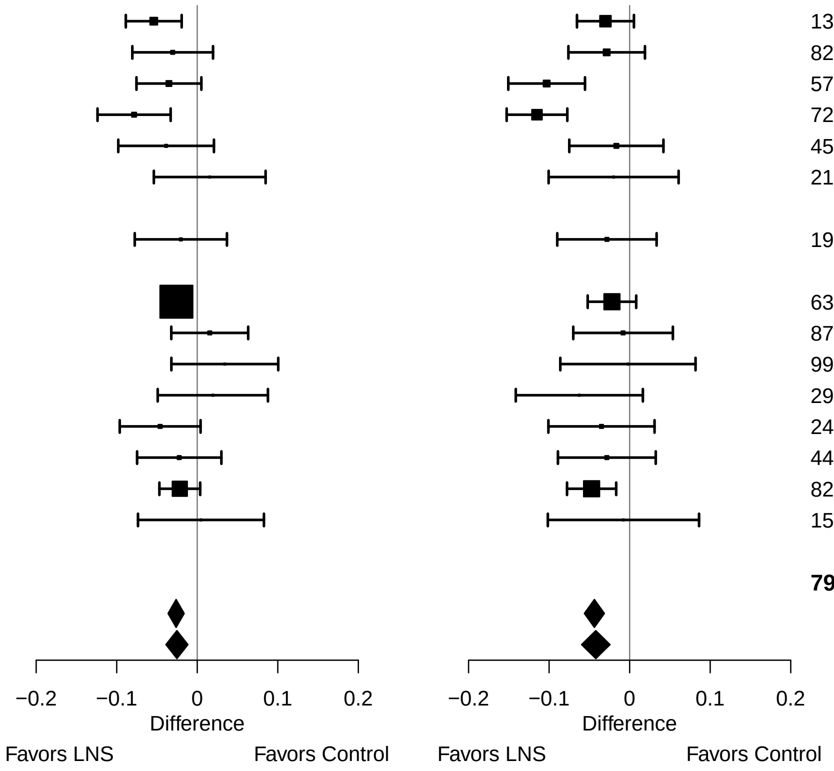

#### Supplemental figure 7N: Underweight prevalence difference

#### 7N2: Stratified by Household food insecurity

**P−for−interaction = 0.068**
**Difference in PDs = −0.02 (−0.04, 0.00)**

| Country | Trial | Mild to secure LNS N | Control N | Control Prevalence | PD (95% CI) | Fixed W | Random W | Moderate to severe LNS N | Control N | Control Prevalence | PD (95% CI) | Fixed W | Random W |
|---|---|---|---|---|---|---|---|---|---|---|---|---|---|
| Bangladesh | JiVitA−4 (34) | 2000 | 851 | 37.0 | −0.05 (−0.08, −0.02) | 0.10 | 0.11 | 813 | 381 | 43.9 | −0.03 (−0.07, 0.02) | 0.13 | 0.11 |
| Bangladesh | RDNS (35) | 1058 | 493 | 32.9 | 0.00 (−0.05, 0.05) | 0.04 | 0.07 | 605 | 322 | 45.3 | −0.07 (−0.13, 0.00) | 0.07 | 0.07 |
| Bangladesh | WASH−B (36) | 923 | 2681 | 30.8 | −0.07 (−0.11, −0.04) | 0.09 | 0.10 | 240 | 780 | 38.1 | −0.04 (−0.12, 0.03) | 0.06 | 0.06 |
| Burkina Faso | iLiNS−Zinc (37) | 1017 | 321 | 27.4 | −0.08 (−0.13, −0.03) | 0.04 | 0.07 | 936 | 344 | 34.0 | −0.12 (−0.16, −0.08) | 0.18 | 0.13 |
| Burkina Faso | PROMIS (38) | 502 | 550 | 19.3 | −0.01 (−0.06, 0.04) | 0.04 | 0.07 | 355 | 361 | 20.2 | −0.05 (−0.12, 0.01) | 0.07 | 0.08 |
| Burkina Faso | PROMIS CS (38) | | | | | | | | | | | | |
| Ghana | GHANA (39) | | | | | | | | | | | | |
| Ghana | iLiNS−DYADG (40) | 244 | 471 | 12.7 | −0.03 (−0.08, 0.02) | 0.04 | 0.07 | 102 | 219 | 13.7 | −0.01 (−0.09, 0.07) | 0.05 | 0.06 |
| Haiti | HAITI (41) | | | | | | | | | | | | |
| Kenya | WASH−B (42) | 1303 | 4621 | 9.8 | −0.02 (−0.04, −0.01) | 0.34 | 0.14 | 156 | 527 | 12.1 | −0.04 (−0.10, 0.02) | 0.08 | 0.09 |
| Madagascar | MAHAY (43) | 989 | 929 | 25.6 | 0.01 (−0.04, 0.06) | 0.04 | 0.07 | 354 | 410 | 27.3 | 0.01 (−0.08, 0.10) | 0.04 | 0.05 |
| Malawi | iLiNS−DYADM (44) | 70 | 122 | 8.2 | 0.05 (−0.04, 0.13) | 0.01 | 0.03 | 148 | 316 | 12.7 | 0.01 (−0.06, 0.07) | 0.07 | 0.07 |
| Malawi | iLiNS−DOSE (45) | 154 | 55 | 14.5 | 0.00 (−0.09, 0.10) | 0.01 | 0.02 | 434 | 146 | 21.9 | −0.03 (−0.09, 0.04) | 0.07 | 0.08 |
| Mali | PROMIS (46) | 327 | 338 | 15.2 | −0.03 (−0.08, 0.01) | 0.04 | 0.07 | 170 | 163 | 19.0 | −0.05 (−0.12, 0.03) | 0.06 | 0.06 |
| Mali | PROMIS CS (46) | | | | | | | | | | | | |
| Zimbabwe | SHINE (HIV−) (47) | 1384 | 1299 | 9.8 | −0.02 (−0.05, 0.00) | 0.19 | 0.13 | 294 | 324 | 15.4 | −0.08 (−0.13, −0.03) | 0.11 | 0.10 |
| Zimbabwe | SHINE (HIV+) (48) | 236 | 238 | 17.6 | 0.01 (−0.06, 0.07) | 0.02 | 0.05 | 85 | 76 | 17.1 | −0.02 (−0.13, 0.09) | 0.02 | 0.03 |
| | | **10207** | **12969** | | **I² = 0.45, Tau² = 0.00** | | | **4692** | **4369** | | **I² = 0.41, Tau² = 0.00** | | |
| **Fixed** | | | | | **−0.03 (−0.04, −0.02)** | | | | | | **−0.05 (−0.07, −0.03)** | | |
| **Random** | | | | | **−0.03 (−0.04, −0.01)** | | | | | | **−0.05 (−0.07, −0.03)** | | |

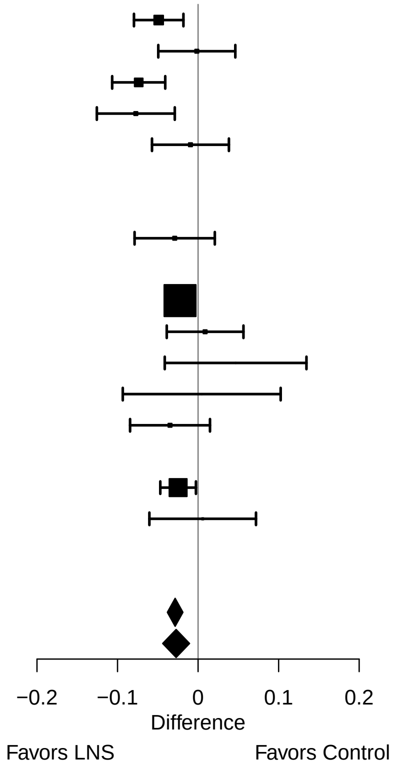

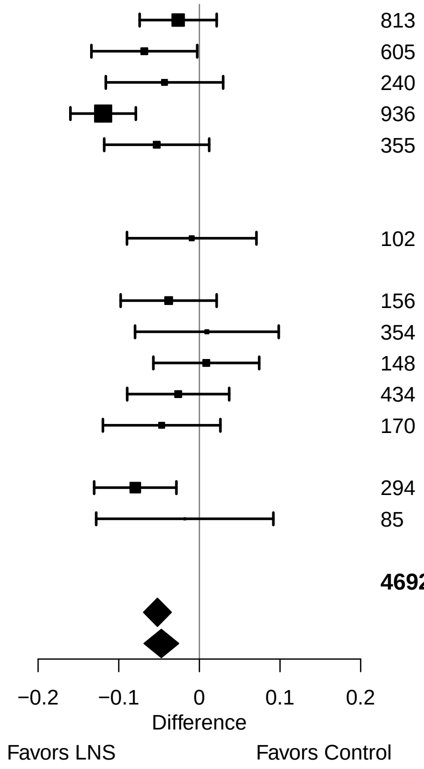

#### Supplemental figure 7N: Underweight prevalence difference

##### 7N3: Stratified by Household source water quality

**P−for−interaction = 0.232**
**Difference in PDs = −0.01 (−0.04, 0.01)**

| Country | Trial | Improved LNS N | Improved Control N | Improved Control Prevalence | Improved PD (95% CI) | Improved Fixed W | Improved Random W | Unimproved LNS N | Unimproved Control N | Unimproved Control Prevalence | Unimproved PD (95% CI) | Unimproved Fixed W | Unimproved Random W |
|---|---|---|---|---|---|---|---|---|---|---|---|---|---|
| Bangladesh | JiVitA−4 (34) | | | | | | | | | | | | |
| Bangladesh | RDNS (35) | | | | | | | | | | | | |
| Bangladesh | WASH−B (36) | 501 | 1002 | 30.7 | −0.06 (−0.10, −0.01) | 0.11 | 0.12 | 70 | 116 | 29.3 | 0.04 (−0.10, 0.17) | 0.02 | 0.04 |
| Burkina Faso | iLiNS−Zinc (37) | 555 | 141 | 27.7 | −0.06 (−0.12, −0.01) | 0.09 | 0.10 | 1397 | 521 | 31.9 | −0.11 (−0.15, −0.08) | 0.25 | 0.14 |
| Burkina Faso | PROMIS (38) | 352 | 379 | 17.9 | 0.00 (−0.05, 0.06) | 0.08 | 0.10 | 358 | 351 | 23.4 | −0.10 (−0.16, −0.04) | 0.08 | 0.11 |
| Burkina Faso | PROMIS CS (38) | 280 | 267 | 27.3 | −0.05 (−0.12, 0.03) | 0.04 | 0.06 | 149 | 169 | 19.5 | 0.07 (−0.01, 0.16) | 0.04 | 0.08 |
| Ghana | GHANA (39) | | | | | | | | | | | | |
| Ghana | iLiNS−DYADG (40) | | | | | | | | | | | | |
| Haiti | HAITI (41) | | | | | | | | | | | | |
| Kenya | WASH−B (42) | 449 | 1555 | 8.8 | −0.01 (−0.04, 0.02) | 0.26 | 0.17 | 248 | 699 | 10.7 | −0.02 (−0.07, 0.03) | 0.14 | 0.13 |
| Madagascar | MAHAY (43) | 418 | 454 | 20.5 | 0.05 (−0.01, 0.12) | 0.06 | 0.08 | 1204 | 1182 | 28.1 | −0.01 (−0.05, 0.03) | 0.16 | 0.13 |
| Malawi | iLiNS−DYADM (44) | | | | | | | | | | | | |
| Malawi | iLiNS−DOSE (45) | | | | | | | | | | | | |
| Mali | PROMIS (46) | 247 | 257 | 14.3 | −0.04 (−0.10, 0.02) | 0.07 | 0.09 | 243 | 240 | 18.5 | −0.04 (−0.10, 0.02) | 0.08 | 0.10 |
| Mali | PROMIS CS (46) | 544 | 563 | 21.5 | −0.03 (−0.08, 0.03) | 0.07 | 0.09 | 363 | 382 | 23.6 | −0.02 (−0.09, 0.05) | 0.06 | 0.10 |
| Zimbabwe | SHINE (HIV−) (47) | 518 | 486 | 9.7 | −0.02 (−0.06, 0.01) | 0.20 | 0.15 | 273 | 301 | 8.6 | 0.00 (−0.04, 0.05) | 0.16 | 0.13 |
| Zimbabwe | SHINE (HIV+) (48) | 85 | 87 | 13.8 | 0.04 (−0.07, 0.15) | 0.02 | 0.04 | 55 | 56 | 21.4 | −0.03 (−0.17, 0.11) | 0.02 | 0.04 |
| | | **3949** | **5191** | | **I² = 0.36, Tau² = 0.00** | | | **4360** | **4017** | | **I² = 0.73, Tau² = 0.00** | | |
| **Fixed** | | | | | **−0.02 (−0.04, 0.00)** | | | | | | **−0.04 (−0.06, −0.02)** | | |
| **Random** | | | | | **−0.02 (−0.04, 0.00)** | | | | | | **−0.03 (−0.06, 0.00)** | | |

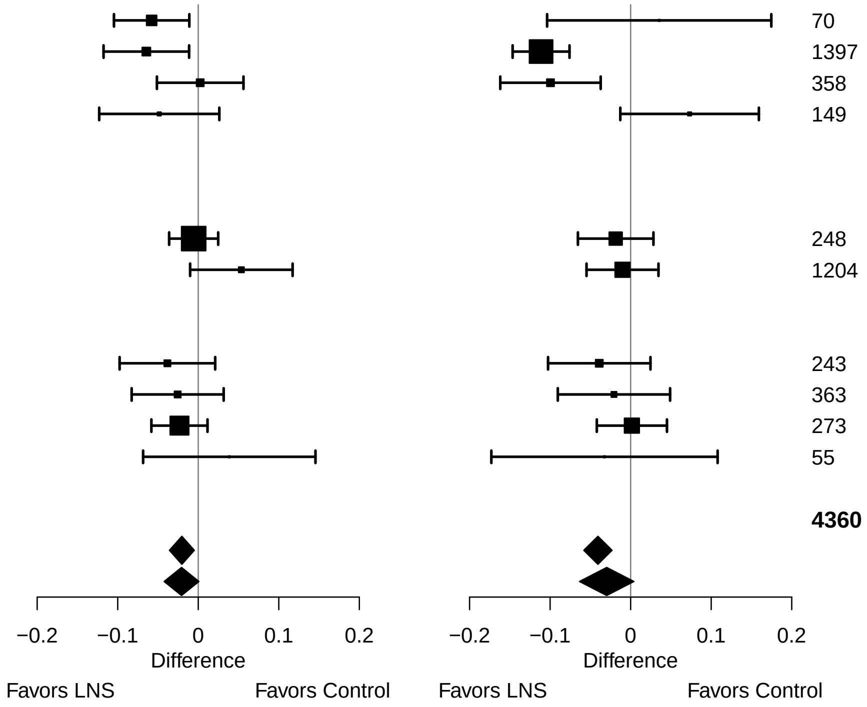

#### Supplemental figure 7N: Underweight prevalence difference

#### 7N4: Stratified by Household sanitation

**P−for−interaction = 0.679**
**Difference in PDs = 0.01 (−0.02, 0.03)**

| | | Improved | | | | | | Unimproved | | | | | |
|---|---|---|---|---|---|---|---|---|---|---|---|---|---|
| **Country** | **Trial** | **LNS N** | **Control N** | **Control Prevalence** | **PD (95% CI)** | **Fixed W** | **Random W** | **LNS N** | **Control N** | **Control Prevalence** | **PD (95% CI)** | **Fixed W** | **Random W** |
| Bangladesh | JiVitA−4 (34) | 2179 | 944 | 37.7 | −0.05 (−0.07, −0.02) | 0.36 | 0.36 | 634 | 288 | 43.9 | −0.03 (−0.08, 0.02) | 0.17 | 0.17 |
| Bangladesh | RDNS (35) | 1179 | 573 | 34.9 | −0.02 (−0.06, 0.01) | 0.19 | 0.19 | 482 | 241 | 44.8 | −0.04 (−0.13, 0.04) | 0.06 | 0.06 |
| Bangladesh | WASH−B (36) | | | | | | | | | | | | |
| Burkina Faso | iLiNS−Zinc (37) | | | | | | | | | | | | |
| Burkina Faso | PROMIS (38) | 392 | 550 | 20.0 | −0.03 (−0.08, 0.02) | 0.10 | 0.10 | 465 | 357 | 19.3 | −0.03 (−0.08, 0.03) | 0.14 | 0.14 |
| Burkina Faso | PROMIS CS (38) | 176 | 165 | 23.0 | 0.01 (−0.09, 0.10) | 0.03 | 0.03 | 252 | 271 | 25.1 | −0.01 (−0.08, 0.06) | 0.09 | 0.09 |
| Ghana | GHANA (39) | | | | | | | | | | | | |
| Ghana | iLiNS−DYADG (40) | | | | | | | | | | | | |
| Haiti | HAITI (41) | | | | | | | | | | | | |
| Kenya | WASH−B (42) | | | | | | | | | | | | |
| Madagascar | MAHAY (43) | | | | | | | | | | | | |
| Malawi | iLiNS−DYADM (44) | | | | | | | | | | | | |
| Malawi | iLiNS−DOSE (45) | | | | | | | | | | | | |
| Mali | PROMIS (46) | 381 | 363 | 16.1 | −0.04 (−0.09, 0.01) | 0.11 | 0.11 | 112 | 134 | 17.0 | −0.03 (−0.10, 0.04) | 0.09 | 0.09 |
| Mali | PROMIS CS (46) | 698 | 692 | 22.5 | −0.03 (−0.09, 0.03) | 0.08 | 0.08 | 217 | 243 | 21.4 | −0.03 (−0.08, 0.03) | 0.13 | 0.13 |
| Zimbabwe | SHINE (HIV−) (47) | 293 | 243 | 7.8 | −0.02 (−0.07, 0.02) | 0.13 | 0.13 | 495 | 543 | 9.9 | −0.01 (−0.04, 0.03) | 0.33 | 0.33 |
| Zimbabwe | SHINE (HIV+) (48) | | | | | | | | | | | | |
| | | **5298** | **3530** | | **I² = 0.00, Tau² = 0.00** | | | **2657** | **2077** | | **I² = 0.00, Tau² = 0.00** | | |
| **Fixed** | | | | | **−0.03 (−0.05, −0.02)** | | | | | | **−0.02 (−0.04, 0.00)** | | |
| **Random** | | | | | **−0.03 (−0.05, −0.02)** | | | | | | **−0.02 (−0.04, 0.00)** | | |

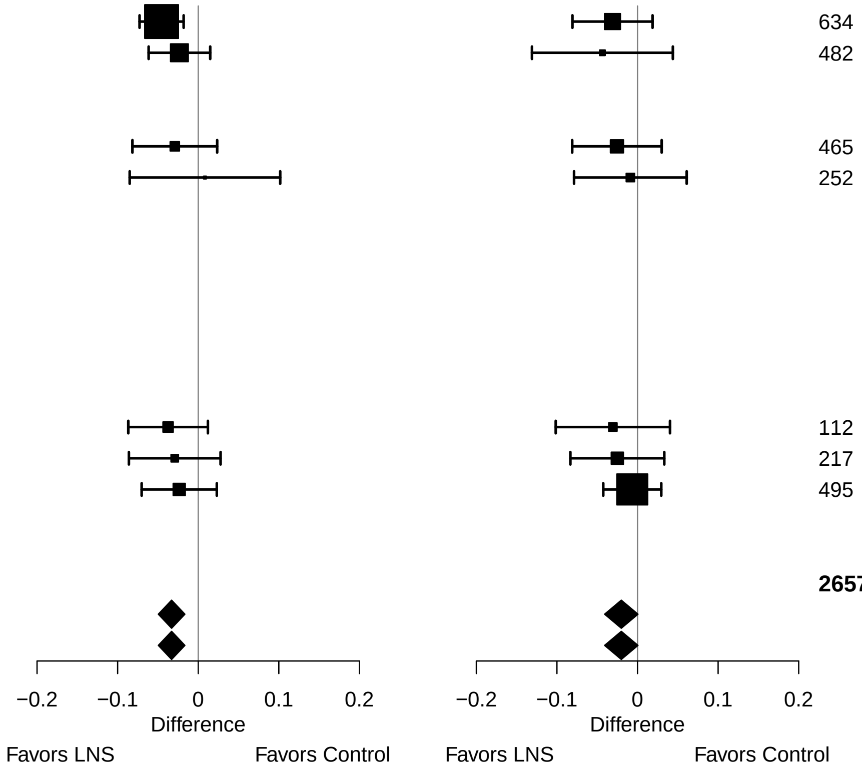

#### Supplemental figure 7N: Underweight prevalence difference

#### 7N5: Stratified by Home environment

**P−for−interaction = 0.750**
**Difference in PDs = 0.00 (−0.03, 0.02)**

| Country | Trial | At least median: LNS N | Control N | Control Prevalence | PD (95% CI) | Fixed W | Random W | Less than median: LNS N | Control N | Control Prevalence | PD (95% CI) | Fixed W | Random W |
|---|---|---|---|---|---|---|---|---|---|---|---|---|---|
| Bangladesh | JiVitA−4 (34) | | | | | | | | | | | | |
| Bangladesh | RDNS (35) | 1192 | 582 | 36.1 | −0.04 (−0.08, 0.01) | 0.09 | 0.13 | 468 | 230 | 41.7 | 0.00 (−0.07, 0.07) | 0.07 | 0.09 |
| Bangladesh | WASH−B (36) | 641 | 1668 | 28.1 | −0.06 (−0.09, −0.02) | 0.13 | 0.14 | 494 | 1702 | 36.5 | −0.07 (−0.12, −0.01) | 0.12 | 0.13 |
| Burkina Faso | iLiNS−Zinc (37) | 473 | 199 | 27.6 | −0.09 (−0.14, −0.05) | 0.08 | 0.12 | 272 | 175 | 31.4 | −0.03 (−0.09, 0.02) | 0.12 | 0.14 |
| Burkina Faso | PROMIS (38) | | | | | | | | | | | | |
| Burkina Faso | PROMIS CS (38) | | | | | | | | | | | | |
| Ghana | GHANA (39) | | | | | | | | | | | | |
| Ghana | iLiNS−DYADG (40) | 213 | 410 | 12.0 | 0.00 (−0.06, 0.05) | 0.06 | 0.11 | 119 | 248 | 15.3 | −0.07 (−0.14, 0.00) | 0.06 | 0.08 |
| Haiti | HAITI (41) | | | | | | | | | | | | |
| Kenya | WASH−B (42) | 840 | 2776 | 8.0 | −0.01 (−0.03, 0.01) | 0.47 | 0.19 | 573 | 2167 | 10.8 | −0.02 (−0.05, 0.01) | 0.42 | 0.30 |
| Madagascar | MAHAY (43) | 915 | 907 | 24.8 | 0.02 (−0.03, 0.06) | 0.08 | 0.12 | 787 | 778 | 27.4 | 0.00 (−0.05, 0.06) | 0.11 | 0.13 |
| Malawi | iLiNS−DYADM (44) | 137 | 289 | 10.4 | −0.01 (−0.07, 0.05) | 0.05 | 0.09 | 78 | 149 | 14.1 | 0.06 (−0.04, 0.17) | 0.03 | 0.05 |
| Malawi | iLiNS−DOSE (45) | 379 | 136 | 22.8 | −0.06 (−0.12, 0.01) | 0.04 | 0.09 | 277 | 86 | 18.6 | 0.00 (−0.08, 0.08) | 0.05 | 0.07 |
| Mali | PROMIS (46) | | | | | | | | | | | | |
| Mali | PROMIS CS (46) | | | | | | | | | | | | |
| Zimbabwe | SHINE (HIV−) (47) | | | | | | | | | | | | |
| Zimbabwe | SHINE (HIV+) (48) | | | | | | | | | | | | |
| | | **4790** | **6967** | | **I² = 0.63, Tau² = 0.00** | | | **3068** | **5535** | | **I² = 0.15, Tau² = 0.00** | | |
| **Fixed** | | | | | **−0.02 (−0.04, −0.01)** | | | | | | **−0.02 (−0.04, −0.01)** | | |
| **Random** | | | | | **−0.03 (−0.06, −0.01)** | | | | | | **−0.02 (−0.05, 0.00)** | | |

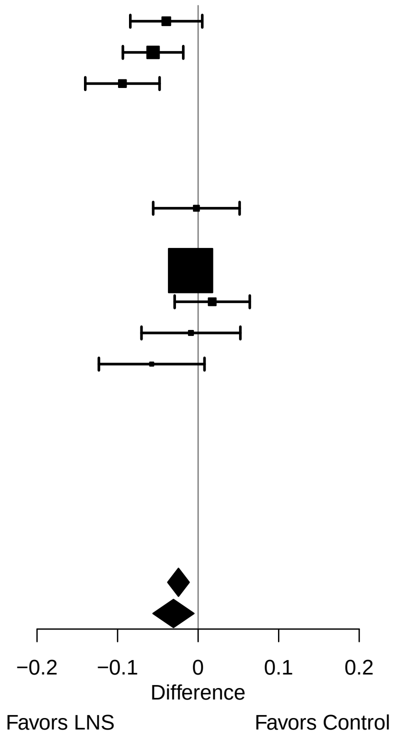

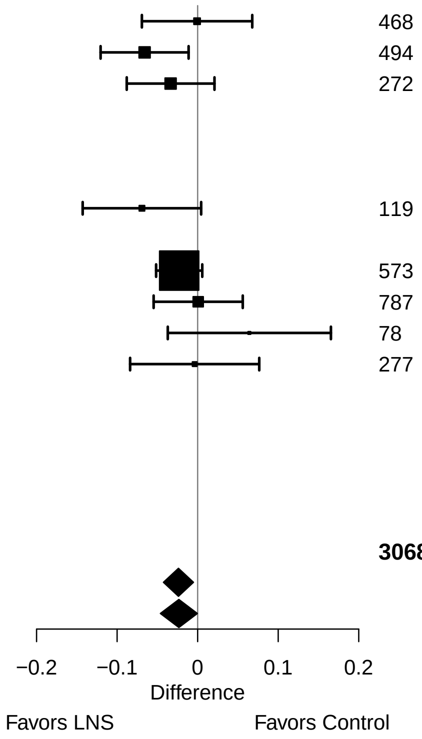

#### Supplemental figure 7N: Underweight prevalence difference

### 7N6: Stratified by Season at the time of assessment

**P−for−interaction = 0.254**
**Difference in PDs = 0.01 (−0.01, 0.03)**

| Country | Trial | Dry LNS N | Dry Control N | Dry Control Prevalence | Dry PD (95% CI) | Dry Fixed W | Dry Random W | Rainy LNS N | Rainy Control N | Rainy Control Prevalence | Rainy PD (95% CI) | Rainy Fixed W | Rainy Random W |
|---|---|---|---|---|---|---|---|---|---|---|---|---|---|
| Bangladesh | JiVitA−4 (34) | 1533 | 656 | 37.5 | −0.04 (−0.08, −0.01) | 0.12 | 0.12 | 1281 | 576 | 41.1 | −0.04 (−0.08, 0.00) | 0.12 | 0.13 |
| Bangladesh | RDNS (35) | 739 | 359 | 37.0 | −0.03 (−0.10, 0.03) | 0.03 | 0.05 | 924 | 456 | 38.4 | −0.03 (−0.08, 0.02) | 0.07 | 0.10 |
| Bangladesh | WASH−B (36) | 645 | 1938 | 28.3 | −0.07 (−0.11, −0.03) | 0.08 | 0.10 | 518 | 1523 | 37.7 | −0.07 (−0.12, −0.03) | 0.08 | 0.11 |
| Burkina Faso | iLiNS−Zinc (37) | 1400 | 477 | 31.7 | −0.10 (−0.14, −0.06) | 0.08 | 0.10 | 560 | 189 | 29.1 | −0.09 (−0.15, −0.03) | 0.05 | 0.09 |
| Burkina Faso | PROMIS (38) | 429 | 447 | 18.3 | −0.04 (−0.08, 0.00) | 0.08 | 0.10 | 428 | 464 | 20.9 | −0.01 (−0.07, 0.05) | 0.05 | 0.09 |
| Burkina Faso | PROMIS CS (38) | 224 | 190 | 23.2 | −0.02 (−0.10, 0.06) | 0.02 | 0.04 | 206 | 246 | 25.2 | 0.02 (−0.08, 0.12) | 0.02 | 0.04 |
| Ghana | GHANA (39) | | | | | | | | | | | | |
| Ghana | iLiNS−DYADG (40) | 204 | 408 | 11.5 | 0.01 (−0.05, 0.06) | 0.05 | 0.07 | 143 | 285 | 15.1 | −0.07 (−0.13, 0.00) | 0.04 | 0.08 |
| Haiti | HAITI (41) | | | | | | | | | | | | |
| Kenya | WASH−B (42) | 348 | 1157 | 10.1 | −0.04 (−0.07, 0.00) | 0.12 | 0.12 | 1113 | 4008 | 10.0 | −0.02 (−0.04, 0.00) | 0.50 | 0.18 |
| Madagascar | MAHAY (43) | | | | | | | | | | | | |
| Malawi | iLiNS−DYADM (44) | 156 | 316 | 11.1 | −0.01 (−0.07, 0.05) | 0.04 | 0.06 | 64 | 128 | 13.3 | 0.07 (−0.04, 0.18) | 0.01 | 0.04 |
| Malawi | iLiNS−DOSE (45) | 421 | 134 | 20.1 | −0.01 (−0.08, 0.05) | 0.03 | 0.06 | 281 | 107 | 19.6 | −0.03 (−0.10, 0.05) | 0.03 | 0.07 |
| Mali | PROMIS (46) | | | | | | | | | | | | |
| Mali | PROMIS CS (46) | | | | | | | | | | | | |
| Zimbabwe | SHINE (HIV−) (47) | 1736 | 1655 | 10.8 | −0.04 (−0.05, −0.02) | 0.36 | 0.17 | 133 | 131 | 8.4 | 0.03 (−0.05, 0.11) | 0.03 | 0.06 |
| Zimbabwe | SHINE (HIV+) (48) | | | | | | | | | | | | |
| | | **7835** | **7737** | | **I² = 0.39, Tau² = 0.00** | | | **5651** | **8113** | | **I² = 0.39, Tau² = 0.00** | | |
| **Fixed** | | | | | **−0.04 (−0.05, −0.03)** | | | | | | **−0.03 (−0.04, −0.02)** | | |
| **Random** | | | | | **−0.04 (−0.06, −0.02)** | | | | | | **−0.03 (−0.05, −0.01)** | | |

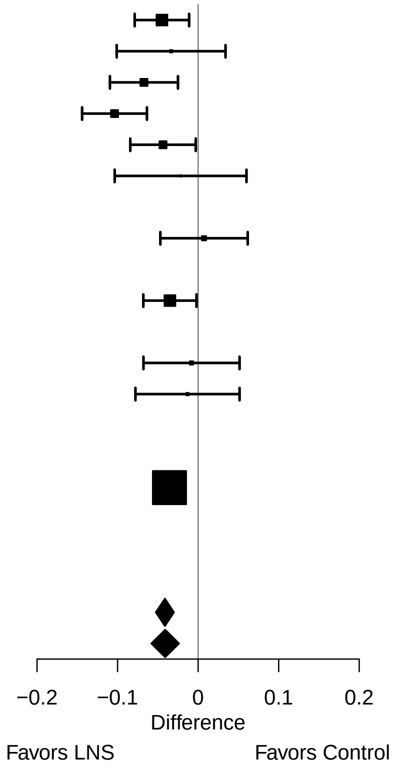

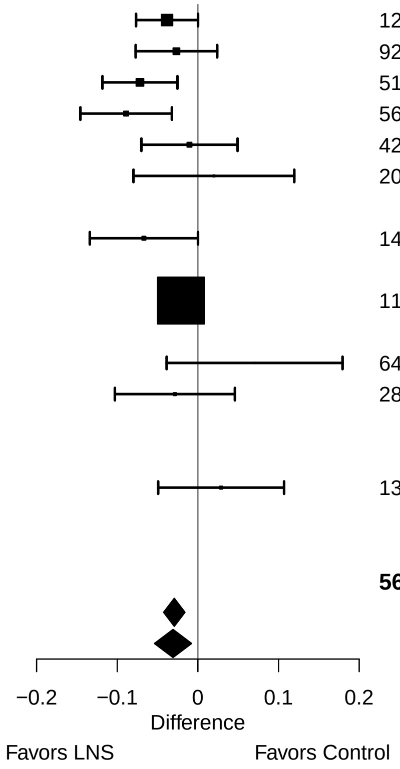

### Supplemental figure 7O: Mean difference in HCZ

#### 7O1: Stratified by Household socio-economic status

**P−for−interaction = 0.842**
**Difference in MDs = 0.00 (−0.04, 0.04)**

| | | At least median | | | | | | Less than median | | | | | |
|---|---|---|---|---|---|---|---|---|---|---|---|---|---|
| Country | Trial | LNS N | Control N | Control Mean | MD (95% CI) | Fixed W | Random W | LNS N | Control N | Control Mean | MD (95% CI) | Fixed W | Random W |
| Bangladesh | JiVitA−4 (34) | 1456 | 640 | −1.36 | 0.07 (0.00, 0.13) | 0.22 | 0.17 | 1428 | 627 | −1.54 | 0.05 (0.00, 0.10) | 0.32 | 0.26 |
| Bangladesh | RDNS (35) | 834 | 407 | −1.78 | 0.06 (−0.04, 0.17) | 0.09 | 0.11 | 829 | 408 | −1.95 | 0.14 (0.05, 0.22) | 0.12 | 0.13 |
| Bangladesh | WASH−B (36) | 583 | 1740 | −1.50 | 0.15 (0.05, 0.24) | 0.11 | 0.12 | 580 | 1723 | −1.73 | 0.10 (0.02, 0.18) | 0.13 | 0.14 |
| Burkina Faso | iLiNS−Zinc (37) | 1226 | 299 | −1.81 | 0.15 (0.09, 0.22) | 0.23 | 0.17 | 713 | 364 | −1.86 | 0.18 (0.09, 0.27) | 0.12 | 0.13 |
| Burkina Faso | PROMIS (38) | | | | | | | | | | | | |
| Burkina Faso | PROMIS CS (38) | | | | | | | | | | | | |
| Ghana | GHANA (39) | 57 | 44 | −0.35 | 0.22 (−0.18, 0.61) | 0.01 | 0.01 | 41 | 46 | −0.79 | 0.00 (−0.30, 0.31) | 0.01 | 0.01 |
| Ghana | iLiNS−DYADG (40) | 157 | 355 | −1.05 | 0.03 (−0.13, 0.20) | 0.04 | 0.06 | 190 | 335 | −1.23 | 0.11 (−0.05, 0.27) | 0.04 | 0.04 |
| Haiti | HAITI (41) | | | | | | | | | | | | |
| Kenya | WASH−B (42) | 827 | 2946 | −0.20 | 0.04 (−0.04, 0.12) | 0.16 | 0.15 | 632 | 2226 | −0.39 | 0.05 (−0.04, 0.14) | 0.12 | 0.13 |
| Madagascar | MAHAY (43) | | | | | | | | | | | | |
| Malawi | iLiNS−DYADM (44) | 121 | 234 | −0.70 | −0.16 (−0.37, 0.04) | 0.02 | 0.04 | 99 | 209 | −0.85 | −0.10 (−0.35, 0.14) | 0.02 | 0.02 |
| Malawi | iLiNS−DOSE (45) | 302 | 102 | −1.09 | 0.11 (−0.08, 0.30) | 0.03 | 0.04 | 290 | 102 | −1.19 | 0.07 (−0.11, 0.26) | 0.03 | 0.03 |
| Mali | PROMIS (46) | | | | | | | | | | | | |
| Mali | PROMIS CS (46) | | | | | | | | | | | | |
| Zimbabwe | SHINE (HIV−) (47) | 905 | 799 | −0.22 | 0.15 (0.05, 0.25) | 0.09 | 0.11 | 826 | 853 | −0.35 | 0.04 (−0.06, 0.14) | 0.09 | 0.10 |
| Zimbabwe | SHINE (HIV+) (48) | 171 | 156 | −0.57 | 0.12 (−0.14, 0.38) | 0.01 | 0.03 | 158 | 167 | −0.53 | 0.08 (−0.16, 0.33) | 0.02 | 0.02 |
| | | **6639** | **7722** | | **I² = 0.36, Tau² = 0.00** | | | **5786** | **7060** | | **I² = 0.15, Tau² = 0.00** | | |
| **Fixed** | | | | | **0.10 (0.06, 0.13)** | | | | | | **0.08 (0.05, 0.11)** | | |
| **Random** | | | | | **0.09 (0.05, 0.13)** | | | | | | **0.08 (0.05, 0.12)** | | |

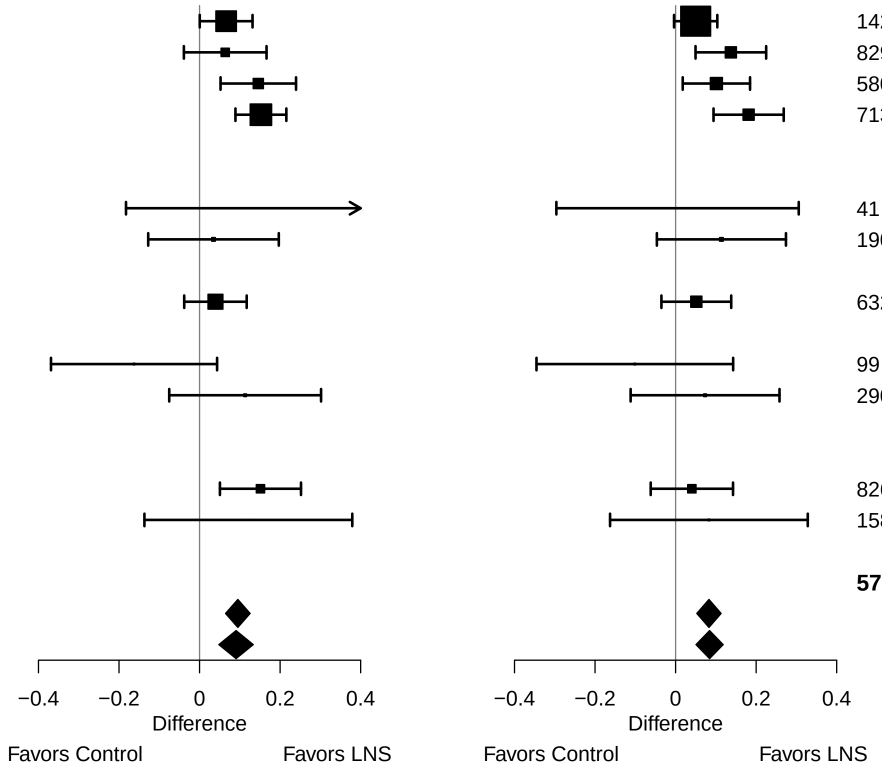

#### Supplemental figure 7O: Mean difference in HCZ

##### 7O2: Stratified by Household food insecurity

**P−for−interaction = 0.603**
**Difference in MDs = −0.01 (−0.06, 0.03)**

| Country | Trial | Mild to secure: LNS N | Control N | Control Mean | MD (95% CI) | Fixed W | Random W | Moderate to severe: LNS N | Control N | Control Mean | MD (95% CI) | Fixed W | Random W |
|---|---|---|---|---|---|---|---|---|---|---|---|---|---|
| Bangladesh | JiVitA−4 (34) | 2045 | 882 | −1.41 | 0.07 (0.02, 0.13) | 0.23 | 0.22 | 839 | 385 | −1.53 | 0.02 (−0.04, 0.09) | 0.34 | 0.18 |
| Bangladesh | RDNS (35) | 1058 | 493 | −1.80 | 0.07 (−0.02, 0.16) | 0.09 | 0.09 | 605 | 322 | −1.96 | 0.14 (0.03, 0.24) | 0.14 | 0.14 |
| Bangladesh | WASH−B (36) | 922 | 2682 | −1.58 | 0.12 (0.04, 0.19) | 0.14 | 0.14 | 241 | 781 | −1.74 | 0.13 (0.00, 0.27) | 0.09 | 0.11 |
| Burkina Faso | iLiNS−Zinc (37) | 1008 | 320 | −1.78 | 0.15 (0.09, 0.22) | 0.19 | 0.19 | 931 | 343 | −1.89 | 0.17 (0.08, 0.27) | 0.19 | 0.16 |
| Burkina Faso | PROMIS (38) | | | | | | | | | | | | |
| Burkina Faso | PROMIS CS (38) | | | | | | | | | | | | |
| Ghana | GHANA (39) | | | | | | | | | | | | |
| Ghana | iLiNS−DYADG (40) | 244 | 470 | −1.12 | 0.08 (−0.06, 0.21) | 0.04 | 0.04 | 102 | 219 | −1.19 | 0.04 (−0.17, 0.25) | 0.03 | 0.06 |
| Haiti | HAITI (41) | | | | | | | | | | | | |
| Kenya | WASH−B (42) | 1303 | 4632 | −0.27 | 0.04 (−0.02, 0.11) | 0.16 | 0.16 | 156 | 527 | −0.33 | 0.04 (−0.15, 0.23) | 0.04 | 0.07 |
| Madagascar | MAHAY (43) | | | | | | | | | | | | |
| Malawi | iLiNS−DYADM (44) | 70 | 122 | −0.77 | −0.06 (−0.34, 0.21) | 0.01 | 0.01 | 148 | 316 | −0.77 | −0.16 (−0.36, 0.03) | 0.04 | 0.07 |
| Malawi | iLiNS−DOSE (45) | 149 | 55 | −1.06 | 0.04 (−0.21, 0.29) | 0.01 | 0.01 | 431 | 143 | −1.18 | 0.12 (−0.04, 0.28) | 0.06 | 0.09 |
| Mali | PROMIS (46) | | | | | | | | | | | | |
| Mali | PROMIS CS (46) | | | | | | | | | | | | |
| Zimbabwe | SHINE (HIV−) (47) | 1385 | 1297 | −0.28 | 0.13 (0.05, 0.22) | 0.11 | 0.11 | 295 | 323 | −0.30 | −0.05 (−0.22, 0.13) | 0.05 | 0.08 |
| Zimbabwe | SHINE (HIV+) (48) | 236 | 239 | −0.52 | 0.05 (−0.17, 0.28) | 0.01 | 0.02 | 86 | 76 | −0.60 | 0.20 (−0.13, 0.53) | 0.01 | 0.03 |
| | | **8420** | **11192** | | **I² = 0.05, Tau² = 0.00** | | | **3834** | **3435** | | **I² = 0.48, Tau² = 0.00** | | |
| **Fixed** | | | | | **0.09 (0.07, 0.12)** | | | | | | **0.08 (0.04, 0.12)** | | |
| **Random** | | | | | **0.09 (0.07, 0.12)** | | | | | | **0.07 (0.01, 0.13)** | | |

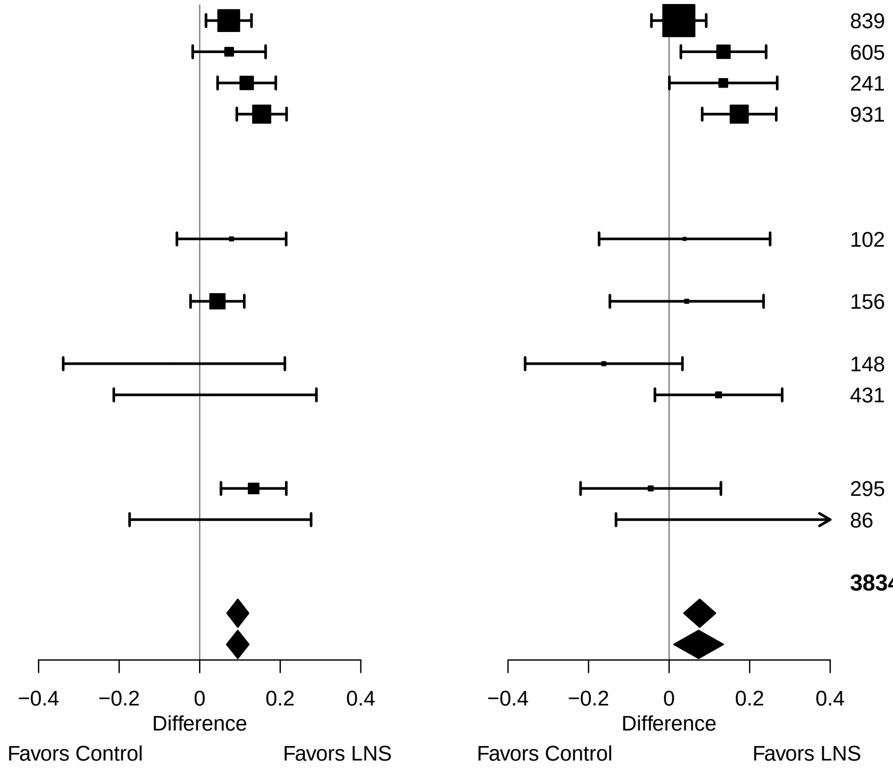

#### Supplemental figure 7O: Mean difference in HCZ

##### 7O3: Stratified by Household source water quality

**P−for−interaction = 0.341**
**Difference in MDs = 0.04 (−0.04, 0.12)**

| Country | Trial | Improved LNS N | Improved Control N | Improved Control Mean | Improved MD (95% CI) | Improved Fixed W | Improved Random W | Unimproved LNS N | Unimproved Control N | Unimproved Control Mean | Unimproved MD (95% CI) | Unimproved Fixed W | Unimproved Random W |
|---|---|---|---|---|---|---|---|---|---|---|---|---|---|
| Bangladesh | JiVitA−4 (34) | | | | | | | | | | | | |
| Bangladesh | RDNS (35) | | | | | | | | | | | | |
| Bangladesh | WASH−B (36) | 500 | 1004 | −1.62 | 0.17 (0.05, 0.28) | 0.18 | 0.17 | 70 | 116 | −1.58 | 0.10 (−0.19, 0.38) | 0.04 | 0.06 |
| Burkina Faso | iLiNS−Zinc (37) | 552 | 141 | −1.77 | 0.11 (0.01, 0.20) | 0.28 | 0.20 | 1386 | 519 | −1.85 | 0.18 (0.11, 0.25) | 0.69 | 0.54 |
| Burkina Faso | PROMIS (38) | | | | | | | | | | | | |
| Burkina Faso | PROMIS CS (38) | | | | | | | | | | | | |
| Ghana | GHANA (39) | | | | | | | | | | | | |
| Ghana | iLiNS−DYADG (40) | | | | | | | | | | | | |
| Haiti | HAITI (41) | | | | | | | | | | | | |
| Kenya | WASH−B (42) | 448 | 1560 | −0.26 | 0.02 (−0.09, 0.14) | 0.18 | 0.17 | 248 | 699 | −0.31 | 0.07 (−0.10, 0.23) | 0.12 | 0.17 |
| Madagascar | MAHAY (43) | | | | | | | | | | | | |
| Malawi | iLiNS−DYADM (44) | 200 | 406 | −0.75 | −0.15 (−0.32, 0.01) | 0.09 | 0.12 | 20 | 36 | −1.01 | 0.11 (−0.48, 0.70) | 0.01 | 0.02 |
| Malawi | iLiNS−DOSE (45) | 553 | 192 | −1.14 | 0.08 (−0.06, 0.22) | 0.12 | 0.14 | 51 | 15 | −1.26 | 0.39 (−0.04, 0.81) | 0.02 | 0.03 |
| Mali | PROMIS (46) | | | | | | | | | | | | |
| Mali | PROMIS CS (46) | | | | | | | | | | | | |
| Zimbabwe | SHINE (HIV−) (47) | 518 | 485 | −0.28 | 0.09 (−0.04, 0.23) | 0.13 | 0.15 | 272 | 299 | −0.22 | −0.05 (−0.24, 0.14) | 0.10 | 0.14 |
| Zimbabwe | SHINE (HIV+) (48) | 85 | 87 | −0.55 | 0.00 (−0.37, 0.36) | 0.02 | 0.04 | 55 | 56 | −0.58 | 0.13 (−0.23, 0.48) | 0.03 | 0.04 |
| | | **2856** | **3875** | | **I² = 0.47, Tau² = 0.00** | | | **2102** | **1740** | | **I² = 0.20, Tau² = 0.00** | | |
| **Fixed** | | | | | **0.07 (0.02, 0.12)** | | | | | | **0.14 (0.09, 0.20)** | | |
| **Random** | | | | | **0.06 (−0.01, 0.13)** | | | | | | **0.13 (0.05, 0.20)** | | |

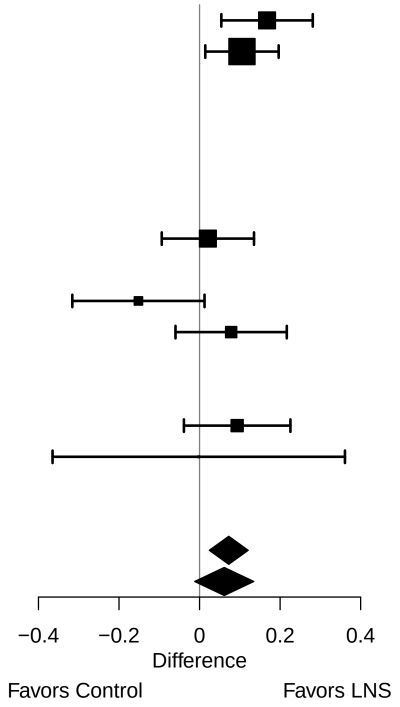

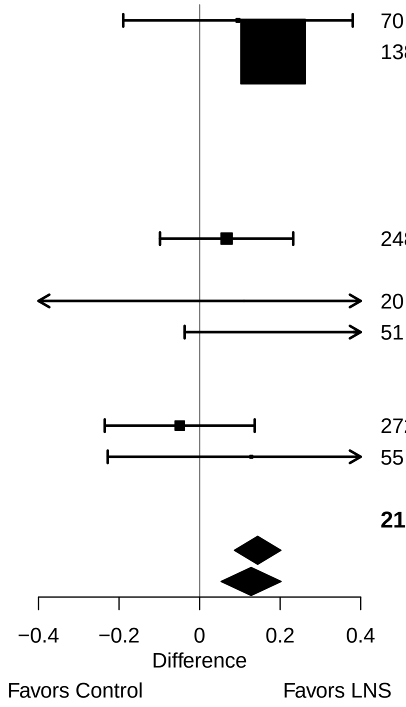

#### Supplemental figure 7O: Mean difference in HCZ

##### 7O4: Stratified by Household sanitation

**P−for−interaction = 0.339**
**Difference in MDs = −0.03 (−0.09, 0.03)**

| | | Improved | | | | | | Unimproved | | | | | |
|---|---|---|---|---|---|---|---|---|---|---|---|---|---|
| Country | Trial | LNS N | Control N | Control Mean | MD (95% CI) | Fixed W | Random W | LNS N | Control N | Control Mean | MD (95% CI) | Fixed W | Random W |
| Bangladesh | JiVitA−4 (34) | 2231 | 972 | −1.41 | 0.07 (0.01, 0.12) | 0.48 | 0.48 | 653 | 295 | −1.57 | 0.02 (−0.05, 0.10) | 0.22 | 0.19 |
| Bangladesh | RDNS (35) | 1179 | 573 | −1.86 | 0.12 (0.04, 0.21) | 0.20 | 0.20 | 482 | 241 | −1.88 | 0.05 (−0.05, 0.15) | 0.12 | 0.14 |
| Bangladesh | WASH−B (36) | 510 | 1017 | −1.61 | 0.16 (0.04, 0.27) | 0.11 | 0.11 | 31 | 55 | −1.56 | 0.04 (−0.32, 0.40) | 0.01 | 0.02 |
| Burkina Faso | iLiNS−Zinc (37) | 43 | 16 | −1.95 | 0.18 (−0.11, 0.47) | 0.02 | 0.02 | 1895 | 644 | −1.83 | 0.16 (0.10, 0.22) | 0.33 | 0.22 |
| Burkina Faso | PROMIS (38) | | | | | | | | | | | | |
| Burkina Faso | PROMIS CS (38) | | | | | | | | | | | | |
| Ghana | GHANA (39) | | | | | | | | | | | | |
| Ghana | iLiNS−DYADG (40) | 336 | 671 | −1.14 | 0.07 (−0.05, 0.19) | 0.11 | 0.11 | 10 | 19 | −1.29 | 0.00 (−0.66, 0.66) | 0.00 | 0.01 |
| Haiti | HAITI (41) | | | | | | | | | | | | |
| Kenya | WASH−B (42) | 100 | 366 | −0.11 | −0.09 (−0.29, 0.11) | 0.04 | 0.04 | 596 | 1894 | −0.31 | 0.06 (−0.04, 0.15) | 0.13 | 0.15 |
| Madagascar | MAHAY (43) | | | | | | | | | | | | |
| Malawi | iLiNS−DYADM (44) | 20 | 42 | −0.61 | −0.16 (−0.58, 0.26) | 0.01 | 0.01 | 200 | 400 | −0.79 | −0.13 (−0.30, 0.04) | 0.04 | 0.06 |
| Malawi | iLiNS−DOSE (45) | 15 | 9 | −1.02 | 0.23 (−0.62, 1.08) | 0.00 | 0.00 | 588 | 198 | −1.15 | 0.11 (−0.02, 0.24) | 0.07 | 0.09 |
| Mali | PROMIS (46) | | | | | | | | | | | | |
| Mali | PROMIS CS (46) | | | | | | | | | | | | |
| Zimbabwe | SHINE (HIV−) (47) | 293 | 243 | −0.21 | 0.05 (−0.15, 0.24) | 0.04 | 0.04 | 494 | 540 | −0.28 | 0.04 (−0.09, 0.17) | 0.07 | 0.10 |
| Zimbabwe | SHINE (HIV+) (48) | 50 | 35 | −0.45 | −0.15 (−0.63, 0.34) | 0.01 | 0.01 | 90 | 107 | −0.59 | 0.12 (−0.17, 0.42) | 0.01 | 0.02 |
| | | **4777** | **3944** | | **I² = 0.00, Tau² = 0.00** | | | **5039** | **4393** | | **I² = 0.48, Tau² = 0.00** | | |
| **Fixed** | | | | | **0.08 (0.04, 0.12)** | | | | | | **0.08 (0.04, 0.11)** | | |
| **Random** | | | | | **0.08 (0.04, 0.12)** | | | | | | **0.07 (0.02, 0.11)** | | |

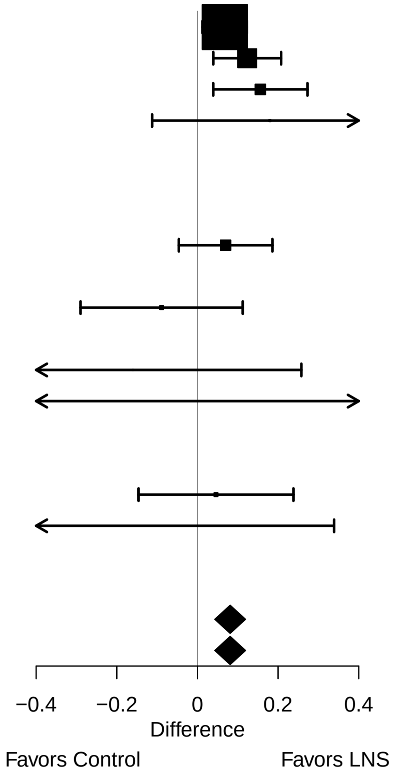

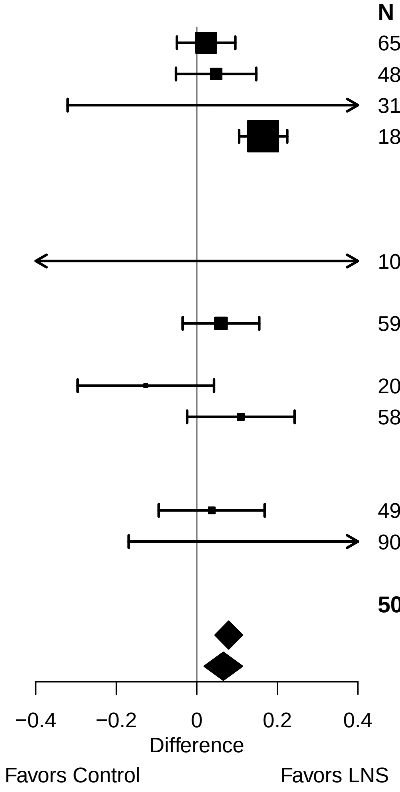

#### Supplemental figure 7O: Mean difference in HCZ

##### 7O5: Stratified by Home environment

**P−for−interaction = 0.416**
**Difference in MDs = 0.03 (−0.04, 0.09)**

| | | At least median | | | | | | Less than median | | | | | |
|---|---|---|---|---|---|---|---|---|---|---|---|---|---|
| **Country** | **Trial** | **LNS N** | **Control N** | **Control Mean** | **MD (95% CI)** | **Fixed W** | **Random W** | **LNS N** | **Control N** | **Control Mean** | **MD (95% CI)** | **Fixed W** | **Random W** |
| Bangladesh | JiVitA−4 (34) | | | | | | | | | | | | |
| Bangladesh | RDNS (35) | 1192 | 582 | −1.85 | 0.10 (0.01, 0.20) | 0.18 | 0.17 | 468 | 230 | −1.90 | 0.09 (0.00, 0.19) | 0.23 | 0.23 |
| Bangladesh | WASH−B (36) | 641 | 1665 | −1.55 | 0.13 (0.04, 0.22) | 0.21 | 0.17 | 494 | 1707 | −1.68 | 0.07 (−0.02, 0.17) | 0.22 | 0.22 |
| Burkina Faso | iLiNS−Zinc (37) | 471 | 199 | −1.77 | 0.14 (0.04, 0.23) | 0.17 | 0.16 | 270 | 174 | −1.95 | 0.16 (0.06, 0.27) | 0.19 | 0.19 |
| Burkina Faso | PROMIS (38) | | | | | | | | | | | | |
| Burkina Faso | PROMIS CS (38) | | | | | | | | | | | | |
| Ghana | GHANA (39) | | | | | | | | | | | | |
| Ghana | iLiNS−DYADG (40) | 213 | 410 | −1.10 | 0.03 (−0.12, 0.17) | 0.08 | 0.12 | 119 | 247 | −1.18 | 0.08 (−0.12, 0.28) | 0.05 | 0.05 |
| Haiti | HAITI (41) | | | | | | | | | | | | |
| Kenya | WASH−B (42) | 839 | 2778 | −0.20 | −0.02 (−0.10, 0.06) | 0.27 | 0.18 | 574 | 2174 | −0.35 | 0.09 (0.00, 0.19) | 0.23 | 0.23 |
| Madagascar | MAHAY (43) | | | | | | | | | | | | |
| Malawi | iLiNS−DYADM (44) | 137 | 289 | −0.73 | −0.16 (−0.35, 0.03) | 0.04 | 0.09 | 78 | 149 | −0.88 | −0.07 (−0.36, 0.22) | 0.02 | 0.02 |
| Malawi | iLiNS−DOSE (45) | 377 | 135 | −1.11 | 0.12 (−0.04, 0.28) | 0.06 | 0.11 | 272 | 85 | −1.10 | 0.07 (−0.14, 0.28) | 0.05 | 0.05 |
| Mali | PROMIS (46) | | | | | | | | | | | | |
| Mali | PROMIS CS (46) | | | | | | | | | | | | |
| Zimbabwe | SHINE (HIV−) (47) | | | | | | | | | | | | |
| Zimbabwe | SHINE (HIV+) (48) | | | | | | | | | | | | |
| | | **3870** | **6058** | | **I² = 0.62, Tau² = 0.01** | | | **2275** | **4766** | | **I² = 0.00, Tau² = 0.00** | | |
| **Fixed** | | | | | **0.06 (0.03, 0.10)** | | | | | | **0.10 (0.05, 0.14)** | | |
| **Random** | | | | | **0.06 (−0.01, 0.13)** | | | | | | **0.10 (0.05, 0.14)** | | |

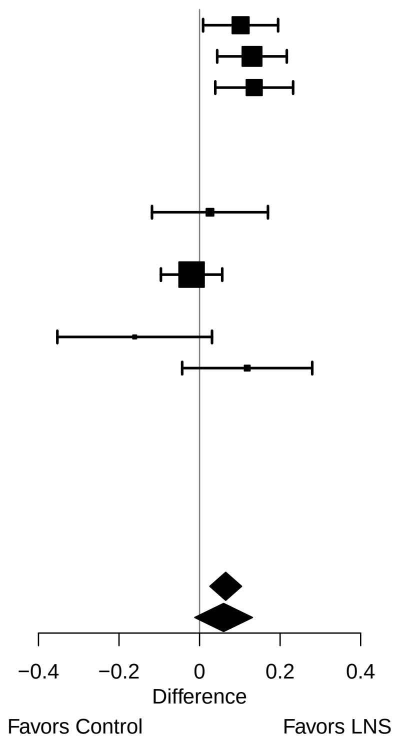

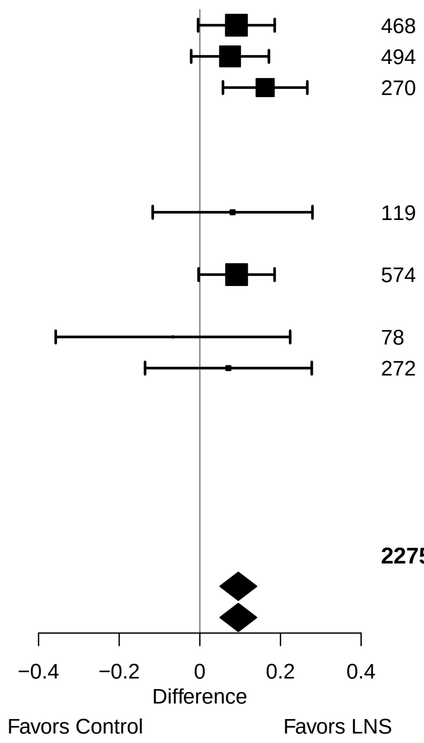

#### Supplemental figure 7O: Mean difference in HCZ

#### 7O6: Stratified by Season at the time of assessment

**P−for−interaction = 0.477**
**Difference in MDs = −0.02 (−0.06, 0.03)**

| Country | Trial | Dry LNS N | Dry Control N | Dry Control Mean | Dry MD (95% CI) | Dry Fixed W | Dry Random W | Rainy LNS N | Rainy Control N | Rainy Control Mean | Rainy MD (95% CI) | Rainy Fixed W | Rainy Random W |
|---|---|---|---|---|---|---|---|---|---|---|---|---|---|
| Bangladesh | JiVitA−4 (34) | 1566 | 674 | −1.44 | 0.06 (0.00, 0.12) | 0.26 | 0.20 | 1319 | 593 | −1.45 | 0.06 (−0.01, 0.12) | 0.27 | 0.17 |
| Bangladesh | RDNS (35) | 739 | 359 | −1.81 | 0.09 (−0.01, 0.19) | 0.08 | 0.10 | 924 | 456 | −1.91 | 0.11 (0.02, 0.20) | 0.13 | 0.14 |
| Bangladesh | WASH−B (36) | 644 | 1936 | −1.56 | 0.08 (−0.01, 0.17) | 0.10 | 0.11 | 519 | 1527 | −1.68 | 0.18 (0.09, 0.26) | 0.17 | 0.15 |
| Burkina Faso | iLiNS−Zinc (37) | 1387 | 477 | −1.86 | 0.17 (0.11, 0.24) | 0.21 | 0.18 | 559 | 187 | −1.80 | 0.14 (0.02, 0.25) | 0.09 | 0.12 |
| Burkina Faso | PROMIS (38) | | | | | | | | | | | | |
| Burkina Faso | PROMIS CS (38) | | | | | | | | | | | | |
| Ghana | GHANA (39) | 57 | 55 | −0.72 | 0.43 (0.06, 0.79) | 0.01 | 0.01 | 41 | 41 | −0.44 | −0.11 (−0.47, 0.24) | 0.01 | 0.02 |
| Ghana | iLiNS−DYADG (40) | 204 | 408 | −1.20 | 0.12 (−0.03, 0.27) | 0.04 | 0.05 | 143 | 284 | −1.05 | −0.01 (−0.18, 0.17) | 0.04 | 0.07 |
| Haiti | HAITI (41) | | | | | | | | | | | | |
| Kenya | WASH−B (42) | 349 | 1161 | −0.34 | 0.10 (−0.02, 0.21) | 0.06 | 0.08 | 1112 | 4015 | −0.26 | 0.03 (−0.04, 0.10) | 0.23 | 0.17 |
| Madagascar | MAHAY (43) | | | | | | | | | | | | |
| Malawi | iLiNS−DYADM (44) | 156 | 316 | −0.75 | −0.05 (−0.24, 0.13) | 0.02 | 0.03 | 64 | 128 | −0.84 | −0.30 (−0.59, −0.01) | 0.01 | 0.03 |
| Malawi | iLiNS−DOSE (45) | 418 | 133 | −1.18 | 0.16 (−0.01, 0.32) | 0.03 | 0.04 | 274 | 105 | −1.06 | 0.04 (−0.15, 0.23) | 0.03 | 0.07 |
| Mali | PROMIS (46) | | | | | | | | | | | | |
| Mali | PROMIS CS (46) | | | | | | | | | | | | |
| Zimbabwe | SHINE (HIV−) (47) | 1738 | 1652 | −0.30 | 0.09 (0.02, 0.16) | 0.16 | 0.15 | 133 | 131 | −0.03 | 0.05 (−0.19, 0.29) | 0.02 | 0.05 |
| Zimbabwe | SHINE (HIV+) (48) | 319 | 305 | −0.58 | 0.12 (−0.06, 0.30) | 0.03 | 0.04 | 18 | 24 | −0.28 | −0.11 (−0.77, 0.56) | 0.00 | 0.01 |
| | | **7577** | **7476** | | **I² = 0.27, Tau² = 0.00** | | | **5106** | **7491** | | **I² = 0.44, Tau² = 0.00** | | |
| **Fixed** | | | | | **0.10 (0.07, 0.13)** | | | | | | **0.08 (0.04, 0.11)** | | |
| **Random** | | | | | **0.10 (0.07, 0.14)** | | | | | | **0.06 (0.01, 0.12)** | | |

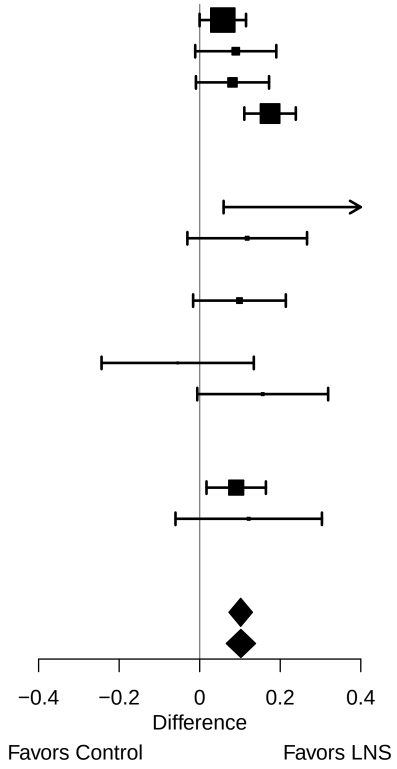

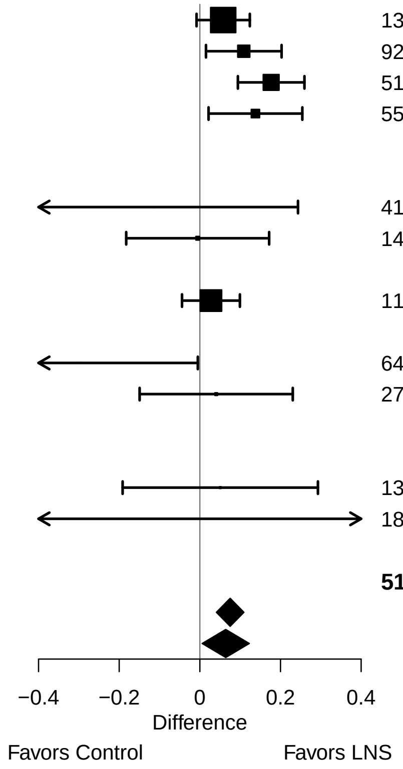

### Supplemental figure 7P: Small head size prevalence ratio

#### 7P1: Stratified by Household socio-economic status

**P−for−interaction = 0.462**
**Ratio of PRs = 1.04 (0.94, 1.14)**

| | | At least median | | | | | | Less than median | | | | | |
|---|---|---|---|---|---|---|---|---|---|---|---|---|---|
| **Country** | **Trial** | **LNS N** | **Control N** | **Control Prevalence** | **PR (95% CI)** | **Fixed W** | **Random W** | **LNS N** | **Control N** | **Control Prevalence** | **PR (95% CI)** | **Fixed W** | **Random W** |
| Bangladesh | JiVitA−4 (34) | 1456 | 640 | 20.2 | 1.00 (0.82, 1.23) | 0.13 | 0.13 | 1428 | 627 | 28.1 | 1.05 (0.90, 1.23) | 0.17 | 0.17 |
| Bangladesh | RDNS (35) | 834 | 407 | 39.1 | 0.89 (0.76, 1.04) | 0.23 | 0.23 | 829 | 408 | 46.8 | 0.90 (0.80, 1.01) | 0.32 | 0.30 |
| Bangladesh | WASH−B (36) | 583 | 1740 | 29.1 | 0.79 (0.67, 0.93) | 0.22 | 0.22 | 580 | 1723 | 36.5 | 1.00 (0.88, 1.13) | 0.27 | 0.25 |
| Burkina Faso | iLiNS−Zinc (37) | 1226 | 299 | 39.3 | 0.94 (0.81, 1.08) | 0.28 | 0.28 | 713 | 364 | 42.2 | 0.82 (0.68, 0.99) | 0.12 | 0.13 |
| Burkina Faso | PROMIS (38) | | | | | | | | | | | | |
| Burkina Faso | PROMIS CS (38) | | | | | | | | | | | | |
| Ghana | GHANA (39) | | | | | | | | | | | | |
| Ghana | iLiNS−DYADG (40) | 157 | 355 | 14.1 | 0.72 (0.43, 1.23) | 0.02 | 0.02 | 190 | 335 | 18.2 | 0.95 (0.65, 1.40) | 0.03 | 0.03 |
| Haiti | HAITI (41) | | | | | | | | | | | | |
| Kenya | WASH−B (42) | 827 | 2946 | 3.5 | 0.90 (0.57, 1.41) | 0.03 | 0.03 | 632 | 2226 | 5.4 | 0.93 (0.61, 1.43) | 0.02 | 0.03 |
| Madagascar | MAHAY (43) | | | | | | | | | | | | |
| Malawi | iLiNS−DYADM (44) | 121 | 234 | 6.8 | 1.69 (0.85, 3.35) | 0.01 | 0.01 | 99 | 209 | 14.8 | 0.48 (0.22, 1.04) | 0.01 | 0.01 |
| Malawi | iLiNS−DOSE (45) | 302 | 102 | 19.4 | 0.82 (0.53, 1.26) | 0.03 | 0.03 | 290 | 102 | 23.5 | 0.81 (0.55, 1.20) | 0.03 | 0.03 |
| Mali | PROMIS (46) | | | | | | | | | | | | |
| Mali | PROMIS CS (46) | | | | | | | | | | | | |
| Zimbabwe | SHINE (HIV−) (47) | 905 | 799 | 4.6 | 0.81 (0.53, 1.23) | 0.03 | 0.03 | 826 | 853 | 6.9 | 0.84 (0.59, 1.19) | 0.03 | 0.04 |
| Zimbabwe | SHINE (HIV+) (48) | 171 | 156 | 10.9 | 0.80 (0.41, 1.58) | 0.01 | 0.01 | 158 | 167 | 8.4 | 1.28 (0.62, 2.64) | 0.01 | 0.01 |
| | | **6582** | **7678** | | **I² = 0.00, Tau² = 0.00** | | | **5745** | **7014** | | **I² = 0.09, Tau² = 0.00** | | |
| **Fixed** | | | | | **0.89 (0.83, 0.96)** | | | | | | **0.93 (0.88, 1.00)** | | |
| **Random** | | | | | **0.89 (0.83, 0.96)** | | | | | | **0.93 (0.87, 1.00)** | | |

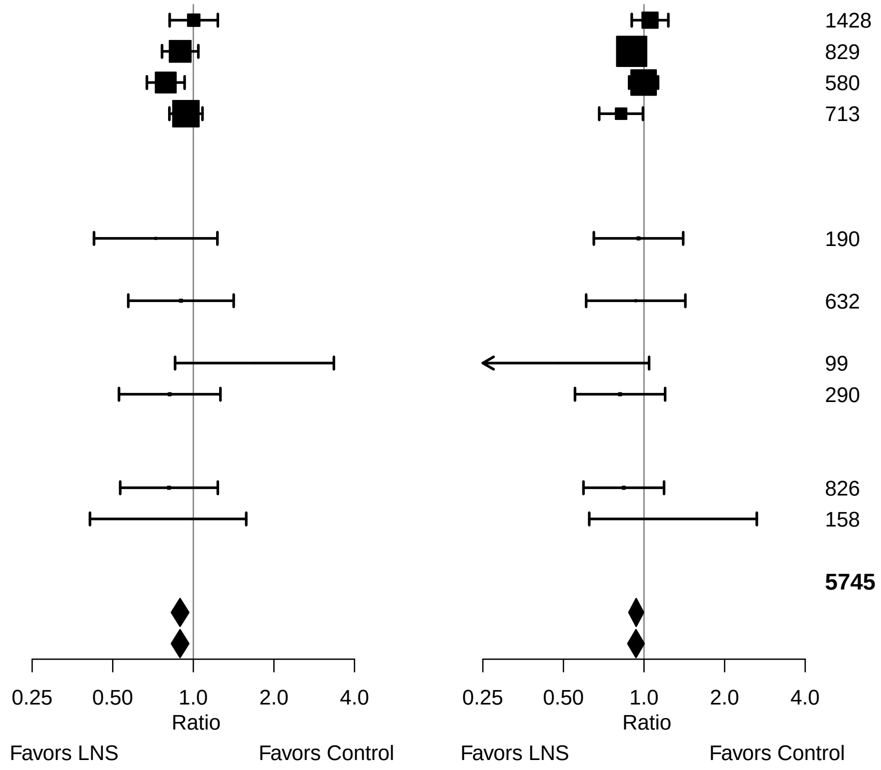

#### Supplemental figure 7P: Small head size prevalence ratio

#### 7P2: Stratified by Household food insecurity

**P−for−interaction = 0.542**
**Ratio of PRs = 1.03 (0.94, 1.13)**

| | | Mild to secure | | | | | | Moderate to severe | | | | | |
|---|---|---|---|---|---|---|---|---|---|---|---|---|---|
| **Country** | **Trial** | **LNS N** | **Control N** | **Control Prevalence** | **PR (95% CI)** | **Fixed W** | **Random W** | **LNS N** | **Control N** | **Control Prevalence** | **PR (95% CI)** | **Fixed W** | **Random W** |
| Bangladesh | JiVitA−4 (34) | 2045 | 882 | 23.6 | 0.88 (0.75, 1.03) | 0.13 | 0.13 | 839 | 385 | 25.2 | 1.33 (1.07, 1.64) | 0.15 | 0.16 |
| Bangladesh | RDNS (35) | 1058 | 493 | 40.8 | 0.91 (0.82, 1.01) | 0.30 | 0.30 | 605 | 322 | 46.3 | 0.88 (0.76, 1.02) | 0.33 | 0.21 |
| Bangladesh | WASH−B (36) | 922 | 2682 | 31.5 | 0.90 (0.80, 1.01) | 0.24 | 0.24 | 241 | 781 | 37.4 | 0.94 (0.77, 1.15) | 0.16 | 0.17 |
| Burkina Faso | iLiNS−Zinc (37) | 1008 | 320 | 38.3 | 0.92 (0.82, 1.03) | 0.24 | 0.24 | 931 | 343 | 43.3 | 0.85 (0.71, 1.02) | 0.20 | 0.18 |
| Burkina Faso | PROMIS (38) | | | | | | | | | | | | |
| Burkina Faso | PROMIS CS (38) | | | | | | | | | | | | |
| Ghana | GHANA (39) | | | | | | | | | | | | |
| Ghana | iLiNS−DYADG (40) | 244 | 470 | 14.7 | 0.81 (0.54, 1.21) | 0.02 | 0.02 | 102 | 219 | 19.2 | 1.02 (0.63, 1.65) | 0.03 | 0.06 |
| Haiti | HAITI (41) | | | | | | | | | | | | |
| Kenya | WASH−B (42) | 1303 | 4632 | 4.1 | 0.96 (0.70, 1.33) | 0.03 | 0.03 | 156 | 527 | 6.6 | 0.68 (0.29, 1.56) | 0.01 | 0.02 |
| Madagascar | MAHAY (43) | | | | | | | | | | | | |
| Malawi | iLiNS−DYADM (44) | 70 | 122 | 9.0 | 0.79 (0.29, 2.19) | 0.00 | 0.00 | 148 | 316 | 11.4 | 0.95 (0.54, 1.65) | 0.02 | 0.04 |
| Malawi | iLiNS−DOSE (45) | 149 | 55 | 16.4 | 1.11 (0.57, 2.16) | 0.01 | 0.01 | 431 | 143 | 22.9 | 0.74 (0.54, 1.03) | 0.06 | 0.10 |
| Mali | PROMIS (46) | | | | | | | | | | | | |
| Mali | PROMIS CS (46) | | | | | | | | | | | | |
| Zimbabwe | SHINE (HIV−) (47) | 1385 | 1297 | 5.7 | 0.71 (0.51, 0.98) | 0.03 | 0.03 | 295 | 323 | 6.2 | 1.37 (0.80, 2.35) | 0.02 | 0.04 |
| Zimbabwe | SHINE (HIV+) (48) | 236 | 239 | 9.6 | 1.10 (0.63, 1.91) | 0.01 | 0.01 | 86 | 76 | 10.5 | 0.77 (0.31, 1.93) | 0.01 | 0.02 |
| | | **8420** | **11192** | | **I² = 0.00, Tau² = 0.00** | | | **3834** | **3435** | | **I² = 0.46, Tau² = 0.01** | | |
| **Fixed** | | | | | **0.90 (0.85, 0.95)** | | | | | | **0.94 (0.87, 1.02)** | | |
| **Random** | | | | | **0.90 (0.85, 0.95)** | | | | | | **0.95 (0.84, 1.08)** | | |

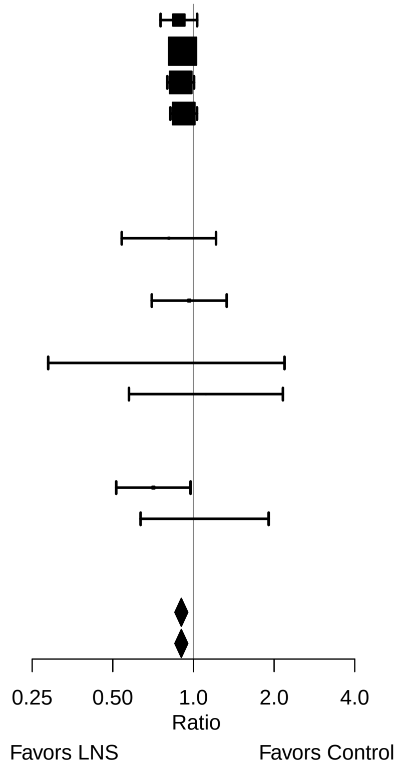

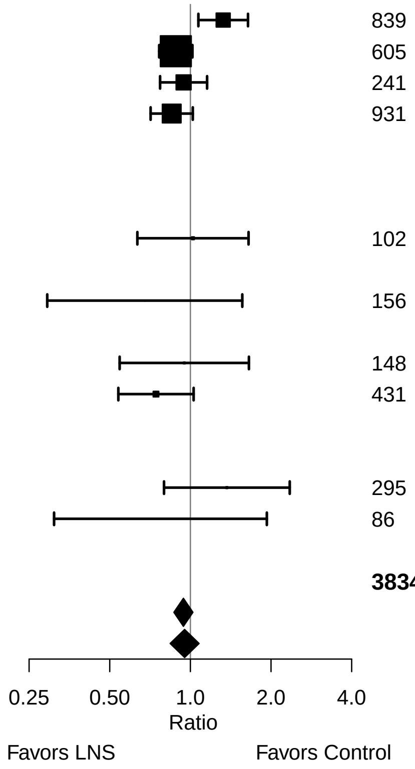

#### Supplemental figure 7P: Small head size prevalence ratio

##### 7P3: Stratified by Household source water quality

**P−for−interaction = 0.505**
**Ratio of PRs = 0.93 (0.74, 1.16)**

| Country | Trial | Improved LNS N | Improved Control N | Improved Control Prevalence | Improved PR (95% CI) | Improved Fixed W | Improved Random W | Unimproved LNS N | Unimproved Control N | Unimproved Control Prevalence | Unimproved PR (95% CI) | Unimproved Fixed W | Unimproved Random W |
|---|---|---|---|---|---|---|---|---|---|---|---|---|---|
| Bangladesh | JiVitA−4 (34) | | | | | | | | | | | | |
| Bangladesh | RDNS (35) | | | | | | | | | | | | |
| Bangladesh | WASH−B (36) | 500 | 1004 | 34.3 | 0.84 (0.70, 1.01) | 0.50 | 0.50 | 70 | 116 | 28.4 | 1.05 (0.69, 1.61) | 0.08 | 0.08 |
| Burkina Faso | iLiNS−Zinc (37) | 552 | 141 | 36.9 | 1.05 (0.78, 1.42) | 0.19 | 0.19 | 1386 | 519 | 41.8 | 0.85 (0.75, 0.96) | 0.86 | 0.86 |
| Burkina Faso | PROMIS (38) | | | | | | | | | | | | |
| Burkina Faso | PROMIS CS (38) | | | | | | | | | | | | |
| Ghana | GHANA (39) | | | | | | | | | | | | |
| Ghana | iLiNS−DYADG (40) | | | | | | | | | | | | |
| Haiti | HAITI (41) | | | | | | | | | | | | |
| Kenya | WASH−B (42) | 448 | 1560 | 4.2 | 0.69 (0.36, 1.30) | 0.04 | 0.04 | 248 | 699 | 5.2 | 0.78 (0.40, 1.55) | 0.03 | 0.03 |
| Madagascar | MAHAY (43) | | | | | | | | | | | | |
| Malawi | iLiNS−DYADM (44) | | | | | | | | | | | | |
| Malawi | iLiNS−DOSE (45) | 553 | 192 | 20.8 | 0.84 (0.62, 1.14) | 0.19 | 0.19 | 51 | 15 | 31.2 | 0.48 (0.20, 1.13) | 0.02 | 0.02 |
| Mali | PROMIS (46) | | | | | | | | | | | | |
| Mali | PROMIS CS (46) | | | | | | | | | | | | |
| Zimbabwe | SHINE (HIV−) (47) | 518 | 485 | 5.8 | 0.90 (0.58, 1.42) | 0.08 | 0.08 | 272 | 299 | 4.0 | 1.28 (0.59, 2.77) | 0.02 | 0.02 |
| Zimbabwe | SHINE (HIV+) (48) | | | | | | | | | | | | |
| | | **2571** | **3382** | | **I² = 0.00, Tau² = 0.00** | | | **2027** | **1648** | | **I² = 0.00, Tau² = 0.00** | | |
| **Fixed** | | | | | **0.88 (0.77, 1.00)** | | | | | | **0.86 (0.77, 0.96)** | | |
| **Random** | | | | | **0.88 (0.77, 1.00)** | | | | | | **0.86 (0.77, 0.96)** | | |

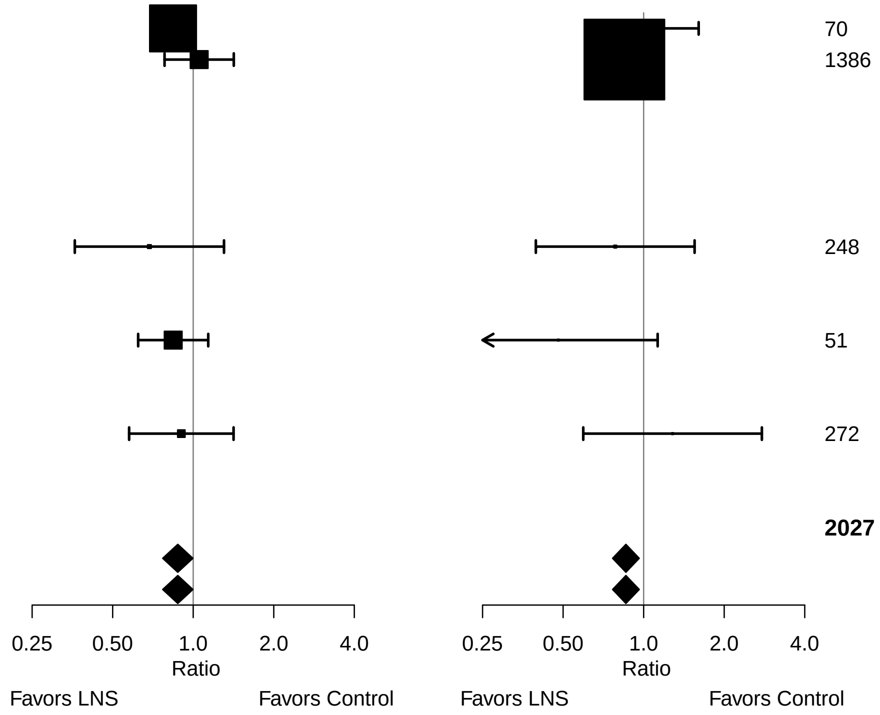

#### Supplemental figure 7P: Small head size prevalence ratio

#### 7P4: Stratified by Household sanitation

**P−for−interaction = 0.109**
**Ratio of PRs = 1.13 (0.97, 1.31)**

| | | Improved | | | | | | Unimproved | | | | | |
|---|---|---|---|---|---|---|---|---|---|---|---|---|---|
| **Country** | **Trial** | **LNS N** | **Control N** | **Control Prevalence** | **PR (95% CI)** | **Fixed W** | **Random W** | **LNS N** | **Control N** | **Control Prevalence** | **PR (95% CI)** | **Fixed W** | **Random W** |
| Bangladesh | JiVitA−4 (34) | 2231 | 972 | 22.9 | 1.01 (0.86, 1.19) | 0.23 | 0.28 | 653 | 295 | 27.9 | 1.09 (0.88, 1.35) | 0.18 | 0.18 |
| Bangladesh | RDNS (35) | 1179 | 573 | 42.8 | 0.86 (0.77, 0.95) | 0.56 | 0.45 | 482 | 241 | 43.6 | 0.98 (0.84, 1.15) | 0.32 | 0.32 |
| Bangladesh | WASH−B (36) | 510 | 1017 | 33.3 | 0.87 (0.72, 1.05) | 0.18 | 0.24 | 31 | 55 | 27.3 | 1.06 (0.54, 2.10) | 0.02 | 0.02 |
| Burkina Faso | iLiNS−Zinc (37) | 43 | 16 | 50.0 | 0.66 (0.34, 1.28) | 0.01 | 0.03 | 1895 | 644 | 40.6 | 0.89 (0.78, 1.02) | 0.45 | 0.45 |
| Burkina Faso | PROMIS (38) | | | | | | | | | | | | |
| Burkina Faso | PROMIS CS (38) | | | | | | | | | | | | |
| Ghana | GHANA (39) | | | | | | | | | | | | |
| Ghana | iLiNS−DYADG (40) | | | | | | | | | | | | |
| Haiti | HAITI (41) | | | | | | | | | | | | |
| Kenya | WASH−B (42) | | | | | | | | | | | | |
| Madagascar | MAHAY (43) | | | | | | | | | | | | |
| Malawi | iLiNS−DYADM (44) | | | | | | | | | | | | |
| Malawi | iLiNS−DOSE (45) | | | | | | | | | | | | |
| Mali | PROMIS (46) | | | | | | | | | | | | |
| Mali | PROMIS CS (46) | | | | | | | | | | | | |
| Zimbabwe | SHINE (HIV−) (47) | 293 | 243 | 3.3 | 1.56 (0.65, 3.69) | 0.01 | 0.02 | 494 | 540 | 5.9 | 0.85 (0.53, 1.37) | 0.04 | 0.04 |
| Zimbabwe | SHINE (HIV+) (48) | | | | | | | | | | | | |
| | | **4256** | **2821** | | **I² = 0.23, Tau² = 0.00** | | | **3555** | **1775** | | **I² = 0.00, Tau² = 0.00** | | |
| **Fixed** | | | | | **0.90 (0.83, 0.97)** | | | | | | **0.96 (0.87, 1.05)** | | |
| **Random** | | | | | **0.90 (0.81, 1.01)** | | | | | | **0.96 (0.87, 1.05)** | | |

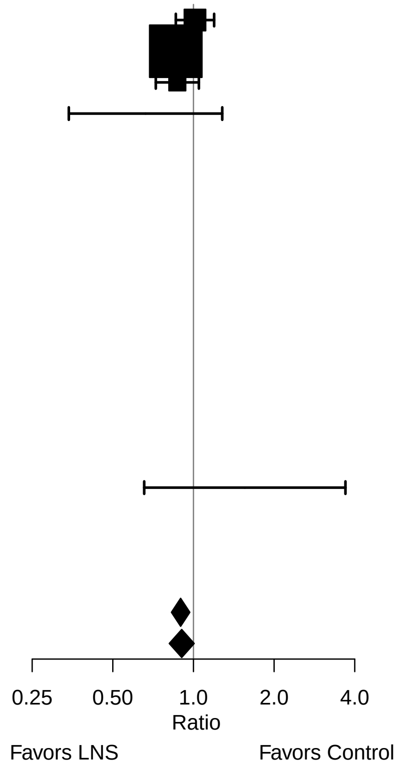

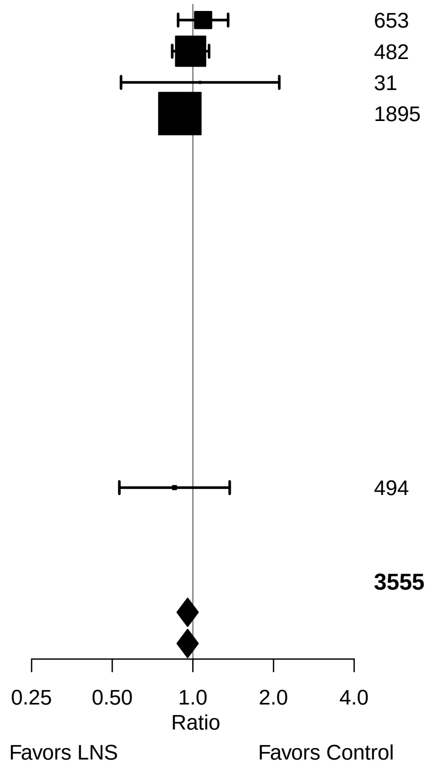

#### Supplemental figure 7P: Small head size prevalence ratio

##### 7P5: Stratified by Home environment

**P−for−interaction = 0.826**
**Ratio of PRs = 0.99 (0.89, 1.10)**

| | | At least median | | | | | | Less than median | | | | | |
|---|---|---|---|---|---|---|---|---|---|---|---|---|---|
| **Country** | **Trial** | **LNS N** | **Control N** | **Control Prevalence** | **PR (95% CI)** | **Fixed W** | **Random W** | **LNS N** | **Control N** | **Control Prevalence** | **PR (95% CI)** | **Fixed W** | **Random W** |
| Bangladesh | JiVitA−4 (34) | | | | | | | | | | | | |
| Bangladesh | RDNS (35) | 1192 | 582 | 41.4 | 0.91 (0.82, 1.01) | 0.53 | 0.53 | 468 | 230 | 46.5 | 0.87 (0.77, 0.98) | 0.36 | 0.36 |
| Bangladesh | WASH−B (36) | 641 | 1665 | 30.9 | 0.86 (0.73, 1.01) | 0.24 | 0.24 | 494 | 1707 | 34.9 | 0.97 (0.85, 1.11) | 0.33 | 0.33 |
| Burkina Faso | iLiNS−Zinc (37) | 471 | 199 | 36.2 | 0.93 (0.73, 1.18) | 0.10 | 0.10 | 270 | 174 | 44.0 | 0.90 (0.76, 1.05) | 0.22 | 0.22 |
| Burkina Faso | PROMIS (38) | | | | | | | | | | | | |
| Burkina Faso | PROMIS CS (38) | | | | | | | | | | | | |
| Ghana | GHANA (39) | | | | | | | | | | | | |
| Ghana | iLiNS−DYADG (40) | 213 | 410 | 14.1 | 0.93 (0.61, 1.41) | 0.03 | 0.03 | 119 | 247 | 18.2 | 0.92 (0.57, 1.49) | 0.02 | 0.02 |
| Haiti | HAITI (41) | | | | | | | | | | | | |
| Kenya | WASH−B (42) | 839 | 2778 | 3.1 | 1.22 (0.82, 1.81) | 0.04 | 0.04 | 574 | 2174 | 5.1 | 0.78 (0.49, 1.25) | 0.03 | 0.03 |
| Madagascar | MAHAY (43) | | | | | | | | | | | | |
| Malawi | iLiNS−DYADM (44) | 137 | 289 | 9.0 | 0.89 (0.45, 1.75) | 0.01 | 0.01 | 78 | 149 | 14.8 | 0.87 (0.43, 1.74) | 0.01 | 0.01 |
| Malawi | iLiNS−DOSE (45) | 377 | 135 | 19.3 | 0.80 (0.54, 1.17) | 0.04 | 0.04 | 272 | 85 | 23.3 | 0.75 (0.49, 1.13) | 0.03 | 0.03 |
| Mali | PROMIS (46) | | | | | | | | | | | | |
| Mali | PROMIS CS (46) | | | | | | | | | | | | |
| Zimbabwe | SHINE (HIV−) (47) | | | | | | | | | | | | |
| Zimbabwe | SHINE (HIV+) (48) | | | | | | | | | | | | |
| | | **3870** | **6058** | | **I² = 0.00, Tau² = 0.00** | | | **2275** | **4766** | | **I² = 0.00, Tau² = 0.00** | | |
| **Fixed** | | | | | **0.90 (0.84, 0.98)** | | | | | | **0.90 (0.84, 0.97)** | | |
| **Random** | | | | | **0.90 (0.84, 0.98)** | | | | | | **0.90 (0.84, 0.97)** | | |

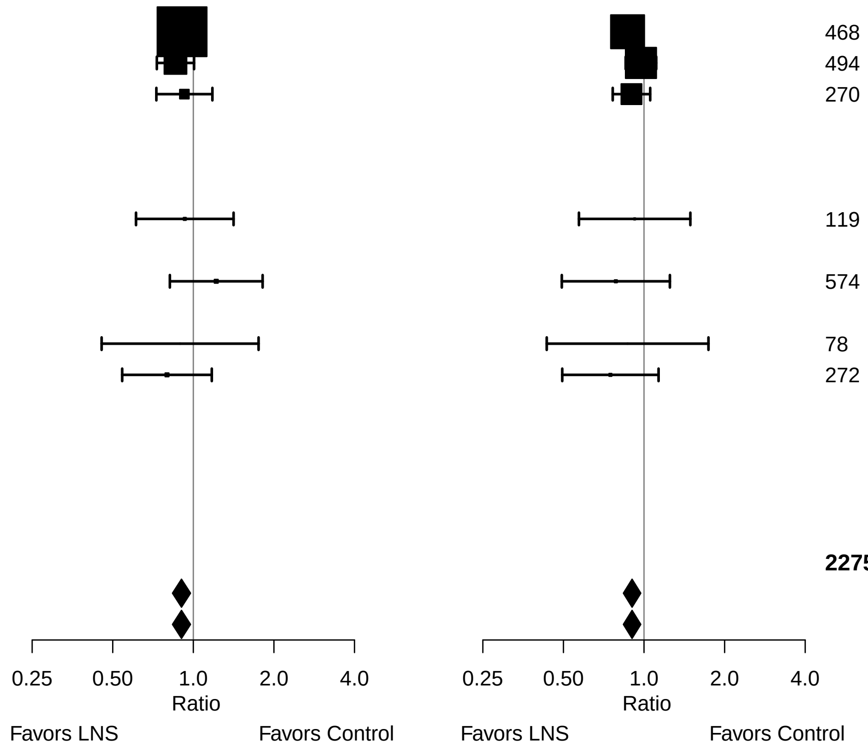

#### Supplemental figure 7P: Small head size prevalence ratio

### 7P6: Stratified by Season at the time of assessment

**P−for−interaction = 0.434**
**Ratio of PRs = 1.04 (0.94, 1.15)**

| Country | Trial | Dry LNS N | Dry Control N | Dry Control Prevalence | Dry PR (95% CI) | Dry Fixed W | Dry Random W | Rainy LNS N | Rainy Control N | Rainy Control Prevalence | Rainy PR (95% CI) | Rainy Fixed W | Rainy Random W |
|---|---|---|---|---|---|---|---|---|---|---|---|---|---|
| Bangladesh | JiVitA−4 (34) | 1566 | 674 | 24.6 | 1.00 (0.85, 1.17) | 0.16 | 0.16 | 1319 | 593 | 23.5 | 1.08 (0.88, 1.32) | 0.13 | 0.13 |
| Bangladesh | RDNS (35) | 739 | 359 | 39.6 | 0.90 (0.80, 1.02) | 0.27 | 0.27 | 924 | 456 | 45.6 | 0.89 (0.79, 1.00) | 0.42 | 0.42 |
| Bangladesh | WASH−B (36) | 644 | 1936 | 31.8 | 0.92 (0.80, 1.06) | 0.23 | 0.23 | 519 | 1527 | 34.1 | 0.88 (0.76, 1.02) | 0.25 | 0.25 |
| Burkina Faso | iLiNS−Zinc (37) | 1387 | 477 | 40.9 | 0.87 (0.77, 0.99) | 0.26 | 0.26 | 559 | 187 | 41.3 | 0.93 (0.72, 1.19) | 0.09 | 0.09 |
| Burkina Faso | PROMIS (38) | | | | | | | | | | | | |
| Burkina Faso | PROMIS CS (38) | | | | | | | | | | | | |
| Ghana | GHANA (39) | | | | | | | | | | | | |
| Ghana | iLiNS−DYADG (40) | 204 | 408 | 18.4 | 0.72 (0.48, 1.08) | 0.03 | 0.03 | 143 | 284 | 12.7 | 1.21 (0.74, 1.98) | 0.02 | 0.02 |
| Haiti | HAITI (41) | | | | | | | | | | | | |
| Kenya | WASH−B (42) | 349 | 1161 | 4.9 | 0.99 (0.55, 1.80) | 0.01 | 0.01 | 1112 | 4015 | 4.2 | 0.89 (0.62, 1.26) | 0.04 | 0.04 |
| Madagascar | MAHAY (43) | | | | | | | | | | | | |
| Malawi | iLiNS−DYADM (44) | 156 | 316 | 10.8 | 0.66 (0.34, 1.26) | 0.01 | 0.01 | 64 | 128 | 10.9 | 1.43 (0.67, 3.04) | 0.01 | 0.01 |
| Malawi | iLiNS−DOSE (45) | 418 | 133 | 25.4 | 0.67 (0.47, 0.94) | 0.04 | 0.04 | 274 | 105 | 17.1 | 0.98 (0.64, 1.51) | 0.03 | 0.03 |
| Mali | PROMIS (46) | | | | | | | | | | | | |
| Mali | PROMIS CS (46) | | | | | | | | | | | | |
| Zimbabwe | SHINE (HIV−) (47) | | | | | | | | | | | | |
| Zimbabwe | SHINE (HIV+) (48) | | | | | | | | | | | | |
| | | **5463** | **5464** | | **I² = 0.00, Tau² = 0.00** | | | **4914** | **7295** | | **I² = 0.00, Tau² = 0.00** | | |
| **Fixed** | | | | | **0.90 (0.84, 0.96)** | | | | | | **0.93 (0.86, 1.00)** | | |
| **Random** | | | | | **0.90 (0.84, 0.96)** | | | | | | **0.93 (0.86, 1.00)** | | |

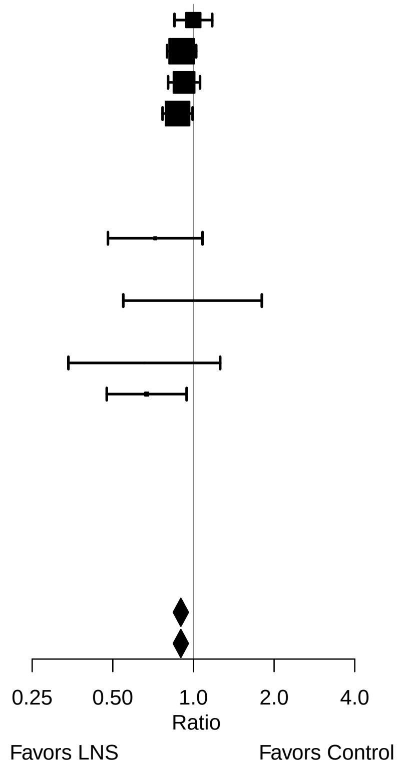

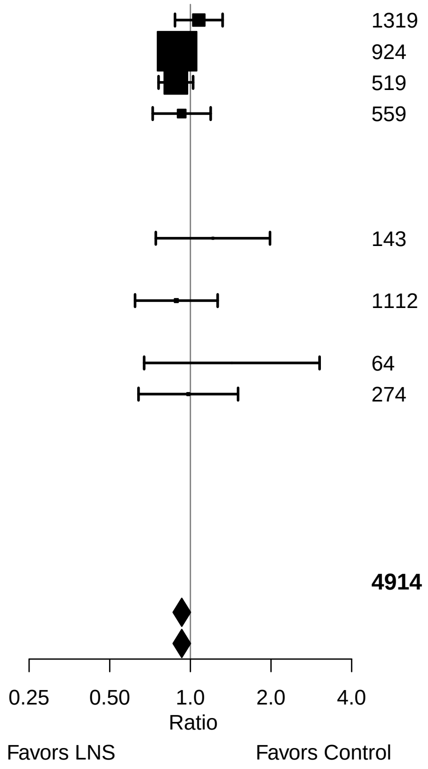

### Supplemental figure 7Q: Small head size prevalence difference

#### 7Q1: Stratified by Household socio-economic status

**P−for−interaction = 0.965**
**Difference in PDs = 0.00 (−0.01, 0.02)**

| Country | Trial | At least median: LNS N | Control N | Control Prevalence | PD (95% CI) | Fixed W | Random W | Less than median: LNS N | Control N | Control Prevalence | PD (95% CI) | Fixed W | Random W |
|---|---|---|---|---|---|---|---|---|---|---|---|---|---|
| Bangladesh | JiVitA−4 (34) | 1456 | 640 | 20.2 | −0.01 (−0.04, 0.03) | 0.07 | 0.11 | 1428 | 627 | 28.1 | 0.00 (−0.03, 0.04) | 0.11 | 0.13 |
| Bangladesh | RDNS (35) | 834 | 407 | 39.1 | −0.04 (−0.10, 0.02) | 0.03 | 0.05 | 829 | 408 | 46.8 | −0.05 (−0.10, 0.00) | 0.06 | 0.07 |
| Bangladesh | WASH−B (36) | 583 | 1740 | 29.1 | −0.06 (−0.10, −0.02) | 0.06 | 0.10 | 580 | 1723 | 36.5 | 0.00 (−0.05, 0.04) | 0.07 | 0.09 |
| Burkina Faso | iLiNS−Zinc (37) | 1226 | 299 | 39.3 | −0.03 (−0.06, 0.01) | 0.07 | 0.11 | 713 | 364 | 42.2 | −0.07 (−0.14, 0.01) | 0.03 | 0.03 |
| Burkina Faso | PROMIS (38) | | | | | | | | | | | | |
| Burkina Faso | PROMIS CS (38) | | | | | | | | | | | | |
| Ghana | GHANA (39) | | | | | | | | | | | | |
| Ghana | iLiNS−DYADG (40) | 157 | 355 | 14.1 | −0.04 (−0.10, 0.02) | 0.02 | 0.05 | 190 | 335 | 18.2 | −0.01 (−0.08, 0.06) | 0.03 | 0.04 |
| Haiti | HAITI (41) | | | | | | | | | | | | |
| Kenya | WASH−B (42) | 827 | 2946 | 3.5 | 0.00 (−0.02, 0.01) | 0.42 | 0.24 | 632 | 2226 | 5.4 | 0.00 (−0.03, 0.02) | 0.31 | 0.27 |
| Madagascar | MAHAY (43) | | | | | | | | | | | | |
| Malawi | iLiNS−DYADM (44) | 121 | 234 | 6.8 | 0.05 (−0.01, 0.11) | 0.02 | 0.05 | 99 | 209 | 14.8 | −0.08 (−0.16, 0.00) | 0.02 | 0.03 |
| Malawi | iLiNS−DOSE (45) | 302 | 102 | 19.4 | −0.02 (−0.10, 0.06) | 0.01 | 0.03 | 290 | 102 | 23.5 | −0.05 (−0.13, 0.03) | 0.02 | 0.03 |
| Mali | PROMIS (46) | | | | | | | | | | | | |
| Mali | PROMIS CS (46) | | | | | | | | | | | | |
| Zimbabwe | SHINE (HIV−) (47) | 905 | 799 | 4.6 | −0.01 (−0.03, 0.01) | 0.28 | 0.22 | 826 | 853 | 6.9 | −0.01 (−0.03, 0.01) | 0.31 | 0.27 |
| Zimbabwe | SHINE (HIV+) (48) | 171 | 156 | 10.9 | −0.02 (−0.09, 0.04) | 0.02 | 0.05 | 158 | 167 | 8.4 | 0.02 (−0.04, 0.09) | 0.03 | 0.04 |
| | | **6582** | **7678** | | **I² = 0.34, Tau² = 0.00** | | | **5745** | **7014** | | **I² = 0.09, Tau² = 0.00** | | |
| **Fixed** | | | | | **−0.01 (−0.02, 0.00)** | | | | | | **−0.01 (−0.02, 0.00)** | | |
| **Random** | | | | | **−0.02 (−0.03, 0.00)** | | | | | | **−0.01 (−0.03, 0.00)** | | |

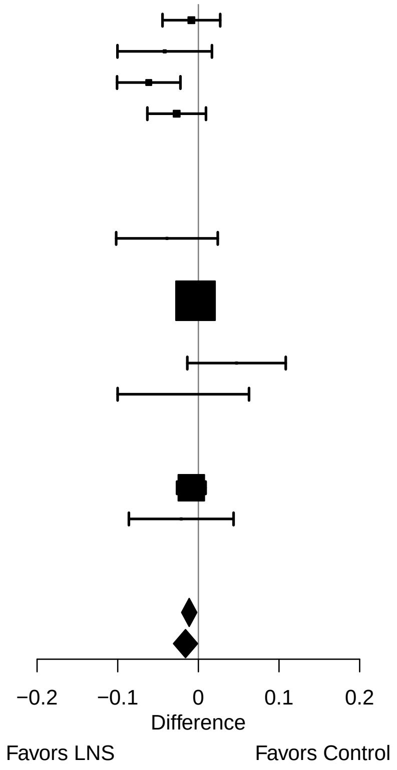

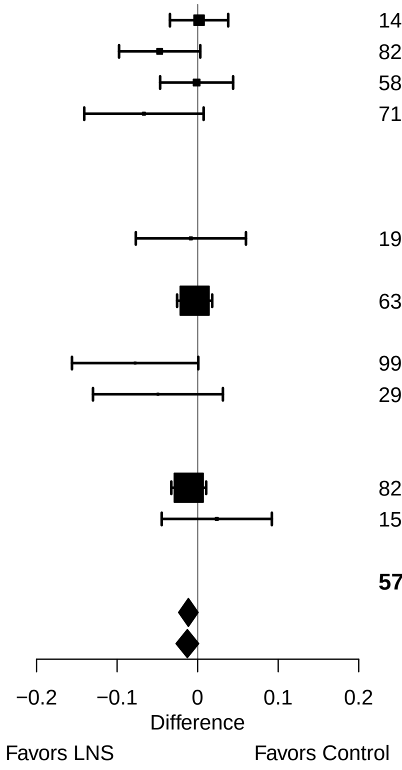

#### Supplemental figure 7Q: Small head size prevalence difference

##### 7Q2: Stratified by Household food insecurity

**P−for−interaction = 0.396**
**Difference in PDs = 0.01 (−0.01, 0.03)**

| | | | | | Mild to secure | | | | | | Moderate to severe | | |
|---|---|---|---|---|---|---|---|---|---|---|---|---|---|
| Country | Trial | LNS N | Control N | Control Prevalence | PD (95% CI) | Fixed W | Random W | LNS N | Control N | Control Prevalence | PD (95% CI) | Fixed W | Random W |
| Bangladesh | JiVitA−4 (34) | 2045 | 882 | 23.6 | −0.02 (−0.06, 0.01) | 0.06 | 0.06 | 839 | 385 | 25.2 | 0.04 (0.00, 0.09) | 0.17 | 0.15 |
| Bangladesh | RDNS (35) | 1058 | 493 | 40.8 | −0.04 (−0.08, 0.00) | 0.04 | 0.04 | 605 | 322 | 46.3 | −0.05 (−0.12, 0.01) | 0.08 | 0.09 |
| Bangladesh | WASH−B (36) | 922 | 2682 | 31.5 | −0.03 (−0.07, 0.00) | 0.06 | 0.06 | 241 | 781 | 37.4 | −0.02 (−0.09, 0.05) | 0.06 | 0.08 |
| Burkina Faso | iLiNS−Zinc (37) | 1008 | 320 | 38.3 | −0.03 (−0.06, 0.01) | 0.05 | 0.05 | 931 | 343 | 43.3 | −0.06 (−0.13, 0.01) | 0.06 | 0.08 |
| Burkina Faso | PROMIS (38) | | | | | | | | | | | | |
| Burkina Faso | PROMIS CS (38) | | | | | | | | | | | | |
| Ghana | GHANA (39) | | | | | | | | | | | | |
| Ghana | iLiNS−DYADG (40) | 244 | 470 | 14.7 | −0.03 (−0.08, 0.03) | 0.02 | 0.02 | 102 | 219 | 19.2 | 0.00 (−0.09, 0.10) | 0.04 | 0.05 |
| Haiti | HAITI (41) | | | | | | | | | | | | |
| Kenya | WASH−B (42) | 1303 | 4632 | 4.1 | 0.00 (−0.01, 0.01) | 0.43 | 0.43 | 156 | 527 | 6.6 | −0.02 (−0.06, 0.02) | 0.19 | 0.16 |
| Madagascar | MAHAY (43) | | | | | | | | | | | | |
| Malawi | iLiNS−DYADM (44) | 70 | 122 | 9.0 | −0.02 (−0.10, 0.06) | 0.01 | 0.01 | 148 | 316 | 11.4 | −0.01 (−0.07, 0.06) | 0.08 | 0.10 |
| Malawi | iLiNS−DOSE (45) | 149 | 55 | 16.4 | 0.03 (−0.08, 0.14) | 0.01 | 0.01 | 431 | 143 | 22.9 | −0.05 (−0.12, 0.01) | 0.07 | 0.08 |
| Mali | PROMIS (46) | | | | | | | | | | | | |
| Mali | PROMIS CS (46) | | | | | | | | | | | | |
| Zimbabwe | SHINE (HIV−) (47) | 1385 | 1297 | 5.7 | −0.02 (−0.03, 0.00) | 0.29 | 0.29 | 295 | 323 | 6.2 | 0.02 (−0.02, 0.06) | 0.20 | 0.16 |
| Zimbabwe | SHINE (HIV+) (48) | 236 | 239 | 9.6 | 0.01 (−0.05, 0.07) | 0.02 | 0.02 | 86 | 76 | 10.5 | −0.02 (−0.11, 0.06) | 0.04 | 0.06 |
| | | **8420** | **11192** | | **I² = 0.00, Tau² = 0.00** | | | **3834** | **3435** | | **I² = 0.37, Tau² = 0.00** | | |
| **Fixed** | | | | | **−0.01 (−0.02, 0.00)** | | | | | | **−0.01 (−0.02, 0.01)** | | |
| **Random** | | | | | **−0.01 (−0.02, 0.00)** | | | | | | **−0.01 (−0.03, 0.01)** | | |

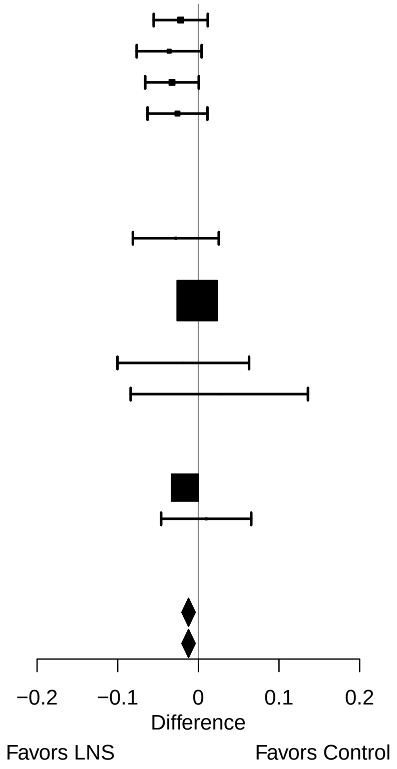

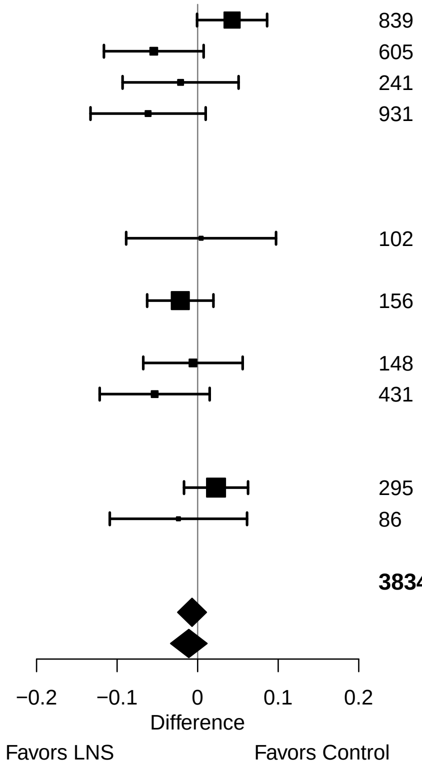

#### Supplemental figure 7Q: Small head size prevalence difference

##### 7Q3: Stratified by Household source water quality

**P−for−interaction = 0.976**
**Difference in PDs = 0.00 (−0.02, 0.03)**

| Country | Trial | LNS N | Control N | Control Prevalence | Improved PD (95% CI) | Fixed W | Random W | LNS N | Control N | Control Prevalence | Unimproved PD (95% CI) | Fixed W | Random W |
|---|---|---|---|---|---|---|---|---|---|---|---|---|---|
| Bangladesh | JiVitA−4 (34) | | | | | | | | | | | | |
| Bangladesh | RDNS (35) | | | | | | | | | | | | |
| Bangladesh | WASH−B (36) | 500 | 1004 | 34.3 | −0.05 (−0.11, 0.00) | 0.07 | 0.07 | 70 | 116 | 28.4 | 0.02 (−0.11, 0.14) | 0.03 | 0.09 |
| Burkina Faso | iLiNS−Zinc (37) | 552 | 141 | 36.9 | 0.00 (−0.09, 0.09) | 0.03 | 0.03 | 1386 | 519 | 41.8 | −0.06 (−0.11, 0.00) | 0.14 | 0.24 |
| Burkina Faso | PROMIS (38) | | | | | | | | | | | | |
| Burkina Faso | PROMIS CS (38) | | | | | | | | | | | | |
| Ghana | GHANA (39) | | | | | | | | | | | | |
| Ghana | iLiNS−DYADG (40) | | | | | | | | | | | | |
| Haiti | HAITI (41) | | | | | | | | | | | | |
| Kenya | WASH−B (42) | 448 | 1560 | 4.2 | −0.01 (−0.03, 0.01) | 0.52 | 0.52 | 248 | 699 | 5.2 | −0.01 (−0.04, 0.02) | 0.50 | 0.33 |
| Madagascar | MAHAY (43) | | | | | | | | | | | | |
| Malawi | iLiNS−DYADM (44) | | | | | | | | | | | | |
| Malawi | iLiNS−DOSE (45) | 553 | 192 | 20.8 | −0.02 (−0.08, 0.04) | 0.06 | 0.06 | 51 | 15 | 31.2 | −0.19 (−0.39, 0.01) | 0.01 | 0.04 |
| Mali | PROMIS (46) | | | | | | | | | | | | |
| Mali | PROMIS CS (46) | | | | | | | | | | | | |
| Zimbabwe | SHINE (HIV−) (47) | 518 | 485 | 5.8 | −0.01 (−0.03, 0.02) | 0.33 | 0.33 | 272 | 299 | 4.0 | 0.01 (−0.02, 0.05) | 0.33 | 0.30 |
| Zimbabwe | SHINE (HIV+) (48) | | | | | | | | | | | | |
| | | **2571** | **3382** | | **I² = 0.00, Tau² = 0.00** | | | **2027** | **1648** | | **I² = 0.45, Tau² = 0.00** | | |
| **Fixed** | | | | | **−0.01 (−0.03, 0.00)** | | | | | | **−0.01 (−0.03, 0.01)** | | |
| **Random** | | | | | **−0.01 (−0.03, 0.00)** | | | | | | **−0.02 (−0.06, 0.02)** | | |

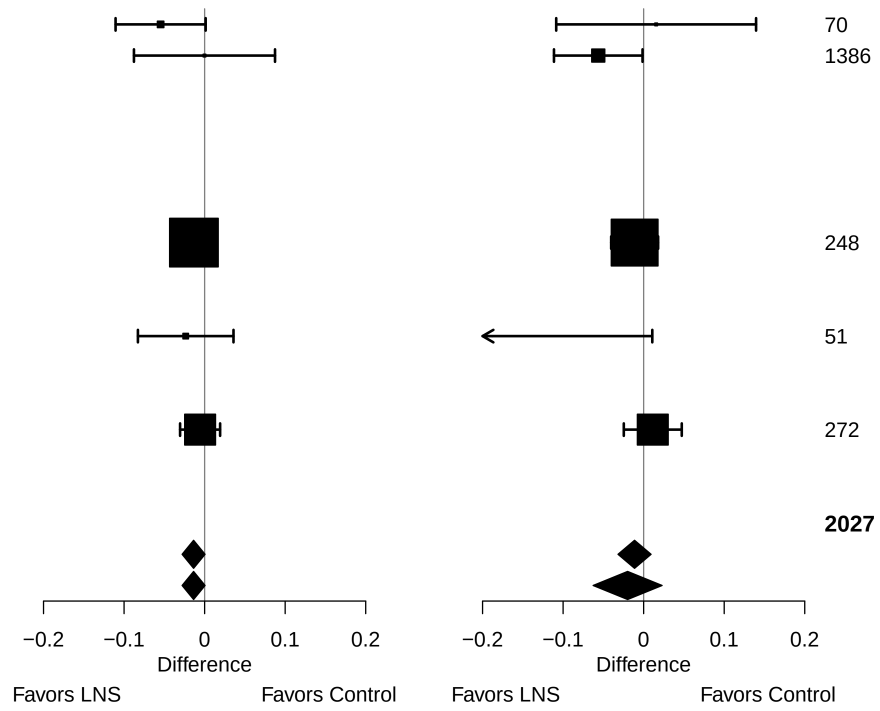

#### Supplemental figure 7Q: Small head size prevalence difference

#### 7Q4: Stratified by Household sanitation

**P−for−interaction = 0.794**
**Difference in PDs = 0.00 (−0.03, 0.04)**

| Country | Trial | LNS N | Control N | Control Prevalence | Improved PD (95% CI) | Fixed W | Random W | LNS N | Control N | Control Prevalence | Unimproved PD (95% CI) | Fixed W | Random W |
|---|---|---|---|---|---|---|---|---|---|---|---|---|---|
| Bangladesh | JiVitA−4 (34) | 2231 | 972 | 22.9 | −0.01 (−0.04, 0.02) | 0.37 | 0.29 | 653 | 295 | 27.9 | 0.02 (−0.03, 0.07) | 0.16 | 0.16 |
| Bangladesh | RDNS (35) | 1179 | 573 | 42.8 | −0.06 (−0.10, −0.02) | 0.20 | 0.23 | 482 | 241 | 43.6 | −0.01 (−0.08, 0.06) | 0.08 | 0.08 |
| Bangladesh | WASH−B (36) | 510 | 1017 | 33.3 | −0.04 (−0.10, 0.01) | 0.11 | 0.18 | 31 | 55 | 27.3 | 0.02 (−0.18, 0.21) | 0.01 | 0.01 |
| Burkina Faso | iLiNS−Zinc (37) | 43 | 16 | 50.0 | −0.11 (−0.33, 0.11) | 0.01 | 0.02 | 1895 | 644 | 40.6 | −0.04 (−0.09, 0.01) | 0.14 | 0.14 |
| Burkina Faso | PROMIS (38) | | | | | | | | | | | | |
| Burkina Faso | PROMIS CS (38) | | | | | | | | | | | | |
| Ghana | GHANA (39) | | | | | | | | | | | | |
| Ghana | iLiNS−DYADG (40) | | | | | | | | | | | | |
| Haiti | HAITI (41) | | | | | | | | | | | | |
| Kenya | WASH−B (42) | | | | | | | | | | | | |
| Madagascar | MAHAY (43) | | | | | | | | | | | | |
| Malawi | iLiNS−DYADM (44) | | | | | | | | | | | | |
| Malawi | iLiNS−DOSE (45) | | | | | | | | | | | | |
| Mali | PROMIS (46) | | | | | | | | | | | | |
| Mali | PROMIS CS (46) | | | | | | | | | | | | |
| Zimbabwe | SHINE (HIV−) (47) | 293 | 243 | 3.3 | 0.02 (−0.02, 0.05) | 0.31 | 0.28 | 494 | 540 | 5.9 | −0.01 (−0.03, 0.02) | 0.60 | 0.60 |
| Zimbabwe | SHINE (HIV+) (48) | | | | | | | | | | | | |
| | | **4256** | **2821** | | **I² = 0.60, Tau² = 0.00** | | | **3555** | **1775** | | **I² = 0.00, Tau² = 0.00** | | |
| **Fixed** | | | | | **−0.02 (−0.03, 0.00)** | | | | | | **−0.01 (−0.03, 0.01)** | | |
| **Random** | | | | | **−0.02 (−0.05, 0.01)** | | | | | | **−0.01 (−0.03, 0.01)** | | |

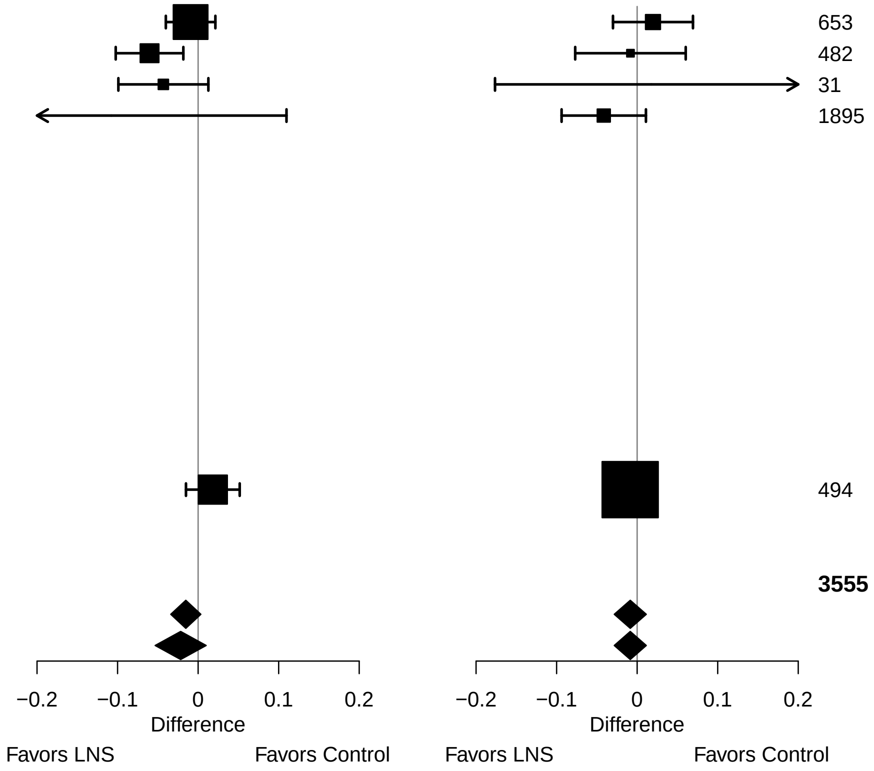

#### Supplemental figure 7Q: Small head size prevalence difference

#### 7Q5: Stratified by Home environment

**P−for−interaction = 0.291**
**Difference in PDs = −0.01 (−0.03, 0.01)**

| | | At least median | | | | | | Less than median | | | | | |
|---|---|---|---|---|---|---|---|---|---|---|---|---|---|
| **Country** | **Trial** | **LNS N** | **Control N** | **Control Prevalence** | **PD (95% CI)** | **Fixed W** | **Random W** | **LNS N** | **Control N** | **Control Prevalence** | **PD (95% CI)** | **Fixed W** | **Random W** |
| Bangladesh | JiVitA−4 (34) | | | | | | | | | | | | |
| Bangladesh | RDNS (35) | 1192 | 582 | 41.4 | −0.04 (−0.08, 0.00) | 0.08 | 0.12 | 468 | 230 | 46.5 | −0.06 (−0.11, −0.01) | 0.09 | 0.09 |
| Bangladesh | WASH−B (36) | 641 | 1665 | 30.9 | −0.04 (−0.09, 0.00) | 0.08 | 0.12 | 494 | 1707 | 34.9 | −0.01 (−0.05, 0.04) | 0.12 | 0.12 |
| Burkina Faso | iLiNS−Zinc (37) | 471 | 199 | 36.2 | −0.03 (−0.10, 0.05) | 0.03 | 0.04 | 270 | 174 | 44.0 | −0.01 (−0.08, 0.06) | 0.05 | 0.05 |
| Burkina Faso | PROMIS (38) | | | | | | | | | | | | |
| Burkina Faso | PROMIS CS (38) | | | | | | | | | | | | |
| Ghana | GHANA (39) | | | | | | | | | | | | |
| Ghana | iLiNS−DYADG (40) | 213 | 410 | 14.1 | −0.01 (−0.07, 0.05) | 0.05 | 0.07 | 119 | 247 | 18.2 | −0.01 (−0.10, 0.07) | 0.03 | 0.03 |
| Haiti | HAITI (41) | | | | | | | | | | | | |
| Kenya | WASH−B (42) | 839 | 2778 | 3.1 | 0.01 (−0.01, 0.02) | 0.69 | 0.52 | 574 | 2174 | 5.1 | −0.01 (−0.03, 0.01) | 0.65 | 0.65 |
| Madagascar | MAHAY (43) | | | | | | | | | | | | |
| Malawi | iLiNS−DYADM (44) | 137 | 289 | 9.0 | −0.01 (−0.07, 0.05) | 0.05 | 0.07 | 78 | 149 | 14.8 | −0.02 (−0.12, 0.08) | 0.03 | 0.03 |
| Malawi | iLiNS−DOSE (45) | 377 | 135 | 19.3 | −0.03 (−0.10, 0.04) | 0.03 | 0.05 | 272 | 85 | 23.3 | −0.06 (−0.15, 0.03) | 0.03 | 0.03 |
| Mali | PROMIS (46) | | | | | | | | | | | | |
| Mali | PROMIS CS (46) | | | | | | | | | | | | |
| Zimbabwe | SHINE (HIV−) (47) | | | | | | | | | | | | |
| Zimbabwe | SHINE (HIV+) (48) | | | | | | | | | | | | |
| | | **3870** | **6058** | | **I² = 0.33, Tau² = 0.00** | | | **2275** | **4766** | | **I² = 0.00, Tau² = 0.00** | | |
| **Fixed** | | | | | **0.00 (−0.02, 0.01)** | | | | | | **−0.02 (−0.03, 0.00)** | | |
| **Random** | | | | | **−0.01 (−0.03, 0.01)** | | | | | | **−0.02 (−0.03, 0.00)** | | |

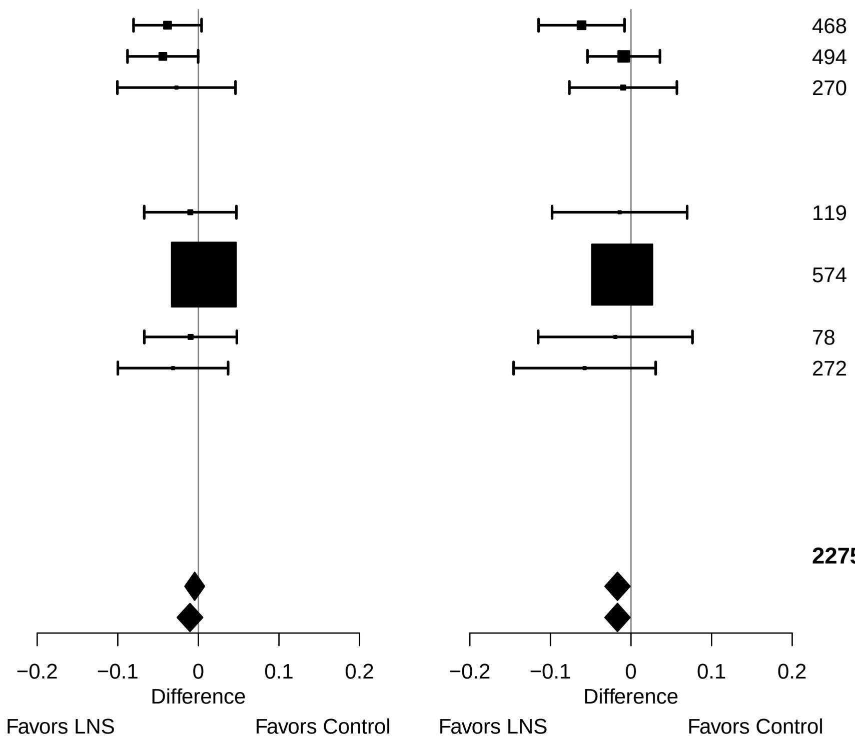

#### Supplemental figure 7Q: Small head size prevalence difference

#### 7Q6: Stratified by Season at the time of assessment

**P−for−interaction = 0.258**
**Difference in PDs = 0.01 (−0.01, 0.03)**

| Country | Trial | Dry LNS N | Dry Control N | Dry Control Prevalence | Dry PD (95% CI) | Dry Fixed W | Dry Random W | Rainy LNS N | Rainy Control N | Rainy Control Prevalence | Rainy PD (95% CI) | Rainy Fixed W | Rainy Random W |
|---|---|---|---|---|---|---|---|---|---|---|---|---|---|
| Bangladesh | JiVitA−4 (34) | 1566 | 674 | 24.6 | −0.01 (−0.05, 0.02) | 0.22 | 0.20 | 1319 | 593 | 23.5 | 0.01 (−0.03, 0.05) | 0.08 | 0.14 |
| Bangladesh | RDNS (35) | 739 | 359 | 39.6 | −0.04 (−0.09, 0.01) | 0.10 | 0.11 | 924 | 456 | 45.6 | −0.05 (−0.10, 0.00) | 0.05 | 0.09 |
| Bangladesh | WASH−B (36) | 644 | 1936 | 31.8 | −0.02 (−0.07, 0.02) | 0.14 | 0.14 | 519 | 1527 | 34.1 | −0.04 (−0.09, 0.01) | 0.06 | 0.11 |
| Burkina Faso | iLiNS−Zinc (37) | 1387 | 477 | 40.9 | −0.05 (−0.10, 0.00) | 0.09 | 0.10 | 559 | 187 | 41.3 | −0.02 (−0.11, 0.06) | 0.02 | 0.04 |
| Burkina Faso | PROMIS (38) | | | | | | | | | | | | |
| Burkina Faso | PROMIS CS (38) | | | | | | | | | | | | |
| Ghana | GHANA (39) | | | | | | | | | | | | |
| Ghana | iLiNS−DYADG (40) | 204 | 408 | 18.4 | −0.05 (−0.11, 0.01) | 0.06 | 0.07 | 143 | 284 | 12.7 | 0.03 (−0.04, 0.10) | 0.03 | 0.05 |
| Haiti | HAITI (41) | | | | | | | | | | | | |
| Kenya | WASH−B (42) | 349 | 1161 | 4.9 | 0.00 (−0.03, 0.03) | 0.27 | 0.24 | 1112 | 4015 | 4.2 | 0.00 (−0.02, 0.01) | 0.73 | 0.50 |
| Madagascar | MAHAY (43) | | | | | | | | | | | | |
| Malawi | iLiNS−DYADM (44) | 156 | 316 | 10.8 | −0.04 (−0.09, 0.02) | 0.07 | 0.08 | 64 | 128 | 10.9 | 0.05 (−0.05, 0.15) | 0.01 | 0.03 |
| Malawi | iLiNS−DOSE (45) | 418 | 133 | 25.4 | −0.09 (−0.16, −0.02) | 0.05 | 0.06 | 274 | 105 | 17.1 | 0.01 (−0.07, 0.09) | 0.02 | 0.04 |
| Mali | PROMIS (46) | | | | | | | | | | | | |
| Mali | PROMIS CS (46) | | | | | | | | | | | | |
| Zimbabwe | SHINE (HIV−) (47) | | | | | | | | | | | | |
| Zimbabwe | SHINE (HIV+) (48) | | | | | | | | | | | | |
| | | **5463** | **5464** | | **I² = 0.15, Tau² = 0.00** | | | **4914** | **7295** | | **I² = 0.10, Tau² = 0.00** | | |
| **Fixed** | | | | | **−0.02 (−0.04, −0.01)** | | | | | | **−0.01 (−0.02, 0.00)** | | |
| **Random** | | | | | **−0.03 (−0.04, −0.01)** | | | | | | **−0.01 (−0.02, 0.01)** | | |

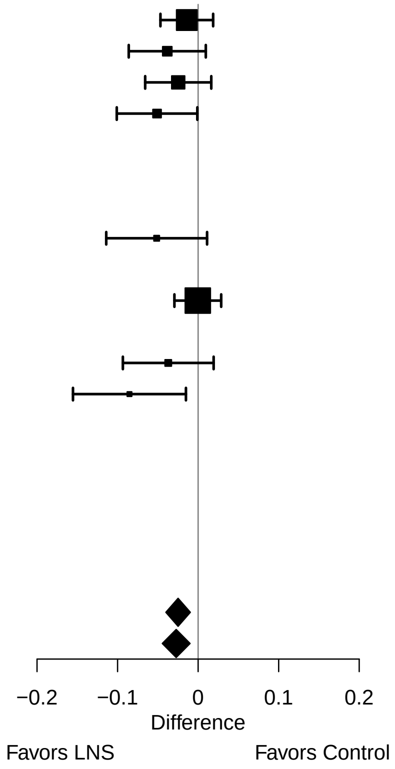

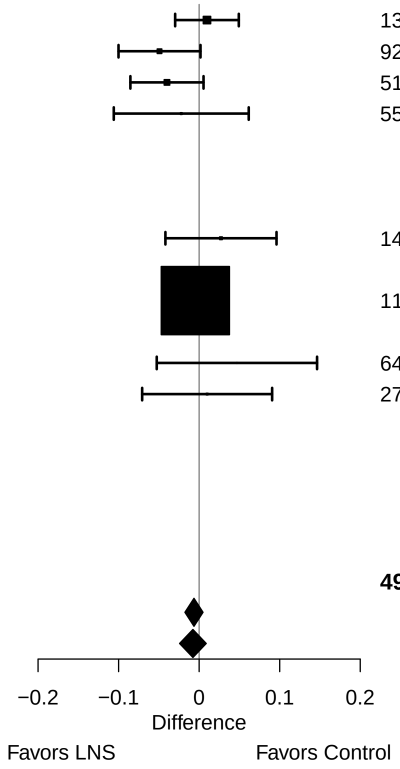
