## Supplemental Methods for "Characteristics that modify the effect of small-quantity lipid-based nutrient supplementation on child growth: an individual participant data meta-analysis of randomized controlled trials"

**Supplemental Methods for SQ-LNS IPD growth paper: Characteristics that modify the effect of small-quantity lipid-based nutrient supplementation on child growth: an individual participant data meta-analysis of randomized controlled trials**

**1. Search terms**

We used the same search terms used by Das et al. (2019) to search 16 international and 7 regional databases for studies published since that review was completed. We did not have access to WHOLIS and IndMed, so those databases were not included

**Cochrane Central Register of Controlled Trials (CENTRAL) in the Cochrane Library**

#1[mh Lipids]

#2(fatty next acid*)

#3((Docosahexaenoic or Eicosapentaenoic) next acid*)

#4(PUFA or PUFAs)

#5lipid*

#6(omega next (3* or 6*))

#7(soy* or peanut or groundnut or whey or sesame or cashew or chickpea or oil*)

#8{or #1-#7}

#9[mh “Dietary Supplements”]

#10[mh “Food, fortified”]

#11((diet* or food*) near/3 (fortif* or enrich* or supplement*))

#12(complement* near/3 (food* or feed*))

#13“Ready to use”

#14“point of use”

#15(RUSF or RUTF)

#16(home* near/2 fortif*)

#17{or #9-#16}

#18#8 and #17

#19(lipid next based)

#20(lipid* near/3 supplement*)

#21(lipid* near/3 nutrient*)

#22(lipid* near/3 fortif*)

#23(lipid* near/3 formulation*)

#24(lipid* near/3 enrich*)

#25(lipid* near/3 emuls*)

#26(lipid* near/3 powder*)

#27(lipid* near/3 spread*)

#28(lipid* near/3 paste*)

#29(Nutributter* or Plumpy*)

#30(LNS or iLiNS)

#31{or #19-#30}

#32#18 or #31

#33[mh Înfant]

#34infant* or toddler* or baby or babies or child*

#35#33 or #34

#36#32 and #35 in Trials

**MEDLINE Ovid**

1 exp Lipids/

2 fatty acid$.tw,kf.

3 Docosahexaenoic acid.tw,kf.

4 Eicosapentaenoic Acid$.tw,kf.

5 PUFA$.tw,kf.

6 lipid.tw,kf.

7 (omega 3$ or omega 6$).tw,kf.

8 (soy$ or peanut or groundnut or whey or sesame or cashew or chickpea or oil$).tw,kf.

9 or/1-8

10 Dietary Supplements/

11 Food, fortified/

12 ((diet$ or food$) adj3 (fortif$ or enrich$ or supplement$)).tw,kf.

13 (complement$ adj3 (food$ or feed$)).tw,kf.

14 “Ready to use”.tw,kf.

15 (RUSF or RUTF).tw,kf.

16 “point of use”.tw,kf.

17 (home$ adj2 fortif$).tw,kf.

18 or/10-17

19 9 and 18

20 (lipid$ adj3 nutrient$).tw,kf.

21 (lipid$ adj3 supplement$).tw,kf.

22 lipid based.tw,kf.

23 (lipid$ adj3 fortif$).tw,kf.

24 (lipid$ adj3 enrich$).tw,kf.

25 (lipid$ adj2 emuls$).tw,kf.

26 (lipid$ adj2 formulation$).tw,kf.

27 (lipid$ adj3 powder$).tw,kf.

28 (lipid adj3 spread$).tw,kf.

29 (lipid$ adj3 paste$).tw,kf.

30 (Nutributter$ or Plumpy$).tw,kf.

31 (LBNS$ or LNS$1 or iLiNS).tw,kf.

32 or/20-31

33 19 or 32

34 Infant/

35 (baby or babies or infant$ or toddler$ or child$).tw.

36 34 or 35

37 33 and 36

38 exp animals/ not humans.sh.

39 37 not 38

**MEDLINE In-Process and Other Non-Indexed Citations Ovid**

1 lipid based.tw,kf.

2 (lipid$ adj3 nutrient$).tw,kf.

3 (lipid$ adj3 supplement$).tw,kf.

4 (lipid$ adj3 fortif$).tw,kf.

5 (lipid$ adj3 enrich$).tw,kf.

6 (lipid$ adj2 emuls$).tw,kf.

7 (lipid$ adj2 formulation$).tw,kf.

8 (Lipid$ adj3 powder$).tw,kf.

9 (lipid adj3 spread$).tw,kf.

10 (lipid$ adj3 paste$).tw,kf.

11 (Nutributter$ or Plumpy$).tw,kf.

12 (LBNS$ or LNS$1 or iLiNS).tw,kf.

13 or/1-12

14 ((diet$ or food$) adj3 (fortif$ or enrich$ or supplement$)).tw,kf.

15 (complement$ adj3 (food$ or feed$)).tw,kf.

16 (RUSF or RUTF).tw,kf.

17 “point of use”.tw,kf.

18 (home$ adj2 fortif$).tw,kf.

19 lipid$.tw,kf.

20 fatty acid$.tw,kf.

21 Docosahexaenoic acid$.tw,kf.

22 Eicosapentaenoic Acid$.tw,kf.

23 PUFA$.tw,kf.

24 (omega 3$ or omega 6$).tw,kf.

25 (soy$ or peanut or groundnut or whey or sesame or cashew or chickpea or oil$).tw,kf.

26 or/14-18

27 or/19-25

28 26 and 27

29 13 or 28

30 (baby or babies or infant$ or toddler$ or child$).tw.

31 29 and 30

**MEDLINE E-pub ahead of print Ovid**

1 lipid based.tw,kf.

2 (lipid$ adj3 nutrient$).tw,kf.

3 (lipid$ adj3 supplement$).tw,kf.

4 (lipid$ adj3 fortif$).tw,kf.

5 (lipid$ adj3 enrich$).tw,kf.

6 (lipid$ adj2 emuls$).tw,kf.

7 (lipid$ adj2 formulation$).tw,kf.

8 (Lipid$ adj3 powder$).tw,kf.

9 (lipid adj3 spread$).tw,kf.

10 (lipid$ adj3 paste$).tw,kf.

11 (Nutributter$ or Plumpy$).tw,kf.

12 (LBNS$ or LNS$1 or iLiNS).tw,kf.

13 or/1-12

14 ((diet$ or food$) adj3 (fortif$ or enrich$ or supplement$)).tw,kf.

15 (complement$ adj3 (food$ or feed$)).tw,kf.

16 (RUSF or RUTF).tw,kf.

17 “point of use”.tw,kf.

18 (home$ adj2 fortif$).tw,kf.

19 lipid$.tw,kf.

20 fatty acid$.tw,kf.

21 Docosahexaenoic acid$.tw,kf.

22 Eicosapentaenoic Acid$.tw,kf.

23 PUFA$.tw,kf.

24 (omega 3$ or omega 6$).tw,kf.

25 (soy$ or peanut or groundnut or whey or sesame or cashew or chickpea or oil$).tw,kf.

26 or/14-18

27 or/19-25

28 26 and 27

29 13 or 28

30 (baby or babies or infant$ or toddler$ or child$).tw.

31 29 and 30

**Embase Ovid**

1 exp Lipids/

2 fatty acid$.tw,kw.

3 Docosahexaenoic acid.tw,kw.

4 Eicosapentaenoic Acid$.tw,kw.

5 PUFA$.tw,kw.

6 lipid$.tw,kw.

7 (omega 3$ or omega 6$).tw,kw.

8 (soy$ or peanut or groundnut or whey or sesame or cashew or chickpea or oil$).tw,kw.

9 or/1-8

10 dietary supplement/

11 fortified food/

12 ((diet$ or food$) adj3 (fortif$ or enrich$ or supplement$)).tw,kw.

13 (complement$ adj3 (food$ or feed$)).tw,kw.

14 “Ready to use”.tw,kw.

15 (RUSF or RUTF).tw,kw.

16 “point of use”.tw,kw.

17 (home$ adj2 fortif$).tw,kw.

18 or/10-17

19 9 and 18

20 lipid based.tw,kw.

21 (lipid$ adj3 nutrient$).tw,kw.

22 (lipid$ adj3 supplement$).tw,kw.

23 (lipid$ adj3 fortif$).tw,kw.

24 (lipid$ adj3 enrich$).tw,kw.

25 (lipid$ adj2 emuls$).tw,kw.

26 (lipid$ adj2 formulation$).tw,kw.

27 (Lipid$ adj3 powder$).tw,kw.

28 (lipid adj3 spread$).tw,kw.

29 (lipid$ adj3 paste$).tw,kw.

30 (Nutributter$ or Plumpy$).tw,kw.

31 (LBNS$ or LNS$1 or iLiNS).tw,kw.

32 or/20-31

33 19 or 32

34 infant/

35 (baby or babies or infant$ or toddler$ or child$).tw.

36 34 or 35

37 33 and 36

38 exp animals/ or exp invertebrate/ or animal experiment/ or animal model/ or animal tissue/ or animal cell/ or nonhuman/

39 human/ or normal human/ or human cell/

40 38 not 39

41 37 not 40

**CINAHL Plus EBSCOhost (Cumulative Index to Nursing and Allied Health Literature)**

S1(MH “Lipids+”)

S2TI (lipid*) or AB (lipid*)

S3TI(Docosahexaenoic acid*) OR AB(Docosahexaenoic acid*)

S4TI( Eicosapentaenoic acid*) OR AB( Eicosapentaenoic acid*)

S5TI(PUFA*) OR AB(PUFA* )

S6TI(omega 3* or omega 6*) OR AB(omega 3* or omega 6* )

S7TI (soy* or peanut or groundnut or whey or sesame or cashew or chickpea or oil*) or AB(soy* or peanut or groundnut or whey or

sesame or cashew or chickpea or oil*)

S8TI(fatty acid*) OR AB(fatty acid* )

S9S1 OR S2 OR S3 OR S4 OR S5 OR S6 OR S7 OR S8

S10(MH “Dietary Supplements”)

S11(MH “Dietary Supplementation”)

S12(MH “Food, Fortified”)

S13TI ((diet* or food*) n3 (fortif* or enrich* or supplement*)) OR AB((diet* or food*) n3 (fortif* or enrich* or supplement*))

S14TI (complement* N3 (food* or feed*)) or AB (complement* N3 (food* or feed*))

S15“Ready to use”

S16(RUSF or RUTF)

S17“point of use”

S18TI (home* N2 fortif*) OR AB(home* N2 fortif*)

S19S10 OR S11 OR S12 OR S13 OR S14 OR S15 OR S16 OR S17 OR S18

S20S9 AND S19

S21TI (lipid based) or AB (lipid based)

S22TI(lipid* N3 supplement*) OR AB( lipid* N3 supplement*)

S23TI(lipid* N3 nutrient*) OR AB(lipid* N3 nutrient*)

S24TI(lipid* N3 fortif*) OR AB (lipid* N3 fortif*)

S25TI(lipid* N3 formulation*) OR AB(lipid* N3 formulation*)

S26TI(lipid* N3 enrich*) OR AB(lipid* N3 enrich* )

S27TI(lipid* N3 emuls*) OR AB(lipid* N3 emuls*)

S28TI(lipid* N3 powder*) OR AB(lipid* N3 powder*)

S29TI(lipid N3 spread*) OR AB(lipid N3 spread*)

S30TI(lipid* N3 paste*) OR AB(lipid* N3 paste*)

S31Nutributter*

S32 Plumpy*

S33TI(LNS*1 or iLiNS) OR AB( LNS*1 or iLiNS)

S34S21 OR S22 OR S23 OR S24 OR S25 OR S26 OR S27 OR S28 OR S29 OR S30 OR S31 OR S32 OR S33

S35S20 OR S34

S36(MH “Infant”)

S37TI(baby or babies or infant* or toddler* or child*) OR AB (baby or babies or infant* or toddler* or child*)

S38S36 OR S37

S39S35 AND S38

**Science Citation Index (SCI) and Social Sciences Citation Index (SSCI) Web of Science**

#5 #4 AND #3

#4 TS=(infant* OR child* OR toddler* or baby or babies)

#3 #2 OR #1

#2 TS=(Nutributter* OR Plumpy* OR LNS OR iLiNS OR “lipid based” )

#1 TS=(( “fatty acid*” OR PUFA OR PUFAs OR “omega 3*” OR “omega 6*” OR soy* OR peanut* OR groundnut* OR whey* OR

sesame* OR cashew* OR chickpea* OR oil* ) Near/3 ( FORTIF* OR ENRICH OR SUPPLEMENT* OR “READY TO USE” OR

“POINT OF USE” OR RUSF OR RUTF OR PASTE* OR SPREAD* OR FORMULAT* OR EMULS* OR NUTRIENT* OR

POWDER*))

**Conference Proceedings Citation Index - Science (CPCI-S) and Conference Proceedings Citation Index -**

**Social Science & Humanities (CPCI-SS&H) Web of Science**

#5 #4 AND #3

DocType=All document types; Language=All languages;

#4 TS=(infant* OR child* OR toddler* or baby or babies)

DocType=All document types; Language=All languages;

#3 #2 OR #1

DocType=All document types; Language=All languages;

#2 TS=(Nutributter* OR Plumpy* OR LNS OR iLiNS OR “lipid based” )

DocType=All document types; Language=All languages;

#1 TS=(( “fatty acid*” OR PUFA OR PUFAs OR “omega 3*” OR “omega 6*” OR soy* OR peanut* OR groundnut* OR whey* OR

sesame* OR cashew* OR chickpea* OR oil* ) Near/3 ( FORTIF* OR ENRICH OR SUPPLEMENT* OR “READY TO USE” OR

“POINT OF USE” OR RUSF OR RUTF OR PASTE* OR SPREAD* OR FORMULAT* OR EMULS* OR NUTRIENT* OR

POWDER*))

DocType=All document types; Language=All languages;

**Cochrane Database of Systematic Reviews (CDSR), part of the Cochrane Library**

#1[mh Lipids]

#3((Docosahexaenoic or Eicosapentaenoic) next acid*):ti,ab

#4(PUFA or PUFAs):ti,ab

#5lipid*:ti,ab 23529

#6(omega next (3* or 6*)):ti,ab

#7(soy* or peanut or groundnut or whey or sesame or cashew or chickpea or oil*):ti,ab

#8{or #1-#7}

#9[mh “Dietary Supplements”]

#10[mh “Food, fortified”]

#11((diet* or food*) near/3 (fortif* or enrich* or supplement*)):ti,ab

#12(complement* near/3 (food* or feed*)):ti,ab

#13“Ready to use”:ti,ab

#14“point of use”:ti,ab

#15(RUSF or RUTF):ti,ab

#16(home* near/2 fortif*):ti,ab

#17{or #9-#16}

#18#8 and #17

#19(lipid next based):ti,ab

#20(lipid* near/3 supplement*):ti,ab

#21(lipid* near/3 nutrient*):ti,ab

#22(lipid* near/3 fortif*):ti,ab

#23(lipid* near/3 formulation*):ti,ab

#24(lipid* near/3 enrich*):ti,ab

#25(lipid* near/3 emuls*):ti,ab

#26(lipid* near/3 powder*):ti,ab

#27(lipid* near/3 spread*):ti,ab

#28(lipid* near/3 paste*):ti,ab

#29(Nutributter* or Plumpy*):ti,ab

#30(LNS*1 or iLiNS):ti,ab

#31{or #19-#30}

#32#18 or #31

#33[mh Înfant]

#34infant* or toddler* or baby or babies or child*:ti,ab

#35#33 or #34

#36#32 and #35 in Cochrane Reviews (Reviews and Protocols)

**Database of Abstracts of Reviews of Effect (DARE), part of the Cochrane Library**

#1[mh Lipids]

#3((Docosahexaenoic or Eicosapentaenoic) next acid*):ti,ab

#4(PUFA or PUFAs):ti,ab

#5lipid*:ti,ab 23529

#6(omega next (3* or 6*)):ti,ab

#7(soy* or peanut or groundnut or whey or sesame or cashew or chickpea or oil*):ti,ab

#8{or #1-#7}

#9[mh “Dietary Supplements”]

#10[mh “Food, fortified”]

#11((diet* or food*) near/3 (fortif* or enrich* or supplement*)):ti,ab

#12(complement* near/3 (food* or feed*)):ti,ab

#13“Ready to use”:ti,ab

#14“point of use”:ti,ab

#15(RUSF or RUTF):ti,ab

#16(home* near/2 fortif*):ti,ab

#17{or #9-#16}

#18#8 and #17

#19(lipid next based):ti,ab

#20(lipid* near/3 supplement*):ti,ab

#21(lipid* near/3 nutrient*):ti,ab

#22(lipid* near/3 fortif*):ti,ab

#23(lipid* near/3 formulation*):ti,ab

#24(lipid* near/3 enrich*):ti,ab

#25(lipid* near/3 emuls*):ti,ab

#26(lipid* near/3 powder*):ti,ab

#27(lipid* near/3 spread*):ti,ab

#28(lipid* near/3 paste*):ti,ab

#29(Nutributter* or Plumpy*):ti,ab

#30(LNS*1 or iLiNS):ti,ab

#31{or #19-#30}

#32#18 or #31

#33[mh Înfant]

#34infant* or toddler* or baby or babies or child*:ti,ab

#35#33 or #34

#36#32 and #35 in Other Reviews

**Epistemonikos (epistemonikos.org)**

(title:(LIPID* OR FATTY ACID* OR OMEGAOR Docosahexaenoic OR Eicosapentaenoic OR soy* OR peanut OR groundnut OR

whey OR sesame OR cashew OR chickpea OR oil*) OR abstract:(LIPID* OR FATTY ACID* OR OMEGA OR Docosahexaenoic

OR Eicosapentaenoic OR soy* OR peanut OR groundnut OR whey OR sesame OR cashew OR chickpea OR oil*)) AND (title:

(fortif* OR enrich* OR supplement* OR “Ready to use” OR “point of use” OR RUSF OR RUTF) OR abstract:(fortif* OR enrich*

OR supplement* OR “Ready to use” OR “point of use” OR RUSF OR RUTF)) AND title:(babies OR children OR infant*)

LIMITED TO

PUBLICATION TYPE: SYSTEMATIC REVIEW

SYSTEMATIC REVIEW QUESTION : INTERVENTIONS

**POPLINE (www.popline.org)**

((ALL FIELDS(lipid* OR “fatty acid*” OR PUFA OR PUFAs OR “omega 3*” OR “omega 6*” OR soy* OR peanut* OR groundnut*

OR whey* OR sesame* OR cashew* OR chickpea* OR oil*) AND ALL FIELDS (FORTIF* OR ENRICH OR SUPPLEMENT* OR

“READY TO USE” OR “POINT OF USE” OR RUSF OR RUTF OR PASTE* OR SPREAD* OR FORMULAT* OR EMULS*

OR NUTRIENT* OR POWDER*)) OR (Nutributter* OR Plumpy* OR LNS OR iLiNS)) AND ALL FIELDS (infan* OR child*

OR baby OR babies OR toddler*)

**ClinicalTrials.gov (clinicaltrials.gov)**

Interventional Studies | LIPID-BASED OR LNS OR iLiNS OR “NUTRIENT SUPPLEMENT” OR PASTE OR SPREAD OR

BLEND OR NUTRIBUTTER OR Plumpy OR PLUMPYNUT | Child

**World Health Organization International Clinical Trials Registry Platform (WHO ICTRP; who.int/trialsearch)**

(LIPID BASED OR LNS OR iLiNS OR NUTRIENT SUPPLEMENT OR SPREAD OR paste OR BLEND OR NUTRIBUTTER

OR Plumpy OR PLUMPYNUT) NOT (teeth OR oral heath OR dentistry)

**IBECS (Índice Bibliográfico Español en Ciencias de la Salud; ibecs.isciii.es)**

WORD| “lipid based” OR LNS OR iLiNS OR “nutrient supplement” OR paste OR spread OR blend OR nutributter OR Plumpy

OR plumpynut

AND

WORD| infan* OR child* OR baby OR babies OR toddler*

**SciELO (Scientific Electronic Library Online; www.scielo.br)**

“lipid based” OR LNS OR iLiNS OR “nutrient supplement” OR paste OR spread OR blend OR nutributter OR Plumpy OR

plumpynut [All indexes]

AND

infan* OR child* OR baby OR babies OR toddler* [All indexes]

**AIM (Africa Index Medicus; search.bvsalud.org/ghl/?lang=en&submit=Search&where=REGIONAL)**

(lipid basedOR LNSOR iLiNS OR nutrient supplementOR paste OR spreadOR blendOR nutributter OR PlumpyOR plumpynut)

AND (infan OR child OR baby OR babies OR toddler)

**IMEMR (Index Medicus for the Eastern Mediterranean Region;**

**search.bvsalud.org/ghl/?lang=en&submit=Search&where=REGIONAL)**

(lipid basedOR LNSOR iLiNS OR nutrient supplementOR paste OR spreadOR blendOR nutributter OR PlumpyOR plumpynut)

[Title]

AND

(infan OR child OR baby OR babies OR toddler) [Title]

**LILACS (Latin American and Caribbean Health Sciences Literature; lilacs.bvsalud.org/en)**

(tw:(“lipid based” OR LNS OR iLiNS OR “nutrient supplement” OR paste OR spread OR blend OR nutributter OR Plumpy OR

plumpynut) AND (tw:(infan* OR child* OR baby OR babies OR toddler*)

**PAHO/WHO Institutional Repository for Information Sharing (iris.paho.org/xmlui)**

(“lipid based” OR LNS OR iLiNS OR “nutrient supplement” OR paste OR spread OR blend OR nutributter OR Plumpy OR

plumpynut) AND (infan* OR child* OR baby OR babies OR toddler*)

**WPRIM (Western Pacific Index Medicus;**

**search.bvsalud.org/ghl/?lang=en&submit=Search&where=REGIONAL)**

(“lipid based” OR LNS OR iLiNS OR “nutrient supplement” OR paste OR spread OR blend OR nutributter OR Plumpy OR

plumpynut) AND (infan* OR child* OR baby OR babies OR toddler*)

**IMSEAR (Index Medicus for the South-East Asian Region;**

**search.bvsalud.org/ghl/?lang=en&submit=Search&where=REGIONAL)**

(“lipid based” OR LNS OR iLiNS OR “nutrient supplement” OR paste OR spread OR blend OR nutributter OR Plumpy OR

plumpynut) [Title] AND (infan* OR child* OR baby OR babies OR toddler*) [Title]

**Native Health Research Database (hscssl.unm.edu/nhd)**

Keywords: (Supplement AND child)

**2. Methods to determine rainy vs. dry season**

For each study we identified the GPS coordinates of an approximately central location within the study area. Using this location we retrieved the historical monthly rainfall data for that area from two online repositories. For rainfall information prior to 2017, data were extracted from the Global Precipitation Climatology Center at 0.5° resolution^1^. Information for more recent years was extracted from the Climate Hazards Group Infrared Precipitation with Stations^2^.

We computed average rainfall for 3-month time periods, defined as the “index” month (when the child assessment was conducted) and the two months prior to the index month. Then a k-means clustering analysis with k=2 groups was conducted to categorize “index” months into time periods with more precipitation vs time periods with less precipitation. These categorizations were then applied to the child-level month of assessment to determine a ‘Rainy’ vs ‘Dry’ season designation for each assessment of a given child.

Plots of average precipitation over time with the k-means categorizations were also visually assessed. The two studies conducted in Mali and Haiti were found to have little variation in precipitation and so were not assigned seasonality values.

**Acknowledgements**

Our approach was motivated by discussions and the preliminary work on related topics^3^ of Ben Arnold, Andrew Mertens, and Jack Colford.

**Sources**

1. Schneider, Udo; Becker, Andreas; Finger, Peter; Meyer-Christoffer, Anja; Ziese, Markus (2018): GPCC Full Data Monthly Product Version 2018 at 0.5°: Monthly Land-Surface Precipitation from Rain-Gauges built on GTS-based and Historical Data. DOI: 10.5676/DWD_GPCC/FD_M_V2018_050 (<https://opendata.dwd.de/climate_environment/GPCC/html/fulldata-monthly_v2018_doi_download.html>)

2. Funk, C.C., Peterson, P.J., Landsfeld, M.F., Pedreros, D.H., Verdin, J.P., Rowland, J.D., Romero, B.E., Husak, G.J., Michaelsen, J.C., and Verdin, A.P., 2014, A quasi-global precipitation time series for drought monitoring: U.S. Geological Survey Data Series 832, 4 p., <https://dx.doi.org/10.3133/ds832>. ISSN 2327-638X (online) (<ftp://ftp.chg.ucsb.edu/pub/org/chg/products/CHIRPS-2.0/africa_monthly/bils/>)

3. Mertens, Andrew, Jade Benjamin-Chung, John M. Colford, Alan E. Hubbard, Mark van der Laan, Jeremy Coyle, Oleg Sofrygin et al. "Child wasting and concurrent stunting in low-and middle-income countries." *medRxiv* (2020).
